# Declines and pronounced state-level variation in tapentadol use in the US

**DOI:** 10.1101/2022.03.03.22271869

**Authors:** Ching Y. Low, Kenneth L. McCall, Brian J. Piper

## Abstract

**Background:** Tapentadol is an opioid approved for the treatment of moderate-to-severe pain in the United States (US). Tapentadol is unique as it is the only Schedule II prescription drug that has dual modes of action as it combines agonist activity at the µ opioid receptor with norepinephrine reuptake inhibition. This descriptive study characterized tapentadol use in the US.

**Methods:** Drug distribution data from 2010 to 2020 were extracted for each state from the Drug Enforcement Administration. Use per state, corrected for population, was analyzed. The percentage of distribution channels (pharmacies, hospitals, and providers), the distributed amount of tapentadol, and the final adjusted quota of tapentadol were obtained. Data on tapentadol use as reported by the Medicare and Medicaid programs for 2010 to 2020 were also analyzed.

**Results:** The distributed amount of tapentadol was 3.5 tons in 2020 and on average, the final adjusted production quota was 207.2% greater than the distributed amount between 2010 and 2020. Distributed tapentadol was 1.3% of all Schedule II opioids distributed in 2020. Tapentadol use decreased by −53.8% between 2012 and 2020 in the US whereas New Hampshire was the only state that had a positive change (+13.1%). There were minor changes in the amounts of tapentadol distributed via various distribution channels (Pharmacies = 98.0%, hospitals = 1.9% in 2020). Tapentadol prescribed by Nurse Practitioners experienced the largest increase of +8.7% among all specialties to 18.0%, the highest percentage of Medicare claims of tapentadol in 2019. Diabetes prevalence was significantly correlated with tapentadol distribution in 2012 (*r*(50) = .44, *p* < .01) and 2020 (*r*(50) = .28, *p* < .05).

**Discussion:** There has been a substantial decline over the past decade in tapentadol distribution and prescribing to Medicaid patients. The unusual tapentadol prescribing pattern in New Hampshire may warrant investigation regarding differing prescribers’ attitudes towards tapentadol or the employment of tapentadol as part of a step-down therapy for opioid addiction.

## 1. Introduction

The largely iatrogenic US opioid epidemic has resulted in over one-million overdoses.^1^ There is a need for other pharmacotherapies to treat acute and chronic pain which are less likely to be misused and diverted. Tapentadol is a synthetic, centrally acting analgesic that combines agonist activity at the µ opioid receptor (MOR) with norepinephrine reuptake inhibition (NRI). Through the MOR mechanism, analgesic effects are proposed to occur through Gi/o-protein-dependent signaling and this interrupts the pre- and postsynaptic ascending pain signals in the central nervous system.^2,3^ In addition, the NRI mechanism raises norepinephrine levels within the synapse and hence increases pain inhibition in the descending pathway.^3^ NRI mechanisms are effective in treating chronic neuropathic pain.^4^ The synergy of these two mechanisms relieves both nociceptive and neuropathic chronic pain on a potentially lower dose, and thereby offers the possible advantage of mitigating the adverse effects of opioid use.^3^ Further, one investigation reported that tapentadol has about equal affinity for the human norepinephrine and serotonin transporters.^5^

Tapentadol immediate release (IR) was approved in 2008 for acute pain severe enough to warrant an opioid analgesic whereas tapentadol extended release (ER) was approved in 2013 for around-the-clock, long-term treatment for severe chronic pain and severe pain associated with diabetic peripheral neuropathy in the US. The US Drug Enforcement Administration (DEA) classifies tapentadol as a Schedule II controlled substance due to its risk of inducing opioid addiction, diversion, and misuse.^6^ Due to the widely recognized public health issue of the misuse of prescription opioids in the US, current strategies to mitigate the opioid crisis and manage chronic pain include focusing on opioids with the potential to treat pain effectively and the possibility of mitigating abuse and addiction.^7^ Tapentadol’s therapeutic potency is comparable to morphine across a variety of preclinical pain models,^8^ and tapentadol is the only Schedule II prescription opioid that has dual MOR and monoamine transporter^5^ mechanisms. Furthermore, the diversion rate of tapentadol ER (0.001 per 100,000 population) was significantly lower than that of other Scheduled II Controlled Substances (1.495 per 100,000 population).^9^ Tapentadol ER costed less than other Schedule II opioid products when sold illicitly.^9^

The purpose of this study was to investigate the trends of tapentadol prescribed adjusted for population 18 and above in the US over the period 2010 through 2020. Three complimentary datasets including the Automation of Reports and Consolidated Orders Systems (ARCOS)^10,11^ published by the DEA, Medicaid, and Medicare Part D^12^ programs were studied to achieve a more complete picture of the prescribing trends of tapentadol. Included in this study was a comparison of the distribution pattern of tapentadol against other Schedule II Controlled Substances in the US over the same period for a more meaningful understanding of tapentadol’s distribution trend against the backdrop of additional opioid prescribing regulations in the US.^13^ Examination of the correlation between the adult prevalence of diabetes^14^ and tapentadol distribution was also determined.

## 2. Methods

### 2.1 Procedures

Tapentadol and other Schedule II controlled substances data were extracted from DEA’s ARCOS, a comprehensive drug reporting database containing an annually updated report of the distribution of DEA controlled substances from manufacturers and distributors. ARCOS has been used in prior pharmacoepidemiology studies and showed a very high (*r* = 0.985) correlation with a state prescription drug monitoring program for oxycodone.^15^ We extracted prescribed tapentadol (grams) by State, Quarter and Business Activity from published reports available on the ARCOS website^10,11^ to compute tapentadol (mg/person aged 18 and above) distributed in the states and in each quarter, and by retail distribution via hospitals, pharmacies, practitioners, and teaching institutions which were collectively grouped under business activity. Schedule II Controlled Substances by state and year were extracted from ARCOS reports computing Schedule II Controlled Substances (mg/person) distributed in each state and year. Annual Final Adjusted Quota (gram) of tapentadol was extracted from the DEA^10,11^. Since the beginning of the Medicaid Drug Rebate Program (MDRP), data for covered outpatient drugs paid by State Medicaid agencies have been reported by various states to the Centers for Medicare and Medicaid Services (CMS) and captured in the State Drug Utilization Data (SDUD) database^16^. We extracted Medicaid claims for tapentadol including the corresponding 11-digit National Drug Code, Units Reimbursed, and State for every tapentadol claim in each quarter. We sourced the milligrams per tablet by the NDC codes from the National Drug Code Directory^17^ to match the milligrams per tablet to each Medicaid entry retrieved and thereafter computed the tapentadol (grams) reimbursed per entry. Total distributed tapentadol in grams using ARCOS and Medicaid data were converted to metric tons as necessary. Prescriber State, Provider Specialty Type, and Aggregate Cost Paid for All Claims were extracted for tapentadol from Medicare Part D Prescriber Data from the Centers for Medicare and Medicaid Services from 2013 through 2019^10^, the most recent year available, to compute Medicare claims on tapentadol aggregated by prescribers’ specialty and the state the prescribers were located in. Tapentadol is legally allowed to be prescribed to users aged 18 and above and we found no off-label prescription orders of tapentadol to anyone younger than 18 at Geisinger’s health system between 2009 and 2020. Therefore, we calculated tapentadol (mg/person aged 18 and above) using estimates of people aged 18 and above (excluding the Virgin Islands and Puerto Rico) from 2010 to 2020 extracted from the US Census Bureau, Population Division.^18^ Mg/person for all opioids in Supplemental Figures 3 and 4 were calculated using the total population, including the Virgin Islands and Puerto Rico, from 2010 to 2020 extracted from the US Census Bureau, Population Division.^18^ We assumed the population estimates in each state were consistent across every quarter within the year. The morphine milligrams equivalent (MME) for each Scheduled II opioid was determined. Conversions^15^ were completed with the following multipliers: codeine 0.15, fentanyl 75, hydrocodone 1, hydromorphone 4, meperidine 0.1, morphine 1, oxycodone 1.5, oxymorphone 3, and tapentadol 0.4. The diabetes prevalence was retrieved to compute adult diabetes diagnosed per 100 people in each state.^14^ Institutional Review Board approval was obtained from Geisinger and the University of New England.

### 2.2. Statistics

The programs GraphPad Prism, and Microsoft Excel were used to graph and analyze the data. We identified the peak year of tapentadol distribution to be 2012 and expressed ARCOS data (mg/person aged 18 and above) as a percent change relative to this peak. The results, excluding the Virgin Islands and Puerto Rico, were presented on a heat map prepared with http://www.heatmapper.ca/geomap^19^. Percentage change values were deemed significant as ≥1.96 standard deviations above and below the mean, accounting for values outside a 95% confidence interval. The amount of tapentadol (grams) prescribed by retail distributors including hospitals, pharmacies, practitioners, and teaching institutions was presented as percentages for each year. An unpaired t-test between distributed tapentadol (grams) and the final adjusted quota was performed. Regional analysis was conducted by dividing ARCOS aggregate data per US region (Midwest, Northwest, South, West), and divisional analysis was conducted among divisions within Census regions (New England, Middle Atlantic, East North Central, West North Central, South Atlantic, East South Central, West South Central, Mountain, Pacific). A *p* < 0.05 was considered statistically significant for all analyses. The distribution trend of tapentadol (mg/person) and that of another Scheduled II Controlled Substance (mg/person) was graphed for visual comparison at the national level and for NH. Analysis of correlation between tapentadol (grams) using ARCOS and Medicaid data was completed. Scatterplot and prescribing trends using these two data sources were plotted to aid in visual comparison. Aggregate tapentadol claims grouped by prescriber specialty and state using Medicare data were presented as percentages of a whole for each year. Adult diabetes diagnosed per 100 persons was correlated with tapentadol (mg/person aged 18 and above) from ARCOS for every state (except the Virgin Islands and Puerto Rico) at the national level, and individually at every state.

## 3. Results

DEA’s ARCOS data was used to generate Figures 1 and 2. Between 2012, the peak year, and 2020, tapentadol saw a 53.8% reduction in mg/person in the US. In the same period, other Scheduled II Controlled Substances also experienced reductions in mg/person with meperidine experiencing the most reduction (−91.8%), and codeine the least reduction (−32.1%, Figure 1A). On average, the final adjusted quota from ARCOS was 207.2% greater than the distributed amount of tapentadol and the difference was statistically significant (Figure 1B). A regional analysis identified a significant difference between the Northeast and West regions (Figure 1C). Distribution of Schedule II Controlled Substances in MME expressed in percentages determined that oxycodone was nearly half of 2020’s distribution while tapentadol was only 1.3%. The percentage distributions of opioids were relatively constant in 2012 and 2020 (Figure 1D). The amount of tapentadol distributed by various business activities showed little variation between 2010 and 2020. Pharmacies remained the largest distribution channel for tapentadol (98.0% in 2010 and 98.1% in 2020). Hospitals inched upwards from 1.1% to 1.9% of tapentadol distributed in 2020, and this gain was achieved from practitioners having lost nearly 1% of tapentadol distributed over the same period (Figure IE).

**Figure 1.**
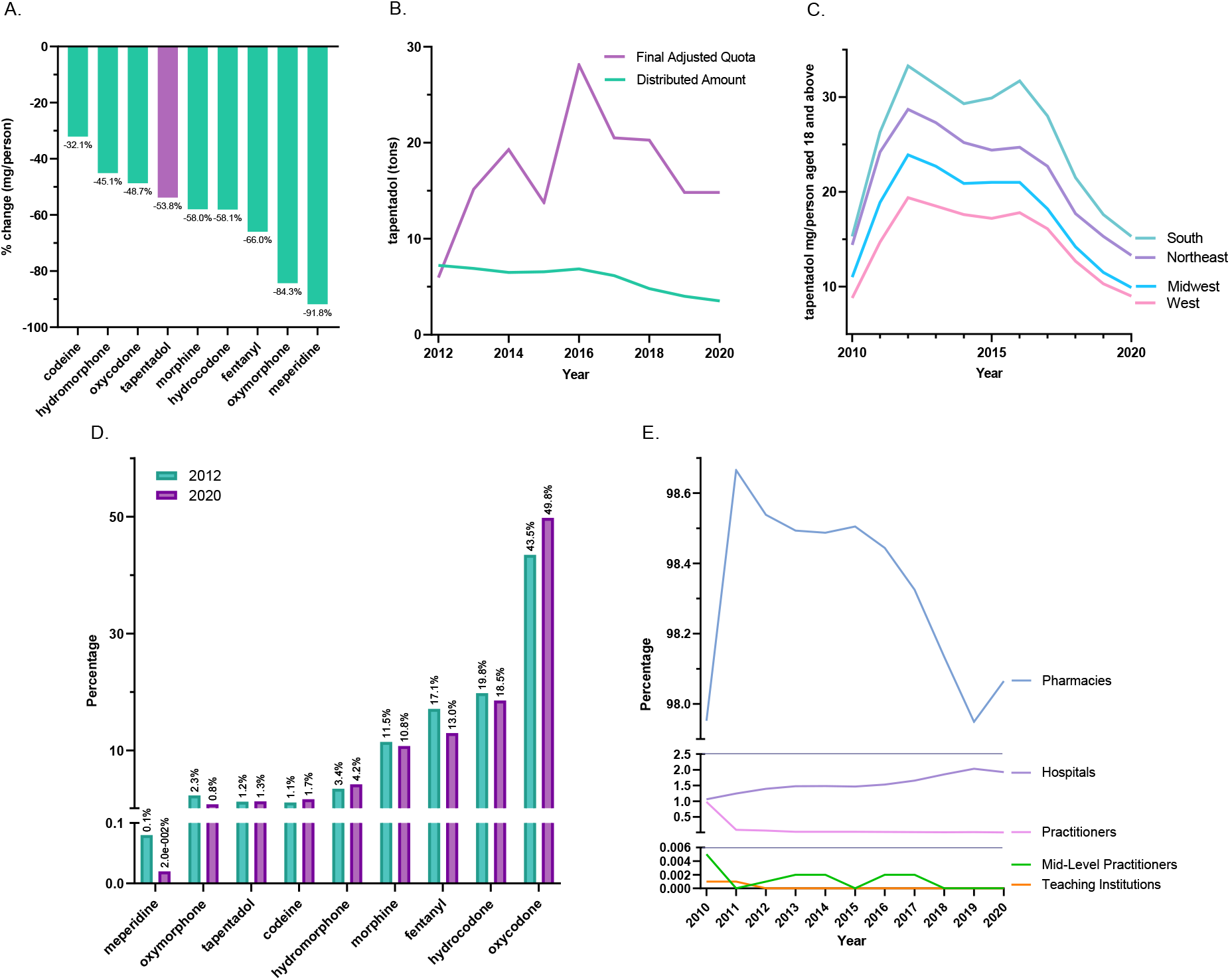
Controlled substance data for tapentadol and other opioids from the Drug Enforcement Administration’s Automated Reports and Consolidated Ordering System. **A**. All Schedule II Controlled Substances distribution declined, and distribution of tapentadol decreased by 53.8% for 2012-2020. **B**. Unpaired t-test between Distributed and Final Adjusted Quota amounts of tapentadol was statistically significant (*p* < 0.05) for 2012-2020. **C**. Tapentadol (mg/person aged 18 and above) in Northeast and West regions were statistically different (*p* < 0.05) for 2010-2020. **D**. Percentages of distributed Scheduled II Controlled Substances in morphine mg equivalents (MME) for 2012 and 2020. Tapentadol was 1.3% of total Scheduled II Controlled Substances in 2020. **E**. Amount of tapentadol (grams) distributed by retail distributors in the US presented as percentages of the total each year for 2010-2020.

**Figure 2.**
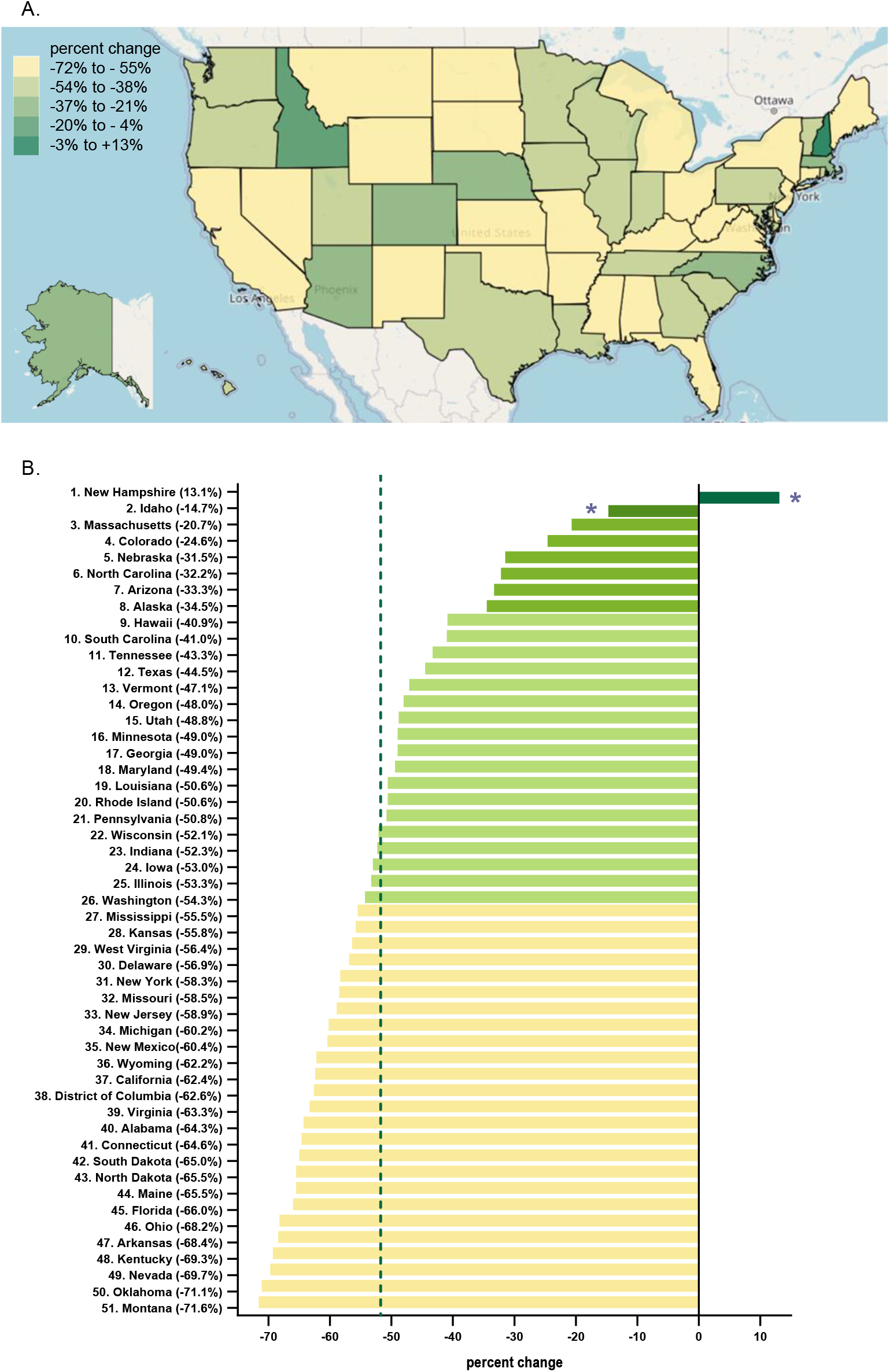
Data from the Drug Enforcement Administration’s Automated Reports and Consolidated Ordering System. **A**. Heatmap of percentage change in tapentadol (mg/person aged 18 and above) between 2012 and 2020. Only NH experienced a positive change (+13.1%). **B**. Percentage change in tapentadol (mg/person aged 18 and above) between 2012 and 2020. The average percentage change in tapentadol milligrams per person aged 18 and above was −51.7% and SD was 16.1%, designated with a vertical dashed line. States with a percentage change that was significantly different than the national average (* p < 0.05).

Further analysis of regional divisional analysis between New England and other regional divisions indicated the data between New England and West North Central as well as the Pacific were nonsignificant, whereas the data between New England and all other divisions were statistically different (Supplemental Figure 1B).

Every state in the US saw a reduction in tapentadol (mg/aged 18 and above) between 2012 and 2020, except for NH which saw a +13.1% increase (Figure 2A). Percentage change values of Idaho (−14.7%) and NH were deemed significant as they were outside the 95% confidence interval (Figure 2B).

For the US, a correlation analysis on ARCOS with Medicaid showed a strong, positive and significant correlation of 0.80 (Figure 3A). Tapentadol claims from Medicaid mirrored that of tapentadol distribution reported by ARCOS. Medicaid peaked in the second quarter of 2011, and there was a 55.2% decrease from the peak quarter until the end of 2020 (Figure 3B).

**Figure 3.**
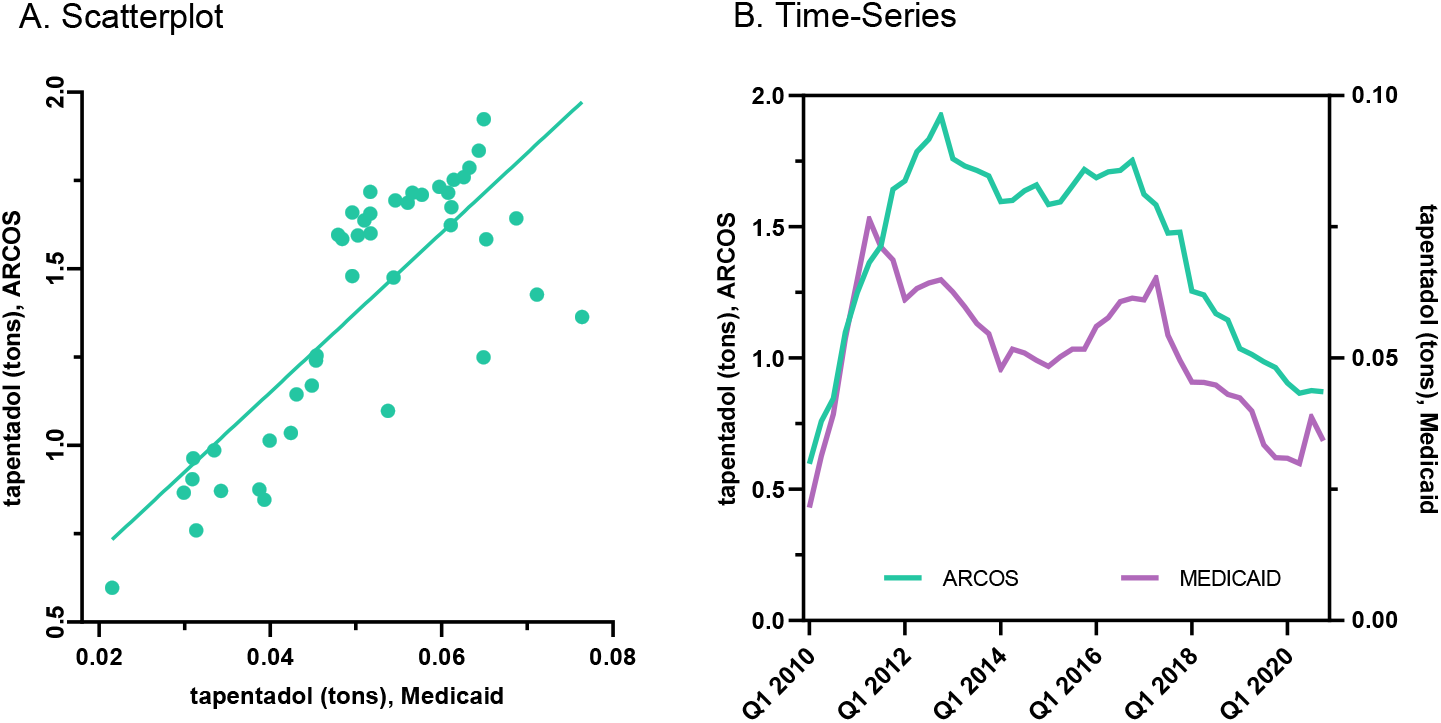
**A**. Scatterplot of prescribed tapentadol (tons) from ARCOS and tapentadol (tons) claims from Medicaid. Correlation 0.80 was statistically significant (p < 0.05). **B**. Quarterly time-series of prescribed tapentadol (tons) from ARCOS and tapentadol (tons) claims from Medicaid.

Amount of tapentadol claims in the US using data from Medicare Part D presented as percentages each year by specialties indicated Nurse Practitioner experienced a strong and steady increase in the share of tapentadol’s prescription and became the specialty with the largest share in 2019. NPs accounted for 9.3% in 2013 and 18.0% in 2019. Physician Assistants experienced a steady but more moderate increase from 9.3% in 2013 to 14.5% in 2019 (Figure 4).

**Figure 4.**
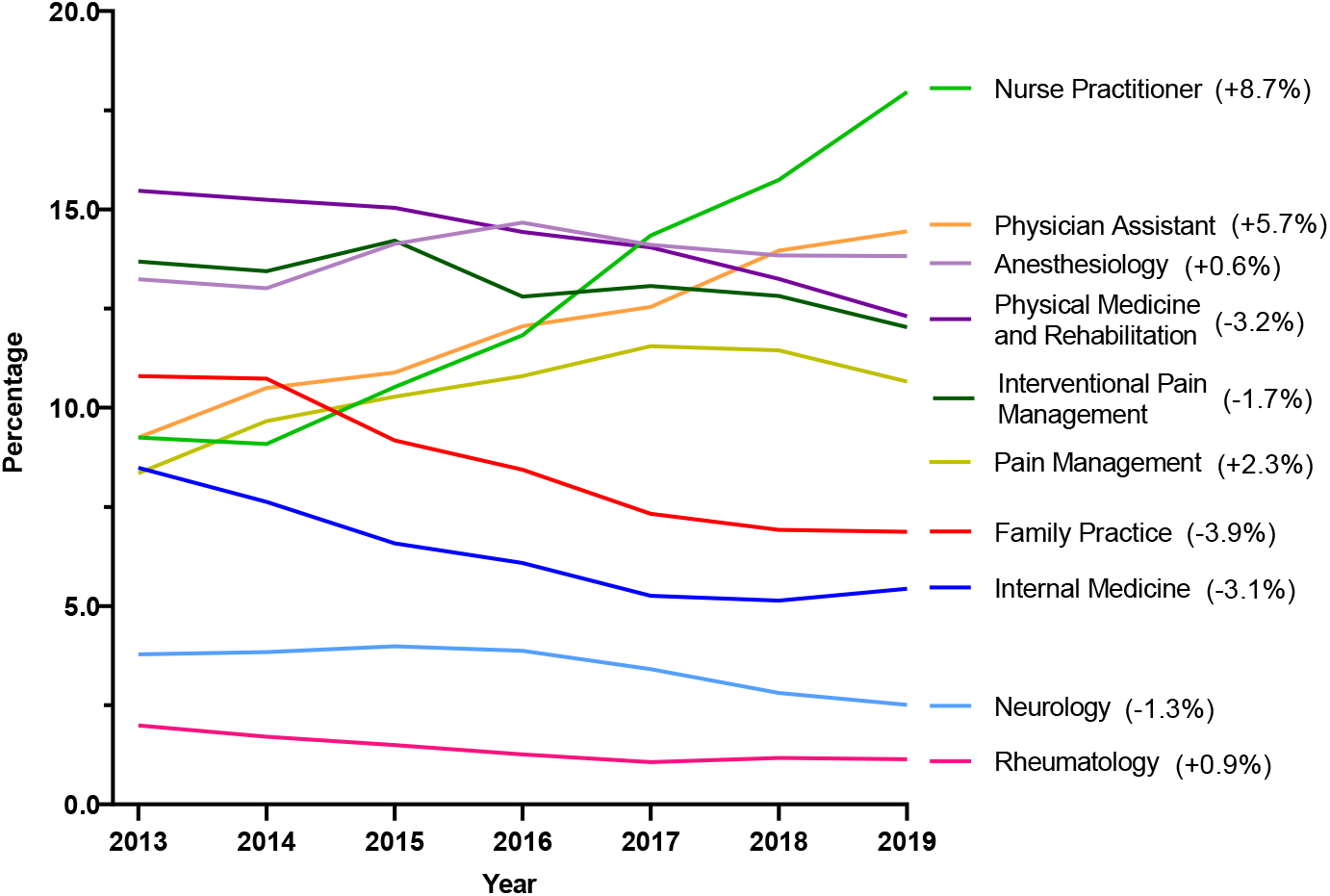
Amount of tapentadol claims in the US using data from Medicare Part D presented as percentages of a whole per year by specialties. Nurse Practitioners prescribed 18.0% of Medicare claims of tapentadol in 2019, followed by Physician Assistants 14.5% and Anesthesiologists 13.8%.

Because NH was atypical in ARCOS, further examination was completed for this state. Narrowing the focus on NH in the same period, Certified Registered Nurse Anesthetist (CRNA), saw a steep increase in percentage from 16.9% in 2013 to 59.7% in 2015 followed by a pronounced decline to 5.3% in 2019. Nurse Practitioners experienced a strong increase in the share of tapentadol’s prescription after the dip between 2013 and 2016 where it became the specialty with the largest share at 36.0% in 2019. Pain Management experienced a seven-fold increase from 1.9% in 2013 to 13.7% in 2019 (Supplemental Figure 2).

At the national level, correlational analysis between tapentadol’s distribution in ARCOS and the diabetes prevalence indicated the correlation coefficient was 0.44 for 2012 and it was 0.28 for 2020. Both correlations were statistically significant (Figure 5). Correlational analysis at the state level indicated Massachusetts, Wyoming, and Kentucky were statistically significant (Figure 6).

**Figure 5.**
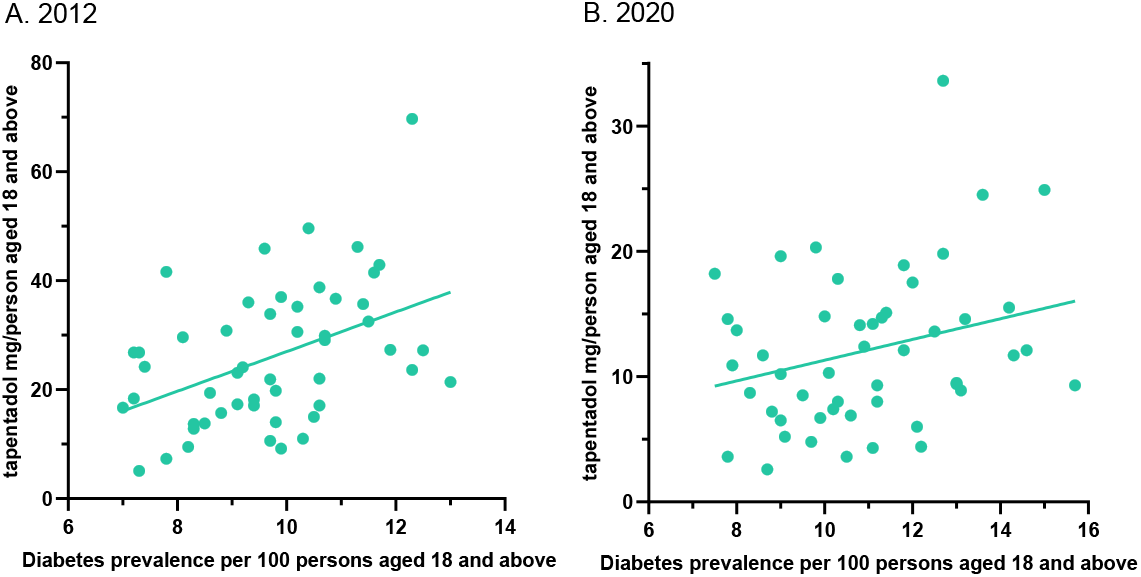
Correlation between tapentadol (mg/person aged 18 and above) and diabetes prevalence per 100 persons aged 18 and above indicated *r*(50) = 0.44, *p* < .01 for 2012 (**A**), and *r*(50) = 0.28, *p* < .05 for 2020 (**B**).

**Figure 6.**
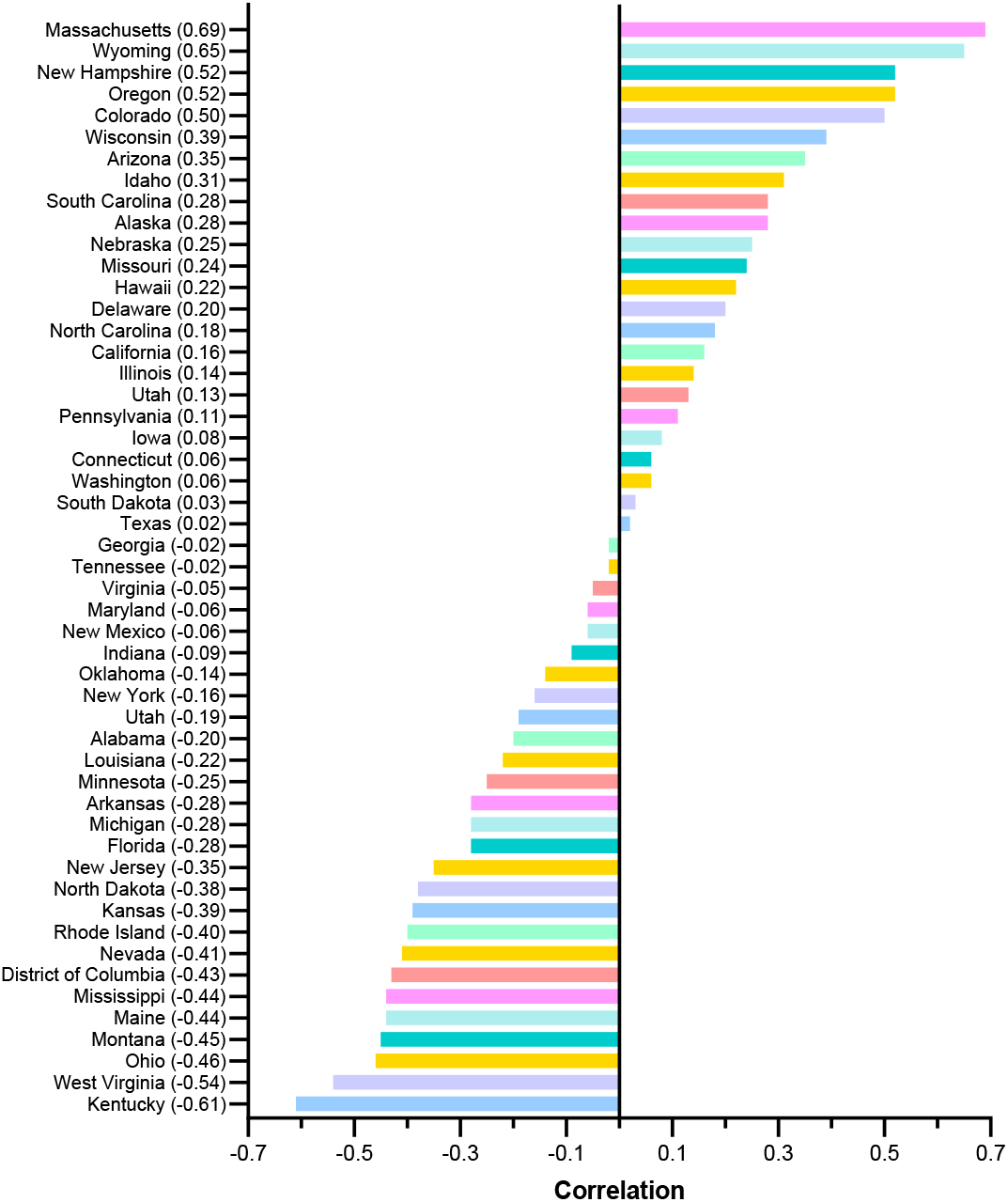
Correlation between tapentadol (mg/person aged 18 and above) and diabetes prevalence per 100 persons aged 18 and above for each state for years 2010 through 2020.

At the national level, the decreasing trend of distributed tapentadol per person compared with the trends other Schedule II Controlled Substances (fentanyl, hydrocodone, hydromorphone, morphine, oxycodone, oxymorphone) were similar such that the distribution of opioids peaked around 2012 and declined thereafter. Meperidine showed a strong and steady decline from 2010 to 2020. Methadone’s overall decline was less consistent than meperidine’s, and codeine had two peaks in 2011 and 2015 and declined thereafter. Buprenorphine, a Schedule III Controlled Substance, showed a steady and strong increase over the same period (Supplemental Figure 3A). For NH, the declining distribution trend of tapentadol compared with other Scheduled II Controlled Substances except methadone were less similar. In fact, the correlation between tapentadol and methadone was statistically significant at 0.81. Tapentadol (mg/person) peaked in 2012 and 2013 followed by a slightly higher peak in 2017, and this was followed by a mild and steady decrease until 2020. On the other hand, other Schedule II Controlled Substances, except codeine and methadone, demonstrated more prominent declines between 2012 and 2020. Like buprenorphine at the national level, buprenorphine at the NH level experienced a consistent and prominent increase over the same period (Supplemental Figure 4A).

## 4. Discussion

This study identified pronounced and regionally dependent changes in the prescribing trends of tapentadol in the US between 2010 and 2020 with a notable peak in 2012. Using ARCOS data, tapentadol decreased −53.8% between 2012 and 2020, and its decrease was ranked fourth among Schedule II opioids (Figure 1A). Given tapentadol is the only Schedule II opioid with a dual mechanism of action and has potentially fewer side effects than many other opioids, we wanted to better understand tapentadol’s use patterns.

According to the study using Medicare Part D data, prescribers may be incentivized to prescribe opioids that they receive opioid-specific payments. Prescribers who received incentives prescribed almost nine thousand daily doses each year more than prescribers who did not receive any opioid-specific, direct-to-physician incentives. In addition, the relationships between incentives and prescribing were largest for hydrocodone and oxycodone and were smallest for fentanyl and tapentadol.^20^ Our analysis based on ARCOS data showed the decrease in the distribution of tapentadol (−53.8%) was closer to the decrease in oxycodone (−48.7%) and hydrocodone (−58.1%) than fentanyl (−66.0%) between 2012 and 2020 (Figure 1A). Since the relationships between incentives and prescribing were small for both tapentadol and fentanyl, we expected tapentadol’s decrease to be more like fentanyl’s. However, tapentadol’s decrease was not as acute as that of fentanyl’s, indicating that the opioid-specific payments for tapentadol may not have played a crucial role in influencing the prescription of tapentadol.

Tapentadol production quotas were 207.2% of distribution. The DEA’s distributed amount was also significantly less than the quota (Figure 1B). The DEA’s final adjusted quota seeks to “ensure an adequate and uninterrupted supply of controlled substances in order to meet the demand of legitimate medical, scientific, and export needs of the United States.”^21^ Our data suggest DEA’s quota was not a determining factor influencing the amount of tapentadol prescribed. Presumably, this was because the DEA’s production quota also included tapentadol that was subsequently exported to other countries.

Investigating the possible factors influencing the distribution trends of opioids in the US, we found there were notable changes in the insurance and policy intervention landscapes in the period surrounding the electoral year 2012, likely in response to a November, 2011 communication from the Centers for Disease Control and Prevention (CDC) stating that overdoses from prescription opioid painkillers had reached epidemic levels.^22^ Additional states adopted state-level policy interventions such as mandated Prescription Drug Monitoring Programs (PDMPs) and pain clinic laws^23^ while the Centers for Medicare and Medicaid Services (CMS) implemented the Overutilization Monitoring System in 2013.^24^ In addition, private insurers began to take actions to limit the reimbursement of opioids.^25^ We recognize success in containing the opioid crisis varies across states as state-level policy interventions, insurance reimbursements, prescription rights, provider prescribing behavior, etc. differed considerably.^26,27^ Opioid overdose issue was once again highlighted in the electoral year 2016 leading to additional states joining the ranks in enacting state-wide policies to combat the opioid crisis and many others signing more stringent regulations such as prescription limit laws to limit the distribution of prescription opioids in 2016 and shortly after.^28^

Our data shows that all states in the US showed a reduction in the percentage change in the amount of tapentadol distributed between 2012 and 2020, except NH (+13.1%) and this was statistically significantly different from the national average at 5.0% alpha (Figure 2B). In general, distributed tapentadol decreased slower than other Schedule II substances except for methadone in NH (Supplemental Figure 4). Such observations were not found at the national level as Supplemental Figure 3 showed that the declining prescribing trend of tapentadol was more like those experienced by other Scheduled II Controlled Substances.

New Hampshire’s governor signed House Bill 1423 into law on June 7, 2016, and the Bill is seen as an initiative to curb the prescription of opioids. The legislation prohibits medical professionals from prescribing opioid prescriptions for more than seven days in an emergency room, urgent care setting, or walk-in clinic. The law also required that patients requiring opioids be prescribed the lowest effective dose of pain medications.^29^ Based on a report by Higgins et. al., we see legislation changes in opioid prescribing in NH undeniably reducing the amounts of opioids prescribed on hospital discharge after gynecologic surgery,^30^ and this indicates the effectiveness of opioid prescribing laws in affecting prescribing practices in NH. Supplemental 4 showed a marked decline in some Schedule II opioids in 2016 such as fentanyl and oxymorphone which may have been attributed to the implementation of House Bill 1423. NH prescription rate changed over the years. New Hampshire providers wrote 46.1 opioid prescriptions for every 100 persons in 2018 relative to the average US rate of 51.4 prescriptions.^31^ However, tapentadol and methadone were two Schedule II opioids that did not show similar decline patterns as other Scheduled II drugs (Supplemental Figure 4). Methadone, together with buprenorphine, a Schedule III opioid, and naloxone, a full opioid antagonist that is not classified as a controlled substance, are three medications approved by the US Food and Drug Administration (FDA) for treating opioid use disorders (OUDs). New Hampshire has been expanding the availability of addiction treatment through medication-assisted treatment (MAT) for OUDs, and all three medications are approved for use in MAT in NH. Methadone, per federal regulation, must be dispensed at certified opioid treatment programs (OTPs) whereas buprenorphine and naloxone may be prescribed in an office setting.^32^ A possible off-label use of tapentadol may include it being used as part of a step-down therapy in treating OUDs.^33^ Our data showed for NH, tapentadol’s trend resembled methadone’s with their correlation statistically significant at 0.81 and statistically nonsignificant with buprenorphine (Supplemental Figures 4A, 4F). On the national level, tapentadol’s correlation with either methadone or buprenorphine was not significant (Supplemental Figures 3A, 3F). This raises the question of tapentadol was used as part of a step-down therapy prior to the use of methadone in treating OUDs in NH. If this conjecture is accurate, tapentadol could have been used as a bridge, and it was prescribed prior to methadone. As such, tapentadol was not required to be administered at certified opioid treatment centers as pharmacies remained the most sizable channel of distribution as more than 97% of tapentadol distributed in the US from 2010 through 2020 were through pharmacies. This indicates that most tapentadol was prescribed in outpatient services and office-setting (Figure 1E). Tapentadol distributed by hospitals steadily increased to nearly 2% of the total in 2020 (Figure 1E), and this observation can be partly explained by Anesthesiology as a specialty increased in its share (13.8%) of the total tapentadol Medicare claims in 2019 (Figure 4). For both US and NH, Nurse Practitioner was the specialty that had the largest share of tapentadol prescribed using Medicare Part D data in 2019. Nurse Practitioners prescribed 18.0% of Medicare claims of tapentadol at the national level (Figure 4), and 36.0% in NH (Supplemental Figure 2). An unusual observation was that tapentadol prescribed by Pain Management as a specialty increased to 13.7% in NH (third-ranking specialty) but increased to only 10.7% (sixth ranking specialty) in the US. In addition, Anesthesiology as a specialty prescribed 13.8% of tapentadol Medicare claims in the US but only 3.0% in NH.

Recent research shows that Nurse Practitioners (NPs) “increase the number of opioids they prescribe following a grant of NP independence”^34^ and NPs and Physician Assistants (PAs) practicing in states with independent prescription authority “had more outliers who prescribed high-frequency, high dose opioids than did MDs.”^35^ Prescriptive rights of NPs differ across states. In NH, NPs were granted full authority to prescribe Scheduled II to IV controlled substances from year 2009.^36^ Our data showed that NPs and CRNAs experienced opposing trends between 2013 and 2019. NP decreased sharply from 57.1% in 2013 to 10.8% in 2016 then rebounded strongly to 36.0% and ranked first among all specialties in 2019, followed by Pain Management 13.7% and Physician Assistant 11.2% (Supplemental Figure 2).

According to the American Association of Nurse Practitioners, 70.2% of all NPs deliver primary care in 2020,^37^ and therefore there is an increased likelihood of NPs caring for chronic but less complex cases. We investigated if diabetes prevalence could have influenced the prescribing trend of tapentadol as this opioid has been marketed for diabetic neuropathy. The correlation between tapentadol (mg/person aged 18 and above) using ARCOS data and diabetes prevalence per 100 persons aged 18 and above at the national level showed the correlation coefficient, R, decreased from 0.44 in 2012 to 0.28 in 2020. Both correlations were statistically significant at 5.0% alpha indicating that diabetes prevalence had influenced the prescribing trend of tapentadol (Figure 5). Upon further investigation, the data showed the correlation in NH was not statistically significant and therefore, we fail to attribute NH’s tapentadol trend with the diabetes prevalence in NH (Figure 6).

The amount of tapentadol reported to DEA’s ARCOS strongly and significantly correlated with the amount of Medicaid claims at 0.80 (Figure 3A). Furthermore, the annual pattern for tapentadol from ARCOS closely mirrored that of Medicaid claims (Fig. 3B). We postulate that Medicaid policies did not particularly favor or disfavor the distribution of tapentadol over other opioids, but rather, tapentadol was distributed in a similar manner to the non-Medicaid patient population. The general ability of outcome detection was limited by having only eleven annual data points for each state in a subset of temporal analyses.

In conclusion, this study quantified the dynamic pattern in tapentadol distribution from 2010 to 2020 at national and state levels. Distributed tapentadol peaked in 2012. Thereafter, there has been a reduction in the distribution of tapentadol at both national and state levels with the only exception was that NH experienced a +13.1% increase. The declining tapentadol pattern is consistent with other opioids because, since 2011, there has been a steady decrease in the distribution of most medications used for pain.^15,38^ As tapentadol is the only Schedule II prescription opioid that has dual MOR and NRI mechanisms, and an opioid with a diversion rate significantly lower than that of other Scheduled II Controlled Substances,^9^ we question if tapentadol should be considered prior to other opioids for adults with a history of prescription opioid misuse. At the national level, the data showed the reduction in tapentadol distributed was not as pronounced as morphine, hydrocodone, fentanyl, oxymorphone, and meperidine (Figure 1A). However, distributed tapentadol was only 1.3% of Scheduled II opioids distributed in 2020 whereas oxycodone was 49.8%, and hydrocodone was 18.5% of the total distributed for Schedule II Controlled Substances (Figure 1D). In addition, the abnormal tapentadol distribution trend experienced by NH (+13.1% between 2012 and 2020) may warrant investigation in future studies such as differing prescribers’ attitudes towards tapentadol or perhaps the employment of tapentadol as part of a step-down therapy for opioid addiction.

## Supporting information

raw DEA data

## Data Availability

All data produced in the present study are available upon reasonable request to the authors.

https://www.deadiversion.usdoj.gov/arcos/retail_drug_summary/index.html

https://www.medicaid.gov/medicaid/prescription-drugs/state-drug-utilization-data/index.html

https://www.cms.gov/Research-Statistics-Data-and-Systems/Statistics-Trends-and-Reports/Medicare-Provider-Charge-Data/Part-D-Prescriber

## Acknowledgments

Jove H. Graham, PhD contributed to examining tapentadol prescribing within the Geisinger Health System. BJP was supported by the Health Resources Services Administration (D34HP31025). The software used for this report was provided by NIEHS (T32-ES007060-31A1).

## Supplemental Figures

**Supplemental Figure 1.**
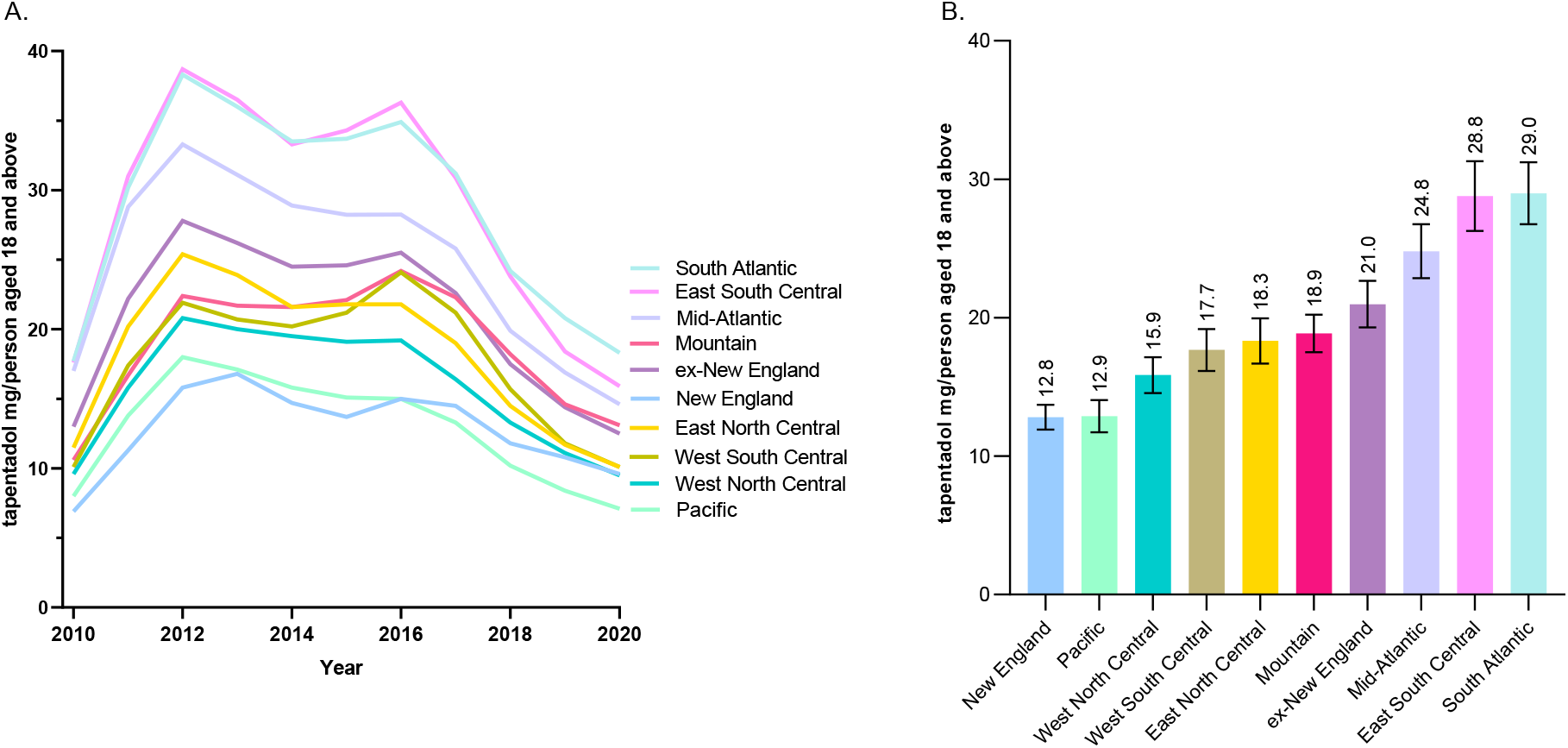
Tapentadol (mg/person aged 18 and above) in various regional divisions using data from the Drug Enforcement Administration’s Automated Reports and Consolidated Ordering System. **A**. Time-Series. **B**. New England and West North Central as well the Pacific were statistically insignificant. New England and all other divisions, as well as ex-New England, were statistically different at alpha 5.0%. Correlations between New England and all other divisions were statistically significant (p < 0.05).

**Supplemental Figure 2.**
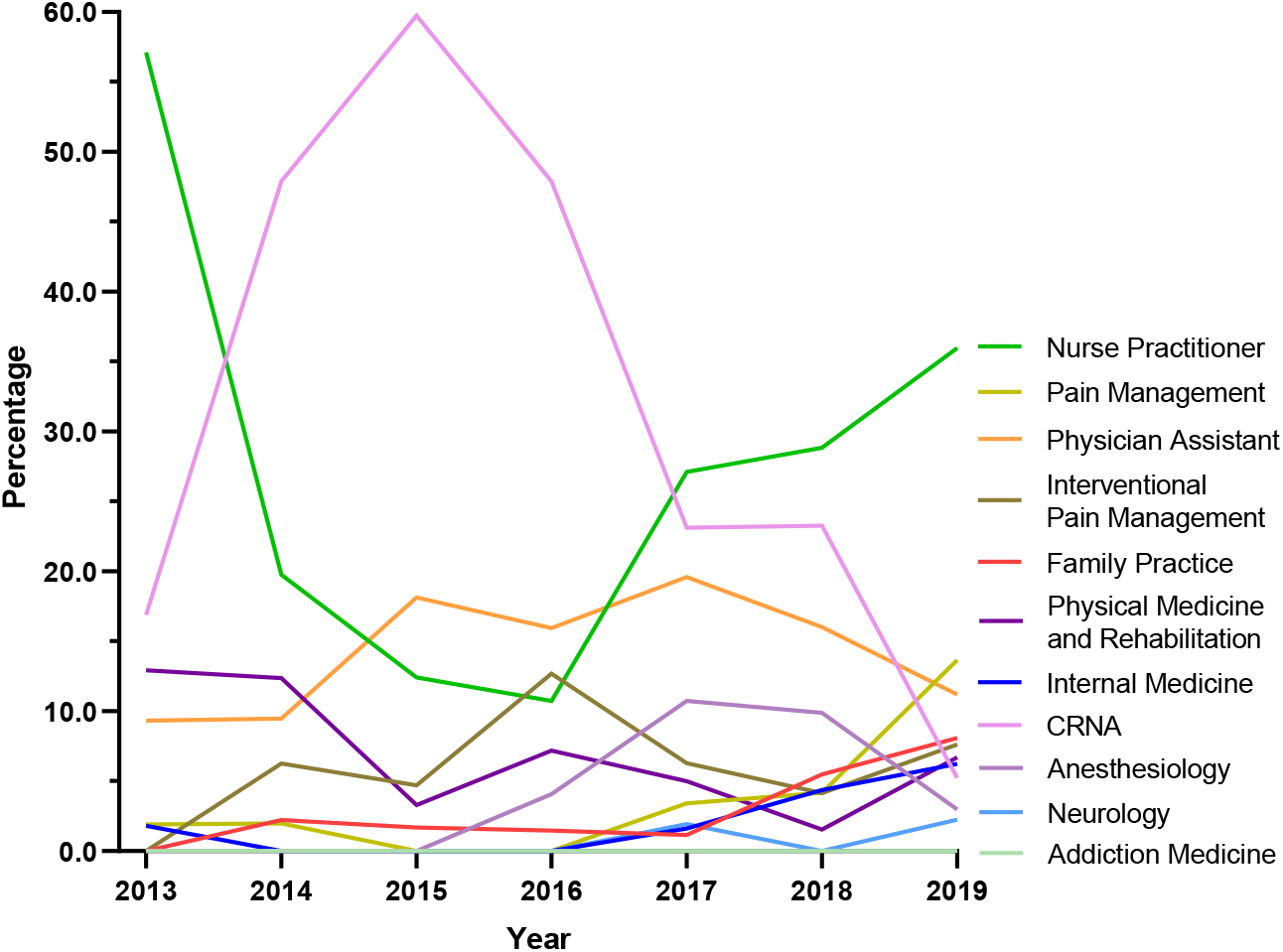
Amount of tapentadol claims in New Hampshire using data from Medicare Part D presented as percentages of a whole per year by specialties. Nurse Practitioners prescribed 36.0% Medicare claims of tapentadol in 2019, followed by Pain Management 13.7% and Physician Assistant 11.2%.

**Supplemental Figure 3.**
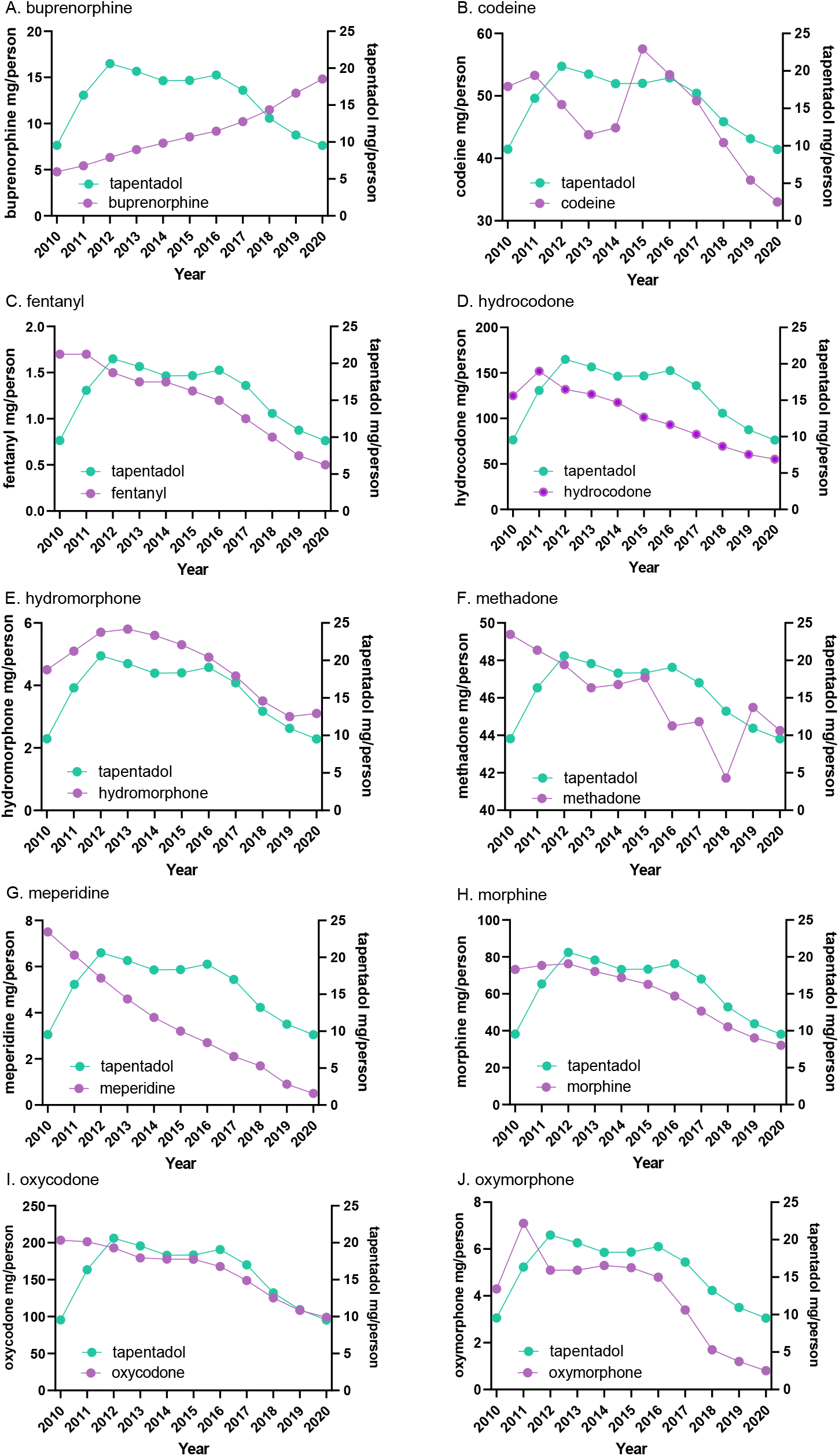
Amounts of tapentadol, Scheduled II Controlled Substances, and buprenorphine (mg/person) distributed in the US using data from the Drug Enforcement Administration’s Automated Reports and Consolidated Ordering System.

**Supplemental Figure 4.**
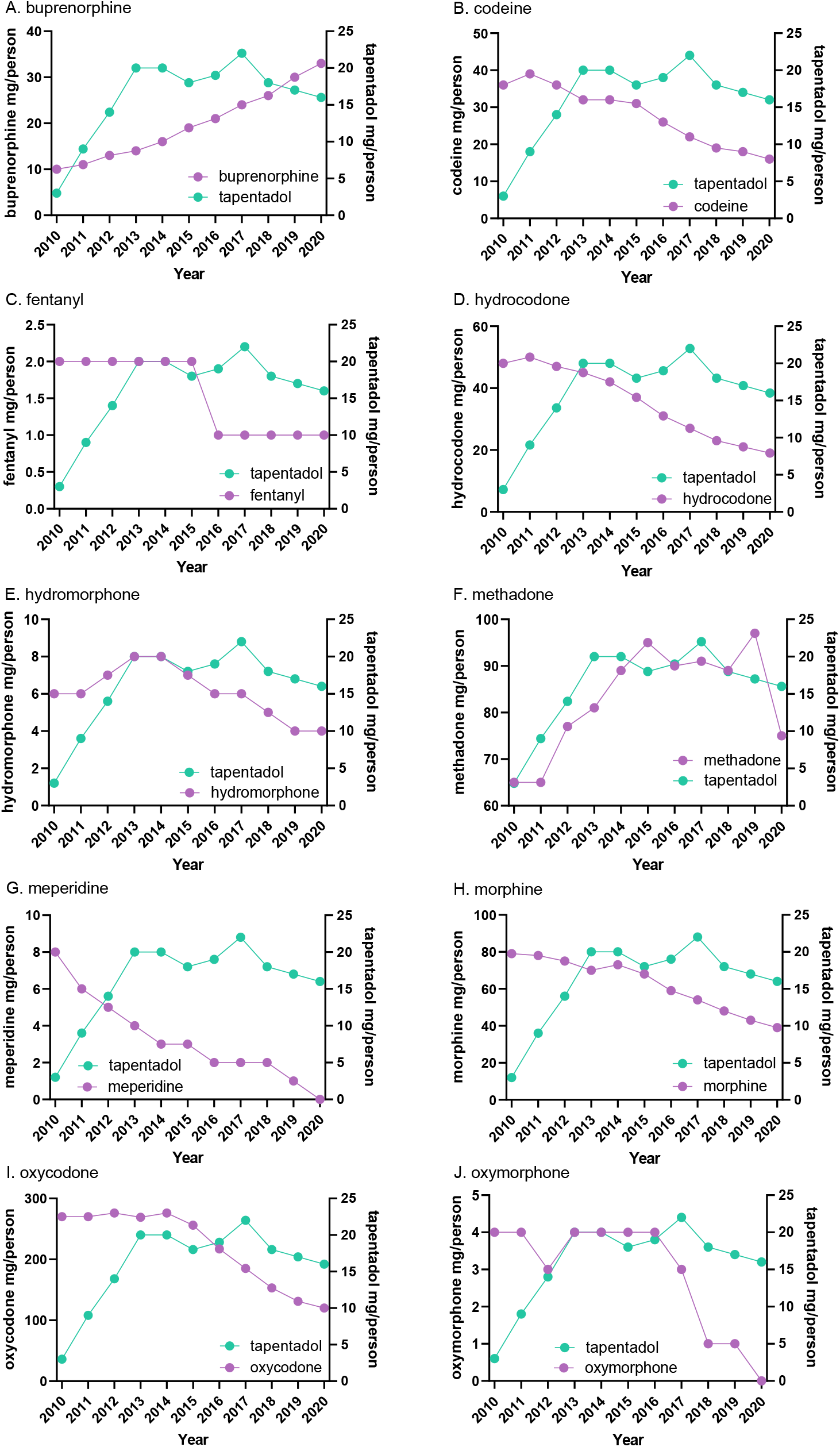
Amounts of tapentadol, Scheduled II Controlled Substances, and buprenorphine (mg/person) distributed in New Hampshire using data from the Drug Enforcement Administration’s Automated Reports and Consolidated Ordering System.

## Notes

Disclosures: BJP was part of an osteoarthritis research team supported by Pfizer and Eli Lilly. The other authors have no disclosures.

### Competing Interest Statement

BJP was part of an osteoarthritis research team supported by Pfizer and Eli Lilly. The other authors have no disclosures.

### Funding Statement

This study did not receive any external funding. BJP was supported by the Health Resources Services Administration (D34HP31025). The software used for this report was provided by NIEHS (T32-ES007060-31A1).

