## Supplementary material for "Declines and pronounced state-level variation in tapentadol use in the US": raw DEA data

### DEPARTMENT OF JUSTICE DRUG ENFORCEMENT ADMINISTRATION

#### DEA DIVERSION CONTROL DIVISION

#### ARCOS DRUG RETAIL SUMMARY REPORTS

REPORT 01 by GRAMS - RETAIL DRUG DISTRIBUTION BY ZIP CODE WITHIN STATE

REPORT 02 by GRAMS - RETAIL DRUG DISTRIBUTION BY STATE WITHIN DRUG CODE

REPORT 03 by GRAMS - QUARTERLY DRUG DISTRIBUTION BY STATE PER 100K POPULATION

REPORT 04 by GRAMS - CUMULATIVE DISTRIBUTION BY STATE PER 100K POPULATION

REPORT 05 by GRAMS - STATISTICAL SUMMARY FOR RETAIL DRUG PURCHASES

REPORT 07 by GRAMS - UNITED STATES SUMMARY FOR RETAIL DRUG PURCHASES

DATE RANGE: 01/01/2020 TO 12/31/2020

**DISCLAIMER**: Automated Reports and Consolidated Ordering System (ARCOS) is a data collection system in which manufacturers and distributors report their controlled substances transactions to the Drug Enforcement Administration (DEA). ARCOS provides an acquisition/distribution transactional records of applicable activities to the DEA involving certain controlled substances, in accordance with Title 21, United States Code, Section 827(d)(1), and Title 21, Code of Federal Regulations, Section 1304.33. This information is collected and compiled by DEA in accordance with law for determining quota, distribution trends, internal audits, and other analyses.

Recipients should be aware, prior to use, analysis, studies, or publication of ARCOS data, the information contained in ARCOS generated reports are complete and accurate as to what was provided by DEA registrants to ARCOS in accordance with reporting requirements. This information is susceptible to future updates and corrections, without notice, as new information is obtained. DEA assumes no liability for analysis, conclusions or policy decisions of third parties based on internal interpretation of provided data.

DRUG: 1100 - AMPHETAMINE

STATE: AK - ALASKA

| REGISTRANT ZIP CODE 3 | 1ST QUARTER | 2ND QUARTER | 3RD QUARTER | 4TH QUARTER | TOTAL GRAMS |
|---|---|---|---|---|---|
| 995 | 5,097.87 | 4,776.84 | 5,091.24 | 5,046.39 | **20,012.34** |
| 996 | 1,787.55 | 1,811.43 | 2,013.64 | 2,011.10 | **7,623.72** |
| 997 | 1,189.97 | 1,245.75 | 1,317.81 | 1,204.19 | **4,957.72** |
| 998 | 515.12 | 518.47 | 610.39 | 590.51 | **2,234.49** |
| 999 | 185.87 | 171.04 | 184.97 | 181.77 | **723.65** |
| **AK - ALASKA - Total** | **8,776.37** | **8,523.53** | **9,218.04** | **9,033.97** | **35,551.91** |

STATE: AL - ALABAMA

| REGISTRANT ZIP CODE 3 | 1ST QUARTER | 2ND QUARTER | 3RD QUARTER | 4TH QUARTER | TOTAL GRAMS |
|---|---|---|---|---|---|
| 350 | 18,752.35 | 18,458.78 | 18,964.73 | 19,684.57 | **75,860.42** |
| 351 | 9,619.61 | 9,678.19 | 9,915.98 | 10,014.38 | **39,228.15** |
| 352 | 20,432.76 | 19,603.33 | 20,020.31 | 21,000.96 | **81,057.37** |
| 354 | 8,417.72 | 7,406.55 | 7,851.25 | 8,229.32 | **31,904.84** |
| 355 | 3,367.90 | 3,651.08 | 3,609.64 | 3,820.11 | **14,448.74** |
| 356 | 10,976.73 | 10,688.37 | 11,577.87 | 11,618.55 | **44,861.51** |
| 357 | 5,417.90 | 5,092.13 | 5,480.09 | 5,729.03 | **21,719.15** |
| 358 | 6,287.92 | 5,945.85 | 6,496.66 | 6,640.31 | **25,370.74** |
| 359 | 5,628.29 | 5,607.45 | 5,760.31 | 5,946.53 | **22,942.58** |
| 360 | 6,791.25 | 6,645.36 | 6,725.17 | 6,922.19 | **27,083.97** |
| 361 | 4,992.55 | 4,788.86 | 4,728.93 | 4,938.70 | **19,449.04** |
| 362 | 2,724.06 | 2,592.14 | 2,622.15 | 2,777.01 | **10,715.35** |
| 363 | 5,106.50 | 4,918.93 | 5,352.84 | 5,100.30 | **20,478.56** |
| 364 | 1,486.20 | 1,407.05 | 1,469.44 | 1,544.48 | **5,907.18** |
| 365 | 12,329.89 | 12,484.30 | 12,607.44 | 13,151.47 | **50,573.09** |
| 366 | 8,256.55 | 7,624.01 | 8,040.81 | 8,434.99 | **32,356.36** |
| 367 | 1,549.27 | 1,455.54 | 1,485.99 | 1,578.22 | **6,069.01** |
| 368 | 6,655.54 | 6,249.50 | 7,033.49 | 7,116.24 | **27,054.77** |
| 369 | 120.31 | 138.49 | 117.16 | 147.60 | **523.55** |
| **AL - ALABAMA - Total** | **138,913.28** | **134,435.90** | **139,860.25** | **144,394.96** | **557,604.40** |

STATE: AR - ARKANSAS

| REGISTRANT ZIP CODE 3 | 1ST QUARTER | 2ND QUARTER | 3RD QUARTER | 4TH QUARTER | TOTAL GRAMS |
|---|---|---|---|---|---|
| 716 | 2,862.38 | 2,637.82 | 2,768.70 | 2,906.03 | **11,174.94** |
| 717 | 1,080.25 | 962.55 | 1,057.47 | 1,157.89 | **4,258.16** |
| 718 | 981.20 | 964.49 | 1,047.21 | 1,105.15 | **4,098.05** |
| 719 | 3,623.86 | 3,678.61 | 3,701.75 | 3,864.50 | **14,868.72** |
| 720 | 8,971.52 | 8,937.58 | 9,253.91 | 9,698.10 | **36,861.10** |
| 721 | 6,657.20 | 6,677.21 | 6,687.36 | 7,123.97 | **27,145.73** |
| 722 | 7,836.08 | 7,578.27 | 7,914.53 | 8,192.18 | **31,521.07** |
| 723 | 1,659.02 | 1,633.42 | 1,628.81 | 1,779.28 | **6,700.53** |
| 724 | 3,203.87 | 2,915.32 | 3,056.54 | 3,374.37 | **12,550.09** |
| 725 | 850.38 | 854.93 | 847.24 | 856.32 | **3,408.87** |
| 726 | 1,663.68 | 1,686.92 | 1,695.55 | 1,772.84 | **6,818.99** |
| 727 | 14,460.30 | 14,644.00 | 15,103.31 | 15,974.34 | **60,181.96** |
| 728 | 1,155.14 | 1,077.34 | 1,122.74 | 1,207.96 | **4,563.18** |
| 729 | 2,036.05 | 2,119.74 | 2,103.28 | 2,308.82 | **8,567.88** |
| **AR - ARKANSAS - Total** | **57,040.92** | **56,368.21** | **57,988.40** | **61,321.73** | **232,719.27** |

STATE: AS - AMERICAN SAMOA

| REGISTRANT ZIP CODE 3 | 1ST QUARTER | 2ND QUARTER | 3RD QUARTER | 4TH QUARTER | TOTAL GRAMS |
|---|---|---|---|---|---|
| 967 | 1.25 | 0 | 1.25 | 1.25 | **3.75** |
| **AS - AMERICAN SAMOA - Total** | **1.25** | **0.00** | **1.25** | **1.25** | **3.75** |

STATE: AZ - ARIZONA

| REGISTRANT ZIP CODE 3 | 1ST QUARTER | 2ND QUARTER | 3RD QUARTER | 4TH QUARTER | TOTAL GRAMS |
|---|---|---|---|---|---|
| 850 | 22,656.82 | 20,779.38 | 21,799.15 | 22,713.51 | **87,948.86** |
| 851 | 4,662.85 | 4,214.24 | 4,574.62 | 4,944.18 | **18,395.90** |
| 852 | 44,037.87 | 42,577.26 | 44,482.48 | 46,976.96 | **178,074.56** |
| 853 | 14,227.00 | 11,798.28 | 13,135.65 | 14,260.15 | **53,421.07** |
| 855 | 578.83 | 454.23 | 542.87 | 521.01 | **2,096.94** |
| 856 | 2,106.89 | 1,754.56 | 2,064.21 | 2,090.65 | **8,016.32** |

| | | | | | |
|---|---|---|---|---|---|
| 857 | 13,140.85 | 10,482.55 | 12,025.38 | 12,755.65 | **48,404.42** |
| 859 | 620.84 | 564.46 | 663.43 | 622.43 | **2,471.16** |
| 860 | 1,565.48 | 1,336.37 | 1,560.01 | 1,626.76 | **6,088.63** |
| 863 | 2,189.06 | 1,973.49 | 2,156.93 | 2,325.71 | **8,645.18** |
| 864 | 1,199.07 | 998.34 | 1,057.26 | 1,155.83 | **4,410.50** |
| 865 | 82.62 | 66.63 | 84.34 | 58.88 | **292.46** |
| **AZ - ARIZONA - Total** | **107,068.17** | **96,999.79** | **104,146.34** | **110,051.71** | **418,266.00** |

**STATE: CA - CALIFORNIA**

| REGISTRANT ZIP CODE 3 | 1ST QUARTER | 2ND QUARTER | 3RD QUARTER | 4TH QUARTER | TOTAL GRAMS |
|---|---|---|---|---|---|
| 900 | 24,368.14 | 21,796.54 | 23,107.46 | 23,021.46 | **92,293.61** |
| 902 | 14,467.99 | 13,450.21 | 13,970.77 | 14,418.77 | **56,307.74** |
| 903 | 312.79 | 313.62 | 300.69 | 367.41 | **1,294.52** |
| 904 | 4,327.92 | 4,132.90 | 4,028.40 | 4,501.89 | **16,991.11** |
| 905 | 2,105.33 | 1,867.94 | 2,160.91 | 2,185.05 | **8,319.24** |
| 906 | 2,933.62 | 2,019.74 | 2,536.63 | 2,773.36 | **10,263.35** |
| 907 | 3,882.54 | 3,272.90 | 3,335.20 | 3,521.38 | **14,012.03** |
| 908 | 3,639.42 | 3,355.15 | 3,559.97 | 3,772.19 | **14,326.73** |
| 910 | 1,592.57 | 1,309.77 | 1,425.25 | 1,519.71 | **5,847.31** |
| 911 | 2,879.90 | 2,771.64 | 2,780.93 | 2,946.61 | **11,379.08** |
| 912 | 1,918.28 | 1,776.94 | 1,967.00 | 1,985.71 | **7,647.93** |
| 913 | 16,179.30 | 15,017.73 | 15,568.22 | 15,794.57 | **62,559.82** |
| 914 | 5,218.82 | 4,397.56 | 4,811.45 | 4,727.26 | **19,155.09** |
| 915 | 1,813.07 | 1,565.89 | 1,660.32 | 1,859.54 | **6,898.81** |
| 916 | 2,066.42 | 1,796.71 | 2,024.64 | 2,056.50 | **7,944.27** |
| 917 | 7,989.21 | 6,777.88 | 7,380.78 | 7,857.22 | **30,005.10** |
| 918 | 539.76 | 465.81 | 530.29 | 585.55 | **2,121.42** |
| 919 | 3,060.98 | 3,175.79 | 3,096.25 | 3,410.36 | **12,743.38** |
| 920 | 28,965.34 | 30,355.13 | 30,231.47 | 31,946.74 | **121,498.68** |
| 921 | 19,536.29 | 18,288.74 | 19,074.79 | 19,443.94 | **76,343.76** |
| 922 | 4,243.41 | 3,541.55 | 3,904.24 | 4,077.87 | **15,767.07** |
| 923 | 4,837.66 | 4,153.40 | 4,311.48 | 4,485.76 | **17,788.30** |
| 924 | 354.31 | 267.65 | 301.39 | 322.35 | **1,245.69** |
| 925 | 7,078.34 | 6,064.46 | 6,379.98 | 6,730.90 | **26,253.67** |
| 926 | 22,314.46 | 20,911.12 | 21,937.74 | 22,484.28 | **87,647.60** |
| 927 | 5,106.72 | 4,570.96 | 4,528.40 | 4,523.27 | **18,729.36** |
| 928 | 9,325.76 | 8,419.50 | 8,898.98 | 9,040.95 | **35,685.19** |
| 930 | 6,113.23 | 5,931.38 | 6,023.92 | 6,419.54 | **24,488.08** |
| 931 | 3,538.19 | 3,247.29 | 3,312.37 | 3,386.54 | **13,484.39** |
| 932 | 2,269.64 | 1,818.55 | 1,995.68 | 2,067.59 | **8,151.45** |
| 933 | 2,358.90 | 1,803.68 | 2,072.15 | 2,002.73 | **8,237.46** |
| 934 | 3,323.62 | 3,231.70 | 3,315.93 | 3,488.36 | **13,359.62** |
| 935 | 2,445.96 | 2,199.61 | 2,343.39 | 2,390.93 | **9,379.90** |
| 936 | 2,507.56 | 2,065.19 | 2,384.67 | 2,425.24 | **9,382.66** |
| 937 | 4,110.07 | 3,344.38 | 3,554.49 | 3,756.59 | **14,765.54** |
| 939 | 1,835.99 | 1,909.86 | 2,016.75 | 2,081.32 | **7,843.93** |
| 940 | 6,741.84 | 6,037.83 | 6,381.77 | 6,602.27 | **25,763.71** |
| 941 | 12,977.67 | 11,533.96 | 11,240.03 | 11,625.24 | **47,376.89** |
| 943 | 961.57 | 846.47 | 884.35 | 856.13 | **3,548.51** |
| 944 | 1,367.10 | 1,417.65 | 1,358.98 | 1,451.05 | **5,594.78** |
| 945 | 17,684.73 | 17,484.09 | 17,856.33 | 18,986.72 | **72,011.87** |
| 946 | 4,002.10 | 3,905.88 | 4,025.46 | 4,189.09 | **16,122.53** |
| 947 | 1,483.78 | 1,361.96 | 1,411.55 | 1,412.10 | **5,669.39** |
| 948 | 816.21 | 713.00 | 721.54 | 751.91 | **3,002.66** |
| 949 | 5,236.85 | 5,159.85 | 5,189.53 | 5,415.23 | **21,001.45** |
| 950 | 7,490.21 | 6,868.13 | 7,023.28 | 7,662.28 | **29,043.90** |
| 951 | 4,308.14 | 4,230.34 | 4,177.39 | 4,576.27 | **17,292.14** |
| 952 | 2,052.89 | 1,984.89 | 1,834.06 | 1,964.69 | **7,836.53** |
| 953 | 5,226.61 | 4,798.28 | 4,880.26 | 5,219.26 | **20,124.41** |
| 954 | 3,899.41 | 3,912.15 | 3,957.08 | 4,075.01 | **15,843.65** |
| 955 | 944.83 | 887.91 | 938.69 | 978.70 | **3,750.13** |

| | | | | | |
|---|---|---|---|---|---|
| 956 | 11,053.41 | 10,652.13 | 11,025.13 | 11,978.75 | **44,709.42** |
| 957 | 2,745.10 | 2,632.30 | 2,648.86 | 2,867.82 | **10,894.08** |
| 958 | 5,525.58 | 5,277.39 | 5,315.81 | 5,762.09 | **21,880.87** |
| 959 | 3,991.71 | 3,777.29 | 3,892.68 | 3,932.24 | **15,593.91** |
| 960 | 2,054.51 | 1,930.74 | 2,124.40 | 2,135.83 | **8,245.48** |
| 961 | 793.66 | 923.48 | 971.70 | 1,019.61 | **3,708.46** |
| **CA - CALIFORNIA - Total** | **330,919.44** | **307,724.58** | **318,691.84** | **331,841.77** | **1,289,177.62** |

**STATE: CO - COLORADO**

| REGISTRANT ZIP CODE 3 | 1ST QUARTER | 2ND QUARTER | 3RD QUARTER | 4TH QUARTER | TOTAL GRAMS |
|---|---|---|---|---|---|
| 800 | 17,652.32 | 15,702.04 | 16,968.28 | 18,244.72 | **68,567.37** |
| 801 | 18,125.74 | 17,213.37 | 18,601.96 | 19,753.72 | **73,694.80** |
| 802 | 24,629.70 | 24,856.96 | 25,983.50 | 26,381.48 | **101,851.64** |
| 803 | 4,072.96 | 3,123.10 | 3,648.83 | 3,811.08 | **14,655.96** |
| 804 | 3,692.42 | 3,603.39 | 3,976.66 | 4,156.85 | **15,429.33** |
| 805 | 10,131.45 | 9,325.23 | 10,290.15 | 11,086.70 | **40,833.53** |
| 806 | 3,053.28 | 2,346.02 | 2,888.22 | 3,120.02 | **11,407.54** |
| 807 | 498.81 | 365.80 | 414.40 | 481.09 | **1,760.10** |
| 808 | 1,534.58 | 1,328.49 | 1,622.41 | 1,482.29 | **5,967.78** |
| 809 | 10,090.00 | 8,176.92 | 9,890.33 | 9,868.78 | **38,026.04** |
| 810 | 1,882.03 | 1,623.05 | 1,699.64 | 1,843.03 | **7,047.75** |
| 811 | 386.92 | 378.04 | 469.66 | 473.12 | **1,707.73** |
| 812 | 725.85 | 688.65 | 831.13 | 845.52 | **3,091.15** |
| 813 | 887.28 | 784.56 | 863.52 | 949.08 | **3,484.44** |
| 814 | 760.57 | 742.29 | 823.86 | 857.23 | **3,183.94** |
| 815 | 2,253.66 | 2,069.81 | 2,217.49 | 2,316.02 | **8,856.98** |
| 816 | 3,006.03 | 2,741.86 | 3,160.49 | 3,019.68 | **11,928.06** |
| **CO - COLORADO - Total** | **103,383.61** | **95,069.58** | **104,350.53** | **108,690.40** | **411,494.11** |

**STATE: CT - CONNECTICUT**

| REGISTRANT ZIP CODE 3 | 1ST QUARTER | 2ND QUARTER | 3RD QUARTER | 4TH QUARTER | TOTAL GRAMS |
|---|---|---|---|---|---|
| 060 | 10,389.23 | 10,204.57 | 10,603.10 | 11,248.08 | **42,444.98** |
| 061 | 3,748.12 | 3,485.66 | 3,471.43 | 3,712.66 | **14,417.87** |
| 062 | 2,969.72 | 2,792.65 | 2,875.81 | 3,020.83 | **11,659.00** |
| 063 | 6,533.82 | 6,343.10 | 6,952.52 | 6,917.72 | **26,747.15** |
| 064 | 14,652.90 | 14,778.41 | 14,917.04 | 15,716.57 | **60,064.92** |
| 065 | 4,752.42 | 4,549.12 | 4,748.48 | 4,811.08 | **18,861.10** |
| 066 | 2,075.82 | 1,993.51 | 2,021.81 | 2,180.36 | **8,271.50** |
| 067 | 4,178.57 | 4,288.97 | 4,475.18 | 4,647.76 | **17,590.48** |
| 068 | 11,172.53 | 11,914.22 | 11,741.39 | 12,115.38 | **46,943.52** |
| 069 | 2,344.71 | 2,421.63 | 2,471.55 | 2,507.39 | **9,745.29** |
| **CT - CONNECTICUT - Total** | **62,817.84** | **62,771.85** | **64,278.31** | **66,877.82** | **256,745.82** |

**STATE: DC - DISTRICT OF COLUMBIA**

| REGISTRANT ZIP CODE 3 | 1ST QUARTER | 2ND QUARTER | 3RD QUARTER | 4TH QUARTER | TOTAL GRAMS |
|---|---|---|---|---|---|
| 200 | 12,635.49 | 10,543.02 | 11,562.76 | 11,395.45 | **46,136.72** |
| 203 | 107.29 | 88.53 | 68.51 | 40.04 | **304.37** |
| 204 | 649.17 | 541.39 | 557.12 | 592.66 | **2,340.35** |
| **DC - DISTRICT OF COLUMBIA - Total** | **13,391.96** | **11,172.94** | **12,188.38** | **12,028.15** | **48,781.43** |

**STATE: DE - DELAWARE**

| REGISTRANT ZIP CODE 3 | 1ST QUARTER | 2ND QUARTER | 3RD QUARTER | 4TH QUARTER | TOTAL GRAMS |
|---|---|---|---|---|---|
| 197 | 8,881.88 | 8,692.38 | 8,711.96 | 9,293.01 | **35,579.23** |
| 198 | 6,636.74 | 6,412.41 | 6,375.04 | 6,877.49 | **26,301.68** |
| 199 | 6,140.19 | 6,666.82 | 6,724.25 | 6,687.31 | **26,218.57** |
| **DE - DELAWARE - Total** | **21,658.81** | **21,771.61** | **21,811.24** | **22,857.81** | **88,099.48** |

**STATE: FL - FLORIDA**

| REGISTRANT ZIP CODE 3 | 1ST QUARTER | 2ND QUARTER | 3RD QUARTER | 4TH QUARTER | TOTAL GRAMS |
|---|---|---|---|---|---|
| 320 | 11,977.66 | 11,150.12 | 12,132.93 | 12,780.11 | **48,040.82** |
| 321 | 10,314.90 | 8,841.39 | 10,141.09 | 10,505.61 | **39,803.00** |
| 322 | 25,833.10 | 23,703.32 | 25,783.07 | 25,970.48 | **101,289.97** |
| 323 | 10,426.44 | 9,343.54 | 9,980.20 | 10,198.89 | **39,949.06** |
| 324 | 8,310.01 | 7,973.88 | 8,424.62 | 8,639.52 | **33,348.02** |
| 325 | 21,451.34 | 20,576.20 | 21,166.71 | 21,854.41 | **85,048.65** |

| | | | | | |
|---|---|---|---|---|---|
| 326 | 7,292.14 | 6,389.62 | 6,913.78 | 7,153.57 | **27,749.11** |
| 327 | 21,392.79 | 19,367.62 | 21,480.57 | 22,384.14 | **84,625.11** |
| 328 | 16,304.39 | 13,535.82 | 15,459.59 | 15,729.76 | **61,029.56** |
| 329 | 16,357.56 | 14,569.54 | 15,434.22 | 15,856.57 | **62,217.89** |
| 330 | 19,059.88 | 15,600.58 | 18,149.56 | 18,620.26 | **71,430.28** |
| 331 | 22,176.33 | 17,179.06 | 21,088.08 | 22,048.28 | **82,491.75** |
| 333 | 15,366.60 | 13,096.95 | 15,303.93 | 15,470.22 | **59,237.71** |
| 334 | 31,282.46 | 27,478.66 | 30,387.93 | 31,255.67 | **120,404.72** |
| 335 | 14,098.71 | 13,160.04 | 14,575.30 | 15,097.40 | **56,931.45** |
| 336 | 18,097.08 | 16,189.43 | 18,072.23 | 18,135.68 | **70,494.41** |
| 337 | 18,514.54 | 17,156.39 | 18,499.18 | 19,692.66 | **73,862.77** |
| 338 | 9,532.41 | 8,121.76 | 9,412.37 | 10,020.87 | **37,087.40** |
| 339 | 16,488.03 | 15,057.03 | 16,083.08 | 16,831.67 | **64,459.80** |
| 341 | 9,483.72 | 8,584.30 | 8,984.54 | 9,581.23 | **36,633.78** |
| 342 | 17,237.81 | 15,376.64 | 17,178.35 | 17,757.22 | **67,550.03** |
| 344 | 5,516.02 | 4,513.45 | 5,250.53 | 5,563.15 | **20,843.15** |
| 346 | 12,566.18 | 10,587.49 | 12,215.21 | 12,652.89 | **48,021.78** |
| 347 | 10,997.38 | 9,195.55 | 10,613.40 | 10,921.16 | **41,727.49** |
| 349 | 8,511.36 | 7,740.34 | 8,882.56 | 8,839.47 | **33,973.73** |
| **FL - FLORIDA - Total** | **378,588.81** | **334,488.72** | **371,613.02** | **383,560.88** | **1,468,251.43** |

**STATE: GA - GEORGIA**

| REGISTRANT ZIP CODE 3 | 1ST QUARTER | 2ND QUARTER | 3RD QUARTER | 4TH QUARTER | TOTAL GRAMS |
|---|---|---|---|---|---|
| 300 | 44,293.07 | 42,260.20 | 44,409.42 | 46,373.97 | **177,336.66** |
| 301 | 25,969.95 | 24,715.90 | 26,490.63 | 27,385.99 | **104,562.47** |
| 302 | 16,166.07 | 14,981.93 | 15,798.85 | 16,636.52 | **63,583.38** |
| 303 | 27,135.95 | 25,068.88 | 26,065.79 | 26,802.95 | **105,073.57** |
| 304 | 7,390.73 | 6,873.97 | 7,289.50 | 7,518.74 | **29,072.93** |
| 305 | 14,103.72 | 13,707.03 | 14,471.02 | 14,756.29 | **57,038.05** |
| 306 | 9,518.07 | 8,645.15 | 9,601.59 | 9,909.76 | **37,674.57** |
| 307 | 6,875.01 | 6,474.51 | 6,810.72 | 7,114.93 | **27,275.18** |
| 308 | 4,527.94 | 4,230.08 | 4,376.26 | 4,519.55 | **17,653.83** |
| 309 | 5,716.78 | 5,232.76 | 5,810.82 | 5,339.38 | **22,099.74** |
| 310 | 8,354.38 | 7,601.88 | 7,979.31 | 8,636.74 | **32,572.31** |
| 312 | 2,731.45 | 2,645.84 | 2,726.19 | 2,720.22 | **10,823.70** |
| 313 | 6,336.06 | 5,932.08 | 6,613.31 | 6,312.00 | **25,193.46** |
| 314 | 7,153.69 | 6,837.10 | 6,946.01 | 7,148.35 | **28,085.14** |
| 315 | 9,568.01 | 8,564.43 | 9,452.79 | 9,548.12 | **37,133.35** |
| 316 | 3,818.76 | 3,200.02 | 3,607.44 | 3,675.57 | **14,301.80** |
| 317 | 8,137.02 | 7,034.93 | 7,632.53 | 8,048.47 | **30,852.95** |
| 318 | 651.86 | 520.82 | 577.95 | 615.01 | **2,365.63** |
| 319 | 6,651.79 | 6,291.69 | 6,698.50 | 6,437.98 | **26,079.96** |
| 398 | 1,778.97 | 1,342.07 | 1,476.30 | 1,678.80 | **6,276.14** |
| **GA - GEORGIA - Total** | **216,879.29** | **202,161.25** | **214,834.94** | **221,179.33** | **855,054.81** |

**STATE: GU - GUAM**

| REGISTRANT ZIP CODE 3 | 1ST QUARTER | 2ND QUARTER | 3RD QUARTER | 4TH QUARTER | TOTAL GRAMS |
|---|---|---|---|---|---|
| 969 | 458.80 | 400.33 | 605.21 | 409.16 | **1,873.50** |
| **GU - GUAM - Total** | **458.80** | **400.33** | **605.21** | **409.16** | **1,873.50** |

**STATE: HI - HAWAII**

| REGISTRANT ZIP CODE 3 | 1ST QUARTER | 2ND QUARTER | 3RD QUARTER | 4TH QUARTER | TOTAL GRAMS |
|---|---|---|---|---|---|
| 967 | 5,870.99 | 5,404.20 | 5,855.80 | 6,421.17 | **23,552.16** |
| 968 | 5,611.98 | 4,690.48 | 5,241.90 | 5,138.07 | **20,682.44** |
| **HI - HAWAII - Total** | **11,482.97** | **10,094.69** | **11,097.70** | **11,559.24** | **44,234.60** |

**STATE: IA - IOWA**

| REGISTRANT ZIP CODE 3 | 1ST QUARTER | 2ND QUARTER | 3RD QUARTER | 4TH QUARTER | TOTAL GRAMS |
|---|---|---|---|---|---|
| 500 | 4,635.01 | 4,342.79 | 4,828.49 | 4,917.91 | **18,724.20** |
| 501 | 2,095.48 | 2,027.73 | 2,116.22 | 2,191.18 | **8,430.61** |
| 502 | 3,472.57 | 3,551.68 | 3,734.84 | 3,851.49 | **14,610.58** |
| 503 | 6,217.80 | 5,940.75 | 6,159.50 | 6,346.29 | **24,664.35** |
| 504 | 1,041.96 | 992.17 | 1,064.90 | 1,079.81 | **4,178.84** |
| 505 | 1,328.41 | 1,331.29 | 1,384.26 | 1,438.04 | **5,482.00** |

| | | | | | |
|---|---|---|---|---|---|
| 506 | 1,753.30 | 1,596.63 | 1,817.17 | 1,757.00 | **6,924.10** |
| 507 | 1,340.35 | 1,300.21 | 1,374.00 | 1,437.77 | **5,452.33** |
| 508 | 197.37 | 178.87 | 198.35 | 188.47 | **763.06** |
| 510 | 1,101.19 | 1,059.18 | 1,107.05 | 1,159.70 | **4,427.10** |
| 511 | 3,071.11 | 2,990.97 | 3,082.60 | 3,134.07 | **12,278.75** |
| 512 | 521.72 | 536.06 | 539.70 | 551.17 | **2,148.65** |
| 513 | 790.48 | 784.74 | 857.59 | 764.56 | **3,197.37** |
| 514 | 440.16 | 416.82 | 447.46 | 452.46 | **1,756.89** |
| 515 | 2,490.90 | 2,407.94 | 2,473.56 | 2,575.91 | **9,948.31** |
| 516 | 195.99 | 171.66 | 176.07 | 196.44 | **740.16** |
| 520 | 3,678.56 | 3,700.73 | 3,787.68 | 3,813.30 | **14,980.26** |
| 521 | 604.91 | 513.21 | 541.38 | 592.22 | **2,251.73** |
| 522 | 5,790.46 | 5,388.10 | 5,620.65 | 6,100.85 | **22,900.06** |
| 523 | 2,749.91 | 2,736.74 | 2,733.88 | 2,808.93 | **11,029.47** |
| 524 | 4,491.36 | 4,501.92 | 4,556.96 | 4,885.84 | **18,436.07** |
| 525 | 981.29 | 955.68 | 1,005.40 | 1,077.44 | **4,019.81** |
| 526 | 1,612.72 | 1,636.74 | 1,687.74 | 1,751.93 | **6,689.13** |
| 527 | 3,299.87 | 3,261.44 | 3,334.13 | 3,363.99 | **13,259.43** |
| 528 | 2,693.02 | 2,614.87 | 2,811.98 | 2,872.25 | **10,992.13** |
| **IA - IOWA - Total** | **56,595.88** | **54,938.91** | **57,441.57** | **59,309.02** | **228,285.38** |

**STATE: ID - IDAHO**

| REGISTRANT ZIP CODE 3 | 1ST QUARTER | 2ND QUARTER | 3RD QUARTER | 4TH QUARTER | TOTAL GRAMS |
|---|---|---|---|---|---|
| 832 | 3,006.93 | 2,805.58 | 3,140.71 | 3,171.31 | **12,124.53** |
| 833 | 2,414.15 | 2,406.56 | 2,496.25 | 2,681.77 | **9,998.73** |
| 834 | 6,236.91 | 5,834.89 | 6,447.99 | 6,835.76 | **25,355.56** |
| 835 | 519.95 | 424.35 | 483.62 | 467.70 | **1,895.62** |
| 836 | 9,428.15 | 9,675.91 | 9,834.97 | 10,793.59 | **39,732.63** |
| 837 | 8,691.98 | 8,669.06 | 8,972.02 | 9,305.93 | **35,638.98** |
| 838 | 2,679.69 | 2,713.93 | 2,837.12 | 2,957.49 | **11,188.23** |
| **ID - IDAHO - Total** | **32,977.75** | **32,530.29** | **34,212.68** | **36,213.55** | **135,934.28** |

**STATE: IL - ILLINOIS**

| REGISTRANT ZIP CODE 3 | 1ST QUARTER | 2ND QUARTER | 3RD QUARTER | 4TH QUARTER | TOTAL GRAMS |
|---|---|---|---|---|---|
| 600 | 36,645.03 | 36,775.53 | 37,321.77 | 38,859.72 | **149,602.05** |
| 601 | 26,676.53 | 26,368.14 | 26,563.54 | 28,196.19 | **107,804.40** |
| 602 | 1,746.85 | 1,591.99 | 1,622.62 | 1,716.97 | **6,678.43** |
| 603 | 1,484.13 | 1,503.43 | 1,458.84 | 1,577.13 | **6,023.53** |
| 604 | 21,606.59 | 21,106.90 | 21,494.09 | 22,328.20 | **86,535.78** |
| 605 | 20,002.61 | 20,091.71 | 20,257.78 | 21,291.23 | **81,643.33** |
| 606 | 43,688.73 | 39,976.59 | 41,008.55 | 43,079.83 | **167,753.71** |
| 607 | 2,632.47 | 2,626.82 | 2,701.93 | 2,808.13 | **10,769.34** |
| 608 | 816.16 | 859.37 | 774.64 | 795.53 | **3,245.70** |
| 609 | 2,311.51 | 2,178.53 | 2,208.29 | 2,428.90 | **9,127.23** |
| 610 | 3,893.77 | 3,838.49 | 3,832.76 | 4,094.80 | **15,659.83** |
| 611 | 4,407.34 | 4,108.29 | 4,178.30 | 4,219.75 | **16,913.68** |
| 612 | 3,288.12 | 3,178.26 | 3,378.53 | 3,369.90 | **13,214.81** |
| 613 | 2,913.74 | 2,853.58 | 2,931.35 | 3,137.13 | **11,835.79** |
| 614 | 2,046.96 | 1,952.55 | 1,939.59 | 2,023.72 | **7,962.82** |
| 615 | 3,504.41 | 3,310.03 | 3,418.53 | 3,673.13 | **13,906.10** |
| 616 | 4,463.42 | 4,231.15 | 4,324.61 | 4,546.84 | **17,566.02** |
| 617 | 4,157.93 | 3,841.02 | 4,170.25 | 4,290.78 | **16,459.98** |
| 618 | 4,129.72 | 3,653.56 | 3,786.68 | 4,120.03 | **15,689.99** |
| 619 | 1,100.73 | 967.52 | 1,117.69 | 1,131.02 | **4,316.97** |
| 620 | 3,881.47 | 3,589.35 | 3,803.32 | 3,880.87 | **15,155.01** |
| 622 | 6,150.40 | 5,834.97 | 6,440.23 | 6,319.84 | **24,745.44** |
| 623 | 2,074.61 | 1,870.42 | 1,925.21 | 1,998.66 | **7,868.90** |
| 624 | 1,298.17 | 1,192.66 | 1,234.06 | 1,246.32 | **4,971.20** |
| 625 | 1,535.19 | 1,555.25 | 1,575.66 | 1,571.69 | **6,237.79** |
| 626 | 1,702.71 | 1,598.21 | 1,631.49 | 1,734.19 | **6,666.61** |
| 627 | 2,504.50 | 2,278.54 | 2,447.69 | 2,464.39 | **9,695.11** |
| 628 | 2,505.15 | 2,506.36 | 2,601.37 | 2,684.26 | **10,297.14** |

| | | | | | |
|---|---|---|---|---|---|
| 629 | 3,920.32 | 3,689.04 | 3,814.28 | 4,043.00 | **15,466.64** |
| **IL - ILLINOIS - Total** | **217,089.27** | **209,128.24** | **213,963.66** | **223,632.15** | **863,813.33** |

**STATE: IN - INDIANA**

| REGISTRANT ZIP CODE 3 | 1ST QUARTER | 2ND QUARTER | 3RD QUARTER | 4TH QUARTER | TOTAL GRAMS |
|---|---|---|---|---|---|
| 460 | 18,717.37 | 17,863.48 | 18,482.63 | 19,231.72 | **74,295.19** |
| 461 | 11,166.61 | 10,879.95 | 11,184.46 | 11,614.83 | **44,845.85** |
| 462 | 18,903.73 | 18,173.73 | 18,471.81 | 19,296.82 | **74,846.09** |
| 463 | 14,644.89 | 14,316.39 | 14,795.67 | 15,694.67 | **59,451.62** |
| 464 | 1,493.65 | 1,427.73 | 1,363.37 | 1,433.83 | **5,718.58** |
| 465 | 18,827.13 | 17,768.36 | 18,582.84 | 19,282.96 | **74,461.29** |
| 466 | 4,873.82 | 4,664.05 | 4,875.97 | 4,929.64 | **19,343.48** |
| 467 | 4,047.03 | 4,092.11 | 4,175.37 | 4,461.80 | **16,776.32** |
| 468 | 7,052.68 | 6,716.97 | 7,183.38 | 7,223.36 | **28,176.39** |
| 469 | 6,218.33 | 6,098.13 | 6,213.80 | 6,390.58 | **24,920.85** |
| 470 | 1,897.82 | 1,933.82 | 2,071.60 | 2,183.84 | **8,087.09** |
| 471 | 6,739.32 | 6,641.39 | 6,751.42 | 7,034.71 | **27,166.84** |
| 472 | 2,793.52 | 2,622.73 | 2,727.78 | 2,804.71 | **10,948.73** |
| 473 | 5,799.62 | 5,604.60 | 5,956.03 | 6,144.40 | **23,504.66** |
| 474 | 4,627.66 | 4,354.07 | 4,429.11 | 4,481.48 | **17,892.32** |
| 475 | 2,734.27 | 2,634.11 | 2,664.99 | 2,831.06 | **10,864.44** |
| 476 | 3,083.38 | 3,090.65 | 3,097.50 | 3,183.36 | **12,454.89** |
| 477 | 6,975.46 | 6,452.94 | 6,644.91 | 6,924.63 | **26,997.94** |
| 478 | 2,542.41 | 2,363.62 | 2,522.27 | 2,476.57 | **9,904.87** |
| 479 | 7,761.94 | 7,324.66 | 7,700.13 | 7,766.43 | **30,553.16** |
| **IN - INDIANA - Total** | **150,900.67** | **145,023.48** | **149,895.04** | **155,391.39** | **601,210.59** |

**STATE: KS - KANSAS**

| REGISTRANT ZIP CODE 3 | 1ST QUARTER | 2ND QUARTER | 3RD QUARTER | 4TH QUARTER | TOTAL GRAMS |
|---|---|---|---|---|---|
| 660 | 16,680.44 | 16,072.58 | 16,898.43 | 17,501.98 | **67,153.43** |
| 661 | 2,391.63 | 2,355.84 | 2,388.09 | 2,422.31 | **9,557.86** |
| 662 | 21,608.04 | 20,840.17 | 21,938.40 | 22,638.29 | **87,024.90** |
| 664 | 1,071.02 | 779.73 | 1,053.02 | 891.95 | **3,795.73** |
| 665 | 1,835.34 | 1,648.59 | 1,819.38 | 1,980.87 | **7,284.17** |
| 666 | 5,415.53 | 5,621.34 | 5,788.57 | 5,962.69 | **22,788.13** |
| 667 | 2,000.21 | 1,976.65 | 2,037.01 | 2,149.66 | **8,163.53** |
| 668 | 675.05 | 683.16 | 696.51 | 783.94 | **2,838.66** |
| 669 | 106.03 | 132.15 | 139.74 | 154.81 | **532.73** |
| 670 | 2,749.60 | 2,935.60 | 2,890.24 | 3,119.78 | **11,695.22** |
| 671 | 1,089.39 | 1,062.48 | 1,115.12 | 1,177.98 | **4,444.97** |
| 672 | 8,273.80 | 8,329.50 | 8,417.58 | 8,976.08 | **33,996.96** |
| 673 | 934.01 | 923.28 | 947.61 | 1,018.51 | **3,823.41** |
| 674 | 1,400.42 | 1,485.16 | 1,494.07 | 1,616.07 | **5,995.73** |
| 675 | 1,353.70 | 1,333.57 | 1,385.13 | 1,375.69 | **5,448.08** |
| 676 | 906.04 | 909.65 | 959.61 | 1,029.06 | **3,804.37** |
| 677 | 414.65 | 397.71 | 409.84 | 436.50 | **1,658.70** |
| 678 | 698.36 | 761.71 | 654.39 | 734.64 | **2,849.10** |
| 679 | 177.01 | 200.88 | 235.39 | 205.37 | **818.66** |
| **KS - KANSAS - Total** | **69,780.28** | **68,449.76** | **71,268.15** | **74,176.17** | **283,674.35** |

**STATE: KY - KENTUCKY**

| REGISTRANT ZIP CODE 3 | 1ST QUARTER | 2ND QUARTER | 3RD QUARTER | 4TH QUARTER | TOTAL GRAMS |
|---|---|---|---|---|---|
| 400 | 4,514.90 | 4,528.85 | 4,735.20 | 4,872.74 | **18,651.70** |
| 401 | 1,881.70 | 1,643.47 | 1,712.80 | 1,766.95 | **7,004.91** |
| 402 | 19,162.40 | 18,453.82 | 18,492.61 | 19,332.21 | **75,441.03** |
| 403 | 5,031.08 | 4,824.54 | 5,030.27 | 5,266.16 | **20,152.04** |
| 404 | 2,218.51 | 1,984.95 | 2,110.69 | 2,334.50 | **8,648.65** |
| 405 | 8,680.48 | 8,234.53 | 8,581.29 | 9,186.42 | **34,682.71** |
| 406 | 866.52 | 854.69 | 853.29 | 904.48 | **3,478.98** |
| 407 | 809.93 | 713.38 | 777.07 | 853.29 | **3,153.66** |
| 408 | 152.89 | 135.40 | 139.56 | 140.66 | **568.51** |
| 409 | 246.95 | 273.00 | 243.08 | 282.87 | **1,045.90** |
| 410 | 11,077.03 | 10,814.97 | 11,022.20 | 11,529.45 | **44,443.66** |

| | | | | | |
|---|---|---|---|---|---|
| 411 | 1,622.32 | 1,314.04 | 1,487.38 | 1,515.91 | **5,939.64** |
| 412 | 435.10 | 363.63 | 391.10 | 403.55 | **1,593.38** |
| 413 | 185.50 | 143.61 | 141.79 | 161.05 | **631.96** |
| 414 | 148.94 | 106.58 | 133.88 | 121.58 | **510.98** |
| 415 | 666.93 | 593.37 | 583.44 | 650.38 | **2,494.13** |
| 416 | 397.98 | 364.14 | 445.81 | 393.61 | **1,601.54** |
| 417 | 396.66 | 338.03 | 369.33 | 367.12 | **1,471.15** |
| 418 | 87.95 | 95.13 | 77.86 | 94.76 | **355.70** |
| 420 | 4,936.72 | 4,873.94 | 4,905.22 | 4,979.78 | **19,695.66** |
| 421 | 3,281.58 | 3,037.57 | 3,217.93 | 3,413.04 | **12,950.13** |
| 422 | 2,192.48 | 1,968.41 | 2,073.13 | 2,031.10 | **8,265.13** |
| 423 | 3,655.20 | 3,222.22 | 3,326.20 | 3,479.66 | **13,683.28** |
| 424 | 3,999.65 | 3,599.82 | 3,938.89 | 3,937.57 | **15,475.94** |
| 425 | 429.56 | 370.90 | 401.91 | 470.34 | **1,672.70** |
| 426 | 328.74 | 232.27 | 193.26 | 231.53 | **985.80** |
| 427 | 3,192.23 | 2,867.57 | 3,101.56 | 3,031.72 | **12,193.08** |
| **KY - KENTUCKY - Total** | **80,599.95** | **75,952.84** | **78,486.75** | **81,752.42** | **316,791.97** |

**STATE: LA - LOUISIANA**

| REGISTRANT ZIP CODE 3 | 1ST QUARTER | 2ND QUARTER | 3RD QUARTER | 4TH QUARTER | TOTAL GRAMS |
|---|---|---|---|---|---|
| 700 | 20,599.29 | 18,954.12 | 19,512.21 | 20,570.42 | **79,636.04** |
| 701 | 12,680.35 | 11,582.39 | 12,151.44 | 13,232.93 | **49,647.10** |
| 703 | 6,838.58 | 6,176.72 | 6,789.59 | 6,913.75 | **26,718.63** |
| 704 | 22,240.88 | 21,581.83 | 22,151.53 | 23,516.21 | **89,490.45** |
| 705 | 36,927.49 | 35,467.94 | 37,570.26 | 39,161.28 | **149,126.97** |
| 706 | 14,322.18 | 13,855.57 | 12,895.61 | 14,046.78 | **55,120.15** |
| 707 | 22,964.64 | 22,662.13 | 23,576.37 | 24,659.11 | **93,862.25** |
| 708 | 17,430.34 | 16,506.35 | 17,064.51 | 18,132.52 | **69,133.72** |
| 710 | 2,242.11 | 2,245.13 | 2,309.48 | 2,571.16 | **9,367.88** |
| 711 | 8,566.13 | 7,906.55 | 8,340.28 | 8,843.12 | **33,656.08** |
| 712 | 8,088.69 | 7,662.53 | 8,170.67 | 8,648.71 | **32,570.60** |
| 713 | 6,242.24 | 6,044.71 | 6,087.46 | 6,411.33 | **24,785.75** |
| 714 | 2,569.93 | 2,455.98 | 2,600.34 | 2,814.12 | **10,440.36** |
| **LA - LOUISIANA - Total** | **181,712.87** | **173,101.92** | **179,219.74** | **189,521.44** | **723,555.98** |

**STATE: MA - MASSACHUSETTS**

| REGISTRANT ZIP CODE 3 | 1ST QUARTER | 2ND QUARTER | 3RD QUARTER | 4TH QUARTER | TOTAL GRAMS |
|---|---|---|---|---|---|
| 010 | 9,432.13 | 8,706.28 | 9,214.00 | 9,640.08 | **36,992.49** |
| 011 | 2,561.57 | 2,260.68 | 2,288.68 | 2,498.08 | **9,609.00** |
| 012 | 2,659.76 | 2,580.45 | 2,684.66 | 2,769.94 | **10,694.81** |
| 013 | 2,751.33 | 2,783.21 | 2,850.02 | 2,819.57 | **11,204.14** |
| 014 | 5,932.47 | 5,762.73 | 5,983.47 | 6,122.72 | **23,801.39** |
| 015 | 8,485.73 | 8,121.32 | 8,449.39 | 8,916.35 | **33,972.79** |
| 016 | 5,656.56 | 5,506.75 | 5,649.17 | 5,957.16 | **22,769.64** |
| 017 | 12,098.00 | 11,431.81 | 11,803.11 | 12,083.17 | **47,416.09** |
| 018 | 21,810.00 | 21,071.99 | 21,352.84 | 21,885.31 | **86,120.15** |
| 019 | 18,761.80 | 18,482.55 | 18,717.61 | 19,977.32 | **75,939.28** |
| 020 | 15,106.62 | 14,816.12 | 15,018.89 | 15,684.58 | **60,626.22** |
| 021 | 36,994.14 | 33,612.22 | 34,527.95 | 35,814.47 | **140,948.78** |
| 022 | 1,739.83 | 1,441.26 | 1,562.10 | 1,568.51 | **6,311.70** |
| 023 | 14,491.97 | 14,314.36 | 14,674.56 | 15,222.39 | **58,703.27** |
| 024 | 11,486.88 | 10,914.33 | 10,962.99 | 11,643.05 | **45,007.24** |
| 025 | 4,287.07 | 4,924.44 | 5,213.96 | 4,883.49 | **19,308.96** |
| 026 | 5,684.50 | 6,211.09 | 6,989.22 | 6,289.35 | **25,174.16** |
| 027 | 14,127.98 | 13,412.33 | 14,041.61 | 14,384.90 | **55,966.82** |
| **MA - MASSACHUSETTS - Total** | **194,068.35** | **186,353.92** | **191,984.25** | **198,160.44** | **770,566.96** |

**STATE: MD - MARYLAND**

| REGISTRANT ZIP CODE 3 | 1ST QUARTER | 2ND QUARTER | 3RD QUARTER | 4TH QUARTER | TOTAL GRAMS |
|---|---|---|---|---|---|
| 206 | 5,044.05 | 4,823.38 | 4,977.73 | 5,258.29 | **20,103.44** |
| 207 | 6,722.56 | 6,393.06 | 6,625.26 | 6,188.64 | **25,929.52** |
| 208 | 15,436.88 | 15,253.33 | 15,053.99 | 15,062.65 | **60,806.84** |
| 209 | 3,264.18 | 2,846.73 | 2,943.31 | 3,091.26 | **12,145.48** |

| | | | | | |
|---|---|---|---|---|---|
| 210 | 20,252.32 | 18,576.64 | 19,456.67 | 20,353.79 | **78,639.42** |
| 211 | 9,438.60 | 9,222.88 | 9,239.12 | 9,711.18 | **37,611.77** |
| 212 | 20,284.50 | 18,612.69 | 19,075.83 | 20,099.15 | **78,072.17** |
| 214 | 3,445.72 | 3,118.00 | 3,277.45 | 3,160.23 | **13,001.40** |
| 215 | 2,175.44 | 1,895.40 | 1,941.99 | 2,070.31 | **8,083.15** |
| 216 | 3,495.12 | 3,660.66 | 3,547.08 | 3,834.63 | **14,537.49** |
| 217 | 12,373.67 | 11,776.99 | 12,393.72 | 12,768.15 | **49,312.54** |
| 218 | 2,358.30 | 2,339.36 | 2,535.39 | 2,563.48 | **9,796.53** |
| 219 | 2,666.50 | 2,692.54 | 2,628.89 | 2,693.37 | **10,681.30** |
| **MD - MARYLAND - Total** | **106,957.83** | **101,211.66** | **103,696.43** | **106,855.15** | **418,721.06** |

**STATE: ME - MAINE**

| REGISTRANT ZIP CODE 3 | 1ST QUARTER | 2ND QUARTER | 3RD QUARTER | 4TH QUARTER | TOTAL GRAMS |
|---|---|---|---|---|---|
| 039 | 1,098.98 | 1,062.76 | 1,181.18 | 959.82 | **4,302.75** |
| 040 | 10,017.95 | 9,440.90 | 10,412.04 | 10,820.34 | **40,691.23** |
| 041 | 4,165.95 | 4,057.84 | 4,191.82 | 4,174.06 | **16,589.67** |
| 042 | 2,768.03 | 2,316.46 | 2,632.65 | 2,778.51 | **10,495.65** |
| 043 | 1,951.39 | 1,849.76 | 2,038.18 | 2,033.72 | **7,873.06** |
| 044 | 2,506.72 | 2,100.37 | 2,402.91 | 2,456.19 | **9,466.19** |
| 045 | 928.51 | 1,008.81 | 1,045.98 | 1,000.46 | **3,983.76** |
| 046 | 830.85 | 783.15 | 912.90 | 889.95 | **3,416.85** |
| 047 | 641.08 | 484.28 | 500.15 | 615.43 | **2,240.93** |
| 048 | 739.94 | 695.31 | 776.97 | 719.03 | **2,931.26** |
| 049 | 2,269.48 | 1,956.59 | 2,126.59 | 2,245.21 | **8,597.88** |
| **ME - MAINE - Total** | **27,918.87** | **25,756.25** | **28,221.38** | **28,692.72** | **110,589.22** |

**STATE: MI - MICHIGAN**

| REGISTRANT ZIP CODE 3 | 1ST QUARTER | 2ND QUARTER | 3RD QUARTER | 4TH QUARTER | TOTAL GRAMS |
|---|---|---|---|---|---|
| 480 | 42,023.90 | 40,527.17 | 41,636.37 | 43,111.16 | **167,298.60** |
| 481 | 44,875.56 | 42,569.01 | 44,749.03 | 45,754.38 | **177,947.99** |
| 482 | 6,863.12 | 6,644.87 | 7,137.38 | 7,112.72 | **27,758.09** |
| 483 | 35,611.87 | 34,253.06 | 35,534.17 | 36,752.52 | **142,151.62** |
| 484 | 9,300.95 | 9,121.34 | 9,530.12 | 9,893.95 | **37,846.36** |
| 485 | 2,751.72 | 2,797.70 | 2,907.24 | 2,950.28 | **11,406.93** |
| 486 | 5,304.77 | 5,206.20 | 5,318.79 | 5,394.17 | **21,223.93** |
| 487 | 2,977.07 | 2,954.87 | 3,018.22 | 3,083.46 | **12,033.62** |
| 488 | 16,306.27 | 14,805.91 | 15,649.88 | 16,437.42 | **63,199.48** |
| 489 | 4,413.26 | 4,366.19 | 4,320.28 | 4,500.38 | **17,600.11** |
| 490 | 18,361.20 | 17,720.25 | 17,839.04 | 18,572.38 | **72,492.87** |
| 491 | 2,347.45 | 2,395.44 | 2,401.08 | 2,502.17 | **9,646.14** |
| 492 | 4,521.14 | 4,178.69 | 4,125.30 | 4,524.48 | **17,349.61** |
| 493 | 7,705.87 | 7,382.58 | 7,712.90 | 7,875.58 | **30,676.92** |
| 494 | 19,527.74 | 19,186.62 | 19,464.04 | 20,332.77 | **78,511.18** |
| 495 | 19,953.41 | 19,310.58 | 20,161.21 | 20,711.60 | **80,136.80** |
| 496 | 6,176.94 | 6,365.26 | 6,400.07 | 6,455.70 | **25,397.97** |
| 497 | 3,860.22 | 3,947.04 | 4,171.35 | 4,182.22 | **16,160.83** |
| 498 | 3,437.73 | 3,429.26 | 3,791.95 | 3,605.30 | **14,264.24** |
| 499 | 1,561.06 | 1,448.21 | 1,603.35 | 1,664.88 | **6,277.51** |
| **MI - MICHIGAN - Total** | **257,881.27** | **248,610.24** | **257,471.78** | **265,417.53** | **1,029,380.82** |

**STATE: MN - MINNESOTA**

| REGISTRANT ZIP CODE 3 | 1ST QUARTER | 2ND QUARTER | 3RD QUARTER | 4TH QUARTER | TOTAL GRAMS |
|---|---|---|---|---|---|
| 550 | 13,924.66 | 14,082.78 | 14,464.47 | 14,834.96 | **57,306.87** |
| 551 | 18,978.00 | 18,946.51 | 18,776.88 | 20,333.68 | **77,035.08** |
| 553 | 24,715.74 | 24,536.85 | 25,164.50 | 26,478.56 | **100,895.65** |
| 554 | 28,823.48 | 28,478.91 | 28,524.65 | 29,684.19 | **115,511.23** |
| 556 | 140.04 | 146.71 | 150.39 | 143.95 | **581.09** |
| 557 | 3,646.87 | 3,864.83 | 4,018.81 | 4,254.92 | **15,785.44** |
| 558 | 2,133.74 | 2,105.61 | 2,141.46 | 2,286.97 | **8,667.78** |
| 559 | 5,640.00 | 5,413.32 | 5,728.77 | 6,008.24 | **22,790.32** |
| 560 | 5,669.62 | 5,555.85 | 5,524.05 | 6,003.66 | **22,753.18** |
| 561 | 920.58 | 867.11 | 863.49 | 953.19 | **3,604.37** |
| 562 | 1,861.53 | 1,710.48 | 1,700.79 | 1,862.48 | **7,135.28** |

| | | | | | |
|---|---|---|---|---|---|
| 563 | 4,586.48 | 4,596.81 | 4,709.49 | 4,840.09 | **18,732.87** |
| 564 | 2,466.33 | 2,501.07 | 2,616.73 | 2,620.67 | **10,204.80** |
| 565 | 3,108.50 | 3,124.98 | 3,180.02 | 3,329.64 | **12,743.13** |
| 566 | 1,674.24 | 1,543.94 | 1,677.31 | 1,597.86 | **6,493.35** |
| 567 | 930.19 | 887.12 | 881.15 | 988.44 | **3,686.91** |
| **MN - MINNESOTA - Total** | **119,220.03** | **118,362.87** | **120,122.93** | **126,221.49** | **483,927.32** |

**STATE: MO - MISSOURI**

| REGISTRANT ZIP CODE 3 | 1ST QUARTER | 2ND QUARTER | 3RD QUARTER | 4TH QUARTER | TOTAL GRAMS |
|---|---|---|---|---|---|
| 630 | 20,677.65 | 20,226.58 | 20,480.07 | 21,379.94 | **82,764.24** |
| 631 | 48,879.29 | 51,972.63 | 62,375.11 | 55,128.82 | **218,355.84** |
| 633 | 16,280.59 | 16,228.47 | 16,618.08 | 17,174.18 | **66,301.32** |
| 634 | 1,236.29 | 1,214.45 | 1,268.07 | 1,298.43 | **5,017.24** |
| 635 | 1,359.48 | 1,324.71 | 1,431.50 | 1,484.80 | **5,600.49** |
| 636 | 2,049.14 | 1,940.07 | 2,086.00 | 2,146.92 | **8,222.13** |
| 637 | 2,157.10 | 2,148.65 | 2,351.31 | 2,260.06 | **8,917.13** |
| 638 | 1,781.94 | 1,755.78 | 1,822.42 | 1,851.59 | **7,211.73** |
| 639 | 565.03 | 577.92 | 603.57 | 711.60 | **2,458.11** |
| 640 | 16,611.11 | 16,537.31 | 17,397.09 | 17,862.23 | **68,407.75** |
| 641 | 16,655.36 | 15,944.59 | 16,923.48 | 17,628.44 | **67,151.87** |
| 644 | 676.28 | 646.02 | 740.93 | 738.41 | **2,801.63** |
| 645 | 1,263.79 | 1,220.11 | 1,367.11 | 1,294.55 | **5,145.57** |
| 646 | 424.32 | 400.15 | 414.16 | 419.72 | **1,658.35** |
| 647 | 1,184.64 | 1,130.91 | 1,145.85 | 1,290.97 | **4,752.38** |
| 648 | 1,775.97 | 1,763.96 | 1,774.26 | 1,853.32 | **7,167.51** |
| 650 | 1,177.90 | 1,242.91 | 1,244.39 | 1,293.48 | **4,958.68** |
| 651 | 1,238.27 | 1,259.53 | 1,345.82 | 1,398.44 | **5,242.06** |
| 652 | 5,572.89 | 5,290.98 | 5,767.01 | 6,056.51 | **22,687.39** |
| 653 | 1,081.03 | 1,043.23 | 1,163.03 | 1,151.96 | **4,439.26** |
| 654 | 875.66 | 825.31 | 921.83 | 955.37 | **3,578.17** |
| 655 | 654.22 | 629.72 | 601.83 | 645.38 | **2,531.15** |
| 656 | 1,469.40 | 1,461.56 | 1,537.07 | 1,556.90 | **6,024.93** |
| 657 | 2,168.72 | 2,152.75 | 2,207.62 | 2,365.90 | **8,894.99** |
| 658 | 3,969.66 | 3,979.48 | 3,968.27 | 4,443.58 | **16,360.99** |
| **MO - MISSOURI - Total** | **151,785.72** | **152,917.79** | **167,555.88** | **164,391.48** | **636,650.88** |

**STATE: MS - MISSISSIPPI**

| REGISTRANT ZIP CODE 3 | 1ST QUARTER | 2ND QUARTER | 3RD QUARTER | 4TH QUARTER | TOTAL GRAMS |
|---|---|---|---|---|---|
| 386 | 10,173.73 | 9,767.42 | 10,373.84 | 11,067.64 | **41,382.64** |
| 387 | 1,572.43 | 1,429.57 | 1,540.27 | 1,572.86 | **6,115.13** |
| 388 | 5,446.99 | 5,417.81 | 5,441.70 | 5,898.93 | **22,205.42** |
| 389 | 1,594.27 | 1,459.04 | 1,647.27 | 1,559.69 | **6,260.27** |
| 390 | 5,281.65 | 5,267.73 | 5,447.64 | 5,738.75 | **21,735.78** |
| 391 | 6,557.62 | 6,518.60 | 6,813.28 | 7,097.47 | **26,986.97** |
| 392 | 5,184.97 | 5,044.61 | 5,086.90 | 5,594.36 | **20,910.84** |
| 393 | 4,058.95 | 3,847.26 | 4,062.77 | 4,169.66 | **16,138.64** |
| 394 | 9,287.39 | 8,968.48 | 9,496.01 | 9,716.86 | **37,468.75** |
| 395 | 9,838.76 | 9,183.89 | 9,637.93 | 9,927.77 | **38,588.35** |
| 396 | 2,841.51 | 2,660.90 | 2,837.16 | 3,023.88 | **11,363.46** |
| 397 | 3,589.19 | 3,142.76 | 3,389.04 | 3,499.19 | **13,620.18** |
| **MS - MISSISSIPPI - Total** | **65,427.47** | **62,708.08** | **65,773.81** | **68,867.07** | **262,776.43** |

**STATE: MT - MONTANA**

| REGISTRANT ZIP CODE 3 | 1ST QUARTER | 2ND QUARTER | 3RD QUARTER | 4TH QUARTER | TOTAL GRAMS |
|---|---|---|---|---|---|
| 590 | 791.01 | 808.86 | 828.25 | 850.29 | **3,278.41** |
| 591 | 2,885.65 | 2,532.95 | 2,779.52 | 2,747.70 | **10,945.81** |
| 592 | 405.43 | 394.11 | 342.43 | 383.98 | **1,525.95** |
| 593 | 451.31 | 435.83 | 430.08 | 432.67 | **1,749.89** |
| 594 | 1,271.67 | 1,327.79 | 1,306.41 | 1,347.29 | **5,253.16** |
| 595 | 235.69 | 191.67 | 233.36 | 263.70 | **924.42** |
| 596 | 1,248.27 | 1,324.75 | 1,405.86 | 1,445.38 | **5,424.26** |
| 597 | 4,109.01 | 4,099.45 | 4,409.89 | 4,533.97 | **17,152.33** |
| 598 | 4,012.47 | 4,039.52 | 4,049.28 | 4,263.39 | **16,364.66** |

| | | | | | |
|---|---|---|---|---|---|
| 599 | 1,557.99 | 1,625.30 | 1,657.56 | 1,765.88 | **6,606.73** |
| **MT - MONTANA - Total** | **16,968.49** | **16,780.24** | **17,442.63** | **18,034.25** | **69,225.61** |

**STATE: NC - NORTH CAROLINA**

| REGISTRANT ZIP CODE 3 | 1ST QUARTER | 2ND QUARTER | 3RD QUARTER | 4TH QUARTER | TOTAL GRAMS |
|---|---|---|---|---|---|
| 270 | 4,829.09 | 4,493.55 | 4,656.43 | 5,023.53 | **19,002.60** |
| 271 | 5,981.43 | 5,596.55 | 5,736.74 | 6,133.20 | **23,447.93** |
| 272 | 12,769.54 | 12,312.80 | 12,904.22 | 13,166.70 | **51,153.27** |
| 273 | 6,931.78 | 6,695.02 | 6,992.19 | 7,229.24 | **27,848.24** |
| 274 | 8,709.59 | 8,122.07 | 8,403.29 | 8,836.46 | **34,071.42** |
| 275 | 28,309.66 | 27,291.86 | 28,294.66 | 30,236.52 | **114,132.70** |
| 276 | 17,461.48 | 15,532.49 | 16,498.06 | 17,320.69 | **66,812.72** |
| 277 | 6,979.49 | 6,235.19 | 6,574.50 | 6,998.98 | **26,788.16** |
| 278 | 10,533.59 | 9,940.27 | 10,497.16 | 10,860.39 | **41,831.41** |
| 279 | 3,692.59 | 3,954.20 | 4,325.23 | 4,364.83 | **16,336.86** |
| 280 | 20,650.76 | 20,239.32 | 20,918.56 | 21,948.47 | **83,757.12** |
| 281 | 14,282.53 | 13,779.87 | 14,345.54 | 15,217.86 | **57,625.80** |
| 282 | 22,847.81 | 21,606.99 | 22,544.59 | 23,817.97 | **90,817.36** |
| 283 | 16,911.53 | 16,226.74 | 17,337.18 | 16,831.61 | **67,307.06** |
| 284 | 20,428.62 | 20,790.04 | 21,533.35 | 21,760.84 | **84,512.85** |
| 285 | 13,019.74 | 12,234.00 | 12,993.19 | 12,929.17 | **51,176.10** |
| 286 | 9,463.96 | 9,110.95 | 9,254.39 | 9,566.12 | **37,395.42** |
| 287 | 8,505.62 | 8,224.78 | 8,751.01 | 9,266.44 | **34,747.85** |
| 288 | 5,035.24 | 5,139.23 | 5,084.42 | 5,296.99 | **20,555.88** |
| 289 | 481.25 | 483.16 | 539.44 | 523.37 | **2,027.22** |
| **NC - NORTH CAROLINA - Total** | **237,825.31** | **228,009.09** | **238,184.17** | **247,329.39** | **951,347.95** |

**STATE: ND - NORTH DAKOTA**

| REGISTRANT ZIP CODE 3 | 1ST QUARTER | 2ND QUARTER | 3RD QUARTER | 4TH QUARTER | TOTAL GRAMS |
|---|---|---|---|---|---|
| 580 | 888.43 | 891.76 | 900.63 | 951.45 | **3,632.27** |
| 581 | 2,590.78 | 2,519.86 | 2,418.49 | 2,903.56 | **10,432.69** |
| 582 | 1,580.93 | 1,518.32 | 1,458.89 | 1,714.44 | **6,272.57** |
| 583 | 363.36 | 360.18 | 407.97 | 389.79 | **1,521.31** |
| 584 | 347.30 | 330.93 | 329.25 | 368.43 | **1,375.91** |
| 585 | 1,696.55 | 1,617.34 | 1,600.10 | 1,770.17 | **6,684.16** |
| 586 | 537.86 | 513.54 | 511.22 | 488.68 | **2,051.30** |
| 587 | 1,580.21 | 1,396.36 | 1,491.26 | 1,529.42 | **5,997.25** |
| 588 | 975.81 | 837.64 | 839.39 | 850.35 | **3,503.19** |
| **ND - NORTH DAKOTA - Total** | **10,561.24** | **9,985.92** | **9,957.21** | **10,966.29** | **41,470.66** |

**STATE: NE - NEBRASKA**

| REGISTRANT ZIP CODE 3 | 1ST QUARTER | 2ND QUARTER | 3RD QUARTER | 4TH QUARTER | TOTAL GRAMS |
|---|---|---|---|---|---|
| 680 | 2,746.26 | 2,772.23 | 2,892.80 | 3,152.83 | **11,564.11** |
| 681 | 15,220.48 | 15,059.86 | 15,111.20 | 16,254.09 | **61,645.62** |
| 683 | 639.97 | 658.27 | 617.32 | 714.67 | **2,630.23** |
| 684 | 512.77 | 500.03 | 530.48 | 549.23 | **2,092.50** |
| 685 | 6,251.82 | 5,953.41 | 6,396.90 | 6,664.70 | **25,266.83** |
| 686 | 540.98 | 541.66 | 547.51 | 553.38 | **2,183.54** |
| 687 | 1,194.99 | 1,137.06 | 1,151.17 | 1,213.45 | **4,696.67** |
| 688 | 1,964.22 | 1,840.30 | 1,989.98 | 2,149.65 | **7,944.15** |
| 689 | 551.74 | 510.32 | 502.12 | 570.36 | **2,134.54** |
| 690 | 204.06 | 195.53 | 226.91 | 231.83 | **858.33** |
| 691 | 780.20 | 736.51 | 713.30 | 841.24 | **3,071.26** |
| 692 | 18.46 | 16.27 | 21.58 | 31.75 | **88.06** |
| 693 | 649.42 | 565.55 | 614.02 | 581.98 | **2,410.97** |
| **NE - NEBRASKA - Total** | **31,275.37** | **30,487.00** | **31,315.27** | **33,509.16** | **126,586.80** |

**STATE: NH - NEW HAMPSHIRE**

| REGISTRANT ZIP CODE 3 | 1ST QUARTER | 2ND QUARTER | 3RD QUARTER | 4TH QUARTER | TOTAL GRAMS |
|---|---|---|---|---|---|
| 030 | 13,421.78 | 13,404.87 | 13,700.94 | 13,833.23 | **54,360.81** |
| 031 | 6,960.52 | 6,943.78 | 6,996.57 | 7,408.58 | **28,309.45** |
| 032 | 3,197.14 | 3,567.28 | 3,588.04 | 3,848.39 | **14,200.86** |
| 033 | 2,362.96 | 2,354.69 | 2,409.65 | 2,556.69 | **9,683.99** |
| 034 | 1,742.89 | 1,857.48 | 1,874.34 | 2,032.69 | **7,507.40** |

| | | | | | |
|---|---|---|---|---|---|
| 035 | 900.25 | 650.72 | 817.54 | 837.33 | **3,205.85** |
| 036 | 129.33 | 123.45 | 118.44 | 122.73 | **493.95** |
| 037 | 1,946.02 | 1,978.20 | 2,032.24 | 2,134.11 | **8,090.57** |
| 038 | 13,355.01 | 13,663.40 | 13,919.75 | 14,361.12 | **55,299.27** |
| **NH - NEW HAMPSHIRE - Total** | **44,015.90** | **44,543.88** | **45,457.51** | **47,134.87** | **181,152.17** |

**STATE: NJ - NEW JERSEY**

| REGISTRANT ZIP CODE 3 | 1ST QUARTER | 2ND QUARTER | 3RD QUARTER | 4TH QUARTER | TOTAL GRAMS |
|---|---|---|---|---|---|
| 070 | 20,365.32 | 19,473.63 | 19,796.05 | 21,488.45 | **81,123.46** |
| 071 | 1,631.75 | 1,469.31 | 1,488.99 | 1,589.58 | **6,179.62** |
| 072 | 739.61 | 651.19 | 654.24 | 749.34 | **2,794.37** |
| 073 | 2,321.70 | 2,250.13 | 2,459.14 | 2,411.83 | **9,442.79** |
| 074 | 7,337.66 | 7,116.14 | 7,369.14 | 7,991.82 | **29,814.75** |
| 075 | 939.89 | 835.71 | 872.00 | 957.02 | **3,604.61** |
| 076 | 6,852.15 | 6,583.24 | 6,786.35 | 7,263.50 | **27,485.24** |
| 077 | 10,321.22 | 10,053.67 | 10,769.16 | 11,285.35 | **42,429.40** |
| 078 | 6,563.08 | 6,156.75 | 6,525.07 | 6,919.61 | **26,164.50** |
| 079 | 5,102.40 | 5,066.02 | 4,995.22 | 5,281.40 | **20,445.03** |
| 080 | 26,247.77 | 24,949.86 | 25,210.01 | 26,933.21 | **103,340.86** |
| 081 | 2,136.62 | 2,024.20 | 1,943.75 | 2,091.99 | **8,196.56** |
| 082 | 4,297.19 | 4,281.24 | 4,952.13 | 4,752.68 | **18,283.24** |
| 083 | 2,900.81 | 2,671.13 | 2,692.21 | 2,858.36 | **11,122.51** |
| 084 | 548.12 | 567.80 | 702.60 | 641.41 | **2,459.93** |
| 085 | 3,982.73 | 3,794.32 | 3,787.95 | 4,098.16 | **15,663.16** |
| 086 | 3,066.75 | 2,758.37 | 2,843.07 | 2,962.93 | **11,631.11** |
| 087 | 7,106.82 | 7,017.20 | 7,445.25 | 7,603.48 | **29,172.75** |
| 088 | 10,295.49 | 10,139.09 | 10,136.13 | 10,633.37 | **41,204.09** |
| 089 | 1,053.09 | 940.50 | 943.65 | 989.16 | **3,926.41** |
| **NJ - NEW JERSEY - Total** | **123,810.17** | **118,799.48** | **122,372.09** | **129,502.66** | **494,484.40** |

**STATE: NM - NEW MEXICO**

| REGISTRANT ZIP CODE 3 | 1ST QUARTER | 2ND QUARTER | 3RD QUARTER | 4TH QUARTER | TOTAL GRAMS |
|---|---|---|---|---|---|
| 870 | 907.02 | 531.36 | 697.93 | 725.30 | **2,861.61** |
| 871 | 7,839.31 | 5,786.03 | 7,204.11 | 7,394.73 | **28,224.19** |
| 873 | 137.22 | 77.69 | 119.02 | 107.53 | **441.46** |
| 874 | 785.05 | 651.63 | 728.36 | 749.28 | **2,914.32** |
| 875 | 2,156.88 | 1,896.87 | 2,092.59 | 2,149.25 | **8,295.58** |
| 877 | 229.21 | 181.93 | 188.05 | 161.31 | **760.50** |
| 878 | 50.20 | 48.87 | 50.18 | 58.70 | **207.94** |
| 879 | 60.98 | 65.76 | 76.76 | 65.29 | **268.80** |
| 880 | 2,491.88 | 1,895.12 | 2,417.08 | 2,397.84 | **9,201.93** |
| 881 | 384.03 | 377.34 | 383.00 | 401.90 | **1,546.27** |
| 882 | 2,439.46 | 1,940.33 | 2,174.35 | 2,193.62 | **8,747.76** |
| 883 | 1,063.51 | 878.38 | 992.45 | 937.20 | **3,871.55** |
| 884 | 44.28 | 57.04 | 46.45 | 61.47 | **209.25** |
| **NM - NEW MEXICO - Total** | **18,589.03** | **14,388.35** | **17,170.33** | **17,403.44** | **67,551.14** |

**STATE: NV - NEVADA**

| REGISTRANT ZIP CODE 3 | 1ST QUARTER | 2ND QUARTER | 3RD QUARTER | 4TH QUARTER | TOTAL GRAMS |
|---|---|---|---|---|---|
| 890 | 7,352.40 | 6,745.51 | 7,198.68 | 7,690.62 | **28,987.22** |
| 891 | 17,581.40 | 15,567.31 | 17,019.84 | 17,131.47 | **67,300.02** |
| 893 | 55.54 | 52.96 | 43.78 | 53.93 | **206.21** |
| 894 | 1,958.04 | 1,967.13 | 2,067.45 | 2,205.76 | **8,198.38** |
| 895 | 3,257.05 | 3,288.27 | 3,401.54 | 3,637.61 | **13,584.47** |
| 897 | 623.38 | 544.76 | 641.07 | 634.98 | **2,444.18** |
| 898 | 489.68 | 510.44 | 517.69 | 578.96 | **2,096.77** |
| **NV - NEVADA - Total** | **31,317.49** | **28,676.38** | **30,890.05** | **31,933.33** | **122,817.25** |

**STATE: NY - NEW YORK**

| REGISTRANT ZIP CODE 3 | 1ST QUARTER | 2ND QUARTER | 3RD QUARTER | 4TH QUARTER | TOTAL GRAMS |
|---|---|---|---|---|---|
| 100 | 55,315.19 | 35,674.43 | 39,928.65 | 44,763.14 | **175,681.41** |
| 101 | 2,416.67 | 1,280.59 | 1,247.76 | 1,437.81 | **6,382.82** |
| 102 | 384.81 | 232.36 | 219.30 | 275.92 | **1,112.39** |
| 103 | 4,171.48 | 4,554.94 | 4,550.55 | 4,600.60 | **17,877.56** |

| | | | | | |
|---|---|---|---|---|---|
| 104 | 4,347.22 | 4,052.19 | 3,765.31 | 4,101.06 | **16,265.78** |
| 105 | 10,957.59 | 10,983.69 | 10,944.05 | 11,492.03 | **44,377.36** |
| 106 | 1,431.55 | 1,193.58 | 1,257.92 | 1,322.60 | **5,205.65** |
| 107 | 2,062.62 | 2,247.44 | 2,374.14 | 2,388.08 | **9,072.28** |
| 108 | 868.94 | 819.17 | 879.96 | 861.57 | **3,429.64** |
| 109 | 7,582.23 | 7,178.75 | 7,640.40 | 8,018.37 | **30,419.75** |
| 110 | 3,040.06 | 2,908.24 | 2,993.89 | 3,159.08 | **12,101.26** |
| 111 | 3,306.19 | 3,025.38 | 3,169.39 | 3,216.80 | **12,717.77** |
| 112 | 18,171.40 | 16,632.50 | 17,758.49 | 18,747.93 | **71,310.31** |
| 113 | 6,266.09 | 5,752.80 | 5,942.08 | 6,345.98 | **24,306.95** |
| 114 | 1,548.82 | 1,404.81 | 1,372.60 | 1,569.17 | **5,895.40** |
| 115 | 8,521.46 | 8,374.88 | 8,616.10 | 9,169.36 | **34,681.79** |
| 116 | 566.67 | 441.80 | 465.89 | 573.93 | **2,048.29** |
| 117 | 24,653.87 | 24,160.36 | 25,563.65 | 26,657.15 | **101,035.04** |
| 118 | 1,121.74 | 1,188.35 | 1,299.00 | 1,231.35 | **4,840.44** |
| 119 | 3,981.32 | 5,444.66 | 5,578.76 | 4,990.98 | **19,995.73** |
| 120 | 6,783.21 | 6,448.02 | 6,740.42 | 6,816.77 | **26,788.42** |
| 121 | 4,964.82 | 4,523.60 | 4,912.73 | 4,955.72 | **19,356.88** |
| 122 | 3,452.94 | 3,096.75 | 3,140.07 | 3,335.69 | **13,025.45** |
| 123 | 3,077.85 | 2,811.21 | 3,076.71 | 3,170.83 | **12,136.61** |
| 124 | 2,568.38 | 2,765.19 | 2,768.88 | 2,994.70 | **11,097.15** |
| 125 | 6,922.96 | 6,923.48 | 7,208.38 | 7,631.06 | **28,685.89** |
| 126 | 1,561.51 | 1,545.62 | 1,628.77 | 1,746.28 | **6,482.18** |
| 127 | 1,540.90 | 1,439.65 | 1,572.70 | 1,593.32 | **6,146.57** |
| 128 | 4,383.55 | 4,008.55 | 4,394.60 | 4,434.78 | **17,221.49** |
| 129 | 2,985.68 | 2,660.15 | 3,014.57 | 3,038.12 | **11,698.52** |
| 130 | 7,432.93 | 6,831.47 | 7,295.17 | 7,627.58 | **29,187.16** |
| 131 | 2,213.02 | 2,097.12 | 2,176.26 | 2,359.75 | **8,846.16** |
| 132 | 4,174.74 | 3,643.37 | 4,008.78 | 4,077.66 | **15,904.55** |
| 133 | 1,340.75 | 1,162.96 | 1,244.47 | 1,316.39 | **5,064.57** |
| 134 | 2,366.54 | 2,196.24 | 2,289.45 | 2,365.35 | **9,217.58** |
| 135 | 1,140.93 | 950.61 | 970.15 | 1,016.88 | **4,078.57** |
| 136 | 3,446.81 | 3,134.15 | 3,355.16 | 3,358.53 | **13,294.64** |
| 137 | 1,369.65 | 1,316.90 | 1,315.59 | 1,343.59 | **5,345.72** |
| 138 | 1,827.90 | 1,610.01 | 1,673.45 | 1,757.48 | **6,868.84** |
| 139 | 1,241.48 | 967.48 | 1,041.39 | 1,061.95 | **4,312.30** |
| 140 | 7,596.22 | 7,476.30 | 7,836.91 | 8,117.74 | **31,027.17** |
| 141 | 3,198.70 | 3,140.54 | 3,171.82 | 3,334.01 | **12,845.06** |
| 142 | 10,864.59 | 11,142.36 | 11,014.15 | 11,427.65 | **44,448.74** |
| 143 | 1,401.72 | 1,302.44 | 1,295.27 | 1,401.97 | **5,401.39** |
| 144 | 6,887.82 | 6,513.50 | 6,611.70 | 7,010.26 | **27,023.28** |
| 145 | 6,074.63 | 5,666.54 | 5,852.15 | 6,091.77 | **23,685.09** |
| 146 | 11,515.22 | 10,694.03 | 11,200.45 | 11,788.36 | **45,198.05** |
| 147 | 1,705.99 | 1,637.29 | 1,633.77 | 1,794.68 | **6,771.73** |
| 148 | 4,014.30 | 3,523.01 | 3,825.92 | 4,071.85 | **15,435.08** |
| 149 | 1,022.95 | 951.08 | 949.91 | 1,024.63 | **3,948.57** |
| **NY - NEW YORK - Total** | **279,794.59** | **249,730.54** | **262,787.64** | **276,988.28** | **1,069,301.05** |

**STATE: OH - OHIO**

| REGISTRANT ZIP CODE 3 | 1ST QUARTER | 2ND QUARTER | 3RD QUARTER | 4TH QUARTER | TOTAL GRAMS |
|---|---|---|---|---|---|
| 430 | 18,721.49 | 18,073.15 | 18,579.92 | 19,617.05 | **74,991.61** |
| 431 | 7,324.26 | 7,213.47 | 7,558.80 | 7,933.14 | **30,029.67** |
| 432 | 16,067.90 | 15,633.44 | 15,889.26 | 16,558.62 | **64,149.23** |
| 433 | 2,526.17 | 2,436.24 | 2,430.69 | 2,591.76 | **9,984.87** |
| 434 | 2,409.83 | 2,457.16 | 2,563.99 | 2,612.87 | **10,043.85** |
| 435 | 6,394.88 | 6,327.20 | 6,449.31 | 6,825.69 | **25,997.08** |
| 436 | 10,026.51 | 9,723.21 | 9,935.61 | 10,295.10 | **39,980.44** |
| 437 | 1,736.14 | 1,733.66 | 1,762.14 | 1,862.86 | **7,094.80** |
| 438 | 286.84 | 243.22 | 253.76 | 290.52 | **1,074.35** |
| 439 | 2,382.29 | 2,263.37 | 2,292.22 | 2,270.71 | **9,208.59** |
| 440 | 17,853.99 | 17,341.90 | 17,856.99 | 18,885.45 | **71,938.34** |

| | | | | | |
|---|---|---|---|---|---|
| 441 | 24,183.71 | 23,229.48 | 23,731.25 | 24,594.96 | **95,739.40** |
| 442 | 9,405.28 | 9,017.64 | 9,205.92 | 9,975.66 | **37,604.50** |
| 443 | 4,233.06 | 4,127.74 | 4,095.14 | 4,342.37 | **16,798.32** |
| 444 | 5,301.00 | 5,175.75 | 5,226.42 | 5,622.14 | **21,325.31** |
| 445 | 5,301.78 | 5,245.98 | 5,268.76 | 5,679.75 | **21,496.27** |
| 446 | 6,261.51 | 6,084.65 | 6,123.13 | 6,545.23 | **25,014.52** |
| 447 | 5,325.16 | 5,193.09 | 5,307.72 | 5,333.40 | **21,159.37** |
| 448 | 3,780.31 | 3,697.11 | 3,934.13 | 4,058.40 | **15,469.95** |
| 449 | 999.28 | 1,001.07 | 1,082.10 | 1,190.68 | **4,273.13** |
| 450 | 17,020.28 | 17,120.40 | 17,770.31 | 18,469.54 | **70,380.53** |
| 451 | 5,889.28 | 5,962.97 | 6,030.11 | 6,257.20 | **24,139.57** |
| 452 | 23,325.00 | 22,356.46 | 23,228.66 | 24,415.84 | **93,325.97** |
| 453 | 5,911.58 | 5,833.86 | 5,962.14 | 6,335.77 | **24,043.35** |
| 454 | 7,034.17 | 6,920.24 | 7,108.76 | 7,633.14 | **28,696.32** |
| 455 | 1,499.34 | 1,445.42 | 1,432.87 | 1,462.96 | **5,840.59** |
| 456 | 4,294.39 | 4,061.89 | 4,310.39 | 4,351.04 | **17,017.71** |
| 457 | 2,757.85 | 2,508.75 | 2,608.12 | 2,829.98 | **10,704.70** |
| 458 | 4,085.38 | 3,847.03 | 3,887.70 | 4,155.77 | **15,975.88** |
| **OH - OHIO - Total** | **222,338.66** | **216,275.59** | **221,886.36** | **232,997.60** | **893,498.21** |

**STATE: OK - OKLAHOMA**

| REGISTRANT ZIP CODE 3 | 1ST QUARTER | 2ND QUARTER | 3RD QUARTER | 4TH QUARTER | **TOTAL GRAMS** |
|---|---|---|---|---|---|
| 730 | 12,362.20 | 11,700.73 | 12,371.21 | 13,099.78 | **49,533.92** |
| 731 | 10,555.79 | 10,646.28 | 10,887.42 | 11,831.67 | **43,921.16** |
| 734 | 1,062.87 | 1,147.86 | 1,214.04 | 1,244.19 | **4,668.96** |
| 735 | 2,684.99 | 2,610.11 | 2,909.70 | 2,440.69 | **10,645.49** |
| 736 | 377.15 | 336.02 | 378.07 | 373.23 | **1,464.47** |
| 737 | 1,161.83 | 1,128.05 | 1,173.84 | 1,250.76 | **4,714.48** |
| 738 | 178.59 | 198.94 | 211.99 | 243.17 | **832.69** |
| 739 | 61.23 | 56.68 | 54.67 | 55.80 | **228.39** |
| 740 | 13,244.86 | 12,787.95 | 13,214.93 | 13,578.02 | **52,825.76** |
| 741 | 9,675.13 | 9,325.99 | 9,467.13 | 9,766.08 | **38,234.33** |
| 743 | 1,115.66 | 1,189.57 | 1,159.37 | 1,127.59 | **4,592.19** |
| 744 | 2,759.64 | 2,632.63 | 2,752.99 | 2,834.03 | **10,979.29** |
| 745 | 477.03 | 497.30 | 518.18 | 524.07 | **2,016.59** |
| 746 | 326.03 | 359.45 | 393.82 | 406.57 | **1,485.87** |
| 747 | 810.84 | 778.05 | 802.80 | 815.68 | **3,207.37** |
| 748 | 1,455.65 | 1,412.34 | 1,406.07 | 1,435.54 | **5,709.60** |
| 749 | 476.65 | 424.65 | 448.44 | 507.76 | **1,857.49** |
| **OK - OKLAHOMA - Total** | **58,786.14** | **57,232.59** | **59,364.66** | **61,534.64** | **236,918.03** |

**STATE: OR - OREGON**

| REGISTRANT ZIP CODE 3 | 1ST QUARTER | 2ND QUARTER | 3RD QUARTER | 4TH QUARTER | **TOTAL GRAMS** |
|---|---|---|---|---|---|
| 970 | 14,569.11 | 14,303.20 | 14,509.90 | 15,362.45 | **58,744.67** |
| 971 | 5,640.08 | 5,249.63 | 5,721.75 | 6,039.55 | **22,651.02** |
| 972 | 23,669.48 | 25,514.09 | 26,049.07 | 27,376.42 | **102,609.07** |
| 973 | 9,469.93 | 8,728.52 | 9,055.22 | 9,723.61 | **36,977.27** |
| 974 | 8,034.42 | 8,114.25 | 8,298.11 | 8,779.12 | **33,225.90** |
| 975 | 2,966.24 | 2,972.65 | 3,074.51 | 3,311.80 | **12,325.19** |
| 976 | 488.45 | 512.29 | 531.25 | 557.82 | **2,089.81** |
| 977 | 4,466.57 | 4,683.39 | 4,806.80 | 4,940.57 | **18,897.32** |
| 978 | 1,145.78 | 1,178.31 | 1,248.20 | 1,317.82 | **4,890.10** |
| 979 | 412.41 | 384.31 | 399.15 | 379.73 | **1,575.61** |
| **OR - OREGON - Total** | **70,862.47** | **71,640.65** | **73,693.95** | **77,788.88** | **293,985.95** |

**STATE: PA - PENNSYLVANIA**

| REGISTRANT ZIP CODE 3 | 1ST QUARTER | 2ND QUARTER | 3RD QUARTER | 4TH QUARTER | **TOTAL GRAMS** |
|---|---|---|---|---|---|
| 150 | 6,978.24 | 6,715.33 | 7,027.55 | 7,327.69 | **28,048.81** |
| 151 | 5,962.09 | 5,950.04 | 6,116.92 | 6,459.64 | **24,488.68** |
| 152 | 12,499.08 | 11,938.38 | 12,478.14 | 13,493.68 | **50,409.27** |
| 153 | 2,607.64 | 2,642.23 | 2,586.08 | 2,825.60 | **10,661.55** |
| 154 | 1,920.09 | 1,815.43 | 1,708.60 | 1,798.05 | **7,242.17** |
| 155 | 1,132.43 | 1,094.69 | 1,090.81 | 1,104.40 | **4,422.32** |

| | | | | | |
|---|---|---|---|---|---|
| 156 | 4,567.70 | 4,447.43 | 4,600.28 | 4,882.20 | **18,497.61** |
| 157 | 1,265.62 | 1,252.61 | 1,293.29 | 1,347.89 | **5,159.42** |
| 158 | 1,425.95 | 1,284.33 | 1,400.96 | 1,447.11 | **5,558.35** |
| 159 | 1,430.71 | 1,383.13 | 1,394.82 | 1,524.89 | **5,733.55** |
| 160 | 3,451.48 | 3,451.49 | 3,433.43 | 3,738.50 | **14,074.91** |
| 161 | 2,773.55 | 2,843.15 | 2,779.28 | 2,977.05 | **11,373.04** |
| 162 | 987.64 | 940.84 | 986.42 | 1,027.53 | **3,942.44** |
| 163 | 1,904.98 | 1,859.16 | 1,888.37 | 2,000.11 | **7,652.62** |
| 164 | 1,099.96 | 1,149.05 | 1,171.78 | 1,173.89 | **4,594.67** |
| 165 | 4,591.85 | 4,487.42 | 4,498.76 | 4,615.38 | **18,193.41** |
| 166 | 2,776.90 | 2,742.43 | 2,820.17 | 2,914.44 | **11,253.94** |
| 167 | 650.85 | 672.58 | 676.85 | 709.80 | **2,710.07** |
| 168 | 2,921.61 | 2,621.32 | 2,783.19 | 2,793.62 | **11,119.74** |
| 169 | 566.82 | 533.22 | 546.48 | 600.82 | **2,247.34** |
| 170 | 12,595.64 | 12,138.66 | 12,387.30 | 12,625.28 | **49,746.88** |
| 171 | 3,124.44 | 3,048.84 | 3,006.82 | 3,088.99 | **12,269.08** |
| 172 | 3,808.73 | 3,655.36 | 3,838.12 | 3,769.48 | **15,071.68** |
| 173 | 6,280.05 | 6,096.97 | 6,333.97 | 6,609.06 | **25,320.06** |
| 174 | 4,090.89 | 4,054.61 | 4,145.96 | 4,297.34 | **16,588.80** |
| 175 | 3,768.88 | 3,742.25 | 4,300.28 | 4,378.28 | **16,189.69** |
| 176 | 4,400.91 | 4,343.67 | 4,391.91 | 4,684.83 | **17,821.32** |
| 177 | 2,553.62 | 2,411.44 | 2,479.33 | 2,645.80 | **10,090.19** |
| 178 | 2,912.54 | 2,601.11 | 2,791.87 | 2,746.45 | **11,051.97** |
| 179 | 858.17 | 861.13 | 882.48 | 936.56 | **3,538.34** |
| 180 | 6,207.22 | 6,190.15 | 6,443.49 | 6,629.33 | **25,470.18** |
| 181 | 2,971.03 | 2,776.10 | 2,805.27 | 2,775.49 | **11,327.90** |
| 182 | 1,003.47 | 966.96 | 1,013.02 | 1,036.49 | **4,019.95** |
| 183 | 1,662.05 | 1,682.30 | 1,790.75 | 1,762.33 | **6,897.42** |
| 184 | 2,466.96 | 2,592.09 | 2,665.19 | 2,667.27 | **10,391.51** |
| 185 | 3,659.52 | 3,455.76 | 3,516.85 | 3,536.96 | **14,169.09** |
| 186 | 1,997.94 | 2,003.24 | 2,022.57 | 2,174.46 | **8,198.21** |
| 187 | 20,930.88 | 25,868.57 | 23,619.86 | 20,888.61 | **91,307.91** |
| 188 | 750.99 | 640.39 | 696.74 | 708.49 | **2,796.60** |
| 189 | 7,862.75 | 7,772.59 | 7,913.58 | 8,235.65 | **31,784.58** |
| 190 | 23,758.21 | 23,375.66 | 23,237.41 | 23,756.66 | **94,127.95** |
| 191 | 21,584.34 | 20,521.08 | 20,314.25 | 21,100.39 | **83,520.06** |
| 193 | 10,924.96 | 10,518.48 | 10,441.65 | 11,001.61 | **42,886.71** |
| 194 | 11,763.97 | 11,445.57 | 11,504.02 | 11,951.48 | **46,665.04** |
| 195 | 2,094.65 | 2,070.25 | 2,094.60 | 2,201.83 | **8,461.33** |
| 196 | 3,460.51 | 3,170.57 | 3,188.10 | 3,270.33 | **13,089.51** |
| **PA - PENNSYLVANIA - Total** | **229,008.50** | **227,828.06** | **229,107.57** | **234,241.73** | **920,185.86** |

**STATE: PR - PUERTO RICO**

| REGISTRANT ZIP CODE 3 | 1ST QUARTER | 2ND QUARTER | 3RD QUARTER | 4TH QUARTER | TOTAL GRAMS |
|---|---|---|---|---|---|
| 006 | 1,880.29 | 1,545.55 | 1,889.55 | 1,835.56 | **7,150.95** |
| 007 | 1,935.79 | 1,455.17 | 2,011.65 | 1,974.66 | **7,377.27** |
| 009 | 4,514.53 | 3,822.90 | 4,607.05 | 4,520.55 | **17,465.02** |
| **PR - PUERTO RICO - Total** | **8,330.60** | **6,823.61** | **8,508.26** | **8,330.77** | **31,993.24** |

**STATE: RI - RHODE ISLAND**

| REGISTRANT ZIP CODE 3 | 1ST QUARTER | 2ND QUARTER | 3RD QUARTER | 4TH QUARTER | TOTAL GRAMS |
|---|---|---|---|---|---|
| 028 | 26,069.85 | 25,498.88 | 26,540.12 | 26,829.62 | **104,938.47** |
| 029 | 14,204.97 | 13,891.13 | 14,075.20 | 14,157.46 | **56,328.76** |
| **RI - RHODE ISLAND - Total** | **40,274.83** | **39,390.00** | **40,615.32** | **40,987.09** | **161,267.23** |

**STATE: SC - SOUTH CAROLINA**

| REGISTRANT ZIP CODE 3 | 1ST QUARTER | 2ND QUARTER | 3RD QUARTER | 4TH QUARTER | TOTAL GRAMS |
|---|---|---|---|---|---|
| 290 | 13,495.73 | 13,290.75 | 13,582.55 | 14,201.88 | **54,570.91** |
| 291 | 10,369.88 | 9,982.55 | 10,357.19 | 10,549.52 | **41,259.14** |
| 292 | 10,995.68 | 9,665.68 | 10,577.17 | 10,920.19 | **42,158.72** |
| 293 | 9,329.23 | 8,791.33 | 9,140.21 | 9,758.55 | **37,019.32** |
| 294 | 37,733.62 | 36,580.12 | 37,795.80 | 39,066.98 | **151,176.52** |
| 295 | 19,815.37 | 19,160.48 | 19,933.80 | 20,836.24 | **79,745.90** |

| | | | | | |
|---|---|---|---|---|---|
| 296 | 31,518.68 | 30,210.18 | 31,739.06 | 33,266.86 | **126,734.77** |
| 297 | 9,469.96 | 8,958.63 | 9,317.16 | 9,841.13 | **37,586.88** |
| 298 | 4,663.18 | 4,546.88 | 4,568.95 | 4,692.37 | **18,471.38** |
| 299 | 4,567.59 | 4,817.71 | 4,901.40 | 4,848.20 | **19,134.90** |
| **SC - SOUTH CAROLINA - Total** | **151,958.92** | **146,004.30** | **151,913.29** | **157,981.93** | **607,858.44** |

**STATE: SD - SOUTH DAKOTA**

| REGISTRANT ZIP CODE 3 | 1ST QUARTER | 2ND QUARTER | 3RD QUARTER | 4TH QUARTER | TOTAL GRAMS |
|---|---|---|---|---|---|
| 570 | 2,409.59 | 2,269.24 | 2,352.96 | 2,511.09 | **9,542.88** |
| 571 | 7,119.88 | 6,521.62 | 6,383.48 | 6,163.26 | **26,188.24** |
| 572 | 753.96 | 712.18 | 781.99 | 798.15 | **3,046.28** |
| 573 | 620.48 | 690.56 | 725.71 | 695.76 | **2,732.51** |
| 574 | 814.36 | 796.76 | 844.32 | 799.41 | **3,254.86** |
| 575 | 272.35 | 231.03 | 283.74 | 262.04 | **1,049.16** |
| 576 | 35.81 | 61.05 | 43.12 | 54.98 | **194.95** |
| 577 | 2,880.52 | 2,869.37 | 2,881.15 | 2,913.65 | **11,544.69** |
| **SD - SOUTH DAKOTA - Total** | **14,906.94** | **14,151.81** | **14,296.48** | **14,198.33** | **57,553.56** |

**STATE: TN - TENNESSEE**

| REGISTRANT ZIP CODE 3 | 1ST QUARTER | 2ND QUARTER | 3RD QUARTER | 4TH QUARTER | TOTAL GRAMS |
|---|---|---|---|---|---|
| 370 | 21,988.03 | 21,821.18 | 22,818.52 | 23,238.37 | **89,866.09** |
| 371 | 14,218.46 | 14,524.26 | 15,117.14 | 16,143.94 | **60,003.80** |
| 372 | 15,841.42 | 15,189.44 | 15,798.56 | 16,096.96 | **62,926.38** |
| 373 | 13,310.52 | 13,065.72 | 13,789.57 | 14,305.71 | **54,471.51** |
| 374 | 9,038.96 | 8,878.72 | 9,184.07 | 9,609.62 | **36,711.37** |
| 376 | 4,666.11 | 4,575.54 | 4,517.69 | 4,862.02 | **18,621.36** |
| 377 | 4,874.18 | 4,770.76 | 4,893.29 | 5,325.36 | **19,863.60** |
| 378 | 7,730.11 | 7,533.04 | 7,768.62 | 8,413.67 | **31,445.44** |
| 379 | 12,681.48 | 12,297.88 | 12,424.88 | 13,318.44 | **50,722.68** |
| 380 | 10,749.09 | 10,730.10 | 11,076.34 | 11,435.20 | **43,990.73** |
| 381 | 13,365.41 | 12,744.13 | 13,092.63 | 13,637.02 | **52,839.18** |
| 382 | 3,160.78 | 3,046.54 | 3,090.21 | 3,273.40 | **12,570.93** |
| 383 | 4,685.00 | 4,592.20 | 4,765.83 | 4,953.81 | **18,996.84** |
| 384 | 3,545.61 | 3,281.76 | 3,527.28 | 3,552.01 | **13,906.66** |
| 385 | 2,353.24 | 2,240.77 | 2,462.16 | 2,519.14 | **9,575.31** |
| **TN - TENNESSEE - Total** | **142,208.40** | **139,292.03** | **144,326.77** | **150,684.68** | **576,511.88** |

**STATE: TX - TEXAS**

| REGISTRANT ZIP CODE 3 | 1ST QUARTER | 2ND QUARTER | 3RD QUARTER | 4TH QUARTER | TOTAL GRAMS |
|---|---|---|---|---|---|
| 750 | 63,556.43 | 62,086.62 | 65,056.06 | 67,339.27 | **258,038.37** |
| 751 | 11,274.26 | 11,503.15 | 12,150.38 | 12,595.21 | **47,523.02** |
| 752 | 31,638.15 | 30,444.89 | 31,586.85 | 32,276.26 | **125,946.14** |
| 753 | 73.82 | 35.56 | 39.16 | 48.19 | **196.74** |
| 754 | 5,184.45 | 5,188.96 | 5,637.88 | 5,801.54 | **21,812.83** |
| 755 | 1,440.68 | 1,334.95 | 1,447.69 | 1,520.05 | **5,743.38** |
| 756 | 4,586.24 | 4,713.21 | 4,882.01 | 5,113.24 | **19,294.70** |
| 757 | 6,232.17 | 6,103.93 | 6,273.54 | 6,400.22 | **25,009.86** |
| 758 | 918.12 | 907.70 | 909.83 | 913.93 | **3,649.58** |
| 759 | 3,047.26 | 2,804.34 | 3,015.79 | 3,190.28 | **12,057.67** |
| 760 | 28,117.87 | 28,005.46 | 29,158.22 | 30,158.79 | **115,440.35** |
| 761 | 17,351.30 | 17,064.25 | 17,908.53 | 18,772.11 | **71,096.19** |
| 762 | 14,653.86 | 15,025.01 | 15,494.82 | 16,253.16 | **61,426.84** |
| 763 | 3,810.61 | 3,845.34 | 3,898.12 | 4,196.48 | **15,750.54** |
| 764 | 1,597.44 | 1,663.59 | 1,795.13 | 1,799.27 | **6,855.44** |
| 765 | 6,770.87 | 6,836.68 | 7,205.81 | 6,902.19 | **27,715.55** |
| 766 | 1,342.50 | 1,302.67 | 1,409.70 | 1,367.51 | **5,422.39** |
| 767 | 4,382.61 | 3,901.58 | 4,201.10 | 4,265.88 | **16,751.16** |
| 768 | 918.52 | 887.72 | 940.74 | 937.74 | **3,684.71** |
| 769 | 2,199.23 | 2,124.25 | 2,109.43 | 2,258.84 | **8,691.75** |
| 770 | 45,025.52 | 43,843.69 | 43,732.60 | 46,301.92 | **178,903.74** |
| 772 | 15.95 | 1.88 | 2.19 | 11.89 | **31.91** |
| 773 | 33,976.99 | 33,763.68 | 34,681.67 | 36,256.03 | **138,678.37** |
| 774 | 25,961.28 | 25,360.74 | 26,222.09 | 27,705.98 | **105,250.09** |

| | | | | | |
|---|---|---|---|---|---|
| 775 | 28,004.66 | 27,041.42 | 28,153.38 | 29,346.65 | **112,546.10** |
| 776 | 5,876.17 | 5,650.69 | 5,780.90 | 6,211.38 | **23,519.13** |
| 777 | 2,723.43 | 2,800.71 | 2,758.91 | 2,945.44 | **11,228.49** |
| 778 | 7,025.78 | 6,265.90 | 6,651.63 | 7,243.29 | **27,186.60** |
| 779 | 2,645.83 | 2,594.66 | 2,649.36 | 2,678.91 | **10,568.76** |
| 780 | 6,732.17 | 6,716.49 | 6,974.53 | 7,308.84 | **27,732.04** |
| 781 | 7,830.97 | 7,657.29 | 7,944.14 | 8,417.89 | **31,850.29** |
| 782 | 31,860.72 | 30,494.22 | 31,318.01 | 32,357.32 | **126,030.27** |
| 783 | 2,232.22 | 2,341.78 | 2,412.88 | 2,343.49 | **9,330.37** |
| 784 | 5,720.26 | 5,694.31 | 5,816.76 | 5,855.55 | **23,086.88** |
| 785 | 4,110.02 | 3,699.70 | 3,794.16 | 4,077.46 | **15,681.35** |
| 786 | 22,378.66 | 22,543.36 | 23,033.03 | 24,197.64 | **92,152.69** |
| 787 | 35,819.91 | 35,381.80 | 35,904.53 | 37,019.64 | **144,125.88** |
| 788 | 774.91 | 709.47 | 697.39 | 741.92 | **2,923.69** |
| 789 | 921.43 | 889.22 | 860.19 | 901.76 | **3,572.60** |
| 790 | 999.13 | 998.76 | 1,077.14 | 1,126.48 | **4,201.50** |
| 791 | 2,510.66 | 2,436.17 | 2,442.28 | 2,562.39 | **9,951.49** |
| 792 | 128.03 | 107.68 | 137.60 | 131.07 | **504.38** |
| 793 | 507.75 | 467.17 | 530.07 | 578.91 | **2,083.90** |
| 794 | 5,065.53 | 4,591.93 | 4,854.70 | 5,175.74 | **19,687.91** |
| 795 | 599.53 | 589.03 | 596.18 | 596.93 | **2,381.67** |
| 796 | 2,729.92 | 2,589.12 | 2,715.81 | 2,804.88 | **10,839.73** |
| 797 | 7,056.95 | 6,964.59 | 6,890.12 | 6,869.60 | **27,781.27** |
| 798 | 296.25 | 250.05 | 263.24 | 266.93 | **1,076.48** |
| 799 | 3,974.12 | 3,852.12 | 4,158.26 | 3,423.59 | **15,408.08** |
| **TX - TEXAS - Total** | **502,601.16** | **492,077.50** | **508,174.54** | **527,569.66** | **2,030,422.85** |

**STATE: UT - UTAH**

| REGISTRANT ZIP CODE 3 | 1ST QUARTER | 2ND QUARTER | 3RD QUARTER | 4TH QUARTER | TOTAL GRAMS |
|---|---|---|---|---|---|
| 840 | 51,875.72 | 52,425.16 | 54,373.19 | 56,651.76 | **215,325.83** |
| 841 | 19,678.48 | 19,805.07 | 20,556.35 | 20,917.10 | **80,957.01** |
| 843 | 5,412.90 | 5,444.43 | 5,306.67 | 5,972.64 | **22,136.64** |
| 844 | 5,474.22 | 5,303.74 | 5,734.88 | 5,841.01 | **22,353.86** |
| 845 | 749.37 | 825.23 | 854.46 | 929.62 | **3,358.68** |
| 846 | 10,388.70 | 10,781.91 | 11,120.31 | 11,604.42 | **43,895.33** |
| 847 | 7,806.78 | 7,718.81 | 8,131.29 | 8,414.52 | **32,071.40** |
| **UT - UTAH - Total** | **101,386.17** | **102,304.35** | **106,077.15** | **110,331.08** | **420,098.76** |

**STATE: VA - VIRGINIA**

| REGISTRANT ZIP CODE 3 | 1ST QUARTER | 2ND QUARTER | 3RD QUARTER | 4TH QUARTER | TOTAL GRAMS |
|---|---|---|---|---|---|
| 201 | 20,797.93 | 19,838.03 | 20,276.28 | 21,164.62 | **82,076.87** |
| 220 | 11,039.77 | 9,600.07 | 10,410.50 | 9,900.17 | **40,950.52** |
| 221 | 8,044.79 | 7,058.96 | 7,698.32 | 7,620.35 | **30,422.43** |
| 222 | 5,373.92 | 5,119.85 | 5,291.29 | 5,208.22 | **20,993.28** |
| 223 | 5,156.03 | 4,918.97 | 5,036.68 | 5,249.12 | **20,360.80** |
| 224 | 5,036.69 | 4,854.78 | 4,854.91 | 5,215.97 | **19,962.34** |
| 225 | 2,459.22 | 2,321.24 | 2,440.48 | 2,728.40 | **9,949.34** |
| 226 | 4,081.01 | 3,813.48 | 3,909.99 | 4,112.46 | **15,916.94** |
| 227 | 1,227.54 | 1,162.00 | 1,255.50 | 1,338.23 | **4,983.27** |
| 228 | 3,363.54 | 2,834.22 | 3,116.37 | 3,316.71 | **12,630.84** |
| 229 | 7,277.53 | 6,778.57 | 7,093.01 | 7,201.25 | **28,350.37** |
| 230 | 5,314.88 | 5,111.38 | 5,139.34 | 5,450.37 | **21,015.97** |
| 231 | 13,415.63 | 13,676.93 | 13,806.31 | 14,509.81 | **55,408.68** |
| 232 | 18,087.64 | 17,177.13 | 17,522.25 | 18,087.12 | **70,874.14** |
| 233 | 9,941.10 | 10,087.81 | 10,001.95 | 10,589.78 | **40,620.63** |
| 234 | 25,578.38 | 25,984.82 | 26,579.52 | 26,134.87 | **104,277.59** |
| 235 | 6,073.65 | 6,250.11 | 6,102.42 | 6,402.86 | **24,829.04** |
| 236 | 10,166.70 | 10,289.73 | 10,797.57 | 10,675.56 | **41,929.56** |
| 237 | 3,927.78 | 3,928.09 | 4,041.72 | 3,941.39 | **15,838.97** |
| 238 | 6,880.35 | 6,596.17 | 6,664.34 | 6,697.24 | **26,838.10** |
| 239 | 1,085.04 | 840.87 | 1,001.77 | 975.44 | **3,903.12** |
| 240 | 5,926.55 | 5,413.47 | 5,596.28 | 5,954.16 | **22,890.46** |

| | | | | | |
|---|---|---|---|---|---|
| 241 | 3,002.63 | 2,872.92 | 2,969.55 | 3,083.91 | **11,929.00** |
| 242 | 1,371.27 | 1,281.39 | 1,337.18 | 1,441.87 | **5,431.71** |
| 243 | 1,041.33 | 958.82 | 907.39 | 914.69 | **3,822.23** |
| 244 | 3,580.83 | 3,588.73 | 3,561.39 | 3,710.63 | **14,441.58** |
| 245 | 5,362.25 | 5,309.99 | 5,424.50 | 5,739.90 | **21,836.63** |
| 246 | 469.79 | 453.33 | 442.29 | 473.70 | **1,839.11** |
| **VA - VIRGINIA - Total** | **195,083.74** | **188,121.88** | **193,279.10** | **197,838.79** | **774,323.52** |

**STATE: VI - VIRGIN ISLANDS**

| REGISTRANT ZIP CODE 3 | 1ST QUARTER | 2ND QUARTER | 3RD QUARTER | 4TH QUARTER | TOTAL GRAMS |
|---|---|---|---|---|---|
| 008 | 447.40 | 337.41 | 356.19 | 345.12 | **1,486.11** |
| **VI - VIRGIN ISLANDS - Total** | **447.40** | **337.41** | **356.19** | **345.12** | **1,486.11** |

**STATE: VT - VERMONT**

| REGISTRANT ZIP CODE 3 | 1ST QUARTER | 2ND QUARTER | 3RD QUARTER | 4TH QUARTER | TOTAL GRAMS |
|---|---|---|---|---|---|
| 050 | 626.15 | 634.58 | 658.84 | 674.46 | **2,594.04** |
| 051 | 561.09 | 552.34 | 606.92 | 583.65 | **2,304.01** |
| 052 | 931.67 | 939.14 | 966.89 | 992.15 | **3,829.86** |
| 053 | 1,671.97 | 1,502.24 | 1,540.75 | 1,580.34 | **6,295.31** |
| 054 | 4,041.16 | 3,530.66 | 3,667.17 | 3,942.84 | **15,181.83** |
| 056 | 1,768.71 | 1,533.42 | 1,673.95 | 1,738.67 | **6,714.76** |
| 057 | 1,046.84 | 949.94 | 962.93 | 1,109.93 | **4,069.64** |
| 058 | 853.94 | 805.63 | 913.19 | 978.31 | **3,551.07** |
| **VT - VERMONT - Total** | **11,501.54** | **10,447.96** | **10,990.64** | **11,600.36** | **44,540.51** |

**STATE: WA - WASHINGTON**

| REGISTRANT ZIP CODE 3 | 1ST QUARTER | 2ND QUARTER | 3RD QUARTER | 4TH QUARTER | TOTAL GRAMS |
|---|---|---|---|---|---|
| 980 | 31,223.74 | 32,280.09 | 33,421.45 | 34,464.92 | **131,390.20** |
| 981 | 20,440.83 | 19,265.65 | 19,729.15 | 20,601.42 | **80,037.04** |
| 982 | 17,527.50 | 18,253.98 | 18,431.54 | 18,967.27 | **73,180.29** |
| 983 | 11,799.38 | 11,560.98 | 12,062.75 | 12,533.49 | **47,956.61** |
| 984 | 7,617.32 | 7,186.51 | 7,277.32 | 7,602.90 | **29,684.05** |
| 985 | 6,447.80 | 6,198.27 | 6,478.66 | 6,760.92 | **25,885.65** |
| 986 | 9,282.33 | 8,797.84 | 9,004.49 | 9,689.23 | **36,773.90** |
| 988 | 2,132.61 | 2,184.25 | 2,227.60 | 2,417.03 | **8,961.48** |
| 989 | 2,794.78 | 2,609.93 | 2,761.28 | 2,821.98 | **10,987.97** |
| 990 | 1,182.84 | 1,228.70 | 1,316.32 | 1,375.42 | **5,103.29** |
| 991 | 1,232.24 | 1,047.13 | 1,157.12 | 1,179.02 | **4,615.51** |
| 992 | 7,179.30 | 6,635.21 | 6,892.28 | 7,117.77 | **27,824.56** |
| 993 | 5,839.75 | 5,727.37 | 5,866.04 | 6,174.02 | **23,607.18** |
| 994 | 270.81 | 255.12 | 270.01 | 299.01 | **1,094.95** |
| **WA - WASHINGTON - Total** | **124,971.25** | **123,231.04** | **126,896.00** | **132,004.39** | **507,102.69** |

**STATE: WI - WISCONSIN**

| REGISTRANT ZIP CODE 3 | 1ST QUARTER | 2ND QUARTER | 3RD QUARTER | 4TH QUARTER | TOTAL GRAMS |
|---|---|---|---|---|---|
| 530 | 15,314.11 | 15,030.94 | 15,363.32 | 16,281.10 | **61,989.47** |
| 531 | 15,727.28 | 15,677.32 | 16,018.72 | 16,395.45 | **63,818.77** |
| 532 | 14,109.81 | 13,496.97 | 13,658.57 | 14,147.50 | **55,412.85** |
| 534 | 4,158.72 | 4,121.56 | 4,064.56 | 4,357.57 | **16,702.42** |
| 535 | 10,744.48 | 10,599.28 | 10,883.80 | 11,213.70 | **43,441.25** |
| 537 | 8,248.88 | 7,653.93 | 7,890.22 | 8,216.22 | **32,009.25** |
| 538 | 1,145.19 | 1,139.94 | 1,196.78 | 1,283.59 | **4,765.50** |
| 539 | 3,699.05 | 3,666.53 | 3,635.87 | 3,901.17 | **14,902.62** |
| 540 | 2,543.22 | 2,494.15 | 2,635.79 | 2,549.79 | **10,222.96** |
| 541 | 6,928.11 | 6,766.72 | 6,878.26 | 6,981.37 | **27,554.46** |
| 542 | 2,505.32 | 2,602.73 | 2,621.06 | 2,635.15 | **10,364.25** |
| 543 | 8,236.19 | 8,208.80 | 8,064.34 | 8,586.07 | **33,095.40** |
| 544 | 8,224.03 | 7,938.00 | 7,908.96 | 8,102.26 | **32,173.24** |
| 545 | 2,260.45 | 2,217.80 | 2,414.05 | 2,336.20 | **9,228.50** |
| 546 | 4,093.91 | 3,804.54 | 3,891.10 | 4,040.86 | **15,830.41** |
| 547 | 4,318.46 | 4,153.89 | 4,251.91 | 4,532.24 | **17,256.50** |
| 548 | 2,441.04 | 2,414.00 | 2,605.48 | 2,597.93 | **10,058.46** |
| 549 | 15,026.60 | 14,932.36 | 15,049.55 | 15,412.86 | **60,421.37** |
| **WI - WISCONSIN - Total** | **129,724.86** | **126,919.43** | **129,032.35** | **133,571.04** | **519,247.69** |

STATE: WV - WEST VIRGINIA

| REGISTRANT ZIP CODE 3 | 1ST QUARTER | 2ND QUARTER | 3RD QUARTER | 4TH QUARTER | TOTAL GRAMS |
|---|---|---|---|---|---|
| 247 | 349.59 | 294.52 | 371.81 | 283.50 | **1,299.42** |
| 248 | 201.53 | 142.45 | 191.80 | 197.35 | **733.14** |
| 249 | 519.26 | 444.28 | 491.80 | 482.04 | **1,937.39** |
| 250 | 1,014.03 | 815.71 | 826.73 | 925.96 | **3,582.42** |
| 251 | 716.26 | 649.91 | 734.23 | 725.35 | **2,825.75** |
| 252 | 388.73 | 370.23 | 407.58 | 403.29 | **1,569.84** |
| 253 | 3,277.16 | 2,996.05 | 3,051.14 | 3,227.04 | **12,551.39** |
| 254 | 2,919.96 | 2,819.88 | 2,893.67 | 3,058.68 | **11,692.19** |
| 255 | 2,489.68 | 2,241.16 | 2,272.22 | 2,331.69 | **9,334.76** |
| 256 | 317.00 | 264.52 | 267.76 | 301.39 | **1,150.66** |
| 257 | 2,119.53 | 1,828.22 | 1,927.64 | 2,036.81 | **7,912.20** |
| 258 | 1,128.03 | 1,141.16 | 1,091.24 | 1,167.70 | **4,528.13** |
| 259 | 235.42 | 204.79 | 214.79 | 218.20 | **873.19** |
| 260 | 1,454.83 | 1,313.68 | 1,416.00 | 1,462.69 | **5,647.21** |
| 261 | 2,151.07 | 1,916.69 | 1,997.79 | 1,925.00 | **7,990.55** |
| 262 | 337.55 | 292.97 | 273.26 | 340.05 | **1,243.84** |
| 263 | 1,128.14 | 1,090.87 | 1,154.20 | 1,167.66 | **4,540.87** |
| 264 | 250.17 | 231.73 | 208.17 | 261.75 | **951.82** |
| 265 | 3,058.71 | 2,637.89 | 2,905.12 | 3,002.19 | **11,603.91** |
| 266 | 168.39 | 144.08 | 146.73 | 154.58 | **613.78** |
| 267 | 395.55 | 325.70 | 350.65 | 379.28 | **1,451.19** |
| 268 | 259.34 | 249.67 | 232.15 | 214.06 | **955.21** |
| **WV - WEST VIRGINIA - Total** | **24,879.94** | **22,416.16** | **23,426.49** | **24,266.27** | **94,988.86** |

STATE: WY - WYOMING

| REGISTRANT ZIP CODE 3 | 1ST QUARTER | 2ND QUARTER | 3RD QUARTER | 4TH QUARTER | TOTAL GRAMS |
|---|---|---|---|---|---|
| 820 | 1,655.57 | 1,417.19 | 1,611.26 | 1,645.35 | **6,329.37** |
| 822 | 97.30 | 101.07 | 107.39 | 113.91 | **419.67** |
| 823 | 117.57 | 140.51 | 129.42 | 164.36 | **551.86** |
| 824 | 926.73 | 860.56 | 989.12 | 1,019.84 | **3,796.26** |
| 825 | 356.27 | 350.17 | 308.47 | 348.42 | **1,363.34** |
| 826 | 1,233.25 | 1,089.96 | 1,218.77 | 1,137.60 | **4,679.59** |
| 827 | 717.60 | 619.75 | 631.59 | 651.67 | **2,620.62** |
| 828 | 570.33 | 462.72 | 526.30 | 600.09 | **2,159.45** |
| 829 | 1,433.96 | 1,355.67 | 1,403.90 | 1,508.97 | **5,702.50** |
| 830 | 663.36 | 731.07 | 742.85 | 788.39 | **2,925.67** |
| 831 | 165.51 | 174.86 | 186.03 | 208.63 | **735.03** |
| **WY - WYOMING - Total** | **7,937.45** | **7,303.55** | **7,855.10** | **8,187.23** | **31,283.34** |

DRUG: 1105 - METHAMPHETAMINE

STATE: AK - ALASKA

| REGISTRANT ZIP CODE 3 | 1ST QUARTER | 2ND QUARTER | 3RD QUARTER | 4TH QUARTER | TOTAL GRAMS |
|---|---|---|---|---|---|
| 997 | 2.01 | 3.21 | 1.21 | 0 | **6.43** |
| **AK - ALASKA - Total** | **2.01** | **3.21** | **1.21** | **0.00** | **6.43** |

STATE: AL - ALABAMA

| REGISTRANT ZIP CODE 3 | 1ST QUARTER | 2ND QUARTER | 3RD QUARTER | 4TH QUARTER | TOTAL GRAMS |
|---|---|---|---|---|---|
| 350 | 2.81 | 2.81 | 2.41 | 2.41 | **10.44** |
| 351 | 0.80 | 0.80 | 1.21 | 1.21 | **4.02** |
| 352 | 8.44 | 8.84 | 11.25 | 7.23 | **35.75** |
| 354 | 1.61 | 2.01 | 1.21 | 1.61 | **6.43** |
| 355 | 3.62 | 5.62 | 4.82 | 4.42 | **18.48** |
| 356 | 6.03 | 5.62 | 4.82 | 0.80 | **17.27** |
| 357 | 0.80 | 2.01 | 2.01 | 1.21 | **6.03** |
| 358 | 0 | 0.80 | 0 | 0 | **0.80** |
| 359 | 0.40 | 1.21 | 0.40 | 0.80 | **2.81** |
| 362 | 4.42 | 2.81 | 4.42 | 2.81 | **14.47** |
| 363 | 0 | 0 | 0 | 0.80 | **0.80** |
| 365 | 0.80 | 1.61 | 2.01 | 1.61 | **6.03** |
| 366 | 0.80 | 0.80 | 0 | 0 | **1.61** |
| **AL - ALABAMA - Total** | **30.53** | **34.95** | **34.55** | **24.91** | **124.94** |

**STATE: AR - ARKANSAS**

| REGISTRANT ZIP CODE 3 | 1ST QUARTER | 2ND QUARTER | 3RD QUARTER | 4TH QUARTER | TOTAL GRAMS |
|---|---|---|---|---|---|
| 720 | 0.40 | 0.40 | 0.80 | 0.40 | **2.01** |
| 722 | 1.21 | 2.81 | 2.01 | 1.61 | **7.63** |
| 723 | 0 | 0 | 0 | 0.40 | **0.40** |
| 727 | 0 | 0 | 0 | 0.80 | **0.80** |
| **AR - ARKANSAS - Total** | **1.61** | **3.21** | **2.81** | **3.21** | **10.85** |

**STATE: AZ - ARIZONA**

| REGISTRANT ZIP CODE 3 | 1ST QUARTER | 2ND QUARTER | 3RD QUARTER | 4TH QUARTER | TOTAL GRAMS |
|---|---|---|---|---|---|
| 850 | 6.83 | 7.23 | 8.84 | 8.03 | **30.93** |
| 851 | 1.61 | 3.21 | 3.21 | 2.41 | **10.45** |
| 852 | 12.46 | 17.28 | 10.85 | 14.06 | **54.64** |
| 853 | 1.61 | 3.21 | 3.62 | 3.21 | **11.65** |
| 856 | 1.21 | 0 | 0 | 2.41 | **3.62** |
| 857 | 0.40 | 1.61 | 0.40 | 0 | **2.41** |
| 864 | 3.21 | 0 | 0.80 | 0.80 | **4.82** |
| **AZ - ARIZONA - Total** | **27.32** | **32.55** | **27.72** | **30.94** | **118.52** |

**STATE: CA - CALIFORNIA**

| REGISTRANT ZIP CODE 3 | 1ST QUARTER | 2ND QUARTER | 3RD QUARTER | 4TH QUARTER | TOTAL GRAMS |
|---|---|---|---|---|---|
| 900 | 8.04 | 10.04 | 9.64 | 7.23 | **34.95** |
| 902 | 5.62 | 5.62 | 5.22 | 4.82 | **21.29** |
| 904 | 4.42 | 6.83 | 5.62 | 7.23 | **24.10** |
| 906 | 0 | 0 | 0.40 | 0 | **0.40** |
| 907 | 0 | 0 | 0.40 | 0 | **0.40** |
| 908 | 2.01 | 2.41 | 1.61 | 2.81 | **8.84** |
| 911 | 2.81 | 1.21 | 0.80 | 0 | **4.82** |
| 912 | 2.41 | 3.21 | 1.21 | 2.81 | **9.64** |
| 913 | 17.68 | 17.27 | 15.67 | 10.85 | **61.46** |
| 914 | 5.22 | 4.02 | 5.63 | 5.62 | **20.49** |
| 915 | 3.62 | 7.23 | 5.63 | 5.22 | **21.70** |
| 916 | 0.40 | 0 | 0 | 0 | **0.40** |
| 917 | 9.64 | 9.64 | 7.23 | 5.22 | **31.74** |
| 918 | 0.40 | 0 | 0 | 0 | **0.40** |
| 919 | 3.62 | 4.02 | 1.61 | 2.01 | **11.25** |
| 920 | 18.88 | 27.72 | 24.50 | 28.92 | **100.03** |
| 921 | 6.83 | 7.23 | 7.23 | 10.04 | **31.34** |
| 922 | 6.83 | 4.02 | 3.62 | 4.02 | **18.48** |
| 923 | 5.62 | 4.02 | 4.42 | 5.22 | **19.28** |
| 925 | 25.32 | 21.70 | 26.12 | 18.88 | **92.02** |
| 926 | 24.51 | 15.67 | 18.08 | 16.87 | **75.13** |
| 927 | 4.42 | 3.62 | 4.82 | 4.42 | **17.27** |
| 928 | 11.25 | 10.45 | 9.24 | 12.05 | **42.98** |
| 930 | 0.40 | 0 | 0 | 0.80 | **1.21** |
| 933 | 0 | 0 | 0 | 0.40 | **0.40** |
| 934 | 0 | 1.21 | 0.80 | 0 | **2.01** |
| 935 | 2.01 | 3.21 | 2.01 | 2.01 | **9.24** |
| 936 | 0.40 | 1.21 | 0 | 0 | **1.61** |
| 937 | 1.21 | 1.61 | 2.41 | 1.61 | **6.83** |
| 939 | 1.21 | 0.40 | 0 | 0 | **1.61** |
| 940 | 4.82 | 4.82 | 6.43 | 3.62 | **19.69** |
| 941 | 4.42 | 6.43 | 6.03 | 6.03 | **22.90** |
| 943 | 0 | 0.40 | 2.01 | 0.40 | **2.81** |
| 945 | 5.63 | 4.82 | 2.81 | 8.04 | **21.29** |
| 946 | 1.21 | 2.41 | 2.41 | 0.80 | **6.83** |
| 949 | 2.01 | 2.81 | 4.02 | 3.21 | **12.06** |
| 950 | 8.03 | 5.62 | 8.44 | 9.64 | **31.74** |
| 951 | 6.03 | 6.83 | 8.04 | 10.45 | **31.34** |
| 952 | 0 | 0 | 1.21 | 0 | **1.21** |
| 953 | 4.42 | 4.82 | 2.01 | 4.02 | **15.27** |
| 954 | 0 | 0 | 3.62 | 0 | **3.62** |

| | | | | | |
|---|---|---|---|---|---|
| 955 | 1.61 | 0.80 | 0.40 | 1.61 | **4.42** |
| 956 | 8.03 | 8.44 | 8.03 | 8.03 | **32.54** |
| 957 | 4.42 | 4.42 | 2.01 | 2.01 | **12.86** |
| 958 | 0 | 0 | 0.80 | 0 | **0.80** |
| 959 | 1.61 | 0 | 0 | 0 | **1.61** |
| 960 | 3.21 | 2.41 | 1.61 | 0.40 | **7.63** |
| 961 | 0 | 0.40 | 0.40 | 0.80 | **1.61** |
| **CA - CALIFORNIA - Total** | **230.21** | **229.00** | **224.18** | **218.16** | **901.54** |

**STATE: CO - COLORADO**

| REGISTRANT ZIP CODE 3 | 1ST QUARTER | 2ND QUARTER | 3RD QUARTER | 4TH QUARTER | TOTAL GRAMS |
|---|---|---|---|---|---|
| 800 | 5.62 | 8.44 | 6.03 | 14.46 | **34.55** |
| 801 | 14.06 | 10.04 | 13.26 | 11.65 | **49.01** |
| 802 | 9.64 | 10.85 | 13.66 | 13.66 | **47.81** |
| 803 | 0 | 0.80 | 0 | 0 | **0.80** |
| 804 | 1.61 | 3.62 | 2.41 | 1.61 | **9.24** |
| 805 | 0.40 | 0.80 | 0.80 | 0.40 | **2.41** |
| 806 | 0.40 | 0 | 0 | 0 | **0.40** |
| 808 | 2.01 | 3.21 | 0.80 | 1.21 | **7.23** |
| 809 | 3.62 | 3.21 | 3.21 | 1.61 | **11.65** |
| 810 | 0.40 | 0.40 | 0.40 | 0.40 | **1.61** |
| **CO - COLORADO - Total** | **37.76** | **41.38** | **40.58** | **44.99** | **164.72** |

**STATE: CT - CONNECTICUT**

| REGISTRANT ZIP CODE 3 | 1ST QUARTER | 2ND QUARTER | 3RD QUARTER | 4TH QUARTER | TOTAL GRAMS |
|---|---|---|---|---|---|
| 060 | 5.62 | 6.43 | 8.44 | 10.04 | **30.53** |
| 061 | 2.01 | 2.41 | 2.01 | 0.80 | **7.23** |
| 064 | 0 | 2.01 | 0 | 1.21 | **3.21** |
| 065 | 2.01 | 2.01 | 2.41 | 2.41 | **8.84** |
| 067 | 2.01 | 2.01 | 3.21 | 1.61 | **8.84** |
| 068 | 4.02 | 6.03 | 4.82 | 5.22 | **20.09** |
| 069 | 0 | 1.21 | 1.21 | 1.61 | **4.02** |
| **CT - CONNECTICUT - Total** | **15.67** | **22.10** | **22.10** | **22.90** | **82.76** |

**STATE: DC - DISTRICT OF COLUMBIA**

| REGISTRANT ZIP CODE 3 | 1ST QUARTER | 2ND QUARTER | 3RD QUARTER | 4TH QUARTER | TOTAL GRAMS |
|---|---|---|---|---|---|
| 200 | 4.82 | 6.43 | 5.22 | 10.45 | **26.92** |
| **DC - DISTRICT OF COLUMBIA - Total** | **4.82** | **6.43** | **5.22** | **10.45** | **26.92** |

**STATE: DE - DELAWARE**

| REGISTRANT ZIP CODE 3 | 1ST QUARTER | 2ND QUARTER | 3RD QUARTER | 4TH QUARTER | TOTAL GRAMS |
|---|---|---|---|---|---|
| 197 | 0.80 | 1.21 | 0.80 | 0.40 | **3.21** |
| 198 | 2.41 | 3.62 | 3.21 | 0 | **9.24** |
| 199 | 2.01 | 0.80 | 2.81 | 3.21 | **8.84** |
| **DE - DELAWARE - Total** | **5.22** | **5.62** | **6.83** | **3.62** | **21.29** |

**STATE: FL - FLORIDA**

| REGISTRANT ZIP CODE 3 | 1ST QUARTER | 2ND QUARTER | 3RD QUARTER | 4TH QUARTER | TOTAL GRAMS |
|---|---|---|---|---|---|
| 320 | 1.21 | 0 | 0 | 1.21 | **2.41** |
| 321 | 1.21 | 1.61 | 1.21 | 0.80 | **4.82** |
| 322 | 3.62 | 6.43 | 5.22 | 3.62 | **18.88** |
| 323 | 0.80 | 0.80 | 0.80 | 0 | **2.41** |
| 324 | 0.40 | 1.21 | 0 | 1.61 | **3.21** |
| 325 | 3.62 | 2.41 | 1.61 | 3.62 | **11.25** |
| 326 | 1.61 | 0.40 | 0.40 | 2.81 | **5.22** |
| 327 | 10.85 | 14.06 | 12.06 | 10.05 | **47.01** |
| 328 | 6.03 | 4.02 | 4.82 | 4.82 | **19.68** |
| 329 | 4.42 | 4.42 | 4.02 | 7.23 | **20.09** |
| 330 | 1.21 | 0 | 0.40 | 1.21 | **2.81** |
| 331 | 3.21 | 0.80 | 3.21 | 2.01 | **9.24** |
| 333 | 0 | 0 | 1.21 | 0.40 | **1.61** |
| 334 | 0.80 | 0.40 | 0.40 | 0.40 | **2.01** |
| 335 | 0.40 | 0 | 2.01 | 2.41 | **4.82** |
| 336 | 1.61 | 2.01 | 2.81 | 1.21 | **7.63** |
| 337 | 0 | 0 | 1.61 | 0.40 | **2.01** |

| | | | | | |
|---|---|---|---|---|---|
| 338 | 0.40 | 0.40 | 0.40 | 0 | **1.21** |
| 339 | 4.02 | 4.42 | 2.41 | 2.41 | **13.26** |
| 341 | 0.80 | 0 | 0 | 0 | **0.80** |
| 342 | 4.42 | 4.02 | 3.62 | 3.21 | **15.27** |
| 344 | 0 | 0.80 | 0.80 | 0.80 | **2.41** |
| 346 | 0.80 | 1.61 | 1.61 | 0.80 | **4.82** |
| 349 | 0 | 1.21 | 1.61 | 1.21 | **4.02** |
| **FL - FLORIDA - Total** | **51.43** | **51.03** | **52.23** | **52.23** | **206.91** |

**STATE: GA - GEORGIA**

| REGISTRANT ZIP CODE 3 | 1ST QUARTER | 2ND QUARTER | 3RD QUARTER | 4TH QUARTER | TOTAL GRAMS |
|---|---|---|---|---|---|
| 300 | 8.04 | 5.63 | 7.63 | 4.82 | **26.12** |
| 301 | 4.02 | 4.82 | 5.22 | 6.03 | **20.09** |
| 302 | 1.61 | 0 | 0 | 0 | **1.61** |
| 303 | 8.44 | 8.44 | 7.23 | 7.23 | **31.34** |
| 304 | 2.81 | 2.41 | 2.81 | 3.62 | **11.65** |
| 305 | 0 | 0 | 0.80 | 0 | **0.80** |
| 315 | 2.01 | 2.81 | 1.21 | 1.61 | **7.63** |
| 317 | 1.61 | 0.40 | 0.80 | 1.21 | **4.02** |
| **GA - GEORGIA - Total** | **28.52** | **24.51** | **25.71** | **24.51** | **103.25** |

**STATE: HI - HAWAII**

| REGISTRANT ZIP CODE 3 | 1ST QUARTER | 2ND QUARTER | 3RD QUARTER | 4TH QUARTER | TOTAL GRAMS |
|---|---|---|---|---|---|
| 967 | 1.61 | 1.61 | 0.80 | 0.80 | **4.82** |
| 968 | 2.41 | 2.41 | 1.61 | 2.41 | **8.84** |
| **HI - HAWAII - Total** | **4.02** | **4.02** | **2.41** | **3.21** | **13.66** |

**STATE: IA - IOWA**

| REGISTRANT ZIP CODE 3 | 1ST QUARTER | 2ND QUARTER | 3RD QUARTER | 4TH QUARTER | TOTAL GRAMS |
|---|---|---|---|---|---|
| 500 | 0.40 | 1.61 | 0 | 0.40 | **2.41** |
| 502 | 5.22 | 5.62 | 7.63 | 10.45 | **28.92** |
| 503 | 1.61 | 1.21 | 2.01 | 0.80 | **5.62** |
| 504 | 2.01 | 2.01 | 2.01 | 2.01 | **8.04** |
| 505 | 0.40 | 1.21 | 0.40 | 0.40 | **2.41** |
| 511 | 0 | 0.40 | 0 | 0 | **0.40** |
| 515 | 0.40 | 0.80 | 1.21 | 0.40 | **2.81** |
| 516 | 0 | 1.61 | 1.21 | 0.80 | **3.62** |
| 528 | 0.40 | 2.41 | 2.01 | 1.21 | **6.03** |
| **IA - IOWA - Total** | **10.44** | **16.87** | **16.47** | **16.47** | **60.26** |

**STATE: ID - IDAHO**

| REGISTRANT ZIP CODE 3 | 1ST QUARTER | 2ND QUARTER | 3RD QUARTER | 4TH QUARTER | TOTAL GRAMS |
|---|---|---|---|---|---|
| 832 | 0 | 0 | 0.40 | 2.01 | **2.41** |
| 836 | 5.62 | 6.43 | 8.44 | 6.43 | **26.91** |
| 837 | 1.21 | 0.80 | 1.61 | 0.80 | **4.42** |
| 838 | 4.42 | 5.22 | 2.81 | 4.82 | **17.28** |
| **ID - IDAHO - Total** | **11.25** | **12.46** | **13.26** | **14.06** | **51.02** |

**STATE: IL - ILLINOIS**

| REGISTRANT ZIP CODE 3 | 1ST QUARTER | 2ND QUARTER | 3RD QUARTER | 4TH QUARTER | TOTAL GRAMS |
|---|---|---|---|---|---|
| 600 | 15.27 | 12.85 | 18.48 | 13.26 | **59.85** |
| 601 | 5.22 | 6.43 | 5.22 | 3.62 | **20.49** |
| 602 | 1.21 | 0.40 | 2.41 | 1.61 | **5.62** |
| 604 | 4.42 | 0.40 | 2.01 | 2.81 | **9.64** |
| 605 | 5.22 | 0.40 | 2.01 | 1.61 | **9.24** |
| 606 | 7.23 | 4.42 | 4.42 | 4.02 | **20.09** |
| 607 | 4.02 | 2.01 | 2.41 | 0 | **8.44** |
| 611 | 0.80 | 1.61 | 1.61 | 2.01 | **6.03** |
| 615 | 0 | 0 | 0.40 | 0 | **0.40** |
| 618 | 0 | 0 | 0 | 0.80 | **0.80** |
| 623 | 0.80 | 0.40 | 0.80 | 0.40 | **2.41** |
| 625 | 0 | 0 | 0 | 0.40 | **0.40** |
| 627 | 0.40 | 1.61 | 1.21 | 1.21 | **4.42** |
| **IL - ILLINOIS - Total** | **44.59** | **30.53** | **40.98** | **31.74** | **147.83** |

**STATE: IN - INDIANA**

| REGISTRANT ZIP CODE 3 | 1ST QUARTER | 2ND QUARTER | 3RD QUARTER | 4TH QUARTER | TOTAL GRAMS |
|---|---|---|---|---|---|
| 460 | 14.46 | 18.88 | 11.65 | 4.82 | **49.82** |
| 461 | 0 | 0 | 0 | 0.40 | **0.40** |
| 462 | 0.80 | 1.21 | 2.01 | 0 | **4.02** |
| 463 | 5.22 | 4.02 | 2.41 | 2.81 | **14.46** |
| 465 | 2.41 | 2.41 | 0 | 0 | **4.82** |
| 466 | 0 | 0 | 0 | 1.21 | **1.21** |
| 468 | 2.01 | 1.21 | 0 | 0 | **3.21** |
| 469 | 0 | 0.80 | 0.80 | 0 | **1.61** |
| 471 | 0.40 | 2.01 | 0.80 | 2.41 | **5.62** |
| 473 | 0 | 0 | 0.40 | 0 | **0.40** |
| 474 | 2.41 | 3.21 | 2.81 | 2.41 | **10.85** |
| 477 | 0 | 2.41 | 3.21 | 3.21 | **8.84** |
| 478 | 0 | 0 | 0 | 0.40 | **0.40** |
| **IN - INDIANA - Total** | **27.72** | **36.16** | **24.11** | **17.68** | **105.66** |

**STATE: KS - KANSAS**

| REGISTRANT ZIP CODE 3 | 1ST QUARTER | 2ND QUARTER | 3RD QUARTER | 4TH QUARTER | TOTAL GRAMS |
|---|---|---|---|---|---|
| 660 | 5.22 | 2.41 | 3.62 | 3.21 | **14.46** |
| 662 | 0.80 | 0 | 0 | 0 | **0.80** |
| 666 | 0.80 | 1.21 | 0.40 | 1.21 | **3.62** |
| 668 | 0 | 0.40 | 0.40 | 0 | **0.80** |
| 670 | 0.80 | 0.80 | 0.40 | 0 | **2.01** |
| 671 | 0.40 | 0 | 0 | 0 | **0.40** |
| 672 | 5.22 | 4.82 | 5.62 | 4.42 | **20.09** |
| **KS - KANSAS - Total** | **13.26** | **9.64** | **10.44** | **8.84** | **42.18** |

**STATE: KY - KENTUCKY**

| REGISTRANT ZIP CODE 3 | 1ST QUARTER | 2ND QUARTER | 3RD QUARTER | 4TH QUARTER | TOTAL GRAMS |
|---|---|---|---|---|---|
| 400 | 0.40 | 0 | 0 | 0 | **0.40** |
| 402 | 0.80 | 1.21 | 2.01 | 2.41 | **6.43** |
| 403 | 0.40 | 0.80 | 0 | 0 | **1.21** |
| 404 | 2.01 | 0.40 | 0.80 | 0 | **3.21** |
| 405 | 2.01 | 4.42 | 4.42 | 4.02 | **14.86** |
| 408 | 3.21 | 1.61 | 1.61 | 1.61 | **8.04** |
| 410 | 1.21 | 0.40 | 0.40 | 0.80 | **2.81** |
| 417 | 0.40 | 0 | 0 | 0 | **0.40** |
| 424 | 0 | 0.80 | 0 | 0.40 | **1.21** |
| 427 | 0 | 0 | 0 | 0.80 | **0.80** |
| **KY - KENTUCKY - Total** | **10.45** | **9.64** | **9.24** | **10.04** | **39.37** |

**STATE: LA - LOUISIANA**

| REGISTRANT ZIP CODE 3 | 1ST QUARTER | 2ND QUARTER | 3RD QUARTER | 4TH QUARTER | TOTAL GRAMS |
|---|---|---|---|---|---|
| 701 | 0 | 0.40 | 0.40 | 0.40 | **1.21** |
| 703 | 0.80 | 0.80 | 1.21 | 1.61 | **4.42** |
| 704 | 0 | 0 | 0 | 0.80 | **0.80** |
| 705 | 0 | 0 | 0 | 0.40 | **0.40** |
| 706 | 1.61 | 0.40 | 0.80 | 1.21 | **4.02** |
| 707 | 0.40 | 0 | 0 | 0 | **0.40** |
| 708 | 4.42 | 7.23 | 4.82 | 4.02 | **20.49** |
| 711 | 3.21 | 0.80 | 6.83 | 4.02 | **14.87** |
| 712 | 0.40 | 1.21 | 1.61 | 0.80 | **4.02** |
| 713 | 0.40 | 1.21 | 0.80 | 1.21 | **3.62** |
| **LA - LOUISIANA - Total** | **11.25** | **12.05** | **16.47** | **14.46** | **54.24** |

**STATE: MA - MASSACHUSETTS**

| REGISTRANT ZIP CODE 3 | 1ST QUARTER | 2ND QUARTER | 3RD QUARTER | 4TH QUARTER | TOTAL GRAMS |
|---|---|---|---|---|---|
| 010 | 0.80 | 0.80 | 1.61 | 0.80 | **4.02** |
| 012 | 0 | 0 | 0 | 0.40 | **0.40** |
| 014 | 1.61 | 1.61 | 2.41 | 0.80 | **6.43** |
| 016 | 0 | 0 | 0.80 | 0 | **0.80** |
| 017 | 3.21 | 3.21 | 3.21 | 3.21 | **12.85** |
| 018 | 5.22 | 4.02 | 4.42 | 1.61 | **15.27** |
| 019 | 4.42 | 1.61 | 2.41 | 1.21 | **9.64** |

| | | | | | |
|---|---|---|---|---|---|
| 021 | 4.82 | 4.42 | 8.44 | 6.43 | **24.11** |
| 023 | 0.80 | 0 | 2.01 | 0.40 | **3.21** |
| 024 | 2.41 | 0.40 | 1.21 | 0 | **4.02** |
| 026 | 0 | 0 | 0 | 0.40 | **0.40** |
| 027 | 2.81 | 2.41 | 1.61 | 2.81 | **9.64** |
| **MA - MASSACHUSETTS - Total** | **26.12** | **18.48** | **28.12** | **18.08** | **90.80** |

**STATE: MD - MARYLAND**

| REGISTRANT ZIP CODE 3 | 1ST QUARTER | 2ND QUARTER | 3RD QUARTER | 4TH QUARTER | TOTAL GRAMS |
|---|---|---|---|---|---|
| 206 | 0.80 | 0 | 0 | 0 | **0.80** |
| 207 | 1.21 | 4.82 | 4.02 | 3.21 | **13.26** |
| 208 | 7.23 | 6.43 | 3.21 | 9.64 | **26.52** |
| 209 | 2.41 | 1.61 | 3.21 | 1.61 | **8.84** |
| 210 | 4.42 | 4.02 | 4.42 | 2.81 | **15.67** |
| 211 | 12.05 | 12.05 | 13.26 | 11.25 | **48.61** |
| 212 | 2.41 | 2.81 | 4.82 | 0.80 | **10.85** |
| 214 | 0.40 | 1.21 | 0.40 | 0.40 | **2.41** |
| 215 | 0 | 0 | 0 | 0.40 | **0.40** |
| 216 | 0 | 0 | 0.40 | 3.21 | **3.62** |
| 217 | 0.80 | 1.21 | 0.80 | 2.01 | **4.82** |
| **MD - MARYLAND - Total** | **31.74** | **34.15** | **34.55** | **35.36** | **135.80** |

**STATE: ME - MAINE**

| REGISTRANT ZIP CODE 3 | 1ST QUARTER | 2ND QUARTER | 3RD QUARTER | 4TH QUARTER | TOTAL GRAMS |
|---|---|---|---|---|---|
| 040 | 0.40 | 1.21 | 1.61 | 1.21 | **4.42** |
| 042 | 1.21 | 3.62 | 2.01 | 3.21 | **10.04** |
| 044 | 0 | 2.01 | 0 | 0 | **2.01** |
| 045 | 6.83 | 2.81 | 6.83 | 2.81 | **19.29** |
| 046 | 0 | 0 | 0 | 0.40 | **0.40** |
| **ME - MAINE - Total** | **8.44** | **9.64** | **10.45** | **7.63** | **36.16** |

**STATE: MI - MICHIGAN**

| REGISTRANT ZIP CODE 3 | 1ST QUARTER | 2ND QUARTER | 3RD QUARTER | 4TH QUARTER | TOTAL GRAMS |
|---|---|---|---|---|---|
| 480 | 0.80 | 1.21 | 0 | 0 | **2.01** |
| 481 | 2.81 | 1.21 | 1.61 | 0 | **5.62** |
| 483 | 3.21 | 1.61 | 0.40 | 0 | **5.22** |
| 484 | 0 | 0 | 1.21 | 2.41 | **3.62** |
| 485 | 0 | 0.40 | 0 | 0 | **0.40** |
| 486 | 4.02 | 6.03 | 2.81 | 8.44 | **21.30** |
| 487 | 0 | 0 | 1.61 | 0 | **1.61** |
| 488 | 1.21 | 0 | 0 | 0.40 | **1.61** |
| 490 | 4.82 | 3.21 | 5.22 | 3.21 | **16.47** |
| 494 | 1.21 | 0 | 0.80 | 0 | **2.01** |
| **MI - MICHIGAN - Total** | **18.08** | **13.66** | **13.66** | **14.46** | **59.86** |

**STATE: MN - MINNESOTA**

| REGISTRANT ZIP CODE 3 | 1ST QUARTER | 2ND QUARTER | 3RD QUARTER | 4TH QUARTER | TOTAL GRAMS |
|---|---|---|---|---|---|
| 550 | 1.21 | 4.02 | 3.21 | 2.81 | **11.25** |
| 551 | 7.23 | 6.43 | 8.84 | 9.64 | **32.14** |
| 553 | 4.82 | 9.24 | 5.63 | 7.63 | **27.32** |
| 554 | 25.31 | 12.86 | 22.50 | 18.88 | **79.55** |
| 557 | 2.41 | 6.43 | 5.62 | 6.03 | **20.49** |
| 559 | 5.62 | 4.82 | 1.21 | 5.22 | **16.87** |
| 560 | 0.80 | 1.61 | 0.40 | 2.41 | **5.22** |
| 563 | 0 | 1.21 | 0 | 0 | **1.21** |
| 565 | 0.40 | 0.40 | 0.40 | 0.80 | **2.01** |
| **MN - MINNESOTA - Total** | **47.81** | **47.01** | **47.81** | **53.43** | **196.06** |

**STATE: MO - MISSOURI**

| REGISTRANT ZIP CODE 3 | 1ST QUARTER | 2ND QUARTER | 3RD QUARTER | 4TH QUARTER | TOTAL GRAMS |
|---|---|---|---|---|---|
| 631 | 32.14 | 39.37 | 40.57 | 51.42 | **163.50** |
| 633 | 0 | 0.80 | 0 | 0 | **0.80** |
| 637 | 0 | 1.61 | 2.01 | 0.40 | **4.02** |
| 640 | 3.21 | 0 | 3.21 | 0.40 | **6.83** |
| 645 | 1.21 | 1.21 | 2.01 | 1.61 | **6.03** |

| | | | | | |
|---|---|---|---|---|---|
| 647 | 0.40 | 0 | 0 | 0 | **0.40** |
| 652 | 2.01 | 0.80 | 2.41 | 2.41 | **7.63** |
| 656 | 0.40 | 0 | 0 | 0.80 | **1.21** |
| **MO - MISSOURI - Total** | **39.37** | **43.79** | **50.22** | **57.04** | **190.42** |

**STATE: MS - MISSISSIPPI**

| REGISTRANT ZIP CODE 3 | 1ST QUARTER | 2ND QUARTER | 3RD QUARTER | 4TH QUARTER | TOTAL GRAMS |
|---|---|---|---|---|---|
| 386 | 0.40 | 0 | 0 | 0 | **0.40** |
| 390 | 0.80 | 0.40 | 1.21 | 1.21 | **3.62** |
| 391 | 4.02 | 7.63 | 6.03 | 6.03 | **23.70** |
| 394 | 0 | 1.21 | 0 | 0 | **1.21** |
| **MS - MISSISSIPPI - Total** | **5.22** | **9.24** | **7.23** | **7.23** | **28.92** |

**STATE: MT - MONTANA**

| REGISTRANT ZIP CODE 3 | 1ST QUARTER | 2ND QUARTER | 3RD QUARTER | 4TH QUARTER | TOTAL GRAMS |
|---|---|---|---|---|---|
| 591 | 0 | 0 | 2.01 | 0 | **2.01** |
| **MT - MONTANA - Total** | **0.00** | **0.00** | **2.01** | **0.00** | **2.01** |

**STATE: NC - NORTH CAROLINA**

| REGISTRANT ZIP CODE 3 | 1ST QUARTER | 2ND QUARTER | 3RD QUARTER | 4TH QUARTER | TOTAL GRAMS |
|---|---|---|---|---|---|
| 272 | 0 | 0.40 | 0.80 | 0.80 | **2.01** |
| 273 | 0.80 | 0 | 0 | 0 | **0.80** |
| 274 | 0.80 | 1.61 | 1.21 | 2.01 | **5.63** |
| 275 | 9.24 | 8.03 | 7.63 | 9.24 | **34.15** |
| 276 | 7.23 | 4.02 | 3.21 | 2.01 | **16.47** |
| 277 | 2.41 | 2.01 | 2.01 | 2.41 | **8.84** |
| 278 | 2.41 | 4.82 | 1.21 | 0.80 | **9.24** |
| 281 | 7.63 | 4.42 | 4.02 | 4.82 | **20.89** |
| 282 | 6.03 | 3.62 | 5.62 | 3.21 | **18.48** |
| 283 | 0 | 0 | 1.21 | 2.01 | **3.21** |
| 285 | 7.23 | 4.42 | 0 | 0 | **11.65** |
| 287 | 2.41 | 0.40 | 0.80 | 1.21 | **4.82** |
| 288 | 2.41 | 0.80 | 1.61 | 2.01 | **6.83** |
| **NC - NORTH CAROLINA - Total** | **48.61** | **34.55** | **29.33** | **30.53** | **143.02** |

**STATE: ND - NORTH DAKOTA**

| REGISTRANT ZIP CODE 3 | 1ST QUARTER | 2ND QUARTER | 3RD QUARTER | 4TH QUARTER | TOTAL GRAMS |
|---|---|---|---|---|---|
| 580 | 0.40 | 1.21 | 0.80 | 0 | **2.41** |
| 581 | 2.81 | 2.81 | 2.01 | 2.41 | **10.04** |
| 585 | 0 | 2.01 | 1.61 | 0.40 | **4.02** |
| **ND - NORTH DAKOTA - Total** | **3.21** | **6.03** | **4.42** | **2.81** | **16.47** |

**STATE: NE - NEBRASKA**

| REGISTRANT ZIP CODE 3 | 1ST QUARTER | 2ND QUARTER | 3RD QUARTER | 4TH QUARTER | TOTAL GRAMS |
|---|---|---|---|---|---|
| 681 | 0 | 1.61 | 0 | 0.40 | **2.01** |
| 684 | 2.41 | 2.01 | 1.61 | 0 | **6.03** |
| 685 | 1.61 | 3.21 | 4.02 | 0.80 | **9.64** |
| 687 | 2.81 | 2.01 | 0 | 0 | **4.82** |
| **NE - NEBRASKA - Total** | **6.83** | **8.84** | **5.62** | **1.21** | **22.50** |

**STATE: NH - NEW HAMPSHIRE**

| REGISTRANT ZIP CODE 3 | 1ST QUARTER | 2ND QUARTER | 3RD QUARTER | 4TH QUARTER | TOTAL GRAMS |
|---|---|---|---|---|---|
| 031 | 4.82 | 3.21 | 3.62 | 4.02 | **15.67** |
| 037 | 2.81 | 2.01 | 2.81 | 1.21 | **8.84** |
| **NH - NEW HAMPSHIRE - Total** | **7.63** | **5.22** | **6.43** | **5.22** | **24.51** |

**STATE: NJ - NEW JERSEY**

| REGISTRANT ZIP CODE 3 | 1ST QUARTER | 2ND QUARTER | 3RD QUARTER | 4TH QUARTER | TOTAL GRAMS |
|---|---|---|---|---|---|
| 070 | 4.82 | 4.02 | 3.62 | 3.62 | **16.07** |
| 074 | 2.41 | 0.40 | 0.40 | 1.21 | **4.42** |
| 076 | 11.65 | 5.63 | 4.82 | 7.23 | **29.33** |
| 077 | 0 | 1.21 | 0.40 | 0.40 | **2.01** |
| 078 | 2.81 | 4.02 | 4.02 | 2.01 | **12.86** |
| 079 | 2.01 | 1.21 | 0.80 | 1.61 | **5.63** |
| 080 | 5.22 | 4.82 | 5.22 | 4.82 | **20.09** |
| 081 | 0.80 | 2.01 | 0 | 0.80 | **3.62** |
| 082 | 0.40 | 0 | 0 | 0.80 | **1.21** |

| | | | | | |
|---|---|---|---|---|---|
| 085 | 1.21 | 0.80 | 0.80 | 0.40 | **3.21** |
| 086 | 0.80 | 0 | 0 | 0 | **0.80** |
| 087 | 3.21 | 2.01 | 1.61 | 2.01 | **8.84** |
| 088 | 0.80 | 0.80 | 0.80 | 1.21 | **3.62** |
| 089 | 0 | 1.21 | 3.62 | 1.61 | **6.43** |
| **NJ - NEW JERSEY - Total** | **36.16** | **28.12** | **26.11** | **27.72** | **118.12** |

**STATE: NM - NEW MEXICO**

| REGISTRANT ZIP CODE 3 | 1ST QUARTER | 2ND QUARTER | 3RD QUARTER | 4TH QUARTER | TOTAL GRAMS |
|---|---|---|---|---|---|
| 870 | 0.40 | 0 | 0 | 0 | **0.40** |
| 871 | 1.21 | 4.02 | 5.22 | 3.62 | **14.06** |
| 875 | 0 | 0.80 | 0 | 0.80 | **1.61** |
| 880 | 3.62 | 2.81 | 4.82 | 4.42 | **15.67** |
| 882 | 0 | 0.80 | 0 | 0.80 | **1.61** |
| 883 | 0.80 | 0.80 | 0.80 | 0.80 | **3.21** |
| **NM - NEW MEXICO - Total** | **6.03** | **9.24** | **10.85** | **10.45** | **36.56** |

**STATE: NV - NEVADA**

| REGISTRANT ZIP CODE 3 | 1ST QUARTER | 2ND QUARTER | 3RD QUARTER | 4TH QUARTER | TOTAL GRAMS |
|---|---|---|---|---|---|
| 891 | 4.02 | 8.44 | 8.44 | 6.83 | **27.72** |
| 894 | 0.80 | 1.21 | 0 | 0 | **2.01** |
| 895 | 1.61 | 0.40 | 0.40 | 1.61 | **4.02** |
| **NV - NEVADA - Total** | **6.43** | **10.04** | **8.84** | **8.44** | **33.75** |

**STATE: NY - NEW YORK**

| REGISTRANT ZIP CODE 3 | 1ST QUARTER | 2ND QUARTER | 3RD QUARTER | 4TH QUARTER | TOTAL GRAMS |
|---|---|---|---|---|---|
| 100 | 20.49 | 18.08 | 20.49 | 21.69 | **80.75** |
| 101 | 0.40 | 0 | 0 | 0.40 | **0.80** |
| 104 | 1.21 | 0.40 | 0.80 | 0 | **2.41** |
| 105 | 1.21 | 2.41 | 5.22 | 2.01 | **10.85** |
| 106 | 5.62 | 6.03 | 2.81 | 2.81 | **17.28** |
| 107 | 0.80 | 1.21 | 0.40 | 0 | **2.41** |
| 109 | 2.41 | 1.21 | 2.01 | 1.21 | **6.83** |
| 110 | 1.61 | 2.81 | 7.23 | 8.03 | **19.68** |
| 111 | 0 | 0.80 | 1.61 | 2.81 | **5.22** |
| 112 | 3.21 | 3.21 | 1.61 | 2.81 | **10.85** |
| 113 | 2.41 | 2.81 | 4.02 | 3.21 | **12.45** |
| 114 | 2.81 | 1.21 | 1.21 | 0.80 | **6.03** |
| 115 | 3.21 | 2.01 | 1.61 | 1.21 | **8.03** |
| 116 | 0.80 | 1.61 | 0.80 | 1.21 | **4.42** |
| 117 | 12.05 | 16.88 | 9.64 | 8.84 | **47.41** |
| 120 | 2.81 | 4.42 | 4.82 | 2.41 | **14.47** |
| 122 | 1.21 | 0 | 0.40 | 2.81 | **4.42** |
| 123 | 3.21 | 4.42 | 3.21 | 4.42 | **15.27** |
| 125 | 1.21 | 4.02 | 4.02 | 0.80 | **10.04** |
| 126 | 0 | 0.80 | 0 | 0.40 | **1.21** |
| 127 | 1.21 | 0.40 | 0.80 | 0 | **2.41** |
| 128 | 0 | 0 | 0 | 0.40 | **0.40** |
| 131 | 0 | 0.40 | 0.40 | 0 | **0.80** |
| 132 | 0.40 | 0 | 0 | 0 | **0.40** |
| 139 | 1.21 | 1.21 | 2.01 | 1.61 | **6.03** |
| 140 | 5.62 | 4.82 | 4.82 | 3.62 | **18.88** |
| 145 | 0 | 0.40 | 0 | 0 | **0.40** |
| 146 | 2.41 | 2.01 | 2.01 | 2.01 | **8.44** |
| 148 | 2.41 | 3.21 | 2.01 | 2.41 | **10.04** |
| 149 | 1.21 | 0.40 | 2.01 | 0.80 | **4.42** |
| **NY - NEW YORK - Total** | **81.15** | **87.18** | **85.98** | **78.74** | **333.06** |

**STATE: OH - OHIO**

| REGISTRANT ZIP CODE 3 | 1ST QUARTER | 2ND QUARTER | 3RD QUARTER | 4TH QUARTER | TOTAL GRAMS |
|---|---|---|---|---|---|
| 430 | 3.62 | 3.21 | 3.21 | 4.02 | **14.06** |
| 431 | 0.80 | 1.61 | 3.21 | 2.01 | **7.63** |
| 432 | 2.81 | 2.41 | 3.62 | 3.62 | **12.46** |
| 434 | 1.61 | 0.80 | 0.80 | 0.80 | **4.02** |

| | | | | | |
|---|---|---|---|---|---|
| 440 | 0 | 0.80 | 0 | 0 | **0.80** |
| 441 | 1.21 | 2.81 | 1.21 | 0 | **5.22** |
| 442 | 0.40 | 1.21 | 1.21 | 2.01 | **4.82** |
| 444 | 0 | 2.41 | 1.21 | 2.41 | **6.03** |
| 445 | 3.21 | 3.62 | 2.81 | 6.03 | **15.67** |
| 447 | 0 | 3.21 | 5.62 | 0 | **8.84** |
| 448 | 0 | 1.21 | 1.21 | 0.40 | **2.81** |
| 450 | 4.42 | 3.62 | 0.40 | 2.81 | **11.25** |
| 452 | 2.01 | 1.21 | 1.61 | 2.01 | **6.83** |
| 453 | 2.01 | 0.40 | 1.61 | 0.80 | **4.82** |
| 454 | 6.03 | 2.41 | 4.82 | 2.81 | **16.07** |
| 457 | 0.40 | 0.40 | 0 | 0 | **0.80** |
| **OH - OHIO - Total** | **28.52** | **31.34** | **32.54** | **29.73** | **122.13** |

**STATE: OK - OKLAHOMA**

| REGISTRANT ZIP CODE 3 | 1ST QUARTER | 2ND QUARTER | 3RD QUARTER | 4TH QUARTER | TOTAL GRAMS |
|---|---|---|---|---|---|
| 730 | 1.61 | 2.01 | 2.41 | 2.01 | **8.03** |
| 731 | 0.80 | 0 | 0.40 | 0 | **1.21** |
| 737 | 0.40 | 0 | 0 | 0 | **0.40** |
| 740 | 1.21 | 4.82 | 4.42 | 5.62 | **16.07** |
| 741 | 0 | 0.40 | 1.21 | 1.61 | **3.21** |
| 743 | 1.61 | 4.02 | 2.41 | 2.81 | **10.85** |
| 747 | 0.80 | 0 | 1.21 | 0.40 | **2.41** |
| 748 | 0.40 | 0.80 | 0 | 0 | **1.21** |
| **OK - OKLAHOMA - Total** | **6.83** | **12.05** | **12.05** | **12.45** | **43.39** |

**STATE: OR - OREGON**

| REGISTRANT ZIP CODE 3 | 1ST QUARTER | 2ND QUARTER | 3RD QUARTER | 4TH QUARTER | TOTAL GRAMS |
|---|---|---|---|---|---|
| 970 | 14.46 | 10.04 | 11.25 | 17.67 | **53.43** |
| 971 | 1.61 | 0.40 | 0 | 0 | **2.01** |
| 972 | 15.67 | 14.06 | 18.08 | 14.46 | **62.27** |
| 973 | 1.61 | 1.61 | 1.21 | 2.01 | **6.43** |
| 974 | 3.62 | 2.41 | 2.41 | 1.21 | **9.64** |
| 975 | 2.41 | 2.41 | 1.61 | 3.21 | **9.64** |
| 978 | 8.04 | 12.06 | 0 | 0 | **20.09** |
| **OR - OREGON - Total** | **47.41** | **42.99** | **34.55** | **38.57** | **163.51** |

**STATE: PA - PENNSYLVANIA**

| REGISTRANT ZIP CODE 3 | 1ST QUARTER | 2ND QUARTER | 3RD QUARTER | 4TH QUARTER | TOTAL GRAMS |
|---|---|---|---|---|---|
| 150 | 5.62 | 4.42 | 3.62 | 5.22 | **18.88** |
| 151 | 4.02 | 4.82 | 4.42 | 5.62 | **18.88** |
| 152 | 6.03 | 6.83 | 5.63 | 6.03 | **24.51** |
| 156 | 1.61 | 0.40 | 1.21 | 1.21 | **4.42** |
| 162 | 0 | 0 | 0 | 1.21 | **1.21** |
| 168 | 2.01 | 0 | 0 | 0 | **2.01** |
| 170 | 0.80 | 1.61 | 1.61 | 0.80 | **4.82** |
| 171 | 3.21 | 2.81 | 2.01 | 2.81 | **10.85** |
| 173 | 0 | 0.40 | 0.80 | 0 | **1.21** |
| 174 | 0.40 | 0.80 | 0 | 0 | **1.21** |
| 176 | 0 | 0 | 0.40 | 2.01 | **2.41** |
| 179 | 2.41 | 2.81 | 1.21 | 1.61 | **8.03** |
| 180 | 2.41 | 1.21 | 4.02 | 1.61 | **9.24** |
| 181 | 2.41 | 1.61 | 1.61 | 1.21 | **6.83** |
| 183 | 1.61 | 0 | 0.40 | 0 | **2.01** |
| 184 | 3.62 | 3.62 | 3.62 | 2.41 | **13.26** |
| 185 | 0.80 | 0.80 | 1.61 | 0 | **3.21** |
| 186 | 2.01 | 2.01 | 2.01 | 0.80 | **6.83** |
| 187 | 28.13 | 37.37 | 46.61 | 49.43 | **161.54** |
| 188 | 0 | 0 | 0.80 | 0 | **0.80** |
| 189 | 4.02 | 2.41 | 2.41 | 2.81 | **11.65** |
| 190 | 7.23 | 6.03 | 3.21 | 2.01 | **18.48** |
| 191 | 1.21 | 4.82 | 4.42 | 4.82 | **15.27** |
| 193 | 2.81 | 2.41 | 2.01 | 0 | **7.23** |

| | | | | | |
|---|---|---|---|---|---|
| 194 | 4.82 | 1.61 | 1.61 | 0.80 | **8.84** |
| 195 | 0.40 | 0.80 | 0 | 0 | **1.21** |
| 196 | 0.80 | 0 | 0.80 | 0.80 | **2.41** |
| **PA - PENNSYLVANIA - Total** | **88.39** | **89.60** | **96.03** | **93.22** | **367.23** |
| **STATE: RI - RHODE ISLAND** | | | | | |
| REGISTRANT ZIP CODE 3 | 1ST QUARTER | 2ND QUARTER | 3RD QUARTER | 4TH QUARTER | **TOTAL GRAMS** |
| 028 | 3.62 | 2.41 | 1.61 | 2.41 | **10.04** |
| 029 | 0 | 5.63 | 2.01 | 0 | **7.64** |
| **RI - RHODE ISLAND - Total** | **3.62** | **8.04** | **3.62** | **2.41** | **17.68** |
| **STATE: SC - SOUTH CAROLINA** | | | | | |
| REGISTRANT ZIP CODE 3 | 1ST QUARTER | 2ND QUARTER | 3RD QUARTER | 4TH QUARTER | **TOTAL GRAMS** |
| 290 | 0 | 0 | 0 | 1.21 | **1.21** |
| 291 | 0 | 0 | 0 | 0.40 | **0.40** |
| 292 | 2.81 | 2.01 | 1.21 | 0 | **6.03** |
| 293 | 0 | 0 | 0.80 | 0 | **0.80** |
| 294 | 2.01 | 3.62 | 3.21 | 1.21 | **10.04** |
| 295 | 1.21 | 0.40 | 0 | 0.40 | **2.01** |
| 296 | 0 | 2.81 | 4.02 | 0.40 | **7.23** |
| 298 | 0 | 0 | 0 | 2.41 | **2.41** |
| **SC - SOUTH CAROLINA - Total** | **6.03** | **8.84** | **9.24** | **6.03** | **30.13** |
| **STATE: SD - SOUTH DAKOTA** | | | | | |
| REGISTRANT ZIP CODE 3 | 1ST QUARTER | 2ND QUARTER | 3RD QUARTER | 4TH QUARTER | **TOTAL GRAMS** |
| 571 | 5.62 | 4.02 | 8.44 | 8.84 | **26.91** |
| **SD - SOUTH DAKOTA - Total** | **5.62** | **4.02** | **8.44** | **8.84** | **26.91** |
| **STATE: TN - TENNESSEE** | | | | | |
| REGISTRANT ZIP CODE 3 | 1ST QUARTER | 2ND QUARTER | 3RD QUARTER | 4TH QUARTER | **TOTAL GRAMS** |
| 371 | 0 | 0 | 0 | 0.40 | **0.40** |
| 372 | 3.21 | 4.82 | 3.62 | 4.02 | **15.67** |
| 374 | 0.40 | 0 | 0 | 0.40 | **0.80** |
| 377 | 0.80 | 0.80 | 0.80 | 0 | **2.41** |
| 378 | 0.80 | 0.80 | 3.62 | 1.21 | **6.43** |
| 379 | 1.61 | 2.41 | 1.61 | 1.61 | **7.23** |
| 380 | 0.80 | 1.61 | 2.41 | 1.61 | **6.43** |
| 381 | 5.22 | 4.42 | 5.22 | 6.83 | **21.69** |
| 383 | 0 | 0.40 | 0 | 0 | **0.40** |
| **TN - TENNESSEE - Total** | **12.86** | **15.27** | **17.28** | **16.07** | **61.47** |
| **STATE: TX - TEXAS** | | | | | |
| REGISTRANT ZIP CODE 3 | 1ST QUARTER | 2ND QUARTER | 3RD QUARTER | 4TH QUARTER | **TOTAL GRAMS** |
| 750 | 23.70 | 18.08 | 22.90 | 21.69 | **86.37** |
| 751 | 0 | 1.21 | 1.61 | 1.21 | **4.02** |
| 752 | 16.87 | 10.04 | 8.44 | 10.45 | **45.80** |
| 754 | 0 | 0.80 | 0.40 | 0 | **1.21** |
| 756 | 1.21 | 2.01 | 0 | 0 | **3.21** |
| 757 | 2.01 | 2.41 | 2.01 | 2.01 | **8.44** |
| 760 | 6.03 | 4.82 | 5.62 | 4.82 | **21.29** |
| 761 | 0 | 0 | 0.40 | 0.80 | **1.21** |
| 762 | 3.62 | 2.81 | 2.41 | 5.62 | **14.46** |
| 764 | 0 | 0 | 0.80 | 0.40 | **1.21** |
| 765 | 0 | 0 | 0 | 0.40 | **0.40** |
| 770 | 5.22 | 6.43 | 10.85 | 7.63 | **30.13** |
| 773 | 2.41 | 5.22 | 8.84 | 8.44 | **24.91** |
| 774 | 3.21 | 1.61 | 4.82 | 4.02 | **13.66** |
| 775 | 3.21 | 2.81 | 5.63 | 3.62 | **15.27** |
| 780 | 0.40 | 1.21 | 0.40 | 0 | **2.01** |
| 782 | 3.62 | 5.22 | 5.22 | 6.43 | **20.49** |
| 784 | 2.81 | 0.80 | 0 | 0 | **3.62** |
| 785 | 0.80 | 0.80 | 2.41 | 1.61 | **5.62** |
| 786 | 3.21 | 2.81 | 3.62 | 4.42 | **14.06** |
| 787 | 10.85 | 7.23 | 4.42 | 6.03 | **28.53** |
| 790 | 2.01 | 1.21 | 2.01 | 1.21 | **6.43** |

| | | | | | |
|---|---|---|---|---|---|
| 791 | 0 | 0 | 0 | 0.80 | **0.80** |
| 797 | 1.21 | 0.40 | 2.01 | 0 | **3.62** |
| **TX - TEXAS - Total** | **92.40** | **77.94** | **94.81** | **91.59** | **356.75** |

**STATE: UT - UTAH**

| REGISTRANT ZIP CODE 3 | 1ST QUARTER | 2ND QUARTER | 3RD QUARTER | 4TH QUARTER | TOTAL GRAMS |
|---|---|---|---|---|---|
| 840 | 6.43 | 5.62 | 12.05 | 11.25 | **35.35** |
| 841 | 2.81 | 0.80 | 2.81 | 2.41 | **8.84** |
| 843 | 0 | 0 | 0 | 1.61 | **1.61** |
| 844 | 1.21 | 0.40 | 1.21 | 0 | **2.81** |
| 846 | 8.44 | 10.85 | 6.03 | 6.03 | **31.33** |
| 847 | 5.22 | 5.62 | 8.03 | 8.03 | **26.91** |
| **UT - UTAH - Total** | **24.10** | **23.30** | **30.13** | **29.33** | **106.85** |

**STATE: VA - VIRGINIA**

| REGISTRANT ZIP CODE 3 | 1ST QUARTER | 2ND QUARTER | 3RD QUARTER | 4TH QUARTER | TOTAL GRAMS |
|---|---|---|---|---|---|
| 201 | 1.21 | 3.21 | 0 | 0.40 | **4.82** |
| 221 | 4.82 | 5.62 | 4.02 | 3.21 | **17.68** |
| 222 | 0 | 0 | 0.40 | 0 | **0.40** |
| 223 | 2.41 | 2.41 | 2.41 | 3.21 | **10.45** |
| 231 | 0.80 | 0.40 | 0.40 | 0.80 | **2.41** |
| 232 | 2.01 | 4.82 | 4.02 | 3.62 | **14.47** |
| 233 | 0 | 0.80 | 2.41 | 0.40 | **3.62** |
| 234 | 0.40 | 1.21 | 1.61 | 1.61 | **4.82** |
| 235 | 2.01 | 2.01 | 2.01 | 2.01 | **8.03** |
| 236 | 0 | 0 | 0.40 | 0 | **0.40** |
| 237 | 1.61 | 1.21 | 2.01 | 1.61 | **6.43** |
| 238 | 2.01 | 2.01 | 2.81 | 3.21 | **10.04** |
| 240 | 2.41 | 0.40 | 0.80 | 0.80 | **4.42** |
| 241 | 0.80 | 2.01 | 0.80 | 1.61 | **5.22** |
| 242 | 0.40 | 0.80 | 0.40 | 0 | **1.61** |
| 245 | 0.40 | 1.21 | 1.21 | 1.21 | **4.02** |
| **VA - VIRGINIA - Total** | **21.29** | **28.12** | **25.71** | **23.70** | **98.84** |

**STATE: VT - VERMONT**

| REGISTRANT ZIP CODE 3 | 1ST QUARTER | 2ND QUARTER | 3RD QUARTER | 4TH QUARTER | TOTAL GRAMS |
|---|---|---|---|---|---|
| 054 | 1.21 | 0.40 | 0.40 | 0.80 | **2.81** |
| 056 | 2.01 | 0.80 | 1.21 | 0.80 | **4.82** |
| **VT - VERMONT - Total** | **3.21** | **1.21** | **1.61** | **1.61** | **7.63** |

**STATE: WA - WASHINGTON**

| REGISTRANT ZIP CODE 3 | 1ST QUARTER | 2ND QUARTER | 3RD QUARTER | 4TH QUARTER | TOTAL GRAMS |
|---|---|---|---|---|---|
| 980 | 23.30 | 21.29 | 22.50 | 23.70 | **90.79** |
| 981 | 18.88 | 15.27 | 9.64 | 8.03 | **51.82** |
| 982 | 11.65 | 12.05 | 8.03 | 10.44 | **42.18** |
| 983 | 11.65 | 10.04 | 11.25 | 8.44 | **41.38** |
| 984 | 4.42 | 0.80 | 0.80 | 2.81 | **8.84** |
| 985 | 4.82 | 4.42 | 2.81 | 0 | **12.05** |
| 986 | 6.83 | 5.22 | 8.03 | 8.03 | **28.12** |
| 988 | 0.80 | 2.01 | 0 | 0 | **2.81** |
| 989 | 0 | 0 | 0 | 0.40 | **0.40** |
| 990 | 1.21 | 0.80 | 0.80 | 1.21 | **4.02** |
| 992 | 4.82 | 2.81 | 0.80 | 2.41 | **10.85** |
| 994 | 4.02 | 1.21 | 1.61 | 2.81 | **9.64** |
| **WA - WASHINGTON - Total** | **92.39** | **75.92** | **66.28** | **68.29** | **302.89** |

**STATE: WI - WISCONSIN**

| REGISTRANT ZIP CODE 3 | 1ST QUARTER | 2ND QUARTER | 3RD QUARTER | 4TH QUARTER | TOTAL GRAMS |
|---|---|---|---|---|---|
| 530 | 0 | 0.40 | 0 | 0 | **0.40** |
| 531 | 1.61 | 1.61 | 1.21 | 0 | **4.42** |
| 532 | 0.80 | 3.21 | 2.81 | 2.01 | **8.84** |
| 535 | 0 | 0 | 0 | 0.40 | **0.40** |
| 537 | 6.83 | 6.83 | 8.44 | 2.81 | **24.91** |
| 539 | 0.80 | 0.80 | 0.80 | 1.61 | **4.02** |
| 541 | 0 | 0 | 0 | 0.80 | **0.80** |

| | | | | | |
|---|---|---|---|---|---|
| 542 | 6.83 | 5.63 | 3.62 | 6.83 | **22.90** |
| 547 | 0 | 0 | 0.40 | 0.40 | **0.80** |
| 549 | 3.62 | 3.62 | 3.62 | 4.42 | **15.27** |
| **WI - WISCONSIN - Total** | **20.49** | **22.10** | **20.89** | **19.29** | **82.77** |

**STATE: WV - WEST VIRGINIA**

| REGISTRANT ZIP CODE 3 | 1ST QUARTER | 2ND QUARTER | 3RD QUARTER | 4TH QUARTER | TOTAL GRAMS |
|---|---|---|---|---|---|
| 258 | 1.21 | 0 | 2.01 | 1.21 | **4.42** |
| 261 | 0 | 0.40 | 0 | 0 | **0.40** |
| 265 | 2.01 | 1.21 | 1.61 | 2.01 | **6.83** |
| **WV - WEST VIRGINIA - Total** | **3.21** | **1.61** | **3.62** | **3.21** | **11.65** |

**STATE: WY - WYOMING**

| REGISTRANT ZIP CODE 3 | 1ST QUARTER | 2ND QUARTER | 3RD QUARTER | 4TH QUARTER | TOTAL GRAMS |
|---|---|---|---|---|---|
| 820 | 3.21 | 1.61 | 4.02 | 2.81 | **11.65** |
| 824 | 0.40 | 0 | 0 | 0 | **0.40** |
| 826 | 1.21 | 1.21 | 0.80 | 0.80 | **4.02** |
| 827 | 0.40 | 0.40 | 0.40 | 0.80 | **2.01** |
| 829 | 0 | 0 | 0 | 1.21 | **1.21** |
| 830 | 0.40 | 0 | 0 | 0 | **0.40** |
| **WY - WYOMING - Total** | **5.62** | **3.21** | **5.22** | **5.62** | **19.68** |

**DRUG: 1205 - LISDEXAMFETAMINE**

**STATE: AK - ALASKA**

| REGISTRANT ZIP CODE 3 | 1ST QUARTER | 2ND QUARTER | 3RD QUARTER | 4TH QUARTER | TOTAL GRAMS |
|---|---|---|---|---|---|
| 995 | 1,496.12 | 1,468.37 | 1,414.61 | 1,541.79 | **5,920.90** |
| 996 | 931.32 | 879.87 | 938.26 | 1,009.36 | **3,758.81** |
| 997 | 323.74 | 328.94 | 490.23 | 359.58 | **1,502.48** |
| 998 | 209.85 | 158.98 | 216.79 | 223.15 | **808.76** |
| 999 | 146.84 | 133.54 | 145.68 | 164.76 | **590.82** |
| **AK - ALASKA - Total** | **3,107.87** | **2,969.70** | **3,205.56** | **3,298.64** | **12,581.77** |

**STATE: AL - ALABAMA**

| REGISTRANT ZIP CODE 3 | 1ST QUARTER | 2ND QUARTER | 3RD QUARTER | 4TH QUARTER | TOTAL GRAMS |
|---|---|---|---|---|---|
| 350 | 7,199.08 | 6,193.76 | 6,668.38 | 6,830.25 | **26,891.48** |
| 351 | 4,970.50 | 4,425.36 | 4,510.34 | 4,846.79 | **18,752.99** |
| 352 | 10,265.32 | 8,893.49 | 9,270.99 | 9,621.32 | **38,051.12** |
| 354 | 4,767.59 | 3,696.95 | 4,221.29 | 4,180.24 | **16,866.07** |
| 355 | 1,440.05 | 1,331.94 | 1,314.60 | 1,395.53 | **5,482.12** |
| 356 | 4,384.31 | 4,008.55 | 4,252.50 | 4,452.53 | **17,097.89** |
| 357 | 2,799.74 | 2,554.05 | 2,815.35 | 2,828.07 | **10,997.20** |
| 358 | 3,011.90 | 2,781.82 | 2,893.97 | 3,173.19 | **11,860.88** |
| 359 | 2,780.66 | 2,415.88 | 2,478.31 | 2,713.60 | **10,388.46** |
| 360 | 3,397.49 | 2,818.24 | 3,105.55 | 3,277.83 | **12,599.11** |
| 361 | 2,534.97 | 2,170.19 | 2,115.85 | 2,296.21 | **9,117.22** |
| 362 | 1,853.97 | 1,662.62 | 1,652.21 | 1,696.15 | **6,864.94** |
| 363 | 4,178.51 | 3,810.84 | 4,144.40 | 4,124.17 | **16,257.91** |
| 364 | 990.86 | 857.90 | 1,014.57 | 922.07 | **3,785.40** |
| 365 | 8,076.06 | 7,602.02 | 7,705.49 | 7,737.87 | **31,121.44** |
| 366 | 5,947.49 | 5,339.91 | 5,541.09 | 5,716.83 | **22,545.32** |
| 367 | 631.29 | 486.76 | 505.26 | 552.66 | **2,175.97** |
| 368 | 4,624.80 | 3,655.33 | 4,191.23 | 4,261.75 | **16,733.10** |
| 369 | 129.49 | 109.26 | 105.21 | 124.29 | **468.26** |
| **AL - ALABAMA - Total** | **73,984.08** | **64,814.84** | **68,506.58** | **70,751.35** | **278,056.85** |

**STATE: AR - ARKANSAS**

| REGISTRANT ZIP CODE 3 | 1ST QUARTER | 2ND QUARTER | 3RD QUARTER | 4TH QUARTER | TOTAL GRAMS |
|---|---|---|---|---|---|
| 716 | 1,253.90 | 870.62 | 1,023.24 | 1,077.00 | **4,224.75** |
| 717 | 652.67 | 502.95 | 608.16 | 570.01 | **2,333.79** |
| 718 | 687.94 | 579.83 | 627.24 | 641.69 | **2,536.70** |
| 719 | 1,793.84 | 1,591.51 | 1,651.63 | 1,742.97 | **6,779.96** |
| 720 | 4,375.06 | 3,779.04 | 3,859.97 | 4,287.19 | **16,301.26** |
| 721 | 3,035.03 | 2,623.42 | 2,722.27 | 2,754.65 | **11,135.36** |
| 722 | 3,434.49 | 2,832.69 | 3,131.57 | 3,174.93 | **12,573.68** |
| 723 | 1,035.96 | 783.90 | 857.90 | 931.32 | **3,609.08** |

| | | | | | |
|---|---|---|---|---|---|
| 724 | 2,485.83 | 2,060.93 | 2,216.44 | 2,290.43 | **9,053.62** |
| 725 | 580.41 | 526.07 | 497.17 | 512.77 | **2,116.42** |
| 726 | 768.29 | 685.63 | 721.47 | 674.64 | **2,850.03** |
| 727 | 5,789.67 | 5,036.99 | 5,547.45 | 5,472.29 | **21,846.40** |
| 728 | 1,072.95 | 834.20 | 953.87 | 1,014.57 | **3,875.58** |
| 729 | 2,392.18 | 1,958.60 | 2,020.46 | 2,106.60 | **8,477.84** |
| **AR - ARKANSAS - Total** | **29,358.23** | **24,666.37** | **26,438.83** | **27,251.06** | **107,714.48** |

**STATE: AZ - ARIZONA**

| REGISTRANT ZIP CODE 3 | 1ST QUARTER | 2ND QUARTER | 3RD QUARTER | 4TH QUARTER | TOTAL GRAMS |
|---|---|---|---|---|---|
| 850 | 7,318.75 | 6,844.13 | 7,089.82 | 7,389.85 | **28,642.54** |
| 851 | 1,851.08 | 1,661.46 | 1,685.16 | 1,759.74 | **6,957.43** |
| 852 | 20,408.09 | 19,368.08 | 19,839.81 | 21,280.44 | **80,896.42** |
| 853 | 5,027.74 | 4,590.11 | 4,788.40 | 5,231.23 | **19,637.48** |
| 855 | 153.77 | 145.68 | 145.10 | 177.48 | **622.04** |
| 856 | 661.92 | 616.25 | 668.86 | 644.58 | **2,591.62** |
| 857 | 5,029.47 | 4,761.81 | 4,842.74 | 5,056.64 | **19,690.66** |
| 859 | 213.32 | 252.63 | 255.52 | 254.94 | **976.41** |
| 860 | 482.14 | 430.68 | 448.61 | 506.42 | **1,867.84** |
| 863 | 983.93 | 1,004.74 | 1,021.50 | 1,119.78 | **4,129.95** |
| 864 | 428.95 | 383.28 | 371.14 | 398.31 | **1,581.68** |
| 865 | 0 | 2.31 | 2.31 | 2.31 | **6.94** |
| **AZ - ARIZONA - Total** | **42,559.14** | **40,061.17** | **41,158.99** | **43,821.71** | **167,601.02** |

**STATE: CA - CALIFORNIA**

| REGISTRANT ZIP CODE 3 | 1ST QUARTER | 2ND QUARTER | 3RD QUARTER | 4TH QUARTER | TOTAL GRAMS |
|---|---|---|---|---|---|
| 900 | 7,177.69 | 6,031.32 | 6,676.48 | 6,793.25 | **26,678.74** |
| 902 | 5,187.87 | 4,617.86 | 4,712.09 | 4,828.29 | **19,346.12** |
| 903 | 87.29 | 126.03 | 97.12 | 100.01 | **410.45** |
| 904 | 1,338.30 | 1,140.59 | 1,226.73 | 1,236.56 | **4,942.18** |
| 905 | 796.62 | 765.98 | 815.12 | 871.77 | **3,249.50** |
| 906 | 1,078.16 | 942.88 | 923.23 | 1,000.69 | **3,944.95** |
| 907 | 1,544.68 | 1,422.70 | 1,309.97 | 1,440.63 | **5,717.99** |
| 908 | 926.12 | 834.78 | 856.17 | 900.68 | **3,517.74** |
| 910 | 681.58 | 718.58 | 763.67 | 722.05 | **2,885.88** |
| 911 | 1,492.65 | 1,415.77 | 1,344.66 | 1,400.74 | **5,653.82** |
| 912 | 581.57 | 526.65 | 558.44 | 548.62 | **2,215.28** |
| 913 | 5,830.14 | 5,332.97 | 5,590.81 | 5,486.17 | **22,240.09** |
| 914 | 1,292.63 | 1,172.96 | 1,231.93 | 1,220.37 | **4,917.90** |
| 915 | 476.35 | 423.75 | 472.89 | 433.00 | **1,805.98** |
| 916 | 704.13 | 650.36 | 622.61 | 675.22 | **2,652.32** |
| 917 | 2,929.23 | 2,690.48 | 2,621.11 | 2,911.31 | **11,152.13** |
| 918 | 146.26 | 160.13 | 128.92 | 167.07 | **602.38** |
| 919 | 909.35 | 916.87 | 932.48 | 960.80 | **3,719.50** |
| 920 | 14,741.55 | 14,963.54 | 14,679.69 | 15,843.41 | **60,228.19** |
| 921 | 5,060.11 | 4,690.13 | 4,648.50 | 4,991.32 | **19,390.05** |
| 922 | 1,185.11 | 1,175.28 | 1,148.11 | 1,275.87 | **4,784.36** |
| 923 | 1,893.86 | 1,587.46 | 1,667.24 | 1,820.44 | **6,969.00** |
| 924 | 120.82 | 154.35 | 128.34 | 159.56 | **563.07** |
| 925 | 2,123.36 | 1,899.06 | 1,934.32 | 2,134.35 | **8,091.09** |
| 926 | 8,043.68 | 7,367.31 | 7,781.23 | 7,910.72 | **31,102.94** |
| 927 | 1,272.40 | 1,097.81 | 1,104.75 | 1,091.45 | **4,566.41** |
| 928 | 3,268.00 | 3,009.59 | 3,046.59 | 2,973.75 | **12,297.92** |
| 930 | 3,032.71 | 2,691.63 | 2,789.33 | 2,875.47 | **11,389.15** |
| 931 | 1,057.34 | 929.01 | 850.96 | 959.65 | **3,796.96** |
| 932 | 1,107.64 | 951.55 | 1,035.38 | 1,071.22 | **4,165.79** |
| 933 | 1,000.11 | 773.50 | 830.73 | 859.63 | **3,463.98** |
| 934 | 2,080.00 | 1,807.14 | 1,827.95 | 1,847.61 | **7,562.70** |
| 935 | 1,044.05 | 863.68 | 978.15 | 973.52 | **3,859.40** |
| 936 | 1,378.77 | 1,140.01 | 1,189.73 | 1,300.73 | **5,009.24** |
| 937 | 1,852.81 | 1,692.10 | 1,583.99 | 1,590.35 | **6,719.26** |
| 939 | 831.89 | 701.24 | 790.26 | 832.46 | **3,155.85** |

| | | | | | |
|---|---|---|---|---|---|
| 940 | 2,665.62 | 2,473.11 | 2,670.82 | 2,675.45 | **10,485.00** |
| 941 | 4,603.99 | 3,721.81 | 3,991.78 | 3,982.53 | **16,300.11** |
| 943 | 447.45 | 353.80 | 382.70 | 389.64 | **1,573.59** |
| 944 | 544.57 | 559.60 | 527.81 | 523.18 | **2,155.16** |
| 945 | 5,401.77 | 5,025.42 | 5,067.05 | 5,296.55 | **20,790.79** |
| 946 | 1,149.26 | 1,160.25 | 1,172.39 | 1,211.70 | **4,693.59** |
| 947 | 571.16 | 510.46 | 543.41 | 543.41 | **2,168.45** |
| 948 | 153.20 | 104.64 | 123.14 | 133.54 | **514.51** |
| 949 | 1,676.49 | 1,566.65 | 1,678.22 | 1,704.24 | **6,625.60** |
| 950 | 2,541.33 | 2,310.67 | 2,348.82 | 2,395.07 | **9,595.88** |
| 951 | 1,338.30 | 1,389.75 | 1,484.56 | 1,474.73 | **5,687.35** |
| 952 | 738.81 | 668.86 | 602.96 | 671.75 | **2,682.38** |
| 953 | 2,106.02 | 1,908.89 | 1,753.38 | 2,011.21 | **7,779.49** |
| 954 | 629.55 | 700.08 | 737.66 | 696.03 | **2,763.32** |
| 955 | 191.35 | 146.84 | 180.37 | 164.76 | **683.31** |
| 956 | 3,446.63 | 3,350.67 | 3,408.48 | 3,577.28 | **13,783.06** |
| 957 | 1,159.09 | 1,079.31 | 1,168.34 | 1,207.07 | **4,613.82** |
| 958 | 1,662.04 | 1,441.20 | 1,582.26 | 1,568.96 | **6,254.46** |
| 959 | 1,224.99 | 1,187.42 | 1,179.32 | 1,189.73 | **4,781.47** |
| 960 | 848.07 | 727.25 | 864.26 | 841.71 | **3,281.30** |
| 961 | 169.96 | 209.85 | 223.72 | 228.35 | **831.89** |
| **CA - CALIFORNIA - Total** | **117,544.49** | **107,981.56** | **110,591.11** | **114,695.62** | **450,812.79** |
| **STATE: CO - COLORADO** | | | | | |
| REGISTRANT ZIP CODE 3 | 1ST QUARTER | 2ND QUARTER | 3RD QUARTER | 4TH QUARTER | **TOTAL GRAMS** |
| 800 | 4,767.59 | 4,332.86 | 4,632.32 | 4,997.10 | **18,729.86** |
| 801 | 5,573.46 | 5,319.10 | 5,478.08 | 5,929.57 | **22,300.21** |
| 802 | 6,635.43 | 6,085.08 | 6,532.53 | 6,882.28 | **26,135.32** |
| 803 | 1,889.23 | 1,509.42 | 1,740.66 | 1,795.58 | **6,934.89** |
| 804 | 1,025.55 | 1,001.85 | 1,115.73 | 1,123.83 | **4,266.96** |
| 805 | 3,719.50 | 3,244.30 | 3,694.64 | 3,827.60 | **14,486.03** |
| 806 | 1,179.32 | 993.75 | 1,188.57 | 1,166.03 | **4,527.68** |
| 807 | 152.62 | 135.28 | 130.65 | 153.77 | **572.32** |
| 808 | 500.63 | 537.63 | 575.79 | 612.21 | **2,226.26** |
| 809 | 3,269.73 | 2,951.20 | 3,110.76 | 3,321.18 | **12,652.87** |
| 810 | 596.60 | 615.68 | 628.39 | 667.13 | **2,507.80** |
| 811 | 134.12 | 136.43 | 123.14 | 130.07 | **523.76** |
| 812 | 171.12 | 156.09 | 203.49 | 203.49 | **734.19** |
| 813 | 211.58 | 225.46 | 242.80 | 264.77 | **944.62** |
| 814 | 143.37 | 153.77 | 211.58 | 186.15 | **694.88** |
| 815 | 624.93 | 672.33 | 686.20 | 693.14 | **2,676.60** |
| 816 | 925.54 | 796.62 | 983.93 | 905.88 | **3,611.97** |
| **CO - COLORADO - Total** | **31,520.32** | **28,866.85** | **31,279.26** | **32,859.78** | **124,526.21** |
| **STATE: CT - CONNECTICUT** | | | | | |
| REGISTRANT ZIP CODE 3 | 1ST QUARTER | 2ND QUARTER | 3RD QUARTER | 4TH QUARTER | **TOTAL GRAMS** |
| 060 | 6,166.01 | 5,927.84 | 6,137.11 | 6,488.02 | **24,718.98** |
| 061 | 2,236.09 | 1,958.02 | 1,900.21 | 2,165.56 | **8,259.89** |
| 062 | 1,836.62 | 1,663.19 | 1,717.54 | 1,775.35 | **6,992.70** |
| 063 | 3,674.40 | 3,660.53 | 3,794.65 | 3,954.78 | **15,084.36** |
| 064 | 7,519.92 | 7,445.93 | 7,517.61 | 7,650.00 | **30,133.46** |
| 065 | 2,055.72 | 1,725.63 | 1,937.21 | 2,052.83 | **7,771.40** |
| 066 | 1,356.22 | 1,155.62 | 1,257.95 | 1,319.22 | **5,089.01** |
| 067 | 2,960.45 | 2,728.05 | 2,763.90 | 2,910.16 | **11,362.56** |
| 068 | 7,740.18 | 7,493.91 | 7,822.27 | 7,902.05 | **30,958.41** |
| 069 | 1,226.15 | 1,079.89 | 1,075.27 | 1,158.51 | **4,539.82** |
| **CT - CONNECTICUT - Total** | **36,771.78** | **34,838.62** | **35,923.71** | **37,376.48** | **144,910.59** |
| **STATE: DC - DISTRICT OF COLUMBIA** | | | | | |
| REGISTRANT ZIP CODE 3 | 1ST QUARTER | 2ND QUARTER | 3RD QUARTER | 4TH QUARTER | **TOTAL GRAMS** |
| 200 | 6,174.69 | 4,991.32 | 5,621.44 | 5,634.16 | **22,421.61** |
| 203 | 28.33 | 0 | 6.94 | 12.72 | **47.98** |
| 204 | 6.94 | 95.39 | 91.92 | 37.00 | **231.24** |

| DC - DISTRICT OF COLUMBIA - Total | 6,209.95 | 5,086.70 | 5,720.30 | 5,683.88 | 22,700.83 |
|---|---|---|---|---|---|

STATE: DE - DELAWARE

| REGISTRANT ZIP CODE 3 | 1ST QUARTER | 2ND QUARTER | 3RD QUARTER | 4TH QUARTER | TOTAL GRAMS |
|---|---|---|---|---|---|
| 197 | 5,081.50 | 4,569.88 | 4,665.27 | 4,915.01 | **19,231.65** |
| 198 | 3,185.33 | 2,939.64 | 2,982.42 | 3,158.16 | **12,265.55** |
| 199 | 5,357.83 | 5,156.07 | 5,177.46 | 5,078.03 | **20,769.40** |
| **DE - DELAWARE - Total** | **13,624.66** | **12,665.59** | **12,825.15** | **13,151.20** | **52,266.60** |

STATE: FL - FLORIDA

| REGISTRANT ZIP CODE 3 | 1ST QUARTER | 2ND QUARTER | 3RD QUARTER | 4TH QUARTER | TOTAL GRAMS |
|---|---|---|---|---|---|
| 320 | 5,154.92 | 4,639.83 | 4,577.97 | 4,695.91 | **19,068.63** |
| 321 | 2,706.09 | 2,448.25 | 2,580.64 | 2,636.71 | **10,371.69** |
| 322 | 8,431.01 | 7,585.83 | 7,767.35 | 7,881.24 | **31,665.43** |
| 323 | 5,563.63 | 4,898.24 | 4,984.38 | 5,284.41 | **20,730.67** |
| 324 | 2,933.28 | 2,673.13 | 2,717.07 | 2,776.61 | **11,100.10** |
| 325 | 8,347.19 | 7,645.37 | 7,943.67 | 8,252.38 | **32,188.61** |
| 326 | 3,392.29 | 2,882.41 | 3,063.35 | 3,284.76 | **12,622.81** |
| 327 | 9,009.69 | 8,186.47 | 8,395.75 | 8,666.30 | **34,258.21** |
| 328 | 7,355.74 | 6,509.41 | 6,594.96 | 6,831.41 | **27,291.52** |
| 329 | 6,044.04 | 5,611.04 | 5,762.50 | 5,943.45 | **23,361.02** |
| 330 | 6,568.37 | 5,490.22 | 5,441.66 | 5,851.53 | **23,351.77** |
| 331 | 8,437.37 | 7,304.29 | 7,533.80 | 8,067.96 | **31,343.43** |
| 333 | 3,691.17 | 3,159.32 | 3,218.28 | 3,288.81 | **13,357.58** |
| 334 | 10,027.14 | 9,104.50 | 8,576.69 | 9,057.09 | **36,765.43** |
| 335 | 7,145.32 | 6,340.02 | 6,480.50 | 7,069.58 | **27,035.42** |
| 336 | 6,648.73 | 5,693.13 | 6,177.58 | 6,326.15 | **24,845.58** |
| 337 | 5,086.70 | 4,660.06 | 4,665.85 | 5,037.56 | **19,450.17** |
| 338 | 5,405.24 | 4,857.20 | 4,835.81 | 5,393.09 | **20,491.33** |
| 339 | 5,441.08 | 5,049.13 | 4,743.89 | 5,323.72 | **20,557.81** |
| 341 | 2,516.47 | 2,429.75 | 2,140.70 | 2,615.32 | **9,702.25** |
| 342 | 6,368.35 | 5,976.40 | 5,980.44 | 6,309.96 | **24,635.15** |
| 344 | 2,407.21 | 2,114.11 | 2,156.31 | 2,266.73 | **8,944.36** |
| 346 | 4,479.12 | 4,021.26 | 4,084.85 | 4,327.66 | **16,912.89** |
| 347 | 5,291.35 | 4,534.04 | 4,551.96 | 4,982.64 | **19,359.99** |
| 349 | 3,008.43 | 2,878.94 | 2,788.18 | 3,139.66 | **11,815.21** |
| **FL - FLORIDA - Total** | **141,459.91** | **126,692.35** | **127,764.15** | **135,310.66** | **531,227.07** |

STATE: GA - GEORGIA

| REGISTRANT ZIP CODE 3 | 1ST QUARTER | 2ND QUARTER | 3RD QUARTER | 4TH QUARTER | TOTAL GRAMS |
|---|---|---|---|---|---|
| 300 | 23,050.00 | 19,857.16 | 20,945.14 | 21,501.27 | **85,353.57** |
| 301 | 12,063.79 | 10,245.67 | 11,273.53 | 11,564.89 | **45,147.88** |
| 302 | 7,289.26 | 6,376.44 | 6,471.25 | 6,810.60 | **26,947.55** |
| 303 | 12,151.66 | 11,082.76 | 11,244.62 | 11,952.22 | **46,431.26** |
| 304 | 2,588.73 | 2,221.06 | 2,445.36 | 2,636.14 | **9,891.29** |
| 305 | 6,119.77 | 5,548.03 | 5,874.65 | 6,137.11 | **23,679.55** |
| 306 | 5,001.14 | 4,079.07 | 4,705.73 | 4,810.37 | **18,596.32** |
| 307 | 2,223.95 | 1,956.29 | 2,041.85 | 2,220.48 | **8,442.57** |
| 308 | 2,003.69 | 1,713.49 | 1,751.64 | 1,889.81 | **7,358.63** |
| 309 | 2,660.42 | 2,162.67 | 2,439.00 | 2,217.01 | **9,479.11** |
| 310 | 5,068.20 | 4,382.58 | 4,718.45 | 4,754.29 | **18,923.53** |
| 312 | 1,941.26 | 1,673.02 | 1,839.51 | 1,989.24 | **7,443.04** |
| 313 | 2,431.49 | 2,116.42 | 2,360.96 | 2,299.68 | **9,208.55** |
| 314 | 2,888.77 | 2,478.31 | 2,540.75 | 2,629.78 | **10,537.61** |
| 315 | 3,257.59 | 2,643.65 | 2,889.92 | 2,848.30 | **11,639.47** |
| 316 | 1,291.48 | 1,069.49 | 1,197.82 | 1,390.91 | **4,949.69** |
| 317 | 2,930.97 | 2,475.42 | 2,541.33 | 2,650.59 | **10,598.31** |
| 318 | 247.43 | 208.69 | 231.82 | 271.13 | **959.07** |
| 319 | 1,884.61 | 1,698.46 | 1,897.90 | 1,827.95 | **7,308.92** |
| 398 | 430.11 | 307.55 | 302.92 | 402.94 | **1,443.52** |
| **GA - GEORGIA - Total** | **97,524.31** | **84,296.23** | **89,714.18** | **92,804.71** | **364,339.43** |

STATE: GU - GUAM

| REGISTRANT ZIP CODE 3 | 1ST QUARTER | 2ND QUARTER | 3RD QUARTER | 4TH QUARTER | TOTAL GRAMS |
|---|---|---|---|---|---|

| | | | | | |
|---|---|---|---|---|---|
| 969 | 21.97 | 27.17 | 13.87 | 12.72 | **75.73** |
| **GU - GUAM - Total** | **21.97** | **27.17** | **13.87** | **12.72** | **75.73** |

**STATE: HI - HAWAII**

| REGISTRANT ZIP CODE 3 | 1ST QUARTER | 2ND QUARTER | 3RD QUARTER | 4TH QUARTER | TOTAL GRAMS |
|---|---|---|---|---|---|
| 967 | 1,003.58 | 923.80 | 990.86 | 1,087.41 | **4,005.65** |
| 968 | 957.91 | 819.17 | 870.04 | 858.48 | **3,505.60** |
| **HI - HAWAII - Total** | **1,961.49** | **1,742.97** | **1,860.90** | **1,945.88** | **7,511.25** |

**STATE: IA - IOWA**

| REGISTRANT ZIP CODE 3 | 1ST QUARTER | 2ND QUARTER | 3RD QUARTER | 4TH QUARTER | TOTAL GRAMS |
|---|---|---|---|---|---|
| 500 | 2,454.61 | 2,010.05 | 2,502.59 | 2,452.30 | **9,419.56** |
| 501 | 1,485.72 | 1,342.35 | 1,327.90 | 1,363.16 | **5,519.12** |
| 502 | 1,538.32 | 1,342.35 | 1,501.90 | 1,552.78 | **5,935.35** |
| 503 | 2,763.90 | 2,473.69 | 2,513.00 | 2,536.70 | **10,287.29** |
| 504 | 911.09 | 760.20 | 831.31 | 838.82 | **3,341.42** |
| 505 | 1,597.87 | 1,323.85 | 1,364.89 | 1,518.67 | **5,805.28** |
| 506 | 1,432.53 | 1,278.76 | 1,399.00 | 1,386.86 | **5,497.15** |
| 507 | 1,190.89 | 1,039.42 | 1,103.01 | 1,159.09 | **4,492.42** |
| 508 | 235.29 | 197.13 | 198.87 | 197.71 | **829.00** |
| 510 | 1,072.95 | 1,070.64 | 1,054.45 | 1,130.76 | **4,328.81** |
| 511 | 2,681.23 | 2,556.94 | 2,518.78 | 2,633.25 | **10,390.19** |
| 512 | 433.00 | 379.81 | 412.76 | 430.11 | **1,655.68** |
| 513 | 539.37 | 501.79 | 554.40 | 513.93 | **2,109.49** |
| 514 | 280.96 | 241.07 | 267.08 | 275.75 | **1,064.86** |
| 515 | 1,952.24 | 1,816.97 | 1,770.72 | 1,860.90 | **7,400.84** |
| 516 | 176.90 | 132.38 | 184.41 | 175.74 | **669.44** |
| 520 | 1,779.39 | 1,675.33 | 1,734.88 | 1,862.64 | **7,052.24** |
| 521 | 255.52 | 211.01 | 232.40 | 235.86 | **934.79** |
| 522 | 2,255.75 | 1,930.85 | 2,041.85 | 2,292.74 | **8,521.19** |
| 523 | 1,731.99 | 1,622.73 | 1,638.91 | 1,682.85 | **6,676.48** |
| 524 | 2,510.11 | 2,360.96 | 2,283.50 | 2,528.03 | **9,682.60** |
| 525 | 1,018.61 | 889.12 | 967.74 | 848.07 | **3,723.54** |
| 526 | 999.53 | 959.07 | 955.60 | 1,018.03 | **3,932.24** |
| 527 | 1,623.30 | 1,504.22 | 1,588.04 | 1,538.90 | **6,254.46** |
| 528 | 1,386.86 | 1,309.97 | 1,375.88 | 1,421.55 | **5,494.26** |
| **IA - IOWA - Total** | **34,307.92** | **30,930.66** | **32,323.88** | **33,455.23** | **131,017.69** |

**STATE: ID - IDAHO**

| REGISTRANT ZIP CODE 3 | 1ST QUARTER | 2ND QUARTER | 3RD QUARTER | 4TH QUARTER | TOTAL GRAMS |
|---|---|---|---|---|---|
| 832 | 788.53 | 738.81 | 829.00 | 879.29 | **3,235.63** |
| 833 | 837.09 | 833.62 | 792.00 | 883.34 | **3,346.04** |
| 834 | 1,825.64 | 1,747.60 | 1,874.78 | 1,955.13 | **7,403.15** |
| 835 | 169.38 | 157.82 | 171.12 | 137.01 | **635.33** |
| 836 | 3,594.05 | 3,453.57 | 3,540.86 | 3,810.84 | **14,399.31** |
| 837 | 2,444.21 | 2,302.57 | 2,587.00 | 2,709.55 | **10,043.33** |
| 838 | 1,190.89 | 1,083.36 | 1,211.70 | 1,281.07 | **4,767.01** |
| **ID - IDAHO - Total** | **10,849.78** | **10,317.35** | **11,006.45** | **11,656.23** | **43,829.81** |

**STATE: IL - ILLINOIS**

| REGISTRANT ZIP CODE 3 | 1ST QUARTER | 2ND QUARTER | 3RD QUARTER | 4TH QUARTER | TOTAL GRAMS |
|---|---|---|---|---|---|
| 600 | 15,696.57 | 14,742.71 | 15,637.61 | 15,890.81 | **61,967.70** |
| 601 | 10,413.32 | 9,634.04 | 9,768.73 | 10,487.31 | **40,303.40** |
| 602 | 688.52 | 718.00 | 706.44 | 759.62 | **2,872.58** |
| 603 | 749.80 | 711.06 | 695.45 | 777.54 | **2,933.86** |
| 604 | 7,602.02 | 7,039.52 | 7,497.38 | 7,591.03 | **29,729.95** |
| 605 | 9,010.27 | 8,596.35 | 8,788.85 | 9,181.96 | **35,577.43** |
| 606 | 13,089.92 | 11,060.79 | 11,863.19 | 12,633.80 | **48,647.69** |
| 607 | 716.84 | 650.94 | 656.14 | 696.03 | **2,719.96** |
| 608 | 163.02 | 158.98 | 178.05 | 220.26 | **720.31** |
| 609 | 1,151.58 | 1,072.38 | 1,144.64 | 1,280.49 | **4,649.08** |
| 610 | 1,019.77 | 942.88 | 949.24 | 1,048.10 | **3,959.99** |
| 611 | 1,114.00 | 972.94 | 1,098.39 | 1,144.06 | **4,329.39** |
| 612 | 765.98 | 710.48 | 780.44 | 785.06 | **3,041.96** |

| | | | | | |
|---|---|---|---|---|---|
| 613 | 628.97 | 597.18 | 627.82 | 658.46 | **2,512.42** |
| 614 | 842.87 | 712.80 | 830.73 | 902.41 | **3,288.81** |
| 615 | 693.14 | 631.29 | 642.85 | 742.28 | **2,709.55** |
| 616 | 1,249.85 | 1,138.86 | 1,218.06 | 1,207.65 | **4,814.42** |
| 617 | 1,446.98 | 1,333.68 | 1,515.20 | 1,605.38 | **5,901.24** |
| 618 | 1,352.75 | 1,186.26 | 1,287.43 | 1,483.98 | **5,310.43** |
| 619 | 265.93 | 253.79 | 305.81 | 339.92 | **1,165.45** |
| 620 | 1,261.99 | 1,170.07 | 1,252.74 | 1,378.19 | **5,063.00** |
| 622 | 1,977.10 | 1,845.87 | 2,054.57 | 2,029.71 | **7,907.25** |
| 623 | 870.62 | 889.12 | 892.59 | 893.74 | **3,546.07** |
| 624 | 326.05 | 242.22 | 308.13 | 374.03 | **1,250.43** |
| 625 | 450.34 | 416.81 | 457.86 | 467.10 | **1,792.11** |
| 626 | 467.10 | 410.45 | 500.63 | 491.96 | **1,870.15** |
| 627 | 693.72 | 622.04 | 726.09 | 724.36 | **2,766.21** |
| 628 | 594.86 | 586.19 | 716.27 | 732.45 | **2,629.78** |
| 629 | 1,285.12 | 1,274.71 | 1,330.21 | 1,313.44 | **5,203.48** |
| **IL - ILLINOIS - Total** | **76,589.00** | **70,322.40** | **74,431.53** | **77,841.17** | **299,184.09** |

**STATE: IN - INDIANA**

| REGISTRANT ZIP CODE 3 | 1ST QUARTER | 2ND QUARTER | 3RD QUARTER | 4TH QUARTER | TOTAL GRAMS |
|---|---|---|---|---|---|
| 460 | 10,506.39 | 9,323.60 | 9,897.07 | 10,001.71 | **39,728.77** |
| 461 | 6,842.97 | 5,877.54 | 6,359.10 | 6,555.08 | **25,634.69** |
| 462 | 8,722.95 | 7,718.79 | 8,137.91 | 8,189.36 | **32,769.02** |
| 463 | 8,515.41 | 7,920.55 | 8,393.43 | 8,599.24 | **33,428.63** |
| 464 | 668.86 | 560.76 | 587.93 | 608.16 | **2,425.71** |
| 465 | 7,606.64 | 6,918.70 | 7,378.29 | 7,630.92 | **29,534.55** |
| 466 | 2,137.81 | 2,070.75 | 2,025.66 | 2,036.65 | **8,270.88** |
| 467 | 2,493.92 | 2,259.21 | 2,282.92 | 2,644.81 | **9,680.86** |
| 468 | 3,791.76 | 3,503.86 | 3,905.07 | 3,928.77 | **15,129.46** |
| 469 | 6,148.67 | 5,528.95 | 5,728.39 | 6,064.27 | **23,470.28** |
| 470 | 664.82 | 598.91 | 649.21 | 709.91 | **2,622.84** |
| 471 | 3,858.24 | 3,318.87 | 3,505.60 | 3,679.61 | **14,362.32** |
| 472 | 1,552.20 | 1,312.29 | 1,388.02 | 1,411.14 | **5,663.65** |
| 473 | 3,293.44 | 2,874.31 | 3,343.15 | 3,420.04 | **12,930.94** |
| 474 | 2,909.58 | 2,320.49 | 2,541.91 | 2,710.71 | **10,482.69** |
| 475 | 1,688.05 | 1,499.59 | 1,645.85 | 1,753.38 | **6,586.87** |
| 476 | 2,459.24 | 2,274.82 | 2,286.96 | 2,324.54 | **9,345.56** |
| 477 | 4,982.64 | 4,331.70 | 4,467.56 | 4,676.83 | **18,458.73** |
| 478 | 1,285.12 | 1,094.92 | 1,166.03 | 1,200.14 | **4,746.20** |
| 479 | 3,233.31 | 2,765.63 | 3,071.45 | 3,300.37 | **12,370.76** |
| **IN - INDIANA - Total** | **83,362.02** | **74,074.27** | **78,761.50** | **81,445.62** | **317,643.40** |

**STATE: KS - KANSAS**

| REGISTRANT ZIP CODE 3 | 1ST QUARTER | 2ND QUARTER | 3RD QUARTER | 4TH QUARTER | TOTAL GRAMS |
|---|---|---|---|---|---|
| 660 | 7,233.19 | 6,592.65 | 7,244.17 | 7,315.86 | **28,385.87** |
| 661 | 886.23 | 863.68 | 851.54 | 792.58 | **3,394.03** |
| 662 | 7,587.56 | 6,784.00 | 7,167.28 | 7,193.30 | **28,732.15** |
| 664 | 480.40 | 490.23 | 515.09 | 488.49 | **1,974.21** |
| 665 | 919.76 | 801.82 | 845.18 | 982.19 | **3,548.96** |
| 666 | 3,965.19 | 4,014.90 | 4,138.62 | 4,188.33 | **16,307.04** |
| 667 | 1,201.87 | 1,145.79 | 1,261.99 | 1,253.32 | **4,862.98** |
| 668 | 600.65 | 499.48 | 523.18 | 568.27 | **2,191.58** |
| 669 | 124.87 | 109.84 | 119.67 | 142.79 | **497.17** |
| 670 | 1,708.29 | 1,616.37 | 1,584.57 | 1,747.02 | **6,656.24** |
| 671 | 603.54 | 567.69 | 593.13 | 575.79 | **2,340.15** |
| 672 | 4,102.20 | 3,746.67 | 3,855.93 | 4,079.07 | **15,783.86** |
| 673 | 590.82 | 569.43 | 602.38 | 637.07 | **2,399.69** |
| 674 | 981.61 | 831.89 | 881.02 | 927.27 | **3,621.80** |
| 675 | 861.37 | 727.83 | 709.91 | 748.06 | **3,047.17** |
| 676 | 516.24 | 510.46 | 533.59 | 561.34 | **2,121.63** |
| 677 | 108.10 | 110.42 | 128.34 | 114.46 | **461.32** |
| 678 | 326.63 | 289.63 | 311.02 | 325.47 | **1,252.74** |

| | | | | | |
|---|---|---|---|---|---|
| 679 | 178.05 | 139.32 | 164.18 | 137.59 | **619.15** |
| **KS - KANSAS - Total** | **32,976.56** | **30,412.11** | **32,030.79** | **32,778.27** | **128,197.72** |

**STATE: KY - KENTUCKY**

| REGISTRANT ZIP CODE 3 | 1ST QUARTER | 2ND QUARTER | 3RD QUARTER | 4TH QUARTER | TOTAL GRAMS |
|---|---|---|---|---|---|
| 400 | 2,756.96 | 2,417.61 | 2,418.77 | 2,579.48 | **10,172.83** |
| 401 | 722.05 | 598.33 | 646.32 | 679.27 | **2,645.96** |
| 402 | 8,874.41 | 7,754.06 | 7,921.13 | 8,164.51 | **32,714.10** |
| 403 | 2,684.70 | 2,352.87 | 2,439.00 | 2,492.19 | **9,968.76** |
| 404 | 980.46 | 997.80 | 963.11 | 978.72 | **3,920.10** |
| 405 | 4,265.19 | 3,716.60 | 3,940.91 | 4,133.42 | **16,056.12** |
| 406 | 435.31 | 387.33 | 400.62 | 394.84 | **1,618.10** |
| 407 | 427.79 | 338.19 | 349.17 | 387.33 | **1,502.48** |
| 408 | 90.18 | 77.47 | 87.87 | 80.93 | **336.45** |
| 409 | 110.42 | 98.86 | 132.38 | 101.75 | **443.40** |
| 410 | 4,231.69 | 3,737.42 | 3,884.83 | 4,217.82 | **16,071.76** |
| 411 | 623.77 | 526.65 | 518.56 | 593.13 | **2,262.11** |
| 412 | 209.27 | 162.45 | 187.88 | 225.46 | **785.06** |
| 413 | 64.17 | 51.45 | 68.22 | 57.81 | **241.65** |
| 414 | 90.76 | 77.47 | 84.98 | 76.89 | **330.10** |
| 415 | 401.20 | 369.98 | 335.30 | 331.25 | **1,437.73** |
| 416 | 226.62 | 205.80 | 220.83 | 207.54 | **860.79** |
| 417 | 283.85 | 212.74 | 241.65 | 323.74 | **1,061.97** |
| 418 | 53.76 | 49.72 | 58.97 | 60.70 | **223.15** |
| 420 | 3,809.68 | 3,577.86 | 3,514.27 | 3,435.65 | **14,337.46** |
| 421 | 1,680.54 | 1,399.00 | 1,585.73 | 1,673.60 | **6,338.87** |
| 422 | 697.77 | 698.34 | 760.20 | 590.24 | **2,746.55** |
| 423 | 2,159.20 | 1,881.72 | 2,016.41 | 2,043.58 | **8,100.92** |
| 424 | 2,237.83 | 2,072.49 | 2,154.58 | 2,071.91 | **8,536.80** |
| 425 | 368.25 | 294.25 | 309.28 | 351.48 | **1,323.27** |
| 426 | 113.89 | 95.96 | 64.17 | 115.04 | **389.06** |
| 427 | 1,212.85 | 1,174.12 | 1,216.90 | 1,175.86 | **4,779.73** |
| **KY - KENTUCKY - Total** | **39,812.56** | **35,326.53** | **36,522.05** | **37,544.13** | **149,205.27** |

**STATE: LA - LOUISIANA**

| REGISTRANT ZIP CODE 3 | 1ST QUARTER | 2ND QUARTER | 3RD QUARTER | 4TH QUARTER | TOTAL GRAMS |
|---|---|---|---|---|---|
| 700 | 7,956.39 | 6,655.67 | 7,284.06 | 7,588.72 | **29,484.83** |
| 701 | 5,000.57 | 4,073.87 | 4,580.29 | 4,769.33 | **18,424.05** |
| 703 | 5,463.05 | 4,476.23 | 5,062.42 | 5,175.73 | **20,177.42** |
| 704 | 9,420.72 | 8,295.74 | 8,505.01 | 9,418.98 | **35,640.44** |
| 705 | 16,833.69 | 14,026.44 | 15,358.38 | 15,760.74 | **61,979.26** |
| 706 | 8,769.78 | 8,054.67 | 7,507.21 | 7,685.84 | **32,017.49** |
| 707 | 11,594.37 | 10,124.27 | 10,998.93 | 11,372.38 | **44,089.95** |
| 708 | 7,824.01 | 6,299.56 | 7,180.00 | 7,518.19 | **28,821.75** |
| 710 | 2,134.92 | 1,805.98 | 1,915.25 | 2,105.44 | **7,961.59** |
| 711 | 6,479.92 | 5,347.43 | 5,959.05 | 6,067.16 | **23,853.56** |
| 712 | 4,615.55 | 3,753.03 | 4,212.61 | 4,464.09 | **17,045.28** |
| 713 | 3,616.02 | 2,951.20 | 3,368.01 | 3,609.08 | **13,544.30** |
| 714 | 1,824.48 | 1,606.54 | 1,829.11 | 1,844.72 | **7,104.85** |
| **LA - LOUISIANA - Total** | **91,533.46** | **77,470.60** | **83,760.33** | **87,380.39** | **340,144.79** |

**STATE: MA - MASSACHUSETTS**

| REGISTRANT ZIP CODE 3 | 1ST QUARTER | 2ND QUARTER | 3RD QUARTER | 4TH QUARTER | TOTAL GRAMS |
|---|---|---|---|---|---|
| 010 | 2,184.64 | 1,936.06 | 2,002.54 | 2,015.83 | **8,139.07** |
| 011 | 894.90 | 663.66 | 626.08 | 783.90 | **2,968.54** |
| 012 | 585.62 | 660.77 | 695.45 | 785.06 | **2,726.90** |
| 013 | 539.37 | 454.39 | 501.21 | 553.24 | **2,048.21** |
| 014 | 1,597.29 | 1,479.36 | 1,540.06 | 1,655.10 | **6,271.81** |
| 015 | 1,794.42 | 1,683.43 | 1,531.97 | 1,873.04 | **6,882.86** |
| 016 | 1,140.59 | 935.94 | 989.13 | 1,081.05 | **4,146.71** |
| 017 | 2,918.25 | 2,639.60 | 2,658.68 | 2,907.26 | **11,123.80** |
| 018 | 4,879.74 | 4,361.76 | 4,592.43 | 4,560.63 | **18,394.56** |
| 019 | 4,209.72 | 3,951.31 | 4,088.32 | 4,342.07 | **16,591.44** |

| | | | | | |
|---|---|---|---|---|---|
| 020 | 3,614.86 | 3,391.71 | 3,363.39 | 3,521.21 | **13,891.16** |
| 021 | 7,184.63 | 6,180.47 | 6,745.27 | 7,115.25 | **27,225.62** |
| 022 | 478.67 | 318.53 | 392.53 | 411.61 | **1,601.34** |
| 023 | 2,919.98 | 2,732.68 | 2,651.74 | 2,782.40 | **11,086.80** |
| 024 | 3,030.98 | 2,902.64 | 3,010.17 | 3,021.73 | **11,965.51** |
| 025 | 835.35 | 892.59 | 1,111.69 | 898.37 | **3,737.99** |
| 026 | 932.48 | 1,282.80 | 1,422.70 | 1,056.19 | **4,694.17** |
| 027 | 2,913.05 | 2,673.13 | 2,507.22 | 2,567.34 | **10,660.74** |
| **MA - MASSACHUSETTS - Total** | **42,654.53** | **39,140.84** | **40,430.58** | **41,931.29** | **164,157.24** |

**STATE: MD - MARYLAND**

| REGISTRANT ZIP CODE 3 | 1ST QUARTER | 2ND QUARTER | 3RD QUARTER | 4TH QUARTER | TOTAL GRAMS |
|---|---|---|---|---|---|
| 206 | 3,061.04 | 2,942.53 | 2,924.61 | 3,012.48 | **11,940.66** |
| 207 | 4,157.70 | 3,683.08 | 3,868.65 | 3,833.96 | **15,543.37** |
| 208 | 7,958.12 | 7,474.83 | 7,407.20 | 7,708.39 | **30,548.54** |
| 209 | 1,531.39 | 1,295.52 | 1,334.25 | 1,558.56 | **5,719.72** |
| 210 | 11,452.74 | 10,371.11 | 10,293.07 | 11,227.86 | **43,344.78** |
| 211 | 5,475.19 | 5,110.98 | 4,968.19 | 5,261.87 | **20,816.22** |
| 212 | 10,343.94 | 9,333.42 | 9,235.15 | 10,095.94 | **39,008.45** |
| 214 | 1,619.26 | 1,452.19 | 1,529.65 | 1,496.12 | **6,097.22** |
| 215 | 1,849.92 | 1,742.97 | 1,849.34 | 1,819.28 | **7,261.51** |
| 216 | 2,370.21 | 2,120.47 | 2,111.22 | 2,280.60 | **8,882.51** |
| 217 | 6,641.21 | 6,231.34 | 6,473.56 | 6,837.77 | **26,183.88** |
| 218 | 2,274.25 | 2,095.03 | 2,198.51 | 2,147.06 | **8,714.86** |
| 219 | 1,338.88 | 1,176.43 | 1,214.59 | 1,299.57 | **5,029.47** |
| **MD - MARYLAND - Total** | **60,073.84** | **55,029.92** | **55,407.99** | **58,579.45** | **229,091.20** |

**STATE: ME - MAINE**

| REGISTRANT ZIP CODE 3 | 1ST QUARTER | 2ND QUARTER | 3RD QUARTER | 4TH QUARTER | TOTAL GRAMS |
|---|---|---|---|---|---|
| 039 | 561.34 | 550.93 | 559.60 | 600.07 | **2,271.93** |
| 040 | 5,717.99 | 5,588.49 | 5,947.49 | 6,120.34 | **23,374.32** |
| 041 | 2,125.10 | 1,964.38 | 2,038.96 | 2,051.68 | **8,180.12** |
| 042 | 2,094.46 | 1,970.16 | 2,082.32 | 2,188.69 | **8,335.62** |
| 043 | 982.19 | 928.43 | 1,094.34 | 1,100.12 | **4,105.09** |
| 044 | 2,038.96 | 1,781.13 | 2,005.43 | 2,104.86 | **7,930.38** |
| 045 | 385.01 | 360.16 | 465.37 | 426.64 | **1,637.18** |
| 046 | 757.89 | 809.34 | 878.13 | 924.96 | **3,370.32** |
| 047 | 766.56 | 704.70 | 764.83 | 776.39 | **3,012.48** |
| 048 | 378.08 | 358.42 | 401.20 | 366.52 | **1,504.22** |
| 049 | 1,817.55 | 1,756.27 | 1,868.42 | 1,936.64 | **7,378.87** |
| **ME - MAINE - Total** | **17,625.11** | **16,772.42** | **18,106.09** | **18,596.90** | **71,100.52** |

**STATE: MI - MICHIGAN**

| REGISTRANT ZIP CODE 3 | 1ST QUARTER | 2ND QUARTER | 3RD QUARTER | 4TH QUARTER | TOTAL GRAMS |
|---|---|---|---|---|---|
| 480 | 8,768.62 | 7,385.23 | 7,455.18 | 8,245.44 | **31,854.47** |
| 481 | 8,439.10 | 7,257.47 | 7,546.52 | 8,247.17 | **31,490.26** |
| 482 | 1,662.62 | 1,538.32 | 1,459.12 | 1,534.28 | **6,194.34** |
| 483 | 7,393.90 | 6,908.30 | 7,093.87 | 7,529.17 | **28,925.23** |
| 484 | 1,944.73 | 1,838.36 | 1,891.54 | 1,977.68 | **7,652.31** |
| 485 | 792.58 | 740.55 | 738.23 | 798.36 | **3,069.71** |
| 486 | 2,501.44 | 2,199.67 | 2,150.53 | 2,255.17 | **9,106.81** |
| 487 | 981.61 | 874.67 | 886.81 | 905.88 | **3,648.97** |
| 488 | 5,154.34 | 4,268.69 | 4,662.38 | 4,872.23 | **18,957.63** |
| 489 | 1,292.63 | 1,190.89 | 1,208.81 | 1,303.62 | **4,995.94** |
| 490 | 6,167.17 | 5,365.92 | 5,569.99 | 5,844.59 | **22,947.68** |
| 491 | 813.39 | 774.08 | 737.66 | 836.51 | **3,161.63** |
| 492 | 1,616.37 | 1,289.74 | 1,348.13 | 1,447.56 | **5,701.80** |
| 493 | 1,914.09 | 1,763.78 | 1,883.45 | 2,090.41 | **7,651.73** |
| 494 | 5,750.36 | 5,034.67 | 5,335.86 | 5,550.92 | **21,671.81** |
| 495 | 4,703.42 | 3,901.02 | 4,205.68 | 4,412.06 | **17,222.18** |
| 496 | 2,659.84 | 2,365.59 | 2,534.97 | 2,665.04 | **10,225.43** |
| 497 | 1,559.14 | 1,413.45 | 1,555.09 | 1,622.15 | **6,149.83** |
| 498 | 1,242.92 | 1,186.84 | 1,275.87 | 1,257.37 | **4,962.99** |

| | | | | | |
|---|---|---|---|---|---|
| 499 | 400.62 | 333.56 | 363.05 | 421.43 | **1,518.67** |
| **MI - MICHIGAN - Total** | **65,758.88** | **57,630.79** | **59,902.72** | **63,817.04** | **247,109.42** |
| **STATE: MN - MINNESOTA** | | | | | |
| REGISTRANT ZIP CODE 3 | 1ST QUARTER | 2ND QUARTER | 3RD QUARTER | 4TH QUARTER | **TOTAL GRAMS** |
| 550 | 3,315.98 | 3,231.00 | 3,320.61 | 3,543.17 | **13,410.76** |
| 551 | 4,632.89 | 4,427.09 | 4,634.63 | 5,095.95 | **18,790.56** |
| 553 | 6,166.01 | 6,053.29 | 6,183.94 | 6,589.76 | **24,993.00** |
| 554 | 7,004.26 | 6,443.50 | 6,698.44 | 7,167.86 | **27,314.07** |
| 556 | 10.41 | 27.17 | 26.59 | 17.92 | **82.09** |
| 557 | 1,314.60 | 1,249.85 | 1,341.19 | 1,449.87 | **5,355.52** |
| 558 | 522.02 | 490.23 | 531.85 | 563.07 | **2,107.17** |
| 559 | 1,457.39 | 1,402.47 | 1,631.40 | 1,686.32 | **6,177.58** |
| 560 | 3,162.21 | 3,014.79 | 3,093.41 | 3,481.90 | **12,752.31** |
| 561 | 684.47 | 695.45 | 625.50 | 693.14 | **2,698.57** |
| 562 | 829.00 | 767.72 | 791.42 | 856.17 | **3,244.30** |
| 563 | 1,537.75 | 1,522.72 | 1,497.86 | 1,566.65 | **6,124.97** |
| 564 | 881.02 | 863.10 | 1,007.63 | 922.65 | **3,674.40** |
| 565 | 1,536.59 | 1,475.31 | 1,573.59 | 1,718.11 | **6,303.60** |
| 566 | 931.90 | 703.55 | 781.59 | 796.04 | **3,213.08** |
| 567 | 502.37 | 519.71 | 498.32 | 574.05 | **2,094.46** |
| **MN - MINNESOTA - Total** | **34,488.87** | **32,886.95** | **34,237.97** | **36,722.65** | **138,336.44** |
| **STATE: MO - MISSOURI** | | | | | |
| REGISTRANT ZIP CODE 3 | 1ST QUARTER | 2ND QUARTER | 3RD QUARTER | 4TH QUARTER | **TOTAL GRAMS** |
| 630 | 7,541.31 | 6,802.50 | 7,003.10 | 7,434.37 | **28,781.29** |
| 631 | 25,328.30 | 25,154.87 | 25,312.11 | 26,219.73 | **102,014.99** |
| 633 | 7,537.85 | 6,720.41 | 7,145.32 | 7,198.50 | **28,602.08** |
| 634 | 585.04 | 557.29 | 671.17 | 626.66 | **2,440.16** |
| 635 | 516.24 | 497.74 | 541.68 | 539.37 | **2,095.03** |
| 636 | 884.49 | 776.39 | 875.82 | 939.41 | **3,476.12** |
| 637 | 541.10 | 452.65 | 517.40 | 569.43 | **2,080.58** |
| 638 | 1,055.61 | 867.15 | 960.80 | 1,018.61 | **3,902.18** |
| 639 | 367.67 | 361.89 | 336.45 | 390.22 | **1,456.23** |
| 640 | 5,750.94 | 5,443.39 | 5,734.17 | 5,786.78 | **22,715.28** |
| 641 | 5,979.87 | 5,550.34 | 5,738.22 | 5,886.21 | **23,154.64** |
| 644 | 404.09 | 364.78 | 389.06 | 364.78 | **1,522.72** |
| 645 | 947.51 | 883.91 | 964.85 | 995.49 | **3,791.76** |
| 646 | 186.73 | 158.98 | 162.45 | 167.07 | **675.22** |
| 647 | 759.05 | 680.42 | 659.03 | 783.33 | **2,881.83** |
| 648 | 1,126.14 | 872.35 | 1,026.71 | 1,032.49 | **4,057.68** |
| 650 | 530.12 | 531.85 | 512.77 | 546.30 | **2,121.05** |
| 651 | 751.53 | 612.79 | 676.96 | 744.59 | **2,785.86** |
| 652 | 2,084.05 | 1,920.45 | 1,941.84 | 2,111.22 | **8,057.56** |
| 653 | 382.70 | 301.19 | 347.44 | 356.69 | **1,388.02** |
| 654 | 476.35 | 363.62 | 379.81 | 441.09 | **1,660.88** |
| 655 | 476.35 | 431.26 | 427.22 | 451.50 | **1,786.33** |
| 656 | 607.58 | 661.92 | 704.70 | 641.69 | **2,615.90** |
| 657 | 1,192.62 | 1,063.13 | 1,082.20 | 1,061.97 | **4,399.92** |
| 658 | 1,708.86 | 1,504.79 | 1,552.20 | 1,609.43 | **6,375.29** |
| **MO - MISSOURI - Total** | **67,722.10** | **63,536.08** | **65,663.49** | **67,916.92** | **264,838.59** |
| **STATE: MS - MISSISSIPPI** | | | | | |
| REGISTRANT ZIP CODE 3 | 1ST QUARTER | 2ND QUARTER | 3RD QUARTER | 4TH QUARTER | **TOTAL GRAMS** |
| 386 | 6,616.35 | 5,533.00 | 5,787.36 | 6,207.64 | **24,144.35** |
| 387 | 497.17 | 393.69 | 420.86 | 458.43 | **1,770.14** |
| 388 | 2,971.43 | 2,515.89 | 2,821.71 | 2,871.42 | **11,180.45** |
| 389 | 797.20 | 586.77 | 640.53 | 671.17 | **2,695.68** |
| 390 | 2,644.81 | 2,203.72 | 2,460.97 | 2,626.31 | **9,935.80** |
| 391 | 3,043.70 | 2,441.89 | 2,782.40 | 2,787.02 | **11,055.01** |
| 392 | 2,868.53 | 2,153.42 | 2,437.85 | 2,569.08 | **10,028.88** |
| 393 | 2,594.51 | 2,012.94 | 2,191.58 | 2,375.41 | **9,174.45** |
| 394 | 6,440.61 | 4,980.33 | 5,556.12 | 5,754.41 | **22,731.47** |

| | | | | | |
|---|---|---|---|---|---|
| 395 | 5,881.01 | 4,618.44 | 5,160.70 | 5,231.23 | **20,891.38** |
| 396 | 1,689.21 | 1,293.21 | 1,358.54 | 1,459.70 | **5,800.66** |
| 397 | 1,708.86 | 1,165.45 | 1,361.43 | 1,419.81 | **5,655.55** |
| **MS - MISSISSIPPI - Total** | **37,753.40** | **29,898.75** | **32,980.03** | **34,431.64** | **135,063.82** |
| **STATE: MT - MONTANA** | | | | | |
| REGISTRANT ZIP CODE 3 | 1ST QUARTER | 2ND QUARTER | 3RD QUARTER | 4TH QUARTER | **TOTAL GRAMS** |
| 590 | 262.46 | 297.14 | 263.04 | 309.86 | **1,132.50** |
| 591 | 1,378.19 | 1,369.52 | 1,401.31 | 1,426.17 | **5,575.20** |
| 592 | 159.56 | 145.68 | 135.85 | 201.76 | **642.85** |
| 593 | 295.41 | 287.32 | 283.27 | 267.08 | **1,133.08** |
| 594 | 772.92 | 828.42 | 770.61 | 813.39 | **3,185.33** |
| 595 | 244.54 | 194.82 | 186.15 | 204.65 | **830.15** |
| 596 | 493.70 | 469.42 | 501.21 | 513.93 | **1,978.26** |
| 597 | 1,488.61 | 1,446.41 | 1,443.52 | 1,514.04 | **5,892.57** |
| 598 | 1,533.12 | 1,503.64 | 1,571.28 | 1,612.32 | **6,220.36** |
| 599 | 548.04 | 546.88 | 542.26 | 576.37 | **2,213.54** |
| **MT - MONTANA - Total** | **7,176.53** | **7,089.24** | **7,098.49** | **7,439.57** | **28,803.83** |
| **STATE: NC - NORTH CAROLINA** | | | | | |
| REGISTRANT ZIP CODE 3 | 1ST QUARTER | 2ND QUARTER | 3RD QUARTER | 4TH QUARTER | **TOTAL GRAMS** |
| 270 | 2,839.05 | 2,530.92 | 2,677.18 | 2,853.50 | **10,900.65** |
| 271 | 3,458.19 | 3,121.16 | 3,114.80 | 3,316.56 | **13,010.72** |
| 272 | 6,614.04 | 5,790.25 | 6,067.74 | 6,356.21 | **24,828.24** |
| 273 | 3,772.10 | 3,362.81 | 3,553.58 | 3,731.06 | **14,419.55** |
| 274 | 4,136.31 | 3,651.86 | 3,812.57 | 4,022.42 | **15,623.15** |
| 275 | 13,211.90 | 11,894.41 | 12,652.30 | 12,996.27 | **50,754.87** |
| 276 | 6,562.01 | 5,715.10 | 6,042.30 | 6,293.77 | **24,613.19** |
| 277 | 2,714.76 | 2,378.30 | 2,480.63 | 2,655.79 | **10,229.48** |
| 278 | 5,132.95 | 4,398.18 | 4,709.78 | 4,698.80 | **18,939.71** |
| 279 | 1,272.40 | 1,141.75 | 1,185.68 | 1,260.84 | **4,860.66** |
| 280 | 9,745.61 | 9,097.56 | 9,547.90 | 9,930.60 | **38,321.67** |
| 281 | 7,380.02 | 6,627.92 | 7,075.37 | 7,272.50 | **28,355.81** |
| 282 | 8,713.12 | 7,814.18 | 8,032.70 | 8,743.76 | **33,303.76** |
| 283 | 8,443.15 | 7,034.90 | 7,300.82 | 7,506.05 | **30,284.92** |
| 284 | 6,766.08 | 6,412.29 | 6,886.91 | 6,570.68 | **26,635.96** |
| 285 | 5,100.00 | 4,639.83 | 5,182.67 | 4,883.21 | **19,805.71** |
| 286 | 9,799.95 | 8,881.93 | 9,583.74 | 9,813.25 | **38,078.87** |
| 287 | 4,137.46 | 4,127.06 | 4,249.61 | 4,499.35 | **17,013.48** |
| 288 | 1,985.77 | 1,872.47 | 1,913.51 | 2,055.72 | **7,827.47** |
| 289 | 288.47 | 275.18 | 282.69 | 250.90 | **1,097.23** |
| **NC - NORTH CAROLINA - Total** | **112,073.36** | **100,768.03** | **106,352.48** | **109,711.24** | **428,905.11** |
| **STATE: ND - NORTH DAKOTA** | | | | | |
| REGISTRANT ZIP CODE 3 | 1ST QUARTER | 2ND QUARTER | 3RD QUARTER | 4TH QUARTER | **TOTAL GRAMS** |
| 580 | 607.58 | 578.10 | 577.52 | 600.07 | **2,363.27** |
| 581 | 1,893.28 | 1,794.42 | 1,647.59 | 1,997.34 | **7,332.62** |
| 582 | 1,290.32 | 1,161.98 | 1,092.03 | 1,359.69 | **4,904.02** |
| 583 | 337.03 | 311.02 | 296.57 | 300.03 | **1,244.65** |
| 584 | 232.40 | 176.32 | 225.46 | 230.66 | **864.84** |
| 585 | 635.91 | 672.91 | 642.27 | 700.08 | **2,651.17** |
| 586 | 186.15 | 216.21 | 197.13 | 189.62 | **789.11** |
| 587 | 1,055.03 | 1,001.27 | 982.19 | 1,083.36 | **4,121.85** |
| 588 | 332.41 | 322.00 | 320.27 | 317.38 | **1,292.05** |
| **ND - NORTH DAKOTA - Total** | **6,570.11** | **6,234.23** | **5,981.02** | **6,778.22** | **25,563.58** |
| **STATE: NE - NEBRASKA** | | | | | |
| REGISTRANT ZIP CODE 3 | 1ST QUARTER | 2ND QUARTER | 3RD QUARTER | 4TH QUARTER | **TOTAL GRAMS** |
| 680 | 1,690.94 | 1,534.86 | 1,650.48 | 1,787.49 | **6,663.76** |
| 681 | 7,491.60 | 6,852.22 | 7,147.05 | 7,509.52 | **29,000.39** |
| 683 | 475.78 | 424.90 | 464.79 | 529.54 | **1,895.01** |
| 684 | 420.86 | 369.98 | 393.69 | 428.37 | **1,612.90** |
| 685 | 4,291.24 | 4,005.65 | 4,366.39 | 4,443.85 | **17,107.14** |
| 686 | 416.23 | 374.61 | 437.62 | 483.87 | **1,712.33** |

| | | | | | |
|---|---|---|---|---|---|
| 687 | 625.50 | 564.23 | 650.94 | 653.25 | **2,493.92** |
| 688 | 1,579.95 | 1,348.13 | 1,511.15 | 1,523.29 | **5,962.52** |
| 689 | 516.24 | 432.42 | 446.29 | 493.12 | **1,888.07** |
| 690 | 194.24 | 169.38 | 186.73 | 174.01 | **724.36** |
| 691 | 398.31 | 328.94 | 389.06 | 389.06 | **1,505.37** |
| 692 | 10.41 | 13.30 | 21.97 | 19.08 | **64.75** |
| 693 | 461.90 | 457.28 | 468.84 | 479.24 | **1,867.26** |
| **NE - NEBRASKA - Total** | **18,573.20** | **16,875.90** | **18,135.00** | **18,913.70** | **72,497.79** |

**STATE: NH - NEW HAMPSHIRE**

| REGISTRANT ZIP CODE 3 | 1ST QUARTER | 2ND QUARTER | 3RD QUARTER | 4TH QUARTER | TOTAL GRAMS |
|---|---|---|---|---|---|
| 030 | 4,025.31 | 4,042.65 | 4,073.87 | 4,241.52 | **16,383.35** |
| 031 | 2,366.74 | 2,241.87 | 2,268.46 | 2,279.45 | **9,156.53** |
| 032 | 1,137.70 | 1,156.20 | 1,252.16 | 1,262.57 | **4,808.64** |
| 033 | 960.80 | 878.71 | 889.12 | 994.33 | **3,722.96** |
| 034 | 421.43 | 393.69 | 412.19 | 440.51 | **1,667.82** |
| 035 | 322.58 | 287.89 | 358.42 | 363.62 | **1,332.52** |
| 036 | 43.36 | 43.36 | 50.87 | 47.98 | **185.57** |
| 037 | 978.15 | 909.35 | 908.77 | 938.83 | **3,735.10** |
| 038 | 4,387.20 | 4,256.55 | 4,499.35 | 4,535.77 | **17,678.88** |
| **NH - NEW HAMPSHIRE - Total** | **14,643.27** | **14,210.28** | **14,713.22** | **15,104.60** | **58,671.37** |

**STATE: NJ - NEW JERSEY**

| REGISTRANT ZIP CODE 3 | 1ST QUARTER | 2ND QUARTER | 3RD QUARTER | 4TH QUARTER | TOTAL GRAMS |
|---|---|---|---|---|---|
| 070 | 9,241.51 | 8,014.78 | 8,354.70 | 8,727.00 | **34,337.98** |
| 071 | 579.83 | 470.00 | 375.77 | 471.15 | **1,896.75** |
| 072 | 329.52 | 291.36 | 283.27 | 260.15 | **1,164.29** |
| 073 | 750.37 | 615.68 | 679.85 | 689.10 | **2,734.99** |
| 074 | 3,521.21 | 3,285.92 | 3,391.13 | 3,674.98 | **13,873.24** |
| 075 | 280.96 | 264.19 | 193.09 | 276.91 | **1,015.14** |
| 076 | 3,627.58 | 3,209.03 | 3,128.68 | 3,411.95 | **13,377.23** |
| 077 | 3,319.45 | 3,280.14 | 3,543.17 | 3,516.58 | **13,659.35** |
| 078 | 2,932.12 | 2,649.43 | 2,676.60 | 2,989.93 | **11,248.09** |
| 079 | 3,217.70 | 3,010.74 | 2,791.07 | 3,139.08 | **12,158.60** |
| 080 | 8,866.90 | 8,042.53 | 7,943.67 | 8,428.12 | **33,281.22** |
| 081 | 601.80 | 534.16 | 502.37 | 561.91 | **2,200.25** |
| 082 | 1,122.67 | 1,209.96 | 1,330.79 | 1,231.93 | **4,895.35** |
| 083 | 818.01 | 716.84 | 734.19 | 720.31 | **2,989.36** |
| 084 | 95.39 | 121.98 | 165.91 | 132.38 | **515.67** |
| 085 | 2,115.27 | 2,103.71 | 1,893.86 | 2,047.63 | **8,160.46** |
| 086 | 1,451.03 | 1,256.79 | 1,271.82 | 1,346.97 | **5,326.61** |
| 087 | 2,219.33 | 2,302.57 | 2,210.65 | 2,280.60 | **9,013.16** |
| 088 | 4,170.41 | 4,151.91 | 3,881.36 | 4,090.64 | **16,294.33** |
| 089 | 423.75 | 339.34 | 344.55 | 394.84 | **1,502.48** |
| **NJ - NEW JERSEY - Total** | **49,684.80** | **45,871.08** | **45,696.49** | **48,392.17** | **189,644.55** |

**STATE: NM - NEW MEXICO**

| REGISTRANT ZIP CODE 3 | 1ST QUARTER | 2ND QUARTER | 3RD QUARTER | 4TH QUARTER | TOTAL GRAMS |
|---|---|---|---|---|---|
| 870 | 153.77 | 184.99 | 199.44 | 171.12 | **709.33** |
| 871 | 1,618.68 | 1,520.98 | 1,594.40 | 1,564.92 | **6,298.98** |
| 873 | 22.55 | 19.08 | 18.50 | 10.98 | **71.11** |
| 874 | 148.57 | 144.53 | 149.73 | 160.13 | **602.96** |
| 875 | 369.98 | 397.73 | 415.08 | 378.66 | **1,561.45** |
| 877 | 44.51 | 56.65 | 27.17 | 54.34 | **182.68** |
| 878 | 30.64 | 38.15 | 37.00 | 24.86 | **130.65** |
| 879 | 12.14 | 22.55 | 20.23 | 12.14 | **67.06** |
| 880 | 469.42 | 461.90 | 556.71 | 543.99 | **2,032.02** |
| 881 | 89.61 | 106.37 | 80.36 | 79.78 | **356.11** |
| 882 | 522.60 | 476.93 | 499.48 | 445.14 | **1,944.15** |
| 883 | 170.54 | 209.85 | 163.02 | 217.94 | **761.36** |
| 884 | 18.50 | 35.26 | 21.39 | 24.86 | **100.01** |
| **NM - NEW MEXICO - Total** | **3,671.51** | **3,674.98** | **3,782.51** | **3,688.86** | **14,817.86** |

**STATE: NV - NEVADA**

| REGISTRANT ZIP CODE 3 | 1ST QUARTER | 2ND QUARTER | 3RD QUARTER | 4TH QUARTER | TOTAL GRAMS |
|---|---|---|---|---|---|
| 890 | 1,978.84 | 1,675.33 | 1,654.52 | 1,641.23 | **6,949.92** |
| 891 | 4,269.27 | 3,769.79 | 3,506.75 | 3,485.94 | **15,031.76** |
| 893 | 2.89 | 5.78 | 2.89 | 2.89 | **14.45** |
| 894 | 718.58 | 708.17 | 700.66 | 763.67 | **2,891.08** |
| 895 | 1,082.78 | 971.79 | 912.82 | 1,043.47 | **4,010.86** |
| 897 | 216.79 | 184.41 | 216.21 | 178.63 | **796.04** |
| 898 | 89.61 | 83.82 | 106.95 | 79.78 | **360.16** |
| **NV - NEVADA - Total** | **8,358.75** | **7,399.10** | **7,100.80** | **7,195.61** | **30,054.26** |

**STATE: NY - NEW YORK**

| REGISTRANT ZIP CODE 3 | 1ST QUARTER | 2ND QUARTER | 3RD QUARTER | 4TH QUARTER | TOTAL GRAMS |
|---|---|---|---|---|---|
| 100 | 19,943.87 | 11,842.96 | 13,441.98 | 16,423.24 | **61,652.05** |
| 101 | 790.26 | 482.14 | 398.31 | 528.38 | **2,199.09** |
| 102 | 130.65 | 66.48 | 64.75 | 79.20 | **341.08** |
| 103 | 1,087.98 | 1,026.13 | 1,012.83 | 987.39 | **4,114.34** |
| 104 | 1,101.28 | 1,021.50 | 932.48 | 1,028.44 | **4,083.70** |
| 105 | 5,582.71 | 5,224.29 | 5,616.82 | 5,716.83 | **22,140.65** |
| 106 | 625.50 | 437.62 | 548.04 | 608.16 | **2,219.33** |
| 107 | 724.94 | 685.63 | 693.14 | 746.91 | **2,850.61** |
| 108 | 472.89 | 421.43 | 450.92 | 430.11 | **1,775.35** |
| 109 | 2,875.47 | 2,647.12 | 2,635.56 | 2,808.41 | **10,966.56** |
| 110 | 1,411.14 | 1,416.35 | 1,392.64 | 1,556.82 | **5,776.95** |
| 111 | 871.20 | 748.06 | 896.06 | 881.02 | **3,396.34** |
| 112 | 6,175.26 | 5,512.18 | 5,307.54 | 6,141.16 | **23,136.14** |
| 113 | 1,854.54 | 1,487.45 | 1,528.50 | 1,764.36 | **6,634.85** |
| 114 | 490.81 | 440.51 | 430.68 | 466.53 | **1,828.53** |
| 115 | 4,446.17 | 4,227.65 | 4,387.20 | 4,793.03 | **17,854.04** |
| 116 | 267.08 | 146.84 | 180.37 | 172.27 | **766.56** |
| 117 | 9,409.73 | 8,691.16 | 8,947.83 | 9,385.45 | **36,434.17** |
| 118 | 598.91 | 466.53 | 526.65 | 561.34 | **2,153.42** |
| 119 | 1,164.87 | 1,942.42 | 2,096.19 | 1,515.78 | **6,719.26** |
| 120 | 1,911.20 | 1,904.26 | 1,985.77 | 1,906.00 | **7,707.23** |
| 121 | 1,190.89 | 1,019.19 | 1,204.76 | 1,176.43 | **4,591.27** |
| 122 | 707.02 | 659.61 | 670.02 | 723.78 | **2,760.43** |
| 123 | 834.78 | 755.58 | 790.84 | 784.48 | **3,165.68** |
| 124 | 499.48 | 637.64 | 536.48 | 526.65 | **2,200.25** |
| 125 | 2,439.58 | 2,473.69 | 2,555.20 | 2,748.87 | **10,217.34** |
| 126 | 508.15 | 453.81 | 517.40 | 549.77 | **2,029.13** |
| 127 | 306.39 | 335.88 | 337.03 | 353.80 | **1,333.10** |
| 128 | 1,516.36 | 1,393.20 | 1,467.22 | 1,517.50 | **5,894.28** |
| 129 | 555.55 | 510.46 | 581.57 | 626.08 | **2,273.67** |
| 130 | 1,849.92 | 1,768.41 | 1,729.68 | 1,775.92 | **7,123.93** |
| 131 | 722.63 | 696.61 | 720.89 | 704.70 | **2,844.83** |
| 132 | 996.07 | 786.79 | 898.95 | 900.68 | **3,582.49** |
| 133 | 434.73 | 417.97 | 430.11 | 477.51 | **1,760.31** |
| 134 | 676.96 | 694.88 | 670.02 | 646.32 | **2,688.17** |
| 135 | 286.74 | 184.41 | 191.93 | 201.76 | **864.84** |
| 136 | 1,319.80 | 1,144.64 | 1,231.35 | 1,204.76 | **4,900.55** |
| 137 | 608.74 | 541.68 | 474.62 | 512.77 | **2,137.81** |
| 138 | 804.72 | 692.56 | 748.64 | 729.56 | **2,975.48** |
| 139 | 585.62 | 437.04 | 515.67 | 534.74 | **2,073.07** |
| 140 | 1,737.19 | 1,652.21 | 1,733.72 | 1,711.18 | **6,834.30** |
| 141 | 687.36 | 660.19 | 617.99 | 611.63 | **2,577.17** |
| 142 | 2,424.55 | 2,322.81 | 2,295.64 | 2,406.05 | **9,449.04** |
| 143 | 204.65 | 150.31 | 183.84 | 157.82 | **696.61** |
| 144 | 2,521.09 | 2,332.06 | 2,386.40 | 2,533.23 | **9,772.78** |
| 145 | 1,856.28 | 1,721.58 | 1,672.44 | 1,863.79 | **7,114.10** |
| 146 | 2,975.48 | 2,751.18 | 2,800.32 | 2,977.22 | **11,504.19** |
| 147 | 566.54 | 519.71 | 574.63 | 582.72 | **2,243.61** |
| 148 | 1,290.32 | 1,038.85 | 1,114.00 | 1,270.66 | **4,713.83** |

| | | | | | |
|---|---|---|---|---|---|
| 149 | 335.88 | 269.97 | 249.16 | 278.07 | **1,133.08** |
| **NY - NEW YORK - Total** | **93,379.91** | **79,861.61** | **83,374.74** | **89,589.30** | **346,205.56** |

**STATE: OH - OHIO**

| REGISTRANT ZIP CODE 3 | 1ST QUARTER | 2ND QUARTER | 3RD QUARTER | 4TH QUARTER | TOTAL GRAMS |
|---|---|---|---|---|---|
| 430 | 9,520.15 | 8,604.44 | 9,128.20 | 9,312.03 | **36,564.83** |
| 431 | 3,791.18 | 3,480.74 | 3,481.90 | 3,577.86 | **14,331.68** |
| 432 | 6,472.99 | 5,995.48 | 6,218.04 | 6,516.92 | **25,203.43** |
| 433 | 890.27 | 804.72 | 922.07 | 869.46 | **3,486.52** |
| 434 | 950.97 | 966.58 | 977.57 | 999.53 | **3,894.66** |
| 435 | 2,229.15 | 2,045.32 | 2,112.38 | 2,142.44 | **8,529.29** |
| 436 | 2,767.36 | 2,450.57 | 2,581.22 | 2,640.18 | **10,439.33** |
| 437 | 1,045.20 | 925.54 | 979.30 | 1,015.14 | **3,965.19** |
| 438 | 216.21 | 162.45 | 195.98 | 176.90 | **751.53** |
| 439 | 1,264.30 | 1,055.61 | 1,124.98 | 1,170.07 | **4,614.97** |
| 440 | 9,212.60 | 8,438.53 | 8,829.90 | 9,062.30 | **35,543.32** |
| 441 | 10,487.89 | 9,367.53 | 9,541.54 | 9,861.23 | **39,258.19** |
| 442 | 4,794.76 | 4,440.96 | 4,577.40 | 4,904.60 | **18,717.72** |
| 443 | 1,793.84 | 1,661.46 | 1,765.52 | 1,829.69 | **7,050.51** |
| 444 | 2,208.92 | 2,033.76 | 1,998.49 | 2,139.55 | **8,380.72** |
| 445 | 1,922.76 | 1,677.65 | 1,762.05 | 1,858.59 | **7,221.05** |
| 446 | 3,084.16 | 2,760.43 | 2,898.59 | 2,986.46 | **11,729.65** |
| 447 | 2,438.43 | 2,203.72 | 2,323.96 | 2,366.16 | **9,332.27** |
| 448 | 1,785.17 | 1,712.33 | 1,752.80 | 1,767.25 | **7,017.56** |
| 449 | 686.78 | 673.49 | 753.26 | 793.73 | **2,907.26** |
| 450 | 6,591.50 | 5,697.18 | 6,268.34 | 6,461.42 | **25,018.43** |
| 451 | 2,406.05 | 2,115.27 | 2,175.39 | 2,412.99 | **9,109.70** |
| 452 | 7,670.81 | 6,356.21 | 6,930.26 | 7,241.86 | **28,199.14** |
| 453 | 3,572.66 | 3,061.62 | 3,216.55 | 3,221.75 | **13,072.58** |
| 454 | 3,474.38 | 2,924.03 | 3,135.04 | 3,425.24 | **12,958.69** |
| 455 | 609.90 | 549.20 | 611.05 | 582.15 | **2,352.29** |
| 456 | 1,871.31 | 1,590.35 | 1,754.53 | 1,770.72 | **6,986.92** |
| 457 | 1,294.94 | 1,059.66 | 1,154.47 | 1,226.73 | **4,735.80** |
| 458 | 1,085.67 | 960.80 | 1,060.24 | 1,036.53 | **4,143.24** |
| **OH - OHIO - Total** | **96,140.34** | **85,775.59** | **90,231.00** | **93,369.51** | **365,516.44** |

**STATE: OK - OKLAHOMA**

| REGISTRANT ZIP CODE 3 | 1ST QUARTER | 2ND QUARTER | 3RD QUARTER | 4TH QUARTER | TOTAL GRAMS |
|---|---|---|---|---|---|
| 730 | 9,417.25 | 8,091.67 | 8,720.64 | 9,373.31 | **35,602.87** |
| 731 | 7,267.87 | 6,396.68 | 6,690.35 | 6,993.28 | **27,348.18** |
| 734 | 677.53 | 580.41 | 631.29 | 638.80 | **2,528.03** |
| 735 | 1,336.57 | 1,087.98 | 1,245.23 | 1,205.34 | **4,875.12** |
| 736 | 346.86 | 287.89 | 316.22 | 337.61 | **1,288.58** |
| 737 | 955.60 | 825.53 | 892.59 | 938.26 | **3,611.97** |
| 738 | 119.67 | 74.57 | 87.87 | 93.65 | **375.77** |
| 739 | 68.79 | 56.08 | 53.19 | 56.08 | **234.13** |
| 740 | 7,480.04 | 6,470.10 | 6,822.16 | 7,263.25 | **28,035.54** |
| 741 | 4,378.53 | 3,879.63 | 4,104.51 | 4,038.61 | **16,401.28** |
| 743 | 1,222.10 | 1,029.02 | 1,022.66 | 1,003.00 | **4,276.78** |
| 744 | 1,760.31 | 1,453.92 | 1,633.71 | 1,688.05 | **6,536.00** |
| 745 | 757.31 | 593.13 | 662.50 | 682.74 | **2,695.68** |
| 746 | 367.67 | 304.66 | 342.81 | 354.95 | **1,370.10** |
| 747 | 704.70 | 571.16 | 586.19 | 644.58 | **2,506.64** |
| 748 | 1,838.94 | 1,364.89 | 1,519.25 | 1,636.02 | **6,359.10** |
| 749 | 556.71 | 437.04 | 489.65 | 456.70 | **1,940.10** |
| **OK - OKLAHOMA - Total** | **39,256.46** | **33,504.36** | **35,820.81** | **37,404.23** | **145,985.86** |

**STATE: OR - OREGON**

| REGISTRANT ZIP CODE 3 | 1ST QUARTER | 2ND QUARTER | 3RD QUARTER | 4TH QUARTER | TOTAL GRAMS |
|---|---|---|---|---|---|
| 970 | 3,738.57 | 3,307.31 | 3,466.87 | 3,690.59 | **14,203.34** |
| 971 | 1,172.39 | 986.82 | 1,085.09 | 1,178.17 | **4,422.47** |
| 972 | 5,249.73 | 4,837.54 | 5,178.04 | 5,464.78 | **20,730.09** |
| 973 | 1,888.07 | 1,707.71 | 1,789.22 | 1,789.22 | **7,174.22** |

| | | | | | |
|---|---|---|---|---|---|
| 974 | 2,073.07 | 1,896.17 | 1,958.60 | 2,011.21 | **7,939.05** |
| 975 | 733.03 | 649.78 | 771.19 | 723.20 | **2,877.20** |
| 976 | 89.61 | 98.86 | 98.28 | 113.89 | **400.62** |
| 977 | 1,278.76 | 1,255.06 | 1,497.28 | 1,484.56 | **5,515.65** |
| 978 | 472.89 | 436.47 | 456.70 | 437.62 | **1,803.67** |
| 979 | 113.89 | 89.03 | 89.61 | 108.10 | **400.62** |
| **OR - OREGON - Total** | **16,809.99** | **15,264.73** | **16,390.87** | **17,001.34** | **65,466.93** |

**STATE: PA - PENNSYLVANIA**

| REGISTRANT ZIP CODE 3 | 1ST QUARTER | 2ND QUARTER | 3RD QUARTER | 4TH QUARTER | TOTAL GRAMS |
|---|---|---|---|---|---|
| 150 | 2,849.45 | 2,717.65 | 2,634.40 | 2,884.14 | **11,085.65** |
| 151 | 2,338.41 | 2,142.44 | 2,273.67 | 2,265.57 | **9,020.09** |
| 152 | 5,086.70 | 4,709.78 | 4,618.44 | 4,923.10 | **19,338.02** |
| 153 | 1,258.52 | 1,061.97 | 1,094.34 | 1,103.59 | **4,518.43** |
| 154 | 818.01 | 800.09 | 784.48 | 781.59 | **3,184.17** |
| 155 | 356.69 | 319.11 | 243.96 | 296.57 | **1,216.32** |
| 156 | 1,952.82 | 1,631.40 | 1,625.04 | 1,748.17 | **6,957.43** |
| 157 | 1,185.68 | 959.07 | 992.60 | 1,031.33 | **4,168.68** |
| 158 | 1,024.39 | 955.02 | 959.65 | 975.83 | **3,914.89** |
| 159 | 824.37 | 720.31 | 759.62 | 724.94 | **3,029.24** |
| 160 | 1,577.06 | 1,501.90 | 1,520.98 | 1,531.97 | **6,131.91** |
| 161 | 1,725.05 | 1,584.57 | 1,700.19 | 1,625.62 | **6,635.43** |
| 162 | 668.28 | 597.76 | 641.11 | 643.43 | **2,550.58** |
| 163 | 1,152.73 | 881.60 | 971.79 | 1,025.55 | **4,031.67** |
| 164 | 725.52 | 624.93 | 597.76 | 632.44 | **2,580.64** |
| 165 | 2,855.81 | 2,700.31 | 2,537.86 | 2,611.86 | **10,705.83** |
| 166 | 1,490.34 | 1,360.85 | 1,386.86 | 1,314.02 | **5,552.07** |
| 167 | 608.16 | 523.76 | 552.66 | 549.77 | **2,234.36** |
| 168 | 1,379.92 | 1,087.98 | 1,237.13 | 1,223.84 | **4,928.88** |
| 169 | 337.03 | 334.14 | 349.75 | 339.92 | **1,360.85** |
| 170 | 4,766.43 | 4,312.05 | 4,524.21 | 4,461.20 | **18,063.89** |
| 171 | 1,198.98 | 1,064.86 | 1,031.33 | 1,055.03 | **4,350.20** |
| 172 | 1,315.76 | 1,249.85 | 1,306.51 | 1,337.72 | **5,209.84** |
| 173 | 2,249.97 | 1,982.88 | 2,099.66 | 2,141.28 | **8,473.79** |
| 174 | 1,280.49 | 1,156.78 | 1,097.23 | 1,210.54 | **4,745.04** |
| 175 | 1,322.11 | 1,074.11 | 1,224.99 | 1,285.12 | **4,906.33** |
| 176 | 1,575.32 | 1,441.78 | 1,443.52 | 1,471.84 | **5,932.46** |
| 177 | 846.34 | 700.66 | 811.65 | 815.12 | **3,173.77** |
| 178 | 822.06 | 731.87 | 801.82 | 731.87 | **3,087.63** |
| 179 | 301.77 | 278.07 | 298.30 | 261.88 | **1,140.01** |
| 180 | 2,179.44 | 1,918.14 | 2,055.72 | 2,083.47 | **8,236.77** |
| 181 | 897.21 | 689.67 | 793.73 | 809.34 | **3,189.96** |
| 182 | 216.21 | 179.21 | 223.15 | 215.63 | **834.20** |
| 183 | 603.54 | 504.68 | 613.36 | 607.58 | **2,329.16** |
| 184 | 613.36 | 694.88 | 734.19 | 698.34 | **2,740.77** |
| 185 | 442.25 | 465.95 | 462.48 | 507.57 | **1,878.25** |
| 186 | 456.12 | 475.20 | 429.53 | 553.82 | **1,914.67** |
| 187 | 13,835.09 | 14,878.56 | 15,087.25 | 15,309.82 | **59,110.73** |
| 188 | 235.29 | 209.85 | 224.88 | 197.13 | **867.15** |
| 189 | 2,373.10 | 2,266.15 | 2,218.17 | 2,401.43 | **9,258.85** |
| 190 | 8,076.06 | 7,378.29 | 7,175.38 | 7,696.25 | **30,325.97** |
| 191 | 7,173.64 | 6,078.14 | 6,263.14 | 6,679.37 | **26,194.29** |
| 193 | 4,610.93 | 4,163.48 | 4,082.54 | 4,612.08 | **17,469.03** |
| 194 | 4,011.44 | 3,837.43 | 3,729.32 | 3,915.47 | **15,493.66** |
| 195 | 955.60 | 849.23 | 949.24 | 924.96 | **3,679.03** |
| 196 | 1,752.80 | 1,468.37 | 1,423.86 | 1,500.75 | **6,145.78** |
| **PA - PENNSYLVANIA - Total** | **94,326.26** | **87,264.77** | **88,587.47** | **91,717.88** | **361,896.38** |

**STATE: PR - PUERTO RICO**

| REGISTRANT ZIP CODE 3 | 1ST QUARTER | 2ND QUARTER | 3RD QUARTER | 4TH QUARTER | TOTAL GRAMS |
|---|---|---|---|---|---|
| 006 | 408.14 | 262.46 | 297.72 | 275.18 | **1,243.49** |
| 007 | 257.25 | 193.09 | 225.46 | 246.85 | **922.65** |

| | | | | | |
|---|---|---|---|---|---|
| 009 | 1,530.81 | 1,255.63 | 1,327.90 | 1,488.61 | **5,602.95** |
| **PR - PUERTO RICO - Total** | **2,196.20** | **1,711.18** | **1,851.08** | **2,010.63** | **7,769.09** |

**STATE: RI - RHODE ISLAND**

| REGISTRANT ZIP CODE 3 | 1ST QUARTER | 2ND QUARTER | 3RD QUARTER | 4TH QUARTER | TOTAL GRAMS |
|---|---|---|---|---|---|
| 028 | 5,407.55 | 5,037.56 | 5,372.28 | 4,982.64 | **20,800.04** |
| 029 | 2,527.45 | 2,357.49 | 2,462.71 | 2,370.21 | **9,717.86** |
| **RI - RHODE ISLAND - Total** | **7,935.00** | **7,395.06** | **7,834.99** | **7,352.85** | **30,517.90** |

**STATE: SC - SOUTH CAROLINA**

| REGISTRANT ZIP CODE 3 | 1ST QUARTER | 2ND QUARTER | 3RD QUARTER | 4TH QUARTER | TOTAL GRAMS |
|---|---|---|---|---|---|
| 290 | 5,613.35 | 4,969.35 | 5,194.81 | 5,367.66 | **21,145.16** |
| 291 | 3,389.40 | 3,175.50 | 3,028.09 | 3,391.71 | **12,984.70** |
| 292 | 4,832.34 | 3,810.84 | 4,355.98 | 4,255.97 | **17,255.13** |
| 293 | 4,919.05 | 4,273.32 | 4,479.70 | 4,675.09 | **18,347.16** |
| 294 | 13,768.38 | 12,106.10 | 12,530.87 | 13,093.23 | **51,498.57** |
| 295 | 8,019.98 | 6,755.68 | 7,002.53 | 7,255.16 | **29,033.34** |
| 296 | 15,286.12 | 13,184.73 | 13,622.93 | 14,599.34 | **56,693.11** |
| 297 | 3,688.86 | 3,343.15 | 3,506.75 | 3,600.41 | **14,139.17** |
| 298 | 2,323.96 | 1,985.77 | 2,100.82 | 2,103.71 | **8,514.26** |
| 299 | 1,919.29 | 1,739.50 | 1,783.44 | 1,820.44 | **7,262.67** |
| **SC - SOUTH CAROLINA - Total** | **63,760.73** | **55,343.94** | **57,605.90** | **60,162.71** | **236,873.27** |

**STATE: SD - SOUTH DAKOTA**

| REGISTRANT ZIP CODE 3 | 1ST QUARTER | 2ND QUARTER | 3RD QUARTER | 4TH QUARTER | TOTAL GRAMS |
|---|---|---|---|---|---|
| 570 | 1,811.19 | 1,641.80 | 1,644.12 | 1,710.60 | **6,807.71** |
| 571 | 4,492.42 | 3,820.08 | 3,876.16 | 3,711.40 | **15,900.06** |
| 572 | 453.81 | 408.14 | 429.53 | 439.93 | **1,731.41** |
| 573 | 981.61 | 807.61 | 893.16 | 970.63 | **3,653.01** |
| 574 | 544.57 | 479.82 | 522.02 | 501.21 | **2,047.63** |
| 575 | 308.71 | 273.44 | 263.04 | 281.53 | **1,126.72** |
| 576 | 24.28 | 38.15 | 23.12 | 35.84 | **121.40** |
| 577 | 1,093.19 | 1,065.44 | 986.82 | 1,122.09 | **4,267.53** |
| **SD - SOUTH DAKOTA - Total** | **9,709.77** | **8,534.49** | **8,637.97** | **8,773.25** | **35,655.47** |

**STATE: TN - TENNESSEE**

| REGISTRANT ZIP CODE 3 | 1ST QUARTER | 2ND QUARTER | 3RD QUARTER | 4TH QUARTER | TOTAL GRAMS |
|---|---|---|---|---|---|
| 370 | 10,669.41 | 9,331.69 | 10,175.72 | 10,449.16 | **40,625.98** |
| 371 | 7,141.27 | 6,275.28 | 6,839.50 | 7,082.88 | **27,338.93** |
| 372 | 7,196.77 | 6,223.25 | 6,771.29 | 6,987.49 | **27,178.79** |
| 373 | 5,532.42 | 4,571.61 | 5,053.17 | 5,336.44 | **20,493.65** |
| 374 | 3,069.13 | 2,607.23 | 2,918.25 | 2,860.44 | **11,455.05** |
| 376 | 2,349.98 | 2,065.55 | 2,087.52 | 2,395.65 | **8,898.69** |
| 377 | 2,957.56 | 2,566.76 | 2,732.10 | 3,099.19 | **11,355.62** |
| 378 | 4,237.47 | 3,792.34 | 4,053.64 | 4,342.11 | **16,425.56** |
| 379 | 5,802.97 | 5,210.99 | 5,733.60 | 5,957.32 | **22,704.88** |
| 380 | 5,008.08 | 4,379.69 | 4,636.94 | 4,814.99 | **18,839.70** |
| 381 | 7,019.87 | 5,976.40 | 6,367.77 | 6,651.04 | **26,015.08** |
| 382 | 826.10 | 657.88 | 736.50 | 764.83 | **2,985.31** |
| 383 | 2,160.36 | 1,697.88 | 1,916.40 | 2,064.40 | **7,839.04** |
| 384 | 1,826.22 | 1,534.28 | 1,652.21 | 1,789.80 | **6,802.50** |
| 385 | 1,596.13 | 1,388.02 | 1,514.04 | 1,601.92 | **6,100.11** |
| **TN - TENNESSEE - Total** | **67,393.74** | **58,278.84** | **63,188.64** | **66,197.65** | **255,058.88** |

**STATE: TX - TEXAS**

| REGISTRANT ZIP CODE 3 | 1ST QUARTER | 2ND QUARTER | 3RD QUARTER | 4TH QUARTER | TOTAL GRAMS |
|---|---|---|---|---|---|
| 750 | 30,286.66 | 26,811.12 | 28,846.61 | 29,877.94 | **115,822.34** |
| 751 | 7,317.01 | 6,413.44 | 6,813.49 | 6,972.46 | **27,516.40** |
| 752 | 14,948.51 | 13,221.73 | 13,679.00 | 14,181.95 | **56,031.19** |
| 753 | 35.84 | 17.92 | 29.48 | 23.12 | **106.37** |
| 754 | 2,678.92 | 2,333.79 | 2,691.06 | 2,666.20 | **10,369.96** |
| 755 | 880.45 | 675.80 | 793.15 | 832.46 | **3,181.86** |
| 756 | 3,450.68 | 2,718.80 | 3,106.13 | 3,078.96 | **12,354.58** |
| 757 | 3,488.83 | 2,998.03 | 3,145.44 | 3,437.38 | **13,069.68** |
| 758 | 760.78 | 645.16 | 652.10 | 730.72 | **2,788.75** |

| | | | | | |
|---|---|---|---|---|---|
| 759 | 1,931.43 | 1,540.64 | 1,744.71 | 1,789.22 | **7,005.99** |
| 760 | 12,770.81 | 11,314.00 | 12,245.31 | 12,571.36 | **48,901.48** |
| 761 | 6,993.28 | 6,095.49 | 6,366.62 | 6,723.30 | **26,178.68** |
| 762 | 6,140.58 | 5,648.04 | 6,096.64 | 6,386.27 | **24,271.53** |
| 763 | 1,669.55 | 1,395.53 | 1,487.45 | 1,519.82 | **6,072.36** |
| 764 | 690.83 | 535.90 | 614.52 | 644.58 | **2,485.83** |
| 765 | 2,958.14 | 2,240.72 | 2,643.07 | 2,502.02 | **10,343.94** |
| 766 | 675.80 | 623.19 | 657.88 | 638.80 | **2,595.67** |
| 767 | 3,109.60 | 2,318.76 | 2,596.25 | 2,663.31 | **10,687.91** |
| 768 | 474.62 | 374.61 | 342.24 | 356.69 | **1,548.15** |
| 769 | 1,489.19 | 1,150.42 | 1,254.48 | 1,337.72 | **5,231.81** |
| 770 | 25,249.10 | 22,217.54 | 22,388.08 | 23,208.40 | **93,063.12** |
| 773 | 19,244.37 | 16,921.57 | 17,490.99 | 18,628.12 | **72,285.05** |
| 774 | 15,521.99 | 13,374.92 | 14,097.55 | 14,906.31 | **57,900.76** |
| 775 | 11,843.53 | 9,800.53 | 10,753.24 | 11,186.81 | **43,584.12** |
| 776 | 3,399.81 | 2,754.65 | 3,006.12 | 3,154.11 | **12,314.69** |
| 777 | 1,424.44 | 1,357.38 | 1,261.99 | 1,319.80 | **5,363.61** |
| 778 | 5,138.73 | 4,032.83 | 4,698.22 | 4,979.18 | **18,848.95** |
| 779 | 1,686.90 | 1,233.09 | 1,471.84 | 1,527.34 | **5,919.17** |
| 780 | 5,127.75 | 4,091.79 | 4,587.22 | 4,597.63 | **18,404.39** |
| 781 | 3,921.25 | 3,401.54 | 3,435.07 | 3,726.43 | **14,484.30** |
| 782 | 16,506.49 | 13,747.80 | 14,416.08 | 14,995.91 | **59,666.28** |
| 783 | 1,751.64 | 1,492.65 | 1,570.70 | 1,423.86 | **6,238.86** |
| 784 | 3,409.06 | 2,716.49 | 2,836.74 | 2,991.09 | **11,953.37** |
| 785 | 7,166.13 | 5,469.40 | 5,637.05 | 5,982.18 | **24,254.76** |
| 786 | 9,809.20 | 8,335.62 | 9,132.82 | 9,476.22 | **36,753.86** |
| 787 | 14,729.41 | 13,489.39 | 14,262.88 | 14,925.39 | **57,407.06** |
| 788 | 641.11 | 448.61 | 515.67 | 604.69 | **2,210.08** |
| 789 | 436.47 | 358.42 | 401.20 | 401.20 | **1,597.29** |
| 790 | 596.60 | 461.32 | 542.26 | 538.21 | **2,138.39** |
| 791 | 1,409.41 | 1,353.91 | 1,366.05 | 1,307.66 | **5,437.03** |
| 792 | 96.54 | 87.29 | 63.59 | 54.34 | **301.77** |
| 793 | 381.55 | 250.32 | 341.08 | 264.77 | **1,237.71** |
| 794 | 3,226.95 | 2,497.97 | 2,970.86 | 2,963.34 | **11,659.12** |
| 795 | 443.98 | 349.17 | 364.78 | 367.67 | **1,525.61** |
| 796 | 1,299.57 | 1,057.92 | 1,238.87 | 1,245.81 | **4,842.17** |
| 797 | 2,841.36 | 2,371.94 | 2,449.41 | 2,521.09 | **10,183.81** |
| 798 | 36.42 | 41.62 | 31.22 | 46.25 | **155.51** |
| 799 | 2,585.26 | 1,978.26 | 2,192.16 | 2,177.70 | **8,933.38** |
| **TX - TEXAS - Total** | **262,676.50** | **224,767.01** | **239,329.35** | **248,455.82** | **975,228.69** |

**STATE: UT - UTAH**

| REGISTRANT ZIP CODE 3 | 1ST QUARTER | 2ND QUARTER | 3RD QUARTER | 4TH QUARTER | **TOTAL GRAMS** |
|---|---|---|---|---|---|
| 840 | 13,853.01 | 13,674.38 | 13,947.82 | 14,625.35 | **56,100.56** |
| 841 | 4,038.03 | 3,998.14 | 4,030.51 | 4,266.38 | **16,333.06** |
| 843 | 849.23 | 831.31 | 820.90 | 939.99 | **3,441.43** |
| 844 | 1,185.11 | 1,126.14 | 1,188.00 | 1,206.49 | **4,705.73** |
| 845 | 147.99 | 140.48 | 116.20 | 164.76 | **569.43** |
| 846 | 2,201.40 | 1,986.35 | 2,140.70 | 2,186.95 | **8,515.41** |
| 847 | 990.86 | 1,005.32 | 1,103.01 | 1,140.01 | **4,239.21** |
| **UT - UTAH - Total** | **23,265.63** | **22,762.11** | **23,347.15** | **24,529.94** | **93,904.83** |

**STATE: VA - VIRGINIA**

| REGISTRANT ZIP CODE 3 | 1ST QUARTER | 2ND QUARTER | 3RD QUARTER | 4TH QUARTER | **TOTAL GRAMS** |
|---|---|---|---|---|---|
| 201 | 8,100.34 | 6,718.10 | 7,310.07 | 7,385.23 | **29,513.74** |
| 220 | 4,205.68 | 3,362.23 | 3,712.56 | 3,497.51 | **14,777.97** |
| 221 | 3,028.09 | 2,610.70 | 3,014.21 | 2,840.78 | **11,493.78** |
| 222 | 2,496.81 | 2,292.17 | 2,415.30 | 2,291.01 | **9,495.29** |
| 223 | 2,112.38 | 1,884.03 | 1,924.49 | 2,134.35 | **8,055.25** |
| 224 | 2,278.87 | 2,072.49 | 2,132.03 | 2,065.55 | **8,548.94** |
| 225 | 1,172.39 | 1,029.02 | 1,067.75 | 1,203.03 | **4,472.18** |
| 226 | 1,645.27 | 1,481.67 | 1,568.39 | 1,583.42 | **6,278.74** |

| | | | | | |
|---|---|---|---|---|---|
| 227 | 512.20 | 533.01 | 525.49 | 496.01 | **2,066.71** |
| 228 | 1,115.15 | 843.45 | 939.41 | 1,024.97 | **3,922.99** |
| 229 | 2,612.43 | 2,150.53 | 2,314.13 | 2,575.44 | **9,652.54** |
| 230 | 1,778.81 | 1,502.48 | 1,685.16 | 1,694.41 | **6,660.87** |
| 231 | 4,894.77 | 4,291.81 | 4,688.97 | 4,786.67 | **18,662.22** |
| 232 | 5,771.17 | 5,065.31 | 5,419.69 | 5,422.00 | **21,678.17** |
| 233 | 2,981.84 | 2,755.22 | 2,806.10 | 2,900.33 | **11,443.49** |
| 234 | 7,109.47 | 6,411.71 | 6,291.46 | 6,502.47 | **26,315.11** |
| 235 | 1,810.03 | 1,524.45 | 1,631.40 | 1,592.09 | **6,557.97** |
| 236 | 3,224.06 | 2,940.22 | 2,873.16 | 2,972.59 | **12,010.03** |
| 237 | 1,090.30 | 1,008.78 | 948.66 | 894.90 | **3,942.64** |
| 238 | 2,255.75 | 1,907.73 | 1,880.56 | 1,980.57 | **8,024.61** |
| 239 | 538.79 | 406.40 | 486.18 | 459.01 | **1,890.39** |
| 240 | 3,402.12 | 2,597.98 | 2,865.06 | 3,133.88 | **11,999.04** |
| 241 | 1,851.08 | 1,564.34 | 1,578.21 | 1,729.10 | **6,722.72** |
| 242 | 862.53 | 774.08 | 724.36 | 829.00 | **3,189.96** |
| 243 | 966.01 | 770.61 | 881.02 | 826.10 | **3,443.74** |
| 244 | 1,315.76 | 1,254.48 | 1,198.40 | 1,203.60 | **4,972.24** |
| 245 | 3,048.90 | 2,611.86 | 2,868.53 | 2,739.62 | **11,268.90** |
| 246 | 506.99 | 384.44 | 423.75 | 437.04 | **1,752.22** |
| **VA - VIRGINIA - Total** | **72,687.98** | **62,749.29** | **66,174.53** | **67,200.66** | **268,812.45** |

**STATE: VI - VIRGIN ISLANDS**

| REGISTRANT ZIP CODE 3 | 1ST QUARTER | 2ND QUARTER | 3RD QUARTER | 4TH QUARTER | TOTAL GRAMS |
|---|---|---|---|---|---|
| 008 | 64.75 | 98.28 | 75.15 | 68.22 | **306.39** |
| **VI - VIRGIN ISLANDS - Total** | **64.75** | **98.28** | **75.15** | **68.22** | **306.39** |

**STATE: VT - VERMONT**

| REGISTRANT ZIP CODE 3 | 1ST QUARTER | 2ND QUARTER | 3RD QUARTER | 4TH QUARTER | TOTAL GRAMS |
|---|---|---|---|---|---|
| 050 | 331.83 | 360.73 | 298.30 | 301.19 | **1,292.05** |
| 051 | 330.10 | 318.53 | 348.02 | 348.59 | **1,345.24** |
| 052 | 728.98 | 731.87 | 712.80 | 723.20 | **2,896.86** |
| 053 | 692.56 | 620.30 | 611.05 | 650.36 | **2,574.28** |
| 054 | 3,190.53 | 2,796.27 | 3,004.96 | 3,148.91 | **12,140.68** |
| 056 | 747.48 | 746.91 | 700.08 | 762.51 | **2,956.98** |
| 057 | 880.45 | 926.69 | 908.77 | 982.19 | **3,698.11** |
| 058 | 953.87 | 975.83 | 1,044.63 | 1,078.16 | **4,052.48** |
| **VT - VERMONT - Total** | **7,855.80** | **7,477.15** | **7,628.61** | **7,995.12** | **30,956.68** |

**STATE: WA - WASHINGTON**

| REGISTRANT ZIP CODE 3 | 1ST QUARTER | 2ND QUARTER | 3RD QUARTER | 4TH QUARTER | TOTAL GRAMS |
|---|---|---|---|---|---|
| 980 | 10,362.44 | 9,762.37 | 10,186.12 | 10,638.77 | **40,949.71** |
| 981 | 6,497.84 | 5,930.15 | 6,286.84 | 6,597.28 | **25,312.11** |
| 982 | 3,683.08 | 3,656.48 | 3,581.33 | 3,732.79 | **14,653.68** |
| 983 | 3,960.56 | 3,583.06 | 3,956.52 | 3,679.61 | **15,179.75** |
| 984 | 2,277.14 | 1,929.12 | 2,004.85 | 2,006.59 | **8,217.69** |
| 985 | 2,174.23 | 1,985.77 | 2,167.88 | 2,230.89 | **8,558.77** |
| 986 | 2,893.39 | 2,622.84 | 2,965.07 | 3,030.40 | **11,511.71** |
| 988 | 390.80 | 369.98 | 446.29 | 420.28 | **1,627.35** |
| 989 | 820.90 | 702.39 | 798.93 | 790.84 | **3,113.07** |
| 990 | 517.98 | 472.89 | 450.92 | 457.86 | **1,899.64** |
| 991 | 441.67 | 369.41 | 405.25 | 394.84 | **1,611.16** |
| 992 | 2,750.02 | 2,441.89 | 2,582.95 | 2,627.46 | **10,402.33** |
| 993 | 2,009.48 | 1,727.94 | 1,881.72 | 1,992.13 | **7,611.26** |
| 994 | 110.42 | 97.70 | 130.65 | 145.10 | **483.87** |
| **WA - WASHINGTON - Total** | **38,889.94** | **35,652.01** | **37,845.32** | **38,744.84** | **151,132.10** |

**STATE: WI - WISCONSIN**

| REGISTRANT ZIP CODE 3 | 1ST QUARTER | 2ND QUARTER | 3RD QUARTER | 4TH QUARTER | TOTAL GRAMS |
|---|---|---|---|---|---|
| 530 | 9,070.39 | 8,446.62 | 8,950.14 | 9,594.15 | **36,061.30** |
| 531 | 9,932.91 | 9,597.04 | 9,736.94 | 10,389.61 | **39,656.50** |
| 532 | 10,541.08 | 9,104.50 | 9,445.58 | 10,033.50 | **39,124.65** |
| 534 | 2,611.86 | 2,544.80 | 2,643.65 | 2,761.01 | **10,561.31** |
| 535 | 6,170.06 | 5,866.56 | 6,132.48 | 6,503.63 | **24,672.73** |

| | | | | | |
|---|---|---|---|---|---|
| 537 | 3,806.47 | 3,335.38 | 3,594.05 | 3,865.64 | **14,601.53** |
| 538 | 976.41 | 871.77 | 934.79 | 1,003.58 | **3,786.56** |
| 539 | 2,580.06 | 2,415.30 | 2,560.40 | 2,703.20 | **10,258.96** |
| 540 | 912.24 | 859.63 | 938.83 | 1,066.59 | **3,777.31** |
| 541 | 4,209.15 | 4,025.89 | 4,091.79 | 4,447.32 | **16,774.15** |
| 542 | 1,845.30 | 1,760.89 | 1,829.69 | 2,045.32 | **7,481.19** |
| 543 | 5,894.89 | 5,569.42 | 5,460.15 | 5,983.34 | **22,907.79** |
| 544 | 5,711.63 | 5,260.71 | 5,376.91 | 5,596.59 | **21,945.83** |
| 545 | 1,379.92 | 1,286.27 | 1,479.36 | 1,404.20 | **5,549.76** |
| 546 | 2,226.26 | 1,965.54 | 2,096.77 | 2,243.61 | **8,532.18** |
| 547 | 1,790.95 | 1,505.37 | 1,678.80 | 1,771.30 | **6,746.43** |
| 548 | 1,790.38 | 1,863.22 | 1,874.78 | 1,994.45 | **7,522.82** |
| 549 | 8,563.97 | 8,011.89 | 8,468.59 | 8,750.12 | **33,794.57** |
| **WI - WISCONSIN - Total** | **80,013.92** | **74,290.79** | **77,293.70** | **82,157.14** | **313,755.57** |

**STATE: WV - WEST VIRGINIA**

| REGISTRANT ZIP CODE 3 | 1ST QUARTER | 2ND QUARTER | 3RD QUARTER | 4TH QUARTER | TOTAL GRAMS |
|---|---|---|---|---|---|
| 247 | 404.09 | 389.06 | 369.41 | 406.40 | **1,568.96** |
| 248 | 252.05 | 261.88 | 240.49 | 242.80 | **997.22** |
| 249 | 380.97 | 317.38 | 313.33 | 331.83 | **1,343.50** |
| 250 | 685.63 | 570.01 | 589.66 | 667.13 | **2,512.42** |
| 251 | 567.69 | 464.79 | 461.90 | 564.80 | **2,059.19** |
| 252 | 420.28 | 327.20 | 337.03 | 356.69 | **1,441.20** |
| 253 | 1,769.56 | 1,491.50 | 1,536.59 | 1,632.55 | **6,430.21** |
| 254 | 1,412.88 | 1,288.58 | 1,375.88 | 1,340.04 | **5,417.38** |
| 255 | 1,763.21 | 1,375.88 | 1,519.25 | 1,629.66 | **6,287.99** |
| 256 | 241.65 | 220.26 | 216.21 | 220.26 | **898.37** |
| 257 | 1,631.98 | 1,493.81 | 1,445.83 | 1,493.23 | **6,064.85** |
| 258 | 1,027.28 | 903.57 | 909.35 | 968.90 | **3,809.10** |
| 259 | 274.02 | 234.13 | 250.32 | 254.36 | **1,012.83** |
| 260 | 1,101.28 | 936.52 | 966.58 | 970.05 | **3,974.44** |
| 261 | 1,704.24 | 1,460.86 | 1,475.89 | 1,472.42 | **6,113.41** |
| 262 | 655.57 | 576.37 | 528.96 | 600.07 | **2,360.96** |
| 263 | 1,121.51 | 1,089.72 | 1,087.41 | 1,115.73 | **4,414.37** |
| 264 | 335.88 | 279.80 | 321.42 | 283.85 | **1,220.95** |
| 265 | 2,216.44 | 1,937.79 | 2,019.88 | 2,133.19 | **8,307.30** |
| 266 | 186.73 | 167.07 | 171.12 | 186.15 | **711.06** |
| 267 | 652.67 | 608.16 | 629.55 | 610.47 | **2,500.86** |
| 268 | 376.34 | 322.58 | 328.94 | 332.41 | **1,360.27** |
| **WV - WEST VIRGINIA - Total** | **19,181.94** | **16,716.92** | **17,095.00** | **17,813.00** | **70,806.84** |

**STATE: WY - WYOMING**

| REGISTRANT ZIP CODE 3 | 1ST QUARTER | 2ND QUARTER | 3RD QUARTER | 4TH QUARTER | TOTAL GRAMS |
|---|---|---|---|---|---|
| 820 | 640.53 | 570.58 | 652.10 | 639.96 | **2,503.17** |
| 822 | 85.56 | 42.20 | 63.01 | 53.19 | **243.96** |
| 823 | 27.75 | 13.87 | 26.59 | 12.14 | **80.36** |
| 824 | 526.07 | 476.35 | 537.05 | 528.38 | **2,067.86** |
| 825 | 104.06 | 90.76 | 100.59 | 113.31 | **408.72** |
| 826 | 405.25 | 389.64 | 425.48 | 423.75 | **1,644.12** |
| 827 | 295.99 | 287.89 | 317.96 | 373.45 | **1,275.29** |
| 828 | 210.43 | 190.19 | 174.01 | 202.91 | **777.54** |
| 829 | 397.73 | 324.89 | 397.73 | 371.72 | **1,492.08** |
| 830 | 104.64 | 134.70 | 160.71 | 164.76 | **564.80** |
| 831 | 31.80 | 42.78 | 65.90 | 58.97 | **199.44** |
| **WY - WYOMING - Total** | **2,829.80** | **2,563.87** | **2,921.14** | **2,942.53** | **11,257.34** |

**DRUG: 1724 - METHYLPHENIDATE (DL;D;L;ISOMERS)**

**STATE: AK - ALASKA**

| REGISTRANT ZIP CODE 3 | 1ST QUARTER | 2ND QUARTER | 3RD QUARTER | 4TH QUARTER | TOTAL GRAMS |
|---|---|---|---|---|---|
| 995 | 4,323.37 | 3,592.81 | 3,826.20 | 3,938.19 | **15,680.58** |
| 996 | 1,569.74 | 1,446.35 | 1,496.32 | 1,603.72 | **6,116.13** |
| 997 | 1,373.40 | 1,161.30 | 1,308.65 | 1,229.29 | **5,072.64** |
| 998 | 586.98 | 560.17 | 544.65 | 526.23 | **2,218.04** |

| | | | | | |
|---|---|---|---|---|---|
| 999 | 360.26 | 332.90 | 317.18 | 422.45 | **1,432.80** |
| **AK - ALASKA - Total** | **8,213.75** | **7,093.54** | **7,493.00** | **7,719.89** | **30,520.19** |

**STATE: AL - ALABAMA**

| REGISTRANT ZIP CODE 3 | 1ST QUARTER | 2ND QUARTER | 3RD QUARTER | 4TH QUARTER | TOTAL GRAMS |
|---|---|---|---|---|---|
| 350 | 9,336.24 | 7,493.18 | 7,815.33 | 8,762.63 | **33,407.39** |
| 351 | 5,157.22 | 3,746.53 | 4,060.71 | 4,534.89 | **17,499.36** |
| 352 | 9,296.25 | 7,147.56 | 7,729.19 | 8,616.54 | **32,789.54** |
| 354 | 2,789.66 | 1,948.77 | 2,323.35 | 2,532.50 | **9,594.29** |
| 355 | 2,444.09 | 2,174.09 | 2,271.38 | 2,542.82 | **9,432.38** |
| 356 | 4,360.00 | 3,404.62 | 3,954.70 | 4,134.40 | **15,853.72** |
| 357 | 3,010.93 | 2,600.90 | 2,732.50 | 3,035.95 | **11,380.28** |
| 358 | 2,949.73 | 2,516.96 | 2,809.31 | 2,988.47 | **11,264.47** |
| 359 | 4,195.54 | 3,295.79 | 3,518.01 | 3,904.55 | **14,913.89** |
| 360 | 3,942.33 | 2,876.88 | 3,064.49 | 3,470.22 | **13,353.91** |
| 361 | 3,091.80 | 2,140.17 | 2,295.69 | 2,432.60 | **9,960.27** |
| 362 | 2,343.60 | 1,822.31 | 2,000.53 | 2,220.16 | **8,386.60** |
| 363 | 5,900.33 | 4,541.26 | 5,158.10 | 5,201.30 | **20,800.98** |
| 364 | 1,689.77 | 1,219.97 | 1,396.78 | 1,501.99 | **5,808.51** |
| 365 | 7,482.49 | 5,982.11 | 6,273.34 | 6,754.18 | **26,492.11** |
| 366 | 5,043.56 | 3,882.94 | 3,827.27 | 4,483.54 | **17,237.32** |
| 367 | 601.94 | 504.25 | 539.06 | 512.21 | **2,157.46** |
| 368 | 3,948.28 | 2,895.26 | 3,462.65 | 3,715.05 | **14,021.25** |
| 369 | 128.89 | 113.59 | 104.03 | 141.04 | **487.54** |
| **AL - ALABAMA - Total** | **77,712.65** | **60,307.15** | **65,336.42** | **71,485.05** | **274,841.27** |

**STATE: AR - ARKANSAS**

| REGISTRANT ZIP CODE 3 | 1ST QUARTER | 2ND QUARTER | 3RD QUARTER | 4TH QUARTER | TOTAL GRAMS |
|---|---|---|---|---|---|
| 716 | 1,497.58 | 1,117.05 | 1,139.24 | 1,261.62 | **5,015.49** |
| 717 | 847.09 | 624.53 | 689.87 | 730.64 | **2,892.13** |
| 718 | 926.24 | 654.17 | 778.74 | 788.02 | **3,147.17** |
| 719 | 2,323.06 | 1,879.16 | 1,928.39 | 2,155.36 | **8,285.97** |
| 720 | 6,600.36 | 5,130.21 | 5,779.33 | 6,286.57 | **23,796.46** |
| 721 | 4,442.52 | 3,667.13 | 3,985.43 | 4,451.92 | **16,547.00** |
| 722 | 4,110.53 | 3,279.51 | 3,520.54 | 3,913.75 | **14,824.33** |
| 723 | 1,239.72 | 916.29 | 1,049.91 | 1,141.81 | **4,347.73** |
| 724 | 3,425.83 | 2,583.85 | 2,821.55 | 2,983.51 | **11,814.74** |
| 725 | 1,120.67 | 939.49 | 949.92 | 1,091.14 | **4,101.22** |
| 726 | 1,109.80 | 923.52 | 987.02 | 1,141.23 | **4,161.57** |
| 727 | 7,464.41 | 6,092.01 | 6,669.82 | 7,197.74 | **27,423.98** |
| 728 | 1,362.20 | 1,052.22 | 1,161.72 | 1,240.22 | **4,816.36** |
| 729 | 2,901.32 | 2,391.15 | 2,515.46 | 2,679.56 | **10,487.48** |
| **AR - ARKANSAS - Total** | **39,371.34** | **31,250.31** | **33,976.92** | **37,063.06** | **141,661.62** |

**STATE: AS - AMERICAN SAMOA**

| REGISTRANT ZIP CODE 3 | 1ST QUARTER | 2ND QUARTER | 3RD QUARTER | 4TH QUARTER | TOTAL GRAMS |
|---|---|---|---|---|---|
| 967 | 1.73 | 4.32 | 6.92 | 2.59 | **15.57** |
| **AS - AMERICAN SAMOA - Total** | **1.73** | **4.32** | **6.92** | **2.59** | **15.57** |

**STATE: AZ - ARIZONA**

| REGISTRANT ZIP CODE 3 | 1ST QUARTER | 2ND QUARTER | 3RD QUARTER | 4TH QUARTER | TOTAL GRAMS |
|---|---|---|---|---|---|
| 850 | 12,237.79 | 10,176.58 | 10,604.27 | 11,131.05 | **44,149.68** |
| 851 | 4,520.08 | 3,780.82 | 3,986.92 | 4,273.10 | **16,560.93** |
| 852 | 29,140.54 | 26,152.38 | 26,787.25 | 29,496.56 | **111,576.73** |
| 853 | 13,082.47 | 10,348.18 | 11,004.29 | 11,895.31 | **46,330.25** |
| 855 | 738.28 | 518.21 | 615.61 | 601.38 | **2,473.47** |
| 856 | 2,795.84 | 2,233.13 | 2,274.90 | 2,356.93 | **9,660.79** |
| 857 | 12,091.27 | 9,923.50 | 10,442.97 | 10,830.54 | **43,288.29** |
| 859 | 653.40 | 599.25 | 646.06 | 628.04 | **2,526.75** |
| 860 | 1,462.07 | 1,154.95 | 1,270.15 | 1,399.48 | **5,286.65** |
| 863 | 2,153.38 | 2,005.04 | 2,089.95 | 2,265.12 | **8,513.50** |
| 864 | 1,116.10 | 1,009.11 | 1,007.53 | 1,043.23 | **4,175.96** |
| 865 | 474.20 | 208.53 | 257.42 | 181.31 | **1,121.46** |
| **AZ - ARIZONA - Total** | **80,465.43** | **68,109.67** | **70,987.30** | **76,102.07** | **295,664.47** |

**STATE: CA - CALIFORNIA**

| REGISTRANT ZIP CODE 3 | 1ST QUARTER | 2ND QUARTER | 3RD QUARTER | 4TH QUARTER | **TOTAL GRAMS** |
|---|---|---|---|---|---|
| 900 | 10,524.96 | 8,489.65 | 9,099.99 | 9,039.34 | **37,153.95** |
| 902 | 7,723.10 | 6,612.35 | 6,566.91 | 6,864.12 | **27,766.48** |
| 903 | 333.12 | 254.61 | 299.24 | 245.30 | **1,132.28** |
| 904 | 1,544.99 | 1,322.10 | 1,340.86 | 1,485.28 | **5,693.22** |
| 905 | 1,257.24 | 1,120.89 | 1,149.46 | 1,247.85 | **4,775.44** |
| 906 | 3,359.13 | 2,232.36 | 2,572.39 | 2,967.39 | **11,131.27** |
| 907 | 3,282.18 | 2,797.56 | 2,772.19 | 2,978.86 | **11,830.80** |
| 908 | 2,143.07 | 1,896.33 | 2,012.98 | 2,154.65 | **8,207.04** |
| 910 | 1,391.73 | 1,165.52 | 1,213.11 | 1,394.33 | **5,164.69** |
| 911 | 2,492.99 | 2,243.63 | 2,279.02 | 2,487.98 | **9,503.63** |
| 912 | 995.72 | 916.16 | 874.42 | 937.61 | **3,723.91** |
| 913 | 11,637.16 | 10,294.74 | 10,118.78 | 10,476.10 | **42,526.78** |
| 914 | 2,779.19 | 2,083.04 | 2,244.48 | 2,350.87 | **9,457.58** |
| 915 | 728.70 | 657.78 | 635.85 | 773.95 | **2,796.29** |
| 916 | 1,017.85 | 779.78 | 908.67 | 872.62 | **3,578.91** |
| 917 | 8,360.44 | 6,876.59 | 6,842.50 | 7,391.10 | **29,470.64** |
| 918 | 326.70 | 301.20 | 266.90 | 317.87 | **1,212.67** |
| 919 | 2,793.06 | 2,798.80 | 2,519.12 | 2,822.18 | **10,933.17** |
| 920 | 26,445.39 | 25,341.29 | 24,980.23 | 26,218.40 | **102,985.31** |
| 921 | 11,800.09 | 9,517.63 | 10,051.31 | 10,419.63 | **41,788.66** |
| 922 | 4,237.35 | 3,512.03 | 3,356.88 | 3,606.71 | **14,712.96** |
| 923 | 7,132.55 | 6,203.05 | 6,029.11 | 6,698.69 | **26,063.39** |
| 924 | 739.88 | 628.83 | 714.19 | 587.34 | **2,670.24** |
| 925 | 9,066.61 | 7,609.29 | 7,896.97 | 8,456.13 | **33,028.99** |
| 926 | 12,299.52 | 11,011.46 | 11,316.73 | 11,864.69 | **46,492.42** |
| 927 | 2,828.14 | 2,492.31 | 2,287.54 | 2,517.36 | **10,125.35** |
| 928 | 8,118.17 | 6,979.18 | 7,175.80 | 7,412.09 | **29,685.24** |
| 930 | 5,447.09 | 4,681.94 | 4,571.76 | 4,768.34 | **19,469.13** |
| 931 | 1,561.88 | 1,313.04 | 1,458.52 | 1,567.48 | **5,900.92** |
| 932 | 3,309.40 | 2,601.69 | 2,686.09 | 2,698.04 | **11,295.21** |
| 933 | 2,911.82 | 2,409.10 | 2,440.73 | 2,375.78 | **10,137.43** |
| 934 | 3,168.93 | 2,754.49 | 2,884.46 | 3,027.15 | **11,835.03** |
| 935 | 3,084.59 | 2,706.16 | 2,800.38 | 2,948.63 | **11,539.77** |
| 936 | 3,688.78 | 3,308.10 | 3,231.85 | 3,456.00 | **13,684.73** |
| 937 | 5,042.20 | 4,005.42 | 4,146.07 | 4,476.24 | **17,669.94** |
| 939 | 2,363.33 | 1,635.29 | 1,812.80 | 1,720.74 | **7,532.17** |
| 940 | 5,814.27 | 5,115.69 | 5,127.60 | 5,647.94 | **21,705.49** |
| 941 | 6,367.59 | 5,549.71 | 5,509.46 | 5,806.87 | **23,233.64** |
| 943 | 1,199.92 | 917.29 | 921.94 | 1,011.98 | **4,051.13** |
| 944 | 1,127.33 | 889.38 | 1,095.21 | 1,069.92 | **4,181.84** |
| 945 | 17,761.60 | 15,151.40 | 15,351.00 | 16,205.11 | **64,469.11** |
| 946 | 3,239.44 | 2,838.69 | 2,701.40 | 2,935.30 | **11,714.82** |
| 947 | 907.85 | 846.20 | 825.07 | 932.03 | **3,511.16** |
| 948 | 678.97 | 522.65 | 479.05 | 586.10 | **2,266.77** |
| 949 | 4,111.87 | 3,394.14 | 3,699.24 | 3,997.12 | **15,202.37** |
| 950 | 6,517.09 | 5,690.78 | 5,417.14 | 6,018.02 | **23,643.03** |
| 951 | 3,993.62 | 3,373.89 | 3,320.70 | 3,742.58 | **14,430.79** |
| 952 | 2,670.50 | 2,216.79 | 2,203.12 | 2,411.67 | **9,502.09** |
| 953 | 5,408.01 | 4,648.71 | 4,563.02 | 4,944.08 | **19,563.82** |
| 954 | 3,401.05 | 3,037.00 | 3,153.93 | 3,228.90 | **12,820.88** |
| 955 | 1,121.37 | 958.15 | 1,027.63 | 990.96 | **4,098.10** |
| 956 | 13,182.60 | 11,044.95 | 11,271.54 | 12,067.63 | **47,566.72** |
| 957 | 3,428.31 | 2,734.39 | 2,903.66 | 3,070.31 | **12,136.68** |
| 958 | 5,934.94 | 4,812.73 | 4,845.54 | 5,407.19 | **21,000.40** |
| 959 | 3,580.05 | 2,914.08 | 2,928.84 | 3,120.59 | **12,543.56** |
| 960 | 2,598.01 | 2,168.21 | 2,420.46 | 2,314.78 | **9,501.46** |
| 961 | 488.70 | 460.12 | 513.89 | 496.94 | **1,959.65** |
| **CA - CALIFORNIA - Total** | **269,470.13** | **230,838.34** | **233,837.75** | **247,632.87** | **981,779.10** |

**STATE: CO - COLORADO**

| REGISTRANT ZIP CODE 3 | 1ST QUARTER | 2ND QUARTER | 3RD QUARTER | 4TH QUARTER | TOTAL GRAMS |
|---|---|---|---|---|---|
| 800 | 11,430.47 | 9,828.10 | 10,071.95 | 11,023.86 | **42,354.38** |
| 801 | 12,429.87 | 10,522.48 | 11,326.77 | 12,349.18 | **46,628.30** |
| 802 | 11,540.05 | 10,786.25 | 10,758.25 | 11,674.02 | **44,758.58** |
| 803 | 2,294.77 | 1,608.57 | 1,938.36 | 2,262.78 | **8,104.49** |
| 804 | 1,896.29 | 1,700.65 | 1,756.11 | 2,050.72 | **7,403.77** |
| 805 | 7,527.86 | 6,280.25 | 7,046.62 | 7,743.29 | **28,598.01** |
| 806 | 2,392.12 | 1,894.68 | 2,263.44 | 2,330.79 | **8,881.03** |
| 807 | 489.66 | 412.96 | 415.11 | 442.30 | **1,760.03** |
| 808 | 1,521.09 | 1,111.04 | 1,604.92 | 1,387.59 | **5,624.64** |
| 809 | 8,058.67 | 6,716.45 | 7,368.86 | 7,029.39 | **29,173.37** |
| 810 | 1,775.10 | 1,715.52 | 1,795.67 | 1,797.98 | **7,084.28** |
| 811 | 268.55 | 276.70 | 284.33 | 286.53 | **1,116.11** |
| 812 | 550.16 | 493.64 | 619.13 | 635.92 | **2,298.85** |
| 813 | 559.76 | 532.79 | 581.51 | 545.43 | **2,219.49** |
| 814 | 698.13 | 663.05 | 658.95 | 685.32 | **2,705.46** |
| 815 | 1,876.33 | 1,776.86 | 1,774.25 | 1,780.08 | **7,207.53** |
| 816 | 1,411.52 | 1,128.31 | 1,322.19 | 1,276.51 | **5,138.54** |
| **CO - COLORADO - Total** | **66,720.41** | **57,448.30** | **61,586.42** | **65,301.70** | **251,056.84** |

**STATE: CT - CONNECTICUT**

| REGISTRANT ZIP CODE 3 | 1ST QUARTER | 2ND QUARTER | 3RD QUARTER | 4TH QUARTER | TOTAL GRAMS |
|---|---|---|---|---|---|
| 060 | 12,037.44 | 10,510.15 | 10,532.57 | 11,292.16 | **44,372.31** |
| 061 | 4,374.27 | 3,490.96 | 3,632.07 | 3,864.19 | **15,361.48** |
| 062 | 2,769.75 | 2,308.39 | 2,391.08 | 2,482.79 | **9,952.02** |
| 063 | 4,911.33 | 4,427.93 | 4,302.35 | 4,648.10 | **18,289.72** |
| 064 | 12,555.47 | 10,837.46 | 11,046.73 | 11,821.67 | **46,261.33** |
| 065 | 3,578.45 | 3,061.61 | 2,894.16 | 3,244.79 | **12,779.01** |
| 066 | 1,645.70 | 1,244.06 | 1,260.66 | 1,460.35 | **5,610.78** |
| 067 | 4,696.18 | 3,946.06 | 4,065.43 | 4,262.30 | **16,969.97** |
| 068 | 8,799.98 | 7,973.15 | 7,879.39 | 8,696.97 | **33,349.50** |
| 069 | 1,119.42 | 1,067.65 | 1,093.95 | 1,147.42 | **4,428.44** |
| **CT - CONNECTICUT - Total** | **56,488.00** | **48,867.42** | **49,098.39** | **52,920.75** | **207,374.56** |

**STATE: DC - DISTRICT OF COLUMBIA**

| REGISTRANT ZIP CODE 3 | 1ST QUARTER | 2ND QUARTER | 3RD QUARTER | 4TH QUARTER | TOTAL GRAMS |
|---|---|---|---|---|---|
| 200 | 8,454.42 | 6,133.08 | 6,159.38 | 6,704.70 | **27,451.59** |
| 203 | 276.94 | 43.60 | 93.55 | 63.00 | **477.10** |
| 204 | 447.27 | 588.24 | 347.13 | 425.40 | **1,808.04** |
| **DC - DISTRICT OF COLUMBIA - Total** | **9,178.64** | **6,764.92** | **6,600.07** | **7,193.09** | **29,736.72** |

**STATE: DE - DELAWARE**

| REGISTRANT ZIP CODE 3 | 1ST QUARTER | 2ND QUARTER | 3RD QUARTER | 4TH QUARTER | TOTAL GRAMS |
|---|---|---|---|---|---|
| 197 | 7,129.11 | 5,700.36 | 5,650.09 | 6,455.77 | **24,935.33** |
| 198 | 5,079.34 | 4,042.27 | 3,932.63 | 4,654.79 | **17,709.04** |
| 199 | 6,529.07 | 5,438.46 | 5,271.50 | 5,737.43 | **22,976.45** |
| **DE - DELAWARE - Total** | **18,737.51** | **15,181.09** | **14,854.22** | **16,847.98** | **65,620.81** |

**STATE: FL - FLORIDA**

| REGISTRANT ZIP CODE 3 | 1ST QUARTER | 2ND QUARTER | 3RD QUARTER | 4TH QUARTER | TOTAL GRAMS |
|---|---|---|---|---|---|
| 320 | 9,179.60 | 7,384.82 | 7,561.37 | 8,459.58 | **32,585.37** |
| 321 | 4,896.64 | 4,229.41 | 4,262.23 | 4,666.70 | **18,054.98** |
| 322 | 12,660.17 | 10,052.38 | 11,016.11 | 11,162.01 | **44,890.67** |
| 323 | 5,147.47 | 3,977.58 | 4,398.56 | 4,822.72 | **18,346.33** |
| 324 | 5,291.18 | 4,604.70 | 4,631.65 | 5,220.20 | **19,747.73** |
| 325 | 13,315.02 | 10,508.80 | 10,734.62 | 11,428.81 | **45,987.25** |
| 326 | 6,658.17 | 5,177.66 | 5,511.48 | 6,055.04 | **23,402.35** |
| 327 | 12,700.61 | 10,662.61 | 10,932.55 | 11,953.02 | **46,248.78** |
| 328 | 8,027.42 | 6,613.68 | 6,743.32 | 7,394.67 | **28,779.09** |
| 329 | 9,639.91 | 7,911.76 | 8,108.92 | 9,010.59 | **34,671.18** |
| 330 | 9,553.41 | 7,461.29 | 7,386.64 | 8,502.64 | **32,903.98** |
| 331 | 11,658.89 | 8,624.24 | 8,821.22 | 10,194.47 | **39,298.83** |
| 333 | 4,804.35 | 4,010.22 | 3,930.14 | 4,122.03 | **16,866.75** |

| | | | | | |
|---|---|---|---|---|---|
| 334 | 13,619.65 | 11,795.35 | 11,165.77 | 12,470.54 | **49,051.31** |
| 335 | 11,027.91 | 8,805.88 | 9,300.32 | 10,559.09 | **39,693.19** |
| 336 | 9,711.28 | 8,078.61 | 8,487.05 | 8,740.97 | **35,017.91** |
| 337 | 7,214.25 | 6,108.96 | 5,985.56 | 6,816.45 | **26,125.22** |
| 338 | 8,465.84 | 7,064.64 | 7,508.37 | 8,559.55 | **31,598.40** |
| 339 | 9,785.56 | 7,854.72 | 7,711.52 | 9,199.06 | **34,550.85** |
| 341 | 3,586.21 | 3,291.83 | 2,991.65 | 3,665.93 | **13,535.63** |
| 342 | 10,480.12 | 8,660.45 | 9,108.01 | 9,906.74 | **38,155.32** |
| 344 | 5,459.26 | 4,254.82 | 4,557.56 | 5,123.80 | **19,395.44** |
| 346 | 8,276.47 | 6,888.10 | 6,890.87 | 7,676.06 | **29,731.49** |
| 347 | 7,453.38 | 5,934.89 | 6,470.42 | 7,291.53 | **27,150.22** |
| 349 | 4,780.16 | 4,003.34 | 4,024.17 | 4,381.82 | **17,189.49** |
| **FL - FLORIDA - Total** | **213,392.92** | **173,960.75** | **178,240.06** | **197,384.04** | **762,977.76** |

**STATE: GA - GEORGIA**

| REGISTRANT ZIP CODE 3 | 1ST QUARTER | 2ND QUARTER | 3RD QUARTER | 4TH QUARTER | TOTAL GRAMS |
|---|---|---|---|---|---|
| 300 | 32,820.35 | 25,283.59 | 27,123.14 | 29,967.60 | **115,194.67** |
| 301 | 20,211.17 | 15,645.49 | 17,384.03 | 18,631.00 | **71,871.68** |
| 302 | 13,565.05 | 10,053.24 | 10,942.04 | 12,296.79 | **46,857.11** |
| 303 | 13,518.88 | 10,573.21 | 10,477.73 | 11,990.76 | **46,560.58** |
| 304 | 4,228.95 | 3,110.83 | 3,350.26 | 3,681.59 | **14,371.63** |
| 305 | 10,368.42 | 8,088.40 | 8,878.88 | 9,668.86 | **37,004.56** |
| 306 | 6,383.91 | 4,782.03 | 5,140.86 | 5,875.56 | **22,182.36** |
| 307 | 6,816.67 | 5,181.53 | 5,601.89 | 6,350.19 | **23,950.29** |
| 308 | 3,574.11 | 2,540.47 | 2,717.21 | 3,177.01 | **12,008.80** |
| 309 | 4,608.94 | 3,163.77 | 3,975.87 | 3,306.86 | **15,055.44** |
| 310 | 11,943.43 | 8,341.16 | 9,399.66 | 10,315.27 | **39,999.53** |
| 312 | 3,589.81 | 2,603.34 | 2,793.93 | 3,045.43 | **12,032.51** |
| 313 | 5,302.79 | 3,545.79 | 4,515.71 | 4,466.51 | **17,830.80** |
| 314 | 4,160.19 | 3,158.09 | 3,184.90 | 3,462.08 | **13,965.26** |
| 315 | 8,670.70 | 5,952.54 | 7,038.42 | 7,679.72 | **29,341.38** |
| 316 | 4,192.64 | 2,705.84 | 3,090.89 | 3,554.48 | **13,543.85** |
| 317 | 8,420.82 | 5,326.76 | 6,379.65 | 6,920.31 | **27,047.54** |
| 318 | 525.89 | 377.80 | 423.88 | 490.49 | **1,818.05** |
| 319 | 5,265.48 | 3,771.30 | 4,606.53 | 4,007.72 | **17,651.03** |
| 398 | 1,522.32 | 991.31 | 989.45 | 1,248.65 | **4,751.73** |
| **GA - GEORGIA - Total** | **169,690.52** | **125,196.48** | **138,014.94** | **150,136.86** | **583,038.80** |

**STATE: GU - GUAM**

| REGISTRANT ZIP CODE 3 | 1ST QUARTER | 2ND QUARTER | 3RD QUARTER | 4TH QUARTER | TOTAL GRAMS |
|---|---|---|---|---|---|
| 969 | 417.60 | 412.90 | 346.18 | 482.07 | **1,658.75** |
| **GU - GUAM - Total** | **417.60** | **412.90** | **346.18** | **482.07** | **1,658.75** |

**STATE: HI - HAWAII**

| REGISTRANT ZIP CODE 3 | 1ST QUARTER | 2ND QUARTER | 3RD QUARTER | 4TH QUARTER | TOTAL GRAMS |
|---|---|---|---|---|---|
| 967 | 5,502.01 | 4,499.52 | 4,738.42 | 5,300.53 | **20,040.48** |
| 968 | 5,365.19 | 4,065.52 | 4,449.16 | 4,242.81 | **18,122.68** |
| **HI - HAWAII - Total** | **10,867.20** | **8,565.04** | **9,187.59** | **9,543.33** | **38,163.16** |

**STATE: IA - IOWA**

| REGISTRANT ZIP CODE 3 | 1ST QUARTER | 2ND QUARTER | 3RD QUARTER | 4TH QUARTER | TOTAL GRAMS |
|---|---|---|---|---|---|
| 500 | 6,928.90 | 5,693.63 | 6,481.88 | 6,883.68 | **25,988.09** |
| 501 | 3,981.14 | 3,319.83 | 3,411.95 | 3,580.28 | **14,293.20** |
| 502 | 4,871.02 | 4,053.71 | 4,276.14 | 4,593.55 | **17,794.42** |
| 503 | 7,583.32 | 6,481.63 | 6,633.02 | 7,345.77 | **28,043.73** |
| 504 | 2,137.47 | 1,721.43 | 1,815.93 | 2,054.92 | **7,729.75** |
| 505 | 2,370.56 | 1,887.11 | 2,053.97 | 2,263.30 | **8,574.93** |
| 506 | 3,331.10 | 2,669.41 | 3,022.22 | 3,201.79 | **12,224.52** |
| 507 | 1,801.76 | 1,582.19 | 1,694.79 | 1,877.09 | **6,955.82** |
| 508 | 628.19 | 516.11 | 561.40 | 600.62 | **2,306.32** |
| 510 | 1,968.16 | 1,655.29 | 1,673.77 | 1,815.93 | **7,113.15** |
| 511 | 4,015.36 | 3,764.85 | 3,844.11 | 4,023.80 | **15,648.12** |
| 512 | 1,035.15 | 919.00 | 942.56 | 1,135.51 | **4,032.21** |
| 513 | 865.85 | 886.73 | 886.61 | 899.47 | **3,538.66** |

| | | | | | |
|---|---|---|---|---|---|
| 514 | 945.71 | 703.24 | 868.18 | 949.22 | **3,466.34** |
| 515 | 3,194.63 | 2,830.98 | 2,829.68 | 3,052.88 | **11,908.16** |
| 516 | 427.05 | 409.44 | 437.90 | 452.89 | **1,727.28** |
| 520 | 3,694.89 | 3,053.17 | 3,307.09 | 3,536.61 | **13,591.77** |
| 521 | 1,223.79 | 974.91 | 1,101.31 | 1,171.17 | **4,471.17** |
| 522 | 6,983.40 | 5,895.25 | 6,278.35 | 6,882.83 | **26,039.82** |
| 523 | 4,164.68 | 3,555.03 | 3,785.87 | 3,869.99 | **15,375.57** |
| 524 | 6,104.16 | 5,351.79 | 5,472.72 | 6,044.82 | **22,973.49** |
| 525 | 2,384.05 | 1,962.14 | 2,076.21 | 2,317.01 | **8,739.41** |
| 526 | 3,058.49 | 2,604.88 | 2,834.06 | 2,811.03 | **11,308.46** |
| 527 | 4,521.22 | 3,688.14 | 3,900.17 | 4,080.75 | **16,190.28** |
| 528 | 3,478.91 | 2,879.33 | 2,809.86 | 3,214.48 | **12,382.58** |
| **IA - IOWA - Total** | **81,698.92** | **69,059.22** | **72,999.72** | **78,659.37** | **302,417.24** |

**STATE: ID - IDAHO**

| REGISTRANT ZIP CODE 3 | 1ST QUARTER | 2ND QUARTER | 3RD QUARTER | 4TH QUARTER | TOTAL GRAMS |
|---|---|---|---|---|---|
| 832 | 2,898.37 | 2,421.13 | 2,685.96 | 2,892.67 | **10,898.12** |
| 833 | 2,795.00 | 2,497.48 | 2,417.60 | 2,672.30 | **10,382.38** |
| 834 | 3,900.43 | 3,563.82 | 3,729.85 | 4,015.95 | **15,210.05** |
| 835 | 795.11 | 738.79 | 737.17 | 836.80 | **3,107.87** |
| 836 | 9,609.05 | 8,478.01 | 8,670.24 | 9,490.39 | **36,247.69** |
| 837 | 6,898.87 | 5,944.25 | 5,914.74 | 6,523.03 | **25,280.90** |
| 838 | 3,129.54 | 2,804.14 | 2,929.77 | 3,210.30 | **12,073.74** |
| **ID - IDAHO - Total** | **30,026.38** | **26,447.61** | **27,085.32** | **29,641.44** | **113,200.75** |

**STATE: IL - ILLINOIS**

| REGISTRANT ZIP CODE 3 | 1ST QUARTER | 2ND QUARTER | 3RD QUARTER | 4TH QUARTER | TOTAL GRAMS |
|---|---|---|---|---|---|
| 600 | 27,166.36 | 23,006.25 | 24,107.20 | 26,668.02 | **100,947.82** |
| 601 | 18,387.94 | 15,328.70 | 15,882.34 | 17,530.75 | **67,129.73** |
| 602 | 1,643.15 | 1,293.10 | 1,503.46 | 1,632.45 | **6,072.16** |
| 603 | 1,398.98 | 1,208.80 | 1,172.84 | 1,477.34 | **5,257.97** |
| 604 | 13,097.99 | 10,863.76 | 11,013.67 | 12,222.26 | **47,197.68** |
| 605 | 15,494.17 | 12,641.31 | 13,220.34 | 14,450.23 | **55,806.06** |
| 606 | 17,568.37 | 13,915.08 | 13,933.50 | 15,862.15 | **61,279.10** |
| 607 | 1,289.35 | 1,047.48 | 1,121.94 | 1,281.03 | **4,739.80** |
| 608 | 595.98 | 548.98 | 529.81 | 628.61 | **2,303.38** |
| 609 | 2,280.97 | 1,703.04 | 1,822.74 | 1,997.21 | **7,803.96** |
| 610 | 3,718.23 | 3,117.80 | 3,173.21 | 3,344.60 | **13,353.83** |
| 611 | 3,793.54 | 2,912.42 | 3,100.74 | 3,497.91 | **13,304.61** |
| 612 | 2,800.13 | 2,230.63 | 2,508.81 | 2,544.61 | **10,084.17** |
| 613 | 2,109.93 | 1,749.46 | 1,764.36 | 1,879.69 | **7,503.43** |
| 614 | 2,155.46 | 1,711.54 | 1,806.25 | 2,028.01 | **7,701.26** |
| 615 | 3,679.72 | 3,208.58 | 3,325.84 | 3,739.93 | **13,954.07** |
| 616 | 4,666.54 | 3,730.47 | 4,084.81 | 4,266.36 | **16,748.17** |
| 617 | 5,609.61 | 4,654.03 | 4,959.32 | 5,153.65 | **20,376.62** |
| 618 | 5,758.85 | 4,645.43 | 4,976.30 | 5,395.02 | **20,775.60** |
| 619 | 2,193.17 | 1,655.93 | 1,780.60 | 1,957.13 | **7,586.82** |
| 620 | 5,462.40 | 4,224.14 | 4,490.83 | 4,807.62 | **18,984.99** |
| 622 | 7,619.98 | 6,180.93 | 6,598.28 | 6,860.15 | **27,259.34** |
| 623 | 2,156.97 | 1,727.50 | 1,826.83 | 1,937.63 | **7,648.94** |
| 624 | 2,112.88 | 1,682.76 | 1,864.79 | 1,902.52 | **7,562.95** |
| 625 | 2,526.10 | 2,010.52 | 2,261.54 | 2,384.48 | **9,182.63** |
| 626 | 3,649.91 | 3,028.37 | 3,249.42 | 3,494.14 | **13,421.83** |
| 627 | 4,159.60 | 3,109.90 | 3,508.17 | 3,627.64 | **14,405.30** |
| 628 | 3,386.08 | 2,617.55 | 2,937.22 | 3,126.39 | **12,067.24** |
| 629 | 3,813.59 | 3,063.87 | 3,382.78 | 3,558.69 | **13,818.93** |
| **IL - ILLINOIS - Total** | **170,295.93** | **138,818.33** | **145,907.92** | **159,256.21** | **614,278.40** |

**STATE: IN - INDIANA**

| REGISTRANT ZIP CODE 3 | 1ST QUARTER | 2ND QUARTER | 3RD QUARTER | 4TH QUARTER | TOTAL GRAMS |
|---|---|---|---|---|---|
| 460 | 15,661.15 | 12,177.88 | 13,687.64 | 14,647.79 | **56,174.46** |
| 461 | 11,887.59 | 9,130.84 | 10,452.81 | 11,214.71 | **42,685.94** |
| 462 | 15,669.07 | 11,968.89 | 13,150.30 | 13,287.85 | **54,076.11** |

| | | | | | |
|---|---|---|---|---|---|
| 463 | 9,793.73 | 8,192.86 | 8,634.14 | 9,664.68 | **36,285.41** |
| 464 | 856.31 | 665.29 | 665.83 | 691.37 | **2,878.79** |
| 465 | 8,888.88 | 7,268.35 | 7,662.98 | 8,465.73 | **32,285.94** |
| 466 | 2,654.29 | 2,317.64 | 2,171.41 | 2,472.59 | **9,615.93** |
| 467 | 4,708.71 | 3,862.03 | 4,358.43 | 4,642.17 | **17,571.34** |
| 468 | 6,989.99 | 5,910.63 | 6,230.29 | 6,869.63 | **26,000.55** |
| 469 | 5,382.57 | 4,318.95 | 4,727.41 | 5,217.27 | **19,646.20** |
| 470 | 1,731.19 | 1,275.13 | 1,552.15 | 1,688.60 | **6,247.07** |
| 471 | 4,992.04 | 4,239.00 | 4,583.04 | 4,899.80 | **18,713.87** |
| 472 | 3,910.19 | 3,091.39 | 3,384.04 | 3,693.51 | **14,079.13** |
| 473 | 5,386.92 | 4,202.89 | 4,686.00 | 5,224.88 | **19,500.69** |
| 474 | 4,423.73 | 3,081.60 | 3,452.13 | 3,948.84 | **14,906.31** |
| 475 | 3,210.79 | 2,597.91 | 2,779.07 | 3,035.77 | **11,623.55** |
| 476 | 3,009.98 | 2,220.60 | 2,621.75 | 2,844.10 | **10,696.42** |
| 477 | 6,037.86 | 4,627.62 | 4,875.92 | 5,431.84 | **20,973.24** |
| 478 | 3,380.83 | 2,560.18 | 2,784.40 | 3,032.28 | **11,757.69** |
| 479 | 6,006.89 | 4,272.80 | 5,009.26 | 5,869.52 | **21,158.48** |
| **IN - INDIANA - Total** | **124,582.71** | **97,982.47** | **107,469.02** | **116,842.93** | **446,877.14** |

**STATE: KS - KANSAS**

| REGISTRANT ZIP CODE 3 | 1ST QUARTER | 2ND QUARTER | 3RD QUARTER | 4TH QUARTER | TOTAL GRAMS |
|---|---|---|---|---|---|
| 660 | 10,575.97 | 8,868.20 | 9,094.25 | 10,442.05 | **38,980.47** |
| 661 | 1,195.02 | 1,185.23 | 1,014.41 | 1,104.91 | **4,499.57** |
| 662 | 9,670.43 | 7,892.38 | 8,159.67 | 9,509.92 | **35,232.40** |
| 664 | 1,555.16 | 1,297.59 | 1,387.07 | 1,290.15 | **5,529.97** |
| 665 | 1,818.22 | 1,483.06 | 1,678.08 | 1,863.45 | **6,842.81** |
| 666 | 5,242.30 | 4,911.06 | 5,084.57 | 5,500.83 | **20,738.75** |
| 667 | 2,017.30 | 1,776.61 | 1,795.89 | 1,952.87 | **7,542.67** |
| 668 | 859.62 | 771.48 | 780.04 | 901.56 | **3,312.70** |
| 669 | 299.42 | 230.57 | 259.81 | 269.85 | **1,059.66** |
| 670 | 2,984.60 | 2,598.82 | 2,601.54 | 2,803.08 | **10,988.05** |
| 671 | 1,305.98 | 1,125.88 | 1,174.22 | 1,347.26 | **4,953.35** |
| 672 | 6,437.90 | 5,595.01 | 5,395.72 | 6,146.11 | **23,574.74** |
| 673 | 1,534.77 | 1,277.18 | 1,350.02 | 1,518.49 | **5,680.46** |
| 674 | 2,094.93 | 1,854.34 | 1,923.94 | 2,060.59 | **7,933.80** |
| 675 | 1,832.50 | 1,438.89 | 1,564.94 | 1,634.50 | **6,470.84** |
| 676 | 1,014.16 | 883.85 | 889.85 | 1,044.01 | **3,831.86** |
| 677 | 391.80 | 317.21 | 383.89 | 387.48 | **1,480.38** |
| 678 | 833.46 | 712.79 | 710.05 | 763.41 | **3,019.71** |
| 679 | 147.53 | 139.84 | 128.51 | 180.74 | **596.63** |
| **KS - KANSAS - Total** | **51,811.08** | **44,359.97** | **45,376.48** | **50,721.27** | **192,268.80** |

**STATE: KY - KENTUCKY**

| REGISTRANT ZIP CODE 3 | 1ST QUARTER | 2ND QUARTER | 3RD QUARTER | 4TH QUARTER | TOTAL GRAMS |
|---|---|---|---|---|---|
| 400 | 4,144.80 | 3,457.92 | 3,587.01 | 4,146.39 | **15,336.13** |
| 401 | 2,018.72 | 1,646.74 | 1,605.60 | 1,451.21 | **6,722.26** |
| 402 | 14,266.82 | 11,454.20 | 11,708.58 | 12,516.32 | **49,945.92** |
| 403 | 6,294.41 | 5,387.99 | 5,356.45 | 5,514.95 | **22,553.80** |
| 404 | 2,550.69 | 2,040.47 | 2,069.55 | 2,329.94 | **8,990.65** |
| 405 | 5,905.25 | 4,781.73 | 4,963.51 | 5,596.43 | **21,246.92** |
| 406 | 997.42 | 834.01 | 864.06 | 1,016.23 | **3,711.71** |
| 407 | 1,875.76 | 1,389.65 | 1,590.53 | 1,614.70 | **6,470.63** |
| 408 | 321.36 | 225.41 | 219.88 | 251.65 | **1,018.29** |
| 409 | 684.53 | 670.75 | 620.05 | 623.90 | **2,599.22** |
| 410 | 10,815.58 | 8,738.81 | 9,086.86 | 10,064.05 | **38,705.29** |
| 411 | 2,414.82 | 1,949.26 | 2,013.74 | 2,145.12 | **8,522.94** |
| 412 | 719.82 | 595.63 | 650.27 | 697.68 | **2,663.40** |
| 413 | 458.06 | 361.41 | 391.54 | 451.12 | **1,662.13** |
| 414 | 297.53 | 204.76 | 231.69 | 235.02 | **969.00** |
| 415 | 1,201.24 | 945.92 | 976.71 | 897.07 | **4,020.94** |
| 416 | 658.67 | 549.00 | 700.32 | 662.41 | **2,570.40** |
| 417 | 877.21 | 547.48 | 666.61 | 654.07 | **2,745.37** |

| | | | | | |
|---|---|---|---|---|---|
| 418 | 332.28 | 294.54 | 272.07 | 280.41 | **1,179.30** |
| 420 | 4,813.99 | 3,695.59 | 3,820.46 | 3,848.01 | **16,178.05** |
| 421 | 3,352.90 | 2,630.98 | 2,671.82 | 3,028.72 | **11,684.41** |
| 422 | 2,802.19 | 2,072.08 | 2,350.08 | 2,260.24 | **9,484.60** |
| 423 | 2,984.38 | 2,169.16 | 2,320.96 | 2,504.44 | **9,978.94** |
| 424 | 3,334.01 | 2,465.04 | 2,758.46 | 2,798.84 | **11,356.36** |
| 425 | 1,037.59 | 799.55 | 893.69 | 936.50 | **3,667.33** |
| 426 | 786.09 | 641.05 | 554.61 | 647.23 | **2,628.98** |
| 427 | 3,094.80 | 2,594.43 | 2,688.21 | 2,782.94 | **11,160.39** |
| **KY - KENTUCKY - Total** | **79,040.91** | **63,143.59** | **65,633.32** | **69,955.57** | **277,773.39** |

**STATE: LA - LOUISIANA**

| REGISTRANT ZIP CODE 3 | 1ST QUARTER | 2ND QUARTER | 3RD QUARTER | 4TH QUARTER | TOTAL GRAMS |
|---|---|---|---|---|---|
| 700 | 9,753.01 | 6,834.64 | 7,454.86 | 8,689.98 | **32,732.48** |
| 701 | 4,462.35 | 3,365.61 | 3,879.53 | 4,172.62 | **15,880.10** |
| 703 | 4,721.19 | 3,093.38 | 3,686.05 | 4,268.77 | **15,769.40** |
| 704 | 9,305.65 | 6,839.68 | 7,287.62 | 8,458.11 | **31,891.05** |
| 705 | 13,113.00 | 9,011.97 | 10,169.10 | 11,648.45 | **43,942.53** |
| 706 | 7,096.98 | 5,320.92 | 4,926.82 | 5,397.38 | **22,742.09** |
| 707 | 10,374.56 | 7,261.86 | 8,729.62 | 9,678.53 | **36,044.56** |
| 708 | 6,849.19 | 4,842.50 | 5,583.35 | 6,040.71 | **23,315.75** |
| 710 | 1,772.45 | 1,415.78 | 1,438.25 | 1,557.43 | **6,183.91** |
| 711 | 4,613.30 | 3,215.00 | 3,637.54 | 4,152.16 | **15,618.00** |
| 712 | 4,870.88 | 3,298.02 | 3,634.76 | 4,190.75 | **15,994.41** |
| 713 | 3,622.58 | 2,593.09 | 2,905.03 | 3,285.64 | **12,406.34** |
| 714 | 2,589.73 | 1,853.17 | 1,978.01 | 2,206.59 | **8,627.49** |
| **LA - LOUISIANA - Total** | **83,144.88** | **58,945.61** | **65,310.52** | **73,747.11** | **281,148.12** |

**STATE: MA - MASSACHUSETTS**

| REGISTRANT ZIP CODE 3 | 1ST QUARTER | 2ND QUARTER | 3RD QUARTER | 4TH QUARTER | TOTAL GRAMS |
|---|---|---|---|---|---|
| 010 | 10,175.79 | 8,511.86 | 8,529.22 | 9,230.15 | **36,447.02** |
| 011 | 4,137.77 | 3,066.33 | 2,977.97 | 3,531.91 | **13,713.97** |
| 012 | 3,989.95 | 3,669.98 | 3,693.28 | 3,767.84 | **15,121.04** |
| 013 | 2,127.96 | 1,749.84 | 1,904.78 | 1,917.03 | **7,699.60** |
| 014 | 6,099.12 | 4,991.25 | 5,366.98 | 5,625.47 | **22,082.81** |
| 015 | 8,144.20 | 7,129.03 | 7,024.66 | 7,943.59 | **30,241.48** |
| 016 | 4,113.80 | 3,367.87 | 3,493.51 | 3,622.42 | **14,597.59** |
| 017 | 10,914.53 | 9,050.69 | 9,292.33 | 10,057.02 | **39,314.57** |
| 018 | 15,917.55 | 12,799.17 | 12,944.55 | 14,301.03 | **55,962.29** |
| 019 | 11,040.93 | 9,568.48 | 9,404.60 | 10,440.31 | **40,454.32** |
| 020 | 10,436.83 | 9,224.05 | 9,131.97 | 10,048.78 | **38,841.63** |
| 021 | 22,000.21 | 17,113.08 | 17,024.37 | 19,727.01 | **75,864.67** |
| 022 | 1,122.10 | 744.64 | 832.02 | 884.36 | **3,583.11** |
| 023 | 9,228.62 | 7,548.92 | 7,726.20 | 8,103.54 | **32,607.28** |
| 024 | 11,948.18 | 10,404.97 | 10,115.93 | 11,437.66 | **43,906.73** |
| 025 | 2,901.39 | 2,713.57 | 2,795.74 | 2,867.18 | **11,277.88** |
| 026 | 3,637.79 | 3,974.53 | 3,895.71 | 3,361.59 | **14,869.63** |
| 027 | 9,405.12 | 7,725.24 | 7,680.27 | 8,028.24 | **32,838.87** |
| **MA - MASSACHUSETTS - Total** | **147,341.82** | **123,353.48** | **123,834.08** | **134,895.12** | **529,424.49** |

**STATE: MD - MARYLAND**

| REGISTRANT ZIP CODE 3 | 1ST QUARTER | 2ND QUARTER | 3RD QUARTER | 4TH QUARTER | TOTAL GRAMS |
|---|---|---|---|---|---|
| 206 | 4,022.75 | 3,119.10 | 3,206.50 | 3,574.31 | **13,922.65** |
| 207 | 7,211.84 | 5,361.35 | 5,965.70 | 5,601.72 | **24,140.61** |
| 208 | 16,280.59 | 13,874.27 | 14,184.54 | 14,750.12 | **59,089.52** |
| 209 | 3,321.63 | 2,670.14 | 2,465.48 | 2,918.44 | **11,375.68** |
| 210 | 18,254.35 | 15,501.23 | 14,895.51 | 17,146.40 | **65,797.50** |
| 211 | 8,943.21 | 7,382.69 | 7,459.63 | 8,379.34 | **32,164.87** |
| 212 | 20,421.81 | 16,802.53 | 15,924.05 | 17,548.98 | **70,697.38** |
| 214 | 2,169.88 | 1,755.31 | 1,549.90 | 1,761.43 | **7,236.52** |
| 215 | 3,883.93 | 3,412.57 | 3,322.08 | 3,422.71 | **14,041.29** |
| 216 | 3,974.00 | 3,232.44 | 3,017.30 | 3,731.83 | **13,955.56** |
| 217 | 12,240.50 | 10,323.54 | 10,497.02 | 11,369.60 | **44,430.65** |

| | | | | | |
|---|---|---|---|---|---|
| 218 | 3,364.17 | 2,802.79 | 2,689.56 | 3,083.70 | **11,940.23** |
| 219 | 1,940.73 | 1,645.93 | 1,601.98 | 1,811.59 | **7,000.23** |
| **MD - MARYLAND - Total** | **106,029.40** | **87,883.90** | **86,779.24** | **95,100.16** | **375,792.69** |

**STATE: ME - MAINE**

| REGISTRANT ZIP CODE 3 | 1ST QUARTER | 2ND QUARTER | 3RD QUARTER | 4TH QUARTER | TOTAL GRAMS |
|---|---|---|---|---|---|
| 039 | 751.45 | 686.97 | 612.34 | 689.01 | **2,739.77** |
| 040 | 7,935.55 | 7,348.75 | 7,605.48 | 7,936.01 | **30,825.79** |
| 041 | 2,986.88 | 2,713.01 | 2,830.22 | 2,836.57 | **11,366.67** |
| 042 | 3,182.73 | 2,802.44 | 2,996.02 | 3,046.67 | **12,027.86** |
| 043 | 2,908.77 | 2,785.59 | 2,832.94 | 2,942.44 | **11,469.75** |
| 044 | 6,906.17 | 5,683.93 | 5,928.17 | 6,169.06 | **24,687.34** |
| 045 | 1,409.33 | 1,477.40 | 1,557.37 | 1,495.33 | **5,939.43** |
| 046 | 1,966.44 | 1,997.81 | 2,097.62 | 2,170.71 | **8,232.59** |
| 047 | 2,032.21 | 1,825.61 | 1,703.80 | 1,845.72 | **7,407.34** |
| 048 | 1,373.20 | 1,419.12 | 1,442.53 | 1,458.25 | **5,693.10** |
| 049 | 5,147.67 | 4,695.52 | 4,993.63 | 4,943.38 | **19,780.21** |
| **ME - MAINE - Total** | **36,600.41** | **33,436.15** | **34,600.14** | **35,533.15** | **140,169.85** |

**STATE: MI - MICHIGAN**

| REGISTRANT ZIP CODE 3 | 1ST QUARTER | 2ND QUARTER | 3RD QUARTER | 4TH QUARTER | TOTAL GRAMS |
|---|---|---|---|---|---|
| 480 | 21,917.62 | 17,883.00 | 17,728.85 | 19,759.39 | **77,288.87** |
| 481 | 26,516.45 | 20,931.06 | 21,990.36 | 24,426.45 | **93,864.31** |
| 482 | 4,109.25 | 3,181.87 | 2,964.86 | 3,370.32 | **13,626.30** |
| 483 | 18,599.16 | 15,506.32 | 15,813.68 | 18,022.64 | **67,941.79** |
| 484 | 6,128.69 | 4,948.93 | 5,173.43 | 5,667.79 | **21,918.84** |
| 485 | 1,915.84 | 1,647.08 | 1,494.89 | 1,807.13 | **6,864.94** |
| 486 | 6,636.34 | 5,462.69 | 5,498.40 | 5,849.23 | **23,446.67** |
| 487 | 3,475.63 | 3,205.71 | 3,151.83 | 3,391.33 | **13,224.50** |
| 488 | 12,692.10 | 10,017.84 | 10,537.60 | 11,745.23 | **44,992.77** |
| 489 | 2,744.28 | 2,380.17 | 2,248.94 | 2,403.49 | **9,776.88** |
| 490 | 13,375.81 | 11,451.21 | 11,578.68 | 12,538.58 | **48,944.28** |
| 491 | 953.01 | 891.29 | 861.96 | 957.80 | **3,664.06** |
| 492 | 5,453.57 | 4,059.23 | 4,229.21 | 4,857.10 | **18,599.11** |
| 493 | 5,446.64 | 4,509.64 | 4,702.65 | 5,050.56 | **19,709.49** |
| 494 | 16,167.69 | 13,522.19 | 14,235.91 | 15,848.72 | **59,774.51** |
| 495 | 11,955.00 | 10,267.07 | 10,498.84 | 11,714.32 | **44,435.23** |
| 496 | 6,186.12 | 5,459.99 | 5,567.64 | 5,978.27 | **23,192.03** |
| 497 | 4,939.05 | 4,206.36 | 4,542.14 | 4,638.91 | **18,326.46** |
| 498 | 6,342.16 | 5,949.69 | 6,101.93 | 6,322.52 | **24,716.31** |
| 499 | 1,373.31 | 1,185.77 | 1,250.87 | 1,358.25 | **5,168.21** |
| **MI - MICHIGAN - Total** | **176,927.74** | **146,667.10** | **150,172.68** | **165,708.03** | **639,475.55** |

**STATE: MN - MINNESOTA**

| REGISTRANT ZIP CODE 3 | 1ST QUARTER | 2ND QUARTER | 3RD QUARTER | 4TH QUARTER | TOTAL GRAMS |
|---|---|---|---|---|---|
| 550 | 11,912.72 | 10,045.72 | 10,527.90 | 11,549.06 | **44,035.40** |
| 551 | 14,787.26 | 13,117.98 | 12,510.41 | 14,226.42 | **54,642.07** |
| 553 | 21,111.65 | 18,070.55 | 18,442.36 | 20,088.89 | **77,713.44** |
| 554 | 19,965.52 | 17,517.44 | 17,278.55 | 19,377.19 | **74,138.69** |
| 556 | 126.10 | 116.86 | 111.90 | 113.79 | **468.65** |
| 557 | 2,533.99 | 2,283.48 | 2,184.80 | 2,529.01 | **9,531.28** |
| 558 | 2,595.26 | 2,312.57 | 2,167.51 | 2,415.76 | **9,491.10** |
| 559 | 8,835.24 | 7,742.57 | 7,807.55 | 8,618.95 | **33,004.31** |
| 560 | 4,808.52 | 4,220.99 | 4,133.58 | 4,683.01 | **17,846.10** |
| 561 | 1,374.82 | 1,287.20 | 1,167.42 | 1,363.29 | **5,192.73** |
| 562 | 2,436.54 | 1,927.61 | 2,036.04 | 2,209.70 | **8,609.90** |
| 563 | 6,007.02 | 5,206.42 | 5,314.28 | 5,545.55 | **22,073.27** |
| 564 | 2,202.59 | 1,843.48 | 1,919.84 | 1,929.58 | **7,895.49** |
| 565 | 3,972.55 | 3,381.50 | 3,463.04 | 3,905.28 | **14,722.37** |
| 566 | 1,113.85 | 863.70 | 1,057.19 | 891.54 | **3,926.29** |
| 567 | 1,016.10 | 941.99 | 913.76 | 931.88 | **3,803.73** |
| **MN - MINNESOTA - Total** | **104,799.72** | **90,880.06** | **91,036.15** | **100,378.88** | **387,094.81** |

**STATE: MO - MISSOURI**

| REGISTRANT ZIP CODE 3 | 1ST QUARTER | 2ND QUARTER | 3RD QUARTER | 4TH QUARTER | TOTAL GRAMS |
|---|---|---|---|---|---|
| 630 | 16,855.66 | 13,511.97 | 14,473.44 | 16,172.48 | **61,013.55** |
| 631 | 51,134.95 | 49,671.96 | 49,327.96 | 57,345.03 | **207,479.91** |
| 633 | 12,835.64 | 10,504.95 | 11,491.51 | 12,717.32 | **47,549.42** |
| 634 | 1,410.73 | 1,075.13 | 1,188.72 | 1,303.89 | **4,978.46** |
| 635 | 789.11 | 647.16 | 685.35 | 741.05 | **2,862.67** |
| 636 | 2,251.55 | 1,838.12 | 2,038.57 | 2,204.98 | **8,333.22** |
| 637 | 2,240.98 | 1,610.67 | 1,894.26 | 1,987.53 | **7,733.44** |
| 638 | 2,178.09 | 1,659.88 | 1,833.04 | 2,000.57 | **7,671.59** |
| 639 | 793.99 | 632.93 | 720.45 | 783.52 | **2,930.88** |
| 640 | 10,185.18 | 8,701.14 | 8,798.14 | 10,138.06 | **37,822.51** |
| 641 | 7,897.57 | 6,661.09 | 6,746.24 | 7,870.31 | **29,175.20** |
| 644 | 1,084.37 | 978.71 | 1,015.23 | 1,102.02 | **4,180.33** |
| 645 | 2,201.32 | 1,619.99 | 1,786.23 | 1,856.99 | **7,464.53** |
| 646 | 502.19 | 417.27 | 432.75 | 531.68 | **1,883.88** |
| 647 | 1,033.40 | 970.21 | 1,007.65 | 1,188.08 | **4,199.34** |
| 648 | 3,203.50 | 2,640.15 | 2,848.13 | 3,093.51 | **11,785.29** |
| 650 | 1,481.99 | 1,323.85 | 1,198.72 | 1,322.79 | **5,327.35** |
| 651 | 1,822.36 | 1,491.23 | 1,606.54 | 1,766.21 | **6,686.34** |
| 652 | 4,111.59 | 3,499.97 | 3,870.96 | 4,140.73 | **15,623.25** |
| 653 | 1,223.30 | 1,077.73 | 1,171.35 | 1,215.69 | **4,688.07** |
| 654 | 1,586.75 | 1,260.88 | 1,163.16 | 1,390.41 | **5,401.20** |
| 655 | 1,350.83 | 1,217.44 | 1,128.73 | 1,250.25 | **4,947.25** |
| 656 | 1,634.79 | 1,504.44 | 1,566.12 | 1,684.94 | **6,390.29** |
| 657 | 3,340.91 | 2,848.29 | 3,209.53 | 3,290.88 | **12,689.60** |
| 658 | 4,091.89 | 3,526.00 | 3,729.45 | 4,039.09 | **15,386.43** |
| **MO - MISSOURI - Total** | **137,242.64** | **120,891.16** | **124,932.22** | **141,138.00** | **524,204.02** |

**STATE: MS - MISSISSIPPI**

| REGISTRANT ZIP CODE 3 | 1ST QUARTER | 2ND QUARTER | 3RD QUARTER | 4TH QUARTER | TOTAL GRAMS |
|---|---|---|---|---|---|
| 386 | 6,582.88 | 4,827.18 | 5,415.13 | 5,954.65 | **22,779.85** |
| 387 | 656.43 | 438.18 | 587.53 | 524.73 | **2,206.87** |
| 388 | 3,553.63 | 2,831.62 | 3,168.98 | 3,590.52 | **13,144.75** |
| 389 | 1,241.98 | 861.73 | 986.50 | 980.15 | **4,070.37** |
| 390 | 3,556.04 | 2,476.47 | 2,799.00 | 3,153.03 | **11,984.54** |
| 391 | 3,650.19 | 2,339.75 | 2,816.47 | 3,212.77 | **12,019.17** |
| 392 | 2,965.84 | 2,203.46 | 2,526.57 | 3,054.21 | **10,750.08** |
| 393 | 2,478.44 | 1,923.82 | 1,961.01 | 2,373.22 | **8,736.49** |
| 394 | 7,661.41 | 5,578.89 | 6,139.87 | 7,075.44 | **26,455.60** |
| 395 | 9,271.08 | 6,254.16 | 7,410.83 | 8,167.63 | **31,103.71** |
| 396 | 1,997.77 | 1,406.58 | 1,647.15 | 1,679.15 | **6,730.65** |
| 397 | 1,749.93 | 1,169.36 | 1,359.20 | 1,541.91 | **5,820.41** |
| **MS - MISSISSIPPI - Total** | **45,365.63** | **32,311.22** | **36,818.24** | **41,307.41** | **155,802.50** |

**STATE: MT - MONTANA**

| REGISTRANT ZIP CODE 3 | 1ST QUARTER | 2ND QUARTER | 3RD QUARTER | 4TH QUARTER | TOTAL GRAMS |
|---|---|---|---|---|---|
| 590 | 853.44 | 797.87 | 845.95 | 903.33 | **3,400.60** |
| 591 | 4,469.34 | 3,824.90 | 4,077.75 | 4,237.79 | **16,609.78** |
| 592 | 592.17 | 479.56 | 421.22 | 510.84 | **2,003.78** |
| 593 | 694.30 | 628.02 | 651.89 | 716.28 | **2,690.49** |
| 594 | 2,477.92 | 2,205.81 | 2,158.08 | 2,385.74 | **9,227.55** |
| 595 | 303.48 | 311.37 | 287.20 | 323.61 | **1,225.66** |
| 596 | 1,320.51 | 1,308.06 | 1,367.44 | 1,458.76 | **5,454.77** |
| 597 | 2,620.31 | 2,278.03 | 2,428.86 | 2,552.19 | **9,879.39** |
| 598 | 2,770.24 | 2,644.64 | 2,673.43 | 2,765.49 | **10,853.80** |
| 599 | 1,479.44 | 1,368.05 | 1,467.24 | 1,604.00 | **5,918.73** |
| **MT - MONTANA - Total** | **17,581.15** | **15,846.31** | **16,379.06** | **17,458.03** | **67,264.56** |

**STATE: NC - NORTH CAROLINA**

| REGISTRANT ZIP CODE 3 | 1ST QUARTER | 2ND QUARTER | 3RD QUARTER | 4TH QUARTER | TOTAL GRAMS |
|---|---|---|---|---|---|
| 270 | 5,029.50 | 4,137.51 | 4,423.35 | 5,026.96 | **18,617.32** |
| 271 | 5,457.94 | 4,288.92 | 4,466.81 | 4,938.54 | **19,152.20** |
| 272 | 11,393.28 | 9,386.36 | 9,519.23 | 10,294.41 | **40,593.29** |

| | | | | | |
|---|---|---|---|---|---|
| 273 | 6,863.25 | 5,660.57 | 5,708.98 | 6,357.37 | **24,590.16** |
| 274 | 5,772.03 | 4,417.54 | 4,675.27 | 5,203.38 | **20,068.22** |
| 275 | 25,886.19 | 20,624.07 | 21,512.68 | 24,087.36 | **92,110.30** |
| 276 | 10,321.62 | 8,416.70 | 8,427.68 | 9,925.04 | **37,091.04** |
| 277 | 5,846.85 | 4,763.95 | 4,913.17 | 5,404.85 | **20,928.82** |
| 278 | 9,346.02 | 7,174.36 | 7,420.08 | 8,071.86 | **32,012.32** |
| 279 | 2,420.59 | 2,129.69 | 2,188.67 | 2,294.31 | **9,033.26** |
| 280 | 17,599.88 | 14,618.27 | 15,099.46 | 16,537.77 | **63,855.39** |
| 281 | 13,489.39 | 10,358.18 | 11,139.43 | 12,232.76 | **47,219.76** |
| 282 | 13,620.57 | 11,086.75 | 11,519.26 | 13,008.59 | **49,235.17** |
| 283 | 16,221.00 | 12,270.49 | 13,050.38 | 13,215.51 | **54,757.38** |
| 284 | 8,565.18 | 7,360.19 | 7,841.99 | 8,305.70 | **32,073.06** |
| 285 | 11,937.94 | 8,864.68 | 10,075.80 | 9,298.40 | **40,176.83** |
| 286 | 11,185.86 | 9,214.36 | 9,354.59 | 10,438.26 | **40,193.06** |
| 287 | 9,145.81 | 8,018.61 | 8,048.53 | 8,883.67 | **34,096.62** |
| 288 | 4,099.74 | 3,686.46 | 3,509.21 | 3,946.95 | **15,242.36** |
| 289 | 559.26 | 463.62 | 470.44 | 528.64 | **2,021.95** |
| **NC - NORTH CAROLINA - Total** | **194,761.90** | **156,941.27** | **163,365.00** | **178,000.33** | **693,068.50** |

**STATE: ND - NORTH DAKOTA**

| REGISTRANT ZIP CODE 3 | 1ST QUARTER | 2ND QUARTER | 3RD QUARTER | 4TH QUARTER | TOTAL GRAMS |
|---|---|---|---|---|---|
| 580 | 1,697.15 | 1,557.19 | 1,474.93 | 1,664.64 | **6,393.92** |
| 581 | 3,899.10 | 3,237.85 | 3,025.77 | 3,586.64 | **13,749.37** |
| 582 | 2,474.11 | 2,024.71 | 1,902.19 | 2,522.93 | **8,923.95** |
| 583 | 956.03 | 814.05 | 695.70 | 836.77 | **3,302.56** |
| 584 | 625.29 | 553.79 | 560.80 | 622.78 | **2,362.66** |
| 585 | 2,861.78 | 2,379.41 | 2,352.08 | 2,813.93 | **10,407.20** |
| 586 | 925.94 | 780.16 | 744.72 | 795.02 | **3,245.85** |
| 587 | 1,943.95 | 1,579.89 | 1,552.47 | 1,765.79 | **6,842.10** |
| 588 | 751.49 | 652.43 | 650.19 | 746.19 | **2,800.30** |
| **ND - NORTH DAKOTA - Total** | **16,134.84** | **13,579.49** | **12,958.86** | **15,354.71** | **58,027.91** |

**STATE: NE - NEBRASKA**

| REGISTRANT ZIP CODE 3 | 1ST QUARTER | 2ND QUARTER | 3RD QUARTER | 4TH QUARTER | TOTAL GRAMS |
|---|---|---|---|---|---|
| 680 | 3,585.42 | 2,849.07 | 3,354.26 | 3,693.32 | **13,482.07** |
| 681 | 13,309.66 | 11,205.05 | 11,819.77 | 13,029.99 | **49,364.47** |
| 683 | 873.49 | 719.69 | 784.76 | 874.91 | **3,252.85** |
| 684 | 780.81 | 675.97 | 768.49 | 828.71 | **3,053.98** |
| 685 | 6,097.12 | 4,919.01 | 5,575.42 | 5,865.71 | **22,457.26** |
| 686 | 845.07 | 662.22 | 737.97 | 801.61 | **3,046.86** |
| 687 | 1,626.14 | 1,360.88 | 1,461.84 | 1,613.84 | **6,062.70** |
| 688 | 2,362.60 | 2,044.48 | 2,203.75 | 2,591.39 | **9,202.21** |
| 689 | 926.90 | 779.40 | 939.12 | 1,014.45 | **3,659.88** |
| 690 | 306.40 | 281.58 | 306.27 | 278.94 | **1,173.19** |
| 691 | 1,112.35 | 965.90 | 899.30 | 1,128.90 | **4,106.44** |
| 692 | 13.49 | 16.09 | 27.93 | 27.50 | **85.01** |
| 693 | 764.12 | 659.25 | 735.87 | 667.35 | **2,826.59** |
| **NE - NEBRASKA - Total** | **32,603.57** | **27,138.59** | **29,614.75** | **32,416.61** | **121,773.51** |

**STATE: NH - NEW HAMPSHIRE**

| REGISTRANT ZIP CODE 3 | 1ST QUARTER | 2ND QUARTER | 3RD QUARTER | 4TH QUARTER | TOTAL GRAMS |
|---|---|---|---|---|---|
| 030 | 8,777.00 | 8,262.03 | 8,132.53 | 8,775.27 | **33,946.83** |
| 031 | 5,136.41 | 4,435.14 | 4,581.16 | 4,868.90 | **19,021.60** |
| 032 | 3,292.39 | 3,154.79 | 3,274.96 | 3,306.47 | **13,028.61** |
| 033 | 2,548.09 | 2,178.95 | 2,238.21 | 2,365.36 | **9,330.60** |
| 034 | 2,060.96 | 1,663.00 | 1,965.46 | 1,959.32 | **7,648.73** |
| 035 | 1,773.99 | 1,673.50 | 1,580.38 | 1,640.31 | **6,668.18** |
| 036 | 72.59 | 89.90 | 113.63 | 84.19 | **360.31** |
| 037 | 3,501.45 | 3,226.49 | 3,259.48 | 3,633.92 | **13,621.34** |
| 038 | 7,875.60 | 7,392.75 | 7,441.74 | 8,015.86 | **30,725.95** |
| **NH - NEW HAMPSHIRE - Total** | **35,038.47** | **32,076.53** | **32,587.55** | **34,649.60** | **134,352.15** |

**STATE: NJ - NEW JERSEY**

| REGISTRANT ZIP CODE 3 | 1ST QUARTER | 2ND QUARTER | 3RD QUARTER | 4TH QUARTER | TOTAL GRAMS |
|---|---|---|---|---|---|

| | | | | | |
|---|---|---|---|---|---|
| 070 | 16,460.75 | 13,924.88 | 13,411.02 | 15,113.17 | **58,909.83** |
| 071 | 2,005.75 | 1,389.74 | 1,338.91 | 1,541.04 | **6,275.44** |
| 072 | 928.09 | 609.35 | 646.06 | 759.69 | **2,943.19** |
| 073 | 1,100.59 | 966.97 | 754.76 | 896.81 | **3,719.13** |
| 074 | 5,115.34 | 4,408.37 | 4,599.76 | 5,413.26 | **19,536.73** |
| 075 | 770.19 | 554.16 | 485.35 | 587.73 | **2,397.42** |
| 076 | 5,202.74 | 4,467.30 | 4,463.12 | 5,013.86 | **19,147.02** |
| 077 | 6,519.95 | 5,754.68 | 5,567.45 | 6,264.08 | **24,106.16** |
| 078 | 4,922.14 | 4,133.75 | 4,036.47 | 4,617.97 | **17,710.33** |
| 079 | 4,972.92 | 4,398.42 | 3,885.41 | 5,104.86 | **18,361.61** |
| 080 | 18,275.05 | 15,313.20 | 14,926.18 | 17,191.02 | **65,705.45** |
| 081 | 1,597.56 | 1,361.04 | 1,216.93 | 1,494.16 | **5,669.69** |
| 082 | 2,675.09 | 2,689.55 | 2,616.69 | 2,728.09 | **10,709.41** |
| 083 | 2,463.53 | 2,079.97 | 1,989.52 | 2,207.52 | **8,740.54** |
| 084 | 218.41 | 214.06 | 328.28 | 233.53 | **994.28** |
| 085 | 3,887.63 | 3,470.01 | 3,234.15 | 3,767.19 | **14,358.98** |
| 086 | 2,952.08 | 2,639.93 | 2,100.55 | 2,600.97 | **10,293.52** |
| 087 | 5,540.97 | 4,778.85 | 4,755.40 | 5,364.97 | **20,440.19** |
| 088 | 8,412.84 | 7,249.18 | 6,966.70 | 7,927.71 | **30,556.43** |
| 089 | 855.63 | 663.52 | 652.74 | 819.10 | **2,990.99** |
| **NJ - NEW JERSEY - Total** | **94,877.24** | **81,066.91** | **77,975.47** | **89,646.71** | **343,566.33** |

**STATE: NM - NEW MEXICO**

| REGISTRANT ZIP CODE 3 | 1ST QUARTER | 2ND QUARTER | 3RD QUARTER | 4TH QUARTER | TOTAL GRAMS |
|---|---|---|---|---|---|
| 870 | 1,362.91 | 1,091.02 | 1,144.67 | 1,253.51 | **4,852.11** |
| 871 | 8,085.34 | 6,923.70 | 7,425.76 | 7,139.82 | **29,574.62** |
| 873 | 462.52 | 327.86 | 334.14 | 332.33 | **1,456.86** |
| 874 | 1,305.82 | 1,242.77 | 1,227.82 | 1,125.61 | **4,902.03** |
| 875 | 1,916.95 | 1,679.62 | 1,637.18 | 1,825.65 | **7,059.41** |
| 877 | 233.41 | 229.42 | 203.14 | 214.06 | **880.03** |
| 878 | 80.43 | 63.48 | 79.04 | 61.49 | **284.43** |
| 879 | 81.20 | 68.15 | 67.54 | 72.90 | **289.79** |
| 880 | 1,848.51 | 1,621.21 | 1,777.19 | 1,871.66 | **7,118.58** |
| 881 | 460.33 | 314.58 | 424.11 | 363.54 | **1,562.56** |
| 882 | 1,835.89 | 1,483.49 | 1,453.46 | 1,392.40 | **6,165.24** |
| 883 | 640.62 | 605.39 | 570.05 | 679.83 | **2,495.89** |
| 884 | 71.43 | 82.07 | 65.64 | 89.16 | **308.30** |
| **NM - NEW MEXICO - Total** | **18,385.37** | **15,732.77** | **16,409.74** | **16,421.96** | **66,949.84** |

**STATE: NV - NEVADA**

| REGISTRANT ZIP CODE 3 | 1ST QUARTER | 2ND QUARTER | 3RD QUARTER | 4TH QUARTER | TOTAL GRAMS |
|---|---|---|---|---|---|
| 890 | 3,727.51 | 3,158.64 | 3,122.66 | 3,487.64 | **13,496.44** |
| 891 | 6,864.15 | 5,706.90 | 5,678.66 | 6,219.90 | **24,469.62** |
| 893 | 25.94 | 16.78 | 13.94 | 25.36 | **82.02** |
| 894 | 2,001.53 | 1,720.28 | 1,788.08 | 1,861.32 | **7,371.21** |
| 895 | 2,325.87 | 2,299.54 | 2,397.44 | 2,500.21 | **9,523.06** |
| 897 | 464.24 | 426.83 | 421.71 | 420.75 | **1,733.53** |
| 898 | 439.67 | 415.79 | 328.43 | 393.74 | **1,577.64** |
| **NV - NEVADA - Total** | **15,848.91** | **13,744.76** | **13,750.92** | **14,908.92** | **58,253.51** |

**STATE: NY - NEW YORK**

| REGISTRANT ZIP CODE 3 | 1ST QUARTER | 2ND QUARTER | 3RD QUARTER | 4TH QUARTER | TOTAL GRAMS |
|---|---|---|---|---|---|
| 100 | 24,386.72 | 15,629.70 | 16,671.41 | 20,563.68 | **77,251.51** |
| 101 | 1,057.05 | 493.72 | 488.27 | 793.66 | **2,832.71** |
| 102 | 161.24 | 99.15 | 87.30 | 98.15 | **445.85** |
| 103 | 2,743.81 | 2,258.97 | 2,075.60 | 2,312.75 | **9,391.13** |
| 104 | 6,773.46 | 5,583.71 | 4,782.11 | 5,475.88 | **22,615.16** |
| 105 | 10,648.69 | 9,034.19 | 8,734.41 | 10,156.09 | **38,573.37** |
| 106 | 1,133.11 | 816.97 | 891.05 | 1,070.63 | **3,911.76** |
| 107 | 1,817.44 | 1,541.11 | 1,560.26 | 1,775.61 | **6,694.42** |
| 108 | 940.87 | 796.72 | 793.21 | 876.55 | **3,407.36** |
| 109 | 7,270.90 | 5,794.36 | 5,827.40 | 6,474.49 | **25,367.15** |
| 110 | 3,299.88 | 2,842.95 | 2,652.22 | 2,989.02 | **11,784.06** |

| | | | | | |
|---|---|---|---|---|---|
| 111 | 1,178.99 | 1,048.08 | 965.73 | 1,133.41 | **4,326.20** |
| 112 | 12,044.06 | 10,196.32 | 9,542.15 | 11,155.50 | **42,938.02** |
| 113 | 5,071.59 | 3,697.49 | 3,433.49 | 4,244.44 | **16,447.01** |
| 114 | 2,089.32 | 1,698.04 | 1,504.67 | 1,760.47 | **7,052.51** |
| 115 | 8,701.47 | 7,273.37 | 7,368.89 | 8,535.31 | **31,879.04** |
| 116 | 636.31 | 572.93 | 479.13 | 567.12 | **2,255.49** |
| 117 | 20,623.75 | 17,589.86 | 16,841.21 | 19,378.33 | **74,433.14** |
| 118 | 1,361.09 | 1,123.37 | 1,199.31 | 1,337.58 | **5,021.35** |
| 119 | 2,786.81 | 3,797.06 | 3,586.42 | 2,966.20 | **13,136.50** |
| 120 | 7,531.53 | 6,663.57 | 6,718.34 | 6,928.26 | **27,841.70** |
| 121 | 4,750.31 | 4,281.68 | 4,125.63 | 4,622.25 | **17,779.86** |
| 122 | 3,112.00 | 2,352.42 | 2,302.31 | 2,560.75 | **10,327.48** |
| 123 | 3,330.29 | 2,574.45 | 2,685.54 | 3,118.65 | **11,708.93** |
| 124 | 1,903.48 | 1,801.48 | 1,724.98 | 1,916.67 | **7,346.62** |
| 125 | 5,516.88 | 4,660.18 | 4,544.13 | 5,044.37 | **19,765.57** |
| 126 | 1,060.68 | 834.98 | 933.51 | 1,024.97 | **3,854.14** |
| 127 | 950.31 | 821.05 | 832.43 | 903.37 | **3,507.17** |
| 128 | 5,050.22 | 4,387.58 | 4,500.63 | 4,795.68 | **18,734.11** |
| 129 | 4,249.04 | 3,913.04 | 4,024.60 | 4,428.22 | **16,614.89** |
| 130 | 6,539.76 | 5,670.63 | 5,314.08 | 6,270.69 | **23,795.16** |
| 131 | 2,215.62 | 1,953.99 | 1,987.11 | 2,029.20 | **8,185.92** |
| 132 | 3,622.66 | 2,724.57 | 2,841.80 | 3,187.07 | **12,376.10** |
| 133 | 1,929.26 | 1,774.81 | 1,839.62 | 1,960.69 | **7,504.37** |
| 134 | 2,770.46 | 2,317.48 | 2,252.17 | 2,504.56 | **9,844.67** |
| 135 | 1,296.32 | 863.15 | 880.09 | 937.94 | **3,977.51** |
| 136 | 5,738.12 | 5,083.31 | 4,959.19 | 5,233.35 | **21,013.96** |
| 137 | 1,456.80 | 1,177.21 | 1,129.17 | 1,369.79 | **5,132.97** |
| 138 | 2,586.26 | 2,305.34 | 2,016.29 | 2,341.70 | **9,249.59** |
| 139 | 1,177.92 | 1,012.12 | 1,012.19 | 1,123.74 | **4,325.96** |
| 140 | 5,703.47 | 4,780.98 | 5,054.85 | 5,563.43 | **21,102.73** |
| 141 | 2,515.20 | 2,196.10 | 2,086.14 | 2,493.72 | **9,291.15** |
| 142 | 7,550.77 | 6,408.48 | 6,483.68 | 6,930.20 | **27,373.13** |
| 143 | 674.57 | 614.71 | 605.23 | 613.84 | **2,508.34** |
| 144 | 7,639.45 | 6,768.54 | 6,578.38 | 7,364.79 | **28,351.16** |
| 145 | 7,431.30 | 6,564.09 | 6,582.75 | 7,063.04 | **27,641.19** |
| 146 | 12,370.98 | 10,285.45 | 10,069.88 | 11,183.48 | **43,909.79** |
| 147 | 2,347.44 | 1,975.40 | 2,017.32 | 2,228.02 | **8,568.18** |
| 148 | 5,505.11 | 4,428.88 | 4,720.95 | 4,845.24 | **19,500.18** |
| 149 | 1,216.09 | 1,079.20 | 1,017.90 | 1,162.03 | **4,475.22** |
| **NY - NEW YORK - Total** | **234,468.84** | **194,162.93** | **191,325.12** | **215,414.57** | **835,371.46** |

**STATE: OH - OHIO**

| REGISTRANT ZIP CODE 3 | 1ST QUARTER | 2ND QUARTER | 3RD QUARTER | 4TH QUARTER | TOTAL GRAMS |
|---|---|---|---|---|---|
| 430 | 17,453.88 | 14,689.46 | 15,352.70 | 16,513.22 | **64,009.26** |
| 431 | 8,288.98 | 6,998.09 | 7,158.58 | 7,871.19 | **30,316.85** |
| 432 | 11,200.24 | 9,232.86 | 9,733.64 | 10,696.52 | **40,863.26** |
| 433 | 2,748.41 | 2,362.10 | 2,564.81 | 2,610.84 | **10,286.16** |
| 434 | 2,895.95 | 2,285.67 | 2,540.84 | 2,784.99 | **10,507.46** |
| 435 | 6,629.08 | 5,358.48 | 5,489.64 | 5,883.15 | **23,360.36** |
| 436 | 7,144.74 | 5,805.08 | 5,600.80 | 6,202.11 | **24,752.74** |
| 437 | 2,416.56 | 2,082.68 | 2,083.90 | 2,267.11 | **8,850.25** |
| 438 | 432.34 | 359.44 | 421.45 | 430.04 | **1,643.27** |
| 439 | 2,076.63 | 1,750.99 | 1,737.84 | 2,030.00 | **7,595.46** |
| 440 | 14,935.14 | 12,192.82 | 12,545.78 | 14,106.83 | **53,780.56** |
| 441 | 16,787.22 | 13,842.60 | 14,240.42 | 15,277.00 | **60,147.23** |
| 442 | 8,654.29 | 7,262.23 | 7,453.38 | 8,515.44 | **31,885.34** |
| 443 | 3,906.56 | 3,306.70 | 3,190.91 | 3,588.31 | **13,992.48** |
| 444 | 3,728.29 | 3,044.06 | 3,217.21 | 3,413.48 | **13,403.04** |
| 445 | 2,686.10 | 2,317.47 | 2,293.96 | 2,545.16 | **9,842.69** |
| 446 | 6,041.74 | 4,976.08 | 5,244.38 | 5,782.54 | **22,044.74** |
| 447 | 4,406.14 | 3,898.93 | 3,836.02 | 4,174.32 | **16,315.41** |

| | | | | | |
|---|---|---|---|---|---|
| 448 | 4,394.56 | 3,453.97 | 3,730.31 | 4,002.28 | **15,581.12** |
| 449 | 1,575.40 | 1,350.28 | 1,417.79 | 1,574.78 | **5,918.25** |
| 450 | 16,468.95 | 13,570.85 | 14,850.55 | 16,169.52 | **61,059.87** |
| 451 | 6,447.59 | 5,634.38 | 5,654.25 | 6,409.05 | **24,145.27** |
| 452 | 19,941.51 | 15,868.33 | 16,790.60 | 19,334.61 | **71,935.06** |
| 453 | 8,408.06 | 7,011.12 | 7,476.11 | 8,123.12 | **31,018.42** |
| 454 | 9,563.59 | 8,049.77 | 8,271.23 | 9,270.60 | **35,155.19** |
| 455 | 2,195.42 | 1,849.88 | 2,000.90 | 2,207.72 | **8,253.92** |
| 456 | 6,789.78 | 5,667.94 | 6,015.65 | 6,421.57 | **24,894.94** |
| 457 | 2,360.90 | 1,811.52 | 1,948.26 | 2,133.79 | **8,254.48** |
| 458 | 5,339.47 | 4,347.82 | 4,710.36 | 5,128.16 | **19,525.82** |
| **OH - OHIO - Total** | **205,917.54** | **170,381.63** | **177,572.28** | **195,467.45** | **749,338.89** |

**STATE: OK - OKLAHOMA**

| REGISTRANT ZIP CODE 3 | 1ST QUARTER | 2ND QUARTER | 3RD QUARTER | 4TH QUARTER | TOTAL GRAMS |
|---|---|---|---|---|---|
| 730 | 10,495.88 | 8,374.21 | 9,207.18 | 9,968.95 | **38,046.22** |
| 731 | 7,597.00 | 6,134.61 | 6,712.34 | 7,081.99 | **27,525.93** |
| 734 | 719.59 | 611.46 | 656.58 | 679.34 | **2,666.98** |
| 735 | 2,847.34 | 2,138.41 | 2,831.39 | 2,184.13 | **10,001.28** |
| 736 | 828.58 | 702.94 | 668.89 | 778.67 | **2,979.07** |
| 737 | 1,059.70 | 776.72 | 953.49 | 991.00 | **3,780.92** |
| 738 | 179.93 | 158.53 | 174.52 | 179.69 | **692.67** |
| 739 | 50.42 | 44.75 | 37.71 | 51.59 | **184.46** |
| 740 | 10,350.52 | 8,718.26 | 9,175.63 | 9,806.11 | **38,050.53** |
| 741 | 5,653.28 | 4,278.15 | 4,740.25 | 4,946.31 | **19,617.99** |
| 743 | 1,622.98 | 1,323.55 | 1,508.08 | 1,471.91 | **5,926.52** |
| 744 | 3,059.34 | 2,429.43 | 2,661.72 | 2,927.66 | **11,078.16** |
| 745 | 697.15 | 614.98 | 702.20 | 616.00 | **2,630.33** |
| 746 | 736.81 | 547.68 | 568.51 | 679.22 | **2,532.22** |
| 747 | 903.76 | 583.20 | 740.82 | 690.26 | **2,918.04** |
| 748 | 1,783.79 | 1,370.84 | 1,540.52 | 1,511.13 | **6,206.27** |
| 749 | 705.15 | 630.44 | 589.74 | 647.94 | **2,573.28** |
| **OK - OKLAHOMA - Total** | **49,291.23** | **39,438.16** | **43,469.57** | **45,211.90** | **177,410.86** |

**STATE: OR - OREGON**

| REGISTRANT ZIP CODE 3 | 1ST QUARTER | 2ND QUARTER | 3RD QUARTER | 4TH QUARTER | TOTAL GRAMS |
|---|---|---|---|---|---|
| 970 | 13,183.61 | 11,047.40 | 11,139.06 | 11,967.37 | **47,337.44** |
| 971 | 5,481.45 | 4,579.16 | 4,740.88 | 5,080.23 | **19,881.73** |
| 972 | 18,367.28 | 18,013.15 | 17,861.06 | 19,397.29 | **73,638.77** |
| 973 | 11,545.88 | 9,317.17 | 9,642.85 | 10,532.87 | **41,038.77** |
| 974 | 8,258.72 | 7,226.15 | 7,380.78 | 7,770.01 | **30,635.65** |
| 975 | 3,518.36 | 3,061.26 | 3,125.69 | 3,179.06 | **12,884.37** |
| 976 | 662.62 | 547.94 | 567.65 | 650.33 | **2,428.55** |
| 977 | 3,184.56 | 3,165.97 | 3,216.43 | 3,453.11 | **13,020.08** |
| 978 | 1,885.13 | 1,590.35 | 1,613.43 | 1,703.52 | **6,792.43** |
| 979 | 610.20 | 594.58 | 523.38 | 504.07 | **2,232.24** |
| **OR - OREGON - Total** | **66,697.81** | **59,143.12** | **59,811.22** | **64,237.88** | **249,890.03** |

**STATE: PA - PENNSYLVANIA**

| REGISTRANT ZIP CODE 3 | 1ST QUARTER | 2ND QUARTER | 3RD QUARTER | 4TH QUARTER | TOTAL GRAMS |
|---|---|---|---|---|---|
| 150 | 7,472.60 | 6,396.90 | 6,724.14 | 7,163.97 | **27,757.61** |
| 151 | 6,434.89 | 5,427.23 | 5,864.12 | 6,381.92 | **24,108.15** |
| 152 | 12,597.82 | 10,630.88 | 11,300.58 | 12,421.38 | **46,950.65** |
| 153 | 3,394.98 | 3,113.52 | 3,225.40 | 3,641.68 | **13,375.57** |
| 154 | 1,568.78 | 1,473.72 | 1,425.69 | 1,507.54 | **5,975.73** |
| 155 | 1,209.83 | 985.06 | 1,063.24 | 1,065.76 | **4,323.89** |
| 156 | 4,943.92 | 4,028.58 | 4,363.75 | 4,707.78 | **18,044.03** |
| 157 | 1,951.95 | 1,778.63 | 1,912.87 | 2,141.51 | **7,784.96** |
| 158 | 2,163.28 | 1,988.77 | 1,991.49 | 2,062.56 | **8,206.11** |
| 159 | 2,136.82 | 1,961.70 | 1,929.78 | 2,042.04 | **8,070.34** |
| 160 | 3,933.75 | 3,512.43 | 3,602.26 | 4,088.74 | **15,137.19** |
| 161 | 4,024.43 | 3,546.51 | 3,616.76 | 3,981.31 | **15,169.02** |
| 162 | 1,401.77 | 1,197.91 | 1,277.47 | 1,369.57 | **5,246.72** |

| | | | | | |
|---|---|---|---|---|---|
| 163 | 2,310.43 | 1,915.86 | 2,067.63 | 2,143.83 | **8,437.74** |
| 164 | 1,227.72 | 1,044.95 | 1,071.74 | 1,174.95 | **4,519.37** |
| 165 | 4,213.62 | 3,733.25 | 3,703.86 | 4,061.73 | **15,712.46** |
| 166 | 3,373.98 | 3,016.25 | 3,180.96 | 3,201.27 | **12,772.46** |
| 167 | 838.38 | 769.75 | 744.48 | 817.19 | **3,169.80** |
| 168 | 3,234.45 | 2,530.20 | 3,016.60 | 3,106.51 | **11,887.76** |
| 169 | 1,013.93 | 715.47 | 787.29 | 746.76 | **3,263.45** |
| 170 | 14,670.17 | 12,619.73 | 13,411.95 | 13,686.59 | **54,388.44** |
| 171 | 2,802.01 | 2,502.59 | 2,496.92 | 2,689.09 | **10,490.61** |
| 172 | 4,107.04 | 3,314.24 | 3,562.58 | 3,618.85 | **14,602.72** |
| 173 | 6,287.39 | 5,246.92 | 5,604.29 | 5,858.45 | **22,997.05** |
| 174 | 4,389.36 | 3,555.14 | 3,744.72 | 4,010.62 | **15,699.84** |
| 175 | 4,713.70 | 3,720.15 | 4,223.19 | 4,227.88 | **16,884.91** |
| 176 | 4,613.15 | 3,911.16 | 3,896.64 | 4,405.60 | **16,826.55** |
| 177 | 2,982.68 | 2,514.29 | 2,605.34 | 2,870.33 | **10,972.64** |
| 178 | 3,467.82 | 2,799.74 | 3,060.59 | 3,198.77 | **12,526.92** |
| 179 | 1,336.69 | 1,013.98 | 1,160.84 | 1,272.55 | **4,784.05** |
| 180 | 6,320.32 | 5,505.07 | 5,705.25 | 5,997.16 | **23,527.79** |
| 181 | 3,008.61 | 2,294.82 | 2,357.10 | 2,498.08 | **10,158.61** |
| 182 | 1,237.47 | 1,079.44 | 1,046.35 | 1,131.52 | **4,494.78** |
| 183 | 1,695.33 | 1,644.15 | 1,487.47 | 1,575.99 | **6,402.93** |
| 184 | 2,278.35 | 2,099.29 | 2,090.71 | 2,377.03 | **8,845.38** |
| 185 | 2,428.02 | 2,084.58 | 2,188.88 | 2,258.29 | **8,959.76** |
| 186 | 2,067.85 | 1,908.87 | 2,053.71 | 2,111.68 | **8,142.12** |
| 187 | 27,425.23 | 27,071.61 | 26,337.98 | 27,198.66 | **108,033.48** |
| 188 | 1,166.70 | 1,055.64 | 1,063.77 | 1,096.52 | **4,382.63** |
| 189 | 5,940.35 | 5,108.26 | 5,152.59 | 5,927.19 | **22,128.40** |
| 190 | 18,552.47 | 15,752.42 | 15,751.18 | 17,474.23 | **67,530.30** |
| 191 | 13,509.38 | 11,016.50 | 10,760.08 | 11,802.60 | **47,088.56** |
| 193 | 8,903.55 | 7,300.22 | 7,395.91 | 7,937.70 | **31,537.38** |
| 194 | 8,898.31 | 7,611.28 | 7,792.83 | 8,321.59 | **32,624.01** |
| 195 | 2,600.16 | 2,333.15 | 2,356.05 | 2,535.92 | **9,825.28** |
| 196 | 3,506.97 | 3,024.84 | 3,007.26 | 3,218.54 | **12,757.61** |
| **PA - PENNSYLVANIA - Total** | **228,356.43** | **197,855.63** | **203,184.30** | **217,129.45** | **846,525.81** |

**STATE: PR - PUERTO RICO**

| REGISTRANT ZIP CODE 3 | 1ST QUARTER | 2ND QUARTER | 3RD QUARTER | 4TH QUARTER | **TOTAL GRAMS** |
|---|---|---|---|---|---|
| 006 | 3,409.04 | 2,221.68 | 2,771.42 | 2,330.62 | **10,732.75** |
| 007 | 3,845.37 | 2,387.46 | 2,826.26 | 2,969.18 | **12,028.27** |
| 009 | 5,856.44 | 3,807.59 | 4,216.03 | 4,572.50 | **18,452.56** |
| **PR - PUERTO RICO - Total** | **13,110.85** | **8,416.74** | **9,813.70** | **9,872.29** | **41,213.58** |

**STATE: RI - RHODE ISLAND**

| REGISTRANT ZIP CODE 3 | 1ST QUARTER | 2ND QUARTER | 3RD QUARTER | 4TH QUARTER | **TOTAL GRAMS** |
|---|---|---|---|---|---|
| 028 | 13,221.79 | 11,478.14 | 11,355.91 | 11,319.60 | **47,375.45** |
| 029 | 6,110.19 | 4,877.70 | 5,121.97 | 5,099.49 | **21,209.36** |
| **RI - RHODE ISLAND - Total** | **19,331.98** | **16,355.84** | **16,477.89** | **16,419.10** | **68,584.81** |

**STATE: SC - SOUTH CAROLINA**

| REGISTRANT ZIP CODE 3 | 1ST QUARTER | 2ND QUARTER | 3RD QUARTER | 4TH QUARTER | **TOTAL GRAMS** |
|---|---|---|---|---|---|
| 290 | 7,506.30 | 6,113.92 | 5,891.68 | 6,593.47 | **26,105.37** |
| 291 | 5,565.75 | 4,271.24 | 4,530.16 | 4,788.74 | **19,155.90** |
| 292 | 6,779.91 | 4,854.03 | 5,217.69 | 5,820.82 | **22,672.46** |
| 293 | 7,861.25 | 6,354.04 | 6,564.99 | 7,515.31 | **28,295.59** |
| 294 | 19,144.29 | 14,398.64 | 14,947.72 | 17,189.44 | **65,680.09** |
| 295 | 11,878.76 | 9,278.84 | 9,128.76 | 10,403.03 | **40,689.38** |
| 296 | 20,765.94 | 16,938.84 | 17,206.28 | 19,974.36 | **74,885.41** |
| 297 | 7,814.78 | 5,869.67 | 6,092.89 | 6,669.09 | **26,446.44** |
| 298 | 3,482.73 | 2,637.89 | 2,693.83 | 2,974.08 | **11,788.52** |
| 299 | 3,002.32 | 2,651.58 | 2,763.94 | 2,635.49 | **11,053.33** |
| **SC - SOUTH CAROLINA - Total** | **93,802.03** | **73,368.68** | **75,037.95** | **84,563.83** | **326,772.50** |

**STATE: SD - SOUTH DAKOTA**

| REGISTRANT ZIP CODE 3 | 1ST QUARTER | 2ND QUARTER | 3RD QUARTER | 4TH QUARTER | **TOTAL GRAMS** |
|---|---|---|---|---|---|

| | | | | | |
|---|---|---|---|---|---|
| 570 | 2,822.79 | 2,409.97 | 2,463.06 | 2,743.94 | **10,439.76** |
| 571 | 8,249.07 | 6,972.76 | 6,885.68 | 7,039.35 | **29,146.86** |
| 572 | 1,205.81 | 987.13 | 984.93 | 1,082.43 | **4,260.31** |
| 573 | 1,782.57 | 1,457.98 | 1,521.82 | 1,729.51 | **6,491.87** |
| 574 | 1,308.09 | 1,060.54 | 1,101.52 | 1,048.62 | **4,518.77** |
| 575 | 981.81 | 698.56 | 712.96 | 926.63 | **3,319.96** |
| 576 | 205.86 | 222.29 | 192.71 | 217.01 | **837.87** |
| 577 | 2,831.42 | 2,635.56 | 2,726.71 | 2,793.47 | **10,987.16** |
| **SD - SOUTH DAKOTA - Total** | **19,387.42** | **16,444.78** | **16,589.38** | **17,580.97** | **70,002.56** |

**STATE: TN - TENNESSEE**

| REGISTRANT ZIP CODE 3 | 1ST QUARTER | 2ND QUARTER | 3RD QUARTER | 4TH QUARTER | TOTAL GRAMS |
|---|---|---|---|---|---|
| 370 | 13,385.19 | 11,072.01 | 11,915.67 | 12,768.61 | **49,141.49** |
| 371 | 8,724.66 | 6,947.02 | 7,768.49 | 8,667.87 | **32,108.04** |
| 372 | 5,570.99 | 4,783.27 | 4,991.49 | 5,459.30 | **20,805.05** |
| 373 | 8,679.57 | 6,868.43 | 7,661.85 | 8,270.77 | **31,480.62** |
| 374 | 3,740.72 | 2,950.26 | 3,405.21 | 3,618.42 | **13,714.62** |
| 376 | 5,034.83 | 4,054.96 | 4,348.75 | 4,581.58 | **18,020.11** |
| 377 | 4,885.51 | 4,159.67 | 4,411.08 | 4,745.03 | **18,201.28** |
| 378 | 7,041.16 | 6,039.74 | 6,379.10 | 6,952.30 | **26,412.30** |
| 379 | 6,475.35 | 5,262.47 | 5,514.79 | 6,654.03 | **23,906.63** |
| 380 | 6,185.02 | 4,601.91 | 5,067.21 | 5,371.26 | **21,225.40** |
| 381 | 7,495.22 | 5,763.23 | 6,307.57 | 6,784.82 | **26,350.84** |
| 382 | 1,546.77 | 1,151.25 | 1,292.92 | 1,390.56 | **5,381.51** |
| 383 | 3,719.73 | 2,673.24 | 3,128.38 | 3,332.91 | **12,854.27** |
| 384 | 1,677.76 | 1,398.12 | 1,430.56 | 1,615.89 | **6,122.34** |
| 385 | 2,151.62 | 1,877.04 | 1,966.86 | 2,260.09 | **8,255.62** |
| **TN - TENNESSEE - Total** | **86,314.12** | **69,602.62** | **75,589.93** | **82,473.43** | **313,980.10** |

**STATE: TX - TEXAS**

| REGISTRANT ZIP CODE 3 | 1ST QUARTER | 2ND QUARTER | 3RD QUARTER | 4TH QUARTER | TOTAL GRAMS |
|---|---|---|---|---|---|
| 750 | 34,000.12 | 26,038.70 | 28,728.06 | 33,411.01 | **122,177.88** |
| 751 | 8,549.18 | 6,404.48 | 6,693.55 | 7,635.50 | **29,282.71** |
| 752 | 11,792.37 | 9,020.69 | 8,927.11 | 10,766.48 | **40,506.65** |
| 753 | 51.72 | 22.00 | 17.44 | 16.04 | **107.20** |
| 754 | 5,421.53 | 3,893.86 | 4,599.73 | 5,319.06 | **19,234.19** |
| 755 | 1,489.73 | 1,065.97 | 1,268.12 | 1,271.63 | **5,095.45** |
| 756 | 4,405.28 | 3,273.09 | 3,725.22 | 4,232.35 | **15,635.94** |
| 757 | 5,591.89 | 3,986.56 | 4,522.01 | 5,068.81 | **19,169.28** |
| 758 | 1,268.21 | 924.01 | 1,034.90 | 1,118.78 | **4,345.90** |
| 759 | 4,302.37 | 3,282.05 | 3,451.78 | 4,034.39 | **15,070.60** |
| 760 | 17,561.45 | 13,375.16 | 14,990.26 | 17,437.67 | **63,364.53** |
| 761 | 10,934.27 | 8,231.01 | 8,591.74 | 10,053.58 | **37,810.60** |
| 762 | 8,540.28 | 6,575.35 | 7,269.23 | 8,655.41 | **31,040.26** |
| 763 | 3,514.09 | 2,784.32 | 2,971.16 | 3,280.64 | **12,550.20** |
| 764 | 1,339.57 | 991.30 | 1,199.98 | 1,287.61 | **4,818.46** |
| 765 | 10,289.62 | 7,316.47 | 8,493.37 | 8,023.14 | **34,122.59** |
| 766 | 1,380.27 | 1,024.50 | 1,120.31 | 1,260.61 | **4,785.69** |
| 767 | 4,383.86 | 2,958.60 | 3,264.31 | 3,892.66 | **14,499.42** |
| 768 | 1,292.17 | 1,041.87 | 975.83 | 1,064.28 | **4,374.15** |
| 769 | 2,381.98 | 1,796.12 | 1,927.02 | 2,049.46 | **8,154.58** |
| 770 | 24,185.89 | 18,135.98 | 18,526.66 | 22,222.73 | **83,071.25** |
| 772 | 6.75 | 0 | 0 | 3.11 | **9.86** |
| 773 | 18,764.20 | 13,629.36 | 15,135.70 | 18,035.85 | **65,565.12** |
| 774 | 18,625.04 | 13,262.06 | 14,391.26 | 17,372.08 | **63,650.44** |
| 775 | 16,911.07 | 11,955.24 | 12,768.47 | 15,859.49 | **57,494.26** |
| 776 | 3,230.52 | 2,382.40 | 2,621.72 | 2,912.73 | **11,147.37** |
| 777 | 1,478.81 | 1,170.45 | 1,184.00 | 1,284.61 | **5,117.87** |
| 778 | 5,198.49 | 3,712.79 | 4,423.53 | 4,933.61 | **18,268.42** |
| 779 | 2,531.41 | 1,653.23 | 1,992.07 | 2,262.18 | **8,438.89** |
| 780 | 6,708.87 | 5,437.87 | 5,536.31 | 6,491.60 | **24,174.65** |
| 781 | 6,700.45 | 4,992.83 | 5,648.44 | 6,390.84 | **23,732.57** |

| | | | | | |
|---|---|---|---|---|---|
| 782 | 28,383.39 | 20,077.35 | 21,243.70 | 24,676.04 | **94,380.49** |
| 783 | 3,311.35 | 2,222.88 | 2,471.57 | 2,599.26 | **10,605.06** |
| 784 | 6,218.78 | 4,358.90 | 4,556.18 | 5,343.46 | **20,477.32** |
| 785 | 11,093.72 | 7,924.75 | 7,942.97 | 8,666.79 | **35,628.22** |
| 786 | 14,601.01 | 10,709.46 | 12,150.57 | 13,917.80 | **51,378.83** |
| 787 | 14,606.09 | 11,647.07 | 12,629.88 | 14,644.34 | **53,527.39** |
| 788 | 1,380.42 | 894.43 | 991.93 | 1,165.45 | **4,432.24** |
| 789 | 795.87 | 520.71 | 602.47 | 639.82 | **2,558.87** |
| 790 | 1,342.13 | 903.02 | 1,008.41 | 1,109.76 | **4,363.32** |
| 791 | 2,543.61 | 1,902.76 | 2,126.57 | 2,179.56 | **8,752.49** |
| 792 | 333.73 | 221.86 | 214.01 | 231.85 | **1,001.46** |
| 793 | 987.71 | 743.35 | 838.60 | 782.24 | **3,351.89** |
| 794 | 4,753.37 | 3,408.38 | 3,854.11 | 4,451.89 | **16,467.75** |
| 795 | 1,221.52 | 759.47 | 932.31 | 1,013.13 | **3,926.43** |
| 796 | 3,695.94 | 2,694.15 | 2,988.25 | 3,338.60 | **12,716.95** |
| 797 | 4,125.91 | 3,186.81 | 3,374.71 | 3,793.96 | **14,481.39** |
| 798 | 186.07 | 124.40 | 141.83 | 150.69 | **602.99** |
| 799 | 8,111.36 | 5,657.56 | 7,112.26 | 5,588.28 | **26,469.47** |
| **TX - TEXAS - Total** | **350,523.40** | **258,295.68** | **281,179.60** | **321,940.89** | **1,211,939.57** |

**STATE: UT - UTAH**

| REGISTRANT ZIP CODE 3 | 1ST QUARTER | 2ND QUARTER | 3RD QUARTER | 4TH QUARTER | TOTAL GRAMS |
|---|---|---|---|---|---|
| 840 | 26,275.40 | 24,031.61 | 24,895.13 | 26,962.13 | **102,164.27** |
| 841 | 9,234.23 | 8,471.41 | 8,319.70 | 9,242.43 | **35,267.77** |
| 843 | 2,495.74 | 2,276.22 | 2,332.46 | 2,478.02 | **9,582.44** |
| 844 | 3,336.12 | 2,962.47 | 3,133.59 | 3,464.13 | **12,896.31** |
| 845 | 614.22 | 570.00 | 552.46 | 650.31 | **2,387.00** |
| 846 | 5,530.69 | 4,857.43 | 5,062.55 | 5,570.88 | **21,021.56** |
| 847 | 3,119.57 | 2,910.55 | 2,910.54 | 3,381.04 | **12,321.70** |
| **UT - UTAH - Total** | **50,605.97** | **46,079.68** | **47,206.44** | **51,748.95** | **195,641.04** |

**STATE: VA - VIRGINIA**

| REGISTRANT ZIP CODE 3 | 1ST QUARTER | 2ND QUARTER | 3RD QUARTER | 4TH QUARTER | TOTAL GRAMS |
|---|---|---|---|---|---|
| 201 | 13,686.37 | 10,829.69 | 11,106.53 | 12,458.20 | **48,080.79** |
| 220 | 9,681.69 | 7,117.26 | 8,837.33 | 7,766.00 | **33,402.28** |
| 221 | 6,295.86 | 4,630.54 | 5,568.53 | 5,224.94 | **21,719.87** |
| 222 | 3,164.50 | 2,752.43 | 2,862.19 | 2,753.50 | **11,532.62** |
| 223 | 3,997.41 | 3,289.98 | 3,356.91 | 3,705.52 | **14,349.82** |
| 224 | 4,901.00 | 3,802.05 | 3,976.42 | 4,305.87 | **16,985.35** |
| 225 | 2,714.24 | 2,010.17 | 2,195.81 | 2,496.27 | **9,416.48** |
| 226 | 3,250.44 | 2,631.58 | 2,712.38 | 2,888.55 | **11,482.95** |
| 227 | 933.42 | 750.62 | 746.64 | 915.49 | **3,346.18** |
| 228 | 2,061.47 | 1,692.35 | 1,745.69 | 1,886.82 | **7,386.33** |
| 229 | 6,447.40 | 5,190.64 | 5,503.00 | 5,571.04 | **22,712.07** |
| 230 | 3,622.99 | 2,964.07 | 2,955.78 | 3,317.31 | **12,860.14** |
| 231 | 9,099.72 | 7,417.84 | 7,507.62 | 8,669.01 | **32,694.19** |
| 232 | 9,622.22 | 7,963.80 | 7,757.62 | 8,784.92 | **34,128.55** |
| 233 | 5,811.17 | 4,558.85 | 4,343.43 | 5,060.24 | **19,773.68** |
| 234 | 12,265.71 | 9,648.76 | 10,259.64 | 10,548.25 | **42,722.35** |
| 235 | 2,869.57 | 2,543.77 | 2,375.57 | 2,511.30 | **10,300.21** |
| 236 | 7,049.42 | 5,238.52 | 5,700.62 | 5,695.85 | **23,684.41** |
| 237 | 2,229.57 | 2,520.55 | 2,409.13 | 1,755.48 | **8,914.74** |
| 238 | 4,673.25 | 3,646.30 | 3,726.32 | 4,089.07 | **16,134.94** |
| 239 | 1,241.31 | 962.28 | 1,044.72 | 1,113.86 | **4,362.17** |
| 240 | 5,847.42 | 4,638.20 | 4,727.64 | 5,027.36 | **20,240.62** |
| 241 | 3,470.27 | 2,923.11 | 3,172.23 | 3,057.44 | **12,623.05** |
| 242 | 2,177.05 | 1,895.67 | 1,988.40 | 2,100.89 | **8,162.02** |
| 243 | 1,934.97 | 1,395.14 | 1,582.41 | 1,683.89 | **6,596.41** |
| 244 | 2,584.62 | 2,259.60 | 2,175.75 | 2,410.93 | **9,430.90** |
| 245 | 5,873.82 | 4,572.41 | 4,977.99 | 5,343.46 | **20,767.68** |
| 246 | 947.99 | 804.20 | 813.85 | 907.70 | **3,473.74** |
| **VA - VIRGINIA - Total** | **138,454.85** | **110,650.38** | **116,130.16** | **122,049.17** | **487,284.55** |

| REGISTRANT ZIP CODE 3 | 1ST QUARTER | 2ND QUARTER | 3RD QUARTER | 4TH QUARTER | TOTAL GRAMS |
|---|---|---|---|---|---|
| **STATE: VI - VIRGIN ISLANDS** | | | | | |
| 008 | 126.17 | 78.47 | 100.47 | 85.25 | **390.37** |
| **VI - VIRGIN ISLANDS - Total** | **126.17** | **78.47** | **100.47** | **85.25** | **390.37** |
| **STATE: VT - VERMONT** | | | | | |
| REGISTRANT ZIP CODE 3 | 1ST QUARTER | 2ND QUARTER | 3RD QUARTER | 4TH QUARTER | TOTAL GRAMS |
| 050 | 1,387.33 | 1,387.62 | 1,331.87 | 1,492.60 | **5,599.41** |
| 051 | 806.54 | 838.95 | 859.11 | 854.23 | **3,358.84** |
| 052 | 1,204.94 | 1,152.27 | 1,049.20 | 1,156.77 | **4,563.18** |
| 053 | 1,196.72 | 1,067.07 | 981.39 | 1,098.19 | **4,343.37** |
| 054 | 7,141.90 | 5,870.64 | 6,376.12 | 6,768.43 | **26,157.09** |
| 056 | 2,958.94 | 2,738.22 | 2,696.75 | 2,809.70 | **11,203.60** |
| 057 | 2,406.16 | 2,324.94 | 2,251.17 | 2,473.44 | **9,455.72** |
| 058 | 2,341.55 | 2,276.36 | 2,397.01 | 2,655.02 | **9,669.94** |
| **VT - VERMONT - Total** | **19,444.07** | **17,656.08** | **17,942.62** | **19,308.38** | **74,351.15** |
| **STATE: WA - WASHINGTON** | | | | | |
| REGISTRANT ZIP CODE 3 | 1ST QUARTER | 2ND QUARTER | 3RD QUARTER | 4TH QUARTER | TOTAL GRAMS |
| 980 | 26,295.71 | 24,877.60 | 24,827.69 | 27,512.91 | **103,513.91** |
| 981 | 14,729.37 | 12,763.18 | 12,811.58 | 13,732.48 | **54,036.60** |
| 982 | 15,847.50 | 14,258.37 | 14,055.30 | 14,887.02 | **59,048.19** |
| 983 | 13,162.71 | 11,143.06 | 11,440.38 | 11,758.36 | **47,504.51** |
| 984 | 7,761.23 | 6,464.32 | 6,350.46 | 6,701.32 | **27,277.33** |
| 985 | 7,736.69 | 6,478.31 | 6,704.73 | 7,225.04 | **28,144.76** |
| 986 | 9,418.98 | 7,517.97 | 7,713.04 | 8,466.39 | **33,116.38** |
| 988 | 2,561.19 | 2,500.88 | 2,322.84 | 2,447.48 | **9,832.40** |
| 989 | 3,473.11 | 3,029.29 | 3,022.15 | 3,112.99 | **12,637.55** |
| 990 | 1,718.11 | 1,659.81 | 1,587.39 | 1,805.68 | **6,770.99** |
| 991 | 1,294.06 | 1,043.95 | 1,059.90 | 1,258.24 | **4,656.15** |
| 992 | 8,748.00 | 7,332.98 | 7,667.61 | 8,044.27 | **31,792.86** |
| 993 | 5,207.36 | 4,482.06 | 4,322.24 | 4,769.28 | **18,780.95** |
| 994 | 605.40 | 507.17 | 504.95 | 564.54 | **2,182.06** |
| **WA - WASHINGTON - Total** | **118,559.42** | **104,058.93** | **104,390.27** | **112,286.02** | **439,294.64** |
| **STATE: WI - WISCONSIN** | | | | | |
| REGISTRANT ZIP CODE 3 | 1ST QUARTER | 2ND QUARTER | 3RD QUARTER | 4TH QUARTER | TOTAL GRAMS |
| 530 | 13,599.82 | 11,862.99 | 12,123.36 | 13,482.10 | **51,068.26** |
| 531 | 11,743.20 | 10,136.38 | 10,711.17 | 11,445.49 | **44,036.24** |
| 532 | 11,799.37 | 9,666.80 | 9,795.07 | 10,757.91 | **42,019.15** |
| 534 | 2,464.71 | 2,028.86 | 1,982.25 | 2,220.96 | **8,696.79** |
| 535 | 11,002.81 | 9,633.78 | 9,801.60 | 10,781.29 | **41,219.47** |
| 537 | 6,579.99 | 5,412.16 | 5,695.57 | 6,243.07 | **23,930.79** |
| 538 | 1,018.12 | 897.43 | 945.85 | 1,104.58 | **3,965.98** |
| 539 | 3,706.37 | 3,343.15 | 3,224.48 | 3,536.55 | **13,810.56** |
| 540 | 2,397.42 | 2,042.32 | 2,234.63 | 2,419.80 | **9,094.18** |
| 541 | 4,157.46 | 3,737.76 | 3,908.00 | 4,129.43 | **15,932.64** |
| 542 | 2,049.92 | 1,927.80 | 1,877.13 | 2,086.35 | **7,941.20** |
| 543 | 4,064.93 | 3,577.91 | 3,524.24 | 3,992.65 | **15,159.73** |
| 544 | 7,429.64 | 6,608.84 | 6,455.75 | 6,871.18 | **27,365.40** |
| 545 | 1,477.71 | 1,219.25 | 1,341.08 | 1,287.65 | **5,325.68** |
| 546 | 5,046.30 | 4,267.15 | 4,491.53 | 4,562.11 | **18,367.09** |
| 547 | 5,999.70 | 4,973.41 | 5,287.96 | 5,707.07 | **21,968.14** |
| 548 | 3,151.88 | 2,656.59 | 2,904.89 | 2,851.82 | **11,565.18** |
| 549 | 10,723.67 | 9,438.23 | 9,745.12 | 10,407.51 | **40,314.53** |
| **WI - WISCONSIN - Total** | **108,413.02** | **93,430.83** | **96,049.66** | **103,887.51** | **401,781.02** |
| **STATE: WV - WEST VIRGINIA** | | | | | |
| REGISTRANT ZIP CODE 3 | 1ST QUARTER | 2ND QUARTER | 3RD QUARTER | 4TH QUARTER | TOTAL GRAMS |
| 247 | 908.92 | 746.72 | 747.97 | 774.83 | **3,178.44** |
| 248 | 357.42 | 253.55 | 261.16 | 273.30 | **1,145.44** |
| 249 | 621.96 | 628.09 | 534.85 | 631.29 | **2,416.19** |
| 250 | 1,186.01 | 921.08 | 925.04 | 1,129.81 | **4,161.93** |
| 251 | 968.18 | 953.37 | 884.16 | 954.23 | **3,759.94** |

| | | | | | |
|---|---|---|---|---|---|
| 252 | 676.87 | 554.95 | 514.00 | 587.40 | **2,333.22** |
| 253 | 2,731.04 | 2,133.10 | 2,202.69 | 2,292.95 | **9,359.78** |
| 254 | 2,757.85 | 2,303.06 | 2,244.78 | 2,424.11 | **9,729.80** |
| 255 | 2,905.21 | 2,300.67 | 2,318.17 | 2,567.53 | **10,091.58** |
| 256 | 729.32 | 638.14 | 639.99 | 712.00 | **2,719.45** |
| 257 | 2,436.46 | 2,122.40 | 2,029.42 | 2,082.26 | **8,670.54** |
| 258 | 1,731.96 | 1,482.73 | 1,627.83 | 1,577.95 | **6,420.47** |
| 259 | 602.21 | 551.41 | 538.09 | 551.11 | **2,242.83** |
| 260 | 2,008.68 | 1,642.63 | 1,547.48 | 1,822.54 | **7,021.33** |
| 261 | 1,977.49 | 1,648.82 | 1,586.09 | 1,545.37 | **6,757.77** |
| 262 | 875.16 | 789.31 | 749.23 | 713.67 | **3,127.36** |
| 263 | 1,550.24 | 1,296.76 | 1,292.99 | 1,366.83 | **5,506.82** |
| 264 | 470.39 | 447.92 | 449.11 | 477.14 | **1,844.57** |
| 265 | 3,111.07 | 2,599.15 | 2,645.04 | 2,887.98 | **11,243.25** |
| 266 | 515.67 | 342.55 | 413.20 | 399.72 | **1,671.14** |
| 267 | 798.57 | 720.24 | 612.40 | 675.97 | **2,807.18** |
| 268 | 560.35 | 470.51 | 378.25 | 509.43 | **1,918.54** |
| **WV - WEST VIRGINIA - Total** | **30,481.01** | **25,547.18** | **25,141.93** | **26,957.44** | **108,127.56** |
| **STATE: WY - WYOMING** | | | | | |
| REGISTRANT ZIP CODE 3 | 1ST QUARTER | 2ND QUARTER | 3RD QUARTER | 4TH QUARTER | **TOTAL GRAMS** |
| 820 | 2,180.21 | 1,810.55 | 1,980.66 | 1,989.25 | **7,960.66** |
| 822 | 243.06 | 226.25 | 175.49 | 194.81 | **839.60** |
| 823 | 102.74 | 107.49 | 84.40 | 119.86 | **414.50** |
| 824 | 761.10 | 692.99 | 766.70 | 810.01 | **3,030.81** |
| 825 | 454.75 | 335.31 | 385.72 | 393.32 | **1,569.10** |
| 826 | 1,336.53 | 1,288.08 | 1,271.93 | 1,315.32 | **5,211.86** |
| 827 | 736.22 | 611.21 | 698.19 | 744.95 | **2,790.57** |
| 828 | 533.76 | 459.21 | 388.20 | 500.36 | **1,881.53** |
| 829 | 803.74 | 878.62 | 750.30 | 770.84 | **3,203.50** |
| 830 | 467.90 | 398.94 | 407.36 | 450.26 | **1,724.46** |
| 831 | 214.73 | 181.78 | 191.08 | 221.28 | **808.87** |
| **WY - WYOMING - Total** | **7,834.74** | **6,990.44** | **7,100.03** | **7,510.26** | **29,435.46** |
| **DRUG: 9050 - CODEINE** | | | | | |
| **STATE: AK - ALASKA** | | | | | |
| REGISTRANT ZIP CODE 3 | 1ST QUARTER | 2ND QUARTER | 3RD QUARTER | 4TH QUARTER | **TOTAL GRAMS** |
| 995 | 1,796.58 | 1,516.86 | 1,827.30 | 1,673.88 | **6,814.62** |
| 996 | 791.53 | 768.52 | 935.92 | 765.20 | **3,261.17** |
| 997 | 1,728.10 | 1,515.53 | 1,636.67 | 1,475.42 | **6,355.73** |
| 998 | 282.52 | 218.98 | 253.78 | 244.76 | **1,000.05** |
| 999 | 209.40 | 159.31 | 221.74 | 237.95 | **828.40** |
| **AK - ALASKA - Total** | **4,808.13** | **4,179.20** | **4,875.41** | **4,397.22** | **18,259.96** |
| **STATE: AL - ALABAMA** | | | | | |
| REGISTRANT ZIP CODE 3 | 1ST QUARTER | 2ND QUARTER | 3RD QUARTER | 4TH QUARTER | **TOTAL GRAMS** |
| 350 | 4,804.50 | 4,932.99 | 4,668.68 | 4,551.14 | **18,957.32** |
| 351 | 3,252.79 | 2,964.61 | 2,753.07 | 2,952.36 | **11,922.83** |
| 352 | 4,271.65 | 4,103.43 | 3,350.49 | 4,239.29 | **15,964.85** |
| 354 | 1,575.80 | 1,424.12 | 1,327.33 | 1,359.68 | **5,686.94** |
| 355 | 1,753.32 | 1,883.55 | 2,075.45 | 2,289.42 | **8,001.74** |
| 356 | 5,006.22 | 5,054.78 | 5,174.82 | 5,569.53 | **20,805.35** |
| 357 | 548.74 | 566.23 | 700.97 | 714.42 | **2,530.36** |
| 358 | 783.91 | 774.55 | 698.89 | 1,000.03 | **3,257.39** |
| 359 | 1,927.82 | 1,631.04 | 1,657.82 | 1,595.03 | **6,811.71** |
| 360 | 2,693.69 | 2,673.44 | 2,971.04 | 2,988.47 | **11,326.64** |
| 361 | 1,823.97 | 1,767.16 | 1,598.81 | 1,619.64 | **6,809.58** |
| 362 | 1,224.83 | 1,442.45 | 1,416.12 | 1,158.46 | **5,241.86** |
| 363 | 1,531.35 | 1,324.21 | 1,471.64 | 1,559.06 | **5,886.26** |
| 364 | 1,311.69 | 1,392.36 | 1,317.40 | 1,503.23 | **5,524.69** |
| 365 | 3,392.70 | 2,949.48 | 2,689.86 | 3,026.81 | **12,058.85** |
| 366 | 3,003.17 | 2,879.94 | 2,352.12 | 2,868.46 | **11,103.70** |
| 367 | 2,017.82 | 1,715.04 | 1,630.31 | 1,852.25 | **7,215.42** |

| | | | | | |
|---|---|---|---|---|---|
| 368 | 1,735.80 | 1,730.62 | 1,748.01 | 1,866.52 | **7,080.96** |
| 369 | 429.86 | 430.97 | 534.84 | 600.04 | **1,995.72** |
| **AL - ALABAMA - Total** | **43,089.65** | **41,641.00** | **40,137.68** | **43,313.83** | **168,182.16** |

**STATE: AR - ARKANSAS**

| REGISTRANT ZIP CODE 3 | 1ST QUARTER | 2ND QUARTER | 3RD QUARTER | 4TH QUARTER | TOTAL GRAMS |
|---|---|---|---|---|---|
| 716 | 2,839.04 | 2,512.88 | 2,890.86 | 2,701.29 | **10,944.07** |
| 717 | 2,700.18 | 2,613.05 | 2,453.37 | 2,603.29 | **10,369.90** |
| 718 | 4,604.50 | 4,167.14 | 4,267.99 | 4,233.99 | **17,273.62** |
| 719 | 3,256.77 | 3,235.94 | 3,382.74 | 3,577.59 | **13,453.04** |
| 720 | 3,986.77 | 4,223.75 | 3,898.16 | 3,686.21 | **15,794.89** |
| 721 | 4,593.71 | 4,233.27 | 4,501.69 | 4,427.46 | **17,756.12** |
| 722 | 2,166.83 | 1,888.10 | 2,034.39 | 1,725.43 | **7,814.75** |
| 723 | 5,198.82 | 4,630.56 | 4,946.23 | 4,793.25 | **19,568.87** |
| 724 | 5,868.40 | 5,785.29 | 5,416.64 | 5,394.01 | **22,464.33** |
| 725 | 3,021.05 | 3,065.59 | 2,826.72 | 2,911.81 | **11,825.17** |
| 726 | 1,686.49 | 1,771.76 | 2,001.79 | 1,709.97 | **7,170.01** |
| 727 | 2,815.42 | 3,063.91 | 2,895.38 | 2,961.99 | **11,736.70** |
| 728 | 2,243.25 | 1,981.54 | 2,056.86 | 2,004.56 | **8,286.21** |
| 729 | 3,127.10 | 3,029.49 | 2,949.00 | 2,778.46 | **11,884.05** |
| **AR - ARKANSAS - Total** | **48,108.33** | **46,202.27** | **46,521.82** | **45,509.30** | **186,341.73** |

**STATE: AS - AMERICAN SAMOA**

| REGISTRANT ZIP CODE 3 | 1ST QUARTER | 2ND QUARTER | 3RD QUARTER | 4TH QUARTER | TOTAL GRAMS |
|---|---|---|---|---|---|
| 967 | 360.25 | 320.46 | 395.61 | 278.47 | **1,354.79** |
| **AS - AMERICAN SAMOA - Total** | **360.25** | **320.46** | **395.61** | **278.47** | **1,354.79** |

**STATE: AZ - ARIZONA**

| REGISTRANT ZIP CODE 3 | 1ST QUARTER | 2ND QUARTER | 3RD QUARTER | 4TH QUARTER | TOTAL GRAMS |
|---|---|---|---|---|---|
| 850 | 13,354.42 | 12,587.11 | 13,097.24 | 12,441.40 | **51,480.16** |
| 851 | 2,132.49 | 1,935.22 | 1,812.75 | 1,968.34 | **7,848.80** |
| 852 | 9,631.40 | 8,601.61 | 8,891.17 | 8,780.90 | **35,905.07** |
| 853 | 8,103.54 | 6,759.01 | 6,757.62 | 6,959.59 | **28,579.76** |
| 855 | 793.17 | 723.59 | 755.33 | 722.62 | **2,994.71** |
| 856 | 1,649.61 | 1,532.02 | 1,574.44 | 1,510.52 | **6,266.58** |
| 857 | 6,503.70 | 5,796.84 | 6,392.66 | 6,053.60 | **24,746.80** |
| 859 | 411.74 | 459.49 | 572.69 | 613.80 | **2,057.72** |
| 860 | 655.89 | 695.09 | 627.31 | 490.76 | **2,469.05** |
| 863 | 1,410.80 | 1,452.92 | 1,633.63 | 1,513.54 | **6,010.89** |
| 864 | 2,021.75 | 1,925.87 | 1,990.87 | 2,060.54 | **7,999.03** |
| 865 | 546.62 | 436.30 | 509.23 | 553.44 | **2,045.59** |
| **AZ - ARIZONA - Total** | **47,215.13** | **42,905.06** | **44,614.94** | **43,669.04** | **178,404.18** |

**STATE: CA - CALIFORNIA**

| REGISTRANT ZIP CODE 3 | 1ST QUARTER | 2ND QUARTER | 3RD QUARTER | 4TH QUARTER | TOTAL GRAMS |
|---|---|---|---|---|---|
| 900 | 16,741.93 | 16,193.38 | 16,745.99 | 16,859.26 | **66,540.56** |
| 902 | 16,985.73 | 18,368.81 | 17,915.35 | 17,245.51 | **70,515.41** |
| 903 | 2,211.21 | 2,290.77 | 2,318.39 | 2,110.87 | **8,931.24** |
| 904 | 838.49 | 876.48 | 843.54 | 845.72 | **3,404.24** |
| 905 | 1,529.09 | 1,390.34 | 1,612.17 | 1,603.51 | **6,135.10** |
| 906 | 5,046.54 | 4,492.13 | 4,975.49 | 5,247.71 | **19,761.88** |
| 907 | 6,145.69 | 8,157.97 | 5,995.04 | 5,763.16 | **26,061.86** |
| 908 | 4,370.93 | 4,348.51 | 4,424.98 | 4,268.91 | **17,413.34** |
| 910 | 2,171.46 | 2,222.49 | 2,158.16 | 2,146.19 | **8,698.31** |
| 911 | 1,510.19 | 1,384.90 | 1,384.18 | 1,470.47 | **5,749.74** |
| 912 | 2,346.27 | 1,927.01 | 2,101.16 | 1,936.37 | **8,310.81** |
| 913 | 9,746.05 | 9,112.85 | 9,955.37 | 10,082.66 | **38,896.93** |
| 914 | 3,252.11 | 2,522.44 | 2,647.90 | 2,591.98 | **11,014.43** |
| 915 | 1,016.83 | 1,133.22 | 1,101.57 | 1,116.54 | **4,368.16** |
| 916 | 1,245.94 | 1,030.53 | 1,087.43 | 1,129.01 | **4,492.91** |
| 917 | 14,733.93 | 12,627.73 | 13,932.67 | 13,121.96 | **54,416.30** |
| 918 | 686.24 | 544.79 | 582.36 | 657.50 | **2,470.89** |
| 919 | 2,777.24 | 2,899.73 | 2,790.77 | 2,903.21 | **11,370.96** |
| 920 | 10,902.82 | 10,620.99 | 9,724.31 | 9,596.95 | **40,845.07** |

| | | | | | |
|---|---|---|---|---|---|
| 921 | 7,184.23 | 5,927.93 | 6,142.82 | 6,160.72 | **25,415.69** |
| 922 | 7,815.25 | 6,716.51 | 7,160.18 | 7,176.45 | **28,868.39** |
| 923 | 12,750.99 | 13,812.35 | 12,750.91 | 12,863.17 | **52,177.43** |
| 924 | 2,143.80 | 2,397.96 | 1,989.09 | 2,066.44 | **8,597.29** |
| 925 | 12,630.15 | 12,217.23 | 13,001.90 | 12,554.57 | **50,403.85** |
| 926 | 8,192.02 | 7,585.75 | 7,698.19 | 7,953.47 | **31,429.43** |
| 927 | 2,197.11 | 2,447.25 | 2,440.12 | 2,443.28 | **9,527.77** |
| 928 | 8,498.95 | 8,000.71 | 8,392.79 | 8,222.12 | **33,114.57** |
| 930 | 4,788.82 | 4,471.57 | 5,089.85 | 4,836.01 | **19,186.25** |
| 931 | 1,250.33 | 890.84 | 1,117.92 | 1,048.11 | **4,307.21** |
| 932 | 6,782.12 | 7,849.90 | 7,027.99 | 6,614.36 | **28,274.37** |
| 933 | 4,665.27 | 4,410.23 | 4,589.39 | 4,767.91 | **18,432.80** |
| 934 | 3,897.85 | 3,941.82 | 3,565.05 | 3,745.19 | **15,149.91** |
| 935 | 7,090.63 | 6,240.29 | 6,641.17 | 6,126.94 | **26,099.02** |
| 936 | 5,497.16 | 5,248.05 | 5,464.63 | 5,127.28 | **21,337.11** |
| 937 | 7,062.35 | 8,272.16 | 6,734.54 | 6,770.14 | **28,839.19** |
| 939 | 3,295.18 | 3,247.96 | 3,171.30 | 3,481.18 | **13,195.63** |
| 940 | 3,528.22 | 3,151.92 | 3,553.12 | 3,310.95 | **13,544.21** |
| 941 | 5,518.25 | 5,010.58 | 4,903.91 | 4,914.60 | **20,347.34** |
| 943 | 445.05 | 498.85 | 444.27 | 387.21 | **1,775.39** |
| 944 | 664.72 | 620.92 | 597.35 | 710.39 | **2,593.38** |
| 945 | 21,642.70 | 23,084.08 | 22,710.60 | 23,509.15 | **90,946.53** |
| 946 | 2,610.02 | 2,741.35 | 2,412.41 | 2,398.41 | **10,162.18** |
| 947 | 702.60 | 690.80 | 778.66 | 623.88 | **2,795.94** |
| 948 | 1,082.39 | 704.94 | 942.97 | 875.78 | **3,606.09** |
| 949 | 2,435.37 | 2,034.61 | 2,260.44 | 2,237.93 | **8,968.34** |
| 950 | 4,149.36 | 3,631.51 | 4,266.88 | 3,995.14 | **16,042.88** |
| 951 | 4,000.11 | 3,772.64 | 4,305.03 | 4,346.46 | **16,424.25** |
| 952 | 5,916.39 | 5,902.05 | 5,861.20 | 5,897.13 | **23,576.77** |
| 953 | 11,707.89 | 10,305.59 | 11,206.59 | 11,366.00 | **44,586.07** |
| 954 | 3,331.54 | 3,087.33 | 3,209.76 | 3,161.84 | **12,790.47** |
| 955 | 2,156.52 | 1,875.55 | 2,032.85 | 1,897.45 | **7,962.37** |
| 956 | 9,747.64 | 8,961.57 | 9,237.96 | 9,287.53 | **37,234.69** |
| 957 | 1,901.14 | 1,753.90 | 2,011.56 | 1,810.35 | **7,476.94** |
| 958 | 5,196.05 | 4,422.47 | 5,101.60 | 5,023.87 | **19,743.99** |
| 959 | 5,192.41 | 4,938.22 | 4,967.34 | 5,149.55 | **20,247.53** |
| 960 | 3,103.48 | 2,982.46 | 2,871.25 | 3,016.28 | **11,973.46** |
| 961 | 782.56 | 738.17 | 669.88 | 623.43 | **2,814.04** |
| **CA - CALIFORNIA - Total** | **305,815.29** | **296,733.11** | **299,620.29** | **297,198.18** | **1,199,366.88** |
| **STATE: CO - COLORADO** | | | | | |
| REGISTRANT ZIP CODE 3 | 1ST QUARTER | 2ND QUARTER | 3RD QUARTER | 4TH QUARTER | **TOTAL GRAMS** |
| 800 | 2,405.79 | 2,300.95 | 2,350.19 | 2,357.13 | **9,414.06** |
| 801 | 1,827.62 | 1,811.61 | 1,866.47 | 1,764.73 | **7,270.42** |
| 802 | 2,700.63 | 2,905.16 | 2,455.33 | 2,418.90 | **10,480.02** |
| 803 | 266.66 | 215.27 | 212.13 | 209.01 | **903.07** |
| 804 | 308.44 | 316.92 | 374.86 | 370.05 | **1,370.26** |
| 805 | 1,452.88 | 1,299.04 | 1,643.47 | 1,650.05 | **6,045.44** |
| 806 | 676.25 | 560.47 | 592.49 | 669.10 | **2,498.31** |
| 807 | 352.51 | 308.49 | 342.75 | 260.97 | **1,264.72** |
| 808 | 393.94 | 402.97 | 513.11 | 394.68 | **1,704.70** |
| 809 | 1,781.76 | 1,506.33 | 1,524.29 | 1,797.02 | **6,609.40** |
| 810 | 1,360.08 | 1,253.93 | 1,467.47 | 1,463.14 | **5,544.62** |
| 811 | 287.50 | 270.92 | 306.10 | 226.16 | **1,090.67** |
| 812 | 453.07 | 258.58 | 423.60 | 266.12 | **1,401.37** |
| 813 | 190.25 | 130.38 | 230.94 | 157.46 | **709.04** |
| 814 | 301.68 | 370.41 | 267.25 | 269.63 | **1,208.97** |
| 815 | 569.46 | 552.84 | 514.95 | 521.56 | **2,158.81** |
| 816 | 240.69 | 283.35 | 233.82 | 287.66 | **1,045.52** |
| **CO - COLORADO - Total** | **15,569.21** | **14,747.60** | **15,319.22** | **15,083.39** | **60,719.42** |

**STATE: CT - CONNECTICUT**

| REGISTRANT ZIP CODE 3 | 1ST QUARTER | 2ND QUARTER | 3RD QUARTER | 4TH QUARTER | TOTAL GRAMS |
|---|---|---|---|---|---|
| 060 | 3,070.33 | 3,020.08 | 2,903.47 | 2,940.54 | **11,934.42** |
| 061 | 1,103.69 | 732.45 | 636.51 | 880.87 | **3,353.52** |
| 062 | 578.67 | 517.35 | 572.31 | 553.34 | **2,221.67** |
| 063 | 1,167.21 | 1,118.91 | 1,166.53 | 1,132.77 | **4,585.42** |
| 064 | 3,348.42 | 3,496.68 | 3,338.42 | 3,698.36 | **13,881.87** |
| 065 | 1,143.70 | 858.87 | 904.64 | 1,025.67 | **3,932.88** |
| 066 | 945.31 | 1,011.32 | 1,063.67 | 832.54 | **3,852.84** |
| 067 | 1,589.88 | 1,568.99 | 1,726.18 | 1,524.89 | **6,409.94** |
| 068 | 1,802.18 | 1,772.28 | 1,804.59 | 1,613.95 | **6,993.01** |
| 069 | 433.36 | 451.32 | 382.06 | 387.95 | **1,654.69** |
| **CT - CONNECTICUT - Total** | **15,182.75** | **14,548.25** | **14,498.38** | **14,590.89** | **58,820.26** |

**STATE: DC - DISTRICT OF COLUMBIA**

| REGISTRANT ZIP CODE 3 | 1ST QUARTER | 2ND QUARTER | 3RD QUARTER | 4TH QUARTER | TOTAL GRAMS |
|---|---|---|---|---|---|
| 200 | 7,152.27 | 6,271.63 | 6,171.04 | 6,686.00 | **26,280.94** |
| 203 | 0 | 0 | 2.21 | 4.78 | **6.99** |
| 204 | 75.51 | 98.71 | 95.40 | 75.14 | **344.76** |
| **DC - DISTRICT OF COLUMBIA - Total** | **7,227.78** | **6,370.34** | **6,268.65** | **6,765.93** | **26,632.70** |

**STATE: DE - DELAWARE**

| REGISTRANT ZIP CODE 3 | 1ST QUARTER | 2ND QUARTER | 3RD QUARTER | 4TH QUARTER | TOTAL GRAMS |
|---|---|---|---|---|---|
| 197 | 2,275.41 | 2,130.42 | 2,188.28 | 2,367.46 | **8,961.57** |
| 198 | 1,572.07 | 1,483.38 | 1,513.58 | 1,485.02 | **6,054.05** |
| 199 | 2,331.62 | 2,207.85 | 2,614.56 | 2,456.18 | **9,610.21** |
| **DE - DELAWARE - Total** | **6,179.10** | **5,821.66** | **6,316.42** | **6,308.66** | **24,625.84** |

**STATE: FL - FLORIDA**

| REGISTRANT ZIP CODE 3 | 1ST QUARTER | 2ND QUARTER | 3RD QUARTER | 4TH QUARTER | TOTAL GRAMS |
|---|---|---|---|---|---|
| 320 | 4,037.74 | 3,788.27 | 4,082.00 | 4,115.05 | **16,023.06** |
| 321 | 4,534.89 | 3,919.77 | 3,792.62 | 4,077.87 | **16,325.15** |
| 322 | 4,470.50 | 3,599.93 | 3,993.45 | 4,400.20 | **16,464.08** |
| 323 | 2,445.10 | 2,439.02 | 2,112.48 | 2,820.26 | **9,816.85** |
| 324 | 2,883.19 | 3,071.34 | 3,074.32 | 3,715.13 | **12,743.97** |
| 325 | 3,604.50 | 2,931.88 | 2,898.25 | 3,462.79 | **12,897.42** |
| 326 | 2,233.41 | 2,062.95 | 2,187.92 | 2,361.04 | **8,845.33** |
| 327 | 5,882.74 | 5,188.55 | 5,638.24 | 5,893.56 | **22,603.09** |
| 328 | 4,088.46 | 3,151.47 | 3,852.91 | 3,860.51 | **14,953.36** |
| 329 | 3,904.94 | 3,977.04 | 3,709.34 | 3,857.24 | **15,448.56** |
| 330 | 9,201.72 | 7,890.58 | 8,842.01 | 8,565.13 | **34,499.44** |
| 331 | 10,825.40 | 8,706.64 | 10,225.32 | 10,348.19 | **40,105.55** |
| 333 | 4,924.60 | 4,470.19 | 4,642.08 | 4,406.30 | **18,443.17** |
| 334 | 9,155.99 | 8,576.95 | 8,481.25 | 8,471.15 | **34,685.35** |
| 335 | 6,058.53 | 5,583.94 | 5,968.64 | 6,083.12 | **23,694.23** |
| 336 | 6,022.30 | 5,895.93 | 5,845.97 | 6,205.64 | **23,969.83** |
| 337 | 6,758.93 | 6,477.44 | 6,176.45 | 7,201.11 | **26,613.92** |
| 338 | 6,340.04 | 6,122.15 | 6,216.91 | 6,123.51 | **24,802.60** |
| 339 | 6,313.15 | 5,464.88 | 5,713.65 | 5,979.54 | **23,471.22** |
| 341 | 2,023.32 | 1,661.54 | 1,354.07 | 1,724.31 | **6,763.24** |
| 342 | 5,316.96 | 4,196.26 | 4,332.47 | 4,764.78 | **18,610.48** |
| 344 | 6,032.03 | 6,105.57 | 6,206.21 | 6,812.54 | **25,156.35** |
| 346 | 6,150.14 | 5,529.36 | 6,247.37 | 6,058.72 | **23,985.58** |
| 347 | 3,785.15 | 3,263.90 | 3,487.81 | 3,610.36 | **14,147.22** |
| 349 | 2,705.63 | 2,508.53 | 2,734.35 | 2,937.99 | **10,886.50** |
| **FL - FLORIDA - Total** | **129,699.36** | **116,584.06** | **121,816.09** | **127,856.02** | **495,955.54** |

**STATE: GA - GEORGIA**

| REGISTRANT ZIP CODE 3 | 1ST QUARTER | 2ND QUARTER | 3RD QUARTER | 4TH QUARTER | TOTAL GRAMS |
|---|---|---|---|---|---|
| 300 | 10,538.02 | 10,345.75 | 10,259.42 | 10,981.35 | **42,124.54** |
| 301 | 7,551.55 | 7,205.97 | 7,054.27 | 7,721.91 | **29,533.70** |
| 302 | 8,498.95 | 8,300.06 | 8,694.64 | 8,834.19 | **34,327.84** |
| 303 | 5,019.96 | 3,947.85 | 4,384.84 | 4,815.99 | **18,168.64** |
| 304 | 2,872.22 | 2,528.98 | 3,033.40 | 3,113.21 | **11,547.80** |
| 305 | 5,313.61 | 5,277.32 | 5,202.24 | 5,408.47 | **21,201.64** |

| | | | | | |
|---|---|---|---|---|---|
| 306 | 4,855.33 | 4,376.91 | 4,347.52 | 4,523.43 | **18,103.19** |
| 307 | 1,755.92 | 1,631.05 | 1,572.58 | 1,730.51 | **6,690.06** |
| 308 | 2,271.79 | 2,536.64 | 2,420.24 | 2,301.08 | **9,529.74** |
| 309 | 2,149.93 | 2,020.57 | 2,409.18 | 2,157.80 | **8,737.48** |
| 310 | 5,521.73 | 5,211.90 | 5,243.69 | 5,807.25 | **21,784.56** |
| 312 | 1,512.17 | 1,613.56 | 1,713.71 | 1,650.20 | **6,489.64** |
| 313 | 1,143.47 | 1,178.65 | 1,150.45 | 1,320.09 | **4,792.65** |
| 314 | 1,125.60 | 1,063.94 | 1,065.71 | 974.17 | **4,229.43** |
| 315 | 4,932.20 | 5,031.55 | 4,655.76 | 5,057.05 | **19,676.56** |
| 316 | 1,820.18 | 1,559.46 | 1,687.57 | 1,948.29 | **7,015.50** |
| 317 | 5,425.77 | 4,938.68 | 5,197.35 | 5,464.89 | **21,026.69** |
| 318 | 419.92 | 393.40 | 461.91 | 366.88 | **1,642.10** |
| 319 | 2,213.35 | 2,034.69 | 2,383.77 | 2,381.67 | **9,013.48** |
| 398 | 1,834.38 | 1,836.59 | 1,661.25 | 1,386.28 | **6,718.51** |
| **GA - GEORGIA - Total** | **76,776.06** | **73,033.50** | **74,599.49** | **77,944.71** | **302,353.76** |

**STATE: GU - GUAM**

| REGISTRANT ZIP CODE 3 | 1ST QUARTER | 2ND QUARTER | 3RD QUARTER | 4TH QUARTER | TOTAL GRAMS |
|---|---|---|---|---|---|
| 969 | 919.77 | 876.63 | 897.74 | 769.11 | **3,463.26** |
| **GU - GUAM - Total** | **919.77** | **876.63** | **897.74** | **769.11** | **3,463.26** |

**STATE: HI - HAWAII**

| REGISTRANT ZIP CODE 3 | 1ST QUARTER | 2ND QUARTER | 3RD QUARTER | 4TH QUARTER | TOTAL GRAMS |
|---|---|---|---|---|---|
| 967 | 6,085.30 | 5,843.73 | 5,880.39 | 5,954.12 | **23,763.54** |
| 968 | 2,931.36 | 2,662.56 | 2,810.33 | 2,737.69 | **11,141.95** |
| **HI - HAWAII - Total** | **9,016.67** | **8,506.29** | **8,690.72** | **8,691.81** | **34,905.49** |

**STATE: IA - IOWA**

| REGISTRANT ZIP CODE 3 | 1ST QUARTER | 2ND QUARTER | 3RD QUARTER | 4TH QUARTER | TOTAL GRAMS |
|---|---|---|---|---|---|
| 500 | 901.73 | 737.70 | 693.20 | 765.80 | **3,098.43** |
| 501 | 655.66 | 836.87 | 688.89 | 807.49 | **2,988.91** |
| 502 | 717.35 | 561.16 | 508.58 | 598.98 | **2,386.07** |
| 503 | 1,307.48 | 1,117.04 | 1,348.62 | 1,237.23 | **5,010.37** |
| 504 | 207.75 | 154.71 | 228.00 | 167.97 | **758.43** |
| 505 | 462.63 | 400.22 | 485.22 | 394.29 | **1,742.35** |
| 506 | 643.14 | 531.24 | 561.54 | 628.58 | **2,364.50** |
| 507 | 455.28 | 350.85 | 416.60 | 351.77 | **1,574.51** |
| 508 | 162.62 | 99.45 | 122.66 | 109.40 | **494.14** |
| 510 | 326.16 | 277.36 | 335.00 | 207.67 | **1,146.19** |
| 511 | 449.01 | 532.23 | 497.48 | 300.38 | **1,779.11** |
| 512 | 76.51 | 77.35 | 114.93 | 57.55 | **326.35** |
| 513 | 172.93 | 157.61 | 196.23 | 209.91 | **736.69** |
| 514 | 155.81 | 251.95 | 152.68 | 191.17 | **751.62** |
| 515 | 601.51 | 626.65 | 494.30 | 586.39 | **2,308.85** |
| 516 | 201.12 | 157.81 | 229.83 | 189.50 | **778.25** |
| 520 | 698.30 | 540.36 | 598.37 | 542.02 | **2,379.05** |
| 521 | 172.39 | 190.07 | 148.08 | 143.66 | **654.19** |
| 522 | 708.22 | 580.39 | 560.47 | 578.15 | **2,427.23** |
| 523 | 483.36 | 351.03 | 406.92 | 370.09 | **1,611.40** |
| 524 | 565.23 | 469.00 | 530.97 | 518.44 | **2,083.63** |
| 525 | 766.63 | 735.83 | 812.72 | 643.93 | **2,959.12** |
| 526 | 530.75 | 584.18 | 590.88 | 624.52 | **2,330.33** |
| 527 | 912.95 | 837.99 | 860.81 | 865.62 | **3,477.37** |
| 528 | 574.54 | 659.64 | 687.54 | 677.95 | **2,599.67** |
| **IA - IOWA - Total** | **12,909.09** | **11,818.69** | **12,270.50** | **11,768.46** | **48,766.74** |

**STATE: ID - IDAHO**

| REGISTRANT ZIP CODE 3 | 1ST QUARTER | 2ND QUARTER | 3RD QUARTER | 4TH QUARTER | TOTAL GRAMS |
|---|---|---|---|---|---|
| 832 | 1,058.42 | 1,141.69 | 1,185.90 | 1,395.16 | **4,781.16** |
| 833 | 886.67 | 942.41 | 975.56 | 908.52 | **3,713.16** |
| 834 | 1,020.50 | 949.03 | 1,011.63 | 1,019.21 | **4,000.36** |
| 835 | 895.48 | 856.77 | 1,063.79 | 928.97 | **3,745.00** |
| 836 | 3,342.99 | 2,900.99 | 3,021.42 | 2,666.05 | **11,931.44** |
| 837 | 1,209.02 | 1,315.76 | 1,209.16 | 1,247.70 | **4,981.64** |

| | | | | | |
|---|---|---|---|---|---|
| 838 | 2,018.41 | 2,095.05 | 2,162.04 | 2,284.86 | **8,560.36** |
| **ID - IDAHO - Total** | **10,431.47** | **10,201.69** | **10,629.50** | **10,450.46** | **41,713.13** |

**STATE: IL - ILLINOIS**

| REGISTRANT ZIP CODE 3 | 1ST QUARTER | 2ND QUARTER | 3RD QUARTER | 4TH QUARTER | TOTAL GRAMS |
|---|---|---|---|---|---|
| 600 | 15,566.58 | 16,496.49 | 16,280.98 | 17,944.60 | **66,288.65** |
| 601 | 10,814.04 | 10,073.58 | 10,609.18 | 10,356.42 | **41,853.22** |
| 602 | 391.19 | 313.83 | 366.88 | 406.01 | **1,477.91** |
| 603 | 788.02 | 747.38 | 809.81 | 776.11 | **3,121.31** |
| 604 | 14,510.56 | 13,446.32 | 14,343.16 | 14,066.37 | **56,366.41** |
| 605 | 7,572.41 | 6,992.34 | 7,288.75 | 7,565.72 | **29,419.21** |
| 606 | 19,786.41 | 18,933.35 | 18,668.98 | 18,552.50 | **75,941.23** |
| 607 | 1,188.68 | 1,231.96 | 1,325.40 | 1,383.54 | **5,129.58** |
| 608 | 1,259.94 | 1,731.42 | 1,650.39 | 1,587.96 | **6,229.70** |
| 609 | 1,404.52 | 1,508.58 | 1,329.56 | 1,573.37 | **5,816.02** |
| 610 | 2,629.24 | 2,438.83 | 2,523.23 | 2,562.33 | **10,153.63** |
| 611 | 3,677.49 | 3,513.49 | 3,476.24 | 3,461.78 | **14,129.01** |
| 612 | 1,539.49 | 1,229.62 | 1,482.39 | 1,655.70 | **5,907.21** |
| 613 | 1,233.77 | 1,289.01 | 1,201.44 | 1,310.49 | **5,034.70** |
| 614 | 1,342.82 | 1,531.67 | 1,385.73 | 1,333.13 | **5,593.35** |
| 615 | 1,599.55 | 1,613.83 | 1,498.44 | 1,747.25 | **6,459.07** |
| 616 | 1,850.70 | 1,882.00 | 1,988.17 | 1,955.11 | **7,675.98** |
| 617 | 2,145.81 | 2,200.21 | 2,090.33 | 1,965.86 | **8,402.21** |
| 618 | 3,069.22 | 2,985.87 | 2,888.95 | 3,062.84 | **12,006.89** |
| 619 | 949.60 | 955.67 | 912.49 | 1,004.30 | **3,822.06** |
| 620 | 3,719.30 | 3,917.70 | 3,850.28 | 3,876.32 | **15,363.60** |
| 622 | 5,081.23 | 5,406.00 | 4,850.65 | 4,966.42 | **20,304.30** |
| 623 | 985.04 | 993.44 | 922.05 | 937.08 | **3,837.62** |
| 624 | 1,109.91 | 1,094.95 | 1,327.17 | 1,175.29 | **4,707.32** |
| 625 | 1,745.15 | 1,606.03 | 1,588.09 | 1,430.34 | **6,369.61** |
| 626 | 1,442.45 | 1,489.66 | 1,557.16 | 1,499.73 | **5,989.01** |
| 627 | 2,238.62 | 2,107.66 | 2,187.04 | 2,170.46 | **8,703.78** |
| 628 | 1,511.89 | 1,516.31 | 1,617.79 | 1,579.21 | **6,225.20** |
| 629 | 3,201.95 | 2,959.11 | 3,200.76 | 3,157.48 | **12,519.30** |
| **IL - ILLINOIS - Total** | **114,355.60** | **112,206.31** | **113,221.47** | **115,063.71** | **454,847.09** |

**STATE: IN - INDIANA**

| REGISTRANT ZIP CODE 3 | 1ST QUARTER | 2ND QUARTER | 3RD QUARTER | 4TH QUARTER | TOTAL GRAMS |
|---|---|---|---|---|---|
| 460 | 1,432.25 | 1,888.70 | 1,802.12 | 2,042.94 | **7,166.02** |
| 461 | 2,396.96 | 2,069.70 | 2,455.38 | 2,329.77 | **9,251.81** |
| 462 | 3,632.85 | 3,537.66 | 3,961.18 | 4,323.49 | **15,455.18** |
| 463 | 6,670.55 | 6,645.90 | 6,690.97 | 6,793.69 | **26,801.12** |
| 464 | 1,548.18 | 1,548.18 | 1,412.25 | 1,474.32 | **5,982.92** |
| 465 | 1,692.38 | 1,675.87 | 1,648.29 | 1,825.62 | **6,842.16** |
| 466 | 594.96 | 592.49 | 548.65 | 592.49 | **2,328.59** |
| 467 | 913.12 | 975.19 | 1,138.35 | 1,165.53 | **4,192.19** |
| 468 | 1,099.07 | 1,028.78 | 1,245.82 | 1,178.01 | **4,551.68** |
| 469 | 1,505.70 | 1,433.80 | 1,337.23 | 1,546.13 | **5,822.86** |
| 470 | 598.01 | 419.92 | 456.75 | 420.65 | **1,895.33** |
| 471 | 1,493.01 | 1,597.42 | 1,353.67 | 1,301.65 | **5,745.76** |
| 472 | 877.00 | 742.36 | 868.69 | 845.57 | **3,333.62** |
| 473 | 1,091.79 | 1,189.03 | 1,118.31 | 1,285.63 | **4,684.76** |
| 474 | 728.77 | 903.55 | 926.40 | 869.30 | **3,428.02** |
| 475 | 899.51 | 544.57 | 730.00 | 665.24 | **2,839.32** |
| 476 | 490.64 | 492.85 | 644.24 | 662.11 | **2,289.85** |
| 477 | 1,099.88 | 1,062.67 | 1,057.88 | 1,259.84 | **4,480.27** |
| 478 | 1,456.07 | 1,575.86 | 1,532.86 | 1,390.13 | **5,954.91** |
| 479 | 938.20 | 1,037.18 | 1,010.37 | 977.66 | **3,963.42** |
| **IN - INDIANA - Total** | **31,158.90** | **30,961.66** | **31,939.43** | **32,949.77** | **127,009.76** |

**STATE: KS - KANSAS**

| REGISTRANT ZIP CODE 3 | 1ST QUARTER | 2ND QUARTER | 3RD QUARTER | 4TH QUARTER | TOTAL GRAMS |
|---|---|---|---|---|---|
| 660 | 2,064.81 | 2,401.43 | 2,155.19 | 2,202.73 | **8,824.16** |

| | | | | | |
|---|---|---|---|---|---|
| 661 | 809.96 | 753.83 | 812.06 | 840.96 | **3,216.80** |
| 662 | 6,927.35 | 7,281.94 | 7,520.49 | 7,825.32 | **29,555.10** |
| 664 | 347.71 | 322.67 | 397.92 | 359.43 | **1,427.74** |
| 665 | 383.96 | 564.94 | 633.13 | 631.40 | **2,213.43** |
| 666 | 821.55 | 763.84 | 692.35 | 748.26 | **3,026.00** |
| 667 | 874.86 | 766.71 | 830.80 | 694.17 | **3,166.54** |
| 668 | 276.59 | 293.87 | 336.04 | 299.67 | **1,206.17** |
| 669 | 278.84 | 340.72 | 271.84 | 408.86 | **1,300.25** |
| 670 | 1,399.27 | 1,411.05 | 1,176.61 | 1,160.23 | **5,147.15** |
| 671 | 629.94 | 733.92 | 766.19 | 623.94 | **2,753.99** |
| 672 | 2,692.05 | 2,466.51 | 2,385.60 | 2,335.63 | **9,879.79** |
| 673 | 673.16 | 699.12 | 738.54 | 791.68 | **2,902.50** |
| 674 | 927.75 | 853.29 | 795.60 | 835.93 | **3,412.57** |
| 675 | 428.38 | 422.86 | 451.95 | 489.16 | **1,792.35** |
| 676 | 274.29 | 273.04 | 192.02 | 307.37 | **1,046.72** |
| 677 | 218.80 | 126.64 | 196.70 | 158.20 | **700.34** |
| 678 | 424.14 | 566.33 | 528.88 | 392.66 | **1,912.01** |
| 679 | 552.71 | 501.87 | 548.10 | 549.30 | **2,151.99** |
| **KS - KANSAS - Total** | **21,006.12** | **21,544.59** | **21,430.00** | **21,654.90** | **85,635.60** |
| **STATE: KY - KENTUCKY** | | | | | |
| REGISTRANT ZIP CODE 3 | 1ST QUARTER | 2ND QUARTER | 3RD QUARTER | 4TH QUARTER | **TOTAL GRAMS** |
| 400 | 1,443.23 | 1,454.16 | 1,453.52 | 1,279.04 | **5,629.94** |
| 401 | 570.75 | 568.80 | 554.70 | 584.38 | **2,278.63** |
| 402 | 3,671.66 | 3,838.44 | 3,556.79 | 3,878.01 | **14,944.89** |
| 403 | 1,428.26 | 1,741.44 | 1,701.52 | 1,782.55 | **6,653.77** |
| 404 | 1,111.24 | 939.84 | 976.86 | 1,044.27 | **4,072.21** |
| 405 | 1,007.08 | 859.82 | 804.52 | 836.18 | **3,507.60** |
| 406 | 125.98 | 64.09 | 55.25 | 161.34 | **406.66** |
| 407 | 1,228.07 | 1,259.56 | 1,221.63 | 1,174.11 | **4,883.37** |
| 408 | 656.40 | 486.22 | 559.34 | 502.80 | **2,204.76** |
| 409 | 1,631.05 | 1,624.42 | 1,753.71 | 1,635.66 | **6,644.85** |
| 410 | 2,789.69 | 2,438.83 | 2,696.49 | 2,498.14 | **10,423.16** |
| 411 | 869.67 | 739.46 | 790.38 | 831.91 | **3,231.43** |
| 412 | 1,376.89 | 1,403.41 | 1,457.56 | 1,503.97 | **5,741.84** |
| 413 | 1,192.57 | 1,173.34 | 1,333.13 | 981.51 | **4,680.55** |
| 414 | 423.90 | 410.64 | 364.89 | 429.64 | **1,629.06** |
| 415 | 2,054.29 | 2,160.37 | 2,071.41 | 2,055.39 | **8,341.46** |
| 416 | 800.06 | 808.90 | 736.63 | 784.59 | **3,130.16** |
| 417 | 539.26 | 583.47 | 663.03 | 519.37 | **2,305.13** |
| 418 | 517.35 | 331.52 | 325.99 | 374.61 | **1,549.46** |
| 420 | 1,663.10 | 1,884.11 | 1,502.31 | 1,869.74 | **6,919.26** |
| 421 | 2,032.96 | 1,931.61 | 1,989.99 | 2,046.07 | **8,000.63** |
| 422 | 1,126.05 | 1,085.34 | 1,210.39 | 1,225.50 | **4,647.28** |
| 423 | 1,610.91 | 1,134.67 | 1,460.68 | 1,377.77 | **5,584.02** |
| 424 | 1,184.34 | 1,337.46 | 1,014.25 | 1,189.06 | **4,725.12** |
| 425 | 422.49 | 342.75 | 544.05 | 437.60 | **1,746.89** |
| 426 | 943.71 | 959.18 | 816.91 | 801.15 | **3,520.95** |
| 427 | 2,188.70 | 1,942.60 | 2,028.81 | 1,861.39 | **8,021.50** |
| **KY - KENTUCKY - Total** | **34,609.66** | **33,504.47** | **33,644.74** | **33,665.74** | **135,424.61** |
| **STATE: LA - LOUISIANA** | | | | | |
| REGISTRANT ZIP CODE 3 | 1ST QUARTER | 2ND QUARTER | 3RD QUARTER | 4TH QUARTER | **TOTAL GRAMS** |
| 700 | 3,332.86 | 3,097.46 | 2,979.98 | 3,089.94 | **12,500.23** |
| 701 | 1,736.31 | 1,464.17 | 1,791.07 | 1,764.47 | **6,756.01** |
| 703 | 1,336.58 | 1,467.51 | 1,550.75 | 1,667.52 | **6,022.35** |
| 704 | 2,495.78 | 2,458.35 | 2,520.53 | 2,534.45 | **10,009.11** |
| 705 | 7,123.66 | 6,733.91 | 7,049.61 | 7,228.51 | **28,135.69** |
| 706 | 2,519.40 | 2,484.36 | 2,374.64 | 2,615.26 | **9,993.66** |
| 707 | 1,336.27 | 1,200.66 | 1,317.90 | 1,502.46 | **5,357.29** |
| 708 | 1,385.25 | 1,084.96 | 1,270.43 | 1,236.64 | **4,977.29** |
| 710 | 954.01 | 1,122.73 | 951.45 | 923.09 | **3,951.29** |

| | | | | | |
|---|---|---|---|---|---|
| 711 | 2,048.67 | 1,803.11 | 1,873.57 | 1,961.89 | **7,687.25** |
| 712 | 2,860.96 | 2,621.99 | 2,971.66 | 2,635.54 | **11,090.15** |
| 713 | 2,044.13 | 2,092.58 | 2,247.94 | 2,335.97 | **8,720.62** |
| 714 | 1,170.25 | 1,360.78 | 1,355.90 | 1,349.73 | **5,236.66** |
| **LA - LOUISIANA - Total** | **30,344.13** | **28,992.56** | **30,255.42** | **30,845.49** | **120,437.60** |

**STATE: MA - MASSACHUSETTS**

| REGISTRANT ZIP CODE 3 | 1ST QUARTER | 2ND QUARTER | 3RD QUARTER | 4TH QUARTER | TOTAL GRAMS |
|---|---|---|---|---|---|
| 010 | 2,186.77 | 2,046.61 | 2,226.89 | 2,109.42 | **8,569.69** |
| 011 | 746.25 | 830.97 | 654.79 | 666.13 | **2,898.14** |
| 012 | 398.91 | 496.52 | 457.92 | 488.06 | **1,841.40** |
| 013 | 476.46 | 252.56 | 365.15 | 367.45 | **1,461.62** |
| 014 | 1,266.14 | 1,153.14 | 1,089.98 | 1,410.18 | **4,919.45** |
| 015 | 2,029.15 | 1,709.89 | 1,944.37 | 2,010.97 | **7,694.39** |
| 016 | 1,454.90 | 1,459.10 | 1,299.04 | 1,412.88 | **5,625.92** |
| 017 | 1,856.12 | 1,701.76 | 1,533.47 | 1,786.51 | **6,877.86** |
| 018 | 6,400.34 | 5,386.09 | 5,533.41 | 5,705.50 | **23,025.33** |
| 019 | 2,231.91 | 1,773.26 | 1,700.18 | 1,913.74 | **7,619.09** |
| 020 | 1,616.81 | 1,528.64 | 1,446.01 | 1,742.92 | **6,334.37** |
| 021 | 4,947.95 | 4,584.13 | 4,542.66 | 4,655.67 | **18,730.40** |
| 022 | 190.42 | 126.77 | 160.57 | 104.67 | **582.43** |
| 023 | 1,800.60 | 1,812.01 | 1,720.66 | 1,759.88 | **7,093.15** |
| 024 | 1,182.56 | 892.81 | 1,173.58 | 1,079.01 | **4,327.96** |
| 025 | 532.43 | 505.06 | 702.03 | 588.86 | **2,328.39** |
| 026 | 918.65 | 838.62 | 824.05 | 915.58 | **3,496.90** |
| 027 | 2,696.44 | 2,491.59 | 2,483.12 | 2,759.38 | **10,430.53** |
| **MA - MASSACHUSETTS - Total** | **32,932.79** | **29,589.55** | **29,857.88** | **31,476.80** | **123,857.02** |

**STATE: MD - MARYLAND**

| REGISTRANT ZIP CODE 3 | 1ST QUARTER | 2ND QUARTER | 3RD QUARTER | 4TH QUARTER | TOTAL GRAMS |
|---|---|---|---|---|---|
| 206 | 3,336.54 | 3,237.82 | 3,322.93 | 3,669.33 | **13,566.62** |
| 207 | 9,969.55 | 8,929.38 | 9,310.60 | 9,849.60 | **38,059.13** |
| 208 | 4,300.63 | 3,756.23 | 4,172.22 | 4,103.04 | **16,332.11** |
| 209 | 1,625.88 | 1,410.80 | 1,573.96 | 1,581.36 | **6,192.00** |
| 210 | 5,081.79 | 4,326.61 | 5,416.25 | 5,399.37 | **20,224.01** |
| 211 | 2,492.47 | 2,200.57 | 2,405.74 | 2,540.35 | **9,639.13** |
| 212 | 7,412.22 | 6,471.57 | 7,188.84 | 6,723.60 | **27,796.22** |
| 214 | 856.91 | 1,176.28 | 959.84 | 1,041.73 | **4,034.76** |
| 215 | 833.01 | 708.23 | 837.43 | 732.27 | **3,110.95** |
| 216 | 1,207.70 | 1,069.07 | 1,170.39 | 1,190.82 | **4,637.98** |
| 217 | 3,540.32 | 3,144.56 | 3,096.40 | 3,249.34 | **13,030.62** |
| 218 | 1,206.73 | 1,104.78 | 1,041.54 | 982.87 | **4,335.92** |
| 219 | 597.83 | 542.02 | 651.95 | 562.64 | **2,354.45** |
| **MD - MARYLAND - Total** | **42,461.58** | **38,077.93** | **41,148.10** | **41,626.31** | **163,313.92** |

**STATE: ME - MAINE**

| REGISTRANT ZIP CODE 3 | 1ST QUARTER | 2ND QUARTER | 3RD QUARTER | 4TH QUARTER | TOTAL GRAMS |
|---|---|---|---|---|---|
| 039 | 89.51 | 40.89 | 100.93 | 92.82 | **324.15** |
| 040 | 1,404.61 | 1,404.85 | 1,373.48 | 1,536.05 | **5,718.99** |
| 041 | 353.41 | 285.60 | 292.24 | 455.64 | **1,386.90** |
| 042 | 1,049.67 | 1,047.57 | 949.49 | 869.77 | **3,916.50** |
| 043 | 448.25 | 588.92 | 541.79 | 518.93 | **2,097.89** |
| 044 | 854.93 | 821.42 | 880.80 | 674.81 | **3,231.96** |
| 045 | 209.00 | 174.12 | 164.46 | 213.74 | **761.32** |
| 046 | 610.53 | 632.82 | 703.70 | 650.86 | **2,597.91** |
| 047 | 746.62 | 663.19 | 674.99 | 726.64 | **2,811.45** |
| 048 | 229.16 | 209.39 | 215.65 | 229.28 | **883.48** |
| 049 | 886.78 | 730.96 | 781.96 | 766.70 | **3,166.40** |
| **ME - MAINE - Total** | **6,882.49** | **6,599.70** | **6,679.49** | **6,735.26** | **26,896.94** |

**STATE: MI - MICHIGAN**

| REGISTRANT ZIP CODE 3 | 1ST QUARTER | 2ND QUARTER | 3RD QUARTER | 4TH QUARTER | TOTAL GRAMS |
|---|---|---|---|---|---|
| 480 | 22,345.34 | 21,277.28 | 21,962.88 | 23,257.15 | **88,842.66** |
| 481 | 34,201.66 | 31,857.32 | 33,561.87 | 33,050.82 | **132,671.67** |

| | | | | | |
|---|---|---|---|---|---|
| 482 | 31,154.08 | 27,854.60 | 29,642.78 | 29,504.80 | **118,156.26** |
| 483 | 11,079.83 | 11,148.51 | 11,680.14 | 12,158.69 | **46,067.18** |
| 484 | 6,064.27 | 5,644.09 | 6,158.46 | 5,950.49 | **23,817.31** |
| 485 | 6,854.07 | 6,162.40 | 6,349.42 | 6,729.47 | **26,095.35** |
| 486 | 8,263.76 | 7,561.14 | 8,048.56 | 8,012.19 | **31,885.65** |
| 487 | 4,634.94 | 4,313.74 | 4,681.26 | 4,289.15 | **17,919.10** |
| 488 | 7,807.87 | 6,905.80 | 7,598.32 | 7,487.55 | **29,799.55** |
| 489 | 2,097.57 | 1,845.62 | 2,145.62 | 2,127.94 | **8,216.74** |
| 490 | 7,097.55 | 6,922.90 | 6,941.62 | 6,946.54 | **27,908.62** |
| 491 | 521.77 | 582.82 | 518.27 | 517.16 | **2,140.01** |
| 492 | 3,029.87 | 2,497.56 | 2,848.12 | 2,940.35 | **11,315.89** |
| 493 | 1,564.37 | 1,610.24 | 1,547.24 | 1,626.82 | **6,348.67** |
| 494 | 4,620.57 | 4,341.62 | 4,675.40 | 4,465.31 | **18,102.90** |
| 495 | 2,936.17 | 2,380.79 | 2,758.39 | 2,696.91 | **10,772.25** |
| 496 | 2,238.57 | 2,257.18 | 2,220.98 | 2,115.57 | **8,832.31** |
| 497 | 2,649.37 | 2,660.31 | 2,804.72 | 2,679.77 | **10,794.17** |
| 498 | 1,813.34 | 1,611.82 | 1,901.53 | 1,463.87 | **6,790.56** |
| 499 | 1,162.12 | 923.12 | 1,111.12 | 950.19 | **4,146.55** |
| **MI - MICHIGAN - Total** | **162,137.09** | **150,358.86** | **159,156.71** | **158,970.75** | **630,623.41** |

**STATE: MN - MINNESOTA**

| REGISTRANT ZIP CODE 3 | 1ST QUARTER | 2ND QUARTER | 3RD QUARTER | 4TH QUARTER | TOTAL GRAMS |
|---|---|---|---|---|---|
| 550 | 2,307.75 | 2,286.37 | 2,428.62 | 2,284.56 | **9,307.29** |
| 551 | 3,872.82 | 3,529.64 | 3,622.66 | 3,600.54 | **14,625.66** |
| 553 | 3,561.90 | 3,301.07 | 3,465.86 | 3,934.99 | **14,263.82** |
| 554 | 4,519.82 | 4,013.95 | 4,326.60 | 4,331.04 | **17,191.40** |
| 556 | 104.06 | 52.91 | 80.10 | 66.66 | **303.72** |
| 557 | 1,170.27 | 1,582.01 | 1,446.11 | 1,437.83 | **5,636.23** |
| 558 | 523.95 | 507.93 | 568.88 | 598.71 | **2,199.47** |
| 559 | 735.38 | 827.26 | 752.96 | 776.16 | **3,091.76** |
| 560 | 1,173.21 | 936.49 | 1,059.82 | 1,077.76 | **4,247.28** |
| 561 | 414.14 | 276.47 | 367.37 | 365.42 | **1,423.41** |
| 562 | 922.16 | 792.22 | 646.84 | 589.94 | **2,951.16** |
| 563 | 1,468.93 | 1,494.23 | 1,477.46 | 1,232.99 | **5,673.60** |
| 564 | 718.42 | 741.29 | 721.89 | 861.95 | **3,043.55** |
| 565 | 864.92 | 819.48 | 722.14 | 984.52 | **3,391.06** |
| 566 | 625.25 | 588.02 | 638.51 | 653.37 | **2,505.15** |
| 567 | 511.08 | 447.55 | 369.42 | 502.02 | **1,830.07** |
| **MN - MINNESOTA - Total** | **23,494.05** | **22,196.88** | **22,695.25** | **23,298.46** | **91,684.63** |

**STATE: MO - MISSOURI**

| REGISTRANT ZIP CODE 3 | 1ST QUARTER | 2ND QUARTER | 3RD QUARTER | 4TH QUARTER | TOTAL GRAMS |
|---|---|---|---|---|---|
| 630 | 5,474.45 | 5,952.28 | 5,524.21 | 5,241.02 | **22,191.96** |
| 631 | 20,732.53 | 18,249.61 | 17,945.94 | 19,786.76 | **76,714.83** |
| 633 | 2,909.54 | 3,335.63 | 2,880.73 | 3,158.88 | **12,284.78** |
| 634 | 327.64 | 404.45 | 430.88 | 394.58 | **1,557.55** |
| 635 | 479.59 | 556.95 | 471.95 | 583.47 | **2,091.95** |
| 636 | 1,529.82 | 1,407.33 | 1,610.15 | 1,505.16 | **6,052.45** |
| 637 | 860.76 | 729.88 | 907.90 | 1,005.58 | **3,504.12** |
| 638 | 1,964.96 | 1,827.75 | 1,928.31 | 1,884.47 | **7,605.50** |
| 639 | 821.42 | 815.71 | 698.39 | 795.73 | **3,131.24** |
| 640 | 4,278.40 | 3,926.93 | 4,039.93 | 4,267.76 | **16,513.03** |
| 641 | 3,278.61 | 2,982.09 | 2,999.21 | 3,111.81 | **12,371.72** |
| 644 | 570.71 | 531.70 | 578.85 | 463.93 | **2,145.19** |
| 645 | 442.46 | 474.51 | 450.16 | 502.58 | **1,869.71** |
| 646 | 501.69 | 554.74 | 406.84 | 460.81 | **1,924.07** |
| 647 | 986.51 | 1,051.55 | 1,123.13 | 926.03 | **4,087.23** |
| 648 | 1,163.06 | 1,192.53 | 1,204.17 | 1,144.39 | **4,704.15** |
| 650 | 923.07 | 782.64 | 1,007.60 | 977.40 | **3,690.70** |
| 651 | 566.15 | 550.42 | 501.80 | 574.77 | **2,193.14** |
| 652 | 1,388.84 | 1,427.25 | 1,563.15 | 1,472.94 | **5,852.18** |
| 653 | 734.75 | 800.41 | 661.55 | 806.12 | **3,002.83** |

| | | | | | |
|---|---|---|---|---|---|
| 654 | 506.67 | 450.86 | 667.45 | 501.69 | **2,126.67** |
| 655 | 1,009.27 | 1,099.36 | 881.99 | 1,000.15 | **3,990.77** |
| 656 | 1,251.46 | 1,192.44 | 1,166.67 | 1,350.37 | **4,960.95** |
| 657 | 2,028.19 | 1,945.66 | 2,323.86 | 2,251.38 | **8,549.08** |
| 658 | 2,004.60 | 1,967.69 | 1,912.30 | 2,037.52 | **7,922.11** |
| **MO - MISSOURI - Total** | **56,735.16** | **54,210.35** | **53,887.13** | **56,205.27** | **221,037.90** |

**STATE: MS - MISSISSIPPI**

| REGISTRANT ZIP CODE 3 | 1ST QUARTER | 2ND QUARTER | 3RD QUARTER | 4TH QUARTER | TOTAL GRAMS |
|---|---|---|---|---|---|
| 386 | 1,853.00 | 1,707.48 | 1,741.30 | 2,059.16 | **7,360.95** |
| 387 | 1,169.14 | 955.87 | 946.59 | 1,012.23 | **4,083.82** |
| 388 | 1,819.32 | 1,944.97 | 1,821.10 | 1,982.34 | **7,567.73** |
| 389 | 955.87 | 939.29 | 969.31 | 984.60 | **3,849.07** |
| 390 | 1,354.69 | 1,554.06 | 1,450.37 | 1,528.65 | **5,887.77** |
| 391 | 1,702.84 | 1,582.89 | 1,599.45 | 1,837.49 | **6,722.67** |
| 392 | 1,219.93 | 819.78 | 976.46 | 1,144.52 | **4,160.70** |
| 393 | 1,647.55 | 1,604.77 | 1,451.52 | 2,057.82 | **6,761.66** |
| 394 | 2,194.23 | 2,149.48 | 2,014.88 | 2,200.79 | **8,559.38** |
| 395 | 1,556.81 | 1,495.30 | 1,437.28 | 1,715.66 | **6,205.05** |
| 396 | 784.87 | 895.16 | 725.00 | 870.78 | **3,275.82** |
| 397 | 1,463.26 | 1,507.26 | 1,606.91 | 1,397.69 | **5,975.13** |
| **MS - MISSISSIPPI - Total** | **17,721.51** | **17,156.32** | **16,740.17** | **18,791.74** | **70,409.74** |

**STATE: MT - MONTANA**

| REGISTRANT ZIP CODE 3 | 1ST QUARTER | 2ND QUARTER | 3RD QUARTER | 4TH QUARTER | TOTAL GRAMS |
|---|---|---|---|---|---|
| 590 | 527.11 | 440.16 | 452.14 | 387.87 | **1,807.28** |
| 591 | 745.56 | 683.55 | 604.56 | 704.47 | **2,738.13** |
| 592 | 337.38 | 248.60 | 244.00 | 241.42 | **1,071.40** |
| 593 | 306.63 | 325.61 | 272.39 | 398.91 | **1,303.55** |
| 594 | 759.46 | 690.98 | 872.87 | 865.42 | **3,188.74** |
| 595 | 238.69 | 165.76 | 161.34 | 240.90 | **806.69** |
| 596 | 629.96 | 583.93 | 530.92 | 601.62 | **2,346.43** |
| 597 | 1,003.19 | 1,078.31 | 1,102.82 | 891.76 | **4,076.08** |
| 598 | 1,749.43 | 1,709.21 | 1,585.88 | 1,546.98 | **6,591.49** |
| 599 | 844.23 | 770.77 | 874.17 | 939.36 | **3,428.53** |
| **MT - MONTANA - Total** | **7,141.64** | **6,696.87** | **6,701.08** | **6,818.72** | **27,358.30** |

**STATE: NC - NORTH CAROLINA**

| REGISTRANT ZIP CODE 3 | 1ST QUARTER | 2ND QUARTER | 3RD QUARTER | 4TH QUARTER | TOTAL GRAMS |
|---|---|---|---|---|---|
| 270 | 834.68 | 875.75 | 753.64 | 854.39 | **3,318.45** |
| 271 | 717.80 | 657.65 | 609.50 | 652.59 | **2,637.54** |
| 272 | 1,762.32 | 1,819.45 | 1,642.20 | 1,704.52 | **6,928.48** |
| 273 | 1,192.90 | 1,213.71 | 1,134.43 | 1,342.25 | **4,883.28** |
| 274 | 989.20 | 926.21 | 936.35 | 885.48 | **3,737.25** |
| 275 | 3,424.65 | 3,323.57 | 3,249.07 | 3,359.95 | **13,357.24** |
| 276 | 1,261.20 | 1,278.30 | 1,364.87 | 1,338.31 | **5,242.68** |
| 277 | 957.04 | 884.08 | 894.54 | 964.50 | **3,700.17** |
| 278 | 1,803.14 | 1,866.87 | 1,781.22 | 1,850.85 | **7,302.08** |
| 279 | 809.96 | 1,027.66 | 932.10 | 1,097.85 | **3,867.57** |
| 280 | 2,332.60 | 1,987.57 | 1,698.32 | 1,990.08 | **8,008.56** |
| 281 | 1,625.30 | 1,477.39 | 1,279.40 | 1,506.64 | **5,888.72** |
| 282 | 1,417.95 | 1,513.17 | 1,360.59 | 1,470.97 | **5,762.67** |
| 283 | 4,866.82 | 4,660.44 | 4,694.02 | 4,770.80 | **18,992.09** |
| 284 | 1,871.53 | 1,677.40 | 1,896.22 | 1,809.28 | **7,254.43** |
| 285 | 2,214.33 | 2,297.57 | 1,803.04 | 2,238.91 | **8,553.84** |
| 286 | 2,230.40 | 2,353.97 | 2,125.74 | 2,329.71 | **9,039.81** |
| 287 | 1,545.00 | 1,385.92 | 1,604.94 | 1,570.80 | **6,106.66** |
| 288 | 528.16 | 519.17 | 519.71 | 464.75 | **2,031.79** |
| 289 | 606.30 | 535.10 | 611.26 | 809.81 | **2,562.47** |
| **NC - NORTH CAROLINA - Total** | **32,991.28** | **32,280.93** | **30,891.15** | **33,012.42** | **129,175.78** |

**STATE: ND - NORTH DAKOTA**

| REGISTRANT ZIP CODE 3 | 1ST QUARTER | 2ND QUARTER | 3RD QUARTER | 4TH QUARTER | TOTAL GRAMS |
|---|---|---|---|---|---|
| 580 | 254.16 | 190.25 | 218.98 | 172.39 | **835.78** |

| | | | | | |
|---|---|---|---|---|---|
| 581 | 532.04 | 350.34 | 406.26 | 495.10 | **1,783.74** |
| 582 | 483.53 | 484.77 | 463.97 | 521.87 | **1,954.13** |
| 583 | 223.22 | 243.11 | 198.91 | 203.33 | **868.57** |
| 584 | 209.30 | 166.20 | 175.92 | 183.22 | **734.64** |
| 585 | 663.73 | 603.67 | 487.63 | 740.23 | **2,495.26** |
| 586 | 277.37 | 193.47 | 301.00 | 260.79 | **1,032.63** |
| 587 | 361.34 | 281.96 | 386.77 | 331.32 | **1,361.38** |
| 588 | 775.34 | 749.22 | 691.35 | 755.85 | **2,971.76** |
| **ND - NORTH DAKOTA - Total** | **3,780.02** | **3,263.00** | **3,330.78** | **3,664.10** | **14,037.91** |

**STATE: NE - NEBRASKA**

| REGISTRANT ZIP CODE 3 | 1ST QUARTER | 2ND QUARTER | 3RD QUARTER | 4TH QUARTER | TOTAL GRAMS |
|---|---|---|---|---|---|
| 680 | 778.66 | 849.68 | 774.66 | 791.23 | **3,194.23** |
| 681 | 2,640.63 | 2,672.66 | 2,597.34 | 2,456.00 | **10,366.63** |
| 683 | 223.77 | 205.54 | 274.05 | 305.36 | **1,008.72** |
| 684 | 134.82 | 146.05 | 158.30 | 168.50 | **607.66** |
| 685 | 1,145.13 | 986.86 | 1,086.34 | 1,213.48 | **4,431.81** |
| 686 | 175.33 | 88.77 | 175.14 | 168.33 | **607.57** |
| 687 | 517.61 | 514.95 | 471.86 | 592.31 | **2,096.72** |
| 688 | 685.13 | 918.21 | 887.11 | 754.43 | **3,244.88** |
| 689 | 221.01 | 213.08 | 218.80 | 169.62 | **822.51** |
| 690 | 122.10 | 139.24 | 64.09 | 112.72 | **438.15** |
| 691 | 731.91 | 732.09 | 625.64 | 759.53 | **2,849.17** |
| 692 | 15.47 | 8.84 | 6.63 | 5.53 | **36.47** |
| 693 | 317.51 | 415.50 | 286.39 | 281.79 | **1,301.19** |
| **NE - NEBRASKA - Total** | **7,709.07** | **7,891.47** | **7,626.35** | **7,778.81** | **31,005.70** |

**STATE: NH - NEW HAMPSHIRE**

| REGISTRANT ZIP CODE 3 | 1ST QUARTER | 2ND QUARTER | 3RD QUARTER | 4TH QUARTER | TOTAL GRAMS |
|---|---|---|---|---|---|
| 030 | 1,482.90 | 1,524.59 | 1,454.33 | 1,742.75 | **6,204.57** |
| 031 | 1,022.27 | 1,067.82 | 900.18 | 910.32 | **3,900.58** |
| 032 | 603.50 | 712.70 | 685.35 | 679.56 | **2,681.11** |
| 033 | 490.07 | 439.24 | 289.68 | 338.68 | **1,557.67** |
| 034 | 321.18 | 325.59 | 224.94 | 321.91 | **1,193.62** |
| 035 | 250.29 | 299.46 | 205.25 | 249.55 | **1,004.54** |
| 036 | 47.52 | 22.10 | 46.41 | 48.62 | **164.65** |
| 037 | 350.11 | 461.70 | 390.34 | 356.26 | **1,558.41** |
| 038 | 1,152.88 | 1,021.94 | 1,060.15 | 1,089.46 | **4,324.43** |
| **NH - NEW HAMPSHIRE - Total** | **5,720.71** | **5,875.14** | **5,256.63** | **5,737.10** | **22,589.58** |

**STATE: NJ - NEW JERSEY**

| REGISTRANT ZIP CODE 3 | 1ST QUARTER | 2ND QUARTER | 3RD QUARTER | 4TH QUARTER | TOTAL GRAMS |
|---|---|---|---|---|---|
| 070 | 9,593.62 | 8,160.58 | 9,414.05 | 9,365.40 | **36,533.65** |
| 071 | 1,687.82 | 1,123.19 | 1,397.53 | 1,522.86 | **5,731.41** |
| 072 | 715.67 | 661.52 | 854.28 | 770.38 | **3,001.85** |
| 073 | 1,590.79 | 1,559.16 | 1,571.91 | 1,400.44 | **6,122.31** |
| 074 | 3,139.65 | 2,969.13 | 3,311.78 | 3,299.58 | **12,720.15** |
| 075 | 1,327.18 | 1,104.11 | 1,207.33 | 1,217.19 | **4,855.81** |
| 076 | 3,247.91 | 2,614.77 | 2,766.37 | 2,912.68 | **11,541.72** |
| 077 | 4,696.60 | 3,748.27 | 4,955.36 | 4,838.24 | **18,238.47** |
| 078 | 2,226.23 | 2,094.24 | 2,042.43 | 2,353.08 | **8,715.99** |
| 079 | 1,201.48 | 669.29 | 926.13 | 981.07 | **3,777.96** |
| 080 | 9,363.03 | 8,461.48 | 9,346.82 | 9,848.02 | **37,019.35** |
| 081 | 1,590.17 | 1,319.88 | 1,437.20 | 1,283.78 | **5,631.04** |
| 082 | 1,756.18 | 1,855.26 | 1,965.85 | 2,121.67 | **7,698.97** |
| 083 | 1,604.38 | 1,373.15 | 1,630.85 | 1,500.70 | **6,109.07** |
| 084 | 277.37 | 357.11 | 430.97 | 463.20 | **1,528.65** |
| 085 | 1,395.71 | 1,226.45 | 1,386.73 | 1,528.09 | **5,536.99** |
| 086 | 1,490.26 | 1,511.41 | 1,603.64 | 1,589.89 | **6,195.20** |
| 087 | 3,682.24 | 3,411.74 | 3,991.12 | 4,112.04 | **15,197.15** |
| 088 | 5,171.34 | 4,372.09 | 5,163.27 | 5,223.64 | **19,930.34** |
| 089 | 476.56 | 374.50 | 474.04 | 360.51 | **1,685.61** |
| **NJ - NEW JERSEY - Total** | **56,234.18** | **48,967.33** | **55,877.69** | **56,692.47** | **217,771.67** |

**STATE: NM - NEW MEXICO**

| REGISTRANT ZIP CODE 3 | 1ST QUARTER | 2ND QUARTER | 3RD QUARTER | 4TH QUARTER | TOTAL GRAMS |
|---|---|---|---|---|---|
| 870 | 1,241.30 | 1,099.01 | 1,218.47 | 1,208.60 | **4,767.38** |
| 871 | 3,713.08 | 3,833.71 | 3,687.97 | 3,883.05 | **15,117.81** |
| 873 | 1,190.47 | 1,011.33 | 934.09 | 925.79 | **4,061.68** |
| 874 | 509.43 | 469.45 | 420.28 | 496.16 | **1,895.32** |
| 875 | 665.37 | 681.52 | 666.20 | 775.05 | **2,788.14** |
| 877 | 264.65 | 304.80 | 323.40 | 267.22 | **1,160.07** |
| 878 | 70.72 | 48.62 | 21.00 | 35.36 | **175.70** |
| 879 | 89.60 | 121.56 | 137.03 | 146.97 | **495.15** |
| 880 | 2,225.52 | 2,068.38 | 1,920.51 | 2,033.86 | **8,248.28** |
| 881 | 877.41 | 737.07 | 866.54 | 773.90 | **3,254.92** |
| 882 | 2,128.25 | 2,016.72 | 1,873.61 | 2,009.12 | **8,027.69** |
| 883 | 575.25 | 714.41 | 798.39 | 758.25 | **2,846.30** |
| 884 | 101.66 | 125.98 | 139.24 | 78.46 | **445.34** |
| **NM - NEW MEXICO - Total** | **13,652.71** | **13,232.55** | **13,006.74** | **13,391.78** | **53,283.78** |

**STATE: NV - NEVADA**

| REGISTRANT ZIP CODE 3 | 1ST QUARTER | 2ND QUARTER | 3RD QUARTER | 4TH QUARTER | TOTAL GRAMS |
|---|---|---|---|---|---|
| 890 | 4,125.69 | 4,021.98 | 4,169.13 | 4,132.28 | **16,449.08** |
| 891 | 8,710.07 | 8,605.24 | 8,356.69 | 8,236.59 | **33,908.59** |
| 893 | 31.60 | 62.99 | 40.44 | 60.34 | **195.37** |
| 894 | 1,114.97 | 1,138.18 | 1,078.69 | 1,070.57 | **4,402.41** |
| 895 | 1,482.33 | 1,324.70 | 1,442.98 | 1,420.75 | **5,670.76** |
| 897 | 395.22 | 420.83 | 374.78 | 346.97 | **1,537.80** |
| 898 | 197.28 | 129.40 | 128.55 | 77.72 | **532.95** |
| **NV - NEVADA - Total** | **16,057.17** | **15,703.30** | **15,591.27** | **15,345.22** | **62,696.95** |

**STATE: NY - NEW YORK**

| REGISTRANT ZIP CODE 3 | 1ST QUARTER | 2ND QUARTER | 3RD QUARTER | 4TH QUARTER | TOTAL GRAMS |
|---|---|---|---|---|---|
| 063 | 0 | 0 | 0 | 2.21 | **2.21** |
| 100 | 8,914.66 | 7,078.22 | 7,451.38 | 7,906.87 | **31,351.13** |
| 101 | 348.45 | 170.86 | 257.86 | 370.61 | **1,147.77** |
| 102 | 41.99 | 20.80 | 17.68 | 35.35 | **115.82** |
| 103 | 2,407.92 | 2,020.03 | 2,046.36 | 2,191.21 | **8,665.52** |
| 104 | 5,986.16 | 4,812.24 | 5,520.92 | 5,773.55 | **22,092.88** |
| 105 | 2,028.67 | 2,187.75 | 1,994.94 | 2,019.38 | **8,230.74** |
| 106 | 408.87 | 343.94 | 444.13 | 452.60 | **1,649.54** |
| 107 | 1,101.13 | 759.65 | 987.50 | 1,035.18 | **3,883.46** |
| 108 | 337.03 | 292.55 | 229.83 | 324.78 | **1,184.19** |
| 109 | 3,489.24 | 3,168.05 | 3,717.08 | 3,719.77 | **14,094.14** |
| 110 | 900.40 | 839.35 | 752.79 | 969.09 | **3,461.62** |
| 111 | 745.90 | 747.18 | 759.27 | 588.16 | **2,840.51** |
| 112 | 12,207.44 | 10,448.06 | 10,380.07 | 10,985.44 | **44,021.01** |
| 113 | 4,756.49 | 3,495.85 | 4,092.29 | 3,566.09 | **15,910.71** |
| 114 | 2,136.43 | 1,598.89 | 1,744.86 | 1,753.33 | **7,233.50** |
| 115 | 3,076.60 | 2,419.24 | 2,537.06 | 2,564.58 | **10,597.49** |
| 116 | 575.82 | 447.55 | 309.78 | 440.36 | **1,773.50** |
| 117 | 6,460.13 | 5,089.44 | 5,865.34 | 5,516.62 | **22,931.52** |
| 118 | 374.76 | 220.88 | 299.43 | 328.16 | **1,223.23** |
| 119 | 828.63 | 939.50 | 1,047.86 | 844.39 | **3,660.39** |
| 120 | 1,468.78 | 1,527.41 | 1,173.99 | 1,178.13 | **5,348.31** |
| 121 | 660.44 | 678.11 | 709.98 | 669.65 | **2,718.18** |
| 122 | 490.24 | 497.03 | 383.86 | 506.79 | **1,877.92** |
| 123 | 515.32 | 455.64 | 475.54 | 499.11 | **1,945.60** |
| 124 | 738.72 | 759.61 | 881.54 | 839.36 | **3,219.23** |
| 125 | 2,122.54 | 2,038.38 | 2,309.39 | 2,375.32 | **8,845.63** |
| 126 | 339.24 | 272.21 | 236.74 | 288.87 | **1,137.06** |
| 127 | 1,644.31 | 1,689.62 | 1,904.37 | 2,108.89 | **7,347.19** |
| 128 | 1,375.20 | 1,069.75 | 1,239.74 | 1,203.09 | **4,887.78** |
| 129 | 1,019.39 | 954.53 | 885.29 | 1,024.90 | **3,884.11** |
| 130 | 2,476.91 | 2,427.34 | 2,301.69 | 2,395.38 | **9,601.33** |

| | | | | | |
|---|---|---|---|---|---|
| 131 | 856.78 | 784.21 | 805.19 | 896.37 | **3,342.54** |
| 132 | 1,189.75 | 1,110.26 | 1,037.43 | 1,114.11 | **4,451.56** |
| 133 | 649.20 | 625.97 | 634.18 | 507.01 | **2,416.36** |
| 134 | 725.46 | 611.55 | 621.68 | 623.25 | **2,581.93** |
| 135 | 353.97 | 333.17 | 281.23 | 297.80 | **1,266.17** |
| 136 | 3,824.01 | 3,589.86 | 3,485.28 | 3,094.19 | **13,993.33** |
| 137 | 669.10 | 502.88 | 559.34 | 628.94 | **2,360.26** |
| 138 | 954.56 | 689.81 | 910.90 | 990.46 | **3,545.73** |
| 139 | 730.80 | 682.80 | 624.89 | 704.46 | **2,742.96** |
| 140 | 4,359.62 | 4,026.94 | 4,149.83 | 4,148.51 | **16,684.90** |
| 141 | 1,858.11 | 1,967.13 | 1,950.38 | 1,959.66 | **7,735.28** |
| 142 | 7,875.97 | 7,320.27 | 7,261.88 | 7,258.37 | **29,716.47** |
| 143 | 624.35 | 596.91 | 592.31 | 635.95 | **2,449.52** |
| 144 | 2,145.03 | 1,816.20 | 1,780.19 | 2,070.72 | **7,812.14** |
| 145 | 1,717.17 | 1,423.79 | 1,500.69 | 1,541.56 | **6,183.21** |
| 146 | 2,925.65 | 2,282.30 | 2,672.29 | 2,611.04 | **10,491.28** |
| 147 | 1,006.86 | 932.10 | 789.72 | 1,089.62 | **3,818.30** |
| 148 | 1,443.72 | 1,116.79 | 1,404.27 | 1,497.93 | **5,462.71** |
| 149 | 226.90 | 207.75 | 203.33 | 258.95 | **896.92** |
| **NY - NEW YORK - Total** | **104,114.82** | **90,090.35** | **94,223.53** | **96,406.11** | **384,834.80** |

**STATE: OH - OHIO**

| REGISTRANT ZIP CODE 3 | 1ST QUARTER | 2ND QUARTER | 3RD QUARTER | 4TH QUARTER | TOTAL GRAMS |
|---|---|---|---|---|---|
| 430 | 2,590.67 | 2,596.45 | 2,536.30 | 2,637.43 | **10,360.85** |
| 431 | 2,153.55 | 1,698.18 | 1,926.74 | 1,985.76 | **7,764.23** |
| 432 | 3,947.81 | 3,707.75 | 3,742.16 | 4,018.97 | **15,416.68** |
| 433 | 470.43 | 668.45 | 553.97 | 588.93 | **2,281.78** |
| 434 | 785.50 | 760.63 | 732.73 | 761.00 | **3,039.85** |
| 435 | 1,681.24 | 1,776.64 | 1,652.95 | 1,750.21 | **6,861.03** |
| 436 | 3,559.74 | 3,371.07 | 3,356.58 | 3,165.76 | **13,453.16** |
| 437 | 1,004.30 | 1,095.78 | 932.16 | 918.71 | **3,950.95** |
| 438 | 218.80 | 183.44 | 192.28 | 284.00 | **878.51** |
| 439 | 1,597.89 | 1,406.34 | 1,352.93 | 1,542.55 | **5,899.71** |
| 440 | 3,842.11 | 3,273.28 | 3,436.88 | 3,380.43 | **13,932.69** |
| 441 | 6,452.29 | 5,832.12 | 6,419.35 | 6,004.72 | **24,708.48** |
| 442 | 1,647.90 | 1,452.10 | 1,743.43 | 1,437.74 | **6,281.18** |
| 443 | 764.97 | 742.56 | 771.62 | 916.02 | **3,195.17** |
| 444 | 1,815.74 | 1,262.29 | 1,708.92 | 1,637.46 | **6,424.42** |
| 445 | 1,033.00 | 903.01 | 860.83 | 1,006.93 | **3,803.78** |
| 446 | 1,655.71 | 1,538.52 | 1,509.19 | 1,629.35 | **6,332.76** |
| 447 | 1,418.12 | 1,292.11 | 1,286.51 | 1,351.06 | **5,347.80** |
| 448 | 1,173.72 | 1,288.35 | 1,376.11 | 1,207.52 | **5,045.70** |
| 449 | 549.30 | 451.97 | 596.44 | 636.87 | **2,234.58** |
| 450 | 2,661.22 | 2,589.04 | 2,741.57 | 2,689.10 | **10,680.93** |
| 451 | 1,206.10 | 1,204.47 | 1,290.67 | 1,177.40 | **4,878.64** |
| 452 | 3,523.69 | 3,399.70 | 3,262.86 | 3,349.86 | **13,536.11** |
| 453 | 2,538.35 | 2,260.54 | 2,467.54 | 2,350.59 | **9,617.02** |
| 454 | 2,788.93 | 2,613.01 | 2,962.19 | 2,759.46 | **11,123.58** |
| 455 | 447.17 | 383.08 | 422.12 | 416.49 | **1,668.85** |
| 456 | 2,525.76 | 2,385.41 | 2,267.63 | 2,417.17 | **9,595.97** |
| 457 | 1,107.25 | 906.86 | 960.00 | 845.73 | **3,819.83** |
| 458 | 1,812.79 | 1,913.51 | 1,707.92 | 1,930.59 | **7,364.81** |
| **OH - OHIO - Total** | **56,974.04** | **52,956.64** | **54,770.58** | **54,797.81** | **219,499.06** |

**STATE: OK - OKLAHOMA**

| REGISTRANT ZIP CODE 3 | 1ST QUARTER | 2ND QUARTER | 3RD QUARTER | 4TH QUARTER | TOTAL GRAMS |
|---|---|---|---|---|---|
| 730 | 9,086.65 | 9,025.32 | 11,458.78 | 11,422.08 | **40,992.84** |
| 731 | 7,010.93 | 6,425.64 | 10,278.42 | 9,819.88 | **33,534.87** |
| 734 | 3,295.54 | 3,233.38 | 3,380.35 | 3,296.36 | **13,205.63** |
| 735 | 6,110.59 | 6,293.04 | 6,546.72 | 5,714.49 | **24,664.84** |
| 736 | 1,522.76 | 1,524.97 | 1,425.51 | 1,448.72 | **5,921.96** |
| 737 | 1,792.39 | 1,927.39 | 1,846.54 | 1,787.97 | **7,354.29** |

| | | | | | |
|---|---|---|---|---|---|
| 738 | 419.94 | 387.68 | 459.70 | 518.81 | **1,786.14** |
| 739 | 397.82 | 296.52 | 403.34 | 497.27 | **1,594.95** |
| 740 | 8,964.26 | 8,216.70 | 8,544.39 | 8,682.90 | **34,408.25** |
| 741 | 3,928.50 | 3,752.14 | 3,706.70 | 3,744.56 | **15,131.91** |
| 743 | 3,183.65 | 3,140.55 | 3,337.25 | 3,234.48 | **12,895.93** |
| 744 | 5,198.20 | 4,441.01 | 4,580.64 | 4,554.51 | **18,774.37** |
| 745 | 1,922.79 | 1,585.30 | 1,713.01 | 1,976.58 | **7,197.68** |
| 746 | 1,538.23 | 1,509.50 | 1,575.14 | 1,529.39 | **6,152.26** |
| 747 | 4,682.38 | 4,722.06 | 4,747.29 | 4,742.87 | **18,894.61** |
| 748 | 7,265.69 | 6,970.28 | 7,963.94 | 7,119.42 | **29,319.33** |
| 749 | 2,051.08 | 2,255.77 | 1,789.17 | 1,922.88 | **8,018.89** |
| **OK - OKLAHOMA - Total** | **68,371.42** | **65,707.25** | **73,756.91** | **72,013.18** | **279,848.77** |

**STATE: OR - OREGON**

| REGISTRANT ZIP CODE 3 | 1ST QUARTER | 2ND QUARTER | 3RD QUARTER | 4TH QUARTER | TOTAL GRAMS |
|---|---|---|---|---|---|
| 970 | 4,096.09 | 3,784.29 | 4,036.70 | 3,944.79 | **15,861.87** |
| 971 | 1,685.80 | 1,627.43 | 1,564.03 | 1,629.28 | **6,506.55** |
| 972 | 5,617.57 | 5,980.79 | 5,561.08 | 5,395.32 | **22,554.77** |
| 973 | 3,435.69 | 3,293.25 | 3,453.86 | 3,321.20 | **13,504.01** |
| 974 | 3,772.16 | 3,524.00 | 3,775.79 | 3,679.28 | **14,751.23** |
| 975 | 2,550.17 | 2,550.31 | 2,854.26 | 2,658.38 | **10,613.12** |
| 976 | 470.35 | 414.54 | 497.81 | 467.85 | **1,850.55** |
| 977 | 1,303.91 | 1,343.43 | 1,272.41 | 1,420.72 | **5,340.47** |
| 978 | 1,062.12 | 1,040.46 | 1,285.55 | 1,041.17 | **4,429.30** |
| 979 | 282.89 | 243.11 | 227.82 | 215.89 | **969.71** |
| **OR - OREGON - Total** | **24,276.75** | **23,801.62** | **24,529.32** | **23,773.89** | **96,381.58** |

**STATE: PA - PENNSYLVANIA**

| REGISTRANT ZIP CODE 3 | 1ST QUARTER | 2ND QUARTER | 3RD QUARTER | 4TH QUARTER | TOTAL GRAMS |
|---|---|---|---|---|---|
| 150 | 1,946.61 | 2,021.70 | 2,156.36 | 2,062.34 | **8,187.00** |
| 151 | 2,301.08 | 2,446.72 | 2,031.97 | 2,449.72 | **9,229.49** |
| 152 | 3,475.45 | 3,529.51 | 3,107.23 | 3,678.77 | **13,790.96** |
| 153 | 777.85 | 824.88 | 951.29 | 1,031.69 | **3,585.71** |
| 154 | 627.23 | 737.95 | 532.19 | 537.24 | **2,434.61** |
| 155 | 3,464.99 | 2,980.98 | 3,255.48 | 2,766.30 | **12,467.76** |
| 156 | 1,761.27 | 1,430.70 | 1,381.90 | 1,728.51 | **6,302.38** |
| 157 | 1,260.19 | 896.61 | 1,406.91 | 1,274.23 | **4,837.93** |
| 158 | 478.67 | 566.88 | 570.56 | 584.38 | **2,200.48** |
| 159 | 997.85 | 816.52 | 775.82 | 795.99 | **3,386.18** |
| 160 | 1,082.35 | 1,006.66 | 1,145.58 | 972.39 | **4,206.99** |
| 161 | 1,064.73 | 1,138.89 | 1,115.54 | 981.83 | **4,301.00** |
| 162 | 605.71 | 535.07 | 667.85 | 527.07 | **2,335.70** |
| 163 | 890.84 | 1,070.86 | 882.18 | 748.83 | **3,592.72** |
| 164 | 390.28 | 382.13 | 400.34 | 391.07 | **1,563.82** |
| 165 | 1,249.58 | 968.90 | 1,080.34 | 1,126.58 | **4,425.39** |
| 166 | 1,006.14 | 926.84 | 1,083.85 | 974.58 | **3,991.40** |
| 167 | 362.46 | 417.71 | 318.25 | 360.43 | **1,458.85** |
| 168 | 360.53 | 388.29 | 390.55 | 451.36 | **1,590.73** |
| 169 | 314.94 | 295.23 | 279.20 | 285.29 | **1,174.66** |
| 170 | 3,050.12 | 2,722.27 | 2,660.41 | 2,880.33 | **11,313.14** |
| 171 | 738.36 | 718.84 | 805.16 | 593.40 | **2,855.76** |
| 172 | 642.57 | 629.11 | 697.82 | 692.65 | **2,662.15** |
| 173 | 1,603.39 | 1,473.09 | 1,608.37 | 1,603.22 | **6,288.07** |
| 174 | 854.56 | 658.95 | 792.45 | 770.73 | **3,076.68** |
| 175 | 896.37 | 895.36 | 883.57 | 929.89 | **3,605.19** |
| 176 | 863.92 | 770.89 | 720.39 | 791.11 | **3,146.32** |
| 177 | 752.57 | 673.00 | 710.51 | 678.52 | **2,814.60** |
| 178 | 970.65 | 1,095.33 | 906.74 | 1,105.88 | **4,078.60** |
| 179 | 885.25 | 933.57 | 932.06 | 943.78 | **3,694.66** |
| 180 | 2,327.55 | 2,004.00 | 2,014.78 | 1,978.28 | **8,324.61** |
| 181 | 1,093.35 | 786.25 | 826.90 | 1,151.08 | **3,857.57** |
| 182 | 715.51 | 636.12 | 639.08 | 745.71 | **2,736.43** |

| | | | | | |
|---|---|---|---|---|---|
| 183 | 1,480.96 | 1,425.10 | 1,587.49 | 1,681.22 | **6,174.77** |
| 184 | 1,385.51 | 1,255.18 | 1,384.98 | 1,338.91 | **5,364.58** |
| 185 | 1,135.06 | 1,011.45 | 1,044.61 | 886.67 | **4,077.78** |
| 186 | 1,356.34 | 1,362.38 | 1,414.98 | 1,418.05 | **5,551.75** |
| 187 | 2,991.92 | 3,052.01 | 3,303.29 | 3,056.83 | **12,404.05** |
| 188 | 1,094.94 | 905.51 | 914.28 | 879.40 | **3,794.13** |
| 189 | 2,325.38 | 2,002.93 | 2,124.97 | 2,345.32 | **8,798.60** |
| 190 | 8,113.73 | 8,425.77 | 8,922.45 | 9,381.29 | **34,843.25** |
| 191 | 14,256.93 | 13,610.80 | 13,621.66 | 14,053.00 | **55,542.40** |
| 193 | 2,371.27 | 2,166.69 | 2,144.67 | 2,168.96 | **8,851.59** |
| 194 | 3,436.50 | 3,045.11 | 3,441.14 | 3,585.34 | **13,508.09** |
| 195 | 1,041.11 | 1,078.83 | 976.27 | 1,098.94 | **4,195.15** |
| 196 | 1,675.24 | 1,313.15 | 1,532.19 | 1,491.25 | **6,011.83** |
| **PA - PENNSYLVANIA - Total** | **82,477.79** | **78,034.72** | **80,144.62** | **81,978.34** | **322,635.48** |
| **STATE: PR - PUERTO RICO** | | | | | |
| REGISTRANT ZIP CODE 3 | 1ST QUARTER | 2ND QUARTER | 3RD QUARTER | 4TH QUARTER | **TOTAL GRAMS** |
| 006 | 3,938.40 | 2,665.38 | 4,002.49 | 3,821.26 | **14,427.53** |
| 007 | 3,186.96 | 1,993.51 | 3,179.23 | 3,024.52 | **11,384.23** |
| 009 | 2,173.63 | 1,615.58 | 2,016.72 | 2,158.16 | **7,964.10** |
| **PR - PUERTO RICO - Total** | **9,299.00** | **6,274.47** | **9,198.44** | **9,003.95** | **33,775.85** |
| **STATE: RI - RHODE ISLAND** | | | | | |
| REGISTRANT ZIP CODE 3 | 1ST QUARTER | 2ND QUARTER | 3RD QUARTER | 4TH QUARTER | **TOTAL GRAMS** |
| 028 | 2,680.30 | 2,613.46 | 2,425.39 | 2,647.26 | **10,366.41** |
| 029 | 1,489.59 | 1,453.38 | 1,273.36 | 1,183.65 | **5,399.98** |
| **RI - RHODE ISLAND - Total** | **4,169.89** | **4,066.84** | **3,698.75** | **3,830.91** | **15,766.40** |
| **STATE: SC - SOUTH CAROLINA** | | | | | |
| REGISTRANT ZIP CODE 3 | 1ST QUARTER | 2ND QUARTER | 3RD QUARTER | 4TH QUARTER | **TOTAL GRAMS** |
| 290 | 1,962.35 | 2,140.76 | 1,817.12 | 2,045.67 | **7,965.89** |
| 291 | 2,362.96 | 2,308.15 | 2,099.60 | 2,234.07 | **9,004.78** |
| 292 | 2,471.88 | 2,424.24 | 2,307.79 | 2,462.76 | **9,666.67** |
| 293 | 2,982.40 | 3,031.90 | 2,651.93 | 3,039.64 | **11,705.87** |
| 294 | 28,350.46 | 28,471.46 | 26,489.82 | 32,579.86 | **115,891.60** |
| 295 | 7,621.59 | 7,349.36 | 7,283.45 | 7,391.20 | **29,645.61** |
| 296 | 4,579.24 | 4,575.98 | 4,402.13 | 4,893.06 | **18,450.41** |
| 297 | 1,223.84 | 1,315.56 | 1,079.17 | 1,256.44 | **4,875.00** |
| 298 | 1,668.07 | 1,969.55 | 1,830.38 | 1,991.91 | **7,459.91** |
| 299 | 1,081.10 | 1,019.04 | 837.99 | 920.03 | **3,858.16** |
| **SC - SOUTH CAROLINA - Total** | **54,303.90** | **54,605.99** | **50,799.37** | **58,814.65** | **218,523.90** |
| **STATE: SD - SOUTH DAKOTA** | | | | | |
| REGISTRANT ZIP CODE 3 | 1ST QUARTER | 2ND QUARTER | 3RD QUARTER | 4TH QUARTER | **TOTAL GRAMS** |
| 570 | 739.79 | 597.53 | 607.19 | 654.55 | **2,599.07** |
| 571 | 955.77 | 906.58 | 1,024.79 | 938.41 | **3,825.55** |
| 572 | 222.12 | 333.73 | 333.91 | 298.36 | **1,188.11** |
| 573 | 426.17 | 399.09 | 350.11 | 364.88 | **1,540.25** |
| 574 | 374.05 | 415.50 | 415.50 | 393.58 | **1,598.63** |
| 575 | 234.27 | 132.61 | 171.28 | 179.56 | **717.72** |
| 576 | 165.76 | 172.39 | 125.98 | 172.39 | **636.51** |
| 577 | 1,143.31 | 1,133.68 | 1,028.40 | 1,071.30 | **4,376.70** |
| **SD - SOUTH DAKOTA - Total** | **4,261.24** | **4,091.11** | **4,057.14** | **4,073.04** | **16,482.53** |
| **STATE: TN - TENNESSEE** | | | | | |
| REGISTRANT ZIP CODE 3 | 1ST QUARTER | 2ND QUARTER | 3RD QUARTER | 4TH QUARTER | **TOTAL GRAMS** |
| 370 | 3,674.21 | 3,408.49 | 3,456.37 | 3,733.61 | **14,272.68** |
| 371 | 23,775.28 | 25,002.19 | 22,002.26 | 17,108.31 | **87,888.04** |
| 372 | 1,589.36 | 1,289.43 | 1,120.92 | 1,306.77 | **5,306.48** |
| 373 | 2,564.45 | 2,558.10 | 2,539.33 | 3,162.93 | **10,824.80** |
| 374 | 1,259.20 | 1,279.36 | 1,218.13 | 1,434.16 | **5,190.85** |
| 376 | 2,111.54 | 2,057.11 | 1,596.69 | 1,974.23 | **7,739.58** |
| 377 | 2,098.65 | 1,875.40 | 1,910.62 | 2,277.59 | **8,162.26** |
| 378 | 2,962.80 | 2,794.56 | 2,984.64 | 3,081.17 | **11,823.16** |
| 379 | 1,742.29 | 1,850.02 | 1,660.68 | 1,854.07 | **7,107.06** |

| | | | | | |
|---|---|---|---|---|---|
| 380 | 1,442.89 | 1,435.19 | 1,615.06 | 1,496.51 | **5,989.65** |
| 381 | 2,558.70 | 3,896.99 | 3,318.98 | 3,089.87 | **12,864.54** |
| 382 | 636.51 | 517.16 | 596.73 | 637.61 | **2,388.01** |
| 383 | 2,693.88 | 3,064.61 | 2,920.57 | 3,436.89 | **12,115.95** |
| 384 | 601.75 | 592.49 | 664.50 | 741.67 | **2,600.41** |
| 385 | 1,438.78 | 1,663.10 | 1,717.25 | 1,705.64 | **6,524.76** |
| **TN - TENNESSEE - Total** | **51,150.27** | **53,284.21** | **49,322.72** | **47,041.03** | **200,798.23** |

**STATE: TX - TEXAS**

| REGISTRANT ZIP CODE 3 | 1ST QUARTER | 2ND QUARTER | 3RD QUARTER | 4TH QUARTER | TOTAL GRAMS |
|---|---|---|---|---|---|
| 750 | 52,257.27 | 48,623.68 | 51,265.49 | 50,454.47 | **202,600.91** |
| 751 | 26,005.52 | 24,247.54 | 25,007.58 | 24,020.78 | **99,281.42** |
| 752 | 28,087.98 | 23,368.77 | 25,266.62 | 25,544.99 | **102,268.35** |
| 753 | 486.40 | 345.14 | 429.20 | 398.00 | **1,658.75** |
| 754 | 17,352.03 | 17,269.95 | 17,582.62 | 17,550.39 | **69,754.99** |
| 755 | 7,019.46 | 6,408.18 | 6,507.64 | 6,520.17 | **26,455.46** |
| 756 | 22,726.93 | 21,101.84 | 23,024.81 | 21,828.42 | **88,681.99** |
| 757 | 25,038.39 | 23,898.83 | 24,070.69 | 24,972.62 | **97,980.54** |
| 758 | 3,882.04 | 3,659.93 | 3,804.69 | 3,331.73 | **14,678.38** |
| 759 | 16,373.83 | 15,250.84 | 15,829.09 | 15,047.63 | **62,501.39** |
| 760 | 39,241.68 | 36,279.98 | 37,824.83 | 38,062.15 | **151,408.64** |
| 761 | 26,425.76 | 23,427.64 | 25,154.93 | 24,909.95 | **99,918.28** |
| 762 | 14,264.00 | 13,285.75 | 13,936.82 | 13,948.08 | **55,434.64** |
| 763 | 9,185.46 | 8,887.90 | 9,389.23 | 9,324.22 | **36,786.81** |
| 764 | 6,264.53 | 6,346.48 | 6,636.93 | 6,371.72 | **25,619.66** |
| 765 | 11,284.66 | 10,052.59 | 10,984.36 | 10,154.93 | **42,476.54** |
| 766 | 4,478.78 | 4,271.20 | 4,748.76 | 4,213.92 | **17,712.66** |
| 767 | 6,564.19 | 6,353.12 | 6,365.20 | 6,153.91 | **25,436.41** |
| 768 | 5,260.04 | 4,651.16 | 4,717.55 | 4,521.96 | **19,150.70** |
| 769 | 3,879.83 | 3,519.87 | 3,416.37 | 3,724.20 | **14,540.27** |
| 770 | 60,790.89 | 52,751.31 | 57,236.70 | 56,999.97 | **227,778.87** |
| 772 | 2.21 | 0 | 2.21 | 0 | **4.42** |
| 773 | 33,531.72 | 31,954.67 | 33,077.20 | 33,010.79 | **131,574.38** |
| 774 | 26,475.34 | 24,619.23 | 26,153.56 | 26,158.17 | **103,406.30** |
| 775 | 32,721.44 | 30,489.75 | 31,081.65 | 32,081.24 | **126,374.08** |
| 776 | 12,939.37 | 11,925.70 | 12,272.78 | 12,061.70 | **49,199.55** |
| 777 | 8,411.60 | 7,890.08 | 8,460.34 | 8,181.32 | **32,943.35** |
| 778 | 9,889.66 | 9,368.61 | 10,369.97 | 10,367.58 | **39,995.83** |
| 779 | 4,376.47 | 4,463.04 | 4,333.17 | 4,580.72 | **17,753.39** |
| 780 | 12,588.47 | 10,853.23 | 11,431.15 | 11,567.99 | **46,440.84** |
| 781 | 11,366.62 | 10,540.51 | 11,000.21 | 11,644.00 | **44,551.35** |
| 782 | 55,697.75 | 49,960.68 | 50,571.22 | 50,689.34 | **206,918.98** |
| 783 | 11,024.32 | 11,660.84 | 11,098.28 | 11,000.95 | **44,784.39** |
| 784 | 14,591.46 | 14,631.57 | 14,384.34 | 14,377.21 | **57,984.57** |
| 785 | 24,470.08 | 21,679.74 | 22,054.98 | 22,401.04 | **90,605.84** |
| 786 | 17,643.74 | 15,941.69 | 17,655.32 | 17,733.87 | **68,974.62** |
| 787 | 13,472.83 | 11,923.53 | 12,572.79 | 12,971.87 | **50,941.03** |
| 788 | 3,456.78 | 3,203.54 | 3,310.17 | 3,596.20 | **13,566.69** |
| 789 | 1,688.88 | 1,483.16 | 1,479.84 | 1,458.67 | **6,110.55** |
| 790 | 7,433.40 | 7,124.61 | 7,181.15 | 7,066.58 | **28,805.73** |
| 791 | 12,028.93 | 10,870.43 | 12,073.15 | 10,785.29 | **45,757.79** |
| 792 | 968.57 | 918.10 | 1,129.73 | 876.30 | **3,892.70** |
| 793 | 4,893.20 | 4,435.67 | 4,709.72 | 4,475.53 | **18,514.12** |
| 794 | 12,554.73 | 11,631.85 | 12,590.47 | 12,038.92 | **48,815.97** |
| 795 | 2,969.27 | 2,682.62 | 2,944.22 | 2,765.94 | **11,362.05** |
| 796 | 9,594.23 | 8,764.15 | 9,220.54 | 8,927.88 | **36,506.80** |
| 797 | 17,687.60 | 16,716.63 | 16,498.84 | 16,712.36 | **67,615.43** |
| 798 | 1,076.50 | 1,156.99 | 993.44 | 1,039.85 | **4,266.78** |
| 799 | 19,046.05 | 16,886.56 | 18,412.55 | 16,326.70 | **70,671.86** |
| **TX - TEXAS - Total** | **769,470.88** | **707,778.85** | **740,263.11** | **732,952.20** | **2,950,465.05** |

**STATE: UT - UTAH**

| REGISTRANT ZIP CODE 3 | 1ST QUARTER | 2ND QUARTER | 3RD QUARTER | 4TH QUARTER | TOTAL GRAMS |
|---|---|---|---|---|---|
| 840 | 5,359.83 | 5,256.28 | 5,385.34 | 5,326.07 | **21,327.53** |
| 841 | 2,420.83 | 2,252.33 | 2,287.77 | 2,196.31 | **9,157.23** |
| 843 | 587.69 | 603.70 | 448.65 | 532.00 | **2,172.04** |
| 844 | 577.20 | 460.07 | 745.17 | 513.85 | **2,296.28** |
| 845 | 346.99 | 424.70 | 406.66 | 321.75 | **1,500.10** |
| 846 | 1,522.90 | 1,537.12 | 1,653.34 | 1,675.62 | **6,388.98** |
| 847 | 1,224.47 | 1,215.72 | 1,305.68 | 1,410.10 | **5,155.97** |
| **UT - UTAH - Total** | **12,039.91** | **11,749.93** | **12,232.60** | **11,975.70** | **47,998.14** |

**STATE: VA - VIRGINIA**

| REGISTRANT ZIP CODE 3 | 1ST QUARTER | 2ND QUARTER | 3RD QUARTER | 4TH QUARTER | TOTAL GRAMS |
|---|---|---|---|---|---|
| 201 | 4,186.36 | 4,623.16 | 4,513.65 | 4,487.22 | **17,810.39** |
| 220 | 2,440.81 | 2,292.17 | 2,089.18 | 2,358.29 | **9,180.46** |
| 221 | 2,352.54 | 2,150.75 | 1,996.71 | 1,692.07 | **8,192.06** |
| 222 | 767.63 | 777.11 | 791.57 | 816.70 | **3,153.01** |
| 223 | 1,420.72 | 1,242.69 | 1,213.12 | 1,239.23 | **5,115.76** |
| 224 | 1,700.11 | 1,785.03 | 1,522.40 | 1,843.09 | **6,850.63** |
| 225 | 737.54 | 709.77 | 789.01 | 708.11 | **2,944.43** |
| 226 | 913.60 | 930.99 | 796.27 | 924.37 | **3,565.23** |
| 227 | 353.97 | 267.41 | 272.39 | 279.20 | **1,172.97** |
| 228 | 806.83 | 848.74 | 770.75 | 826.84 | **3,253.17** |
| 229 | 949.38 | 1,002.45 | 962.02 | 1,015.25 | **3,929.11** |
| 230 | 1,033.86 | 864.50 | 858.88 | 942.41 | **3,699.66** |
| 231 | 1,437.72 | 1,655.08 | 1,429.89 | 1,493.76 | **6,016.46** |
| 232 | 3,031.91 | 2,683.42 | 2,302.80 | 2,877.65 | **10,895.79** |
| 233 | 1,168.38 | 1,141.84 | 1,087.33 | 1,179.56 | **4,577.10** |
| 234 | 2,603.15 | 2,799.49 | 2,870.34 | 2,951.41 | **11,224.39** |
| 235 | 876.49 | 1,014.98 | 868.71 | 1,001.99 | **3,762.18** |
| 236 | 1,838.02 | 2,036.19 | 1,805.97 | 1,740.46 | **7,420.63** |
| 237 | 432.96 | 438.51 | 516.54 | 420.06 | **1,808.07** |
| 238 | 1,745.06 | 1,513.89 | 1,421.43 | 1,557.96 | **6,238.34** |
| 239 | 418.81 | 395.97 | 308.67 | 402.24 | **1,525.70** |
| 240 | 1,070.34 | 906.81 | 908.42 | 870.99 | **3,756.56** |
| 241 | 996.95 | 808.22 | 659.52 | 837.07 | **3,301.76** |
| 242 | 1,016.79 | 1,294.46 | 1,107.18 | 1,317.84 | **4,736.27** |
| 243 | 716.80 | 685.68 | 600.86 | 549.29 | **2,552.63** |
| 244 | 430.03 | 450.30 | 422.66 | 555.83 | **1,858.83** |
| 245 | 1,384.80 | 1,150.30 | 1,064.23 | 1,186.36 | **4,785.69** |
| 246 | 942.01 | 920.69 | 858.62 | 1,004.98 | **3,726.31** |
| **VA - VIRGINIA - Total** | **37,773.56** | **37,390.60** | **34,809.14** | **37,080.24** | **147,053.55** |

**STATE: VI - VIRGIN ISLANDS**

| REGISTRANT ZIP CODE 3 | 1ST QUARTER | 2ND QUARTER | 3RD QUARTER | 4TH QUARTER | TOTAL GRAMS |
|---|---|---|---|---|---|
| 008 | 644.24 | 537.05 | 690.66 | 539.26 | **2,411.22** |
| **VI - VIRGIN ISLANDS - Total** | **644.24** | **537.05** | **690.66** | **539.26** | **2,411.22** |

**STATE: VT - VERMONT**

| REGISTRANT ZIP CODE 3 | 1ST QUARTER | 2ND QUARTER | 3RD QUARTER | 4TH QUARTER | TOTAL GRAMS |
|---|---|---|---|---|---|
| 050 | 130.55 | 233.62 | 172.42 | 124.63 | **661.22** |
| 051 | 106.08 | 102.58 | 106.63 | 91.78 | **407.08** |
| 052 | 132.97 | 123.48 | 223.77 | 141.43 | **621.65** |
| 053 | 105.15 | 64.99 | 181.39 | 95.39 | **446.91** |
| 054 | 1,084.72 | 756.78 | 970.04 | 946.29 | **3,757.83** |
| 056 | 323.04 | 321.19 | 318.25 | 265.94 | **1,228.43** |
| 057 | 239.42 | 208.02 | 316.23 | 255.63 | **1,019.30** |
| 058 | 196.13 | 244.37 | 213.03 | 161.32 | **814.85** |
| **VT - VERMONT - Total** | **2,318.06** | **2,055.04** | **2,501.75** | **2,082.42** | **8,957.27** |

**STATE: WA - WASHINGTON**

| REGISTRANT ZIP CODE 3 | 1ST QUARTER | 2ND QUARTER | 3RD QUARTER | 4TH QUARTER | TOTAL GRAMS |
|---|---|---|---|---|---|
| 980 | 7,794.97 | 7,999.95 | 8,202.77 | 7,542.46 | **31,540.15** |
| 981 | 4,799.90 | 4,631.57 | 3,981.80 | 4,127.16 | **17,540.44** |
| 982 | 5,726.97 | 5,618.48 | 5,560.06 | 5,276.77 | **22,182.28** |

| | | | | | |
|---|---|---|---|---|---|
| 983 | 4,469.90 | 4,178.53 | 4,243.80 | 4,192.33 | **17,084.56** |
| 984 | 2,703.82 | 2,604.73 | 2,496.84 | 2,429.01 | **10,234.40** |
| 985 | 2,760.23 | 2,398.60 | 2,959.26 | 2,841.97 | **10,960.06** |
| 986 | 3,522.03 | 3,227.01 | 3,317.74 | 3,316.13 | **13,382.93** |
| 988 | 1,067.40 | 1,253.04 | 1,142.18 | 1,151.39 | **4,614.01** |
| 989 | 1,340.09 | 1,370.98 | 1,499.54 | 1,472.80 | **5,683.41** |
| 990 | 991.58 | 818.09 | 1,031.36 | 892.67 | **3,733.71** |
| 991 | 891.75 | 878.22 | 822.48 | 1,019.46 | **3,611.91** |
| 992 | 4,025.19 | 3,967.05 | 4,210.94 | 3,637.81 | **15,840.99** |
| 993 | 1,691.73 | 1,679.39 | 1,849.30 | 1,819.17 | **7,039.59** |
| 994 | 525.45 | 474.61 | 432.26 | 446.80 | **1,879.12** |
| **WA - WASHINGTON - Total** | **42,311.00** | **41,100.27** | **41,750.34** | **40,165.95** | **165,327.56** |

**STATE: WI - WISCONSIN**

| REGISTRANT ZIP CODE 3 | 1ST QUARTER | 2ND QUARTER | 3RD QUARTER | 4TH QUARTER | TOTAL GRAMS |
|---|---|---|---|---|---|
| 530 | 3,580.38 | 3,272.60 | 3,572.23 | 3,217.91 | **13,643.11** |
| 531 | 4,034.03 | 3,776.62 | 4,354.98 | 3,980.28 | **16,145.91** |
| 532 | 8,226.87 | 7,343.68 | 7,210.51 | 6,391.46 | **29,172.51** |
| 534 | 1,138.01 | 1,065.06 | 1,214.44 | 1,178.34 | **4,595.84** |
| 535 | 3,183.32 | 2,903.11 | 2,881.92 | 3,266.20 | **12,234.55** |
| 537 | 1,259.69 | 1,198.78 | 1,347.63 | 1,267.93 | **5,074.02** |
| 538 | 255.03 | 273.68 | 318.95 | 300.94 | **1,148.60** |
| 539 | 1,053.24 | 1,168.12 | 1,173.91 | 1,078.73 | **4,474.01** |
| 540 | 554.92 | 501.50 | 499.11 | 460.99 | **2,016.51** |
| 541 | 971.04 | 935.90 | 848.86 | 973.52 | **3,729.32** |
| 542 | 653.84 | 736.21 | 621.66 | 697.17 | **2,708.87** |
| 543 | 1,149.34 | 1,076.46 | 1,351.73 | 1,298.62 | **4,876.15** |
| 544 | 1,759.78 | 1,965.10 | 1,825.21 | 1,728.49 | **7,278.58** |
| 545 | 772.51 | 859.13 | 713.08 | 677.56 | **3,022.28** |
| 546 | 849.96 | 723.42 | 698.95 | 758.77 | **3,031.10** |
| 547 | 1,041.37 | 1,133.39 | 1,104.91 | 1,176.41 | **4,456.09** |
| 548 | 1,133.24 | 1,338.82 | 1,357.65 | 1,366.11 | **5,195.83** |
| 549 | 2,618.54 | 2,410.35 | 2,584.12 | 2,518.25 | **10,131.26** |
| **WI - WISCONSIN - Total** | **34,235.11** | **32,681.92** | **33,679.83** | **32,337.66** | **132,934.53** |

**STATE: WV - WEST VIRGINIA**

| REGISTRANT ZIP CODE 3 | 1ST QUARTER | 2ND QUARTER | 3RD QUARTER | 4TH QUARTER | TOTAL GRAMS |
|---|---|---|---|---|---|
| 247 | 1,021.61 | 823.44 | 827.12 | 755.58 | **3,427.76** |
| 248 | 3,030.05 | 2,871.10 | 3,240.19 | 3,158.23 | **12,299.57** |
| 249 | 392.66 | 338.15 | 304.99 | 309.41 | **1,345.21** |
| 250 | 1,207.00 | 1,274.41 | 1,050.90 | 1,233.24 | **4,765.55** |
| 251 | 2,102.90 | 2,113.95 | 2,095.90 | 2,149.31 | **8,462.06** |
| 252 | 1,048.87 | 1,353.31 | 1,129.54 | 1,287.57 | **4,819.29** |
| 253 | 2,447.88 | 2,298.04 | 2,309.74 | 2,352.30 | **9,407.96** |
| 254 | 1,392.36 | 1,301.31 | 1,320.72 | 1,518.61 | **5,533.00** |
| 255 | 2,355.88 | 2,499.62 | 2,520.62 | 2,481.50 | **9,857.62** |
| 256 | 5,912.02 | 5,724.16 | 6,140.76 | 5,965.06 | **23,742.00** |
| 257 | 996.74 | 950.10 | 1,109.50 | 1,002.45 | **4,058.79** |
| 258 | 2,062.02 | 1,597.15 | 1,922.41 | 1,920.76 | **7,502.34** |
| 259 | 934.31 | 713.85 | 1,010.37 | 914.77 | **3,573.31** |
| 260 | 1,015.15 | 1,164.14 | 1,150.15 | 952.42 | **4,281.87** |
| 261 | 1,362.33 | 1,293.27 | 1,374.49 | 1,256.81 | **5,286.90** |
| 262 | 1,197.13 | 1,389.76 | 1,342.81 | 1,270.25 | **5,199.95** |
| 263 | 1,066.00 | 1,035.61 | 1,014.42 | 1,005.22 | **4,121.25** |
| 264 | 397.82 | 381.24 | 434.28 | 409.97 | **1,623.32** |
| 265 | 2,806.48 | 2,591.38 | 2,515.19 | 2,588.21 | **10,501.25** |
| 266 | 926.03 | 700.60 | 921.61 | 970.23 | **3,518.48** |
| 267 | 408.12 | 451.21 | 503.15 | 385.65 | **1,748.13** |
| 268 | 245.32 | 190.07 | 225.43 | 195.59 | **856.41** |
| **WV - WEST VIRGINIA - Total** | **34,328.68** | **33,055.88** | **34,464.29** | **34,083.16** | **135,932.01** |

**STATE: WY - WYOMING**

| REGISTRANT ZIP CODE 3 | 1ST QUARTER | 2ND QUARTER | 3RD QUARTER | 4TH QUARTER | TOTAL GRAMS |
|---|---|---|---|---|---|

| | | | | | |
|---|---|---|---|---|---|
| 820 | 585.86 | 602.36 | 612.00 | 739.06 | **2,539.28** |
| 822 | 119.35 | 72.93 | 106.08 | 81.77 | **380.14** |
| 823 | 50.83 | 17.68 | 39.78 | 4.42 | **112.72** |
| 824 | 525.16 | 497.61 | 493.83 | 384.90 | **1,901.50** |
| 825 | 256.37 | 236.48 | 162.44 | 254.16 | **909.46** |
| 826 | 401.95 | 452.12 | 461.70 | 496.61 | **1,812.39** |
| 827 | 264.65 | 199.82 | 251.38 | 212.52 | **928.37** |
| 828 | 186.17 | 166.81 | 178.42 | 239.39 | **770.80** |
| 829 | 293.94 | 377.93 | 380.14 | 348.09 | **1,400.10** |
| 830 | 113.26 | 97.24 | 130.40 | 103.87 | **444.78** |
| 831 | 40.22 | 41.99 | 52.16 | 39.78 | **174.16** |
| **WY - WYOMING - Total** | **2,837.77** | **2,762.98** | **2,868.34** | **2,904.58** | **11,373.68** |

**DRUG: 9064 - BUPRENORPHINE**

**STATE: AK - ALASKA**

| REGISTRANT ZIP CODE 3 | 1ST QUARTER | 2ND QUARTER | 3RD QUARTER | 4TH QUARTER | TOTAL GRAMS |
|---|---|---|---|---|---|
| 995 | 1,098.55 | 1,114.05 | 1,166.13 | 1,264.80 | **4,643.52** |
| 996 | 754.12 | 751.88 | 779.70 | 793.77 | **3,079.48** |
| 997 | 163.39 | 155.68 | 176.73 | 180.35 | **676.15** |
| 998 | 364.74 | 332.11 | 369.71 | 369.79 | **1,436.35** |
| 999 | 307.05 | 309.82 | 321.61 | 350.23 | **1,288.71** |
| **AK - ALASKA - Total** | **2,687.85** | **2,663.53** | **2,813.89** | **2,958.94** | **11,124.21** |

**STATE: AL - ALABAMA**

| REGISTRANT ZIP CODE 3 | 1ST QUARTER | 2ND QUARTER | 3RD QUARTER | 4TH QUARTER | TOTAL GRAMS |
|---|---|---|---|---|---|
| 350 | 4,631.14 | 4,982.64 | 5,194.80 | 5,052.25 | **19,860.82** |
| 351 | 2,660.44 | 2,726.18 | 2,906.44 | 2,936.40 | **11,229.46** |
| 352 | 3,827.21 | 3,695.74 | 3,724.32 | 3,885.37 | **15,132.65** |
| 354 | 762.04 | 836.69 | 882.94 | 839.55 | **3,321.22** |
| 355 | 2,575.82 | 2,697.82 | 2,827.71 | 2,806.49 | **10,907.84** |
| 356 | 3,923.58 | 4,276.50 | 4,344.55 | 4,382.01 | **16,926.63** |
| 357 | 1,181.98 | 1,254.19 | 1,294.11 | 1,289.24 | **5,019.52** |
| 358 | 1,406.14 | 1,461.15 | 1,492.37 | 1,526.79 | **5,886.45** |
| 359 | 3,501.84 | 3,789.06 | 3,677.69 | 3,751.65 | **14,720.24** |
| 360 | 889.66 | 1,014.57 | 977.30 | 986.32 | **3,867.86** |
| 361 | 516.79 | 523.08 | 549.73 | 583.37 | **2,172.97** |
| 362 | 1,358.47 | 1,435.22 | 1,489.23 | 1,532.43 | **5,815.34** |
| 363 | 1,035.65 | 1,087.55 | 1,093.46 | 1,146.54 | **4,363.19** |
| 364 | 467.20 | 467.34 | 487.55 | 539.78 | **1,961.86** |
| 365 | 2,229.64 | 2,381.16 | 2,563.37 | 2,576.85 | **9,751.03** |
| 366 | 2,030.40 | 2,046.82 | 2,144.06 | 2,362.43 | **8,583.71** |
| 367 | 272.78 | 276.58 | 273.95 | 293.48 | **1,116.79** |
| 368 | 670.25 | 745.33 | 749.51 | 731.82 | **2,896.91** |
| 369 | 34.11 | 39.70 | 41.91 | 32.31 | **148.03** |
| **AL - ALABAMA - Total** | **33,975.14** | **35,737.30** | **36,715.00** | **37,255.07** | **143,682.51** |

**STATE: AR - ARKANSAS**

| REGISTRANT ZIP CODE 3 | 1ST QUARTER | 2ND QUARTER | 3RD QUARTER | 4TH QUARTER | TOTAL GRAMS |
|---|---|---|---|---|---|
| 716 | 208.35 | 265.57 | 287.90 | 314.92 | **1,076.74** |
| 717 | 313.10 | 332.70 | 347.85 | 367.41 | **1,361.06** |
| 718 | 253.30 | 261.76 | 284.74 | 280.73 | **1,080.53** |
| 719 | 1,060.92 | 1,148.32 | 1,199.40 | 1,182.79 | **4,591.43** |
| 720 | 889.54 | 1,039.52 | 1,064.65 | 1,091.63 | **4,085.34** |
| 721 | 1,353.31 | 1,439.67 | 1,438.77 | 1,480.83 | **5,712.58** |
| 722 | 1,070.39 | 951.96 | 1,006.58 | 968.32 | **3,997.26** |
| 723 | 165.48 | 202.14 | 197.87 | 212.65 | **778.13** |
| 724 | 1,211.61 | 1,285.19 | 1,361.75 | 1,431.82 | **5,290.38** |
| 725 | 418.90 | 477.16 | 511.58 | 533.55 | **1,941.19** |
| 726 | 545.73 | 623.84 | 624.94 | 638.60 | **2,433.10** |
| 727 | 1,379.22 | 1,502.31 | 1,629.48 | 1,656.55 | **6,167.56** |
| 728 | 264.03 | 294.62 | 348.56 | 370.38 | **1,277.60** |
| 729 | 775.12 | 820.84 | 958.60 | 983.46 | **3,538.03** |
| **AR - ARKANSAS - Total** | **9,909.01** | **10,645.60** | **11,262.67** | **11,513.63** | **43,330.91** |

STATE: AZ - ARIZONA

| REGISTRANT ZIP CODE 3 | 1ST QUARTER | 2ND QUARTER | 3RD QUARTER | 4TH QUARTER | TOTAL GRAMS |
|---|---|---|---|---|---|
| 850 | 4,313.73 | 4,135.35 | 4,293.31 | 4,299.46 | **17,041.84** |
| 851 | 820.35 | 757.53 | 793.04 | 789.49 | **3,160.42** |
| 852 | 3,518.87 | 3,413.93 | 3,591.96 | 3,783.69 | **14,308.45** |
| 853 | 2,396.92 | 2,434.00 | 2,557.43 | 2,580.07 | **9,968.41** |
| 855 | 215.66 | 228.23 | 232.10 | 238.51 | **914.50** |
| 856 | 244.79 | 230.98 | 251.96 | 276.16 | **1,003.89** |
| 857 | 2,515.78 | 2,429.12 | 2,429.56 | 2,547.33 | **9,921.79** |
| 859 | 222.31 | 246.57 | 265.05 | 288.64 | **1,022.57** |
| 860 | 218.36 | 212.71 | 213.21 | 239.09 | **883.38** |
| 863 | 981.05 | 868.64 | 920.85 | 1,020.25 | **3,790.78** |
| 864 | 401.07 | 404.31 | 380.91 | 406.50 | **1,592.79** |
| 865 | 44.28 | 45.78 | 41.76 | 36.60 | **168.42** |
| **AZ - ARIZONA - Total** | **15,893.16** | **15,407.16** | **15,971.14** | **16,505.78** | **63,777.24** |

STATE: CA - CALIFORNIA

| REGISTRANT ZIP CODE 3 | 1ST QUARTER | 2ND QUARTER | 3RD QUARTER | 4TH QUARTER | TOTAL GRAMS |
|---|---|---|---|---|---|
| 900 | 1,378.10 | 1,325.11 | 1,392.80 | 1,522.40 | **5,618.40** |
| 902 | 1,082.01 | 1,060.11 | 1,042.92 | 1,049.01 | **4,234.04** |
| 903 | 36.52 | 43.52 | 47.81 | 51.38 | **179.23** |
| 904 | 253.38 | 244.16 | 226.21 | 216.80 | **940.55** |
| 905 | 233.82 | 208.82 | 240.74 | 248.84 | **932.22** |
| 906 | 322.33 | 297.26 | 337.12 | 346.49 | **1,303.20** |
| 907 | 889.68 | 831.70 | 851.60 | 858.18 | **3,431.15** |
| 908 | 359.68 | 373.36 | 378.90 | 390.52 | **1,502.45** |
| 910 | 124.64 | 117.58 | 121.20 | 125.48 | **488.91** |
| 911 | 186.11 | 179.08 | 180.85 | 198.33 | **744.37** |
| 912 | 310.80 | 312.81 | 311.95 | 326.94 | **1,262.49** |
| 913 | 1,622.21 | 1,584.18 | 1,736.75 | 1,729.49 | **6,672.63** |
| 914 | 722.81 | 647.83 | 679.07 | 671.39 | **2,721.09** |
| 915 | 169.07 | 151.66 | 143.18 | 136.13 | **600.04** |
| 916 | 194.24 | 207.31 | 225.29 | 221.81 | **848.66** |
| 917 | 1,139.93 | 1,089.49 | 1,205.39 | 1,301.38 | **4,736.18** |
| 918 | 16.32 | 20.34 | 19.11 | 18.57 | **74.33** |
| 919 | 409.92 | 413.46 | 399.11 | 439.35 | **1,661.84** |
| 920 | 1,997.44 | 1,943.86 | 1,896.27 | 1,908.19 | **7,745.76** |
| 921 | 1,851.00 | 1,863.14 | 1,984.20 | 2,456.75 | **8,155.10** |
| 922 | 911.81 | 1,301.63 | 1,535.38 | 2,342.12 | **6,090.94** |
| 923 | 986.31 | 989.84 | 1,180.33 | 1,221.35 | **4,377.83** |
| 924 | 118.16 | 127.42 | 150.89 | 180.43 | **576.90** |
| 925 | 2,054.56 | 1,972.10 | 2,057.13 | 2,124.40 | **8,208.19** |
| 926 | 2,164.08 | 2,163.47 | 2,232.13 | 2,290.34 | **8,850.02** |
| 927 | 1,277.18 | 1,119.80 | 1,314.20 | 1,475.39 | **5,186.58** |
| 928 | 1,132.69 | 1,670.95 | 1,324.16 | 1,497.00 | **5,624.80** |
| 930 | 910.63 | 903.53 | 984.27 | 1,024.54 | **3,822.96** |
| 931 | 578.89 | 488.45 | 507.23 | 478.88 | **2,053.45** |
| 932 | 861.02 | 1,571.92 | 2,211.64 | 3,397.48 | **8,042.06** |
| 933 | 501.62 | 515.79 | 525.31 | 580.50 | **2,123.23** |
| 934 | 1,040.61 | 1,100.76 | 1,203.23 | 1,229.82 | **4,574.42** |
| 935 | 715.50 | 965.25 | 1,458.82 | 1,870.34 | **5,009.91** |
| 936 | 228.60 | 314.84 | 407.04 | 477.20 | **1,427.68** |
| 937 | 491.84 | 454.49 | 525.05 | 521.56 | **1,992.93** |
| 939 | 1,045.91 | 1,196.92 | 1,433.36 | 1,640.31 | **5,316.51** |
| 940 | 928.80 | 843.01 | 879.34 | 912.34 | **3,563.50** |
| 941 | 1,727.14 | 1,754.99 | 1,592.04 | 1,685.40 | **6,759.57** |
| 943 | 187.57 | 211.13 | 238.74 | 219.70 | **857.15** |
| 944 | 231.95 | 223.30 | 224.89 | 227.19 | **907.34** |
| 945 | 4,498.59 | 4,441.11 | 4,570.82 | 4,595.06 | **18,105.57** |
| 946 | 724.24 | 748.61 | 714.18 | 751.67 | **2,938.71** |
| 947 | 161.57 | 136.38 | 133.08 | 150.30 | **581.33** |

| | | | | | |
|---|---|---|---|---|---|
| 948 | 195.88 | 171.15 | 184.20 | 169.41 | **720.64** |
| 949 | 1,206.45 | 1,200.69 | 1,211.73 | 1,219.76 | **4,838.63** |
| 950 | 1,567.35 | 1,478.82 | 1,565.16 | 1,650.57 | **6,261.90** |
| 951 | 974.91 | 949.16 | 956.50 | 1,008.73 | **3,889.30** |
| 952 | 544.99 | 560.97 | 654.46 | 774.82 | **2,535.25** |
| 953 | 1,657.29 | 1,831.59 | 1,954.77 | 2,149.12 | **7,592.77** |
| 954 | 2,944.84 | 2,940.66 | 2,998.69 | 3,035.36 | **11,919.55** |
| 955 | 1,939.89 | 1,978.62 | 2,013.76 | 2,065.62 | **7,997.89** |
| 956 | 3,579.38 | 3,810.33 | 4,461.29 | 5,113.23 | **16,964.23** |
| 957 | 414.07 | 457.94 | 514.33 | 558.48 | **1,944.81** |
| 958 | 3,080.39 | 3,285.56 | 3,235.07 | 3,357.53 | **12,958.54** |
| 959 | 2,446.03 | 2,423.05 | 2,574.60 | 2,690.07 | **10,133.76** |
| 960 | 1,612.72 | 1,676.94 | 1,713.14 | 1,771.70 | **6,774.49** |
| 961 | 379.01 | 526.78 | 803.74 | 794.73 | **2,504.25** |
| **CA - CALIFORNIA - Total** | **59,322.45** | **61,422.71** | **65,665.16** | **71,469.89** | **257,880.21** |

**STATE: CO - COLORADO**

| REGISTRANT ZIP CODE 3 | 1ST QUARTER | 2ND QUARTER | 3RD QUARTER | 4TH QUARTER | TOTAL GRAMS |
|---|---|---|---|---|---|
| 800 | 1,889.69 | 1,795.19 | 1,810.29 | 1,875.53 | **7,370.71** |
| 801 | 983.45 | 1,008.49 | 1,012.73 | 1,098.42 | **4,103.09** |
| 802 | 2,749.84 | 2,585.93 | 2,681.07 | 2,713.07 | **10,729.91** |
| 803 | 187.77 | 160.07 | 151.03 | 171.94 | **670.80** |
| 804 | 187.37 | 201.89 | 190.88 | 219.55 | **799.68** |
| 805 | 1,759.41 | 1,607.50 | 1,615.37 | 1,625.64 | **6,607.93** |
| 806 | 648.07 | 636.53 | 668.92 | 711.51 | **2,665.04** |
| 807 | 91.57 | 68.20 | 71.58 | 85.50 | **316.85** |
| 808 | 134.15 | 157.82 | 162.89 | 178.19 | **633.05** |
| 809 | 1,415.23 | 1,428.37 | 1,477.93 | 1,587.50 | **5,909.03** |
| 810 | 1,180.27 | 1,104.46 | 1,186.59 | 1,204.80 | **4,676.11** |
| 811 | 164.11 | 195.93 | 185.46 | 188.88 | **734.38** |
| 812 | 153.80 | 170.50 | 199.65 | 194.19 | **718.14** |
| 813 | 172.22 | 142.65 | 161.99 | 171.41 | **648.27** |
| 814 | 74.88 | 84.89 | 83.26 | 117.82 | **360.85** |
| 815 | 350.29 | 272.56 | 348.43 | 333.07 | **1,304.36** |
| 816 | 163.72 | 166.97 | 183.70 | 189.03 | **703.43** |
| **CO - COLORADO - Total** | **12,305.85** | **11,787.94** | **12,191.77** | **12,666.06** | **48,951.62** |

**STATE: CT - CONNECTICUT**

| REGISTRANT ZIP CODE 3 | 1ST QUARTER | 2ND QUARTER | 3RD QUARTER | 4TH QUARTER | TOTAL GRAMS |
|---|---|---|---|---|---|
| 060 | 3,587.49 | 3,763.98 | 3,688.63 | 3,792.88 | **14,832.97** |
| 061 | 1,877.90 | 1,745.19 | 1,627.53 | 1,691.01 | **6,941.64** |
| 062 | 1,045.05 | 999.28 | 962.46 | 994.84 | **4,001.63** |
| 063 | 2,285.80 | 2,279.21 | 2,258.54 | 2,428.27 | **9,251.83** |
| 064 | 4,437.32 | 4,647.05 | 4,528.11 | 4,627.74 | **18,240.22** |
| 065 | 2,402.45 | 2,267.70 | 2,296.95 | 2,335.24 | **9,302.34** |
| 066 | 425.28 | 411.90 | 428.15 | 458.40 | **1,723.73** |
| 067 | 2,048.42 | 2,099.81 | 2,088.42 | 2,066.60 | **8,303.25** |
| 068 | 889.25 | 922.46 | 888.12 | 902.37 | **3,602.21** |
| 069 | 161.83 | 167.00 | 156.14 | 155.71 | **640.67** |
| **CT - CONNECTICUT - Total** | **19,160.79** | **19,303.56** | **18,923.07** | **19,453.06** | **76,840.48** |

**STATE: DC - DISTRICT OF COLUMBIA**

| REGISTRANT ZIP CODE 3 | 1ST QUARTER | 2ND QUARTER | 3RD QUARTER | 4TH QUARTER | TOTAL GRAMS |
|---|---|---|---|---|---|
| 200 | 3,595.35 | 3,574.93 | 3,888.96 | 3,953.33 | **15,012.57** |
| 204 | 42.02 | 32.96 | 40.90 | 46.79 | **162.67** |
| **DC - DISTRICT OF COLUMBIA - Total** | **3,637.37** | **3,607.89** | **3,929.87** | **4,000.11** | **15,175.24** |

**STATE: DE - DELAWARE**

| REGISTRANT ZIP CODE 3 | 1ST QUARTER | 2ND QUARTER | 3RD QUARTER | 4TH QUARTER | TOTAL GRAMS |
|---|---|---|---|---|---|
| 197 | 1,476.02 | 1,454.23 | 1,458.43 | 1,488.48 | **5,877.17** |
| 198 | 1,112.51 | 1,149.63 | 1,143.99 | 1,144.84 | **4,550.97** |
| 199 | 2,050.65 | 2,112.79 | 2,197.16 | 2,137.91 | **8,498.52** |
| **DE - DELAWARE - Total** | **4,639.19** | **4,716.66** | **4,799.58** | **4,771.24** | **18,926.66** |

**STATE: FL - FLORIDA**

| REGISTRANT ZIP CODE 3 | 1ST QUARTER | 2ND QUARTER | 3RD QUARTER | 4TH QUARTER | TOTAL GRAMS |
|---|---|---|---|---|---|
| 320 | 3,907.55 | 4,340.06 | 4,404.14 | 4,486.59 | **17,138.34** |
| 321 | 3,227.50 | 3,362.29 | 3,465.50 | 3,511.95 | **13,567.24** |
| 322 | 5,217.20 | 5,106.78 | 5,021.90 | 5,072.01 | **20,417.89** |
| 323 | 1,012.42 | 1,062.13 | 1,082.27 | 1,122.17 | **4,278.99** |
| 324 | 1,208.29 | 1,255.61 | 1,340.86 | 1,371.28 | **5,176.05** |
| 325 | 3,196.24 | 3,017.92 | 3,210.38 | 3,375.16 | **12,799.70** |
| 326 | 3,083.53 | 3,310.37 | 3,224.49 | 3,447.83 | **13,066.23** |
| 327 | 2,878.89 | 2,946.75 | 2,961.72 | 3,110.26 | **11,897.62** |
| 328 | 1,912.61 | 1,950.66 | 2,101.72 | 2,060.03 | **8,025.02** |
| 329 | 3,847.70 | 3,801.92 | 4,090.32 | 4,245.73 | **15,985.66** |
| 330 | 1,775.55 | 1,771.49 | 1,811.83 | 1,808.04 | **7,166.91** |
| 331 | 1,127.17 | 1,155.70 | 1,062.31 | 1,175.50 | **4,520.69** |
| 333 | 1,572.19 | 1,484.06 | 1,506.77 | 1,423.38 | **5,986.39** |
| 334 | 4,814.51 | 4,615.46 | 4,660.69 | 4,993.15 | **19,083.81** |
| 335 | 1,858.92 | 1,854.79 | 1,918.38 | 1,958.53 | **7,590.63** |
| 336 | 1,726.65 | 1,699.39 | 1,742.22 | 1,790.92 | **6,959.18** |
| 337 | 2,739.01 | 2,732.42 | 2,687.13 | 2,850.25 | **11,008.80** |
| 338 | 1,426.51 | 1,515.82 | 1,557.61 | 1,681.10 | **6,181.03** |
| 339 | 3,113.57 | 3,060.51 | 3,229.35 | 3,298.00 | **12,701.43** |
| 341 | 1,563.85 | 1,429.63 | 1,343.32 | 1,421.83 | **5,758.62** |
| 342 | 4,132.97 | 4,226.69 | 4,366.96 | 4,586.61 | **17,313.23** |
| 344 | 1,747.07 | 1,807.34 | 1,904.65 | 2,033.47 | **7,492.53** |
| 346 | 2,608.26 | 2,739.03 | 2,738.92 | 2,941.62 | **11,027.84** |
| 347 | 1,589.72 | 1,574.43 | 1,618.10 | 1,678.62 | **6,460.86** |
| 349 | 1,972.58 | 1,949.63 | 2,032.09 | 2,019.65 | **7,973.95** |
| **FL - FLORIDA - Total** | **63,260.47** | **63,770.88** | **65,083.61** | **67,463.68** | **259,578.64** |

**STATE: GA - GEORGIA**

| REGISTRANT ZIP CODE 3 | 1ST QUARTER | 2ND QUARTER | 3RD QUARTER | 4TH QUARTER | TOTAL GRAMS |
|---|---|---|---|---|---|
| 300 | 2,947.51 | 3,035.26 | 3,073.40 | 3,201.56 | **12,257.72** |
| 301 | 2,873.93 | 3,126.86 | 3,166.40 | 3,335.41 | **12,502.61** |
| 302 | 1,819.73 | 1,987.00 | 1,971.71 | 2,074.65 | **7,853.10** |
| 303 | 974.49 | 915.80 | 941.99 | 914.59 | **3,746.88** |
| 304 | 832.68 | 913.21 | 915.45 | 938.90 | **3,600.24** |
| 305 | 1,344.90 | 1,526.77 | 1,536.08 | 1,604.39 | **6,012.14** |
| 306 | 1,136.75 | 1,235.01 | 1,215.91 | 1,319.67 | **4,907.34** |
| 307 | 699.45 | 701.28 | 759.46 | 832.24 | **2,992.43** |
| 308 | 149.58 | 141.24 | 129.54 | 137.51 | **557.87** |
| 309 | 444.19 | 441.01 | 376.47 | 429.32 | **1,690.99** |
| 310 | 1,322.62 | 1,424.97 | 1,432.47 | 1,466.61 | **5,646.67** |
| 312 | 491.20 | 516.65 | 529.49 | 543.87 | **2,081.20** |
| 313 | 803.71 | 734.97 | 815.50 | 824.49 | **3,178.68** |
| 314 | 871.13 | 887.05 | 883.51 | 899.67 | **3,541.36** |
| 315 | 2,640.76 | 2,720.87 | 2,734.30 | 2,325.05 | **10,420.97** |
| 316 | 251.01 | 275.16 | 309.50 | 267.02 | **1,102.69** |
| 317 | 688.18 | 666.53 | 734.50 | 757.04 | **2,846.24** |
| 318 | 18.95 | 17.68 | 20.12 | 24.28 | **81.03** |
| 319 | 282.60 | 293.34 | 270.83 | 282.77 | **1,129.55** |
| 398 | 204.50 | 225.38 | 169.31 | 175.42 | **774.61** |
| **GA - GEORGIA - Total** | **20,797.85** | **21,786.05** | **21,985.93** | **22,354.48** | **86,924.31** |

**STATE: GU - GUAM**

| REGISTRANT ZIP CODE 3 | 1ST QUARTER | 2ND QUARTER | 3RD QUARTER | 4TH QUARTER | TOTAL GRAMS |
|---|---|---|---|---|---|
| 969 | 0.01 | 0 | 2.99 | 0.48 | **3.47** |
| **GU - GUAM - Total** | **0.01** | **0.00** | **2.99** | **0.48** | **3.47** |

**STATE: HI - HAWAII**

| REGISTRANT ZIP CODE 3 | 1ST QUARTER | 2ND QUARTER | 3RD QUARTER | 4TH QUARTER | TOTAL GRAMS |
|---|---|---|---|---|---|
| 967 | 1,869.21 | 1,828.27 | 2,020.42 | 2,044.18 | **7,762.09** |
| 968 | 477.97 | 478.47 | 499.80 | 458.88 | **1,915.11** |
| **HI - HAWAII - Total** | **2,347.18** | **2,306.74** | **2,520.22** | **2,503.06** | **9,677.20** |

**STATE: IA - IOWA**

| REGISTRANT ZIP CODE 3 | 1ST QUARTER | 2ND QUARTER | 3RD QUARTER | 4TH QUARTER | TOTAL GRAMS |
|---|---|---|---|---|---|
| 500 | 128.06 | 125.62 | 144.49 | 145.65 | **543.81** |
| 501 | 151.94 | 121.74 | 121.57 | 136.09 | **531.34** |
| 502 | 121.84 | 132.08 | 209.81 | 273.47 | **737.20** |
| 503 | 533.70 | 478.06 | 396.94 | 381.81 | **1,790.51** |
| 504 | 227.04 | 216.44 | 206.76 | 224.93 | **875.18** |
| 505 | 84.52 | 91.74 | 91.28 | 86.96 | **354.50** |
| 506 | 113.56 | 120.42 | 129.22 | 118.10 | **481.30** |
| 507 | 72.20 | 76.94 | 76.15 | 76.85 | **302.13** |
| 508 | 5.68 | 8.33 | 4.35 | 7.51 | **25.87** |
| 510 | 114.83 | 96.64 | 109.55 | 94.58 | **415.59** |
| 511 | 241.99 | 255.75 | 249.49 | 254.71 | **1,001.94** |
| 512 | 60.61 | 52.76 | 50.87 | 46.49 | **210.73** |
| 513 | 33.86 | 36.95 | 42.14 | 46.12 | **159.08** |
| 514 | 12.71 | 9.14 | 8.74 | 11.16 | **41.75** |
| 515 | 108.65 | 108.98 | 116.18 | 129.70 | **463.51** |
| 516 | 14.35 | 14.15 | 9.63 | 12.23 | **50.37** |
| 520 | 56.96 | 50.86 | 75.65 | 81.33 | **264.81** |
| 521 | 10.94 | 11.73 | 13.32 | 14.79 | **50.78** |
| 522 | 281.24 | 292.59 | 337.05 | 325.21 | **1,236.10** |
| 523 | 34.88 | 48.45 | 42.19 | 42.16 | **167.68** |
| 524 | 152.76 | 137.51 | 151.04 | 148.27 | **589.57** |
| 525 | 109.87 | 112.72 | 105.70 | 111.85 | **440.14** |
| 526 | 82.89 | 86.69 | 95.30 | 98.56 | **363.44** |
| 527 | 158.17 | 132.56 | 148.31 | 148.20 | **587.24** |
| 528 | 114.60 | 125.52 | 133.19 | 144.36 | **517.67** |
| **IA - IOWA - Total** | **3,027.86** | **2,944.40** | **3,068.91** | **3,161.09** | **12,202.26** |

**STATE: ID - IDAHO**

| REGISTRANT ZIP CODE 3 | 1ST QUARTER | 2ND QUARTER | 3RD QUARTER | 4TH QUARTER | TOTAL GRAMS |
|---|---|---|---|---|---|
| 832 | 637.30 | 633.42 | 642.37 | 657.38 | **2,570.48** |
| 833 | 429.03 | 504.11 | 500.01 | 480.96 | **1,914.12** |
| 834 | 262.50 | 302.96 | 339.62 | 351.05 | **1,256.13** |
| 835 | 558.56 | 602.67 | 610.74 | 619.84 | **2,391.80** |
| 836 | 872.45 | 952.36 | 988.99 | 988.49 | **3,802.29** |
| 837 | 848.41 | 883.76 | 927.27 | 944.76 | **3,604.21** |
| 838 | 1,858.48 | 1,972.24 | 1,916.06 | 1,997.34 | **7,744.12** |
| **ID - IDAHO - Total** | **5,466.73** | **5,851.54** | **5,925.07** | **6,039.82** | **23,283.15** |

**STATE: IL - ILLINOIS**

| REGISTRANT ZIP CODE 3 | 1ST QUARTER | 2ND QUARTER | 3RD QUARTER | 4TH QUARTER | TOTAL GRAMS |
|---|---|---|---|---|---|
| 600 | 3,014.08 | 2,946.37 | 3,127.63 | 3,237.93 | **12,326.01** |
| 601 | 1,507.99 | 1,567.98 | 1,575.19 | 1,739.80 | **6,390.96** |
| 602 | 36.40 | 39.98 | 46.04 | 42.54 | **164.96** |
| 603 | 349.92 | 264.45 | 137.80 | 168.43 | **920.60** |
| 604 | 2,481.22 | 2,602.84 | 2,691.51 | 2,703.65 | **10,479.22** |
| 605 | 1,190.81 | 1,085.85 | 1,183.11 | 1,173.39 | **4,633.15** |
| 606 | 3,141.01 | 3,085.87 | 3,021.39 | 3,070.77 | **12,319.05** |
| 607 | 122.08 | 157.50 | 142.95 | 145.33 | **567.86** |
| 608 | 99.59 | 117.30 | 99.13 | 104.66 | **420.68** |
| 609 | 280.94 | 324.73 | 334.18 | 366.86 | **1,306.71** |
| 610 | 300.37 | 332.40 | 337.67 | 385.81 | **1,356.25** |
| 611 | 826.18 | 849.17 | 867.09 | 938.37 | **3,480.81** |
| 612 | 214.96 | 237.94 | 248.85 | 262.60 | **964.35** |
| 613 | 406.61 | 389.86 | 423.41 | 449.56 | **1,669.46** |
| 614 | 139.25 | 151.82 | 172.98 | 189.99 | **654.03** |
| 615 | 325.00 | 336.00 | 340.64 | 362.55 | **1,364.19** |
| 616 | 602.72 | 600.35 | 672.33 | 647.43 | **2,522.83** |
| 617 | 265.41 | 298.60 | 337.75 | 355.57 | **1,257.33** |
| 618 | 621.71 | 644.11 | 685.68 | 736.17 | **2,687.66** |
| 619 | 101.79 | 132.26 | 147.89 | 150.52 | **532.46** |
| 620 | 634.18 | 678.98 | 743.29 | 816.54 | **2,872.98** |

| | | | | | |
|---|---|---|---|---|---|
| 622 | 428.06 | 434.70 | 431.71 | 441.70 | **1,736.16** |
| 623 | 349.32 | 364.85 | 362.85 | 341.94 | **1,418.96** |
| 624 | 214.87 | 246.79 | 287.19 | 309.64 | **1,058.50** |
| 625 | 398.61 | 396.34 | 430.35 | 416.74 | **1,642.05** |
| 626 | 368.54 | 363.04 | 340.11 | 381.21 | **1,452.89** |
| 627 | 543.16 | 500.36 | 499.00 | 485.31 | **2,027.84** |
| 628 | 550.43 | 607.67 | 607.54 | 660.73 | **2,426.38** |
| 629 | 1,866.93 | 1,918.52 | 1,992.62 | 2,047.42 | **7,825.50** |
| **IL - ILLINOIS - Total** | **21,382.14** | **21,676.64** | **22,287.89** | **23,133.16** | **88,479.83** |

**STATE: IN - INDIANA**

| REGISTRANT ZIP CODE 3 | 1ST QUARTER | 2ND QUARTER | 3RD QUARTER | 4TH QUARTER | TOTAL GRAMS |
|---|---|---|---|---|---|
| 460 | 3,032.90 | 3,193.45 | 3,391.41 | 3,494.76 | **13,112.51** |
| 461 | 3,506.16 | 4,508.38 | 4,654.91 | 4,803.78 | **17,473.23** |
| 462 | 4,634.26 | 4,574.26 | 4,653.50 | 4,753.02 | **18,615.04** |
| 463 | 2,788.60 | 2,826.27 | 2,836.40 | 2,982.77 | **11,434.05** |
| 464 | 350.70 | 313.30 | 361.33 | 389.31 | **1,414.63** |
| 465 | 1,116.19 | 1,207.03 | 1,240.75 | 1,282.21 | **4,846.18** |
| 466 | 396.19 | 400.37 | 384.46 | 438.55 | **1,619.57** |
| 467 | 415.10 | 555.73 | 573.46 | 599.39 | **2,143.69** |
| 468 | 943.87 | 956.21 | 984.53 | 962.66 | **3,847.27** |
| 469 | 1,834.99 | 1,872.53 | 1,999.02 | 2,078.85 | **7,785.39** |
| 470 | 905.97 | 956.47 | 1,072.84 | 1,082.63 | **4,017.91** |
| 471 | 3,038.63 | 3,145.98 | 3,234.54 | 3,229.15 | **12,648.30** |
| 472 | 1,927.63 | 2,035.60 | 2,219.49 | 2,292.09 | **8,474.82** |
| 473 | 3,169.47 | 3,301.79 | 3,299.87 | 3,538.40 | **13,309.52** |
| 474 | 2,315.85 | 2,257.29 | 2,268.44 | 2,254.07 | **9,095.64** |
| 475 | 777.75 | 877.48 | 960.61 | 1,040.33 | **3,656.17** |
| 476 | 237.01 | 264.22 | 293.98 | 327.94 | **1,123.16** |
| 477 | 1,306.84 | 1,126.43 | 1,304.57 | 1,381.21 | **5,119.05** |
| 478 | 636.74 | 621.64 | 674.48 | 730.17 | **2,663.03** |
| 479 | 933.55 | 951.89 | 990.13 | 971.56 | **3,847.12** |
| **IN - INDIANA - Total** | **34,268.41** | **35,946.33** | **37,398.72** | **38,632.83** | **146,246.28** |

**STATE: KS - KANSAS**

| REGISTRANT ZIP CODE 3 | 1ST QUARTER | 2ND QUARTER | 3RD QUARTER | 4TH QUARTER | TOTAL GRAMS |
|---|---|---|---|---|---|
| 660 | 693.89 | 665.72 | 687.42 | 705.64 | **2,752.66** |
| 661 | 212.10 | 160.40 | 158.78 | 186.99 | **718.28** |
| 662 | 869.38 | 905.31 | 1,007.73 | 1,058.96 | **3,841.38** |
| 664 | 29.88 | 33.80 | 43.44 | 50.60 | **157.71** |
| 665 | 56.83 | 56.58 | 63.31 | 65.58 | **242.30** |
| 666 | 143.96 | 165.95 | 194.29 | 180.26 | **684.45** |
| 667 | 204.79 | 189.40 | 195.04 | 208.50 | **797.73** |
| 668 | 13.38 | 16.39 | 18.81 | 21.52 | **70.10** |
| 669 | 9.09 | 9.37 | 9.55 | 6.95 | **34.95** |
| 670 | 89.66 | 103.95 | 96.11 | 102.99 | **392.71** |
| 671 | 48.06 | 55.96 | 57.68 | 61.29 | **222.99** |
| 672 | 545.59 | 595.59 | 621.03 | 635.53 | **2,397.74** |
| 673 | 92.31 | 76.54 | 91.86 | 93.65 | **354.36** |
| 674 | 189.25 | 187.17 | 210.39 | 223.72 | **810.53** |
| 675 | 62.51 | 75.75 | 79.44 | 85.79 | **303.49** |
| 676 | 84.03 | 86.66 | 94.24 | 89.50 | **354.44** |
| 677 | 16.82 | 21.32 | 20.85 | 31.89 | **90.88** |
| 678 | 67.84 | 77.90 | 75.26 | 75.44 | **296.44** |
| 679 | 10.36 | 18.96 | 23.01 | 22.40 | **74.74** |
| **KS - KANSAS - Total** | **3,439.72** | **3,502.72** | **3,748.25** | **3,907.19** | **14,597.88** |

**STATE: KY - KENTUCKY**

| REGISTRANT ZIP CODE 3 | 1ST QUARTER | 2ND QUARTER | 3RD QUARTER | 4TH QUARTER | TOTAL GRAMS |
|---|---|---|---|---|---|
| 400 | 910.61 | 1,015.79 | 1,113.40 | 1,025.45 | **4,065.25** |
| 401 | 423.65 | 471.04 | 488.94 | 486.75 | **1,870.38** |
| 402 | 4,535.36 | 5,522.37 | 6,036.96 | 6,418.09 | **22,512.77** |
| 403 | 5,163.55 | 5,304.01 | 5,301.34 | 5,382.68 | **21,151.58** |

| | | | | | |
|---|---|---|---|---|---|
| 404 | 2,470.16 | 2,594.87 | 2,637.93 | 2,634.23 | **10,337.18** |
| 405 | 3,227.39 | 3,174.19 | 3,467.48 | 3,150.93 | **13,019.98** |
| 406 | 675.14 | 692.37 | 699.84 | 674.82 | **2,742.17** |
| 407 | 2,882.61 | 2,663.21 | 2,970.07 | 3,182.90 | **11,698.80** |
| 408 | 604.74 | 664.40 | 612.76 | 597.90 | **2,479.80** |
| 409 | 2,189.12 | 2,361.88 | 2,421.84 | 2,396.80 | **9,369.66** |
| 410 | 2,978.28 | 3,055.58 | 3,192.86 | 3,125.33 | **12,352.05** |
| 411 | 1,991.81 | 2,044.70 | 2,259.67 | 2,291.70 | **8,587.88** |
| 412 | 1,587.66 | 1,583.41 | 1,660.14 | 1,396.84 | **6,228.05** |
| 413 | 1,092.07 | 1,074.67 | 1,143.20 | 1,073.63 | **4,383.57** |
| 414 | 743.78 | 504.62 | 725.87 | 780.01 | **2,754.27** |
| 415 | 3,112.07 | 2,967.47 | 3,068.73 | 3,133.48 | **12,281.76** |
| 416 | 1,759.27 | 1,871.54 | 2,435.74 | 2,924.93 | **8,991.48** |
| 417 | 1,496.03 | 1,386.25 | 1,393.13 | 1,423.78 | **5,699.18** |
| 418 | 736.17 | 619.52 | 730.20 | 795.87 | **2,881.76** |
| 420 | 1,869.91 | 2,057.03 | 2,157.25 | 2,157.21 | **8,241.40** |
| 421 | 1,748.98 | 1,715.41 | 1,923.65 | 2,061.99 | **7,450.04** |
| 422 | 522.14 | 579.06 | 583.78 | 617.94 | **2,302.92** |
| 423 | 1,675.28 | 1,303.92 | 1,357.03 | 1,537.59 | **5,873.82** |
| 424 | 688.19 | 766.52 | 782.52 | 774.66 | **3,011.90** |
| 425 | 1,208.37 | 1,139.19 | 1,383.04 | 1,444.24 | **5,174.84** |
| 426 | 586.76 | 697.00 | 762.06 | 822.96 | **2,868.78** |
| 427 | 1,648.64 | 1,873.11 | 2,060.95 | 2,041.07 | **7,623.77** |
| **KY - KENTUCKY - Total** | **48,527.73** | **49,703.12** | **53,370.40** | **54,353.78** | **205,955.04** |

**STATE: LA - LOUISIANA**

| REGISTRANT ZIP CODE 3 | 1ST QUARTER | 2ND QUARTER | 3RD QUARTER | 4TH QUARTER | TOTAL GRAMS |
|---|---|---|---|---|---|
| 700 | 4,300.35 | 4,281.65 | 4,499.14 | 4,430.50 | **17,511.64** |
| 701 | 1,791.55 | 1,724.13 | 1,937.95 | 1,952.44 | **7,406.08** |
| 703 | 1,334.01 | 1,389.89 | 1,419.67 | 1,420.28 | **5,563.85** |
| 704 | 4,439.26 | 4,817.11 | 5,000.36 | 4,849.40 | **19,106.13** |
| 705 | 3,052.77 | 3,370.00 | 3,580.10 | 3,655.22 | **13,658.10** |
| 706 | 1,530.34 | 1,636.38 | 1,488.58 | 1,434.76 | **6,090.06** |
| 707 | 2,031.79 | 2,223.88 | 2,216.45 | 2,197.79 | **8,669.91** |
| 708 | 1,259.98 | 1,253.03 | 1,299.68 | 1,280.50 | **5,093.20** |
| 710 | 356.80 | 497.06 | 481.43 | 531.98 | **1,867.27** |
| 711 | 2,150.20 | 2,440.69 | 2,319.39 | 2,495.71 | **9,405.99** |
| 712 | 1,690.72 | 1,850.85 | 2,020.35 | 2,091.38 | **7,653.31** |
| 713 | 1,285.99 | 1,475.63 | 1,652.36 | 1,748.16 | **6,162.14** |
| 714 | 658.50 | 737.33 | 782.29 | 819.39 | **2,997.52** |
| **LA - LOUISIANA - Total** | **25,882.27** | **27,697.63** | **28,697.76** | **28,907.54** | **111,185.20** |

**STATE: MA - MASSACHUSETTS**

| REGISTRANT ZIP CODE 3 | 1ST QUARTER | 2ND QUARTER | 3RD QUARTER | 4TH QUARTER | TOTAL GRAMS |
|---|---|---|---|---|---|
| 010 | 3,932.54 | 3,767.89 | 3,697.27 | 3,791.32 | **15,189.02** |
| 011 | 1,561.54 | 1,459.92 | 1,460.14 | 1,489.34 | **5,970.94** |
| 012 | 1,634.39 | 1,708.21 | 1,673.35 | 1,752.69 | **6,768.64** |
| 013 | 1,179.73 | 1,182.98 | 1,161.66 | 1,154.51 | **4,678.87** |
| 014 | 1,400.90 | 1,447.77 | 1,449.17 | 1,527.86 | **5,825.71** |
| 015 | 1,902.76 | 1,923.45 | 1,827.72 | 1,970.85 | **7,624.76** |
| 016 | 2,614.10 | 2,450.30 | 2,401.06 | 2,479.60 | **9,945.06** |
| 017 | 1,536.48 | 1,542.00 | 1,600.25 | 1,728.68 | **6,407.42** |
| 018 | 5,014.62 | 4,993.98 | 4,965.39 | 4,846.71 | **19,820.70** |
| 019 | 3,651.02 | 3,673.31 | 3,722.74 | 3,889.59 | **14,936.66** |
| 020 | 1,937.60 | 2,047.06 | 1,970.83 | 2,005.91 | **7,961.41** |
| 021 | 10,731.46 | 10,176.23 | 9,909.42 | 10,133.02 | **40,950.13** |
| 022 | 173.72 | 125.17 | 133.35 | 133.42 | **565.66** |
| 023 | 3,663.13 | 3,841.18 | 3,822.96 | 3,957.89 | **15,285.17** |
| 024 | 1,105.46 | 1,102.69 | 1,104.63 | 1,126.90 | **4,439.68** |
| 025 | 1,117.19 | 1,263.55 | 1,195.50 | 1,244.03 | **4,820.26** |
| 026 | 1,567.41 | 1,680.49 | 1,668.75 | 1,651.46 | **6,568.12** |
| 027 | 6,399.70 | 6,557.32 | 6,567.23 | 6,715.08 | **26,239.33** |

| | | | | | |
|---|---|---|---|---|---|
| **MA - MASSACHUSETTS - Total** | **51,123.74** | **50,943.53** | **50,331.42** | **51,598.85** | **203,997.54** |
| **STATE: MD - MARYLAND** | | | | | |
| REGISTRANT ZIP CODE 3 | 1ST QUARTER | 2ND QUARTER | 3RD QUARTER | 4TH QUARTER | **TOTAL GRAMS** |
| 206 | 2,738.54 | 2,636.95 | 2,873.49 | 3,014.22 | **11,263.21** |
| 207 | 1,512.72 | 1,548.31 | 1,639.16 | 1,741.04 | **6,441.23** |
| 208 | 1,234.64 | 1,141.57 | 1,143.11 | 1,179.54 | **4,698.86** |
| 209 | 413.06 | 374.16 | 369.33 | 366.84 | **1,523.39** |
| 210 | 5,091.85 | 4,788.88 | 4,919.75 | 5,297.18 | **20,097.66** |
| 211 | 1,817.20 | 1,836.44 | 1,942.88 | 1,972.37 | **7,568.90** |
| 212 | 13,690.90 | 13,498.58 | 14,016.97 | 14,544.94 | **55,751.39** |
| 214 | 1,102.41 | 1,001.69 | 1,102.79 | 1,123.71 | **4,330.59** |
| 215 | 1,935.23 | 1,893.69 | 1,973.66 | 1,999.70 | **7,802.28** |
| 216 | 1,057.50 | 1,098.86 | 1,152.74 | 1,265.63 | **4,574.73** |
| 217 | 3,095.16 | 2,927.13 | 2,941.76 | 3,035.24 | **11,999.28** |
| 218 | 994.17 | 1,066.69 | 1,127.92 | 1,126.08 | **4,314.87** |
| 219 | 1,457.69 | 1,477.35 | 1,447.20 | 1,471.37 | **5,853.61** |
| **MD - MARYLAND - Total** | **36,141.08** | **35,290.28** | **36,650.75** | **38,137.87** | **146,219.99** |
| **STATE: ME - MAINE** | | | | | |
| REGISTRANT ZIP CODE 3 | 1ST QUARTER | 2ND QUARTER | 3RD QUARTER | 4TH QUARTER | **TOTAL GRAMS** |
| 039 | 131.89 | 120.66 | 117.40 | 105.82 | **475.77** |
| 040 | 3,398.30 | 3,478.03 | 3,535.45 | 3,627.30 | **14,039.08** |
| 041 | 1,280.44 | 1,109.49 | 1,134.42 | 1,184.03 | **4,708.37** |
| 042 | 2,018.96 | 2,045.10 | 2,113.35 | 2,210.65 | **8,388.06** |
| 043 | 1,334.95 | 1,333.69 | 1,378.65 | 1,441.20 | **5,488.50** |
| 044 | 1,745.86 | 1,640.70 | 1,727.75 | 1,683.19 | **6,797.50** |
| 045 | 575.27 | 598.06 | 639.32 | 658.77 | **2,471.42** |
| 046 | 1,081.36 | 1,063.68 | 1,147.15 | 1,118.08 | **4,410.26** |
| 047 | 531.80 | 586.85 | 618.53 | 669.31 | **2,406.49** |
| 048 | 1,127.16 | 1,183.37 | 1,216.64 | 1,143.91 | **4,671.08** |
| 049 | 1,981.54 | 2,041.64 | 2,114.03 | 2,227.53 | **8,364.75** |
| **ME - MAINE - Total** | **15,207.53** | **15,201.26** | **15,742.69** | **16,069.80** | **62,221.29** |
| **STATE: MI - MICHIGAN** | | | | | |
| REGISTRANT ZIP CODE 3 | 1ST QUARTER | 2ND QUARTER | 3RD QUARTER | 4TH QUARTER | **TOTAL GRAMS** |
| 480 | 4,971.19 | 5,104.29 | 5,312.37 | 5,468.65 | **20,856.49** |
| 481 | 9,556.92 | 9,432.05 | 9,849.56 | 10,167.30 | **39,005.83** |
| 482 | 1,082.61 | 1,040.51 | 1,079.83 | 1,117.78 | **4,320.73** |
| 483 | 3,546.67 | 3,483.71 | 3,503.68 | 3,686.04 | **14,220.10** |
| 484 | 1,293.37 | 1,373.05 | 1,396.79 | 1,410.70 | **5,473.91** |
| 485 | 1,045.59 | 1,062.65 | 1,033.04 | 1,074.86 | **4,216.15** |
| 486 | 1,416.26 | 1,507.67 | 1,595.78 | 1,632.20 | **6,151.92** |
| 487 | 843.20 | 902.17 | 946.22 | 1,005.16 | **3,696.75** |
| 488 | 1,710.35 | 1,825.56 | 1,845.63 | 1,950.77 | **7,332.31** |
| 489 | 310.17 | 347.43 | 343.60 | 361.12 | **1,362.32** |
| 490 | 1,805.57 | 1,838.25 | 1,884.11 | 1,906.25 | **7,434.19** |
| 491 | 124.53 | 145.82 | 167.91 | 186.97 | **625.23** |
| 492 | 1,030.68 | 1,159.07 | 1,154.89 | 1,228.23 | **4,572.87** |
| 493 | 568.39 | 601.15 | 663.56 | 657.58 | **2,490.67** |
| 494 | 1,227.57 | 1,318.41 | 1,378.14 | 1,483.58 | **5,407.70** |
| 495 | 1,340.58 | 1,315.30 | 1,362.31 | 1,412.80 | **5,431.00** |
| 496 | 1,689.17 | 1,788.67 | 1,880.27 | 1,922.33 | **7,280.44** |
| 497 | 1,772.03 | 1,920.27 | 2,007.63 | 2,073.61 | **7,773.55** |
| 498 | 1,089.20 | 1,136.48 | 1,241.41 | 1,291.88 | **4,758.97** |
| 499 | 297.62 | 294.41 | 326.25 | 361.37 | **1,279.64** |
| **MI - MICHIGAN - Total** | **36,721.68** | **37,596.92** | **38,972.98** | **40,399.20** | **153,690.78** |
| **STATE: MN - MINNESOTA** | | | | | |
| REGISTRANT ZIP CODE 3 | 1ST QUARTER | 2ND QUARTER | 3RD QUARTER | 4TH QUARTER | **TOTAL GRAMS** |
| 550 | 1,291.95 | 1,321.34 | 1,347.43 | 1,355.40 | **5,316.12** |
| 551 | 1,971.47 | 1,974.48 | 2,061.38 | 2,131.77 | **8,139.10** |
| 553 | 1,544.20 | 1,505.24 | 1,563.98 | 1,668.64 | **6,282.06** |
| 554 | 3,192.51 | 3,000.93 | 3,034.16 | 3,225.32 | **12,452.92** |

| | | | | | |
|---|---|---|---|---|---|
| 556 | 34.86 | 27.06 | 23.00 | 23.17 | **108.09** |
| 557 | 488.83 | 529.15 | 551.33 | 563.13 | **2,132.44** |
| 558 | 651.19 | 641.95 | 677.76 | 702.14 | **2,673.03** |
| 559 | 304.24 | 315.25 | 374.54 | 398.95 | **1,392.99** |
| 560 | 344.65 | 371.03 | 352.66 | 362.88 | **1,431.21** |
| 561 | 21.21 | 22.34 | 26.28 | 27.43 | **97.27** |
| 562 | 167.80 | 163.34 | 184.78 | 191.38 | **707.30** |
| 563 | 601.08 | 588.59 | 600.03 | 613.53 | **2,403.23** |
| 564 | 298.81 | 317.48 | 334.96 | 384.82 | **1,336.07** |
| 565 | 582.65 | 571.61 | 578.26 | 635.98 | **2,368.50** |
| 566 | 769.59 | 822.96 | 883.05 | 858.81 | **3,334.42** |
| 567 | 95.72 | 100.60 | 112.67 | 134.66 | **443.66** |
| **MN - MINNESOTA - Total** | **12,360.78** | **12,273.37** | **12,706.26** | **13,278.01** | **50,618.41** |

**STATE: MO - MISSOURI**

| REGISTRANT ZIP CODE 3 | 1ST QUARTER | 2ND QUARTER | 3RD QUARTER | 4TH QUARTER | TOTAL GRAMS |
|---|---|---|---|---|---|
| 630 | 1,177.40 | 1,179.18 | 1,320.21 | 1,361.28 | **5,038.07** |
| 631 | 3,364.37 | 3,529.36 | 3,274.46 | 3,313.40 | **13,481.59** |
| 633 | 910.23 | 905.45 | 876.80 | 938.45 | **3,630.93** |
| 634 | 216.64 | 216.24 | 227.61 | 248.77 | **909.26** |
| 635 | 111.87 | 112.68 | 129.63 | 150.00 | **504.19** |
| 636 | 892.00 | 1,012.90 | 1,089.59 | 1,090.59 | **4,085.08** |
| 637 | 417.06 | 442.52 | 427.46 | 457.91 | **1,744.95** |
| 638 | 341.15 | 367.22 | 403.96 | 403.90 | **1,516.23** |
| 639 | 462.80 | 469.96 | 466.36 | 511.92 | **1,911.04** |
| 640 | 1,038.16 | 1,117.63 | 1,097.87 | 1,184.80 | **4,438.46** |
| 641 | 1,048.34 | 1,133.56 | 1,091.26 | 1,200.52 | **4,473.68** |
| 644 | 52.99 | 60.11 | 67.67 | 75.73 | **256.50** |
| 645 | 680.39 | 696.33 | 800.76 | 764.55 | **2,942.03** |
| 646 | 64.27 | 86.31 | 88.44 | 102.24 | **341.26** |
| 647 | 218.17 | 250.53 | 266.68 | 283.61 | **1,019.00** |
| 648 | 731.70 | 676.83 | 708.44 | 723.38 | **2,840.34** |
| 650 | 271.72 | 279.00 | 289.04 | 306.78 | **1,146.54** |
| 651 | 216.55 | 231.13 | 243.04 | 285.48 | **976.20** |
| 652 | 1,054.23 | 997.43 | 1,049.72 | 1,081.18 | **4,182.56** |
| 653 | 360.79 | 385.05 | 415.44 | 404.11 | **1,565.39** |
| 654 | 506.02 | 524.69 | 579.35 | 539.50 | **2,149.56** |
| 655 | 470.15 | 521.32 | 586.92 | 613.22 | **2,191.61** |
| 656 | 472.64 | 596.94 | 612.55 | 686.35 | **2,368.48** |
| 657 | 1,016.64 | 1,005.60 | 1,099.16 | 1,219.18 | **4,340.57** |
| 658 | 3,244.57 | 3,004.41 | 3,157.19 | 3,388.24 | **12,794.42** |
| **MO - MISSOURI - Total** | **19,340.84** | **19,802.37** | **20,369.60** | **21,335.10** | **80,847.92** |

**STATE: MS - MISSISSIPPI**

| REGISTRANT ZIP CODE 3 | 1ST QUARTER | 2ND QUARTER | 3RD QUARTER | 4TH QUARTER | TOTAL GRAMS |
|---|---|---|---|---|---|
| 386 | 1,532.36 | 1,617.12 | 1,635.67 | 1,694.35 | **6,479.50** |
| 387 | 136.89 | 180.25 | 164.74 | 170.50 | **652.39** |
| 388 | 1,965.46 | 2,134.82 | 2,057.03 | 2,183.94 | **8,341.25** |
| 389 | 196.53 | 219.09 | 222.61 | 241.67 | **879.89** |
| 390 | 708.77 | 707.00 | 742.62 | 742.67 | **2,901.05** |
| 391 | 747.15 | 717.33 | 750.50 | 752.01 | **2,966.98** |
| 392 | 716.15 | 658.42 | 700.16 | 687.20 | **2,761.94** |
| 393 | 629.39 | 675.99 | 663.28 | 658.18 | **2,626.83** |
| 394 | 2,117.34 | 2,258.29 | 2,308.02 | 2,328.69 | **9,012.35** |
| 395 | 2,806.60 | 2,840.42 | 2,935.48 | 2,969.17 | **11,551.68** |
| 396 | 394.50 | 398.86 | 431.28 | 421.50 | **1,646.14** |
| 397 | 376.79 | 405.55 | 420.63 | 438.07 | **1,641.04** |
| **MS - MISSISSIPPI - Total** | **12,327.92** | **12,813.16** | **13,032.02** | **13,287.94** | **51,461.04** |

**STATE: MT - MONTANA**

| REGISTRANT ZIP CODE 3 | 1ST QUARTER | 2ND QUARTER | 3RD QUARTER | 4TH QUARTER | TOTAL GRAMS |
|---|---|---|---|---|---|
| 590 | 131.08 | 141.74 | 160.75 | 151.99 | **585.57** |
| 591 | 560.25 | 556.83 | 588.25 | 625.30 | **2,330.63** |

| | | | | | |
|---|---|---|---|---|---|
| 592 | 172.74 | 195.12 | 194.53 | 196.65 | **759.05** |
| 593 | 86.89 | 91.68 | 98.19 | 105.39 | **382.16** |
| 594 | 976.95 | 1,088.41 | 1,135.05 | 1,202.19 | **4,402.60** |
| 595 | 137.23 | 146.51 | 173.79 | 163.35 | **620.86** |
| 596 | 426.90 | 443.98 | 383.44 | 447.86 | **1,702.18** |
| 597 | 381.85 | 393.64 | 436.38 | 425.49 | **1,637.37** |
| 598 | 1,017.61 | 991.49 | 1,009.90 | 1,066.79 | **4,085.80** |
| 599 | 451.27 | 435.35 | 469.75 | 473.79 | **1,830.15** |
| **MT - MONTANA - Total** | **4,342.76** | **4,484.77** | **4,650.04** | **4,858.79** | **18,336.37** |

**STATE: NC - NORTH CAROLINA**

| REGISTRANT ZIP CODE 3 | 1ST QUARTER | 2ND QUARTER | 3RD QUARTER | 4TH QUARTER | TOTAL GRAMS |
|---|---|---|---|---|---|
| 270 | 1,321.01 | 1,485.62 | 1,514.00 | 1,520.91 | **5,841.54** |
| 271 | 1,060.67 | 1,127.74 | 1,241.79 | 1,273.09 | **4,703.29** |
| 272 | 2,893.97 | 3,130.25 | 3,326.78 | 3,651.98 | **13,002.98** |
| 273 | 1,946.51 | 2,031.11 | 2,103.71 | 2,130.64 | **8,211.98** |
| 274 | 943.04 | 895.05 | 908.85 | 966.91 | **3,713.85** |
| 275 | 3,596.86 | 3,651.59 | 3,789.58 | 3,928.43 | **14,966.47** |
| 276 | 1,642.32 | 1,419.83 | 1,508.22 | 1,509.49 | **6,079.87** |
| 277 | 2,434.11 | 2,408.51 | 2,427.00 | 2,453.72 | **9,723.35** |
| 278 | 2,257.47 | 2,223.22 | 2,205.63 | 2,316.32 | **9,002.64** |
| 279 | 711.93 | 728.11 | 771.99 | 748.76 | **2,960.80** |
| 280 | 4,728.74 | 4,828.94 | 5,004.14 | 5,059.13 | **19,620.95** |
| 281 | 2,931.73 | 2,971.31 | 2,942.99 | 3,002.20 | **11,848.23** |
| 282 | 2,434.49 | 2,475.39 | 2,505.70 | 2,574.86 | **9,990.44** |
| 283 | 5,203.09 | 5,214.28 | 5,609.86 | 5,875.94 | **21,903.17** |
| 284 | 4,798.74 | 5,162.51 | 5,126.45 | 5,148.40 | **20,236.09** |
| 285 | 2,249.43 | 2,349.75 | 2,501.75 | 2,528.11 | **9,629.04** |
| 286 | 7,585.28 | 7,427.48 | 8,204.71 | 7,927.34 | **31,144.81** |
| 287 | 3,989.69 | 4,202.81 | 4,219.27 | 4,396.05 | **16,807.82** |
| 288 | 2,364.33 | 2,177.37 | 2,291.83 | 2,432.57 | **9,266.10** |
| 289 | 263.76 | 299.51 | 306.00 | 340.77 | **1,210.04** |
| **NC - NORTH CAROLINA - Total** | **55,357.18** | **56,210.40** | **58,510.26** | **59,785.61** | **229,863.45** |

**STATE: ND - NORTH DAKOTA**

| REGISTRANT ZIP CODE 3 | 1ST QUARTER | 2ND QUARTER | 3RD QUARTER | 4TH QUARTER | TOTAL GRAMS |
|---|---|---|---|---|---|
| 580 | 36.74 | 38.74 | 34.35 | 35.20 | **145.03** |
| 581 | 371.88 | 277.87 | 349.14 | 343.54 | **1,342.43** |
| 582 | 244.04 | 267.30 | 249.66 | 280.21 | **1,041.20** |
| 583 | 237.66 | 221.68 | 232.86 | 201.61 | **893.81** |
| 584 | 36.70 | 46.06 | 58.60 | 51.16 | **192.52** |
| 585 | 451.83 | 446.52 | 467.19 | 502.49 | **1,868.02** |
| 586 | 107.91 | 111.20 | 114.79 | 100.18 | **434.08** |
| 587 | 151.02 | 141.88 | 135.41 | 148.18 | **576.49** |
| 588 | 188.90 | 174.24 | 173.68 | 158.73 | **695.56** |
| **ND - NORTH DAKOTA - Total** | **1,826.67** | **1,725.49** | **1,815.68** | **1,821.29** | **7,189.14** |

**STATE: NE - NEBRASKA**

| REGISTRANT ZIP CODE 3 | 1ST QUARTER | 2ND QUARTER | 3RD QUARTER | 4TH QUARTER | TOTAL GRAMS |
|---|---|---|---|---|---|
| 680 | 103.61 | 98.78 | 119.80 | 124.19 | **446.39** |
| 681 | 753.43 | 766.95 | 759.01 | 830.32 | **3,109.72** |
| 683 | 55.12 | 62.17 | 60.66 | 79.78 | **257.73** |
| 684 | 42.42 | 39.42 | 46.64 | 53.00 | **181.48** |
| 685 | 428.37 | 426.62 | 466.87 | 484.96 | **1,806.82** |
| 686 | 52.30 | 61.05 | 54.08 | 52.33 | **219.75** |
| 687 | 84.53 | 97.49 | 97.72 | 117.77 | **397.51** |
| 688 | 145.28 | 148.91 | 147.25 | 152.73 | **594.17** |
| 689 | 33.07 | 28.03 | 31.03 | 33.61 | **125.74** |
| 690 | 1.36 | 2.41 | 3.71 | 3.00 | **10.49** |
| 691 | 356.81 | 336.63 | 348.77 | 351.74 | **1,393.95** |
| 692 | 3.58 | 4.26 | 5.12 | 8.76 | **21.72** |
| 693 | 61.76 | 76.49 | 93.41 | 105.23 | **336.88** |
| **NE - NEBRASKA - Total** | **2,121.64** | **2,149.20** | **2,234.06** | **2,397.42** | **8,902.33** |

**STATE: NH - NEW HAMPSHIRE**

| REGISTRANT ZIP CODE 3 | 1ST QUARTER | 2ND QUARTER | 3RD QUARTER | 4TH QUARTER | TOTAL GRAMS |
|---|---|---|---|---|---|
| 030 | 1,900.02 | 1,957.46 | 1,983.37 | 2,117.71 | **7,958.56** |
| 031 | 1,706.49 | 1,645.43 | 1,721.60 | 1,751.01 | **6,824.53** |
| 032 | 1,589.89 | 1,650.25 | 1,699.13 | 1,841.06 | **6,780.32** |
| 033 | 1,127.76 | 1,054.31 | 1,112.95 | 857.14 | **4,152.15** |
| 034 | 576.93 | 532.45 | 520.79 | 582.51 | **2,212.68** |
| 035 | 576.30 | 545.53 | 581.62 | 638.74 | **2,342.18** |
| 036 | 13.81 | 17.09 | 14.58 | 18.15 | **63.62** |
| 037 | 1,299.64 | 1,247.34 | 1,271.19 | 1,422.36 | **5,240.53** |
| 038 | 2,340.39 | 2,331.68 | 2,375.57 | 2,483.16 | **9,530.80** |
| **NH - NEW HAMPSHIRE - Total** | **11,131.21** | **10,981.54** | **11,280.80** | **11,711.83** | **45,105.37** |

**STATE: NJ - NEW JERSEY**

| REGISTRANT ZIP CODE 3 | 1ST QUARTER | 2ND QUARTER | 3RD QUARTER | 4TH QUARTER | TOTAL GRAMS |
|---|---|---|---|---|---|
| 070 | 3,117.79 | 3,120.41 | 3,148.61 | 3,225.59 | **12,612.40** |
| 071 | 1,442.64 | 1,461.80 | 1,466.25 | 1,391.08 | **5,761.77** |
| 072 | 280.90 | 266.62 | 266.53 | 266.19 | **1,080.24** |
| 073 | 235.65 | 245.12 | 251.49 | 289.95 | **1,022.21** |
| 074 | 1,295.51 | 1,411.57 | 1,381.38 | 1,477.90 | **5,566.37** |
| 075 | 476.33 | 408.66 | 518.73 | 498.82 | **1,902.54** |
| 076 | 1,077.11 | 1,007.65 | 1,093.83 | 1,182.23 | **4,360.83** |
| 077 | 2,604.51 | 2,474.60 | 2,476.15 | 2,652.58 | **10,207.85** |
| 078 | 1,145.81 | 1,207.41 | 1,246.80 | 1,332.84 | **4,932.86** |
| 079 | 464.03 | 382.22 | 453.65 | 395.47 | **1,695.37** |
| 080 | 8,795.17 | 8,527.79 | 8,696.39 | 9,112.85 | **35,132.19** |
| 081 | 2,551.37 | 2,419.89 | 1,597.83 | 1,748.78 | **8,317.87** |
| 082 | 2,355.07 | 2,372.37 | 2,429.26 | 2,590.01 | **9,746.71** |
| 083 | 1,604.32 | 1,731.86 | 1,670.69 | 1,683.78 | **6,690.66** |
| 084 | 407.82 | 463.85 | 418.91 | 460.11 | **1,750.68** |
| 085 | 830.88 | 755.09 | 761.48 | 799.31 | **3,146.75** |
| 086 | 1,153.68 | 1,176.92 | 1,160.35 | 1,196.87 | **4,687.81** |
| 087 | 3,161.54 | 2,956.44 | 3,043.80 | 3,173.53 | **12,335.30** |
| 088 | 2,384.66 | 2,273.39 | 2,390.17 | 2,433.91 | **9,482.13** |
| 089 | 257.90 | 242.46 | 255.57 | 219.44 | **975.38** |
| **NJ - NEW JERSEY - Total** | **35,642.68** | **34,906.13** | **34,727.87** | **36,131.25** | **141,407.92** |

**STATE: NM - NEW MEXICO**

| REGISTRANT ZIP CODE 3 | 1ST QUARTER | 2ND QUARTER | 3RD QUARTER | 4TH QUARTER | TOTAL GRAMS |
|---|---|---|---|---|---|
| 870 | 579.00 | 627.32 | 621.85 | 614.94 | **2,443.10** |
| 871 | 4,832.07 | 4,795.81 | 4,848.32 | 4,653.48 | **19,129.68** |
| 873 | 138.50 | 156.15 | 145.94 | 174.11 | **614.70** |
| 874 | 343.69 | 342.46 | 341.37 | 370.28 | **1,397.80** |
| 875 | 2,302.88 | 2,398.60 | 2,317.74 | 2,344.34 | **9,363.57** |
| 877 | 250.76 | 289.37 | 280.77 | 259.01 | **1,079.92** |
| 878 | 238.45 | 222.43 | 219.67 | 233.06 | **913.62** |
| 879 | 27.29 | 38.21 | 42.78 | 37.33 | **145.61** |
| 880 | 561.87 | 553.08 | 573.00 | 569.89 | **2,257.84** |
| 881 | 51.13 | 58.53 | 55.02 | 60.10 | **224.78** |
| 882 | 557.16 | 565.90 | 599.86 | 605.43 | **2,328.35** |
| 883 | 199.06 | 187.02 | 198.65 | 221.61 | **806.34** |
| 884 | 26.93 | 21.85 | 28.15 | 23.29 | **100.23** |
| **NM - NEW MEXICO - Total** | **10,108.79** | **10,256.74** | **10,273.13** | **10,166.87** | **40,805.54** |

**STATE: NV - NEVADA**

| REGISTRANT ZIP CODE 3 | 1ST QUARTER | 2ND QUARTER | 3RD QUARTER | 4TH QUARTER | TOTAL GRAMS |
|---|---|---|---|---|---|
| 890 | 1,000.90 | 913.87 | 1,024.96 | 1,091.81 | **4,031.55** |
| 891 | 2,329.44 | 2,230.84 | 2,365.11 | 2,465.09 | **9,390.49** |
| 893 | 17.44 | 23.50 | 25.60 | 32.96 | **99.51** |
| 894 | 518.51 | 533.40 | 550.20 | 554.33 | **2,156.44** |
| 895 | 715.09 | 679.10 | 748.85 | 798.77 | **2,941.81** |
| 897 | 231.15 | 236.00 | 259.74 | 256.21 | **983.10** |
| 898 | 92.91 | 96.65 | 88.74 | 93.92 | **372.22** |

| NV - NEVADA - Total | 4,905.44 | 4,713.37 | 5,063.22 | 5,293.08 | 19,975.11 |
|---|---|---|---|---|---|

STATE: NY - NEW YORK

| REGISTRANT ZIP CODE 3 | 1ST QUARTER | 2ND QUARTER | 3RD QUARTER | 4TH QUARTER | TOTAL GRAMS |
|---|---|---|---|---|---|
| 100 | 3,611.18 | 3,193.49 | 3,047.16 | 2,979.63 | **12,831.45** |
| 101 | 84.70 | 53.08 | 70.09 | 66.52 | **274.39** |
| 102 | 2.27 | 1.14 | 3.70 | 2.17 | **9.27** |
| 103 | 2,136.87 | 2,157.38 | 2,097.68 | 2,139.36 | **8,531.30** |
| 104 | 2,729.93 | 2,643.15 | 2,639.93 | 2,583.39 | **10,596.41** |
| 105 | 1,466.22 | 1,471.63 | 1,445.56 | 1,524.98 | **5,908.39** |
| 106 | 243.68 | 227.58 | 215.03 | 222.47 | **908.76** |
| 107 | 502.69 | 469.43 | 468.45 | 453.02 | **1,893.59** |
| 108 | 113.09 | 103.59 | 101.40 | 104.35 | **422.43** |
| 109 | 1,812.45 | 1,793.73 | 1,743.20 | 1,786.20 | **7,135.58** |
| 110 | 624.17 | 562.75 | 540.75 | 568.06 | **2,295.73** |
| 111 | 335.81 | 320.01 | 322.66 | 318.98 | **1,297.47** |
| 112 | 4,183.63 | 3,997.95 | 4,176.64 | 4,249.55 | **16,607.77** |
| 113 | 1,161.23 | 1,161.87 | 1,138.29 | 1,190.37 | **4,651.76** |
| 114 | 574.70 | 544.71 | 574.03 | 575.74 | **2,269.18** |
| 115 | 2,039.67 | 1,813.79 | 1,967.48 | 2,026.14 | **7,847.08** |
| 116 | 120.03 | 139.62 | 142.71 | 154.28 | **556.64** |
| 117 | 7,048.43 | 7,089.14 | 7,097.95 | 7,327.44 | **28,562.96** |
| 118 | 473.89 | 385.73 | 432.77 | 441.48 | **1,733.87** |
| 119 | 1,471.46 | 1,579.17 | 1,530.11 | 1,556.06 | **6,136.80** |
| 120 | 2,370.55 | 2,520.45 | 2,513.06 | 2,609.34 | **10,013.41** |
| 121 | 1,312.52 | 1,426.05 | 1,404.56 | 1,413.53 | **5,556.66** |
| 122 | 1,279.81 | 1,126.78 | 1,211.24 | 1,235.10 | **4,852.92** |
| 123 | 1,009.93 | 1,021.74 | 1,029.37 | 1,017.51 | **4,078.54** |
| 124 | 1,311.49 | 1,368.71 | 1,420.77 | 1,412.76 | **5,513.73** |
| 125 | 2,231.52 | 2,264.86 | 2,369.04 | 2,344.61 | **9,210.03** |
| 126 | 612.49 | 670.87 | 680.75 | 678.56 | **2,642.67** |
| 127 | 1,215.11 | 1,254.65 | 1,224.04 | 1,205.88 | **4,899.68** |
| 128 | 1,866.19 | 2,075.00 | 2,039.86 | 2,026.03 | **8,007.07** |
| 129 | 1,906.95 | 2,028.10 | 2,223.72 | 2,203.22 | **8,361.99** |
| 130 | 1,836.37 | 2,010.74 | 1,997.28 | 2,096.13 | **7,940.53** |
| 131 | 604.50 | 640.63 | 664.90 | 665.64 | **2,575.66** |
| 132 | 1,677.13 | 1,653.06 | 1,603.74 | 1,635.45 | **6,569.38** |
| 133 | 605.54 | 625.21 | 687.81 | 676.71 | **2,595.27** |
| 134 | 1,208.19 | 1,204.18 | 1,287.54 | 1,289.50 | **4,989.41** |
| 135 | 505.60 | 463.12 | 473.69 | 489.92 | **1,932.34** |
| 136 | 1,996.47 | 2,218.44 | 2,358.81 | 2,428.85 | **9,002.57** |
| 137 | 655.25 | 700.31 | 701.29 | 720.41 | **2,777.26** |
| 138 | 1,132.02 | 1,138.51 | 1,160.85 | 1,133.98 | **4,565.38** |
| 139 | 1,088.86 | 1,025.64 | 1,041.10 | 1,002.30 | **4,157.90** |
| 140 | 3,247.12 | 3,441.25 | 3,445.61 | 3,524.03 | **13,658.01** |
| 141 | 1,408.57 | 1,496.31 | 1,504.42 | 1,541.53 | **5,950.83** |
| 142 | 5,090.58 | 5,129.68 | 5,358.64 | 5,366.84 | **20,945.74** |
| 143 | 1,266.74 | 1,196.97 | 1,232.54 | 1,349.65 | **5,045.90** |
| 144 | 1,445.53 | 1,503.92 | 1,560.02 | 1,670.56 | **6,180.03** |
| 145 | 1,132.34 | 1,280.11 | 1,304.41 | 1,332.07 | **5,048.93** |
| 146 | 3,514.82 | 3,472.56 | 3,452.82 | 3,634.34 | **14,074.54** |
| 147 | 1,087.33 | 1,184.22 | 1,143.80 | 1,176.18 | **4,591.52** |
| 148 | 1,733.10 | 1,813.99 | 1,821.53 | 1,920.40 | **7,289.03** |
| 149 | 509.50 | 494.58 | 488.25 | 495.07 | **1,987.40** |
| **NY - NEW YORK - Total** | **77,598.23** | **78,159.57** | **79,161.05** | **80,566.31** | **315,485.16** |

STATE: OH - OHIO

| REGISTRANT ZIP CODE 3 | 1ST QUARTER | 2ND QUARTER | 3RD QUARTER | 4TH QUARTER | TOTAL GRAMS |
|---|---|---|---|---|---|
| 430 | 3,163.79 | 3,148.24 | 3,279.37 | 3,195.76 | **12,787.16** |
| 431 | 2,899.95 | 2,954.83 | 3,125.58 | 2,972.19 | **11,952.55** |
| 432 | 6,387.34 | 6,303.31 | 6,540.96 | 6,584.08 | **25,815.70** |
| 433 | 1,526.17 | 1,646.43 | 1,729.28 | 1,708.54 | **6,610.42** |

| | | | | | |
|---|---|---|---|---|---|
| 434 | 383.45 | 423.50 | 422.61 | 452.61 | **1,682.17** |
| 435 | 673.62 | 694.11 | 741.09 | 844.18 | **2,953.00** |
| 436 | 2,419.24 | 2,353.20 | 2,371.78 | 2,416.65 | **9,560.87** |
| 437 | 1,777.65 | 1,896.77 | 1,948.12 | 2,015.10 | **7,637.63** |
| 438 | 122.41 | 132.72 | 137.27 | 154.93 | **547.32** |
| 439 | 1,952.73 | 2,038.44 | 2,059.54 | 1,987.18 | **8,037.89** |
| 440 | 2,696.72 | 2,848.71 | 2,927.10 | 2,943.46 | **11,416.00** |
| 441 | 4,007.24 | 4,294.59 | 4,256.72 | 4,538.68 | **17,097.23** |
| 442 | 2,111.44 | 2,062.61 | 2,131.30 | 2,216.32 | **8,521.66** |
| 443 | 1,409.13 | 1,401.06 | 1,396.10 | 1,380.98 | **5,587.27** |
| 444 | 1,780.32 | 1,755.29 | 1,865.52 | 1,974.09 | **7,375.22** |
| 445 | 2,022.59 | 1,944.62 | 1,953.30 | 2,049.14 | **7,969.65** |
| 446 | 1,109.99 | 1,259.79 | 1,306.85 | 1,217.64 | **4,894.27** |
| 447 | 1,048.62 | 875.02 | 935.25 | 937.60 | **3,796.48** |
| 448 | 1,427.63 | 1,497.64 | 1,468.80 | 1,567.69 | **5,961.76** |
| 449 | 992.77 | 1,057.98 | 976.31 | 1,080.50 | **4,107.56** |
| 450 | 3,776.37 | 3,757.87 | 3,886.55 | 4,007.06 | **15,427.84** |
| 451 | 3,519.88 | 3,413.84 | 3,570.09 | 3,629.98 | **14,133.79** |
| 452 | 4,752.39 | 4,153.20 | 4,470.73 | 4,284.38 | **17,660.69** |
| 453 | 1,513.73 | 1,578.00 | 1,633.26 | 1,732.52 | **6,457.51** |
| 454 | 2,584.58 | 2,454.08 | 2,586.81 | 2,751.76 | **10,377.23** |
| 455 | 918.19 | 1,023.76 | 1,003.22 | 968.00 | **3,913.18** |
| 456 | 5,658.71 | 5,993.37 | 5,898.10 | 5,989.54 | **23,539.72** |
| 457 | 1,439.72 | 1,516.03 | 1,485.61 | 1,397.58 | **5,838.94** |
| 458 | 1,037.36 | 1,081.85 | 1,106.38 | 1,199.45 | **4,425.05** |
| **OH - OHIO - Total** | **65,113.70** | **65,560.84** | **67,213.60** | **68,197.61** | **266,085.76** |

**STATE: OK - OKLAHOMA**

| REGISTRANT ZIP CODE 3 | 1ST QUARTER | 2ND QUARTER | 3RD QUARTER | 4TH QUARTER | TOTAL GRAMS |
|---|---|---|---|---|---|
| 730 | 2,993.73 | 3,111.14 | 3,192.76 | 3,213.37 | **12,511.00** |
| 731 | 3,424.99 | 3,332.85 | 3,723.91 | 3,722.28 | **14,204.02** |
| 734 | 203.01 | 239.02 | 250.30 | 250.19 | **942.52** |
| 735 | 752.57 | 830.88 | 853.25 | 798.00 | **3,234.71** |
| 736 | 159.22 | 194.04 | 174.89 | 175.54 | **703.69** |
| 737 | 293.52 | 275.87 | 284.21 | 277.78 | **1,131.38** |
| 738 | 105.39 | 125.46 | 122.54 | 120.40 | **473.79** |
| 739 | 18.74 | 27.34 | 25.62 | 23.17 | **94.87** |
| 740 | 3,625.51 | 3,701.43 | 3,800.78 | 3,945.67 | **15,073.39** |
| 741 | 2,672.16 | 2,472.72 | 2,667.45 | 2,723.21 | **10,535.53** |
| 743 | 529.00 | 589.81 | 634.40 | 658.99 | **2,412.20** |
| 744 | 1,611.65 | 1,739.44 | 1,880.49 | 1,953.44 | **7,185.02** |
| 745 | 439.94 | 481.40 | 513.11 | 528.01 | **1,962.46** |
| 746 | 244.60 | 253.91 | 257.68 | 277.37 | **1,033.57** |
| 747 | 302.35 | 310.36 | 296.79 | 301.01 | **1,210.51** |
| 748 | 997.28 | 994.63 | 1,049.58 | 1,028.60 | **4,070.10** |
| 749 | 590.32 | 673.92 | 689.03 | 721.96 | **2,675.24** |
| **OK - OKLAHOMA - Total** | **18,963.98** | **19,354.23** | **20,416.80** | **20,718.99** | **79,453.99** |

**STATE: OR - OREGON**

| REGISTRANT ZIP CODE 3 | 1ST QUARTER | 2ND QUARTER | 3RD QUARTER | 4TH QUARTER | TOTAL GRAMS |
|---|---|---|---|---|---|
| 970 | 2,675.26 | 2,740.26 | 2,858.62 | 2,756.02 | **11,030.17** |
| 971 | 1,471.19 | 1,342.34 | 1,436.68 | 1,364.33 | **5,614.54** |
| 972 | 4,772.71 | 4,516.25 | 4,561.79 | 4,521.63 | **18,372.38** |
| 973 | 2,355.82 | 2,351.78 | 2,448.53 | 2,467.97 | **9,624.11** |
| 974 | 2,640.48 | 2,727.75 | 2,727.07 | 2,699.60 | **10,794.90** |
| 975 | 1,732.42 | 1,717.15 | 1,876.15 | 1,858.65 | **7,184.38** |
| 976 | 156.21 | 172.78 | 168.97 | 184.50 | **682.45** |
| 977 | 697.50 | 703.02 | 790.05 | 756.04 | **2,946.60** |
| 978 | 1,109.42 | 1,134.43 | 1,196.45 | 1,147.82 | **4,588.12** |
| 979 | 91.86 | 93.40 | 97.97 | 87.61 | **370.84** |
| **OR - OREGON - Total** | **17,702.87** | **17,499.17** | **18,162.29** | **17,844.17** | **71,208.49** |

**STATE: PA - PENNSYLVANIA**

| REGISTRANT ZIP CODE 3 | 1ST QUARTER | 2ND QUARTER | 3RD QUARTER | 4TH QUARTER | TOTAL GRAMS |
|---|---|---|---|---|---|
| 150 | 4,615.73 | 4,540.07 | 4,709.19 | 4,791.91 | **18,656.90** |
| 151 | 3,514.13 | 3,681.91 | 3,637.21 | 3,712.55 | **14,545.80** |
| 152 | 6,675.94 | 6,597.29 | 6,701.82 | 6,797.28 | **26,772.33** |
| 153 | 2,139.89 | 2,330.37 | 2,401.84 | 2,387.05 | **9,259.14** |
| 154 | 2,392.76 | 2,473.08 | 2,738.42 | 2,790.09 | **10,394.33** |
| 155 | 1,419.82 | 1,054.31 | 847.57 | 922.26 | **4,243.96** |
| 156 | 3,747.88 | 3,728.32 | 3,847.82 | 3,739.45 | **15,063.47** |
| 157 | 1,212.61 | 1,297.94 | 1,305.84 | 1,504.81 | **5,321.20** |
| 158 | 770.49 | 864.79 | 881.50 | 924.72 | **3,441.50** |
| 159 | 1,504.88 | 1,464.50 | 1,579.16 | 1,538.78 | **6,087.32** |
| 160 | 1,996.59 | 1,986.80 | 2,016.87 | 1,939.59 | **7,939.86** |
| 161 | 1,697.50 | 1,876.41 | 1,884.06 | 1,914.35 | **7,372.31** |
| 162 | 1,068.71 | 1,111.56 | 1,168.32 | 1,135.34 | **4,483.93** |
| 163 | 1,178.81 | 1,238.61 | 1,299.15 | 1,335.36 | **5,051.93** |
| 164 | 344.72 | 418.80 | 411.56 | 429.38 | **1,604.47** |
| 165 | 2,037.93 | 1,974.99 | 2,037.02 | 2,137.46 | **8,187.40** |
| 166 | 2,456.04 | 2,392.05 | 2,441.22 | 2,404.02 | **9,693.34** |
| 167 | 397.03 | 420.92 | 434.61 | 475.19 | **1,727.75** |
| 168 | 1,239.84 | 1,284.58 | 1,185.04 | 1,182.18 | **4,891.64** |
| 169 | 339.26 | 364.96 | 340.63 | 375.65 | **1,420.51** |
| 170 | 2,499.84 | 2,748.04 | 2,708.38 | 2,803.99 | **10,760.25** |
| 171 | 813.72 | 818.69 | 806.41 | 902.19 | **3,341.01** |
| 172 | 1,226.41 | 1,254.91 | 1,227.53 | 1,216.33 | **4,925.17** |
| 173 | 1,384.51 | 1,394.40 | 1,390.12 | 1,446.38 | **5,615.41** |
| 174 | 788.57 | 777.64 | 819.59 | 870.34 | **3,256.14** |
| 175 | 601.12 | 768.74 | 905.72 | 1,032.25 | **3,307.82** |
| 176 | 748.43 | 755.48 | 772.93 | 867.13 | **3,143.98** |
| 177 | 2,200.55 | 2,271.97 | 2,355.09 | 2,444.47 | **9,272.07** |
| 178 | 1,356.71 | 1,494.05 | 1,515.22 | 1,587.75 | **5,953.74** |
| 179 | 668.38 | 701.04 | 684.20 | 686.01 | **2,739.62** |
| 180 | 1,119.69 | 1,226.74 | 1,175.64 | 1,308.08 | **4,830.14** |
| 181 | 785.17 | 860.26 | 826.49 | 831.44 | **3,303.36** |
| 182 | 525.37 | 568.60 | 561.90 | 605.81 | **2,261.68** |
| 183 | 649.21 | 638.38 | 669.26 | 688.10 | **2,644.96** |
| 184 | 1,527.89 | 1,611.22 | 1,632.54 | 1,661.36 | **6,433.02** |
| 185 | 2,527.51 | 2,441.62 | 2,342.03 | 2,409.30 | **9,720.46** |
| 186 | 1,297.95 | 1,374.38 | 1,352.51 | 1,393.91 | **5,418.75** |
| 187 | 2,348.87 | 2,385.18 | 2,321.13 | 2,306.57 | **9,361.75** |
| 188 | 588.93 | 626.91 | 621.11 | 630.41 | **2,467.37** |
| 189 | 1,465.08 | 1,485.64 | 1,648.07 | 1,830.37 | **6,429.16** |
| 190 | 6,377.21 | 6,562.39 | 6,619.21 | 7,585.61 | **27,144.42** |
| 191 | 12,487.74 | 12,912.92 | 12,893.55 | 13,434.08 | **51,728.29** |
| 193 | 1,281.92 | 1,227.17 | 1,268.49 | 1,261.85 | **5,039.43** |
| 194 | 2,068.32 | 2,017.99 | 2,133.29 | 2,225.56 | **8,445.16** |
| 195 | 459.93 | 465.97 | 496.70 | 584.92 | **2,007.53** |
| 196 | 778.69 | 827.09 | 835.39 | 849.60 | **3,290.77** |
| **PA - PENNSYLVANIA - Total** | **89,328.27** | **91,319.68** | **92,451.36** | **95,901.23** | **369,000.54** |

**STATE: PR - PUERTO RICO**

| REGISTRANT ZIP CODE 3 | 1ST QUARTER | 2ND QUARTER | 3RD QUARTER | 4TH QUARTER | TOTAL GRAMS |
|---|---|---|---|---|---|
| 006 | 527.02 | 440.48 | 522.48 | 527.47 | **2,017.45** |
| 007 | 1,288.33 | 1,078.97 | 1,263.79 | 1,265.15 | **4,896.24** |
| 009 | 517.23 | 430.50 | 458.88 | 516.13 | **1,922.75** |
| **PR - PUERTO RICO - Total** | **2,332.59** | **1,949.95** | **2,245.15** | **2,308.75** | **8,836.44** |

**STATE: RI - RHODE ISLAND**

| REGISTRANT ZIP CODE 3 | 1ST QUARTER | 2ND QUARTER | 3RD QUARTER | 4TH QUARTER | TOTAL GRAMS |
|---|---|---|---|---|---|
| 028 | 5,319.21 | 5,599.35 | 5,125.96 | 5,669.20 | **21,713.72** |
| 029 | 3,930.68 | 3,796.25 | 3,601.55 | 3,860.98 | **15,189.46** |
| **RI - RHODE ISLAND - Total** | **9,249.89** | **9,395.60** | **8,727.50** | **9,530.18** | **36,903.18** |

**STATE: SC - SOUTH CAROLINA**

| REGISTRANT ZIP CODE 3 | 1ST QUARTER | 2ND QUARTER | 3RD QUARTER | 4TH QUARTER | TOTAL GRAMS |
|---|---|---|---|---|---|
| 290 | 621.45 | 694.27 | 677.71 | 759.44 | **2,752.86** |
| 291 | 680.11 | 754.53 | 735.94 | 762.06 | **2,932.64** |
| 292 | 594.01 | 644.39 | 656.27 | 670.89 | **2,565.57** |
| 293 | 2,527.62 | 2,725.33 | 2,710.42 | 2,817.24 | **10,780.61** |
| 294 | 2,466.80 | 2,686.00 | 2,519.92 | 3,115.81 | **10,788.53** |
| 295 | 4,036.49 | 4,221.89 | 4,286.22 | 4,509.84 | **17,054.44** |
| 296 | 4,221.10 | 4,442.19 | 4,558.24 | 4,719.15 | **17,940.68** |
| 297 | 1,028.99 | 1,073.99 | 1,043.43 | 1,028.05 | **4,174.47** |
| 298 | 305.03 | 315.60 | 365.87 | 372.31 | **1,358.81** |
| 299 | 508.82 | 518.61 | 566.00 | 532.53 | **2,125.96** |
| **SC - SOUTH CAROLINA - Total** | **16,990.43** | **18,076.80** | **18,120.02** | **19,287.30** | **72,474.56** |

**STATE: SD - SOUTH DAKOTA**

| REGISTRANT ZIP CODE 3 | 1ST QUARTER | 2ND QUARTER | 3RD QUARTER | 4TH QUARTER | TOTAL GRAMS |
|---|---|---|---|---|---|
| 570 | 80.21 | 80.19 | 67.20 | 82.61 | **310.21** |
| 571 | 387.80 | 389.43 | 403.22 | 408.21 | **1,588.65** |
| 572 | 62.56 | 58.43 | 85.90 | 90.54 | **297.44** |
| 573 | 42.76 | 61.47 | 73.61 | 69.54 | **247.37** |
| 574 | 96.16 | 99.49 | 112.72 | 102.43 | **410.80** |
| 575 | 53.07 | 55.65 | 64.93 | 84.73 | **258.39** |
| 576 | 6.36 | 8.46 | 8.58 | 11.52 | **34.92** |
| 577 | 482.60 | 510.28 | 541.47 | 551.94 | **2,086.29** |
| **SD - SOUTH DAKOTA - Total** | **1,211.52** | **1,263.40** | **1,357.62** | **1,401.52** | **5,234.06** |

**STATE: TN - TENNESSEE**

| REGISTRANT ZIP CODE 3 | 1ST QUARTER | 2ND QUARTER | 3RD QUARTER | 4TH QUARTER | TOTAL GRAMS |
|---|---|---|---|---|---|
| 370 | 4,815.35 | 4,754.95 | 4,884.13 | 4,789.38 | **19,243.80** |
| 371 | 5,548.05 | 6,459.18 | 6,357.39 | 5,356.08 | **23,720.70** |
| 372 | 2,887.62 | 2,992.73 | 3,241.15 | 3,963.90 | **13,085.40** |
| 373 | 2,846.02 | 3,142.18 | 3,253.81 | 3,405.12 | **12,647.13** |
| 374 | 871.30 | 855.36 | 865.66 | 941.02 | **3,533.34** |
| 376 | 7,463.99 | 7,958.93 | 8,018.30 | 8,275.10 | **31,716.31** |
| 377 | 3,474.44 | 3,616.48 | 3,640.37 | 3,714.20 | **14,445.49** |
| 378 | 5,558.21 | 5,921.40 | 5,985.00 | 5,974.95 | **23,439.57** |
| 379 | 3,184.67 | 3,443.48 | 3,692.19 | 3,748.35 | **14,068.68** |
| 380 | 1,171.31 | 1,270.40 | 1,333.60 | 1,326.07 | **5,101.38** |
| 381 | 2,364.22 | 2,544.67 | 3,001.64 | 3,022.88 | **10,933.41** |
| 382 | 619.77 | 675.95 | 655.02 | 644.95 | **2,595.68** |
| 383 | 2,033.51 | 2,154.40 | 2,290.62 | 2,250.40 | **8,728.94** |
| 384 | 1,444.66 | 1,538.98 | 1,559.13 | 1,524.50 | **6,067.27** |
| 385 | 1,492.37 | 1,648.12 | 1,647.13 | 1,636.58 | **6,424.21** |
| **TN - TENNESSEE - Total** | **45,775.51** | **48,977.21** | **50,425.13** | **50,573.48** | **195,751.34** |

**STATE: TX - TEXAS**

| REGISTRANT ZIP CODE 3 | 1ST QUARTER | 2ND QUARTER | 3RD QUARTER | 4TH QUARTER | TOTAL GRAMS |
|---|---|---|---|---|---|
| 750 | 3,190.43 | 3,133.75 | 3,108.61 | 3,114.48 | **12,547.28** |
| 751 | 940.45 | 971.11 | 1,069.07 | 1,108.83 | **4,089.46** |
| 752 | 1,500.92 | 1,438.82 | 1,563.34 | 1,610.07 | **6,113.15** |
| 753 | 3.52 | 5.06 | 8.44 | 3.31 | **20.33** |
| 754 | 621.67 | 660.34 | 682.51 | 756.97 | **2,721.49** |
| 755 | 177.32 | 160.76 | 168.45 | 190.62 | **697.14** |
| 756 | 653.79 | 715.36 | 751.54 | 802.36 | **2,923.06** |
| 757 | 668.45 | 694.19 | 677.34 | 745.67 | **2,785.64** |
| 758 | 91.49 | 114.26 | 120.27 | 125.70 | **451.71** |
| 759 | 365.06 | 363.93 | 393.31 | 400.07 | **1,522.37** |
| 760 | 2,327.36 | 2,301.33 | 2,496.20 | 2,631.22 | **9,756.12** |
| 761 | 1,362.80 | 1,300.93 | 1,429.65 | 1,426.53 | **5,519.91** |
| 762 | 630.86 | 614.39 | 637.93 | 619.82 | **2,503.00** |
| 763 | 382.34 | 364.29 | 360.61 | 386.63 | **1,493.86** |
| 764 | 171.73 | 162.39 | 142.46 | 151.87 | **628.45** |
| 765 | 436.76 | 407.53 | 457.44 | 450.13 | **1,751.86** |
| 766 | 95.92 | 111.72 | 117.14 | 118.67 | **443.44** |

| | | | | | |
|---|---|---|---|---|---|
| 767 | 271.60 | 291.78 | 349.85 | 379.66 | **1,292.89** |
| 768 | 59.50 | 68.00 | 61.91 | 73.91 | **263.33** |
| 769 | 124.34 | 129.41 | 133.43 | 120.92 | **508.11** |
| 770 | 3,064.57 | 3,126.43 | 3,088.17 | 3,305.61 | **12,584.77** |
| 772 | 0 | 0.00 | 0.00 | 0 | **0.00** |
| 773 | 2,476.48 | 2,506.86 | 2,659.40 | 2,644.38 | **10,287.12** |
| 774 | 987.47 | 984.95 | 1,035.23 | 1,067.17 | **4,074.82** |
| 775 | 2,607.42 | 2,629.16 | 2,667.64 | 2,719.34 | **10,623.56** |
| 776 | 1,010.80 | 1,017.03 | 1,054.85 | 1,050.77 | **4,133.45** |
| 777 | 378.66 | 338.24 | 379.83 | 422.27 | **1,519.00** |
| 778 | 345.46 | 347.88 | 325.42 | 321.49 | **1,340.25** |
| 779 | 238.22 | 253.88 | 270.42 | 279.06 | **1,041.58** |
| 780 | 408.12 | 427.62 | 418.88 | 468.17 | **1,722.79** |
| 781 | 396.89 | 396.32 | 422.68 | 444.94 | **1,660.84** |
| 782 | 1,305.66 | 1,300.73 | 1,180.98 | 1,291.01 | **5,078.39** |
| 783 | 186.61 | 192.50 | 177.61 | 191.82 | **748.53** |
| 784 | 403.77 | 388.02 | 378.60 | 403.99 | **1,574.38** |
| 785 | 142.62 | 123.32 | 148.06 | 157.66 | **571.67** |
| 786 | 1,561.22 | 1,591.52 | 1,627.82 | 1,738.30 | **6,518.85** |
| 787 | 1,719.59 | 1,693.36 | 1,749.94 | 1,875.39 | **7,038.28** |
| 788 | 68.51 | 69.72 | 71.37 | 72.13 | **281.72** |
| 789 | 39.88 | 37.39 | 32.82 | 43.25 | **153.34** |
| 790 | 59.25 | 61.23 | 82.80 | 85.12 | **288.40** |
| 791 | 195.68 | 189.45 | 182.97 | 193.12 | **761.22** |
| 792 | 8.60 | 10.68 | 8.36 | 9.26 | **36.90** |
| 793 | 43.28 | 44.94 | 46.17 | 50.66 | **185.05** |
| 794 | 233.39 | 217.82 | 235.10 | 248.86 | **935.17** |
| 795 | 40.65 | 50.19 | 73.44 | 68.66 | **232.95** |
| 796 | 400.03 | 402.36 | 429.23 | 454.04 | **1,685.66** |
| 797 | 712.27 | 653.94 | 681.55 | 627.58 | **2,675.34** |
| 798 | 11.32 | 12.84 | 10.45 | 11.76 | **46.38** |
| 799 | 194.17 | 191.47 | 239.20 | 229.96 | **854.79** |
| **TX - TEXAS - Total** | **33,316.90** | **33,269.17** | **34,408.50** | **35,693.23** | **136,687.79** |

**STATE: UT - UTAH**

| REGISTRANT ZIP CODE 3 | 1ST QUARTER | 2ND QUARTER | 3RD QUARTER | 4TH QUARTER | **TOTAL GRAMS** |
|---|---|---|---|---|---|
| 840 | 5,212.84 | 5,195.19 | 5,191.12 | 5,269.69 | **20,868.84** |
| 841 | 3,221.08 | 3,019.67 | 3,124.79 | 3,297.97 | **12,663.52** |
| 843 | 412.47 | 438.66 | 418.25 | 450.53 | **1,719.91** |
| 844 | 962.99 | 985.50 | 993.27 | 1,019.62 | **3,961.37** |
| 845 | 351.05 | 365.54 | 397.42 | 367.23 | **1,481.23** |
| 846 | 1,406.49 | 1,375.80 | 1,421.44 | 1,436.55 | **5,640.29** |
| 847 | 658.20 | 656.60 | 673.34 | 665.72 | **2,653.85** |
| **UT - UTAH - Total** | **12,225.11** | **12,036.96** | **12,219.63** | **12,507.31** | **48,989.01** |

**STATE: VA - VIRGINIA**

| REGISTRANT ZIP CODE 3 | 1ST QUARTER | 2ND QUARTER | 3RD QUARTER | 4TH QUARTER | **TOTAL GRAMS** |
|---|---|---|---|---|---|
| 201 | 1,728.01 | 1,838.35 | 1,786.28 | 1,837.58 | **7,190.22** |
| 220 | 743.28 | 752.14 | 711.54 | 728.45 | **2,935.41** |
| 221 | 603.13 | 613.19 | 619.30 | 613.49 | **2,449.12** |
| 222 | 225.14 | 157.00 | 133.06 | 126.50 | **641.70** |
| 223 | 320.93 | 235.85 | 217.00 | 234.03 | **1,007.82** |
| 224 | 647.57 | 690.97 | 644.07 | 738.68 | **2,721.30** |
| 225 | 257.97 | 313.41 | 310.52 | 343.93 | **1,225.82** |
| 226 | 1,291.24 | 1,314.18 | 1,393.32 | 1,500.72 | **5,499.47** |
| 227 | 275.62 | 278.07 | 285.27 | 329.26 | **1,168.23** |
| 228 | 318.28 | 324.53 | 299.58 | 363.52 | **1,305.91** |
| 229 | 741.93 | 730.65 | 691.46 | 722.71 | **2,886.74** |
| 230 | 470.81 | 526.06 | 502.99 | 518.99 | **2,018.85** |
| 231 | 1,234.00 | 1,332.47 | 1,344.73 | 1,373.07 | **5,284.27** |
| 232 | 2,782.17 | 2,594.04 | 2,674.44 | 2,765.33 | **10,815.97** |
| 233 | 456.50 | 440.07 | 441.36 | 456.94 | **1,794.87** |

| | | | | | |
|---|---|---|---|---|---|
| 234 | 743.18 | 820.78 | 811.62 | 846.40 | **3,221.99** |
| 235 | 324.54 | 376.81 | 382.60 | 406.04 | **1,489.99** |
| 236 | 932.72 | 955.45 | 899.37 | 1,003.14 | **3,790.68** |
| 237 | 171.51 | 201.97 | 210.90 | 211.69 | **796.07** |
| 238 | 575.62 | 589.68 | 619.76 | 650.37 | **2,435.43** |
| 239 | 148.04 | 201.57 | 217.18 | 235.89 | **802.68** |
| 240 | 2,692.21 | 2,572.54 | 2,732.05 | 2,652.01 | **10,648.81** |
| 241 | 1,136.69 | 1,361.43 | 1,393.21 | 1,369.11 | **5,260.43** |
| 242 | 4,744.90 | 4,982.09 | 5,170.55 | 5,056.75 | **19,954.28** |
| 243 | 2,009.95 | 1,895.03 | 2,011.15 | 2,053.91 | **7,970.04** |
| 244 | 267.63 | 360.53 | 349.48 | 388.84 | **1,366.48** |
| 245 | 1,399.51 | 1,388.81 | 1,462.99 | 1,548.41 | **5,799.72** |
| 246 | 1,311.90 | 1,567.96 | 1,636.01 | 1,736.79 | **6,252.66** |
| **VA - VIRGINIA - Total** | **28,554.98** | **29,415.62** | **29,951.80** | **30,812.56** | **118,734.95** |

**STATE: VI - VIRGIN ISLANDS**

| REGISTRANT ZIP CODE 3 | 1ST QUARTER | 2ND QUARTER | 3RD QUARTER | 4TH QUARTER | TOTAL GRAMS |
|---|---|---|---|---|---|
| 008 | 28.43 | 13.14 | 29.81 | 31.94 | **103.33** |
| **VI - VIRGIN ISLANDS - Total** | **28.43** | **13.14** | **29.81** | **31.94** | **103.33** |

**STATE: VT - VERMONT**

| REGISTRANT ZIP CODE 3 | 1ST QUARTER | 2ND QUARTER | 3RD QUARTER | 4TH QUARTER | TOTAL GRAMS |
|---|---|---|---|---|---|
| 050 | 760.54 | 742.69 | 788.20 | 748.41 | **3,039.83** |
| 051 | 430.73 | 430.46 | 610.94 | 651.44 | **2,123.57** |
| 052 | 644.44 | 651.32 | 629.05 | 683.81 | **2,608.61** |
| 053 | 366.00 | 328.26 | 341.77 | 353.90 | **1,389.92** |
| 054 | 3,457.09 | 3,371.79 | 3,579.11 | 3,672.85 | **14,080.84** |
| 056 | 946.83 | 839.71 | 855.44 | 853.31 | **3,495.29** |
| 057 | 1,127.28 | 1,107.92 | 1,217.78 | 1,281.68 | **4,734.66** |
| 058 | 573.29 | 596.22 | 890.09 | 910.59 | **2,970.19** |
| **VT - VERMONT - Total** | **8,306.21** | **8,068.36** | **8,912.38** | **9,155.98** | **34,442.93** |

**STATE: WA - WASHINGTON**

| REGISTRANT ZIP CODE 3 | 1ST QUARTER | 2ND QUARTER | 3RD QUARTER | 4TH QUARTER | TOTAL GRAMS |
|---|---|---|---|---|---|
| 980 | 4,601.75 | 5,095.82 | 5,306.58 | 5,460.23 | **20,464.39** |
| 981 | 3,984.34 | 3,646.82 | 3,699.73 | 3,624.61 | **14,955.51** |
| 982 | 6,239.38 | 6,242.67 | 6,290.70 | 6,329.09 | **25,101.85** |
| 983 | 3,232.90 | 3,199.55 | 3,227.01 | 3,224.22 | **12,883.68** |
| 984 | 1,994.79 | 1,822.08 | 2,025.10 | 1,993.88 | **7,835.85** |
| 985 | 3,221.03 | 3,286.85 | 3,216.82 | 3,366.35 | **13,091.06** |
| 986 | 3,225.79 | 3,049.91 | 3,185.97 | 3,155.05 | **12,616.72** |
| 988 | 1,273.23 | 1,252.70 | 1,154.42 | 1,117.71 | **4,798.06** |
| 989 | 1,393.19 | 1,389.80 | 1,420.37 | 1,470.42 | **5,673.78** |
| 990 | 368.01 | 344.36 | 370.54 | 409.39 | **1,492.29** |
| 991 | 639.94 | 614.73 | 608.18 | 597.74 | **2,460.58** |
| 992 | 3,062.73 | 3,126.33 | 3,049.60 | 3,170.04 | **12,408.70** |
| 993 | 1,715.72 | 1,767.07 | 1,687.04 | 1,800.86 | **6,970.69** |
| 994 | 358.71 | 395.89 | 404.30 | 404.95 | **1,563.85** |
| **WA - WASHINGTON - Total** | **35,311.51** | **35,234.59** | **35,646.34** | **36,124.56** | **142,317.00** |

**STATE: WI - WISCONSIN**

| REGISTRANT ZIP CODE 3 | 1ST QUARTER | 2ND QUARTER | 3RD QUARTER | 4TH QUARTER | TOTAL GRAMS |
|---|---|---|---|---|---|
| 530 | 1,531.52 | 1,557.75 | 1,661.53 | 1,662.97 | **6,413.77** |
| 531 | 2,106.74 | 2,131.97 | 2,232.77 | 2,308.49 | **8,779.97** |
| 532 | 3,560.31 | 3,538.96 | 3,607.29 | 3,688.14 | **14,394.71** |
| 534 | 408.55 | 437.68 | 465.14 | 486.90 | **1,798.26** |
| 535 | 1,451.07 | 1,484.80 | 1,617.52 | 1,734.68 | **6,288.07** |
| 537 | 990.28 | 959.85 | 966.68 | 984.69 | **3,901.50** |
| 538 | 45.12 | 50.98 | 52.91 | 56.31 | **205.32** |
| 539 | 689.85 | 683.50 | 684.80 | 701.79 | **2,759.93** |
| 540 | 154.72 | 192.04 | 167.14 | 187.17 | **701.07** |
| 541 | 593.95 | 683.53 | 722.53 | 707.08 | **2,707.09** |
| 542 | 172.16 | 179.61 | 196.52 | 199.18 | **747.46** |
| 543 | 467.27 | 478.49 | 454.02 | 503.82 | **1,903.61** |

| | | | | | |
|---|---|---|---|---|---|
| 544 | 559.50 | 590.49 | 663.10 | 642.11 | **2,455.20** |
| 545 | 337.45 | 359.83 | 365.46 | 416.98 | **1,479.72** |
| 546 | 410.02 | 415.57 | 457.14 | 487.37 | **1,770.09** |
| 547 | 435.95 | 389.62 | 453.95 | 451.86 | **1,731.38** |
| 548 | 429.17 | 466.32 | 532.24 | 566.20 | **1,993.92** |
| 549 | 1,330.00 | 1,248.78 | 1,379.52 | 1,469.61 | **5,427.90** |
| **WI - WISCONSIN - Total** | **15,673.63** | **15,849.75** | **16,680.25** | **17,255.36** | **65,458.99** |

**STATE: WV - WEST VIRGINIA**

| REGISTRANT ZIP CODE 3 | 1ST QUARTER | 2ND QUARTER | 3RD QUARTER | 4TH QUARTER | TOTAL GRAMS |
|---|---|---|---|---|---|
| 247 | 792.82 | 829.71 | 883.95 | 1,039.23 | **3,545.71** |
| 248 | 570.11 | 663.83 | 919.18 | 994.93 | **3,148.05** |
| 249 | 108.53 | 164.15 | 199.61 | 225.55 | **697.84** |
| 250 | 489.73 | 520.26 | 557.17 | 629.95 | **2,197.10** |
| 251 | 500.33 | 542.92 | 567.43 | 628.56 | **2,239.24** |
| 252 | 317.40 | 342.25 | 405.42 | 378.90 | **1,443.97** |
| 253 | 1,603.81 | 1,537.92 | 1,637.26 | 1,691.99 | **6,470.97** |
| 254 | 1,736.06 | 1,826.95 | 1,920.84 | 1,974.76 | **7,458.61** |
| 255 | 1,636.60 | 1,708.29 | 1,902.22 | 2,026.24 | **7,273.35** |
| 256 | 1,497.10 | 1,637.52 | 1,827.98 | 1,940.60 | **6,903.20** |
| 257 | 3,205.78 | 3,730.79 | 3,488.73 | 3,115.08 | **13,540.38** |
| 258 | 2,462.64 | 2,334.93 | 2,444.93 | 2,315.35 | **9,557.86** |
| 259 | 269.23 | 323.27 | 351.77 | 393.29 | **1,337.57** |
| 260 | 1,807.75 | 1,886.50 | 1,880.17 | 1,869.22 | **7,443.64** |
| 261 | 803.59 | 769.80 | 801.17 | 779.21 | **3,153.77** |
| 262 | 244.86 | 279.85 | 312.47 | 337.47 | **1,174.66** |
| 263 | 926.47 | 879.49 | 980.34 | 1,021.66 | **3,807.95** |
| 264 | 166.00 | 159.03 | 183.41 | 199.88 | **708.31** |
| 265 | 1,389.55 | 1,320.47 | 1,443.78 | 1,557.93 | **5,711.73** |
| 266 | 193.95 | 200.78 | 221.63 | 223.20 | **839.56** |
| 267 | 386.76 | 431.88 | 465.98 | 476.33 | **1,760.94** |
| 268 | 24.88 | 28.65 | 35.03 | 46.59 | **135.15** |
| **WV - WEST VIRGINIA - Total** | **21,133.96** | **22,119.25** | **23,430.45** | **23,865.89** | **90,549.56** |

**STATE: WY - WYOMING**

| REGISTRANT ZIP CODE 3 | 1ST QUARTER | 2ND QUARTER | 3RD QUARTER | 4TH QUARTER | TOTAL GRAMS |
|---|---|---|---|---|---|
| 820 | 258.28 | 245.45 | 295.28 | 271.47 | **1,070.48** |
| 822 | 22.70 | 22.98 | 27.96 | 20.89 | **94.53** |
| 823 | 11.78 | 14.72 | 14.71 | 10.97 | **52.19** |
| 824 | 186.14 | 187.50 | 194.76 | 201.33 | **769.73** |
| 825 | 140.50 | 128.35 | 134.91 | 153.70 | **557.46** |
| 826 | 494.00 | 488.85 | 495.02 | 447.59 | **1,925.46** |
| 827 | 104.20 | 92.92 | 97.40 | 94.33 | **388.85** |
| 828 | 114.15 | 96.78 | 106.39 | 112.71 | **430.04** |
| 829 | 200.58 | 206.91 | 205.45 | 215.23 | **828.17** |
| 830 | 19.00 | 22.93 | 24.79 | 24.92 | **91.64** |
| 831 | 32.64 | 44.72 | 47.36 | 49.14 | **173.86** |
| **WY - WYOMING - Total** | **1,583.97** | **1,552.11** | **1,644.04** | **1,602.28** | **6,382.40** |

**DRUG: 9143 - OXYCODONE**

**STATE: AK - ALASKA**

| REGISTRANT ZIP CODE 3 | 1ST QUARTER | 2ND QUARTER | 3RD QUARTER | 4TH QUARTER | TOTAL GRAMS |
|---|---|---|---|---|---|
| 995 | 10,137.56 | 9,444.34 | 9,664.39 | 9,732.36 | **38,978.65** |
| 996 | 6,425.52 | 6,323.22 | 6,642.08 | 6,572.49 | **25,963.31** |
| 997 | 2,110.97 | 2,087.82 | 2,192.61 | 2,082.37 | **8,473.77** |
| 998 | 705.05 | 630.89 | 728.96 | 651.64 | **2,716.54** |
| 999 | 316.49 | 385.50 | 383.59 | 381.37 | **1,466.94** |
| **AK - ALASKA - Total** | **19,695.59** | **18,871.76** | **19,611.63** | **19,420.23** | **77,599.20** |

**STATE: AL - ALABAMA**

| REGISTRANT ZIP CODE 3 | 1ST QUARTER | 2ND QUARTER | 3RD QUARTER | 4TH QUARTER | TOTAL GRAMS |
|---|---|---|---|---|---|
| 350 | 19,257.03 | 18,426.69 | 19,518.94 | 19,526.49 | **76,729.15** |
| 351 | 11,611.56 | 11,913.94 | 11,949.92 | 11,981.19 | **47,456.61** |
| 352 | 16,454.67 | 14,704.21 | 15,538.20 | 16,013.54 | **62,710.63** |

| | | | | | |
|---|---|---|---|---|---|
| 354 | 6,583.80 | 5,736.17 | 6,228.20 | 5,822.54 | **24,370.70** |
| 355 | 8,892.13 | 9,010.74 | 9,077.69 | 8,354.47 | **35,335.03** |
| 356 | 13,355.04 | 12,029.41 | 12,779.02 | 12,062.45 | **50,225.91** |
| 357 | 7,358.14 | 7,118.83 | 7,247.78 | 7,069.94 | **28,794.69** |
| 358 | 8,850.62 | 8,282.11 | 8,696.57 | 8,217.85 | **34,047.15** |
| 359 | 15,963.30 | 15,267.26 | 15,971.14 | 15,473.22 | **62,674.92** |
| 360 | 5,725.30 | 5,613.62 | 5,880.01 | 5,934.19 | **23,153.12** |
| 361 | 4,115.68 | 3,704.13 | 4,000.93 | 4,002.98 | **15,823.71** |
| 362 | 6,414.36 | 6,313.58 | 6,045.72 | 6,256.45 | **25,030.11** |
| 363 | 7,650.60 | 7,559.48 | 7,687.64 | 7,520.87 | **30,418.59** |
| 364 | 4,030.59 | 3,775.87 | 3,850.10 | 3,868.76 | **15,525.32** |
| 365 | 11,008.59 | 10,542.57 | 10,838.91 | 11,205.07 | **43,595.14** |
| 366 | 9,330.43 | 8,639.96 | 9,252.96 | 9,436.35 | **36,659.70** |
| 367 | 1,256.41 | 1,358.06 | 1,361.35 | 1,377.60 | **5,353.42** |
| 368 | 5,634.71 | 5,648.94 | 5,841.48 | 5,828.08 | **22,953.22** |
| 369 | 368.26 | 370.57 | 422.39 | 355.28 | **1,516.50** |
| **AL - ALABAMA - Total** | **163,861.22** | **156,016.14** | **162,188.95** | **160,307.32** | **642,373.62** |

**STATE: AR - ARKANSAS**

| REGISTRANT ZIP CODE 3 | 1ST QUARTER | 2ND QUARTER | 3RD QUARTER | 4TH QUARTER | TOTAL GRAMS |
|---|---|---|---|---|---|
| 716 | 2,728.10 | 2,686.85 | 2,626.24 | 2,717.25 | **10,758.44** |
| 717 | 1,870.87 | 1,814.37 | 1,836.72 | 1,825.14 | **7,347.10** |
| 718 | 1,474.67 | 1,567.90 | 1,542.83 | 1,564.88 | **6,150.27** |
| 719 | 3,667.36 | 3,836.63 | 3,760.43 | 3,662.88 | **14,927.30** |
| 720 | 9,344.23 | 9,066.76 | 9,096.62 | 9,240.82 | **36,748.44** |
| 721 | 8,811.09 | 8,685.19 | 8,588.90 | 8,764.39 | **34,849.56** |
| 722 | 7,864.01 | 6,874.64 | 7,193.40 | 7,373.52 | **29,305.57** |
| 723 | 3,462.00 | 3,086.77 | 2,967.67 | 2,961.71 | **12,478.15** |
| 724 | 4,164.82 | 3,852.72 | 4,048.92 | 4,104.00 | **16,170.47** |
| 725 | 3,142.08 | 3,103.41 | 3,070.04 | 2,973.77 | **12,289.31** |
| 726 | 4,033.42 | 3,920.57 | 3,997.52 | 4,033.16 | **15,984.67** |
| 727 | 16,215.17 | 14,905.69 | 14,805.37 | 14,023.59 | **59,949.82** |
| 728 | 3,219.78 | 3,109.06 | 3,149.34 | 2,955.15 | **12,433.33** |
| 729 | 9,656.68 | 9,523.24 | 9,731.74 | 9,653.56 | **38,565.21** |
| **AR - ARKANSAS - Total** | **79,654.28** | **76,033.80** | **76,415.74** | **75,853.81** | **307,957.64** |

**STATE: AS - AMERICAN SAMOA**

| REGISTRANT ZIP CODE 3 | 1ST QUARTER | 2ND QUARTER | 3RD QUARTER | 4TH QUARTER | TOTAL GRAMS |
|---|---|---|---|---|---|
| 967 | 20.62 | 14.34 | 7.17 | 8.97 | **51.10** |
| **AS - AMERICAN SAMOA - Total** | **20.62** | **14.34** | **7.17** | **8.97** | **51.10** |

**STATE: AZ - ARIZONA**

| REGISTRANT ZIP CODE 3 | 1ST QUARTER | 2ND QUARTER | 3RD QUARTER | 4TH QUARTER | TOTAL GRAMS |
|---|---|---|---|---|---|
| 850 | 57,408.89 | 54,890.62 | 54,450.19 | 54,440.00 | **221,189.69** |
| 851 | 13,982.78 | 14,002.89 | 13,940.53 | 14,229.00 | **56,155.20** |
| 852 | 61,378.25 | 59,355.40 | 58,836.52 | 60,586.26 | **240,156.43** |
| 853 | 50,415.99 | 48,333.83 | 49,269.85 | 50,227.86 | **198,247.53** |
| 855 | 5,114.17 | 4,940.79 | 5,088.13 | 4,783.97 | **19,927.06** |
| 856 | 8,739.77 | 8,641.42 | 8,790.98 | 8,429.34 | **34,601.51** |
| 857 | 42,296.60 | 41,006.77 | 40,649.02 | 40,482.54 | **164,434.93** |
| 859 | 4,597.37 | 4,840.87 | 5,112.02 | 4,635.31 | **19,185.56** |
| 860 | 2,891.75 | 2,943.63 | 2,934.53 | 2,948.94 | **11,718.85** |
| 863 | 10,013.17 | 9,733.73 | 10,321.37 | 10,213.52 | **40,281.79** |
| 864 | 11,738.65 | 11,479.45 | 11,476.11 | 11,602.34 | **46,296.55** |
| 865 | 183.33 | 139.85 | 148.15 | 180.20 | **651.53** |
| **AZ - ARIZONA - Total** | **268,760.73** | **260,309.26** | **261,017.39** | **262,759.27** | **1,052,846.65** |

**STATE: CA - CALIFORNIA**

| REGISTRANT ZIP CODE 3 | 1ST QUARTER | 2ND QUARTER | 3RD QUARTER | 4TH QUARTER | TOTAL GRAMS |
|---|---|---|---|---|---|
| 900 | 17,244.53 | 16,332.62 | 16,293.70 | 15,786.63 | **65,657.47** |
| 902 | 12,244.47 | 11,962.27 | 11,957.40 | 11,574.74 | **47,738.88** |
| 903 | 1,146.68 | 1,113.93 | 1,009.20 | 797.26 | **4,067.07** |
| 904 | 3,167.89 | 3,002.62 | 3,149.06 | 3,115.34 | **12,434.90** |
| 905 | 3,248.51 | 3,020.53 | 3,384.01 | 3,283.47 | **12,936.52** |

| | | | | | |
|---|---|---|---|---|---|
| 906 | 5,786.87 | 5,104.60 | 5,386.83 | 5,451.00 | **21,729.30** |
| 907 | 5,659.61 | 5,532.86 | 5,621.21 | 5,536.11 | **22,349.79** |
| 908 | 3,662.99 | 3,652.58 | 3,685.06 | 3,416.35 | **14,416.97** |
| 910 | 3,105.46 | 2,922.76 | 3,003.18 | 3,138.20 | **12,169.60** |
| 911 | 2,276.48 | 2,122.75 | 2,184.27 | 2,159.26 | **8,742.76** |
| 912 | 2,292.73 | 2,209.26 | 2,361.28 | 2,350.22 | **9,213.49** |
| 913 | 22,702.02 | 21,638.48 | 21,620.10 | 20,950.36 | **86,910.96** |
| 914 | 5,454.97 | 4,455.59 | 4,696.55 | 4,482.34 | **19,089.45** |
| 915 | 2,215.20 | 2,027.57 | 2,180.41 | 2,013.54 | **8,436.72** |
| 916 | 1,764.70 | 1,651.08 | 1,665.92 | 1,654.10 | **6,735.81** |
| 917 | 20,809.76 | 19,123.17 | 19,212.86 | 19,032.90 | **78,178.69** |
| 918 | 560.77 | 396.84 | 372.67 | 405.11 | **1,735.39** |
| 919 | 7,980.98 | 8,341.46 | 7,805.31 | 7,765.24 | **31,892.99** |
| 920 | 33,823.73 | 32,552.47 | 33,813.12 | 33,072.49 | **133,261.82** |
| 921 | 20,039.88 | 18,437.27 | 19,607.18 | 19,312.52 | **77,396.85** |
| 922 | 19,225.25 | 18,340.53 | 18,495.75 | 18,580.23 | **74,641.76** |
| 923 | 23,041.42 | 22,148.42 | 22,276.16 | 22,568.53 | **90,034.54** |
| 924 | 2,929.34 | 2,741.61 | 2,750.42 | 2,794.26 | **11,215.63** |
| 925 | 23,501.34 | 23,723.18 | 23,892.08 | 23,286.57 | **94,403.17** |
| 926 | 18,876.56 | 18,305.91 | 18,457.70 | 18,335.30 | **73,975.48** |
| 927 | 4,713.24 | 4,666.80 | 4,584.19 | 4,390.63 | **18,354.86** |
| 928 | 15,062.41 | 14,962.57 | 15,522.95 | 14,493.76 | **60,041.69** |
| 930 | 10,845.83 | 9,813.29 | 10,505.67 | 10,013.28 | **41,178.07** |
| 931 | 1,719.91 | 1,577.20 | 1,603.02 | 1,714.78 | **6,614.91** |
| 932 | 8,565.72 | 7,978.03 | 8,321.58 | 8,418.38 | **33,283.71** |
| 933 | 10,549.81 | 10,042.11 | 10,238.32 | 9,702.31 | **40,532.55** |
| 934 | 10,297.08 | 9,877.70 | 10,565.40 | 10,384.59 | **41,124.77** |
| 935 | 14,471.35 | 13,804.35 | 13,976.45 | 13,487.28 | **55,739.42** |
| 936 | 7,025.36 | 6,831.43 | 6,699.25 | 6,823.63 | **27,379.67** |
| 937 | 9,935.04 | 9,539.85 | 9,716.07 | 9,014.60 | **38,205.56** |
| 939 | 3,574.19 | 3,441.15 | 3,380.71 | 3,277.15 | **13,673.20** |
| 940 | 9,395.10 | 8,409.16 | 9,091.58 | 8,930.49 | **35,826.33** |
| 941 | 14,629.98 | 13,661.04 | 13,774.86 | 14,246.20 | **56,312.08** |
| 943 | 2,456.10 | 2,040.08 | 2,302.06 | 2,264.29 | **9,062.53** |
| 944 | 1,337.40 | 1,244.27 | 1,181.43 | 1,276.91 | **5,040.00** |
| 945 | 32,575.11 | 30,308.07 | 31,515.46 | 31,145.67 | **125,544.31** |
| 946 | 3,287.22 | 2,880.81 | 3,063.27 | 3,393.26 | **12,624.57** |
| 947 | 780.12 | 748.17 | 802.40 | 777.92 | **3,108.61** |
| 948 | 1,328.70 | 1,222.25 | 1,087.92 | 1,064.65 | **4,703.51** |
| 949 | 6,703.17 | 6,609.41 | 6,638.17 | 6,415.98 | **26,366.72** |
| 950 | 8,413.00 | 7,560.83 | 8,088.95 | 8,096.56 | **32,159.34** |
| 951 | 6,197.38 | 5,489.54 | 6,099.05 | 5,824.60 | **23,610.57** |
| 952 | 5,612.59 | 5,479.06 | 5,390.86 | 5,486.13 | **21,968.64** |
| 953 | 21,684.64 | 20,750.04 | 20,558.33 | 20,192.32 | **83,185.33** |
| 954 | 13,900.24 | 13,642.59 | 13,830.21 | 13,974.23 | **55,347.26** |
| 955 | 3,723.44 | 3,497.42 | 3,633.06 | 3,438.57 | **14,292.49** |
| 956 | 23,263.05 | 21,910.79 | 22,545.84 | 22,196.62 | **89,916.30** |
| 957 | 4,084.92 | 3,821.74 | 3,782.88 | 3,868.85 | **15,558.38** |
| 958 | 13,693.57 | 13,110.65 | 13,761.13 | 14,108.82 | **54,674.17** |
| 959 | 10,081.16 | 9,647.51 | 9,474.48 | 9,271.39 | **38,474.54** |
| 960 | 6,696.39 | 6,217.77 | 6,499.95 | 6,473.32 | **25,887.42** |
| 961 | 1,816.45 | 1,783.04 | 1,736.16 | 1,685.78 | **7,021.43** |
| **CA - CALIFORNIA - Total** | **551,181.82** | **523,429.08** | **534,853.07** | **526,715.01** | **2,136,178.98** |

**STATE: CO - COLORADO**

| REGISTRANT ZIP CODE 3 | 1ST QUARTER | 2ND QUARTER | 3RD QUARTER | 4TH QUARTER | **TOTAL GRAMS** |
|---|---|---|---|---|---|
| 800 | 27,571.52 | 26,109.13 | 26,579.48 | 26,959.30 | **107,219.43** |
| 801 | 21,014.47 | 19,759.66 | 20,187.27 | 20,499.73 | **81,461.13** |
| 802 | 27,873.11 | 27,057.44 | 27,256.70 | 26,272.61 | **108,459.86** |
| 803 | 2,483.52 | 2,217.87 | 2,259.83 | 2,189.92 | **9,151.13** |
| 804 | 4,062.32 | 3,893.07 | 3,941.96 | 4,001.02 | **15,898.38** |

| | | | | | |
|---|---|---|---|---|---|
| 805 | 12,757.26 | 11,940.30 | 12,688.66 | 12,573.86 | **49,960.09** |
| 806 | 6,610.60 | 6,205.49 | 6,610.89 | 6,483.61 | **25,910.59** |
| 807 | 1,712.26 | 1,719.62 | 1,729.12 | 1,637.58 | **6,798.58** |
| 808 | 3,611.36 | 3,699.31 | 3,900.75 | 3,646.22 | **14,857.63** |
| 809 | 20,496.80 | 19,786.49 | 20,550.19 | 20,462.74 | **81,296.22** |
| 810 | 14,845.19 | 14,537.63 | 14,997.52 | 14,807.22 | **59,187.56** |
| 811 | 1,552.69 | 1,583.77 | 1,682.61 | 1,656.43 | **6,475.50** |
| 812 | 2,479.82 | 2,660.83 | 2,719.89 | 2,766.59 | **10,627.13** |
| 813 | 949.38 | 978.90 | 1,001.12 | 1,017.06 | **3,946.45** |
| 814 | 1,702.30 | 1,652.01 | 1,682.87 | 1,720.77 | **6,757.94** |
| 815 | 3,412.39 | 3,202.33 | 3,332.87 | 3,235.84 | **13,183.44** |
| 816 | 2,676.50 | 2,543.61 | 2,674.51 | 2,738.37 | **10,632.99** |
| **CO - COLORADO - Total** | **155,811.48** | **149,547.46** | **153,796.25** | **152,668.87** | **611,824.06** |

**STATE: CT - CONNECTICUT**

| REGISTRANT ZIP CODE 3 | 1ST QUARTER | 2ND QUARTER | 3RD QUARTER | 4TH QUARTER | TOTAL GRAMS |
|---|---|---|---|---|---|
| 060 | 19,526.26 | 18,932.17 | 19,630.74 | 19,076.32 | **77,165.48** |
| 061 | 5,791.72 | 5,267.80 | 5,897.15 | 5,990.78 | **22,947.45** |
| 062 | 3,429.25 | 3,426.34 | 3,582.38 | 3,737.41 | **14,175.38** |
| 063 | 8,878.39 | 9,082.11 | 9,576.95 | 9,103.07 | **36,640.52** |
| 064 | 31,887.70 | 30,292.07 | 30,285.49 | 30,685.27 | **123,150.53** |
| 065 | 13,128.80 | 12,365.71 | 12,924.79 | 13,025.19 | **51,444.50** |
| 066 | 7,788.15 | 7,601.21 | 7,932.34 | 7,811.05 | **31,132.75** |
| 067 | 12,319.64 | 12,074.88 | 12,437.11 | 12,008.94 | **48,840.57** |
| 068 | 11,762.75 | 10,608.05 | 11,099.77 | 11,053.77 | **44,524.34** |
| 069 | 2,275.58 | 2,175.81 | 2,084.43 | 2,117.46 | **8,653.28** |
| **CT - CONNECTICUT - Total** | **116,788.24** | **111,826.14** | **115,451.16** | **114,609.27** | **458,674.80** |

**STATE: DC - DISTRICT OF COLUMBIA**

| REGISTRANT ZIP CODE 3 | 1ST QUARTER | 2ND QUARTER | 3RD QUARTER | 4TH QUARTER | TOTAL GRAMS |
|---|---|---|---|---|---|
| 200 | 9,959.57 | 8,593.08 | 9,252.53 | 9,104.95 | **36,910.14** |
| 203 | 17.48 | 0 | 7.62 | 8.07 | **33.17** |
| 204 | 820.57 | 671.84 | 632.21 | 773.32 | **2,897.94** |
| 205 | 0 | 4.03 | 0 | 0.90 | **4.93** |
| **DC - DISTRICT OF COLUMBIA - Total** | **10,797.62** | **9,268.95** | **9,892.37** | **9,887.24** | **39,846.18** |

**STATE: DE - DELAWARE**

| REGISTRANT ZIP CODE 3 | 1ST QUARTER | 2ND QUARTER | 3RD QUARTER | 4TH QUARTER | TOTAL GRAMS |
|---|---|---|---|---|---|
| 197 | 15,077.62 | 14,644.93 | 14,944.20 | 15,179.43 | **59,846.18** |
| 198 | 10,170.29 | 9,904.73 | 10,168.20 | 10,315.64 | **40,558.86** |
| 199 | 19,864.14 | 19,414.64 | 19,758.22 | 19,432.46 | **78,469.46** |
| **DE - DELAWARE - Total** | **45,112.05** | **43,964.30** | **44,870.62** | **44,927.53** | **178,874.50** |

**STATE: FL - FLORIDA**

| REGISTRANT ZIP CODE 3 | 1ST QUARTER | 2ND QUARTER | 3RD QUARTER | 4TH QUARTER | TOTAL GRAMS |
|---|---|---|---|---|---|
| 320 | 27,392.81 | 26,698.99 | 28,120.02 | 27,438.82 | **109,650.64** |
| 321 | 27,956.18 | 27,102.72 | 28,699.11 | 28,557.51 | **112,315.51** |
| 322 | 49,861.33 | 47,999.64 | 50,332.55 | 50,373.60 | **198,567.13** |
| 323 | 11,352.63 | 10,794.85 | 11,418.85 | 11,301.05 | **44,867.37** |
| 324 | 13,665.37 | 12,093.74 | 12,411.19 | 12,614.54 | **50,784.84** |
| 325 | 23,855.66 | 23,081.11 | 24,145.95 | 24,326.91 | **95,409.63** |
| 326 | 18,962.32 | 18,080.78 | 18,497.96 | 18,244.90 | **73,785.95** |
| 327 | 38,357.19 | 36,838.64 | 37,975.68 | 37,482.86 | **150,654.37** |
| 328 | 25,322.06 | 24,608.20 | 26,521.68 | 26,055.70 | **102,507.64** |
| 329 | 40,480.44 | 38,821.38 | 39,222.14 | 39,270.23 | **157,794.19** |
| 330 | 54,454.45 | 52,829.93 | 55,982.37 | 56,332.27 | **219,599.03** |
| 331 | 45,217.03 | 44,310.18 | 46,621.57 | 45,942.47 | **182,091.25** |
| 333 | 34,985.38 | 34,252.03 | 35,094.01 | 35,633.43 | **139,964.86** |
| 334 | 77,709.26 | 75,030.23 | 78,513.80 | 78,433.13 | **309,686.41** |
| 335 | 33,481.54 | 32,276.44 | 34,076.16 | 34,715.99 | **134,550.12** |
| 336 | 48,018.63 | 46,390.77 | 47,202.15 | 47,031.58 | **188,643.14** |
| 337 | 42,235.07 | 41,664.23 | 42,058.49 | 43,048.97 | **169,006.76** |
| 338 | 25,867.58 | 25,890.96 | 26,505.45 | 26,689.96 | **104,953.95** |
| 339 | 37,699.17 | 35,465.71 | 36,815.22 | 38,160.43 | **148,140.53** |

| | | | | | |
|---|---|---|---|---|---|
| 341 | 15,556.62 | 14,987.87 | 14,862.03 | 15,315.15 | **60,721.67** |
| 342 | 38,094.52 | 37,268.94 | 37,618.25 | 38,245.92 | **151,227.63** |
| 344 | 24,587.26 | 23,903.24 | 24,617.24 | 24,838.49 | **97,946.23** |
| 346 | 45,648.38 | 43,542.42 | 46,279.04 | 47,298.77 | **182,768.61** |
| 347 | 21,809.50 | 21,455.74 | 22,583.52 | 22,810.47 | **88,659.23** |
| 349 | 27,082.25 | 26,224.58 | 26,721.72 | 26,421.75 | **106,450.31** |
| **FL - FLORIDA - Total** | **849,652.64** | **821,613.32** | **852,896.15** | **856,584.90** | **3,380,747.01** |

**STATE: GA - GEORGIA**

| REGISTRANT ZIP CODE 3 | 1ST QUARTER | 2ND QUARTER | 3RD QUARTER | 4TH QUARTER | TOTAL GRAMS |
|---|---|---|---|---|---|
| 300 | 62,480.17 | 59,404.59 | 61,048.92 | 61,682.17 | **244,615.85** |
| 301 | 52,246.12 | 51,113.44 | 52,677.65 | 51,872.27 | **207,909.48** |
| 302 | 33,384.26 | 32,516.39 | 33,491.50 | 33,945.42 | **133,337.56** |
| 303 | 22,720.07 | 21,237.83 | 22,385.05 | 22,660.84 | **89,003.79** |
| 304 | 7,327.71 | 7,181.00 | 7,237.64 | 7,395.67 | **29,142.01** |
| 305 | 33,122.12 | 32,075.66 | 33,193.48 | 33,559.56 | **131,950.81** |
| 306 | 22,414.84 | 20,803.62 | 21,332.73 | 20,920.53 | **85,471.72** |
| 307 | 11,809.91 | 11,180.32 | 11,440.81 | 11,478.89 | **45,909.93** |
| 308 | 6,416.78 | 5,994.37 | 5,588.79 | 5,722.64 | **23,722.57** |
| 309 | 11,434.59 | 10,459.79 | 10,652.23 | 10,027.89 | **42,574.50** |
| 310 | 17,875.73 | 17,308.48 | 18,091.30 | 17,979.11 | **71,254.62** |
| 312 | 7,739.82 | 7,720.60 | 8,044.43 | 7,714.00 | **31,218.85** |
| 313 | 5,826.15 | 5,271.13 | 5,757.34 | 5,426.47 | **22,281.09** |
| 314 | 7,026.02 | 6,491.19 | 6,650.56 | 6,557.91 | **26,725.68** |
| 315 | 13,882.89 | 13,006.33 | 14,027.25 | 13,156.18 | **54,072.64** |
| 316 | 6,946.45 | 6,291.69 | 6,706.77 | 6,921.40 | **26,866.31** |
| 317 | 12,088.42 | 11,368.22 | 12,078.83 | 11,875.90 | **47,411.38** |
| 318 | 949.66 | 848.60 | 928.61 | 894.69 | **3,621.57** |
| 319 | 6,273.40 | 6,030.48 | 6,577.77 | 6,115.51 | **24,997.16** |
| 398 | 2,545.36 | 2,646.31 | 2,753.41 | 2,759.50 | **10,704.59** |
| **GA - GEORGIA - Total** | **344,510.47** | **328,950.02** | **340,665.07** | **338,666.54** | **1,352,792.10** |

**STATE: GU - GUAM**

| REGISTRANT ZIP CODE 3 | 1ST QUARTER | 2ND QUARTER | 3RD QUARTER | 4TH QUARTER | TOTAL GRAMS |
|---|---|---|---|---|---|
| 969 | 672.29 | 609.08 | 751.00 | 708.50 | **2,740.87** |
| **GU - GUAM - Total** | **672.29** | **609.08** | **751.00** | **708.50** | **2,740.87** |

**STATE: HI - HAWAII**

| REGISTRANT ZIP CODE 3 | 1ST QUARTER | 2ND QUARTER | 3RD QUARTER | 4TH QUARTER | TOTAL GRAMS |
|---|---|---|---|---|---|
| 967 | 19,518.09 | 18,661.38 | 18,488.71 | 17,515.89 | **74,184.07** |
| 968 | 8,721.10 | 7,708.79 | 7,772.75 | 7,299.08 | **31,501.71** |
| **HI - HAWAII - Total** | **28,239.19** | **26,370.16** | **26,261.46** | **24,814.97** | **105,685.78** |

**STATE: IA - IOWA**

| REGISTRANT ZIP CODE 3 | 1ST QUARTER | 2ND QUARTER | 3RD QUARTER | 4TH QUARTER | TOTAL GRAMS |
|---|---|---|---|---|---|
| 500 | 3,008.95 | 2,856.06 | 3,061.03 | 2,937.06 | **11,863.10** |
| 501 | 2,149.65 | 2,085.86 | 2,152.79 | 2,064.96 | **8,453.25** |
| 502 | 2,311.27 | 2,360.34 | 2,369.21 | 2,404.55 | **9,445.38** |
| 503 | 6,648.61 | 6,336.93 | 6,596.49 | 6,630.41 | **26,212.44** |
| 504 | 1,300.91 | 1,253.63 | 1,246.57 | 1,302.94 | **5,104.06** |
| 505 | 1,073.26 | 993.45 | 1,017.15 | 1,042.68 | **4,126.54** |
| 506 | 1,084.02 | 983.12 | 1,223.76 | 1,072.35 | **4,363.24** |
| 507 | 1,013.68 | 949.34 | 1,045.14 | 1,117.66 | **4,125.81** |
| 508 | 340.88 | 324.32 | 369.91 | 357.48 | **1,392.58** |
| 510 | 575.85 | 547.99 | 583.67 | 596.83 | **2,304.34** |
| 511 | 1,399.91 | 1,346.31 | 1,252.19 | 1,292.26 | **5,290.67** |
| 512 | 275.31 | 316.38 | 279.38 | 306.00 | **1,177.07** |
| 513 | 406.55 | 349.51 | 381.17 | 358.12 | **1,495.34** |
| 514 | 452.15 | 481.53 | 486.63 | 433.42 | **1,853.74** |
| 515 | 1,926.93 | 1,986.40 | 2,031.52 | 2,140.86 | **8,085.70** |
| 516 | 303.16 | 350.97 | 332.83 | 352.59 | **1,339.54** |
| 520 | 975.37 | 871.04 | 894.43 | 1,072.20 | **3,813.04** |
| 521 | 322.70 | 290.06 | 328.39 | 307.88 | **1,249.03** |
| 522 | 2,271.79 | 1,883.51 | 2,163.76 | 2,129.65 | **8,448.71** |

| | | | | | |
|---|---|---|---|---|---|
| 523 | 817.15 | 713.80 | 832.88 | 760.03 | **3,123.87** |
| 524 | 1,619.34 | 1,399.92 | 1,486.59 | 1,574.96 | **6,080.82** |
| 525 | 1,374.77 | 1,386.42 | 1,529.29 | 1,458.29 | **5,748.76** |
| 526 | 1,489.57 | 1,387.96 | 1,500.03 | 1,485.79 | **5,863.36** |
| 527 | 1,003.38 | 903.85 | 941.50 | 992.87 | **3,841.61** |
| 528 | 969.89 | 826.86 | 883.74 | 845.28 | **3,525.77** |
| **IA - IOWA - Total** | **35,115.06** | **33,185.56** | **34,990.03** | **35,037.12** | **138,327.77** |

**STATE: ID - IDAHO**

| REGISTRANT ZIP CODE 3 | 1ST QUARTER | 2ND QUARTER | 3RD QUARTER | 4TH QUARTER | TOTAL GRAMS |
|---|---|---|---|---|---|
| 832 | 6,017.07 | 5,757.38 | 5,921.55 | 5,895.91 | **23,591.92** |
| 833 | 5,582.25 | 5,006.35 | 5,243.06 | 5,071.90 | **20,903.55** |
| 834 | 4,681.30 | 4,555.03 | 5,066.77 | 5,173.88 | **19,476.97** |
| 835 | 1,548.00 | 1,461.72 | 1,522.07 | 1,437.76 | **5,969.55** |
| 836 | 13,195.38 | 12,679.77 | 12,931.77 | 12,852.44 | **51,659.36** |
| 837 | 9,130.76 | 8,624.15 | 8,961.90 | 8,554.35 | **35,271.16** |
| 838 | 5,505.85 | 5,612.83 | 5,751.90 | 5,985.72 | **22,856.31** |
| **ID - IDAHO - Total** | **45,660.61** | **43,697.23** | **45,399.03** | **44,971.95** | **179,728.81** |

**STATE: IL - ILLINOIS**

| REGISTRANT ZIP CODE 3 | 1ST QUARTER | 2ND QUARTER | 3RD QUARTER | 4TH QUARTER | TOTAL GRAMS |
|---|---|---|---|---|---|
| 600 | 16,077.80 | 14,758.51 | 15,262.79 | 15,375.59 | **61,474.68** |
| 601 | 10,820.02 | 10,284.58 | 10,365.60 | 10,791.75 | **42,261.95** |
| 602 | 333.52 | 409.20 | 425.73 | 391.13 | **1,559.58** |
| 603 | 481.73 | 470.25 | 481.72 | 581.99 | **2,015.69** |
| 604 | 14,191.29 | 13,524.11 | 13,937.50 | 13,755.17 | **55,408.08** |
| 605 | 7,042.45 | 6,506.97 | 6,553.39 | 6,876.41 | **26,979.22** |
| 606 | 9,185.10 | 8,501.45 | 8,866.05 | 9,172.14 | **35,724.74** |
| 607 | 1,289.69 | 1,220.52 | 1,184.63 | 1,119.51 | **4,814.35** |
| 608 | 449.01 | 494.09 | 564.61 | 525.25 | **2,032.96** |
| 609 | 1,878.37 | 1,805.40 | 1,922.77 | 1,908.86 | **7,515.40** |
| 610 | 1,954.37 | 1,963.72 | 2,108.45 | 1,954.12 | **7,980.66** |
| 611 | 2,563.05 | 2,472.93 | 2,637.04 | 2,580.45 | **10,253.47** |
| 612 | 1,031.13 | 867.02 | 1,071.13 | 1,024.11 | **3,993.38** |
| 613 | 1,241.83 | 1,195.22 | 1,250.81 | 1,184.20 | **4,872.07** |
| 614 | 765.42 | 734.50 | 723.55 | 796.50 | **3,019.97** |
| 615 | 1,401.11 | 1,440.80 | 1,404.35 | 1,451.13 | **5,697.39** |
| 616 | 2,092.23 | 2,081.07 | 2,057.67 | 2,016.07 | **8,247.04** |
| 617 | 1,438.74 | 1,415.53 | 1,576.55 | 1,545.35 | **5,976.16** |
| 618 | 3,022.81 | 2,939.13 | 2,900.13 | 2,823.23 | **11,685.29** |
| 619 | 582.85 | 550.22 | 561.99 | 570.34 | **2,265.40** |
| 620 | 5,176.56 | 4,831.67 | 5,084.72 | 4,940.07 | **20,033.03** |
| 622 | 6,633.56 | 6,250.05 | 6,629.33 | 6,499.81 | **26,012.75** |
| 623 | 1,401.04 | 1,409.73 | 1,387.28 | 1,484.52 | **5,682.57** |
| 624 | 1,471.93 | 1,393.33 | 1,477.52 | 1,350.29 | **5,693.08** |
| 625 | 1,325.02 | 1,253.32 | 1,355.73 | 1,365.08 | **5,299.15** |
| 626 | 1,111.66 | 1,055.40 | 1,215.17 | 1,132.20 | **4,514.42** |
| 627 | 1,075.05 | 977.12 | 1,105.55 | 1,113.43 | **4,271.15** |
| 628 | 3,546.26 | 3,322.84 | 3,505.45 | 3,580.65 | **13,955.19** |
| 629 | 5,401.45 | 5,121.83 | 5,098.47 | 5,302.99 | **20,924.74** |
| **IL - ILLINOIS - Total** | **104,985.04** | **99,250.52** | **102,715.68** | **103,212.35** | **410,163.59** |

**STATE: IN - INDIANA**

| REGISTRANT ZIP CODE 3 | 1ST QUARTER | 2ND QUARTER | 3RD QUARTER | 4TH QUARTER | TOTAL GRAMS |
|---|---|---|---|---|---|
| 460 | 10,452.84 | 10,122.46 | 10,735.39 | 10,841.47 | **42,152.16** |
| 461 | 15,430.41 | 14,953.02 | 14,703.02 | 14,619.34 | **59,705.79** |
| 462 | 28,966.75 | 27,575.87 | 28,541.13 | 28,949.13 | **114,032.88** |
| 463 | 12,788.72 | 12,174.43 | 12,677.68 | 12,411.47 | **50,052.29** |
| 464 | 1,637.75 | 1,566.53 | 1,660.77 | 1,601.37 | **6,466.43** |
| 465 | 11,693.64 | 11,622.05 | 11,953.31 | 12,028.02 | **47,297.02** |
| 466 | 4,172.92 | 4,058.90 | 4,055.82 | 3,958.24 | **16,245.88** |
| 467 | 7,086.78 | 6,836.11 | 7,161.13 | 7,249.68 | **28,333.69** |
| 468 | 10,369.30 | 10,409.75 | 10,990.32 | 11,365.83 | **43,135.20** |

| | | | | | |
|---|---|---|---|---|---|
| 469 | 7,230.50 | 7,298.44 | 7,304.06 | 7,856.29 | **29,689.29** |
| 470 | 5,076.70 | 4,943.39 | 5,111.31 | 5,099.99 | **20,231.39** |
| 471 | 11,758.70 | 10,927.48 | 11,163.25 | 11,238.14 | **45,087.56** |
| 472 | 4,875.50 | 4,662.19 | 4,853.94 | 5,017.46 | **19,409.09** |
| 473 | 10,071.40 | 10,231.70 | 10,639.59 | 10,744.32 | **41,687.00** |
| 474 | 5,546.36 | 5,343.18 | 5,414.03 | 5,400.75 | **21,704.32** |
| 475 | 3,861.21 | 3,587.75 | 3,666.39 | 3,836.79 | **14,952.14** |
| 476 | 2,999.10 | 2,963.06 | 3,195.49 | 3,254.96 | **12,412.60** |
| 477 | 8,131.19 | 7,673.60 | 7,929.35 | 7,542.23 | **31,276.38** |
| 478 | 2,426.54 | 2,141.16 | 2,216.54 | 2,344.09 | **9,128.33** |
| 479 | 4,486.37 | 4,193.59 | 4,476.27 | 4,346.03 | **17,502.26** |
| **IN - INDIANA - Total** | **169,062.67** | **163,284.66** | **168,448.77** | **169,705.60** | **670,501.70** |

**STATE: KS - KANSAS**

| REGISTRANT ZIP CODE 3 | 1ST QUARTER | 2ND QUARTER | 3RD QUARTER | 4TH QUARTER | TOTAL GRAMS |
|---|---|---|---|---|---|
| 660 | 11,989.22 | 11,514.81 | 12,103.59 | 11,963.00 | **47,570.62** |
| 661 | 4,872.23 | 4,563.73 | 4,833.62 | 4,974.86 | **19,244.43** |
| 662 | 11,220.16 | 10,580.29 | 10,915.92 | 11,135.26 | **43,851.63** |
| 664 | 1,771.32 | 1,764.11 | 1,769.67 | 1,541.82 | **6,846.91** |
| 665 | 2,251.61 | 2,074.20 | 2,105.90 | 2,132.52 | **8,564.22** |
| 666 | 4,824.85 | 4,650.60 | 4,846.62 | 4,817.59 | **19,139.66** |
| 667 | 3,620.57 | 3,315.97 | 3,467.75 | 3,360.39 | **13,764.68** |
| 668 | 956.33 | 931.97 | 981.58 | 960.04 | **3,829.92** |
| 669 | 561.04 | 534.63 | 541.36 | 541.11 | **2,178.13** |
| 670 | 6,555.30 | 6,332.98 | 6,438.85 | 6,481.28 | **25,808.41** |
| 671 | 2,935.98 | 2,745.86 | 2,912.70 | 2,910.82 | **11,505.35** |
| 672 | 18,218.15 | 18,070.12 | 18,514.08 | 18,402.76 | **73,205.11** |
| 673 | 1,698.78 | 1,607.53 | 1,717.13 | 1,816.71 | **6,840.15** |
| 674 | 3,866.49 | 3,860.52 | 4,083.81 | 3,950.06 | **15,760.87** |
| 675 | 4,363.50 | 4,150.22 | 4,509.38 | 4,467.65 | **17,490.75** |
| 676 | 1,724.80 | 1,633.83 | 1,648.79 | 1,631.76 | **6,639.18** |
| 677 | 1,113.73 | 1,017.57 | 1,059.28 | 965.57 | **4,156.15** |
| 678 | 3,168.11 | 2,956.26 | 3,051.35 | 2,819.58 | **11,995.31** |
| 679 | 958.89 | 824.55 | 867.48 | 848.30 | **3,499.23** |
| **KS - KANSAS - Total** | **86,671.05** | **83,129.73** | **86,368.85** | **85,721.08** | **341,890.72** |

**STATE: KY - KENTUCKY**

| REGISTRANT ZIP CODE 3 | 1ST QUARTER | 2ND QUARTER | 3RD QUARTER | 4TH QUARTER | TOTAL GRAMS |
|---|---|---|---|---|---|
| 400 | 6,672.99 | 6,649.22 | 6,887.36 | 6,786.26 | **26,995.84** |
| 401 | 4,003.76 | 3,600.85 | 3,858.72 | 3,894.40 | **15,357.72** |
| 402 | 27,513.49 | 25,019.37 | 26,885.96 | 26,497.80 | **105,916.63** |
| 403 | 11,700.63 | 10,890.98 | 11,521.87 | 10,932.87 | **45,046.34** |
| 404 | 6,604.11 | 5,889.59 | 6,207.46 | 6,173.38 | **24,874.54** |
| 405 | 8,241.03 | 7,953.87 | 8,285.32 | 8,002.07 | **32,482.29** |
| 406 | 1,198.17 | 1,218.90 | 1,220.45 | 1,194.97 | **4,832.49** |
| 407 | 5,210.37 | 4,946.54 | 4,969.01 | 4,612.82 | **19,738.74** |
| 408 | 1,259.71 | 1,105.45 | 1,240.52 | 1,353.16 | **4,958.83** |
| 409 | 3,286.76 | 3,121.67 | 3,068.10 | 2,884.28 | **12,360.81** |
| 410 | 21,421.56 | 20,709.15 | 21,781.54 | 21,641.02 | **85,553.27** |
| 411 | 3,490.13 | 3,031.67 | 3,221.47 | 3,204.11 | **12,947.38** |
| 412 | 1,895.26 | 1,832.91 | 1,752.83 | 1,597.11 | **7,078.11** |
| 413 | 1,461.45 | 1,290.22 | 1,288.73 | 1,292.06 | **5,332.45** |
| 414 | 861.54 | 769.53 | 757.73 | 744.43 | **3,133.23** |
| 415 | 3,435.33 | 2,945.40 | 3,155.02 | 3,189.91 | **12,725.65** |
| 416 | 3,381.20 | 3,365.49 | 3,365.00 | 3,262.68 | **13,374.38** |
| 417 | 3,831.06 | 3,670.37 | 3,665.01 | 3,887.41 | **15,053.84** |
| 418 | 1,688.20 | 1,487.78 | 1,481.52 | 1,558.15 | **6,215.65** |
| 420 | 9,929.71 | 9,399.97 | 9,835.80 | 9,652.13 | **38,817.61** |
| 421 | 7,920.85 | 7,991.16 | 8,152.76 | 8,102.35 | **32,167.12** |
| 422 | 4,363.86 | 3,941.52 | 4,167.68 | 3,802.13 | **16,275.19** |
| 423 | 5,193.65 | 4,637.66 | 5,019.94 | 4,956.78 | **19,808.03** |
| 424 | 6,245.73 | 5,689.86 | 6,072.00 | 5,723.99 | **23,731.57** |

| | | | | | |
|---|---|---|---|---|---|
| 425 | 2,377.94 | 2,188.97 | 2,270.52 | 2,323.61 | **9,161.05** |
| 426 | 2,676.06 | 2,646.35 | 2,401.27 | 2,403.53 | **10,127.20** |
| 427 | 6,270.58 | 6,093.79 | 6,145.58 | 6,221.56 | **24,731.51** |
| **KY - KENTUCKY - Total** | **162,135.14** | **152,088.24** | **158,679.16** | **155,894.94** | **628,797.49** |

**STATE: LA - LOUISIANA**

| REGISTRANT ZIP CODE 3 | 1ST QUARTER | 2ND QUARTER | 3RD QUARTER | 4TH QUARTER | **TOTAL GRAMS** |
|---|---|---|---|---|---|
| 700 | 19,649.52 | 18,163.24 | 18,913.44 | 18,853.62 | **75,579.83** |
| 701 | 11,947.28 | 10,724.80 | 11,838.29 | 12,086.39 | **46,596.77** |
| 703 | 6,266.33 | 5,860.21 | 6,137.17 | 6,189.75 | **24,453.47** |
| 704 | 19,566.11 | 19,162.36 | 19,934.27 | 20,042.09 | **78,704.84** |
| 705 | 15,668.83 | 14,804.01 | 15,300.40 | 14,843.56 | **60,616.81** |
| 706 | 4,552.27 | 4,106.91 | 4,034.40 | 4,240.65 | **16,934.23** |
| 707 | 15,763.46 | 15,563.10 | 15,511.59 | 15,374.16 | **62,212.30** |
| 708 | 11,694.15 | 10,428.92 | 11,179.27 | 11,359.57 | **44,661.92** |
| 710 | 3,832.80 | 3,758.62 | 3,845.07 | 3,916.90 | **15,353.39** |
| 711 | 11,659.83 | 11,466.83 | 12,031.37 | 12,304.11 | **47,462.14** |
| 712 | 5,721.14 | 5,513.03 | 5,700.90 | 5,529.07 | **22,464.13** |
| 713 | 7,925.75 | 7,582.51 | 7,552.81 | 7,624.21 | **30,685.28** |
| 714 | 4,680.74 | 4,740.86 | 4,686.69 | 4,647.38 | **18,755.67** |
| **LA - LOUISIANA - Total** | **138,928.22** | **131,875.41** | **136,665.68** | **137,011.46** | **544,480.77** |

**STATE: MA - MASSACHUSETTS**

| REGISTRANT ZIP CODE 3 | 1ST QUARTER | 2ND QUARTER | 3RD QUARTER | 4TH QUARTER | **TOTAL GRAMS** |
|---|---|---|---|---|---|
| 010 | 17,360.26 | 16,699.01 | 17,130.02 | 16,622.74 | **67,812.03** |
| 011 | 6,929.53 | 6,438.72 | 6,970.47 | 6,894.09 | **27,232.82** |
| 012 | 3,254.02 | 3,297.17 | 3,339.91 | 3,371.41 | **13,262.51** |
| 013 | 3,160.76 | 2,985.02 | 3,035.96 | 3,038.48 | **12,220.23** |
| 014 | 5,700.89 | 5,318.75 | 5,943.25 | 5,800.37 | **22,763.26** |
| 015 | 7,946.21 | 7,527.60 | 7,857.60 | 8,179.79 | **31,511.20** |
| 016 | 6,878.15 | 6,586.56 | 6,777.54 | 6,479.76 | **26,722.01** |
| 017 | 7,544.67 | 6,307.10 | 7,134.98 | 7,224.66 | **28,211.41** |
| 018 | 20,364.71 | 19,005.81 | 19,696.53 | 19,248.99 | **78,316.04** |
| 019 | 10,195.65 | 9,104.91 | 9,748.71 | 9,943.71 | **38,992.98** |
| 020 | 7,722.31 | 7,326.57 | 7,712.84 | 7,525.34 | **30,287.07** |
| 021 | 25,824.66 | 22,924.12 | 25,403.76 | 25,084.79 | **99,237.34** |
| 022 | 2,023.24 | 1,811.64 | 1,902.63 | 1,928.36 | **7,665.88** |
| 023 | 12,342.85 | 12,273.39 | 12,269.10 | 12,089.96 | **48,975.29** |
| 024 | 3,968.82 | 3,764.01 | 3,939.25 | 3,885.82 | **15,557.91** |
| 025 | 3,709.97 | 3,759.61 | 3,801.58 | 3,933.61 | **15,204.77** |
| 026 | 4,739.85 | 4,523.75 | 4,908.26 | 4,561.98 | **18,733.84** |
| 027 | 22,779.37 | 22,427.07 | 23,186.28 | 22,567.31 | **90,960.03** |
| **MA - MASSACHUSETTS - Total** | **172,445.93** | **162,080.81** | **170,758.69** | **168,381.18** | **673,666.61** |

**STATE: MD - MARYLAND**

| REGISTRANT ZIP CODE 3 | 1ST QUARTER | 2ND QUARTER | 3RD QUARTER | 4TH QUARTER | **TOTAL GRAMS** |
|---|---|---|---|---|---|
| 206 | 17,832.62 | 17,191.70 | 17,797.70 | 17,624.92 | **70,446.94** |
| 207 | 20,377.67 | 19,515.26 | 20,302.39 | 19,386.42 | **79,581.75** |
| 208 | 15,363.62 | 14,043.46 | 14,422.16 | 13,836.88 | **57,666.11** |
| 209 | 3,253.11 | 3,090.94 | 3,090.94 | 3,104.20 | **12,539.19** |
| 210 | 38,314.18 | 36,879.52 | 37,159.37 | 36,925.14 | **149,278.21** |
| 211 | 15,191.29 | 15,229.23 | 15,639.99 | 15,362.70 | **61,423.20** |
| 212 | 68,761.95 | 65,652.13 | 66,795.39 | 67,020.88 | **268,230.35** |
| 214 | 3,630.77 | 3,231.85 | 3,255.69 | 3,490.37 | **13,608.68** |
| 215 | 6,878.05 | 6,602.37 | 6,581.31 | 6,347.63 | **26,409.36** |
| 216 | 7,807.18 | 7,573.43 | 7,518.09 | 7,850.10 | **30,748.79** |
| 217 | 22,404.34 | 21,673.16 | 21,883.22 | 21,639.07 | **87,599.80** |
| 218 | 6,777.54 | 6,529.63 | 6,840.25 | 6,689.93 | **26,837.35** |
| 219 | 5,013.78 | 4,546.23 | 4,522.69 | 4,454.44 | **18,537.14** |
| **MD - MARYLAND - Total** | **231,606.11** | **221,758.90** | **225,809.20** | **223,732.67** | **902,906.87** |

**STATE: ME - MAINE**

| REGISTRANT ZIP CODE 3 | 1ST QUARTER | 2ND QUARTER | 3RD QUARTER | 4TH QUARTER | **TOTAL GRAMS** |
|---|---|---|---|---|---|
| 039 | 646.93 | 622.82 | 538.29 | 554.89 | **2,362.93** |

| | | | | | |
|---|---|---|---|---|---|
| 040 | 9,648.43 | 9,723.45 | 10,141.82 | 10,061.20 | **39,574.90** |
| 041 | 2,520.24 | 2,361.63 | 2,465.69 | 2,485.38 | **9,832.94** |
| 042 | 4,590.47 | 4,372.61 | 4,729.70 | 4,478.35 | **18,171.13** |
| 043 | 4,485.22 | 4,367.50 | 4,405.44 | 4,338.04 | **17,596.20** |
| 044 | 3,708.71 | 3,379.36 | 3,746.67 | 3,698.33 | **14,533.06** |
| 045 | 1,281.56 | 1,219.65 | 1,246.16 | 1,276.52 | **5,023.90** |
| 046 | 1,458.72 | 1,515.74 | 1,742.79 | 1,713.00 | **6,430.24** |
| 047 | 1,344.82 | 1,252.54 | 1,364.42 | 1,295.82 | **5,257.60** |
| 048 | 940.82 | 894.79 | 995.35 | 1,009.00 | **3,839.96** |
| 049 | 5,978.53 | 5,826.27 | 5,810.48 | 5,835.06 | **23,450.35** |
| **ME - MAINE - Total** | **36,604.45** | **35,536.35** | **37,186.81** | **36,745.60** | **146,073.21** |

**STATE: MI - MICHIGAN**

| REGISTRANT ZIP CODE 3 | 1ST QUARTER | 2ND QUARTER | 3RD QUARTER | 4TH QUARTER | TOTAL GRAMS |
|---|---|---|---|---|---|
| 480 | 35,347.23 | 32,535.28 | 33,136.67 | 34,604.13 | **135,623.31** |
| 481 | 34,050.22 | 31,299.99 | 32,402.08 | 32,407.27 | **130,159.55** |
| 482 | 23,286.52 | 21,660.14 | 21,181.26 | 20,876.52 | **87,004.45** |
| 483 | 22,805.72 | 21,623.58 | 22,415.88 | 22,576.52 | **89,421.71** |
| 484 | 5,565.64 | 5,435.45 | 5,457.99 | 5,584.31 | **22,043.38** |
| 485 | 2,236.70 | 2,136.10 | 2,322.43 | 2,185.62 | **8,880.86** |
| 486 | 6,108.92 | 5,677.59 | 5,828.79 | 5,752.35 | **23,367.64** |
| 487 | 2,914.84 | 2,906.48 | 2,762.01 | 2,867.48 | **11,450.80** |
| 488 | 7,331.78 | 6,990.40 | 7,415.69 | 7,564.81 | **29,302.68** |
| 489 | 3,046.12 | 2,935.93 | 2,954.80 | 3,001.96 | **11,938.80** |
| 490 | 11,745.92 | 11,038.01 | 12,096.97 | 12,073.09 | **46,953.99** |
| 491 | 1,601.62 | 1,646.12 | 1,476.97 | 1,583.05 | **6,307.77** |
| 492 | 5,683.85 | 5,271.26 | 5,441.04 | 5,412.98 | **21,809.13** |
| 493 | 2,276.22 | 2,080.79 | 2,262.98 | 2,333.46 | **8,953.46** |
| 494 | 4,911.52 | 4,816.40 | 4,968.66 | 5,054.24 | **19,750.82** |
| 495 | 5,304.62 | 4,696.54 | 5,334.26 | 5,056.48 | **20,391.91** |
| 496 | 6,108.51 | 6,052.18 | 6,185.73 | 6,121.33 | **24,467.75** |
| 497 | 4,557.65 | 4,920.75 | 5,031.10 | 4,952.83 | **19,462.32** |
| 498 | 2,824.91 | 2,605.42 | 2,766.13 | 2,609.80 | **10,806.27** |
| 499 | 868.13 | 857.16 | 869.67 | 846.70 | **3,441.66** |
| **MI - MICHIGAN - Total** | **188,576.62** | **177,185.58** | **182,311.12** | **183,464.92** | **731,538.25** |

**STATE: MN - MINNESOTA**

| REGISTRANT ZIP CODE 3 | 1ST QUARTER | 2ND QUARTER | 3RD QUARTER | 4TH QUARTER | TOTAL GRAMS |
|---|---|---|---|---|---|
| 550 | 11,450.84 | 11,330.72 | 11,756.03 | 11,690.56 | **46,228.16** |
| 551 | 16,553.82 | 15,678.90 | 15,847.55 | 16,127.58 | **64,207.85** |
| 553 | 14,301.42 | 13,735.23 | 14,245.53 | 14,327.88 | **56,610.05** |
| 554 | 24,519.14 | 22,789.37 | 24,119.67 | 23,931.83 | **95,360.01** |
| 556 | 172.35 | 172.58 | 171.12 | 173.34 | **689.39** |
| 557 | 3,412.90 | 3,288.83 | 3,546.30 | 3,502.90 | **13,750.93** |
| 558 | 1,623.13 | 1,503.03 | 1,722.10 | 1,509.95 | **6,358.21** |
| 559 | 3,919.17 | 3,381.31 | 4,331.99 | 3,682.52 | **15,314.98** |
| 560 | 3,080.68 | 2,883.38 | 3,132.26 | 3,207.56 | **12,303.87** |
| 561 | 757.03 | 605.60 | 544.49 | 616.32 | **2,523.43** |
| 562 | 1,391.80 | 1,326.18 | 1,326.07 | 1,310.89 | **5,354.92** |
| 563 | 4,883.79 | 4,416.97 | 4,840.87 | 4,798.32 | **18,939.94** |
| 564 | 2,118.86 | 2,064.59 | 2,357.13 | 2,238.57 | **8,779.15** |
| 565 | 1,629.03 | 1,625.31 | 1,668.95 | 1,687.18 | **6,610.47** |
| 566 | 966.05 | 954.16 | 983.83 | 968.84 | **3,872.88** |
| 567 | 1,128.19 | 987.35 | 946.34 | 965.99 | **4,027.87** |
| **MN - MINNESOTA - Total** | **91,908.18** | **86,743.51** | **91,540.20** | **90,740.23** | **360,932.12** |

**STATE: MO - MISSOURI**

| REGISTRANT ZIP CODE 3 | 1ST QUARTER | 2ND QUARTER | 3RD QUARTER | 4TH QUARTER | TOTAL GRAMS |
|---|---|---|---|---|---|
| 630 | 19,458.85 | 18,163.80 | 18,750.83 | 18,462.76 | **74,836.24** |
| 631 | 32,999.90 | 31,883.41 | 32,444.55 | 31,591.28 | **128,919.15** |
| 633 | 10,870.34 | 10,270.91 | 10,708.76 | 10,507.55 | **42,357.55** |
| 634 | 1,146.46 | 1,009.69 | 1,203.68 | 1,044.77 | **4,404.60** |
| 635 | 1,366.60 | 1,375.70 | 1,486.49 | 1,374.46 | **5,603.26** |

| | | | | | |
|---|---|---|---|---|---|
| 636 | 5,704.71 | 5,511.87 | 5,775.09 | 5,804.34 | **22,796.00** |
| 637 | 2,701.72 | 2,527.60 | 2,680.34 | 2,635.78 | **10,545.45** |
| 638 | 3,274.65 | 3,053.88 | 3,175.80 | 3,244.85 | **12,749.18** |
| 639 | 1,409.92 | 1,413.93 | 1,411.24 | 1,542.53 | **5,777.63** |
| 640 | 17,495.32 | 16,798.59 | 17,444.87 | 17,330.08 | **69,068.86** |
| 641 | 13,314.79 | 12,057.66 | 12,989.53 | 13,058.23 | **51,420.21** |
| 644 | 1,302.97 | 1,309.61 | 1,288.41 | 1,331.00 | **5,231.99** |
| 645 | 3,696.42 | 3,552.73 | 3,692.86 | 3,648.87 | **14,590.88** |
| 646 | 1,555.68 | 1,592.70 | 1,789.22 | 1,612.83 | **6,550.43** |
| 647 | 3,791.26 | 3,588.83 | 3,701.23 | 3,604.67 | **14,685.99** |
| 648 | 4,940.43 | 4,663.84 | 4,721.08 | 4,567.28 | **18,892.64** |
| 650 | 2,740.63 | 2,570.94 | 2,604.09 | 2,576.63 | **10,492.28** |
| 651 | 1,305.99 | 1,263.51 | 1,363.56 | 1,358.25 | **5,291.31** |
| 652 | 8,611.50 | 8,797.83 | 8,868.95 | 8,873.77 | **35,152.05** |
| 653 | 2,588.96 | 2,517.92 | 2,608.56 | 2,639.17 | **10,354.61** |
| 654 | 3,728.58 | 3,747.20 | 3,926.18 | 3,735.52 | **15,137.48** |
| 655 | 5,403.09 | 5,240.60 | 5,280.40 | 5,109.89 | **21,033.98** |
| 656 | 7,449.65 | 7,344.72 | 7,610.68 | 7,390.94 | **29,795.98** |
| 657 | 10,450.78 | 10,412.60 | 10,491.39 | 10,243.22 | **41,597.98** |
| 658 | 10,154.88 | 9,496.88 | 9,680.20 | 9,652.43 | **38,984.40** |
| **MO - MISSOURI - Total** | **177,464.07** | **170,166.97** | **175,697.99** | **172,941.10** | **696,270.13** |

**STATE: MS - MISSISSIPPI**

| REGISTRANT ZIP CODE 3 | 1ST QUARTER | 2ND QUARTER | 3RD QUARTER | 4TH QUARTER | TOTAL GRAMS |
|---|---|---|---|---|---|
| 386 | 11,418.82 | 10,959.42 | 11,277.01 | 11,543.64 | **45,198.89** |
| 387 | 1,403.66 | 1,343.52 | 1,391.34 | 1,339.52 | **5,478.05** |
| 388 | 7,471.50 | 7,297.71 | 7,467.88 | 7,459.66 | **29,696.75** |
| 389 | 1,473.28 | 1,434.84 | 1,367.53 | 1,380.27 | **5,655.91** |
| 390 | 3,999.80 | 3,839.15 | 4,000.85 | 3,943.50 | **15,783.30** |
| 391 | 3,983.93 | 3,688.13 | 3,836.00 | 3,923.52 | **15,431.58** |
| 392 | 4,652.57 | 4,232.54 | 4,579.84 | 4,826.82 | **18,291.77** |
| 393 | 4,573.15 | 4,415.05 | 4,633.94 | 4,759.25 | **18,381.39** |
| 394 | 17,046.94 | 16,703.02 | 17,782.57 | 17,799.25 | **69,331.77** |
| 395 | 15,317.90 | 14,032.19 | 14,780.28 | 14,902.93 | **59,033.30** |
| 396 | 3,436.20 | 3,511.74 | 3,544.10 | 3,627.82 | **14,119.86** |
| 397 | 2,291.97 | 2,174.91 | 2,346.14 | 2,252.52 | **9,065.55** |
| **MS - MISSISSIPPI - Total** | **77,069.73** | **73,632.22** | **77,007.47** | **77,758.70** | **305,468.13** |

**STATE: MT - MONTANA**

| REGISTRANT ZIP CODE 3 | 1ST QUARTER | 2ND QUARTER | 3RD QUARTER | 4TH QUARTER | TOTAL GRAMS |
|---|---|---|---|---|---|
| 590 | 1,778.25 | 1,777.51 | 1,790.45 | 1,752.69 | **7,098.90** |
| 591 | 6,160.26 | 5,869.73 | 6,256.53 | 6,169.63 | **24,456.14** |
| 592 | 858.70 | 835.71 | 780.68 | 927.99 | **3,403.07** |
| 593 | 950.92 | 923.21 | 932.81 | 920.30 | **3,727.25** |
| 594 | 2,760.53 | 2,799.98 | 2,883.08 | 2,779.87 | **11,223.46** |
| 595 | 415.12 | 308.69 | 409.60 | 424.02 | **1,557.43** |
| 596 | 1,974.33 | 1,957.78 | 1,958.35 | 1,954.59 | **7,845.05** |
| 597 | 3,099.79 | 3,165.36 | 3,249.81 | 3,219.69 | **12,734.64** |
| 598 | 4,596.29 | 4,265.69 | 4,490.13 | 4,511.93 | **17,864.04** |
| 599 | 4,032.96 | 3,770.96 | 4,160.49 | 4,088.18 | **16,052.58** |
| **MT - MONTANA - Total** | **26,627.13** | **25,674.61** | **26,911.93** | **26,748.88** | **105,962.55** |

**STATE: NC - NORTH CAROLINA**

| REGISTRANT ZIP CODE 3 | 1ST QUARTER | 2ND QUARTER | 3RD QUARTER | 4TH QUARTER | TOTAL GRAMS |
|---|---|---|---|---|---|
| 270 | 12,865.44 | 12,592.67 | 12,610.92 | 12,734.70 | **50,803.72** |
| 271 | 9,428.68 | 8,684.24 | 9,235.59 | 9,515.49 | **36,864.00** |
| 272 | 24,939.33 | 24,520.42 | 25,627.06 | 25,655.56 | **100,742.37** |
| 273 | 18,664.78 | 18,500.02 | 19,005.13 | 18,739.38 | **74,909.32** |
| 274 | 10,697.28 | 10,354.00 | 11,175.70 | 11,746.47 | **43,973.45** |
| 275 | 38,945.68 | 38,096.89 | 39,170.20 | 38,705.44 | **154,918.21** |
| 276 | 11,157.22 | 9,933.61 | 10,585.30 | 10,800.14 | **42,476.26** |
| 277 | 11,396.62 | 10,156.21 | 10,429.96 | 11,125.51 | **43,108.30** |
| 278 | 21,737.89 | 20,910.48 | 22,332.80 | 21,965.34 | **86,946.51** |

| | | | | | |
|---|---|---|---|---|---|
| 279 | 9,294.14 | 9,112.47 | 9,454.12 | 8,938.71 | **36,799.44** |
| 280 | 29,608.61 | 28,054.96 | 29,458.81 | 29,088.66 | **116,211.04** |
| 281 | 22,739.09 | 22,308.26 | 22,956.83 | 23,626.29 | **91,630.47** |
| 282 | 18,000.07 | 17,206.24 | 17,412.27 | 17,644.74 | **70,263.32** |
| 283 | 59,973.59 | 56,596.43 | 59,222.97 | 57,865.43 | **233,658.43** |
| 284 | 28,252.50 | 27,202.63 | 27,556.79 | 27,343.92 | **110,355.83** |
| 285 | 25,723.04 | 25,023.86 | 26,156.18 | 25,220.40 | **102,123.48** |
| 286 | 40,684.40 | 39,973.80 | 40,200.99 | 39,817.01 | **160,676.20** |
| 287 | 19,762.13 | 19,766.73 | 20,334.40 | 20,478.30 | **80,341.56** |
| 288 | 7,419.43 | 6,812.69 | 7,094.15 | 7,156.83 | **28,483.10** |
| 289 | 3,243.69 | 3,294.32 | 3,164.61 | 2,877.34 | **12,579.96** |
| **NC - NORTH CAROLINA - Total** | **424,533.59** | **409,100.93** | **423,184.77** | **421,045.68** | **1,677,864.97** |

**STATE: ND - NORTH DAKOTA**

| REGISTRANT ZIP CODE 3 | 1ST QUARTER | 2ND QUARTER | 3RD QUARTER | 4TH QUARTER | TOTAL GRAMS |
|---|---|---|---|---|---|
| 580 | 470.58 | 451.61 | 459.99 | 490.30 | **1,872.49** |
| 581 | 1,705.39 | 1,593.43 | 1,735.97 | 1,825.86 | **6,860.64** |
| 582 | 1,490.01 | 1,440.48 | 1,510.98 | 1,597.21 | **6,038.69** |
| 583 | 807.79 | 787.89 | 827.25 | 779.46 | **3,202.39** |
| 584 | 460.33 | 489.75 | 492.96 | 499.48 | **1,942.51** |
| 585 | 2,633.73 | 2,464.35 | 2,389.60 | 2,493.56 | **9,981.24** |
| 586 | 748.12 | 706.74 | 706.92 | 705.74 | **2,867.53** |
| 587 | 952.67 | 998.83 | 998.18 | 1,159.76 | **4,109.43** |
| 588 | 790.14 | 773.53 | 708.20 | 764.23 | **3,036.10** |
| **ND - NORTH DAKOTA - Total** | **10,058.75** | **9,706.62** | **9,830.06** | **10,315.60** | **39,911.02** |

**STATE: NE - NEBRASKA**

| REGISTRANT ZIP CODE 3 | 1ST QUARTER | 2ND QUARTER | 3RD QUARTER | 4TH QUARTER | TOTAL GRAMS |
|---|---|---|---|---|---|
| 680 | 3,096.13 | 2,877.24 | 2,842.11 | 2,898.42 | **11,713.90** |
| 681 | 12,026.67 | 11,123.26 | 11,763.51 | 11,668.12 | **46,581.56** |
| 683 | 928.85 | 833.12 | 873.49 | 867.91 | **3,503.37** |
| 684 | 855.88 | 771.49 | 733.85 | 721.31 | **3,082.53** |
| 685 | 3,318.54 | 3,085.10 | 3,381.11 | 3,207.01 | **12,991.76** |
| 686 | 383.90 | 402.56 | 394.09 | 430.83 | **1,611.38** |
| 687 | 988.93 | 942.88 | 1,001.01 | 980.52 | **3,913.34** |
| 688 | 2,352.93 | 2,194.62 | 2,346.93 | 2,399.73 | **9,294.21** |
| 689 | 996.63 | 1,012.77 | 894.23 | 921.40 | **3,825.03** |
| 690 | 489.47 | 439.91 | 482.02 | 492.59 | **1,903.99** |
| 691 | 1,367.60 | 1,272.83 | 1,241.97 | 1,316.05 | **5,198.45** |
| 692 | 80.24 | 70.51 | 75.22 | 71.76 | **297.73** |
| 693 | 1,345.62 | 1,265.70 | 1,374.47 | 1,324.32 | **5,310.10** |
| **NE - NEBRASKA - Total** | **28,231.39** | **26,291.98** | **27,404.00** | **27,299.99** | **109,227.36** |

**STATE: NH - NEW HAMPSHIRE**

| REGISTRANT ZIP CODE 3 | 1ST QUARTER | 2ND QUARTER | 3RD QUARTER | 4TH QUARTER | TOTAL GRAMS |
|---|---|---|---|---|---|
| 030 | 10,194.81 | 9,789.33 | 10,092.96 | 9,965.44 | **40,042.53** |
| 031 | 5,995.37 | 5,273.32 | 5,891.24 | 5,962.20 | **23,122.13** |
| 032 | 4,582.74 | 4,553.16 | 4,865.94 | 4,926.22 | **18,928.06** |
| 033 | 3,355.28 | 3,086.92 | 3,120.66 | 3,091.92 | **12,654.78** |
| 034 | 1,877.06 | 1,992.13 | 1,973.07 | 2,016.90 | **7,859.16** |
| 035 | 1,948.70 | 1,750.81 | 1,825.75 | 1,829.35 | **7,354.61** |
| 036 | 151.51 | 151.51 | 175.27 | 179.75 | **658.03** |
| 037 | 2,655.50 | 2,475.58 | 2,699.40 | 2,744.33 | **10,574.80** |
| 038 | 11,265.64 | 10,775.43 | 11,252.29 | 11,227.52 | **44,520.88** |
| **NH - NEW HAMPSHIRE - Total** | **42,026.61** | **39,848.19** | **41,896.56** | **41,943.63** | **165,714.99** |

**STATE: NJ - NEW JERSEY**

| REGISTRANT ZIP CODE 3 | 1ST QUARTER | 2ND QUARTER | 3RD QUARTER | 4TH QUARTER | TOTAL GRAMS |
|---|---|---|---|---|---|
| 070 | 48,373.78 | 44,802.10 | 46,647.59 | 46,407.58 | **186,231.05** |
| 071 | 11,811.36 | 10,841.14 | 11,580.30 | 11,406.52 | **45,639.33** |
| 072 | 5,091.35 | 4,464.91 | 4,945.69 | 5,116.23 | **19,618.17** |
| 073 | 7,074.11 | 7,325.43 | 7,358.44 | 7,478.30 | **29,236.28** |
| 074 | 14,128.96 | 13,250.81 | 14,149.97 | 13,854.55 | **55,384.29** |
| 075 | 5,854.57 | 5,235.15 | 5,673.25 | 5,451.68 | **22,214.65** |

| | | | | | |
|---|---|---|---|---|---|
| 076 | 18,515.42 | 16,325.98 | 17,456.93 | 17,140.16 | **69,438.49** |
| 077 | 22,276.14 | 21,545.84 | 22,509.47 | 22,289.75 | **88,621.20** |
| 078 | 10,124.22 | 9,439.45 | 9,625.12 | 9,922.17 | **39,110.96** |
| 079 | 4,888.36 | 4,106.26 | 4,431.63 | 4,601.89 | **18,028.14** |
| 080 | 75,615.55 | 73,017.44 | 74,330.31 | 74,993.53 | **297,956.83** |
| 081 | 12,006.20 | 12,137.08 | 12,198.23 | 12,164.72 | **48,506.22** |
| 082 | 14,830.76 | 14,881.83 | 15,481.26 | 15,176.40 | **60,370.25** |
| 083 | 10,693.64 | 10,438.63 | 10,527.08 | 10,615.44 | **42,274.79** |
| 084 | 3,039.67 | 2,982.35 | 3,165.62 | 3,110.37 | **12,298.01** |
| 085 | 8,934.73 | 8,454.58 | 8,544.11 | 8,608.09 | **34,541.51** |
| 086 | 11,198.81 | 11,271.22 | 11,523.03 | 11,141.83 | **45,134.88** |
| 087 | 18,499.48 | 17,876.14 | 18,963.35 | 18,398.93 | **73,737.90** |
| 088 | 31,020.43 | 28,720.08 | 29,340.88 | 28,916.89 | **117,998.29** |
| 089 | 2,712.64 | 2,835.53 | 2,781.62 | 2,872.97 | **11,202.76** |
| **NJ - NEW JERSEY - Total** | **336,690.17** | **319,951.95** | **331,233.89** | **329,668.00** | **1,317,544.01** |

**STATE: NM - NEW MEXICO**

| REGISTRANT ZIP CODE 3 | 1ST QUARTER | 2ND QUARTER | 3RD QUARTER | 4TH QUARTER | TOTAL GRAMS |
|---|---|---|---|---|---|
| 870 | 5,654.51 | 5,499.61 | 5,706.64 | 5,713.42 | **22,574.18** |
| 871 | 28,923.71 | 26,944.40 | 27,695.76 | 27,030.47 | **110,594.34** |
| 873 | 939.89 | 849.75 | 742.84 | 790.76 | **3,323.24** |
| 874 | 1,963.00 | 1,868.20 | 1,968.57 | 1,926.57 | **7,726.34** |
| 875 | 11,439.81 | 11,044.39 | 10,875.58 | 10,638.92 | **43,998.70** |
| 877 | 1,486.13 | 1,616.21 | 1,471.98 | 1,481.78 | **6,056.10** |
| 878 | 546.42 | 526.02 | 564.96 | 562.58 | **2,199.98** |
| 879 | 325.52 | 315.79 | 280.38 | 322.11 | **1,243.80** |
| 880 | 3,967.61 | 3,856.31 | 4,051.04 | 3,837.30 | **15,712.26** |
| 881 | 723.25 | 678.65 | 700.70 | 805.74 | **2,908.35** |
| 882 | 5,798.32 | 5,422.37 | 5,360.14 | 5,207.30 | **21,788.12** |
| 883 | 1,940.79 | 1,747.56 | 1,907.25 | 1,823.94 | **7,419.54** |
| 884 | 192.30 | 181.77 | 198.80 | 214.26 | **787.13** |
| **NM - NEW MEXICO - Total** | **63,901.26** | **60,551.02** | **61,524.64** | **60,355.14** | **246,332.07** |

**STATE: NV - NEVADA**

| REGISTRANT ZIP CODE 3 | 1ST QUARTER | 2ND QUARTER | 3RD QUARTER | 4TH QUARTER | TOTAL GRAMS |
|---|---|---|---|---|---|
| 890 | 32,224.37 | 31,423.66 | 32,014.88 | 32,293.20 | **127,956.11** |
| 891 | 74,883.10 | 73,847.50 | 73,878.54 | 73,236.82 | **295,845.96** |
| 893 | 334.89 | 310.46 | 285.54 | 329.02 | **1,259.90** |
| 894 | 9,510.62 | 9,196.96 | 9,503.52 | 9,363.00 | **37,574.11** |
| 895 | 9,637.78 | 9,195.54 | 9,832.03 | 9,506.51 | **38,171.86** |
| 897 | 3,236.61 | 3,060.74 | 3,171.84 | 2,993.54 | **12,462.74** |
| 898 | 1,053.31 | 999.59 | 1,044.69 | 1,098.98 | **4,196.57** |
| **NV - NEVADA - Total** | **130,880.68** | **128,034.45** | **129,731.04** | **128,821.07** | **517,467.24** |

**STATE: NY - NEW YORK**

| REGISTRANT ZIP CODE 3 | 1ST QUARTER | 2ND QUARTER | 3RD QUARTER | 4TH QUARTER | TOTAL GRAMS |
|---|---|---|---|---|---|
| 063 | 0.22 | 0 | 0 | 0.45 | **0.67** |
| 100 | 43,238.71 | 38,960.90 | 41,650.56 | 41,437.37 | **165,287.54** |
| 101 | 974.77 | 811.84 | 909.43 | 958.92 | **3,654.96** |
| 102 | 34.53 | 25.33 | 26.67 | 29.81 | **116.33** |
| 103 | 19,409.00 | 19,086.49 | 19,297.37 | 18,769.71 | **76,562.58** |
| 104 | 35,309.67 | 32,555.14 | 33,766.19 | 34,719.50 | **136,350.50** |
| 105 | 13,918.25 | 12,493.25 | 13,306.09 | 12,905.07 | **52,622.66** |
| 106 | 1,899.29 | 1,698.62 | 1,886.48 | 1,764.67 | **7,249.06** |
| 107 | 5,950.71 | 5,498.06 | 6,033.14 | 6,202.36 | **23,684.27** |
| 108 | 1,558.74 | 1,502.62 | 1,532.02 | 1,535.57 | **6,128.94** |
| 109 | 17,107.64 | 16,431.38 | 16,838.29 | 16,371.33 | **66,748.64** |
| 110 | 7,949.28 | 6,971.58 | 7,212.90 | 7,120.22 | **29,253.98** |
| 111 | 4,837.76 | 4,112.32 | 4,161.61 | 4,172.48 | **17,284.17** |
| 112 | 41,355.64 | 38,164.96 | 39,662.14 | 39,482.50 | **158,665.24** |
| 113 | 19,989.29 | 18,275.01 | 18,372.62 | 18,151.63 | **74,788.55** |
| 114 | 9,540.11 | 8,599.88 | 8,953.13 | 9,211.06 | **36,304.17** |
| 115 | 19,018.88 | 18,411.22 | 19,208.65 | 18,527.41 | **75,166.15** |

| | | | | | |
|---|---|---|---|---|---|
| 116 | 2,720.08 | 2,421.39 | 2,540.19 | 2,428.24 | **10,109.90** |
| 117 | 63,495.16 | 58,455.65 | 61,257.69 | 61,568.40 | **244,776.91** |
| 118 | 2,480.91 | 2,605.85 | 2,619.10 | 2,573.41 | **10,279.27** |
| 119 | 7,587.86 | 7,552.42 | 7,696.09 | 7,565.98 | **30,402.35** |
| 120 | 7,988.57 | 7,603.38 | 8,162.85 | 7,817.46 | **31,572.27** |
| 121 | 4,777.55 | 4,690.49 | 5,019.43 | 5,017.23 | **19,504.70** |
| 122 | 4,203.45 | 3,377.41 | 3,899.19 | 3,703.35 | **15,183.40** |
| 123 | 2,494.41 | 2,542.68 | 2,739.21 | 2,747.07 | **10,523.37** |
| 124 | 9,676.51 | 9,553.90 | 9,415.14 | 9,600.02 | **38,245.57** |
| 125 | 14,980.05 | 14,436.61 | 14,972.41 | 14,625.54 | **59,014.60** |
| 126 | 4,607.69 | 4,501.75 | 4,775.82 | 4,751.40 | **18,636.66** |
| 127 | 5,650.20 | 5,461.02 | 5,349.35 | 5,339.06 | **21,799.63** |
| 128 | 6,889.77 | 6,842.29 | 7,322.70 | 7,157.18 | **28,211.94** |
| 129 | 5,133.86 | 4,966.36 | 5,192.33 | 5,377.00 | **20,669.55** |
| 130 | 8,377.18 | 8,077.76 | 8,398.08 | 8,448.74 | **33,301.76** |
| 131 | 2,339.27 | 2,241.07 | 2,404.77 | 2,362.64 | **9,347.75** |
| 132 | 5,349.77 | 4,869.75 | 5,392.88 | 5,275.34 | **20,887.74** |
| 133 | 2,264.05 | 2,200.16 | 2,320.76 | 2,422.70 | **9,207.67** |
| 134 | 2,983.06 | 2,834.20 | 2,905.33 | 2,859.34 | **11,581.93** |
| 135 | 1,284.87 | 1,202.43 | 1,254.72 | 1,281.58 | **5,023.61** |
| 136 | 4,921.92 | 4,988.33 | 5,144.28 | 5,129.40 | **20,183.94** |
| 137 | 2,832.95 | 2,895.52 | 2,769.09 | 2,748.32 | **11,245.88** |
| 138 | 5,562.87 | 5,055.75 | 5,061.84 | 5,060.37 | **20,740.84** |
| 139 | 2,951.14 | 2,788.18 | 2,737.69 | 2,848.61 | **11,325.63** |
| 140 | 9,207.44 | 8,679.81 | 8,834.59 | 8,735.84 | **35,457.67** |
| 141 | 4,006.64 | 4,018.81 | 4,069.08 | 4,010.83 | **16,105.36** |
| 142 | 11,868.96 | 11,470.21 | 11,957.68 | 11,447.52 | **46,744.37** |
| 143 | 1,824.40 | 1,623.56 | 1,688.77 | 1,712.77 | **6,849.49** |
| 144 | 6,242.52 | 6,081.80 | 6,244.87 | 6,360.40 | **24,929.58** |
| 145 | 4,898.78 | 4,764.94 | 4,849.57 | 4,887.13 | **19,400.42** |
| 146 | 14,228.13 | 13,156.64 | 13,640.97 | 13,734.05 | **54,759.80** |
| 147 | 2,718.13 | 2,668.36 | 2,741.53 | 2,726.20 | **10,854.23** |
| 148 | 7,188.07 | 7,130.02 | 7,388.96 | 6,914.72 | **28,621.77** |
| 149 | 2,330.32 | 2,374.00 | 2,288.48 | 2,454.78 | **9,447.58** |
| **NY - NEW YORK - Total** | **488,159.02** | **457,731.11** | **475,872.75** | **473,052.67** | **1,894,815.55** |

**STATE: OH - OHIO**

| REGISTRANT ZIP CODE 3 | 1ST QUARTER | 2ND QUARTER | 3RD QUARTER | 4TH QUARTER | TOTAL GRAMS |
|---|---|---|---|---|---|
| 430 | 19,664.33 | 18,658.72 | 19,597.45 | 19,517.38 | **77,437.86** |
| 431 | 14,790.87 | 14,514.29 | 14,811.29 | 14,796.64 | **58,913.08** |
| 432 | 27,054.79 | 25,028.19 | 26,400.12 | 26,189.85 | **104,672.95** |
| 433 | 5,284.26 | 5,051.73 | 5,213.84 | 5,110.81 | **20,660.64** |
| 434 | 2,549.74 | 2,674.79 | 2,759.32 | 2,729.43 | **10,713.29** |
| 435 | 5,749.71 | 5,631.25 | 5,760.38 | 5,967.63 | **23,108.97** |
| 436 | 15,561.97 | 14,489.27 | 15,379.63 | 15,418.13 | **60,849.00** |
| 437 | 6,903.09 | 6,522.50 | 6,669.47 | 6,688.24 | **26,783.31** |
| 438 | 1,064.92 | 1,035.54 | 1,021.84 | 1,027.25 | **4,149.54** |
| 439 | 5,031.17 | 4,731.02 | 5,102.95 | 5,098.94 | **19,964.07** |
| 440 | 14,603.48 | 13,782.39 | 14,944.74 | 15,066.77 | **58,397.38** |
| 441 | 23,285.61 | 21,189.60 | 23,118.38 | 23,767.15 | **91,360.73** |
| 442 | 10,768.57 | 10,117.30 | 10,699.79 | 10,942.97 | **42,528.63** |
| 443 | 7,504.43 | 7,061.17 | 7,596.99 | 7,244.30 | **29,406.88** |
| 444 | 10,276.11 | 9,935.56 | 9,856.42 | 9,600.93 | **39,669.02** |
| 445 | 6,211.39 | 5,809.15 | 5,967.32 | 5,983.66 | **23,971.52** |
| 446 | 8,080.90 | 7,508.26 | 8,258.10 | 8,211.74 | **32,058.99** |
| 447 | 5,192.03 | 5,012.23 | 5,536.28 | 5,203.27 | **20,943.80** |
| 448 | 5,714.16 | 5,399.42 | 5,848.90 | 5,866.23 | **22,828.70** |
| 449 | 2,038.58 | 1,939.74 | 2,042.78 | 2,112.89 | **8,133.98** |
| 450 | 21,215.71 | 21,188.61 | 21,962.68 | 21,700.14 | **86,067.14** |
| 451 | 10,572.13 | 10,353.68 | 10,460.96 | 10,704.52 | **42,091.30** |
| 452 | 25,708.21 | 24,324.11 | 25,609.56 | 25,801.50 | **101,443.38** |

| | | | | | |
|---|---|---|---|---|---|
| 453 | 10,336.96 | 9,878.75 | 10,358.12 | 10,280.83 | **40,854.65** |
| 454 | 11,649.88 | 11,143.52 | 11,976.95 | 11,952.52 | **46,722.87** |
| 455 | 4,345.65 | 4,208.39 | 4,293.45 | 4,022.66 | **16,870.15** |
| 456 | 16,263.30 | 15,607.71 | 15,956.83 | 15,800.27 | **63,628.10** |
| 457 | 4,251.05 | 4,200.91 | 4,565.32 | 4,328.77 | **17,346.05** |
| 458 | 5,072.47 | 4,656.54 | 5,028.60 | 5,224.85 | **19,982.45** |
| **OH - OHIO - Total** | **306,745.45** | **291,654.33** | **306,798.41** | **306,360.25** | **1,211,558.44** |

**STATE: OK - OKLAHOMA**

| REGISTRANT ZIP CODE 3 | 1ST QUARTER | 2ND QUARTER | 3RD QUARTER | 4TH QUARTER | TOTAL GRAMS |
|---|---|---|---|---|---|
| 730 | 23,230.64 | 22,880.13 | 22,586.33 | 22,652.76 | **91,349.86** |
| 731 | 26,775.23 | 25,329.17 | 25,358.30 | 26,068.68 | **103,531.38** |
| 734 | 3,089.39 | 2,788.72 | 3,149.31 | 3,067.53 | **12,094.96** |
| 735 | 7,472.51 | 7,538.17 | 8,131.07 | 7,420.55 | **30,562.29** |
| 736 | 2,115.28 | 2,097.68 | 2,103.39 | 1,982.06 | **8,298.41** |
| 737 | 2,896.18 | 2,802.31 | 2,847.37 | 2,937.22 | **11,483.08** |
| 738 | 708.76 | 704.48 | 680.24 | 648.23 | **2,741.70** |
| 739 | 169.93 | 156.22 | 134.48 | 96.15 | **556.77** |
| 740 | 22,019.35 | 21,556.30 | 21,936.18 | 21,533.09 | **87,044.91** |
| 741 | 14,948.19 | 14,238.28 | 13,889.88 | 13,840.81 | **56,917.15** |
| 743 | 3,558.20 | 3,676.31 | 3,676.44 | 3,601.38 | **14,512.33** |
| 744 | 11,285.47 | 11,006.94 | 11,315.79 | 11,206.43 | **44,814.62** |
| 745 | 2,085.82 | 2,123.65 | 2,242.87 | 2,199.19 | **8,651.54** |
| 746 | 1,612.77 | 1,678.32 | 1,619.39 | 1,738.84 | **6,649.32** |
| 747 | 1,830.29 | 1,801.01 | 1,834.25 | 1,731.83 | **7,197.38** |
| 748 | 6,716.29 | 6,551.52 | 6,649.43 | 6,405.68 | **26,322.93** |
| 749 | 4,666.30 | 4,644.22 | 4,699.46 | 4,755.15 | **18,765.13** |
| **OK - OKLAHOMA - Total** | **135,180.60** | **131,573.42** | **132,854.17** | **131,885.58** | **531,493.77** |

**STATE: OR - OREGON**

| REGISTRANT ZIP CODE 3 | 1ST QUARTER | 2ND QUARTER | 3RD QUARTER | 4TH QUARTER | TOTAL GRAMS |
|---|---|---|---|---|---|
| 970 | 19,721.39 | 18,762.41 | 19,753.90 | 19,886.15 | **78,123.85** |
| 971 | 8,647.47 | 7,889.11 | 8,822.28 | 8,344.54 | **33,703.40** |
| 972 | 25,570.29 | 25,107.06 | 26,343.96 | 26,412.88 | **103,434.19** |
| 973 | 15,098.12 | 14,322.58 | 15,170.10 | 14,817.46 | **59,408.27** |
| 974 | 20,386.21 | 19,915.46 | 20,774.72 | 20,538.46 | **81,614.85** |
| 975 | 16,544.43 | 16,192.18 | 16,628.68 | 16,470.71 | **65,836.00** |
| 976 | 1,830.81 | 1,830.77 | 1,962.24 | 2,057.75 | **7,681.57** |
| 977 | 6,195.39 | 5,954.32 | 6,399.29 | 6,277.46 | **24,826.46** |
| 978 | 3,985.07 | 3,978.93 | 3,891.46 | 3,964.52 | **15,819.98** |
| 979 | 1,120.49 | 1,121.15 | 1,150.27 | 1,161.23 | **4,553.14** |
| **OR - OREGON - Total** | **119,099.66** | **115,073.98** | **120,896.89** | **119,931.17** | **475,001.70** |

**STATE: PA - PENNSYLVANIA**

| REGISTRANT ZIP CODE 3 | 1ST QUARTER | 2ND QUARTER | 3RD QUARTER | 4TH QUARTER | TOTAL GRAMS |
|---|---|---|---|---|---|
| 150 | 17,079.73 | 16,192.23 | 17,335.90 | 12,334.25 | **62,942.11** |
| 151 | 8,757.61 | 8,260.31 | 9,083.98 | 9,080.05 | **35,181.95** |
| 152 | 21,519.09 | 20,630.10 | 21,444.48 | 21,312.80 | **84,906.47** |
| 153 | 5,076.82 | 4,974.89 | 5,301.33 | 5,196.12 | **20,549.17** |
| 154 | 7,219.86 | 7,184.06 | 7,015.80 | 6,972.41 | **28,392.12** |
| 155 | 2,054.60 | 1,885.44 | 2,084.10 | 2,054.17 | **8,078.32** |
| 156 | 8,534.94 | 7,329.04 | 7,173.85 | 7,318.83 | **30,356.66** |
| 157 | 3,469.77 | 3,217.70 | 3,337.89 | 3,050.41 | **13,075.77** |
| 158 | 3,439.25 | 2,911.28 | 3,195.85 | 3,013.36 | **12,559.74** |
| 159 | 6,284.87 | 5,710.83 | 5,724.36 | 5,622.62 | **23,342.68** |
| 160 | 5,600.84 | 5,548.69 | 5,646.50 | 5,498.35 | **22,294.39** |
| 161 | 7,048.80 | 6,830.45 | 7,234.15 | 7,388.81 | **28,502.21** |
| 162 | 1,976.42 | 1,786.58 | 1,939.51 | 1,841.41 | **7,543.93** |
| 163 | 5,177.90 | 4,959.94 | 5,076.52 | 5,166.77 | **20,381.13** |
| 164 | 2,065.90 | 1,951.53 | 2,115.90 | 2,198.91 | **8,332.25** |
| 165 | 3,978.00 | 3,957.65 | 3,993.62 | 3,984.58 | **15,913.85** |
| 166 | 6,346.43 | 6,111.01 | 6,607.71 | 6,384.19 | **25,449.34** |
| 167 | 1,184.93 | 1,228.03 | 1,287.49 | 1,299.76 | **5,000.21** |

| | | | | | |
|---|---|---|---|---|---|
| 168 | 2,521.18 | 2,251.71 | 2,589.77 | 2,510.97 | **9,873.63** |
| 169 | 1,478.88 | 1,530.33 | 1,432.78 | 1,488.51 | **5,930.49** |
| 170 | 18,372.02 | 16,903.55 | 17,598.66 | 17,376.86 | **70,251.09** |
| 171 | 4,883.74 | 4,627.18 | 4,602.21 | 4,845.71 | **18,958.83** |
| 172 | 5,546.03 | 5,091.14 | 5,135.91 | 5,029.53 | **20,802.61** |
| 173 | 8,547.36 | 8,116.84 | 8,431.36 | 8,475.77 | **33,571.33** |
| 174 | 4,788.20 | 4,804.18 | 4,687.99 | 4,793.88 | **19,074.25** |
| 175 | 7,462.15 | 7,274.98 | 7,858.68 | 7,588.17 | **30,183.98** |
| 176 | 6,574.60 | 6,140.69 | 6,336.96 | 6,223.55 | **25,275.80** |
| 177 | 5,448.47 | 5,239.21 | 5,430.46 | 5,421.70 | **21,539.83** |
| 178 | 5,302.31 | 4,819.80 | 5,315.23 | 4,823.96 | **20,261.29** |
| 179 | 3,384.97 | 3,131.18 | 3,221.36 | 3,112.94 | **12,850.45** |
| 180 | 12,083.71 | 11,838.67 | 12,548.02 | 11,891.77 | **48,362.18** |
| 181 | 7,467.60 | 6,530.92 | 7,001.55 | 6,749.04 | **27,749.11** |
| 182 | 4,162.07 | 3,996.58 | 4,168.04 | 4,330.52 | **16,657.20** |
| 183 | 6,318.28 | 6,397.25 | 6,727.45 | 6,507.58 | **25,950.57** |
| 184 | 6,908.19 | 6,654.03 | 6,665.65 | 6,479.49 | **26,707.36** |
| 185 | 7,193.46 | 6,843.72 | 6,712.36 | 6,206.38 | **26,955.91** |
| 186 | 5,618.70 | 5,355.27 | 5,542.08 | 5,433.58 | **21,949.63** |
| 187 | 21,332.38 | 23,526.79 | 23,376.18 | 23,536.48 | **91,771.84** |
| 188 | 2,340.23 | 2,390.31 | 2,414.88 | 2,489.28 | **9,634.70** |
| 189 | 10,948.69 | 10,593.44 | 11,051.59 | 10,427.49 | **43,021.21** |
| 190 | 65,555.81 | 69,083.38 | 72,715.07 | 65,435.39 | **272,789.65** |
| 191 | 80,904.83 | 78,974.60 | 78,883.32 | 78,663.11 | **317,425.86** |
| 193 | 12,837.77 | 11,698.86 | 11,996.05 | 12,140.96 | **48,673.64** |
| 194 | 17,340.86 | 16,345.81 | 16,560.00 | 16,886.13 | **67,132.79** |
| 195 | 3,745.10 | 3,635.87 | 3,945.28 | 4,081.36 | **15,407.61** |
| 196 | 5,144.52 | 4,915.18 | 4,908.31 | 5,018.29 | **19,986.29** |
| **PA - PENNSYLVANIA - Total** | **461,027.84** | **449,381.25** | **463,456.14** | **447,686.20** | **1,821,551.43** |
| **STATE: PR - PUERTO RICO** | | | | | |
| REGISTRANT ZIP CODE 3 | 1ST QUARTER | 2ND QUARTER | 3RD QUARTER | 4TH QUARTER | **TOTAL GRAMS** |
| 006 | 3,482.45 | 2,856.92 | 3,661.93 | 3,385.32 | **13,386.63** |
| 007 | 3,481.42 | 2,612.85 | 3,524.05 | 3,308.53 | **12,926.86** |
| 009 | 4,483.22 | 3,657.72 | 4,127.98 | 4,048.19 | **16,317.11** |
| **PR - PUERTO RICO - Total** | **11,447.09** | **9,127.49** | **11,313.96** | **10,742.04** | **42,630.59** |
| **STATE: RI - RHODE ISLAND** | | | | | |
| REGISTRANT ZIP CODE 3 | 1ST QUARTER | 2ND QUARTER | 3RD QUARTER | 4TH QUARTER | **TOTAL GRAMS** |
| 028 | 17,435.39 | 16,125.73 | 16,902.38 | 15,758.38 | **66,221.88** |
| 029 | 10,962.95 | 10,425.04 | 11,409.86 | 10,210.58 | **43,008.43** |
| **RI - RHODE ISLAND - Total** | **28,398.35** | **26,550.77** | **28,312.24** | **25,968.96** | **109,230.32** |
| **STATE: SC - SOUTH CAROLINA** | | | | | |
| REGISTRANT ZIP CODE 3 | 1ST QUARTER | 2ND QUARTER | 3RD QUARTER | 4TH QUARTER | **TOTAL GRAMS** |
| 290 | 10,109.69 | 10,147.65 | 9,959.04 | 10,092.86 | **40,309.24** |
| 291 | 11,484.50 | 11,209.12 | 11,629.44 | 11,790.88 | **46,113.94** |
| 292 | 8,008.22 | 7,614.72 | 7,994.11 | 8,230.10 | **31,847.15** |
| 293 | 14,368.81 | 13,840.94 | 14,283.90 | 14,912.65 | **57,406.29** |
| 294 | 27,272.98 | 25,764.47 | 26,929.92 | 26,865.41 | **106,832.77** |
| 295 | 38,390.68 | 35,166.04 | 36,556.17 | 36,580.65 | **146,693.55** |
| 296 | 28,123.53 | 27,395.36 | 28,188.32 | 28,196.59 | **111,903.79** |
| 297 | 7,888.39 | 7,363.00 | 7,884.53 | 7,922.92 | **31,058.84** |
| 298 | 7,394.39 | 7,267.53 | 7,088.84 | 7,070.99 | **28,821.75** |
| 299 | 6,469.09 | 6,336.53 | 6,782.54 | 6,952.30 | **26,540.47** |
| **SC - SOUTH CAROLINA - Total** | **159,510.27** | **152,105.35** | **157,296.80** | **158,615.35** | **627,527.78** |
| **STATE: SD - SOUTH DAKOTA** | | | | | |
| REGISTRANT ZIP CODE 3 | 1ST QUARTER | 2ND QUARTER | 3RD QUARTER | 4TH QUARTER | **TOTAL GRAMS** |
| 570 | 2,040.13 | 1,857.39 | 1,936.02 | 1,910.54 | **7,744.08** |
| 571 | 2,949.64 | 2,606.51 | 2,800.68 | 2,728.11 | **11,084.93** |
| 572 | 540.33 | 504.53 | 530.14 | 528.91 | **2,103.91** |
| 573 | 581.74 | 609.99 | 590.84 | 585.22 | **2,367.78** |
| 574 | 682.87 | 553.83 | 629.89 | 622.40 | **2,488.99** |

| | | | | | |
|---|---|---|---|---|---|
| 575 | 518.07 | 491.06 | 486.29 | 528.17 | **2,023.59** |
| 576 | 100.77 | 119.46 | 122.46 | 132.91 | **475.59** |
| 577 | 3,040.24 | 2,751.23 | 3,097.84 | 2,987.72 | **11,877.04** |
| **SD - SOUTH DAKOTA - Total** | **10,453.79** | **9,493.98** | **10,194.15** | **10,023.99** | **40,165.91** |

**STATE: TN - TENNESSEE**

| REGISTRANT ZIP CODE 3 | 1ST QUARTER | 2ND QUARTER | 3RD QUARTER | 4TH QUARTER | TOTAL GRAMS |
|---|---|---|---|---|---|
| 370 | 40,184.40 | 38,668.93 | 39,887.46 | 38,517.40 | **157,258.19** |
| 371 | 33,871.08 | 33,028.30 | 34,210.61 | 34,239.82 | **135,349.81** |
| 372 | 23,157.32 | 21,898.03 | 23,070.77 | 22,517.26 | **90,643.37** |
| 373 | 31,515.37 | 30,319.40 | 30,566.31 | 30,709.07 | **123,110.15** |
| 374 | 11,427.88 | 11,043.95 | 11,547.10 | 11,398.71 | **45,417.64** |
| 376 | 27,915.05 | 26,738.82 | 26,975.80 | 26,334.06 | **107,963.73** |
| 377 | 22,794.39 | 22,373.47 | 22,252.61 | 22,226.19 | **89,646.65** |
| 378 | 32,324.28 | 31,671.80 | 31,755.99 | 32,214.71 | **127,966.77** |
| 379 | 18,127.29 | 17,407.27 | 17,405.83 | 17,642.49 | **70,582.88** |
| 380 | 11,590.94 | 11,395.36 | 11,702.03 | 11,354.61 | **46,042.94** |
| 381 | 20,828.93 | 27,232.40 | 27,113.70 | 21,438.08 | **96,613.10** |
| 382 | 4,825.36 | 4,683.61 | 4,629.19 | 4,439.45 | **18,577.61** |
| 383 | 10,727.73 | 10,501.99 | 10,326.27 | 10,533.50 | **42,089.49** |
| 384 | 10,225.95 | 9,455.67 | 9,732.95 | 9,108.41 | **38,522.97** |
| 385 | 18,535.34 | 17,519.45 | 17,989.28 | 17,520.55 | **71,564.63** |
| **TN - TENNESSEE - Total** | **318,051.32** | **313,938.44** | **319,165.89** | **310,194.28** | **1,261,349.93** |

**STATE: TX - TEXAS**

| REGISTRANT ZIP CODE 3 | 1ST QUARTER | 2ND QUARTER | 3RD QUARTER | 4TH QUARTER | TOTAL GRAMS |
|---|---|---|---|---|---|
| 750 | 24,491.96 | 23,740.97 | 24,256.10 | 24,206.47 | **96,695.51** |
| 751 | 6,205.09 | 6,110.05 | 6,812.67 | 6,480.16 | **25,607.96** |
| 752 | 6,909.64 | 6,609.78 | 7,113.08 | 7,822.96 | **28,455.46** |
| 753 | 199.67 | 159.27 | 246.51 | 293.95 | **899.41** |
| 754 | 3,658.39 | 3,604.98 | 3,860.46 | 3,939.16 | **15,062.99** |
| 755 | 976.00 | 940.26 | 1,049.16 | 1,005.90 | **3,971.32** |
| 756 | 2,117.68 | 2,282.08 | 2,333.30 | 2,343.43 | **9,076.48** |
| 757 | 3,900.13 | 3,636.12 | 3,804.98 | 3,696.34 | **15,037.58** |
| 758 | 553.96 | 557.95 | 558.93 | 523.40 | **2,194.22** |
| 759 | 1,290.18 | 1,338.52 | 1,349.62 | 1,346.75 | **5,325.06** |
| 760 | 15,514.41 | 15,531.42 | 16,010.75 | 16,142.34 | **63,198.92** |
| 761 | 10,857.87 | 10,662.70 | 10,798.65 | 10,722.46 | **43,041.67** |
| 762 | 5,847.89 | 5,401.08 | 5,502.12 | 5,759.58 | **22,510.68** |
| 763 | 1,628.68 | 1,603.74 | 1,632.62 | 1,609.46 | **6,474.50** |
| 764 | 1,031.14 | 1,059.39 | 1,035.30 | 1,086.81 | **4,212.64** |
| 765 | 3,758.62 | 3,359.60 | 3,871.38 | 3,502.09 | **14,491.70** |
| 766 | 860.27 | 828.96 | 911.13 | 935.38 | **3,535.74** |
| 767 | 1,700.88 | 1,557.10 | 1,704.90 | 1,513.87 | **6,476.76** |
| 768 | 621.15 | 555.70 | 560.06 | 563.85 | **2,300.77** |
| 769 | 572.68 | 496.86 | 548.96 | 523.26 | **2,141.75** |
| 770 | 42,972.62 | 43,482.47 | 40,841.77 | 30,620.45 | **157,917.31** |
| 773 | 10,138.91 | 11,491.90 | 10,133.26 | 10,710.06 | **42,474.13** |
| 774 | 13,404.64 | 14,925.20 | 15,786.18 | 14,836.94 | **58,952.95** |
| 775 | 14,529.19 | 16,515.60 | 18,116.05 | 15,903.71 | **65,064.55** |
| 776 | 2,287.20 | 2,009.99 | 2,269.35 | 2,201.48 | **8,768.02** |
| 777 | 1,083.35 | 1,070.34 | 1,178.96 | 982.82 | **4,315.47** |
| 778 | 1,071.88 | 1,089.16 | 1,105.63 | 1,028.47 | **4,295.13** |
| 779 | 2,510.76 | 2,390.36 | 2,341.69 | 2,266.39 | **9,509.20** |
| 780 | 2,406.09 | 2,413.55 | 2,246.80 | 2,344.27 | **9,410.71** |
| 781 | 3,156.64 | 2,842.81 | 3,006.26 | 2,931.07 | **11,936.78** |
| 782 | 14,306.67 | 13,303.84 | 13,760.56 | 13,340.86 | **54,711.94** |
| 783 | 975.85 | 915.38 | 950.20 | 1,019.24 | **3,860.68** |
| 784 | 1,522.36 | 1,396.22 | 1,487.34 | 1,443.96 | **5,849.88** |
| 785 | 2,383.88 | 2,129.68 | 2,157.23 | 2,169.36 | **8,840.16** |
| 786 | 11,301.49 | 10,647.17 | 11,361.76 | 11,514.32 | **44,824.73** |
| 787 | 8,899.88 | 7,973.45 | 8,116.84 | 8,632.19 | **33,622.36** |

| | | | | | |
|---|---|---|---|---|---|
| 788 | 342.28 | 323.02 | 370.60 | 332.67 | **1,368.56** |
| 789 | 651.96 | 645.63 | 578.01 | 588.27 | **2,463.86** |
| 790 | 956.66 | 933.82 | 869.80 | 868.39 | **3,628.68** |
| 791 | 2,266.97 | 2,156.28 | 2,292.75 | 2,080.72 | **8,796.72** |
| 792 | 158.01 | 173.16 | 181.41 | 161.19 | **673.76** |
| 793 | 409.92 | 344.33 | 452.71 | 424.04 | **1,630.99** |
| 794 | 2,274.16 | 2,294.41 | 2,259.46 | 2,386.36 | **9,214.38** |
| 795 | 454.38 | 454.16 | 631.29 | 412.71 | **1,952.54** |
| 796 | 939.16 | 906.65 | 922.56 | 826.95 | **3,595.32** |
| 797 | 1,297.91 | 1,214.94 | 1,325.73 | 1,466.43 | **5,305.02** |
| 798 | 106.95 | 101.12 | 80.52 | 85.18 | **373.77** |
| 799 | 3,269.99 | 3,014.63 | 3,730.24 | 2,851.26 | **12,866.12** |
| **TX - TEXAS - Total** | **238,776.06** | **237,195.76** | **242,515.64** | **228,447.39** | **946,934.86** |

**STATE: UT - UTAH**

| REGISTRANT ZIP CODE 3 | 1ST QUARTER | 2ND QUARTER | 3RD QUARTER | 4TH QUARTER | TOTAL GRAMS |
|---|---|---|---|---|---|
| 840 | 51,280.18 | 51,177.66 | 51,817.87 | 50,781.44 | **205,057.15** |
| 841 | 30,686.77 | 29,735.72 | 29,280.71 | 28,745.02 | **118,448.22** |
| 843 | 4,381.44 | 4,375.30 | 4,502.21 | 4,476.67 | **17,735.62** |
| 844 | 11,274.24 | 11,080.41 | 11,419.52 | 10,981.26 | **44,755.43** |
| 845 | 4,037.66 | 4,271.28 | 4,448.64 | 4,197.37 | **16,954.95** |
| 846 | 10,632.77 | 10,303.15 | 10,466.05 | 10,262.41 | **41,664.37** |
| 847 | 15,497.20 | 15,253.09 | 15,101.89 | 15,418.70 | **61,270.89** |
| **UT - UTAH - Total** | **127,790.26** | **126,196.61** | **127,036.89** | **124,862.87** | **505,886.63** |

**STATE: VA - VIRGINIA**

| REGISTRANT ZIP CODE 3 | 1ST QUARTER | 2ND QUARTER | 3RD QUARTER | 4TH QUARTER | TOTAL GRAMS |
|---|---|---|---|---|---|
| 201 | 14,866.08 | 14,068.69 | 14,108.96 | 14,681.35 | **57,725.08** |
| 220 | 8,523.60 | 7,727.89 | 8,578.53 | 7,286.53 | **32,116.55** |
| 221 | 7,231.60 | 6,607.66 | 7,039.75 | 7,048.39 | **27,927.41** |
| 222 | 2,395.62 | 2,343.10 | 2,484.27 | 2,069.89 | **9,292.88** |
| 223 | 4,283.10 | 3,874.49 | 4,215.15 | 4,209.52 | **16,582.25** |
| 224 | 7,045.75 | 6,948.08 | 6,989.71 | 7,064.88 | **28,048.43** |
| 225 | 2,981.01 | 2,987.19 | 3,068.36 | 3,235.41 | **12,271.97** |
| 226 | 6,117.09 | 5,807.61 | 6,198.47 | 5,771.88 | **23,895.05** |
| 227 | 1,664.62 | 1,581.97 | 1,639.04 | 1,635.76 | **6,521.39** |
| 228 | 4,291.59 | 4,283.71 | 4,349.18 | 4,162.26 | **17,086.73** |
| 229 | 6,529.17 | 6,392.54 | 6,559.84 | 6,253.29 | **25,734.84** |
| 230 | 4,500.09 | 4,288.18 | 4,293.04 | 4,023.40 | **17,104.71** |
| 231 | 9,333.76 | 8,740.66 | 8,954.27 | 8,788.56 | **35,817.25** |
| 232 | 18,054.61 | 16,915.88 | 16,747.08 | 16,993.63 | **68,711.21** |
| 233 | 7,342.96 | 7,277.82 | 7,157.59 | 6,837.06 | **28,615.43** |
| 234 | 12,905.80 | 12,305.21 | 12,325.04 | 12,303.42 | **49,839.48** |
| 235 | 4,767.51 | 4,468.32 | 4,577.07 | 4,498.67 | **18,311.57** |
| 236 | 8,361.37 | 8,230.88 | 8,617.95 | 8,300.16 | **33,510.37** |
| 237 | 3,550.31 | 3,373.31 | 3,739.64 | 3,100.70 | **13,763.96** |
| 238 | 11,614.31 | 10,668.07 | 10,182.63 | 9,468.08 | **41,933.09** |
| 239 | 2,252.53 | 2,351.33 | 2,257.47 | 2,256.36 | **9,117.68** |
| 240 | 12,807.27 | 12,209.95 | 12,522.89 | 12,674.65 | **50,214.76** |
| 241 | 11,769.57 | 11,470.98 | 11,763.65 | 11,829.50 | **46,833.69** |
| 242 | 17,379.04 | 16,386.60 | 17,098.98 | 16,328.06 | **67,192.69** |
| 243 | 7,696.42 | 7,408.10 | 7,159.10 | 7,117.62 | **29,381.24** |
| 244 | 3,845.26 | 3,785.38 | 3,750.28 | 3,745.25 | **15,126.17** |
| 245 | 10,235.09 | 10,024.76 | 10,279.95 | 10,259.13 | **40,798.92** |
| 246 | 4,294.20 | 4,237.59 | 4,147.83 | 4,204.32 | **16,883.94** |
| **VA - VIRGINIA - Total** | **216,639.31** | **206,765.97** | **210,805.73** | **206,147.72** | **840,358.73** |

**STATE: VI - VIRGIN ISLANDS**

| REGISTRANT ZIP CODE 3 | 1ST QUARTER | 2ND QUARTER | 3RD QUARTER | 4TH QUARTER | TOTAL GRAMS |
|---|---|---|---|---|---|
| 008 | 431.22 | 409.25 | 374.06 | 340.00 | **1,554.53** |
| **VI - VIRGIN ISLANDS - Total** | **431.22** | **409.25** | **374.06** | **340.00** | **1,554.53** |

**STATE: VT - VERMONT**

| REGISTRANT ZIP CODE 3 | 1ST QUARTER | 2ND QUARTER | 3RD QUARTER | 4TH QUARTER | TOTAL GRAMS |
|---|---|---|---|---|---|

| | | | | | |
|---|---|---|---|---|---|
| 050 | 1,433.73 | 1,384.64 | 1,451.69 | 1,360.85 | **5,630.91** |
| 051 | 1,065.32 | 1,084.18 | 1,083.25 | 1,092.83 | **4,325.58** |
| 052 | 1,122.04 | 1,040.72 | 1,059.65 | 1,007.42 | **4,229.82** |
| 053 | 1,135.57 | 1,075.98 | 1,048.11 | 1,061.86 | **4,321.52** |
| 054 | 6,597.68 | 6,081.49 | 6,324.23 | 6,180.88 | **25,184.29** |
| 056 | 1,508.39 | 1,446.49 | 1,565.38 | 1,507.96 | **6,028.22** |
| 057 | 1,445.10 | 1,430.95 | 1,419.26 | 1,543.38 | **5,838.70** |
| 058 | 1,472.73 | 1,628.90 | 1,669.76 | 1,680.93 | **6,452.32** |
| **VT - VERMONT - Total** | **15,780.56** | **15,173.37** | **15,621.33** | **15,436.11** | **62,011.36** |

**STATE: WA - WASHINGTON**

| REGISTRANT ZIP CODE 3 | 1ST QUARTER | 2ND QUARTER | 3RD QUARTER | 4TH QUARTER | TOTAL GRAMS |
|---|---|---|---|---|---|
| 980 | 35,011.35 | 35,193.15 | 36,078.62 | 35,361.80 | **141,644.91** |
| 981 | 20,753.72 | 18,342.97 | 19,407.02 | 18,513.95 | **77,017.66** |
| 982 | 26,582.40 | 26,112.94 | 27,378.93 | 26,747.05 | **106,821.31** |
| 983 | 24,647.86 | 24,092.45 | 24,911.17 | 24,785.82 | **98,437.30** |
| 984 | 16,830.36 | 15,965.17 | 16,411.94 | 16,179.12 | **65,386.59** |
| 985 | 13,928.54 | 13,124.53 | 14,446.10 | 14,114.83 | **55,614.00** |
| 986 | 21,002.93 | 19,457.31 | 19,516.86 | 20,013.86 | **79,990.96** |
| 988 | 6,404.56 | 6,238.99 | 6,349.17 | 6,397.69 | **25,390.41** |
| 989 | 7,753.56 | 7,692.39 | 7,912.11 | 7,741.40 | **31,099.46** |
| 990 | 2,427.03 | 2,321.70 | 2,476.39 | 2,444.79 | **9,669.91** |
| 991 | 3,544.93 | 3,699.90 | 3,709.46 | 3,748.86 | **14,703.15** |
| 992 | 12,524.28 | 12,122.58 | 12,619.02 | 12,354.98 | **49,620.85** |
| 993 | 10,244.08 | 10,065.06 | 10,273.12 | 10,379.91 | **40,962.18** |
| 994 | 714.20 | 741.87 | 783.33 | 782.99 | **3,022.38** |
| **WA - WASHINGTON - Total** | **202,369.80** | **195,171.01** | **202,273.24** | **199,567.03** | **799,381.08** |

**STATE: WI - WISCONSIN**

| REGISTRANT ZIP CODE 3 | 1ST QUARTER | 2ND QUARTER | 3RD QUARTER | 4TH QUARTER | TOTAL GRAMS |
|---|---|---|---|---|---|
| 530 | 12,245.86 | 11,621.42 | 11,951.66 | 12,352.81 | **48,171.76** |
| 531 | 13,745.42 | 13,770.46 | 14,008.64 | 13,900.30 | **55,424.82** |
| 532 | 33,912.20 | 31,069.85 | 31,968.56 | 31,443.40 | **128,394.01** |
| 534 | 3,248.39 | 3,017.93 | 3,092.34 | 3,239.65 | **12,598.31** |
| 535 | 10,895.66 | 10,493.42 | 11,190.99 | 10,887.33 | **43,467.41** |
| 537 | 7,786.96 | 6,838.50 | 7,555.79 | 7,290.26 | **29,471.51** |
| 538 | 797.71 | 760.72 | 830.33 | 853.16 | **3,241.92** |
| 539 | 4,245.59 | 4,077.30 | 3,963.28 | 3,960.96 | **16,247.12** |
| 540 | 2,156.53 | 2,145.62 | 2,333.56 | 2,247.94 | **8,883.64** |
| 541 | 2,173.65 | 2,065.94 | 2,186.96 | 2,226.50 | **8,653.04** |
| 542 | 1,119.02 | 1,213.22 | 1,304.06 | 1,223.97 | **4,860.28** |
| 543 | 2,432.47 | 2,196.07 | 2,344.60 | 2,398.51 | **9,371.65** |
| 544 | 4,208.27 | 3,948.55 | 4,129.17 | 4,112.82 | **16,398.82** |
| 545 | 1,735.70 | 1,770.32 | 1,811.59 | 1,807.34 | **7,124.95** |
| 546 | 4,149.44 | 3,801.09 | 4,067.75 | 4,044.85 | **16,063.13** |
| 547 | 2,988.48 | 2,581.98 | 2,736.06 | 2,776.97 | **11,083.48** |
| 548 | 3,368.28 | 3,578.92 | 3,700.33 | 3,649.83 | **14,297.35** |
| 549 | 7,281.19 | 6,825.59 | 7,209.67 | 6,850.72 | **28,167.17** |
| **WI - WISCONSIN - Total** | **118,490.82** | **111,776.90** | **116,385.34** | **115,267.31** | **461,920.37** |

**STATE: WV - WEST VIRGINIA**

| REGISTRANT ZIP CODE 3 | 1ST QUARTER | 2ND QUARTER | 3RD QUARTER | 4TH QUARTER | TOTAL GRAMS |
|---|---|---|---|---|---|
| 247 | 1,642.17 | 1,705.99 | 1,954.20 | 1,725.23 | **7,027.59** |
| 248 | 873.01 | 903.54 | 994.72 | 894.37 | **3,665.64** |
| 249 | 1,538.00 | 1,497.74 | 1,374.79 | 1,445.03 | **5,855.56** |
| 250 | 1,356.94 | 1,255.25 | 1,256.98 | 1,358.15 | **5,227.32** |
| 251 | 1,142.98 | 1,200.47 | 1,172.55 | 1,116.53 | **4,632.53** |
| 252 | 664.81 | 523.74 | 583.51 | 573.09 | **2,345.15** |
| 253 | 3,413.79 | 3,106.33 | 3,113.85 | 2,901.50 | **12,535.47** |
| 254 | 8,564.75 | 8,480.64 | 8,761.27 | 8,246.90 | **34,053.55** |
| 255 | 3,334.98 | 3,040.15 | 3,072.04 | 2,960.62 | **12,407.79** |
| 256 | 863.65 | 840.09 | 840.61 | 843.66 | **3,388.01** |
| 257 | 2,971.59 | 2,782.13 | 2,833.66 | 2,841.62 | **11,429.00** |

| | | | | | |
|---|---|---|---|---|---|
| 258 | 2,067.97 | 2,102.01 | 2,176.31 | 2,008.26 | **8,354.56** |
| 259 | 937.28 | 948.47 | 930.77 | 974.09 | **3,790.62** |
| 260 | 5,184.08 | 4,887.18 | 4,893.23 | 4,880.79 | **19,845.28** |
| 261 | 1,745.39 | 1,467.10 | 1,634.52 | 1,592.51 | **6,439.53** |
| 262 | 1,543.81 | 1,687.23 | 1,756.99 | 1,622.17 | **6,610.20** |
| 263 | 3,151.96 | 2,804.91 | 2,974.55 | 2,747.32 | **11,678.74** |
| 264 | 944.84 | 937.29 | 1,007.44 | 972.79 | **3,862.36** |
| 265 | 4,215.07 | 3,738.40 | 3,964.92 | 4,076.50 | **15,994.90** |
| 266 | 855.44 | 829.52 | 762.62 | 818.41 | **3,265.99** |
| 267 | 2,009.61 | 1,830.01 | 1,973.11 | 2,076.20 | **7,888.94** |
| 268 | 554.57 | 533.28 | 552.53 | 506.63 | **2,147.01** |
| **WV - WEST VIRGINIA - Total** | **49,576.70** | **47,101.48** | **48,585.18** | **47,182.39** | **192,445.76** |

**STATE: WY - WYOMING**

| REGISTRANT ZIP CODE 3 | 1ST QUARTER | 2ND QUARTER | 3RD QUARTER | 4TH QUARTER | TOTAL GRAMS |
|---|---|---|---|---|---|
| 820 | 3,111.07 | 2,804.31 | 2,843.56 | 2,737.44 | **11,496.39** |
| 822 | 518.04 | 445.78 | 468.42 | 474.29 | **1,906.54** |
| 823 | 290.87 | 341.26 | 383.70 | 374.51 | **1,390.34** |
| 824 | 2,452.74 | 2,183.01 | 2,272.89 | 2,107.89 | **9,016.53** |
| 825 | 1,590.92 | 1,379.81 | 1,405.49 | 1,386.39 | **5,762.62** |
| 826 | 3,752.07 | 3,570.51 | 3,682.38 | 3,534.03 | **14,538.99** |
| 827 | 955.48 | 897.31 | 869.83 | 896.68 | **3,619.29** |
| 828 | 966.47 | 947.56 | 1,067.60 | 1,245.77 | **4,227.39** |
| 829 | 4,809.50 | 4,944.85 | 4,842.51 | 5,008.38 | **19,605.24** |
| 830 | 160.67 | 133.64 | 171.16 | 153.33 | **618.80** |
| 831 | 442.74 | 474.33 | 553.56 | 570.28 | **2,040.91** |
| **WY - WYOMING - Total** | **19,050.56** | **18,122.38** | **18,561.10** | **18,489.01** | **74,223.05** |

**DRUG: 9150 - HYDROMORPHONE**

**STATE: AK - ALASKA**

| REGISTRANT ZIP CODE 3 | 1ST QUARTER | 2ND QUARTER | 3RD QUARTER | 4TH QUARTER | TOTAL GRAMS |
|---|---|---|---|---|---|
| 995 | 281.98 | 242.69 | 252.02 | 252.22 | **1,028.92** |
| 996 | 188.03 | 226.76 | 249.72 | 203.25 | **867.75** |
| 997 | 31.55 | 30.59 | 41.47 | 32.52 | **136.13** |
| 998 | 19.82 | 15.35 | 14.01 | 11.99 | **61.16** |
| 999 | 4.92 | 2.61 | 5.25 | 2.76 | **15.53** |
| **AK - ALASKA - Total** | **526.30** | **517.99** | **562.47** | **502.73** | **2,109.49** |

**STATE: AL - ALABAMA**

| REGISTRANT ZIP CODE 3 | 1ST QUARTER | 2ND QUARTER | 3RD QUARTER | 4TH QUARTER | TOTAL GRAMS |
|---|---|---|---|---|---|
| 350 | 146.86 | 147.40 | 126.62 | 156.55 | **577.44** |
| 351 | 83.67 | 95.44 | 86.48 | 75.39 | **340.98** |
| 352 | 329.71 | 483.13 | 598.92 | 558.50 | **1,970.26** |
| 354 | 121.16 | 75.62 | 67.49 | 81.11 | **345.38** |
| 355 | 30.64 | 36.12 | 22.70 | 29.44 | **118.90** |
| 356 | 129.13 | 113.50 | 131.56 | 130.88 | **505.08** |
| 357 | 84.49 | 102.38 | 85.92 | 92.23 | **365.01** |
| 358 | 188.36 | 165.24 | 174.92 | 149.74 | **678.25** |
| 359 | 118.25 | 87.42 | 100.38 | 105.50 | **411.56** |
| 360 | 102.58 | 76.81 | 62.93 | 69.37 | **311.69** |
| 361 | 80.25 | 72.14 | 78.43 | 75.71 | **306.53** |
| 362 | 78.29 | 56.34 | 49.62 | 62.76 | **247.01** |
| 363 | 162.16 | 153.19 | 165.42 | 137.34 | **618.11** |
| 364 | 64.71 | 49.91 | 54.55 | 46.98 | **216.15** |
| 365 | 147.96 | 159.56 | 175.51 | 186.31 | **669.34** |
| 366 | 258.83 | 209.73 | 252.14 | 282.49 | **1,003.18** |
| 367 | 18.59 | 20.12 | 19.56 | 18.02 | **76.28** |
| 368 | 95.90 | 70.66 | 80.29 | 73.82 | **320.68** |
| 369 | 4.52 | 3.77 | 3.24 | 2.79 | **14.32** |
| **AL - ALABAMA - Total** | **2,246.08** | **2,178.47** | **2,336.67** | **2,334.94** | **9,096.16** |

**STATE: AR - ARKANSAS**

| REGISTRANT ZIP CODE 3 | 1ST QUARTER | 2ND QUARTER | 3RD QUARTER | 4TH QUARTER | TOTAL GRAMS |
|---|---|---|---|---|---|
| 716 | 30.71 | 28.37 | 42.72 | 34.16 | **135.97** |

| | | | | | |
|---|---|---|---|---|---|
| 717 | 32.27 | 35.94 | 36.69 | 31.47 | **136.38** |
| 718 | 49.54 | 48.74 | 45.41 | 42.24 | **185.92** |
| 719 | 122.49 | 136.94 | 133.53 | 124.04 | **516.99** |
| 720 | 137.02 | 130.86 | 119.24 | 132.57 | **519.70** |
| 721 | 147.45 | 156.46 | 154.60 | 174.87 | **633.37** |
| 722 | 292.71 | 267.72 | 220.92 | 250.73 | **1,032.08** |
| 723 | 31.73 | 29.89 | 28.66 | 29.26 | **119.54** |
| 724 | 116.90 | 100.05 | 125.03 | 118.76 | **460.74** |
| 725 | 106.28 | 82.56 | 87.99 | 81.66 | **358.49** |
| 726 | 80.53 | 74.90 | 84.81 | 82.62 | **322.85** |
| 727 | 495.89 | 468.67 | 444.10 | 459.41 | **1,868.07** |
| 728 | 44.81 | 44.61 | 38.62 | 48.42 | **176.46** |
| 729 | 196.05 | 214.37 | 198.71 | 186.88 | **796.01** |
| **AR - ARKANSAS - Total** | **1,884.37** | **1,820.07** | **1,761.02** | **1,797.10** | **7,262.57** |

**STATE: AS - AMERICAN SAMOA**

| REGISTRANT ZIP CODE 3 | 1ST QUARTER | 2ND QUARTER | 3RD QUARTER | 4TH QUARTER | TOTAL GRAMS |
|---|---|---|---|---|---|
| 967 | 0.53 | 0.53 | 0.18 | 0.44 | **1.68** |
| **AS - AMERICAN SAMOA - Total** | **0.53** | **0.53** | **0.18** | **0.44** | **1.68** |

**STATE: AZ - ARIZONA**

| REGISTRANT ZIP CODE 3 | 1ST QUARTER | 2ND QUARTER | 3RD QUARTER | 4TH QUARTER | TOTAL GRAMS |
|---|---|---|---|---|---|
| 850 | 866.46 | 986.44 | 983.98 | 991.78 | **3,828.66** |
| 851 | 156.71 | 163.27 | 161.71 | 141.05 | **622.74** |
| 852 | 1,584.71 | 1,334.37 | 1,570.07 | 1,518.80 | **6,007.96** |
| 853 | 802.71 | 858.83 | 842.91 | 910.24 | **3,414.68** |
| 855 | 57.13 | 64.71 | 58.00 | 58.91 | **238.75** |
| 856 | 137.26 | 159.59 | 153.82 | 143.10 | **593.76** |
| 857 | 1,003.69 | 1,092.84 | 998.84 | 905.15 | **4,000.51** |
| 859 | 72.11 | 70.51 | 75.14 | 73.55 | **291.31** |
| 860 | 95.31 | 83.02 | 83.74 | 70.07 | **332.14** |
| 863 | 288.65 | 236.21 | 257.38 | 255.19 | **1,037.44** |
| 864 | 320.57 | 306.95 | 288.77 | 270.58 | **1,186.88** |
| 865 | 0.73 | 0.66 | 0.77 | 0.35 | **2.51** |
| **AZ - ARIZONA - Total** | **5,386.03** | **5,357.41** | **5,475.13** | **5,338.77** | **21,557.34** |

**STATE: CA - CALIFORNIA**

| REGISTRANT ZIP CODE 3 | 1ST QUARTER | 2ND QUARTER | 3RD QUARTER | 4TH QUARTER | TOTAL GRAMS |
|---|---|---|---|---|---|
| 900 | 1,591.13 | 1,556.26 | 1,608.92 | 1,600.71 | **6,357.02** |
| 902 | 540.23 | 462.42 | 519.27 | 466.54 | **1,988.46** |
| 903 | 28.63 | 29.49 | 29.24 | 28.34 | **115.70** |
| 904 | 229.16 | 162.67 | 157.63 | 165.19 | **714.64** |
| 905 | 446.44 | 439.57 | 449.33 | 372.61 | **1,707.96** |
| 906 | 288.80 | 240.66 | 270.18 | 263.94 | **1,063.58** |
| 907 | 307.59 | 211.49 | 259.93 | 236.92 | **1,015.93** |
| 908 | 882.70 | 1,189.33 | 792.28 | 1,245.19 | **4,109.51** |
| 910 | 283.51 | 331.09 | 303.25 | 370.44 | **1,288.29** |
| 911 | 150.40 | 215.26 | 191.53 | 305.34 | **862.53** |
| 912 | 143.61 | 146.51 | 124.70 | 139.34 | **554.16** |
| 913 | 929.85 | 896.73 | 902.34 | 885.27 | **3,614.19** |
| 914 | 265.20 | 190.64 | 251.22 | 250.47 | **957.53** |
| 915 | 175.56 | 91.74 | 151.66 | 118.94 | **537.90** |
| 916 | 78.69 | 80.37 | 92.38 | 59.93 | **311.37** |
| 917 | 1,299.63 | 1,053.66 | 1,112.64 | 1,116.58 | **4,582.51** |
| 918 | 18.96 | 25.16 | 26.09 | 25.96 | **96.17** |
| 919 | 304.68 | 359.19 | 213.52 | 236.10 | **1,113.49** |
| 920 | 1,311.41 | 1,278.28 | 1,277.95 | 1,201.98 | **5,069.62** |
| 921 | 899.83 | 1,069.75 | 809.98 | 802.00 | **3,581.56** |
| 922 | 537.18 | 585.84 | 547.33 | 607.80 | **2,278.15** |
| 923 | 895.73 | 826.60 | 860.06 | 833.15 | **3,415.54** |
| 924 | 101.91 | 102.74 | 93.19 | 96.90 | **394.73** |
| 925 | 900.63 | 810.73 | 774.25 | 732.36 | **3,217.97** |
| 926 | 812.53 | 1,092.65 | 1,119.73 | 728.78 | **3,753.70** |

| | | | | | |
|---|---|---|---|---|---|
| 927 | 268.72 | 239.16 | 260.21 | 253.35 | **1,021.45** |
| 928 | 1,142.55 | 946.51 | 1,041.23 | 956.83 | **4,087.13** |
| 930 | 426.00 | 360.66 | 366.83 | 368.99 | **1,522.47** |
| 931 | 117.33 | 102.33 | 95.77 | 86.01 | **401.44** |
| 932 | 375.55 | 379.35 | 312.73 | 294.18 | **1,361.81** |
| 933 | 485.10 | 394.71 | 435.30 | 389.48 | **1,704.59** |
| 934 | 596.84 | 576.29 | 559.57 | 568.37 | **2,301.07** |
| 935 | 797.00 | 761.52 | 720.19 | 686.57 | **2,965.29** |
| 936 | 202.28 | 179.98 | 203.55 | 191.58 | **777.39** |
| 937 | 406.18 | 385.23 | 382.06 | 409.14 | **1,582.61** |
| 939 | 157.30 | 146.54 | 182.01 | 142.06 | **627.92** |
| 940 | 442.66 | 319.68 | 381.47 | 440.70 | **1,584.50** |
| 941 | 651.79 | 581.89 | 616.47 | 539.08 | **2,389.23** |
| 943 | 281.48 | 275.82 | 278.00 | 258.24 | **1,093.54** |
| 944 | 51.79 | 73.99 | 41.20 | 51.50 | **218.48** |
| 945 | 1,789.05 | 1,728.50 | 1,826.37 | 1,628.14 | **6,972.06** |
| 946 | 330.90 | 306.40 | 336.78 | 243.50 | **1,217.59** |
| 947 | 212.82 | 107.47 | 73.40 | 41.18 | **434.87** |
| 948 | 57.77 | 52.43 | 60.65 | 44.95 | **215.81** |
| 949 | 383.47 | 347.61 | 382.90 | 359.46 | **1,473.43** |
| 950 | 432.08 | 383.28 | 404.07 | 406.77 | **1,626.20** |
| 951 | 285.67 | 257.43 | 306.03 | 236.25 | **1,085.38** |
| 952 | 206.01 | 213.14 | 199.37 | 185.12 | **803.65** |
| 953 | 639.04 | 666.67 | 637.51 | 583.81 | **2,527.03** |
| 954 | 573.87 | 550.72 | 449.78 | 451.63 | **2,026.01** |
| 955 | 176.93 | 165.84 | 159.34 | 151.47 | **653.59** |
| 956 | 1,058.25 | 1,018.84 | 1,058.47 | 984.36 | **4,119.91** |
| 957 | 114.80 | 136.04 | 136.95 | 137.44 | **525.23** |
| 958 | 693.16 | 692.70 | 812.31 | 693.34 | **2,891.51** |
| 959 | 483.12 | 460.51 | 414.99 | 415.72 | **1,774.34** |
| 960 | 241.99 | 240.45 | 211.65 | 233.31 | **927.40** |
| 961 | 58.50 | 36.02 | 39.63 | 57.67 | **191.82** |
| **CA - CALIFORNIA - Total** | **27,563.99** | **26,536.57** | **26,325.41** | **25,381.01** | **105,806.98** |

**STATE: CO - COLORADO**

| REGISTRANT ZIP CODE 3 | 1ST QUARTER | 2ND QUARTER | 3RD QUARTER | 4TH QUARTER | TOTAL GRAMS |
|---|---|---|---|---|---|
| 800 | 1,074.24 | 1,023.25 | 906.84 | 950.79 | **3,955.12** |
| 801 | 1,369.20 | 1,291.32 | 1,275.80 | 1,146.31 | **5,082.63** |
| 802 | 942.12 | 1,005.25 | 1,079.94 | 802.02 | **3,829.33** |
| 803 | 99.01 | 102.06 | 73.63 | 72.54 | **347.23** |
| 804 | 217.56 | 178.41 | 202.40 | 182.33 | **780.71** |
| 805 | 448.08 | 500.11 | 455.74 | 434.63 | **1,838.56** |
| 806 | 279.52 | 197.09 | 226.33 | 186.71 | **889.66** |
| 807 | 59.51 | 51.53 | 63.45 | 63.69 | **238.19** |
| 808 | 110.58 | 111.93 | 126.80 | 120.88 | **470.19** |
| 809 | 594.66 | 628.12 | 660.68 | 633.95 | **2,517.40** |
| 810 | 262.81 | 236.61 | 270.71 | 265.32 | **1,035.46** |
| 811 | 13.31 | 16.75 | 17.29 | 17.07 | **64.42** |
| 812 | 78.00 | 96.58 | 93.34 | 80.44 | **348.35** |
| 813 | 40.80 | 56.95 | 58.59 | 40.20 | **196.55** |
| 814 | 71.90 | 53.44 | 61.63 | 41.58 | **228.56** |
| 815 | 139.31 | 180.61 | 128.24 | 105.63 | **553.79** |
| 816 | 104.45 | 105.40 | 96.61 | 83.34 | **389.80** |
| **CO - COLORADO - Total** | **5,905.07** | **5,835.41** | **5,798.01** | **5,227.46** | **22,765.94** |

**STATE: CT - CONNECTICUT**

| REGISTRANT ZIP CODE 3 | 1ST QUARTER | 2ND QUARTER | 3RD QUARTER | 4TH QUARTER | TOTAL GRAMS |
|---|---|---|---|---|---|
| 060 | 898.39 | 785.95 | 872.43 | 838.67 | **3,395.43** |
| 061 | 446.26 | 392.09 | 461.78 | 476.17 | **1,776.30** |
| 062 | 120.94 | 116.29 | 112.54 | 129.78 | **479.56** |
| 063 | 227.84 | 218.03 | 219.57 | 242.86 | **908.31** |
| 064 | 1,524.68 | 1,236.20 | 1,152.62 | 1,101.14 | **5,014.64** |

| | | | | | |
|---|---|---|---|---|---|
| 065 | 460.00 | 598.80 | 345.62 | 383.05 | **1,787.47** |
| 066 | 236.91 | 170.87 | 119.23 | 113.96 | **640.96** |
| 067 | 308.02 | 313.24 | 365.88 | 345.37 | **1,332.51** |
| 068 | 597.14 | 559.85 | 552.15 | 526.69 | **2,235.82** |
| 069 | 170.15 | 161.54 | 104.90 | 108.52 | **545.11** |
| **CT - CONNECTICUT - Total** | **4,990.33** | **4,552.84** | **4,306.73** | **4,266.20** | **18,116.11** |

**STATE: DC - DISTRICT OF COLUMBIA**

| REGISTRANT ZIP CODE 3 | 1ST QUARTER | 2ND QUARTER | 3RD QUARTER | 4TH QUARTER | TOTAL GRAMS |
|---|---|---|---|---|---|
| 200 | 434.78 | 353.85 | 376.63 | 376.43 | **1,541.69** |
| 204 | 11.53 | 10.11 | 2.70 | 4.85 | **29.18** |
| **DC - DISTRICT OF COLUMBIA - Total** | **446.31** | **363.96** | **379.32** | **381.28** | **1,570.87** |

**STATE: DE - DELAWARE**

| REGISTRANT ZIP CODE 3 | 1ST QUARTER | 2ND QUARTER | 3RD QUARTER | 4TH QUARTER | TOTAL GRAMS |
|---|---|---|---|---|---|
| 197 | 532.21 | 524.12 | 517.43 | 472.13 | **2,045.89** |
| 198 | 255.95 | 222.41 | 193.40 | 222.22 | **893.98** |
| 199 | 476.84 | 468.03 | 468.07 | 449.91 | **1,862.84** |
| **DE - DELAWARE - Total** | **1,265.00** | **1,214.55** | **1,178.90** | **1,144.26** | **4,802.71** |

**STATE: FL - FLORIDA**

| REGISTRANT ZIP CODE 3 | 1ST QUARTER | 2ND QUARTER | 3RD QUARTER | 4TH QUARTER | TOTAL GRAMS |
|---|---|---|---|---|---|
| 320 | 552.35 | 564.29 | 656.02 | 582.42 | **2,355.07** |
| 321 | 820.37 | 808.59 | 780.05 | 844.94 | **3,253.95** |
| 322 | 1,181.49 | 1,200.73 | 1,106.77 | 1,182.01 | **4,671.00** |
| 323 | 469.92 | 398.57 | 455.01 | 537.79 | **1,861.30** |
| 324 | 227.08 | 192.81 | 157.27 | 172.36 | **749.51** |
| 325 | 561.70 | 508.04 | 461.62 | 518.70 | **2,050.06** |
| 326 | 534.44 | 471.29 | 595.96 | 586.19 | **2,187.88** |
| 327 | 2,459.91 | 2,103.03 | 2,245.53 | 1,829.73 | **8,638.20** |
| 328 | 1,350.41 | 1,455.20 | 1,365.47 | 1,456.39 | **5,627.47** |
| 329 | 1,123.79 | 1,053.65 | 1,059.81 | 1,057.93 | **4,295.17** |
| 330 | 1,004.48 | 834.79 | 994.30 | 1,073.51 | **3,907.09** |
| 331 | 903.14 | 1,119.87 | 849.24 | 808.08 | **3,680.34** |
| 333 | 772.61 | 618.49 | 628.49 | 673.10 | **2,692.70** |
| 334 | 1,557.49 | 1,401.36 | 1,399.87 | 1,312.90 | **5,671.63** |
| 335 | 1,523.98 | 1,395.71 | 1,493.70 | 1,327.79 | **5,741.19** |
| 336 | 7,637.01 | 15,314.35 | 3,014.94 | 2,826.78 | **28,793.08** |
| 337 | 2,013.37 | 2,056.42 | 2,073.88 | 2,039.61 | **8,183.27** |
| 338 | 751.08 | 865.14 | 869.07 | 712.11 | **3,197.39** |
| 339 | 1,374.72 | 1,171.55 | 1,236.61 | 1,269.62 | **5,052.50** |
| 341 | 491.34 | 516.26 | 504.13 | 567.49 | **2,079.21** |
| 342 | 1,669.35 | 1,658.43 | 1,886.34 | 1,758.71 | **6,972.83** |
| 344 | 623.24 | 595.36 | 625.84 | 598.74 | **2,443.18** |
| 346 | 2,851.14 | 2,679.18 | 2,604.76 | 2,559.91 | **10,694.99** |
| 347 | 880.83 | 816.95 | 861.63 | 832.74 | **3,392.16** |
| 349 | 963.13 | 897.91 | 861.19 | 860.10 | **3,582.32** |
| **FL - FLORIDA - Total** | **34,298.37** | **40,697.97** | **28,787.52** | **27,989.64** | **131,773.51** |

**STATE: GA - GEORGIA**

| REGISTRANT ZIP CODE 3 | 1ST QUARTER | 2ND QUARTER | 3RD QUARTER | 4TH QUARTER | TOTAL GRAMS |
|---|---|---|---|---|---|
| 300 | 1,500.52 | 1,417.02 | 1,099.49 | 1,086.41 | **5,103.45** |
| 301 | 813.27 | 784.78 | 800.72 | 729.69 | **3,128.47** |
| 302 | 387.15 | 389.98 | 367.91 | 351.00 | **1,496.03** |
| 303 | 786.41 | 951.86 | 617.33 | 634.57 | **2,990.17** |
| 304 | 95.42 | 114.30 | 80.76 | 92.07 | **382.54** |
| 305 | 465.49 | 465.78 | 476.47 | 431.70 | **1,839.43** |
| 306 | 406.59 | 393.81 | 471.01 | 377.32 | **1,648.74** |
| 307 | 155.32 | 138.40 | 143.95 | 219.25 | **656.93** |
| 308 | 115.49 | 104.79 | 102.18 | 111.59 | **434.05** |
| 309 | 219.16 | 213.17 | 209.44 | 217.10 | **858.87** |
| 310 | 165.60 | 147.42 | 157.18 | 162.21 | **632.41** |
| 312 | 42.28 | 62.48 | 61.38 | 53.11 | **219.25** |
| 313 | 88.11 | 86.74 | 86.61 | 80.97 | **342.44** |

| | | | | | |
|---|---|---|---|---|---|
| 314 | 213.22 | 125.08 | 146.03 | 131.60 | **615.93** |
| 315 | 176.49 | 215.23 | 200.10 | 220.92 | **812.74** |
| 316 | 82.67 | 78.16 | 78.81 | 91.47 | **331.11** |
| 317 | 419.59 | 356.04 | 342.77 | 350.08 | **1,468.48** |
| 318 | 7.71 | 5.32 | 5.98 | 5.23 | **24.25** |
| 319 | 78.76 | 139.91 | 61.22 | 88.72 | **368.62** |
| 398 | 48.46 | 36.80 | 42.48 | 32.87 | **160.60** |
| **GA - GEORGIA - Total** | **6,267.70** | **6,227.08** | **5,551.83** | **5,467.90** | **23,514.51** |

**STATE: GU - GUAM**

| REGISTRANT ZIP CODE 3 | 1ST QUARTER | 2ND QUARTER | 3RD QUARTER | 4TH QUARTER | TOTAL GRAMS |
|---|---|---|---|---|---|
| 969 | 28.81 | 31.78 | 28.73 | 17.12 | **106.44** |
| **GU - GUAM - Total** | **28.81** | **31.78** | **28.73** | **17.12** | **106.44** |

**STATE: HI - HAWAII**

| REGISTRANT ZIP CODE 3 | 1ST QUARTER | 2ND QUARTER | 3RD QUARTER | 4TH QUARTER | TOTAL GRAMS |
|---|---|---|---|---|---|
| 967 | 255.69 | 264.63 | 261.03 | 226.59 | **1,007.94** |
| 968 | 303.97 | 261.99 | 222.95 | 232.76 | **1,021.67** |
| **HI - HAWAII - Total** | **559.66** | **526.62** | **483.98** | **459.36** | **2,029.61** |

**STATE: IA - IOWA**

| REGISTRANT ZIP CODE 3 | 1ST QUARTER | 2ND QUARTER | 3RD QUARTER | 4TH QUARTER | TOTAL GRAMS |
|---|---|---|---|---|---|
| 500 | 195.19 | 221.65 | 198.26 | 196.70 | **811.80** |
| 501 | 98.53 | 70.63 | 86.86 | 90.84 | **346.85** |
| 502 | 149.11 | 143.78 | 126.03 | 116.09 | **535.01** |
| 503 | 409.69 | 404.86 | 345.37 | 361.59 | **1,521.51** |
| 504 | 63.23 | 71.17 | 59.63 | 60.21 | **254.23** |
| 505 | 38.91 | 33.49 | 28.56 | 37.12 | **138.09** |
| 506 | 54.60 | 50.53 | 61.29 | 61.02 | **227.44** |
| 507 | 44.72 | 31.24 | 43.69 | 42.36 | **162.00** |
| 508 | 15.56 | 12.08 | 15.18 | 13.94 | **56.76** |
| 510 | 14.08 | 16.40 | 15.67 | 13.51 | **59.66** |
| 511 | 55.00 | 40.38 | 46.29 | 53.84 | **195.52** |
| 512 | 7.00 | 4.94 | 4.91 | 4.85 | **21.70** |
| 513 | 18.83 | 24.92 | 14.06 | 18.37 | **76.17** |
| 514 | 32.06 | 28.41 | 31.79 | 26.78 | **119.03** |
| 515 | 51.23 | 47.06 | 51.25 | 51.60 | **201.14** |
| 516 | 6.56 | 11.19 | 7.13 | 11.40 | **36.28** |
| 520 | 39.38 | 45.48 | 52.21 | 42.09 | **179.16** |
| 521 | 10.56 | 7.20 | 6.00 | 9.89 | **33.65** |
| 522 | 309.03 | 265.57 | 304.92 | 331.62 | **1,211.13** |
| 523 | 37.18 | 42.98 | 43.77 | 49.12 | **173.06** |
| 524 | 88.14 | 87.54 | 66.25 | 55.54 | **297.47** |
| 525 | 120.83 | 100.20 | 118.64 | 119.98 | **459.66** |
| 526 | 66.26 | 56.29 | 71.54 | 58.47 | **252.56** |
| 527 | 65.74 | 49.82 | 47.99 | 47.16 | **210.72** |
| 528 | 53.99 | 40.53 | 41.97 | 38.72 | **175.22** |
| **IA - IOWA - Total** | **2,045.41** | **1,908.33** | **1,889.26** | **1,912.81** | **7,755.82** |

**STATE: ID - IDAHO**

| REGISTRANT ZIP CODE 3 | 1ST QUARTER | 2ND QUARTER | 3RD QUARTER | 4TH QUARTER | TOTAL GRAMS |
|---|---|---|---|---|---|
| 832 | 141.91 | 124.29 | 136.15 | 138.96 | **541.32** |
| 833 | 109.61 | 123.45 | 102.58 | 100.26 | **435.90** |
| 834 | 85.88 | 94.66 | 129.13 | 124.17 | **433.84** |
| 835 | 81.34 | 83.63 | 79.70 | 74.76 | **319.43** |
| 836 | 478.09 | 447.82 | 438.32 | 409.02 | **1,773.25** |
| 837 | 408.16 | 395.55 | 404.69 | 357.31 | **1,565.71** |
| 838 | 162.43 | 167.74 | 169.90 | 182.03 | **682.10** |
| **ID - IDAHO - Total** | **1,467.43** | **1,437.15** | **1,460.46** | **1,386.51** | **5,751.55** |

**STATE: IL - ILLINOIS**

| REGISTRANT ZIP CODE 3 | 1ST QUARTER | 2ND QUARTER | 3RD QUARTER | 4TH QUARTER | TOTAL GRAMS |
|---|---|---|---|---|---|
| 600 | 1,657.55 | 1,559.91 | 1,612.99 | 1,422.16 | **6,252.61** |
| 601 | 1,022.57 | 825.61 | 774.82 | 732.35 | **3,355.36** |
| 602 | 77.28 | 70.31 | 58.22 | 51.92 | **257.73** |

| | | | | | |
|---|---|---|---|---|---|
| 603 | 43.71 | 53.09 | 41.97 | 48.12 | **186.89** |
| 604 | 1,036.33 | 965.72 | 881.05 | 897.62 | **3,780.72** |
| 605 | 632.71 | 532.85 | 503.19 | 485.91 | **2,154.67** |
| 606 | 1,359.35 | 1,675.74 | 1,276.10 | 1,390.13 | **5,701.32** |
| 607 | 62.27 | 39.16 | 48.21 | 49.96 | **199.60** |
| 608 | 66.33 | 75.78 | 65.92 | 54.66 | **262.69** |
| 609 | 107.92 | 99.81 | 95.61 | 100.25 | **403.59** |
| 610 | 70.12 | 86.42 | 100.17 | 80.75 | **337.46** |
| 611 | 161.79 | 162.38 | 153.04 | 120.33 | **597.55** |
| 612 | 53.81 | 52.39 | 53.41 | 52.85 | **212.47** |
| 613 | 41.66 | 55.05 | 51.39 | 56.66 | **204.76** |
| 614 | 31.22 | 34.20 | 35.47 | 38.04 | **138.93** |
| 615 | 36.39 | 43.79 | 37.42 | 44.40 | **161.99** |
| 616 | 136.18 | 142.96 | 102.19 | 98.19 | **479.52** |
| 617 | 129.08 | 141.21 | 145.22 | 133.63 | **549.14** |
| 618 | 180.44 | 164.33 | 151.00 | 157.31 | **653.08** |
| 619 | 35.82 | 33.20 | 35.50 | 34.23 | **138.76** |
| 620 | 107.27 | 99.29 | 84.40 | 96.28 | **387.23** |
| 622 | 180.74 | 151.76 | 164.40 | 139.92 | **636.81** |
| 623 | 50.39 | 38.24 | 51.06 | 47.07 | **186.75** |
| 624 | 84.20 | 65.31 | 72.09 | 65.24 | **286.86** |
| 625 | 147.49 | 162.56 | 169.10 | 178.34 | **657.48** |
| 626 | 45.15 | 42.86 | 44.21 | 49.59 | **181.81** |
| 627 | 140.06 | 119.33 | 127.78 | 119.27 | **506.44** |
| 628 | 66.90 | 62.91 | 71.21 | 58.37 | **259.40** |
| 629 | 87.03 | 81.76 | 85.27 | 93.65 | **347.71** |
| **IL - ILLINOIS - Total** | **7,851.79** | **7,637.95** | **7,092.41** | **6,897.18** | **29,479.33** |

**STATE: IN - INDIANA**

| REGISTRANT ZIP CODE 3 | 1ST QUARTER | 2ND QUARTER | 3RD QUARTER | 4TH QUARTER | TOTAL GRAMS |
|---|---|---|---|---|---|
| 460 | 385.99 | 360.29 | 489.57 | 358.42 | **1,594.26** |
| 461 | 384.73 | 266.14 | 290.85 | 266.64 | **1,208.37** |
| 462 | 1,297.80 | 1,927.87 | 980.83 | 979.86 | **5,186.36** |
| 463 | 359.95 | 302.28 | 297.34 | 315.51 | **1,275.08** |
| 464 | 39.19 | 27.57 | 30.99 | 37.02 | **134.76** |
| 465 | 269.32 | 301.59 | 287.65 | 282.34 | **1,140.90** |
| 466 | 76.38 | 167.39 | 86.13 | 53.85 | **383.75** |
| 467 | 213.10 | 227.80 | 195.26 | 260.23 | **896.38** |
| 468 | 462.09 | 455.30 | 371.47 | 354.49 | **1,643.35** |
| 469 | 115.96 | 157.85 | 140.48 | 119.42 | **533.72** |
| 470 | 13.49 | 19.75 | 21.11 | 26.14 | **80.50** |
| 471 | 144.62 | 132.87 | 113.09 | 108.19 | **498.78** |
| 472 | 113.06 | 128.07 | 130.40 | 109.25 | **480.79** |
| 473 | 197.88 | 214.34 | 299.29 | 244.44 | **955.95** |
| 474 | 162.83 | 189.08 | 184.64 | 165.87 | **702.42** |
| 475 | 183.44 | 188.08 | 206.91 | 143.13 | **721.56** |
| 476 | 107.43 | 82.94 | 90.87 | 77.64 | **358.88** |
| 477 | 483.52 | 196.61 | 646.64 | 208.14 | **1,534.92** |
| 478 | 94.11 | 102.18 | 83.08 | 73.05 | **352.42** |
| 479 | 132.06 | 118.21 | 137.80 | 139.64 | **527.71** |
| **IN - INDIANA - Total** | **5,236.95** | **5,566.22** | **5,084.40** | **4,323.29** | **20,210.86** |

**STATE: KS - KANSAS**

| REGISTRANT ZIP CODE 3 | 1ST QUARTER | 2ND QUARTER | 3RD QUARTER | 4TH QUARTER | TOTAL GRAMS |
|---|---|---|---|---|---|
| 660 | 321.66 | 327.14 | 288.57 | 306.43 | **1,243.81** |
| 661 | 108.86 | 177.82 | 115.51 | 120.63 | **522.81** |
| 662 | 681.40 | 525.02 | 487.37 | 447.19 | **2,140.98** |
| 664 | 71.45 | 77.57 | 85.68 | 68.31 | **303.02** |
| 665 | 90.86 | 101.50 | 144.80 | 124.58 | **461.74** |
| 666 | 106.77 | 151.50 | 121.70 | 111.16 | **491.12** |
| 667 | 75.12 | 56.32 | 69.12 | 71.10 | **271.67** |
| 668 | 17.90 | 17.98 | 15.24 | 13.12 | **64.24** |

| | | | | | |
|---|---|---|---|---|---|
| 669 | 10.04 | 6.21 | 11.73 | 13.36 | **41.33** |
| 670 | 107.71 | 108.70 | 107.03 | 109.22 | **432.67** |
| 671 | 59.11 | 55.87 | 63.09 | 75.11 | **253.18** |
| 672 | 515.32 | 391.39 | 458.88 | 471.43 | **1,837.02** |
| 673 | 28.06 | 33.34 | 30.90 | 28.37 | **120.67** |
| 674 | 125.70 | 114.66 | 124.65 | 119.59 | **484.60** |
| 675 | 224.22 | 210.38 | 210.65 | 232.52 | **877.77** |
| 676 | 62.00 | 65.41 | 70.39 | 69.03 | **266.83** |
| 677 | 13.96 | 13.83 | 11.33 | 13.45 | **52.56** |
| 678 | 48.48 | 44.37 | 46.78 | 42.30 | **181.93** |
| 679 | 13.25 | 18.58 | 14.58 | 13.17 | **59.58** |
| **KS - KANSAS - Total** | **2,681.87** | **2,497.59** | **2,478.02** | **2,450.06** | **10,107.53** |

**STATE: KY - KENTUCKY**

| REGISTRANT ZIP CODE 3 | 1ST QUARTER | 2ND QUARTER | 3RD QUARTER | 4TH QUARTER | TOTAL GRAMS |
|---|---|---|---|---|---|
| 400 | 90.25 | 87.51 | 83.87 | 73.69 | **335.31** |
| 401 | 36.38 | 32.65 | 37.39 | 25.43 | **131.84** |
| 402 | 777.11 | 573.26 | 643.28 | 681.11 | **2,674.76** |
| 403 | 58.24 | 70.97 | 59.14 | 59.17 | **247.52** |
| 404 | 32.97 | 43.96 | 34.14 | 15.01 | **126.08** |
| 405 | 337.10 | 304.86 | 219.11 | 265.77 | **1,126.85** |
| 406 | 22.72 | 16.73 | 18.37 | 15.18 | **73.02** |
| 407 | 24.33 | 16.42 | 9.67 | 15.56 | **65.98** |
| 408 | 5.30 | 6.54 | 9.84 | 9.40 | **31.08** |
| 409 | 6.87 | 6.39 | 11.15 | 5.22 | **29.63** |
| 410 | 133.90 | 208.45 | 168.72 | 100.21 | **611.29** |
| 411 | 39.56 | 22.80 | 36.52 | 62.72 | **161.59** |
| 412 | 5.02 | 2.49 | 2.69 | 3.40 | **13.60** |
| 413 | 4.70 | 5.15 | 5.70 | 4.18 | **19.73** |
| 414 | 0.78 | 0.38 | 1.03 | 0.47 | **2.66** |
| 415 | 19.17 | 11.07 | 10.10 | 8.67 | **49.01** |
| 416 | 2.61 | 1.90 | 3.14 | 3.05 | **10.70** |
| 417 | 31.60 | 45.74 | 35.21 | 47.30 | **159.86** |
| 418 | 2.35 | 3.29 | 3.66 | 3.77 | **13.06** |
| 420 | 161.61 | 168.54 | 155.58 | 214.93 | **700.65** |
| 421 | 234.55 | 152.71 | 171.11 | 121.72 | **680.09** |
| 422 | 87.54 | 93.51 | 103.06 | 104.84 | **388.95** |
| 423 | 100.74 | 92.55 | 106.94 | 99.95 | **400.18** |
| 424 | 170.19 | 177.93 | 174.35 | 191.30 | **713.76** |
| 425 | 11.53 | 12.11 | 7.91 | 24.14 | **55.68** |
| 426 | 8.47 | 6.27 | 6.37 | 6.69 | **27.80** |
| 427 | 74.66 | 77.42 | 74.65 | 75.98 | **302.71** |
| **KY - KENTUCKY - Total** | **2,480.24** | **2,241.61** | **2,192.71** | **2,238.84** | **9,153.39** |

**STATE: LA - LOUISIANA**

| REGISTRANT ZIP CODE 3 | 1ST QUARTER | 2ND QUARTER | 3RD QUARTER | 4TH QUARTER | TOTAL GRAMS |
|---|---|---|---|---|---|
| 700 | 218.53 | 185.38 | 194.82 | 183.50 | **782.23** |
| 701 | 211.46 | 171.22 | 265.80 | 161.14 | **809.63** |
| 703 | 74.12 | 59.44 | 62.45 | 65.93 | **261.94** |
| 704 | 301.47 | 295.55 | 298.28 | 315.97 | **1,211.27** |
| 705 | 449.91 | 413.05 | 421.85 | 599.71 | **1,884.51** |
| 706 | 186.02 | 126.71 | 127.18 | 143.64 | **583.55** |
| 707 | 303.65 | 114.91 | 292.41 | 133.20 | **844.18** |
| 708 | 219.05 | 219.04 | 207.42 | 190.47 | **835.98** |
| 710 | 27.37 | 31.86 | 30.29 | 41.63 | **131.15** |
| 711 | 101.08 | 107.16 | 101.80 | 131.14 | **441.17** |
| 712 | 152.66 | 74.49 | 76.71 | 94.94 | **398.80** |
| 713 | 83.15 | 85.52 | 85.46 | 72.81 | **326.95** |
| 714 | 64.32 | 42.90 | 54.82 | 40.48 | **202.52** |
| **LA - LOUISIANA - Total** | **2,392.79** | **1,927.24** | **2,219.28** | **2,174.57** | **8,713.88** |

**STATE: MA - MASSACHUSETTS**

| REGISTRANT ZIP CODE 3 | 1ST QUARTER | 2ND QUARTER | 3RD QUARTER | 4TH QUARTER | TOTAL GRAMS |
|---|---|---|---|---|---|

| | | | | | |
|---|---|---|---|---|---|
| 010 | 445.93 | 427.06 | 413.56 | 374.48 | **1,661.04** |
| 011 | 344.19 | 268.23 | 322.63 | 275.15 | **1,210.21** |
| 012 | 100.89 | 96.43 | 106.53 | 101.00 | **404.85** |
| 013 | 87.88 | 101.47 | 79.78 | 95.81 | **364.95** |
| 014 | 70.68 | 116.58 | 109.20 | 96.67 | **393.14** |
| 015 | 185.15 | 157.50 | 145.21 | 146.83 | **634.69** |
| 016 | 158.39 | 167.47 | 151.94 | 181.97 | **659.76** |
| 017 | 341.38 | 295.27 | 360.20 | 390.04 | **1,386.89** |
| 018 | 1,011.31 | 1,110.21 | 682.87 | 837.83 | **3,642.22** |
| 019 | 325.92 | 283.41 | 306.26 | 276.69 | **1,192.29** |
| 020 | 389.97 | 408.24 | 336.11 | 326.55 | **1,460.87** |
| 021 | 1,676.38 | 2,562.47 | 1,622.82 | 2,097.25 | **7,958.91** |
| 022 | 252.56 | 249.98 | 228.82 | 246.55 | **977.92** |
| 023 | 283.24 | 275.01 | 299.81 | 349.54 | **1,207.60** |
| 024 | 173.25 | 183.40 | 198.57 | 161.24 | **716.46** |
| 025 | 82.75 | 77.29 | 95.67 | 84.10 | **339.81** |
| 026 | 127.07 | 159.51 | 125.79 | 120.95 | **533.33** |
| 027 | 443.66 | 432.49 | 417.90 | 401.54 | **1,695.59** |
| **MA - MASSACHUSETTS - Total** | **6,500.63** | **7,372.03** | **6,003.67** | **6,564.19** | **26,440.52** |
| **STATE: MD - MARYLAND** | | | | | |
| REGISTRANT ZIP CODE 3 | 1ST QUARTER | 2ND QUARTER | 3RD QUARTER | 4TH QUARTER | **TOTAL GRAMS** |
| 206 | 318.08 | 286.14 | 314.20 | 295.82 | **1,214.24** |
| 207 | 457.34 | 439.63 | 419.92 | 398.84 | **1,715.73** |
| 208 | 809.34 | 796.72 | 710.28 | 740.18 | **3,056.52** |
| 209 | 171.81 | 236.44 | 133.30 | 109.85 | **651.40** |
| 210 | 887.46 | 715.40 | 858.73 | 847.83 | **3,309.42** |
| 211 | 337.55 | 320.11 | 343.45 | 351.32 | **1,352.43** |
| 212 | 1,762.67 | 1,769.86 | 1,440.66 | 1,344.48 | **6,317.67** |
| 214 | 111.72 | 97.23 | 138.87 | 127.31 | **475.14** |
| 215 | 67.49 | 93.31 | 78.41 | 66.81 | **306.01** |
| 216 | 115.05 | 104.12 | 103.38 | 104.83 | **427.38** |
| 217 | 550.80 | 524.03 | 544.23 | 481.76 | **2,100.82** |
| 218 | 319.18 | 298.79 | 281.66 | 338.05 | **1,237.69** |
| 219 | 167.29 | 124.14 | 126.30 | 124.27 | **542.00** |
| **MD - MARYLAND - Total** | **6,075.78** | **5,805.93** | **5,493.39** | **5,331.35** | **22,706.44** |
| **STATE: ME - MAINE** | | | | | |
| REGISTRANT ZIP CODE 3 | 1ST QUARTER | 2ND QUARTER | 3RD QUARTER | 4TH QUARTER | **TOTAL GRAMS** |
| 039 | 12.45 | 19.82 | 15.18 | 18.22 | **65.67** |
| 040 | 284.80 | 355.50 | 332.42 | 319.77 | **1,292.49** |
| 041 | 359.19 | 365.15 | 343.48 | 471.69 | **1,539.51** |
| 042 | 97.32 | 102.17 | 116.35 | 112.11 | **427.94** |
| 043 | 315.78 | 296.61 | 298.39 | 315.97 | **1,226.74** |
| 044 | 212.87 | 162.91 | 173.19 | 166.30 | **715.28** |
| 045 | 37.37 | 57.47 | 50.05 | 39.76 | **184.65** |
| 046 | 97.34 | 77.84 | 60.38 | 49.24 | **284.81** |
| 047 | 22.08 | 37.29 | 21.56 | 39.04 | **119.97** |
| 048 | 37.61 | 27.01 | 29.42 | 18.79 | **112.83** |
| 049 | 126.52 | 144.37 | 141.59 | 140.70 | **553.18** |
| **ME - MAINE - Total** | **1,603.34** | **1,646.13** | **1,582.01** | **1,691.59** | **6,523.07** |
| **STATE: MI - MICHIGAN** | | | | | |
| REGISTRANT ZIP CODE 3 | 1ST QUARTER | 2ND QUARTER | 3RD QUARTER | 4TH QUARTER | **TOTAL GRAMS** |
| 480 | 1,107.99 | 937.98 | 954.82 | 765.52 | **3,766.30** |
| 481 | 922.12 | 758.32 | 719.85 | 664.72 | **3,065.01** |
| 482 | 608.90 | 523.51 | 479.49 | 548.90 | **2,160.80** |
| 483 | 986.10 | 965.99 | 1,012.74 | 784.78 | **3,749.61** |
| 484 | 226.46 | 210.89 | 245.62 | 264.08 | **947.06** |
| 485 | 121.48 | 257.05 | 103.13 | 107.92 | **589.58** |
| 486 | 324.16 | 300.61 | 263.42 | 301.14 | **1,189.32** |
| 487 | 112.15 | 96.61 | 108.14 | 96.14 | **413.04** |
| 488 | 275.97 | 254.57 | 292.67 | 273.22 | **1,096.42** |

| | | | | | |
|---|---|---|---|---|---|
| 489 | 110.36 | 181.39 | 130.28 | 140.38 | **562.41** |
| 490 | 398.51 | 352.83 | 348.92 | 311.13 | **1,411.39** |
| 491 | 15.98 | 29.88 | 23.47 | 24.91 | **94.25** |
| 492 | 115.48 | 98.20 | 86.88 | 92.33 | **392.88** |
| 493 | 82.67 | 90.61 | 88.25 | 79.38 | **340.91** |
| 494 | 242.05 | 195.11 | 180.39 | 178.09 | **795.64** |
| 495 | 298.10 | 368.44 | 350.82 | 300.77 | **1,318.13** |
| 496 | 263.87 | 237.63 | 265.83 | 224.37 | **991.70** |
| 497 | 142.57 | 139.93 | 151.88 | 139.66 | **574.05** |
| 498 | 163.75 | 175.61 | 101.83 | 98.76 | **539.95** |
| 499 | 27.74 | 33.63 | 35.63 | 36.73 | **133.73** |
| **MI - MICHIGAN - Total** | **6,546.42** | **6,208.78** | **5,944.06** | **5,432.92** | **24,132.18** |

**STATE: MN - MINNESOTA**

| REGISTRANT ZIP CODE 3 | 1ST QUARTER | 2ND QUARTER | 3RD QUARTER | 4TH QUARTER | **TOTAL GRAMS** |
|---|---|---|---|---|---|
| 550 | 495.48 | 485.22 | 484.10 | 440.12 | **1,904.93** |
| 551 | 876.72 | 1,012.40 | 797.95 | 1,038.13 | **3,725.20** |
| 553 | 553.42 | 565.77 | 592.12 | 495.54 | **2,206.84** |
| 554 | 1,603.25 | 1,562.62 | 1,593.16 | 1,438.13 | **6,197.17** |
| 556 | 5.87 | 6.03 | 6.03 | 6.03 | **23.96** |
| 557 | 152.44 | 139.92 | 137.69 | 137.15 | **567.21** |
| 558 | 107.51 | 162.67 | 172.14 | 143.97 | **586.28** |
| 559 | 487.24 | 406.51 | 437.54 | 448.99 | **1,780.28** |
| 560 | 157.37 | 119.28 | 148.19 | 138.69 | **563.52** |
| 561 | 10.84 | 19.44 | 15.42 | 15.69 | **61.39** |
| 562 | 43.85 | 28.77 | 38.24 | 43.32 | **154.18** |
| 563 | 113.70 | 152.71 | 122.55 | 144.71 | **533.67** |
| 564 | 64.91 | 53.74 | 65.54 | 71.89 | **256.08** |
| 565 | 62.79 | 71.51 | 82.20 | 58.64 | **275.13** |
| 566 | 53.61 | 38.16 | 41.07 | 34.96 | **167.80** |
| 567 | 20.99 | 14.39 | 12.64 | 14.63 | **62.66** |
| **MN - MINNESOTA - Total** | **4,809.99** | **4,839.14** | **4,746.60** | **4,670.59** | **19,066.30** |

**STATE: MO - MISSOURI**

| REGISTRANT ZIP CODE 3 | 1ST QUARTER | 2ND QUARTER | 3RD QUARTER | 4TH QUARTER | **TOTAL GRAMS** |
|---|---|---|---|---|---|
| 630 | 483.90 | 324.52 | 354.35 | 370.93 | **1,533.70** |
| 631 | 729.22 | 898.68 | 738.33 | 986.93 | **3,353.16** |
| 633 | 238.66 | 198.00 | 184.11 | 159.94 | **780.71** |
| 634 | 25.86 | 18.88 | 21.32 | 20.48 | **86.55** |
| 635 | 35.03 | 35.31 | 32.74 | 30.78 | **133.87** |
| 636 | 121.38 | 74.33 | 102.53 | 69.32 | **367.56** |
| 637 | 55.18 | 49.15 | 47.88 | 68.57 | **220.78** |
| 638 | 51.42 | 54.18 | 40.67 | 44.82 | **191.10** |
| 639 | 28.66 | 25.71 | 34.37 | 42.57 | **131.32** |
| 640 | 304.13 | 311.39 | 294.14 | 281.22 | **1,190.88** |
| 641 | 323.18 | 388.36 | 291.89 | 308.12 | **1,311.55** |
| 644 | 26.90 | 26.29 | 33.16 | 30.54 | **116.89** |
| 645 | 108.32 | 97.19 | 101.70 | 82.01 | **389.22** |
| 646 | 26.15 | 32.29 | 24.26 | 37.54 | **120.24** |
| 647 | 44.91 | 42.03 | 41.25 | 64.80 | **192.98** |
| 648 | 126.95 | 92.18 | 101.41 | 86.33 | **406.86** |
| 650 | 44.64 | 53.15 | 49.82 | 53.24 | **200.85** |
| 651 | 32.40 | 24.59 | 46.73 | 27.34 | **131.07** |
| 652 | 337.63 | 235.37 | 205.58 | 225.27 | **1,003.85** |
| 653 | 35.29 | 28.24 | 33.87 | 32.63 | **130.02** |
| 654 | 18.46 | 26.47 | 34.07 | 31.87 | **110.87** |
| 655 | 53.21 | 54.99 | 50.04 | 62.28 | **220.51** |
| 656 | 181.18 | 204.31 | 187.12 | 185.06 | **757.67** |
| 657 | 221.83 | 237.70 | 222.17 | 219.55 | **901.25** |
| 658 | 246.18 | 325.83 | 230.97 | 218.83 | **1,021.82** |
| **MO - MISSOURI - Total** | **3,900.69** | **3,859.12** | **3,504.50** | **3,740.98** | **15,005.29** |

**STATE: MS - MISSISSIPPI**

| REGISTRANT ZIP CODE 3 | 1ST QUARTER | 2ND QUARTER | 3RD QUARTER | 4TH QUARTER | TOTAL GRAMS |
|---|---|---|---|---|---|
| 386 | 99.03 | 105.99 | 87.44 | 88.66 | **381.12** |
| 387 | 14.63 | 8.71 | 10.96 | 16.34 | **50.64** |
| 388 | 82.02 | 92.18 | 38.85 | 45.92 | **258.97** |
| 389 | 19.81 | 16.15 | 17.01 | 17.33 | **70.30** |
| 390 | 54.27 | 51.39 | 42.04 | 55.58 | **203.28** |
| 391 | 2,930.23 | 4,600.67 | 4,631.12 | 4,592.95 | **16,754.97** |
| 392 | 183.22 | 252.68 | 182.82 | 198.29 | **817.01** |
| 393 | 52.73 | 53.93 | 51.14 | 46.44 | **204.23** |
| 394 | 211.69 | 201.69 | 205.48 | 229.49 | **848.36** |
| 395 | 318.78 | 348.18 | 427.80 | 408.76 | **1,503.52** |
| 396 | 19.63 | 19.56 | 46.18 | 21.23 | **106.60** |
| 397 | 25.08 | 32.00 | 31.31 | 32.14 | **120.52** |
| **MS - MISSISSIPPI - Total** | **4,011.13** | **5,783.13** | **5,772.14** | **5,753.14** | **21,319.53** |

**STATE: MT - MONTANA**

| REGISTRANT ZIP CODE 3 | 1ST QUARTER | 2ND QUARTER | 3RD QUARTER | 4TH QUARTER | TOTAL GRAMS |
|---|---|---|---|---|---|
| 590 | 96.36 | 104.18 | 101.72 | 107.37 | **409.62** |
| 591 | 446.07 | 390.49 | 425.21 | 406.71 | **1,668.47** |
| 592 | 25.45 | 22.17 | 24.54 | 17.36 | **89.52** |
| 593 | 54.06 | 56.45 | 52.44 | 40.51 | **203.47** |
| 594 | 79.67 | 89.47 | 95.90 | 86.70 | **351.74** |
| 595 | 9.31 | 11.38 | 6.24 | 8.45 | **35.38** |
| 596 | 59.49 | 67.49 | 82.63 | 100.42 | **310.03** |
| 597 | 188.21 | 240.34 | 203.76 | 181.69 | **813.99** |
| 598 | 332.59 | 272.62 | 279.21 | 252.48 | **1,136.91** |
| 599 | 145.00 | 164.62 | 178.15 | 143.82 | **631.59** |
| **MT - MONTANA - Total** | **1,436.22** | **1,419.21** | **1,449.80** | **1,345.51** | **5,650.73** |

**STATE: NC - NORTH CAROLINA**

| REGISTRANT ZIP CODE 3 | 1ST QUARTER | 2ND QUARTER | 3RD QUARTER | 4TH QUARTER | TOTAL GRAMS |
|---|---|---|---|---|---|
| 270 | 190.83 | 197.27 | 160.64 | 175.58 | **724.31** |
| 271 | 371.46 | 340.15 | 419.30 | 337.87 | **1,468.77** |
| 272 | 566.99 | 489.29 | 522.37 | 453.02 | **2,031.68** |
| 273 | 279.18 | 314.56 | 291.68 | 277.88 | **1,163.31** |
| 274 | 253.10 | 277.13 | 265.50 | 224.63 | **1,020.35** |
| 275 | 1,291.60 | 1,133.45 | 1,268.76 | 1,140.81 | **4,834.60** |
| 276 | 423.52 | 438.88 | 369.57 | 363.10 | **1,595.08** |
| 277 | 754.49 | 579.45 | 416.18 | 399.07 | **2,149.19** |
| 278 | 352.66 | 300.28 | 349.65 | 311.29 | **1,313.87** |
| 279 | 206.21 | 171.21 | 192.01 | 194.26 | **763.69** |
| 280 | 587.47 | 613.31 | 662.21 | 591.45 | **2,454.43** |
| 281 | 463.98 | 465.46 | 437.57 | 451.86 | **1,818.86** |
| 282 | 702.13 | 689.89 | 770.49 | 688.94 | **2,851.44** |
| 283 | 730.32 | 684.77 | 670.82 | 631.06 | **2,716.98** |
| 284 | 455.83 | 445.95 | 459.69 | 472.06 | **1,833.52** |
| 285 | 426.85 | 400.14 | 385.35 | 374.33 | **1,586.68** |
| 286 | 729.02 | 660.18 | 612.53 | 631.22 | **2,632.95** |
| 287 | 343.51 | 330.20 | 380.61 | 352.97 | **1,407.29** |
| 288 | 287.24 | 229.80 | 220.92 | 238.77 | **976.74** |
| 289 | 36.95 | 34.62 | 36.82 | 25.42 | **133.81** |
| **NC - NORTH CAROLINA - Total** | **9,453.32** | **8,795.99** | **8,892.66** | **8,335.58** | **35,477.56** |

**STATE: ND - NORTH DAKOTA**

| REGISTRANT ZIP CODE 3 | 1ST QUARTER | 2ND QUARTER | 3RD QUARTER | 4TH QUARTER | TOTAL GRAMS |
|---|---|---|---|---|---|
| 580 | 27.70 | 32.79 | 36.37 | 26.62 | **123.47** |
| 581 | 180.84 | 123.51 | 133.49 | 119.24 | **557.08** |
| 582 | 71.81 | 50.81 | 28.58 | 30.59 | **181.80** |
| 583 | 12.36 | 14.31 | 12.40 | 14.02 | **53.10** |
| 584 | 14.36 | 15.26 | 22.23 | 15.87 | **67.72** |
| 585 | 145.43 | 127.52 | 116.97 | 120.90 | **510.82** |
| 586 | 20.48 | 22.25 | 14.06 | 20.67 | **77.47** |
| 587 | 86.28 | 38.58 | 33.71 | 36.95 | **195.52** |

| | | | | | |
|---|---|---|---|---|---|
| 588 | 28.72 | 35.46 | 33.56 | 31.73 | **129.48** |
| **ND - NORTH DAKOTA - Total** | **587.99** | **460.48** | **431.37** | **416.61** | **1,896.45** |

**STATE: NE - NEBRASKA**

| REGISTRANT ZIP CODE 3 | 1ST QUARTER | 2ND QUARTER | 3RD QUARTER | 4TH QUARTER | TOTAL GRAMS |
|---|---|---|---|---|---|
| 680 | 166.94 | 168.53 | 184.91 | 183.33 | **703.71** |
| 681 | 390.93 | 414.15 | 393.32 | 450.26 | **1,648.65** |
| 683 | 10.57 | 9.97 | 15.39 | 12.90 | **48.82** |
| 684 | 20.32 | 21.40 | 16.76 | 16.13 | **74.61** |
| 685 | 102.61 | 152.34 | 104.86 | 148.00 | **507.81** |
| 686 | 13.73 | 13.85 | 13.07 | 16.11 | **56.77** |
| 687 | 31.03 | 27.66 | 28.66 | 19.83 | **107.18** |
| 688 | 93.96 | 80.54 | 80.57 | 90.19 | **345.25** |
| 689 | 20.23 | 24.71 | 23.96 | 25.16 | **94.06** |
| 690 | 8.51 | 11.25 | 7.38 | 4.61 | **31.75** |
| 691 | 21.62 | 24.77 | 23.20 | 26.93 | **96.52** |
| 692 | 4.95 | 4.52 | 7.04 | 3.72 | **20.23** |
| 693 | 28.71 | 23.11 | 35.00 | 22.60 | **109.42** |
| **NE - NEBRASKA - Total** | **914.10** | **976.79** | **934.11** | **1,019.78** | **3,844.78** |

**STATE: NH - NEW HAMPSHIRE**

| REGISTRANT ZIP CODE 3 | 1ST QUARTER | 2ND QUARTER | 3RD QUARTER | 4TH QUARTER | TOTAL GRAMS |
|---|---|---|---|---|---|
| 030 | 319.53 | 255.61 | 301.53 | 294.67 | **1,171.34** |
| 031 | 224.65 | 190.73 | 210.56 | 193.64 | **819.57** |
| 032 | 177.80 | 141.58 | 166.46 | 165.35 | **651.20** |
| 033 | 196.07 | 141.49 | 113.36 | 133.95 | **584.88** |
| 034 | 60.77 | 50.04 | 63.73 | 50.64 | **225.17** |
| 035 | 99.58 | 75.69 | 93.39 | 88.46 | **357.13** |
| 036 | 12.59 | 5.14 | 8.16 | 1.77 | **27.66** |
| 037 | 213.32 | 198.15 | 212.16 | 219.55 | **843.18** |
| 038 | 305.89 | 277.89 | 270.28 | 267.25 | **1,121.31** |
| **NH - NEW HAMPSHIRE - Total** | **1,610.20** | **1,336.32** | **1,439.63** | **1,415.28** | **5,801.43** |

**STATE: NJ - NEW JERSEY**

| REGISTRANT ZIP CODE 3 | 1ST QUARTER | 2ND QUARTER | 3RD QUARTER | 4TH QUARTER | TOTAL GRAMS |
|---|---|---|---|---|---|
| 070 | 1,078.07 | 1,014.38 | 890.44 | 936.42 | **3,919.31** |
| 071 | 171.46 | 261.90 | 174.33 | 153.25 | **760.94** |
| 072 | 76.67 | 70.93 | 94.53 | 79.75 | **321.88** |
| 073 | 47.22 | 36.81 | 44.08 | 46.20 | **174.31** |
| 074 | 897.78 | 897.99 | 651.53 | 777.01 | **3,224.32** |
| 075 | 160.54 | 101.69 | 116.68 | 119.19 | **498.10** |
| 076 | 868.82 | 727.54 | 712.56 | 714.25 | **3,023.17** |
| 077 | 529.14 | 677.58 | 534.95 | 491.80 | **2,233.46** |
| 078 | 316.97 | 321.80 | 300.52 | 305.96 | **1,245.24** |
| 079 | 340.96 | 550.31 | 289.80 | 323.97 | **1,505.04** |
| 080 | 826.13 | 926.77 | 858.26 | 818.85 | **3,430.02** |
| 081 | 116.54 | 256.06 | 165.65 | 157.85 | **696.09** |
| 082 | 134.10 | 130.80 | 143.72 | 136.34 | **544.97** |
| 083 | 96.29 | 107.91 | 100.82 | 91.45 | **396.48** |
| 084 | 17.36 | 29.32 | 18.76 | 25.07 | **90.52** |
| 085 | 157.16 | 162.66 | 186.09 | 133.70 | **639.61** |
| 086 | 97.33 | 106.31 | 92.36 | 96.02 | **392.02** |
| 087 | 338.40 | 373.70 | 307.75 | 330.44 | **1,350.30** |
| 088 | 762.82 | 665.19 | 705.42 | 711.09 | **2,844.52** |
| 089 | 104.53 | 149.06 | 99.94 | 111.97 | **465.50** |
| **NJ - NEW JERSEY - Total** | **7,138.29** | **7,568.71** | **6,488.22** | **6,560.59** | **27,755.80** |

**STATE: NM - NEW MEXICO**

| REGISTRANT ZIP CODE 3 | 1ST QUARTER | 2ND QUARTER | 3RD QUARTER | 4TH QUARTER | TOTAL GRAMS |
|---|---|---|---|---|---|
| 870 | 72.12 | 73.67 | 48.69 | 61.64 | **256.12** |
| 871 | 687.90 | 427.77 | 455.95 | 491.97 | **2,063.59** |
| 873 | 19.24 | 10.73 | 12.70 | 18.59 | **61.26** |
| 874 | 92.68 | 40.36 | 42.96 | 65.11 | **241.11** |
| 875 | 143.72 | 178.77 | 181.75 | 156.44 | **660.69** |

| | | | | | |
|---|---|---|---|---|---|
| 877 | 27.81 | 30.45 | 25.17 | 27.30 | **110.73** |
| 878 | 3.91 | 1.06 | 1.11 | 2.48 | **8.56** |
| 879 | 4.43 | 3.35 | 0.98 | 3.99 | **12.74** |
| 880 | 91.25 | 96.55 | 80.05 | 79.08 | **346.93** |
| 881 | 24.81 | 18.17 | 38.81 | 25.41 | **107.20** |
| 882 | 82.71 | 80.72 | 63.93 | 95.49 | **322.84** |
| 883 | 33.69 | 27.86 | 25.96 | 27.54 | **115.06** |
| 884 | 0.48 | 0.81 | 2.24 | 0.62 | **4.14** |
| **NM - NEW MEXICO - Total** | **1,284.76** | **990.26** | **980.30** | **1,055.65** | **4,310.97** |

**STATE: NV - NEVADA**

| REGISTRANT ZIP CODE 3 | 1ST QUARTER | 2ND QUARTER | 3RD QUARTER | 4TH QUARTER | TOTAL GRAMS |
|---|---|---|---|---|---|
| 890 | 452.19 | 515.16 | 455.92 | 483.27 | **1,906.53** |
| 891 | 1,311.35 | 1,238.96 | 1,335.43 | 1,244.99 | **5,130.74** |
| 893 | 1.20 | 1.53 | 1.68 | 1.91 | **6.32** |
| 894 | 204.04 | 223.23 | 185.52 | 175.60 | **788.40** |
| 895 | 257.57 | 332.02 | 274.88 | 294.67 | **1,159.15** |
| 897 | 68.89 | 78.77 | 79.42 | 71.37 | **298.44** |
| 898 | 19.02 | 24.00 | 23.77 | 21.95 | **88.74** |
| **NV - NEVADA - Total** | **2,314.26** | **2,413.68** | **2,356.63** | **2,293.76** | **9,378.32** |

**STATE: NY - NEW YORK**

| REGISTRANT ZIP CODE 3 | 1ST QUARTER | 2ND QUARTER | 3RD QUARTER | 4TH QUARTER | TOTAL GRAMS |
|---|---|---|---|---|---|
| 100 | 3,040.74 | 5,092.66 | 2,087.05 | 2,109.03 | **12,329.48** |
| 101 | 26.69 | 32.27 | 32.38 | 15.78 | **107.12** |
| 102 | 2.66 | 3.37 | 2.48 | 5.32 | **13.83** |
| 103 | 389.45 | 330.02 | 283.77 | 480.41 | **1,483.65** |
| 104 | 700.41 | 1,493.05 | 484.55 | 666.11 | **3,344.12** |
| 105 | 465.94 | 534.48 | 365.08 | 393.46 | **1,758.96** |
| 106 | 90.24 | 114.73 | 64.85 | 79.20 | **349.01** |
| 107 | 95.26 | 86.55 | 79.55 | 88.78 | **350.13** |
| 108 | 42.83 | 46.75 | 37.33 | 54.92 | **181.84** |
| 109 | 441.29 | 553.92 | 393.27 | 353.95 | **1,742.43** |
| 110 | 1,000.73 | 1,096.49 | 720.37 | 570.61 | **3,388.20** |
| 111 | 105.46 | 128.25 | 92.80 | 98.24 | **424.74** |
| 112 | 1,039.02 | 1,290.23 | 823.48 | 898.93 | **4,051.65** |
| 113 | 539.57 | 614.02 | 357.30 | 423.99 | **1,934.88** |
| 114 | 203.36 | 309.60 | 235.72 | 198.74 | **947.41** |
| 115 | 721.94 | 834.51 | 670.39 | 714.89 | **2,941.73** |
| 116 | 30.92 | 46.28 | 28.73 | 36.54 | **142.48** |
| 117 | 1,871.24 | 1,986.43 | 1,586.44 | 1,665.77 | **7,109.87** |
| 118 | 188.36 | 116.45 | 183.34 | 185.38 | **673.53** |
| 119 | 168.52 | 215.71 | 142.61 | 138.80 | **665.63** |
| 120 | 225.12 | 308.31 | 260.29 | 244.60 | **1,038.32** |
| 121 | 89.83 | 106.32 | 105.49 | 126.72 | **428.36** |
| 122 | 198.14 | 155.81 | 125.90 | 167.53 | **647.38** |
| 123 | 78.90 | 68.27 | 47.81 | 49.85 | **244.83** |
| 124 | 137.78 | 140.36 | 144.46 | 152.26 | **574.87** |
| 125 | 360.34 | 340.25 | 284.88 | 282.08 | **1,267.55** |
| 126 | 132.60 | 106.79 | 94.74 | 96.98 | **431.10** |
| 127 | 84.64 | 78.34 | 47.76 | 40.13 | **250.87** |
| 128 | 169.67 | 194.33 | 182.72 | 164.05 | **710.77** |
| 129 | 262.52 | 246.46 | 257.11 | 266.76 | **1,032.84** |
| 130 | 171.40 | 150.24 | 171.27 | 154.08 | **646.99** |
| 131 | 81.31 | 74.47 | 83.50 | 62.24 | **301.53** |
| 132 | 106.37 | 106.65 | 108.17 | 129.47 | **450.67** |
| 133 | 61.46 | 46.89 | 53.94 | 55.25 | **217.54** |
| 134 | 68.45 | 47.84 | 51.52 | 42.73 | **210.54** |
| 135 | 29.04 | 36.39 | 29.87 | 29.13 | **124.43** |
| 136 | 132.58 | 135.77 | 126.19 | 121.30 | **515.84** |
| 137 | 46.91 | 48.55 | 41.53 | 42.57 | **179.56** |
| 138 | 78.99 | 105.70 | 71.18 | 73.06 | **328.94** |

| | | | | | |
|---|---|---|---|---|---|
| 139 | 46.59 | 21.46 | 18.70 | 20.47 | **107.21** |
| 140 | 210.98 | 221.28 | 200.66 | 168.79 | **801.71** |
| 141 | 136.09 | 136.07 | 119.99 | 112.84 | **504.99** |
| 142 | 535.60 | 457.65 | 476.26 | 466.53 | **1,936.03** |
| 143 | 47.97 | 50.18 | 51.59 | 40.68 | **190.42** |
| 144 | 287.62 | 303.39 | 251.28 | 248.16 | **1,090.46** |
| 145 | 211.14 | 221.92 | 218.66 | 175.06 | **826.79** |
| 146 | 939.53 | 1,042.82 | 924.43 | 884.75 | **3,791.54** |
| 147 | 54.28 | 47.54 | 53.21 | 57.97 | **213.00** |
| 148 | 215.40 | 179.84 | 176.02 | 204.47 | **775.74** |
| 149 | 53.74 | 47.26 | 40.21 | 40.97 | **182.17** |
| **NY - NEW YORK - Total** | **16,419.63** | **20,152.93** | **13,490.84** | **13,900.32** | **63,963.71** |

**STATE: OH - OHIO**

| REGISTRANT ZIP CODE 3 | 1ST QUARTER | 2ND QUARTER | 3RD QUARTER | 4TH QUARTER | TOTAL GRAMS |
|---|---|---|---|---|---|
| 430 | 571.95 | 655.49 | 740.82 | 751.89 | **2,720.14** |
| 431 | 225.14 | 252.10 | 232.29 | 207.64 | **917.17** |
| 432 | 1,087.16 | 885.55 | 723.38 | 830.54 | **3,526.62** |
| 433 | 117.36 | 126.38 | 109.13 | 116.93 | **469.79** |
| 434 | 32.42 | 34.81 | 43.12 | 38.57 | **148.92** |
| 435 | 197.94 | 122.67 | 368.17 | 175.16 | **863.95** |
| 436 | 212.76 | 249.40 | 209.94 | 241.62 | **913.73** |
| 437 | 77.81 | 104.61 | 85.62 | 73.69 | **341.73** |
| 438 | 24.91 | 19.42 | 17.73 | 24.67 | **86.73** |
| 439 | 95.46 | 111.52 | 122.99 | 71.95 | **401.92** |
| 440 | 321.54 | 278.01 | 276.86 | 224.57 | **1,100.98** |
| 441 | 844.34 | 750.29 | 729.84 | 834.34 | **3,158.80** |
| 442 | 299.68 | 273.75 | 320.30 | 253.70 | **1,147.43** |
| 443 | 226.12 | 197.59 | 233.62 | 233.97 | **891.31** |
| 444 | 96.10 | 80.16 | 106.33 | 80.28 | **362.87** |
| 445 | 111.91 | 81.39 | 110.19 | 70.13 | **373.63** |
| 446 | 294.27 | 274.49 | 285.51 | 278.16 | **1,132.42** |
| 447 | 247.07 | 228.33 | 254.37 | 210.75 | **940.52** |
| 448 | 101.45 | 120.99 | 108.77 | 108.07 | **439.28** |
| 449 | 29.80 | 45.19 | 36.08 | 35.84 | **146.91** |
| 450 | 313.22 | 345.83 | 364.42 | 497.70 | **1,521.17** |
| 451 | 108.05 | 102.01 | 99.94 | 86.92 | **396.91** |
| 452 | 812.17 | 578.21 | 522.63 | 534.65 | **2,447.65** |
| 453 | 169.70 | 138.24 | 136.44 | 147.96 | **592.34** |
| 454 | 229.01 | 181.36 | 238.87 | 248.64 | **897.87** |
| 455 | 66.67 | 58.49 | 49.00 | 35.04 | **209.20** |
| 456 | 186.89 | 120.09 | 117.53 | 98.77 | **523.28** |
| 457 | 75.17 | 73.74 | 78.08 | 97.95 | **324.93** |
| 458 | 180.03 | 174.00 | 143.87 | 164.42 | **662.32** |
| **OH - OHIO - Total** | **7,356.09** | **6,664.11** | **6,865.84** | **6,774.50** | **27,660.54** |

**STATE: OK - OKLAHOMA**

| REGISTRANT ZIP CODE 3 | 1ST QUARTER | 2ND QUARTER | 3RD QUARTER | 4TH QUARTER | TOTAL GRAMS |
|---|---|---|---|---|---|
| 730 | 467.74 | 403.12 | 351.81 | 563.44 | **1,786.10** |
| 731 | 610.15 | 721.95 | 710.39 | 636.35 | **2,678.84** |
| 734 | 41.40 | 37.24 | 45.18 | 53.02 | **176.84** |
| 735 | 159.07 | 128.28 | 147.07 | 125.18 | **559.59** |
| 736 | 14.53 | 11.46 | 11.57 | 18.23 | **55.80** |
| 737 | 43.09 | 44.89 | 45.24 | 47.69 | **180.91** |
| 738 | 22.00 | 24.94 | 16.88 | 14.87 | **78.69** |
| 739 | 0.45 | 0.82 | 0.44 | 2.65 | **4.37** |
| 740 | 601.92 | 396.98 | 420.90 | 386.19 | **1,805.98** |
| 741 | 398.40 | 364.79 | 397.24 | 319.60 | **1,480.04** |
| 743 | 75.08 | 64.72 | 70.96 | 56.49 | **267.24** |
| 744 | 132.33 | 127.51 | 140.72 | 136.53 | **537.09** |
| 745 | 56.55 | 66.30 | 76.19 | 72.67 | **271.71** |
| 746 | 16.91 | 14.18 | 5.15 | 17.59 | **53.83** |

| | | | | | |
|---|---|---|---|---|---|
| 747 | 65.76 | 63.83 | 60.10 | 58.49 | **248.18** |
| 748 | 75.90 | 71.65 | 72.52 | 69.78 | **289.86** |
| 749 | 143.45 | 134.54 | 134.10 | 150.81 | **562.90** |
| **OK - OKLAHOMA - Total** | **2,924.73** | **2,677.19** | **2,706.47** | **2,729.58** | **11,037.97** |

**STATE: OR - OREGON**

| REGISTRANT ZIP CODE 3 | 1ST QUARTER | 2ND QUARTER | 3RD QUARTER | 4TH QUARTER | TOTAL GRAMS |
|---|---|---|---|---|---|
| 970 | 554.93 | 568.20 | 600.34 | 576.87 | **2,300.33** |
| 971 | 262.49 | 243.19 | 247.15 | 232.11 | **984.95** |
| 972 | 1,246.65 | 1,298.70 | 1,177.05 | 1,137.58 | **4,859.98** |
| 973 | 560.26 | 462.16 | 479.87 | 462.87 | **1,965.17** |
| 974 | 603.62 | 526.24 | 686.93 | 634.38 | **2,451.16** |
| 975 | 424.64 | 432.98 | 426.20 | 449.87 | **1,733.70** |
| 976 | 47.89 | 58.83 | 49.51 | 44.37 | **200.61** |
| 977 | 158.03 | 155.22 | 185.22 | 153.85 | **652.32** |
| 978 | 90.72 | 110.36 | 109.99 | 106.89 | **417.95** |
| 979 | 16.85 | 10.28 | 11.90 | 16.48 | **55.51** |
| **OR - OREGON - Total** | **3,966.07** | **3,866.17** | **3,974.15** | **3,815.28** | **15,621.67** |

**STATE: PA - PENNSYLVANIA**

| REGISTRANT ZIP CODE 3 | 1ST QUARTER | 2ND QUARTER | 3RD QUARTER | 4TH QUARTER | TOTAL GRAMS |
|---|---|---|---|---|---|
| 150 | 265.01 | 297.83 | 288.11 | 247.60 | **1,098.54** |
| 151 | 153.84 | 145.97 | 165.26 | 143.87 | **608.94** |
| 152 | 750.53 | 683.42 | 568.08 | 813.40 | **2,815.43** |
| 153 | 55.53 | 57.85 | 90.07 | 58.50 | **261.94** |
| 154 | 38.40 | 45.10 | 42.13 | 43.29 | **168.92** |
| 155 | 39.65 | 30.77 | 23.54 | 33.81 | **127.77** |
| 156 | 117.13 | 92.52 | 130.04 | 121.97 | **461.66** |
| 157 | 30.12 | 45.19 | 49.78 | 47.38 | **172.47** |
| 158 | 61.23 | 58.26 | 63.54 | 69.00 | **252.03** |
| 159 | 50.53 | 40.45 | 38.08 | 45.39 | **174.45** |
| 160 | 81.98 | 53.76 | 77.50 | 83.43 | **296.67** |
| 161 | 72.63 | 79.90 | 71.51 | 85.67 | **309.71** |
| 162 | 33.21 | 25.24 | 31.92 | 28.82 | **119.18** |
| 163 | 171.00 | 171.81 | 207.01 | 177.45 | **727.27** |
| 164 | 114.27 | 114.18 | 94.77 | 79.49 | **402.70** |
| 165 | 90.07 | 97.68 | 96.91 | 77.43 | **362.10** |
| 166 | 117.80 | 106.68 | 159.51 | 167.54 | **551.52** |
| 167 | 18.56 | 17.11 | 14.55 | 16.87 | **67.09** |
| 168 | 50.89 | 68.05 | 52.06 | 55.53 | **226.52** |
| 169 | 31.08 | 31.56 | 35.24 | 33.52 | **131.40** |
| 170 | 406.45 | 327.25 | 362.07 | 320.96 | **1,416.73** |
| 171 | 106.57 | 127.02 | 102.46 | 95.18 | **431.23** |
| 172 | 121.86 | 98.37 | 81.13 | 94.67 | **396.03** |
| 173 | 204.24 | 172.24 | 203.66 | 170.76 | **750.89** |
| 174 | 120.79 | 130.00 | 185.88 | 255.64 | **692.31** |
| 175 | 184.40 | 178.11 | 175.44 | 175.72 | **713.67** |
| 176 | 427.80 | 318.20 | 301.83 | 299.17 | **1,347.00** |
| 177 | 96.56 | 88.18 | 101.11 | 119.02 | **404.87** |
| 178 | 207.25 | 163.48 | 194.59 | 167.26 | **732.57** |
| 179 | 86.89 | 70.56 | 57.90 | 50.71 | **266.07** |
| 180 | 465.61 | 309.99 | 406.83 | 378.12 | **1,560.55** |
| 181 | 169.39 | 242.07 | 223.68 | 161.95 | **797.10** |
| 182 | 38.40 | 48.73 | 40.19 | 52.03 | **179.35** |
| 183 | 152.08 | 205.79 | 144.77 | 125.84 | **628.49** |
| 184 | 136.89 | 124.67 | 148.29 | 133.25 | **543.10** |
| 185 | 115.83 | 144.40 | 144.10 | 99.95 | **504.28** |
| 186 | 54.91 | 62.50 | 67.29 | 64.19 | **248.89** |
| 187 | 469.09 | 471.52 | 493.69 | 428.01 | **1,862.31** |
| 188 | 36.64 | 31.34 | 52.01 | 42.82 | **162.80** |
| 189 | 294.20 | 332.62 | 330.95 | 271.35 | **1,229.12** |
| 190 | 2,218.73 | 3,833.04 | 2,236.85 | 2,127.34 | **10,415.95** |

| | | | | | |
|---|---|---|---|---|---|
| 191 | 1,235.23 | 1,865.17 | 1,124.53 | 992.43 | **5,217.35** |
| 193 | 310.18 | 256.90 | 335.21 | 320.27 | **1,222.57** |
| 194 | 465.07 | 444.01 | 508.75 | 575.61 | **1,993.45** |
| 195 | 92.69 | 103.02 | 95.39 | 100.79 | **391.90** |
| 196 | 109.70 | 119.36 | 118.09 | 103.81 | **450.95** |
| **PA - PENNSYLVANIA - Total** | **10,670.90** | **12,531.85** | **10,536.30** | **10,156.82** | **43,895.86** |
| **STATE: PR - PUERTO RICO** | | | | | |
| REGISTRANT ZIP CODE 3 | 1ST QUARTER | 2ND QUARTER | 3RD QUARTER | 4TH QUARTER | **TOTAL GRAMS** |
| 006 | 9.04 | 12.41 | 3.72 | 5.32 | **30.50** |
| 007 | 3.19 | 4.96 | 4.61 | 5.14 | **17.91** |
| 009 | 35.07 | 24.12 | 17.29 | 22.52 | **98.99** |
| **PR - PUERTO RICO - Total** | **47.30** | **41.49** | **25.62** | **32.98** | **147.40** |
| **STATE: RI - RHODE ISLAND** | | | | | |
| REGISTRANT ZIP CODE 3 | 1ST QUARTER | 2ND QUARTER | 3RD QUARTER | 4TH QUARTER | **TOTAL GRAMS** |
| 028 | 717.18 | 562.12 | 621.85 | 423.68 | **2,324.82** |
| 029 | 271.42 | 326.16 | 254.86 | 383.72 | **1,236.17** |
| **RI - RHODE ISLAND - Total** | **988.61** | **888.28** | **876.71** | **807.39** | **3,560.99** |
| **STATE: SC - SOUTH CAROLINA** | | | | | |
| REGISTRANT ZIP CODE 3 | 1ST QUARTER | 2ND QUARTER | 3RD QUARTER | 4TH QUARTER | **TOTAL GRAMS** |
| 290 | 344.26 | 347.98 | 325.49 | 336.26 | **1,353.98** |
| 291 | 525.91 | 455.00 | 470.61 | 466.36 | **1,917.87** |
| 292 | 495.49 | 435.84 | 417.58 | 382.35 | **1,731.26** |
| 293 | 148.47 | 134.47 | 165.31 | 156.07 | **604.32** |
| 294 | 1,850.17 | 2,351.16 | 3,270.16 | 1,971.50 | **9,442.99** |
| 295 | 710.26 | 631.71 | 701.86 | 660.38 | **2,704.22** |
| 296 | 742.33 | 757.77 | 682.61 | 613.42 | **2,796.13** |
| 297 | 220.45 | 168.42 | 197.24 | 165.98 | **752.10** |
| 298 | 227.21 | 232.27 | 209.73 | 223.04 | **892.26** |
| 299 | 265.25 | 347.29 | 355.78 | 258.26 | **1,226.58** |
| **SC - SOUTH CAROLINA - Total** | **5,529.80** | **5,861.92** | **6,796.37** | **5,233.61** | **23,421.69** |
| **STATE: SD - SOUTH DAKOTA** | | | | | |
| REGISTRANT ZIP CODE 3 | 1ST QUARTER | 2ND QUARTER | 3RD QUARTER | 4TH QUARTER | **TOTAL GRAMS** |
| 570 | 81.41 | 75.85 | 93.49 | 74.43 | **325.19** |
| 571 | 199.30 | 269.95 | 224.06 | 172.65 | **865.95** |
| 572 | 20.31 | 20.10 | 19.92 | 22.71 | **83.04** |
| 573 | 21.70 | 24.10 | 20.30 | 17.77 | **83.88** |
| 574 | 11.49 | 11.98 | 8.85 | 12.55 | **44.86** |
| 575 | 7.62 | 11.57 | 8.12 | 10.17 | **37.48** |
| 576 | 2.48 | 2.16 | 1.63 | 2.53 | **8.80** |
| 577 | 61.44 | 68.09 | 68.62 | 81.71 | **279.86** |
| **SD - SOUTH DAKOTA - Total** | **405.75** | **483.80** | **444.99** | **394.52** | **1,729.06** |
| **STATE: TN - TENNESSEE** | | | | | |
| REGISTRANT ZIP CODE 3 | 1ST QUARTER | 2ND QUARTER | 3RD QUARTER | 4TH QUARTER | **TOTAL GRAMS** |
| 370 | 422.14 | 289.80 | 386.67 | 298.02 | **1,396.63** |
| 371 | 282.45 | 328.32 | 252.91 | 267.85 | **1,131.53** |
| 372 | 624.05 | 571.88 | 588.77 | 515.42 | **2,300.12** |
| 373 | 393.86 | 417.02 | 387.25 | 467.42 | **1,665.56** |
| 374 | 263.66 | 271.19 | 270.65 | 334.73 | **1,140.23** |
| 376 | 217.71 | 184.77 | 186.07 | 197.78 | **786.34** |
| 377 | 109.77 | 103.10 | 97.14 | 83.27 | **393.28** |
| 378 | 168.55 | 160.35 | 161.05 | 168.24 | **658.20** |
| 379 | 689.59 | 768.75 | 229.50 | 160.61 | **1,848.45** |
| 380 | 179.21 | 170.45 | 129.06 | 117.34 | **596.07** |
| 381 | 1,415.23 | 2,738.76 | 843.11 | 1,364.08 | **6,361.19** |
| 382 | 33.55 | 34.13 | 26.70 | 35.47 | **129.86** |
| 383 | 91.47 | 79.98 | 70.39 | 69.78 | **311.61** |
| 384 | 75.30 | 63.93 | 89.36 | 41.19 | **269.78** |
| 385 | 117.38 | 82.79 | 69.47 | 85.93 | **355.58** |
| **TN - TENNESSEE - Total** | **5,083.91** | **6,265.23** | **3,788.12** | **4,207.15** | **19,344.41** |
| **STATE: TX - TEXAS** | | | | | |

| REGISTRANT ZIP CODE 3 | 1ST QUARTER | 2ND QUARTER | 3RD QUARTER | 4TH QUARTER | TOTAL GRAMS |
|---|---|---|---|---|---|
| 750 | 1,551.21 | 1,477.27 | 1,549.29 | 1,445.78 | **6,023.55** |
| 751 | 339.78 | 347.20 | 344.05 | 334.53 | **1,365.56** |
| 752 | 3,120.91 | 1,894.98 | 3,606.84 | 3,004.81 | **11,627.54** |
| 753 | 93.18 | 84.21 | 91.91 | 96.17 | **365.47** |
| 754 | 344.22 | 325.45 | 342.91 | 316.78 | **1,329.36** |
| 755 | 85.27 | 135.59 | 70.51 | 63.68 | **355.06** |
| 756 | 798.56 | 686.43 | 687.71 | 694.22 | **2,866.91** |
| 757 | 223.70 | 211.16 | 238.59 | 212.15 | **885.59** |
| 758 | 36.44 | 44.06 | 33.18 | 37.39 | **151.07** |
| 759 | 104.00 | 89.62 | 112.13 | 100.08 | **405.84** |
| 760 | 767.03 | 720.70 | 640.06 | 709.80 | **2,837.58** |
| 761 | 809.35 | 952.63 | 757.77 | 706.14 | **3,225.89** |
| 762 | 599.63 | 479.60 | 501.39 | 484.23 | **2,064.85** |
| 763 | 202.69 | 152.52 | 176.82 | 125.11 | **657.14** |
| 764 | 77.09 | 67.44 | 68.22 | 63.36 | **276.10** |
| 765 | 352.24 | 376.41 | 302.27 | 270.86 | **1,301.77** |
| 766 | 48.09 | 44.28 | 42.40 | 56.26 | **191.04** |
| 767 | 85.81 | 105.85 | 90.50 | 79.53 | **361.69** |
| 768 | 143.63 | 138.42 | 309.42 | 129.74 | **721.20** |
| 769 | 23.91 | 20.21 | 22.26 | 26.10 | **92.48** |
| 770 | 1,728.02 | 1,721.84 | 1,784.36 | 1,564.94 | **6,799.16** |
| 773 | 752.61 | 760.26 | 687.88 | 731.34 | **2,932.08** |
| 774 | 502.49 | 493.28 | 617.73 | 510.47 | **2,123.97** |
| 775 | 654.24 | 637.00 | 615.24 | 722.72 | **2,629.21** |
| 776 | 326.77 | 298.56 | 296.76 | 312.64 | **1,234.72** |
| 777 | 340.08 | 336.50 | 327.10 | 334.75 | **1,338.43** |
| 778 | 111.41 | 99.77 | 121.30 | 115.16 | **447.65** |
| 779 | 269.33 | 244.13 | 236.60 | 230.55 | **980.61** |
| 780 | 191.51 | 204.85 | 181.71 | 203.50 | **781.57** |
| 781 | 222.77 | 236.97 | 199.54 | 219.28 | **878.55** |
| 782 | 1,419.28 | 1,419.30 | 1,136.63 | 1,062.95 | **5,038.17** |
| 783 | 73.05 | 98.29 | 81.32 | 76.31 | **328.97** |
| 784 | 197.81 | 167.98 | 208.14 | 167.87 | **741.80** |
| 785 | 260.74 | 258.62 | 306.21 | 283.24 | **1,108.81** |
| 786 | 568.88 | 521.50 | 570.46 | 566.75 | **2,227.58** |
| 787 | 555.23 | 882.20 | 564.23 | 739.67 | **2,741.34** |
| 788 | 27.61 | 29.65 | 28.12 | 23.69 | **109.08** |
| 789 | 23.08 | 19.51 | 24.14 | 25.82 | **92.55** |
| 790 | 50.28 | 51.02 | 41.46 | 34.89 | **177.65** |
| 791 | 78.89 | 120.79 | 92.16 | 65.80 | **357.64** |
| 792 | 8.10 | 11.61 | 6.89 | 9.53 | **36.13** |
| 793 | 63.94 | 60.00 | 60.02 | 60.28 | **244.23** |
| 794 | 336.50 | 354.47 | 350.96 | 313.91 | **1,355.84** |
| 795 | 29.39 | 28.64 | 29.48 | 28.84 | **116.34** |
| 796 | 63.82 | 57.18 | 64.60 | 63.84 | **249.44** |
| 797 | 127.72 | 97.25 | 123.99 | 118.65 | **467.61** |
| 798 | 2.88 | 1.37 | 6.16 | 5.28 | **15.70** |
| 799 | 338.87 | 311.73 | 319.74 | 294.17 | **1,264.51** |
| **TX - TEXAS - Total** | **19,132.03** | **17,878.33** | **19,071.14** | **17,843.57** | **73,925.07** |

**STATE: UT - UTAH**

| REGISTRANT ZIP CODE 3 | 1ST QUARTER | 2ND QUARTER | 3RD QUARTER | 4TH QUARTER | TOTAL GRAMS |
|---|---|---|---|---|---|
| 840 | 816.15 | 842.41 | 893.45 | 923.07 | **3,475.07** |
| 841 | 1,085.60 | 821.74 | 716.40 | 582.21 | **3,205.95** |
| 843 | 131.23 | 111.90 | 125.62 | 135.70 | **504.45** |
| 844 | 227.65 | 174.81 | 173.33 | 178.06 | **753.86** |
| 845 | 26.50 | 16.62 | 18.13 | 20.72 | **81.97** |
| 846 | 186.92 | 209.24 | 197.88 | 205.72 | **799.76** |
| 847 | 263.11 | 243.10 | 235.87 | 284.69 | **1,026.77** |
| **UT - UTAH - Total** | **2,737.14** | **2,419.82** | **2,360.69** | **2,330.18** | **9,847.83** |

**STATE: VA - VIRGINIA**

| REGISTRANT ZIP CODE 3 | 1ST QUARTER | 2ND QUARTER | 3RD QUARTER | 4TH QUARTER | TOTAL GRAMS |
|---|---|---|---|---|---|
| 201 | 737.74 | 675.53 | 731.34 | 697.35 | **2,841.96** |
| 220 | 447.38 | 397.01 | 429.46 | 391.32 | **1,665.17** |
| 221 | 292.24 | 291.89 | 245.10 | 235.57 | **1,064.80** |
| 222 | 155.16 | 146.86 | 125.75 | 136.47 | **564.24** |
| 223 | 170.72 | 177.62 | 133.45 | 145.58 | **627.37** |
| 224 | 258.25 | 245.19 | 252.13 | 266.28 | **1,021.85** |
| 225 | 83.46 | 83.92 | 98.44 | 104.37 | **370.18** |
| 226 | 352.12 | 286.88 | 282.34 | 265.29 | **1,186.63** |
| 227 | 54.24 | 67.22 | 65.54 | 92.33 | **279.32** |
| 228 | 238.50 | 237.21 | 209.88 | 215.28 | **900.88** |
| 229 | 521.47 | 479.89 | 488.93 | 498.40 | **1,988.69** |
| 230 | 186.22 | 189.24 | 179.23 | 163.92 | **718.61** |
| 231 | 394.22 | 437.97 | 448.37 | 411.34 | **1,691.89** |
| 232 | 942.76 | 732.36 | 806.58 | 916.35 | **3,398.06** |
| 233 | 419.56 | 387.35 | 417.40 | 420.15 | **1,644.46** |
| 234 | 661.41 | 680.79 | 637.41 | 616.02 | **2,595.63** |
| 235 | 297.08 | 235.20 | 274.82 | 270.74 | **1,077.84** |
| 236 | 356.87 | 331.61 | 355.58 | 341.91 | **1,385.97** |
| 237 | 145.56 | 107.86 | 132.95 | 102.91 | **489.28** |
| 238 | 184.44 | 187.85 | 202.89 | 203.55 | **778.73** |
| 239 | 49.91 | 46.10 | 45.52 | 53.12 | **194.64** |
| 240 | 403.53 | 378.31 | 353.29 | 381.13 | **1,516.26** |
| 241 | 270.61 | 261.58 | 279.28 | 267.47 | **1,078.94** |
| 242 | 167.68 | 119.55 | 109.64 | 89.95 | **486.82** |
| 243 | 88.73 | 89.12 | 91.20 | 87.76 | **356.82** |
| 244 | 150.69 | 138.13 | 135.52 | 135.54 | **559.88** |
| 245 | 298.28 | 235.66 | 244.13 | 244.86 | **1,022.93** |
| 246 | 31.62 | 31.74 | 44.05 | 27.42 | **134.83** |
| **VA - VIRGINIA - Total** | **8,360.44** | **7,679.64** | **7,820.22** | **7,782.38** | **31,642.67** |

**STATE: VI - VIRGIN ISLANDS**

| REGISTRANT ZIP CODE 3 | 1ST QUARTER | 2ND QUARTER | 3RD QUARTER | 4TH QUARTER | TOTAL GRAMS |
|---|---|---|---|---|---|
| 008 | 17.42 | 10.52 | 18.08 | 16.98 | **62.99** |
| **VI - VIRGIN ISLANDS - Total** | **17.42** | **10.52** | **18.08** | **16.98** | **62.99** |

**STATE: VT - VERMONT**

| REGISTRANT ZIP CODE 3 | 1ST QUARTER | 2ND QUARTER | 3RD QUARTER | 4TH QUARTER | TOTAL GRAMS |
|---|---|---|---|---|---|
| 050 | 49.06 | 48.21 | 51.76 | 43.18 | **192.21** |
| 051 | 37.11 | 27.98 | 34.28 | 32.30 | **131.67** |
| 052 | 29.57 | 30.30 | 29.99 | 36.65 | **126.51** |
| 053 | 19.16 | 21.16 | 22.19 | 18.25 | **80.76** |
| 054 | 590.94 | 511.62 | 521.71 | 473.49 | **2,097.75** |
| 056 | 123.81 | 118.06 | 106.83 | 97.39 | **446.09** |
| 057 | 113.83 | 103.14 | 108.29 | 114.42 | **439.68** |
| 058 | 83.19 | 67.52 | 55.20 | 56.10 | **262.01** |
| **VT - VERMONT - Total** | **1,046.68** | **927.99** | **930.24** | **871.77** | **3,776.68** |

**STATE: WA - WASHINGTON**

| REGISTRANT ZIP CODE 3 | 1ST QUARTER | 2ND QUARTER | 3RD QUARTER | 4TH QUARTER | TOTAL GRAMS |
|---|---|---|---|---|---|
| 980 | 1,419.66 | 1,396.68 | 1,458.31 | 1,402.57 | **5,677.22** |
| 981 | 1,261.28 | 1,145.90 | 1,231.72 | 1,273.97 | **4,912.87** |
| 982 | 902.42 | 973.58 | 1,091.23 | 1,043.39 | **4,010.62** |
| 983 | 768.22 | 761.08 | 854.58 | 740.19 | **3,124.07** |
| 984 | 488.42 | 437.00 | 478.88 | 419.81 | **1,824.10** |
| 985 | 584.19 | 552.42 | 607.67 | 602.33 | **2,346.62** |
| 986 | 846.24 | 829.92 | 836.25 | 806.68 | **3,319.09** |
| 988 | 192.08 | 188.48 | 295.86 | 199.09 | **875.51** |
| 989 | 241.91 | 223.64 | 247.16 | 239.89 | **952.59** |
| 990 | 110.72 | 111.36 | 108.42 | 113.53 | **444.03** |
| 991 | 121.87 | 108.09 | 150.06 | 112.18 | **492.21** |
| 992 | 656.34 | 687.32 | 761.85 | 637.72 | **2,743.24** |

| | | | | | |
|---|---|---|---|---|---|
| 993 | 353.40 | 432.92 | 413.05 | 383.68 | **1,583.04** |
| 994 | 42.94 | 40.33 | 133.89 | 77.45 | **294.61** |
| **WA - WASHINGTON - Total** | **7,989.69** | **7,888.71** | **8,668.94** | **8,052.48** | **32,599.82** |

**STATE: WI - WISCONSIN**

| REGISTRANT ZIP CODE 3 | 1ST QUARTER | 2ND QUARTER | 3RD QUARTER | 4TH QUARTER | TOTAL GRAMS |
|---|---|---|---|---|---|
| 530 | 299.19 | 299.95 | 234.56 | 224.71 | **1,058.41** |
| 531 | 549.88 | 343.85 | 340.93 | 318.33 | **1,553.00** |
| 532 | 854.65 | 549.87 | 660.91 | 596.01 | **2,661.45** |
| 534 | 55.12 | 46.77 | 59.95 | 54.03 | **215.88** |
| 535 | 353.41 | 460.15 | 340.66 | 227.61 | **1,381.83** |
| 537 | 381.52 | 359.49 | 368.06 | 479.81 | **1,588.88** |
| 538 | 32.06 | 21.86 | 18.97 | 21.05 | **93.94** |
| 539 | 73.40 | 71.05 | 64.83 | 62.92 | **272.19** |
| 540 | 79.47 | 111.64 | 95.70 | 106.71 | **393.52** |
| 541 | 67.51 | 52.99 | 52.30 | 40.51 | **213.31** |
| 542 | 14.30 | 23.74 | 32.85 | 20.30 | **91.18** |
| 543 | 182.30 | 131.69 | 76.58 | 86.13 | **476.70** |
| 544 | 186.22 | 150.10 | 186.19 | 104.28 | **626.78** |
| 545 | 54.80 | 78.83 | 68.89 | 72.26 | **274.78** |
| 546 | 166.27 | 110.14 | 126.16 | 93.31 | **495.88** |
| 547 | 123.81 | 76.39 | 80.71 | 82.90 | **363.81** |
| 548 | 114.56 | 157.90 | 159.63 | 147.29 | **579.38** |
| 549 | 320.71 | 297.95 | 330.67 | 183.92 | **1,133.26** |
| **WI - WISCONSIN - Total** | **3,909.17** | **3,344.37** | **3,298.57** | **2,922.07** | **13,474.18** |

**STATE: WV - WEST VIRGINIA**

| REGISTRANT ZIP CODE 3 | 1ST QUARTER | 2ND QUARTER | 3RD QUARTER | 4TH QUARTER | TOTAL GRAMS |
|---|---|---|---|---|---|
| 247 | 29.16 | 36.32 | 42.72 | 41.40 | **149.61** |
| 248 | 7.27 | 6.92 | 6.38 | 6.92 | **27.48** |
| 249 | 13.37 | 11.48 | 14.72 | 15.96 | **55.53** |
| 250 | 8.51 | 7.98 | 8.69 | 4.61 | **29.79** |
| 251 | 18.97 | 9.35 | 6.91 | 5.47 | **40.71** |
| 252 | 6.66 | 7.38 | 8.82 | 2.10 | **24.96** |
| 253 | 120.51 | 91.20 | 123.53 | 89.21 | **424.45** |
| 254 | 225.17 | 191.44 | 161.36 | 135.23 | **713.20** |
| 255 | 36.40 | 39.43 | 38.37 | 32.55 | **146.75** |
| 256 | 4.43 | 2.53 | 2.95 | 1.95 | **11.86** |
| 257 | 79.56 | 70.51 | 62.93 | 91.51 | **304.51** |
| 258 | 107.98 | 116.04 | 91.24 | 83.84 | **399.10** |
| 259 | 6.74 | 3.95 | 7.54 | 7.10 | **25.32** |
| 260 | 39.80 | 59.81 | 48.39 | 45.33 | **193.33** |
| 261 | 22.77 | 22.41 | 27.47 | 22.64 | **95.28** |
| 262 | 10.99 | 15.73 | 16.17 | 20.26 | **63.15** |
| 263 | 46.53 | 47.60 | 47.28 | 49.03 | **190.44** |
| 264 | 13.44 | 13.65 | 13.64 | 15.13 | **55.86** |
| 265 | 165.77 | 133.29 | 194.44 | 179.79 | **673.29** |
| 266 | 2.74 | 0.70 | 0.38 | 6.40 | **10.23** |
| 267 | 31.59 | 37.52 | 39.18 | 47.42 | **155.72** |
| 268 | 24.58 | 22.40 | 29.13 | 28.99 | **105.09** |
| **WV - WEST VIRGINIA - Total** | **1,022.95** | **947.63** | **992.25** | **932.82** | **3,895.65** |

**STATE: WY - WYOMING**

| REGISTRANT ZIP CODE 3 | 1ST QUARTER | 2ND QUARTER | 3RD QUARTER | 4TH QUARTER | TOTAL GRAMS |
|---|---|---|---|---|---|
| 820 | 83.57 | 105.56 | 121.97 | 119.73 | **430.83** |
| 822 | 7.40 | 2.15 | 1.75 | 3.22 | **14.52** |
| 823 | 3.81 | 5.16 | 3.28 | 5.28 | **17.53** |
| 824 | 106.13 | 138.20 | 118.79 | 116.88 | **480.00** |
| 825 | 18.75 | 13.70 | 13.45 | 10.33 | **56.23** |
| 826 | 106.18 | 109.71 | 103.20 | 84.29 | **403.39** |
| 827 | 34.44 | 57.84 | 39.88 | 39.09 | **171.25** |
| 828 | 52.41 | 58.76 | 47.96 | 45.91 | **205.04** |
| 829 | 69.41 | 79.81 | 75.21 | 71.99 | **296.42** |

| | | | | | |
|---|---|---|---|---|---|
| 830 | 7.40 | 3.68 | 10.50 | 11.58 | **33.15** |
| 831 | 9.22 | 14.85 | 10.59 | 7.40 | **42.07** |
| **WY - WYOMING - Total** | **498.71** | **589.42** | **546.58** | **515.70** | **2,150.41** |

**DRUG: 9193 - HYDROCODONE**

**STATE: AK - ALASKA**

| REGISTRANT ZIP CODE 3 | 1ST QUARTER | 2ND QUARTER | 3RD QUARTER | 4TH QUARTER | TOTAL GRAMS |
|---|---|---|---|---|---|
| 995 | 3,430.10 | 3,158.16 | 3,313.77 | 3,279.83 | **13,181.85** |
| 996 | 2,627.47 | 2,605.60 | 2,819.00 | 2,751.62 | **10,803.69** |
| 997 | 729.57 | 645.57 | 845.72 | 730.97 | **2,951.83** |
| 998 | 331.08 | 296.91 | 336.21 | 306.11 | **1,270.32** |
| 999 | 313.38 | 304.06 | 320.14 | 335.42 | **1,273.01** |
| **AK - ALASKA - Total** | **7,431.60** | **7,010.30** | **7,634.84** | **7,403.95** | **29,480.70** |

**STATE: AL - ALABAMA**

| REGISTRANT ZIP CODE 3 | 1ST QUARTER | 2ND QUARTER | 3RD QUARTER | 4TH QUARTER | TOTAL GRAMS |
|---|---|---|---|---|---|
| 350 | 20,353.83 | 19,229.89 | 19,424.05 | 19,339.88 | **78,347.65** |
| 351 | 11,507.92 | 10,937.03 | 11,332.21 | 11,062.97 | **44,840.13** |
| 352 | 16,722.78 | 16,032.56 | 16,219.96 | 15,968.23 | **64,943.53** |
| 354 | 8,080.20 | 7,310.72 | 7,458.93 | 7,285.97 | **30,135.81** |
| 355 | 8,822.66 | 8,253.18 | 8,718.56 | 8,523.97 | **34,318.36** |
| 356 | 19,068.38 | 17,378.36 | 18,534.94 | 17,734.65 | **72,716.34** |
| 357 | 6,770.43 | 6,613.34 | 6,824.51 | 6,780.88 | **26,989.16** |
| 358 | 6,818.25 | 6,430.57 | 6,606.48 | 6,666.41 | **26,521.72** |
| 359 | 16,814.69 | 16,406.62 | 16,559.51 | 16,330.05 | **66,110.86** |
| 360 | 7,588.41 | 7,341.14 | 7,769.18 | 7,650.42 | **30,349.15** |
| 361 | 5,987.66 | 5,574.02 | 5,629.99 | 5,520.68 | **22,712.34** |
| 362 | 7,707.56 | 7,035.47 | 7,166.10 | 7,273.89 | **29,183.02** |
| 363 | 7,403.06 | 6,985.28 | 7,407.19 | 7,445.02 | **29,240.54** |
| 364 | 4,725.48 | 4,435.69 | 4,479.55 | 4,661.43 | **18,302.15** |
| 365 | 13,856.77 | 13,017.90 | 13,310.68 | 13,261.06 | **53,446.41** |
| 366 | 10,644.22 | 9,623.83 | 9,896.24 | 9,994.90 | **40,159.19** |
| 367 | 2,330.13 | 2,276.59 | 2,304.77 | 2,476.40 | **9,387.90** |
| 368 | 6,154.90 | 5,793.09 | 6,110.12 | 6,083.43 | **24,141.53** |
| 369 | 600.22 | 627.89 | 639.52 | 750.45 | **2,618.08** |
| **AL - ALABAMA - Total** | **181,957.55** | **171,303.17** | **176,392.46** | **174,810.69** | **704,463.87** |

**STATE: AR - ARKANSAS**

| REGISTRANT ZIP CODE 3 | 1ST QUARTER | 2ND QUARTER | 3RD QUARTER | 4TH QUARTER | TOTAL GRAMS |
|---|---|---|---|---|---|
| 716 | 4,546.88 | 4,205.20 | 4,391.65 | 4,551.76 | **17,695.49** |
| 717 | 4,279.34 | 4,202.48 | 4,405.21 | 4,481.56 | **17,368.60** |
| 718 | 6,021.97 | 5,943.36 | 6,139.08 | 5,988.00 | **24,092.41** |
| 719 | 8,078.66 | 8,001.79 | 8,123.23 | 8,299.31 | **32,503.00** |
| 720 | 9,639.62 | 9,212.38 | 9,444.05 | 9,659.53 | **37,955.59** |
| 721 | 11,101.47 | 10,632.86 | 10,699.23 | 10,779.96 | **43,213.52** |
| 722 | 5,355.06 | 4,923.68 | 5,194.23 | 5,300.65 | **20,773.62** |
| 723 | 5,683.70 | 5,606.31 | 5,624.44 | 5,592.55 | **22,507.00** |
| 724 | 9,032.25 | 8,443.33 | 8,920.01 | 8,954.16 | **35,349.75** |
| 725 | 4,697.73 | 4,686.38 | 4,740.40 | 4,683.13 | **18,807.64** |
| 726 | 4,436.50 | 4,372.29 | 4,528.11 | 4,607.70 | **17,944.61** |
| 727 | 11,521.18 | 10,966.48 | 11,030.55 | 10,983.08 | **44,501.29** |
| 728 | 3,493.03 | 3,282.79 | 3,455.71 | 3,426.07 | **13,657.59** |
| 729 | 10,779.51 | 10,341.26 | 11,078.20 | 10,968.28 | **43,167.26** |
| **AR - ARKANSAS - Total** | **98,666.90** | **94,820.61** | **97,774.11** | **98,275.73** | **389,537.35** |

**STATE: AS - AMERICAN SAMOA**

| REGISTRANT ZIP CODE 3 | 1ST QUARTER | 2ND QUARTER | 3RD QUARTER | 4TH QUARTER | TOTAL GRAMS |
|---|---|---|---|---|---|
| 967 | 1.82 | 3.63 | 4.84 | 0 | **10.29** |
| **AS - AMERICAN SAMOA - Total** | **1.82** | **3.63** | **4.84** | **0.00** | **10.29** |

**STATE: AZ - ARIZONA**

| REGISTRANT ZIP CODE 3 | 1ST QUARTER | 2ND QUARTER | 3RD QUARTER | 4TH QUARTER | TOTAL GRAMS |
|---|---|---|---|---|---|
| 850 | 10,066.30 | 9,328.31 | 9,674.12 | 9,703.17 | **38,771.90** |
| 851 | 4,200.76 | 3,989.35 | 3,945.25 | 4,095.64 | **16,231.01** |
| 852 | 16,305.48 | 15,759.05 | 15,912.65 | 16,297.27 | **64,274.46** |

| | | | | | |
|---|---|---|---|---|---|
| 853 | 13,319.98 | 12,437.17 | 12,476.54 | 13,142.08 | **51,375.77** |
| 855 | 1,559.64 | 1,520.24 | 1,520.74 | 1,499.41 | **6,100.03** |
| 856 | 2,950.02 | 2,687.56 | 2,828.36 | 2,745.40 | **11,211.35** |
| 857 | 9,298.22 | 8,884.82 | 9,054.87 | 8,887.10 | **36,125.00** |
| 859 | 1,811.78 | 1,950.07 | 1,969.74 | 1,891.15 | **7,622.74** |
| 860 | 1,206.64 | 1,222.37 | 1,266.88 | 1,269.56 | **4,965.45** |
| 863 | 5,240.11 | 5,258.08 | 5,467.24 | 5,320.16 | **21,285.59** |
| 864 | 6,843.99 | 6,833.34 | 6,599.02 | 6,484.69 | **26,761.03** |
| 865 | 41.96 | 65.38 | 73.83 | 59.53 | **240.70** |
| **AZ - ARIZONA - Total** | **72,844.89** | **69,935.74** | **70,789.24** | **71,395.16** | **284,965.03** |
| **STATE: CA - CALIFORNIA** | | | | | |
| REGISTRANT ZIP CODE 3 | 1ST QUARTER | 2ND QUARTER | 3RD QUARTER | 4TH QUARTER | **TOTAL GRAMS** |
| 900 | 13,482.67 | 12,006.59 | 13,079.17 | 12,463.58 | **51,032.01** |
| 902 | 11,870.32 | 10,965.73 | 11,365.47 | 11,171.46 | **45,372.98** |
| 903 | 1,250.60 | 1,190.84 | 1,243.13 | 1,165.95 | **4,850.51** |
| 904 | 1,175.27 | 1,049.64 | 1,150.92 | 1,130.72 | **4,506.54** |
| 905 | 2,908.29 | 2,764.43 | 2,887.02 | 2,813.26 | **11,373.00** |
| 906 | 9,645.89 | 8,532.30 | 9,561.12 | 9,307.82 | **37,047.12** |
| 907 | 8,794.51 | 8,038.78 | 8,063.12 | 8,222.31 | **33,118.73** |
| 908 | 5,356.91 | 5,158.19 | 5,385.19 | 5,241.53 | **21,141.82** |
| 910 | 2,916.63 | 2,604.11 | 2,803.57 | 2,764.46 | **11,088.77** |
| 911 | 1,913.54 | 1,769.30 | 1,900.88 | 1,901.12 | **7,484.83** |
| 912 | 2,534.73 | 2,341.16 | 2,581.98 | 2,561.03 | **10,018.91** |
| 913 | 17,676.15 | 16,590.28 | 17,205.54 | 17,008.95 | **68,480.92** |
| 914 | 4,645.73 | 3,965.32 | 4,370.39 | 4,260.86 | **17,242.31** |
| 915 | 1,434.71 | 1,308.20 | 1,317.28 | 1,310.80 | **5,370.98** |
| 916 | 1,226.07 | 1,030.27 | 1,130.93 | 1,102.38 | **4,489.65** |
| 917 | 25,700.53 | 23,213.37 | 24,770.49 | 24,367.77 | **98,052.16** |
| 918 | 792.61 | 659.23 | 721.18 | 674.49 | **2,847.52** |
| 919 | 5,882.61 | 6,243.61 | 6,074.61 | 5,796.66 | **23,997.49** |
| 920 | 20,014.00 | 18,565.28 | 19,772.51 | 19,581.95 | **77,933.74** |
| 921 | 12,430.32 | 11,484.07 | 11,954.34 | 11,728.57 | **47,597.30** |
| 922 | 15,667.72 | 14,988.72 | 15,038.26 | 15,231.33 | **60,926.02** |
| 923 | 33,712.54 | 32,656.69 | 33,562.91 | 32,875.26 | **132,807.41** |
| 924 | 5,124.44 | 4,720.90 | 4,864.68 | 4,835.80 | **19,545.83** |
| 925 | 30,106.81 | 29,693.00 | 30,445.72 | 30,064.27 | **120,309.80** |
| 926 | 16,848.02 | 15,306.03 | 16,351.98 | 16,132.03 | **64,638.06** |
| 927 | 5,287.28 | 5,459.44 | 5,149.94 | 4,929.54 | **20,826.20** |
| 928 | 18,298.03 | 18,001.45 | 17,780.34 | 17,571.24 | **71,651.05** |
| 930 | 12,075.01 | 11,306.21 | 11,682.59 | 11,696.49 | **46,760.30** |
| 931 | 1,781.98 | 1,639.16 | 1,726.30 | 1,695.07 | **6,842.51** |
| 932 | 17,467.23 | 17,654.89 | 17,717.97 | 17,833.53 | **70,673.62** |
| 933 | 15,895.41 | 15,609.22 | 16,168.62 | 15,927.38 | **63,600.62** |
| 934 | 10,781.37 | 10,532.09 | 10,723.34 | 10,604.59 | **42,641.39** |
| 935 | 13,202.53 | 12,929.36 | 12,699.74 | 12,740.80 | **51,572.43** |
| 936 | 12,185.42 | 11,960.29 | 12,004.04 | 11,934.99 | **48,084.73** |
| 937 | 15,258.67 | 14,911.26 | 14,691.81 | 14,684.21 | **59,545.96** |
| 939 | 5,020.37 | 4,592.52 | 4,874.36 | 4,704.55 | **19,191.80** |
| 940 | 6,371.09 | 5,764.01 | 6,205.11 | 6,199.41 | **24,539.61** |
| 941 | 5,527.81 | 5,004.88 | 5,158.77 | 5,245.99 | **20,937.46** |
| 943 | 1,275.32 | 1,228.96 | 1,283.78 | 1,203.09 | **4,991.14** |
| 944 | 1,155.68 | 1,044.40 | 1,023.39 | 1,041.37 | **4,264.85** |
| 945 | 42,940.74 | 41,578.90 | 41,197.03 | 41,552.61 | **167,269.28** |
| 946 | 5,096.15 | 4,756.41 | 4,827.25 | 4,824.82 | **19,504.64** |
| 947 | 755.67 | 818.39 | 825.12 | 748.47 | **3,147.66** |
| 948 | 2,190.15 | 2,079.42 | 2,032.85 | 2,057.93 | **8,360.36** |
| 949 | 5,929.38 | 5,720.25 | 5,885.35 | 5,866.99 | **23,401.97** |
| 950 | 8,427.52 | 7,900.30 | 8,370.87 | 8,273.39 | **32,972.08** |
| 951 | 5,742.80 | 5,265.24 | 5,606.84 | 5,467.66 | **22,082.54** |
| 952 | 11,670.10 | 11,419.60 | 11,179.56 | 11,301.73 | **45,570.99** |

| | | | | | |
|---|---|---|---|---|---|
| 953 | 33,439.22 | 32,689.55 | 32,763.48 | 32,781.41 | **131,673.65** |
| 954 | 11,720.59 | 11,652.69 | 11,662.09 | 11,642.25 | **46,677.63** |
| 955 | 5,070.37 | 4,741.78 | 4,784.32 | 4,594.15 | **19,190.63** |
| 956 | 37,841.40 | 36,418.51 | 36,763.31 | 36,557.15 | **147,580.37** |
| 957 | 6,654.54 | 6,333.74 | 6,466.38 | 6,485.03 | **25,939.70** |
| 958 | 21,389.90 | 20,793.84 | 20,935.42 | 20,874.10 | **83,993.26** |
| 959 | 18,080.40 | 17,322.82 | 17,236.95 | 17,166.96 | **69,807.12** |
| 960 | 13,046.02 | 12,625.28 | 12,446.47 | 12,269.10 | **50,386.88** |
| 961 | 2,070.69 | 1,943.66 | 1,921.72 | 1,849.81 | **7,785.88** |
| **CA - CALIFORNIA - Total** | **626,760.44** | **598,544.57** | **611,397.50** | **606,040.19** | **2,442,742.71** |

**STATE: CO - COLORADO**

| REGISTRANT ZIP CODE 3 | 1ST QUARTER | 2ND QUARTER | 3RD QUARTER | 4TH QUARTER | **TOTAL GRAMS** |
|---|---|---|---|---|---|
| 800 | 6,842.46 | 6,462.95 | 6,572.56 | 6,620.54 | **26,498.51** |
| 801 | 5,690.79 | 5,208.05 | 5,445.95 | 5,508.32 | **21,853.11** |
| 802 | 6,199.60 | 6,011.78 | 5,939.55 | 6,016.65 | **24,167.58** |
| 803 | 618.71 | 568.88 | 597.62 | 536.18 | **2,321.39** |
| 804 | 1,320.28 | 1,224.04 | 1,330.80 | 1,251.07 | **5,126.19** |
| 805 | 4,651.62 | 4,282.05 | 4,605.27 | 4,749.52 | **18,288.46** |
| 806 | 1,857.23 | 1,661.58 | 1,810.48 | 1,869.29 | **7,198.59** |
| 807 | 667.77 | 618.56 | 665.31 | 634.72 | **2,586.35** |
| 808 | 1,173.58 | 1,170.43 | 1,226.44 | 1,178.16 | **4,748.61** |
| 809 | 5,582.34 | 5,457.12 | 5,557.75 | 5,493.72 | **22,090.93** |
| 810 | 4,193.55 | 3,980.49 | 4,096.94 | 4,041.21 | **16,312.19** |
| 811 | 531.46 | 546.99 | 554.33 | 527.65 | **2,160.43** |
| 812 | 1,093.39 | 1,043.69 | 1,149.59 | 1,064.78 | **4,351.46** |
| 813 | 750.84 | 750.75 | 773.32 | 800.05 | **3,074.95** |
| 814 | 770.39 | 762.83 | 779.97 | 772.85 | **3,086.05** |
| 815 | 1,781.13 | 1,793.47 | 1,733.28 | 1,703.69 | **7,011.57** |
| 816 | 1,005.08 | 857.71 | 945.98 | 954.99 | **3,763.75** |
| **CO - COLORADO - Total** | **44,730.22** | **42,401.37** | **43,785.14** | **43,723.39** | **174,640.12** |

**STATE: CT - CONNECTICUT**

| REGISTRANT ZIP CODE 3 | 1ST QUARTER | 2ND QUARTER | 3RD QUARTER | 4TH QUARTER | **TOTAL GRAMS** |
|---|---|---|---|---|---|
| 060 | 3,040.29 | 2,715.04 | 2,828.11 | 2,781.46 | **11,364.91** |
| 061 | 752.01 | 614.11 | 640.27 | 616.65 | **2,623.03** |
| 062 | 718.75 | 688.09 | 700.92 | 704.68 | **2,812.42** |
| 063 | 1,425.31 | 1,465.67 | 1,521.05 | 1,413.12 | **5,825.15** |
| 064 | 3,455.40 | 3,063.51 | 3,223.18 | 3,203.80 | **12,945.89** |
| 065 | 806.62 | 730.96 | 818.61 | 813.19 | **3,169.38** |
| 066 | 606.78 | 575.60 | 573.30 | 571.34 | **2,327.03** |
| 067 | 1,750.95 | 1,750.60 | 1,850.12 | 1,736.18 | **7,087.85** |
| 068 | 1,088.70 | 946.08 | 1,047.92 | 935.81 | **4,018.53** |
| 069 | 198.25 | 169.49 | 174.67 | 179.99 | **722.39** |
| **CT - CONNECTICUT - Total** | **13,843.06** | **12,719.15** | **13,378.14** | **12,956.23** | **52,896.58** |

**STATE: DC - DISTRICT OF COLUMBIA**

| REGISTRANT ZIP CODE 3 | 1ST QUARTER | 2ND QUARTER | 3RD QUARTER | 4TH QUARTER | **TOTAL GRAMS** |
|---|---|---|---|---|---|
| 200 | 800.89 | 602.29 | 711.72 | 619.88 | **2,734.78** |
| 203 | 7.26 | 0 | 1.82 | 0 | **9.08** |
| 204 | 83.24 | 79.91 | 66.59 | 79.91 | **309.66** |
| **DC - DISTRICT OF COLUMBIA - Total** | **891.40** | **682.21** | **780.13** | **699.79** | **3,053.53** |

**STATE: DE - DELAWARE**

| REGISTRANT ZIP CODE 3 | 1ST QUARTER | 2ND QUARTER | 3RD QUARTER | 4TH QUARTER | **TOTAL GRAMS** |
|---|---|---|---|---|---|
| 197 | 1,152.88 | 1,094.76 | 1,092.74 | 1,244.48 | **4,584.86** |
| 198 | 742.84 | 626.69 | 661.02 | 639.66 | **2,670.21** |
| 199 | 2,163.43 | 2,110.05 | 2,339.14 | 2,288.70 | **8,901.31** |
| **DE - DELAWARE - Total** | **4,059.16** | **3,831.49** | **4,092.90** | **4,172.84** | **16,156.38** |

**STATE: FL - FLORIDA**

| REGISTRANT ZIP CODE 3 | 1ST QUARTER | 2ND QUARTER | 3RD QUARTER | 4TH QUARTER | **TOTAL GRAMS** |
|---|---|---|---|---|---|
| 320 | 14,155.07 | 13,380.10 | 13,871.67 | 13,623.52 | **55,030.37** |
| 321 | 16,513.90 | 15,760.11 | 16,093.00 | 15,977.65 | **64,344.67** |
| 322 | 24,910.99 | 23,620.62 | 24,280.81 | 23,760.93 | **96,573.35** |

| | | | | | |
|---|---|---|---|---|---|
| 323 | 6,400.91 | 5,968.65 | 6,327.62 | 6,279.35 | **24,976.53** |
| 324 | 12,024.46 | 10,373.56 | 10,807.92 | 10,722.60 | **43,928.54** |
| 325 | 20,315.69 | 19,136.68 | 19,603.32 | 19,032.07 | **78,087.75** |
| 326 | 4,697.60 | 4,530.32 | 4,509.50 | 4,328.85 | **18,066.27** |
| 327 | 13,983.43 | 13,145.23 | 13,331.42 | 13,453.20 | **53,913.27** |
| 328 | 5,744.72 | 5,211.61 | 5,735.11 | 5,586.56 | **22,278.00** |
| 329 | 11,874.47 | 11,076.40 | 11,271.33 | 11,296.30 | **45,518.51** |
| 330 | 6,312.07 | 5,582.66 | 5,627.71 | 5,543.04 | **23,065.47** |
| 331 | 3,101.85 | 2,977.73 | 3,175.49 | 3,176.44 | **12,431.52** |
| 333 | 4,482.29 | 4,039.79 | 4,225.72 | 4,173.72 | **16,921.53** |
| 334 | 10,859.23 | 10,071.66 | 10,187.61 | 10,192.32 | **41,310.81** |
| 335 | 9,781.36 | 9,162.63 | 9,601.13 | 9,524.70 | **38,069.81** |
| 336 | 8,567.07 | 7,961.76 | 8,037.74 | 7,781.56 | **32,348.12** |
| 337 | 13,197.67 | 11,961.82 | 12,155.43 | 12,438.46 | **49,753.37** |
| 338 | 8,983.17 | 8,530.10 | 8,692.05 | 8,830.31 | **35,035.64** |
| 339 | 12,038.07 | 10,961.99 | 10,919.20 | 11,267.79 | **45,187.04** |
| 341 | 3,695.13 | 3,210.22 | 2,823.69 | 3,101.46 | **12,830.50** |
| 342 | 13,438.45 | 12,590.56 | 12,491.28 | 12,809.20 | **51,329.48** |
| 344 | 10,121.72 | 9,830.91 | 10,078.54 | 10,041.33 | **40,072.51** |
| 346 | 13,502.51 | 12,687.32 | 13,084.30 | 13,214.51 | **52,488.64** |
| 347 | 6,642.87 | 6,140.60 | 6,317.19 | 6,200.23 | **25,300.89** |
| 349 | 5,742.60 | 5,445.35 | 5,650.78 | 5,449.03 | **22,287.76** |
| **FL - FLORIDA - Total** | **261,087.28** | **243,358.40** | **248,899.54** | **247,805.11** | **1,001,150.34** |

**STATE: GA - GEORGIA**

| REGISTRANT ZIP CODE 3 | 1ST QUARTER | 2ND QUARTER | 3RD QUARTER | 4TH QUARTER | TOTAL GRAMS |
|---|---|---|---|---|---|
| 300 | 21,513.27 | 19,519.44 | 20,683.31 | 21,008.98 | **82,725.00** |
| 301 | 25,336.92 | 23,749.18 | 25,267.10 | 24,611.15 | **98,964.36** |
| 302 | 14,873.75 | 14,200.11 | 14,587.96 | 14,341.36 | **58,003.19** |
| 303 | 4,353.91 | 3,727.86 | 3,930.85 | 4,105.66 | **16,118.28** |
| 304 | 5,013.19 | 4,718.16 | 4,911.98 | 4,799.85 | **19,443.18** |
| 305 | 15,998.96 | 15,156.18 | 15,780.79 | 15,892.70 | **62,828.64** |
| 306 | 10,503.81 | 10,062.65 | 10,232.52 | 10,054.36 | **40,853.34** |
| 307 | 8,497.30 | 8,365.81 | 8,302.37 | 8,334.09 | **33,499.58** |
| 308 | 2,302.62 | 2,027.53 | 2,072.41 | 2,052.35 | **8,454.92** |
| 309 | 3,892.34 | 3,125.02 | 3,498.74 | 3,046.42 | **13,562.52** |
| 310 | 16,693.72 | 15,972.14 | 16,218.24 | 16,507.17 | **65,391.27** |
| 312 | 6,543.66 | 6,186.58 | 6,301.59 | 6,214.83 | **25,246.67** |
| 313 | 3,554.08 | 3,383.06 | 3,487.48 | 3,299.77 | **13,724.39** |
| 314 | 3,407.90 | 3,055.15 | 3,176.27 | 3,153.17 | **12,792.49** |
| 315 | 13,099.40 | 11,937.56 | 12,526.19 | 11,906.00 | **49,469.15** |
| 316 | 7,842.62 | 7,061.42 | 7,642.75 | 7,414.75 | **29,961.53** |
| 317 | 9,452.94 | 8,649.86 | 9,168.96 | 9,088.76 | **36,360.52** |
| 318 | 1,070.62 | 955.82 | 999.76 | 981.29 | **4,007.50** |
| 319 | 4,736.54 | 4,370.83 | 4,757.91 | 4,604.90 | **18,470.17** |
| 398 | 2,890.03 | 2,781.12 | 2,805.39 | 2,747.28 | **11,223.81** |
| **GA - GEORGIA - Total** | **181,577.60** | **169,005.49** | **176,352.55** | **174,164.86** | **701,100.50** |

**STATE: GU - GUAM**

| REGISTRANT ZIP CODE 3 | 1ST QUARTER | 2ND QUARTER | 3RD QUARTER | 4TH QUARTER | TOTAL GRAMS |
|---|---|---|---|---|---|
| 969 | 103.05 | 88.09 | 108.52 | 111.39 | **411.05** |
| **GU - GUAM - Total** | **103.05** | **88.09** | **108.52** | **111.39** | **411.05** |

**STATE: HI - HAWAII**

| REGISTRANT ZIP CODE 3 | 1ST QUARTER | 2ND QUARTER | 3RD QUARTER | 4TH QUARTER | TOTAL GRAMS |
|---|---|---|---|---|---|
| 967 | 9,302.06 | 9,109.39 | 8,676.83 | 8,590.72 | **35,679.00** |
| 968 | 2,901.58 | 2,648.87 | 2,612.77 | 2,584.35 | **10,747.57** |
| **HI - HAWAII - Total** | **12,203.65** | **11,758.26** | **11,289.60** | **11,175.07** | **46,426.58** |

**STATE: IA - IOWA**

| REGISTRANT ZIP CODE 3 | 1ST QUARTER | 2ND QUARTER | 3RD QUARTER | 4TH QUARTER | TOTAL GRAMS |
|---|---|---|---|---|---|
| 500 | 3,009.88 | 2,869.61 | 3,002.79 | 2,954.53 | **11,836.80** |
| 501 | 2,768.85 | 2,623.69 | 2,670.31 | 2,644.44 | **10,707.29** |
| 502 | 2,638.10 | 2,503.86 | 2,637.52 | 2,718.00 | **10,497.47** |

| | | | | | |
|---|---|---|---|---|---|
| 503 | 6,236.54 | 5,925.81 | 6,104.58 | 6,152.47 | **24,419.41** |
| 504 | 1,227.20 | 1,199.42 | 1,225.17 | 1,241.94 | **4,893.73** |
| 505 | 1,545.02 | 1,479.31 | 1,557.27 | 1,574.67 | **6,156.28** |
| 506 | 1,269.67 | 1,167.78 | 1,279.23 | 1,260.68 | **4,977.35** |
| 507 | 1,074.97 | 1,003.72 | 1,124.96 | 1,116.38 | **4,320.04** |
| 508 | 508.80 | 409.98 | 474.37 | 444.58 | **1,837.72** |
| 510 | 748.03 | 748.07 | 714.30 | 784.67 | **2,995.07** |
| 511 | 1,726.42 | 1,554.64 | 1,635.04 | 1,630.59 | **6,546.70** |
| 512 | 318.75 | 280.41 | 312.66 | 314.14 | **1,225.95** |
| 513 | 480.42 | 436.16 | 500.17 | 500.42 | **1,917.17** |
| 514 | 473.32 | 490.50 | 482.77 | 467.94 | **1,914.53** |
| 515 | 1,545.63 | 1,462.16 | 1,522.51 | 1,552.99 | **6,083.30** |
| 516 | 277.33 | 276.15 | 276.93 | 290.50 | **1,120.91** |
| 520 | 1,170.69 | 1,080.92 | 1,150.98 | 1,172.03 | **4,574.62** |
| 521 | 354.11 | 330.37 | 354.59 | 345.46 | **1,384.54** |
| 522 | 1,540.61 | 1,425.32 | 1,501.21 | 1,548.34 | **6,015.49** |
| 523 | 911.09 | 848.00 | 930.17 | 903.18 | **3,592.43** |
| 524 | 1,464.99 | 1,322.91 | 1,352.94 | 1,372.85 | **5,513.69** |
| 525 | 2,006.47 | 1,852.88 | 1,978.69 | 1,958.11 | **7,796.15** |
| 526 | 1,456.71 | 1,441.76 | 1,489.12 | 1,432.34 | **5,819.93** |
| 527 | 1,343.54 | 1,194.54 | 1,336.90 | 1,293.72 | **5,168.70** |
| 528 | 1,702.17 | 1,657.59 | 1,603.31 | 1,590.96 | **6,554.03** |
| **IA - IOWA - Total** | **37,799.33** | **35,585.55** | **37,218.48** | **37,265.94** | **147,869.30** |

**STATE: ID - IDAHO**

| REGISTRANT ZIP CODE 3 | 1ST QUARTER | 2ND QUARTER | 3RD QUARTER | 4TH QUARTER | **TOTAL GRAMS** |
|---|---|---|---|---|---|
| 832 | 4,803.44 | 4,687.37 | 4,680.49 | 4,743.60 | **18,914.90** |
| 833 | 5,342.67 | 5,163.61 | 5,397.65 | 5,260.21 | **21,164.13** |
| 834 | 3,744.37 | 3,741.88 | 3,877.29 | 3,923.18 | **15,286.72** |
| 835 | 2,748.87 | 2,828.92 | 2,709.37 | 2,841.34 | **11,128.51** |
| 836 | 11,907.20 | 11,690.54 | 11,860.55 | 12,071.79 | **47,530.09** |
| 837 | 7,030.05 | 6,835.86 | 6,992.45 | 6,961.07 | **27,819.43** |
| 838 | 7,506.60 | 7,362.14 | 7,619.84 | 7,598.95 | **30,087.54** |
| **ID - IDAHO - Total** | **43,083.20** | **42,310.34** | **43,137.64** | **43,400.14** | **171,931.31** |

**STATE: IL - ILLINOIS**

| REGISTRANT ZIP CODE 3 | 1ST QUARTER | 2ND QUARTER | 3RD QUARTER | 4TH QUARTER | **TOTAL GRAMS** |
|---|---|---|---|---|---|
| 600 | 24,611.34 | 22,069.23 | 23,600.46 | 23,938.79 | **94,219.82** |
| 601 | 22,175.87 | 20,408.67 | 21,758.53 | 21,746.46 | **86,089.53** |
| 602 | 672.78 | 635.03 | 680.72 | 690.14 | **2,678.67** |
| 603 | 937.89 | 888.55 | 957.13 | 912.12 | **3,695.69** |
| 604 | 28,151.78 | 26,600.33 | 28,036.87 | 27,768.38 | **110,557.37** |
| 605 | 12,928.13 | 12,162.00 | 13,096.47 | 12,909.44 | **51,096.03** |
| 606 | 21,412.60 | 19,305.92 | 20,355.89 | 20,017.63 | **81,092.03** |
| 607 | 2,291.90 | 2,189.50 | 2,271.32 | 2,218.63 | **8,971.35** |
| 608 | 1,797.37 | 1,816.92 | 1,965.83 | 1,870.67 | **7,450.78** |
| 609 | 4,075.71 | 4,009.56 | 4,056.07 | 4,021.93 | **16,163.27** |
| 610 | 4,807.89 | 4,678.46 | 4,856.42 | 4,809.67 | **19,152.43** |
| 611 | 7,720.21 | 6,977.91 | 7,569.67 | 7,392.11 | **29,659.90** |
| 612 | 2,096.57 | 1,839.49 | 2,063.09 | 1,939.48 | **7,938.64** |
| 613 | 3,170.49 | 3,069.86 | 3,280.18 | 3,370.57 | **12,891.10** |
| 614 | 2,763.91 | 2,570.07 | 2,588.11 | 2,655.39 | **10,577.47** |
| 615 | 3,876.88 | 3,600.86 | 3,741.18 | 3,807.39 | **15,026.30** |
| 616 | 5,899.56 | 5,573.93 | 5,646.21 | 5,341.20 | **22,460.90** |
| 617 | 3,346.72 | 3,098.10 | 3,369.56 | 3,258.66 | **13,073.03** |
| 618 | 6,099.99 | 5,834.12 | 5,933.17 | 5,997.48 | **23,864.77** |
| 619 | 1,776.67 | 1,530.14 | 1,741.45 | 1,739.33 | **6,787.59** |
| 620 | 8,274.78 | 7,830.76 | 8,161.09 | 8,002.58 | **32,269.21** |
| 622 | 7,934.18 | 7,502.98 | 8,051.95 | 7,822.99 | **31,312.11** |
| 623 | 2,682.82 | 2,643.98 | 2,623.41 | 2,765.42 | **10,715.64** |
| 624 | 3,190.74 | 2,904.74 | 3,175.12 | 2,980.73 | **12,251.33** |
| 625 | 3,756.15 | 3,582.66 | 3,950.14 | 3,854.10 | **15,143.05** |

| | | | | | |
|---|---|---|---|---|---|
| 626 | 3,010.88 | 3,029.43 | 3,086.75 | 3,189.11 | **12,316.17** |
| 627 | 3,942.81 | 3,544.96 | 3,841.09 | 3,754.51 | **15,083.36** |
| 628 | 5,772.77 | 5,560.84 | 5,664.64 | 5,609.89 | **22,608.13** |
| 629 | 11,927.14 | 11,501.59 | 11,906.18 | 11,460.91 | **46,795.83** |
| **IL - ILLINOIS - Total** | **211,106.51** | **196,960.59** | **208,028.68** | **205,845.72** | **821,941.50** |

**STATE: IN - INDIANA**

| REGISTRANT ZIP CODE 3 | 1ST QUARTER | 2ND QUARTER | 3RD QUARTER | 4TH QUARTER | TOTAL GRAMS |
|---|---|---|---|---|---|
| 460 | 11,153.83 | 10,372.52 | 11,047.83 | 11,330.74 | **43,904.93** |
| 461 | 15,443.57 | 14,285.88 | 14,482.47 | 14,468.68 | **58,680.59** |
| 462 | 22,119.48 | 20,215.87 | 21,045.96 | 21,053.83 | **84,435.15** |
| 463 | 20,476.90 | 19,847.47 | 20,642.39 | 20,355.93 | **81,322.69** |
| 464 | 2,663.26 | 2,508.78 | 2,563.18 | 2,459.64 | **10,194.87** |
| 465 | 7,064.97 | 6,799.26 | 7,166.38 | 7,111.97 | **28,142.58** |
| 466 | 2,173.06 | 2,154.87 | 2,206.90 | 2,107.80 | **8,642.63** |
| 467 | 4,638.18 | 4,629.26 | 4,774.84 | 4,983.90 | **19,026.18** |
| 468 | 5,877.97 | 5,670.53 | 6,020.30 | 5,876.15 | **23,444.95** |
| 469 | 9,959.52 | 9,599.16 | 9,983.99 | 10,113.11 | **39,655.78** |
| 470 | 2,492.42 | 2,377.77 | 2,440.36 | 2,403.24 | **9,713.78** |
| 471 | 12,499.60 | 12,185.17 | 12,138.24 | 12,127.35 | **48,950.37** |
| 472 | 5,046.81 | 4,780.24 | 5,191.92 | 5,332.26 | **20,351.23** |
| 473 | 10,738.28 | 10,231.79 | 10,760.03 | 10,614.32 | **42,344.42** |
| 474 | 6,523.58 | 6,198.97 | 6,454.97 | 6,246.94 | **25,424.45** |
| 475 | 5,515.01 | 5,146.08 | 5,436.62 | 5,434.12 | **21,531.83** |
| 476 | 3,681.53 | 3,561.22 | 3,772.72 | 3,690.27 | **14,705.73** |
| 477 | 9,195.47 | 8,787.43 | 8,912.58 | 8,827.21 | **35,722.68** |
| 478 | 3,659.47 | 3,473.31 | 3,719.59 | 3,764.23 | **14,616.59** |
| 479 | 6,649.35 | 6,425.39 | 6,736.54 | 6,640.11 | **26,451.39** |
| **IN - INDIANA - Total** | **167,572.24** | **159,250.97** | **165,497.80** | **164,941.79** | **657,262.81** |

**STATE: KS - KANSAS**

| REGISTRANT ZIP CODE 3 | 1ST QUARTER | 2ND QUARTER | 3RD QUARTER | 4TH QUARTER | TOTAL GRAMS |
|---|---|---|---|---|---|
| 660 | 7,755.81 | 7,471.04 | 7,736.30 | 7,757.41 | **30,720.57** |
| 661 | 2,040.20 | 2,014.09 | 2,086.65 | 1,965.91 | **8,106.86** |
| 662 | 6,143.96 | 5,405.91 | 5,780.26 | 5,882.74 | **23,212.87** |
| 664 | 980.31 | 923.27 | 1,018.16 | 911.78 | **3,833.52** |
| 665 | 1,447.73 | 1,349.69 | 1,404.76 | 1,407.74 | **5,609.92** |
| 666 | 3,141.80 | 2,935.90 | 3,101.52 | 3,000.00 | **12,179.22** |
| 667 | 3,708.75 | 3,465.17 | 3,692.44 | 3,542.94 | **14,409.30** |
| 668 | 891.58 | 836.22 | 847.91 | 835.37 | **3,411.08** |
| 669 | 469.22 | 439.03 | 489.34 | 478.21 | **1,875.80** |
| 670 | 4,523.38 | 4,552.41 | 4,565.68 | 4,541.54 | **18,183.02** |
| 671 | 2,452.47 | 2,303.05 | 2,310.49 | 2,308.58 | **9,374.59** |
| 672 | 12,621.60 | 12,708.73 | 12,759.11 | 12,569.48 | **50,658.92** |
| 673 | 1,932.42 | 1,734.10 | 1,858.16 | 1,925.46 | **7,450.14** |
| 674 | 3,409.16 | 3,343.60 | 3,452.36 | 3,467.37 | **13,672.49** |
| 675 | 2,757.19 | 2,591.15 | 2,776.57 | 2,628.59 | **10,753.50** |
| 676 | 995.01 | 871.62 | 949.75 | 859.83 | **3,676.21** |
| 677 | 492.81 | 471.73 | 476.89 | 455.78 | **1,897.21** |
| 678 | 1,803.59 | 1,734.92 | 1,697.80 | 1,627.30 | **6,863.62** |
| 679 | 727.09 | 668.05 | 679.53 | 621.94 | **2,696.61** |
| **KS - KANSAS - Total** | **58,294.09** | **55,819.69** | **57,683.70** | **56,787.97** | **228,585.45** |

**STATE: KY - KENTUCKY**

| REGISTRANT ZIP CODE 3 | 1ST QUARTER | 2ND QUARTER | 3RD QUARTER | 4TH QUARTER | TOTAL GRAMS |
|---|---|---|---|---|---|
| 400 | 6,476.22 | 6,205.74 | 6,529.92 | 6,370.83 | **25,582.71** |
| 401 | 3,811.23 | 3,769.34 | 3,950.20 | 3,850.40 | **15,381.18** |
| 402 | 27,667.45 | 26,088.84 | 27,418.61 | 26,712.13 | **107,887.03** |
| 403 | 9,175.86 | 9,087.44 | 9,243.20 | 9,176.04 | **36,682.55** |
| 404 | 5,979.69 | 5,680.39 | 5,806.11 | 5,830.19 | **23,296.39** |
| 405 | 5,965.82 | 5,758.16 | 5,708.17 | 5,741.53 | **23,173.68** |
| 406 | 1,496.55 | 1,437.04 | 1,405.08 | 1,376.41 | **5,715.09** |
| 407 | 5,596.16 | 5,324.51 | 5,479.68 | 5,576.94 | **21,977.29** |

| | | | | | |
|---|---|---|---|---|---|
| 408 | 1,638.44 | 1,357.17 | 1,504.55 | 1,550.94 | **6,051.10** |
| 409 | 6,975.96 | 6,592.12 | 7,092.70 | 6,846.31 | **27,507.09** |
| 410 | 7,799.71 | 7,446.76 | 7,716.23 | 7,805.27 | **30,767.98** |
| 411 | 2,856.50 | 2,516.51 | 2,703.40 | 2,749.64 | **10,826.06** |
| 412 | 2,606.61 | 2,587.38 | 2,697.86 | 2,751.20 | **10,643.05** |
| 413 | 2,828.84 | 2,715.52 | 2,828.95 | 2,766.86 | **11,140.17** |
| 414 | 1,120.94 | 1,078.32 | 1,095.57 | 1,125.88 | **4,420.71** |
| 415 | 4,733.89 | 4,607.28 | 4,747.60 | 4,866.81 | **18,955.58** |
| 416 | 3,615.94 | 3,605.29 | 3,776.11 | 3,781.28 | **14,778.63** |
| 417 | 4,232.26 | 4,068.64 | 4,260.84 | 4,209.49 | **16,771.23** |
| 418 | 1,760.28 | 1,725.21 | 1,754.64 | 1,765.08 | **7,005.20** |
| 420 | 11,365.52 | 10,995.99 | 11,064.24 | 11,012.29 | **44,438.04** |
| 421 | 10,666.72 | 9,805.49 | 10,377.38 | 10,209.10 | **41,058.69** |
| 422 | 5,445.52 | 5,366.42 | 5,538.15 | 5,300.17 | **21,650.26** |
| 423 | 7,788.59 | 7,552.52 | 7,451.61 | 7,742.91 | **30,535.63** |
| 424 | 7,523.29 | 7,195.77 | 7,500.27 | 7,170.11 | **29,389.43** |
| 425 | 2,618.44 | 2,500.38 | 2,516.54 | 2,641.22 | **10,276.58** |
| 426 | 3,269.26 | 3,249.33 | 3,179.14 | 3,183.82 | **12,881.55** |
| 427 | 7,417.69 | 7,042.88 | 7,619.21 | 7,309.16 | **29,388.93** |
| **KY - KENTUCKY - Total** | **162,433.38** | **155,360.44** | **160,965.97** | **159,422.02** | **638,181.81** |

**STATE: LA - LOUISIANA**

| REGISTRANT ZIP CODE 3 | 1ST QUARTER | 2ND QUARTER | 3RD QUARTER | 4TH QUARTER | **TOTAL GRAMS** |
|---|---|---|---|---|---|
| 700 | 10,797.77 | 9,923.86 | 10,308.22 | 9,961.61 | **40,991.46** |
| 701 | 5,133.44 | 4,601.87 | 4,993.22 | 5,134.49 | **19,863.02** |
| 703 | 6,531.88 | 5,929.80 | 6,193.88 | 6,152.14 | **24,807.71** |
| 704 | 12,879.12 | 12,151.17 | 12,678.52 | 12,819.89 | **50,528.70** |
| 705 | 19,999.19 | 18,568.31 | 19,470.66 | 19,153.67 | **77,191.83** |
| 706 | 5,595.80 | 5,187.01 | 4,806.98 | 4,950.73 | **20,540.52** |
| 707 | 10,558.81 | 9,997.66 | 10,532.29 | 10,258.93 | **41,347.69** |
| 708 | 7,832.23 | 7,280.21 | 7,638.94 | 7,568.72 | **30,320.10** |
| 710 | 3,919.78 | 3,617.57 | 3,884.29 | 3,895.81 | **15,317.45** |
| 711 | 10,222.38 | 9,823.09 | 10,266.56 | 10,248.81 | **40,560.84** |
| 712 | 9,359.34 | 8,718.48 | 9,471.66 | 8,931.18 | **36,480.66** |
| 713 | 9,190.35 | 8,621.84 | 8,656.65 | 8,580.05 | **35,048.89** |
| 714 | 4,507.50 | 4,288.04 | 4,322.80 | 4,257.75 | **17,376.09** |
| **LA - LOUISIANA - Total** | **116,527.59** | **108,708.91** | **113,224.68** | **111,913.78** | **450,374.96** |

**STATE: MA - MASSACHUSETTS**

| REGISTRANT ZIP CODE 3 | 1ST QUARTER | 2ND QUARTER | 3RD QUARTER | 4TH QUARTER | **TOTAL GRAMS** |
|---|---|---|---|---|---|
| 010 | 1,498.72 | 1,425.82 | 1,568.17 | 1,500.59 | **5,993.29** |
| 011 | 366.77 | 303.34 | 365.45 | 379.26 | **1,414.81** |
| 012 | 517.73 | 489.66 | 545.76 | 479.06 | **2,032.22** |
| 013 | 290.03 | 269.27 | 286.53 | 293.59 | **1,139.43** |
| 014 | 912.43 | 877.41 | 953.14 | 955.61 | **3,698.59** |
| 015 | 1,308.31 | 1,198.15 | 1,256.72 | 1,261.43 | **5,024.61** |
| 016 | 799.08 | 727.56 | 762.44 | 736.19 | **3,025.27** |
| 017 | 1,046.21 | 856.53 | 938.58 | 938.96 | **3,780.27** |
| 018 | 4,014.50 | 3,483.68 | 3,476.19 | 3,437.98 | **14,412.35** |
| 019 | 2,045.31 | 1,739.91 | 1,856.30 | 1,832.02 | **7,473.54** |
| 020 | 1,036.36 | 905.77 | 1,052.46 | 962.28 | **3,956.87** |
| 021 | 2,248.58 | 2,050.21 | 2,198.58 | 2,163.89 | **8,661.25** |
| 022 | 51.25 | 41.41 | 36.63 | 35.63 | **164.91** |
| 023 | 1,381.86 | 1,343.60 | 1,389.63 | 1,271.66 | **5,386.76** |
| 024 | 539.03 | 547.32 | 563.34 | 551.10 | **2,200.78** |
| 025 | 396.44 | 384.20 | 403.83 | 375.25 | **1,559.73** |
| 026 | 693.41 | 652.29 | 700.40 | 633.44 | **2,679.55** |
| 027 | 2,446.14 | 2,252.82 | 2,370.30 | 2,192.50 | **9,261.75** |
| **MA - MASSACHUSETTS - Total** | **21,592.15** | **19,548.95** | **20,724.45** | **20,000.43** | **81,865.98** |

**STATE: MD - MARYLAND**

| REGISTRANT ZIP CODE 3 | 1ST QUARTER | 2ND QUARTER | 3RD QUARTER | 4TH QUARTER | **TOTAL GRAMS** |
|---|---|---|---|---|---|
| 206 | 2,024.04 | 1,891.46 | 2,078.45 | 1,904.78 | **7,898.73** |

| | | | | | |
|---|---|---|---|---|---|
| 207 | 2,192.52 | 1,933.67 | 2,098.28 | 1,990.62 | **8,215.10** |
| 208 | 1,715.09 | 1,413.45 | 1,543.42 | 1,578.03 | **6,249.98** |
| 209 | 413.49 | 355.56 | 351.73 | 371.34 | **1,492.12** |
| 210 | 4,303.87 | 3,819.24 | 4,194.57 | 3,967.46 | **16,285.14** |
| 211 | 2,048.77 | 1,807.41 | 2,017.91 | 1,970.10 | **7,844.19** |
| 212 | 4,223.28 | 3,795.58 | 3,977.98 | 3,975.59 | **15,972.43** |
| 214 | 465.31 | 445.62 | 460.42 | 482.09 | **1,853.44** |
| 215 | 1,542.18 | 1,429.25 | 1,476.49 | 1,435.88 | **5,883.78** |
| 216 | 1,162.22 | 1,045.02 | 1,165.28 | 1,134.30 | **4,506.82** |
| 217 | 2,808.34 | 2,641.62 | 2,761.54 | 2,674.22 | **10,885.73** |
| 218 | 1,033.48 | 1,006.53 | 1,010.11 | 1,002.50 | **4,052.62** |
| 219 | 440.22 | 427.94 | 414.87 | 416.43 | **1,699.47** |
| **MD - MARYLAND - Total** | **24,372.82** | **22,012.34** | **23,551.05** | **22,903.33** | **92,839.54** |

**STATE: ME - MAINE**

| REGISTRANT ZIP CODE 3 | 1ST QUARTER | 2ND QUARTER | 3RD QUARTER | 4TH QUARTER | TOTAL GRAMS |
|---|---|---|---|---|---|
| 039 | 156.79 | 123.70 | 116.05 | 120.15 | **516.69** |
| 040 | 3,123.71 | 3,092.57 | 3,231.08 | 3,265.46 | **12,712.82** |
| 041 | 674.30 | 623.92 | 649.94 | 657.92 | **2,606.08** |
| 042 | 1,595.60 | 1,493.83 | 1,527.56 | 1,532.11 | **6,149.10** |
| 043 | 1,466.06 | 1,378.77 | 1,454.25 | 1,392.96 | **5,692.04** |
| 044 | 1,343.03 | 1,252.73 | 1,322.01 | 1,288.36 | **5,206.13** |
| 045 | 448.21 | 463.20 | 479.44 | 490.78 | **1,881.63** |
| 046 | 789.70 | 796.78 | 832.10 | 814.96 | **3,233.54** |
| 047 | 882.46 | 879.21 | 870.13 | 867.22 | **3,499.03** |
| 048 | 384.61 | 356.66 | 386.16 | 358.28 | **1,485.72** |
| 049 | 1,745.23 | 1,669.63 | 1,657.18 | 1,647.06 | **6,719.11** |
| **ME - MAINE - Total** | **12,609.69** | **12,131.00** | **12,525.92** | **12,435.28** | **49,701.88** |

**STATE: MI - MICHIGAN**

| REGISTRANT ZIP CODE 3 | 1ST QUARTER | 2ND QUARTER | 3RD QUARTER | 4TH QUARTER | TOTAL GRAMS |
|---|---|---|---|---|---|
| 480 | 34,106.53 | 31,822.32 | 33,193.80 | 33,638.97 | **132,761.63** |
| 481 | 42,887.42 | 39,470.67 | 41,363.93 | 40,679.05 | **164,401.06** |
| 482 | 15,747.59 | 14,039.22 | 14,467.54 | 14,569.92 | **58,824.27** |
| 483 | 21,687.32 | 20,394.43 | 21,495.90 | 21,743.84 | **85,321.49** |
| 484 | 11,916.62 | 11,942.73 | 12,266.39 | 12,216.40 | **48,342.14** |
| 485 | 6,729.13 | 6,340.07 | 6,761.54 | 6,658.22 | **26,488.95** |
| 486 | 12,741.87 | 12,241.96 | 12,691.22 | 12,583.07 | **50,258.12** |
| 487 | 6,416.76 | 6,334.56 | 6,481.71 | 6,335.04 | **25,568.07** |
| 488 | 12,513.89 | 12,333.32 | 12,606.30 | 12,627.01 | **50,080.53** |
| 489 | 4,835.43 | 4,735.84 | 4,897.30 | 4,919.81 | **19,388.38** |
| 490 | 17,185.45 | 16,311.33 | 17,111.79 | 16,852.48 | **67,461.06** |
| 491 | 1,656.52 | 1,649.50 | 1,576.49 | 1,597.70 | **6,480.21** |
| 492 | 7,043.13 | 6,717.01 | 6,842.07 | 6,977.98 | **27,580.19** |
| 493 | 5,694.76 | 5,509.38 | 5,546.73 | 5,625.90 | **22,376.78** |
| 494 | 13,033.53 | 12,850.33 | 12,901.80 | 13,125.13 | **51,910.78** |
| 495 | 9,154.16 | 8,987.72 | 9,175.41 | 9,112.36 | **36,429.65** |
| 496 | 6,618.37 | 6,517.89 | 6,864.87 | 6,594.76 | **26,595.88** |
| 497 | 5,501.59 | 5,508.26 | 5,845.47 | 5,702.62 | **22,557.94** |
| 498 | 5,050.70 | 4,725.51 | 4,869.59 | 4,723.33 | **19,369.13** |
| 499 | 1,436.57 | 1,420.70 | 1,484.51 | 1,552.84 | **5,894.62** |
| **MI - MICHIGAN - Total** | **241,957.37** | **229,852.72** | **238,444.37** | **237,836.43** | **948,090.89** |

**STATE: MN - MINNESOTA**

| REGISTRANT ZIP CODE 3 | 1ST QUARTER | 2ND QUARTER | 3RD QUARTER | 4TH QUARTER | TOTAL GRAMS |
|---|---|---|---|---|---|
| 550 | 3,029.80 | 2,930.72 | 3,070.59 | 3,035.17 | **12,066.28** |
| 551 | 3,066.64 | 3,007.57 | 3,076.03 | 3,202.96 | **12,353.19** |
| 553 | 4,008.45 | 3,867.43 | 4,030.95 | 4,045.53 | **15,952.36** |
| 554 | 4,872.84 | 4,609.55 | 4,828.53 | 4,795.66 | **19,106.59** |
| 556 | 157.66 | 154.31 | 131.80 | 142.54 | **586.31** |
| 557 | 2,329.08 | 2,221.71 | 2,369.05 | 2,272.82 | **9,192.65** |
| 558 | 1,446.83 | 1,332.35 | 1,506.92 | 1,350.42 | **5,636.53** |
| 559 | 1,158.37 | 1,092.78 | 1,185.33 | 1,158.54 | **4,595.02** |

| | | | | | |
|---|---|---|---|---|---|
| 560 | 1,189.26 | 1,153.38 | 1,196.71 | 1,217.49 | **4,756.83** |
| 561 | 480.61 | 384.56 | 407.27 | 429.99 | **1,702.42** |
| 562 | 958.06 | 836.19 | 901.08 | 926.24 | **3,621.57** |
| 563 | 2,240.89 | 2,003.70 | 2,156.29 | 2,082.17 | **8,483.04** |
| 564 | 1,134.63 | 1,097.93 | 1,167.12 | 1,174.25 | **4,573.94** |
| 565 | 1,263.34 | 1,171.84 | 1,250.17 | 1,248.03 | **4,933.39** |
| 566 | 859.88 | 797.57 | 884.76 | 848.99 | **3,391.20** |
| 567 | 559.97 | 510.39 | 532.16 | 514.98 | **2,117.50** |
| **MN - MINNESOTA - Total** | **28,756.30** | **27,171.98** | **28,694.76** | **28,445.78** | **113,068.82** |

**STATE: MO - MISSOURI**

| REGISTRANT ZIP CODE 3 | 1ST QUARTER | 2ND QUARTER | 3RD QUARTER | 4TH QUARTER | TOTAL GRAMS |
|---|---|---|---|---|---|
| 630 | 11,594.09 | 10,625.09 | 11,377.88 | 11,320.73 | **44,917.79** |
| 631 | 18,010.85 | 13,931.31 | 15,418.58 | 15,731.73 | **63,092.47** |
| 633 | 6,868.52 | 6,614.40 | 6,950.66 | 6,786.66 | **27,220.24** |
| 634 | 1,527.51 | 1,538.71 | 1,524.14 | 1,409.32 | **5,999.68** |
| 635 | 1,162.80 | 1,143.34 | 1,200.76 | 1,133.60 | **4,640.50** |
| 636 | 4,520.48 | 4,481.81 | 4,574.42 | 4,457.21 | **18,033.91** |
| 637 | 3,008.91 | 2,848.88 | 3,109.61 | 2,945.99 | **11,913.39** |
| 638 | 6,399.55 | 6,191.00 | 6,241.52 | 6,271.84 | **25,103.92** |
| 639 | 3,975.74 | 3,893.53 | 3,904.51 | 3,914.80 | **15,688.59** |
| 640 | 12,369.97 | 11,880.25 | 12,485.60 | 12,083.19 | **48,819.02** |
| 641 | 8,083.37 | 7,488.74 | 7,808.38 | 7,584.60 | **30,965.09** |
| 644 | 1,437.27 | 1,335.66 | 1,416.24 | 1,408.65 | **5,597.81** |
| 645 | 2,851.54 | 2,692.73 | 2,824.36 | 2,746.10 | **11,114.72** |
| 646 | 1,004.46 | 964.16 | 981.83 | 938.14 | **3,888.59** |
| 647 | 3,000.21 | 2,875.36 | 2,990.52 | 2,952.14 | **11,818.23** |
| 648 | 4,695.56 | 4,563.85 | 4,663.74 | 4,559.20 | **18,482.36** |
| 650 | 2,311.82 | 2,291.06 | 2,391.84 | 2,286.52 | **9,281.24** |
| 651 | 1,017.99 | 976.94 | 1,108.81 | 1,046.35 | **4,150.08** |
| 652 | 3,128.16 | 2,998.43 | 3,125.23 | 3,160.02 | **12,411.84** |
| 653 | 1,721.37 | 1,622.78 | 1,744.73 | 1,714.51 | **6,803.38** |
| 654 | 1,740.96 | 1,626.29 | 1,714.75 | 1,719.64 | **6,801.64** |
| 655 | 2,828.38 | 2,896.58 | 2,795.69 | 2,852.09 | **11,372.74** |
| 656 | 5,308.48 | 5,233.37 | 5,498.33 | 5,436.33 | **21,476.50** |
| 657 | 7,737.87 | 7,638.58 | 7,719.67 | 7,638.52 | **30,734.63** |
| 658 | 6,882.53 | 6,415.17 | 6,544.27 | 6,730.38 | **26,572.35** |
| **MO - MISSOURI - Total** | **123,188.40** | **114,768.01** | **120,116.06** | **118,828.24** | **476,900.71** |

**STATE: MS - MISSISSIPPI**

| REGISTRANT ZIP CODE 3 | 1ST QUARTER | 2ND QUARTER | 3RD QUARTER | 4TH QUARTER | TOTAL GRAMS |
|---|---|---|---|---|---|
| 386 | 7,111.95 | 6,660.49 | 6,726.17 | 6,937.81 | **27,436.42** |
| 387 | 2,087.17 | 1,952.15 | 2,012.48 | 2,077.94 | **8,129.74** |
| 388 | 9,303.32 | 8,984.13 | 8,968.10 | 9,050.46 | **36,306.00** |
| 389 | 2,349.33 | 2,288.68 | 2,370.45 | 2,318.00 | **9,326.47** |
| 390 | 4,801.44 | 4,515.22 | 4,787.51 | 4,723.44 | **18,827.61** |
| 391 | 5,644.29 | 5,225.02 | 5,481.56 | 5,359.85 | **21,710.72** |
| 392 | 4,378.62 | 4,032.89 | 4,404.82 | 4,392.50 | **17,208.83** |
| 393 | 6,772.07 | 6,424.39 | 6,786.14 | 6,764.54 | **26,747.15** |
| 394 | 13,809.30 | 13,462.25 | 14,000.28 | 13,820.38 | **55,092.20** |
| 395 | 11,359.08 | 10,354.44 | 10,731.44 | 10,884.11 | **43,329.08** |
| 396 | 3,651.80 | 3,423.33 | 3,690.45 | 3,716.76 | **14,482.33** |
| 397 | 3,277.14 | 3,034.23 | 3,274.25 | 3,240.66 | **12,826.28** |
| **MS - MISSISSIPPI - Total** | **74,545.51** | **70,357.22** | **73,233.65** | **73,286.45** | **291,422.83** |

**STATE: MT - MONTANA**

| REGISTRANT ZIP CODE 3 | 1ST QUARTER | 2ND QUARTER | 3RD QUARTER | 4TH QUARTER | TOTAL GRAMS |
|---|---|---|---|---|---|
| 590 | 942.53 | 945.48 | 929.98 | 919.90 | **3,737.89** |
| 591 | 2,717.66 | 2,504.49 | 2,656.36 | 2,659.77 | **10,538.28** |
| 592 | 425.06 | 372.28 | 405.66 | 401.81 | **1,604.81** |
| 593 | 551.30 | 577.71 | 585.12 | 555.04 | **2,269.17** |
| 594 | 2,427.48 | 2,375.75 | 2,427.31 | 2,312.56 | **9,543.10** |
| 595 | 449.98 | 439.48 | 405.82 | 447.75 | **1,743.03** |

| | | | | | |
|---|---|---|---|---|---|
| 596 | 1,738.68 | 1,820.56 | 1,730.87 | 1,750.16 | **7,040.27** |
| 597 | 2,206.97 | 2,073.22 | 2,220.96 | 2,273.14 | **8,774.28** |
| 598 | 4,617.12 | 4,499.98 | 4,429.24 | 4,432.12 | **17,978.46** |
| 599 | 3,136.21 | 3,015.33 | 3,202.65 | 3,144.34 | **12,498.52** |
| **MT - MONTANA - Total** | **19,213.00** | **18,624.27** | **18,993.96** | **18,896.59** | **75,727.82** |

**STATE: NC - NORTH CAROLINA**

| REGISTRANT ZIP CODE 3 | 1ST QUARTER | 2ND QUARTER | 3RD QUARTER | 4TH QUARTER | TOTAL GRAMS |
|---|---|---|---|---|---|
| 270 | 5,591.61 | 5,083.72 | 5,248.71 | 5,271.85 | **21,195.90** |
| 271 | 3,720.32 | 3,294.13 | 3,452.42 | 3,420.19 | **13,887.06** |
| 272 | 9,167.35 | 8,326.10 | 8,683.80 | 8,686.25 | **34,863.50** |
| 273 | 5,543.75 | 5,081.05 | 5,211.48 | 5,229.24 | **21,065.52** |
| 274 | 3,406.05 | 3,022.42 | 3,253.04 | 3,479.19 | **13,160.70** |
| 275 | 10,502.77 | 9,279.10 | 9,805.34 | 9,892.86 | **39,480.07** |
| 276 | 3,260.48 | 2,718.86 | 2,870.42 | 2,976.78 | **11,826.54** |
| 277 | 2,376.25 | 2,295.46 | 2,033.17 | 2,243.19 | **8,948.07** |
| 278 | 6,786.66 | 6,056.20 | 6,410.15 | 6,363.42 | **25,616.43** |
| 279 | 2,023.02 | 1,882.61 | 2,027.96 | 1,963.64 | **7,897.23** |
| 280 | 15,349.17 | 14,173.42 | 14,573.91 | 14,669.75 | **58,766.26** |
| 281 | 11,149.75 | 10,223.36 | 10,465.03 | 10,868.33 | **42,706.47** |
| 282 | 7,806.66 | 6,739.54 | 6,893.35 | 6,696.37 | **28,135.91** |
| 283 | 12,398.99 | 12,083.85 | 12,412.55 | 12,011.47 | **48,906.85** |
| 284 | 7,906.64 | 7,283.98 | 7,643.03 | 7,537.38 | **30,371.02** |
| 285 | 6,375.98 | 5,747.21 | 5,772.26 | 5,749.03 | **23,644.49** |
| 286 | 13,864.75 | 13,245.23 | 13,147.02 | 13,184.86 | **53,441.86** |
| 287 | 7,459.21 | 7,113.15 | 7,293.04 | 7,361.16 | **29,226.56** |
| 288 | 3,366.91 | 2,902.43 | 3,021.00 | 3,041.13 | **12,331.48** |
| 289 | 1,368.51 | 1,434.48 | 1,428.38 | 1,385.04 | **5,616.40** |
| **NC - NORTH CAROLINA - Total** | **139,424.82** | **127,986.31** | **131,646.05** | **132,031.14** | **531,088.32** |

**STATE: ND - NORTH DAKOTA**

| REGISTRANT ZIP CODE 3 | 1ST QUARTER | 2ND QUARTER | 3RD QUARTER | 4TH QUARTER | TOTAL GRAMS |
|---|---|---|---|---|---|
| 580 | 419.01 | 368.28 | 369.84 | 387.75 | **1,544.88** |
| 581 | 1,241.76 | 1,109.63 | 1,111.25 | 1,131.07 | **4,593.70** |
| 582 | 808.09 | 712.84 | 672.78 | 764.98 | **2,958.69** |
| 583 | 485.10 | 458.84 | 455.34 | 465.54 | **1,864.81** |
| 584 | 419.86 | 358.37 | 370.23 | 385.61 | **1,534.06** |
| 585 | 2,195.24 | 2,066.21 | 2,114.86 | 2,173.93 | **8,550.25** |
| 586 | 548.38 | 526.14 | 534.51 | 552.51 | **2,161.54** |
| 587 | 566.80 | 517.10 | 552.56 | 572.67 | **2,209.13** |
| 588 | 456.42 | 441.26 | 410.32 | 453.10 | **1,761.10** |
| **ND - NORTH DAKOTA - Total** | **7,140.67** | **6,558.66** | **6,591.69** | **6,887.15** | **27,178.17** |

**STATE: NE - NEBRASKA**

| REGISTRANT ZIP CODE 3 | 1ST QUARTER | 2ND QUARTER | 3RD QUARTER | 4TH QUARTER | TOTAL GRAMS |
|---|---|---|---|---|---|
| 680 | 2,560.19 | 2,415.90 | 2,391.24 | 2,373.73 | **9,741.06** |
| 681 | 7,041.94 | 6,469.20 | 6,776.83 | 6,707.86 | **26,995.83** |
| 683 | 1,004.83 | 952.10 | 908.33 | 979.66 | **3,844.92** |
| 684 | 763.49 | 680.64 | 671.98 | 729.66 | **2,845.77** |
| 685 | 3,433.75 | 3,204.12 | 3,464.49 | 3,203.69 | **13,306.06** |
| 686 | 566.32 | 536.07 | 569.97 | 534.67 | **2,207.03** |
| 687 | 1,309.32 | 1,202.09 | 1,318.63 | 1,268.59 | **5,098.64** |
| 688 | 2,104.54 | 1,943.57 | 2,113.52 | 2,151.85 | **8,313.49** |
| 689 | 888.70 | 829.92 | 848.42 | 877.73 | **3,444.76** |
| 690 | 350.05 | 313.32 | 348.45 | 312.56 | **1,324.39** |
| 691 | 997.90 | 984.73 | 1,014.37 | 988.11 | **3,985.10** |
| 692 | 63.42 | 67.27 | 62.22 | 57.01 | **249.91** |
| 693 | 810.61 | 811.47 | 833.89 | 806.24 | **3,262.22** |
| **NE - NEBRASKA - Total** | **21,895.07** | **20,410.41** | **21,322.35** | **20,991.36** | **84,619.19** |

**STATE: NH - NEW HAMPSHIRE**

| REGISTRANT ZIP CODE 3 | 1ST QUARTER | 2ND QUARTER | 3RD QUARTER | 4TH QUARTER | TOTAL GRAMS |
|---|---|---|---|---|---|
| 030 | 1,592.96 | 1,444.34 | 1,540.59 | 1,534.34 | **6,112.22** |
| 031 | 1,006.24 | 849.00 | 879.56 | 929.33 | **3,664.14** |

| | | | | | |
|---|---|---|---|---|---|
| 032 | 832.55 | 756.47 | 802.76 | 801.86 | **3,193.65** |
| 033 | 533.39 | 455.32 | 500.09 | 506.73 | **1,995.53** |
| 034 | 404.65 | 414.65 | 410.36 | 417.70 | **1,647.36** |
| 035 | 272.17 | 252.35 | 267.95 | 259.68 | **1,052.16** |
| 036 | 27.51 | 26.21 | 26.41 | 20.97 | **101.11** |
| 037 | 321.04 | 289.30 | 319.08 | 334.30 | **1,263.72** |
| 038 | 1,802.43 | 1,706.19 | 1,768.39 | 1,751.45 | **7,028.46** |
| **NH - NEW HAMPSHIRE - Total** | **6,792.94** | **6,193.85** | **6,515.19** | **6,556.37** | **26,058.35** |

**STATE: NJ - NEW JERSEY**

| REGISTRANT ZIP CODE 3 | 1ST QUARTER | 2ND QUARTER | 3RD QUARTER | 4TH QUARTER | TOTAL GRAMS |
|---|---|---|---|---|---|
| 070 | 2,677.35 | 2,198.19 | 2,332.99 | 2,337.35 | **9,545.88** |
| 071 | 220.54 | 195.04 | 217.41 | 216.87 | **849.86** |
| 072 | 167.17 | 129.72 | 140.73 | 116.13 | **553.75** |
| 073 | 209.83 | 209.73 | 210.59 | 187.23 | **817.37** |
| 074 | 1,587.52 | 1,371.38 | 1,382.53 | 1,375.24 | **5,716.67** |
| 075 | 191.41 | 121.35 | 170.21 | 145.44 | **628.41** |
| 076 | 1,098.43 | 856.46 | 907.89 | 935.18 | **3,797.96** |
| 077 | 2,041.05 | 1,808.72 | 2,001.00 | 1,932.64 | **7,783.40** |
| 078 | 1,361.27 | 1,199.09 | 1,247.95 | 1,241.44 | **5,049.75** |
| 079 | 467.29 | 380.10 | 393.16 | 405.39 | **1,645.94** |
| 080 | 5,954.01 | 5,665.42 | 5,923.92 | 6,044.46 | **23,587.82** |
| 081 | 389.70 | 358.49 | 358.24 | 354.39 | **1,460.83** |
| 082 | 1,527.11 | 1,428.34 | 1,566.41 | 1,411.40 | **5,933.26** |
| 083 | 1,383.25 | 1,293.02 | 1,349.01 | 1,340.37 | **5,365.65** |
| 084 | 224.79 | 214.42 | 228.87 | 215.26 | **883.35** |
| 085 | 642.21 | 593.72 | 565.91 | 555.37 | **2,357.20** |
| 086 | 717.59 | 659.93 | 686.00 | 711.69 | **2,775.21** |
| 087 | 1,747.68 | 1,590.05 | 1,723.52 | 1,704.84 | **6,766.10** |
| 088 | 1,944.22 | 1,584.24 | 1,749.24 | 1,708.29 | **6,985.99** |
| 089 | 120.59 | 93.88 | 89.85 | 90.51 | **394.83** |
| **NJ - NEW JERSEY - Total** | **24,673.01** | **21,951.28** | **23,245.44** | **23,029.50** | **92,899.23** |

**STATE: NM - NEW MEXICO**

| REGISTRANT ZIP CODE 3 | 1ST QUARTER | 2ND QUARTER | 3RD QUARTER | 4TH QUARTER | TOTAL GRAMS |
|---|---|---|---|---|---|
| 870 | 1,926.28 | 1,873.75 | 1,915.38 | 1,869.49 | **7,584.90** |
| 871 | 8,456.81 | 7,912.04 | 8,151.31 | 8,023.58 | **32,543.74** |
| 873 | 450.95 | 394.92 | 382.57 | 405.08 | **1,633.52** |
| 874 | 1,138.34 | 1,015.20 | 1,133.02 | 1,171.08 | **4,457.63** |
| 875 | 3,053.40 | 2,897.59 | 3,014.26 | 2,910.29 | **11,875.53** |
| 877 | 713.21 | 746.67 | 725.59 | 718.08 | **2,903.55** |
| 878 | 210.68 | 195.48 | 205.38 | 202.69 | **814.24** |
| 879 | 176.06 | 192.18 | 174.32 | 184.17 | **726.72** |
| 880 | 2,189.41 | 2,115.89 | 2,166.19 | 2,162.34 | **8,633.84** |
| 881 | 1,064.46 | 1,034.55 | 1,080.86 | 1,075.89 | **4,255.75** |
| 882 | 4,783.68 | 4,580.74 | 4,727.33 | 4,669.52 | **18,761.27** |
| 883 | 1,251.20 | 1,172.97 | 1,238.72 | 1,290.78 | **4,953.67** |
| 884 | 238.61 | 243.16 | 229.59 | 243.49 | **954.86** |
| **NM - NEW MEXICO - Total** | **25,653.08** | **24,375.13** | **25,144.51** | **24,926.49** | **100,099.20** |

**STATE: NV - NEVADA**

| REGISTRANT ZIP CODE 3 | 1ST QUARTER | 2ND QUARTER | 3RD QUARTER | 4TH QUARTER | TOTAL GRAMS |
|---|---|---|---|---|---|
| 890 | 18,002.05 | 17,758.90 | 18,316.06 | 17,071.74 | **71,148.75** |
| 891 | 32,292.92 | 31,863.89 | 32,383.68 | 31,626.64 | **128,167.13** |
| 893 | 367.16 | 339.35 | 370.49 | 386.36 | **1,463.35** |
| 894 | 5,987.06 | 5,852.86 | 5,981.43 | 5,929.06 | **23,750.42** |
| 895 | 5,537.21 | 5,340.42 | 5,491.33 | 5,456.70 | **21,825.65** |
| 897 | 1,944.28 | 1,865.49 | 1,914.04 | 1,888.60 | **7,612.41** |
| 898 | 768.45 | 687.82 | 720.44 | 722.21 | **2,898.92** |
| **NV - NEVADA - Total** | **64,899.14** | **63,708.72** | **65,177.46** | **63,081.32** | **256,866.63** |

**STATE: NY - NEW YORK**

| REGISTRANT ZIP CODE 3 | 1ST QUARTER | 2ND QUARTER | 3RD QUARTER | 4TH QUARTER | TOTAL GRAMS |
|---|---|---|---|---|---|
| 063 | 0 | 0.30 | 0 | 0.30 | **0.61** |

| | | | | | |
|---|---|---|---|---|---|
| 100 | 2,605.51 | 1,845.94 | 2,019.85 | 2,034.81 | **8,506.11** |
| 101 | 275.63 | 57.57 | 129.73 | 71.55 | **534.49** |
| 102 | 6.32 | 5.02 | 2.42 | 8.29 | **22.06** |
| 103 | 1,928.94 | 1,765.01 | 1,839.10 | 1,786.06 | **7,319.11** |
| 104 | 895.09 | 829.06 | 829.38 | 763.24 | **3,316.77** |
| 105 | 1,746.57 | 1,534.63 | 1,636.11 | 1,587.97 | **6,505.27** |
| 106 | 218.01 | 162.41 | 168.79 | 182.04 | **731.25** |
| 107 | 476.82 | 501.14 | 497.76 | 512.66 | **1,988.38** |
| 108 | 154.54 | 153.18 | 138.02 | 127.06 | **572.80** |
| 109 | 1,886.90 | 1,706.55 | 1,723.96 | 1,775.16 | **7,092.56** |
| 110 | 694.86 | 620.08 | 645.48 | 621.84 | **2,582.27** |
| 111 | 282.07 | 251.10 | 232.03 | 230.26 | **995.45** |
| 112 | 2,606.49 | 2,231.97 | 2,326.97 | 2,149.36 | **9,314.79** |
| 113 | 1,367.95 | 1,265.20 | 1,276.96 | 1,232.02 | **5,142.12** |
| 114 | 648.90 | 562.97 | 617.29 | 587.64 | **2,416.80** |
| 115 | 2,036.58 | 1,781.56 | 1,889.03 | 1,912.51 | **7,619.68** |
| 116 | 188.07 | 157.33 | 175.06 | 161.09 | **681.54** |
| 117 | 9,114.96 | 8,141.69 | 8,398.86 | 8,448.67 | **34,104.17** |
| 118 | 274.09 | 272.93 | 275.71 | 272.08 | **1,094.81** |
| 119 | 1,455.97 | 1,394.49 | 1,432.64 | 1,411.17 | **5,694.28** |
| 120 | 4,611.63 | 4,433.35 | 4,482.09 | 4,317.54 | **17,844.62** |
| 121 | 2,810.75 | 2,752.91 | 2,802.46 | 2,771.46 | **11,137.57** |
| 122 | 2,014.03 | 1,793.81 | 1,869.87 | 1,741.78 | **7,419.49** |
| 123 | 1,950.18 | 1,920.24 | 2,080.02 | 1,982.22 | **7,932.66** |
| 124 | 1,789.83 | 1,783.84 | 1,681.36 | 1,730.73 | **6,985.76** |
| 125 | 3,230.68 | 2,975.82 | 3,088.86 | 2,918.49 | **12,213.85** |
| 126 | 498.99 | 438.06 | 478.17 | 471.88 | **1,887.09** |
| 127 | 599.66 | 622.85 | 635.09 | 623.84 | **2,481.44** |
| 128 | 3,091.84 | 2,947.89 | 3,071.26 | 2,888.41 | **11,999.40** |
| 129 | 2,183.60 | 2,170.24 | 2,245.80 | 2,274.89 | **8,874.54** |
| 130 | 4,163.83 | 3,848.65 | 4,061.75 | 4,077.81 | **16,152.05** |
| 131 | 1,465.44 | 1,419.69 | 1,454.39 | 1,458.68 | **5,798.20** |
| 132 | 2,592.66 | 2,508.51 | 2,396.28 | 2,538.92 | **10,036.37** |
| 133 | 1,728.53 | 1,626.56 | 1,664.81 | 1,706.66 | **6,726.56** |
| 134 | 2,364.26 | 2,182.20 | 2,231.28 | 2,235.80 | **9,013.54** |
| 135 | 1,421.79 | 1,173.81 | 1,320.76 | 1,276.35 | **5,192.71** |
| 136 | 2,645.77 | 2,520.01 | 2,593.24 | 2,563.13 | **10,322.15** |
| 137 | 1,102.52 | 1,052.86 | 1,043.82 | 1,043.81 | **4,243.03** |
| 138 | 2,451.55 | 2,150.99 | 2,203.75 | 2,227.54 | **9,033.83** |
| 139 | 1,068.03 | 1,013.04 | 1,038.17 | 1,027.73 | **4,146.97** |
| 140 | 8,550.29 | 8,340.54 | 8,546.70 | 8,524.11 | **33,961.64** |
| 141 | 4,039.55 | 4,117.59 | 4,135.89 | 4,076.71 | **16,369.74** |
| 142 | 16,197.40 | 15,596.77 | 15,985.37 | 15,623.25 | **63,402.79** |
| 143 | 2,104.67 | 2,022.09 | 2,035.25 | 2,012.94 | **8,174.95** |
| 144 | 2,970.69 | 2,853.04 | 2,907.58 | 2,815.74 | **11,547.05** |
| 145 | 2,339.06 | 2,137.19 | 2,235.59 | 2,133.90 | **8,845.73** |
| 146 | 4,009.79 | 3,792.71 | 3,911.52 | 3,840.26 | **15,554.29** |
| 147 | 3,539.54 | 3,321.42 | 3,508.09 | 3,536.47 | **13,905.53** |
| 148 | 2,860.74 | 2,835.25 | 2,846.21 | 2,843.38 | **11,385.60** |
| 149 | 692.69 | 694.13 | 648.16 | 649.59 | **2,684.56** |
| **NY - NEW YORK - Total** | **119,954.27** | **112,286.20** | **115,458.76** | **113,809.81** | **461,509.04** |

**STATE: OH - OHIO**

| REGISTRANT ZIP CODE 3 | 1ST QUARTER | 2ND QUARTER | 3RD QUARTER | 4TH QUARTER | **TOTAL GRAMS** |
|---|---|---|---|---|---|
| 430 | 6,824.55 | 6,302.47 | 6,769.78 | 6,713.33 | **26,610.12** |
| 431 | 5,324.15 | 5,083.42 | 5,326.73 | 5,240.72 | **20,975.02** |
| 432 | 7,701.86 | 7,074.18 | 7,327.12 | 7,499.04 | **29,602.20** |
| 433 | 2,052.47 | 2,015.36 | 2,123.83 | 2,160.27 | **8,351.94** |
| 434 | 1,091.49 | 1,095.72 | 1,142.67 | 1,130.52 | **4,460.40** |
| 435 | 2,559.59 | 2,468.23 | 2,614.48 | 2,658.54 | **10,300.84** |
| 436 | 4,450.75 | 4,173.98 | 4,457.14 | 4,493.44 | **17,575.30** |

| | | | | | |
|---|---|---|---|---|---|
| 437 | 2,557.09 | 2,453.28 | 2,559.30 | 2,443.42 | **10,013.08** |
| 438 | 400.98 | 354.01 | 422.35 | 375.62 | **1,552.96** |
| 439 | 1,902.27 | 1,842.31 | 1,986.20 | 1,886.76 | **7,617.54** |
| 440 | 5,429.70 | 5,101.41 | 5,351.47 | 5,386.24 | **21,268.82** |
| 441 | 6,952.12 | 6,421.73 | 6,788.14 | 6,780.12 | **26,942.11** |
| 442 | 4,144.92 | 3,977.51 | 4,127.78 | 4,226.96 | **16,477.17** |
| 443 | 1,932.97 | 1,771.48 | 1,941.83 | 1,861.13 | **7,507.42** |
| 444 | 4,780.42 | 4,616.45 | 4,590.30 | 4,570.18 | **18,557.35** |
| 445 | 3,162.53 | 2,956.95 | 3,075.78 | 3,084.96 | **12,280.23** |
| 446 | 3,806.85 | 3,580.80 | 3,811.31 | 3,768.51 | **14,967.47** |
| 447 | 2,324.07 | 2,185.39 | 2,273.08 | 2,239.58 | **9,022.13** |
| 448 | 2,205.84 | 2,257.39 | 2,286.22 | 2,298.45 | **9,047.89** |
| 449 | 672.23 | 661.99 | 728.95 | 699.99 | **2,763.17** |
| 450 | 11,061.30 | 11,013.36 | 11,702.90 | 11,630.83 | **45,408.40** |
| 451 | 3,856.42 | 3,690.17 | 3,825.91 | 3,792.33 | **15,164.84** |
| 452 | 7,460.00 | 6,972.11 | 7,364.52 | 7,395.26 | **29,191.89** |
| 453 | 7,095.34 | 6,791.28 | 7,045.68 | 7,005.60 | **27,937.89** |
| 454 | 7,072.20 | 6,896.34 | 7,016.06 | 7,020.15 | **28,004.74** |
| 455 | 1,968.63 | 1,879.50 | 1,919.70 | 1,835.56 | **7,603.39** |
| 456 | 6,085.85 | 5,809.79 | 6,216.96 | 5,951.55 | **24,064.15** |
| 457 | 2,116.11 | 2,038.42 | 2,187.36 | 2,070.40 | **8,412.29** |
| 458 | 3,304.54 | 3,134.82 | 3,338.31 | 3,371.85 | **13,149.51** |
| **OH - OHIO - Total** | **120,297.26** | **114,619.85** | **120,321.86** | **119,591.30** | **474,830.27** |

**STATE: OK - OKLAHOMA**

| REGISTRANT ZIP CODE 3 | 1ST QUARTER | 2ND QUARTER | 3RD QUARTER | 4TH QUARTER | TOTAL GRAMS |
|---|---|---|---|---|---|
| 730 | 18,453.75 | 17,989.06 | 18,447.18 | 18,410.84 | **73,300.84** |
| 731 | 25,075.35 | 23,950.06 | 24,874.65 | 24,612.73 | **98,512.79** |
| 734 | 3,919.17 | 3,615.17 | 3,908.59 | 3,834.92 | **15,277.84** |
| 735 | 7,139.14 | 7,066.44 | 7,333.55 | 7,078.08 | **28,617.21** |
| 736 | 1,365.39 | 1,351.65 | 1,389.41 | 1,385.00 | **5,491.45** |
| 737 | 2,955.65 | 2,849.32 | 3,077.53 | 2,921.29 | **11,803.79** |
| 738 | 662.81 | 613.56 | 636.74 | 577.01 | **2,490.12** |
| 739 | 250.37 | 241.21 | 244.87 | 230.96 | **967.41** |
| 740 | 19,706.33 | 18,721.40 | 19,296.02 | 19,089.55 | **76,813.29** |
| 741 | 12,820.60 | 12,276.54 | 12,554.32 | 12,502.25 | **50,153.71** |
| 743 | 3,202.85 | 3,177.19 | 3,202.30 | 3,199.93 | **12,782.26** |
| 744 | 10,852.66 | 10,515.57 | 10,693.23 | 10,373.43 | **42,434.89** |
| 745 | 2,879.52 | 2,820.46 | 2,808.33 | 2,819.20 | **11,327.51** |
| 746 | 1,500.14 | 1,409.62 | 1,523.42 | 1,519.65 | **5,952.83** |
| 747 | 4,122.87 | 4,155.74 | 4,097.44 | 4,175.50 | **16,551.54** |
| 748 | 6,266.97 | 6,031.11 | 6,126.72 | 5,991.28 | **24,416.09** |
| 749 | 3,803.22 | 3,517.14 | 3,698.29 | 3,648.34 | **14,666.99** |
| **OK - OKLAHOMA - Total** | **124,976.79** | **120,301.25** | **123,912.60** | **122,369.92** | **491,560.56** |

**STATE: OR - OREGON**

| REGISTRANT ZIP CODE 3 | 1ST QUARTER | 2ND QUARTER | 3RD QUARTER | 4TH QUARTER | TOTAL GRAMS |
|---|---|---|---|---|---|
| 970 | 10,337.98 | 9,816.80 | 10,123.60 | 10,048.89 | **40,327.27** |
| 971 | 4,397.27 | 3,914.10 | 4,268.43 | 4,178.71 | **16,758.51** |
| 972 | 12,645.68 | 12,804.41 | 13,094.73 | 13,022.30 | **51,567.11** |
| 973 | 8,429.89 | 7,939.30 | 8,185.37 | 8,155.35 | **32,709.90** |
| 974 | 12,738.47 | 12,467.92 | 12,770.91 | 12,706.22 | **50,683.52** |
| 975 | 9,322.57 | 9,222.33 | 9,241.11 | 9,173.14 | **36,959.15** |
| 976 | 1,926.72 | 1,843.67 | 2,005.85 | 1,855.82 | **7,632.06** |
| 977 | 4,086.08 | 3,871.34 | 4,012.45 | 3,959.98 | **15,929.85** |
| 978 | 2,979.81 | 2,878.60 | 2,908.04 | 2,885.95 | **11,652.40** |
| 979 | 1,405.64 | 1,379.74 | 1,378.00 | 1,382.59 | **5,545.97** |
| **OR - OREGON - Total** | **68,270.10** | **66,138.20** | **67,988.50** | **67,368.96** | **269,765.75** |

**STATE: PA - PENNSYLVANIA**

| REGISTRANT ZIP CODE 3 | 1ST QUARTER | 2ND QUARTER | 3RD QUARTER | 4TH QUARTER | TOTAL GRAMS |
|---|---|---|---|---|---|
| 150 | 4,989.38 | 5,129.92 | 5,066.55 | 5,101.21 | **20,287.05** |
| 151 | 4,196.21 | 3,877.44 | 3,863.65 | 3,847.18 | **15,784.47** |

| | | | | | |
|---|---|---|---|---|---|
| 152 | 7,130.14 | 6,560.80 | 6,755.93 | 6,868.62 | **27,315.49** |
| 153 | 2,358.78 | 2,351.45 | 2,972.30 | 2,432.17 | **10,114.70** |
| 154 | 3,372.47 | 3,277.67 | 3,303.61 | 3,201.45 | **13,155.21** |
| 155 | 884.36 | 783.13 | 816.48 | 830.90 | **3,314.87** |
| 156 | 3,334.49 | 3,094.11 | 3,186.53 | 3,206.95 | **12,822.08** |
| 157 | 1,471.67 | 1,443.77 | 1,379.79 | 1,257.87 | **5,553.10** |
| 158 | 1,497.89 | 1,432.58 | 1,498.95 | 1,520.27 | **5,949.69** |
| 159 | 1,522.82 | 1,435.25 | 1,499.81 | 1,446.69 | **5,904.57** |
| 160 | 2,042.83 | 1,952.76 | 2,019.55 | 2,011.39 | **8,026.52** |
| 161 | 2,758.77 | 2,621.20 | 2,688.64 | 2,712.70 | **10,781.31** |
| 162 | 920.59 | 836.13 | 853.49 | 821.55 | **3,431.75** |
| 163 | 2,353.79 | 2,271.30 | 2,301.72 | 2,330.35 | **9,257.16** |
| 164 | 1,503.33 | 1,499.28 | 1,550.62 | 1,633.20 | **6,186.43** |
| 165 | 5,220.32 | 5,067.30 | 5,306.81 | 5,307.27 | **20,901.70** |
| 166 | 3,176.74 | 3,004.70 | 3,124.28 | 3,044.09 | **12,349.81** |
| 167 | 860.67 | 892.17 | 873.44 | 906.42 | **3,532.69** |
| 168 | 893.10 | 791.15 | 883.84 | 875.41 | **3,443.50** |
| 169 | 589.02 | 593.84 | 592.58 | 596.47 | **2,371.90** |
| 170 | 5,093.99 | 4,778.87 | 5,018.38 | 4,923.33 | **19,814.57** |
| 171 | 1,139.62 | 1,126.68 | 1,123.42 | 1,102.98 | **4,492.70** |
| 172 | 969.30 | 843.79 | 930.33 | 891.72 | **3,635.14** |
| 173 | 1,733.95 | 1,617.42 | 1,725.70 | 1,672.15 | **6,749.22** |
| 174 | 936.40 | 910.63 | 887.30 | 936.36 | **3,670.70** |
| 175 | 1,700.40 | 1,617.01 | 1,716.01 | 1,636.69 | **6,670.11** |
| 176 | 1,356.16 | 1,271.49 | 1,417.30 | 1,347.95 | **5,392.90** |
| 177 | 2,029.79 | 1,935.65 | 1,981.87 | 1,944.42 | **7,891.72** |
| 178 | 1,032.29 | 952.80 | 975.10 | 962.70 | **3,922.89** |
| 179 | 807.88 | 757.38 | 794.83 | 782.47 | **3,142.57** |
| 180 | 2,166.22 | 1,956.64 | 1,997.30 | 1,993.79 | **8,113.96** |
| 181 | 847.72 | 713.94 | 751.63 | 818.30 | **3,131.59** |
| 182 | 1,074.36 | 1,043.87 | 991.87 | 1,050.88 | **4,160.97** |
| 183 | 969.61 | 932.27 | 967.16 | 962.73 | **3,831.76** |
| 184 | 1,881.65 | 1,847.52 | 1,858.61 | 1,829.70 | **7,417.48** |
| 185 | 2,192.25 | 2,172.70 | 2,129.96 | 2,080.12 | **8,575.02** |
| 186 | 1,773.93 | 1,653.75 | 1,785.77 | 1,665.19 | **6,878.64** |
| 187 | 4,629.79 | 6,361.90 | 5,836.60 | 5,444.65 | **22,272.94** |
| 188 | 700.53 | 733.09 | 749.32 | 729.51 | **2,912.45** |
| 189 | 1,870.16 | 1,724.18 | 1,911.29 | 1,866.09 | **7,371.71** |
| 190 | 8,473.25 | 5,349.08 | 6,252.62 | 6,543.40 | **26,618.34** |
| 191 | 3,813.64 | 3,659.38 | 3,624.47 | 3,551.89 | **14,649.37** |
| 193 | 1,701.47 | 1,614.81 | 1,566.11 | 1,606.24 | **6,488.65** |
| 194 | 2,498.18 | 2,358.23 | 2,408.45 | 2,370.35 | **9,635.21** |
| 195 | 1,087.02 | 1,053.61 | 1,116.23 | 1,110.27 | **4,367.13** |
| 196 | 1,287.20 | 1,267.17 | 1,271.44 | 1,256.79 | **5,082.59** |
| **PA - PENNSYLVANIA - Total** | **104,844.13** | **99,169.77** | **102,327.61** | **101,032.83** | **407,374.34** |

**STATE: PR - PUERTO RICO**

| REGISTRANT ZIP CODE 3 | 1ST QUARTER | 2ND QUARTER | 3RD QUARTER | 4TH QUARTER | TOTAL GRAMS |
|---|---|---|---|---|---|
| 006 | 173.77 | 110.48 | 145.60 | 130.95 | **560.80** |
| 007 | 88.73 | 49.14 | 61.11 | 65.68 | **264.67** |
| 009 | 122.16 | 73.44 | 70.62 | 67.21 | **333.43** |
| **PR - PUERTO RICO - Total** | **384.66** | **233.06** | **277.33** | **263.84** | **1,158.90** |

**STATE: RI - RHODE ISLAND**

| REGISTRANT ZIP CODE 3 | 1ST QUARTER | 2ND QUARTER | 3RD QUARTER | 4TH QUARTER | TOTAL GRAMS |
|---|---|---|---|---|---|
| 028 | 4,332.52 | 4,147.01 | 4,353.24 | 4,122.77 | **16,955.54** |
| 029 | 2,785.38 | 2,638.15 | 2,774.43 | 2,626.69 | **10,824.65** |
| **RI - RHODE ISLAND - Total** | **7,117.89** | **6,785.16** | **7,127.67** | **6,749.46** | **27,780.18** |

**STATE: SC - SOUTH CAROLINA**

| REGISTRANT ZIP CODE 3 | 1ST QUARTER | 2ND QUARTER | 3RD QUARTER | 4TH QUARTER | TOTAL GRAMS |
|---|---|---|---|---|---|
| 290 | 5,861.54 | 5,506.07 | 5,570.91 | 5,759.04 | **22,697.56** |
| 291 | 6,250.03 | 5,715.75 | 5,826.52 | 6,025.38 | **23,817.68** |

| | | | | | |
|---|---|---|---|---|---|
| 292 | 4,431.79 | 3,882.31 | 4,119.65 | 4,208.21 | **16,641.96** |
| 293 | 13,399.98 | 12,887.52 | 13,203.79 | 13,273.08 | **52,764.37** |
| 294 | 13,806.11 | 12,611.13 | 13,090.78 | 13,124.89 | **52,632.91** |
| 295 | 16,144.15 | 14,497.23 | 15,268.94 | 14,940.33 | **60,850.65** |
| 296 | 25,333.78 | 23,760.33 | 24,437.04 | 24,770.05 | **98,301.19** |
| 297 | 4,592.75 | 4,016.84 | 4,461.83 | 4,417.10 | **17,488.52** |
| 298 | 2,706.44 | 2,537.48 | 2,525.99 | 2,483.34 | **10,253.25** |
| 299 | 2,441.21 | 2,159.79 | 2,303.73 | 2,292.01 | **9,196.75** |
| **SC - SOUTH CAROLINA - Total** | **94,967.78** | **87,574.46** | **90,809.17** | **91,293.44** | **364,644.86** |

**STATE: SD - SOUTH DAKOTA**

| REGISTRANT ZIP CODE 3 | 1ST QUARTER | 2ND QUARTER | 3RD QUARTER | 4TH QUARTER | TOTAL GRAMS |
|---|---|---|---|---|---|
| 570 | 1,710.89 | 1,362.34 | 1,540.92 | 1,538.26 | **6,152.41** |
| 571 | 2,851.52 | 2,563.14 | 2,753.07 | 2,574.88 | **10,742.61** |
| 572 | 682.54 | 623.44 | 649.52 | 669.18 | **2,624.68** |
| 573 | 798.23 | 773.59 | 758.97 | 801.49 | **3,132.27** |
| 574 | 571.14 | 499.99 | 507.85 | 540.92 | **2,119.90** |
| 575 | 704.53 | 645.26 | 646.91 | 659.74 | **2,656.44** |
| 576 | 166.60 | 144.04 | 128.41 | 146.23 | **585.28** |
| 577 | 2,064.97 | 1,879.89 | 2,014.60 | 1,931.34 | **7,890.80** |
| **SD - SOUTH DAKOTA - Total** | **9,550.42** | **8,491.70** | **9,000.24** | **8,862.03** | **35,904.38** |

**STATE: TN - TENNESSEE**

| REGISTRANT ZIP CODE 3 | 1ST QUARTER | 2ND QUARTER | 3RD QUARTER | 4TH QUARTER | TOTAL GRAMS |
|---|---|---|---|---|---|
| 370 | 20,427.42 | 19,925.93 | 20,490.86 | 20,052.71 | **80,896.91** |
| 371 | 20,984.22 | 20,799.54 | 21,001.96 | 21,127.02 | **83,912.74** |
| 372 | 8,602.54 | 7,985.23 | 8,208.27 | 8,260.54 | **33,056.58** |
| 373 | 20,029.23 | 19,169.28 | 19,635.77 | 19,518.78 | **78,353.06** |
| 374 | 6,900.48 | 6,710.74 | 6,983.98 | 6,820.41 | **27,415.61** |
| 376 | 12,419.92 | 11,803.08 | 11,975.03 | 11,835.04 | **48,033.06** |
| 377 | 10,348.39 | 9,944.69 | 9,925.67 | 10,207.91 | **40,426.66** |
| 378 | 15,895.77 | 15,477.06 | 15,656.09 | 15,867.77 | **62,896.69** |
| 379 | 7,841.32 | 7,449.32 | 7,566.91 | 7,489.72 | **30,347.26** |
| 380 | 9,121.34 | 8,922.83 | 8,889.20 | 8,997.61 | **35,930.98** |
| 381 | 11,887.71 | 8,377.29 | 9,772.17 | 10,425.91 | **40,463.07** |
| 382 | 5,086.25 | 4,862.79 | 4,926.60 | 4,649.70 | **19,525.34** |
| 383 | 11,413.60 | 10,789.42 | 10,974.85 | 11,240.51 | **44,418.39** |
| 384 | 6,270.47 | 6,105.44 | 5,923.15 | 5,844.73 | **24,143.78** |
| 385 | 5,418.84 | 5,199.33 | 5,316.68 | 5,263.89 | **21,198.74** |
| **TN - TENNESSEE - Total** | **172,647.48** | **163,521.98** | **167,247.19** | **167,602.22** | **671,018.86** |

**STATE: TX - TEXAS**

| REGISTRANT ZIP CODE 3 | 1ST QUARTER | 2ND QUARTER | 3RD QUARTER | 4TH QUARTER | TOTAL GRAMS |
|---|---|---|---|---|---|
| 750 | 38,162.87 | 36,659.30 | 37,622.64 | 37,935.70 | **150,380.50** |
| 751 | 17,449.72 | 17,527.65 | 18,135.54 | 18,046.34 | **71,159.26** |
| 752 | 18,657.21 | 17,460.53 | 17,998.35 | 17,921.37 | **72,037.46** |
| 753 | 238.61 | 146.84 | 175.21 | 203.23 | **763.89** |
| 754 | 11,378.72 | 11,082.03 | 11,345.98 | 11,420.09 | **45,226.83** |
| 755 | 3,490.72 | 3,411.83 | 3,501.13 | 3,365.01 | **13,768.69** |
| 756 | 8,640.23 | 8,761.29 | 8,839.16 | 8,777.17 | **35,017.85** |
| 757 | 12,073.18 | 11,407.91 | 11,917.50 | 11,720.05 | **47,118.64** |
| 758 | 2,192.78 | 2,224.99 | 2,297.18 | 2,269.42 | **8,984.37** |
| 759 | 5,796.55 | 5,505.53 | 5,713.47 | 5,619.85 | **22,635.40** |
| 760 | 30,647.92 | 29,789.48 | 31,026.51 | 30,952.66 | **122,416.58** |
| 761 | 24,454.42 | 22,825.06 | 23,645.51 | 23,609.36 | **94,534.36** |
| 762 | 10,787.93 | 10,512.63 | 10,928.65 | 11,055.39 | **43,284.61** |
| 763 | 4,742.91 | 4,227.58 | 4,537.25 | 4,692.12 | **18,199.87** |
| 764 | 3,430.39 | 3,360.04 | 3,577.71 | 3,513.26 | **13,881.40** |
| 765 | 9,006.87 | 8,594.49 | 8,848.50 | 8,604.37 | **35,054.23** |
| 766 | 2,731.09 | 2,692.37 | 2,888.44 | 2,759.16 | **11,071.06** |
| 767 | 7,576.12 | 7,257.94 | 7,238.62 | 7,164.64 | **29,237.33** |
| 768 | 2,432.13 | 2,428.44 | 2,410.15 | 2,431.69 | **9,702.41** |
| 769 | 2,508.44 | 2,422.33 | 2,486.16 | 2,372.70 | **9,789.64** |

| | | | | | |
|---|---|---|---|---|---|
| 770 | 39,664.08 | 37,051.25 | 36,655.55 | 34,524.44 | **147,895.32** |
| 772 | 0.91 | 0 | 0.61 | 0.91 | **2.42** |
| 773 | 22,359.67 | 21,861.97 | 22,304.97 | 22,338.50 | **88,865.11** |
| 774 | 14,856.17 | 15,152.98 | 15,744.21 | 15,892.86 | **61,646.23** |
| 775 | 24,296.02 | 22,940.12 | 23,544.60 | 23,441.54 | **94,222.28** |
| 776 | 7,086.85 | 6,685.34 | 6,706.56 | 6,846.79 | **27,325.54** |
| 777 | 3,977.30 | 3,982.07 | 4,044.66 | 3,756.49 | **15,760.53** |
| 778 | 4,493.99 | 3,882.92 | 4,107.16 | 4,176.05 | **16,660.12** |
| 779 | 5,632.99 | 5,321.37 | 5,453.21 | 5,186.21 | **21,593.77** |
| 780 | 7,723.41 | 7,461.59 | 7,636.21 | 7,504.45 | **30,325.66** |
| 781 | 8,632.77 | 8,345.87 | 8,793.13 | 8,490.70 | **34,262.47** |
| 782 | 35,288.37 | 32,774.05 | 33,983.61 | 33,177.15 | **135,223.18** |
| 783 | 4,870.25 | 4,467.78 | 4,493.32 | 4,336.87 | **18,168.22** |
| 784 | 8,423.31 | 7,914.88 | 8,066.21 | 7,669.32 | **32,073.72** |
| 785 | 6,428.85 | 6,091.42 | 6,002.89 | 5,850.01 | **24,373.16** |
| 786 | 16,823.95 | 15,699.29 | 16,624.52 | 16,557.05 | **65,704.80** |
| 787 | 13,278.51 | 12,142.27 | 12,327.89 | 12,332.02 | **50,080.70** |
| 788 | 1,404.51 | 1,401.90 | 1,376.61 | 1,400.70 | **5,583.73** |
| 789 | 1,331.16 | 1,195.25 | 1,257.79 | 1,198.19 | **4,982.38** |
| 790 | 3,322.49 | 3,063.51 | 3,226.83 | 3,121.45 | **12,734.27** |
| 791 | 6,325.29 | 6,165.31 | 6,212.11 | 5,736.96 | **24,439.67** |
| 792 | 774.14 | 666.19 | 661.35 | 666.99 | **2,768.67** |
| 793 | 2,455.64 | 2,277.94 | 2,466.27 | 2,323.85 | **9,523.71** |
| 794 | 8,892.61 | 8,456.59 | 8,835.06 | 8,523.35 | **34,707.62** |
| 795 | 2,212.83 | 2,094.65 | 2,139.80 | 2,038.11 | **8,485.39** |
| 796 | 4,152.15 | 3,838.48 | 4,062.13 | 4,048.33 | **16,101.09** |
| 797 | 5,295.80 | 4,869.78 | 5,057.01 | 5,095.80 | **20,318.39** |
| 798 | 269.47 | 223.14 | 244.44 | 235.62 | **972.67** |
| 799 | 5,066.96 | 4,846.18 | 5,198.70 | 4,653.98 | **19,765.82** |
| **TX - TEXAS - Total** | **481,739.26** | **459,172.40** | **472,361.04** | **465,558.29** | **1,878,831.00** |

**STATE: UT - UTAH**

| REGISTRANT ZIP CODE 3 | 1ST QUARTER | 2ND QUARTER | 3RD QUARTER | 4TH QUARTER | TOTAL GRAMS |
|---|---|---|---|---|---|
| 840 | 16,767.91 | 16,232.94 | 16,407.29 | 16,303.97 | **65,712.11** |
| 841 | 10,174.06 | 9,746.33 | 9,452.06 | 9,283.41 | **38,655.85** |
| 843 | 2,092.58 | 2,072.55 | 2,040.62 | 2,060.11 | **8,265.86** |
| 844 | 3,267.09 | 3,058.12 | 3,173.60 | 3,127.59 | **12,626.40** |
| 845 | 1,396.87 | 1,492.46 | 1,463.99 | 1,430.55 | **5,783.88** |
| 846 | 4,139.06 | 3,932.13 | 4,032.57 | 3,967.83 | **16,071.59** |
| 847 | 6,112.51 | 5,996.53 | 6,118.94 | 6,145.61 | **24,373.59** |
| **UT - UTAH - Total** | **43,950.07** | **42,531.06** | **42,689.07** | **42,319.09** | **171,489.29** |

**STATE: VA - VIRGINIA**

| REGISTRANT ZIP CODE 3 | 1ST QUARTER | 2ND QUARTER | 3RD QUARTER | 4TH QUARTER | TOTAL GRAMS |
|---|---|---|---|---|---|
| 201 | 3,092.01 | 2,677.28 | 2,851.36 | 2,860.96 | **11,481.61** |
| 220 | 1,250.47 | 968.95 | 1,053.74 | 999.28 | **4,272.45** |
| 221 | 1,279.25 | 1,045.33 | 1,201.68 | 1,126.47 | **4,652.72** |
| 222 | 328.07 | 260.31 | 272.61 | 273.41 | **1,134.39** |
| 223 | 653.51 | 525.79 | 582.02 | 619.78 | **2,381.10** |
| 224 | 1,999.55 | 1,770.32 | 1,843.68 | 1,928.03 | **7,541.58** |
| 225 | 831.34 | 800.75 | 809.36 | 827.93 | **3,269.38** |
| 226 | 1,913.05 | 1,689.89 | 1,780.40 | 1,744.62 | **7,127.96** |
| 227 | 352.30 | 330.20 | 343.68 | 357.73 | **1,383.91** |
| 228 | 1,650.49 | 1,536.81 | 1,596.26 | 1,543.05 | **6,326.61** |
| 229 | 1,741.72 | 1,580.42 | 1,590.83 | 1,547.99 | **6,460.95** |
| 230 | 1,555.83 | 1,501.97 | 1,493.33 | 1,475.00 | **6,026.13** |
| 231 | 2,716.82 | 2,431.63 | 2,548.53 | 2,522.65 | **10,219.62** |
| 232 | 3,942.60 | 3,353.70 | 3,576.01 | 3,424.70 | **14,297.01** |
| 233 | 1,942.86 | 1,836.04 | 1,796.47 | 1,817.98 | **7,393.35** |
| 234 | 4,247.16 | 3,805.48 | 3,794.04 | 3,904.72 | **15,751.40** |
| 235 | 1,422.59 | 1,218.61 | 1,223.92 | 1,297.10 | **5,162.21** |
| 236 | 2,493.63 | 2,124.93 | 2,302.28 | 2,138.86 | **9,059.69** |

| | | | | | |
|---|---|---|---|---|---|
| 237 | 1,148.51 | 1,009.52 | 1,084.21 | 968.81 | **4,211.05** |
| 238 | 2,138.77 | 1,941.31 | 2,008.27 | 1,969.84 | **8,058.20** |
| 239 | 893.20 | 805.61 | 850.05 | 799.94 | **3,348.80** |
| 240 | 6,424.52 | 6,143.94 | 6,169.83 | 6,253.96 | **24,992.24** |
| 241 | 6,258.54 | 5,842.98 | 6,105.53 | 5,884.62 | **24,091.66** |
| 242 | 9,160.58 | 8,777.20 | 9,055.57 | 8,617.58 | **35,610.93** |
| 243 | 3,775.10 | 3,513.11 | 3,510.41 | 3,483.49 | **14,282.12** |
| 244 | 2,077.49 | 1,969.62 | 2,042.44 | 1,984.84 | **8,074.40** |
| 245 | 5,290.25 | 4,929.69 | 5,075.91 | 5,054.98 | **20,350.83** |
| 246 | 3,063.57 | 2,811.26 | 3,002.30 | 2,981.30 | **11,858.43** |
| **VA - VIRGINIA - Total** | **73,643.77** | **67,202.64** | **69,564.74** | **68,409.58** | **278,820.72** |
| **STATE: VI - VIRGIN ISLANDS** | | | | | |
| REGISTRANT ZIP CODE 3 | 1ST QUARTER | 2ND QUARTER | 3RD QUARTER | 4TH QUARTER | **TOTAL GRAMS** |
| 008 | 250.73 | 138.69 | 161.69 | 151.27 | **702.39** |
| **VI - VIRGIN ISLANDS - Total** | **250.73** | **138.69** | **161.69** | **151.27** | **702.39** |
| **STATE: VT - VERMONT** | | | | | |
| REGISTRANT ZIP CODE 3 | 1ST QUARTER | 2ND QUARTER | 3RD QUARTER | 4TH QUARTER | **TOTAL GRAMS** |
| 050 | 267.41 | 262.04 | 282.49 | 281.92 | **1,093.86** |
| 051 | 139.85 | 137.68 | 136.27 | 138.84 | **552.64** |
| 052 | 209.00 | 220.56 | 212.97 | 220.61 | **863.14** |
| 053 | 188.75 | 192.60 | 168.97 | 160.95 | **711.26** |
| 054 | 1,391.63 | 1,281.35 | 1,338.20 | 1,309.24 | **5,320.42** |
| 056 | 604.12 | 558.98 | 552.04 | 543.45 | **2,258.59** |
| 057 | 316.26 | 280.12 | 309.18 | 315.18 | **1,220.74** |
| 058 | 442.20 | 464.28 | 441.51 | 446.53 | **1,794.53** |
| **VT - VERMONT - Total** | **3,559.23** | **3,397.60** | **3,441.64** | **3,416.70** | **13,815.17** |
| **STATE: WA - WASHINGTON** | | | | | |
| REGISTRANT ZIP CODE 3 | 1ST QUARTER | 2ND QUARTER | 3RD QUARTER | 4TH QUARTER | **TOTAL GRAMS** |
| 980 | 11,636.21 | 11,736.47 | 11,993.64 | 11,789.76 | **47,156.09** |
| 981 | 5,228.39 | 4,432.65 | 4,752.97 | 4,444.61 | **18,858.62** |
| 982 | 10,187.11 | 9,938.88 | 10,331.82 | 10,088.74 | **40,546.54** |
| 983 | 9,347.99 | 8,697.95 | 9,134.01 | 9,035.97 | **36,215.92** |
| 984 | 4,536.88 | 4,266.61 | 4,503.04 | 4,402.15 | **17,708.69** |
| 985 | 6,076.81 | 5,617.20 | 6,104.08 | 6,087.48 | **23,885.57** |
| 986 | 9,370.35 | 8,736.23 | 8,904.17 | 8,904.63 | **35,915.37** |
| 988 | 3,188.45 | 3,176.63 | 3,182.75 | 3,213.44 | **12,761.27** |
| 989 | 3,803.33 | 3,758.67 | 3,848.19 | 3,788.77 | **15,198.97** |
| 990 | 2,840.98 | 2,837.53 | 3,007.41 | 2,860.44 | **11,546.36** |
| 991 | 3,368.51 | 3,424.32 | 3,463.98 | 3,430.70 | **13,687.51** |
| 992 | 14,048.19 | 12,981.63 | 13,455.59 | 13,429.03 | **53,914.44** |
| 993 | 7,482.14 | 7,276.13 | 7,345.48 | 7,380.90 | **29,484.64** |
| 994 | 1,671.29 | 1,542.27 | 1,566.84 | 1,565.72 | **6,346.12** |
| **WA - WASHINGTON - Total** | **92,786.62** | **88,423.17** | **91,593.97** | **90,422.34** | **363,226.10** |
| **STATE: WI - WISCONSIN** | | | | | |
| REGISTRANT ZIP CODE 3 | 1ST QUARTER | 2ND QUARTER | 3RD QUARTER | 4TH QUARTER | **TOTAL GRAMS** |
| 530 | 5,680.80 | 5,064.04 | 5,573.53 | 5,704.82 | **22,023.18** |
| 531 | 7,511.99 | 7,123.58 | 7,598.98 | 7,484.55 | **29,719.10** |
| 532 | 7,556.24 | 6,791.82 | 7,277.39 | 7,018.05 | **28,643.50** |
| 534 | 1,676.65 | 1,572.10 | 1,606.77 | 1,681.11 | **6,536.64** |
| 535 | 5,634.40 | 5,351.37 | 5,761.70 | 5,650.41 | **22,397.88** |
| 537 | 2,581.80 | 2,508.93 | 2,572.63 | 2,484.26 | **10,147.62** |
| 538 | 592.63 | 565.23 | 600.78 | 604.72 | **2,363.35** |
| 539 | 2,718.66 | 2,596.56 | 2,717.02 | 2,644.85 | **10,677.09** |
| 540 | 636.06 | 589.01 | 647.09 | 636.95 | **2,509.11** |
| 541 | 1,743.33 | 1,701.00 | 1,800.32 | 1,734.07 | **6,978.73** |
| 542 | 1,057.45 | 1,038.17 | 1,086.65 | 1,112.54 | **4,294.81** |
| 543 | 1,799.92 | 1,659.61 | 1,658.44 | 1,755.69 | **6,873.65** |
| 544 | 3,471.05 | 3,401.22 | 3,317.02 | 3,257.55 | **13,446.84** |
| 545 | 1,088.22 | 1,045.62 | 1,172.29 | 1,035.07 | **4,341.20** |
| 546 | 2,936.43 | 2,681.98 | 2,776.37 | 2,723.44 | **11,118.23** |

| | | | | | |
|---|---|---|---|---|---|
| 547 | 2,005.79 | 1,895.36 | 2,029.89 | 2,000.70 | **7,931.73** |
| 548 | 2,293.62 | 2,363.75 | 2,503.85 | 2,343.97 | **9,505.21** |
| 549 | 4,844.09 | 4,566.26 | 4,879.01 | 4,735.12 | **19,024.49** |
| **WI - WISCONSIN - Total** | **55,829.13** | **52,515.61** | **55,579.75** | **54,607.87** | **218,532.35** |

**STATE: WV - WEST VIRGINIA**

| REGISTRANT ZIP CODE 3 | 1ST QUARTER | 2ND QUARTER | 3RD QUARTER | 4TH QUARTER | TOTAL GRAMS |
|---|---|---|---|---|---|
| 247 | 1,024.09 | 1,076.53 | 1,112.87 | 1,025.62 | **4,239.11** |
| 248 | 452.36 | 391.33 | 374.33 | 425.29 | **1,643.30** |
| 249 | 823.27 | 752.25 | 805.77 | 795.30 | **3,176.60** |
| 250 | 1,685.84 | 1,581.96 | 1,630.16 | 1,643.76 | **6,541.72** |
| 251 | 1,428.59 | 1,400.61 | 1,408.36 | 1,404.72 | **5,642.28** |
| 252 | 683.57 | 667.92 | 610.52 | 583.87 | **2,545.87** |
| 253 | 3,237.60 | 2,897.81 | 2,970.16 | 2,806.20 | **11,911.77** |
| 254 | 1,614.01 | 1,595.07 | 1,576.14 | 1,631.70 | **6,416.91** |
| 255 | 3,447.61 | 3,283.50 | 3,344.40 | 3,177.68 | **13,253.19** |
| 256 | 1,427.73 | 1,461.70 | 1,452.51 | 1,382.12 | **5,724.06** |
| 257 | 2,914.82 | 2,724.88 | 2,798.84 | 2,709.41 | **11,147.95** |
| 258 | 2,372.80 | 2,231.97 | 2,209.16 | 2,217.92 | **9,031.85** |
| 259 | 648.77 | 643.24 | 653.52 | 695.59 | **2,641.11** |
| 260 | 2,209.01 | 2,322.53 | 2,129.14 | 2,165.79 | **8,826.48** |
| 261 | 1,667.87 | 1,552.20 | 1,687.35 | 1,527.81 | **6,435.23** |
| 262 | 1,421.67 | 1,380.13 | 1,416.16 | 1,333.64 | **5,551.60** |
| 263 | 3,354.35 | 3,052.73 | 3,265.46 | 3,007.33 | **12,679.88** |
| 264 | 850.11 | 860.96 | 813.02 | 860.89 | **3,384.98** |
| 265 | 2,452.16 | 2,282.75 | 2,363.51 | 2,297.69 | **9,396.11** |
| 266 | 749.93 | 778.91 | 785.73 | 762.75 | **3,077.31** |
| 267 | 612.28 | 588.45 | 578.05 | 585.85 | **2,364.63** |
| 268 | 249.88 | 242.96 | 254.59 | 231.03 | **978.46** |
| **WV - WEST VIRGINIA - Total** | **35,328.32** | **33,770.38** | **34,239.73** | **33,271.98** | **136,610.40** |

**STATE: WY - WYOMING**

| REGISTRANT ZIP CODE 3 | 1ST QUARTER | 2ND QUARTER | 3RD QUARTER | 4TH QUARTER | TOTAL GRAMS |
|---|---|---|---|---|---|
| 820 | 1,429.32 | 1,370.79 | 1,398.52 | 1,377.26 | **5,575.89** |
| 822 | 340.39 | 354.14 | 365.47 | 364.29 | **1,424.29** |
| 823 | 143.50 | 151.61 | 155.44 | 166.24 | **616.79** |
| 824 | 908.42 | 924.38 | 877.49 | 885.26 | **3,595.55** |
| 825 | 597.57 | 541.94 | 580.72 | 588.96 | **2,309.19** |
| 826 | 1,548.04 | 1,472.73 | 1,512.76 | 1,418.11 | **5,951.63** |
| 827 | 611.75 | 580.45 | 609.08 | 618.33 | **2,419.61** |
| 828 | 771.49 | 783.60 | 713.44 | 784.68 | **3,053.21** |
| 829 | 1,309.62 | 1,273.56 | 1,275.57 | 1,297.15 | **5,155.91** |
| 830 | 138.85 | 127.88 | 151.25 | 137.61 | **555.60** |
| 831 | 271.26 | 339.44 | 386.57 | 332.61 | **1,329.88** |
| **WY - WYOMING - Total** | **8,070.20** | **7,920.53** | **8,026.31** | **7,970.49** | **31,987.54** |

**DRUG: 9230 - MEPERIDINE (PETHIDINE)**

**STATE: AK - ALASKA**

| REGISTRANT ZIP CODE 3 | 1ST QUARTER | 2ND QUARTER | 3RD QUARTER | 4TH QUARTER | TOTAL GRAMS |
|---|---|---|---|---|---|
| 995 | 73.97 | 54.25 | 82.90 | 119.29 | **330.41** |
| 996 | 28.65 | 51.53 | 32.25 | 31.16 | **143.58** |
| 997 | 0.54 | 1.96 | 2.29 | 4.79 | **9.59** |
| 998 | 2.72 | 0 | 0.54 | 0 | **3.27** |
| 999 | 0 | 0 | 0.44 | 0 | **0.44** |
| **AK - ALASKA - Total** | **105.89** | **107.74** | **118.42** | **155.24** | **487.28** |

**STATE: AL - ALABAMA**

| REGISTRANT ZIP CODE 3 | 1ST QUARTER | 2ND QUARTER | 3RD QUARTER | 4TH QUARTER | TOTAL GRAMS |
|---|---|---|---|---|---|
| 350 | 62.64 | 135.74 | 152.40 | 147.61 | **498.39** |
| 351 | 6.54 | 39.00 | 46.41 | 49.68 | **141.62** |
| 352 | 101.75 | 184.00 | 237.59 | 194.89 | **718.22** |
| 354 | 138.46 | 156.00 | 178.22 | 167.76 | **640.44** |
| 355 | 51.96 | 51.96 | 37.91 | 27.45 | **169.29** |
| 356 | 145.32 | 102.84 | 220.38 | 84.97 | **553.51** |

| | | | | | |
|---|---|---|---|---|---|
| 357 | 46.73 | 30.07 | 31.59 | 34.42 | **142.82** |
| 358 | 142.93 | 107.85 | 114.28 | 132.47 | **497.52** |
| 359 | 68.63 | 59.15 | 79.52 | 64.60 | **271.91** |
| 360 | 45.43 | 12.85 | 65.47 | 43.14 | **166.89** |
| 361 | 68.19 | 68.41 | 93.80 | 66.89 | **297.29** |
| 362 | 46.41 | 33.44 | 39.65 | 20.70 | **140.20** |
| 363 | 110.46 | 124.41 | 117.33 | 130.40 | **482.59** |
| 364 | 28.87 | 46.84 | 83.01 | 54.69 | **213.41** |
| 365 | 109.59 | 142.82 | 135.52 | 62.64 | **450.57** |
| 366 | 177.57 | 130.40 | 287.60 | 53.49 | **649.05** |
| 367 | 40.85 | 49.57 | 49.57 | 14.71 | **154.69** |
| 368 | 33.77 | 64.27 | 42.38 | 69.61 | **210.03** |
| 369 | 0 | 0 | 0 | 1.63 | **1.63** |
| **AL - ALABAMA - Total** | **1,426.10** | **1,539.61** | **2,012.62** | **1,421.74** | **6,400.08** |
| **STATE: AR - ARKANSAS** | | | | | |
| REGISTRANT ZIP CODE 3 | 1ST QUARTER | 2ND QUARTER | 3RD QUARTER | 4TH QUARTER | **TOTAL GRAMS** |
| 716 | 87.80 | 116.78 | 115.47 | 79.20 | **399.26** |
| 717 | 62.86 | 53.38 | 50.11 | 35.73 | **202.08** |
| 718 | 19.61 | 8.61 | 18.52 | 21.79 | **68.52** |
| 719 | 44.12 | 41.40 | 85.19 | 44.01 | **214.72** |
| 720 | 170.38 | 103.71 | 166.24 | 128.98 | **569.31** |
| 721 | 251.86 | 250.23 | 264.06 | 253.50 | **1,019.66** |
| 722 | 162.53 | 210.03 | 245.87 | 155.67 | **774.11** |
| 723 | 55.99 | 62.53 | 34.86 | 74.30 | **227.68** |
| 724 | 67.54 | 68.63 | 89.00 | 124.95 | **350.13** |
| 725 | 13.40 | 20.92 | 34.42 | 26.15 | **94.88** |
| 726 | 57.08 | 32.35 | 46.84 | 30.07 | **166.35** |
| 727 | 213.41 | 114.93 | 191.95 | 217.11 | **737.40** |
| 728 | 19.61 | 18.30 | 33.23 | 29.09 | **100.22** |
| 729 | 27.23 | 37.04 | 39.44 | 45.75 | **149.46** |
| **AR - ARKANSAS - Total** | **1,253.43** | **1,138.83** | **1,415.21** | **1,266.29** | **5,073.76** |
| **STATE: AZ - ARIZONA** | | | | | |
| REGISTRANT ZIP CODE 3 | 1ST QUARTER | 2ND QUARTER | 3RD QUARTER | 4TH QUARTER | **TOTAL GRAMS** |
| 850 | 351.91 | 284.26 | 332.93 | 410.00 | **1,379.11** |
| 851 | 37.91 | 49.24 | 72.77 | 72.55 | **232.47** |
| 852 | 521.88 | 476.06 | 687.68 | 676.37 | **2,361.98** |
| 853 | 195.11 | 211.49 | 239.77 | 228.33 | **874.70** |
| 855 | 13.62 | 9.26 | 44.66 | 45.21 | **112.75** |
| 856 | 90.09 | 9.59 | 70.59 | 22.88 | **193.15** |
| 857 | 103.36 | 190.88 | 174.47 | 156.63 | **625.34** |
| 859 | 15.14 | 1.31 | 2.18 | 3.81 | **22.44** |
| 860 | 12.53 | 9.15 | 25.97 | 11.33 | **58.98** |
| 863 | 7.52 | 12.20 | 23.86 | 39.33 | **82.90** |
| 864 | 42.59 | 26.91 | 46.19 | 52.73 | **168.42** |
| 865 | 0 | 0 | 0.65 | 0 | **0.65** |
| **AZ - ARIZONA - Total** | **1,391.65** | **1,280.34** | **1,721.74** | **1,719.16** | **6,112.90** |
| **STATE: CA - CALIFORNIA** | | | | | |
| REGISTRANT ZIP CODE 3 | 1ST QUARTER | 2ND QUARTER | 3RD QUARTER | 4TH QUARTER | **TOTAL GRAMS** |
| 900 | 169.83 | 147.83 | 414.18 | 220.82 | **952.66** |
| 902 | 206.44 | 266.46 | 439.56 | 297.07 | **1,209.53** |
| 903 | 17.43 | 5.45 | 20.70 | 27.23 | **70.81** |
| 904 | 45.75 | 26.80 | 51.09 | 41.72 | **165.37** |
| 905 | 203.39 | 22.33 | 51.09 | 112.86 | **389.67** |
| 906 | 11.98 | 37.37 | 71.57 | 17.87 | **138.79** |
| 907 | 13.07 | 9.48 | 30.07 | 20.59 | **73.21** |
| 908 | 56.10 | 23.42 | 45.21 | 50.44 | **175.17** |
| 910 | 25.06 | 36.71 | 38.24 | 31.48 | **131.49** |
| 911 | 28.32 | 39.65 | 39.22 | 39.22 | **146.41** |
| 912 | 111.12 | 58.83 | 134.43 | 163.84 | **468.21** |
| 913 | 273.22 | 335.53 | 475.62 | 263.96 | **1,348.32** |

| | | | | | |
|---|---|---|---|---|---|
| 914 | 52.18 | 31.05 | 140.53 | 134.10 | **357.86** |
| 915 | 20.48 | 12.96 | 100.44 | 63.18 | **197.07** |
| 916 | 0 | 1.09 | 0 | 0 | **1.09** |
| 917 | 500.57 | 137.70 | 317.23 | 346.64 | **1,302.13** |
| 918 | 44.23 | 28.32 | 23.97 | 13.07 | **109.59** |
| 919 | 16.99 | 15.14 | 18.74 | 17.97 | **68.85** |
| 920 | 219.73 | 271.15 | 523.77 | 462.98 | **1,477.63** |
| 921 | 381.06 | 298.38 | 522.79 | 462.77 | **1,665.00** |
| 922 | 125.82 | 106.11 | 119.94 | 112.21 | **464.07** |
| 923 | 286.61 | 226.81 | 90.42 | 124.08 | **727.92** |
| 924 | 12.20 | 2.72 | 16.89 | 16.34 | **48.15** |
| 925 | 294.35 | 161.12 | 519.52 | 444.90 | **1,419.89** |
| 926 | 241.84 | 157.96 | 355.90 | 311.78 | **1,067.48** |
| 927 | 111.01 | 62.09 | 156.33 | 83.88 | **413.31** |
| 928 | 100.22 | 176.48 | 286.18 | 164.06 | **726.94** |
| 930 | 89.87 | 46.95 | 121.14 | 57.95 | **315.92** |
| 931 | 215.04 | 121.25 | 238.90 | 123.86 | **699.05** |
| 932 | 93.25 | 50.22 | 168.74 | 130.29 | **442.50** |
| 933 | 275.72 | 243.15 | 414.51 | 463.64 | **1,397.01** |
| 934 | 67.54 | 42.59 | 144.56 | 98.04 | **352.74** |
| 935 | 26.58 | 50.00 | 30.50 | 25.06 | **132.14** |
| 936 | 56.97 | 64.38 | 64.82 | 41.29 | **227.46** |
| 937 | 119.50 | 139.00 | 182.36 | 150.66 | **591.53** |
| 939 | 11.98 | 11.98 | 11.66 | 13.73 | **49.35** |
| 940 | 129.53 | 41.83 | 85.95 | 72.23 | **329.54** |
| 941 | 85.95 | 60.35 | 172.77 | 172.34 | **491.42** |
| 943 | 33.01 | 25.06 | 23.31 | 59.92 | **141.29** |
| 944 | 5.56 | 1.74 | 7.19 | 2.61 | **17.10** |
| 945 | 98.59 | 93.47 | 189.77 | 93.90 | **475.73** |
| 946 | 3.81 | 7.08 | 24.95 | 3.27 | **39.11** |
| 947 | 2.18 | 1.20 | 0.54 | 1.09 | **5.01** |
| 948 | 0 | 0 | 0 | 2.72 | **2.72** |
| 949 | 36.49 | 5.01 | 26.36 | 5.56 | **73.42** |
| 950 | 38.02 | 41.29 | 51.85 | 36.06 | **167.22** |
| 951 | 22.33 | 13.62 | 54.36 | 35.62 | **125.93** |
| 952 | 24.73 | 24.62 | 34.86 | 25.93 | **110.14** |
| 953 | 198.27 | 257.75 | 522.68 | 314.39 | **1,293.09** |
| 954 | 20.15 | 30.50 | 33.23 | 21.24 | **105.12** |
| 955 | 6.75 | 8.72 | 2.18 | 0 | **17.65** |
| 956 | 55.01 | 47.17 | 188.68 | 130.51 | **421.37** |
| 957 | 5.45 | 2.18 | 0.76 | 0.22 | **8.61** |
| 958 | 98.70 | 127.46 | 234.65 | 130.18 | **590.99** |
| 959 | 35.40 | 49.57 | 57.19 | 32.90 | **175.06** |
| 960 | 33.33 | 41.83 | 39.54 | 21.46 | **136.17** |
| 961 | 4.25 | 4.36 | 2.18 | 2.18 | **12.96** |
| **CA - CALIFORNIA - Total** | **5,463.00** | **4,353.25** | **8,133.82** | **6,313.91** | **24,263.98** |

**STATE: CO - COLORADO**

| REGISTRANT ZIP CODE 3 | 1ST QUARTER | 2ND QUARTER | 3RD QUARTER | 4TH QUARTER | **TOTAL GRAMS** |
|---|---|---|---|---|---|
| 800 | 25.60 | 24.73 | 30.61 | 44.66 | **125.60** |
| 801 | 20.15 | 20.59 | 56.43 | 35.73 | **132.90** |
| 802 | 55.56 | 22.33 | 63.40 | 45.86 | **187.15** |
| 803 | 0.54 | 4.58 | 16.34 | 6.86 | **28.32** |
| 804 | 2.18 | 29.74 | 30.57 | 23.31 | **85.80** |
| 805 | 6.86 | 5.12 | 22.33 | 15.90 | **50.22** |
| 806 | 1.09 | 7.30 | 9.59 | 2.18 | **20.15** |
| 807 | 1.09 | 0 | 1.31 | 0.54 | **2.94** |
| 808 | 3.27 | 0.87 | 1.09 | 0.87 | **6.10** |
| 809 | 8.72 | 33.88 | 48.48 | 34.10 | **125.17** |
| 810 | 8.93 | 1.63 | 15.90 | 16.34 | **42.81** |
| 811 | 5.45 | 9.26 | 9.80 | 5.45 | **29.96** |

| | | | | | |
|---|---|---|---|---|---|
| 812 | 1.09 | 0 | 0 | 0.54 | **1.63** |
| 813 | 2.18 | 1.09 | 2.18 | 0.54 | **5.99** |
| 814 | 17.43 | 25.93 | 27.67 | 11.33 | **82.36** |
| 815 | 2.72 | 10.35 | 5.45 | 1.53 | **20.04** |
| 816 | 3.16 | 1.09 | 3.27 | 23.64 | **31.16** |
| **CO - COLORADO - Total** | **166.02** | **198.48** | **344.42** | **269.40** | **978.32** |

**STATE: CT - CONNECTICUT**

| REGISTRANT ZIP CODE 3 | 1ST QUARTER | 2ND QUARTER | 3RD QUARTER | 4TH QUARTER | TOTAL GRAMS |
|---|---|---|---|---|---|
| 060 | 0.54 | 2.72 | 4.58 | 5.45 | **13.29** |
| 061 | 8.72 | 4.90 | 15.47 | 16.34 | **45.43** |
| 062 | 3.27 | 1.63 | 0.98 | 1.09 | **6.97** |
| 063 | 9.26 | 3.81 | 2.72 | 9.80 | **25.60** |
| 064 | 1.09 | 1.53 | 5.45 | 10.24 | **18.30** |
| 065 | 3.81 | 3.81 | 10.35 | 11.33 | **29.30** |
| 066 | 2.72 | 3.27 | 6.54 | 2.72 | **15.25** |
| 067 | 0 | 0 | 6.54 | 0 | **6.54** |
| 068 | 0 | 1.09 | 8.93 | 2.18 | **12.20** |
| 069 | 0.22 | 2.40 | 2.18 | 1.09 | **5.88** |
| **CT - CONNECTICUT - Total** | **29.63** | **25.16** | **63.73** | **60.24** | **178.77** |

**STATE: DC - DISTRICT OF COLUMBIA**

| REGISTRANT ZIP CODE 3 | 1ST QUARTER | 2ND QUARTER | 3RD QUARTER | 4TH QUARTER | TOTAL GRAMS |
|---|---|---|---|---|---|
| 200 | 72.99 | 31.05 | 68.63 | 89.76 | **262.43** |
| 204 | 0 | 0 | 0 | 1.74 | **1.74** |
| **DC - DISTRICT OF COLUMBIA - Total** | **72.99** | **31.05** | **68.63** | **91.51** | **264.17** |

**STATE: DE - DELAWARE**

| REGISTRANT ZIP CODE 3 | 1ST QUARTER | 2ND QUARTER | 3RD QUARTER | 4TH QUARTER | TOTAL GRAMS |
|---|---|---|---|---|---|
| 197 | 0 | 7.30 | 5.45 | 3.27 | **16.01** |
| 198 | 4.36 | 0.54 | 1.09 | 2.18 | **8.17** |
| 199 | 41.83 | 11.55 | 80.07 | 10.24 | **143.69** |
| **DE - DELAWARE - Total** | **46.19** | **19.39** | **86.61** | **15.69** | **167.87** |

**STATE: FL - FLORIDA**

| REGISTRANT ZIP CODE 3 | 1ST QUARTER | 2ND QUARTER | 3RD QUARTER | 4TH QUARTER | TOTAL GRAMS |
|---|---|---|---|---|---|
| 320 | 47.93 | 32.68 | 36.60 | 29.96 | **147.17** |
| 321 | 44.66 | 45.10 | 44.77 | 36.06 | **170.60** |
| 322 | 20.04 | 42.38 | 37.47 | 26.69 | **126.59** |
| 323 | 104.58 | 58.61 | 73.64 | 60.24 | **297.07** |
| 324 | 46.30 | 49.13 | 99.57 | 71.90 | **266.90** |
| 325 | 61.55 | 67.43 | 66.89 | 84.75 | **280.62** |
| 326 | 43.03 | 23.09 | 25.27 | 40.31 | **131.71** |
| 327 | 60.13 | 36.71 | 37.80 | 55.78 | **190.42** |
| 328 | 20.70 | 19.17 | 26.91 | 28.32 | **95.10** |
| 329 | 88.57 | 134.76 | 66.45 | 67.21 | **356.99** |
| 330 | 141.73 | 139.22 | 207.74 | 106.87 | **595.56** |
| 331 | 182.69 | 353.18 | 473.66 | 317.66 | **1,327.19** |
| 333 | 58.83 | 159.27 | 224.52 | 161.66 | **604.28** |
| 334 | 91.51 | 69.18 | 133.45 | 97.39 | **391.52** |
| 335 | 22.33 | 40.09 | 44.66 | 25.82 | **132.90** |
| 336 | 194.13 | 133.45 | 161.01 | 138.24 | **626.83** |
| 337 | 80.29 | 51.75 | 80.18 | 58.06 | **270.27** |
| 338 | 196.20 | 42.70 | 104.04 | 139.33 | **482.27** |
| 339 | 152.19 | 158.29 | 181.60 | 160.03 | **652.10** |
| 341 | 26.04 | 20.37 | 16.56 | 40.09 | **103.05** |
| 342 | 40.31 | 52.73 | 46.52 | 43.25 | **182.80** |
| 344 | 11.00 | 8.50 | 14.27 | 21.79 | **55.56** |
| 346 | 30.50 | 54.90 | 59.81 | 29.41 | **174.63** |
| 347 | 20.15 | 39.54 | 11.98 | 13.07 | **84.75** |
| 349 | 52.40 | 72.44 | 83.12 | 87.48 | **295.44** |
| **FL - FLORIDA - Total** | **1,837.78** | **1,904.66** | **2,358.50** | **1,941.38** | **8,042.31** |

**STATE: GA - GEORGIA**

| REGISTRANT ZIP CODE 3 | 1ST QUARTER | 2ND QUARTER | 3RD QUARTER | 4TH QUARTER | TOTAL GRAMS |
|---|---|---|---|---|---|

| | | | | | |
|---|---|---|---|---|---|
| 300 | 126.48 | 123.97 | 419.85 | 181.82 | **852.11** |
| 301 | 51.31 | 69.72 | 109.81 | 94.12 | **324.96** |
| 302 | 52.73 | 32.79 | 94.45 | 58.94 | **238.90** |
| 303 | 168.42 | 143.58 | 197.50 | 190.75 | **700.25** |
| 304 | 42.49 | 43.68 | 45.43 | 36.06 | **167.65** |
| 305 | 56.76 | 63.51 | 46.52 | 82.03 | **248.81** |
| 306 | 40.52 | 13.18 | 40.31 | 34.42 | **128.44** |
| 307 | 171.79 | 83.56 | 113.51 | 87.04 | **455.90** |
| 308 | 17.43 | 9.37 | 15.25 | 9.26 | **51.31** |
| 309 | 75.93 | 59.26 | 62.09 | 61.77 | **259.05** |
| 310 | 110.24 | 72.44 | 182.58 | 129.64 | **494.90** |
| 312 | 7.08 | 14.60 | 14.92 | 18.96 | **55.56** |
| 313 | 7.08 | 10.89 | 22.01 | 21.90 | **61.88** |
| 314 | 121.68 | 106.98 | 91.94 | 78.11 | **398.71** |
| 315 | 61.88 | 65.47 | 87.37 | 72.66 | **287.38** |
| 316 | 78.98 | 173.10 | 110.46 | 105.34 | **467.89** |
| 317 | 60.68 | 147.94 | 129.64 | 157.63 | **495.88** |
| 318 | 0 | 0 | 0 | 1.09 | **1.09** |
| 319 | 18.74 | 19.72 | 38.35 | 75.71 | **152.51** |
| 398 | 2.40 | 15.80 | 14.60 | 23.86 | **56.65** |
| **GA - GEORGIA - Total** | **1,272.61** | **1,269.56** | **1,836.58** | **1,521.09** | **5,899.84** |

**STATE: GU - GUAM**

| REGISTRANT ZIP CODE 3 | 1ST QUARTER | 2ND QUARTER | 3RD QUARTER | 4TH QUARTER | TOTAL GRAMS |
|---|---|---|---|---|---|
| 969 | 11.44 | 67.54 | 39.22 | 26.15 | **144.34** |
| **GU - GUAM - Total** | **11.44** | **67.54** | **39.22** | **26.15** | **144.34** |

**STATE: HI - HAWAII**

| REGISTRANT ZIP CODE 3 | 1ST QUARTER | 2ND QUARTER | 3RD QUARTER | 4TH QUARTER | TOTAL GRAMS |
|---|---|---|---|---|---|
| 967 | 33.23 | 25.60 | 60.79 | 44.23 | **163.84** |
| 968 | 18.74 | 25.71 | 61.88 | 25.82 | **132.14** |
| **HI - HAWAII - Total** | **51.96** | **51.31** | **122.66** | **70.05** | **295.98** |

**STATE: IA - IOWA**

| REGISTRANT ZIP CODE 3 | 1ST QUARTER | 2ND QUARTER | 3RD QUARTER | 4TH QUARTER | TOTAL GRAMS |
|---|---|---|---|---|---|
| 500 | 18.74 | 4.36 | 7.63 | 27.23 | **57.95** |
| 501 | 0 | 2.29 | 1.09 | 3.38 | **6.75** |
| 502 | 32.79 | 4.25 | 35.51 | 4.58 | **77.13** |
| 503 | 207.42 | 89.00 | 339.34 | 261.89 | **897.65** |
| 504 | 0.54 | 0.44 | 7.63 | 7.08 | **15.69** |
| 505 | 28.32 | 27.02 | 90.31 | 45.75 | **191.40** |
| 506 | 0.98 | 0.87 | 0.87 | 0 | **2.72** |
| 507 | 96.95 | 54.47 | 61.33 | 67.21 | **279.97** |
| 508 | 1.09 | 0.22 | 1.96 | 7.84 | **11.11** |
| 510 | 0.87 | 2.18 | 0.44 | 2.61 | **6.10** |
| 511 | 32.68 | 7.84 | 27.45 | 16.56 | **84.54** |
| 512 | 0.54 | 1.20 | 0 | 0 | **1.74** |
| 513 | 5.23 | 1.74 | 3.05 | 2.18 | **12.20** |
| 514 | 2.18 | 5.23 | 0 | 1.63 | **9.04** |
| 515 | 1.63 | 2.40 | 4.14 | 3.81 | **11.98** |
| 516 | 0 | 1.85 | 0.54 | 5.45 | **7.84** |
| 520 | 0.54 | 5.66 | 3.16 | 3.16 | **12.53** |
| 522 | 22.88 | 46.95 | 50.11 | 50.00 | **169.94** |
| 523 | 0 | 0 | 2.61 | 2.72 | **5.34** |
| 524 | 100.66 | 34.86 | 94.56 | 106.65 | **336.73** |
| 525 | 0.54 | 1.63 | 8.17 | 7.52 | **17.87** |
| 526 | 10.46 | 0.87 | 1.31 | 26.15 | **38.78** |
| 527 | 3.16 | 17.87 | 2.07 | 5.99 | **29.09** |
| 528 | 23.97 | 7.52 | 15.25 | 8.06 | **54.80** |
| **IA - IOWA - Total** | **592.18** | **320.71** | **758.53** | **667.46** | **2,338.89** |

**STATE: ID - IDAHO**

| REGISTRANT ZIP CODE 3 | 1ST QUARTER | 2ND QUARTER | 3RD QUARTER | 4TH QUARTER | TOTAL GRAMS |
|---|---|---|---|---|---|
| 832 | 14.16 | 7.63 | 17.32 | 16.34 | **55.45** |

| | | | | | |
|---|---|---|---|---|---|
| 833 | 13.94 | 4.25 | 14.60 | 21.24 | **54.03** |
| 834 | 68.63 | 90.42 | 36.39 | 44.77 | **240.21** |
| 835 | 5.45 | 1.09 | 2.72 | 0.98 | **10.24** |
| 836 | 17.97 | 17.97 | 10.57 | 12.96 | **59.48** |
| 837 | 54.03 | 82.36 | 43.14 | 34.97 | **214.50** |
| 838 | 13.94 | 9.15 | 30.18 | 17.54 | **70.81** |
| **ID - IDAHO - Total** | **188.14** | **212.86** | **154.91** | **148.81** | **704.72** |

**STATE: IL - ILLINOIS**

| REGISTRANT ZIP CODE 3 | 1ST QUARTER | 2ND QUARTER | 3RD QUARTER | 4TH QUARTER | TOTAL GRAMS |
|---|---|---|---|---|---|
| 600 | 52.62 | 56.54 | 76.47 | 57.52 | **243.15** |
| 601 | 32.68 | 25.27 | 37.37 | 56.97 | **152.29** |
| 602 | 0.54 | 0 | 1.09 | 0.54 | **2.18** |
| 603 | 0.87 | 0 | 0 | 0.44 | **1.31** |
| 604 | 31.48 | 14.16 | 54.69 | 37.04 | **137.37** |
| 605 | 20.15 | 19.50 | 40.52 | 27.45 | **107.63** |
| 606 | 22.44 | 44.99 | 43.47 | 49.24 | **160.14** |
| 607 | 0.54 | 0 | 0 | 0 | **0.54** |
| 609 | 0 | 0.65 | 0.54 | 0.22 | **1.42** |
| 610 | 39.22 | 4.36 | 3.49 | 4.79 | **51.85** |
| 611 | 5.99 | 7.73 | 3.70 | 11.00 | **28.43** |
| 612 | 6.21 | 4.47 | 6.54 | 4.36 | **21.57** |
| 613 | 0.54 | 0.54 | 23.86 | 18.85 | **43.79** |
| 614 | 10.35 | 9.26 | 30.50 | 14.16 | **64.27** |
| 615 | 0 | 1.31 | 3.27 | 0 | **4.58** |
| 616 | 13.94 | 6.54 | 30.28 | 40.31 | **91.07** |
| 617 | 1.09 | 4.68 | 5.77 | 3.70 | **15.25** |
| 618 | 33.23 | 3.92 | 24.84 | 29.85 | **91.83** |
| 619 | 15.25 | 1.09 | 0 | 4.36 | **20.70** |
| 620 | 0.54 | 4.36 | 2.83 | 8.06 | **15.80** |
| 622 | 29.85 | 22.44 | 61.01 | 14.92 | **128.22** |
| 623 | 9.26 | 4.79 | 11.98 | 9.80 | **35.84** |
| 624 | 0.87 | 3.38 | 2.72 | 0 | **6.97** |
| 625 | 7.84 | 1.09 | 2.83 | 3.81 | **15.58** |
| 626 | 4.36 | 13.07 | 5.66 | 2.18 | **25.27** |
| 627 | 11.98 | 8.06 | 44.66 | 10.68 | **75.38** |
| 628 | 35.95 | 24.18 | 22.88 | 33.23 | **116.24** |
| 629 | 22.44 | 57.52 | 76.80 | 101.20 | **257.96** |
| **IL - ILLINOIS - Total** | **410.26** | **343.92** | **617.78** | **544.69** | **1,916.65** |

**STATE: IN - INDIANA**

| REGISTRANT ZIP CODE 3 | 1ST QUARTER | 2ND QUARTER | 3RD QUARTER | 4TH QUARTER | TOTAL GRAMS |
|---|---|---|---|---|---|
| 460 | 70.26 | 49.57 | 110.03 | 166.24 | **396.10** |
| 461 | 85.52 | 62.75 | 115.80 | 121.79 | **385.86** |
| 462 | 179.53 | 71.03 | 279.10 | 605.47 | **1,135.13** |
| 463 | 23.31 | 15.14 | 27.13 | 15.90 | **81.49** |
| 464 | 23.42 | 10.89 | 10.89 | 29.41 | **74.62** |
| 465 | 22.33 | 16.89 | 11.66 | 11.66 | **62.53** |
| 466 | 19.06 | 30.50 | 41.40 | 10.89 | **101.86** |
| 467 | 2.51 | 3.92 | 4.25 | 5.45 | **16.12** |
| 468 | 15.80 | 9.15 | 61.44 | 11.98 | **98.37** |
| 469 | 2.18 | 8.61 | 15.90 | 11.77 | **38.45** |
| 470 | 14.71 | 28.00 | 25.60 | 32.90 | **101.20** |
| 471 | 33.77 | 39.11 | 33.23 | 50.22 | **156.33** |
| 472 | 73.21 | 35.40 | 47.17 | 35.40 | **191.19** |
| 473 | 47.93 | 20.15 | 69.39 | 47.50 | **184.98** |
| 474 | 9.70 | 13.84 | 11.66 | 6.54 | **41.72** |
| 475 | 8.17 | 5.99 | 14.71 | 11.44 | **40.31** |
| 476 | 23.86 | 13.40 | 29.41 | 15.25 | **81.92** |
| 477 | 26.04 | 5.45 | 11.98 | 11.44 | **54.90** |
| 478 | 6.43 | 2.29 | 1.85 | 8.82 | **19.39** |
| 479 | 1.09 | 24.08 | 14.92 | 9.70 | **49.78** |

| IN - INDIANA - Total | 688.81 | 466.14 | 937.52 | 1,219.77 | 3,312.24 |
|---|---|---|---|---|---|

STATE: KS - KANSAS

| REGISTRANT ZIP CODE 3 | 1ST QUARTER | 2ND QUARTER | 3RD QUARTER | 4TH QUARTER | TOTAL GRAMS |
|---|---|---|---|---|---|
| 660 | 98.48 | 13.07 | 111.55 | 99.57 | **322.67** |
| 661 | 0 | 0 | 4.36 | 7.84 | **12.20** |
| 662 | 11.98 | 14.60 | 26.69 | 42.49 | **95.76** |
| 664 | 0.54 | 9.26 | 14.60 | 1.63 | **26.04** |
| 665 | 0 | 6.97 | 8.82 | 2.94 | **18.74** |
| 666 | 14.16 | 5.99 | 13.84 | 15.69 | **49.68** |
| 667 | 7.63 | 3.92 | 12.42 | 5.12 | **29.09** |
| 668 | 3.27 | 1.09 | 6.75 | 3.05 | **14.16** |
| 669 | 4.90 | 4.58 | 10.89 | 4.47 | **24.84** |
| 670 | 4.79 | 7.41 | 7.08 | 10.89 | **30.18** |
| 671 | 6.54 | 7.41 | 11.00 | 6.86 | **31.81** |
| 672 | 81.16 | 82.25 | 88.46 | 62.75 | **314.61** |
| 673 | 1.09 | 11.66 | 22.66 | 2.18 | **37.58** |
| 674 | 6.54 | 5.66 | 24.84 | 9.26 | **46.30** |
| 675 | 12.64 | 12.09 | 17.43 | 11.22 | **53.38** |
| 676 | 8.72 | 5.99 | 6.43 | 1.31 | **22.44** |
| 677 | 1.09 | 3.49 | 2.61 | 1.09 | **8.28** |
| 678 | 29.30 | 53.82 | 39.98 | 23.09 | **146.19** |
| 679 | 2.72 | 13.07 | 15.25 | 13.07 | **44.12** |
| **KS - KANSAS - Total** | **295.55** | **262.32** | **445.66** | **324.52** | **1,328.06** |

STATE: KY - KENTUCKY

| REGISTRANT ZIP CODE 3 | 1ST QUARTER | 2ND QUARTER | 3RD QUARTER | 4TH QUARTER | TOTAL GRAMS |
|---|---|---|---|---|---|
| 400 | 7.63 | 13.07 | 4.36 | 3.27 | **28.32** |
| 401 | 3.92 | 1.20 | 1.74 | 0 | **6.86** |
| 402 | 38.67 | 59.04 | 77.02 | 49.57 | **224.30** |
| 403 | 3.81 | 10.89 | 5.99 | 13.84 | **34.53** |
| 404 | 13.62 | 15.80 | 16.34 | 13.40 | **59.15** |
| 405 | 57.52 | 22.99 | 45.32 | 65.47 | **191.29** |
| 406 | 0 | 1.09 | 1.09 | 1.09 | **3.27** |
| 407 | 2.18 | 4.79 | 19.61 | 14.16 | **40.74** |
| 408 | 10.46 | 29.74 | 22.88 | 21.79 | **84.86** |
| 409 | 13.62 | 34.86 | 41.40 | 21.13 | **111.01** |
| 410 | 25.06 | 16.67 | 53.27 | 33.12 | **128.11** |
| 412 | 0 | 0 | 0 | 0.54 | **0.54** |
| 413 | 2.18 | 4.36 | 2.61 | 1.63 | **10.78** |
| 414 | 1.09 | 0 | 0 | 0 | **1.09** |
| 415 | 8.72 | 8.06 | 20.15 | 18.52 | **55.45** |
| 416 | 8.93 | 11.98 | 8.17 | 2.94 | **32.03** |
| 417 | 4.90 | 8.72 | 7.08 | 16.23 | **36.93** |
| 418 | 39.76 | 37.15 | 26.47 | 23.97 | **127.35** |
| 420 | 27.23 | 63.62 | 81.27 | 40.74 | **212.86** |
| 421 | 44.12 | 78.44 | 113.19 | 55.99 | **291.73** |
| 422 | 20.70 | 10.57 | 18.08 | 6.54 | **55.88** |
| 423 | 40.31 | 30.72 | 22.88 | 16.34 | **110.24** |
| 424 | 26.91 | 12.96 | 17.43 | 16.56 | **73.86** |
| 425 | 9.26 | 31.27 | 11.98 | 18.52 | **71.03** |
| 426 | 4.36 | 3.16 | 7.08 | 9.48 | **24.08** |
| 427 | 34.32 | 33.44 | 46.08 | 36.39 | **150.22** |
| **KY - KENTUCKY - Total** | **449.26** | **544.58** | **671.49** | **501.22** | **2,166.55** |

STATE: LA - LOUISIANA

| REGISTRANT ZIP CODE 3 | 1ST QUARTER | 2ND QUARTER | 3RD QUARTER | 4TH QUARTER | TOTAL GRAMS |
|---|---|---|---|---|---|
| 700 | 51.09 | 93.47 | 118.31 | 54.58 | **317.44** |
| 701 | 23.42 | 23.09 | 64.27 | 50.33 | **161.12** |
| 703 | 52.83 | 58.83 | 66.56 | 51.31 | **229.53** |
| 704 | 212.54 | 182.58 | 198.16 | 185.08 | **778.36** |
| 705 | 144.56 | 263.74 | 311.67 | 255.68 | **975.64** |
| 706 | 91.40 | 109.26 | 81.92 | 100.88 | **383.46** |

| | | | | | |
|---|---|---|---|---|---|
| 707 | 75.38 | 46.84 | 51.96 | 87.37 | **261.56** |
| 708 | 106.76 | 74.08 | 112.31 | 123.43 | **416.58** |
| 710 | 11.77 | 54.69 | 70.92 | 57.19 | **194.56** |
| 711 | 124.95 | 154.04 | 149.03 | 87.04 | **515.06** |
| 712 | 143.36 | 138.02 | 114.49 | 73.21 | **469.08** |
| 713 | 56.32 | 76.15 | 101.53 | 111.66 | **345.66** |
| 714 | 30.83 | 40.63 | 66.45 | 35.40 | **173.32** |
| **LA - LOUISIANA - Total** | **1,125.22** | **1,315.42** | **1,507.59** | **1,273.15** | **5,221.37** |

**STATE: MA - MASSACHUSETTS**

| REGISTRANT ZIP CODE 3 | 1ST QUARTER | 2ND QUARTER | 3RD QUARTER | 4TH QUARTER | TOTAL GRAMS |
|---|---|---|---|---|---|
| 010 | 50.11 | 52.83 | 52.29 | 39.22 | **194.45** |
| 011 | 3.81 | 0.87 | 31.81 | 31.37 | **67.87** |
| 012 | 0 | 0.54 | 0 | 0 | **0.54** |
| 013 | 0 | 0 | 1.31 | 0 | **1.31** |
| 014 | 2.18 | 0.44 | 2.29 | 0.76 | **5.66** |
| 015 | 17.43 | 7.08 | 15.25 | 5.45 | **45.21** |
| 016 | 0.54 | 4.36 | 1.63 | 1.85 | **8.39** |
| 017 | 7.84 | 5.12 | 13.29 | 6.97 | **33.23** |
| 018 | 16.89 | 2.40 | 24.18 | 6.10 | **49.57** |
| 019 | 84.97 | 42.59 | 36.17 | 62.86 | **226.59** |
| 020 | 2.72 | 0 | 0.54 | 5.12 | **8.39** |
| 021 | 86.06 | 38.67 | 142.05 | 64.60 | **331.39** |
| 022 | 3.81 | 14.82 | 9.91 | 7.63 | **36.17** |
| 023 | 29.96 | 21.79 | 35.95 | 31.81 | **119.50** |
| 024 | 0 | 11.44 | 6.54 | 3.27 | **21.24** |
| 025 | 0 | 0 | 2.40 | 3.27 | **5.66** |
| 026 | 0 | 0 | 2.18 | 0 | **2.18** |
| 027 | 11.55 | 9.15 | 16.67 | 10.13 | **47.50** |
| **MA - MASSACHUSETTS - Total** | **317.88** | **212.10** | **394.46** | **280.41** | **1,204.85** |

**STATE: MD - MARYLAND**

| REGISTRANT ZIP CODE 3 | 1ST QUARTER | 2ND QUARTER | 3RD QUARTER | 4TH QUARTER | TOTAL GRAMS |
|---|---|---|---|---|---|
| 206 | 46.73 | 36.71 | 47.71 | 57.30 | **188.46** |
| 207 | 98.59 | 84.10 | 55.88 | 68.30 | **306.88** |
| 208 | 43.25 | 21.24 | 30.61 | 44.12 | **139.22** |
| 209 | 4.36 | 0 | 1.09 | 9.26 | **14.71** |
| 210 | 45.97 | 59.92 | 75.82 | 62.75 | **244.46** |
| 211 | 5.99 | 17.65 | 52.62 | 27.34 | **103.60** |
| 212 | 146.52 | 106.65 | 145.87 | 111.88 | **510.92** |
| 214 | 13.07 | 6.54 | 11.44 | 17.32 | **48.37** |
| 215 | 0.54 | 0 | 1.31 | 13.62 | **15.47** |
| 216 | 28.32 | 5.99 | 10.02 | 12.64 | **56.97** |
| 217 | 9.26 | 10.89 | 22.22 | 8.93 | **51.31** |
| 218 | 0 | 2.18 | 12.85 | 2.61 | **17.65** |
| **MD - MARYLAND - Total** | **442.61** | **351.87** | **467.45** | **436.08** | **1,698.01** |

**STATE: ME - MAINE**

| REGISTRANT ZIP CODE 3 | 1ST QUARTER | 2ND QUARTER | 3RD QUARTER | 4TH QUARTER | TOTAL GRAMS |
|---|---|---|---|---|---|
| 039 | 0.44 | 0 | 1.31 | 21.79 | **23.53** |
| 040 | 9.26 | 2.29 | 5.88 | 1.85 | **19.28** |
| 041 | 32.68 | 33.33 | 1.09 | 4.58 | **71.68** |
| 042 | 6.54 | 9.26 | 1.09 | 14.05 | **30.94** |
| 043 | 0 | 2.61 | 0.87 | 1.09 | **4.58** |
| 044 | 17.43 | 6.21 | 6.75 | 9.80 | **40.20** |
| 045 | 0 | 0.44 | 0 | 0 | **0.44** |
| 046 | 17.97 | 4.14 | 4.90 | 10.89 | **37.91** |
| 047 | 0.98 | 2.18 | 2.72 | 1.09 | **6.97** |
| 049 | 0 | 0 | 0.54 | 4.36 | **4.90** |
| **ME - MAINE - Total** | **85.30** | **60.46** | **25.16** | **69.50** | **240.43** |

**STATE: MI - MICHIGAN**

| REGISTRANT ZIP CODE 3 | 1ST QUARTER | 2ND QUARTER | 3RD QUARTER | 4TH QUARTER | TOTAL GRAMS |
|---|---|---|---|---|---|
| 480 | 39.98 | 46.08 | 73.75 | 55.45 | **215.26** |

| | | | | | |
|---|---|---|---|---|---|
| 481 | 39.33 | 44.01 | 68.30 | 45.97 | **197.61** |
| 482 | 0 | 7.73 | 31.81 | 9.80 | **49.35** |
| 483 | 15.25 | 7.63 | 68.30 | 24.62 | **115.80** |
| 484 | 4.36 | 4.58 | 10.02 | 7.95 | **26.91** |
| 485 | 1.85 | 3.05 | 10.24 | 12.96 | **28.11** |
| 486 | 96.74 | 74.62 | 88.78 | 62.64 | **322.78** |
| 487 | 1.09 | 4.36 | 16.34 | 13.62 | **35.40** |
| 488 | 3.16 | 5.66 | 4.79 | 15.25 | **28.87** |
| 489 | 22.88 | 11.77 | 64.60 | 30.39 | **129.64** |
| 490 | 8.72 | 9.15 | 18.96 | 17.97 | **54.80** |
| 491 | 0 | 1.09 | 0 | 1.09 | **2.18** |
| 492 | 6.32 | 15.90 | 20.04 | 7.30 | **49.57** |
| 493 | 0.54 | 2.18 | 11.55 | 2.18 | **16.45** |
| 494 | 20.92 | 13.62 | 15.80 | 5.88 | **56.21** |
| 495 | 8.17 | 6.75 | 32.25 | 26.69 | **73.86** |
| 496 | 30.94 | 37.69 | 48.91 | 61.77 | **179.31** |
| 497 | 43.03 | 42.92 | 59.15 | 42.70 | **187.81** |
| 498 | 9.80 | 53.71 | 32.35 | 26.80 | **122.66** |
| 499 | 0 | 3.27 | 1.09 | 1.09 | **5.45** |
| **MI - MICHIGAN - Total** | **353.07** | **395.77** | **677.05** | **472.14** | **1,898.02** |

**STATE: MN - MINNESOTA**

| REGISTRANT ZIP CODE 3 | 1ST QUARTER | 2ND QUARTER | 3RD QUARTER | 4TH QUARTER | **TOTAL GRAMS** |
|---|---|---|---|---|---|
| 550 | 12.64 | 3.70 | 12.42 | 6.97 | **35.73** |
| 551 | 5.34 | 16.34 | 11.33 | 4.90 | **37.91** |
| 553 | 19.83 | 23.75 | 18.19 | 12.09 | **73.86** |
| 554 | 35.62 | 34.97 | 23.31 | 32.35 | **126.26** |
| 557 | 8.72 | 10.89 | 17.32 | 10.68 | **47.61** |
| 558 | 2.51 | 1.63 | 9.80 | 1.31 | **15.25** |
| 559 | 13.62 | 3.49 | 13.29 | 10.02 | **40.42** |
| 560 | 5.12 | 0.54 | 4.58 | 6.10 | **16.34** |
| 561 | 2.18 | 0 | 2.18 | 3.05 | **7.41** |
| 562 | 0 | 4.03 | 6.65 | 3.92 | **14.60** |
| 563 | 11.11 | 12.53 | 16.23 | 10.35 | **50.22** |
| 564 | 4.58 | 1.53 | 4.14 | 3.27 | **13.51** |
| 565 | 2.18 | 0 | 2.18 | 1.85 | **6.21** |
| 566 | 0 | 1.63 | 1.31 | 0.54 | **3.49** |
| 567 | 15.25 | 2.18 | 3.49 | 2.18 | **23.09** |
| **MN - MINNESOTA - Total** | **138.68** | **117.22** | **146.41** | **109.59** | **511.90** |

**STATE: MO - MISSOURI**

| REGISTRANT ZIP CODE 3 | 1ST QUARTER | 2ND QUARTER | 3RD QUARTER | 4TH QUARTER | **TOTAL GRAMS** |
|---|---|---|---|---|---|
| 630 | 59.37 | 79.52 | 84.43 | 79.42 | **302.74** |
| 631 | 83.99 | 120.05 | 141.95 | 132.03 | **478.02** |
| 633 | 4.90 | 8.17 | 11.00 | 10.89 | **34.97** |
| 634 | 6.54 | 3.49 | 13.18 | 3.70 | **26.91** |
| 635 | 9.91 | 22.22 | 20.48 | 22.01 | **74.62** |
| 636 | 11.98 | 3.27 | 10.35 | 10.13 | **35.73** |
| 637 | 7.08 | 7.63 | 11.44 | 6.54 | **32.68** |
| 638 | 8.28 | 13.18 | 32.90 | 8.72 | **63.07** |
| 639 | 16.89 | 12.42 | 14.38 | 13.07 | **56.76** |
| 640 | 8.72 | 45.64 | 31.05 | 20.48 | **105.89** |
| 641 | 79.63 | 32.90 | 67.43 | 41.83 | **221.80** |
| 644 | 8.17 | 16.99 | 13.29 | 14.92 | **53.38** |
| 645 | 20.70 | 20.70 | 42.92 | 9.80 | **94.12** |
| 646 | 2.18 | 4.36 | 3.27 | 0 | **9.80** |
| 647 | 5.45 | 8.17 | 13.07 | 6.54 | **33.23** |
| 648 | 66.13 | 13.62 | 39.22 | 32.68 | **151.64** |
| 650 | 1.31 | 0 | 0.54 | 0 | **1.85** |
| 651 | 28.32 | 24.18 | 32.35 | 20.37 | **105.23** |
| 652 | 15.14 | 34.86 | 26.25 | 14.60 | **90.85** |
| 653 | 35.95 | 2.29 | 18.52 | 18.96 | **75.71** |

| | | | | | |
|---|---|---|---|---|---|
| 655 | 40.52 | 53.60 | 53.60 | 14.38 | **162.10** |
| 656 | 1.09 | 4.36 | 0 | 0 | **5.45** |
| 657 | 13.62 | 4.25 | 2.40 | 10.02 | **30.28** |
| 658 | 5.99 | 19.50 | 33.12 | 52.83 | **111.44** |
| **MO - MISSOURI - Total** | **541.86** | **555.36** | **717.14** | **543.92** | **2,358.28** |

**STATE: MS - MISSISSIPPI**

| REGISTRANT ZIP CODE 3 | 1ST QUARTER | 2ND QUARTER | 3RD QUARTER | 4TH QUARTER | TOTAL GRAMS |
|---|---|---|---|---|---|
| 386 | 108.83 | 114.82 | 87.15 | 121.68 | **432.48** |
| 387 | 107.30 | 134.76 | 129.20 | 113.30 | **484.55** |
| 388 | 45.10 | 58.61 | 65.58 | 61.44 | **230.73** |
| 389 | 34.32 | 44.99 | 46.84 | 25.06 | **151.21** |
| 390 | 35.95 | 66.02 | 52.94 | 71.57 | **226.48** |
| 391 | 47.93 | 29.41 | 76.80 | 75.28 | **229.42** |
| 392 | 175.50 | 156.76 | 196.85 | 229.75 | **758.86** |
| 393 | 70.16 | 53.71 | 92.60 | 26.69 | **243.15** |
| 394 | 82.03 | 143.14 | 157.20 | 131.16 | **513.53** |
| 395 | 295.77 | 158.61 | 316.79 | 177.89 | **949.06** |
| 396 | 27.23 | 34.53 | 24.18 | 35.30 | **121.25** |
| 397 | 25.60 | 59.92 | 45.86 | 55.78 | **187.15** |
| **MS - MISSISSIPPI - Total** | **1,055.71** | **1,055.28** | **1,292.00** | **1,124.89** | **4,527.88** |

**STATE: MT - MONTANA**

| REGISTRANT ZIP CODE 3 | 1ST QUARTER | 2ND QUARTER | 3RD QUARTER | 4TH QUARTER | TOTAL GRAMS |
|---|---|---|---|---|---|
| 590 | 0 | 0 | 0.44 | 0 | **0.44** |
| 591 | 21.57 | 7.41 | 25.06 | 27.02 | **81.05** |
| 592 | 4.36 | 0 | 2.72 | 0 | **7.08** |
| 593 | 0.54 | 0 | 0 | 0 | **0.54** |
| 594 | 38.78 | 29.63 | 60.13 | 29.85 | **158.40** |
| 595 | 0 | 0.44 | 1.31 | 0 | **1.74** |
| 596 | 10.78 | 4.14 | 6.21 | 10.02 | **31.16** |
| 597 | 6.86 | 7.95 | 8.61 | 7.73 | **31.16** |
| 598 | 23.64 | 3.81 | 3.59 | 8.17 | **39.22** |
| 599 | 0.54 | 8.61 | 7.95 | 5.77 | **22.88** |
| **MT - MONTANA - Total** | **107.09** | **61.99** | **116.02** | **88.57** | **373.66** |

**STATE: NC - NORTH CAROLINA**

| REGISTRANT ZIP CODE 3 | 1ST QUARTER | 2ND QUARTER | 3RD QUARTER | 4TH QUARTER | TOTAL GRAMS |
|---|---|---|---|---|---|
| 270 | 8.72 | 4.36 | 5.66 | 21.35 | **40.09** |
| 271 | 36.17 | 42.38 | 56.65 | 45.21 | **180.40** |
| 272 | 28.54 | 32.03 | 36.71 | 31.59 | **128.87** |
| 273 | 35.30 | 20.15 | 28.98 | 44.88 | **129.31** |
| 274 | 12.75 | 20.92 | 6.75 | 24.18 | **64.60** |
| 275 | 23.97 | 34.32 | 46.84 | 42.70 | **147.83** |
| 276 | 34.97 | 39.00 | 47.93 | 44.66 | **166.57** |
| 277 | 0.54 | 6.97 | 5.34 | 5.12 | **17.97** |
| 278 | 27.13 | 20.59 | 79.74 | 52.51 | **179.96** |
| 279 | 45.75 | 27.45 | 27.89 | 26.80 | **127.89** |
| 280 | 25.06 | 30.72 | 58.61 | 56.97 | **171.36** |
| 281 | 32.68 | 51.09 | 52.18 | 28.65 | **164.60** |
| 282 | 36.93 | 39.44 | 25.06 | 44.12 | **145.54** |
| 283 | 41.07 | 36.28 | 87.69 | 95.32 | **260.36** |
| 284 | 49.24 | 34.64 | 42.59 | 37.37 | **163.84** |
| 285 | 19.28 | 7.08 | 26.15 | 22.01 | **74.51** |
| 286 | 47.82 | 40.31 | 55.99 | 61.33 | **205.46** |
| 287 | 42.70 | 49.89 | 48.91 | 37.04 | **178.55** |
| 288 | 10.35 | 27.23 | 46.63 | 50.66 | **134.86** |
| 289 | 0 | 0 | 1.31 | 1.09 | **2.40** |
| **NC - NORTH CAROLINA - Total** | **558.96** | **564.84** | **787.62** | **773.57** | **2,684.98** |

**STATE: ND - NORTH DAKOTA**

| REGISTRANT ZIP CODE 3 | 1ST QUARTER | 2ND QUARTER | 3RD QUARTER | 4TH QUARTER | TOTAL GRAMS |
|---|---|---|---|---|---|
| 580 | 9.26 | 0 | 0 | 0 | **9.26** |
| 581 | 9.15 | 6.86 | 5.56 | 1.53 | **23.09** |

| | | | | | |
|---|---|---|---|---|---|
| 582 | 0.22 | 0 | 0.44 | 0 | **0.65** |
| 583 | 3.27 | 4.36 | 1.09 | 4.36 | **13.07** |
| 584 | 1.31 | 1.31 | 0 | 3.27 | **5.88** |
| 585 | 18.52 | 10.46 | 25.06 | 15.47 | **69.50** |
| 586 | 0 | 0 | 2.18 | 0 | **2.18** |
| 587 | 0 | 0 | 2.18 | 12.20 | **14.38** |
| 588 | 0.54 | 1.85 | 1.31 | 0 | **3.70** |
| **ND - NORTH DAKOTA - Total** | **42.27** | **24.84** | **37.80** | **36.82** | **141.73** |

STATE: NE - NEBRASKA

| REGISTRANT ZIP CODE 3 | 1ST QUARTER | 2ND QUARTER | 3RD QUARTER | 4TH QUARTER | TOTAL GRAMS |
|---|---|---|---|---|---|
| 680 | 1.74 | 0.98 | 5.23 | 1.42 | **9.37** |
| 681 | 34.64 | 41.29 | 60.46 | 47.28 | **183.67** |
| 683 | 2.18 | 2.51 | 5.66 | 0.54 | **10.89** |
| 684 | 5.23 | 3.05 | 8.50 | 8.06 | **24.84** |
| 685 | 7.84 | 7.52 | 21.68 | 11.66 | **48.70** |
| 686 | 4.36 | 3.92 | 4.79 | 3.49 | **16.56** |
| 687 | 3.49 | 6.54 | 8.72 | 4.25 | **22.99** |
| 688 | 8.50 | 11.98 | 31.81 | 19.72 | **72.01** |
| 689 | 25.82 | 19.94 | 14.92 | 19.61 | **80.29** |
| 690 | 0 | 0 | 5.45 | 5.99 | **11.44** |
| 691 | 7.63 | 1.74 | 11.66 | 13.62 | **34.64** |
| 692 | 2.18 | 1.09 | 0 | 0.87 | **4.14** |
| 693 | 2.18 | 0 | 8.06 | 3.59 | **13.84** |
| **NE - NEBRASKA - Total** | **105.78** | **100.55** | **186.94** | **140.09** | **533.36** |

STATE: NH - NEW HAMPSHIRE

| REGISTRANT ZIP CODE 3 | 1ST QUARTER | 2ND QUARTER | 3RD QUARTER | 4TH QUARTER | TOTAL GRAMS |
|---|---|---|---|---|---|
| 030 | 4.25 | 19.61 | 3.49 | 5.45 | **32.79** |
| 031 | 0.54 | 2.18 | 2.18 | 1.63 | **6.54** |
| 032 | 11.98 | 4.36 | 8.72 | 10.89 | **35.95** |
| 033 | 2.18 | 0.54 | 1.63 | 1.09 | **5.45** |
| 034 | 2.72 | 2.18 | 2.40 | 2.18 | **9.48** |
| 037 | 6.10 | 17.43 | 0 | 0 | **23.53** |
| 038 | 13.51 | 1.09 | 10.57 | 5.34 | **30.50** |
| **NH - NEW HAMPSHIRE - Total** | **41.29** | **47.39** | **28.98** | **26.58** | **144.23** |

STATE: NJ - NEW JERSEY

| REGISTRANT ZIP CODE 3 | 1ST QUARTER | 2ND QUARTER | 3RD QUARTER | 4TH QUARTER | TOTAL GRAMS |
|---|---|---|---|---|---|
| 070 | 40.74 | 35.51 | 44.34 | 50.11 | **170.71** |
| 071 | 3.70 | 0.54 | 4.36 | 17.43 | **26.04** |
| 072 | 8.93 | 9.59 | 10.89 | 40.96 | **70.37** |
| 073 | 4.36 | 0 | 0 | 0.54 | **4.90** |
| 074 | 20.92 | 26.15 | 21.13 | 30.07 | **98.26** |
| 075 | 14.71 | 4.36 | 17.97 | 11.98 | **49.02** |
| 076 | 17.97 | 42.05 | 60.46 | 49.02 | **169.51** |
| 077 | 26.04 | 6.32 | 13.73 | 8.39 | **54.47** |
| 078 | 10.78 | 5.88 | 8.93 | 6.65 | **32.25** |
| 079 | 32.14 | 13.07 | 11.44 | 13.07 | **69.72** |
| 080 | 23.97 | 11.77 | 37.80 | 21.24 | **94.78** |
| 081 | 0 | 0.44 | 8.17 | 1.63 | **10.24** |
| 082 | 2.18 | 1.31 | 1.09 | 2.18 | **6.75** |
| 083 | 0.54 | 3.92 | 0 | 4.25 | **8.72** |
| 084 | 0 | 1.63 | 7.08 | 14.16 | **22.88** |
| 085 | 0.54 | 4.79 | 2.61 | 21.79 | **29.74** |
| 086 | 6.54 | 0 | 0.22 | 2.83 | **9.59** |
| 087 | 32.79 | 0.76 | 25.71 | 17.21 | **76.47** |
| 088 | 8.93 | 13.40 | 28.65 | 25.82 | **76.80** |
| 089 | 3.16 | 4.36 | 5.45 | 4.90 | **17.87** |
| **NJ - NEW JERSEY - Total** | **258.94** | **185.85** | **310.04** | **344.24** | **1,099.07** |

STATE: NM - NEW MEXICO

| REGISTRANT ZIP CODE 3 | 1ST QUARTER | 2ND QUARTER | 3RD QUARTER | 4TH QUARTER | TOTAL GRAMS |
|---|---|---|---|---|---|
| 870 | 0.87 | 9.15 | 2.61 | 3.49 | **16.12** |

| | | | | | |
|---|---|---|---|---|---|
| 871 | 111.01 | 126.69 | 130.73 | 107.30 | **475.73** |
| 874 | 4.03 | 3.92 | 1.09 | 1.63 | **10.68** |
| 875 | 6.10 | 6.97 | 25.27 | 17.65 | **55.99** |
| 877 | 6.54 | 6.54 | 6.54 | 2.18 | **21.79** |
| 879 | 0.54 | 0.87 | 0 | 0 | **1.42** |
| 880 | 19.94 | 12.53 | 26.25 | 37.37 | **96.08** |
| 881 | 4.36 | 4.90 | 1.09 | 4.58 | **14.92** |
| 882 | 8.72 | 10.02 | 9.91 | 8.72 | **37.37** |
| 883 | 0 | 4.47 | 3.70 | 8.06 | **16.23** |
| 884 | 0 | 0 | 1.09 | 0 | **1.09** |
| **NM - NEW MEXICO - Total** | **162.10** | **186.07** | **208.29** | **190.97** | **747.42** |

**STATE: NV - NEVADA**

| REGISTRANT ZIP CODE 3 | 1ST QUARTER | 2ND QUARTER | 3RD QUARTER | 4TH QUARTER | TOTAL GRAMS |
|---|---|---|---|---|---|
| 890 | 78.00 | 126.04 | 144.78 | 113.19 | **462.00** |
| 891 | 283.89 | 136.28 | 327.25 | 258.73 | **1,006.15** |
| 893 | 0 | 0.87 | 0 | 0 | **0.87** |
| 894 | 2.18 | 10.57 | 15.47 | 34.42 | **62.64** |
| 895 | 34.97 | 46.30 | 74.84 | 102.62 | **258.73** |
| 897 | 1.31 | 6.54 | 14.16 | 2.18 | **24.18** |
| 898 | 12.53 | 9.91 | 2.72 | 3.81 | **28.98** |
| **NV - NEVADA - Total** | **412.87** | **336.51** | **579.22** | **514.95** | **1,843.55** |

**STATE: NY - NEW YORK**

| REGISTRANT ZIP CODE 3 | 1ST QUARTER | 2ND QUARTER | 3RD QUARTER | 4TH QUARTER | TOTAL GRAMS |
|---|---|---|---|---|---|
| 100 | 29.41 | 39.11 | 86.82 | 89.33 | **244.67** |
| 101 | 0 | 0 | 0.54 | 1.09 | **1.63** |
| 103 | 59.04 | 21.46 | 39.00 | 30.72 | **150.22** |
| 104 | 38.56 | 19.83 | 24.18 | 27.23 | **109.81** |
| 105 | 10.02 | 10.46 | 8.28 | 25.16 | **53.92** |
| 106 | 0 | 2.18 | 2.72 | 3.27 | **8.17** |
| 107 | 0 | 0.54 | 0 | 8.93 | **9.48** |
| 108 | 0 | 6.54 | 0 | 0 | **6.54** |
| 109 | 18.74 | 26.58 | 20.59 | 21.24 | **87.15** |
| 110 | 7.08 | 7.63 | 10.35 | 8.72 | **33.77** |
| 111 | 0 | 0 | 0.54 | 0 | **0.54** |
| 112 | 4.03 | 11.11 | 15.25 | 16.89 | **47.28** |
| 113 | 50.55 | 6.54 | 15.25 | 19.61 | **91.94** |
| 114 | 6.54 | 13.07 | 15.80 | 20.70 | **56.10** |
| 115 | 5.45 | 1.09 | 11.44 | 12.53 | **30.50** |
| 117 | 29.96 | 25.93 | 119.72 | 72.66 | **248.27** |
| 118 | 0 | 0 | 0 | 1.09 | **1.09** |
| 119 | 1.63 | 2.72 | 4.90 | 10.35 | **19.61** |
| 120 | 0 | 0.54 | 1.31 | 0.54 | **2.40** |
| 121 | 14.16 | 2.72 | 1.63 | 1.09 | **19.61** |
| 122 | 16.23 | 29.74 | 42.16 | 34.42 | **122.55** |
| 123 | 0 | 2.29 | 4.25 | 0 | **6.54** |
| 124 | 0 | 0 | 0 | 1.09 | **1.09** |
| 125 | 5.45 | 5.56 | 25.60 | 10.68 | **47.28** |
| 126 | 1.63 | 0 | 4.90 | 4.36 | **10.89** |
| 128 | 50.11 | 37.47 | 124.84 | 107.19 | **319.62** |
| 129 | 0.54 | 1.42 | 0 | 0.54 | **2.51** |
| 130 | 10.46 | 11.55 | 18.30 | 20.26 | **60.57** |
| 131 | 14.27 | 4.36 | 0 | 13.07 | **31.70** |
| 132 | 16.23 | 23.09 | 27.13 | 31.37 | **97.83** |
| 133 | 4.03 | 1.53 | 2.18 | 4.36 | **12.09** |
| 134 | 6.54 | 21.90 | 20.92 | 8.82 | **58.17** |
| 135 | 9.26 | 20.70 | 0.54 | 1.09 | **31.59** |
| 136 | 1.63 | 0.54 | 0.54 | 0.54 | **3.27** |
| 137 | 2.72 | 0.54 | 0.54 | 0.54 | **4.36** |
| 138 | 2.18 | 0 | 0 | 3.49 | **5.66** |
| 139 | 0 | 0 | 0 | 0.54 | **0.54** |

| | | | | | |
|---|---|---|---|---|---|
| 140 | 26.15 | 9.48 | 13.07 | 3.27 | **51.96** |
| 141 | 0.54 | 45.32 | 29.85 | 39.11 | **114.82** |
| 142 | 38.02 | 56.76 | 92.92 | 35.40 | **223.10** |
| 143 | 7.63 | 7.84 | 2.18 | 0 | **17.65** |
| 144 | 13.62 | 0 | 4.36 | 3.16 | **21.13** |
| 146 | 46.73 | 19.06 | 39.87 | 53.16 | **158.83** |
| 147 | 2.18 | 0 | 2.72 | 3.27 | **8.17** |
| 148 | 43.47 | 26.15 | 46.84 | 50.76 | **167.22** |
| 149 | 15.25 | 18.63 | 27.23 | 30.50 | **91.62** |
| **NY - NEW YORK - Total** | **610.05** | **541.96** | **909.30** | **832.17** | **2,893.49** |

**STATE: OH - OHIO**

| REGISTRANT ZIP CODE 3 | 1ST QUARTER | 2ND QUARTER | 3RD QUARTER | 4TH QUARTER | TOTAL GRAMS |
|---|---|---|---|---|---|
| 430 | 13.40 | 17.10 | 40.09 | 13.40 | **83.99** |
| 431 | 62.64 | 64.49 | 141.07 | 102.95 | **371.15** |
| 432 | 21.79 | 37.26 | 49.02 | 46.08 | **154.15** |
| 433 | 0 | 0 | 1.74 | 3.81 | **5.56** |
| 434 | 4.90 | 2.18 | 15.25 | 13.29 | **35.62** |
| 435 | 18.52 | 8.06 | 16.89 | 17.65 | **61.11** |
| 436 | 11.98 | 20.04 | 26.15 | 4.58 | **62.75** |
| 437 | 86.71 | 124.73 | 138.79 | 160.14 | **510.37** |
| 439 | 3.81 | 1.63 | 2.72 | 4.36 | **12.53** |
| 440 | 3.92 | 31.16 | 127.35 | 55.88 | **218.31** |
| 441 | 226.26 | 115.04 | 456.12 | 320.06 | **1,117.48** |
| 442 | 5.45 | 10.78 | 23.42 | 21.68 | **61.33** |
| 443 | 65.25 | 20.81 | 25.27 | 30.94 | **142.27** |
| 444 | 8.17 | 30.61 | 22.55 | 14.38 | **75.71** |
| 445 | 2.72 | 23.09 | 12.75 | 12.64 | **51.20** |
| 446 | 5.34 | 22.22 | 21.02 | 32.68 | **81.27** |
| 447 | 22.88 | 5.34 | 24.40 | 11.44 | **64.06** |
| 448 | 16.34 | 27.89 | 20.59 | 26.69 | **91.51** |
| 449 | 0 | 0 | 4.36 | 10.24 | **14.60** |
| 450 | 8.61 | 15.69 | 22.11 | 57.63 | **104.04** |
| 451 | 30.50 | 29.85 | 12.31 | 2.61 | **75.28** |
| 452 | 246.53 | 37.69 | 441.41 | 232.04 | **957.67** |
| 453 | 14.16 | 17.43 | 27.23 | 14.38 | **73.21** |
| 454 | 24.18 | 14.82 | 18.96 | 35.73 | **93.69** |
| 455 | 5.45 | 3.70 | 5.45 | 4.36 | **18.96** |
| 456 | 4.58 | 8.06 | 9.26 | 8.93 | **30.83** |
| 457 | 0.54 | 4.36 | 3.81 | 6.21 | **14.92** |
| 458 | 55.01 | 94.34 | 32.03 | 31.92 | **213.30** |
| **OH - OHIO - Total** | **969.65** | **788.38** | **1,742.13** | **1,296.68** | **4,796.84** |

**STATE: OK - OKLAHOMA**

| REGISTRANT ZIP CODE 3 | 1ST QUARTER | 2ND QUARTER | 3RD QUARTER | 4TH QUARTER | TOTAL GRAMS |
|---|---|---|---|---|---|
| 730 | 106.21 | 78.65 | 192.60 | 190.42 | **567.89** |
| 731 | 136.06 | 101.53 | 285.74 | 236.83 | **760.17** |
| 734 | 0.44 | 0.54 | 5.45 | 0 | **6.43** |
| 735 | 60.57 | 38.45 | 121.25 | 72.12 | **292.39** |
| 736 | 13.62 | 3.49 | 6.21 | 8.39 | **31.70** |
| 737 | 8.17 | 10.78 | 11.33 | 8.06 | **38.35** |
| 738 | 0.54 | 1.09 | 0 | 3.49 | **5.12** |
| 739 | 6.54 | 6.32 | 1.09 | 3.27 | **17.21** |
| 740 | 42.70 | 55.23 | 47.93 | 63.95 | **209.81** |
| 741 | 107.09 | 75.93 | 153.28 | 118.09 | **454.38** |
| 743 | 10.68 | 9.26 | 7.52 | 1.09 | **28.54** |
| 744 | 25.16 | 17.65 | 35.62 | 46.19 | **124.62** |
| 745 | 28.00 | 28.98 | 5.45 | 22.33 | **84.75** |
| 746 | 1.63 | 6.97 | 0.54 | 0 | **9.15** |
| 747 | 9.80 | 11.98 | 9.80 | 13.84 | **45.43** |
| 748 | 50.11 | 67.54 | 39.44 | 32.68 | **189.77** |
| 749 | 32.14 | 5.88 | 19.61 | 10.02 | **67.65** |

| OK - OKLAHOMA - Total | 639.46 | 520.29 | 942.85 | 830.76 | 2,933.36 |
|---|---|---|---|---|---|

STATE: OR - OREGON

| REGISTRANT ZIP CODE 3 | 1ST QUARTER | 2ND QUARTER | 3RD QUARTER | 4TH QUARTER | TOTAL GRAMS |
|---|---|---|---|---|---|
| 970 | 13.62 | 30.61 | 46.41 | 22.01 | **112.64** |
| 971 | 15.36 | 7.08 | 23.09 | 25.38 | **70.92** |
| 972 | 35.19 | 38.78 | 45.64 | 44.88 | **164.50** |
| 973 | 52.73 | 53.71 | 63.07 | 70.48 | **239.99** |
| 974 | 77.56 | 74.51 | 83.77 | 126.15 | **362.00** |
| 975 | 26.25 | 8.39 | 69.83 | 32.68 | **137.15** |
| 976 | 9.04 | 2.18 | 11.22 | 11.77 | **34.21** |
| 977 | 9.26 | 17.10 | 33.66 | 45.43 | **105.45** |
| 978 | 9.04 | 20.04 | 11.98 | 6.21 | **47.28** |
| 979 | 0 | 0 | 0 | 0.54 | **0.54** |
| OR - OREGON - Total | 248.05 | 252.41 | 388.69 | 385.53 | 1,274.68 |

STATE: PA - PENNSYLVANIA

| REGISTRANT ZIP CODE 3 | 1ST QUARTER | 2ND QUARTER | 3RD QUARTER | 4TH QUARTER | TOTAL GRAMS |
|---|---|---|---|---|---|
| 150 | 1.09 | 6.54 | 17.10 | 11.98 | **36.71** |
| 151 | 4.36 | 19.17 | 8.17 | 9.80 | **41.51** |
| 152 | 25.27 | 52.94 | 34.32 | 27.67 | **140.20** |
| 153 | 21.79 | 17.43 | 0 | 0 | **39.22** |
| 154 | 0 | 1.31 | 7.63 | 19.61 | **28.54** |
| 155 | 9.80 | 2.18 | 6.54 | 2.18 | **20.70** |
| 156 | 0.54 | 0.44 | 1.31 | 15.47 | **17.76** |
| 157 | 7.63 | 0 | 9.70 | 2.18 | **19.50** |
| 158 | 0 | 0 | 2.18 | 1.09 | **3.27** |
| 159 | 2.94 | 0.54 | 0.54 | 3.16 | **7.19** |
| 160 | 11.98 | 16.89 | 10.89 | 7.84 | **47.61** |
| 161 | 7.08 | 4.58 | 4.90 | 2.72 | **19.28** |
| 162 | 0 | 0 | 1.53 | 0 | **1.53** |
| 163 | 1.42 | 0.44 | 1.09 | 0 | **2.94** |
| 164 | 0 | 0 | 0.54 | 0 | **0.54** |
| 165 | 0.54 | 1.74 | 8.72 | 13.07 | **24.08** |
| 166 | 6.54 | 5.88 | 10.35 | 12.09 | **34.86** |
| 168 | 0.54 | 13.07 | 22.88 | 0 | **36.49** |
| 169 | 0.54 | 0.54 | 0 | 1.09 | **2.18** |
| 170 | 17.10 | 7.41 | 13.18 | 21.90 | **59.59** |
| 171 | 25.60 | 7.52 | 28.87 | 13.07 | **75.06** |
| 172 | 0 | 0 | 5.45 | 0 | **5.45** |
| 173 | 35.30 | 42.49 | 67.76 | 57.30 | **202.84** |
| 174 | 1.63 | 0.54 | 8.17 | 10.35 | **20.70** |
| 175 | 2.72 | 4.90 | 14.16 | 6.86 | **28.65** |
| 176 | 1.63 | 0 | 2.72 | 1.09 | **5.45** |
| 177 | 5.23 | 0.76 | 4.90 | 19.39 | **30.28** |
| 178 | 0 | 0.87 | 6.21 | 0.54 | **7.63** |
| 179 | 5.45 | 3.16 | 4.36 | 0 | **12.96** |
| 180 | 22.33 | 2.51 | 10.68 | 1.96 | **37.47** |
| 181 | 12.64 | 10.24 | 8.61 | 9.26 | **40.74** |
| 182 | 0 | 9.80 | 0 | 1.09 | **10.89** |
| 183 | 0.54 | 0 | 1.09 | 1.74 | **3.38** |
| 184 | 8.72 | 3.92 | 1.31 | 0 | **13.94** |
| 185 | 0 | 9.80 | 5.45 | 9.70 | **24.95** |
| 187 | 22.22 | 4.90 | 7.95 | 31.59 | **66.67** |
| 188 | 25.60 | 23.97 | 39.22 | 15.25 | **104.04** |
| 189 | 1.31 | 12.31 | 16.12 | 18.30 | **48.04** |
| 190 | 80.72 | 120.70 | 163.19 | 95.76 | **460.37** |
| 191 | 89.87 | 119.18 | 114.28 | 101.75 | **425.07** |
| 193 | 28.32 | 35.08 | 70.92 | 77.45 | **211.77** |
| 194 | 8.72 | 0.87 | 2.18 | 0 | **11.77** |
| 195 | 8.72 | 4.36 | 0.22 | 4.36 | **17.65** |
| 196 | 1.09 | 0 | 1.63 | 3.27 | **5.99** |

| REGISTRANT ZIP CODE 3 | 1ST QUARTER | 2ND QUARTER | 3RD QUARTER | 4TH QUARTER | TOTAL GRAMS |
|---|---|---|---|---|---|
| **PA - PENNSYLVANIA - Total** | **507.54** | **568.98** | **746.98** | **631.95** | **2,455.45** |
| **STATE: PR - PUERTO RICO** | | | | | |
| REGISTRANT ZIP CODE 3 | 1ST QUARTER | 2ND QUARTER | 3RD QUARTER | 4TH QUARTER | **TOTAL GRAMS** |
| 006 | 626.94 | 441.63 | 1,149.84 | 645.56 | **2,863.97** |
| 007 | 264.72 | 212.54 | 1,204.41 | 565.93 | **2,247.60** |
| 009 | 720.62 | 683.80 | 1,390.70 | 942.09 | **3,737.21** |
| **PR - PUERTO RICO - Total** | **1,612.28** | **1,337.97** | **3,744.94** | **2,153.59** | **8,848.78** |
| **STATE: RI - RHODE ISLAND** | | | | | |
| REGISTRANT ZIP CODE 3 | 1ST QUARTER | 2ND QUARTER | 3RD QUARTER | 4TH QUARTER | **TOTAL GRAMS** |
| 028 | 4.36 | 11.66 | 12.53 | 26.91 | **55.45** |
| 029 | 45.75 | 11.33 | 29.30 | 36.49 | **122.88** |
| **RI - RHODE ISLAND - Total** | **50.11** | **22.99** | **41.83** | **63.40** | **178.33** |
| **STATE: SC - SOUTH CAROLINA** | | | | | |
| REGISTRANT ZIP CODE 3 | 1ST QUARTER | 2ND QUARTER | 3RD QUARTER | 4TH QUARTER | **TOTAL GRAMS** |
| 290 | 34.53 | 10.02 | 29.85 | 18.74 | **93.14** |
| 291 | 44.45 | 34.42 | 39.44 | 39.87 | **158.18** |
| 292 | 92.81 | 62.31 | 72.88 | 93.47 | **321.47** |
| 293 | 15.80 | 15.90 | 32.25 | 18.30 | **82.25** |
| 294 | 75.71 | 70.26 | 92.81 | 103.49 | **342.28** |
| 295 | 69.94 | 92.38 | 157.20 | 75.60 | **395.12** |
| 296 | 97.50 | 84.32 | 108.28 | 104.91 | **395.01** |
| 297 | 5.66 | 20.59 | 8.17 | 11.87 | **46.30** |
| 298 | 48.37 | 30.94 | 19.17 | 32.68 | **131.16** |
| 299 | 0.54 | 10.35 | 7.73 | 4.90 | **23.53** |
| **SC - SOUTH CAROLINA - Total** | **485.32** | **431.50** | **567.78** | **503.84** | **1,988.44** |
| **STATE: SD - SOUTH DAKOTA** | | | | | |
| REGISTRANT ZIP CODE 3 | 1ST QUARTER | 2ND QUARTER | 3RD QUARTER | 4TH QUARTER | **TOTAL GRAMS** |
| 570 | 9.80 | 4.79 | 10.89 | 3.70 | **29.20** |
| 571 | 25.71 | 20.48 | 17.54 | 11.23 | **74.96** |
| 572 | 20.15 | 7.41 | 10.02 | 14.27 | **51.85** |
| 573 | 8.17 | 26.47 | 10.02 | 6.54 | **51.20** |
| 574 | 7.19 | 6.10 | 13.62 | 8.28 | **35.19** |
| 575 | 2.18 | 0 | 0 | 2.18 | **4.36** |
| 576 | 0 | 0 | 1.09 | 0 | **1.09** |
| 577 | 15.47 | 4.79 | 34.86 | 29.20 | **84.32** |
| **SD - SOUTH DAKOTA - Total** | **88.68** | **70.05** | **98.04** | **75.39** | **332.16** |
| **STATE: TN - TENNESSEE** | | | | | |
| REGISTRANT ZIP CODE 3 | 1ST QUARTER | 2ND QUARTER | 3RD QUARTER | 4TH QUARTER | **TOTAL GRAMS** |
| 370 | 85.08 | 122.12 | 137.15 | 131.81 | **476.17** |
| 371 | 229.53 | 143.47 | 214.06 | 205.02 | **792.08** |
| 372 | 105.78 | 71.25 | 139.66 | 159.92 | **476.60** |
| 373 | 69.39 | 114.60 | 129.74 | 118.85 | **432.59** |
| 374 | 91.18 | 63.51 | 124.52 | 106.43 | **385.64** |
| 376 | 33.77 | 29.85 | 40.96 | 38.13 | **142.71** |
| 377 | 30.07 | 24.51 | 31.81 | 24.40 | **110.79** |
| 378 | 35.30 | 45.21 | 60.02 | 56.87 | **197.39** |
| 379 | 55.56 | 84.54 | 64.06 | 54.36 | **258.51** |
| 380 | 18.30 | 20.70 | 35.08 | 73.86 | **147.94** |
| 381 | 125.82 | 112.42 | 173.65 | 118.85 | **530.74** |
| 382 | 16.23 | 10.78 | 9.59 | 10.46 | **47.06** |
| 383 | 87.15 | 49.46 | 83.77 | 104.25 | **324.63** |
| 384 | 26.15 | 36.71 | 46.30 | 45.97 | **155.13** |
| 385 | 32.46 | 14.71 | 37.04 | 33.99 | **118.20** |
| **TN - TENNESSEE - Total** | **1,041.77** | **943.83** | **1,327.40** | **1,283.17** | **4,596.18** |
| **STATE: TX - TEXAS** | | | | | |
| REGISTRANT ZIP CODE 3 | 1ST QUARTER | 2ND QUARTER | 3RD QUARTER | 4TH QUARTER | **TOTAL GRAMS** |
| 750 | 227.35 | 202.41 | 365.70 | 323.22 | **1,118.68** |
| 751 | 21.24 | 17.97 | 19.17 | 42.27 | **100.66** |
| 752 | 90.42 | 216.24 | 216.46 | 97.50 | **620.62** |
| 753 | 3.81 | 2.18 | 3.27 | 7.63 | **16.89** |

| | | | | | |
|---|---|---|---|---|---|
| 754 | 113.51 | 3.27 | 109.37 | 38.24 | **264.39** |
| 755 | 20.70 | 24.95 | 16.01 | 31.05 | **92.71** |
| 756 | 61.55 | 84.86 | 95.43 | 62.42 | **304.26** |
| 757 | 46.84 | 63.07 | 136.50 | 38.13 | **284.54** |
| 758 | 2.72 | 7.73 | 21.79 | 7.63 | **39.87** |
| 759 | 17.43 | 20.48 | 42.27 | 42.81 | **122.99** |
| 760 | 90.31 | 77.35 | 139.33 | 167.87 | **474.86** |
| 761 | 113.62 | 124.30 | 92.71 | 82.79 | **413.42** |
| 762 | 25.06 | 30.83 | 64.16 | 45.97 | **166.02** |
| 763 | 28.00 | 30.94 | 64.49 | 36.60 | **160.03** |
| 764 | 13.40 | 21.46 | 27.56 | 13.51 | **75.93** |
| 765 | 20.15 | 12.53 | 27.56 | 32.35 | **92.60** |
| 766 | 5.99 | 1.53 | 1.42 | 36.28 | **45.21** |
| 767 | 10.35 | 15.80 | 34.86 | 32.68 | **93.69** |
| 768 | 11.44 | 16.12 | 20.48 | 4.90 | **52.94** |
| 769 | 25.93 | 39.98 | 39.00 | 39.44 | **144.34** |
| 770 | 790.78 | 329.54 | 740.56 | 471.48 | **2,332.35** |
| 773 | 104.14 | 177.46 | 175.39 | 187.59 | **644.58** |
| 774 | 127.46 | 92.16 | 136.28 | 92.05 | **447.95** |
| 775 | 84.43 | 56.10 | 114.82 | 129.74 | **385.09** |
| 776 | 11.33 | 9.26 | 22.66 | 15.25 | **58.50** |
| 777 | 51.75 | 109.05 | 208.40 | 27.23 | **396.42** |
| 778 | 52.29 | 23.86 | 67.54 | 40.85 | **184.54** |
| 779 | 21.24 | 28.21 | 15.25 | 22.88 | **87.59** |
| 780 | 72.33 | 65.58 | 80.50 | 55.78 | **274.20** |
| 781 | 5.88 | 38.78 | 41.61 | 38.78 | **125.06** |
| 782 | 248.05 | 228.33 | 583.47 | 391.09 | **1,450.94** |
| 783 | 18.52 | 0 | 13.07 | 14.16 | **45.75** |
| 784 | 17.43 | 61.77 | 66.13 | 70.26 | **215.59** |
| 785 | 324.52 | 406.66 | 276.70 | 197.72 | **1,205.61** |
| 786 | 52.73 | 30.18 | 64.38 | 78.54 | **225.83** |
| 787 | 76.26 | 62.53 | 115.26 | 133.23 | **387.27** |
| 788 | 15.03 | 31.05 | 25.27 | 25.60 | **96.95** |
| 789 | 4.90 | 2.18 | 7.63 | 2.18 | **16.89** |
| 790 | 19.83 | 7.19 | 8.06 | 7.41 | **42.49** |
| 791 | 44.56 | 25.60 | 30.18 | 31.16 | **131.49** |
| 792 | 0 | 1.63 | 5.12 | 8.72 | **15.47** |
| 793 | 37.26 | 31.59 | 51.20 | 22.66 | **142.71** |
| 794 | 71.35 | 102.51 | 125.39 | 61.11 | **360.37** |
| 795 | 9.04 | 12.09 | 16.01 | 5.56 | **42.70** |
| 796 | 68.30 | 67.98 | 41.61 | 53.49 | **231.38** |
| 797 | 115.15 | 82.68 | 238.03 | 167.00 | **602.86** |
| 798 | 0.54 | 0 | 17.65 | 0.65 | **18.85** |
| 799 | 93.69 | 20.15 | 100.77 | 77.56 | **292.17** |
| **TX - TEXAS - Total** | **3,488.61** | **3,118.12** | **4,926.48** | **3,613.02** | **15,146.23** |

**STATE: UT - UTAH**

| REGISTRANT ZIP CODE 3 | 1ST QUARTER | 2ND QUARTER | 3RD QUARTER | 4TH QUARTER | TOTAL GRAMS |
|---|---|---|---|---|---|
| 840 | 179.09 | 231.71 | 249.03 | 245.87 | **905.71** |
| 841 | 78.33 | 203.60 | 122.99 | 181.38 | **586.30** |
| 843 | 14.16 | 10.24 | 11.33 | 19.72 | **55.45** |
| 844 | 17.43 | 30.72 | 116.78 | 358.62 | **523.55** |
| 845 | 64.16 | 45.10 | 52.51 | 46.73 | **208.51** |
| 846 | 65.25 | 115.91 | 45.75 | 51.85 | **278.77** |
| 847 | 105.23 | 73.53 | 97.50 | 108.61 | **384.88** |
| **UT - UTAH - Total** | **523.66** | **710.82** | **695.89** | **1,012.79** | **2,943.16** |

**STATE: VA - VIRGINIA**

| REGISTRANT ZIP CODE 3 | 1ST QUARTER | 2ND QUARTER | 3RD QUARTER | 4TH QUARTER | TOTAL GRAMS |
|---|---|---|---|---|---|
| 201 | 29.63 | 19.83 | 31.81 | 11.87 | **93.14** |
| 220 | 112.97 | 65.91 | 83.01 | 82.14 | **344.02** |
| 221 | 41.40 | 38.35 | 15.36 | 49.02 | **144.12** |

| | | | | | |
|---|---|---|---|---|---|
| 222 | 15.25 | 3.27 | 5.99 | 24.51 | **49.02** |
| 223 | 92.49 | 52.73 | 53.38 | 48.80 | **247.40** |
| 224 | 37.37 | 25.49 | 67.98 | 53.38 | **184.21** |
| 225 | 0.54 | 1.31 | 2.72 | 0.87 | **5.45** |
| 226 | 0.44 | 0 | 10.02 | 0.54 | **11.00** |
| 227 | 4.36 | 6.54 | 17.43 | 5.45 | **33.77** |
| 228 | 4.90 | 0 | 4.36 | 0 | **9.26** |
| 229 | 36.49 | 9.37 | 5.99 | 7.52 | **59.37** |
| 230 | 0 | 2.61 | 6.97 | 0 | **9.59** |
| 231 | 37.80 | 49.02 | 29.85 | 53.92 | **170.60** |
| 232 | 51.20 | 47.61 | 98.37 | 116.13 | **313.30** |
| 233 | 11.87 | 5.01 | 25.82 | 12.53 | **55.23** |
| 234 | 57.19 | 26.36 | 70.16 | 67.54 | **221.25** |
| 235 | 10.13 | 11.00 | 9.15 | 20.37 | **50.66** |
| 236 | 34.86 | 12.64 | 45.97 | 25.82 | **119.29** |
| 237 | 0.54 | 8.28 | 18.08 | 1.31 | **28.21** |
| 238 | 14.38 | 37.91 | 111.99 | 14.38 | **178.66** |
| 239 | 9.37 | 12.64 | 5.45 | 2.72 | **30.18** |
| 240 | 76.80 | 63.18 | 75.93 | 67.54 | **283.46** |
| 241 | 6.54 | 0 | 2.40 | 3.27 | **12.20** |
| 242 | 13.62 | 8.93 | 11.66 | 9.91 | **44.12** |
| 243 | 1.09 | 0 | 1.63 | 3.27 | **5.99** |
| 244 | 9.80 | 0 | 5.45 | 0 | **15.25** |
| 245 | 36.71 | 53.38 | 35.19 | 57.95 | **183.23** |
| 246 | 0 | 0.44 | 0 | 0.54 | **0.98** |
| **VA - VIRGINIA - Total** | **747.75** | **561.79** | **852.11** | **741.32** | **2,902.97** |

**STATE: VI - VIRGIN ISLANDS**

| REGISTRANT ZIP CODE 3 | 1ST QUARTER | 2ND QUARTER | 3RD QUARTER | 4TH QUARTER | TOTAL GRAMS |
|---|---|---|---|---|---|
| 008 | 2.18 | 6.54 | 16.89 | 22.88 | **48.48** |
| **VI - VIRGIN ISLANDS - Total** | **2.18** | **6.54** | **16.89** | **22.88** | **48.48** |

**STATE: VT - VERMONT**

| REGISTRANT ZIP CODE 3 | 1ST QUARTER | 2ND QUARTER | 3RD QUARTER | 4TH QUARTER | TOTAL GRAMS |
|---|---|---|---|---|---|
| 050 | 0 | 0.44 | 0 | 1.09 | **1.53** |
| 051 | 0 | 0 | 1.09 | 0 | **1.09** |
| 054 | 101.86 | 96.30 | 187.70 | 182.36 | **568.22** |
| 056 | 0 | 0 | 0 | 1.09 | **1.09** |
| 057 | 33.77 | 16.34 | 34.86 | 17.43 | **102.40** |
| 058 | 0 | 1.09 | 0 | 0 | **1.09** |
| **VT - VERMONT - Total** | **135.63** | **114.17** | **223.65** | **201.97** | **675.41** |

**STATE: WA - WASHINGTON**

| REGISTRANT ZIP CODE 3 | 1ST QUARTER | 2ND QUARTER | 3RD QUARTER | 4TH QUARTER | TOTAL GRAMS |
|---|---|---|---|---|---|
| 980 | 68.09 | 96.19 | 109.48 | 95.76 | **369.52** |
| 981 | 48.91 | 32.14 | 33.77 | 40.74 | **155.56** |
| 982 | 154.36 | 98.04 | 147.17 | 93.03 | **492.62** |
| 983 | 13.73 | 33.33 | 29.09 | 32.25 | **108.39** |
| 984 | 14.60 | 7.63 | 16.23 | 18.63 | **57.08** |
| 985 | 102.40 | 56.43 | 57.30 | 82.79 | **298.92** |
| 986 | 30.28 | 20.37 | 33.12 | 19.72 | **103.49** |
| 988 | 3.27 | 28.54 | 65.58 | 57.30 | **154.69** |
| 989 | 27.67 | 15.36 | 19.83 | 15.36 | **78.22** |
| 990 | 0 | 0 | 3.92 | 6.10 | **10.02** |
| 991 | 15.80 | 8.93 | 9.80 | 28.11 | **62.64** |
| 992 | 32.25 | 20.04 | 68.09 | 38.89 | **159.27** |
| 993 | 42.05 | 26.04 | 27.67 | 45.86 | **141.62** |
| 994 | 13.07 | 34.86 | 41.40 | 28.76 | **118.09** |
| **WA - WASHINGTON - Total** | **566.48** | **477.91** | **662.45** | **603.30** | **2,310.13** |

**STATE: WI - WISCONSIN**

| REGISTRANT ZIP CODE 3 | 1ST QUARTER | 2ND QUARTER | 3RD QUARTER | 4TH QUARTER | TOTAL GRAMS |
|---|---|---|---|---|---|
| 530 | 26.69 | 33.01 | 38.13 | 28.54 | **126.37** |
| 531 | 7.52 | 7.30 | 19.39 | 17.43 | **51.64** |

| | | | | | |
|---|---|---|---|---|---|
| 532 | 45.32 | 34.64 | 24.73 | 22.01 | **126.69** |
| 534 | 5.66 | 0 | 7.63 | 2.18 | **15.47** |
| 535 | 48.15 | 11.44 | 39.76 | 46.63 | **145.98** |
| 537 | 31.48 | 16.78 | 34.75 | 38.24 | **121.25** |
| 538 | 0 | 0 | 4.36 | 0 | **4.36** |
| 539 | 0 | 0 | 0.54 | 1.63 | **2.18** |
| 540 | 0.98 | 0.87 | 0 | 1.63 | **3.49** |
| 541 | 10.02 | 0 | 15.25 | 15.25 | **40.52** |
| 542 | 26.58 | 23.53 | 17.76 | 0.54 | **68.41** |
| 543 | 68.09 | 106.76 | 44.12 | 47.93 | **266.90** |
| 544 | 18.52 | 24.95 | 50.11 | 38.13 | **131.71** |
| 545 | 1.09 | 0.44 | 1.53 | 0.54 | **3.59** |
| 546 | 7.63 | 8.39 | 12.96 | 4.79 | **33.77** |
| 547 | 5.88 | 3.92 | 9.26 | 3.49 | **22.55** |
| 548 | 4.90 | 0 | 4.03 | 15.80 | **24.73** |
| 549 | 6.65 | 11.66 | 11.98 | 14.16 | **44.45** |
| **WI - WISCONSIN - Total** | **315.16** | **283.67** | **336.29** | **298.92** | **1,234.04** |

**STATE: WV - WEST VIRGINIA**

| REGISTRANT ZIP CODE 3 | 1ST QUARTER | 2ND QUARTER | 3RD QUARTER | 4TH QUARTER | TOTAL GRAMS |
|---|---|---|---|---|---|
| 247 | 4.90 | 8.50 | 5.66 | 4.36 | **23.42** |
| 248 | 8.72 | 6.54 | 21.35 | 13.07 | **49.68** |
| 249 | 3.81 | 3.81 | 4.36 | 13.62 | **25.60** |
| 251 | 1.09 | 0 | 0 | 0 | **1.09** |
| 253 | 10.35 | 13.84 | 32.57 | 22.01 | **78.76** |
| 254 | 1.42 | 1.09 | 1.09 | 1.53 | **5.12** |
| 255 | 8.72 | 3.27 | 0 | 0.87 | **12.85** |
| 256 | 0 | 10.46 | 0 | 0 | **10.46** |
| 257 | 41.94 | 25.60 | 13.62 | 8.72 | **89.87** |
| 258 | 0 | 0 | 3.27 | 1.09 | **4.36** |
| 259 | 4.36 | 1.09 | 1.63 | 2.18 | **9.26** |
| 260 | 5.99 | 3.92 | 1.63 | 2.18 | **13.73** |
| 261 | 2.18 | 0.54 | 11.98 | 5.99 | **20.70** |
| 262 | 0 | 0.54 | 0.54 | 1.63 | **2.72** |
| 263 | 2.18 | 8.82 | 13.62 | 10.35 | **34.97** |
| 264 | 0.54 | 0 | 0.54 | 0 | **1.09** |
| 265 | 22.33 | 1.96 | 2.83 | 3.16 | **30.28** |
| 268 | 0 | 0.44 | 0 | 1.09 | **1.53** |
| **WV - WEST VIRGINIA - Total** | **118.52** | **90.42** | **114.71** | **91.83** | **415.49** |

**STATE: WY - WYOMING**

| REGISTRANT ZIP CODE 3 | 1ST QUARTER | 2ND QUARTER | 3RD QUARTER | 4TH QUARTER | TOTAL GRAMS |
|---|---|---|---|---|---|
| 820 | 1.96 | 6.54 | 4.36 | 3.49 | **16.34** |
| 824 | 0 | 2.18 | 1.09 | 1.63 | **4.90** |
| 825 | 2.72 | 8.17 | 3.81 | 2.94 | **17.65** |
| 826 | 4.14 | 1.63 | 3.27 | 14.49 | **23.53** |
| 827 | 2.18 | 2.72 | 7.84 | 4.79 | **17.54** |
| 828 | 0 | 2.18 | 2.18 | 0.87 | **5.23** |
| 829 | 0.54 | 4.14 | 0.54 | 4.58 | **9.80** |
| 830 | 0 | 0 | 1.09 | 1.09 | **2.18** |
| 831 | 0 | 0 | 0 | 1.09 | **1.09** |
| **WY - WYOMING - Total** | **11.55** | **27.56** | **24.18** | **34.97** | **98.26** |

**DRUG: 9250B - METHADONE**

**STATE: AK - ALASKA**

| REGISTRANT ZIP CODE 3 | 1ST QUARTER | 2ND QUARTER | 3RD QUARTER | 4TH QUARTER | TOTAL GRAMS |
|---|---|---|---|---|---|
| 995 | 6,573.39 | 5,588.12 | 4,689.16 | 5,076.17 | **21,926.84** |
| 996 | 4,215.40 | 2,534.64 | 3,405.61 | 3,354.72 | **13,510.36** |
| 997 | 944.93 | 670.62 | 384.77 | 637.89 | **2,638.21** |
| 998 | 59.48 | 41.14 | 50.53 | 35.33 | **186.48** |
| 999 | 35.78 | 70.66 | 31.30 | 36.22 | **173.96** |
| **AK - ALASKA - Total** | **11,828.98** | **8,905.18** | **8,561.38** | **9,140.32** | **38,435.86** |

**STATE: AL - ALABAMA**

| REGISTRANT ZIP CODE 3 | 1ST QUARTER | 2ND QUARTER | 3RD QUARTER | 4TH QUARTER | TOTAL GRAMS |
|---|---|---|---|---|---|
| 350 | 14,149.68 | 10,269.50 | 12,093.99 | 12,754.59 | **49,267.76** |
| 351 | 753.08 | 733.41 | 734.66 | 684.13 | **2,905.28** |
| 352 | 10,259.57 | 11,523.27 | 816.59 | 838.59 | **23,438.02** |
| 354 | 4,758.03 | 3,659.97 | 3,347.02 | 3,824.90 | **15,589.93** |
| 355 | 5,751.44 | 5,316.31 | 4,810.53 | 5,676.31 | **21,554.59** |
| 356 | 3,866.22 | 2,157.38 | 2,662.00 | 3,299.44 | **11,985.05** |
| 357 | 1,181.06 | 841.18 | 847.44 | 836.71 | **3,706.39** |
| 358 | 10,489.25 | 11,833.54 | 9,361.95 | 6,150.07 | **37,834.82** |
| 359 | 3,723.57 | 2,582.13 | 1,995.41 | 3,466.25 | **11,767.35** |
| 360 | 340.14 | 310.80 | 337.64 | 333.16 | **1,321.74** |
| 361 | 2,174.82 | 2,303.97 | 145.34 | 130.14 | **4,754.27** |
| 362 | 4,230.51 | 3,913.98 | 3,500.68 | 3,630.19 | **15,275.37** |
| 363 | 2,257.47 | 1,717.70 | 2,293.24 | 1,873.77 | **8,142.17** |
| 364 | 234.78 | 251.77 | 205.26 | 205.09 | **896.90** |
| 365 | 1,985.12 | 1,974.84 | 2,140.48 | 2,666.39 | **8,766.82** |
| 366 | 15,790.63 | 10,773.14 | 3,979.28 | 3,614.45 | **34,157.49** |
| 367 | 189.97 | 174.23 | 159.20 | 167.70 | **691.10** |
| 368 | 514.10 | 440.85 | 469.02 | 457.84 | **1,881.82** |
| 369 | 8.05 | 12.52 | 8.94 | 8.94 | **38.46** |
| **AL - ALABAMA - Total** | **82,657.50** | **70,790.51** | **49,908.68** | **50,618.66** | **253,975.34** |

**STATE: AR - ARKANSAS**

| REGISTRANT ZIP CODE 3 | 1ST QUARTER | 2ND QUARTER | 3RD QUARTER | 4TH QUARTER | TOTAL GRAMS |
|---|---|---|---|---|---|
| 716 | 123.87 | 99.73 | 83.45 | 95.25 | **402.30** |
| 717 | 181.56 | 152.94 | 163.23 | 158.31 | **656.04** |
| 718 | 2,250.76 | 707.02 | 1,860.80 | 1,862.59 | **6,681.17** |
| 719 | 209.02 | 207.50 | 210.18 | 214.66 | **841.36** |
| 720 | 2,095.67 | 831.34 | 1,361.72 | 1,710.09 | **5,998.83** |
| 721 | 1,841.21 | 1,042.24 | 1,840.76 | 1,272.73 | **5,996.95** |
| 722 | 691.91 | 1,087.05 | 709.53 | 269.12 | **2,757.61** |
| 723 | 230.31 | 186.93 | 187.82 | 200.79 | **805.85** |
| 724 | 2,641.16 | 2,188.15 | 2,184.66 | 2,159.08 | **9,173.06** |
| 725 | 172.62 | 163.23 | 168.59 | 158.31 | **662.75** |
| 726 | 163.23 | 173.96 | 168.59 | 173.07 | **678.85** |
| 727 | 3,357.31 | 2,860.65 | 2,408.44 | 2,903.67 | **11,530.07** |
| 728 | 93.02 | 81.84 | 79.60 | 86.76 | **341.21** |
| 729 | 371.00 | 321.98 | 337.64 | 279.05 | **1,309.67** |
| **AR - ARKANSAS - Total** | **14,422.65** | **10,104.57** | **11,765.03** | **11,543.48** | **47,835.73** |

**STATE: AZ - ARIZONA**

| REGISTRANT ZIP CODE 3 | 1ST QUARTER | 2ND QUARTER | 3RD QUARTER | 4TH QUARTER | TOTAL GRAMS |
|---|---|---|---|---|---|
| 850 | 39,971.45 | 37,731.70 | 36,946.14 | 38,953.18 | **153,602.47** |
| 851 | 2,112.30 | 1,619.22 | 2,100.68 | 1,420.58 | **7,252.78** |
| 852 | 24,443.86 | 16,468.59 | 21,153.63 | 22,488.44 | **84,554.52** |
| 853 | 18,100.42 | 15,132.53 | 15,207.57 | 17,903.83 | **66,344.36** |
| 855 | 1,451.16 | 812.03 | 1,801.32 | 907.01 | **4,971.52** |
| 856 | 1,778.34 | 792.53 | 1,553.57 | 1,999.34 | **6,123.78** |
| 857 | 33,604.58 | 26,492.22 | 32,455.09 | 32,349.55 | **124,901.44** |
| 859 | 1,101.54 | 203.03 | 1,084.19 | 165.29 | **2,554.05** |
| 860 | 826.42 | 833.13 | 828.03 | 825.89 | **3,313.47** |
| 863 | 1,672.53 | 1,002.89 | 957.72 | 1,653.75 | **5,286.89** |
| 864 | 4,413.86 | 2,902.78 | 3,799.86 | 4,053.42 | **15,169.92** |
| 865 | 4.47 | 0 | 3.58 | 5.37 | **13.42** |
| **AZ - ARIZONA - Total** | **129,480.94** | **103,990.64** | **117,891.39** | **122,725.63** | **474,088.60** |

**STATE: CA - CALIFORNIA**

| REGISTRANT ZIP CODE 3 | 1ST QUARTER | 2ND QUARTER | 3RD QUARTER | 4TH QUARTER | TOTAL GRAMS |
|---|---|---|---|---|---|
| 900 | 14,963.57 | 11,689.76 | 13,199.53 | 12,461.90 | **52,314.76** |
| 902 | 4,737.82 | 2,714.95 | 3,199.36 | 3,006.70 | **13,658.83** |
| 903 | 1,994.51 | 875.62 | 875.62 | 1,010.22 | **4,755.97** |
| 904 | 180.95 | 143.11 | 168.42 | 191.59 | **684.07** |
| 905 | 246.76 | 246.94 | 276.64 | 265.01 | **1,035.36** |

| | | | | | |
|---|---|---|---|---|---|
| 906 | 6,524.29 | 5,132.07 | 5,441.53 | 4,833.34 | **21,931.22** |
| 907 | 4,603.48 | 3,824.01 | 4,624.05 | 3,142.92 | **16,194.45** |
| 908 | 1,444.30 | 1,283.55 | 1,379.52 | 1,278.90 | **5,386.28** |
| 910 | 286.21 | 282.54 | 287.01 | 326.28 | **1,182.04** |
| 911 | 1,016.40 | 1,011.48 | 580.20 | 1,005.22 | **3,613.29** |
| 912 | 1,595.88 | 1,029.45 | 1,547.31 | 952.09 | **5,124.73** |
| 913 | 5,255.94 | 3,958.97 | 4,912.85 | 4,211.55 | **18,339.31** |
| 914 | 3,236.21 | 2,926.75 | 2,802.60 | 3,569.55 | **12,535.11** |
| 915 | 190.78 | 150.26 | 127.90 | 144.09 | **613.02** |
| 916 | 923.92 | 988.94 | 639.32 | 780.27 | **3,332.44** |
| 917 | 8,856.35 | 7,668.76 | 8,475.51 | 6,572.86 | **31,573.48** |
| 918 | 81.84 | 66.63 | 59.48 | 67.08 | **275.03** |
| 919 | 3,226.37 | 2,518.81 | 2,891.86 | 2,370.34 | **11,007.38** |
| 920 | 12,274.84 | 10,821.61 | 10,362.61 | 11,318.81 | **44,777.87** |
| 921 | 10,139.72 | 7,974.29 | 9,856.02 | 7,912.67 | **35,882.70** |
| 922 | 7,869.02 | 4,486.94 | 6,398.09 | 6,885.63 | **25,639.68** |
| 923 | 8,912.25 | 7,848.81 | 6,755.05 | 7,818.13 | **31,334.23** |
| 924 | 110.01 | 136.75 | 109.74 | 114.84 | **471.35** |
| 925 | 8,516.83 | 7,743.18 | 8,159.61 | 7,459.47 | **31,879.10** |
| 926 | 1,827.17 | 1,523.25 | 1,842.29 | 2,132.25 | **7,324.96** |
| 927 | 4,058.25 | 3,678.13 | 3,529.03 | 4,198.49 | **15,463.91** |
| 928 | 4,907.70 | 4,481.72 | 4,528.30 | 4,427.82 | **18,345.54** |
| 930 | 12,115.72 | 8,920.30 | 10,902.47 | 9,146.76 | **41,085.25** |
| 931 | 2,397.71 | 2,652.88 | 2,355.67 | 2,561.11 | **9,967.37** |
| 932 | 6,907.81 | 7,023.54 | 6,360.88 | 7,450.80 | **27,743.04** |
| 933 | 10,401.96 | 9,928.29 | 9,120.64 | 9,098.91 | **38,549.80** |
| 934 | 6,919.35 | 4,905.34 | 6,530.28 | 4,793.72 | **23,148.68** |
| 935 | 3,570.89 | 2,598.32 | 3,613.02 | 2,779.26 | **12,561.49** |
| 936 | 1,657.38 | 1,349.94 | 1,540.82 | 1,283.39 | **5,831.52** |
| 937 | 15,394.60 | 16,081.85 | 14,564.32 | 15,840.94 | **61,881.70** |
| 939 | 2,741.78 | 1,298.85 | 2,386.62 | 2,347.80 | **8,775.05** |
| 940 | 4,142.59 | 4,411.36 | 4,115.22 | 4,012.55 | **16,681.72** |
| 941 | 18,289.24 | 17,790.11 | 16,378.49 | 17,579.89 | **70,037.72** |
| 943 | 399.17 | 366.68 | 356.02 | 340.63 | **1,462.50** |
| 944 | 188.00 | 189.08 | 179.24 | 184.25 | **740.56** |
| 945 | 20,591.95 | 18,061.96 | 18,333.77 | 19,619.65 | **76,607.33** |
| 946 | 9,094.44 | 8,747.68 | 8,050.58 | 8,225.80 | **34,118.50** |
| 947 | 1,256.81 | 888.59 | 890.64 | 902.63 | **3,938.67** |
| 948 | 3,574.11 | 2,453.88 | 1,840.68 | 3,442.01 | **11,310.67** |
| 949 | 2,689.55 | 2,596.53 | 2,806.63 | 2,489.56 | **10,582.27** |
| 950 | 5,814.40 | 6,209.46 | 5,915.83 | 5,928.98 | **23,868.67** |
| 951 | 1,587.92 | 3,217.51 | 2,364.79 | 2,349.23 | **9,519.46** |
| 952 | 16,086.77 | 10,939.32 | 12,289.59 | 11,876.38 | **51,192.06** |
| 953 | 15,345.94 | 14,983.79 | 13,897.01 | 15,001.32 | **59,228.06** |
| 954 | 6,074.41 | 4,984.31 | 5,755.02 | 4,964.64 | **21,778.37** |
| 955 | 838.14 | 1,463.24 | 800.22 | 2,269.54 | **5,371.14** |
| 956 | 12,566.14 | 11,003.27 | 12,832.14 | 10,674.57 | **47,076.12** |
| 957 | 265.99 | 254.46 | 278.87 | 271.45 | **1,070.78** |
| 958 | 17,432.61 | 17,072.86 | 16,899.31 | 14,408.87 | **65,813.65** |
| 959 | 10,853.90 | 8,752.38 | 9,830.63 | 8,673.65 | **38,110.56** |
| 960 | 3,914.43 | 3,990.37 | 4,673.15 | 4,305.10 | **16,883.05** |
| 961 | 160.99 | 149.63 | 141.05 | 154.28 | **605.96** |
| **CA - CALIFORNIA - Total** | **331,256.05** | **288,493.06** | **299,033.04** | **291,435.70** | **1,210,217.84** |

**STATE: CO - COLORADO**

| REGISTRANT ZIP CODE 3 | 1ST QUARTER | 2ND QUARTER | 3RD QUARTER | 4TH QUARTER | TOTAL GRAMS |
|---|---|---|---|---|---|
| 800 | 13,567.50 | 8,114.88 | 9,230.28 | 10,579.40 | **41,492.06** |
| 801 | 4,566.81 | 4,190.44 | 4,779.76 | 4,571.55 | **18,108.56** |
| 802 | 22,556.14 | 16,724.48 | 15,923.99 | 15,757.00 | **70,961.61** |
| 803 | 1,401.08 | 1,091.17 | 1,043.14 | 1,273.09 | **4,808.47** |
| 804 | 120.30 | 139.97 | 111.35 | 104.91 | **476.54** |

| | | | | | |
|---|---|---|---|---|---|
| 805 | 3,602.02 | 2,800.81 | 3,126.46 | 2,855.10 | **12,384.40** |
| 806 | 2,886.68 | 1,276.40 | 2,494.75 | 2,587.32 | **9,245.14** |
| 807 | 42.48 | 42.48 | 37.56 | 36.58 | **159.11** |
| 808 | 122.53 | 143.55 | 140.87 | 147.58 | **554.53** |
| 809 | 7,081.95 | 5,398.51 | 4,954.17 | 6,258.56 | **23,693.19** |
| 810 | 6,295.14 | 5,418.72 | 4,707.58 | 5,431.87 | **21,853.32** |
| 811 | 1,027.67 | 567.77 | 575.10 | 851.02 | **3,021.55** |
| 812 | 108.31 | 128.97 | 123.34 | 129.24 | **489.86** |
| 813 | 678.85 | 463.30 | 680.64 | 251.77 | **2,074.56** |
| 814 | 43.92 | 35.33 | 40.61 | 40.52 | **160.37** |
| 815 | 2,801.62 | 1,886.02 | 2,724.61 | 3,158.75 | **10,571.00** |
| 816 | 84.52 | 68.15 | 79.96 | 92.84 | **325.47** |
| **CO - COLORADO - Total** | **66,987.51** | **48,490.95** | **50,774.18** | **54,127.11** | **220,379.75** |

**STATE: CT - CONNECTICUT**

| REGISTRANT ZIP CODE 3 | 1ST QUARTER | 2ND QUARTER | 3RD QUARTER | 4TH QUARTER | **TOTAL GRAMS** |
|---|---|---|---|---|---|
| 060 | 19,180.14 | 15,378.40 | 10,697.11 | 19,068.61 | **64,324.26** |
| 061 | 17,108.71 | 12,715.95 | 13,504.72 | 13,319.23 | **56,648.61** |
| 062 | 10,364.31 | 1,761.97 | 6,048.83 | 6,304.63 | **24,479.73** |
| 063 | 9,568.56 | 9,254.00 | 9,292.91 | 9,074.22 | **37,189.69** |
| 064 | 16,901.83 | 7,745.24 | 16,661.60 | 7,864.64 | **49,173.31** |
| 065 | 28,918.37 | 34,843.14 | 17,615.03 | 26,641.76 | **108,018.30** |
| 066 | 29,205.29 | 5,531.24 | 15,481.17 | 22,222.71 | **72,440.41** |
| 067 | 15,858.43 | 10,904.35 | 9,347.37 | 12,723.11 | **48,833.26** |
| 068 | 7,118.44 | 6,367.14 | 6,340.76 | 6,332.44 | **26,158.79** |
| 069 | 1,898.10 | 3,670.08 | 1,885.84 | 1,870.10 | **9,324.12** |
| **CT - CONNECTICUT - Total** | **156,122.17** | **108,171.51** | **106,875.34** | **125,421.44** | **496,590.47** |

**STATE: DC - DISTRICT OF COLUMBIA**

| REGISTRANT ZIP CODE 3 | 1ST QUARTER | 2ND QUARTER | 3RD QUARTER | 4TH QUARTER | **TOTAL GRAMS** |
|---|---|---|---|---|---|
| 200 | 9,843.89 | 8,839.43 | 9,564.55 | 9,166.62 | **37,414.50** |
| 204 | 627.87 | 866.67 | 630.55 | 574.20 | **2,699.30** |
| **DC - DISTRICT OF COLUMBIA - Total** | **10,471.76** | **9,706.11** | **10,195.11** | **9,740.83** | **40,113.80** |

**STATE: DE - DELAWARE**

| REGISTRANT ZIP CODE 3 | 1ST QUARTER | 2ND QUARTER | 3RD QUARTER | 4TH QUARTER | **TOTAL GRAMS** |
|---|---|---|---|---|---|
| 197 | 16,081.13 | 9,869.97 | 14,614.76 | 13,074.16 | **53,640.03** |
| 198 | 10,502.40 | 6,539.85 | 10,130.15 | 8,298.51 | **35,470.92** |
| 199 | 18,103.19 | 15,212.22 | 17,731.57 | 16,361.80 | **67,408.78** |
| **DE - DELAWARE - Total** | **44,686.73** | **31,622.05** | **42,476.49** | **37,734.47** | **156,519.73** |

**STATE: FL - FLORIDA**

| REGISTRANT ZIP CODE 3 | 1ST QUARTER | 2ND QUARTER | 3RD QUARTER | 4TH QUARTER | **TOTAL GRAMS** |
|---|---|---|---|---|---|
| 320 | 7,588.27 | 6,887.95 | 2,589.65 | 1,931.64 | **18,997.50** |
| 321 | 7,747.65 | 10,506.43 | 1,559.48 | 1,493.65 | **21,307.20** |
| 322 | 27,592.29 | 24,123.33 | 8,968.86 | 9,384.40 | **70,068.88** |
| 323 | 3,560.79 | 3,817.03 | 855.40 | 759.97 | **8,993.19** |
| 324 | 4,238.56 | 4,196.52 | 461.06 | 466.88 | **9,363.03** |
| 325 | 16,865.97 | 8,996.68 | 6,036.93 | 5,707.79 | **37,607.37** |
| 326 | 4,080.07 | 4,301.08 | 1,433.28 | 1,323.35 | **11,137.78** |
| 327 | 1,587.20 | 1,449.55 | 1,448.57 | 1,315.93 | **5,801.26** |
| 328 | 15,754.95 | 12,164.85 | 3,175.16 | 2,943.43 | **34,038.39** |
| 329 | 4,997.28 | 5,516.75 | 2,765.31 | 3,733.94 | **17,013.28** |
| 330 | 7,439.08 | 5,010.61 | 1,153.24 | 1,221.93 | **14,824.86** |
| 331 | 2,437.78 | 2,116.98 | 2,532.18 | 2,110.29 | **9,197.23** |
| 333 | 3,669.45 | 2,460.76 | 1,429.34 | 1,590.06 | **9,149.62** |
| 334 | 10,138.65 | 8,680.15 | 4,516.36 | 6,219.84 | **29,555.00** |
| 335 | 1,445.17 | 1,311.19 | 1,370.76 | 1,281.41 | **5,408.53** |
| 336 | 12,300.03 | 10,816.60 | 6,407.43 | 5,489.34 | **35,013.40** |
| 337 | 22,243.73 | 17,690.83 | 12,950.01 | 12,582.61 | **65,467.17** |
| 338 | 3,006.70 | 2,398.15 | 2,504.32 | 2,850.54 | **10,759.72** |
| 339 | 16,263.77 | 14,185.01 | 14,441.43 | 15,092.11 | **59,982.31** |
| 341 | 3,941.62 | 3,297.38 | 1,090.36 | 1,101.01 | **9,430.37** |
| 342 | 16,012.98 | 12,161.43 | 10,904.97 | 10,432.28 | **49,511.66** |

| | | | | | |
|---|---|---|---|---|---|
| 344 | 11,584.09 | 9,048.47 | 1,985.75 | 1,937.27 | **24,555.57** |
| 346 | 18,752.44 | 18,442.98 | 13,987.43 | 15,650.03 | **66,832.88** |
| 347 | 4,503.13 | 4,504.02 | 860.59 | 800.04 | **10,667.78** |
| 349 | 2,951.52 | 3,311.52 | 1,003.96 | 1,674.32 | **8,941.32** |
| **FL - FLORIDA - Total** | **230,703.17** | **197,396.24** | **106,431.84** | **109,094.06** | **643,625.30** |

**STATE: GA - GEORGIA**

| REGISTRANT ZIP CODE 3 | 1ST QUARTER | 2ND QUARTER | 3RD QUARTER | 4TH QUARTER | TOTAL GRAMS |
|---|---|---|---|---|---|
| 300 | 18,031.19 | 12,022.26 | 12,493.25 | 11,925.84 | **54,472.54** |
| 301 | 15,417.58 | 11,024.73 | 11,725.49 | 12,494.95 | **50,662.75** |
| 302 | 8,913.23 | 7,313.51 | 7,661.88 | 6,344.43 | **30,233.05** |
| 303 | 4,050.38 | 3,468.39 | 3,258.12 | 4,151.98 | **14,928.88** |
| 304 | 1,686.93 | 1,134.55 | 1,157.26 | 1,704.73 | **5,683.46** |
| 305 | 12,961.56 | 13,383.18 | 11,840.25 | 13,477.53 | **51,662.51** |
| 306 | 6,304.18 | 5,416.49 | 6,847.53 | 7,430.41 | **25,998.60** |
| 307 | 28,953.07 | 22,593.89 | 23,332.48 | 22,277.36 | **97,156.79** |
| 308 | 1,183.74 | 878.75 | 276.82 | 281.29 | **2,620.59** |
| 309 | 3,695.12 | 4,309.22 | 432.35 | 392.19 | **8,828.89** |
| 310 | 6,108.57 | 3,609.89 | 4,806.60 | 3,642.53 | **18,167.59** |
| 312 | 2,744.02 | 2,559.33 | 1,844.70 | 2,773.71 | **9,921.76** |
| 313 | 204.46 | 175.30 | 194.53 | 180.94 | **755.23** |
| 314 | 4,282.03 | 4,414.31 | 3,700.58 | 2,994.45 | **15,391.37** |
| 315 | 3,166.18 | 2,313.81 | 1,014.25 | 945.83 | **7,440.07** |
| 316 | 1,979.04 | 1,411.09 | 272.61 | 261.16 | **3,923.91** |
| 317 | 4,144.65 | 1,828.24 | 2,708.87 | 2,361.66 | **11,043.43** |
| 318 | 81.57 | 73.52 | 66.28 | 58.05 | **279.41** |
| 319 | 3,665.70 | 2,603.78 | 1,881.10 | 2,138.33 | **10,288.91** |
| 398 | 745.04 | 758.90 | 1,277.20 | 726.70 | **3,507.84** |
| **GA - GEORGIA - Total** | **128,318.23** | **101,293.13** | **96,792.15** | **96,564.07** | **422,967.57** |

**STATE: GU - GUAM**

| REGISTRANT ZIP CODE 3 | 1ST QUARTER | 2ND QUARTER | 3RD QUARTER | 4TH QUARTER | TOTAL GRAMS |
|---|---|---|---|---|---|
| 969 | 10.73 | 10.73 | 20.12 | 10.73 | **52.32** |
| **GU - GUAM - Total** | **10.73** | **10.73** | **20.12** | **10.73** | **52.32** |

**STATE: HI - HAWAII**

| REGISTRANT ZIP CODE 3 | 1ST QUARTER | 2ND QUARTER | 3RD QUARTER | 4TH QUARTER | TOTAL GRAMS |
|---|---|---|---|---|---|
| 967 | 3,497.28 | 2,610.75 | 2,967.80 | 3,171.99 | **12,247.82** |
| 968 | 4,667.69 | 3,929.28 | 3,879.73 | 4,126.58 | **16,603.28** |
| **HI - HAWAII - Total** | **8,164.98** | **6,540.03** | **6,847.53** | **7,298.57** | **28,851.11** |

**STATE: IA - IOWA**

| REGISTRANT ZIP CODE 3 | 1ST QUARTER | 2ND QUARTER | 3RD QUARTER | 4TH QUARTER | TOTAL GRAMS |
|---|---|---|---|---|---|
| 500 | 946.72 | 720.26 | 423.77 | 668.83 | **2,759.58** |
| 501 | 509.45 | 141.67 | 487.45 | 480.74 | **1,619.31** |
| 502 | 332.27 | 141.40 | 1,030.35 | 2,732.39 | **4,236.42** |
| 503 | 4,881.64 | 3,771.68 | 3,017.97 | 2,292.79 | **13,964.09** |
| 504 | 134.96 | 119.40 | 134.61 | 120.74 | **509.72** |
| 505 | 150.89 | 395.32 | 132.46 | 126.56 | **805.23** |
| 506 | 72.54 | 58.58 | 74.68 | 78.98 | **284.78** |
| 507 | 944.67 | 1,046.45 | 601.04 | 1,229.08 | **3,821.23** |
| 508 | 56.79 | 47.40 | 51.43 | 50.98 | **206.61** |
| 510 | 115.82 | 135.50 | 114.48 | 114.93 | **480.74** |
| 511 | 248.64 | 264.30 | 233.89 | 212.87 | **959.69** |
| 512 | 6.26 | 4.92 | 5.37 | 3.58 | **20.12** |
| 513 | 19.23 | 17.44 | 19.68 | 20.12 | **76.47** |
| 514 | 109.56 | 19.68 | 29.96 | 23.25 | **182.46** |
| 515 | 195.43 | 299.62 | 275.92 | 145.34 | **916.31** |
| 516 | 12.52 | 14.31 | 10.29 | 7.16 | **44.27** |
| 520 | 83.81 | 77.81 | 81.84 | 99.90 | **343.36** |
| 521 | 187.20 | 4.92 | 6.26 | 10.73 | **209.11** |
| 522 | 149.90 | 125.84 | 142.03 | 168.33 | **586.10** |
| 523 | 96.60 | 87.92 | 87.56 | 90.96 | **363.04** |
| 524 | 2,983.54 | 1,950.42 | 2,771.12 | 2,551.01 | **10,256.08** |

| | | | | | |
|---|---|---|---|---|---|
| 525 | 366.26 | 428.42 | 397.56 | 388.17 | **1,580.40** |
| 526 | 239.43 | 406.68 | 314.11 | 302.84 | **1,263.07** |
| 527 | 273.24 | 78.26 | 285.31 | 93.91 | **730.72** |
| 528 | 2,525.25 | 934.38 | 2,496.00 | 1,715.91 | **7,671.54** |
| **IA - IOWA - Total** | **15,642.61** | **11,292.60** | **13,225.14** | **13,730.11** | **53,890.46** |

**STATE: ID - IDAHO**

| REGISTRANT ZIP CODE 3 | 1ST QUARTER | 2ND QUARTER | 3RD QUARTER | 4TH QUARTER | TOTAL GRAMS |
|---|---|---|---|---|---|
| 832 | 610.16 | 604.97 | 589.95 | 568.39 | **2,373.47** |
| 833 | 200.35 | 168.59 | 191.40 | 173.96 | **734.30** |
| 834 | 266.08 | 256.07 | 283.52 | 267.96 | **1,073.64** |
| 835 | 272.79 | 230.31 | 242.38 | 237.02 | **982.50** |
| 836 | 4,005.21 | 3,536.55 | 3,089.62 | 4,192.05 | **14,823.43** |
| 837 | 5,977.81 | 4,494.27 | 5,744.64 | 5,677.03 | **21,893.75** |
| 838 | 748.17 | 713.11 | 647.10 | 656.40 | **2,764.77** |
| **ID - IDAHO - Total** | **12,080.57** | **10,003.86** | **10,788.61** | **11,772.81** | **44,645.85** |

**STATE: IL - ILLINOIS**

| REGISTRANT ZIP CODE 3 | 1ST QUARTER | 2ND QUARTER | 3RD QUARTER | 4TH QUARTER | TOTAL GRAMS |
|---|---|---|---|---|---|
| 600 | 12,045.15 | 8,370.15 | 9,173.77 | 8,855.36 | **38,444.44** |
| 601 | 8,362.28 | 6,842.96 | 7,234.09 | 7,421.11 | **29,860.44** |
| 602 | 4,330.33 | 3,134.07 | 3,555.15 | 3,534.31 | **14,553.86** |
| 603 | 59.03 | 65.29 | 55.90 | 74.68 | **254.90** |
| 604 | 8,999.36 | 8,721.29 | 6,562.39 | 9,092.29 | **33,375.34** |
| 605 | 7,257.43 | 6,140.23 | 6,336.29 | 6,814.17 | **26,548.12** |
| 606 | 63,182.56 | 50,529.75 | 52,717.90 | 54,893.62 | **221,323.84** |
| 607 | 238.63 | 413.57 | 448.36 | 425.29 | **1,525.85** |
| 608 | 3,919.71 | 3,561.05 | 2,978.80 | 4,099.48 | **14,559.04** |
| 609 | 1,569.05 | 974.81 | 911.75 | 1,291.78 | **4,747.39** |
| 610 | 274.13 | 602.02 | 1,113.98 | 605.96 | **2,596.09** |
| 611 | 3,601.39 | 1,965.62 | 2,732.48 | 2,720.85 | **11,020.35** |
| 612 | 891.27 | 584.94 | 701.21 | 840.11 | **3,017.53** |
| 613 | 1,527.46 | 170.38 | 173.07 | 1,515.56 | **3,386.47** |
| 614 | 106.43 | 117.61 | 105.99 | 112.87 | **442.91** |
| 615 | 118.51 | 114.93 | 105.99 | 113.59 | **453.01** |
| 616 | 8,588.36 | 6,516.83 | 6,948.56 | 7,244.28 | **29,298.02** |
| 617 | 1,532.55 | 1,280.07 | 1,390.17 | 1,707.77 | **5,910.55** |
| 618 | 2,407.72 | 2,399.05 | 449.88 | 241.40 | **5,498.06** |
| 619 | 51.34 | 52.05 | 54.02 | 49.28 | **206.70** |
| 620 | 4,694.35 | 3,886.35 | 4,004.77 | 4,068.63 | **16,654.09** |
| 622 | 1,276.13 | 454.89 | 1,267.90 | 749.15 | **3,748.07** |
| 623 | 269.75 | 386.56 | 460.17 | 453.01 | **1,569.49** |
| 624 | 123.87 | 93.46 | 94.81 | 76.02 | **388.17** |
| 625 | 2,283.94 | 2,296.82 | 2,287.88 | 2,272.49 | **9,141.13** |
| 626 | 99.73 | 115.38 | 103.30 | 105.99 | **424.39** |
| 627 | 3,638.42 | 3,622.32 | 4,022.12 | 3,902.71 | **15,185.57** |
| 628 | 185.14 | 160.10 | 184.69 | 172.62 | **702.55** |
| 629 | 871.15 | 894.40 | 1,440.25 | 857.82 | **4,063.62** |
| **IL - ILLINOIS - Total** | **142,505.17** | **114,466.97** | **117,615.62** | **124,312.20** | **498,899.97** |

**STATE: IN - INDIANA**

| REGISTRANT ZIP CODE 3 | 1ST QUARTER | 2ND QUARTER | 3RD QUARTER | 4TH QUARTER | TOTAL GRAMS |
|---|---|---|---|---|---|
| 460 | 315.54 | 309.91 | 338.80 | 370.82 | **1,335.07** |
| 461 | 5,551.27 | 3,418.40 | 4,596.77 | 5,789.27 | **19,355.71** |
| 462 | 16,150.78 | 15,104.98 | 15,658.44 | 14,267.13 | **61,181.34** |
| 463 | 3,455.96 | 4,126.31 | 3,133.62 | 3,818.64 | **14,534.54** |
| 464 | 8,425.25 | 5,873.08 | 3,657.65 | 2,808.42 | **20,764.39** |
| 465 | 1,041.98 | 961.57 | 1,002.00 | 974.63 | **3,980.17** |
| 466 | 3,965.50 | 3,982.32 | 3,994.84 | 4,885.57 | **16,828.23** |
| 467 | 648.35 | 570.45 | 621.07 | 606.76 | **2,446.63** |
| 468 | 9,471.96 | 7,087.32 | 8,018.83 | 7,145.99 | **31,724.10** |
| 469 | 9,018.86 | 10,827.70 | 8,363.36 | 10,171.12 | **38,381.03** |
| 470 | 6,588.60 | 3,385.30 | 5,321.68 | 3,378.15 | **18,673.73** |

| | | | | | |
|---|---|---|---|---|---|
| 471 | 12,973.63 | 9,144.17 | 10,693.98 | 8,978.17 | **41,789.95** |
| 472 | 253.20 | 210.18 | 213.58 | 202.13 | **879.11** |
| 473 | 6,940.63 | 5,321.05 | 6,260.89 | 6,209.10 | **24,731.68** |
| 474 | 1,830.39 | 2,056.23 | 435.13 | 1,878.51 | **6,200.25** |
| 475 | 155.63 | 149.45 | 531.01 | 136.75 | **972.84** |
| 476 | 178.43 | 171.28 | 174.41 | 183.35 | **707.47** |
| 477 | 4,115.04 | 3,189.79 | 3,530.20 | 2,968.96 | **13,803.99** |
| 478 | 1,870.64 | 978.92 | 1,877.79 | 1,973.94 | **6,701.29** |
| 479 | 1,954.26 | 2,212.21 | 218.05 | 2,092.99 | **6,477.51** |
| **IN - INDIANA - Total** | **94,905.92** | **79,080.61** | **78,642.09** | **78,840.40** | **331,469.02** |

**STATE: KS - KANSAS**

| REGISTRANT ZIP CODE 3 | 1ST QUARTER | 2ND QUARTER | 3RD QUARTER | 4TH QUARTER | TOTAL GRAMS |
|---|---|---|---|---|---|
| 660 | 1,220.95 | 1,168.80 | 1,129.72 | 1,585.06 | **5,104.52** |
| 661 | 3,322.34 | 3,451.31 | 3,211.25 | 3,029.78 | **13,014.68** |
| 662 | 1,833.07 | 1,610.99 | 1,378.72 | 1,616.36 | **6,439.14** |
| 664 | 69.32 | 41.14 | 69.32 | 53.66 | **233.44** |
| 665 | 119.49 | 114.39 | 104.64 | 117.61 | **456.14** |
| 666 | 1,819.93 | 1,613.50 | 1,919.47 | 1,592.93 | **6,945.82** |
| 667 | 279.95 | 243.28 | 226.28 | 258.48 | **1,007.99** |
| 668 | 75.58 | 67.08 | 59.48 | 57.24 | **259.38** |
| 669 | 49.19 | 51.43 | 45.61 | 49.64 | **195.87** |
| 670 | 419.92 | 419.92 | 405.16 | 395.32 | **1,640.33** |
| 671 | 154.73 | 144.89 | 155.18 | 154.73 | **609.53** |
| 672 | 10,221.47 | 9,834.64 | 10,492.26 | 9,793.92 | **40,342.30** |
| 673 | 105.54 | 93.91 | 67.97 | 65.74 | **333.16** |
| 674 | 130.14 | 153.39 | 150.26 | 141.76 | **575.55** |
| 675 | 145.34 | 129.24 | 137.29 | 126.11 | **537.98** |
| 676 | 46.06 | 41.14 | 63.95 | 77.81 | **228.97** |
| 677 | 22.36 | 19.68 | 20.57 | 22.36 | **84.97** |
| 678 | 123.43 | 107.78 | 111.62 | 100.89 | **443.71** |
| 679 | 26.83 | 33.09 | 25.94 | 19.68 | **105.54** |
| **KS - KANSAS - Total** | **20,185.62** | **19,339.61** | **19,774.70** | **19,259.09** | **78,559.02** |

**STATE: KY - KENTUCKY**

| REGISTRANT ZIP CODE 3 | 1ST QUARTER | 2ND QUARTER | 3RD QUARTER | 4TH QUARTER | TOTAL GRAMS |
|---|---|---|---|---|---|
| 400 | 1,200.73 | 1,165.22 | 889.03 | 1,402.42 | **4,657.41** |
| 401 | 178.43 | 151.60 | 158.31 | 131.92 | **620.27** |
| 402 | 13,556.78 | 10,682.45 | 11,047.72 | 13,168.88 | **48,455.82** |
| 403 | 2,029.93 | 1,824.13 | 2,397.08 | 2,457.81 | **8,708.95** |
| 404 | 2,900.09 | 1,518.96 | 1,867.69 | 2,775.59 | **9,062.33** |
| 405 | 5,789.67 | 3,886.82 | 5,670.55 | 5,589.46 | **20,936.50** |
| 406 | 1,606.52 | 1,592.03 | 1,575.66 | 2,282.06 | **7,056.28** |
| 407 | 3,528.86 | 1,139.02 | 2,198.44 | 1,639.44 | **8,505.74** |
| 408 | 57.69 | 43.83 | 40.70 | 42.93 | **185.14** |
| 409 | 109.56 | 97.04 | 96.15 | 94.81 | **397.56** |
| 410 | 13,614.47 | 11,251.28 | 9,613.82 | 10,690.76 | **45,170.33** |
| 411 | 8,135.91 | 2,970.75 | 5,095.84 | 5,819.41 | **22,021.92** |
| 412 | 2,317.84 | 858.62 | 1,237.85 | 859.97 | **5,274.28** |
| 413 | 116.27 | 115.38 | 108.22 | 104.20 | **444.07** |
| 414 | 67.97 | 65.74 | 61.71 | 60.82 | **256.25** |
| 415 | 1,904.18 | 781.71 | 1,169.43 | 1,519.14 | **5,374.45** |
| 416 | 139.97 | 106.88 | 105.54 | 117.61 | **470.01** |
| 417 | 4,040.00 | 1,233.38 | 3,086.57 | 2,164.45 | **10,524.40** |
| 418 | 35.33 | 37.56 | 39.35 | 49.64 | **161.89** |
| 420 | 2,617.10 | 2,423.38 | 1,765.55 | 1,025.07 | **7,831.10** |
| 421 | 2,891.42 | 1,357.61 | 2,242.08 | 2,089.59 | **8,580.69** |
| 422 | 1,685.94 | 1,693.55 | 1,154.67 | 1,757.50 | **6,291.66** |
| 423 | 925.26 | 911.39 | 675.27 | 775.89 | **3,287.81** |
| 424 | 255.80 | 237.91 | 220.92 | 196.77 | **911.39** |
| 425 | 1,637.65 | 1,300.46 | 2,352.72 | 1,600.08 | **6,890.90** |
| 426 | 55.45 | 54.56 | 58.58 | 54.11 | **222.71** |

| | | | | | |
|---|---|---|---|---|---|
| 427 | 2,323.65 | 879.64 | 1,587.56 | 1,592.48 | **6,383.33** |
| **KY - KENTUCKY - Total** | **73,722.48** | **48,380.90** | **56,517.01** | **60,062.81** | **238,683.19** |
| **STATE: LA - LOUISIANA** | | | | | |
| REGISTRANT ZIP CODE 3 | 1ST QUARTER | 2ND QUARTER | 3RD QUARTER | 4TH QUARTER | **TOTAL GRAMS** |
| 700 | 10,630.84 | 7,727.26 | 5,759.40 | 9,257.04 | **33,374.54** |
| 701 | 6,223.77 | 4,389.72 | 6,601.39 | 4,841.03 | **22,055.90** |
| 703 | 101.51 | 110.01 | 97.04 | 125.22 | **433.78** |
| 704 | 5,398.69 | 2,718.44 | 2,673.63 | 3,750.85 | **14,541.60** |
| 705 | 3,878.92 | 2,865.03 | 3,793.78 | 4,637.02 | **15,174.75** |
| 706 | 2,481.51 | 1,732.90 | 1,431.93 | 275.92 | **5,922.27** |
| 707 | 280.39 | 294.26 | 276.37 | 286.66 | **1,137.68** |
| 708 | 4,545.34 | 4,023.28 | 3,803.79 | 4,472.89 | **16,845.31** |
| 710 | 280.84 | 274.85 | 261.61 | 292.47 | **1,109.77** |
| 711 | 4,179.76 | 2,820.10 | 3,410.51 | 3,941.62 | **14,351.99** |
| 712 | 1,529.33 | 245.69 | 1,528.35 | 854.15 | **4,157.53** |
| 713 | 5,705.38 | 2,454.68 | 3,540.93 | 4,578.88 | **16,279.87** |
| 714 | 300.07 | 302.31 | 274.58 | 249.09 | **1,126.05** |
| **LA - LOUISIANA - Total** | **45,536.37** | **29,958.52** | **33,453.31** | **37,562.83** | **146,511.04** |
| **STATE: MA - MASSACHUSETTS** | | | | | |
| REGISTRANT ZIP CODE 3 | 1ST QUARTER | 2ND QUARTER | 3RD QUARTER | 4TH QUARTER | **TOTAL GRAMS** |
| 010 | 14,112.92 | 12,335.12 | 14,586.95 | 14,169.08 | **55,204.07** |
| 011 | 10,687.65 | 8,081.53 | 8,858.24 | 9,464.71 | **37,092.13** |
| 012 | 7,029.27 | 5,311.84 | 5,121.96 | 6,808.35 | **24,271.42** |
| 013 | 1,761.61 | 1,885.84 | 2,073.67 | 2,172.05 | **7,893.17** |
| 014 | 7,871.61 | 7,629.50 | 7,684.68 | 8,659.13 | **31,844.93** |
| 015 | 5,530.34 | 3,926.86 | 4,393.56 | 6,089.97 | **19,940.74** |
| 016 | 10,056.98 | 9,076.46 | 9,469.73 | 10,202.21 | **38,805.38** |
| 017 | 3,673.57 | 3,436.73 | 3,896.19 | 3,706.13 | **14,712.61** |
| 018 | 20,077.28 | 15,974.07 | 17,859.83 | 18,388.15 | **72,299.33** |
| 019 | 18,811.48 | 15,006.06 | 13,659.72 | 17,208.70 | **64,685.96** |
| 020 | 667.04 | 451.58 | 485.12 | 679.30 | **2,283.05** |
| 021 | 31,615.40 | 25,430.04 | 25,244.98 | 25,655.59 | **107,946.02** |
| 022 | 337.64 | 316.71 | 318.32 | 291.31 | **1,263.97** |
| 023 | 13,434.34 | 7,650.52 | 7,752.03 | 9,166.62 | **38,003.50** |
| 024 | 1,730.66 | 1,145.64 | 1,310.83 | 1,637.02 | **5,824.15** |
| 025 | 3,398.90 | 3,250.52 | 3,745.66 | 3,267.69 | **13,662.76** |
| 026 | 3,986.79 | 3,254.90 | 3,087.74 | 3,169.40 | **13,498.82** |
| 027 | 25,233.89 | 18,793.76 | 20,336.33 | 20,047.98 | **84,411.95** |
| **MA - MASSACHUSETTS - Total** | **180,017.37** | **142,957.70** | **149,885.53** | **160,783.38** | **633,643.98** |
| **STATE: MD - MARYLAND** | | | | | |
| REGISTRANT ZIP CODE 3 | 1ST QUARTER | 2ND QUARTER | 3RD QUARTER | 4TH QUARTER | **TOTAL GRAMS** |
| 206 | 4,525.71 | 4,535.01 | 3,299.71 | 5,621.04 | **17,981.46** |
| 207 | 2,240.34 | 2,236.63 | 2,477.67 | 2,429.55 | **9,384.18** |
| 208 | 3,706.66 | 3,161.79 | 2,963.15 | 3,109.02 | **12,940.63** |
| 209 | 775.44 | 1,414.94 | 414.11 | 848.79 | **3,453.28** |
| 210 | 40,252.83 | 30,079.30 | 25,460.17 | 28,581.45 | **124,373.74** |
| 211 | 6,468.12 | 3,551.84 | 2,632.22 | 2,622.29 | **15,274.47** |
| 212 | 136,588.76 | 102,596.01 | 103,171.21 | 103,727.08 | **446,083.06** |
| 214 | 2,601.90 | 2,292.62 | 3,129.77 | 2,502.71 | **10,527.00** |
| 215 | 5,039.85 | 2,483.39 | 2,786.50 | 3,950.03 | **14,259.78** |
| 216 | 3,717.13 | 2,962.25 | 3,102.05 | 3,088.99 | **12,870.42** |
| 217 | 19,497.70 | 11,448.86 | 7,977.24 | 12,151.14 | **51,074.94** |
| 218 | 6,243.90 | 4,368.07 | 6,400.86 | 5,708.87 | **22,721.70** |
| 219 | 9,732.68 | 7,433.36 | 8,137.25 | 8,110.78 | **33,414.07** |
| **MD - MARYLAND - Total** | **241,391.02** | **178,564.06** | **171,951.91** | **182,451.72** | **774,358.72** |
| **STATE: ME - MAINE** | | | | | |
| REGISTRANT ZIP CODE 3 | 1ST QUARTER | 2ND QUARTER | 3RD QUARTER | 4TH QUARTER | **TOTAL GRAMS** |
| 039 | 56.79 | 54.56 | 48.30 | 39.35 | **199.00** |
| 040 | 4,542.57 | 3,155.98 | 3,476.53 | 3,478.50 | **14,653.58** |
| 041 | 8,219.98 | 5,452.08 | 7,312.44 | 7,673.06 | **28,657.56** |

| | | | | | |
|---|---|---|---|---|---|
| 042 | 3,378.42 | 2,512.55 | 3,097.13 | 3,108.31 | **12,096.40** |
| 043 | 452.75 | 392.82 | 415.72 | 432.17 | **1,693.46** |
| 044 | 13,490.68 | 6,866.76 | 6,898.95 | 6,371.80 | **33,628.19** |
| 045 | 91.68 | 109.56 | 120.30 | 99.73 | **421.26** |
| 046 | 1,192.15 | 720.08 | 1,151.54 | 1,052.71 | **4,116.48** |
| 047 | 69.76 | 76.92 | 67.53 | 81.39 | **295.60** |
| 048 | 1,664.39 | 1,443.65 | 87.20 | 85.15 | **3,280.39** |
| 049 | 5,103.36 | 4,811.07 | 5,771.92 | 4,825.47 | **20,511.81** |
| **ME - MAINE - Total** | **38,262.52** | **25,596.03** | **28,447.55** | **27,247.63** | **119,553.73** |
| **STATE: MI - MICHIGAN** | | | | | |
| REGISTRANT ZIP CODE 3 | 1ST QUARTER | 2ND QUARTER | 3RD QUARTER | 4TH QUARTER | **TOTAL GRAMS** |
| 480 | 19,748.40 | 14,378.94 | 15,571.00 | 15,568.51 | **65,266.85** |
| 481 | 12,626.74 | 7,730.61 | 8,987.25 | 11,076.58 | **40,421.18** |
| 482 | 17,077.49 | 17,024.28 | 14,845.52 | 15,903.33 | **64,850.62** |
| 483 | 12,719.08 | 8,656.81 | 10,355.72 | 9,366.16 | **41,097.77** |
| 484 | 744.59 | 780.54 | 722.05 | 722.32 | **2,969.50** |
| 485 | 11,169.27 | 7,646.67 | 9,183.97 | 9,553.35 | **37,553.26** |
| 486 | 5,143.60 | 5,100.41 | 5,078.58 | 5,053.27 | **20,375.86** |
| 487 | 326.19 | 308.12 | 307.23 | 292.02 | **1,233.56** |
| 488 | 5,643.75 | 5,656.01 | 5,245.12 | 5,634.90 | **22,179.78** |
| 489 | 8,830.14 | 7,061.08 | 7,939.05 | 9,466.69 | **33,296.96** |
| 490 | 11,868.15 | 8,468.98 | 10,551.24 | 9,720.97 | **40,609.34** |
| 491 | 288.00 | 271.00 | 241.76 | 235.32 | **1,036.07** |
| 492 | 3,970.69 | 2,866.55 | 3,876.78 | 3,890.64 | **14,604.66** |
| 493 | 530.38 | 526.80 | 504.89 | 498.90 | **2,060.97** |
| 494 | 11,008.19 | 7,206.27 | 9,217.24 | 8,050.67 | **35,482.37** |
| 495 | 9,908.70 | 7,604.73 | 9,102.13 | 8,329.19 | **34,944.74** |
| 496 | 1,546.86 | 1,540.07 | 1,511.54 | 1,460.02 | **6,058.49** |
| 497 | 3,691.01 | 4,016.57 | 3,405.07 | 4,181.59 | **15,294.24** |
| 498 | 370.28 | 328.69 | 347.92 | 338.98 | **1,385.87** |
| 499 | 67.97 | 51.88 | 64.40 | 42.93 | **227.18** |
| **MI - MICHIGAN - Total** | **137,279.49** | **107,225.01** | **117,058.44** | **119,386.32** | **480,949.26** |
| **STATE: MN - MINNESOTA** | | | | | |
| REGISTRANT ZIP CODE 3 | 1ST QUARTER | 2ND QUARTER | 3RD QUARTER | 4TH QUARTER | **TOTAL GRAMS** |
| 550 | 518.30 | 546.93 | 567.68 | 535.57 | **2,168.47** |
| 551 | 14,895.16 | 13,664.55 | 11,016.41 | 11,480.79 | **51,056.91** |
| 553 | 3,552.38 | 2,103.54 | 652.64 | 655.95 | **6,964.51** |
| 554 | 32,433.54 | 23,937.10 | 23,683.82 | 25,436.32 | **105,490.78** |
| 556 | 15.20 | 22.36 | 10.73 | 14.76 | **63.06** |
| 557 | 222.62 | 225.66 | 207.59 | 231.20 | **887.07** |
| 558 | 2,663.34 | 2,667.28 | 2,684.54 | 3,935.99 | **11,951.15** |
| 559 | 2,132.97 | 1,467.17 | 2,143.34 | 2,381.16 | **8,124.64** |
| 560 | 177.99 | 181.56 | 121.55 | 140.15 | **621.25** |
| 561 | 29.52 | 27.73 | 31.30 | 24.15 | **112.69** |
| 562 | 83.72 | 98.83 | 72.27 | 78.08 | **332.90** |
| 563 | 2,658.43 | 1,440.43 | 230.58 | 230.84 | **4,560.28** |
| 564 | 3,769.27 | 2,859.93 | 3,771.77 | 2,861.90 | **13,262.88** |
| 565 | 123.87 | 137.74 | 143.19 | 131.92 | **536.73** |
| 566 | 71.55 | 55.45 | 61.27 | 67.26 | **255.53** |
| 567 | 62.70 | 63.32 | 61.27 | 50.98 | **238.27** |
| **MN - MINNESOTA - Total** | **63,410.55** | **49,499.58** | **45,459.96** | **48,257.03** | **206,627.12** |
| **STATE: MO - MISSOURI** | | | | | |
| REGISTRANT ZIP CODE 3 | 1ST QUARTER | 2ND QUARTER | 3RD QUARTER | 4TH QUARTER | **TOTAL GRAMS** |
| 630 | 4,340.61 | 4,148.23 | 5,123.30 | 4,201.62 | **17,813.76** |
| 631 | 9,263.92 | 8,743.23 | 3,900.01 | 5,090.92 | **26,998.08** |
| 633 | 2,443.50 | 1,775.38 | 691.91 | 633.59 | **5,544.39** |
| 634 | 186.48 | 215.55 | 192.74 | 177.09 | **771.87** |
| 635 | 19.23 | 14.13 | 19.68 | 13.42 | **66.45** |
| 636 | 114.48 | 141.76 | 169.94 | 159.65 | **585.83** |
| 637 | 1,935.03 | 559.18 | 89.98 | 98.92 | **2,683.11** |

| | | | | | |
|---|---|---|---|---|---|
| 638 | 97.49 | 82.73 | 91.68 | 113.86 | **385.75** |
| 639 | 1,345.89 | 1,264.77 | 922.84 | 1,344.28 | **4,877.79** |
| 640 | 704.79 | 692.27 | 650.85 | 631.00 | **2,678.91** |
| 641 | 5,265.53 | 4,507.58 | 3,498.61 | 5,467.12 | **18,738.84** |
| 644 | 37.12 | 31.75 | 32.20 | 45.61 | **146.68** |
| 645 | 2,064.28 | 1,524.06 | 73.79 | 55.01 | **3,717.13** |
| 646 | 42.48 | 39.35 | 33.99 | 26.38 | **142.21** |
| 647 | 130.58 | 135.95 | 151.15 | 136.84 | **554.53** |
| 648 | 2,430.08 | 1,096.09 | 1,560.28 | 1,529.87 | **6,616.32** |
| 650 | 81.84 | 75.58 | 81.39 | 72.45 | **311.25** |
| 651 | 77.81 | 76.92 | 79.15 | 66.19 | **300.07** |
| 652 | 1,278.54 | 978.92 | 1,354.93 | 808.72 | **4,421.11** |
| 653 | 72.45 | 49.91 | 77.72 | 54.20 | **254.28** |
| 654 | 85.42 | 101.07 | 92.57 | 94.81 | **373.86** |
| 655 | 128.35 | 122.98 | 115.82 | 127.90 | **495.05** |
| 656 | 242.47 | 233.08 | 252.31 | 240.06 | **967.92** |
| 657 | 1,392.85 | 755.77 | 860.68 | 1,128.73 | **4,138.03** |
| 658 | 4,235.52 | 3,510.07 | 3,309.28 | 3,514.63 | **14,569.51** |
| **MO - MISSOURI - Total** | **38,016.74** | **30,876.31** | **23,426.80** | **25,832.87** | **118,152.73** |

**STATE: MS - MISSISSIPPI**

| REGISTRANT ZIP CODE 3 | 1ST QUARTER | 2ND QUARTER | 3RD QUARTER | 4TH QUARTER | TOTAL GRAMS |
|---|---|---|---|---|---|
| 386 | 2,233.76 | 2,169.81 | 2,282.96 | 2,062.93 | **8,749.47** |
| 387 | 122.98 | 60.37 | 64.40 | 70.66 | **318.41** |
| 388 | 77.63 | 81.12 | 64.31 | 58.58 | **281.65** |
| 389 | 37.12 | 51.88 | 38.46 | 49.19 | **176.64** |
| 390 | 148.47 | 144.89 | 132.82 | 122.98 | **549.16** |
| 391 | 116.72 | 174.86 | 158.22 | 135.95 | **585.74** |
| 392 | 2,090.66 | 1,477.73 | 1,697.79 | 1,987.54 | **7,253.72** |
| 393 | 923.47 | 915.42 | 56.79 | 55.01 | **1,950.69** |
| 394 | 179.77 | 161.44 | 154.28 | 165.02 | **660.51** |
| 395 | 3,288.26 | 2,140.57 | 205.71 | 220.20 | **5,854.74** |
| 396 | 93.46 | 80.05 | 79.60 | 76.92 | **330.03** |
| 397 | 67.97 | 92.75 | 93.46 | 81.39 | **335.58** |
| **MS - MISSISSIPPI - Total** | **9,380.29** | **7,550.88** | **5,028.81** | **5,086.36** | **27,046.34** |

**STATE: MT - MONTANA**

| REGISTRANT ZIP CODE 3 | 1ST QUARTER | 2ND QUARTER | 3RD QUARTER | 4TH QUARTER | TOTAL GRAMS |
|---|---|---|---|---|---|
| 590 | 52.32 | 57.24 | 42.48 | 48.30 | **200.35** |
| 591 | 2,385.81 | 1,603.66 | 2,365.69 | 1,596.50 | **7,951.66** |
| 592 | 19.68 | 17.44 | 14.76 | 16.99 | **68.87** |
| 593 | 18.78 | 18.78 | 16.10 | 16.10 | **69.76** |
| 594 | 92.12 | 86.58 | 108.31 | 112.52 | **399.53** |
| 595 | 16.10 | 13.86 | 16.55 | 15.20 | **61.71** |
| 596 | 106.88 | 110.01 | 72.00 | 94.81 | **383.70** |
| 597 | 1,724.13 | 1,696.23 | 1,978.14 | 1,557.33 | **6,955.84** |
| 598 | 2,506.47 | 2,478.20 | 2,314.08 | 2,482.14 | **9,780.89** |
| 599 | 2,767.81 | 1,870.46 | 2,770.94 | 2,303.97 | **9,713.18** |
| **MT - MONTANA - Total** | **9,690.11** | **7,952.47** | **9,699.05** | **8,243.86** | **35,585.49** |

**STATE: NC - NORTH CAROLINA**

| REGISTRANT ZIP CODE 3 | 1ST QUARTER | 2ND QUARTER | 3RD QUARTER | 4TH QUARTER | TOTAL GRAMS |
|---|---|---|---|---|---|
| 270 | 368.94 | 392.64 | 362.95 | 357.31 | **1,481.84** |
| 271 | 5,733.73 | 3,228.52 | 5,017.76 | 3,942.60 | **17,922.61** |
| 272 | 7,021.89 | 6,337.81 | 1,830.89 | 1,783.98 | **16,974.56** |
| 273 | 6,296.58 | 6,193.09 | 2,582.58 | 3,677.33 | **18,749.58** |
| 274 | 7,437.47 | 5,331.97 | 5,242.26 | 5,256.84 | **23,268.53** |
| 275 | 8,738.99 | 9,341.40 | 7,294.09 | 8,082.09 | **33,456.56** |
| 276 | 7,667.33 | 6,367.23 | 5,695.36 | 5,580.97 | **25,310.89** |
| 277 | 6,974.62 | 6,408.55 | 3,610.25 | 4,423.43 | **21,416.86** |
| 278 | 7,239.63 | 6,547.63 | 6,255.08 | 5,422.57 | **25,464.91** |
| 279 | 1,616.63 | 2,059.09 | 1,919.74 | 2,676.31 | **8,271.77** |
| 280 | 8,982.73 | 6,138.54 | 4,226.22 | 1,484.44 | **20,831.92** |

| | | | | | |
|---|---|---|---|---|---|
| 281 | 9,140.05 | 5,893.20 | 2,672.20 | 2,615.32 | **20,320.77** |
| 282 | 6,889.38 | 5,908.05 | 2,854.66 | 1,165.49 | **16,817.58** |
| 283 | 14,713.86 | 13,209.75 | 12,550.67 | 10,990.30 | **51,464.58** |
| 284 | 7,956.43 | 7,488.59 | 3,872.10 | 4,936.35 | **24,253.48** |
| 285 | 6,289.60 | 4,842.55 | 6,392.01 | 5,700.37 | **23,224.53** |
| 286 | 13,967.40 | 12,379.03 | 8,206.84 | 6,501.30 | **41,054.57** |
| 287 | 11,209.52 | 8,679.88 | 10,572.34 | 7,964.72 | **38,426.46** |
| 288 | 15,693.86 | 13,753.99 | 15,451.92 | 14,895.52 | **59,795.29** |
| 289 | 1,056.73 | 1,047.70 | 879.64 | 651.75 | **3,635.83** |
| **NC - NORTH CAROLINA - Total** | **154,995.37** | **131,549.22** | **107,489.54** | **98,108.99** | **492,143.12** |

**STATE: ND - NORTH DAKOTA**

| REGISTRANT ZIP CODE 3 | 1ST QUARTER | 2ND QUARTER | 3RD QUARTER | 4TH QUARTER | TOTAL GRAMS |
|---|---|---|---|---|---|
| 580 | 52.77 | 43.29 | 45.70 | 48.74 | **190.51** |
| 581 | 2,198.35 | 2,882.20 | 2,162.21 | 2,870.22 | **10,112.98** |
| 582 | 154.37 | 130.14 | 96.42 | 120.12 | **501.04** |
| 583 | 18.78 | 20.12 | 22.36 | 20.57 | **81.84** |
| 584 | 21.91 | 25.49 | 26.38 | 21.47 | **95.25** |
| 585 | 1,849.53 | 991.44 | 1,844.97 | 1,426.03 | **6,111.97** |
| 586 | 20.57 | 25.49 | 23.70 | 22.81 | **92.57** |
| 587 | 1,473.08 | 1,452.95 | 1,820.55 | 1,461.00 | **6,207.58** |
| 588 | 46.33 | 40.52 | 45.17 | 36.67 | **168.68** |
| **ND - NORTH DAKOTA - Total** | **5,835.69** | **5,611.64** | **6,087.47** | **6,027.63** | **23,562.43** |

**STATE: NE - NEBRASKA**

| REGISTRANT ZIP CODE 3 | 1ST QUARTER | 2ND QUARTER | 3RD QUARTER | 4TH QUARTER | TOTAL GRAMS |
|---|---|---|---|---|---|
| 680 | 242.29 | 225.66 | 203.48 | 204.91 | **876.33** |
| 681 | 6,140.09 | 3,935.57 | 2,873.02 | 2,833.31 | **15,781.99** |
| 683 | 51.07 | 33.72 | 33.90 | 26.21 | **144.89** |
| 684 | 30.86 | 34.43 | 32.65 | 24.60 | **122.53** |
| 685 | 1,593.19 | 1,268.62 | 1,697.57 | 1,410.47 | **5,969.85** |
| 686 | 33.99 | 38.91 | 36.22 | 39.80 | **148.92** |
| 687 | 118.51 | 99.28 | 95.70 | 108.67 | **422.16** |
| 688 | 114.93 | 106.88 | 120.30 | 114.04 | **456.14** |
| 689 | 39.80 | 37.56 | 32.65 | 37.03 | **147.04** |
| 690 | 6.26 | 7.16 | 6.26 | 6.71 | **26.38** |
| 691 | 74.95 | 65.83 | 67.97 | 66.90 | **275.65** |
| 692 | 4.47 | 8.05 | 8.94 | 6.26 | **27.73** |
| 693 | 80.85 | 81.03 | 72.00 | 64.84 | **298.73** |
| **NE - NEBRASKA - Total** | **8,531.27** | **5,942.70** | **5,280.65** | **4,943.73** | **24,698.36** |

**STATE: NH - NEW HAMPSHIRE**

| REGISTRANT ZIP CODE 3 | 1ST QUARTER | 2ND QUARTER | 3RD QUARTER | 4TH QUARTER | TOTAL GRAMS |
|---|---|---|---|---|---|
| 030 | 3,020.75 | 3,043.29 | 3,596.65 | 3,020.84 | **12,681.52** |
| 031 | 14,340.00 | 9,644.40 | 3,874.45 | 4,310.02 | **32,168.88** |
| 032 | 3,451.49 | 2,062.75 | 445.68 | 398.01 | **6,357.93** |
| 033 | 5,007.92 | 5,639.73 | 289.43 | 264.47 | **11,201.56** |
| 034 | 3,277.53 | 2,370.16 | 98.38 | 88.55 | **5,834.62** |
| 035 | 143.10 | 124.77 | 124.32 | 129.24 | **521.44** |
| 036 | 24.15 | 28.17 | 26.38 | 23.25 | **101.96** |
| 037 | 3,415.51 | 2,773.48 | 3,416.72 | 3,162.41 | **12,768.11** |
| 038 | 6,397.64 | 4,809.55 | 5,266.58 | 5,275.35 | **21,749.12** |
| **NH - NEW HAMPSHIRE - Total** | **39,078.10** | **30,496.30** | **17,138.60** | **16,672.14** | **103,385.14** |

**STATE: NJ - NEW JERSEY**

| REGISTRANT ZIP CODE 3 | 1ST QUARTER | 2ND QUARTER | 3RD QUARTER | 4TH QUARTER | TOTAL GRAMS |
|---|---|---|---|---|---|
| 070 | 10,049.21 | 8,514.51 | 8,491.17 | 6,028.35 | **33,083.23** |
| 071 | 17,868.14 | 11,570.67 | 10,583.61 | 13,472.88 | **53,495.32** |
| 072 | 980.71 | 1,858.12 | 971.77 | 968.19 | **4,778.78** |
| 073 | 7,241.51 | 5,064.63 | 7,031.95 | 6,745.03 | **26,083.12** |
| 074 | 542.01 | 482.98 | 499.61 | 515.08 | **2,039.68** |
| 075 | 3,889.03 | 3,240.50 | 3,527.51 | 3,670.17 | **14,327.21** |
| 076 | 2,627.57 | 1,809.46 | 2,305.23 | 1,821.98 | **8,564.24** |
| 077 | 17,706.79 | 15,161.33 | 13,164.41 | 11,039.49 | **57,072.02** |

| | | | | | |
|---|---|---|---|---|---|
| 078 | 3,887.96 | 5,626.94 | 4,731.82 | 3,653.09 | **17,899.81** |
| 079 | 227.09 | 170.83 | 192.30 | 232.90 | **823.12** |
| 080 | 5,593.22 | 5,309.96 | 6,035.23 | 7,763.75 | **24,702.17** |
| 081 | 28,213.04 | 23,653.66 | 27,169.28 | 24,510.58 | **103,546.57** |
| 082 | 11,576.76 | 7,808.20 | 8,947.04 | 8,532.93 | **36,864.93** |
| 083 | 11,616.47 | 9,416.24 | 11,401.36 | 11,711.27 | **44,145.35** |
| 084 | 166.36 | 137.29 | 128.35 | 113.59 | **545.58** |
| 085 | 376.10 | 346.94 | 321.27 | 355.88 | **1,400.18** |
| 086 | 7,817.95 | 5,516.21 | 4,258.15 | 3,896.90 | **21,489.21** |
| 087 | 14,476.31 | 12,481.80 | 12,556.03 | 14,014.44 | **53,528.59** |
| 088 | 4,795.42 | 5,082.52 | 7,283.99 | 5,394.75 | **22,556.68** |
| 089 | 5,101.30 | 3,283.79 | 4,050.74 | 3,680.28 | **16,116.10** |
| **NJ - NEW JERSEY - Total** | **154,752.93** | **126,536.58** | **133,650.82** | **128,121.55** | **543,061.88** |

**STATE: NM - NEW MEXICO**

| REGISTRANT ZIP CODE 3 | 1ST QUARTER | 2ND QUARTER | 3RD QUARTER | 4TH QUARTER | TOTAL GRAMS |
|---|---|---|---|---|---|
| 870 | 2,084.22 | 1,938.16 | 1,513.15 | 2,225.27 | **7,760.80** |
| 871 | 44,898.34 | 29,816.17 | 30,038.69 | 26,006.83 | **130,760.03** |
| 873 | 25.94 | 34.88 | 16.10 | 27.73 | **104.64** |
| 874 | 1,467.08 | 799.59 | 1,527.90 | 1,532.11 | **5,326.69** |
| 875 | 8,793.03 | 8,167.93 | 7,345.98 | 8,766.28 | **33,073.21** |
| 877 | 25.94 | 21.47 | 12.97 | 8.50 | **68.87** |
| 878 | 14.31 | 9.84 | 18.78 | 19.23 | **62.16** |
| 879 | 1.79 | 2.68 | 0.89 | 1.79 | **7.16** |
| 880 | 2,322.31 | 1,899.80 | 1,935.03 | 1,916.97 | **8,074.11** |
| 881 | 29.96 | 28.62 | 30.86 | 40.25 | **129.69** |
| 882 | 1,283.02 | 1,473.43 | 1,476.21 | 1,483.81 | **5,716.47** |
| 883 | 124.77 | 127.00 | 76.92 | 93.02 | **421.71** |
| 884 | 0.89 | 2.68 | 2.24 | 4.47 | **10.29** |
| **NM - NEW MEXICO - Total** | **61,071.60** | **44,322.26** | **43,995.71** | **42,126.24** | **191,515.81** |

**STATE: NV - NEVADA**

| REGISTRANT ZIP CODE 3 | 1ST QUARTER | 2ND QUARTER | 3RD QUARTER | 4TH QUARTER | TOTAL GRAMS |
|---|---|---|---|---|---|
| 890 | 11,982.90 | 10,646.76 | 9,761.21 | 10,373.43 | **42,764.30** |
| 891 | 17,139.21 | 13,424.59 | 13,199.47 | 15,084.59 | **58,847.85** |
| 893 | 23.25 | 16.99 | 24.15 | 18.78 | **83.18** |
| 894 | 4,267.27 | 3,240.50 | 3,743.15 | 3,132.55 | **14,383.47** |
| 895 | 4,453.66 | 3,911.93 | 4,719.66 | 3,930.80 | **17,016.05** |
| 897 | 1,370.49 | 1,225.33 | 1,383.19 | 1,386.77 | **5,365.77** |
| 898 | 41.59 | 34.88 | 45.52 | 44.63 | **166.63** |
| **NV - NEVADA - Total** | **39,278.38** | **32,500.98** | **32,876.36** | **33,971.55** | **138,627.26** |

**STATE: NY - NEW YORK**

| REGISTRANT ZIP CODE 3 | 1ST QUARTER | 2ND QUARTER | 3RD QUARTER | 4TH QUARTER | TOTAL GRAMS |
|---|---|---|---|---|---|
| 100 | 97,206.86 | 70,876.00 | 83,527.52 | 77,621.16 | **329,231.55** |
| 101 | 1,340.88 | 1,548.56 | 1,111.29 | 1,409.31 | **5,410.05** |
| 102 | 12.97 | 8.94 | 18.78 | 12.88 | **53.57** |
| 103 | 12,520.97 | 11,099.95 | 8,331.07 | 9,605.59 | **41,557.58** |
| 104 | 76,188.75 | 67,152.98 | 58,229.20 | 65,096.49 | **266,667.42** |
| 105 | 6,605.05 | 7,515.82 | 7,142.95 | 7,002.88 | **28,266.71** |
| 106 | 144.54 | 98.12 | 112.43 | 105.81 | **460.88** |
| 107 | 7,038.48 | 3,746.46 | 4,792.46 | 6,609.88 | **22,187.29** |
| 108 | 1,719.93 | 1,123.81 | 2,460.05 | 890.82 | **6,194.61** |
| 109 | 4,227.83 | 3,488.70 | 3,513.47 | 2,646.17 | **13,876.17** |
| 110 | 3,220.29 | 3,565.55 | 3,115.10 | 3,604.67 | **13,505.61** |
| 111 | 151.60 | 123.43 | 120.74 | 117.79 | **513.56** |
| 112 | 40,394.59 | 36,407.25 | 36,163.44 | 40,466.14 | **153,431.42** |
| 113 | 5,322.13 | 3,921.41 | 4,046.44 | 5,912.79 | **19,202.77** |
| 114 | 6,739.21 | 5,957.60 | 6,077.90 | 7,123.54 | **25,898.25** |
| 115 | 6,217.42 | 2,570.86 | 4,632.63 | 4,350.81 | **17,771.73** |
| 116 | 78.26 | 75.13 | 74.32 | 55.99 | **283.70** |
| 117 | 11,758.23 | 10,458.85 | 9,926.41 | 9,239.06 | **41,382.55** |
| 118 | 126.56 | 103.66 | 110.01 | 87.56 | **427.79** |

| | | | | | |
|---|---|---|---|---|---|
| 119 | 2,404.59 | 2,412.38 | 4,528.70 | 5,250.13 | **14,595.80** |
| 120 | 1,858.65 | 1,696.05 | 1,136.34 | 2,095.13 | **6,786.17** |
| 121 | 2,007.30 | 1,616.81 | 2,047.64 | 2,607.80 | **8,279.55** |
| 122 | 8,494.65 | 6,841.44 | 7,785.66 | 7,068.53 | **30,190.29** |
| 123 | 1,416.64 | 972.12 | 1,328.45 | 1,824.58 | **5,541.79** |
| 124 | 3,266.62 | 2,809.78 | 2,915.57 | 2,690.80 | **11,682.76** |
| 125 | 6,001.51 | 7,270.94 | 5,127.15 | 5,712.18 | **24,111.77** |
| 126 | 4,693.81 | 4,405.81 | 4,595.43 | 5,099.87 | **18,794.92** |
| 127 | 171.64 | 166.81 | 169.04 | 173.78 | **681.26** |
| 128 | 551.40 | 534.67 | 495.59 | 509.09 | **2,090.75** |
| 129 | 1,769.93 | 769.81 | 1,224.08 | 1,432.83 | **5,196.64** |
| 130 | 2,520.60 | 2,086.64 | 2,603.42 | 2,392.70 | **9,603.35** |
| 131 | 1,999.43 | 1,392.13 | 1,588.90 | 1,548.21 | **6,528.67** |
| 132 | 12,687.60 | 10,530.31 | 9,550.31 | 11,672.19 | **44,440.41** |
| 133 | 128.79 | 139.88 | 134.61 | 128.26 | **531.54** |
| 134 | 2,313.28 | 2,280.90 | 2,281.88 | 2,284.12 | **9,160.18** |
| 135 | 1,187.05 | 1,198.41 | 2,118.74 | 2,770.58 | **7,274.78** |
| 136 | 1,559.83 | 1,419.86 | 1,557.60 | 1,600.17 | **6,137.46** |
| 137 | 181.56 | 146.68 | 154.28 | 158.76 | **641.28** |
| 138 | 349.71 | 334.06 | 302.75 | 300.70 | **1,287.22** |
| 139 | 2,581.42 | 1,929.04 | 2,413.18 | 1,988.70 | **8,912.34** |
| 140 | 2,480.17 | 1,954.00 | 2,296.91 | 1,593.37 | **8,324.45** |
| 141 | 1,948.00 | 888.32 | 1,688.27 | 2,476.24 | **7,000.83** |
| 142 | 21,125.00 | 18,479.04 | 18,876.29 | 17,478.40 | **75,958.73** |
| 143 | 2,102.02 | 2,064.10 | 2,070.18 | 2,074.47 | **8,310.76** |
| 144 | 601.39 | 527.07 | 514.91 | 471.80 | **2,115.17** |
| 145 | 253.12 | 284.42 | 272.79 | 270.56 | **1,080.88** |
| 146 | 8,698.14 | 5,830.05 | 6,020.62 | 8,031.67 | **28,580.48** |
| 147 | 295.51 | 263.58 | 239.52 | 230.84 | **1,029.45** |
| 148 | 1,025.70 | 630.55 | 1,223.09 | 1,125.24 | **4,004.59** |
| 149 | 107.78 | 113.59 | 109.56 | 112.25 | **443.18** |
| **NY - NEW YORK - Total** | **377,797.40** | **311,832.33** | **320,877.68** | **335,133.28** | **1,345,640.69** |

**STATE: OH - OHIO**

| REGISTRANT ZIP CODE 3 | 1ST QUARTER | 2ND QUARTER | 3RD QUARTER | 4TH QUARTER | **TOTAL GRAMS** |
|---|---|---|---|---|---|
| 430 | 980.26 | 965.33 | 892.52 | 951.10 | **3,789.22** |
| 431 | 604.88 | 668.92 | 556.50 | 741.90 | **2,572.20** |
| 432 | 17,348.63 | 15,455.36 | 16,387.06 | 19,153.84 | **68,344.89** |
| 433 | 456.77 | 428.69 | 644.15 | 620.45 | **2,150.05** |
| 434 | 102.41 | 88.99 | 97.94 | 91.14 | **380.48** |
| 435 | 184.34 | 211.79 | 198.02 | 227.54 | **821.69** |
| 436 | 7,837.27 | 9,002.67 | 9,443.16 | 10,311.90 | **36,595.00** |
| 437 | 1,429.16 | 1,535.15 | 1,221.57 | 1,291.19 | **5,477.07** |
| 438 | 71.55 | 58.58 | 59.92 | 56.88 | **246.94** |
| 439 | 218.41 | 158.31 | 198.65 | 212.51 | **787.88** |
| 440 | 1,546.51 | 1,264.14 | 2,363.54 | 3,147.21 | **8,321.41** |
| 441 | 9,318.84 | 6,734.12 | 7,248.40 | 8,048.71 | **31,350.06** |
| 442 | 2,677.57 | 3,568.48 | 3,023.07 | 3,116.27 | **12,385.38** |
| 443 | 4,099.75 | 3,842.97 | 4,071.22 | 4,006.64 | **16,020.58** |
| 444 | 1,521.37 | 1,292.41 | 1,190.71 | 1,142.86 | **5,147.36** |
| 445 | 7,254.59 | 7,406.71 | 7,716.34 | 7,626.91 | **30,004.54** |
| 446 | 597.46 | 478.41 | 555.42 | 763.64 | **2,394.93** |
| 447 | 1,730.13 | 1,527.81 | 1,429.79 | 1,528.62 | **6,216.35** |
| 448 | 290.50 | 265.91 | 266.17 | 231.83 | **1,054.41** |
| 449 | 300.07 | 377.44 | 311.25 | 663.20 | **1,651.96** |
| 450 | 2,571.49 | 2,685.61 | 2,528.20 | 2,918.96 | **10,704.27** |
| 451 | 2,142.98 | 1,871.08 | 2,786.06 | 2,947.94 | **9,748.07** |
| 452 | 15,989.12 | 13,658.29 | 17,060.32 | 15,940.98 | **62,648.72** |
| 453 | 591.74 | 542.54 | 518.57 | 544.60 | **2,197.45** |
| 454 | 6,195.33 | 6,159.20 | 6,060.08 | 6,196.85 | **24,611.46** |
| 455 | 139.35 | 123.87 | 104.47 | 102.68 | **470.36** |

| | | | | | |
|---|---|---|---|---|---|
| 456 | 6,048.20 | 4,360.56 | 6,231.28 | 6,040.24 | **22,680.28** |
| 457 | 413.93 | 250.52 | 361.34 | 370.10 | **1,395.89** |
| 458 | 1,454.38 | 945.02 | 1,508.58 | 941.00 | **4,848.99** |
| **OH - OHIO - Total** | **94,116.99** | **85,928.89** | **95,034.31** | **99,937.70** | **375,017.89** |

**STATE: OK - OKLAHOMA**

| REGISTRANT ZIP CODE 3 | 1ST QUARTER | 2ND QUARTER | 3RD QUARTER | 4TH QUARTER | TOTAL GRAMS |
|---|---|---|---|---|---|
| 730 | 540.13 | 495.77 | 437.81 | 492.37 | **1,966.07** |
| 731 | 13,358.76 | 11,114.62 | 11,506.55 | 11,514.60 | **47,494.52** |
| 734 | 2,171.60 | 1,375.94 | 913.81 | 926.33 | **5,387.69** |
| 735 | 1,353.67 | 713.82 | 807.64 | 989.12 | **3,864.26** |
| 736 | 16.55 | 17.44 | 27.28 | 24.60 | **85.86** |
| 737 | 45.61 | 56.53 | 61.71 | 60.82 | **224.67** |
| 738 | 40.25 | 36.22 | 38.91 | 34.43 | **149.81** |
| 739 | 13.86 | 15.20 | 19.23 | 7.60 | **55.90** |
| 740 | 2,774.88 | 1,820.10 | 1,894.34 | 2,379.10 | **8,868.42** |
| 741 | 13,819.02 | 8,898.03 | 10,149.20 | 12,696.54 | **45,562.79** |
| 743 | 1,698.02 | 1,156.01 | 628.76 | 1,151.09 | **4,633.89** |
| 744 | 234.78 | 210.63 | 219.84 | 218.23 | **883.49** |
| 745 | 747.72 | 537.09 | 882.42 | 724.02 | **2,891.24** |
| 746 | 1,266.02 | 1,039.74 | 929.73 | 1,382.74 | **4,618.23** |
| 747 | 83.45 | 88.28 | 76.83 | 69.14 | **317.69** |
| 748 | 105.09 | 120.30 | 120.74 | 133.27 | **479.40** |
| 749 | 4,863.75 | 3,328.51 | 4,223.36 | 3,942.52 | **16,358.13** |
| **OK - OKLAHOMA - Total** | **43,133.16** | **31,024.23** | **32,938.16** | **36,746.51** | **143,842.06** |

**STATE: OR - OREGON**

| REGISTRANT ZIP CODE 3 | 1ST QUARTER | 2ND QUARTER | 3RD QUARTER | 4TH QUARTER | TOTAL GRAMS |
|---|---|---|---|---|---|
| 970 | 1,469.59 | 1,390.08 | 1,405.01 | 1,372.19 | **5,636.87** |
| 971 | 3,051.07 | 796.02 | 1,609.65 | 827.95 | **6,284.68** |
| 972 | 36,127.75 | 32,721.61 | 29,115.83 | 28,771.49 | **126,736.69** |
| 973 | 11,072.14 | 9,852.53 | 9,906.91 | 10,034.72 | **40,866.30** |
| 974 | 10,271.65 | 7,398.66 | 8,613.88 | 8,709.13 | **34,993.31** |
| 975 | 6,512.31 | 5,404.86 | 7,188.83 | 5,802.06 | **24,908.06** |
| 976 | 161.35 | 151.60 | 167.43 | 149.81 | **630.19** |
| 977 | 1,088.57 | 1,067.20 | 1,251.89 | 1,260.75 | **4,668.41** |
| 978 | 817.03 | 850.57 | 1,500.00 | 756.66 | **3,924.27** |
| 979 | 61.53 | 71.82 | 62.43 | 71.10 | **266.89** |
| **OR - OREGON - Total** | **70,632.99** | **59,704.95** | **60,821.87** | **57,755.87** | **248,915.66** |

**STATE: PA - PENNSYLVANIA**

| REGISTRANT ZIP CODE 3 | 1ST QUARTER | 2ND QUARTER | 3RD QUARTER | 4TH QUARTER | TOTAL GRAMS |
|---|---|---|---|---|---|
| 150 | 6,383.96 | 4,341.15 | 4,716.17 | 5,391.17 | **20,832.45** |
| 151 | 384.32 | 349.35 | 343.09 | 350.16 | **1,426.93** |
| 152 | 28,642.09 | 23,127.31 | 22,790.47 | 26,778.87 | **101,338.74** |
| 153 | 6,508.73 | 3,662.03 | 4,759.46 | 5,094.23 | **20,024.45** |
| 154 | 3,969.35 | 3,180.93 | 4,762.23 | 3,970.15 | **15,882.67** |
| 155 | 77.37 | 58.14 | 32.20 | 35.60 | **203.30** |
| 156 | 8,114.62 | 6,913.80 | 7,169.78 | 8,147.09 | **30,345.29** |
| 157 | 1,029.10 | 1,181.06 | 221.10 | 1,070.06 | **3,501.31** |
| 158 | 72.09 | 63.50 | 58.58 | 63.14 | **257.32** |
| 159 | 3,708.81 | 2,986.76 | 3,717.22 | 2,999.55 | **13,412.33** |
| 160 | 4,611.53 | 3,861.12 | 4,189.64 | 4,235.07 | **16,897.36** |
| 161 | 7,908.91 | 5,893.20 | 5,471.13 | 8,926.65 | **28,199.90** |
| 162 | 76.47 | 66.63 | 70.66 | 70.21 | **283.97** |
| 163 | 2,047.28 | 1,453.94 | 1,447.41 | 1,436.85 | **6,385.48** |
| 164 | 94.18 | 86.76 | 138.90 | 81.12 | **400.96** |
| 165 | 4,085.62 | 3,654.43 | 4,081.50 | 4,069.97 | **15,891.52** |
| 166 | 6,354.71 | 4,227.20 | 5,016.60 | 5,282.15 | **20,880.66** |
| 167 | 114.93 | 126.11 | 90.33 | 88.55 | **419.92** |
| 168 | 2,679.62 | 2,088.87 | 2,774.88 | 2,013.29 | **9,556.66** |
| 169 | 56.62 | 56.35 | 58.58 | 48.30 | **219.84** |
| 170 | 10,070.41 | 4,578.70 | 6,888.40 | 6,545.13 | **28,082.64** |

| | | | | | |
|---|---|---|---|---|---|
| 171 | 6,073.87 | 3,703.26 | 5,750.58 | 6,695.07 | **22,222.78** |
| 172 | 305.62 | 301.86 | 290.50 | 296.76 | **1,194.74** |
| 173 | 2,875.94 | 1,707.86 | 2,240.47 | 1,646.59 | **8,470.86** |
| 174 | 4,792.20 | 3,874.45 | 3,883.84 | 3,884.38 | **16,434.87** |
| 175 | 259.29 | 313.93 | 537.09 | 1,767.51 | **2,877.82** |
| 176 | 5,657.62 | 6,039.61 | 6,035.50 | 5,580.34 | **23,313.07** |
| 177 | 4,932.62 | 3,897.88 | 3,364.29 | 4,603.92 | **16,798.71** |
| 178 | 818.11 | 900.48 | 849.14 | 869.27 | **3,437.00** |
| 179 | 125.39 | 111.80 | 97.04 | 101.07 | **435.30** |
| 180 | 6,234.95 | 5,553.78 | 5,405.40 | 6,201.41 | **23,395.54** |
| 181 | 384.14 | 307.14 | 314.20 | 298.64 | **1,304.12** |
| 182 | 115.38 | 138.63 | 122.98 | 93.55 | **470.54** |
| 183 | 3,661.32 | 2,525.88 | 2,519.70 | 2,511.48 | **11,218.37** |
| 184 | 196.95 | 167.07 | 166.72 | 173.07 | **703.80** |
| 185 | 3,975.34 | 2,744.65 | 2,729.26 | 3,515.71 | **12,964.95** |
| 186 | 249.09 | 235.50 | 252.67 | 240.77 | **978.03** |
| 187 | 6,271.80 | 5,153.35 | 5,715.31 | 5,766.38 | **22,906.84** |
| 188 | 100.62 | 90.16 | 142.75 | 137.02 | **470.54** |
| 189 | 7,729.94 | 6,204.63 | 6,554.07 | 6,264.38 | **26,753.02** |
| 190 | 14,983.44 | 14,231.42 | 14,666.37 | 15,106.51 | **58,987.74** |
| 191 | 50,583.96 | 37,243.16 | 37,308.41 | 39,177.31 | **164,312.84** |
| 193 | 4,426.39 | 3,847.71 | 4,037.50 | 4,527.54 | **16,839.14** |
| 194 | 7,291.77 | 4,940.13 | 4,805.61 | 6,044.71 | **23,082.23** |
| 195 | 133.27 | 161.44 | 127.45 | 114.93 | **537.09** |
| 196 | 5,249.02 | 3,250.50 | 3,551.63 | 6,260.64 | **18,311.78** |
| **PA - PENNSYLVANIA - Total** | **234,418.73** | **179,603.62** | **190,266.82** | **208,576.28** | **812,865.45** |

**STATE: PR - PUERTO RICO**

| REGISTRANT ZIP CODE 3 | 1ST QUARTER | 2ND QUARTER | 3RD QUARTER | 4TH QUARTER | TOTAL GRAMS |
|---|---|---|---|---|---|
| 006 | 1,829.94 | 1,831.73 | 1,220.86 | 1,719.93 | **6,602.46** |
| 007 | 5,372.66 | 5,373.56 | 3,577.60 | 5,368.19 | **19,692.00** |
| 009 | 20,007.73 | 15,481.17 | 11,061.94 | 16,346.50 | **62,897.34** |
| **PR - PUERTO RICO - Total** | **27,210.33** | **22,686.46** | **15,860.40** | **23,434.62** | **89,191.80** |

**STATE: RI - RHODE ISLAND**

| REGISTRANT ZIP CODE 3 | 1ST QUARTER | 2ND QUARTER | 3RD QUARTER | 4TH QUARTER | TOTAL GRAMS |
|---|---|---|---|---|---|
| 028 | 27,040.93 | 20,417.90 | 23,211.56 | 21,343.34 | **92,013.73** |
| 029 | 34,653.53 | 12,803.60 | 20,540.25 | 18,706.38 | **86,703.76** |
| **RI - RHODE ISLAND - Total** | **61,694.46** | **33,221.50** | **43,751.81** | **40,049.71** | **178,717.49** |

**STATE: SC - SOUTH CAROLINA**

| REGISTRANT ZIP CODE 3 | 1ST QUARTER | 2ND QUARTER | 3RD QUARTER | 4TH QUARTER | TOTAL GRAMS |
|---|---|---|---|---|---|
| 290 | 602.29 | 569.29 | 555.96 | 565.26 | **2,292.79** |
| 291 | 5,237.16 | 4,442.04 | 4,289.36 | 2,470.78 | **16,439.34** |
| 292 | 2,322.58 | 2,333.04 | 2,043.70 | 2,017.50 | **8,716.82** |
| 293 | 6,780.27 | 5,773.08 | 5,861.09 | 6,344.69 | **24,759.14** |
| 294 | 9,748.51 | 8,775.19 | 6,156.80 | 6,451.57 | **31,132.07** |
| 295 | 15,414.89 | 14,275.88 | 16,292.21 | 14,354.58 | **60,337.57** |
| 296 | 12,741.44 | 12,738.13 | 14,543.12 | 12,159.28 | **52,181.98** |
| 297 | 6,229.50 | 3,877.22 | 2,892.04 | 2,528.47 | **15,527.23** |
| 298 | 1,842.02 | 926.33 | 2,553.15 | 1,469.95 | **6,791.45** |
| 299 | 1,316.11 | 1,260.30 | 1,577.81 | 1,301.80 | **5,456.02** |
| **SC - SOUTH CAROLINA - Total** | **62,234.77** | **54,970.50** | **56,765.26** | **49,663.88** | **223,634.41** |

**STATE: SD - SOUTH DAKOTA**

| REGISTRANT ZIP CODE 3 | 1ST QUARTER | 2ND QUARTER | 3RD QUARTER | 4TH QUARTER | TOTAL GRAMS |
|---|---|---|---|---|---|
| 570 | 240.33 | 195.87 | 200.08 | 171.28 | **807.55** |
| 571 | 1,154.13 | 756.39 | 1,232.93 | 729.29 | **3,872.75** |
| 572 | 28.62 | 18.96 | 21.02 | 22.99 | **91.59** |
| 573 | 53.22 | 76.02 | 69.14 | 73.79 | **272.17** |
| 574 | 9.39 | 7.16 | 6.26 | 7.60 | **30.41** |
| 575 | 18.42 | 18.34 | 16.28 | 17.17 | **70.21** |
| 576 | 0 | 3.58 | 0 | 0 | **3.58** |
| 577 | 431.64 | 413.48 | 472.69 | 566.33 | **1,884.14** |

| SD - SOUTH DAKOTA - Total | 1,935.75 | 1,489.80 | 2,018.39 | 1,588.45 | 7,032.40 |
|---|---|---|---|---|---|

STATE: TN - TENNESSEE

| REGISTRANT ZIP CODE 3 | 1ST QUARTER | 2ND QUARTER | 3RD QUARTER | 4TH QUARTER | TOTAL GRAMS |
|---|---|---|---|---|---|
| 370 | 697.99 | 503.37 | 670.89 | 1,164.42 | **3,036.67** |
| 371 | 714.80 | 695.93 | 632.43 | 719.01 | **2,762.18** |
| 372 | 8,053.42 | 6,812.79 | 6,802.95 | 7,598.82 | **29,267.99** |
| 373 | 581.45 | 493.26 | 510.26 | 514.28 | **2,099.25** |
| 374 | 6,346.84 | 6,389.33 | 7,118.17 | 7,045.01 | **26,899.35** |
| 376 | 2,255.23 | 1,891.66 | 2,205.05 | 2,067.85 | **8,419.79** |
| 377 | 94.36 | 117.17 | 122.98 | 94.63 | **429.13** |
| 378 | 115.38 | 141.32 | 167.07 | 525.82 | **949.58** |
| 379 | 13,849.78 | 8,791.86 | 10,542.92 | 11,807.69 | **44,992.26** |
| 380 | 3,938.49 | 2,028.95 | 2,577.21 | 3,523.94 | **12,068.59** |
| 381 | 8,595.90 | 7,088.03 | 9,436.28 | 8,442.06 | **33,562.27** |
| 382 | 2,931.84 | 2,218.11 | 2,216.77 | 2,903.67 | **10,270.40** |
| 383 | 3,925.34 | 2,997.58 | 3,234.78 | 2,349.59 | **12,507.29** |
| 384 | 1,589.44 | 1,155.21 | 1,294.55 | 1,284.36 | **5,323.56** |
| 385 | 127.00 | 115.38 | 117.17 | 116.72 | **476.27** |
| TN - TENNESSEE - Total | 53,817.27 | 41,439.94 | 47,649.49 | 50,157.86 | 193,064.56 |

STATE: TX - TEXAS

| REGISTRANT ZIP CODE 3 | 1ST QUARTER | 2ND QUARTER | 3RD QUARTER | 4TH QUARTER | TOTAL GRAMS |
|---|---|---|---|---|---|
| 750 | 8,541.79 | 5,961.71 | 7,779.04 | 6,642.08 | **28,924.63** |
| 751 | 1,225.33 | 794.05 | 1,022.30 | 1,230.25 | **4,271.92** |
| 752 | 10,513.36 | 7,878.77 | 6,859.42 | 6,887.86 | **32,139.41** |
| 753 | 22.00 | 16.73 | 21.64 | 27.73 | **88.10** |
| 754 | 879.73 | 506.95 | 824.28 | 834.92 | **3,045.88** |
| 755 | 131.48 | 130.31 | 129.33 | 138.72 | **529.84** |
| 756 | 3,367.42 | 2,889.90 | 3,039.98 | 2,867.45 | **12,164.73** |
| 757 | 5,807.52 | 1,942.46 | 4,202.96 | 4,127.30 | **16,080.24** |
| 758 | 126.11 | 126.38 | 135.50 | 150.71 | **538.70** |
| 759 | 2,189.67 | 1,439.27 | 1,643.01 | 1,194.02 | **6,465.98** |
| 760 | 2,577.84 | 1,688.09 | 1,892.37 | 2,921.02 | **9,079.32** |
| 761 | 13,001.18 | 11,600.19 | 6,460.07 | 9,701.56 | **40,763.00** |
| 762 | 3,026.56 | 2,449.14 | 435.57 | 472.24 | **6,383.51** |
| 763 | 1,040.46 | 811.67 | 1,007.27 | 818.91 | **3,678.31** |
| 764 | 105.54 | 110.01 | 101.51 | 95.70 | **412.77** |
| 765 | 734.03 | 626.35 | 685.56 | 304.63 | **2,350.57** |
| 766 | 127.00 | 127.27 | 124.59 | 124.14 | **503.01** |
| 767 | 1,710.54 | 1,839.78 | 2,054.35 | 1,441.06 | **7,045.73** |
| 768 | 180.22 | 157.41 | 136.04 | 149.36 | **623.04** |
| 769 | 935.99 | 633.41 | 695.93 | 813.19 | **3,078.52** |
| 770 | 15,299.48 | 11,357.93 | 14,682.72 | 12,932.81 | **54,272.94** |
| 772 | 429.31 | 429.31 | 429.31 | 429.31 | **1,717.25** |
| 773 | 5,733.99 | 1,854.42 | 3,861.90 | 3,826.57 | **15,276.88** |
| 774 | 578.32 | 600.77 | 586.10 | 566.68 | **2,331.87** |
| 775 | 4,636.21 | 3,688.06 | 4,486.04 | 3,876.15 | **16,686.46** |
| 776 | 1,162.27 | 788.86 | 789.58 | 779.83 | **3,520.54** |
| 777 | 2,708.69 | 2,038.34 | 294.26 | 929.82 | **5,971.10** |
| 778 | 444.07 | 426.63 | 264.30 | 383.07 | **1,518.07** |
| 779 | 1,079.63 | 1,054.05 | 619.46 | 1,032.94 | **3,786.08** |
| 780 | 2,290.11 | 2,226.25 | 2,532.31 | 2,563.26 | **9,611.94** |
| 781 | 1,970.18 | 1,356.18 | 1,667.88 | 1,643.10 | **6,637.34** |
| 782 | 33,879.25 | 27,348.43 | 17,359.41 | 18,317.58 | **96,904.66** |
| 783 | 193.64 | 219.13 | 197.66 | 203.03 | **813.46** |
| 784 | 6,951.28 | 5,936.22 | 6,247.21 | 6,909.78 | **26,044.48** |
| 785 | 1,925.11 | 998.78 | 994.13 | 1,352.60 | **5,270.61** |
| 786 | 2,413.45 | 2,239.67 | 2,422.30 | 2,534.91 | **9,610.33** |
| 787 | 7,819.74 | 7,829.22 | 8,850.09 | 8,241.45 | **32,740.50** |
| 788 | 46.96 | 55.01 | 54.11 | 46.96 | **203.03** |
| 789 | 51.43 | 47.40 | 43.83 | 54.56 | **197.22** |

| | | | | | |
|---|---|---|---|---|---|
| 790 | 219.13 | 163.23 | 174.86 | 178.43 | **735.64** |
| 791 | 1,093.23 | 1,098.32 | 1,019.17 | 1,002.18 | **4,212.89** |
| 792 | 41.14 | 40.96 | 40.70 | 35.78 | **158.58** |
| 793 | 88.10 | 82.73 | 97.31 | 103.30 | **371.44** |
| 794 | 2,267.57 | 1,895.68 | 789.84 | 1,230.34 | **6,183.43** |
| 795 | 65.74 | 67.08 | 61.53 | 46.96 | **241.31** |
| 796 | 578.59 | 354.00 | 298.37 | 294.70 | **1,525.67** |
| 797 | 2,033.24 | 1,904.18 | 1,353.94 | 1,499.01 | **6,790.37** |
| 798 | 13.42 | 9.39 | 12.07 | 16.99 | **51.88** |
| 799 | 2,812.62 | 2,680.52 | 2,212.57 | 2,675.33 | **10,381.03** |
| **TX - TEXAS - Total** | **155,069.64** | **120,520.59** | **111,693.69** | **114,650.29** | **501,934.21** |

**STATE: UT - UTAH**

| REGISTRANT ZIP CODE 3 | 1ST QUARTER | 2ND QUARTER | 3RD QUARTER | 4TH QUARTER | TOTAL GRAMS |
|---|---|---|---|---|---|
| 840 | 8,364.34 | 6,892.60 | 6,477.87 | 6,487.17 | **28,221.99** |
| 841 | 16,147.68 | 13,899.69 | 13,459.74 | 14,722.09 | **58,229.20** |
| 843 | 567.94 | 562.58 | 560.79 | 542.72 | **2,234.03** |
| 844 | 3,772.22 | 3,152.40 | 2,602.53 | 4,218.44 | **13,745.59** |
| 845 | 495.50 | 517.32 | 500.51 | 871.50 | **2,384.83** |
| 846 | 1,591.23 | 548.09 | 1,097.25 | 1,037.95 | **4,274.52** |
| 847 | 3,401.49 | 2,898.48 | 1,092.24 | 1,065.41 | **8,457.63** |
| **UT - UTAH - Total** | **34,340.40** | **28,471.17** | **25,790.92** | **28,945.29** | **117,547.77** |

**STATE: VA - VIRGINIA**

| REGISTRANT ZIP CODE 3 | 1ST QUARTER | 2ND QUARTER | 3RD QUARTER | 4TH QUARTER | TOTAL GRAMS |
|---|---|---|---|---|---|
| 201 | 3,204.72 | 3,101.96 | 1,891.48 | 3,220.29 | **11,418.45** |
| 220 | 2,520.60 | 2,911.09 | 844.58 | 1,721.72 | **7,997.99** |
| 221 | 274.04 | 211.97 | 234.96 | 242.83 | **963.81** |
| 222 | 81.48 | 87.83 | 82.28 | 56.53 | **308.12** |
| 223 | 683.14 | 174.68 | 150.26 | 124.59 | **1,132.67** |
| 224 | 4,799.71 | 3,638.51 | 4,777.44 | 4,768.85 | **17,984.51** |
| 225 | 107.15 | 118.06 | 104.64 | 122.09 | **451.94** |
| 226 | 7,167.54 | 6,101.51 | 6,749.77 | 5,916.10 | **25,934.92** |
| 227 | 177.54 | 192.30 | 194.98 | 143.55 | **708.36** |
| 228 | 3,176.80 | 3,818.81 | 382.45 | 354.81 | **7,732.87** |
| 229 | 5,933.54 | 5,256.84 | 5,298.52 | 6,165.64 | **22,654.53** |
| 230 | 4,336.59 | 2,860.29 | 3,566.24 | 3,590.84 | **14,353.96** |
| 231 | 1,193.67 | 1,514.58 | 1,327.20 | 956.11 | **4,991.56** |
| 232 | 21,735.98 | 18,786.25 | 9,624.55 | 11,576.22 | **61,722.99** |
| 233 | 5,548.95 | 4,661.43 | 4,850.60 | 5,138.69 | **20,199.67** |
| 234 | 9,249.59 | 7,440.75 | 8,762.42 | 8,553.40 | **34,006.15** |
| 235 | 1,014.88 | 722.28 | 1,256.67 | 700.63 | **3,694.46** |
| 236 | 7,774.21 | 6,322.69 | 7,534.25 | 6,650.40 | **28,281.55** |
| 237 | 1,242.77 | 1,217.10 | 689.31 | 1,190.09 | **4,339.27** |
| 238 | 2,432.77 | 2,435.72 | 3,347.29 | 2,899.82 | **11,115.60** |
| 239 | 93.55 | 80.05 | 102.77 | 91.68 | **368.05** |
| 240 | 4,298.90 | 4,778.08 | 4,820.62 | 5,398.21 | **19,295.80** |
| 241 | 640.21 | 611.77 | 618.92 | 578.41 | **2,449.31** |
| 242 | 535.75 | 440.49 | 396.67 | 361.61 | **1,734.51** |
| 243 | 9,223.14 | 6,092.21 | 7,510.37 | 8,313.00 | **31,138.71** |
| 244 | 1,405.19 | 1,052.62 | 1,038.31 | 1,454.38 | **4,950.50** |
| 245 | 7,511.62 | 4,530.49 | 5,771.92 | 5,645.27 | **23,459.31** |
| 246 | 4,203.77 | 3,247.12 | 4,133.47 | 2,803.14 | **14,387.50** |
| **VA - VIRGINIA - Total** | **110,567.80** | **92,407.46** | **86,062.93** | **88,738.87** | **377,777.06** |

**STATE: VI - VIRGIN ISLANDS**

| REGISTRANT ZIP CODE 3 | 1ST QUARTER | 2ND QUARTER | 3RD QUARTER | 4TH QUARTER | TOTAL GRAMS |
|---|---|---|---|---|---|
| 008 | 9.84 | 13.42 | 8.94 | 17.89 | **50.09** |
| **VI - VIRGIN ISLANDS - Total** | **9.84** | **13.42** | **8.94** | **17.89** | **50.09** |

**STATE: VT - VERMONT**

| REGISTRANT ZIP CODE 3 | 1ST QUARTER | 2ND QUARTER | 3RD QUARTER | 4TH QUARTER | TOTAL GRAMS |
|---|---|---|---|---|---|
| 050 | 209.29 | 238.63 | 241.04 | 221.01 | **909.96** |
| 051 | 134.16 | 140.87 | 134.61 | 130.58 | **540.22** |

| | | | | | |
|---|---|---|---|---|---|
| 052 | 152.05 | 145.34 | 140.87 | 116.99 | **555.24** |
| 053 | 2,683.11 | 2,033.42 | 2,120.00 | 2,326.33 | **9,162.86** |
| 054 | 8,969.13 | 7,177.38 | 7,698.64 | 8,879.87 | **32,725.02** |
| 056 | 2,708.24 | 2,583.47 | 2,604.49 | 2,857.16 | **10,753.37** |
| 057 | 2,533.92 | 2,103.63 | 2,528.02 | 2,099.25 | **9,264.82** |
| 058 | 5,050.95 | 4,405.64 | 5,186.63 | 5,003.09 | **19,646.30** |
| **VT - VERMONT - Total** | **22,440.85** | **18,828.37** | **20,654.29** | **21,634.28** | **83,557.80** |

**STATE: WA - WASHINGTON**

| REGISTRANT ZIP CODE 3 | 1ST QUARTER | 2ND QUARTER | 3RD QUARTER | 4TH QUARTER | TOTAL GRAMS |
|---|---|---|---|---|---|
| 980 | 19,834.57 | 16,845.22 | 14,324.35 | 13,181.94 | **64,186.08** |
| 981 | 25,455.79 | 22,926.16 | 12,337.80 | 12,288.34 | **73,008.08** |
| 982 | 19,016.64 | 14,880.58 | 15,575.62 | 17,774.77 | **67,247.61** |
| 983 | 2,821.65 | 3,581.09 | 3,903.52 | 4,981.27 | **15,287.53** |
| 984 | 17,734.52 | 15,142.82 | 13,357.77 | 16,501.59 | **62,736.70** |
| 985 | 11,736.41 | 8,217.03 | 3,978.02 | 3,939.12 | **27,870.58** |
| 986 | 10,229.70 | 8,651.08 | 8,652.34 | 8,985.59 | **36,518.71** |
| 988 | 377.44 | 366.79 | 367.96 | 391.12 | **1,503.31** |
| 989 | 2,272.58 | 1,896.13 | 2,252.99 | 2,246.02 | **8,667.72** |
| 990 | 215.10 | 198.56 | 185.14 | 190.95 | **789.76** |
| 991 | 320.20 | 317.06 | 307.23 | 306.33 | **1,250.82** |
| 992 | 15,743.14 | 13,241.50 | 13,750.51 | 12,968.35 | **55,703.50** |
| 993 | 470.90 | 433.61 | 460.44 | 409.46 | **1,774.40** |
| 994 | 191.40 | 170.83 | 189.61 | 164.57 | **716.41** |
| **WA - WASHINGTON - Total** | **126,420.04** | **106,868.46** | **89,643.30** | **94,329.42** | **417,261.21** |

**STATE: WI - WISCONSIN**

| REGISTRANT ZIP CODE 3 | 1ST QUARTER | 2ND QUARTER | 3RD QUARTER | 4TH QUARTER | TOTAL GRAMS |
|---|---|---|---|---|---|
| 530 | 2,596.17 | 2,457.99 | 2,744.65 | 2,544.75 | **10,343.56** |
| 531 | 4,994.69 | 3,850.75 | 4,349.11 | 4,973.49 | **18,168.04** |
| 532 | 22,207.50 | 18,419.09 | 18,408.00 | 21,792.41 | **80,827.02** |
| 534 | 4,224.70 | 2,870.76 | 3,275.38 | 3,241.31 | **13,612.14** |
| 535 | 3,518.66 | 2,745.45 | 3,355.79 | 3,291.39 | **12,911.29** |
| 537 | 12,389.58 | 9,947.22 | 10,762.14 | 8,806.58 | **41,905.51** |
| 538 | 80.05 | 72.89 | 66.19 | 63.50 | **282.63** |
| 539 | 287.10 | 267.43 | 258.12 | 257.86 | **1,070.51** |
| 540 | 175.75 | 166.81 | 178.88 | 169.58 | **691.01** |
| 541 | 154.55 | 136.84 | 127.90 | 135.77 | **555.06** |
| 542 | 46.51 | 42.93 | 38.91 | 42.48 | **170.83** |
| 543 | 4,927.97 | 3,931.25 | 4,305.55 | 4,234.45 | **17,399.21** |
| 544 | 2,245.12 | 1,544.18 | 1,847.11 | 1,552.05 | **7,188.47** |
| 545 | 25.49 | 16.55 | 19.68 | 31.30 | **93.02** |
| 546 | 3,587.44 | 2,133.77 | 3,262.77 | 3,219.66 | **12,203.64** |
| 547 | 2,958.59 | 1,738.27 | 2,332.95 | 2,588.48 | **9,618.29** |
| 548 | 213.58 | 228.98 | 240.15 | 222.08 | **904.78** |
| 549 | 8,448.77 | 5,925.04 | 7,824.48 | 8,419.70 | **30,618.00** |
| **WI - WISCONSIN - Total** | **73,082.22** | **56,496.18** | **63,397.75** | **65,586.85** | **258,563.01** |

**STATE: WV - WEST VIRGINIA**

| REGISTRANT ZIP CODE 3 | 1ST QUARTER | 2ND QUARTER | 3RD QUARTER | 4TH QUARTER | TOTAL GRAMS |
|---|---|---|---|---|---|
| 247 | 44.72 | 33.72 | 33.27 | 25.40 | **137.11** |
| 248 | 22.36 | 13.42 | 16.99 | 8.94 | **61.71** |
| 249 | 29.96 | 25.49 | 22.36 | 15.65 | **93.46** |
| 250 | 21.47 | 34.08 | 25.49 | 36.22 | **117.26** |
| 251 | 21.91 | 12.52 | 17.89 | 9.84 | **62.16** |
| 252 | 10.64 | 11.18 | 5.81 | 10.73 | **38.37** |
| 253 | 4,561.62 | 2,888.64 | 2,054.44 | 3,395.95 | **12,900.65** |
| 254 | 3,476.53 | 3,858.44 | 4,186.69 | 3,452.21 | **14,973.87** |
| 255 | 57.24 | 73.34 | 77.81 | 76.47 | **284.87** |
| 256 | 2,955.54 | 2,269.54 | 2,595.55 | 2,668.44 | **10,489.08** |
| 257 | 5,524.08 | 5,304.87 | 4,454.02 | 5,349.94 | **20,632.91** |
| 258 | 3,239.07 | 3,029.78 | 3,490.40 | 3,245.78 | **13,005.02** |
| 259 | 38.91 | 27.73 | 33.54 | 25.94 | **126.11** |

| | | | | | |
|---|---|---|---|---|---|
| 260 | 1,921.89 | 451.31 | 877.23 | 1,245.63 | **4,496.06** |
| 261 | 1,590.51 | 638.60 | 1,355.46 | 923.47 | **4,508.04** |
| 262 | 57.69 | 59.92 | 52.77 | 50.09 | **220.47** |
| 263 | 2,004.35 | 1,383.64 | 1,431.93 | 1,381.85 | **6,201.77** |
| 264 | 44.27 | 38.46 | 38.46 | 30.41 | **151.60** |
| 265 | 2,823.17 | 2,809.13 | 3,376.99 | 2,276.52 | **11,285.81** |
| 266 | 6.71 | 8.05 | 13.42 | 13.42 | **41.59** |
| 267 | 54.74 | 42.93 | 44.72 | 40.70 | **183.08** |
| 268 | 17.89 | 22.81 | 25.94 | 21.91 | **88.55** |
| **WV - WEST VIRGINIA - Total** | **28,525.28** | **23,037.60** | **24,231.17** | **24,305.50** | **100,099.55** |

**STATE: WY - WYOMING**

| REGISTRANT ZIP CODE 3 | 1ST QUARTER | 2ND QUARTER | 3RD QUARTER | 4TH QUARTER | TOTAL GRAMS |
|---|---|---|---|---|---|
| 820 | 95.88 | 77.37 | 75.58 | 66.19 | **315.01** |
| 822 | 29.34 | 25.76 | 22.36 | 25.49 | **102.95** |
| 823 | 9.84 | 13.42 | 17.89 | 22.36 | **63.50** |
| 824 | 42.48 | 38.46 | 46.51 | 33.99 | **161.44** |
| 825 | 51.43 | 65.29 | 67.53 | 62.61 | **246.85** |
| 826 | 147.04 | 131.21 | 131.92 | 116.63 | **526.80** |
| 827 | 25.49 | 22.36 | 24.60 | 19.23 | **91.68** |
| 828 | 56.35 | 49.64 | 39.80 | 36.22 | **182.01** |
| 829 | 191.40 | 186.48 | 170.83 | 151.15 | **699.87** |
| 830 | 24.15 | 16.10 | 19.23 | 26.83 | **86.31** |
| 831 | 25.49 | 22.36 | 25.49 | 23.70 | **97.04** |
| **WY - WYOMING - Total** | **698.88** | **648.44** | **641.73** | **584.40** | **2,573.46** |

**DRUG: 9300 - MORPHINE**

**STATE: AK - ALASKA**

| REGISTRANT ZIP CODE 3 | 1ST QUARTER | 2ND QUARTER | 3RD QUARTER | 4TH QUARTER | TOTAL GRAMS |
|---|---|---|---|---|---|
| 995 | 3,034.86 | 2,966.44 | 2,879.82 | 2,735.69 | **11,616.81** |
| 996 | 2,694.39 | 2,622.33 | 2,554.17 | 2,525.14 | **10,396.02** |
| 997 | 703.39 | 686.36 | 782.71 | 651.18 | **2,823.65** |
| 998 | 337.65 | 310.79 | 358.50 | 282.02 | **1,288.97** |
| 999 | 191.89 | 154.30 | 177.61 | 153.30 | **677.10** |
| **AK - ALASKA - Total** | **6,962.18** | **6,740.22** | **6,752.82** | **6,347.33** | **26,802.55** |

**STATE: AL - ALABAMA**

| REGISTRANT ZIP CODE 3 | 1ST QUARTER | 2ND QUARTER | 3RD QUARTER | 4TH QUARTER | TOTAL GRAMS |
|---|---|---|---|---|---|
| 350 | 5,458.41 | 4,770.88 | 5,134.00 | 4,950.35 | **20,313.64** |
| 351 | 2,725.19 | 2,677.22 | 2,601.22 | 2,639.56 | **10,643.20** |
| 352 | 4,514.43 | 4,162.40 | 4,066.06 | 4,121.70 | **16,864.59** |
| 354 | 3,069.57 | 2,770.44 | 2,777.72 | 2,533.72 | **11,151.46** |
| 355 | 1,705.22 | 1,548.47 | 1,465.40 | 1,471.50 | **6,190.60** |
| 356 | 3,558.60 | 3,249.78 | 3,287.67 | 3,133.30 | **13,229.35** |
| 357 | 1,847.03 | 1,882.29 | 1,988.21 | 1,869.90 | **7,587.43** |
| 358 | 2,655.60 | 2,360.68 | 2,522.65 | 2,573.02 | **10,111.95** |
| 359 | 4,490.94 | 4,102.18 | 4,303.25 | 4,082.90 | **16,979.27** |
| 360 | 2,871.97 | 2,573.57 | 2,646.07 | 2,585.45 | **10,677.06** |
| 361 | 2,283.30 | 2,138.97 | 1,848.24 | 1,699.75 | **7,970.25** |
| 362 | 1,819.64 | 1,867.68 | 1,832.75 | 1,894.85 | **7,414.93** |
| 363 | 1,592.41 | 1,611.74 | 1,690.47 | 1,778.42 | **6,673.04** |
| 364 | 851.00 | 888.89 | 792.05 | 724.30 | **3,256.23** |
| 365 | 3,648.59 | 3,452.86 | 3,694.97 | 3,527.29 | **14,323.71** |
| 366 | 3,236.21 | 2,976.24 | 2,903.73 | 2,933.91 | **12,050.10** |
| 367 | 410.96 | 465.73 | 439.11 | 463.33 | **1,779.14** |
| 368 | 2,541.78 | 2,602.83 | 2,722.92 | 2,667.58 | **10,535.11** |
| 369 | 75.49 | 78.36 | 80.99 | 65.21 | **300.05** |
| **AL - ALABAMA - Total** | **49,356.36** | **46,181.20** | **46,797.49** | **45,716.05** | **188,051.10** |

**STATE: AR - ARKANSAS**

| REGISTRANT ZIP CODE 3 | 1ST QUARTER | 2ND QUARTER | 3RD QUARTER | 4TH QUARTER | TOTAL GRAMS |
|---|---|---|---|---|---|
| 716 | 771.71 | 668.47 | 711.22 | 699.92 | **2,851.32** |
| 717 | 798.04 | 809.19 | 758.13 | 759.67 | **3,125.03** |
| 718 | 577.40 | 562.91 | 533.03 | 583.44 | **2,256.79** |

| | | | | | |
|---|---|---|---|---|---|
| 719 | 1,886.52 | 1,948.61 | 1,929.19 | 1,969.88 | **7,734.19** |
| 720 | 2,834.32 | 2,488.09 | 2,587.36 | 2,458.00 | **10,367.77** |
| 721 | 2,962.78 | 2,914.53 | 2,816.40 | 2,756.22 | **11,449.92** |
| 722 | 1,977.57 | 1,726.15 | 1,728.25 | 1,691.32 | **7,123.28** |
| 723 | 912.46 | 994.97 | 919.26 | 986.25 | **3,812.94** |
| 724 | 1,986.78 | 1,913.81 | 2,030.19 | 1,789.56 | **7,720.33** |
| 725 | 1,095.12 | 1,159.19 | 1,052.71 | 1,079.29 | **4,386.30** |
| 726 | 1,605.24 | 1,697.60 | 1,738.98 | 1,797.82 | **6,839.64** |
| 727 | 4,624.44 | 4,017.19 | 4,164.07 | 4,045.87 | **16,851.58** |
| 728 | 953.09 | 940.26 | 955.25 | 909.54 | **3,758.14** |
| 729 | 2,249.02 | 2,221.14 | 2,252.52 | 2,174.30 | **8,896.98** |
| **AR - ARKANSAS - Total** | **25,234.47** | **24,062.10** | **24,176.56** | **23,701.08** | **97,174.21** |

**STATE: AS - AMERICAN SAMOA**

| REGISTRANT ZIP CODE 3 | 1ST QUARTER | 2ND QUARTER | 3RD QUARTER | 4TH QUARTER | TOTAL GRAMS |
|---|---|---|---|---|---|
| 967 | 6.77 | 9.85 | 0.30 | 5.79 | **22.71** |
| **AS - AMERICAN SAMOA - Total** | **6.77** | **9.85** | **0.30** | **5.79** | **22.71** |

**STATE: AZ - ARIZONA**

| REGISTRANT ZIP CODE 3 | 1ST QUARTER | 2ND QUARTER | 3RD QUARTER | 4TH QUARTER | TOTAL GRAMS |
|---|---|---|---|---|---|
| 850 | 13,475.41 | 12,361.11 | 12,203.10 | 11,564.58 | **49,604.20** |
| 851 | 3,370.96 | 3,254.95 | 3,251.36 | 3,080.96 | **12,958.22** |
| 852 | 19,542.13 | 18,574.85 | 18,655.24 | 18,117.95 | **74,890.16** |
| 853 | 12,795.04 | 12,121.43 | 11,559.30 | 11,881.79 | **48,357.57** |
| 855 | 1,962.64 | 1,881.20 | 1,790.89 | 1,619.99 | **7,254.72** |
| 856 | 3,105.35 | 3,170.18 | 3,141.90 | 3,116.88 | **12,534.31** |
| 857 | 18,246.03 | 17,766.88 | 17,319.90 | 16,785.35 | **70,118.16** |
| 859 | 1,925.80 | 1,826.83 | 2,040.54 | 1,764.02 | **7,557.19** |
| 860 | 1,471.78 | 1,485.44 | 1,479.86 | 1,482.09 | **5,919.16** |
| 863 | 4,499.43 | 4,014.29 | 4,325.32 | 4,212.00 | **17,051.04** |
| 864 | 4,803.03 | 4,398.04 | 4,359.79 | 4,416.04 | **17,976.91** |
| 865 | 137.16 | 99.51 | 126.16 | 123.88 | **486.72** |
| **AZ - ARIZONA - Total** | **85,334.77** | **80,954.71** | **80,253.35** | **78,165.53** | **324,708.36** |

**STATE: CA - CALIFORNIA**

| REGISTRANT ZIP CODE 3 | 1ST QUARTER | 2ND QUARTER | 3RD QUARTER | 4TH QUARTER | TOTAL GRAMS |
|---|---|---|---|---|---|
| 900 | 4,922.65 | 4,611.98 | 4,229.39 | 4,146.78 | **17,910.80** |
| 902 | 3,217.21 | 3,106.35 | 2,839.66 | 2,861.43 | **12,024.65** |
| 903 | 216.10 | 172.70 | 213.09 | 157.84 | **759.74** |
| 904 | 688.96 | 657.74 | 658.75 | 716.62 | **2,722.08** |
| 905 | 1,540.46 | 1,346.81 | 1,259.76 | 1,324.35 | **5,471.38** |
| 906 | 5,531.80 | 4,908.18 | 4,932.40 | 5,182.23 | **20,554.62** |
| 907 | 2,959.01 | 2,856.60 | 2,793.83 | 2,691.35 | **11,300.79** |
| 908 | 2,767.35 | 2,680.41 | 2,861.34 | 2,801.98 | **11,111.08** |
| 910 | 1,585.98 | 1,395.84 | 1,525.71 | 1,369.66 | **5,877.19** |
| 911 | 952.61 | 864.53 | 916.89 | 838.92 | **3,572.94** |
| 912 | 2,413.22 | 2,394.49 | 2,598.41 | 2,434.43 | **9,840.56** |
| 913 | 8,073.00 | 7,808.84 | 7,611.55 | 7,356.70 | **30,850.09** |
| 914 | 2,230.34 | 1,674.77 | 1,676.68 | 1,985.22 | **7,567.01** |
| 915 | 1,140.42 | 1,061.07 | 825.10 | 904.75 | **3,931.33** |
| 916 | 416.09 | 398.36 | 307.96 | 376.46 | **1,498.87** |
| 917 | 10,589.67 | 9,493.15 | 9,708.58 | 9,685.74 | **39,477.14** |
| 918 | 132.43 | 156.72 | 141.89 | 127.39 | **558.43** |
| 919 | 2,810.78 | 2,731.55 | 2,864.17 | 2,718.89 | **11,125.39** |
| 920 | 13,162.96 | 12,167.70 | 12,432.31 | 11,691.41 | **49,454.38** |
| 921 | 8,167.32 | 7,514.02 | 7,404.47 | 7,476.67 | **30,562.49** |
| 922 | 6,145.93 | 5,933.08 | 5,619.60 | 5,916.35 | **23,614.95** |
| 923 | 12,260.10 | 11,829.00 | 11,380.70 | 11,405.65 | **46,875.46** |
| 924 | 1,107.73 | 949.66 | 1,007.54 | 1,020.61 | **4,085.56** |
| 925 | 11,075.79 | 9,785.72 | 10,261.26 | 9,867.08 | **40,989.85** |
| 926 | 8,365.25 | 8,262.59 | 7,958.64 | 8,155.49 | **32,741.96** |
| 927 | 1,928.30 | 1,778.06 | 1,893.78 | 1,815.82 | **7,415.97** |
| 928 | 8,483.65 | 8,212.08 | 7,717.20 | 7,824.50 | **32,237.43** |

| | | | | | |
|---|---|---|---|---|---|
| 930 | 4,226.22 | 3,655.58 | 4,020.55 | 3,969.20 | **15,871.54** |
| 931 | 707.88 | 613.83 | 574.08 | 685.43 | **2,581.21** |
| 932 | 4,900.80 | 4,392.23 | 4,671.86 | 4,526.57 | **18,491.45** |
| 933 | 6,079.66 | 5,738.26 | 5,752.50 | 5,637.50 | **23,207.92** |
| 934 | 5,511.55 | 5,651.24 | 5,502.99 | 5,490.01 | **22,155.80** |
| 935 | 6,077.04 | 5,576.16 | 5,554.15 | 5,254.05 | **22,461.39** |
| 936 | 3,988.71 | 3,780.66 | 3,623.89 | 3,586.71 | **14,979.97** |
| 937 | 6,379.50 | 6,176.40 | 6,180.58 | 6,039.13 | **24,775.60** |
| 939 | 2,290.07 | 2,089.65 | 2,145.47 | 2,110.43 | **8,635.62** |
| 940 | 5,859.18 | 5,033.93 | 4,966.68 | 5,183.63 | **21,043.42** |
| 941 | 8,501.16 | 7,782.84 | 7,350.27 | 7,111.05 | **30,745.32** |
| 943 | 1,108.50 | 855.06 | 1,044.70 | 999.99 | **4,008.25** |
| 944 | 626.94 | 753.13 | 692.00 | 655.70 | **2,727.77** |
| 945 | 21,187.41 | 21,905.58 | 20,782.88 | 20,268.66 | **84,144.52** |
| 946 | 2,315.26 | 2,010.79 | 2,213.12 | 1,956.42 | **8,495.60** |
| 947 | 2,659.75 | 731.94 | 1,138.37 | 1,837.82 | **6,367.87** |
| 948 | 821.23 | 681.79 | 677.15 | 676.85 | **2,857.02** |
| 949 | 3,491.88 | 3,910.17 | 3,493.63 | 3,144.70 | **14,040.39** |
| 950 | 3,504.91 | 3,078.51 | 3,455.23 | 3,197.66 | **13,236.31** |
| 951 | 2,161.38 | 1,935.76 | 1,947.81 | 2,023.22 | **8,068.17** |
| 952 | 4,945.30 | 4,262.50 | 4,420.05 | 4,506.47 | **18,134.32** |
| 953 | 13,517.17 | 13,253.12 | 13,198.09 | 12,669.95 | **52,638.33** |
| 954 | 7,002.00 | 6,680.24 | 6,873.84 | 6,488.84 | **27,044.92** |
| 955 | 2,679.26 | 2,687.79 | 2,678.27 | 2,617.49 | **10,662.81** |
| 956 | 15,934.41 | 14,413.86 | 14,674.19 | 13,904.40 | **58,926.85** |
| 957 | 2,417.17 | 2,173.91 | 2,172.58 | 2,164.74 | **8,928.39** |
| 958 | 8,856.66 | 8,749.95 | 8,452.59 | 7,900.09 | **33,959.29** |
| 959 | 5,907.13 | 5,526.59 | 5,221.77 | 5,254.26 | **21,909.75** |
| 960 | 4,953.92 | 4,638.30 | 4,676.73 | 4,973.38 | **19,242.33** |
| 961 | 986.51 | 931.94 | 868.50 | 838.40 | **3,625.35** |
| **CA - CALIFORNIA - Total** | **282,483.68** | **264,423.71** | **262,665.18** | **258,527.06** | **1,068,099.63** |

**STATE: CO - COLORADO**

| REGISTRANT ZIP CODE 3 | 1ST QUARTER | 2ND QUARTER | 3RD QUARTER | 4TH QUARTER | TOTAL GRAMS |
|---|---|---|---|---|---|
| 800 | 9,711.05 | 8,331.13 | 8,282.83 | 8,358.92 | **34,683.93** |
| 801 | 9,017.30 | 8,645.28 | 8,753.96 | 8,799.51 | **35,216.04** |
| 802 | 7,765.99 | 7,910.01 | 7,694.51 | 6,945.44 | **30,315.94** |
| 803 | 689.46 | 561.10 | 661.71 | 650.16 | **2,562.43** |
| 804 | 1,796.45 | 1,730.76 | 1,713.99 | 1,674.03 | **6,915.23** |
| 805 | 4,805.23 | 4,184.16 | 4,062.59 | 4,469.66 | **17,521.64** |
| 806 | 2,006.86 | 1,793.72 | 2,002.03 | 2,069.72 | **7,872.33** |
| 807 | 733.01 | 771.57 | 720.61 | 772.71 | **2,997.90** |
| 808 | 1,007.02 | 1,099.95 | 1,172.12 | 1,209.94 | **4,489.03** |
| 809 | 5,712.72 | 5,938.90 | 5,928.87 | 5,672.48 | **23,252.98** |
| 810 | 2,919.29 | 3,079.76 | 3,209.67 | 3,181.80 | **12,390.52** |
| 811 | 416.85 | 454.66 | 398.92 | 439.47 | **1,709.90** |
| 812 | 884.38 | 899.04 | 983.89 | 941.38 | **3,708.69** |
| 813 | 408.97 | 418.53 | 453.55 | 518.09 | **1,799.14** |
| 814 | 434.28 | 416.95 | 442.07 | 442.69 | **1,735.99** |
| 815 | 1,378.79 | 1,385.19 | 1,312.96 | 1,486.56 | **5,563.49** |
| 816 | 928.20 | 812.44 | 741.98 | 731.46 | **3,214.08** |
| **CO - COLORADO - Total** | **50,615.85** | **48,433.15** | **48,536.27** | **48,364.01** | **195,949.27** |

**STATE: CT - CONNECTICUT**

| REGISTRANT ZIP CODE 3 | 1ST QUARTER | 2ND QUARTER | 3RD QUARTER | 4TH QUARTER | TOTAL GRAMS |
|---|---|---|---|---|---|
| 060 | 4,651.84 | 4,628.34 | 4,452.68 | 4,337.18 | **18,070.05** |
| 061 | 1,240.05 | 1,192.68 | 1,211.88 | 1,279.67 | **4,924.28** |
| 062 | 1,116.23 | 1,123.94 | 1,198.54 | 1,231.00 | **4,669.71** |
| 063 | 1,480.95 | 1,557.59 | 1,629.17 | 1,561.86 | **6,229.57** |
| 064 | 8,165.10 | 8,035.91 | 7,258.22 | 7,574.63 | **31,033.85** |
| 065 | 2,611.19 | 3,203.49 | 2,149.21 | 2,267.87 | **10,231.77** |
| 066 | 1,266.25 | 1,309.50 | 1,237.41 | 1,259.27 | **5,072.44** |

| | | | | | |
|---|---|---|---|---|---|
| 067 | 2,235.76 | 2,082.52 | 2,140.15 | 2,188.61 | **8,647.04** |
| 068 | 2,420.60 | 2,406.93 | 2,234.87 | 2,198.24 | **9,260.63** |
| 069 | 144.03 | 212.78 | 218.94 | 206.52 | **782.28** |
| **CT - CONNECTICUT - Total** | **25,332.00** | **25,753.67** | **23,731.07** | **24,104.86** | **98,921.60** |
| **STATE: DC - DISTRICT OF COLUMBIA** | | | | | |
| REGISTRANT ZIP CODE 3 | 1ST QUARTER | 2ND QUARTER | 3RD QUARTER | 4TH QUARTER | **TOTAL GRAMS** |
| 200 | 1,944.69 | 1,790.40 | 1,883.79 | 1,760.86 | **7,379.75** |
| 203 | 16.92 | 0 | 0.45 | 0 | **17.37** |
| 204 | 747.71 | 1,076.09 | 546.91 | 556.73 | **2,927.44** |
| 205 | 0.38 | 0.68 | 0 | 0.38 | **1.43** |
| **DC - DISTRICT OF COLUMBIA - Total** | **2,709.70** | **2,867.17** | **2,431.15** | **2,317.96** | **10,325.98** |
| **STATE: DE - DELAWARE** | | | | | |
| REGISTRANT ZIP CODE 3 | 1ST QUARTER | 2ND QUARTER | 3RD QUARTER | 4TH QUARTER | **TOTAL GRAMS** |
| 197 | 5,401.71 | 5,264.62 | 5,532.05 | 5,636.68 | **21,835.05** |
| 198 | 3,155.44 | 2,877.78 | 2,869.49 | 2,995.83 | **11,898.55** |
| 199 | 5,808.24 | 5,304.15 | 5,490.84 | 5,341.76 | **21,945.00** |
| **DE - DELAWARE - Total** | **14,365.39** | **13,446.55** | **13,892.38** | **13,974.27** | **55,678.60** |
| **STATE: FL - FLORIDA** | | | | | |
| REGISTRANT ZIP CODE 3 | 1ST QUARTER | 2ND QUARTER | 3RD QUARTER | 4TH QUARTER | **TOTAL GRAMS** |
| 320 | 7,800.06 | 7,796.91 | 7,521.67 | 7,220.07 | **30,338.71** |
| 321 | 11,002.28 | 10,595.67 | 10,729.92 | 10,430.92 | **42,758.79** |
| 322 | 12,985.03 | 12,487.45 | 12,805.94 | 12,351.25 | **50,629.67** |
| 323 | 3,982.46 | 4,104.01 | 3,948.04 | 3,906.13 | **15,940.64** |
| 324 | 3,383.92 | 3,196.84 | 2,955.77 | 2,952.12 | **12,488.65** |
| 325 | 5,694.52 | 5,233.49 | 5,128.42 | 5,442.00 | **21,498.43** |
| 326 | 6,113.54 | 6,073.35 | 5,907.04 | 5,624.05 | **23,717.99** |
| 327 | 15,901.52 | 14,868.86 | 14,749.61 | 14,126.93 | **59,646.92** |
| 328 | 7,664.93 | 7,360.22 | 8,262.37 | 7,913.12 | **31,200.63** |
| 329 | 12,899.19 | 12,372.47 | 11,999.15 | 12,423.33 | **49,694.15** |
| 330 | 14,531.03 | 13,874.54 | 13,816.31 | 14,113.13 | **56,335.01** |
| 331 | 5,753.69 | 5,618.74 | 5,913.79 | 5,710.30 | **22,996.51** |
| 333 | 11,543.16 | 11,506.67 | 11,267.30 | 11,191.41 | **45,508.55** |
| 334 | 22,183.89 | 22,093.85 | 21,452.52 | 21,318.51 | **87,048.77** |
| 335 | 11,455.52 | 11,169.12 | 11,312.72 | 11,094.37 | **45,031.73** |
| 336 | 19,526.11 | 19,122.10 | 17,793.89 | 15,044.86 | **71,486.96** |
| 337 | 18,030.08 | 17,219.33 | 17,046.98 | 17,561.98 | **69,858.37** |
| 338 | 5,841.96 | 5,746.70 | 5,529.34 | 5,562.19 | **22,680.18** |
| 339 | 13,957.54 | 13,248.67 | 13,565.48 | 14,042.48 | **54,814.16** |
| 341 | 4,519.05 | 4,453.39 | 4,358.60 | 4,459.11 | **17,790.15** |
| 342 | 14,862.65 | 14,660.79 | 14,383.55 | 14,542.50 | **58,449.48** |
| 344 | 8,887.35 | 8,839.71 | 8,785.59 | 8,623.01 | **35,135.66** |
| 346 | 19,060.31 | 18,492.06 | 19,074.38 | 18,475.33 | **75,102.07** |
| 347 | 7,628.53 | 7,110.64 | 7,187.97 | 6,903.91 | **28,831.05** |
| 349 | 12,235.37 | 11,522.64 | 11,281.89 | 10,994.20 | **46,034.09** |
| **FL - FLORIDA - Total** | **277,443.67** | **268,768.21** | **266,778.24** | **262,027.20** | **1,075,017.32** |
| **STATE: GA - GEORGIA** | | | | | |
| REGISTRANT ZIP CODE 3 | 1ST QUARTER | 2ND QUARTER | 3RD QUARTER | 4TH QUARTER | **TOTAL GRAMS** |
| 300 | 15,983.69 | 15,511.95 | 15,155.97 | 15,421.95 | **62,073.57** |
| 301 | 11,858.54 | 11,715.49 | 11,863.54 | 11,497.92 | **46,935.49** |
| 302 | 7,257.63 | 7,503.13 | 7,814.49 | 7,406.17 | **29,981.43** |
| 303 | 4,490.21 | 4,142.68 | 4,273.06 | 4,156.36 | **17,062.32** |
| 304 | 1,751.84 | 1,677.36 | 1,683.08 | 1,600.48 | **6,712.76** |
| 305 | 6,774.46 | 6,491.33 | 6,867.66 | 6,470.61 | **26,604.06** |
| 306 | 3,009.69 | 2,820.47 | 2,913.86 | 2,725.32 | **11,469.35** |
| 307 | 2,306.78 | 2,134.45 | 2,154.53 | 2,117.34 | **8,713.11** |
| 308 | 1,686.40 | 1,640.80 | 1,482.82 | 1,581.36 | **6,391.38** |
| 309 | 2,829.00 | 2,804.88 | 2,725.81 | 2,320.69 | **10,680.39** |
| 310 | 4,585.14 | 4,309.55 | 4,503.35 | 4,207.52 | **17,605.56** |
| 312 | 1,719.06 | 1,747.52 | 1,484.36 | 1,414.35 | **6,365.29** |
| 313 | 1,558.99 | 1,452.29 | 1,533.57 | 1,363.64 | **5,908.50** |

| | | | | | |
|---|---|---|---|---|---|
| 314 | 1,921.66 | 1,831.89 | 1,783.86 | 1,914.34 | **7,451.75** |
| 315 | 2,926.08 | 2,948.82 | 3,037.20 | 2,976.58 | **11,888.68** |
| 316 | 2,734.21 | 2,408.94 | 2,512.64 | 2,509.28 | **10,165.07** |
| 317 | 2,706.80 | 2,541.43 | 2,487.54 | 2,612.34 | **10,348.10** |
| 318 | 366.18 | 349.83 | 405.06 | 362.74 | **1,483.82** |
| 319 | 2,291.53 | 2,457.35 | 2,519.45 | 2,190.60 | **9,458.92** |
| 398 | 660.61 | 627.92 | 734.28 | 693.86 | **2,716.67** |
| **GA - GEORGIA - Total** | **79,418.53** | **77,118.09** | **77,936.13** | **75,543.45** | **310,016.19** |

**STATE: GU - GUAM**

| REGISTRANT ZIP CODE 3 | 1ST QUARTER | 2ND QUARTER | 3RD QUARTER | 4TH QUARTER | TOTAL GRAMS |
|---|---|---|---|---|---|
| 969 | 673.81 | 663.97 | 682.85 | 670.71 | **2,691.33** |
| **GU - GUAM - Total** | **673.81** | **663.97** | **682.85** | **670.71** | **2,691.33** |

**STATE: HI - HAWAII**

| REGISTRANT ZIP CODE 3 | 1ST QUARTER | 2ND QUARTER | 3RD QUARTER | 4TH QUARTER | TOTAL GRAMS |
|---|---|---|---|---|---|
| 967 | 8,103.78 | 7,530.09 | 7,359.72 | 7,336.57 | **30,330.17** |
| 968 | 5,021.57 | 4,408.33 | 3,918.60 | 4,084.68 | **17,433.18** |
| **HI - HAWAII - Total** | **13,125.34** | **11,938.42** | **11,278.33** | **11,421.26** | **47,763.35** |

**STATE: IA - IOWA**

| REGISTRANT ZIP CODE 3 | 1ST QUARTER | 2ND QUARTER | 3RD QUARTER | 4TH QUARTER | TOTAL GRAMS |
|---|---|---|---|---|---|
| 500 | 1,818.94 | 1,586.31 | 1,684.22 | 1,666.85 | **6,756.32** |
| 501 | 1,553.23 | 1,564.54 | 1,458.68 | 1,618.82 | **6,195.27** |
| 502 | 1,322.57 | 1,391.06 | 1,338.12 | 1,352.48 | **5,404.23** |
| 503 | 3,364.61 | 3,149.53 | 3,390.67 | 3,463.90 | **13,368.72** |
| 504 | 770.21 | 724.04 | 764.49 | 749.82 | **3,008.57** |
| 505 | 1,155.13 | 1,042.02 | 1,073.23 | 1,189.04 | **4,459.42** |
| 506 | 751.19 | 778.97 | 628.08 | 776.51 | **2,934.74** |
| 507 | 664.32 | 698.89 | 648.27 | 673.09 | **2,684.56** |
| 508 | 330.03 | 350.27 | 365.80 | 312.31 | **1,358.41** |
| 510 | 365.35 | 445.27 | 394.52 | 392.24 | **1,597.37** |
| 511 | 964.69 | 984.25 | 982.90 | 921.20 | **3,853.04** |
| 512 | 240.95 | 211.75 | 212.20 | 256.22 | **921.12** |
| 513 | 354.67 | 314.74 | 304.00 | 427.70 | **1,401.11** |
| 514 | 457.28 | 407.92 | 487.80 | 407.09 | **1,760.09** |
| 515 | 801.09 | 813.57 | 824.27 | 831.52 | **3,270.45** |
| 516 | 127.14 | 110.77 | 110.78 | 136.91 | **485.60** |
| 520 | 1,061.95 | 875.16 | 1,057.52 | 928.68 | **3,923.30** |
| 521 | 301.30 | 274.69 | 279.56 | 288.11 | **1,143.66** |
| 522 | 1,264.55 | 1,157.63 | 1,162.02 | 1,291.86 | **4,876.05** |
| 523 | 513.74 | 468.88 | 574.64 | 546.57 | **2,103.84** |
| 524 | 1,083.80 | 1,095.09 | 969.35 | 1,075.34 | **4,223.58** |
| 525 | 1,084.40 | 1,042.77 | 1,051.60 | 1,079.40 | **4,258.17** |
| 526 | 878.09 | 924.97 | 974.53 | 1,057.19 | **3,834.77** |
| 527 | 952.00 | 787.84 | 848.85 | 877.79 | **3,466.48** |
| 528 | 1,292.51 | 1,263.93 | 1,212.71 | 1,135.98 | **4,905.12** |
| **IA - IOWA - Total** | **23,473.75** | **22,464.85** | **22,798.79** | **23,456.61** | **92,194.00** |

**STATE: ID - IDAHO**

| REGISTRANT ZIP CODE 3 | 1ST QUARTER | 2ND QUARTER | 3RD QUARTER | 4TH QUARTER | TOTAL GRAMS |
|---|---|---|---|---|---|
| 832 | 2,277.90 | 2,068.94 | 2,154.44 | 2,214.91 | **8,716.18** |
| 833 | 1,874.46 | 1,750.45 | 1,852.24 | 1,705.92 | **7,183.07** |
| 834 | 1,468.80 | 1,338.00 | 1,432.87 | 1,412.83 | **5,652.50** |
| 835 | 999.58 | 1,009.75 | 1,049.45 | 1,050.52 | **4,109.31** |
| 836 | 5,637.77 | 5,729.95 | 5,305.69 | 5,457.79 | **22,131.20** |
| 837 | 3,796.25 | 3,662.11 | 3,920.44 | 3,752.03 | **15,130.84** |
| 838 | 3,031.02 | 3,219.46 | 3,039.08 | 3,137.66 | **12,427.21** |
| **ID - IDAHO - Total** | **19,085.79** | **18,778.67** | **18,754.20** | **18,731.66** | **75,350.32** |

**STATE: IL - ILLINOIS**

| REGISTRANT ZIP CODE 3 | 1ST QUARTER | 2ND QUARTER | 3RD QUARTER | 4TH QUARTER | TOTAL GRAMS |
|---|---|---|---|---|---|
| 600 | 16,073.37 | 15,399.24 | 14,300.38 | 15,013.84 | **60,786.83** |
| 601 | 6,921.86 | 6,380.99 | 6,169.59 | 6,276.79 | **25,749.23** |
| 602 | 188.73 | 256.47 | 200.94 | 185.88 | **832.02** |

| | | | | | |
|---|---|---|---|---|---|
| 603 | 395.52 | 362.20 | 348.79 | 338.54 | **1,445.05** |
| 604 | 8,246.23 | 8,335.62 | 7,755.77 | 8,384.23 | **32,721.84** |
| 605 | 4,570.89 | 4,203.25 | 4,172.95 | 4,282.61 | **17,229.71** |
| 606 | 8,150.45 | 7,780.62 | 7,625.09 | 7,657.36 | **31,213.52** |
| 607 | 754.29 | 683.22 | 707.72 | 672.66 | **2,817.89** |
| 608 | 403.21 | 455.41 | 396.43 | 385.42 | **1,640.46** |
| 609 | 1,431.09 | 1,231.51 | 1,362.16 | 1,253.34 | **5,278.10** |
| 610 | 2,281.55 | 2,206.92 | 2,232.21 | 2,195.80 | **8,916.48** |
| 611 | 2,694.52 | 2,515.84 | 2,590.33 | 2,502.49 | **10,303.18** |
| 612 | 1,219.78 | 1,172.28 | 1,186.78 | 1,339.36 | **4,918.21** |
| 613 | 905.08 | 780.99 | 892.17 | 897.72 | **3,475.95** |
| 614 | 693.46 | 658.36 | 753.00 | 873.00 | **2,977.81** |
| 615 | 1,414.70 | 1,187.43 | 1,440.27 | 1,190.17 | **5,232.57** |
| 616 | 2,952.24 | 2,583.83 | 2,577.95 | 2,527.36 | **10,641.38** |
| 617 | 1,416.18 | 1,298.98 | 1,392.11 | 1,556.38 | **5,663.65** |
| 618 | 2,049.35 | 1,934.30 | 1,934.20 | 2,053.26 | **7,971.12** |
| 619 | 686.92 | 649.26 | 607.69 | 577.58 | **2,521.45** |
| 620 | 2,418.53 | 2,383.48 | 2,561.45 | 2,537.60 | **9,901.07** |
| 622 | 2,876.78 | 2,716.67 | 2,664.77 | 2,629.86 | **10,888.08** |
| 623 | 946.88 | 881.06 | 926.49 | 979.21 | **3,733.64** |
| 624 | 822.72 | 860.78 | 842.75 | 859.13 | **3,385.38** |
| 625 | 1,445.83 | 1,428.72 | 1,157.07 | 1,346.76 | **5,378.39** |
| 626 | 978.87 | 1,156.15 | 1,072.54 | 1,072.16 | **4,279.73** |
| 627 | 1,298.84 | 1,175.89 | 1,352.56 | 1,364.65 | **5,191.94** |
| 628 | 1,491.26 | 1,493.99 | 1,553.82 | 1,622.64 | **6,161.71** |
| 629 | 2,849.55 | 2,678.61 | 2,744.28 | 3,095.15 | **11,367.59** |
| **IL - ILLINOIS - Total** | **78,578.71** | **74,852.06** | **73,522.26** | **75,670.94** | **302,623.97** |

**STATE: IN - INDIANA**

| REGISTRANT ZIP CODE 3 | 1ST QUARTER | 2ND QUARTER | 3RD QUARTER | 4TH QUARTER | TOTAL GRAMS |
|---|---|---|---|---|---|
| 460 | 4,247.29 | 4,375.63 | 4,116.65 | 4,352.70 | **17,092.27** |
| 461 | 5,874.66 | 5,479.67 | 5,715.37 | 5,835.24 | **22,904.94** |
| 462 | 10,308.93 | 9,610.59 | 9,468.46 | 9,217.23 | **38,605.21** |
| 463 | 5,184.45 | 4,967.53 | 5,091.24 | 4,599.88 | **19,843.10** |
| 464 | 432.29 | 407.89 | 467.07 | 378.07 | **1,685.33** |
| 465 | 3,760.86 | 3,569.07 | 3,598.79 | 3,643.80 | **14,572.52** |
| 466 | 1,223.07 | 1,218.31 | 1,082.54 | 1,283.54 | **4,807.46** |
| 467 | 2,355.74 | 2,201.40 | 2,214.42 | 2,205.65 | **8,977.21** |
| 468 | 3,322.12 | 3,496.19 | 3,381.84 | 3,380.52 | **13,580.66** |
| 469 | 3,324.75 | 3,194.25 | 3,036.11 | 3,054.75 | **12,609.86** |
| 470 | 1,049.82 | 1,065.46 | 1,036.99 | 1,040.69 | **4,192.97** |
| 471 | 2,665.99 | 2,686.31 | 2,659.33 | 2,621.40 | **10,633.04** |
| 472 | 1,471.12 | 1,540.91 | 1,481.43 | 1,585.28 | **6,078.75** |
| 473 | 4,166.05 | 4,026.71 | 3,900.61 | 3,989.33 | **16,082.70** |
| 474 | 1,707.20 | 1,630.62 | 1,774.28 | 1,887.12 | **6,999.21** |
| 475 | 1,328.31 | 1,312.06 | 1,272.33 | 1,323.91 | **5,236.61** |
| 476 | 960.73 | 935.02 | 756.72 | 864.29 | **3,516.76** |
| 477 | 2,719.14 | 2,324.95 | 2,531.11 | 2,237.69 | **9,812.90** |
| 478 | 1,292.43 | 1,123.70 | 1,011.61 | 1,080.91 | **4,508.65** |
| 479 | 1,692.61 | 1,427.27 | 1,497.98 | 1,571.16 | **6,189.02** |
| **IN - INDIANA - Total** | **59,087.56** | **56,593.54** | **56,094.89** | **56,153.17** | **227,929.16** |

**STATE: KS - KANSAS**

| REGISTRANT ZIP CODE 3 | 1ST QUARTER | 2ND QUARTER | 3RD QUARTER | 4TH QUARTER | TOTAL GRAMS |
|---|---|---|---|---|---|
| 660 | 4,325.69 | 4,075.11 | 4,335.71 | 4,301.22 | **17,037.72** |
| 661 | 973.62 | 946.39 | 879.02 | 922.01 | **3,721.05** |
| 662 | 5,034.07 | 4,754.06 | 4,736.10 | 5,200.11 | **19,724.34** |
| 664 | 494.29 | 426.32 | 512.18 | 428.96 | **1,861.75** |
| 665 | 959.61 | 913.25 | 882.25 | 924.03 | **3,679.15** |
| 666 | 2,324.57 | 2,341.15 | 2,332.10 | 2,578.92 | **9,576.74** |
| 667 | 1,709.31 | 1,603.11 | 1,548.61 | 1,625.56 | **6,486.58** |
| 668 | 550.04 | 542.82 | 589.66 | 539.40 | **2,221.93** |

| | | | | | |
|---|---|---|---|---|---|
| 669 | 467.00 | 394.36 | 410.53 | 422.08 | **1,693.97** |
| 670 | 1,639.27 | 1,663.57 | 1,594.37 | 1,660.50 | **6,557.71** |
| 671 | 1,014.14 | 930.84 | 893.49 | 1,039.75 | **3,878.22** |
| 672 | 5,279.97 | 5,207.68 | 5,091.92 | 5,263.68 | **20,843.25** |
| 673 | 1,075.28 | 992.01 | 927.93 | 976.17 | **3,971.39** |
| 674 | 1,717.00 | 1,557.55 | 1,595.99 | 1,641.08 | **6,511.63** |
| 675 | 1,082.56 | 1,097.69 | 1,018.99 | 1,085.94 | **4,285.18** |
| 676 | 704.82 | 710.12 | 681.95 | 671.55 | **2,768.45** |
| 677 | 420.99 | 443.33 | 430.63 | 409.65 | **1,704.60** |
| 678 | 731.40 | 661.83 | 720.98 | 694.92 | **2,809.13** |
| 679 | 296.81 | 254.17 | 280.47 | 300.27 | **1,131.72** |
| **KS - KANSAS - Total** | **30,800.45** | **29,515.34** | **29,462.90** | **30,685.81** | **120,464.50** |

**STATE: KY - KENTUCKY**

| REGISTRANT ZIP CODE 3 | 1ST QUARTER | 2ND QUARTER | 3RD QUARTER | 4TH QUARTER | TOTAL GRAMS |
|---|---|---|---|---|---|
| 400 | 1,069.75 | 1,094.87 | 1,047.16 | 1,078.30 | **4,290.07** |
| 401 | 843.22 | 709.87 | 764.03 | 690.09 | **3,007.21** |
| 402 | 5,030.07 | 4,565.91 | 5,084.96 | 5,055.15 | **19,736.09** |
| 403 | 2,060.46 | 1,979.63 | 1,997.39 | 1,952.35 | **7,989.83** |
| 404 | 933.49 | 711.49 | 823.69 | 870.04 | **3,338.71** |
| 405 | 2,634.28 | 2,688.32 | 2,314.53 | 2,342.01 | **9,979.14** |
| 406 | 272.86 | 231.44 | 268.38 | 240.56 | **1,013.24** |
| 407 | 1,093.69 | 824.11 | 855.60 | 807.95 | **3,581.35** |
| 408 | 274.86 | 241.38 | 187.36 | 231.09 | **934.69** |
| 409 | 451.86 | 466.80 | 453.49 | 450.49 | **1,822.65** |
| 410 | 3,918.25 | 3,776.58 | 3,725.59 | 3,843.68 | **15,264.09** |
| 411 | 591.47 | 538.37 | 537.75 | 632.64 | **2,300.23** |
| 412 | 197.67 | 185.78 | 191.80 | 208.02 | **783.27** |
| 413 | 250.48 | 206.60 | 230.98 | 214.43 | **902.47** |
| 414 | 162.02 | 150.25 | 152.39 | 166.23 | **630.89** |
| 415 | 339.75 | 320.64 | 298.06 | 251.63 | **1,210.07** |
| 416 | 373.76 | 294.93 | 296.33 | 259.32 | **1,224.34** |
| 417 | 852.95 | 935.01 | 915.45 | 849.35 | **3,552.77** |
| 418 | 164.42 | 106.48 | 128.22 | 108.95 | **508.07** |
| 420 | 2,155.56 | 2,183.38 | 2,047.30 | 1,793.33 | **8,179.58** |
| 421 | 1,975.25 | 1,877.46 | 1,762.86 | 1,780.80 | **7,396.36** |
| 422 | 950.65 | 914.67 | 954.57 | 865.18 | **3,685.07** |
| 423 | 1,364.97 | 1,170.12 | 1,242.65 | 1,228.29 | **5,006.02** |
| 424 | 1,334.33 | 1,107.38 | 1,240.83 | 1,254.21 | **4,936.75** |
| 425 | 498.23 | 555.15 | 466.17 | 436.62 | **1,956.17** |
| 426 | 418.58 | 422.29 | 410.59 | 372.14 | **1,623.60** |
| 427 | 1,560.24 | 1,554.61 | 1,522.23 | 1,582.84 | **6,219.91** |
| **KY - KENTUCKY - Total** | **31,773.11** | **29,813.49** | **29,920.36** | **29,565.68** | **121,072.64** |

**STATE: LA - LOUISIANA**

| REGISTRANT ZIP CODE 3 | 1ST QUARTER | 2ND QUARTER | 3RD QUARTER | 4TH QUARTER | TOTAL GRAMS |
|---|---|---|---|---|---|
| 700 | 3,766.60 | 3,243.54 | 3,495.59 | 3,269.45 | **13,775.18** |
| 701 | 3,560.17 | 2,665.73 | 3,490.13 | 2,612.94 | **12,328.97** |
| 703 | 1,225.66 | 1,058.82 | 1,107.62 | 1,055.08 | **4,447.18** |
| 704 | 3,916.39 | 4,120.88 | 3,989.41 | 3,964.44 | **15,991.12** |
| 705 | 4,316.90 | 4,116.61 | 4,424.76 | 3,911.37 | **16,769.64** |
| 706 | 1,809.32 | 1,480.43 | 1,427.85 | 1,466.72 | **6,184.32** |
| 707 | 4,139.14 | 3,759.77 | 3,921.27 | 3,686.82 | **15,507.01** |
| 708 | 3,107.35 | 2,913.62 | 2,974.42 | 2,783.51 | **11,778.91** |
| 710 | 1,282.20 | 1,279.47 | 1,191.76 | 1,212.85 | **4,966.29** |
| 711 | 5,110.74 | 4,565.76 | 4,867.79 | 4,588.79 | **19,133.07** |
| 712 | 1,840.32 | 2,050.26 | 1,833.34 | 1,632.73 | **7,356.65** |
| 713 | 2,866.97 | 2,757.93 | 2,596.74 | 2,825.97 | **11,047.60** |
| 714 | 1,600.94 | 1,601.30 | 1,630.28 | 1,606.46 | **6,438.98** |
| **LA - LOUISIANA - Total** | **38,542.70** | **35,614.13** | **36,950.95** | **34,617.14** | **145,724.93** |

**STATE: MA - MASSACHUSETTS**

| REGISTRANT ZIP CODE 3 | 1ST QUARTER | 2ND QUARTER | 3RD QUARTER | 4TH QUARTER | TOTAL GRAMS |
|---|---|---|---|---|---|

| | | | | | |
|---|---|---|---|---|---|
| 010 | 5,529.55 | 5,429.79 | 5,185.16 | 5,102.46 | **21,246.95** |
| 011 | 2,247.83 | 2,106.03 | 2,190.57 | 2,276.99 | **8,821.42** |
| 012 | 887.08 | 925.48 | 874.92 | 928.90 | **3,616.39** |
| 013 | 1,083.49 | 1,044.36 | 1,038.77 | 1,058.55 | **4,225.16** |
| 014 | 1,793.52 | 1,680.44 | 1,627.12 | 1,653.49 | **6,754.58** |
| 015 | 2,342.97 | 2,142.54 | 2,040.01 | 2,019.20 | **8,544.72** |
| 016 | 1,492.70 | 1,453.25 | 1,382.76 | 1,490.32 | **5,819.04** |
| 017 | 3,178.65 | 3,248.14 | 2,555.07 | 2,993.77 | **11,975.63** |
| 018 | 6,200.11 | 6,382.88 | 7,441.99 | 10,247.02 | **30,272.00** |
| 019 | 2,417.42 | 2,465.60 | 2,175.97 | 2,278.10 | **9,337.09** |
| 020 | 1,759.89 | 1,927.36 | 1,771.11 | 1,772.92 | **7,231.28** |
| 021 | 5,645.34 | 5,005.86 | 5,001.83 | 4,961.25 | **20,614.27** |
| 022 | 701.13 | 828.97 | 740.13 | 537.43 | **2,807.64** |
| 023 | 2,941.46 | 3,128.44 | 2,861.81 | 3,059.08 | **11,990.78** |
| 024 | 747.41 | 821.90 | 777.47 | 873.63 | **3,220.41** |
| 025 | 931.55 | 1,101.48 | 913.63 | 996.04 | **3,942.69** |
| 026 | 1,332.26 | 1,310.64 | 1,410.86 | 1,300.86 | **5,354.62** |
| 027 | 6,945.19 | 6,941.23 | 6,870.88 | 6,403.59 | **27,160.89** |
| **MA - MASSACHUSETTS - Total** | **48,177.56** | **47,944.37** | **46,860.05** | **49,953.59** | **192,935.58** |

**STATE: MD - MARYLAND**

| REGISTRANT ZIP CODE 3 | 1ST QUARTER | 2ND QUARTER | 3RD QUARTER | 4TH QUARTER | TOTAL GRAMS |
|---|---|---|---|---|---|
| 206 | 3,033.39 | 3,049.10 | 3,026.94 | 2,968.01 | **12,077.45** |
| 207 | 3,822.47 | 3,499.08 | 3,422.54 | 3,431.37 | **14,175.45** |
| 208 | 3,616.68 | 4,369.89 | 3,646.22 | 3,236.37 | **14,869.16** |
| 209 | 687.55 | 772.48 | 653.85 | 611.22 | **2,725.09** |
| 210 | 8,910.58 | 7,986.34 | 8,233.47 | 8,005.99 | **33,136.38** |
| 211 | 3,431.85 | 3,603.19 | 3,528.18 | 3,639.55 | **14,202.78** |
| 212 | 11,358.81 | 10,562.84 | 10,680.34 | 10,649.20 | **43,251.19** |
| 214 | 817.32 | 686.74 | 767.40 | 689.89 | **2,961.36** |
| 215 | 1,349.07 | 1,276.82 | 1,215.31 | 1,189.30 | **5,030.49** |
| 216 | 1,891.31 | 1,938.31 | 1,960.64 | 1,898.33 | **7,688.59** |
| 217 | 5,562.99 | 5,920.85 | 5,213.78 | 5,052.21 | **21,749.83** |
| 218 | 1,765.54 | 1,534.74 | 1,659.71 | 1,701.18 | **6,661.18** |
| 219 | 1,785.92 | 1,784.85 | 1,799.02 | 1,566.45 | **6,936.25** |
| **MD - MARYLAND - Total** | **48,033.49** | **46,985.23** | **45,807.40** | **44,639.08** | **185,465.20** |

**STATE: ME - MAINE**

| REGISTRANT ZIP CODE 3 | 1ST QUARTER | 2ND QUARTER | 3RD QUARTER | 4TH QUARTER | TOTAL GRAMS |
|---|---|---|---|---|---|
| 039 | 198.32 | 220.21 | 178.92 | 188.61 | **786.06** |
| 040 | 2,694.29 | 2,720.11 | 2,880.75 | 3,017.68 | **11,312.83** |
| 041 | 887.84 | 996.28 | 855.96 | 1,020.50 | **3,760.58** |
| 042 | 1,215.86 | 1,259.61 | 1,224.60 | 1,235.38 | **4,935.46** |
| 043 | 1,850.50 | 1,818.70 | 1,839.13 | 1,888.56 | **7,396.89** |
| 044 | 1,070.47 | 1,030.73 | 1,102.52 | 1,089.04 | **4,292.75** |
| 045 | 343.86 | 373.36 | 416.24 | 354.48 | **1,487.94** |
| 046 | 512.99 | 535.29 | 555.23 | 528.58 | **2,132.09** |
| 047 | 568.84 | 489.38 | 501.23 | 516.51 | **2,075.96** |
| 048 | 335.05 | 445.00 | 384.18 | 384.24 | **1,548.46** |
| 049 | 1,983.49 | 1,992.63 | 1,986.40 | 1,918.72 | **7,881.24** |
| **ME - MAINE - Total** | **11,661.50** | **11,881.31** | **11,925.17** | **12,142.30** | **47,610.28** |

**STATE: MI - MICHIGAN**

| REGISTRANT ZIP CODE 3 | 1ST QUARTER | 2ND QUARTER | 3RD QUARTER | 4TH QUARTER | TOTAL GRAMS |
|---|---|---|---|---|---|
| 480 | 14,454.22 | 13,830.96 | 13,739.59 | 13,539.42 | **55,564.20** |
| 481 | 15,071.99 | 13,877.21 | 13,790.07 | 14,561.78 | **57,301.06** |
| 482 | 6,956.36 | 7,683.81 | 7,827.23 | 7,153.24 | **29,620.63** |
| 483 | 9,901.14 | 10,022.12 | 9,845.26 | 9,969.11 | **39,737.63** |
| 484 | 5,014.08 | 4,699.02 | 4,733.21 | 4,446.33 | **18,892.63** |
| 485 | 2,267.75 | 2,093.93 | 1,989.73 | 2,026.84 | **8,378.25** |
| 486 | 7,847.88 | 7,397.33 | 7,657.24 | 7,389.97 | **30,292.42** |
| 487 | 3,726.33 | 3,672.49 | 3,686.86 | 3,549.33 | **14,635.01** |
| 488 | 6,875.14 | 6,405.56 | 6,272.94 | 6,333.74 | **25,887.37** |

| | | | | | |
|---|---|---|---|---|---|
| 489 | 2,858.85 | 3,047.30 | 2,785.77 | 2,956.59 | **11,648.51** |
| 490 | 10,340.41 | 9,608.99 | 9,646.29 | 9,871.41 | **39,467.10** |
| 491 | 1,245.16 | 1,276.93 | 1,311.30 | 1,275.09 | **5,108.49** |
| 492 | 3,644.81 | 3,388.16 | 3,254.15 | 3,214.10 | **13,501.21** |
| 493 | 2,525.31 | 2,352.80 | 2,457.79 | 2,547.83 | **9,883.73** |
| 494 | 6,272.59 | 6,014.64 | 5,825.84 | 6,006.74 | **24,119.80** |
| 495 | 4,853.55 | 4,563.60 | 4,567.05 | 5,056.65 | **19,040.85** |
| 496 | 5,773.95 | 5,150.03 | 5,313.81 | 5,132.20 | **21,369.99** |
| 497 | 4,691.13 | 4,675.70 | 5,034.49 | 4,789.45 | **19,190.77** |
| 498 | 2,491.23 | 2,382.26 | 2,433.91 | 2,433.84 | **9,741.24** |
| 499 | 778.72 | 756.34 | 729.04 | 708.93 | **2,973.04** |
| **MI - MICHIGAN - Total** | **117,590.60** | **112,899.18** | **112,901.55** | **112,962.60** | **456,353.92** |

**STATE: MN - MINNESOTA**

| REGISTRANT ZIP CODE 3 | 1ST QUARTER | 2ND QUARTER | 3RD QUARTER | 4TH QUARTER | TOTAL GRAMS |
|---|---|---|---|---|---|
| 550 | 3,359.37 | 3,339.04 | 3,311.41 | 3,323.08 | **13,332.91** |
| 551 | 4,343.59 | 5,562.05 | 3,300.69 | 4,057.67 | **17,263.99** |
| 553 | 3,753.74 | 3,943.26 | 3,832.43 | 3,863.10 | **15,392.52** |
| 554 | 6,912.18 | 6,891.66 | 6,806.73 | 7,157.49 | **27,768.06** |
| 556 | 124.79 | 105.13 | 124.23 | 129.15 | **483.31** |
| 557 | 1,127.89 | 1,168.28 | 1,227.94 | 1,222.17 | **4,746.28** |
| 558 | 813.84 | 871.45 | 918.08 | 910.85 | **3,514.22** |
| 559 | 1,464.46 | 1,085.77 | 1,210.98 | 1,372.29 | **5,133.51** |
| 560 | 1,036.70 | 1,009.73 | 974.18 | 1,101.81 | **4,122.42** |
| 561 | 551.00 | 447.52 | 414.73 | 530.28 | **1,943.52** |
| 562 | 652.21 | 640.24 | 575.33 | 718.21 | **2,585.99** |
| 563 | 2,586.81 | 2,545.51 | 2,471.45 | 2,722.27 | **10,326.05** |
| 564 | 1,067.84 | 921.47 | 950.82 | 1,101.74 | **4,041.86** |
| 565 | 859.79 | 865.94 | 852.83 | 860.47 | **3,439.03** |
| 566 | 542.09 | 458.38 | 546.98 | 488.60 | **2,036.06** |
| 567 | 442.49 | 381.07 | 378.47 | 432.63 | **1,634.67** |
| **MN - MINNESOTA - Total** | **29,638.80** | **30,236.52** | **27,897.28** | **29,991.80** | **117,764.39** |

**STATE: MO - MISSOURI**

| REGISTRANT ZIP CODE 3 | 1ST QUARTER | 2ND QUARTER | 3RD QUARTER | 4TH QUARTER | TOTAL GRAMS |
|---|---|---|---|---|---|
| 630 | 6,136.32 | 5,967.89 | 5,831.96 | 5,847.99 | **23,784.15** |
| 631 | 10,626.57 | 10,993.48 | 10,386.06 | 10,526.51 | **42,532.62** |
| 633 | 3,858.36 | 3,861.00 | 3,875.00 | 3,644.89 | **15,239.25** |
| 634 | 481.69 | 455.40 | 509.03 | 498.09 | **1,944.21** |
| 635 | 891.29 | 844.24 | 894.24 | 911.35 | **3,541.11** |
| 636 | 1,630.67 | 1,560.46 | 1,622.83 | 1,590.30 | **6,404.26** |
| 637 | 1,003.82 | 894.06 | 928.11 | 995.01 | **3,821.00** |
| 638 | 1,026.67 | 975.68 | 994.94 | 955.45 | **3,952.74** |
| 639 | 840.55 | 841.24 | 921.03 | 825.05 | **3,427.87** |
| 640 | 5,394.04 | 5,305.74 | 5,227.60 | 5,480.15 | **21,407.54** |
| 641 | 3,856.61 | 3,401.53 | 3,532.99 | 3,503.81 | **14,294.94** |
| 644 | 543.85 | 409.69 | 517.96 | 504.88 | **1,976.39** |
| 645 | 995.85 | 911.23 | 989.44 | 918.39 | **3,814.92** |
| 646 | 498.28 | 534.38 | 512.50 | 530.64 | **2,075.79** |
| 647 | 1,408.41 | 1,371.54 | 1,298.14 | 1,434.18 | **5,512.27** |
| 648 | 2,220.33 | 2,116.93 | 2,085.46 | 2,078.87 | **8,501.59** |
| 650 | 1,163.08 | 1,221.63 | 1,114.73 | 1,119.43 | **4,618.87** |
| 651 | 798.75 | 753.34 | 819.51 | 902.91 | **3,274.52** |
| 652 | 3,135.14 | 3,168.73 | 2,907.13 | 3,307.10 | **12,518.10** |
| 653 | 602.70 | 603.43 | 605.06 | 564.63 | **2,375.82** |
| 654 | 1,014.67 | 913.48 | 1,073.75 | 985.71 | **3,987.61** |
| 655 | 1,423.14 | 1,453.87 | 1,664.38 | 1,609.65 | **6,151.04** |
| 656 | 2,890.85 | 2,847.99 | 2,872.25 | 2,907.78 | **11,518.88** |
| 657 | 4,945.09 | 5,571.84 | 5,096.06 | 5,006.69 | **20,619.68** |
| 658 | 4,513.40 | 4,351.39 | 4,161.46 | 4,335.27 | **17,361.52** |
| **MO - MISSOURI - Total** | **61,900.12** | **61,330.21** | **60,441.64** | **60,984.73** | **244,656.70** |

**STATE: MS - MISSISSIPPI**

| REGISTRANT ZIP CODE 3 | 1ST QUARTER | 2ND QUARTER | 3RD QUARTER | 4TH QUARTER | TOTAL GRAMS |
|---|---|---|---|---|---|
| 386 | 2,883.86 | 2,638.36 | 2,520.83 | 2,675.35 | **10,718.40** |
| 387 | 273.83 | 334.48 | 340.50 | 290.26 | **1,239.07** |
| 388 | 2,574.47 | 2,493.66 | 2,401.16 | 2,336.83 | **9,806.13** |
| 389 | 448.90 | 446.43 | 444.50 | 480.39 | **1,820.22** |
| 390 | 1,534.72 | 1,607.79 | 1,653.53 | 1,459.01 | **6,255.05** |
| 391 | 3,746.64 | 5,450.67 | 4,363.69 | 6,656.52 | **20,217.51** |
| 392 | 2,332.82 | 2,112.72 | 2,151.93 | 2,081.86 | **8,679.33** |
| 393 | 1,888.42 | 1,898.49 | 1,821.38 | 1,766.96 | **7,375.24** |
| 394 | 3,609.08 | 3,344.57 | 3,551.15 | 3,821.34 | **14,326.14** |
| 395 | 3,418.60 | 3,005.11 | 2,983.43 | 2,923.07 | **12,330.21** |
| 396 | 1,303.95 | 1,223.91 | 1,122.28 | 982.09 | **4,632.22** |
| 397 | 743.44 | 697.55 | 693.28 | 595.72 | **2,729.98** |
| **MS - MISSISSIPPI - Total** | **24,758.71** | **25,253.74** | **24,047.66** | **26,069.39** | **100,129.51** |

**STATE: MT - MONTANA**

| REGISTRANT ZIP CODE 3 | 1ST QUARTER | 2ND QUARTER | 3RD QUARTER | 4TH QUARTER | TOTAL GRAMS |
|---|---|---|---|---|---|
| 590 | 719.80 | 790.26 | 675.90 | 713.12 | **2,899.08** |
| 591 | 2,305.36 | 2,082.76 | 2,386.35 | 2,201.11 | **8,975.57** |
| 592 | 462.69 | 399.91 | 368.79 | 382.21 | **1,613.60** |
| 593 | 345.07 | 326.21 | 401.16 | 410.82 | **1,483.26** |
| 594 | 1,329.38 | 1,326.93 | 1,417.45 | 1,163.40 | **5,237.16** |
| 595 | 234.44 | 225.08 | 273.26 | 237.48 | **970.26** |
| 596 | 1,151.12 | 1,128.40 | 1,091.63 | 1,122.95 | **4,494.10** |
| 597 | 1,498.71 | 1,485.04 | 1,388.98 | 1,531.47 | **5,904.20** |
| 598 | 1,965.41 | 1,881.22 | 1,898.65 | 1,804.19 | **7,549.46** |
| 599 | 1,793.38 | 1,838.09 | 1,776.98 | 1,768.40 | **7,176.86** |
| **MT - MONTANA - Total** | **11,805.37** | **11,483.89** | **11,679.15** | **11,335.15** | **46,303.56** |

**STATE: NC - NORTH CAROLINA**

| REGISTRANT ZIP CODE 3 | 1ST QUARTER | 2ND QUARTER | 3RD QUARTER | 4TH QUARTER | TOTAL GRAMS |
|---|---|---|---|---|---|
| 270 | 2,811.68 | 2,647.76 | 2,471.31 | 2,634.22 | **10,564.98** |
| 271 | 2,520.90 | 2,235.35 | 2,333.00 | 2,291.73 | **9,380.98** |
| 272 | 5,236.10 | 5,055.71 | 5,135.55 | 4,924.97 | **20,352.34** |
| 273 | 3,462.61 | 3,405.12 | 3,453.69 | 3,205.26 | **13,526.68** |
| 274 | 2,414.03 | 2,327.84 | 2,323.22 | 2,332.38 | **9,397.47** |
| 275 | 9,146.41 | 8,983.52 | 9,217.63 | 9,194.72 | **36,542.29** |
| 276 | 3,210.01 | 2,985.52 | 2,782.70 | 2,869.99 | **11,848.23** |
| 277 | 4,202.41 | 2,930.20 | 2,941.66 | 3,473.75 | **13,548.02** |
| 278 | 5,873.80 | 5,540.60 | 5,654.17 | 5,498.56 | **22,567.13** |
| 279 | 1,917.58 | 1,828.12 | 1,987.65 | 2,007.28 | **7,740.63** |
| 280 | 6,778.56 | 6,533.43 | 6,644.94 | 6,306.52 | **26,263.45** |
| 281 | 5,836.37 | 5,487.45 | 5,485.37 | 5,881.81 | **22,691.00** |
| 282 | 4,520.25 | 4,448.61 | 4,257.81 | 4,132.62 | **17,359.29** |
| 283 | 6,520.25 | 6,282.63 | 6,579.41 | 6,026.54 | **25,408.83** |
| 284 | 5,088.53 | 4,900.20 | 5,058.53 | 4,732.32 | **19,779.58** |
| 285 | 5,621.39 | 5,643.61 | 5,799.90 | 5,508.61 | **22,573.50** |
| 286 | 8,780.70 | 8,694.12 | 8,325.84 | 8,098.12 | **33,898.79** |
| 287 | 5,250.10 | 5,373.08 | 5,483.56 | 5,692.19 | **21,798.93** |
| 288 | 2,673.79 | 2,664.81 | 2,230.93 | 2,377.75 | **9,947.28** |
| 289 | 724.66 | 684.77 | 706.28 | 636.64 | **2,752.36** |
| **NC - NORTH CAROLINA - Total** | **92,590.15** | **88,652.46** | **88,873.16** | **87,825.97** | **357,941.74** |

**STATE: ND - NORTH DAKOTA**

| REGISTRANT ZIP CODE 3 | 1ST QUARTER | 2ND QUARTER | 3RD QUARTER | 4TH QUARTER | TOTAL GRAMS |
|---|---|---|---|---|---|
| 580 | 231.64 | 270.25 | 231.04 | 259.93 | **992.87** |
| 581 | 977.00 | 1,031.85 | 923.96 | 1,134.24 | **4,067.07** |
| 582 | 706.60 | 631.07 | 503.67 | 702.71 | **2,544.05** |
| 583 | 326.91 | 342.93 | 376.58 | 384.40 | **1,430.83** |
| 584 | 226.54 | 194.45 | 211.57 | 218.71 | **851.27** |
| 585 | 1,266.44 | 1,109.65 | 1,056.32 | 1,120.48 | **4,552.89** |
| 586 | 235.26 | 204.07 | 218.55 | 239.81 | **897.68** |
| 587 | 323.17 | 257.04 | 199.49 | 317.48 | **1,097.18** |

| | | | | | |
|---|---|---|---|---|---|
| 588 | 212.20 | 186.01 | 187.20 | 194.25 | **779.67** |
| **ND - NORTH DAKOTA - Total** | **4,505.77** | **4,227.34** | **3,908.38** | **4,572.03** | **17,213.51** |

**STATE: NE - NEBRASKA**

| REGISTRANT ZIP CODE 3 | 1ST QUARTER | 2ND QUARTER | 3RD QUARTER | 4TH QUARTER | TOTAL GRAMS |
|---|---|---|---|---|---|
| 680 | 2,171.80 | 2,045.63 | 1,865.59 | 1,939.56 | **8,022.59** |
| 681 | 4,509.28 | 4,334.20 | 4,616.05 | 4,523.89 | **17,983.42** |
| 683 | 474.29 | 498.29 | 498.63 | 493.06 | **1,964.27** |
| 684 | 457.27 | 439.45 | 419.34 | 496.15 | **1,812.21** |
| 685 | 1,860.79 | 2,161.02 | 2,032.53 | 2,190.79 | **8,245.13** |
| 686 | 283.90 | 265.39 | 256.79 | 323.33 | **1,129.41** |
| 687 | 854.33 | 820.07 | 824.57 | 811.92 | **3,310.89** |
| 688 | 1,484.76 | 1,259.51 | 1,344.49 | 1,469.74 | **5,558.50** |
| 689 | 559.67 | 499.67 | 540.42 | 565.33 | **2,165.08** |
| 690 | 269.39 | 269.29 | 247.32 | 286.06 | **1,072.06** |
| 691 | 421.38 | 383.61 | 405.25 | 435.06 | **1,645.30** |
| 692 | 23.76 | 23.61 | 35.20 | 31.62 | **114.19** |
| 693 | 634.84 | 565.25 | 604.78 | 593.98 | **2,398.86** |
| **NE - NEBRASKA - Total** | **14,005.46** | **13,564.99** | **13,690.96** | **14,160.51** | **55,421.92** |

**STATE: NH - NEW HAMPSHIRE**

| REGISTRANT ZIP CODE 3 | 1ST QUARTER | 2ND QUARTER | 3RD QUARTER | 4TH QUARTER | TOTAL GRAMS |
|---|---|---|---|---|---|
| 030 | 3,433.55 | 3,261.40 | 3,161.04 | 3,211.00 | **13,066.99** |
| 031 | 1,941.26 | 1,859.21 | 1,868.54 | 2,025.03 | **7,694.04** |
| 032 | 1,711.38 | 1,697.67 | 1,687.10 | 1,682.15 | **6,778.30** |
| 033 | 1,421.56 | 1,605.30 | 1,351.90 | 1,519.77 | **5,898.53** |
| 034 | 626.15 | 550.66 | 683.58 | 647.27 | **2,507.66** |
| 035 | 452.05 | 386.07 | 387.48 | 373.12 | **1,598.73** |
| 036 | 77.83 | 84.60 | 93.62 | 67.68 | **323.74** |
| 037 | 605.18 | 621.98 | 652.38 | 696.25 | **2,575.78** |
| 038 | 3,156.47 | 3,184.98 | 3,108.86 | 3,224.26 | **12,674.56** |
| **NH - NEW HAMPSHIRE - Total** | **13,425.45** | **13,251.87** | **12,994.49** | **13,446.52** | **53,118.33** |

**STATE: NJ - NEW JERSEY**

| REGISTRANT ZIP CODE 3 | 1ST QUARTER | 2ND QUARTER | 3RD QUARTER | 4TH QUARTER | TOTAL GRAMS |
|---|---|---|---|---|---|
| 070 | 7,720.39 | 7,636.49 | 7,119.81 | 7,152.49 | **29,629.18** |
| 071 | 894.87 | 1,006.60 | 1,026.28 | 827.66 | **3,755.41** |
| 072 | 476.58 | 387.03 | 445.54 | 442.78 | **1,751.92** |
| 073 | 582.76 | 459.19 | 494.09 | 489.19 | **2,025.22** |
| 074 | 4,063.57 | 3,639.52 | 3,683.92 | 3,408.76 | **14,795.77** |
| 075 | 594.32 | 443.30 | 536.82 | 578.74 | **2,153.18** |
| 076 | 4,667.22 | 4,245.89 | 3,926.33 | 4,178.25 | **17,017.69** |
| 077 | 4,253.52 | 4,174.27 | 4,042.09 | 3,971.80 | **16,441.67** |
| 078 | 2,042.68 | 2,067.56 | 1,936.33 | 2,203.38 | **8,249.94** |
| 079 | 1,749.46 | 1,515.29 | 1,887.66 | 1,559.35 | **6,711.77** |
| 080 | 12,872.48 | 12,167.69 | 12,827.81 | 12,732.44 | **50,600.42** |
| 081 | 1,060.54 | 1,028.03 | 1,076.51 | 1,142.77 | **4,307.84** |
| 082 | 1,775.42 | 1,868.28 | 1,900.30 | 1,909.15 | **7,453.15** |
| 083 | 1,443.60 | 1,261.95 | 1,366.15 | 1,318.51 | **5,390.21** |
| 084 | 351.79 | 320.72 | 347.25 | 361.58 | **1,381.34** |
| 085 | 1,705.35 | 1,750.74 | 1,560.34 | 1,679.72 | **6,696.15** |
| 086 | 1,394.21 | 1,366.60 | 1,232.07 | 1,136.04 | **5,128.92** |
| 087 | 4,108.52 | 3,807.27 | 3,805.23 | 3,635.89 | **15,356.92** |
| 088 | 6,971.45 | 6,595.00 | 6,283.25 | 6,144.51 | **25,994.22** |
| 089 | 540.59 | 593.95 | 638.36 | 716.95 | **2,489.85** |
| **NJ - NEW JERSEY - Total** | **59,269.30** | **56,335.36** | **56,136.15** | **55,589.96** | **227,330.77** |

**STATE: NM - NEW MEXICO**

| REGISTRANT ZIP CODE 3 | 1ST QUARTER | 2ND QUARTER | 3RD QUARTER | 4TH QUARTER | TOTAL GRAMS |
|---|---|---|---|---|---|
| 870 | 1,230.54 | 1,109.08 | 1,239.98 | 1,174.33 | **4,753.92** |
| 871 | 8,153.80 | 7,599.15 | 7,806.65 | 7,799.98 | **31,359.58** |
| 873 | 739.88 | 670.75 | 676.06 | 762.42 | **2,849.12** |
| 874 | 682.64 | 735.90 | 714.55 | 790.73 | **2,923.82** |
| 875 | 1,883.84 | 1,783.52 | 1,876.16 | 1,974.45 | **7,517.97** |

| | | | | | |
|---|---|---|---|---|---|
| 877 | 304.63 | 282.53 | 280.24 | 295.71 | **1,163.12** |
| 878 | 62.08 | 57.23 | 84.19 | 88.21 | **291.70** |
| 879 | 154.24 | 145.81 | 175.89 | 110.36 | **586.30** |
| 880 | 1,741.54 | 1,701.21 | 1,705.03 | 1,568.60 | **6,716.37** |
| 881 | 394.46 | 362.42 | 407.03 | 421.27 | **1,585.18** |
| 882 | 1,874.84 | 1,796.16 | 1,757.55 | 1,862.41 | **7,290.97** |
| 883 | 560.55 | 466.11 | 437.67 | 468.25 | **1,932.58** |
| 884 | 35.84 | 39.05 | 38.16 | 39.91 | **152.96** |
| **NM - NEW MEXICO - Total** | **17,818.87** | **16,748.92** | **17,199.16** | **17,356.64** | **69,123.58** |

**STATE: NV - NEVADA**

| REGISTRANT ZIP CODE 3 | 1ST QUARTER | 2ND QUARTER | 3RD QUARTER | 4TH QUARTER | TOTAL GRAMS |
|---|---|---|---|---|---|
| 890 | 9,335.08 | 8,681.26 | 8,929.08 | 8,681.22 | **35,626.64** |
| 891 | 19,695.43 | 18,374.06 | 18,108.55 | 17,213.12 | **73,391.17** |
| 893 | 42.54 | 49.45 | 45.63 | 38.91 | **176.53** |
| 894 | 2,750.99 | 2,722.80 | 2,844.84 | 2,576.13 | **10,894.75** |
| 895 | 3,391.57 | 3,272.28 | 3,695.67 | 3,503.68 | **13,863.20** |
| 897 | 869.35 | 871.07 | 850.25 | 919.28 | **3,509.95** |
| 898 | 289.82 | 258.20 | 262.41 | 252.90 | **1,063.33** |
| **NV - NEVADA - Total** | **36,374.78** | **34,229.12** | **34,736.43** | **33,185.23** | **138,525.56** |

**STATE: NY - NEW YORK**

| REGISTRANT ZIP CODE 3 | 1ST QUARTER | 2ND QUARTER | 3RD QUARTER | 4TH QUARTER | TOTAL GRAMS |
|---|---|---|---|---|---|
| 100 | 8,415.59 | 9,049.88 | 7,864.28 | 7,646.41 | **32,976.16** |
| 101 | 165.67 | 152.35 | 101.32 | 123.65 | **543.00** |
| 102 | 1.13 | 4.51 | 0 | 1.13 | **6.77** |
| 103 | 4,423.25 | 4,382.36 | 4,174.60 | 4,250.67 | **17,230.89** |
| 104 | 3,290.41 | 3,547.15 | 3,149.60 | 3,621.04 | **13,608.19** |
| 105 | 2,045.13 | 1,952.36 | 1,958.52 | 1,901.65 | **7,857.66** |
| 106 | 337.43 | 545.78 | 471.43 | 367.77 | **1,722.42** |
| 107 | 660.14 | 792.10 | 648.18 | 729.16 | **2,829.58** |
| 108 | 192.04 | 342.70 | 177.60 | 206.05 | **918.39** |
| 109 | 2,341.68 | 2,311.24 | 2,226.06 | 2,505.57 | **9,384.55** |
| 110 | 2,823.41 | 2,225.22 | 2,787.73 | 2,079.71 | **9,916.07** |
| 111 | 512.60 | 464.65 | 379.05 | 379.31 | **1,735.61** |
| 112 | 4,794.25 | 5,109.50 | 4,849.68 | 4,762.48 | **19,515.91** |
| 113 | 2,471.19 | 2,167.25 | 1,993.81 | 2,134.28 | **8,766.53** |
| 114 | 1,027.43 | 1,010.89 | 1,030.64 | 921.95 | **3,990.90** |
| 115 | 3,228.34 | 3,129.63 | 3,018.77 | 3,047.72 | **12,424.46** |
| 116 | 267.52 | 335.50 | 329.88 | 360.35 | **1,293.24** |
| 117 | 13,019.60 | 12,231.71 | 11,921.27 | 11,835.29 | **49,007.87** |
| 118 | 429.80 | 485.50 | 443.57 | 456.21 | **1,815.08** |
| 119 | 1,544.35 | 1,573.92 | 1,560.56 | 1,552.52 | **6,231.36** |
| 120 | 3,014.14 | 2,733.64 | 2,855.95 | 2,555.38 | **11,159.11** |
| 121 | 1,126.13 | 1,209.04 | 1,198.36 | 1,214.09 | **4,747.62** |
| 122 | 1,969.69 | 1,691.72 | 1,531.73 | 1,505.80 | **6,698.94** |
| 123 | 920.05 | 851.10 | 938.43 | 874.90 | **3,584.48** |
| 124 | 1,810.04 | 1,821.87 | 1,584.40 | 1,639.23 | **6,855.54** |
| 125 | 2,582.73 | 2,663.81 | 2,697.20 | 2,605.42 | **10,549.16** |
| 126 | 866.15 | 730.44 | 750.57 | 642.35 | **2,989.51** |
| 127 | 826.16 | 751.33 | 638.89 | 611.63 | **2,828.01** |
| 128 | 2,773.69 | 2,649.18 | 2,648.75 | 2,769.83 | **10,841.45** |
| 129 | 1,766.35 | 1,754.03 | 1,618.51 | 1,704.57 | **6,843.46** |
| 130 | 3,233.48 | 3,041.39 | 3,027.17 | 3,026.03 | **12,328.07** |
| 131 | 1,178.15 | 1,153.42 | 1,190.12 | 1,234.02 | **4,755.70** |
| 132 | 2,284.35 | 2,001.21 | 2,058.06 | 2,213.21 | **8,556.83** |
| 133 | 957.20 | 948.98 | 1,002.10 | 895.24 | **3,803.52** |
| 134 | 1,222.01 | 1,382.38 | 1,243.03 | 1,467.35 | **5,314.77** |
| 135 | 518.42 | 400.98 | 458.73 | 404.90 | **1,783.03** |
| 136 | 1,888.03 | 1,635.56 | 1,900.60 | 1,895.97 | **7,320.16** |
| 137 | 598.30 | 598.54 | 503.86 | 593.60 | **2,294.31** |
| 138 | 2,366.18 | 2,333.97 | 2,202.25 | 2,015.55 | **8,917.95** |

| | | | | | |
|---|---|---|---|---|---|
| 139 | 539.89 | 499.01 | 505.15 | 574.78 | **2,118.83** |
| 140 | 3,369.49 | 3,340.76 | 3,201.11 | 3,224.87 | **13,136.23** |
| 141 | 1,702.38 | 1,541.40 | 1,583.67 | 1,484.49 | **6,311.94** |
| 142 | 4,813.00 | 4,850.52 | 4,628.60 | 4,732.62 | **19,024.74** |
| 143 | 655.19 | 582.57 | 639.23 | 590.50 | **2,467.49** |
| 144 | 2,418.64 | 2,443.14 | 2,368.93 | 2,343.36 | **9,574.07** |
| 145 | 2,054.91 | 2,076.42 | 1,918.13 | 2,067.18 | **8,116.64** |
| 146 | 4,303.19 | 4,358.28 | 4,163.54 | 4,331.08 | **17,156.09** |
| 147 | 1,439.79 | 1,405.34 | 1,471.74 | 1,491.82 | **5,808.69** |
| 148 | 3,025.99 | 3,017.92 | 2,924.22 | 2,771.04 | **11,739.17** |
| 149 | 627.41 | 669.52 | 630.10 | 691.38 | **2,618.41** |
| **NY - NEW YORK - Total** | **108,842.08** | **106,951.65** | **103,169.69** | **103,055.13** | **422,018.55** |

**STATE: OH - OHIO**

| REGISTRANT ZIP CODE 3 | 1ST QUARTER | 2ND QUARTER | 3RD QUARTER | 4TH QUARTER | TOTAL GRAMS |
|---|---|---|---|---|---|
| 430 | 5,211.60 | 5,020.51 | 4,899.59 | 5,124.01 | **20,255.70** |
| 431 | 2,770.16 | 2,777.54 | 2,758.76 | 2,724.58 | **11,031.04** |
| 432 | 6,105.39 | 5,773.67 | 5,812.97 | 5,855.71 | **23,547.75** |
| 433 | 1,078.14 | 1,077.69 | 1,052.24 | 1,035.84 | **4,243.91** |
| 434 | 779.48 | 741.09 | 724.23 | 688.21 | **2,933.01** |
| 435 | 2,353.37 | 2,155.32 | 2,344.71 | 2,022.37 | **8,875.78** |
| 436 | 3,276.44 | 3,068.51 | 3,076.19 | 3,118.08 | **12,539.21** |
| 437 | 1,220.82 | 1,253.42 | 1,330.05 | 1,435.30 | **5,239.59** |
| 438 | 239.21 | 271.55 | 278.78 | 330.54 | **1,120.08** |
| 439 | 1,345.25 | 1,442.66 | 1,568.64 | 1,550.94 | **5,907.49** |
| 440 | 3,798.80 | 3,665.21 | 3,557.59 | 3,961.63 | **14,983.23** |
| 441 | 6,465.27 | 6,116.23 | 6,195.61 | 6,475.15 | **25,252.26** |
| 442 | 3,554.99 | 3,483.39 | 3,541.93 | 3,741.85 | **14,322.16** |
| 443 | 1,751.97 | 1,752.73 | 1,809.01 | 1,931.97 | **7,245.69** |
| 444 | 2,151.71 | 2,040.81 | 2,158.76 | 1,937.48 | **8,288.76** |
| 445 | 1,924.94 | 1,839.43 | 1,806.03 | 1,815.11 | **7,385.51** |
| 446 | 2,542.71 | 2,277.23 | 2,437.79 | 2,553.46 | **9,811.18** |
| 447 | 1,944.12 | 1,964.50 | 1,909.79 | 1,935.90 | **7,754.31** |
| 448 | 1,731.62 | 1,676.29 | 1,831.76 | 1,831.58 | **7,071.26** |
| 449 | 632.99 | 716.36 | 638.31 | 836.66 | **2,824.32** |
| 450 | 5,734.08 | 5,775.68 | 6,224.02 | 6,001.49 | **23,735.26** |
| 451 | 1,946.25 | 1,854.43 | 2,045.02 | 1,983.46 | **7,829.15** |
| 452 | 5,763.89 | 5,460.54 | 5,419.20 | 6,081.75 | **22,725.38** |
| 453 | 3,839.82 | 3,782.76 | 3,627.65 | 3,880.20 | **15,130.43** |
| 454 | 3,257.13 | 3,365.72 | 2,979.45 | 3,134.62 | **12,736.92** |
| 455 | 686.96 | 633.70 | 644.76 | 663.37 | **2,628.80** |
| 456 | 3,752.65 | 3,810.83 | 3,671.81 | 3,580.11 | **14,815.39** |
| 457 | 1,031.01 | 997.15 | 1,130.56 | 1,196.59 | **4,355.31** |
| 458 | 2,143.55 | 1,918.34 | 1,840.70 | 2,069.45 | **7,972.04** |
| **OH - OHIO - Total** | **79,034.32** | **76,713.28** | **77,315.92** | **79,497.42** | **312,560.94** |

**STATE: OK - OKLAHOMA**

| REGISTRANT ZIP CODE 3 | 1ST QUARTER | 2ND QUARTER | 3RD QUARTER | 4TH QUARTER | TOTAL GRAMS |
|---|---|---|---|---|---|
| 730 | 7,822.61 | 7,600.32 | 7,357.90 | 7,407.64 | **30,188.48** |
| 731 | 11,627.59 | 11,063.25 | 10,306.87 | 10,819.90 | **43,817.60** |
| 734 | 1,225.44 | 1,100.24 | 1,161.36 | 1,122.89 | **4,609.92** |
| 735 | 2,901.36 | 2,847.91 | 2,877.75 | 2,656.70 | **11,283.73** |
| 736 | 519.14 | 529.51 | 574.88 | 565.30 | **2,188.83** |
| 737 | 964.41 | 866.57 | 850.47 | 819.68 | **3,501.14** |
| 738 | 236.81 | 206.80 | 223.77 | 163.74 | **831.13** |
| 739 | 27.37 | 64.26 | 60.57 | 43.06 | **195.25** |
| 740 | 8,335.93 | 7,968.05 | 8,061.39 | 7,865.04 | **32,230.41** |
| 741 | 4,851.62 | 4,419.60 | 4,626.23 | 4,648.52 | **18,545.97** |
| 743 | 1,036.49 | 1,044.41 | 985.74 | 1,003.73 | **4,070.37** |
| 744 | 2,660.24 | 2,633.21 | 2,907.00 | 2,542.58 | **10,743.04** |
| 745 | 693.82 | 638.43 | 777.50 | 758.04 | **2,867.80** |
| 746 | 643.01 | 565.36 | 679.34 | 473.69 | **2,361.40** |

| | | | | | |
|---|---|---|---|---|---|
| 747 | 652.83 | 660.24 | 622.00 | 667.18 | **2,602.24** |
| 748 | 1,665.00 | 1,710.63 | 1,569.45 | 1,479.25 | **6,424.33** |
| 749 | 662.02 | 653.94 | 786.14 | 1,002.49 | **3,104.59** |
| **OK - OKLAHOMA - Total** | **46,525.69** | **44,572.73** | **44,428.37** | **44,039.43** | **179,566.23** |
| **STATE: OR - OREGON** | | | | | |
| REGISTRANT ZIP CODE 3 | 1ST QUARTER | 2ND QUARTER | 3RD QUARTER | 4TH QUARTER | **TOTAL GRAMS** |
| 970 | 7,907.99 | 7,297.76 | 7,508.17 | 7,353.59 | **30,067.52** |
| 971 | 4,336.96 | 3,667.82 | 4,128.69 | 3,948.56 | **16,082.04** |
| 972 | 12,195.68 | 12,505.65 | 12,369.73 | 11,759.49 | **48,830.54** |
| 973 | 5,122.95 | 4,956.69 | 4,991.52 | 4,945.53 | **20,016.69** |
| 974 | 7,908.07 | 7,581.60 | 7,673.02 | 7,615.23 | **30,777.92** |
| 975 | 5,357.70 | 5,276.05 | 5,357.65 | 5,268.60 | **21,260.00** |
| 976 | 1,011.00 | 1,117.15 | 1,266.15 | 1,258.99 | **4,653.30** |
| 977 | 3,098.52 | 2,868.03 | 2,768.13 | 2,702.26 | **11,436.94** |
| 978 | 1,351.55 | 1,338.26 | 1,353.66 | 1,449.12 | **5,492.60** |
| 979 | 407.62 | 366.25 | 347.28 | 313.22 | **1,434.37** |
| **OR - OREGON - Total** | **48,698.05** | **46,975.27** | **47,764.00** | **46,614.59** | **190,051.91** |
| **STATE: PA - PENNSYLVANIA** | | | | | |
| REGISTRANT ZIP CODE 3 | 1ST QUARTER | 2ND QUARTER | 3RD QUARTER | 4TH QUARTER | **TOTAL GRAMS** |
| 150 | 7,683.53 | 7,339.60 | 7,424.78 | 6,706.27 | **29,154.18** |
| 151 | 2,466.08 | 2,447.98 | 2,583.69 | 2,445.59 | **9,943.34** |
| 152 | 6,422.26 | 5,833.17 | 6,118.50 | 5,747.90 | **24,121.83** |
| 153 | 1,557.32 | 1,706.73 | 1,639.30 | 1,416.60 | **6,319.95** |
| 154 | 2,055.48 | 1,954.34 | 1,934.25 | 1,897.31 | **7,841.38** |
| 155 | 931.42 | 678.40 | 633.50 | 667.39 | **2,910.72** |
| 156 | 4,070.70 | 3,606.30 | 3,530.00 | 3,799.21 | **15,006.21** |
| 157 | 1,640.16 | 1,695.02 | 1,642.97 | 1,452.57 | **6,430.73** |
| 158 | 1,584.45 | 1,544.42 | 1,329.88 | 1,423.89 | **5,882.64** |
| 159 | 1,605.64 | 1,517.29 | 1,483.24 | 1,439.97 | **6,046.14** |
| 160 | 2,096.30 | 2,175.63 | 2,186.73 | 2,092.84 | **8,551.51** |
| 161 | 2,556.39 | 2,305.97 | 2,383.09 | 2,495.52 | **9,740.97** |
| 162 | 843.13 | 759.45 | 752.62 | 781.84 | **3,137.04** |
| 163 | 3,183.07 | 3,004.53 | 3,120.94 | 3,095.08 | **12,403.61** |
| 164 | 1,088.41 | 1,178.96 | 1,133.97 | 1,103.45 | **4,504.79** |
| 165 | 2,702.99 | 2,608.70 | 2,469.46 | 2,449.83 | **10,230.98** |
| 166 | 3,167.42 | 3,102.86 | 3,086.01 | 2,710.15 | **12,066.44** |
| 167 | 989.36 | 955.32 | 1,043.62 | 993.26 | **3,981.57** |
| 168 | 722.90 | 734.20 | 699.96 | 678.67 | **2,835.73** |
| 169 | 518.77 | 505.05 | 470.65 | 414.92 | **1,909.39** |
| 170 | 7,342.81 | 7,443.06 | 7,229.18 | 7,044.63 | **29,059.68** |
| 171 | 1,188.51 | 1,348.27 | 1,235.18 | 1,374.17 | **5,146.13** |
| 172 | 1,788.21 | 1,655.79 | 1,719.30 | 1,666.13 | **6,829.43** |
| 173 | 2,862.56 | 2,886.70 | 2,692.91 | 2,608.85 | **11,051.02** |
| 174 | 1,944.73 | 1,714.20 | 1,769.12 | 1,819.42 | **7,247.47** |
| 175 | 2,677.34 | 2,712.71 | 2,638.94 | 2,674.64 | **10,703.63** |
| 176 | 2,651.90 | 2,614.32 | 2,386.28 | 2,641.30 | **10,293.80** |
| 177 | 1,347.29 | 1,234.43 | 1,232.06 | 1,259.15 | **5,072.93** |
| 178 | 1,915.93 | 1,919.33 | 1,958.47 | 1,861.63 | **7,655.36** |
| 179 | 994.83 | 924.21 | 920.04 | 1,099.65 | **3,938.72** |
| 180 | 3,217.72 | 3,252.16 | 3,590.20 | 3,412.53 | **13,472.61** |
| 181 | 2,649.17 | 2,364.71 | 2,181.53 | 2,028.30 | **9,223.71** |
| 182 | 1,034.32 | 1,121.16 | 1,073.33 | 1,147.49 | **4,376.30** |
| 183 | 1,716.78 | 1,454.90 | 1,585.58 | 1,622.89 | **6,380.15** |
| 184 | 1,712.80 | 1,499.71 | 1,559.12 | 1,454.14 | **6,225.77** |
| 185 | 2,073.76 | 2,033.75 | 1,970.40 | 1,981.95 | **8,059.86** |
| 186 | 1,596.06 | 1,566.97 | 1,454.37 | 1,417.51 | **6,034.92** |
| 187 | 5,013.69 | 5,736.12 | 5,377.49 | 5,137.64 | **21,264.94** |
| 188 | 727.62 | 736.40 | 755.86 | 888.98 | **3,108.86** |
| 189 | 2,641.75 | 2,710.00 | 2,658.11 | 2,566.54 | **10,576.40** |
| 190 | 27,973.74 | 38,544.70 | 30,813.00 | 31,905.25 | **129,236.70** |

| | | | | | |
|---|---|---|---|---|---|
| 191 | 8,682.42 | 8,262.24 | 8,197.05 | 7,969.60 | **33,111.31** |
| 193 | 3,926.73 | 3,580.89 | 3,544.59 | 3,703.18 | **14,755.40** |
| 194 | 5,020.51 | 4,752.91 | 4,504.33 | 4,392.32 | **18,670.07** |
| 195 | 1,333.56 | 1,550.51 | 1,419.23 | 1,418.69 | **5,721.99** |
| 196 | 1,632.67 | 1,574.60 | 1,646.36 | 1,610.08 | **6,463.71** |
| **PA - PENNSYLVANIA - Total** | **143,553.21** | **150,848.67** | **141,779.20** | **140,518.92** | **576,699.99** |

**STATE: PR - PUERTO RICO**

| REGISTRANT ZIP CODE 3 | 1ST QUARTER | 2ND QUARTER | 3RD QUARTER | 4TH QUARTER | TOTAL GRAMS |
|---|---|---|---|---|---|
| 006 | 368.61 | 291.65 | 307.66 | 326.74 | **1,294.67** |
| 007 | 330.48 | 201.00 | 359.91 | 329.81 | **1,221.20** |
| 009 | 852.84 | 701.64 | 817.72 | 754.73 | **3,126.94** |
| **PR - PUERTO RICO - Total** | **1,551.93** | **1,194.30** | **1,485.29** | **1,411.28** | **5,642.81** |

**STATE: RI - RHODE ISLAND**

| REGISTRANT ZIP CODE 3 | 1ST QUARTER | 2ND QUARTER | 3RD QUARTER | 4TH QUARTER | TOTAL GRAMS |
|---|---|---|---|---|---|
| 028 | 5,242.24 | 4,946.09 | 5,061.08 | 4,810.75 | **20,060.16** |
| 029 | 2,856.62 | 2,960.76 | 3,176.50 | 2,735.68 | **11,729.57** |
| **RI - RHODE ISLAND - Total** | **8,098.86** | **7,906.86** | **8,237.58** | **7,546.43** | **31,789.73** |

**STATE: SC - SOUTH CAROLINA**

| REGISTRANT ZIP CODE 3 | 1ST QUARTER | 2ND QUARTER | 3RD QUARTER | 4TH QUARTER | TOTAL GRAMS |
|---|---|---|---|---|---|
| 290 | 4,579.80 | 4,435.95 | 4,326.70 | 4,276.83 | **17,619.28** |
| 291 | 4,015.19 | 3,942.38 | 4,131.92 | 4,022.58 | **16,112.07** |
| 292 | 4,066.23 | 4,002.85 | 3,914.85 | 3,938.72 | **15,922.65** |
| 293 | 3,894.93 | 3,792.21 | 3,696.17 | 3,984.88 | **15,368.19** |
| 294 | 9,874.99 | 9,130.91 | 9,073.12 | 9,033.89 | **37,112.90** |
| 295 | 7,900.22 | 7,018.52 | 7,462.34 | 7,634.21 | **30,015.29** |
| 296 | 9,310.25 | 9,059.79 | 8,643.06 | 9,229.44 | **36,242.53** |
| 297 | 2,357.07 | 2,298.22 | 2,288.63 | 2,125.89 | **9,069.80** |
| 298 | 2,158.85 | 1,857.90 | 1,903.34 | 1,806.98 | **7,727.07** |
| 299 | 2,043.22 | 1,827.27 | 2,074.59 | 1,903.72 | **7,848.81** |
| **SC - SOUTH CAROLINA - Total** | **50,200.75** | **47,366.00** | **47,514.72** | **47,957.14** | **193,038.60** |

**STATE: SD - SOUTH DAKOTA**

| REGISTRANT ZIP CODE 3 | 1ST QUARTER | 2ND QUARTER | 3RD QUARTER | 4TH QUARTER | TOTAL GRAMS |
|---|---|---|---|---|---|
| 570 | 1,113.21 | 1,043.39 | 1,011.73 | 1,075.32 | **4,243.65** |
| 571 | 1,824.94 | 1,586.60 | 1,606.39 | 1,656.07 | **6,674.01** |
| 572 | 253.50 | 245.17 | 324.72 | 250.34 | **1,073.73** |
| 573 | 319.66 | 314.01 | 340.52 | 413.31 | **1,387.50** |
| 574 | 272.01 | 268.09 | 249.99 | 281.64 | **1,071.73** |
| 575 | 274.86 | 284.06 | 234.66 | 277.06 | **1,070.64** |
| 576 | 60.53 | 65.53 | 43.59 | 45.57 | **215.22** |
| 577 | 1,276.28 | 1,233.63 | 1,214.03 | 1,196.94 | **4,920.88** |
| **SD - SOUTH DAKOTA - Total** | **5,394.99** | **5,040.47** | **5,025.65** | **5,196.25** | **20,657.36** |

**STATE: TN - TENNESSEE**

| REGISTRANT ZIP CODE 3 | 1ST QUARTER | 2ND QUARTER | 3RD QUARTER | 4TH QUARTER | TOTAL GRAMS |
|---|---|---|---|---|---|
| 370 | 8,959.25 | 8,755.98 | 8,842.03 | 8,190.07 | **34,747.33** |
| 371 | 9,595.61 | 9,241.99 | 9,410.39 | 9,125.32 | **37,373.30** |
| 372 | 6,053.54 | 5,621.27 | 5,743.07 | 5,745.25 | **23,163.13** |
| 373 | 7,286.94 | 6,742.77 | 6,389.44 | 6,559.87 | **26,979.02** |
| 374 | 1,942.50 | 1,900.22 | 1,954.28 | 1,913.94 | **7,710.94** |
| 376 | 6,638.07 | 6,648.39 | 6,579.76 | 6,353.71 | **26,219.93** |
| 377 | 5,308.62 | 5,368.92 | 5,160.45 | 5,363.93 | **21,201.91** |
| 378 | 8,112.27 | 8,113.56 | 7,774.38 | 8,091.34 | **32,091.55** |
| 379 | 5,695.23 | 5,501.93 | 5,191.71 | 5,455.73 | **21,844.59** |
| 380 | 2,869.73 | 2,940.05 | 2,789.52 | 2,879.79 | **11,479.10** |
| 381 | 21,226.28 | 26,964.00 | 28,365.95 | 26,504.50 | **103,060.74** |
| 382 | 1,284.99 | 1,203.74 | 1,128.45 | 1,131.42 | **4,748.60** |
| 383 | 3,815.40 | 3,280.52 | 3,469.14 | 3,478.51 | **14,043.57** |
| 384 | 2,358.26 | 2,030.83 | 2,270.58 | 2,026.71 | **8,686.38** |
| 385 | 4,223.38 | 3,860.34 | 4,383.19 | 3,847.64 | **16,314.55** |
| **TN - TENNESSEE - Total** | **95,370.06** | **98,174.50** | **99,452.35** | **96,667.75** | **389,664.66** |

**STATE: TX - TEXAS**

| REGISTRANT ZIP CODE 3 | 1ST QUARTER | 2ND QUARTER | 3RD QUARTER | 4TH QUARTER | TOTAL GRAMS |
|---|---|---|---|---|---|
| 750 | 11,601.32 | 10,791.63 | 11,029.36 | 10,824.35 | **44,246.66** |
| 751 | 3,896.22 | 3,734.03 | 3,839.37 | 3,689.02 | **15,158.63** |
| 752 | 6,974.26 | 5,718.18 | 6,564.07 | 6,311.72 | **25,568.24** |
| 753 | 200.23 | 265.87 | 222.90 | 201.90 | **890.91** |
| 754 | 2,951.56 | 2,837.92 | 2,825.41 | 2,789.94 | **11,404.82** |
| 755 | 927.01 | 912.95 | 947.29 | 920.92 | **3,708.17** |
| 756 | 3,340.01 | 2,322.48 | 2,394.53 | 2,638.03 | **10,695.04** |
| 757 | 2,799.47 | 2,674.87 | 2,845.98 | 2,908.28 | **11,228.60** |
| 758 | 622.68 | 656.17 | 666.12 | 590.50 | **2,535.47** |
| 759 | 1,531.16 | 1,456.00 | 1,442.65 | 1,657.63 | **6,087.44** |
| 760 | 7,612.78 | 7,309.61 | 7,260.58 | 7,319.55 | **29,502.52** |
| 761 | 8,797.73 | 8,464.66 | 8,656.05 | 8,627.33 | **34,545.77** |
| 762 | 2,958.65 | 2,742.97 | 2,858.70 | 2,897.06 | **11,457.38** |
| 763 | 1,650.72 | 1,472.46 | 1,584.26 | 1,516.42 | **6,223.85** |
| 764 | 766.25 | 656.93 | 702.42 | 656.09 | **2,781.69** |
| 765 | 2,760.11 | 3,076.49 | 2,723.55 | 2,740.13 | **11,300.29** |
| 766 | 680.55 | 840.60 | 740.78 | 770.18 | **3,032.12** |
| 767 | 1,974.75 | 1,995.14 | 1,893.99 | 1,752.76 | **7,616.65** |
| 768 | 838.36 | 751.62 | 820.28 | 845.34 | **3,255.60** |
| 769 | 991.52 | 903.43 | 749.88 | 782.84 | **3,427.67** |
| 770 | 10,662.34 | 9,120.45 | 9,824.95 | 9,503.71 | **39,111.45** |
| 773 | 6,562.00 | 6,057.20 | 6,319.43 | 6,276.93 | **25,215.56** |
| 774 | 4,175.45 | 4,244.90 | 4,485.78 | 4,022.55 | **16,928.68** |
| 775 | 8,615.73 | 7,351.55 | 7,430.71 | 7,684.47 | **31,082.47** |
| 776 | 1,740.01 | 1,601.46 | 1,741.97 | 1,698.76 | **6,782.20** |
| 777 | 2,270.67 | 1,717.22 | 2,116.27 | 1,703.49 | **7,807.65** |
| 778 | 1,702.58 | 1,636.85 | 1,619.43 | 1,704.69 | **6,663.55** |
| 779 | 1,784.17 | 1,513.59 | 1,625.75 | 1,397.72 | **6,321.22** |
| 780 | 2,491.50 | 2,599.22 | 2,849.13 | 2,573.81 | **10,513.66** |
| 781 | 1,875.40 | 1,974.20 | 1,985.04 | 1,981.17 | **7,815.82** |
| 782 | 10,253.82 | 9,240.65 | 9,598.28 | 9,585.20 | **38,677.95** |
| 783 | 1,220.97 | 1,244.55 | 1,121.74 | 1,267.38 | **4,854.65** |
| 784 | 2,438.24 | 2,186.90 | 2,369.01 | 2,276.93 | **9,271.08** |
| 785 | 2,169.82 | 1,889.65 | 2,163.90 | 2,076.98 | **8,300.35** |
| 786 | 5,649.39 | 5,556.87 | 5,992.24 | 6,076.72 | **23,275.22** |
| 787 | 4,566.79 | 4,688.50 | 4,546.04 | 4,515.07 | **18,316.41** |
| 788 | 270.77 | 261.08 | 286.15 | 309.73 | **1,127.73** |
| 789 | 343.26 | 343.69 | 419.29 | 432.91 | **1,539.15** |
| 790 | 1,328.13 | 1,061.14 | 906.94 | 961.08 | **4,257.29** |
| 791 | 2,456.82 | 2,522.98 | 2,574.77 | 2,333.08 | **9,887.64** |
| 792 | 269.48 | 222.97 | 201.72 | 201.57 | **895.74** |
| 793 | 556.36 | 581.81 | 576.54 | 586.29 | **2,300.99** |
| 794 | 2,766.61 | 2,682.61 | 2,764.57 | 3,024.43 | **11,238.22** |
| 795 | 365.28 | 329.18 | 326.81 | 310.41 | **1,331.68** |
| 796 | 1,180.17 | 1,165.13 | 1,061.96 | 1,112.59 | **4,519.85** |
| 797 | 1,789.36 | 1,605.87 | 1,571.78 | 1,847.16 | **6,814.16** |
| 798 | 105.91 | 55.11 | 90.04 | 131.46 | **382.52** |
| 799 | 2,722.24 | 2,305.19 | 2,536.31 | 2,698.91 | **10,262.64** |
| **TX - TEXAS - Total** | **146,208.62** | **135,344.55** | **139,874.73** | **138,735.18** | **560,163.09** |

**STATE: UT - UTAH**

| REGISTRANT ZIP CODE 3 | 1ST QUARTER | 2ND QUARTER | 3RD QUARTER | 4TH QUARTER | TOTAL GRAMS |
|---|---|---|---|---|---|
| 840 | 13,866.02 | 13,707.14 | 13,877.78 | 13,304.33 | **54,755.27** |
| 841 | 9,405.37 | 8,719.84 | 8,370.77 | 8,094.00 | **34,589.98** |
| 843 | 1,461.48 | 1,592.70 | 1,527.03 | 1,449.42 | **6,030.63** |
| 844 | 3,203.19 | 3,215.40 | 3,156.34 | 3,085.88 | **12,660.80** |
| 845 | 900.69 | 997.27 | 1,015.57 | 948.67 | **3,862.21** |
| 846 | 2,429.38 | 2,444.54 | 2,523.99 | 2,421.39 | **9,819.30** |
| 847 | 4,284.79 | 4,395.45 | 4,599.43 | 4,624.73 | **17,904.41** |
| **UT - UTAH - Total** | **35,550.93** | **35,072.34** | **35,070.91** | **33,928.43** | **139,622.61** |

**STATE: VA - VIRGINIA**

| REGISTRANT ZIP CODE 3 | 1ST QUARTER | 2ND QUARTER | 3RD QUARTER | 4TH QUARTER | TOTAL GRAMS |
|---|---|---|---|---|---|
| 201 | 4,564.63 | 4,574.68 | 4,311.37 | 4,504.58 | **17,955.25** |
| 220 | 2,197.46 | 1,946.09 | 2,317.41 | 1,736.02 | **8,196.97** |
| 221 | 2,192.33 | 2,134.59 | 2,077.38 | 2,172.09 | **8,576.38** |
| 222 | 446.56 | 433.82 | 387.93 | 398.29 | **1,666.61** |
| 223 | 1,159.94 | 1,074.03 | 1,139.79 | 1,076.68 | **4,450.43** |
| 224 | 2,015.78 | 1,934.50 | 2,025.83 | 2,031.85 | **8,007.96** |
| 225 | 799.92 | 780.44 | 866.24 | 864.04 | **3,310.64** |
| 226 | 2,119.64 | 2,058.13 | 2,114.33 | 1,974.11 | **8,266.20** |
| 227 | 451.79 | 420.25 | 437.42 | 415.31 | **1,724.78** |
| 228 | 1,558.51 | 1,528.45 | 1,529.00 | 1,608.45 | **6,224.41** |
| 229 | 1,925.62 | 1,950.88 | 2,153.01 | 1,942.76 | **7,972.27** |
| 230 | 1,589.87 | 1,685.08 | 1,485.40 | 1,433.50 | **6,193.85** |
| 231 | 2,675.58 | 2,477.91 | 2,939.17 | 2,818.48 | **10,911.14** |
| 232 | 4,794.24 | 4,849.22 | 4,404.39 | 4,400.33 | **18,448.18** |
| 233 | 2,218.74 | 2,092.69 | 2,017.72 | 1,962.14 | **8,291.28** |
| 234 | 4,239.44 | 4,182.94 | 4,059.51 | 4,129.83 | **16,611.71** |
| 235 | 1,592.28 | 1,330.18 | 1,354.73 | 1,411.48 | **5,688.67** |
| 236 | 2,674.62 | 2,558.22 | 2,665.09 | 2,637.58 | **10,535.51** |
| 237 | 991.57 | 859.41 | 1,081.86 | 1,015.83 | **3,948.67** |
| 238 | 2,366.00 | 2,249.47 | 2,472.62 | 2,241.01 | **9,329.10** |
| 239 | 617.42 | 607.59 | 618.53 | 616.87 | **2,460.41** |
| 240 | 3,453.96 | 3,172.78 | 3,007.19 | 3,068.24 | **12,702.16** |
| 241 | 3,062.66 | 2,928.68 | 3,158.30 | 3,047.63 | **12,197.27** |
| 242 | 2,930.34 | 2,847.89 | 2,886.43 | 2,849.06 | **11,513.73** |
| 243 | 1,654.47 | 1,602.72 | 1,573.81 | 1,567.78 | **6,398.77** |
| 244 | 1,005.00 | 958.90 | 1,008.22 | 987.08 | **3,959.20** |
| 245 | 2,676.91 | 2,576.04 | 2,661.70 | 2,765.77 | **10,680.41** |
| 246 | 524.29 | 675.00 | 533.57 | 531.85 | **2,264.70** |
| **VA - VIRGINIA - Total** | **58,499.56** | **56,490.56** | **57,287.93** | **56,208.62** | **228,486.67** |

**STATE: VI - VIRGIN ISLANDS**

| REGISTRANT ZIP CODE 3 | 1ST QUARTER | 2ND QUARTER | 3RD QUARTER | 4TH QUARTER | TOTAL GRAMS |
|---|---|---|---|---|---|
| 008 | 71.55 | 64.07 | 69.14 | 68.21 | **272.97** |
| **VI - VIRGIN ISLANDS - Total** | **71.55** | **64.07** | **69.14** | **68.21** | **272.97** |

**STATE: VT - VERMONT**

| REGISTRANT ZIP CODE 3 | 1ST QUARTER | 2ND QUARTER | 3RD QUARTER | 4TH QUARTER | TOTAL GRAMS |
|---|---|---|---|---|---|
| 050 | 786.70 | 736.58 | 704.47 | 664.41 | **2,892.17** |
| 051 | 353.79 | 319.24 | 389.86 | 380.33 | **1,443.23** |
| 052 | 404.90 | 407.72 | 379.13 | 422.61 | **1,614.36** |
| 053 | 458.13 | 514.68 | 471.16 | 518.78 | **1,962.75** |
| 054 | 3,231.70 | 2,882.44 | 2,766.13 | 2,988.57 | **11,868.84** |
| 056 | 575.20 | 663.81 | 543.63 | 659.47 | **2,442.12** |
| 057 | 1,156.93 | 1,203.22 | 1,092.22 | 998.99 | **4,451.36** |
| 058 | 649.01 | 578.26 | 556.22 | 652.86 | **2,436.35** |
| **VT - VERMONT - Total** | **7,616.38** | **7,305.95** | **6,902.82** | **7,286.03** | **29,111.18** |

**STATE: WA - WASHINGTON**

| REGISTRANT ZIP CODE 3 | 1ST QUARTER | 2ND QUARTER | 3RD QUARTER | 4TH QUARTER | TOTAL GRAMS |
|---|---|---|---|---|---|
| 980 | 10,900.66 | 11,286.83 | 11,955.63 | 10,766.52 | **44,909.63** |
| 981 | 6,957.11 | 6,079.49 | 6,463.90 | 6,142.53 | **25,643.04** |
| 982 | 8,477.78 | 8,442.31 | 8,410.29 | 8,236.28 | **33,566.66** |
| 983 | 8,712.56 | 8,450.67 | 8,883.99 | 8,453.61 | **34,500.84** |
| 984 | 4,452.79 | 4,263.85 | 4,263.48 | 4,233.21 | **17,213.32** |
| 985 | 4,785.77 | 4,522.47 | 4,890.06 | 4,523.94 | **18,722.25** |
| 986 | 10,041.75 | 9,318.47 | 9,395.47 | 9,351.51 | **38,107.20** |
| 988 | 2,644.23 | 2,358.27 | 2,086.43 | 2,324.49 | **9,413.41** |
| 989 | 2,409.59 | 2,229.17 | 2,266.28 | 2,260.58 | **9,165.63** |
| 990 | 1,191.08 | 1,142.80 | 1,184.13 | 1,146.97 | **4,664.97** |
| 991 | 1,519.42 | 1,596.45 | 1,559.78 | 1,632.25 | **6,307.90** |
| 992 | 7,016.40 | 6,656.07 | 6,762.73 | 6,447.86 | **26,883.06** |

| | | | | | |
|---|---|---|---|---|---|
| 993 | 3,555.01 | 3,351.32 | 3,391.65 | 3,263.11 | **13,561.09** |
| 994 | 827.64 | 889.06 | 970.23 | 947.72 | **3,634.66** |
| **WA - WASHINGTON - Total** | **73,491.80** | **70,587.23** | **72,484.04** | **69,730.59** | **286,293.65** |

**STATE: WI - WISCONSIN**

| REGISTRANT ZIP CODE 3 | 1ST QUARTER | 2ND QUARTER | 3RD QUARTER | 4TH QUARTER | TOTAL GRAMS |
|---|---|---|---|---|---|
| 530 | 5,542.63 | 5,130.03 | 5,012.82 | 5,424.00 | **21,109.48** |
| 531 | 5,353.41 | 5,308.84 | 5,032.98 | 4,983.83 | **20,679.05** |
| 532 | 7,569.87 | 6,960.22 | 7,143.97 | 6,762.85 | **28,436.92** |
| 534 | 1,138.30 | 1,114.84 | 1,144.73 | 1,184.17 | **4,582.04** |
| 535 | 5,371.68 | 5,281.46 | 5,632.78 | 5,412.54 | **21,698.47** |
| 537 | 3,911.97 | 3,742.40 | 4,014.94 | 3,747.76 | **15,417.07** |
| 538 | 522.27 | 508.96 | 510.99 | 528.77 | **2,070.98** |
| 539 | 2,757.21 | 2,704.55 | 2,529.01 | 2,678.75 | **10,669.52** |
| 540 | 725.06 | 689.16 | 650.69 | 671.18 | **2,736.09** |
| 541 | 1,257.62 | 1,350.40 | 1,279.15 | 1,332.78 | **5,219.94** |
| 542 | 863.07 | 833.77 | 849.31 | 787.08 | **3,333.22** |
| 543 | 1,572.55 | 1,434.55 | 1,681.42 | 2,036.86 | **6,725.37** |
| 544 | 2,653.99 | 2,478.89 | 2,469.63 | 2,594.61 | **10,197.12** |
| 545 | 761.12 | 792.64 | 837.46 | 767.98 | **3,159.20** |
| 546 | 2,458.49 | 2,328.46 | 2,320.70 | 2,363.12 | **9,470.77** |
| 547 | 2,154.34 | 2,059.95 | 2,163.49 | 2,343.27 | **8,721.05** |
| 548 | 1,637.62 | 1,571.98 | 1,621.96 | 1,524.10 | **6,355.66** |
| 549 | 4,517.75 | 4,463.82 | 4,400.10 | 4,220.47 | **17,602.14** |
| **WI - WISCONSIN - Total** | **50,768.96** | **48,754.92** | **49,296.13** | **49,364.10** | **198,184.11** |

**STATE: WV - WEST VIRGINIA**

| REGISTRANT ZIP CODE 3 | 1ST QUARTER | 2ND QUARTER | 3RD QUARTER | 4TH QUARTER | TOTAL GRAMS |
|---|---|---|---|---|---|
| 247 | 496.54 | 479.63 | 466.50 | 383.94 | **1,826.61** |
| 248 | 225.00 | 186.80 | 164.65 | 239.93 | **816.37** |
| 249 | 267.34 | 231.49 | 259.18 | 243.89 | **1,001.90** |
| 250 | 505.49 | 383.37 | 406.31 | 489.06 | **1,784.23** |
| 251 | 214.47 | 220.49 | 250.27 | 200.03 | **885.25** |
| 252 | 250.24 | 245.08 | 194.49 | 220.86 | **910.67** |
| 253 | 946.62 | 970.90 | 1,032.92 | 941.25 | **3,891.68** |
| 254 | 2,314.17 | 2,379.31 | 2,296.92 | 2,157.35 | **9,147.76** |
| 255 | 952.51 | 853.74 | 815.55 | 865.10 | **3,486.89** |
| 256 | 223.98 | 195.44 | 235.66 | 181.61 | **836.70** |
| 257 | 695.55 | 871.96 | 758.14 | 680.91 | **3,006.56** |
| 258 | 591.24 | 785.17 | 616.99 | 806.10 | **2,799.51** |
| 259 | 157.17 | 162.88 | 191.31 | 210.45 | **721.81** |
| 260 | 1,351.38 | 1,303.71 | 1,338.07 | 1,329.40 | **5,322.56** |
| 261 | 713.06 | 670.34 | 601.42 | 592.44 | **2,577.26** |
| 262 | 435.42 | 485.26 | 598.76 | 465.10 | **1,984.54** |
| 263 | 1,122.47 | 1,085.44 | 1,141.78 | 1,195.40 | **4,545.10** |
| 264 | 380.76 | 342.70 | 302.33 | 371.37 | **1,397.17** |
| 265 | 1,125.11 | 1,201.57 | 1,269.86 | 1,440.13 | **5,036.67** |
| 266 | 239.83 | 255.29 | 268.24 | 213.77 | **977.13** |
| 267 | 452.70 | 445.97 | 528.62 | 506.95 | **1,934.23** |
| 268 | 260.10 | 238.90 | 252.41 | 248.86 | **1,000.27** |
| **WV - WEST VIRGINIA - Total** | **13,921.15** | **13,995.43** | **13,990.38** | **13,983.88** | **55,890.85** |

**STATE: WY - WYOMING**

| REGISTRANT ZIP CODE 3 | 1ST QUARTER | 2ND QUARTER | 3RD QUARTER | 4TH QUARTER | TOTAL GRAMS |
|---|---|---|---|---|---|
| 820 | 1,036.97 | 792.10 | 833.93 | 902.33 | **3,565.33** |
| 822 | 137.46 | 183.31 | 169.75 | 157.03 | **647.55** |
| 823 | 80.76 | 81.07 | 78.97 | 77.27 | **318.07** |
| 824 | 666.67 | 612.87 | 635.45 | 589.69 | **2,504.68** |
| 825 | 586.08 | 490.77 | 512.15 | 508.74 | **2,097.74** |
| 826 | 1,009.76 | 1,007.84 | 1,028.26 | 1,014.47 | **4,060.33** |
| 827 | 376.23 | 356.43 | 323.85 | 348.26 | **1,404.77** |
| 828 | 745.99 | 574.58 | 672.40 | 801.43 | **2,794.40** |
| 829 | 1,354.84 | 1,410.24 | 1,434.52 | 1,434.22 | **5,633.82** |

| | | | | | |
|---|---|---|---|---|---|
| 830 | 52.45 | 75.69 | 48.52 | 41.77 | **218.44** |
| 831 | 207.66 | 239.64 | 289.83 | 275.72 | **1,012.86** |
| **WY - WYOMING - Total** | **6,254.88** | **5,824.53** | **6,027.63** | **6,150.94** | **24,257.98** |

**DRUG: 9652 - OXYMORPHONE**

**STATE: AK - ALASKA**

| REGISTRANT ZIP CODE 3 | 1ST QUARTER | 2ND QUARTER | 3RD QUARTER | 4TH QUARTER | TOTAL GRAMS |
|---|---|---|---|---|---|
| 995 | 11.42 | 7.76 | 9.81 | 9.10 | **38.09** |
| 996 | 25.25 | 20.34 | 15.34 | 12.85 | **73.78** |
| 997 | 1.07 | 1.61 | 1.07 | 0 | **3.75** |
| **AK - ALASKA - Total** | **37.74** | **29.71** | **26.23** | **21.95** | **115.62** |

**STATE: AL - ALABAMA**

| REGISTRANT ZIP CODE 3 | 1ST QUARTER | 2ND QUARTER | 3RD QUARTER | 4TH QUARTER | TOTAL GRAMS |
|---|---|---|---|---|---|
| 350 | 66.86 | 61.73 | 70.34 | 60.22 | **259.16** |
| 351 | 38.63 | 29.26 | 22.48 | 16.95 | **107.32** |
| 352 | 41.66 | 46.52 | 40.72 | 28.55 | **157.46** |
| 354 | 69.27 | 62.89 | 56.02 | 62.09 | **250.28** |
| 355 | 40.37 | 49.69 | 34.84 | 31.67 | **156.56** |
| 356 | 74.40 | 59.68 | 64.68 | 27.83 | **226.59** |
| 357 | 41.30 | 34.61 | 39.88 | 54.15 | **169.95** |
| 358 | 78.86 | 61.55 | 75.03 | 48.17 | **263.62** |
| 359 | 158.44 | 169.99 | 176.06 | 139.08 | **643.56** |
| 360 | 25.69 | 13.74 | 10.35 | 13.47 | **63.25** |
| 361 | 14.99 | 11.69 | 14.01 | 6.33 | **47.01** |
| 362 | 50.36 | 39.61 | 47.55 | 38.90 | **176.41** |
| 363 | 67.26 | 57.54 | 49.33 | 49.24 | **223.38** |
| 364 | 8.21 | 8.65 | 2.85 | 1.78 | **21.50** |
| 365 | 60.48 | 53.17 | 47.10 | 48.04 | **208.80** |
| 366 | 93.22 | 89.75 | 82.61 | 74.40 | **339.98** |
| 367 | 3.39 | 5.62 | 10.71 | 11.24 | **30.96** |
| 368 | 105.27 | 82.79 | 73.51 | 87.16 | **348.72** |
| **AL - ALABAMA - Total** | **1,038.67** | **938.49** | **918.06** | **799.28** | **3,694.50** |

**STATE: AR - ARKANSAS**

| REGISTRANT ZIP CODE 3 | 1ST QUARTER | 2ND QUARTER | 3RD QUARTER | 4TH QUARTER | TOTAL GRAMS |
|---|---|---|---|---|---|
| 716 | 2.68 | 2.23 | 2.68 | 1.78 | **9.37** |
| 717 | 7.58 | 7.58 | 7.14 | 5.35 | **27.66** |
| 719 | 2.81 | 1.78 | 2.32 | 0.27 | **7.18** |
| 720 | 29.66 | 22.61 | 21.01 | 17.40 | **90.68** |
| 721 | 11.29 | 11.86 | 21.95 | 8.03 | **53.12** |
| 722 | 20.25 | 15.97 | 16.50 | 13.69 | **66.42** |
| 724 | 5.17 | 3.21 | 3.21 | 5.89 | **17.49** |
| 725 | 1.34 | 5.53 | 2.41 | 2.41 | **11.69** |
| 726 | 2.50 | 2.05 | 1.25 | 2.05 | **7.85** |
| 727 | 31.67 | 34.61 | 34.52 | 23.19 | **124.00** |
| 729 | 12.49 | 8.21 | 1.43 | 4.91 | **27.03** |
| **AR - ARKANSAS - Total** | **127.44** | **115.66** | **114.41** | **84.97** | **442.48** |

**STATE: AZ - ARIZONA**

| REGISTRANT ZIP CODE 3 | 1ST QUARTER | 2ND QUARTER | 3RD QUARTER | 4TH QUARTER | TOTAL GRAMS |
|---|---|---|---|---|---|
| 850 | 211.87 | 183.42 | 144.21 | 146.53 | **686.02** |
| 851 | 20.52 | 23.55 | 32.47 | 25.34 | **101.88** |
| 852 | 472.28 | 441.63 | 446.41 | 459.43 | **1,819.75** |
| 853 | 272.31 | 227.57 | 264.33 | 208.53 | **972.75** |
| 855 | 10.62 | 9.81 | 12.31 | 12.04 | **44.78** |
| 856 | 19.54 | 23.91 | 24.44 | 20.25 | **88.14** |
| 857 | 113.83 | 96.70 | 90.37 | 87.43 | **388.33** |
| 859 | 6.60 | 2.23 | 1.78 | 4.37 | **14.99** |
| 860 | 6.42 | 4.55 | 5.98 | 3.03 | **19.98** |
| 863 | 27.74 | 28.37 | 26.41 | 24.80 | **107.32** |
| 864 | 19.27 | 12.49 | 15.97 | 10.88 | **58.61** |
| **AZ - ARIZONA - Total** | **1,181.01** | **1,054.24** | **1,064.68** | **1,002.63** | **4,302.55** |

**STATE: CA - CALIFORNIA**

| REGISTRANT ZIP CODE 3 | 1ST QUARTER | 2ND QUARTER | 3RD QUARTER | 4TH QUARTER | TOTAL GRAMS |
|---|---|---|---|---|---|
| 900 | 160.31 | 112.49 | 152.19 | 195.01 | **620.01** |
| 902 | 114.99 | 133.99 | 116.15 | 110.44 | **475.58** |
| 903 | 0.54 | 0 | 0 | 0 | **0.54** |
| 904 | 27.83 | 20.70 | 22.30 | 15.43 | **86.27** |
| 905 | 2.14 | 3.03 | 0 | 0 | **5.17** |
| 906 | 49.69 | 40.41 | 31.13 | 14.63 | **135.87** |
| 907 | 25.25 | 18.82 | 21.50 | 47.37 | **112.94** |
| 908 | 22.57 | 31.49 | 25.34 | 34.52 | **113.92** |
| 910 | 5.35 | 10.88 | 10.53 | 13.38 | **40.14** |
| 911 | 58.34 | 76.81 | 73.78 | 74.67 | **283.60** |
| 912 | 14.45 | 12.67 | 13.56 | 9.10 | **49.78** |
| 913 | 86.13 | 89.34 | 79.84 | 57.18 | **312.50** |
| 914 | 139.79 | 107.50 | 87.07 | 103.48 | **437.84** |
| 915 | 5.35 | 3.57 | 3.84 | 3.75 | **16.50** |
| 916 | 23.82 | 17.75 | 16.06 | 19.00 | **76.63** |
| 917 | 58.52 | 56.65 | 47.73 | 38.27 | **201.17** |
| 919 | 36.04 | 47.10 | 23.02 | 30.78 | **136.94** |
| 920 | 411.70 | 428.83 | 418.04 | 387.89 | **1,646.46** |
| 921 | 88.85 | 71.68 | 73.87 | 67.31 | **301.71** |
| 922 | 83.14 | 53.44 | 66.10 | 59.50 | **262.19** |
| 923 | 68.16 | 43.09 | 58.43 | 40.41 | **210.09** |
| 924 | 13.38 | 23.55 | 15.52 | 16.41 | **68.87** |
| 925 | 64.32 | 57.99 | 54.51 | 59.86 | **236.67** |
| 926 | 172.18 | 137.96 | 143.18 | 141.31 | **594.63** |
| 927 | 76.90 | 111.87 | 72.13 | 24.09 | **284.98** |
| 928 | 132.30 | 93.05 | 125.16 | 92.96 | **443.46** |
| 930 | 65.30 | 50.58 | 55.04 | 46.57 | **217.49** |
| 931 | 0.89 | 0.45 | 1.34 | 1.69 | **4.37** |
| 932 | 47.28 | 26.23 | 32.56 | 31.13 | **137.20** |
| 933 | 5.35 | 10.71 | 3.57 | 7.05 | **26.67** |
| 934 | 54.77 | 46.12 | 50.76 | 69.94 | **221.60** |
| 935 | 55.93 | 38.18 | 41.66 | 39.07 | **174.85** |
| 936 | 56.29 | 59.50 | 46.30 | 45.14 | **207.23** |
| 937 | 108.30 | 143.00 | 95.05 | 109.51 | **455.86** |
| 939 | 4.28 | 9.90 | 9.99 | 8.12 | **32.29** |
| 940 | 141.58 | 124.58 | 132.70 | 147.42 | **546.28** |
| 941 | 24.89 | 24.44 | 18.24 | 19.18 | **86.76** |
| 943 | 13.74 | 11.60 | 16.68 | 14.45 | **56.47** |
| 944 | 14.63 | 15.17 | 2.94 | 11.15 | **43.89** |
| 945 | 180.29 | 160.22 | 153.53 | 190.64 | **684.69** |
| 946 | 1.34 | 0.45 | 0 | 0.54 | **2.32** |
| 947 | 0 | 0 | 0.27 | 0 | **0.27** |
| 948 | 1.78 | 3.57 | 0 | 1.78 | **7.14** |
| 949 | 46.21 | 37.02 | 39.79 | 35.51 | **158.53** |
| 950 | 42.02 | 17.57 | 18.11 | 15.97 | **93.67** |
| 951 | 15.75 | 11.73 | 14.85 | 12.62 | **54.95** |
| 952 | 21.50 | 24.80 | 21.59 | 24.18 | **92.06** |
| 953 | 44.43 | 42.29 | 45.23 | 43.09 | **175.03** |
| 954 | 43.45 | 51.12 | 39.70 | 59.15 | **193.41** |
| 955 | 15.70 | 0 | 0 | 2.68 | **18.38** |
| 956 | 59.15 | 49.51 | 49.15 | 48.08 | **205.90** |
| 957 | 18.11 | 18.47 | 16.15 | 10.62 | **63.34** |
| 958 | 19.89 | 21.59 | 17.66 | 11.86 | **71.01** |
| 959 | 40.59 | 25.87 | 27.12 | 19.27 | **112.85** |
| 960 | 10.08 | 5.53 | 10.35 | 9.63 | **35.59** |
| 961 | 0 | 0 | 0.54 | 1.07 | **1.61** |
| **CA - CALIFORNIA - Total** | **3,095.59** | **2,834.87** | **2,711.85** | **2,693.87** | **11,336.18** |

**STATE: CO - COLORADO**

| REGISTRANT ZIP CODE 3 | 1ST QUARTER | 2ND QUARTER | 3RD QUARTER | 4TH QUARTER | TOTAL GRAMS |
|---|---|---|---|---|---|

| | | | | | |
|---|---|---|---|---|---|
| 800 | 142.47 | 129.49 | 145.41 | 160.00 | **577.37** |
| 801 | 166.73 | 176.46 | 193.14 | 188.95 | **725.28** |
| 802 | 160.40 | 144.34 | 129.40 | 95.37 | **529.51** |
| 803 | 29.97 | 9.99 | 18.91 | 13.29 | **72.17** |
| 804 | 4.73 | 6.33 | 8.12 | 1.87 | **21.05** |
| 805 | 77.61 | 56.02 | 52.72 | 50.63 | **236.99** |
| 806 | 57.14 | 70.03 | 65.12 | 66.37 | **258.66** |
| 807 | 12.40 | 4.91 | 4.01 | 6.69 | **28.01** |
| 808 | 10.79 | 8.30 | 17.84 | 8.74 | **45.68** |
| 809 | 89.21 | 78.33 | 92.56 | 78.86 | **338.95** |
| 810 | 64.14 | 27.92 | 39.34 | 42.29 | **173.69** |
| 811 | 12.76 | 8.74 | 7.14 | 7.14 | **35.77** |
| 812 | 16.95 | 14.99 | 8.47 | 12.49 | **52.90** |
| 813 | 0 | 0 | 0 | 0.54 | **0.54** |
| 815 | 2.41 | 2.94 | 2.68 | 2.41 | **10.44** |
| 816 | 0 | 9.72 | 1.78 | 0.45 | **11.95** |
| **CO - COLORADO - Total** | **847.72** | **748.52** | **786.65** | **736.07** | **3,118.96** |

**STATE: CT - CONNECTICUT**

| REGISTRANT ZIP CODE 3 | 1ST QUARTER | 2ND QUARTER | 3RD QUARTER | 4TH QUARTER | TOTAL GRAMS |
|---|---|---|---|---|---|
| 060 | 91.66 | 95.19 | 93.76 | 106.29 | **386.90** |
| 061 | 8.83 | 11.33 | 11.69 | 8.30 | **40.14** |
| 062 | 14.27 | 11.60 | 15.08 | 21.86 | **62.80** |
| 063 | 18.47 | 21.95 | 10.26 | 6.60 | **57.27** |
| 064 | 367.46 | 350.10 | 334.89 | 321.29 | **1,373.74** |
| 065 | 96.21 | 101.65 | 78.19 | 73.20 | **349.26** |
| 066 | 71.99 | 67.35 | 44.96 | 48.08 | **232.39** |
| 067 | 41.13 | 46.48 | 30.24 | 39.25 | **157.10** |
| 068 | 98.67 | 90.19 | 68.69 | 74.27 | **331.82** |
| 069 | 38.18 | 28.37 | 25.87 | 21.95 | **114.37** |
| **CT - CONNECTICUT - Total** | **846.87** | **824.21** | **713.64** | **721.08** | **3,105.80** |

**STATE: DC - DISTRICT OF COLUMBIA**

| REGISTRANT ZIP CODE 3 | 1ST QUARTER | 2ND QUARTER | 3RD QUARTER | 4TH QUARTER | TOTAL GRAMS |
|---|---|---|---|---|---|
| 200 | 52.28 | 34.08 | 37.65 | 30.96 | **154.96** |
| **DC - DISTRICT OF COLUMBIA - Total** | **52.28** | **34.08** | **37.65** | **30.96** | **154.96** |

**STATE: DE - DELAWARE**

| REGISTRANT ZIP CODE 3 | 1ST QUARTER | 2ND QUARTER | 3RD QUARTER | 4TH QUARTER | TOTAL GRAMS |
|---|---|---|---|---|---|
| 197 | 291.76 | 209.96 | 232.17 | 216.38 | **950.26** |
| 198 | 191.44 | 206.08 | 167.71 | 187.16 | **752.40** |
| 199 | 367.32 | 316.52 | 278.07 | 261.56 | **1,223.47** |
| **DE - DELAWARE - Total** | **850.53** | **732.55** | **677.95** | **665.11** | **2,926.13** |

**STATE: FL - FLORIDA**

| REGISTRANT ZIP CODE 3 | 1ST QUARTER | 2ND QUARTER | 3RD QUARTER | 4TH QUARTER | TOTAL GRAMS |
|---|---|---|---|---|---|
| 320 | 206.43 | 137.29 | 140.02 | 132.34 | **616.08** |
| 321 | 83.10 | 77.61 | 68.42 | 76.63 | **305.77** |
| 322 | 447.79 | 373.79 | 366.70 | 384.99 | **1,573.26** |
| 323 | 212.94 | 195.95 | 196.26 | 205.00 | **810.16** |
| 324 | 100.27 | 70.39 | 64.50 | 64.45 | **299.61** |
| 325 | 37.47 | 30.15 | 36.84 | 29.35 | **133.82** |
| 326 | 164.50 | 155.49 | 129.71 | 132.57 | **582.27** |
| 327 | 215.00 | 193.54 | 199.52 | 185.69 | **793.75** |
| 328 | 154.73 | 154.56 | 147.15 | 134.71 | **591.15** |
| 329 | 347.65 | 314.87 | 348.81 | 321.51 | **1,332.84** |
| 330 | 280.74 | 214.28 | 201.79 | 208.13 | **904.95** |
| 331 | 90.55 | 73.96 | 57.63 | 59.41 | **281.55** |
| 333 | 221.37 | 190.20 | 155.27 | 143.85 | **710.69** |
| 334 | 497.61 | 426.07 | 395.11 | 363.00 | **1,681.79** |
| 335 | 213.30 | 236.94 | 203.67 | 163.88 | **817.79** |
| 336 | 321.20 | 283.73 | 240.11 | 243.54 | **1,088.59** |
| 337 | 329.99 | 274.86 | 303.85 | 284.67 | **1,193.36** |
| 338 | 64.94 | 78.42 | 57.18 | 63.34 | **263.88** |

| | | | | | |
|---|---|---|---|---|---|
| 339 | 121.19 | 107.14 | 97.77 | 64.77 | **390.87** |
| 341 | 88.23 | 82.16 | 69.76 | 71.19 | **311.34** |
| 342 | 429.37 | 384.67 | 383.34 | 389.49 | **1,586.87** |
| 344 | 40.86 | 38.81 | 40.32 | 38.72 | **158.70** |
| 346 | 755.88 | 715.46 | 715.64 | 722.96 | **2,909.94** |
| 347 | 152.28 | 156.83 | 131.23 | 113.65 | **553.99** |
| 349 | 398.72 | 358.58 | 329.59 | 286.01 | **1,372.90** |
| **FL - FLORIDA - Total** | **5,976.13** | **5,325.75** | **5,080.20** | **4,883.85** | **21,265.92** |

**STATE: GA - GEORGIA**

| REGISTRANT ZIP CODE 3 | 1ST QUARTER | 2ND QUARTER | 3RD QUARTER | 4TH QUARTER | TOTAL GRAMS |
|---|---|---|---|---|---|
| 300 | 395.24 | 463.09 | 376.73 | 345.24 | **1,580.31** |
| 301 | 337.97 | 291.63 | 297.43 | 240.42 | **1,167.45** |
| 302 | 168.61 | 147.87 | 168.70 | 140.28 | **625.45** |
| 303 | 108.93 | 88.50 | 79.75 | 82.34 | **359.52** |
| 304 | 27.70 | 34.52 | 12.67 | 12.40 | **87.29** |
| 305 | 140.02 | 117.49 | 131.76 | 122.84 | **512.11** |
| 306 | 67.62 | 59.24 | 62.71 | 52.10 | **241.67** |
| 307 | 40.46 | 48.84 | 51.61 | 47.86 | **188.77** |
| 308 | 25.34 | 9.01 | 3.21 | 1.43 | **38.98** |
| 309 | 18.73 | 21.95 | 24.27 | 6.07 | **71.01** |
| 310 | 65.84 | 69.54 | 58.52 | 58.61 | **252.51** |
| 312 | 23.19 | 15.43 | 7.00 | 6.24 | **51.88** |
| 313 | 64.41 | 26.76 | 15.43 | 19.89 | **126.50** |
| 314 | 20.52 | 9.81 | 11.33 | 7.14 | **48.80** |
| 315 | 67.93 | 72.97 | 54.24 | 23.19 | **218.34** |
| 316 | 71.99 | 57.45 | 49.07 | 47.10 | **225.61** |
| 317 | 74.89 | 52.46 | 48.57 | 54.51 | **230.43** |
| 318 | 1.78 | 0 | 0.80 | 0.80 | **3.39** |
| 319 | 63.87 | 60.75 | 63.52 | 58.79 | **246.93** |
| 398 | 14.10 | 16.41 | 15.08 | 16.59 | **62.18** |
| **GA - GEORGIA - Total** | **1,799.14** | **1,663.72** | **1,532.40** | **1,343.86** | **6,339.13** |

**STATE: HI - HAWAII**

| REGISTRANT ZIP CODE 3 | 1ST QUARTER | 2ND QUARTER | 3RD QUARTER | 4TH QUARTER | TOTAL GRAMS |
|---|---|---|---|---|---|
| 967 | 79.84 | 81.18 | 68.20 | 71.37 | **300.59** |
| 968 | 56.02 | 78.33 | 47.95 | 59.77 | **242.07** |
| **HI - HAWAII - Total** | **135.87** | **159.51** | **116.15** | **131.14** | **542.66** |

**STATE: IA - IOWA**

| REGISTRANT ZIP CODE 3 | 1ST QUARTER | 2ND QUARTER | 3RD QUARTER | 4TH QUARTER | TOTAL GRAMS |
|---|---|---|---|---|---|
| 500 | 25.16 | 27.30 | 24.98 | 21.59 | **99.02** |
| 501 | 8.30 | 9.10 | 8.47 | 4.37 | **30.24** |
| 502 | 36.67 | 27.57 | 31.40 | 31.85 | **127.48** |
| 503 | 62.71 | 45.68 | 52.01 | 39.97 | **200.37** |
| 504 | 14.99 | 16.06 | 11.33 | 14.01 | **56.38** |
| 505 | 1.61 | 0.54 | 2.68 | 2.14 | **6.96** |
| 506 | 0 | 0.54 | 0 | 0 | **0.54** |
| 508 | 1.20 | 0.80 | 1.61 | 1.20 | **4.82** |
| 510 | 1.07 | 2.14 | 5.71 | 1.87 | **10.79** |
| 511 | 20.34 | 15.88 | 21.59 | 20.16 | **77.97** |
| 512 | 1.07 | 1.61 | 1.61 | 2.14 | **6.42** |
| 513 | 2.14 | 1.61 | 1.61 | 1.61 | **6.96** |
| 514 | 3.12 | 1.78 | 1.78 | 1.78 | **8.47** |
| 515 | 9.28 | 24.89 | 23.82 | 20.52 | **78.50** |
| 516 | 0 | 0.54 | 0.80 | 0.80 | **2.14** |
| 520 | 2.68 | 2.68 | 2.68 | 0 | **8.03** |
| 523 | 2.59 | 4.10 | 3.21 | 3.66 | **13.56** |
| 524 | 0.89 | 1.78 | 2.23 | 0.45 | **5.35** |
| 526 | 0.80 | 2.41 | 0.80 | 2.41 | **6.42** |
| 527 | 1.78 | 11.60 | 6.69 | 8.92 | **28.99** |
| 528 | 2.68 | 3.12 | 2.23 | 2.68 | **10.71** |
| **IA - IOWA - Total** | **199.07** | **201.70** | **207.23** | **182.12** | **790.13** |

**STATE: ID - IDAHO**

| REGISTRANT ZIP CODE 3 | 1ST QUARTER | 2ND QUARTER | 3RD QUARTER | 4TH QUARTER | TOTAL GRAMS |
|---|---|---|---|---|---|
| 832 | 42.37 | 47.64 | 43.71 | 33.99 | **167.71** |
| 833 | 23.28 | 20.07 | 26.67 | 38.63 | **108.66** |
| 834 | 27.83 | 27.39 | 23.28 | 34.70 | **113.21** |
| 835 | 8.03 | 8.03 | 8.92 | 9.81 | **34.79** |
| 836 | 192.56 | 155.31 | 165.35 | 162.01 | **675.23** |
| 837 | 167.49 | 154.15 | 127.39 | 105.36 | **554.40** |
| 838 | 33.45 | 24.98 | 40.14 | 24.00 | **122.57** |
| **ID - IDAHO - Total** | **495.03** | **437.58** | **435.48** | **408.49** | **1,776.57** |

**STATE: IL - ILLINOIS**

| REGISTRANT ZIP CODE 3 | 1ST QUARTER | 2ND QUARTER | 3RD QUARTER | 4TH QUARTER | TOTAL GRAMS |
|---|---|---|---|---|---|
| 600 | 431.78 | 311.08 | 323.56 | 335.70 | **1,402.11** |
| 601 | 176.10 | 137.29 | 141.93 | 156.92 | **612.25** |
| 602 | 32.34 | 23.64 | 28.37 | 18.20 | **102.55** |
| 603 | 15.79 | 18.02 | 14.45 | 17.49 | **65.75** |
| 604 | 217.09 | 165.35 | 179.22 | 178.20 | **739.86** |
| 605 | 152.77 | 108.17 | 105.58 | 102.10 | **468.62** |
| 606 | 105.89 | 79.93 | 86.53 | 90.37 | **362.73** |
| 607 | 4.64 | 3.66 | 2.50 | 0.80 | **11.60** |
| 608 | 0 | 0 | 0 | 0.89 | **0.89** |
| 609 | 5.62 | 6.29 | 5.71 | 2.99 | **20.61** |
| 610 | 18.24 | 11.73 | 9.37 | 8.74 | **48.08** |
| 611 | 2.94 | 2.14 | 5.89 | 2.94 | **13.92** |
| 612 | 1.12 | 2.99 | 3.57 | 3.52 | **11.20** |
| 613 | 23.64 | 19.45 | 28.46 | 20.34 | **91.89** |
| 614 | 3.75 | 6.96 | 0 | 0.45 | **11.15** |
| 615 | 8.56 | 9.63 | 3.57 | 3.57 | **25.34** |
| 616 | 4.28 | 5.35 | 6.69 | 7.49 | **23.82** |
| 617 | 10.35 | 7.14 | 10.26 | 7.94 | **35.68** |
| 618 | 14.18 | 14.90 | 4.64 | 12.40 | **46.12** |
| 619 | 1.07 | 0.54 | 0.54 | 0.80 | **2.94** |
| 620 | 6.07 | 9.46 | 9.01 | 7.76 | **32.29** |
| 622 | 9.05 | 10.97 | 4.46 | 2.59 | **27.08** |
| 623 | 23.77 | 24.27 | 18.51 | 21.50 | **88.05** |
| 624 | 5.71 | 4.01 | 2.23 | 2.23 | **14.18** |
| 625 | 7.14 | 9.63 | 9.63 | 12.85 | **39.25** |
| 626 | 4.10 | 0 | 0 | 0 | **4.10** |
| 627 | 7.23 | 7.67 | 7.94 | 11.24 | **34.08** |
| 628 | 0 | 0 | 0.54 | 1.07 | **1.61** |
| 629 | 18.38 | 19.00 | 21.50 | 15.52 | **74.40** |
| **IL - ILLINOIS - Total** | **1,311.61** | **1,019.27** | **1,034.66** | **1,046.61** | **4,412.15** |

**STATE: IN - INDIANA**

| REGISTRANT ZIP CODE 3 | 1ST QUARTER | 2ND QUARTER | 3RD QUARTER | 4TH QUARTER | TOTAL GRAMS |
|---|---|---|---|---|---|
| 460 | 82.43 | 72.53 | 47.28 | 57.09 | **259.33** |
| 461 | 110.98 | 92.15 | 97.33 | 97.68 | **398.14** |
| 462 | 199.07 | 173.65 | 173.60 | 137.29 | **683.62** |
| 463 | 138.01 | 107.85 | 80.65 | 99.29 | **425.80** |
| 464 | 3.57 | 2.41 | 2.05 | 2.41 | **10.44** |
| 465 | 109.37 | 88.14 | 100.72 | 75.47 | **373.70** |
| 466 | 0.89 | 1.78 | 3.48 | 0 | **6.16** |
| 467 | 26.32 | 16.95 | 21.41 | 28.28 | **92.96** |
| 468 | 68.25 | 52.72 | 43.45 | 46.84 | **211.25** |
| 469 | 37.60 | 39.52 | 32.52 | 28.46 | **138.10** |
| 470 | 26.41 | 24.09 | 23.19 | 33.36 | **107.05** |
| 471 | 41.57 | 43.62 | 43.45 | 53.26 | **181.90** |
| 472 | 34.70 | 28.81 | 25.25 | 39.16 | **127.93** |
| 473 | 45.54 | 16.41 | 36.40 | 46.92 | **145.28** |
| 474 | 51.96 | 61.38 | 36.49 | 38.36 | **188.19** |
| 475 | 45.14 | 32.12 | 25.60 | 24.53 | **127.39** |

| | | | | | |
|---|---|---|---|---|---|
| 476 | 8.83 | 9.28 | 3.48 | 4.82 | **26.41** |
| 477 | 86.00 | 65.30 | 61.02 | 52.01 | **264.33** |
| 478 | 46.12 | 29.71 | 24.44 | 25.07 | **125.34** |
| 479 | 15.08 | 10.35 | 14.81 | 13.47 | **53.70** |
| **IN - INDIANA - Total** | **1,177.84** | **968.78** | **896.61** | **903.79** | **3,947.01** |

**STATE: KS - KANSAS**

| REGISTRANT ZIP CODE 3 | 1ST QUARTER | 2ND QUARTER | 3RD QUARTER | 4TH QUARTER | TOTAL GRAMS |
|---|---|---|---|---|---|
| 660 | 168.03 | 153.71 | 154.56 | 104.42 | **580.71** |
| 661 | 22.93 | 34.79 | 31.13 | 28.64 | **117.49** |
| 662 | 84.08 | 55.44 | 76.45 | 60.35 | **276.33** |
| 664 | 15.79 | 11.42 | 14.10 | 9.37 | **50.67** |
| 665 | 0.54 | 1.43 | 0.54 | 0.54 | **3.03** |
| 666 | 20.07 | 20.43 | 17.75 | 23.64 | **81.89** |
| 667 | 10.44 | 13.29 | 13.02 | 14.45 | **51.21** |
| 668 | 2.14 | 1.07 | 1.61 | 1.61 | **6.42** |
| 669 | 4.46 | 3.57 | 2.68 | 0 | **10.71** |
| 670 | 26.85 | 21.50 | 19.72 | 22.84 | **90.90** |
| 671 | 12.67 | 12.67 | 10.53 | 9.37 | **45.23** |
| 672 | 50.49 | 52.81 | 44.34 | 51.56 | **199.21** |
| 673 | 0 | 0 | 0.54 | 0 | **0.54** |
| 674 | 20.07 | 22.30 | 23.02 | 15.17 | **80.56** |
| 675 | 27.12 | 21.23 | 20.43 | 22.66 | **91.44** |
| 676 | 17.49 | 13.92 | 25.16 | 28.73 | **85.28** |
| 678 | 15.43 | 12.40 | 14.54 | 8.03 | **50.40** |
| 679 | 3.57 | 1.78 | 3.57 | 0 | **8.92** |
| **KS - KANSAS - Total** | **502.16** | **453.77** | **473.66** | **401.36** | **1,830.95** |

**STATE: KY - KENTUCKY**

| REGISTRANT ZIP CODE 3 | 1ST QUARTER | 2ND QUARTER | 3RD QUARTER | 4TH QUARTER | TOTAL GRAMS |
|---|---|---|---|---|---|
| 400 | 15.34 | 16.41 | 26.94 | 35.33 | **94.03** |
| 401 | 13.56 | 19.80 | 10.79 | 8.92 | **53.08** |
| 402 | 121.06 | 104.02 | 108.66 | 120.17 | **453.90** |
| 403 | 138.28 | 86.36 | 88.72 | 100.63 | **413.98** |
| 404 | 52.37 | 42.78 | 40.23 | 43.85 | **179.22** |
| 405 | 88.94 | 123.60 | 96.61 | 98.62 | **407.78** |
| 406 | 7.05 | 4.73 | 7.49 | 1.47 | **20.74** |
| 407 | 205.67 | 183.19 | 100.63 | 53.35 | **542.84** |
| 408 | 6.24 | 5.35 | 5.35 | 6.24 | **23.19** |
| 409 | 150.01 | 160.93 | 78.42 | 79.40 | **468.75** |
| 410 | 72.84 | 59.86 | 53.53 | 52.28 | **238.50** |
| 411 | 52.28 | 35.51 | 20.43 | 19.27 | **127.48** |
| 412 | 6.16 | 1.61 | 1.07 | 4.28 | **13.11** |
| 413 | 1.69 | 2.32 | 1.78 | 1.78 | **7.58** |
| 414 | 1.87 | 2.14 | 1.87 | 1.74 | **7.63** |
| 415 | 49.96 | 54.60 | 37.56 | 42.37 | **184.49** |
| 416 | 134.26 | 121.37 | 114.01 | 90.41 | **460.06** |
| 417 | 9.19 | 12.18 | 5.08 | 6.29 | **32.74** |
| 420 | 18.38 | 20.61 | 12.22 | 14.10 | **65.30** |
| 421 | 41.48 | 38.58 | 40.68 | 55.53 | **176.28** |
| 422 | 50.76 | 40.14 | 33.19 | 35.73 | **159.82** |
| 423 | 7.58 | 3.75 | 4.37 | 6.07 | **21.77** |
| 424 | 15.70 | 15.34 | 9.28 | 13.29 | **53.62** |
| 425 | 40.10 | 33.77 | 16.06 | 10.53 | **100.45** |
| 426 | 16.77 | 12.58 | 12.04 | 11.37 | **52.77** |
| 427 | 37.91 | 37.56 | 42.91 | 38.81 | **157.19** |
| **KY - KENTUCKY - Total** | **1,355.46** | **1,239.08** | **969.94** | **951.83** | **4,516.30** |

**STATE: LA - LOUISIANA**

| REGISTRANT ZIP CODE 3 | 1ST QUARTER | 2ND QUARTER | 3RD QUARTER | 4TH QUARTER | TOTAL GRAMS |
|---|---|---|---|---|---|
| 700 | 309.02 | 169.32 | 208.62 | 203.58 | **890.54** |
| 701 | 84.21 | 78.42 | 79.84 | 64.14 | **306.61** |
| 703 | 67.53 | 60.75 | 58.79 | 43.00 | **230.07** |

| | | | | | |
|---|---|---|---|---|---|
| 704 | 434.45 | 399.04 | 364.69 | 305.46 | **1,503.63** |
| 705 | 121.86 | 123.91 | 118.78 | 103.26 | **467.82** |
| 706 | 13.29 | 22.93 | 14.99 | 20.70 | **71.90** |
| 707 | 581.65 | 536.24 | 384.05 | 325.79 | **1,827.73** |
| 708 | 387.17 | 304.03 | 402.25 | 375.48 | **1,468.93** |
| 710 | 22.75 | 26.85 | 29.53 | 25.60 | **104.73** |
| 711 | 105.31 | 89.08 | 100.05 | 95.77 | **390.20** |
| 712 | 5.71 | 11.60 | 2.14 | 5.98 | **25.42** |
| 713 | 17.22 | 13.47 | 18.56 | 14.90 | **64.14** |
| 714 | 4.46 | 4.46 | 1.78 | 2.68 | **13.38** |
| **LA - LOUISIANA - Total** | **2,154.64** | **1,840.09** | **1,784.07** | **1,586.33** | **7,365.13** |

**STATE: MA - MASSACHUSETTS**

| REGISTRANT ZIP CODE 3 | 1ST QUARTER | 2ND QUARTER | 3RD QUARTER | 4TH QUARTER | TOTAL GRAMS |
|---|---|---|---|---|---|
| 010 | 40.59 | 35.06 | 34.44 | 42.20 | **152.28** |
| 011 | 11.60 | 7.67 | 19.72 | 6.33 | **45.32** |
| 012 | 21.41 | 19.27 | 23.55 | 30.87 | **95.10** |
| 013 | 10.44 | 11.51 | 7.76 | 13.11 | **42.82** |
| 014 | 0.45 | 0.98 | 2.32 | 1.16 | **4.91** |
| 015 | 46.92 | 44.34 | 48.35 | 47.91 | **187.52** |
| 016 | 4.46 | 2.68 | 3.57 | 1.78 | **12.49** |
| 017 | 2.32 | 0.89 | 3.57 | 3.57 | **10.35** |
| 018 | 120.39 | 145.06 | 139.21 | 123.20 | **527.86** |
| 019 | 52.41 | 62.36 | 53.62 | 61.29 | **229.67** |
| 020 | 37.38 | 35.06 | 20.96 | 33.28 | **126.68** |
| 021 | 38.27 | 27.92 | 40.86 | 32.38 | **139.44** |
| 022 | 20.52 | 18.20 | 33.19 | 19.63 | **91.53** |
| 023 | 19.72 | 10.97 | 12.13 | 14.63 | **57.45** |
| 024 | 19.80 | 18.64 | 13.29 | 15.43 | **67.18** |
| 025 | 0.89 | 0 | 0 | 0 | **0.89** |
| 026 | 10.26 | 8.65 | 13.20 | 13.29 | **45.41** |
| 027 | 222.94 | 192.34 | 170.57 | 187.25 | **773.09** |
| **MA - MASSACHUSETTS - Total** | **680.76** | **641.60** | **640.30** | **647.31** | **2,609.97** |

**STATE: MD - MARYLAND**

| REGISTRANT ZIP CODE 3 | 1ST QUARTER | 2ND QUARTER | 3RD QUARTER | 4TH QUARTER | TOTAL GRAMS |
|---|---|---|---|---|---|
| 206 | 281.68 | 202.46 | 203.89 | 202.46 | **890.49** |
| 207 | 391.72 | 309.56 | 283.11 | 274.95 | **1,259.33** |
| 208 | 285.65 | 222.36 | 228.11 | 195.28 | **931.40** |
| 209 | 57.72 | 72.22 | 47.28 | 46.03 | **223.25** |
| 210 | 543.11 | 423.48 | 368.70 | 348.94 | **1,684.24** |
| 211 | 207.28 | 196.13 | 200.14 | 211.29 | **814.84** |
| 212 | 289.40 | 215.35 | 248.94 | 243.28 | **996.97** |
| 214 | 68.78 | 56.16 | 52.19 | 42.55 | **219.68** |
| 215 | 100.54 | 100.54 | 80.56 | 83.63 | **365.27** |
| 216 | 89.43 | 55.62 | 57.90 | 77.21 | **280.16** |
| 217 | 531.96 | 434.01 | 368.08 | 344.84 | **1,678.89** |
| 218 | 36.31 | 40.59 | 45.27 | 31.62 | **153.80** |
| 219 | 70.48 | 45.23 | 25.60 | 36.40 | **177.71** |
| **MD - MARYLAND - Total** | **2,954.06** | **2,373.70** | **2,209.78** | **2,138.50** | **9,676.03** |

**STATE: ME - MAINE**

| REGISTRANT ZIP CODE 3 | 1ST QUARTER | 2ND QUARTER | 3RD QUARTER | 4TH QUARTER | TOTAL GRAMS |
|---|---|---|---|---|---|
| 039 | 0 | 0 | 0 | 0.27 | **0.27** |
| 040 | 40.41 | 39.83 | 39.97 | 46.12 | **166.33** |
| 041 | 2.68 | 4.82 | 7.58 | 7.23 | **22.30** |
| 042 | 38.81 | 40.77 | 45.32 | 34.52 | **159.42** |
| 043 | 97.11 | 98.93 | 81.54 | 81.45 | **359.03** |
| 045 | 17.22 | 6.24 | 5.80 | 5.89 | **35.15** |
| 046 | 2.59 | 4.37 | 2.23 | 1.34 | **10.53** |
| 048 | 3.12 | 2.50 | 4.91 | 4.64 | **15.17** |
| 049 | 45.41 | 44.78 | 40.06 | 37.29 | **167.54** |
| **ME - MAINE - Total** | **247.33** | **242.25** | **227.40** | **218.74** | **935.72** |

STATE: MI - MICHIGAN

| REGISTRANT ZIP CODE 3 | 1ST QUARTER | 2ND QUARTER | 3RD QUARTER | 4TH QUARTER | TOTAL GRAMS |
|---|---|---|---|---|---|
| 480 | 983.58 | 941.43 | 757.84 | 859.81 | **3,542.66** |
| 481 | 1,326.82 | 1,198.27 | 813.24 | 807.08 | **4,145.41** |
| 482 | 1,798.96 | 1,608.23 | 1,317.27 | 1,038.67 | **5,763.14** |
| 483 | 547.39 | 510.10 | 446.05 | 382.44 | **1,885.99** |
| 484 | 73.60 | 87.07 | 79.22 | 68.42 | **308.31** |
| 485 | 33.72 | 37.83 | 36.04 | 26.41 | **133.99** |
| 486 | 45.68 | 32.29 | 30.87 | 28.37 | **137.20** |
| 487 | 18.56 | 17.40 | 18.56 | 19.27 | **73.78** |
| 488 | 85.73 | 72.44 | 72.13 | 63.16 | **293.46** |
| 489 | 25.51 | 28.99 | 32.12 | 28.01 | **114.63** |
| 490 | 129.85 | 139.70 | 195.24 | 143.23 | **608.01** |
| 491 | 18.56 | 6.60 | 12.40 | 8.39 | **45.94** |
| 492 | 39.61 | 38.98 | 28.19 | 24.09 | **130.87** |
| 493 | 27.39 | 20.07 | 21.86 | 25.69 | **95.01** |
| 494 | 25.78 | 36.80 | 24.98 | 21.23 | **108.79** |
| 495 | 44.61 | 30.51 | 26.23 | 29.71 | **131.05** |
| 496 | 89.03 | 65.66 | 70.83 | 57.01 | **282.53** |
| 497 | 37.38 | 42.64 | 41.39 | 20.79 | **142.20** |
| 498 | 0 | 0.89 | 3.21 | 2.68 | **6.78** |
| **MI - MICHIGAN - Total** | **5,351.75** | **4,915.92** | **4,027.65** | **3,654.44** | **17,949.77** |

STATE: MN - MINNESOTA

| REGISTRANT ZIP CODE 3 | 1ST QUARTER | 2ND QUARTER | 3RD QUARTER | 4TH QUARTER | TOTAL GRAMS |
|---|---|---|---|---|---|
| 550 | 34.79 | 40.50 | 38.81 | 32.12 | **146.22** |
| 551 | 44.52 | 34.70 | 53.62 | 34.26 | **167.09** |
| 553 | 20.96 | 16.24 | 17.93 | 23.37 | **78.50** |
| 554 | 15.70 | 14.10 | 8.39 | 7.76 | **45.94** |
| 556 | 0 | 0 | 0.80 | 0 | **0.80** |
| 557 | 15.08 | 20.43 | 16.41 | 15.17 | **67.09** |
| 558 | 8.56 | 4.28 | 4.28 | 3.21 | **20.34** |
| 559 | 3.12 | 3.57 | 4.46 | 2.68 | **13.83** |
| 560 | 4.55 | 8.21 | 3.93 | 6.96 | **23.64** |
| 563 | 10.35 | 6.60 | 5.71 | 4.82 | **27.48** |
| 564 | 0.45 | 0 | 0.45 | 0 | **0.89** |
| 566 | 0.45 | 0 | 0 | 0 | **0.45** |
| 567 | 6.42 | 6.42 | 8.56 | 4.55 | **25.96** |
| **MN - MINNESOTA - Total** | **164.95** | **155.05** | **163.34** | **134.89** | **618.23** |

STATE: MO - MISSOURI

| REGISTRANT ZIP CODE 3 | 1ST QUARTER | 2ND QUARTER | 3RD QUARTER | 4TH QUARTER | TOTAL GRAMS |
|---|---|---|---|---|---|
| 630 | 62.18 | 71.01 | 74.76 | 51.83 | **259.78** |
| 631 | 610.60 | 655.92 | 587.67 | 603.33 | **2,457.51** |
| 633 | 90.46 | 60.66 | 67.00 | 56.83 | **274.95** |
| 634 | 16.24 | 23.19 | 21.23 | 21.50 | **82.16** |
| 635 | 11.42 | 5.89 | 6.24 | 4.46 | **28.01** |
| 636 | 17.75 | 16.77 | 16.24 | 12.13 | **62.89** |
| 637 | 7.67 | 6.24 | 5.17 | 7.14 | **26.23** |
| 638 | 2.23 | 1.78 | 1.78 | 2.68 | **8.47** |
| 640 | 70.07 | 64.77 | 52.19 | 57.90 | **244.93** |
| 641 | 73.60 | 43.71 | 65.57 | 41.22 | **224.10** |
| 644 | 8.47 | 13.38 | 8.30 | 9.90 | **40.06** |
| 645 | 20.07 | 14.99 | 12.31 | 10.71 | **58.08** |
| 646 | 8.39 | 9.99 | 8.39 | 1.78 | **28.55** |
| 647 | 21.46 | 24.84 | 27.74 | 22.03 | **96.08** |
| 648 | 1.43 | 0 | 0 | 0.71 | **2.14** |
| 650 | 28.15 | 25.56 | 17.13 | 9.72 | **80.56** |
| 651 | 22.03 | 25.29 | 28.95 | 25.74 | **102.01** |
| 652 | 25.07 | 18.82 | 22.48 | 24.09 | **90.46** |
| 653 | 2.68 | 0.89 | 0 | 1.78 | **5.35** |
| 654 | 4.68 | 4.95 | 4.42 | 6.29 | **20.34** |

| | | | | | |
|---|---|---|---|---|---|
| 655 | 2.68 | 2.50 | 1.07 | 0 | **6.24** |
| 656 | 9.28 | 9.37 | 17.22 | 14.63 | **50.49** |
| 657 | 53.44 | 39.30 | 46.43 | 35.59 | **174.76** |
| 658 | 14.45 | 15.79 | 10.17 | 17.66 | **58.08** |
| **MO - MISSOURI - Total** | **1,184.49** | **1,155.63** | **1,102.46** | **1,039.65** | **4,482.22** |

**STATE: MS - MISSISSIPPI**

| REGISTRANT ZIP CODE 3 | 1ST QUARTER | 2ND QUARTER | 3RD QUARTER | 4TH QUARTER | TOTAL GRAMS |
|---|---|---|---|---|---|
| 386 | 37.47 | 31.22 | 43.62 | 31.05 | **143.36** |
| 387 | 2.01 | 1.20 | 1.20 | 1.20 | **5.62** |
| 388 | 110.09 | 104.91 | 102.01 | 96.39 | **413.40** |
| 389 | 8.39 | 6.96 | 9.10 | 8.39 | **32.83** |
| 390 | 7.58 | 4.28 | 5.35 | 5.35 | **22.57** |
| 391 | 27.74 | 27.21 | 34.08 | 30.33 | **119.36** |
| 392 | 38.00 | 35.86 | 39.70 | 34.70 | **148.27** |
| 393 | 28.19 | 27.83 | 27.66 | 21.05 | **104.73** |
| 394 | 38.98 | 31.49 | 36.49 | 31.31 | **138.28** |
| 395 | 286.19 | 252.82 | 273.25 | 280.12 | **1,092.38** |
| 396 | 13.56 | 13.65 | 14.81 | 14.10 | **56.11** |
| 397 | 6.24 | 12.67 | 4.10 | 13.92 | **36.93** |
| **MS - MISSISSIPPI - Total** | **604.44** | **550.11** | **591.37** | **567.91** | **2,313.84** |

**STATE: MT - MONTANA**

| REGISTRANT ZIP CODE 3 | 1ST QUARTER | 2ND QUARTER | 3RD QUARTER | 4TH QUARTER | TOTAL GRAMS |
|---|---|---|---|---|---|
| 590 | 4.46 | 11.60 | 8.03 | 7.58 | **31.67** |
| 591 | 4.91 | 6.24 | 8.03 | 4.46 | **23.64** |
| 593 | 0 | 0 | 0 | 0.89 | **0.89** |
| 594 | 4.37 | 4.28 | 2.23 | 1.61 | **12.49** |
| 595 | 1.34 | 2.68 | 2.41 | 0.54 | **6.96** |
| 597 | 8.03 | 8.65 | 11.24 | 7.67 | **35.59** |
| 598 | 7.94 | 5.17 | 5.62 | 1.43 | **20.16** |
| 599 | 52.28 | 40.14 | 38.45 | 34.44 | **165.31** |
| **MT - MONTANA - Total** | **83.32** | **78.77** | **76.01** | **58.61** | **296.71** |

**STATE: NC - NORTH CAROLINA**

| REGISTRANT ZIP CODE 3 | 1ST QUARTER | 2ND QUARTER | 3RD QUARTER | 4TH QUARTER | TOTAL GRAMS |
|---|---|---|---|---|---|
| 270 | 122.22 | 93.14 | 90.90 | 95.81 | **402.07** |
| 271 | 124.36 | 103.66 | 87.87 | 116.24 | **432.13** |
| 272 | 299.48 | 220.97 | 242.61 | 211.70 | **974.75** |
| 273 | 156.30 | 147.82 | 103.57 | 109.73 | **517.42** |
| 274 | 127.84 | 125.43 | 145.32 | 114.72 | **513.31** |
| 275 | 475.53 | 484.23 | 466.08 | 438.67 | **1,864.52** |
| 276 | 153.89 | 146.75 | 155.23 | 175.39 | **631.25** |
| 277 | 140.24 | 131.41 | 129.62 | 114.37 | **515.63** |
| 278 | 652.75 | 518.62 | 465.85 | 401.27 | **2,038.49** |
| 279 | 95.81 | 77.48 | 71.86 | 67.18 | **312.32** |
| 280 | 782.37 | 648.51 | 619.21 | 665.15 | **2,715.24** |
| 281 | 390.83 | 380.44 | 328.25 | 308.18 | **1,407.69** |
| 282 | 289.31 | 289.13 | 246.17 | 254.83 | **1,079.44** |
| 283 | 330.34 | 282.17 | 267.99 | 244.08 | **1,124.58** |
| 284 | 222.94 | 193.50 | 180.83 | 178.60 | **775.86** |
| 285 | 345.29 | 319.73 | 268.61 | 245.06 | **1,178.69** |
| 286 | 686.56 | 654.36 | 624.43 | 606.67 | **2,572.01** |
| 287 | 192.07 | 170.84 | 192.07 | 158.44 | **713.41** |
| 288 | 22.39 | 18.69 | 16.59 | 17.22 | **74.89** |
| 289 | 64.32 | 58.97 | 32.29 | 34.70 | **190.28** |
| **NC - NORTH CAROLINA - Total** | **5,674.83** | **5,065.83** | **4,735.36** | **4,557.99** | **20,034.01** |

**STATE: ND - NORTH DAKOTA**

| REGISTRANT ZIP CODE 3 | 1ST QUARTER | 2ND QUARTER | 3RD QUARTER | 4TH QUARTER | TOTAL GRAMS |
|---|---|---|---|---|---|
| 581 | 3.57 | 3.84 | 1.34 | 3.66 | **12.40** |
| 583 | 0 | 0 | 1.07 | 0 | **1.07** |
| 585 | 29.62 | 24.71 | 29.08 | 37.47 | **120.88** |
| 586 | 1.78 | 1.78 | 2.68 | 1.78 | **8.03** |

| | | | | | |
|---|---|---|---|---|---|
| 587 | 9.28 | 4.55 | 9.28 | 4.82 | **27.92** |
| 588 | 0 | 0 | 6.78 | 21.77 | **28.55** |
| **ND - NORTH DAKOTA - Total** | **44.25** | **34.88** | **50.23** | **69.49** | **198.85** |
| **STATE: NE - NEBRASKA** | | | | | |
| REGISTRANT ZIP CODE 3 | 1ST QUARTER | 2ND QUARTER | 3RD QUARTER | 4TH QUARTER | **TOTAL GRAMS** |
| 680 | 15.34 | 22.17 | 12.00 | 12.76 | **62.27** |
| 681 | 130.69 | 110.40 | 121.06 | 113.74 | **475.89** |
| 684 | 12.62 | 11.78 | 9.90 | 10.71 | **45.01** |
| 685 | 72.17 | 63.12 | 61.55 | 54.20 | **251.04** |
| 686 | 23.46 | 14.72 | 11.11 | 9.37 | **58.66** |
| 687 | 27.21 | 20.79 | 21.23 | 23.55 | **92.78** |
| 688 | 16.28 | 13.02 | 15.79 | 8.65 | **53.75** |
| 689 | 0.98 | 0 | 0 | 0 | **0.98** |
| 690 | 1.78 | 0.89 | 0 | 0 | **2.68** |
| 691 | 1.61 | 1.61 | 1.61 | 1.61 | **6.42** |
| 693 | 1.78 | 6.24 | 4.64 | 7.49 | **20.16** |
| **NE - NEBRASKA - Total** | **303.94** | **264.73** | **258.89** | **242.07** | **1,069.63** |
| **STATE: NH - NEW HAMPSHIRE** | | | | | |
| REGISTRANT ZIP CODE 3 | 1ST QUARTER | 2ND QUARTER | 3RD QUARTER | 4TH QUARTER | **TOTAL GRAMS** |
| 030 | 49.69 | 35.33 | 40.64 | 22.21 | **147.87** |
| 031 | 27.92 | 16.24 | 22.35 | 20.25 | **86.76** |
| 032 | 20.96 | 17.13 | 30.06 | 25.25 | **93.40** |
| 033 | 29.71 | 28.90 | 23.64 | 22.39 | **104.64** |
| 034 | 4.91 | 0 | 1.34 | 1.34 | **7.58** |
| 035 | 0 | 1.61 | 1.87 | 4.01 | **7.49** |
| 038 | 43.45 | 58.61 | 51.92 | 53.70 | **207.68** |
| **NH - NEW HAMPSHIRE - Total** | **176.64** | **157.81** | **171.82** | **149.16** | **655.43** |
| **STATE: NJ - NEW JERSEY** | | | | | |
| REGISTRANT ZIP CODE 3 | 1ST QUARTER | 2ND QUARTER | 3RD QUARTER | 4TH QUARTER | **TOTAL GRAMS** |
| 070 | 287.70 | 269.15 | 252.95 | 220.21 | **1,030.02** |
| 071 | 19.89 | 14.10 | 11.42 | 11.15 | **56.56** |
| 072 | 13.38 | 13.65 | 14.99 | 13.92 | **55.93** |
| 073 | 12.13 | 14.10 | 9.10 | 12.13 | **47.46** |
| 074 | 184.84 | 134.53 | 145.06 | 149.11 | **613.54** |
| 075 | 39.70 | 18.73 | 9.37 | 4.10 | **71.90** |
| 076 | 131.94 | 121.77 | 135.33 | 105.00 | **494.04** |
| 077 | 174.94 | 157.28 | 143.27 | 145.14 | **620.63** |
| 078 | 54.51 | 60.31 | 62.45 | 51.92 | **229.18** |
| 079 | 36.67 | 29.89 | 30.24 | 37.78 | **134.57** |
| 080 | 453.77 | 406.57 | 431.82 | 443.24 | **1,735.40** |
| 081 | 17.75 | 5.89 | 12.13 | 7.76 | **43.53** |
| 082 | 46.21 | 45.14 | 53.79 | 48.57 | **193.72** |
| 083 | 32.47 | 30.78 | 35.51 | 30.60 | **129.35** |
| 084 | 1.96 | 2.14 | 0 | 0 | **4.10** |
| 085 | 72.53 | 67.09 | 33.90 | 53.88 | **227.40** |
| 086 | 35.51 | 28.10 | 37.02 | 37.29 | **137.92** |
| 087 | 143.90 | 136.94 | 139.70 | 108.84 | **529.37** |
| 088 | 267.59 | 229.80 | 219.28 | 199.21 | **915.87** |
| 089 | 16.15 | 13.20 | 12.85 | 18.47 | **60.66** |
| **NJ - NEW JERSEY - Total** | **2,043.53** | **1,799.14** | **1,790.18** | **1,698.34** | **7,331.19** |
| **STATE: NM - NEW MEXICO** | | | | | |
| REGISTRANT ZIP CODE 3 | 1ST QUARTER | 2ND QUARTER | 3RD QUARTER | 4TH QUARTER | **TOTAL GRAMS** |
| 870 | 28.37 | 38.36 | 24.00 | 35.19 | **125.92** |
| 871 | 116.06 | 108.30 | 112.14 | 107.23 | **443.73** |
| 873 | 6.24 | 0 | 3.21 | 4.82 | **14.27** |
| 874 | 1.87 | 2.01 | 1.20 | 4.06 | **9.14** |
| 875 | 21.59 | 35.42 | 17.49 | 22.66 | **97.15** |
| 877 | 8.56 | 5.35 | 4.73 | 4.46 | **23.11** |
| 879 | 4.46 | 0.89 | 1.78 | 2.68 | **9.81** |
| 880 | 11.51 | 10.53 | 14.36 | 13.29 | **49.69** |

| | | | | | |
|---|---|---|---|---|---|
| 881 | 0.45 | 0.27 | 0 | 0 | **0.71** |
| 882 | 12.58 | 12.98 | 17.89 | 16.19 | **59.64** |
| 883 | 15.97 | 13.38 | 14.36 | 6.07 | **49.78** |
| **NM - NEW MEXICO - Total** | **227.66** | **227.49** | **211.16** | **216.65** | **882.96** |

**STATE: NV - NEVADA**

| REGISTRANT ZIP CODE 3 | 1ST QUARTER | 2ND QUARTER | 3RD QUARTER | 4TH QUARTER | TOTAL GRAMS |
|---|---|---|---|---|---|
| 890 | 298.36 | 226.50 | 237.57 | 235.96 | **998.39** |
| 891 | 508.72 | 416.66 | 388.42 | 422.99 | **1,736.78** |
| 894 | 26.05 | 20.70 | 16.41 | 13.74 | **76.90** |
| 895 | 34.88 | 26.32 | 30.51 | 20.88 | **112.58** |
| 897 | 6.24 | 6.78 | 7.14 | 4.01 | **24.18** |
| 898 | 2.68 | 0 | 0 | 0.27 | **2.94** |
| **NV - NEVADA - Total** | **876.93** | **696.95** | **680.05** | **697.85** | **2,951.78** |

**STATE: NY - NEW YORK**

| REGISTRANT ZIP CODE 3 | 1ST QUARTER | 2ND QUARTER | 3RD QUARTER | 4TH QUARTER | TOTAL GRAMS |
|---|---|---|---|---|---|
| 100 | 334.45 | 220.44 | 228.11 | 194.03 | **977.03** |
| 101 | 14.63 | 24.53 | 34.52 | 31.94 | **105.62** |
| 102 | 0.89 | 0.89 | 0 | 0 | **1.78** |
| 103 | 320.26 | 202.15 | 196.84 | 198.63 | **917.88** |
| 104 | 175.57 | 168.79 | 157.90 | 166.38 | **668.63** |
| 105 | 119.32 | 107.23 | 118.65 | 92.20 | **437.40** |
| 106 | 0 | 8.47 | 0 | 0.89 | **9.37** |
| 107 | 30.24 | 24.89 | 22.12 | 19.09 | **96.35** |
| 108 | 25.34 | 13.38 | 18.20 | 24.09 | **81.00** |
| 109 | 168.92 | 122.22 | 127.93 | 126.77 | **545.83** |
| 110 | 43.80 | 50.05 | 46.84 | 15.43 | **156.12** |
| 111 | 38.54 | 1.96 | 1.78 | 0.89 | **43.18** |
| 112 | 522.19 | 490.12 | 429.81 | 449.26 | **1,891.39** |
| 113 | 174.63 | 150.14 | 105.04 | 105.98 | **535.80** |
| 114 | 92.15 | 37.29 | 20.25 | 21.86 | **171.55** |
| 115 | 91.08 | 66.82 | 67.44 | 83.68 | **309.02** |
| 116 | 18.02 | 23.55 | 17.66 | 37.11 | **96.35** |
| 117 | 394.22 | 290.96 | 289.93 | 254.16 | **1,229.27** |
| 118 | 21.95 | 10.26 | 18.91 | 11.51 | **62.63** |
| 119 | 50.58 | 31.58 | 46.48 | 39.43 | **168.07** |
| 120 | 48.44 | 32.52 | 41.66 | 32.07 | **154.69** |
| 121 | 12.49 | 5.35 | 10.62 | 12.58 | **41.04** |
| 122 | 11.42 | 9.10 | 8.56 | 10.17 | **39.25** |
| 123 | 21.05 | 22.39 | 8.92 | 16.68 | **69.05** |
| 124 | 134.13 | 109.95 | 90.50 | 87.34 | **421.92** |
| 125 | 153.89 | 162.01 | 182.79 | 183.59 | **682.28** |
| 126 | 128.82 | 86.27 | 130.43 | 97.77 | **443.28** |
| 127 | 24.80 | 40.14 | 17.13 | 16.77 | **98.84** |
| 128 | 146.39 | 130.51 | 122.66 | 127.57 | **527.14** |
| 129 | 134.31 | 106.12 | 121.19 | 110.71 | **472.32** |
| 130 | 200.99 | 186.27 | 175.88 | 199.25 | **762.39** |
| 131 | 63.03 | 52.14 | 46.30 | 43.94 | **205.41** |
| 132 | 39.83 | 37.20 | 33.81 | 39.61 | **150.45** |
| 133 | 30.96 | 26.32 | 30.24 | 31.80 | **119.32** |
| 134 | 44.03 | 33.59 | 21.46 | 33.36 | **132.43** |
| 135 | 11.51 | 2.68 | 6.96 | 1.07 | **22.21** |
| 136 | 85.86 | 80.20 | 85.64 | 80.38 | **332.08** |
| 137 | 56.87 | 28.46 | 33.28 | 37.02 | **155.63** |
| 138 | 108.21 | 119.10 | 111.24 | 102.06 | **440.61** |
| 139 | 83.59 | 71.64 | 85.06 | 97.42 | **337.70** |
| 140 | 298.81 | 332.00 | 394.31 | 347.52 | **1,372.63** |
| 141 | 159.11 | 184.00 | 188.19 | 214.01 | **745.30** |
| 142 | 379.95 | 359.56 | 341.99 | 359.25 | **1,440.74** |
| 143 | 151.34 | 119.32 | 108.26 | 128.37 | **507.29** |
| 144 | 48.44 | 45.14 | 64.77 | 40.77 | **199.12** |

| | | | | | |
|---|---|---|---|---|---|
| 145 | 15.92 | 16.77 | 33.54 | 23.46 | **89.70** |
| 146 | 49.96 | 26.05 | 31.05 | 20.43 | **127.48** |
| 147 | 39.21 | 33.28 | 31.31 | 32.03 | **135.82** |
| 148 | 100.18 | 102.77 | 95.32 | 75.34 | **373.61** |
| 149 | 8.56 | 12.49 | 9.72 | 9.81 | **40.59** |
| **NY - NEW YORK - Total** | **5,428.87** | **4,619.03** | **4,611.22** | **4,485.48** | **19,144.60** |

**STATE: OH - OHIO**

| REGISTRANT ZIP CODE 3 | 1ST QUARTER | 2ND QUARTER | 3RD QUARTER | 4TH QUARTER | TOTAL GRAMS |
|---|---|---|---|---|---|
| 430 | 116.02 | 89.39 | 87.34 | 88.90 | **381.64** |
| 431 | 73.06 | 72.71 | 61.91 | 61.47 | **269.15** |
| 432 | 47.14 | 67.53 | 65.93 | 54.51 | **235.11** |
| 433 | 22.30 | 39.97 | 25.96 | 28.37 | **116.60** |
| 434 | 40.59 | 34.35 | 35.06 | 28.64 | **138.63** |
| 435 | 55.00 | 46.39 | 50.58 | 40.50 | **192.47** |
| 436 | 146.66 | 112.81 | 94.38 | 78.73 | **432.58** |
| 437 | 4.82 | 9.99 | 11.78 | 10.71 | **37.29** |
| 438 | 10.35 | 10.53 | 12.49 | 9.55 | **42.91** |
| 439 | 39.61 | 37.56 | 28.10 | 31.58 | **136.85** |
| 440 | 111.11 | 72.84 | 62.22 | 71.28 | **317.45** |
| 441 | 42.46 | 28.37 | 35.33 | 28.01 | **134.17** |
| 442 | 83.95 | 78.42 | 63.52 | 62.27 | **288.15** |
| 443 | 13.65 | 23.46 | 27.21 | 27.92 | **92.24** |
| 444 | 61.33 | 34.26 | 51.16 | 61.24 | **207.99** |
| 445 | 44.74 | 34.66 | 19.98 | 17.04 | **116.42** |
| 446 | 123.65 | 109.19 | 106.78 | 97.60 | **437.22** |
| 447 | 52.10 | 62.49 | 55.93 | 52.10 | **222.62** |
| 448 | 21.68 | 15.88 | 28.01 | 17.75 | **83.32** |
| 449 | 4.46 | 3.57 | 3.57 | 6.24 | **17.84** |
| 450 | 246.35 | 280.92 | 253.85 | 212.72 | **993.84** |
| 451 | 119.63 | 94.83 | 93.76 | 105.71 | **413.93** |
| 452 | 99.74 | 93.00 | 91.26 | 87.74 | **371.74** |
| 453 | 73.82 | 74.89 | 74.49 | 62.89 | **286.10** |
| 454 | 96.03 | 69.58 | 82.16 | 69.41 | **317.19** |
| 455 | 15.61 | 12.13 | 16.15 | 3.84 | **47.73** |
| 456 | 350.60 | 289.22 | 281.81 | 283.87 | **1,205.49** |
| 457 | 37.29 | 32.92 | 39.70 | 35.15 | **145.06** |
| 458 | 7.40 | 5.89 | 4.82 | 5.35 | **23.46** |
| **OH - OHIO - Total** | **2,161.15** | **1,937.73** | **1,865.25** | **1,741.07** | **7,705.20** |

**STATE: OK - OKLAHOMA**

| REGISTRANT ZIP CODE 3 | 1ST QUARTER | 2ND QUARTER | 3RD QUARTER | 4TH QUARTER | TOTAL GRAMS |
|---|---|---|---|---|---|
| 730 | 241.13 | 236.14 | 141.22 | 128.64 | **747.13** |
| 731 | 170.97 | 158.84 | 85.69 | 79.58 | **495.07** |
| 734 | 17.49 | 8.83 | 12.85 | 10.17 | **49.33** |
| 735 | 23.82 | 29.97 | 30.06 | 30.96 | **114.81** |
| 737 | 15.43 | 17.22 | 16.86 | 12.49 | **62.00** |
| 738 | 5.35 | 3.57 | 1.87 | 1.87 | **12.67** |
| 740 | 81.40 | 72.53 | 77.66 | 70.21 | **301.80** |
| 741 | 69.54 | 54.37 | 59.95 | 49.38 | **233.24** |
| 743 | 19.00 | 16.15 | 16.59 | 18.02 | **69.76** |
| 744 | 22.39 | 19.80 | 14.63 | 15.97 | **72.80** |
| 745 | 6.24 | 3.57 | 4.10 | 3.57 | **17.49** |
| 746 | 2.77 | 3.93 | 2.59 | 3.21 | **12.49** |
| 747 | 8.30 | 6.33 | 7.23 | 6.87 | **28.73** |
| 748 | 22.12 | 17.04 | 5.53 | 21.95 | **66.64** |
| 749 | 11.15 | 10.08 | 6.42 | 8.03 | **35.68** |
| **OK - OKLAHOMA - Total** | **717.11** | **658.37** | **483.25** | **460.90** | **2,319.64** |

**STATE: OR - OREGON**

| REGISTRANT ZIP CODE 3 | 1ST QUARTER | 2ND QUARTER | 3RD QUARTER | 4TH QUARTER | TOTAL GRAMS |
|---|---|---|---|---|---|
| 970 | 106.83 | 97.42 | 103.89 | 86.04 | **394.17** |
| 971 | 37.38 | 8.12 | 12.67 | 17.75 | **75.92** |

| | | | | | |
|---|---|---|---|---|---|
| 972 | 86.71 | 99.83 | 93.85 | 83.37 | **363.75** |
| 973 | 66.82 | 47.55 | 51.92 | 50.23 | **216.51** |
| 974 | 114.01 | 103.97 | 87.69 | 94.87 | **400.55** |
| 975 | 30.69 | 22.84 | 42.73 | 29.89 | **126.14** |
| 976 | 3.21 | 0 | 0 | 0 | **3.21** |
| 977 | 61.47 | 72.35 | 55.85 | 58.16 | **247.83** |
| 978 | 13.11 | 14.90 | 6.07 | 5.00 | **39.07** |
| 979 | 4.19 | 4.28 | 1.07 | 1.78 | **11.33** |
| **OR - OREGON - Total** | **524.42** | **471.25** | **455.73** | **427.09** | **1,878.49** |

**STATE: PA - PENNSYLVANIA**

| REGISTRANT ZIP CODE 3 | 1ST QUARTER | 2ND QUARTER | 3RD QUARTER | 4TH QUARTER | **TOTAL GRAMS** |
|---|---|---|---|---|---|
| 150 | 273.29 | 199.34 | 190.55 | 179.49 | **842.68** |
| 151 | 263.21 | 215.17 | 172.98 | 201.21 | **852.58** |
| 152 | 638.65 | 629.91 | 626.48 | 592.58 | **2,487.62** |
| 153 | 121.50 | 109.77 | 115.04 | 104.87 | **451.18** |
| 154 | 370.31 | 310.41 | 281.59 | 263.62 | **1,225.92** |
| 155 | 80.82 | 56.20 | 48.35 | 53.53 | **238.90** |
| 156 | 326.95 | 264.91 | 193.32 | 178.96 | **964.14** |
| 157 | 131.45 | 104.64 | 109.51 | 116.73 | **462.33** |
| 158 | 132.12 | 82.70 | 77.17 | 75.47 | **367.46** |
| 159 | 189.84 | 147.02 | 159.46 | 151.66 | **647.98** |
| 160 | 182.48 | 158.53 | 158.26 | 143.18 | **642.45** |
| 161 | 234.67 | 224.50 | 153.08 | 165.40 | **777.64** |
| 162 | 94.83 | 85.51 | 87.92 | 85.91 | **354.16** |
| 163 | 274.14 | 193.72 | 200.23 | 173.65 | **841.74** |
| 164 | 70.30 | 57.76 | 46.92 | 39.30 | **214.28** |
| 165 | 80.78 | 81.89 | 88.36 | 74.18 | **325.22** |
| 166 | 128.86 | 98.31 | 96.79 | 91.35 | **415.32** |
| 167 | 37.91 | 31.31 | 18.64 | 14.45 | **102.32** |
| 168 | 16.77 | 12.04 | 15.61 | 10.62 | **55.04** |
| 169 | 0.89 | 0.89 | 0.89 | 1.78 | **4.46** |
| 170 | 192.38 | 173.29 | 160.22 | 162.36 | **688.26** |
| 171 | 53.62 | 50.85 | 47.19 | 50.40 | **202.06** |
| 172 | 86.13 | 67.75 | 80.38 | 73.60 | **307.86** |
| 173 | 98.35 | 73.42 | 85.55 | 73.78 | **331.10** |
| 174 | 75.83 | 43.18 | 55.67 | 50.23 | **224.90** |
| 175 | 95.54 | 77.26 | 99.56 | 79.22 | **351.58** |
| 176 | 116.60 | 84.70 | 92.29 | 63.52 | **357.11** |
| 177 | 10.21 | 0.80 | 0.27 | 0.45 | **11.73** |
| 178 | 9.37 | 10.88 | 12.22 | 15.30 | **47.77** |
| 179 | 4.82 | 4.01 | 3.39 | 5.00 | **17.22** |
| 180 | 43.53 | 36.04 | 28.64 | 28.01 | **136.22** |
| 181 | 24.31 | 22.12 | 23.06 | 23.77 | **93.27** |
| 182 | 32.83 | 5.35 | 16.33 | 6.24 | **60.75** |
| 183 | 9.72 | 16.50 | 16.15 | 9.28 | **51.65** |
| 184 | 53.44 | 47.64 | 40.14 | 24.27 | **165.48** |
| 185 | 6.87 | 0.89 | 1.96 | 2.68 | **12.40** |
| 186 | 17.31 | 35.24 | 31.67 | 36.49 | **120.70** |
| 187 | 489.23 | 587.45 | 684.78 | 678.09 | **2,439.54** |
| 188 | 13.02 | 11.78 | 4.64 | 4.46 | **33.90** |
| 189 | 100.27 | 75.56 | 85.20 | 79.84 | **340.87** |
| 190 | 395.16 | 286.59 | 310.58 | 308.80 | **1,301.13** |
| 191 | 91.98 | 83.50 | 70.39 | 71.64 | **317.50** |
| 193 | 107.59 | 136.85 | 99.02 | 97.15 | **440.61** |
| 194 | 92.06 | 50.58 | 66.91 | 55.49 | **265.04** |
| 195 | 22.03 | 23.82 | 25.16 | 20.96 | **91.98** |
| 196 | 39.34 | 46.03 | 35.06 | 34.52 | **154.96** |
| **PA - PENNSYLVANIA - Total** | **5,931.35** | **5,116.64** | **5,017.57** | **4,773.45** | **20,839.01** |

**STATE: RI - RHODE ISLAND**

| REGISTRANT ZIP CODE 3 | 1ST QUARTER | 2ND QUARTER | 3RD QUARTER | 4TH QUARTER | **TOTAL GRAMS** |
|---|---|---|---|---|---|

| | | | | | |
|---|---|---|---|---|---|
| 028 | 115.88 | 46.48 | 54.86 | 72.44 | **289.66** |
| 029 | 39.97 | 34.70 | 61.73 | 52.10 | **188.50** |
| **RI - RHODE ISLAND - Total** | **155.85** | **81.18** | **116.60** | **124.54** | **478.17** |

**STATE: SC - SOUTH CAROLINA**

| REGISTRANT ZIP CODE 3 | 1ST QUARTER | 2ND QUARTER | 3RD QUARTER | 4TH QUARTER | TOTAL GRAMS |
|---|---|---|---|---|---|
| 290 | 282.22 | 302.69 | 299.30 | 272.89 | **1,157.10** |
| 291 | 271.29 | 253.27 | 238.90 | 200.72 | **964.18** |
| 292 | 412.55 | 307.33 | 326.15 | 345.87 | **1,391.90** |
| 293 | 35.06 | 22.30 | 31.22 | 24.18 | **112.76** |
| 294 | 426.60 | 367.55 | 416.34 | 385.74 | **1,596.23** |
| 295 | 167.18 | 159.15 | 182.17 | 171.02 | **679.51** |
| 296 | 207.90 | 175.70 | 188.37 | 172.40 | **744.37** |
| 297 | 101.48 | 119.54 | 97.86 | 84.62 | **403.50** |
| 298 | 26.00 | 28.90 | 31.45 | 18.82 | **105.18** |
| 299 | 60.93 | 54.51 | 66.28 | 58.97 | **240.69** |
| **SC - SOUTH CAROLINA - Total** | **1,991.21** | **1,790.94** | **1,878.05** | **1,735.22** | **7,395.42** |

**STATE: SD - SOUTH DAKOTA**

| REGISTRANT ZIP CODE 3 | 1ST QUARTER | 2ND QUARTER | 3RD QUARTER | 4TH QUARTER | TOTAL GRAMS |
|---|---|---|---|---|---|
| 570 | 13.11 | 10.17 | 6.91 | 15.08 | **45.27** |
| 571 | 48.26 | 48.98 | 42.02 | 32.74 | **172.00** |
| 574 | 8.03 | 6.24 | 7.14 | 6.24 | **27.66** |
| 575 | 9.77 | 12.58 | 6.02 | 10.71 | **39.07** |
| 576 | 5.35 | 3.57 | 4.46 | 4.46 | **17.84** |
| 577 | 82.70 | 45.50 | 73.15 | 68.51 | **269.86** |
| **SD - SOUTH DAKOTA - Total** | **167.22** | **127.04** | **139.70** | **137.74** | **571.70** |

**STATE: TN - TENNESSEE**

| REGISTRANT ZIP CODE 3 | 1ST QUARTER | 2ND QUARTER | 3RD QUARTER | 4TH QUARTER | TOTAL GRAMS |
|---|---|---|---|---|---|
| 370 | 564.97 | 469.60 | 437.80 | 411.79 | **1,884.16** |
| 371 | 379.19 | 286.14 | 273.03 | 267.94 | **1,206.30** |
| 372 | 495.38 | 500.07 | 482.22 | 392.39 | **1,870.06** |
| 373 | 443.24 | 412.55 | 389.54 | 388.38 | **1,633.70** |
| 374 | 383.11 | 316.96 | 251.84 | 275.61 | **1,227.53** |
| 376 | 493.42 | 453.90 | 509.08 | 455.95 | **1,912.35** |
| 377 | 1,080.65 | 789.37 | 679.38 | 644.72 | **3,194.12** |
| 378 | 1,228.69 | 909.81 | 833.44 | 861.55 | **3,833.49** |
| 379 | 481.91 | 342.21 | 343.10 | 354.92 | **1,522.15** |
| 380 | 21.32 | 11.78 | 14.27 | 10.44 | **57.81** |
| 381 | 27.30 | 22.21 | 16.99 | 13.69 | **80.20** |
| 382 | 52.86 | 38.18 | 38.63 | 35.33 | **164.99** |
| 383 | 41.08 | 47.28 | 39.16 | 35.15 | **162.67** |
| 384 | 48.84 | 24.35 | 21.68 | 18.82 | **113.70** |
| 385 | 228.65 | 188.41 | 169.50 | 145.81 | **732.37** |
| **TN - TENNESSEE - Total** | **5,970.60** | **4,812.83** | **4,499.66** | **4,312.50** | **19,595.60** |

**STATE: TX - TEXAS**

| REGISTRANT ZIP CODE 3 | 1ST QUARTER | 2ND QUARTER | 3RD QUARTER | 4TH QUARTER | TOTAL GRAMS |
|---|---|---|---|---|---|
| 750 | 399.57 | 371.07 | 338.73 | 313.71 | **1,423.08** |
| 751 | 31.05 | 30.06 | 41.04 | 54.86 | **157.01** |
| 752 | 47.37 | 44.43 | 45.76 | 48.89 | **186.45** |
| 754 | 27.66 | 20.38 | 22.08 | 9.10 | **79.22** |
| 757 | 12.31 | 11.15 | 7.23 | 3.66 | **34.35** |
| 758 | 12.67 | 9.46 | 16.41 | 8.03 | **46.57** |
| 759 | 22.12 | 17.22 | 22.39 | 14.45 | **76.19** |
| 760 | 141.98 | 143.32 | 123.15 | 146.35 | **554.80** |
| 761 | 55.31 | 64.05 | 66.10 | 47.19 | **232.66** |
| 762 | 32.65 | 29.89 | 20.25 | 23.46 | **106.25** |
| 763 | 98.49 | 71.81 | 93.14 | 58.79 | **322.23** |
| 764 | 12.67 | 5.17 | 3.75 | 3.21 | **24.80** |
| 765 | 7.14 | 8.12 | 3.12 | 6.60 | **24.98** |
| 767 | 14.63 | 6.78 | 11.78 | 11.60 | **44.78** |
| 768 | 1.78 | 1.78 | 0.89 | 0 | **4.46** |

| | | | | | |
|---|---|---|---|---|---|
| 769 | 1.20 | 2.10 | 3.12 | 0 | **6.42** |
| 770 | 200.63 | 199.83 | 187.43 | 147.20 | **735.09** |
| 773 | 104.91 | 121.68 | 113.39 | 97.06 | **437.04** |
| 774 | 138.45 | 123.11 | 130.96 | 122.75 | **515.28** |
| 775 | 149.25 | 159.33 | 131.41 | 113.12 | **553.10** |
| 776 | 60.31 | 34.70 | 47.10 | 34.08 | **176.19** |
| 777 | 12.22 | 5.62 | 11.60 | 11.95 | **41.39** |
| 778 | 11.78 | 11.42 | 9.28 | 12.85 | **45.32** |
| 779 | 5.17 | 9.37 | 6.33 | 5.98 | **26.85** |
| 780 | 50.49 | 48.89 | 50.76 | 45.85 | **195.99** |
| 781 | 44.61 | 47.55 | 54.69 | 46.92 | **193.76** |
| 782 | 222.13 | 188.77 | 209.64 | 175.83 | **796.38** |
| 783 | 11.95 | 9.37 | 4.46 | 3.57 | **29.35** |
| 784 | 38.27 | 29.89 | 37.65 | 28.64 | **134.44** |
| 785 | 4.82 | 4.01 | 0.80 | 2.41 | **12.04** |
| 786 | 121.19 | 102.32 | 116.11 | 120.12 | **459.74** |
| 787 | 98.35 | 90.86 | 80.38 | 94.16 | **363.75** |
| 788 | 5.35 | 0 | 0 | 0 | **5.35** |
| 789 | 13.38 | 8.92 | 15.17 | 9.81 | **47.28** |
| 790 | 6.96 | 3.84 | 5.44 | 4.19 | **20.43** |
| 791 | 34.35 | 42.78 | 46.66 | 31.67 | **155.45** |
| 792 | 8.56 | 4.28 | 6.07 | 3.57 | **22.48** |
| 794 | 4.37 | 3.12 | 1.34 | 3.57 | **12.40** |
| 795 | 6.69 | 6.16 | 5.62 | 8.43 | **26.90** |
| 796 | 7.36 | 3.30 | 2.32 | 2.05 | **15.03** |
| 797 | 3.12 | 3.75 | 8.03 | 2.68 | **17.57** |
| 799 | 22.48 | 22.66 | 13.29 | 23.11 | **81.54** |
| **TX - TEXAS - Total** | **2,305.77** | **2,122.31** | **2,114.86** | **1,901.47** | **8,444.40** |

**STATE: UT - UTAH**

| REGISTRANT ZIP CODE 3 | 1ST QUARTER | 2ND QUARTER | 3RD QUARTER | 4TH QUARTER | TOTAL GRAMS |
|---|---|---|---|---|---|
| 840 | 878.18 | 747.67 | 764.13 | 667.51 | **3,057.49** |
| 841 | 280.79 | 249.25 | 249.92 | 210.76 | **990.72** |
| 843 | 88.14 | 87.78 | 89.75 | 84.30 | **349.97** |
| 844 | 209.73 | 185.07 | 155.63 | 139.17 | **689.59** |
| 845 | 51.92 | 54.95 | 58.25 | 45.68 | **210.80** |
| 846 | 216.25 | 174.05 | 190.37 | 182.12 | **762.79** |
| 847 | 193.76 | 191.58 | 197.38 | 183.42 | **766.14** |
| **UT - UTAH - Total** | **1,918.77** | **1,690.35** | **1,705.43** | **1,512.96** | **6,827.51** |

**STATE: VA - VIRGINIA**

| REGISTRANT ZIP CODE 3 | 1ST QUARTER | 2ND QUARTER | 3RD QUARTER | 4TH QUARTER | TOTAL GRAMS |
|---|---|---|---|---|---|
| 201 | 487.44 | 361.79 | 413.80 | 382.44 | **1,645.48** |
| 220 | 421.25 | 293.86 | 262.14 | 230.65 | **1,207.90** |
| 221 | 159.33 | 141.58 | 136.31 | 167.71 | **604.93** |
| 222 | 84.84 | 58.70 | 83.41 | 34.52 | **261.47** |
| 223 | 64.23 | 48.35 | 76.90 | 58.52 | **248.00** |
| 224 | 242.34 | 180.11 | 212.59 | 159.42 | **794.46** |
| 225 | 80.20 | 63.07 | 77.26 | 53.88 | **274.41** |
| 226 | 113.34 | 70.70 | 63.70 | 63.38 | **311.12** |
| 227 | 15.17 | 13.11 | 10.79 | 20.16 | **59.24** |
| 228 | 11.51 | 8.92 | 6.07 | 8.12 | **34.61** |
| 229 | 25.69 | 13.47 | 7.14 | 13.29 | **59.59** |
| 230 | 41.35 | 18.20 | 20.25 | 16.15 | **95.95** |
| 231 | 132.92 | 121.37 | 115.71 | 99.74 | **469.74** |
| 232 | 110.62 | 95.19 | 97.77 | 99.02 | **402.60** |
| 233 | 139.30 | 131.99 | 117.98 | 112.72 | **501.98** |
| 234 | 249.74 | 307.42 | 282.62 | 214.55 | **1,054.33** |
| 235 | 101.79 | 75.11 | 65.79 | 48.80 | **291.49** |
| 236 | 76.50 | 69.32 | 77.79 | 73.51 | **297.11** |
| 237 | 51.83 | 36.04 | 42.73 | 41.57 | **172.18** |
| 238 | 86.09 | 58.79 | 52.46 | 58.43 | **255.77** |

| | | | | | |
|---|---|---|---|---|---|
| 239 | 6.24 | 5.26 | 6.69 | 7.14 | **25.34** |
| 240 | 102.06 | 93.05 | 85.64 | 66.24 | **346.98** |
| 241 | 36.17 | 14.99 | 30.33 | 31.13 | **112.63** |
| 242 | 195.82 | 175.79 | 142.74 | 132.30 | **646.64** |
| 243 | 24.53 | 32.83 | 30.96 | 16.77 | **105.09** |
| 244 | 18.20 | 16.95 | 24.35 | 20.16 | **79.66** |
| 245 | 83.86 | 80.20 | 86.53 | 98.84 | **349.44** |
| 246 | 39.16 | 22.12 | 19.72 | 21.23 | **102.23** |
| **VA - VIRGINIA - Total** | **3,201.52** | **2,608.28** | **2,650.16** | **2,350.42** | **10,810.38** |

**STATE: VT - VERMONT**

| REGISTRANT ZIP CODE 3 | 1ST QUARTER | 2ND QUARTER | 3RD QUARTER | 4TH QUARTER | TOTAL GRAMS |
|---|---|---|---|---|---|
| 052 | 7.14 | 5.35 | 5.35 | 9.81 | **27.66** |
| 054 | 54.42 | 56.65 | 61.02 | 46.12 | **218.21** |
| 056 | 7.49 | 6.42 | 14.45 | 12.31 | **40.68** |
| 057 | 9.63 | 4.82 | 5.89 | 4.82 | **25.16** |
| 058 | 4.82 | 2.23 | 2.23 | 3.75 | **13.02** |
| **VT - VERMONT - Total** | **83.50** | **75.47** | **88.94** | **76.81** | **324.72** |

**STATE: WA - WASHINGTON**

| REGISTRANT ZIP CODE 3 | 1ST QUARTER | 2ND QUARTER | 3RD QUARTER | 4TH QUARTER | TOTAL GRAMS |
|---|---|---|---|---|---|
| 980 | 183.33 | 180.20 | 167.13 | 158.39 | **689.06** |
| 981 | 30.96 | 29.57 | 33.72 | 43.45 | **137.70** |
| 982 | 160.71 | 142.42 | 155.05 | 147.91 | **606.09** |
| 983 | 53.93 | 84.88 | 111.16 | 77.03 | **327.00** |
| 984 | 70.88 | 37.74 | 37.65 | 36.75 | **183.01** |
| 985 | 68.11 | 68.83 | 59.82 | 84.88 | **281.64** |
| 986 | 126.90 | 116.64 | 114.28 | 128.06 | **485.88** |
| 988 | 7.58 | 9.01 | 16.59 | 12.22 | **45.41** |
| 989 | 31.13 | 35.24 | 25.07 | 26.94 | **118.38** |
| 990 | 9.90 | 9.28 | 14.54 | 11.78 | **45.50** |
| 991 | 1.16 | 0.89 | 0.54 | 3.57 | **6.16** |
| 992 | 70.57 | 72.08 | 90.82 | 73.96 | **307.42** |
| 993 | 48.75 | 49.11 | 46.92 | 50.23 | **195.01** |
| 994 | 7.32 | 8.03 | 11.60 | 8.03 | **34.97** |
| **WA - WASHINGTON - Total** | **871.22** | **843.93** | **884.87** | **863.20** | **3,463.22** |

**STATE: WI - WISCONSIN**

| REGISTRANT ZIP CODE 3 | 1ST QUARTER | 2ND QUARTER | 3RD QUARTER | 4TH QUARTER | TOTAL GRAMS |
|---|---|---|---|---|---|
| 530 | 91.98 | 65.39 | 59.24 | 64.41 | **281.01** |
| 531 | 63.25 | 61.91 | 86.98 | 68.16 | **280.30** |
| 532 | 48.08 | 39.34 | 51.92 | 37.56 | **176.90** |
| 534 | 35.24 | 30.87 | 24.35 | 23.55 | **114.01** |
| 535 | 30.87 | 11.33 | 9.10 | 9.90 | **61.20** |
| 537 | 11.60 | 4.01 | 2.23 | 23.64 | **41.48** |
| 538 | 0 | 0 | 0 | 0.54 | **0.54** |
| 539 | 21.23 | 29.08 | 10.08 | 13.47 | **73.87** |
| 540 | 2.94 | 0.80 | 0.80 | 0.80 | **5.35** |
| 541 | 2.77 | 3.03 | 1.61 | 2.85 | **10.26** |
| 542 | 9.81 | 11.15 | 9.72 | 10.44 | **41.13** |
| 543 | 8.70 | 3.48 | 0 | 2.68 | **14.85** |
| 544 | 50.67 | 50.85 | 50.14 | 46.84 | **198.49** |
| 545 | 0.80 | 3.21 | 5.98 | 9.10 | **19.09** |
| 548 | 23.02 | 19.45 | 17.13 | 15.97 | **75.56** |
| 549 | 67.00 | 70.39 | 64.05 | 50.94 | **252.38** |
| **WI - WISCONSIN - Total** | **467.95** | **404.30** | **393.33** | **380.84** | **1,646.42** |

**STATE: WV - WEST VIRGINIA**

| REGISTRANT ZIP CODE 3 | 1ST QUARTER | 2ND QUARTER | 3RD QUARTER | 4TH QUARTER | TOTAL GRAMS |
|---|---|---|---|---|---|
| 247 | 15.70 | 23.37 | 24.62 | 21.95 | **85.64** |
| 248 | 35.51 | 38.45 | 33.72 | 44.07 | **151.75** |
| 249 | 11.95 | 15.52 | 11.42 | 4.64 | **43.53** |
| 250 | 14.10 | 23.19 | 15.70 | 13.11 | **66.10** |
| 251 | 3.21 | 1.07 | 0 | 0 | **4.28** |

| | | | | | |
|---|---|---|---|---|---|
| 252 | 0 | 1.78 | 0 | 1.78 | **3.57** |
| 253 | 57.90 | 56.92 | 54.95 | 58.08 | **227.84** |
| 254 | 170.97 | 153.44 | 132.74 | 115.79 | **572.95** |
| 255 | 51.61 | 39.61 | 33.19 | 23.11 | **147.51** |
| 256 | 12.85 | 2.68 | 3.12 | 4.19 | **22.84** |
| 257 | 10.71 | 14.50 | 13.83 | 15.30 | **54.33** |
| 259 | 2.41 | 4.01 | 2.68 | 1.34 | **10.44** |
| 260 | 49.33 | 41.66 | 46.03 | 34.61 | **171.64** |
| 261 | 8.30 | 3.35 | 2.68 | 2.05 | **16.37** |
| 263 | 9.28 | 0 | 0 | 0 | **9.28** |
| 264 | 3.93 | 1.78 | 2.68 | 2.68 | **11.06** |
| 265 | 7.67 | 3.70 | 3.39 | 2.59 | **17.35** |
| 267 | 30.96 | 28.19 | 13.02 | 21.86 | **94.03** |
| 268 | 3.57 | 0 | 0 | 0 | **3.57** |
| **WV - WEST VIRGINIA - Total** | **499.93** | **453.23** | **393.77** | **367.14** | **1,714.08** |

**STATE: WY - WYOMING**

| REGISTRANT ZIP CODE 3 | 1ST QUARTER | 2ND QUARTER | 3RD QUARTER | 4TH QUARTER | TOTAL GRAMS |
|---|---|---|---|---|---|
| 820 | 13.11 | 11.51 | 14.18 | 7.49 | **46.30** |
| 824 | 0.89 | 2.68 | 8.21 | 3.84 | **15.61** |
| 825 | 11.06 | 19.05 | 11.55 | 19.85 | **61.51** |
| 826 | 11.95 | 10.79 | 7.49 | 6.07 | **36.31** |
| 827 | 14.36 | 10.53 | 8.39 | 3.39 | **36.67** |
| 828 | 6.33 | 6.33 | 2.59 | 4.28 | **19.54** |
| 829 | 6.24 | 7.05 | 7.94 | 8.92 | **30.15** |
| 831 | 0.80 | 0 | 0 | 0 | **0.80** |
| **WY - WYOMING - Total** | **64.77** | **67.93** | **60.35** | **53.84** | **246.89** |

**DRUG: 9780 - TAPENTADOL**

**STATE: AK - ALASKA**

| REGISTRANT ZIP CODE 3 | 1ST QUARTER | 2ND QUARTER | 3RD QUARTER | 4TH QUARTER | TOTAL GRAMS |
|---|---|---|---|---|---|
| 995 | 829.00 | 960.50 | 1,136.50 | 1,043.00 | **3,969.00** |
| 996 | 381.50 | 383.00 | 345.00 | 415.50 | **1,525.00** |
| 997 | 122.00 | 83.00 | 126.50 | 119.00 | **450.50** |
| **AK - ALASKA - Total** | **1,332.50** | **1,426.50** | **1,608.00** | **1,577.50** | **5,944.50** |

**STATE: AL - ALABAMA**

| REGISTRANT ZIP CODE 3 | 1ST QUARTER | 2ND QUARTER | 3RD QUARTER | 4TH QUARTER | TOTAL GRAMS |
|---|---|---|---|---|---|
| 350 | 3,690.50 | 3,482.00 | 3,305.00 | 3,070.00 | **13,547.50** |
| 351 | 1,718.50 | 1,693.00 | 1,598.50 | 1,610.50 | **6,620.50** |
| 352 | 2,263.00 | 1,977.00 | 2,292.00 | 1,960.50 | **8,492.50** |
| 354 | 1,885.50 | 1,680.00 | 1,733.50 | 1,749.50 | **7,048.50** |
| 355 | 1,333.50 | 1,369.00 | 1,123.00 | 1,107.50 | **4,933.00** |
| 356 | 1,556.50 | 1,454.00 | 1,665.00 | 1,427.50 | **6,103.00** |
| 357 | 1,058.50 | 965.00 | 1,056.50 | 1,132.50 | **4,212.50** |
| 358 | 1,329.50 | 1,280.50 | 1,270.00 | 1,093.00 | **4,973.00** |
| 359 | 902.50 | 939.00 | 902.00 | 952.00 | **3,695.50** |
| 360 | 2,340.00 | 2,100.00 | 2,016.50 | 1,767.00 | **8,223.50** |
| 361 | 1,435.00 | 1,427.50 | 1,309.50 | 1,161.00 | **5,333.00** |
| 362 | 544.00 | 471.50 | 470.00 | 453.50 | **1,939.00** |
| 363 | 649.00 | 607.50 | 527.00 | 569.50 | **2,353.00** |
| 364 | 283.50 | 302.50 | 284.00 | 208.50 | **1,078.50** |
| 365 | 1,523.50 | 1,394.00 | 1,512.00 | 1,442.50 | **5,872.00** |
| 366 | 1,266.00 | 1,165.00 | 1,275.50 | 1,195.00 | **4,901.50** |
| 367 | 353.50 | 313.00 | 288.00 | 332.00 | **1,286.50** |
| 368 | 1,568.50 | 1,728.50 | 1,656.50 | 1,537.50 | **6,491.00** |
| 369 | 129.00 | 158.00 | 120.00 | 113.00 | **520.00** |
| **AL - ALABAMA - Total** | **25,830.00** | **24,507.00** | **24,404.50** | **22,882.50** | **97,624.00** |

**STATE: AR - ARKANSAS**

| REGISTRANT ZIP CODE 3 | 1ST QUARTER | 2ND QUARTER | 3RD QUARTER | 4TH QUARTER | TOTAL GRAMS |
|---|---|---|---|---|---|
| 716 | 166.00 | 157.50 | 202.50 | 217.00 | **743.00** |
| 717 | 489.00 | 512.50 | 468.50 | 567.50 | **2,037.50** |
| 718 | 32.00 | 56.00 | 84.00 | 89.50 | **261.50** |

| | | | | | |
|---|---|---|---|---|---|
| 719 | 298.00 | 318.50 | 372.50 | 343.00 | **1,332.00** |
| 720 | 1,568.50 | 1,545.00 | 1,536.50 | 1,906.00 | **6,556.00** |
| 721 | 1,042.50 | 893.50 | 817.50 | 942.50 | **3,696.00** |
| 722 | 776.50 | 497.00 | 590.50 | 731.00 | **2,595.00** |
| 723 | 59.50 | 170.50 | 167.00 | 153.00 | **550.00** |
| 724 | 704.00 | 707.00 | 665.50 | 598.00 | **2,674.50** |
| 725 | 306.50 | 284.50 | 287.00 | 253.00 | **1,131.00** |
| 726 | 582.50 | 483.00 | 642.50 | 634.00 | **2,342.00** |
| 727 | 1,518.00 | 1,439.50 | 1,437.50 | 1,413.00 | **5,808.00** |
| 728 | 283.50 | 327.00 | 333.00 | 382.00 | **1,325.50** |
| 729 | 649.00 | 687.00 | 740.50 | 685.50 | **2,762.00** |
| **AR - ARKANSAS - Total** | **8,475.50** | **8,078.50** | **8,345.00** | **8,915.00** | **33,814.00** |

**STATE: AZ - ARIZONA**

| REGISTRANT ZIP CODE 3 | 1ST QUARTER | 2ND QUARTER | 3RD QUARTER | 4TH QUARTER | TOTAL GRAMS |
|---|---|---|---|---|---|
| 850 | 2,890.00 | 2,877.50 | 2,561.50 | 2,839.50 | **11,168.50** |
| 851 | 642.50 | 399.50 | 380.00 | 539.00 | **1,961.00** |
| 852 | 9,403.50 | 8,364.50 | 8,595.50 | 8,896.00 | **35,259.50** |
| 853 | 3,757.50 | 3,672.50 | 3,688.00 | 3,709.00 | **14,827.00** |
| 855 | 191.00 | 221.50 | 201.00 | 129.50 | **743.00** |
| 856 | 725.00 | 664.00 | 712.50 | 617.50 | **2,719.00** |
| 857 | 3,051.00 | 2,687.00 | 2,702.50 | 3,218.00 | **11,658.50** |
| 859 | 27.00 | 71.00 | 102.00 | 56.00 | **256.00** |
| 860 | 121.50 | 157.00 | 117.50 | 196.50 | **592.50** |
| 863 | 456.00 | 381.00 | 411.00 | 471.00 | **1,719.00** |
| 864 | 124.00 | 165.00 | 138.50 | 115.00 | **542.50** |
| 865 | 0 | 0 | 21.00 | 0 | **21.00** |
| **AZ - ARIZONA - Total** | **21,389.00** | **19,660.50** | **19,631.00** | **20,787.00** | **81,467.50** |

**STATE: CA - CALIFORNIA**

| REGISTRANT ZIP CODE 3 | 1ST QUARTER | 2ND QUARTER | 3RD QUARTER | 4TH QUARTER | TOTAL GRAMS |
|---|---|---|---|---|---|
| 900 | 1,256.50 | 939.50 | 1,019.00 | 1,194.00 | **4,409.00** |
| 902 | 1,278.00 | 1,255.00 | 1,215.00 | 1,160.50 | **4,908.50** |
| 903 | 32.50 | 40.00 | 55.00 | 35.00 | **162.50** |
| 904 | 1,017.00 | 696.00 | 769.00 | 830.50 | **3,312.50** |
| 905 | 417.00 | 189.50 | 349.50 | 278.00 | **1,234.00** |
| 906 | 443.00 | 496.50 | 413.50 | 526.50 | **1,879.50** |
| 907 | 778.50 | 715.50 | 637.50 | 808.00 | **2,939.50** |
| 908 | 414.50 | 332.50 | 317.50 | 285.00 | **1,349.50** |
| 910 | 220.00 | 222.50 | 194.50 | 230.50 | **867.50** |
| 911 | 369.00 | 355.00 | 396.00 | 371.00 | **1,491.00** |
| 912 | 149.00 | 163.50 | 228.50 | 181.00 | **722.00** |
| 913 | 2,986.00 | 2,930.00 | 2,935.50 | 2,860.00 | **11,711.50** |
| 914 | 633.00 | 587.00 | 605.50 | 536.00 | **2,361.50** |
| 915 | 72.50 | 47.50 | 30.00 | 30.00 | **180.00** |
| 916 | 52.50 | 126.00 | 89.00 | 115.00 | **382.50** |
| 917 | 3,243.00 | 2,720.50 | 2,643.00 | 2,584.50 | **11,191.00** |
| 918 | 7.50 | 40.00 | 10.00 | 30.00 | **87.50** |
| 919 | 978.50 | 930.50 | 873.00 | 465.00 | **3,247.00** |
| 920 | 5,487.50 | 5,334.50 | 5,271.50 | 4,860.00 | **20,953.50** |
| 921 | 1,935.00 | 2,080.00 | 1,724.50 | 1,870.50 | **7,610.00** |
| 922 | 1,399.50 | 1,395.50 | 1,470.50 | 1,345.50 | **5,611.00** |
| 923 | 2,016.50 | 2,209.00 | 1,977.50 | 1,740.50 | **7,943.50** |
| 924 | 127.00 | 198.00 | 121.00 | 136.50 | **582.50** |
| 925 | 4,515.00 | 4,006.00 | 4,287.50 | 4,079.00 | **16,887.50** |
| 926 | 3,826.00 | 3,676.00 | 3,832.90 | 3,884.00 | **15,218.90** |
| 927 | 529.00 | 529.50 | 576.00 | 399.50 | **2,034.00** |
| 928 | 2,056.00 | 2,434.50 | 2,289.00 | 2,355.50 | **9,135.00** |
| 930 | 1,731.00 | 1,583.50 | 1,576.50 | 1,565.00 | **6,456.00** |
| 931 | 180.00 | 296.50 | 161.00 | 161.00 | **798.50** |
| 932 | 2,424.50 | 2,263.00 | 2,240.50 | 2,258.00 | **9,186.00** |
| 933 | 776.50 | 811.00 | 745.50 | 760.00 | **3,093.00** |

| | | | | | |
|---|---|---|---|---|---|
| 934 | 1,325.00 | 1,279.00 | 1,232.50 | 1,142.00 | **4,978.50** |
| 935 | 764.50 | 711.00 | 738.00 | 650.50 | **2,864.00** |
| 936 | 1,914.00 | 1,828.00 | 1,788.50 | 1,660.00 | **7,190.50** |
| 937 | 2,298.00 | 2,101.00 | 2,242.50 | 2,134.50 | **8,776.00** |
| 939 | 482.00 | 491.50 | 525.00 | 563.00 | **2,061.50** |
| 940 | 1,034.00 | 873.50 | 1,171.50 | 1,303.50 | **4,382.50** |
| 941 | 781.50 | 505.00 | 555.50 | 586.00 | **2,428.00** |
| 943 | 0 | 15.00 | 28.00 | 24.00 | **67.00** |
| 944 | 126.00 | 273.00 | 243.50 | 222.50 | **865.00** |
| 945 | 1,793.50 | 1,810.50 | 1,930.00 | 1,864.50 | **7,398.50** |
| 946 | 104.00 | 152.50 | 167.00 | 147.00 | **570.50** |
| 947 | 30.00 | 20.00 | 15.00 | 23.00 | **88.00** |
| 948 | 16.00 | 5.00 | 20.00 | 35.00 | **76.00** |
| 949 | 166.00 | 267.00 | 185.50 | 183.50 | **802.00** |
| 950 | 374.50 | 372.50 | 428.00 | 409.00 | **1,584.00** |
| 951 | 189.50 | 199.00 | 222.50 | 261.00 | **872.00** |
| 952 | 280.50 | 287.50 | 371.50 | 392.00 | **1,331.50** |
| 953 | 1,093.50 | 1,227.00 | 1,098.00 | 1,176.50 | **4,595.00** |
| 954 | 371.00 | 403.50 | 432.00 | 456.50 | **1,663.00** |
| 955 | 23.00 | 6.00 | 28.00 | 8.00 | **65.00** |
| 956 | 1,799.00 | 1,871.50 | 1,862.50 | 1,866.50 | **7,399.50** |
| 957 | 501.50 | 463.00 | 466.00 | 436.50 | **1,867.00** |
| 958 | 1,028.50 | 856.50 | 742.50 | 998.00 | **3,625.50** |
| 959 | 1,020.00 | 992.50 | 897.00 | 1,021.00 | **3,930.50** |
| 960 | 378.00 | 412.00 | 425.00 | 439.00 | **1,654.00** |
| 961 | 18.00 | 59.00 | 62.00 | 90.00 | **229.00** |
| **CA - CALIFORNIA - Total** | **59,263.00** | **57,086.00** | **56,932.40** | **56,028.50** | **229,309.90** |

**STATE: CO - COLORADO**

| REGISTRANT ZIP CODE 3 | 1ST QUARTER | 2ND QUARTER | 3RD QUARTER | 4TH QUARTER | TOTAL GRAMS |
|---|---|---|---|---|---|
| 800 | 2,859.00 | 2,500.50 | 2,633.50 | 3,068.00 | **11,061.00** |
| 801 | 5,026.00 | 4,635.50 | 4,731.50 | 4,613.50 | **19,006.50** |
| 802 | 2,484.50 | 2,446.50 | 2,567.50 | 2,627.50 | **10,126.00** |
| 803 | 560.50 | 596.50 | 658.50 | 537.50 | **2,353.00** |
| 804 | 542.00 | 536.00 | 592.00 | 484.50 | **2,154.50** |
| 805 | 2,316.00 | 2,044.50 | 2,113.00 | 2,244.00 | **8,717.50** |
| 806 | 898.00 | 1,012.00 | 884.50 | 820.50 | **3,615.00** |
| 807 | 138.00 | 160.00 | 186.00 | 141.00 | **625.00** |
| 808 | 451.00 | 380.50 | 479.00 | 485.00 | **1,795.50** |
| 809 | 2,642.00 | 2,510.00 | 2,842.00 | 3,044.50 | **11,038.50** |
| 810 | 1,660.00 | 1,342.00 | 1,353.50 | 1,749.50 | **6,105.00** |
| 811 | 126.00 | 179.00 | 218.00 | 208.50 | **731.50** |
| 812 | 457.00 | 508.50 | 540.00 | 675.00 | **2,180.50** |
| 813 | 141.50 | 131.00 | 167.50 | 183.50 | **623.50** |
| 814 | 110.00 | 118.50 | 158.00 | 148.00 | **534.50** |
| 815 | 221.00 | 362.00 | 243.00 | 224.00 | **1,050.00** |
| 816 | 131.00 | 149.00 | 176.50 | 121.00 | **577.50** |
| **CO - COLORADO - Total** | **20,763.50** | **19,612.00** | **20,544.00** | **21,375.50** | **82,295.00** |

**STATE: CT - CONNECTICUT**

| REGISTRANT ZIP CODE 3 | 1ST QUARTER | 2ND QUARTER | 3RD QUARTER | 4TH QUARTER | TOTAL GRAMS |
|---|---|---|---|---|---|
| 060 | 1,010.00 | 1,152.00 | 974.50 | 1,001.50 | **4,138.00** |
| 061 | 293.50 | 268.00 | 305.00 | 242.00 | **1,108.50** |
| 062 | 414.00 | 256.00 | 287.00 | 219.00 | **1,176.00** |
| 063 | 555.50 | 611.00 | 557.50 | 577.50 | **2,301.50** |
| 064 | 1,723.00 | 1,546.00 | 1,662.50 | 2,013.50 | **6,945.00** |
| 065 | 532.00 | 418.00 | 405.50 | 443.00 | **1,798.50** |
| 066 | 249.50 | 256.00 | 251.50 | 381.50 | **1,138.50** |
| 067 | 675.50 | 560.00 | 592.00 | 563.00 | **2,390.50** |
| 068 | 850.50 | 792.50 | 819.00 | 643.50 | **3,105.50** |
| 069 | 75.00 | 95.00 | 97.00 | 118.50 | **385.50** |
| **CT - CONNECTICUT - Total** | **6,378.50** | **5,954.50** | **5,951.50** | **6,203.00** | **24,487.50** |

STATE: DC - DISTRICT OF COLUMBIA

| REGISTRANT ZIP CODE 3 | 1ST QUARTER | 2ND QUARTER | 3RD QUARTER | 4TH QUARTER | TOTAL GRAMS |
|---|---|---|---|---|---|
| 200 | 536.50 | 576.00 | 441.00 | 492.00 | **2,045.50** |
| **DC - DISTRICT OF COLUMBIA - Total** | **536.50** | **576.00** | **441.00** | **492.00** | **2,045.50** |

STATE: DE - DELAWARE

| REGISTRANT ZIP CODE 3 | 1ST QUARTER | 2ND QUARTER | 3RD QUARTER | 4TH QUARTER | TOTAL GRAMS |
|---|---|---|---|---|---|
| 197 | 1,907.00 | 1,637.00 | 1,618.00 | 1,619.50 | **6,781.50** |
| 198 | 1,019.00 | 757.50 | 656.50 | 886.50 | **3,319.50** |
| 199 | 1,504.00 | 1,324.00 | 1,234.00 | 1,370.50 | **5,432.50** |
| **DE - DELAWARE - Total** | **4,430.00** | **3,718.50** | **3,508.50** | **3,876.50** | **15,533.50** |

STATE: FL - FLORIDA

| REGISTRANT ZIP CODE 3 | 1ST QUARTER | 2ND QUARTER | 3RD QUARTER | 4TH QUARTER | TOTAL GRAMS |
|---|---|---|---|---|---|
| 320 | 1,819.00 | 1,943.00 | 1,976.50 | 1,940.50 | **7,679.00** |
| 321 | 1,313.00 | 1,269.00 | 1,281.50 | 1,275.50 | **5,139.00** |
| 322 | 2,212.00 | 2,254.50 | 2,398.00 | 2,120.50 | **8,985.00** |
| 323 | 2,129.00 | 2,268.50 | 2,027.00 | 2,161.50 | **8,586.00** |
| 324 | 1,016.00 | 996.50 | 1,033.50 | 1,072.00 | **4,118.00** |
| 325 | 1,670.00 | 1,511.50 | 1,380.50 | 1,445.50 | **6,007.50** |
| 326 | 969.00 | 801.00 | 873.50 | 741.50 | **3,385.00** |
| 327 | 2,981.50 | 3,230.00 | 3,176.50 | 3,129.50 | **12,517.50** |
| 328 | 2,215.00 | 1,833.50 | 1,905.50 | 1,963.00 | **7,917.00** |
| 329 | 2,551.00 | 2,288.00 | 2,259.00 | 2,252.50 | **9,350.50** |
| 330 | 1,712.00 | 1,580.50 | 1,390.50 | 1,656.50 | **6,339.50** |
| 331 | 509.50 | 431.00 | 442.00 | 477.50 | **1,860.00** |
| 333 | 1,520.50 | 1,297.00 | 1,329.00 | 1,471.50 | **5,618.00** |
| 334 | 4,660.00 | 3,890.00 | 4,290.50 | 4,382.00 | **17,222.50** |
| 335 | 3,504.00 | 3,417.00 | 3,210.00 | 3,463.00 | **13,594.00** |
| 336 | 1,990.50 | 1,924.50 | 2,207.50 | 2,104.50 | **8,227.00** |
| 337 | 2,433.00 | 2,443.00 | 2,040.00 | 2,201.00 | **9,117.00** |
| 338 | 1,778.50 | 1,631.50 | 1,864.50 | 1,817.00 | **7,091.50** |
| 339 | 5,342.00 | 5,026.00 | 4,757.00 | 5,042.50 | **20,167.50** |
| 341 | 2,320.50 | 2,071.50 | 1,888.50 | 1,824.50 | **8,105.00** |
| 342 | 3,251.50 | 2,961.00 | 3,035.50 | 3,129.00 | **12,377.00** |
| 344 | 1,040.00 | 1,080.50 | 1,010.50 | 1,083.50 | **4,214.50** |
| 346 | 2,432.00 | 2,448.00 | 2,519.50 | 2,548.50 | **9,948.00** |
| 347 | 1,739.50 | 1,660.00 | 1,632.00 | 1,419.50 | **6,451.00** |
| 349 | 1,671.50 | 1,789.50 | 1,533.00 | 1,490.50 | **6,484.50** |
| **FL - FLORIDA - Total** | **54,780.50** | **52,046.50** | **51,461.50** | **52,213.00** | **210,501.50** |

STATE: GA - GEORGIA

| REGISTRANT ZIP CODE 3 | 1ST QUARTER | 2ND QUARTER | 3RD QUARTER | 4TH QUARTER | TOTAL GRAMS |
|---|---|---|---|---|---|
| 300 | 7,548.50 | 6,752.50 | 6,982.00 | 7,091.50 | **28,374.50** |
| 301 | 5,027.50 | 4,774.00 | 4,611.50 | 4,733.00 | **19,146.00** |
| 302 | 3,214.00 | 2,871.50 | 3,212.50 | 2,998.50 | **12,296.50** |
| 303 | 2,024.50 | 1,876.00 | 1,826.00 | 1,939.00 | **7,665.50** |
| 304 | 2,004.50 | 2,176.00 | 2,008.00 | 1,859.00 | **8,047.50** |
| 305 | 2,576.50 | 2,632.50 | 2,585.50 | 2,637.50 | **10,432.00** |
| 306 | 1,423.50 | 1,441.00 | 1,494.50 | 1,295.00 | **5,654.00** |
| 307 | 796.50 | 533.00 | 525.50 | 579.50 | **2,434.50** |
| 308 | 601.50 | 576.50 | 439.50 | 419.50 | **2,037.00** |
| 309 | 579.00 | 482.00 | 822.00 | 569.00 | **2,452.00** |
| 310 | 2,451.50 | 2,123.00 | 2,263.50 | 2,126.50 | **8,964.50** |
| 312 | 479.00 | 388.00 | 496.00 | 476.50 | **1,839.50** |
| 313 | 3,129.00 | 3,120.50 | 2,955.50 | 2,815.00 | **12,020.00** |
| 314 | 1,434.00 | 1,344.00 | 1,364.50 | 1,312.00 | **5,454.50** |
| 315 | 4,951.00 | 4,694.50 | 4,791.00 | 4,777.50 | **19,214.00** |
| 316 | 762.00 | 557.00 | 659.00 | 615.00 | **2,593.00** |
| 317 | 997.00 | 919.00 | 980.00 | 948.00 | **3,844.00** |
| 318 | 126.50 | 73.00 | 76.50 | 71.00 | **347.00** |
| 319 | 394.50 | 349.50 | 386.50 | 302.50 | **1,433.00** |
| 398 | 185.00 | 187.00 | 257.50 | 158.50 | **788.00** |

| GA - GEORGIA - Total | 40,705.50 | 37,870.50 | 38,737.00 | 37,724.00 | 155,037.00 |
|---|---|---|---|---|---|

**STATE: HI - HAWAII**

| REGISTRANT ZIP CODE 3 | 1ST QUARTER | 2ND QUARTER | 3RD QUARTER | 4TH QUARTER | TOTAL GRAMS |
|---|---|---|---|---|---|
| 967 | 707.50 | 674.50 | 681.50 | 587.50 | **2,651.00** |
| 968 | 609.00 | 575.50 | 509.50 | 640.50 | **2,334.50** |
| HI - HAWAII - Total | 1,316.50 | 1,250.00 | 1,191.00 | 1,228.00 | 4,985.50 |

**STATE: IA - IOWA**

| REGISTRANT ZIP CODE 3 | 1ST QUARTER | 2ND QUARTER | 3RD QUARTER | 4TH QUARTER | TOTAL GRAMS |
|---|---|---|---|---|---|
| 500 | 698.50 | 698.50 | 662.50 | 537.50 | **2,597.00** |
| 501 | 585.50 | 503.50 | 480.50 | 480.00 | **2,049.50** |
| 502 | 1,003.50 | 893.00 | 747.00 | 729.00 | **3,372.50** |
| 503 | 1,208.00 | 962.50 | 1,056.50 | 1,040.00 | **4,267.00** |
| 504 | 65.00 | 65.00 | 54.00 | 55.00 | **239.00** |
| 505 | 125.50 | 144.00 | 161.50 | 114.00 | **545.00** |
| 506 | 344.00 | 250.50 | 300.00 | 293.50 | **1,188.00** |
| 507 | 196.50 | 239.50 | 312.50 | 309.50 | **1,058.00** |
| 508 | 151.50 | 167.50 | 142.00 | 123.00 | **584.00** |
| 510 | 199.00 | 211.00 | 227.00 | 262.00 | **899.00** |
| 511 | 242.00 | 356.50 | 289.50 | 257.00 | **1,145.00** |
| 512 | 99.50 | 116.00 | 75.50 | 112.50 | **403.50** |
| 513 | 231.50 | 61.00 | 77.00 | 85.00 | **454.50** |
| 514 | 50.00 | 48.00 | 75.00 | 70.00 | **243.00** |
| 515 | 382.50 | 353.50 | 356.00 | 271.00 | **1,363.00** |
| 516 | 161.00 | 160.50 | 207.00 | 167.50 | **696.00** |
| 520 | 433.50 | 348.00 | 396.50 | 314.50 | **1,492.50** |
| 521 | 0 | 5.00 | 5.00 | 5.00 | **15.00** |
| 522 | 24.00 | 64.00 | 51.50 | 49.50 | **189.00** |
| 523 | 147.00 | 125.50 | 86.50 | 113.50 | **472.50** |
| 524 | 103.50 | 95.00 | 95.00 | 173.50 | **467.00** |
| 525 | 257.50 | 194.00 | 301.00 | 225.50 | **978.00** |
| 526 | 7.50 | 0 | 7.50 | 0 | **15.00** |
| 527 | 76.00 | 39.50 | 67.00 | 94.00 | **276.50** |
| 528 | 48.00 | 33.00 | 56.00 | 41.00 | **178.00** |
| IA - IOWA - Total | 6,840.50 | 6,134.50 | 6,289.50 | 5,923.00 | 25,187.50 |

**STATE: ID - IDAHO**

| REGISTRANT ZIP CODE 3 | 1ST QUARTER | 2ND QUARTER | 3RD QUARTER | 4TH QUARTER | TOTAL GRAMS |
|---|---|---|---|---|---|
| 832 | 786.00 | 875.00 | 872.00 | 862.50 | **3,395.50** |
| 833 | 254.00 | 300.00 | 248.00 | 282.00 | **1,084.00** |
| 834 | 644.00 | 625.50 | 777.00 | 886.00 | **2,932.50** |
| 835 | 134.00 | 132.00 | 191.00 | 173.00 | **630.00** |
| 836 | 941.50 | 891.50 | 1,078.00 | 970.00 | **3,881.00** |
| 837 | 745.50 | 531.00 | 480.00 | 614.50 | **2,371.00** |
| 838 | 481.00 | 446.50 | 448.00 | 496.00 | **1,871.50** |
| ID - IDAHO - Total | 3,986.00 | 3,801.50 | 4,094.00 | 4,284.00 | 16,165.50 |

**STATE: IL - ILLINOIS**

| REGISTRANT ZIP CODE 3 | 1ST QUARTER | 2ND QUARTER | 3RD QUARTER | 4TH QUARTER | TOTAL GRAMS |
|---|---|---|---|---|---|
| 600 | 3,947.00 | 3,286.00 | 3,341.50 | 3,405.50 | **13,980.00** |
| 601 | 1,977.50 | 1,752.00 | 1,933.00 | 1,823.00 | **7,485.50** |
| 602 | 145.50 | 164.50 | 153.00 | 201.00 | **664.00** |
| 603 | 223.50 | 233.00 | 243.50 | 347.50 | **1,047.50** |
| 604 | 2,487.00 | 2,186.00 | 2,179.50 | 2,235.50 | **9,088.00** |
| 605 | 1,761.00 | 2,028.00 | 1,946.00 | 2,148.50 | **7,883.50** |
| 606 | 1,097.00 | 923.00 | 988.00 | 964.00 | **3,972.00** |
| 607 | 219.50 | 185.00 | 228.00 | 272.50 | **905.00** |
| 608 | 150.50 | 170.50 | 151.00 | 118.00 | **590.00** |
| 609 | 525.00 | 457.50 | 436.00 | 583.50 | **2,002.00** |
| 610 | 671.50 | 664.00 | 745.50 | 898.00 | **2,979.00** |
| 611 | 721.50 | 542.50 | 581.00 | 482.50 | **2,327.50** |
| 612 | 139.00 | 96.00 | 104.50 | 124.50 | **464.00** |
| 613 | 78.00 | 48.00 | 76.50 | 99.50 | **302.00** |

| | | | | | |
|---|---|---|---|---|---|
| 614 | 93.00 | 81.00 | 117.50 | 70.00 | **361.50** |
| 615 | 305.00 | 260.00 | 194.00 | 336.00 | **1,095.00** |
| 616 | 212.00 | 156.00 | 179.00 | 257.00 | **804.00** |
| 617 | 442.00 | 397.50 | 355.00 | 424.00 | **1,618.50** |
| 618 | 1,326.00 | 1,126.00 | 1,190.00 | 1,335.00 | **4,977.00** |
| 619 | 293.00 | 277.50 | 224.50 | 273.00 | **1,068.00** |
| 620 | 979.50 | 890.50 | 826.00 | 795.00 | **3,491.00** |
| 622 | 1,688.00 | 1,257.00 | 1,331.00 | 1,347.50 | **5,623.50** |
| 623 | 245.00 | 257.00 | 150.00 | 172.00 | **824.00** |
| 624 | 108.00 | 114.50 | 148.00 | 170.50 | **541.00** |
| 625 | 467.50 | 557.50 | 495.00 | 571.00 | **2,091.00** |
| 626 | 187.00 | 165.50 | 212.00 | 257.00 | **821.50** |
| 627 | 99.00 | 60.00 | 141.00 | 145.50 | **445.50** |
| 628 | 427.50 | 277.50 | 308.50 | 415.00 | **1,428.50** |
| 629 | 281.50 | 229.50 | 196.00 | 188.00 | **895.00** |
| **IL - ILLINOIS - Total** | **21,297.50** | **18,843.00** | **19,174.50** | **20,460.00** | **79,775.00** |

**STATE: IN - INDIANA**

| REGISTRANT ZIP CODE 3 | 1ST QUARTER | 2ND QUARTER | 3RD QUARTER | 4TH QUARTER | TOTAL GRAMS |
|---|---|---|---|---|---|
| 460 | 945.00 | 977.50 | 978.50 | 1,028.50 | **3,929.50** |
| 461 | 1,998.00 | 1,875.50 | 1,693.00 | 1,740.50 | **7,307.00** |
| 462 | 1,309.00 | 1,062.00 | 1,006.00 | 1,120.50 | **4,497.50** |
| 463 | 2,443.00 | 2,313.00 | 2,330.00 | 2,320.00 | **9,406.00** |
| 464 | 336.00 | 335.50 | 226.50 | 174.50 | **1,072.50** |
| 465 | 1,986.50 | 1,959.50 | 2,067.50 | 2,306.50 | **8,320.00** |
| 466 | 209.50 | 234.00 | 355.00 | 365.00 | **1,163.50** |
| 467 | 2,448.50 | 2,365.50 | 2,379.50 | 2,130.00 | **9,323.50** |
| 468 | 3,557.00 | 3,104.00 | 3,104.00 | 2,936.00 | **12,701.00** |
| 469 | 933.00 | 768.00 | 822.50 | 828.50 | **3,352.00** |
| 470 | 281.50 | 254.50 | 231.50 | 182.50 | **950.00** |
| 471 | 523.00 | 380.00 | 461.50 | 412.00 | **1,776.50** |
| 472 | 406.00 | 388.00 | 350.50 | 236.50 | **1,381.00** |
| 473 | 431.00 | 512.50 | 477.00 | 544.50 | **1,965.00** |
| 474 | 504.50 | 492.00 | 514.50 | 683.50 | **2,194.50** |
| 475 | 400.00 | 377.50 | 457.50 | 524.00 | **1,759.00** |
| 476 | 378.50 | 387.50 | 419.50 | 529.50 | **1,715.00** |
| 477 | 816.50 | 877.00 | 809.50 | 706.00 | **3,209.00** |
| 478 | 3,145.50 | 3,111.50 | 3,294.00 | 3,328.00 | **12,879.00** |
| 479 | 443.50 | 441.50 | 476.00 | 476.00 | **1,837.00** |
| **IN - INDIANA - Total** | **23,495.50** | **22,216.50** | **22,454.00** | **22,572.50** | **90,738.50** |

**STATE: KS - KANSAS**

| REGISTRANT ZIP CODE 3 | 1ST QUARTER | 2ND QUARTER | 3RD QUARTER | 4TH QUARTER | TOTAL GRAMS |
|---|---|---|---|---|---|
| 660 | 1,229.00 | 1,175.00 | 1,255.50 | 1,184.00 | **4,843.50** |
| 661 | 86.00 | 105.00 | 128.50 | 96.50 | **416.00** |
| 662 | 830.00 | 885.50 | 827.00 | 964.00 | **3,506.50** |
| 664 | 133.00 | 152.00 | 136.00 | 123.00 | **544.00** |
| 665 | 160.00 | 223.00 | 220.00 | 180.00 | **783.00** |
| 666 | 514.00 | 517.50 | 529.00 | 516.00 | **2,076.50** |
| 667 | 173.00 | 157.00 | 243.00 | 236.50 | **809.50** |
| 668 | 48.00 | 49.00 | 42.50 | 36.00 | **175.50** |
| 669 | 20.00 | 10.00 | 55.00 | 25.00 | **110.00** |
| 670 | 105.00 | 74.00 | 123.00 | 122.00 | **424.00** |
| 671 | 73.00 | 50.00 | 77.00 | 60.00 | **260.00** |
| 672 | 458.00 | 433.50 | 379.00 | 434.00 | **1,704.50** |
| 673 | 101.00 | 80.50 | 103.00 | 122.00 | **406.50** |
| 674 | 236.00 | 242.00 | 202.00 | 256.00 | **936.00** |
| 675 | 101.00 | 73.00 | 115.00 | 132.00 | **421.00** |
| 676 | 15.00 | 18.00 | 45.00 | 35.00 | **113.00** |
| 677 | 44.00 | 50.00 | 61.00 | 63.00 | **218.00** |
| 678 | 54.00 | 45.00 | 18.00 | 18.00 | **135.00** |
| 679 | 20.00 | 10.00 | 17.50 | 15.00 | **62.50** |

| KS - KANSAS - Total | 4,400.00 | 4,350.00 | 4,577.00 | 4,618.00 | 17,945.00 |
|---|---|---|---|---|---|

STATE: KY - KENTUCKY

| REGISTRANT ZIP CODE 3 | 1ST QUARTER | 2ND QUARTER | 3RD QUARTER | 4TH QUARTER | TOTAL GRAMS |
|---|---|---|---|---|---|
| 400 | 306.50 | 259.50 | 241.50 | 177.50 | **985.00** |
| 401 | 183.50 | 191.50 | 130.00 | 163.00 | **668.00** |
| 402 | 1,323.50 | 1,202.50 | 1,244.50 | 1,294.00 | **5,064.50** |
| 403 | 350.50 | 264.00 | 375.50 | 321.50 | **1,311.50** |
| 404 | 125.00 | 80.00 | 146.00 | 120.00 | **471.00** |
| 405 | 318.00 | 336.00 | 395.00 | 353.00 | **1,402.00** |
| 406 | 6.00 | 22.00 | 17.50 | 20.00 | **65.50** |
| 407 | 79.50 | 102.50 | 103.00 | 86.00 | **371.00** |
| 409 | 31.00 | 32.50 | 21.50 | 55.00 | **140.00** |
| 410 | 1,405.00 | 1,486.50 | 1,422.50 | 1,494.00 | **5,808.00** |
| 411 | 61.00 | 61.00 | 48.00 | 61.00 | **231.00** |
| 412 | 15.00 | 10.00 | 27.50 | 3.00 | **55.50** |
| 413 | 15.00 | 5.00 | 5.00 | 10.00 | **35.00** |
| 414 | 0 | 0 | 5.00 | 0 | **5.00** |
| 415 | 99.50 | 93.50 | 103.00 | 113.00 | **409.00** |
| 416 | 118.50 | 88.50 | 117.00 | 96.00 | **420.00** |
| 417 | 56.00 | 41.00 | 53.00 | 42.50 | **192.50** |
| 418 | 39.00 | 33.00 | 43.00 | 48.00 | **163.00** |
| 420 | 1,055.50 | 957.50 | 1,055.50 | 953.50 | **4,022.00** |
| 421 | 312.00 | 355.00 | 413.00 | 381.00 | **1,461.00** |
| 422 | 329.00 | 338.00 | 590.50 | 313.50 | **1,571.00** |
| 423 | 696.50 | 737.50 | 603.00 | 682.50 | **2,719.50** |
| 424 | 463.00 | 424.00 | 448.00 | 433.50 | **1,768.50** |
| 425 | 10.00 | 26.00 | 32.00 | 15.00 | **83.00** |
| 426 | 114.00 | 35.00 | 55.00 | 70.00 | **274.00** |
| 427 | 290.00 | 379.00 | 438.50 | 341.00 | **1,448.50** |
| KY - KENTUCKY - Total | 7,802.50 | 7,561.00 | 8,134.00 | 7,647.50 | 31,145.00 |

STATE: LA - LOUISIANA

| REGISTRANT ZIP CODE 3 | 1ST QUARTER | 2ND QUARTER | 3RD QUARTER | 4TH QUARTER | TOTAL GRAMS |
|---|---|---|---|---|---|
| 700 | 1,152.00 | 1,040.50 | 1,107.00 | 1,203.50 | **4,503.00** |
| 701 | 259.50 | 209.00 | 258.00 | 198.50 | **925.00** |
| 703 | 691.00 | 592.00 | 665.00 | 454.00 | **2,402.00** |
| 704 | 2,462.00 | 2,514.50 | 2,475.50 | 2,375.00 | **9,827.00** |
| 705 | 1,206.50 | 1,051.50 | 1,174.50 | 1,192.50 | **4,625.00** |
| 706 | 428.00 | 422.50 | 336.00 | 311.50 | **1,498.00** |
| 707 | 1,755.00 | 1,541.50 | 1,542.50 | 1,492.00 | **6,331.00** |
| 708 | 771.00 | 855.00 | 834.50 | 730.50 | **3,191.00** |
| 710 | 201.00 | 254.00 | 223.00 | 251.50 | **929.50** |
| 711 | 839.50 | 705.50 | 745.50 | 831.00 | **3,121.50** |
| 712 | 303.50 | 249.00 | 231.00 | 291.50 | **1,075.00** |
| 713 | 547.50 | 605.00 | 636.50 | 638.00 | **2,427.00** |
| 714 | 186.50 | 214.00 | 162.50 | 196.50 | **759.50** |
| LA - LOUISIANA - Total | 10,803.00 | 10,254.00 | 10,391.50 | 10,166.00 | 41,614.50 |

STATE: MA - MASSACHUSETTS

| REGISTRANT ZIP CODE 3 | 1ST QUARTER | 2ND QUARTER | 3RD QUARTER | 4TH QUARTER | TOTAL GRAMS |
|---|---|---|---|---|---|
| 010 | 1,113.50 | 1,023.50 | 796.50 | 884.00 | **3,817.50** |
| 011 | 158.00 | 143.00 | 147.00 | 170.00 | **618.00** |
| 012 | 70.00 | 95.00 | 70.00 | 92.50 | **327.50** |
| 013 | 61.00 | 83.00 | 55.50 | 73.00 | **272.50** |
| 014 | 292.00 | 244.00 | 356.00 | 303.00 | **1,195.00** |
| 015 | 554.00 | 427.00 | 611.00 | 551.00 | **2,143.00** |
| 016 | 199.50 | 248.50 | 239.50 | 163.50 | **851.00** |
| 017 | 456.00 | 445.00 | 456.00 | 439.50 | **1,796.50** |
| 018 | 7,548.00 | 7,303.50 | 6,773.00 | 6,408.50 | **28,033.00** |
| 019 | 1,043.00 | 1,132.50 | 1,087.00 | 980.50 | **4,243.00** |
| 020 | 216.00 | 295.50 | 315.50 | 289.50 | **1,116.50** |
| 021 | 688.50 | 638.00 | 724.00 | 658.50 | **2,709.00** |

| | | | | | |
|---|---|---|---|---|---|
| 022 | 91.00 | 100.50 | 120.00 | 86.50 | **398.00** |
| 023 | 494.50 | 492.50 | 498.50 | 439.00 | **1,924.50** |
| 024 | 248.00 | 211.00 | 247.00 | 375.50 | **1,081.50** |
| 025 | 79.00 | 167.00 | 111.00 | 127.00 | **484.00** |
| 026 | 175.50 | 225.00 | 248.00 | 226.00 | **874.50** |
| 027 | 1,602.00 | 1,493.00 | 1,366.50 | 1,295.00 | **5,756.50** |
| **MA - MASSACHUSETTS - Total** | **15,089.50** | **14,767.50** | **14,222.00** | **13,562.50** | **57,641.50** |

**STATE: MD - MARYLAND**

| REGISTRANT ZIP CODE 3 | 1ST QUARTER | 2ND QUARTER | 3RD QUARTER | 4TH QUARTER | TOTAL GRAMS |
|---|---|---|---|---|---|
| 206 | 1,473.50 | 1,453.50 | 1,583.00 | 1,533.50 | **6,043.50** |
| 207 | 1,795.50 | 1,574.00 | 1,662.00 | 1,572.00 | **6,603.50** |
| 208 | 2,342.50 | 2,673.00 | 2,536.00 | 2,710.00 | **10,261.50** |
| 209 | 276.00 | 237.00 | 265.00 | 276.00 | **1,054.00** |
| 210 | 4,442.50 | 4,481.00 | 4,297.00 | 4,626.00 | **17,846.50** |
| 211 | 2,196.00 | 1,897.50 | 2,087.00 | 1,851.50 | **8,032.00** |
| 212 | 2,869.00 | 2,709.00 | 2,928.50 | 2,636.00 | **11,142.50** |
| 214 | 688.50 | 625.50 | 569.00 | 609.00 | **2,492.00** |
| 215 | 510.00 | 452.50 | 509.00 | 534.50 | **2,006.00** |
| 216 | 1,098.00 | 999.00 | 1,125.50 | 901.50 | **4,124.00** |
| 217 | 3,438.50 | 2,846.00 | 3,013.50 | 2,738.50 | **12,036.50** |
| 218 | 756.50 | 890.50 | 962.50 | 798.00 | **3,407.50** |
| 219 | 230.50 | 154.00 | 193.00 | 187.00 | **764.50** |
| **MD - MARYLAND - Total** | **22,117.00** | **20,992.50** | **21,731.00** | **20,973.50** | **85,814.00** |

**STATE: ME - MAINE**

| REGISTRANT ZIP CODE 3 | 1ST QUARTER | 2ND QUARTER | 3RD QUARTER | 4TH QUARTER | TOTAL GRAMS |
|---|---|---|---|---|---|
| 039 | 93.00 | 114.00 | 72.00 | 54.00 | **333.00** |
| 040 | 211.00 | 250.50 | 296.00 | 237.50 | **995.00** |
| 041 | 23.00 | 34.00 | 33.00 | 22.00 | **112.00** |
| 042 | 142.00 | 135.00 | 145.00 | 133.00 | **555.00** |
| 043 | 48.00 | 31.00 | 32.50 | 29.00 | **140.50** |
| 044 | 87.00 | 67.00 | 52.00 | 34.00 | **240.00** |
| 045 | 78.50 | 116.00 | 136.50 | 102.50 | **433.50** |
| 046 | 17.00 | 0 | 15.00 | 20.00 | **52.00** |
| 047 | 133.00 | 116.00 | 113.00 | 92.00 | **454.00** |
| 048 | 27.00 | 27.00 | 27.00 | 27.00 | **108.00** |
| 049 | 155.00 | 127.00 | 186.00 | 158.50 | **626.50** |
| **ME - MAINE - Total** | **1,014.50** | **1,017.50** | **1,108.00** | **909.50** | **4,049.50** |

**STATE: MI - MICHIGAN**

| REGISTRANT ZIP CODE 3 | 1ST QUARTER | 2ND QUARTER | 3RD QUARTER | 4TH QUARTER | TOTAL GRAMS |
|---|---|---|---|---|---|
| 480 | 1,374.50 | 1,421.00 | 1,386.50 | 1,335.00 | **5,517.00** |
| 481 | 1,787.00 | 1,742.00 | 1,800.50 | 1,670.50 | **7,000.00** |
| 482 | 236.00 | 208.50 | 210.50 | 212.50 | **867.50** |
| 483 | 1,422.50 | 1,229.00 | 1,224.00 | 1,063.00 | **4,938.50** |
| 484 | 431.00 | 536.00 | 448.00 | 380.00 | **1,795.00** |
| 485 | 332.50 | 313.00 | 320.00 | 333.50 | **1,299.00** |
| 486 | 603.50 | 558.00 | 529.50 | 619.50 | **2,310.50** |
| 487 | 284.50 | 271.00 | 222.50 | 283.00 | **1,061.00** |
| 488 | 1,176.00 | 1,024.50 | 1,221.00 | 1,090.00 | **4,511.50** |
| 489 | 247.50 | 215.00 | 204.00 | 198.00 | **864.50** |
| 490 | 1,462.00 | 1,388.50 | 1,260.00 | 1,445.00 | **5,555.50** |
| 491 | 237.50 | 284.50 | 290.50 | 409.00 | **1,221.50** |
| 492 | 366.50 | 305.50 | 274.50 | 263.50 | **1,210.00** |
| 493 | 606.00 | 553.00 | 486.50 | 502.00 | **2,147.50** |
| 494 | 470.00 | 397.50 | 578.50 | 524.50 | **1,970.50** |
| 495 | 512.50 | 375.00 | 470.00 | 475.00 | **1,832.50** |
| 496 | 368.00 | 450.00 | 480.00 | 440.50 | **1,738.50** |
| 497 | 318.00 | 379.50 | 255.50 | 195.50 | **1,148.50** |
| 498 | 80.00 | 60.00 | 70.00 | 75.00 | **285.00** |
| 499 | 60.00 | 20.00 | 5.00 | 0 | **85.00** |
| **MI - MICHIGAN - Total** | **12,375.50** | **11,731.50** | **11,737.00** | **11,515.00** | **47,359.00** |

**STATE: MN - MINNESOTA**

| REGISTRANT ZIP CODE 3 | 1ST QUARTER | 2ND QUARTER | 3RD QUARTER | 4TH QUARTER | TOTAL GRAMS |
|---|---|---|---|---|---|
| 550 | 404.50 | 356.00 | 461.50 | 470.00 | **1,692.00** |
| 551 | 483.50 | 521.50 | 376.50 | 344.00 | **1,725.50** |
| 553 | 322.00 | 345.50 | 338.50 | 480.00 | **1,486.00** |
| 554 | 487.00 | 581.00 | 565.50 | 530.50 | **2,164.00** |
| 556 | 10.00 | 0 | 0 | 0 | **10.00** |
| 557 | 10.00 | 5.00 | 10.00 | 15.00 | **40.00** |
| 558 | 96.00 | 72.00 | 84.00 | 61.00 | **313.00** |
| 559 | 70.00 | 53.00 | 65.00 | 45.00 | **233.00** |
| 560 | 119.00 | 113.00 | 108.50 | 127.50 | **468.00** |
| 561 | 177.50 | 235.00 | 220.00 | 213.50 | **846.00** |
| 562 | 40.00 | 17.50 | 40.00 | 33.00 | **130.50** |
| 563 | 192.00 | 141.00 | 146.00 | 176.00 | **655.00** |
| 564 | 98.50 | 123.50 | 135.00 | 123.00 | **480.00** |
| 565 | 125.00 | 80.00 | 115.00 | 106.00 | **426.00** |
| 566 | 68.50 | 52.50 | 15.00 | 52.50 | **188.50** |
| 567 | 130.00 | 133.00 | 173.50 | 124.00 | **560.50** |
| **MN - MINNESOTA - Total** | **2,833.50** | **2,829.50** | **2,854.00** | **2,901.00** | **11,418.00** |

**STATE: MO - MISSOURI**

| REGISTRANT ZIP CODE 3 | 1ST QUARTER | 2ND QUARTER | 3RD QUARTER | 4TH QUARTER | TOTAL GRAMS |
|---|---|---|---|---|---|
| 630 | 1,509.50 | 1,096.00 | 1,230.00 | 1,329.00 | **5,164.50** |
| 631 | 4,718.00 | 4,825.00 | 4,581.00 | 4,364.00 | **18,488.00** |
| 633 | 1,512.50 | 1,461.50 | 1,503.00 | 1,507.00 | **5,984.00** |
| 634 | 286.50 | 225.00 | 280.50 | 216.50 | **1,008.50** |
| 635 | 128.50 | 98.00 | 71.50 | 111.00 | **409.00** |
| 636 | 161.50 | 203.50 | 177.00 | 133.00 | **675.00** |
| 637 | 390.50 | 283.00 | 362.50 | 339.00 | **1,375.00** |
| 638 | 351.00 | 333.00 | 318.50 | 368.00 | **1,370.50** |
| 639 | 124.00 | 157.50 | 129.50 | 122.00 | **533.00** |
| 640 | 1,016.50 | 1,024.00 | 1,003.00 | 967.00 | **4,010.50** |
| 641 | 335.50 | 358.00 | 335.00 | 329.00 | **1,357.50** |
| 644 | 95.00 | 95.00 | 95.00 | 75.00 | **360.00** |
| 645 | 242.00 | 248.00 | 301.00 | 194.00 | **985.00** |
| 646 | 38.00 | 35.00 | 62.00 | 38.00 | **173.00** |
| 647 | 219.50 | 200.50 | 306.00 | 217.00 | **943.00** |
| 648 | 764.50 | 560.00 | 599.00 | 634.50 | **2,558.00** |
| 650 | 422.50 | 510.00 | 512.00 | 528.50 | **1,973.00** |
| 651 | 378.50 | 467.50 | 624.00 | 733.50 | **2,203.50** |
| 652 | 393.50 | 463.50 | 425.00 | 353.50 | **1,635.50** |
| 653 | 190.00 | 216.00 | 193.50 | 227.50 | **827.00** |
| 654 | 226.50 | 282.00 | 359.00 | 279.00 | **1,146.50** |
| 655 | 181.00 | 210.00 | 213.00 | 202.00 | **806.00** |
| 656 | 332.00 | 352.00 | 369.00 | 353.50 | **1,406.50** |
| 657 | 500.50 | 468.00 | 519.00 | 583.50 | **2,071.00** |
| 658 | 523.50 | 446.50 | 445.00 | 367.50 | **1,782.50** |
| **MO - MISSOURI - Total** | **15,041.00** | **14,618.50** | **15,014.00** | **14,572.50** | **59,246.00** |

**STATE: MS - MISSISSIPPI**

| REGISTRANT ZIP CODE 3 | 1ST QUARTER | 2ND QUARTER | 3RD QUARTER | 4TH QUARTER | TOTAL GRAMS |
|---|---|---|---|---|---|
| 386 | 849.00 | 820.50 | 879.50 | 839.50 | **3,388.50** |
| 387 | 206.00 | 184.00 | 219.50 | 154.00 | **763.50** |
| 388 | 1,117.50 | 1,107.50 | 981.00 | 1,040.00 | **4,246.00** |
| 389 | 80.50 | 79.50 | 86.00 | 39.00 | **285.00** |
| 390 | 645.50 | 586.00 | 651.50 | 682.50 | **2,565.50** |
| 391 | 517.00 | 520.50 | 547.50 | 489.50 | **2,074.50** |
| 392 | 665.50 | 562.00 | 724.00 | 733.50 | **2,685.00** |
| 393 | 452.00 | 445.00 | 445.00 | 412.00 | **1,754.00** |
| 394 | 852.00 | 844.00 | 728.00 | 746.50 | **3,170.50** |
| 395 | 897.50 | 825.00 | 845.00 | 875.50 | **3,443.00** |
| 396 | 242.50 | 227.00 | 312.50 | 244.00 | **1,026.00** |

| | | | | | |
|---|---|---|---|---|---|
| 397 | 597.50 | 513.50 | 522.50 | 543.50 | **2,177.00** |
| **MS - MISSISSIPPI - Total** | **7,122.50** | **6,714.50** | **6,942.00** | **6,799.50** | **27,578.50** |

**STATE: MT - MONTANA**

| REGISTRANT ZIP CODE 3 | 1ST QUARTER | 2ND QUARTER | 3RD QUARTER | 4TH QUARTER | TOTAL GRAMS |
|---|---|---|---|---|---|
| 590 | 112.00 | 89.00 | 97.00 | 105.00 | **403.00** |
| 591 | 279.00 | 325.50 | 293.50 | 250.50 | **1,148.50** |
| 593 | 34.00 | 23.00 | 18.00 | 34.00 | **109.00** |
| 594 | 125.00 | 47.00 | 70.00 | 74.00 | **316.00** |
| 595 | 48.00 | 58.50 | 63.00 | 36.00 | **205.50** |
| 596 | 128.00 | 99.00 | 150.50 | 127.00 | **504.50** |
| 597 | 250.50 | 290.00 | 231.00 | 203.50 | **975.00** |
| 598 | 151.50 | 108.50 | 109.00 | 129.00 | **498.00** |
| 599 | 77.00 | 65.00 | 65.00 | 71.50 | **278.50** |
| **MT - MONTANA - Total** | **1,205.00** | **1,105.50** | **1,097.00** | **1,030.50** | **4,438.00** |

**STATE: NC - NORTH CAROLINA**

| REGISTRANT ZIP CODE 3 | 1ST QUARTER | 2ND QUARTER | 3RD QUARTER | 4TH QUARTER | TOTAL GRAMS |
|---|---|---|---|---|---|
| 270 | 3,166.00 | 2,938.00 | 3,299.50 | 3,080.00 | **12,483.50** |
| 271 | 3,291.50 | 3,302.00 | 3,253.00 | 3,280.00 | **13,126.50** |
| 272 | 5,032.50 | 5,066.00 | 5,316.00 | 4,984.00 | **20,398.50** |
| 273 | 3,321.00 | 3,346.00 | 3,173.50 | 3,372.00 | **13,212.50** |
| 274 | 2,454.00 | 2,565.50 | 2,613.50 | 3,111.00 | **10,744.00** |
| 275 | 9,204.00 | 8,566.40 | 8,389.85 | 8,150.50 | **34,310.75** |
| 276 | 3,229.00 | 3,279.50 | 3,113.00 | 3,299.50 | **12,921.00** |
| 277 | 2,025.00 | 1,981.00 | 2,229.00 | 1,994.00 | **8,229.00** |
| 278 | 3,310.00 | 3,285.00 | 3,692.50 | 3,153.00 | **13,440.50** |
| 279 | 1,140.00 | 1,167.00 | 1,214.50 | 1,147.00 | **4,668.50** |
| 280 | 3,593.50 | 3,479.50 | 3,280.50 | 3,389.00 | **13,742.50** |
| 281 | 3,720.00 | 3,586.50 | 3,462.00 | 3,754.50 | **14,523.00** |
| 282 | 2,165.00 | 2,020.00 | 2,262.50 | 2,303.50 | **8,751.00** |
| 283 | 5,539.50 | 5,157.50 | 5,581.00 | 5,183.50 | **21,461.50** |
| 284 | 3,961.00 | 3,650.00 | 3,623.50 | 3,066.50 | **14,301.00** |
| 285 | 5,018.00 | 5,163.50 | 5,666.50 | 5,425.50 | **21,273.50** |
| 286 | 5,008.00 | 4,543.50 | 4,858.00 | 4,830.50 | **19,240.00** |
| 287 | 3,339.50 | 3,286.00 | 2,958.50 | 3,214.00 | **12,798.00** |
| 288 | 1,037.50 | 1,005.50 | 1,109.00 | 993.50 | **4,145.50** |
| 289 | 92.00 | 110.00 | 84.00 | 80.00 | **366.00** |
| **NC - NORTH CAROLINA - Total** | **69,647.00** | **67,498.40** | **69,179.85** | **67,811.50** | **274,136.75** |

**STATE: ND - NORTH DAKOTA**

| REGISTRANT ZIP CODE 3 | 1ST QUARTER | 2ND QUARTER | 3RD QUARTER | 4TH QUARTER | TOTAL GRAMS |
|---|---|---|---|---|---|
| 580 | 94.00 | 64.50 | 93.00 | 102.50 | **354.00** |
| 581 | 66.00 | 87.00 | 19.00 | 25.00 | **197.00** |
| 582 | 243.50 | 195.50 | 206.00 | 236.00 | **881.00** |
| 583 | 12.50 | 22.50 | 17.50 | 21.50 | **74.00** |
| 584 | 39.00 | 61.00 | 10.00 | 30.00 | **140.00** |
| 585 | 478.00 | 443.00 | 399.50 | 493.50 | **1,814.00** |
| 586 | 92.00 | 77.50 | 91.50 | 81.50 | **342.50** |
| 587 | 35.00 | 45.00 | 10.00 | 60.00 | **150.00** |
| 588 | 0 | 11.00 | 0 | 32.00 | **43.00** |
| **ND - NORTH DAKOTA - Total** | **1,060.00** | **1,007.00** | **846.50** | **1,082.00** | **3,995.50** |

**STATE: NE - NEBRASKA**

| REGISTRANT ZIP CODE 3 | 1ST QUARTER | 2ND QUARTER | 3RD QUARTER | 4TH QUARTER | TOTAL GRAMS |
|---|---|---|---|---|---|
| 680 | 1,004.00 | 1,037.00 | 1,116.50 | 1,052.50 | **4,210.00** |
| 681 | 3,281.00 | 3,328.50 | 3,517.00 | 3,602.50 | **13,729.00** |
| 683 | 246.00 | 240.00 | 261.00 | 274.00 | **1,021.00** |
| 684 | 102.00 | 88.00 | 57.00 | 147.00 | **394.00** |
| 685 | 997.00 | 969.00 | 1,170.50 | 1,257.00 | **4,393.50** |
| 686 | 385.50 | 353.50 | 449.50 | 535.00 | **1,723.50** |
| 687 | 221.50 | 191.00 | 199.50 | 215.00 | **827.00** |
| 688 | 598.50 | 502.50 | 498.00 | 542.50 | **2,141.50** |
| 689 | 112.00 | 61.00 | 44.00 | 88.50 | **305.50** |

| | | | | | |
|---|---|---|---|---|---|
| 690 | 31.00 | 67.00 | 66.00 | 49.00 | **213.00** |
| 691 | 189.00 | 219.00 | 178.50 | 197.50 | **784.00** |
| 692 | 15.00 | 15.00 | 22.50 | 30.00 | **82.50** |
| 693 | 9.00 | 16.50 | 18.50 | 66.50 | **110.50** |
| **NE - NEBRASKA - Total** | **7,191.50** | **7,088.00** | **7,598.50** | **8,057.00** | **29,935.00** |
| **STATE: NH - NEW HAMPSHIRE** | | | | | |
| REGISTRANT ZIP CODE 3 | 1ST QUARTER | 2ND QUARTER | 3RD QUARTER | 4TH QUARTER | **TOTAL GRAMS** |
| 030 | 1,255.00 | 1,346.50 | 1,141.50 | 1,199.00 | **4,942.00** |
| 031 | 500.00 | 527.00 | 554.00 | 591.50 | **2,172.50** |
| 032 | 559.50 | 669.00 | 692.00 | 743.00 | **2,663.50** |
| 033 | 647.00 | 663.50 | 720.50 | 624.00 | **2,655.00** |
| 034 | 187.00 | 283.00 | 132.50 | 336.00 | **938.50** |
| 035 | 173.00 | 219.00 | 179.00 | 161.00 | **732.00** |
| 036 | 52.00 | 80.00 | 50.00 | 60.00 | **242.00** |
| 037 | 255.00 | 292.00 | 298.50 | 226.00 | **1,071.50** |
| 038 | 1,548.50 | 1,753.50 | 1,531.00 | 1,740.00 | **6,573.00** |
| **NH - NEW HAMPSHIRE - Total** | **5,177.00** | **5,833.50** | **5,299.00** | **5,680.50** | **21,990.00** |
| **STATE: NJ - NEW JERSEY** | | | | | |
| REGISTRANT ZIP CODE 3 | 1ST QUARTER | 2ND QUARTER | 3RD QUARTER | 4TH QUARTER | **TOTAL GRAMS** |
| 070 | 3,880.50 | 3,843.50 | 3,804.50 | 3,681.00 | **15,209.50** |
| 071 | 455.50 | 300.50 | 353.00 | 378.50 | **1,487.50** |
| 072 | 249.00 | 265.00 | 205.50 | 185.50 | **905.00** |
| 073 | 178.50 | 176.50 | 201.50 | 198.50 | **755.00** |
| 074 | 1,628.00 | 1,351.00 | 1,449.50 | 1,695.50 | **6,124.00** |
| 075 | 190.50 | 172.00 | 203.00 | 285.00 | **850.50** |
| 076 | 1,547.50 | 1,572.00 | 1,682.00 | 1,638.00 | **6,439.50** |
| 077 | 2,850.50 | 2,592.50 | 2,884.00 | 2,521.50 | **10,848.50** |
| 078 | 1,556.00 | 1,272.50 | 1,540.50 | 1,469.50 | **5,838.50** |
| 079 | 619.00 | 675.50 | 564.50 | 550.00 | **2,409.00** |
| 080 | 5,386.00 | 5,296.50 | 5,076.00 | 5,166.50 | **20,925.00** |
| 081 | 315.00 | 298.00 | 314.00 | 317.50 | **1,244.50** |
| 082 | 864.50 | 1,005.00 | 1,059.50 | 958.50 | **3,887.50** |
| 083 | 845.50 | 788.00 | 825.50 | 839.50 | **3,298.50** |
| 084 | 51.00 | 80.00 | 33.00 | 42.00 | **206.00** |
| 085 | 1,269.00 | 1,200.50 | 1,281.50 | 1,185.00 | **4,936.00** |
| 086 | 1,140.50 | 1,057.50 | 948.00 | 1,011.00 | **4,157.00** |
| 087 | 1,584.00 | 1,617.50 | 1,540.50 | 1,639.50 | **6,381.50** |
| 088 | 2,583.00 | 2,554.50 | 2,603.00 | 2,600.50 | **10,341.00** |
| 089 | 422.50 | 317.00 | 295.50 | 331.00 | **1,366.00** |
| **NJ - NEW JERSEY - Total** | **27,616.00** | **26,435.50** | **26,864.50** | **26,694.00** | **107,610.00** |
| **STATE: NM - NEW MEXICO** | | | | | |
| REGISTRANT ZIP CODE 3 | 1ST QUARTER | 2ND QUARTER | 3RD QUARTER | 4TH QUARTER | **TOTAL GRAMS** |
| 870 | 149.00 | 90.00 | 144.00 | 99.00 | **482.00** |
| 871 | 857.50 | 804.00 | 803.50 | 823.00 | **3,288.00** |
| 874 | 61.00 | 91.00 | 51.00 | 39.00 | **242.00** |
| 875 | 90.00 | 136.50 | 132.00 | 198.50 | **557.00** |
| 877 | 34.00 | 19.00 | 24.00 | 21.00 | **98.00** |
| 878 | 20.00 | 60.00 | 55.00 | 35.00 | **170.00** |
| 880 | 206.00 | 220.00 | 201.00 | 203.00 | **830.00** |
| 881 | 91.00 | 81.00 | 81.00 | 71.00 | **324.00** |
| 882 | 203.00 | 124.00 | 202.00 | 136.00 | **665.00** |
| 883 | 110.50 | 144.50 | 98.00 | 71.50 | **424.50** |
| 884 | 18.00 | 13.00 | 18.50 | 43.50 | **93.00** |
| **NM - NEW MEXICO - Total** | **1,840.00** | **1,783.00** | **1,810.00** | **1,740.50** | **7,173.50** |
| **STATE: NV - NEVADA** | | | | | |
| REGISTRANT ZIP CODE 3 | 1ST QUARTER | 2ND QUARTER | 3RD QUARTER | 4TH QUARTER | **TOTAL GRAMS** |
| 890 | 1,853.50 | 1,923.00 | 1,882.50 | 1,780.00 | **7,439.00** |
| 891 | 3,109.50 | 2,894.50 | 2,760.00 | 2,846.50 | **11,610.50** |
| 893 | 9.00 | 0 | 0 | 0 | **9.00** |
| 894 | 191.50 | 319.00 | 241.50 | 339.50 | **1,091.50** |

| | | | | | |
|---|---|---|---|---|---|
| 895 | 307.50 | 355.00 | 418.50 | 545.50 | **1,626.50** |
| 897 | 74.00 | 153.00 | 105.50 | 132.50 | **465.00** |
| 898 | 72.50 | 87.50 | 64.50 | 80.00 | **304.50** |
| **NV - NEVADA - Total** | **5,617.50** | **5,732.00** | **5,472.50** | **5,724.00** | **22,546.00** |

**STATE: NY - NEW YORK**

| REGISTRANT ZIP CODE 3 | 1ST QUARTER | 2ND QUARTER | 3RD QUARTER | 4TH QUARTER | TOTAL GRAMS |
|---|---|---|---|---|---|
| 100 | 2,445.50 | 2,068.00 | 2,140.50 | 2,321.50 | **8,975.50** |
| 101 | 30.00 | 50.00 | 30.00 | 10.00 | **120.00** |
| 102 | 5.00 | 0 | 8.00 | 0 | **13.00** |
| 103 | 1,548.50 | 1,641.00 | 1,334.00 | 1,551.50 | **6,075.00** |
| 104 | 854.00 | 787.50 | 805.50 | 858.50 | **3,305.50** |
| 105 | 864.00 | 825.50 | 839.50 | 963.00 | **3,492.00** |
| 106 | 90.50 | 66.50 | 60.00 | 104.50 | **321.50** |
| 107 | 316.00 | 345.00 | 302.50 | 314.50 | **1,278.00** |
| 108 | 164.50 | 115.50 | 74.00 | 76.50 | **430.50** |
| 109 | 1,235.00 | 1,211.00 | 1,219.00 | 1,302.00 | **4,967.00** |
| 110 | 1,273.00 | 1,272.50 | 1,121.50 | 1,343.50 | **5,010.50** |
| 111 | 549.50 | 450.50 | 340.00 | 422.00 | **1,762.00** |
| 112 | 1,883.00 | 1,790.00 | 1,856.50 | 1,924.00 | **7,453.50** |
| 113 | 1,815.50 | 1,725.00 | 1,665.00 | 1,849.50 | **7,055.00** |
| 114 | 559.50 | 633.50 | 604.00 | 534.00 | **2,331.00** |
| 115 | 3,103.50 | 2,825.00 | 3,272.00 | 3,330.00 | **12,530.50** |
| 116 | 56.00 | 47.00 | 47.00 | 53.00 | **203.00** |
| 117 | 9,523.50 | 9,231.50 | 10,041.50 | 9,679.50 | **38,476.00** |
| 118 | 470.00 | 632.00 | 670.50 | 500.50 | **2,273.00** |
| 119 | 1,009.50 | 1,026.50 | 1,041.50 | 1,188.50 | **4,266.00** |
| 120 | 414.50 | 399.50 | 422.00 | 345.50 | **1,581.50** |
| 121 | 284.00 | 258.50 | 346.00 | 378.00 | **1,266.50** |
| 122 | 574.50 | 507.00 | 641.50 | 535.00 | **2,258.00** |
| 123 | 181.00 | 195.00 | 142.50 | 89.00 | **607.50** |
| 124 | 461.00 | 427.00 | 405.50 | 403.00 | **1,696.50** |
| 125 | 1,281.00 | 1,167.50 | 1,290.50 | 1,220.00 | **4,959.00** |
| 126 | 331.00 | 130.00 | 250.00 | 247.00 | **958.00** |
| 127 | 360.50 | 360.00 | 312.50 | 325.00 | **1,358.00** |
| 128 | 901.50 | 837.00 | 857.50 | 786.50 | **3,382.50** |
| 129 | 524.00 | 498.50 | 468.00 | 448.50 | **1,939.00** |
| 130 | 3,197.50 | 3,136.00 | 2,914.00 | 3,078.50 | **12,326.00** |
| 131 | 607.00 | 568.00 | 505.00 | 540.50 | **2,220.50** |
| 132 | 1,041.00 | 1,183.50 | 1,225.00 | 1,229.00 | **4,678.50** |
| 133 | 842.50 | 880.50 | 855.50 | 839.00 | **3,417.50** |
| 134 | 1,075.00 | 977.00 | 998.00 | 1,083.50 | **4,133.50** |
| 135 | 434.00 | 398.00 | 413.50 | 377.50 | **1,623.00** |
| 136 | 1,988.00 | 1,909.50 | 1,764.50 | 1,702.50 | **7,364.50** |
| 137 | 608.50 | 642.00 | 494.00 | 501.00 | **2,245.50** |
| 138 | 1,262.50 | 1,114.50 | 1,043.00 | 954.00 | **4,374.00** |
| 139 | 634.00 | 521.00 | 543.00 | 598.50 | **2,296.50** |
| 140 | 3,140.50 | 3,099.50 | 3,088.50 | 2,916.50 | **12,245.00** |
| 141 | 1,181.50 | 1,146.00 | 1,115.00 | 1,146.50 | **4,589.00** |
| 142 | 3,208.00 | 3,239.50 | 3,070.00 | 3,078.00 | **12,595.50** |
| 143 | 267.00 | 285.00 | 281.00 | 281.00 | **1,114.00** |
| 144 | 1,441.00 | 1,341.00 | 1,270.50 | 1,438.50 | **5,491.00** |
| 145 | 965.50 | 860.00 | 931.50 | 899.50 | **3,656.50** |
| 146 | 1,349.50 | 1,467.00 | 1,430.00 | 1,761.00 | **6,007.50** |
| 147 | 413.50 | 363.50 | 454.00 | 376.00 | **1,607.00** |
| 148 | 1,090.50 | 1,200.50 | 1,224.50 | 1,173.00 | **4,688.50** |
| 149 | 161.50 | 142.00 | 125.50 | 116.50 | **545.50** |
| **NY - NEW YORK - Total** | **58,017.50** | **55,997.50** | **56,354.50** | **57,194.50** | **227,564.00** |

**STATE: OH - OHIO**

| REGISTRANT ZIP CODE 3 | 1ST QUARTER | 2ND QUARTER | 3RD QUARTER | 4TH QUARTER | TOTAL GRAMS |
|---|---|---|---|---|---|
| 430 | 2,189.50 | 1,901.50 | 2,016.50 | 1,964.00 | **8,071.50** |

| | | | | | |
|---|---|---|---|---|---|
| 431 | 1,522.00 | 1,445.00 | 1,570.50 | 1,391.00 | **5,928.50** |
| 432 | 1,741.00 | 1,640.50 | 1,841.00 | 1,625.00 | **6,847.50** |
| 433 | 444.00 | 527.50 | 466.00 | 410.00 | **1,847.50** |
| 434 | 554.50 | 624.00 | 617.00 | 493.00 | **2,288.50** |
| 435 | 1,648.50 | 1,763.50 | 1,822.50 | 1,604.50 | **6,839.00** |
| 436 | 2,134.00 | 1,825.50 | 1,914.50 | 2,017.00 | **7,891.00** |
| 437 | 170.00 | 233.00 | 234.00 | 189.50 | **826.50** |
| 438 | 119.00 | 145.00 | 132.00 | 167.00 | **563.00** |
| 439 | 653.00 | 587.50 | 533.00 | 564.00 | **2,337.50** |
| 440 | 2,851.50 | 2,528.50 | 2,619.00 | 2,731.50 | **10,730.50** |
| 441 | 1,976.50 | 2,047.50 | 2,042.50 | 2,097.00 | **8,163.50** |
| 442 | 1,833.50 | 1,750.50 | 1,677.00 | 1,968.50 | **7,229.50** |
| 443 | 610.00 | 470.50 | 623.00 | 618.00 | **2,321.50** |
| 444 | 756.00 | 596.00 | 662.00 | 562.00 | **2,576.00** |
| 445 | 396.00 | 363.00 | 372.50 | 387.00 | **1,518.50** |
| 446 | 2,245.50 | 2,350.50 | 2,163.00 | 2,338.50 | **9,097.50** |
| 447 | 1,099.00 | 955.50 | 808.50 | 1,004.50 | **3,867.50** |
| 448 | 235.50 | 237.50 | 231.00 | 231.50 | **935.50** |
| 449 | 144.00 | 167.00 | 170.00 | 132.00 | **613.00** |
| 450 | 2,114.00 | 2,009.00 | 2,049.00 | 2,005.00 | **8,177.00** |
| 451 | 740.50 | 839.50 | 633.00 | 581.00 | **2,794.00** |
| 452 | 1,824.50 | 1,842.00 | 1,693.50 | 1,529.00 | **6,889.00** |
| 453 | 1,350.00 | 1,313.00 | 1,348.00 | 1,317.00 | **5,328.00** |
| 454 | 1,006.00 | 1,006.00 | 1,027.50 | 856.50 | **3,896.00** |
| 455 | 442.50 | 494.00 | 408.00 | 466.50 | **1,811.00** |
| 456 | 824.50 | 765.50 | 855.00 | 868.50 | **3,313.50** |
| 457 | 118.50 | 153.00 | 142.50 | 198.00 | **612.00** |
| 458 | 583.50 | 604.50 | 527.50 | 528.50 | **2,244.00** |
| OH - OHIO - Total | 32,327.00 | 31,186.00 | 31,199.50 | 30,845.50 | 125,558.00 |

**STATE: OK - OKLAHOMA**

| REGISTRANT ZIP CODE 3 | 1ST QUARTER | 2ND QUARTER | 3RD QUARTER | 4TH QUARTER | TOTAL GRAMS |
|---|---|---|---|---|---|
| 730 | 1,823.00 | 1,711.50 | 1,586.50 | 1,618.00 | **6,739.00** |
| 731 | 1,173.50 | 1,325.00 | 1,230.00 | 1,140.50 | **4,869.00** |
| 734 | 311.00 | 324.50 | 277.00 | 287.50 | **1,200.00** |
| 735 | 469.00 | 497.00 | 384.50 | 371.00 | **1,721.50** |
| 736 | 57.00 | 34.00 | 34.00 | 39.00 | **164.00** |
| 737 | 55.00 | 30.00 | 36.00 | 78.00 | **199.00** |
| 738 | 29.00 | 25.50 | 18.00 | 0 | **72.50** |
| 739 | 10.00 | 10.00 | 0 | 0 | **20.00** |
| 740 | 1,747.50 | 1,873.00 | 1,606.50 | 1,581.50 | **6,808.50** |
| 741 | 706.00 | 711.00 | 723.00 | 637.00 | **2,777.00** |
| 743 | 147.00 | 139.00 | 144.00 | 174.00 | **604.00** |
| 744 | 425.00 | 387.50 | 450.50 | 399.50 | **1,662.50** |
| 745 | 28.50 | 37.50 | 19.50 | 24.00 | **109.50** |
| 746 | 38.00 | 41.00 | 42.00 | 24.00 | **145.00** |
| 747 | 49.00 | 43.00 | 32.00 | 17.00 | **141.00** |
| 748 | 192.00 | 186.00 | 238.50 | 236.00 | **852.50** |
| 749 | 25.00 | 31.00 | 32.00 | 33.00 | **121.00** |
| OK - OKLAHOMA - Total | 7,285.50 | 7,406.50 | 6,854.00 | 6,660.00 | 28,206.00 |

**STATE: OR - OREGON**

| REGISTRANT ZIP CODE 3 | 1ST QUARTER | 2ND QUARTER | 3RD QUARTER | 4TH QUARTER | TOTAL GRAMS |
|---|---|---|---|---|---|
| 970 | 1,208.50 | 1,102.00 | 1,001.00 | 1,008.50 | **4,320.00** |
| 971 | 81.00 | 55.00 | 86.50 | 97.00 | **319.50** |
| 972 | 542.00 | 486.00 | 440.00 | 445.00 | **1,913.00** |
| 973 | 803.00 | 712.50 | 700.50 | 700.00 | **2,916.00** |
| 974 | 622.50 | 594.00 | 664.00 | 577.00 | **2,457.50** |
| 975 | 677.50 | 628.50 | 570.50 | 654.00 | **2,530.50** |
| 976 | 86.00 | 57.00 | 106.00 | 163.00 | **412.00** |
| 977 | 221.50 | 193.50 | 255.50 | 246.50 | **917.00** |
| 978 | 43.00 | 90.00 | 91.50 | 60.50 | **285.00** |

| | | | | | |
|---|---|---|---|---|---|
| 979 | 3.00 | 0 | 0 | 9.00 | **12.00** |
| **OR - OREGON - Total** | **4,288.00** | **3,918.50** | **3,915.50** | **3,960.50** | **16,082.50** |

**STATE: PA - PENNSYLVANIA**

| REGISTRANT ZIP CODE 3 | 1ST QUARTER | 2ND QUARTER | 3RD QUARTER | 4TH QUARTER | TOTAL GRAMS |
|---|---|---|---|---|---|
| 150 | 732.00 | 641.00 | 723.00 | 643.00 | **2,739.00** |
| 151 | 610.00 | 569.50 | 606.50 | 578.00 | **2,364.00** |
| 152 | 545.50 | 442.50 | 546.00 | 450.50 | **1,984.50** |
| 153 | 461.00 | 496.00 | 501.00 | 566.50 | **2,024.50** |
| 154 | 166.50 | 140.50 | 160.00 | 160.00 | **627.00** |
| 155 | 208.50 | 187.50 | 185.00 | 217.50 | **798.50** |
| 156 | 271.00 | 242.00 | 294.00 | 219.00 | **1,026.00** |
| 157 | 183.00 | 221.00 | 232.00 | 207.00 | **843.00** |
| 158 | 159.00 | 169.50 | 124.50 | 168.50 | **621.50** |
| 159 | 162.50 | 84.00 | 139.00 | 105.00 | **490.50** |
| 160 | 163.50 | 128.50 | 184.50 | 139.50 | **616.00** |
| 161 | 373.50 | 393.50 | 445.00 | 420.50 | **1,632.50** |
| 162 | 219.00 | 222.50 | 172.50 | 154.50 | **768.50** |
| 163 | 407.50 | 322.00 | 375.00 | 328.00 | **1,432.50** |
| 164 | 175.00 | 150.00 | 168.00 | 119.00 | **612.00** |
| 165 | 649.50 | 647.50 | 609.50 | 599.50 | **2,506.00** |
| 166 | 404.50 | 470.00 | 478.50 | 525.00 | **1,878.00** |
| 167 | 38.50 | 27.00 | 27.50 | 15.00 | **108.00** |
| 168 | 413.00 | 363.50 | 397.00 | 391.50 | **1,565.00** |
| 169 | 90.00 | 73.00 | 82.00 | 94.00 | **339.00** |
| 170 | 1,774.00 | 1,593.50 | 1,458.50 | 1,507.00 | **6,333.00** |
| 171 | 491.50 | 346.00 | 491.00 | 543.00 | **1,871.50** |
| 172 | 575.50 | 564.00 | 567.50 | 629.50 | **2,336.50** |
| 173 | 1,802.00 | 1,582.50 | 1,624.50 | 1,781.50 | **6,790.50** |
| 174 | 545.50 | 385.50 | 347.50 | 345.50 | **1,624.00** |
| 175 | 339.00 | 285.50 | 282.00 | 352.00 | **1,258.50** |
| 176 | 386.50 | 516.50 | 385.50 | 429.00 | **1,717.50** |
| 177 | 115.00 | 126.00 | 92.00 | 124.00 | **457.00** |
| 178 | 122.50 | 157.00 | 196.50 | 185.00 | **661.00** |
| 179 | 208.00 | 270.50 | 261.00 | 281.00 | **1,020.50** |
| 180 | 3,390.50 | 3,422.50 | 3,429.00 | 3,205.50 | **13,447.50** |
| 181 | 1,906.00 | 1,520.00 | 1,718.50 | 1,369.00 | **6,513.50** |
| 182 | 582.00 | 465.50 | 587.00 | 476.50 | **2,111.00** |
| 183 | 1,701.00 | 1,457.00 | 1,588.50 | 1,573.00 | **6,319.50** |
| 184 | 722.00 | 834.00 | 810.00 | 804.50 | **3,170.50** |
| 185 | 560.00 | 451.50 | 443.50 | 435.00 | **1,890.00** |
| 186 | 290.00 | 262.00 | 272.50 | 299.50 | **1,124.00** |
| 187 | 5,451.00 | 5,507.00 | 5,572.00 | 4,899.00 | **21,429.00** |
| 188 | 99.00 | 145.00 | 99.00 | 130.00 | **473.00** |
| 189 | 1,452.00 | 1,290.00 | 1,302.50 | 1,267.50 | **5,312.00** |
| 190 | 5,614.50 | 5,377.00 | 5,714.50 | 5,426.00 | **22,132.00** |
| 191 | 1,169.00 | 1,122.00 | 1,162.00 | 1,055.00 | **4,508.00** |
| 193 | 1,482.50 | 1,390.50 | 1,466.00 | 1,568.50 | **5,907.50** |
| 194 | 1,575.50 | 1,739.50 | 1,604.50 | 1,627.50 | **6,547.00** |
| 195 | 896.50 | 845.00 | 709.50 | 687.50 | **3,138.50** |
| 196 | 662.00 | 669.00 | 728.00 | 727.00 | **2,786.00** |
| **PA - PENNSYLVANIA - Total** | **40,346.00** | **38,315.50** | **39,363.50** | **37,830.00** | **155,855.00** |

**STATE: PR - PUERTO RICO**

| REGISTRANT ZIP CODE 3 | 1ST QUARTER | 2ND QUARTER | 3RD QUARTER | 4TH QUARTER | TOTAL GRAMS |
|---|---|---|---|---|---|
| 006 | 60.00 | 40.00 | 52.50 | 65.00 | **217.50** |
| 007 | 11.00 | 12.00 | 17.00 | 29.00 | **69.00** |
| 009 | 219.50 | 202.50 | 259.00 | 275.50 | **956.50** |
| **PR - PUERTO RICO - Total** | **290.50** | **254.50** | **328.50** | **369.50** | **1,243.00** |

**STATE: RI - RHODE ISLAND**

| REGISTRANT ZIP CODE 3 | 1ST QUARTER | 2ND QUARTER | 3RD QUARTER | 4TH QUARTER | TOTAL GRAMS |
|---|---|---|---|---|---|
| 028 | 1,216.50 | 1,184.50 | 1,081.00 | 1,079.50 | **4,561.50** |

| | | | | | |
|---|---|---|---|---|---|
| 029 | 333.00 | 427.00 | 477.00 | 360.00 | **1,597.00** |
| **RI - RHODE ISLAND - Total** | **1,549.50** | **1,611.50** | **1,558.00** | **1,439.50** | **6,158.50** |

**STATE: SC - SOUTH CAROLINA**

| REGISTRANT ZIP CODE 3 | 1ST QUARTER | 2ND QUARTER | 3RD QUARTER | 4TH QUARTER | TOTAL GRAMS |
|---|---|---|---|---|---|
| 290 | 2,193.00 | 1,958.00 | 1,943.00 | 2,170.00 | **8,264.00** |
| 291 | 1,809.00 | 1,824.00 | 1,778.00 | 1,880.00 | **7,291.00** |
| 292 | 1,610.50 | 1,865.00 | 1,660.00 | 2,315.00 | **7,450.50** |
| 293 | 1,750.00 | 1,887.50 | 1,743.50 | 1,757.00 | **7,138.00** |
| 294 | 2,814.00 | 2,221.50 | 2,493.50 | 2,127.50 | **9,656.50** |
| 295 | 3,302.50 | 2,895.50 | 3,094.50 | 3,151.50 | **12,444.00** |
| 296 | 7,921.50 | 7,643.00 | 7,705.00 | 8,009.50 | **31,279.00** |
| 297 | 1,078.00 | 1,084.50 | 1,440.00 | 1,238.00 | **4,840.50** |
| 298 | 1,057.50 | 999.50 | 1,078.50 | 833.50 | **3,969.00** |
| 299 | 1,552.50 | 1,402.50 | 1,441.00 | 1,616.50 | **6,012.50** |
| **SC - SOUTH CAROLINA - Total** | **25,088.50** | **23,781.00** | **24,377.00** | **25,098.50** | **98,345.00** |

**STATE: SD - SOUTH DAKOTA**

| REGISTRANT ZIP CODE 3 | 1ST QUARTER | 2ND QUARTER | 3RD QUARTER | 4TH QUARTER | TOTAL GRAMS |
|---|---|---|---|---|---|
| 570 | 486.00 | 484.00 | 526.00 | 486.00 | **1,982.00** |
| 571 | 741.00 | 664.00 | 776.50 | 539.00 | **2,720.50** |
| 572 | 97.00 | 97.00 | 111.00 | 124.00 | **429.00** |
| 573 | 226.00 | 203.00 | 179.00 | 171.00 | **779.00** |
| 574 | 419.00 | 442.00 | 419.50 | 356.50 | **1,637.00** |
| 575 | 91.00 | 59.00 | 45.00 | 80.50 | **275.50** |
| 576 | 70.00 | 86.00 | 5.00 | 55.00 | **216.00** |
| 577 | 463.50 | 447.00 | 400.00 | 415.00 | **1,725.50** |
| **SD - SOUTH DAKOTA - Total** | **2,593.50** | **2,482.00** | **2,462.00** | **2,227.00** | **9,764.50** |

**STATE: TN - TENNESSEE**

| REGISTRANT ZIP CODE 3 | 1ST QUARTER | 2ND QUARTER | 3RD QUARTER | 4TH QUARTER | TOTAL GRAMS |
|---|---|---|---|---|---|
| 370 | 3,693.50 | 3,542.00 | 3,436.50 | 3,403.50 | **14,075.50** |
| 371 | 3,829.00 | 4,020.00 | 3,958.00 | 4,120.00 | **15,927.00** |
| 372 | 1,755.50 | 1,419.50 | 1,120.50 | 1,150.50 | **5,446.00** |
| 373 | 1,639.00 | 1,748.00 | 1,813.50 | 1,785.00 | **6,985.50** |
| 374 | 1,119.50 | 1,108.00 | 1,125.50 | 1,169.00 | **4,522.00** |
| 376 | 2,442.00 | 2,196.50 | 2,168.00 | 2,170.50 | **8,977.00** |
| 377 | 678.50 | 599.00 | 509.50 | 645.00 | **2,432.00** |
| 378 | 1,264.50 | 1,275.00 | 1,136.00 | 1,293.50 | **4,969.00** |
| 379 | 724.50 | 653.50 | 614.00 | 564.50 | **2,556.50** |
| 380 | 828.50 | 917.50 | 995.50 | 1,040.00 | **3,781.50** |
| 381 | 1,161.00 | 1,168.00 | 1,066.50 | 1,217.00 | **4,612.50** |
| 382 | 275.00 | 368.00 | 355.50 | 316.50 | **1,315.00** |
| 383 | 690.50 | 664.50 | 756.00 | 832.50 | **2,943.50** |
| 384 | 588.00 | 699.50 | 610.00 | 659.00 | **2,556.50** |
| 385 | 674.50 | 556.00 | 564.00 | 515.50 | **2,310.00** |
| **TN - TENNESSEE - Total** | **21,363.50** | **20,935.00** | **20,229.00** | **20,882.00** | **83,409.50** |

**STATE: TX - TEXAS**

| REGISTRANT ZIP CODE 3 | 1ST QUARTER | 2ND QUARTER | 3RD QUARTER | 4TH QUARTER | TOTAL GRAMS |
|---|---|---|---|---|---|
| 750 | 7,529.00 | 7,188.00 | 6,809.50 | 6,255.00 | **27,781.50** |
| 751 | 2,128.50 | 1,873.50 | 2,241.00 | 2,402.00 | **8,645.00** |
| 752 | 1,780.50 | 1,544.00 | 1,852.00 | 2,077.00 | **7,253.50** |
| 754 | 670.00 | 812.00 | 773.50 | 755.50 | **3,011.00** |
| 755 | 70.50 | 57.00 | 60.00 | 102.00 | **289.50** |
| 756 | 238.00 | 210.50 | 216.00 | 246.50 | **911.00** |
| 757 | 478.00 | 553.50 | 556.00 | 535.00 | **2,122.50** |
| 758 | 156.00 | 211.00 | 198.00 | 129.50 | **694.50** |
| 759 | 163.00 | 122.00 | 121.50 | 105.00 | **511.50** |
| 760 | 3,068.50 | 2,755.00 | 2,867.50 | 2,937.50 | **11,628.50** |
| 761 | 2,242.00 | 1,939.00 | 2,023.00 | 1,936.00 | **8,140.00** |
| 762 | 1,243.50 | 1,505.00 | 1,403.00 | 1,688.50 | **5,840.00** |
| 763 | 719.00 | 782.00 | 737.00 | 792.00 | **3,030.00** |
| 764 | 126.50 | 159.50 | 190.50 | 153.50 | **630.00** |

| | | | | | |
|---|---|---|---|---|---|
| 765 | 1,157.00 | 1,006.00 | 906.00 | 879.00 | **3,948.00** |
| 766 | 166.00 | 73.00 | 112.00 | 89.00 | **440.00** |
| 767 | 210.00 | 177.00 | 246.00 | 222.00 | **855.00** |
| 768 | 49.00 | 54.50 | 47.00 | 46.00 | **196.50** |
| 769 | 162.00 | 134.00 | 152.00 | 139.00 | **587.00** |
| 770 | 2,472.00 | 2,223.00 | 2,466.50 | 2,229.50 | **9,391.00** |
| 773 | 2,717.00 | 2,447.50 | 2,568.50 | 2,941.00 | **10,674.00** |
| 774 | 3,508.00 | 3,434.00 | 3,175.50 | 2,844.50 | **12,962.00** |
| 775 | 3,072.50 | 3,066.50 | 3,287.50 | 2,970.50 | **12,397.00** |
| 776 | 435.50 | 399.00 | 446.00 | 416.00 | **1,696.50** |
| 777 | 194.50 | 165.00 | 162.50 | 164.00 | **686.00** |
| 778 | 320.00 | 250.50 | 365.00 | 366.50 | **1,302.00** |
| 779 | 448.00 | 458.00 | 546.00 | 506.00 | **1,958.00** |
| 780 | 594.00 | 568.00 | 693.00 | 797.50 | **2,652.50** |
| 781 | 1,336.00 | 1,143.50 | 1,150.00 | 1,216.00 | **4,845.50** |
| 782 | 5,166.00 | 5,494.00 | 5,696.50 | 5,671.50 | **22,028.00** |
| 783 | 422.50 | 389.50 | 504.00 | 364.00 | **1,680.00** |
| 784 | 785.00 | 648.50 | 578.00 | 620.50 | **2,632.00** |
| 785 | 286.50 | 318.00 | 290.00 | 329.00 | **1,223.50** |
| 786 | 2,564.00 | 2,544.50 | 2,737.00 | 2,503.00 | **10,348.50** |
| 787 | 2,643.50 | 2,342.00 | 2,657.00 | 2,477.00 | **10,119.50** |
| 788 | 62.00 | 36.00 | 66.00 | 76.00 | **240.00** |
| 789 | 98.00 | 106.00 | 78.00 | 102.00 | **384.00** |
| 790 | 243.00 | 235.00 | 110.00 | 206.50 | **794.50** |
| 791 | 700.00 | 471.50 | 654.00 | 530.50 | **2,356.00** |
| 792 | 0 | 0 | 9.00 | 5.00 | **14.00** |
| 793 | 60.00 | 60.00 | 77.50 | 85.00 | **282.50** |
| 794 | 805.00 | 671.50 | 740.00 | 795.00 | **3,011.50** |
| 795 | 15.00 | 20.00 | 15.00 | 20.00 | **70.00** |
| 796 | 99.00 | 61.50 | 89.50 | 71.50 | **321.50** |
| 797 | 87.00 | 81.00 | 172.50 | 258.50 | **599.00** |
| 798 | 30.00 | 48.00 | 48.00 | 44.00 | **170.00** |
| 799 | 1,429.00 | 1,629.00 | 1,439.00 | 1,505.50 | **6,002.50** |
| **TX - TEXAS - Total** | **52,950.00** | **50,467.50** | **52,333.00** | **51,606.00** | **207,356.50** |

**STATE: UT - UTAH**

| REGISTRANT ZIP CODE 3 | 1ST QUARTER | 2ND QUARTER | 3RD QUARTER | 4TH QUARTER | TOTAL GRAMS |
|---|---|---|---|---|---|
| 840 | 3,820.50 | 3,942.00 | 3,826.50 | 3,405.00 | **14,994.00** |
| 841 | 959.00 | 962.50 | 929.50 | 1,015.50 | **3,866.50** |
| 843 | 241.00 | 159.00 | 122.00 | 190.50 | **712.50** |
| 844 | 817.00 | 831.00 | 838.50 | 822.00 | **3,308.50** |
| 845 | 453.50 | 429.50 | 467.50 | 485.00 | **1,835.50** |
| 846 | 635.50 | 722.00 | 656.00 | 680.00 | **2,693.50** |
| 847 | 1,147.00 | 1,132.50 | 1,091.00 | 1,132.50 | **4,503.00** |
| **UT - UTAH - Total** | **8,073.50** | **8,178.50** | **7,931.00** | **7,730.50** | **31,913.50** |

**STATE: VA - VIRGINIA**

| REGISTRANT ZIP CODE 3 | 1ST QUARTER | 2ND QUARTER | 3RD QUARTER | 4TH QUARTER | TOTAL GRAMS |
|---|---|---|---|---|---|
| 201 | 2,775.50 | 2,663.50 | 2,911.50 | 2,670.00 | **11,020.50** |
| 220 | 1,288.00 | 1,288.50 | 1,466.50 | 1,054.00 | **5,097.00** |
| 221 | 1,089.00 | 1,171.50 | 1,219.50 | 1,174.00 | **4,654.00** |
| 222 | 879.00 | 635.00 | 674.00 | 675.00 | **2,863.00** |
| 223 | 813.50 | 898.00 | 766.00 | 750.00 | **3,227.50** |
| 224 | 894.00 | 940.50 | 836.00 | 962.50 | **3,633.00** |
| 225 | 438.50 | 426.00 | 455.50 | 335.00 | **1,655.00** |
| 226 | 1,069.00 | 1,097.50 | 1,188.50 | 1,094.50 | **4,449.50** |
| 227 | 172.00 | 135.50 | 170.50 | 158.50 | **636.50** |
| 228 | 574.50 | 544.00 | 624.00 | 618.50 | **2,361.00** |
| 229 | 301.00 | 280.50 | 241.00 | 307.50 | **1,130.00** |
| 230 | 207.00 | 201.00 | 152.00 | 222.00 | **782.00** |
| 231 | 745.00 | 707.50 | 646.50 | 614.50 | **2,713.50** |
| 232 | 835.00 | 753.50 | 771.50 | 607.00 | **2,967.00** |

| | | | | | |
|---|---|---|---|---|---|
| 233 | 1,534.50 | 1,694.50 | 1,481.00 | 1,519.00 | **6,229.00** |
| 234 | 3,358.00 | 3,163.50 | 3,040.50 | 3,265.50 | **12,827.50** |
| 235 | 678.50 | 870.00 | 839.50 | 753.00 | **3,141.00** |
| 236 | 914.00 | 936.00 | 851.00 | 891.50 | **3,592.50** |
| 237 | 351.00 | 426.00 | 525.00 | 415.00 | **1,717.00** |
| 238 | 717.00 | 761.50 | 757.50 | 664.00 | **2,900.00** |
| 239 | 164.00 | 192.00 | 177.50 | 218.00 | **751.50** |
| 240 | 988.00 | 829.00 | 891.50 | 979.50 | **3,688.00** |
| 241 | 753.00 | 904.50 | 915.00 | 1,008.00 | **3,580.50** |
| 242 | 701.50 | 681.50 | 584.50 | 541.00 | **2,508.50** |
| 243 | 637.50 | 568.00 | 709.00 | 572.50 | **2,487.00** |
| 244 | 411.00 | 292.00 | 343.00 | 354.50 | **1,400.50** |
| 245 | 946.00 | 1,042.00 | 925.00 | 775.00 | **3,688.00** |
| 246 | 96.00 | 77.50 | 69.00 | 63.00 | **305.50** |
| **VA - VIRGINIA - Total** | **24,331.00** | **24,180.50** | **24,232.00** | **23,262.50** | **96,006.00** |

**STATE: VI - VIRGIN ISLANDS**

| REGISTRANT ZIP CODE 3 | 1ST QUARTER | 2ND QUARTER | 3RD QUARTER | 4TH QUARTER | TOTAL GRAMS |
|---|---|---|---|---|---|
| 008 | 7.50 | 15.00 | 15.00 | 7.50 | **45.00** |
| **VI - VIRGIN ISLANDS - Total** | **7.50** | **15.00** | **15.00** | **7.50** | **45.00** |

**STATE: VT - VERMONT**

| REGISTRANT ZIP CODE 3 | 1ST QUARTER | 2ND QUARTER | 3RD QUARTER | 4TH QUARTER | TOTAL GRAMS |
|---|---|---|---|---|---|
| 050 | 60.00 | 0 | 0 | 35.00 | **95.00** |
| 051 | 99.50 | 38.50 | 0 | 7.50 | **145.50** |
| 052 | 148.00 | 150.00 | 134.00 | 189.00 | **621.00** |
| 054 | 225.50 | 233.50 | 229.50 | 266.00 | **954.50** |
| 056 | 44.00 | 26.00 | 49.00 | 40.00 | **159.00** |
| 057 | 167.00 | 192.00 | 211.00 | 287.50 | **857.50** |
| 058 | 15.00 | 35.00 | 40.00 | 30.00 | **120.00** |
| **VT - VERMONT - Total** | **759.00** | **675.00** | **663.50** | **855.00** | **2,952.50** |

**STATE: WA - WASHINGTON**

| REGISTRANT ZIP CODE 3 | 1ST QUARTER | 2ND QUARTER | 3RD QUARTER | 4TH QUARTER | TOTAL GRAMS |
|---|---|---|---|---|---|
| 980 | 1,440.50 | 1,336.00 | 1,340.50 | 1,256.00 | **5,373.00** |
| 981 | 886.50 | 672.50 | 782.50 | 693.50 | **3,035.00** |
| 982 | 697.00 | 734.00 | 716.50 | 769.50 | **2,917.00** |
| 983 | 1,404.00 | 1,525.50 | 1,696.50 | 1,695.00 | **6,321.00** |
| 984 | 469.00 | 493.50 | 403.50 | 545.00 | **1,911.00** |
| 985 | 558.50 | 508.50 | 551.00 | 503.50 | **2,121.50** |
| 986 | 989.00 | 1,110.00 | 1,046.50 | 981.50 | **4,127.00** |
| 988 | 386.00 | 301.00 | 310.50 | 272.00 | **1,269.50** |
| 989 | 341.50 | 255.50 | 280.00 | 398.00 | **1,275.00** |
| 990 | 302.00 | 314.00 | 338.00 | 354.00 | **1,308.00** |
| 991 | 60.00 | 100.00 | 98.00 | 142.00 | **400.00** |
| 992 | 2,610.50 | 2,389.50 | 2,443.50 | 2,110.50 | **9,554.00** |
| 993 | 828.00 | 770.00 | 924.00 | 878.00 | **3,400.00** |
| 994 | 38.00 | 18.00 | 17.00 | 57.00 | **130.00** |
| **WA - WASHINGTON - Total** | **11,010.50** | **10,528.00** | **10,948.00** | **10,655.50** | **43,142.00** |

**STATE: WI - WISCONSIN**

| REGISTRANT ZIP CODE 3 | 1ST QUARTER | 2ND QUARTER | 3RD QUARTER | 4TH QUARTER | TOTAL GRAMS |
|---|---|---|---|---|---|
| 530 | 1,047.50 | 944.00 | 1,136.00 | 979.00 | **4,106.50** |
| 531 | 1,125.00 | 1,240.50 | 1,303.00 | 1,511.00 | **5,179.50** |
| 532 | 1,260.50 | 1,220.50 | 1,316.00 | 1,191.50 | **4,988.50** |
| 534 | 367.50 | 400.50 | 422.00 | 471.00 | **1,661.00** |
| 535 | 503.00 | 531.00 | 538.50 | 506.00 | **2,078.50** |
| 537 | 141.25 | 123.00 | 119.50 | 90.75 | **474.50** |
| 538 | 0 | 0 | 5.00 | 5.00 | **10.00** |
| 539 | 223.00 | 174.50 | 224.00 | 132.50 | **754.00** |
| 540 | 98.00 | 99.00 | 115.00 | 190.00 | **502.00** |
| 541 | 328.00 | 392.50 | 332.50 | 382.00 | **1,435.00** |
| 542 | 94.50 | 123.50 | 126.00 | 127.00 | **471.00** |
| 543 | 353.00 | 325.50 | 344.50 | 352.50 | **1,375.50** |

| | | | | | |
|---|---|---|---|---|---|
| 544 | 539.50 | 516.00 | 461.50 | 430.50 | **1,947.50** |
| 545 | 114.50 | 163.50 | 164.00 | 114.50 | **556.50** |
| 546 | 52.00 | 33.00 | 46.00 | 52.00 | **183.00** |
| 547 | 156.00 | 117.00 | 155.00 | 132.00 | **560.00** |
| 548 | 93.00 | 56.00 | 57.00 | 69.00 | **275.00** |
| 549 | 953.00 | 843.50 | 910.00 | 921.00 | **3,627.50** |
| **WI - WISCONSIN - Total** | **7,449.25** | **7,303.50** | **7,775.50** | **7,657.25** | **30,185.50** |

**STATE: WV - WEST VIRGINIA**

| REGISTRANT ZIP CODE 3 | 1ST QUARTER | 2ND QUARTER | 3RD QUARTER | 4TH QUARTER | TOTAL GRAMS |
|---|---|---|---|---|---|
| 247 | 134.00 | 129.00 | 129.00 | 71.00 | **463.00** |
| 249 | 36.00 | 36.00 | 36.00 | 36.00 | **144.00** |
| 250 | 95.00 | 82.00 | 100.00 | 95.00 | **372.00** |
| 251 | 120.00 | 95.00 | 80.00 | 90.00 | **385.00** |
| 252 | 47.00 | 29.00 | 48.00 | 71.00 | **195.00** |
| 253 | 54.00 | 74.00 | 77.00 | 73.00 | **278.00** |
| 254 | 907.50 | 765.50 | 796.00 | 747.00 | **3,216.00** |
| 255 | 369.50 | 326.00 | 342.50 | 317.00 | **1,355.00** |
| 256 | 36.00 | 27.00 | 71.00 | 39.00 | **173.00** |
| 257 | 261.00 | 308.00 | 337.00 | 258.00 | **1,164.00** |
| 258 | 40.00 | 85.00 | 50.00 | 108.00 | **283.00** |
| 259 | 5.00 | 15.00 | 5.00 | 10.00 | **35.00** |
| 260 | 897.00 | 842.00 | 784.50 | 845.50 | **3,369.00** |
| 261 | 158.00 | 158.00 | 130.00 | 131.00 | **577.00** |
| 262 | 0 | 0 | 5.00 | 0 | **5.00** |
| 263 | 30.00 | 18.00 | 12.00 | 6.00 | **66.00** |
| 265 | 90.00 | 120.00 | 107.00 | 74.50 | **391.50** |
| 266 | 47.00 | 54.00 | 28.50 | 30.00 | **159.50** |
| 267 | 123.50 | 159.50 | 201.00 | 188.00 | **672.00** |
| 268 | 26.00 | 25.00 | 13.00 | 15.00 | **79.00** |
| **WV - WEST VIRGINIA - Total** | **3,476.50** | **3,348.00** | **3,352.50** | **3,205.00** | **13,382.00** |

**STATE: WY - WYOMING**

| REGISTRANT ZIP CODE 3 | 1ST QUARTER | 2ND QUARTER | 3RD QUARTER | 4TH QUARTER | TOTAL GRAMS |
|---|---|---|---|---|---|
| 820 | 543.50 | 349.00 | 410.00 | 369.00 | **1,671.50** |
| 822 | 59.00 | 79.00 | 52.00 | 53.00 | **243.00** |
| 824 | 39.00 | 68.00 | 53.00 | 29.00 | **189.00** |
| 825 | 0 | 0 | 10.00 | 5.00 | **15.00** |
| 826 | 139.00 | 187.00 | 184.00 | 159.00 | **669.00** |
| 827 | 152.50 | 141.00 | 141.50 | 171.50 | **606.50** |
| 828 | 53.00 | 73.00 | 79.00 | 68.00 | **273.00** |
| 829 | 34.00 | 43.00 | 31.00 | 40.00 | **148.00** |
| 830 | 0 | 0 | 7.50 | 0 | **7.50** |
| 831 | 0 | 0 | 12.00 | 12.00 | **24.00** |
| **WY - WYOMING - Total** | **1,020.00** | **940.00** | **980.00** | **906.50** | **3,846.50** |

**DRUG: 9801 - FENTANYL BASE**

**STATE: AK - ALASKA**

| REGISTRANT ZIP CODE 3 | 1ST QUARTER | 2ND QUARTER | 3RD QUARTER | 4TH QUARTER | TOTAL GRAMS |
|---|---|---|---|---|---|
| 995 | 35.40 | 30.41 | 28.25 | 30.80 | **124.86** |
| 996 | 39.60 | 34.58 | 34.60 | 32.89 | **141.68** |
| 997 | 11.67 | 10.26 | 10.06 | 8.28 | **40.27** |
| 998 | 7.56 | 6.31 | 6.56 | 5.09 | **25.53** |
| 999 | 4.49 | 3.06 | 2.37 | 2.70 | **12.63** |
| **AK - ALASKA - Total** | **98.72** | **84.63** | **81.84** | **79.76** | **344.96** |

**STATE: AL - ALABAMA**

| REGISTRANT ZIP CODE 3 | 1ST QUARTER | 2ND QUARTER | 3RD QUARTER | 4TH QUARTER | TOTAL GRAMS |
|---|---|---|---|---|---|
| 350 | 78.33 | 82.44 | 72.85 | 73.59 | **307.20** |
| 351 | 49.24 | 47.66 | 43.16 | 43.53 | **183.59** |
| 352 | 88.39 | 91.79 | 91.21 | 93.53 | **364.92** |
| 354 | 56.62 | 51.43 | 50.92 | 50.55 | **209.53** |
| 355 | 26.92 | 27.12 | 23.93 | 25.31 | **103.28** |
| 356 | 49.89 | 45.07 | 48.16 | 49.16 | **192.28** |

| | | | | | |
|---|---|---|---|---|---|
| 357 | 13.75 | 14.22 | 15.92 | 17.43 | **61.31** |
| 358 | 34.91 | 36.02 | 33.55 | 36.04 | **140.51** |
| 359 | 58.31 | 55.67 | 56.47 | 57.34 | **227.80** |
| 360 | 35.92 | 34.57 | 35.53 | 35.85 | **141.87** |
| 361 | 61.27 | 66.89 | 58.39 | 60.96 | **247.51** |
| 362 | 22.65 | 19.66 | 20.06 | 20.24 | **82.60** |
| 363 | 52.66 | 51.94 | 54.00 | 52.71 | **211.32** |
| 364 | 17.09 | 14.45 | 17.19 | 17.56 | **66.28** |
| 365 | 53.41 | 44.77 | 43.32 | 43.09 | **184.59** |
| 366 | 38.14 | 41.28 | 35.38 | 41.84 | **156.64** |
| 367 | 13.43 | 15.33 | 14.64 | 15.19 | **58.58** |
| 368 | 38.02 | 34.40 | 36.65 | 37.51 | **146.58** |
| 369 | 2.00 | 2.12 | 1.83 | 2.27 | **8.23** |
| **AL - ALABAMA - Total** | **790.96** | **776.82** | **753.16** | **773.68** | **3,094.63** |

**STATE: AR - ARKANSAS**

| REGISTRANT ZIP CODE 3 | 1ST QUARTER | 2ND QUARTER | 3RD QUARTER | 4TH QUARTER | TOTAL GRAMS |
|---|---|---|---|---|---|
| 716 | 19.72 | 18.69 | 16.98 | 15.91 | **71.31** |
| 717 | 17.95 | 17.85 | 20.38 | 19.04 | **75.21** |
| 718 | 10.30 | 10.54 | 10.52 | 12.40 | **43.76** |
| 719 | 38.82 | 38.09 | 36.14 | 32.60 | **145.64** |
| 720 | 35.09 | 35.84 | 37.69 | 37.28 | **145.90** |
| 721 | 58.68 | 54.78 | 56.52 | 58.79 | **228.77** |
| 722 | 52.54 | 45.39 | 49.13 | 54.16 | **201.22** |
| 723 | 13.94 | 12.85 | 12.72 | 12.77 | **52.27** |
| 724 | 36.56 | 34.54 | 33.89 | 33.73 | **138.73** |
| 725 | 19.11 | 17.06 | 19.60 | 17.66 | **73.42** |
| 726 | 22.49 | 23.96 | 21.32 | 20.79 | **88.56** |
| 727 | 65.38 | 56.50 | 61.52 | 58.20 | **241.60** |
| 728 | 15.46 | 13.89 | 14.08 | 15.43 | **58.85** |
| 729 | 30.52 | 30.41 | 32.53 | 30.15 | **123.61** |
| **AR - ARKANSAS - Total** | **436.55** | **410.38** | **423.02** | **418.90** | **1,688.86** |

**STATE: AS - AMERICAN SAMOA**

| REGISTRANT ZIP CODE 3 | 1ST QUARTER | 2ND QUARTER | 3RD QUARTER | 4TH QUARTER | TOTAL GRAMS |
|---|---|---|---|---|---|
| 967 | 0.11 | 0.08 | 0.03 | 0.03 | **0.24** |
| **AS - AMERICAN SAMOA - Total** | **0.11** | **0.08** | **0.03** | **0.03** | **0.24** |

**STATE: AZ - ARIZONA**

| REGISTRANT ZIP CODE 3 | 1ST QUARTER | 2ND QUARTER | 3RD QUARTER | 4TH QUARTER | TOTAL GRAMS |
|---|---|---|---|---|---|
| 850 | 137.28 | 145.08 | 116.76 | 127.23 | **526.35** |
| 851 | 28.04 | 23.61 | 21.71 | 22.05 | **95.41** |
| 852 | 260.50 | 252.52 | 222.87 | 250.92 | **986.82** |
| 853 | 130.12 | 120.38 | 113.80 | 119.94 | **484.24** |
| 855 | 10.09 | 8.44 | 10.11 | 8.98 | **37.61** |
| 856 | 30.08 | 29.05 | 27.40 | 26.08 | **112.61** |
| 857 | 137.13 | 136.08 | 120.73 | 129.98 | **523.91** |
| 859 | 8.29 | 7.93 | 7.94 | 7.93 | **32.09** |
| 860 | 12.41 | 11.07 | 11.54 | 9.68 | **44.71** |
| 863 | 33.34 | 31.98 | 28.53 | 32.53 | **126.38** |
| 864 | 35.85 | 33.10 | 29.87 | 33.44 | **132.27** |
| 865 | 1.32 | 1.48 | 0.46 | 0.50 | **3.77** |
| **AZ - ARIZONA - Total** | **824.45** | **800.72** | **711.74** | **769.25** | **3,106.16** |

**STATE: CA - CALIFORNIA**

| REGISTRANT ZIP CODE 3 | 1ST QUARTER | 2ND QUARTER | 3RD QUARTER | 4TH QUARTER | TOTAL GRAMS |
|---|---|---|---|---|---|
| 900 | 113.03 | 158.74 | 108.05 | 135.77 | **515.59** |
| 902 | 55.49 | 63.07 | 58.57 | 63.28 | **240.41** |
| 903 | 2.93 | 5.51 | 5.57 | 5.19 | **19.20** |
| 904 | 61.53 | 57.94 | 47.81 | 100.67 | **267.95** |
| 905 | 33.22 | 31.79 | 25.79 | 30.14 | **120.94** |
| 906 | 56.31 | 49.00 | 47.17 | 46.93 | **199.41** |
| 907 | 37.60 | 31.25 | 28.37 | 35.54 | **132.76** |
| 908 | 141.24 | 217.33 | 207.92 | 112.18 | **678.66** |

| | | | | | |
|---|---|---|---|---|---|
| 910 | 37.07 | 31.21 | 34.91 | 36.71 | **139.90** |
| 911 | 16.26 | 16.11 | 15.72 | 15.34 | **63.42** |
| 912 | 30.18 | 34.79 | 34.04 | 42.99 | **142.01** |
| 913 | 110.56 | 113.22 | 105.79 | 110.11 | **439.67** |
| 914 | 34.47 | 28.37 | 33.49 | 33.60 | **129.93** |
| 915 | 15.32 | 12.54 | 8.42 | 13.33 | **49.62** |
| 916 | 3.64 | 4.03 | 3.49 | 2.37 | **13.53** |
| 917 | 169.34 | 175.79 | 144.44 | 190.22 | **679.79** |
| 918 | 3.05 | 2.87 | 3.02 | 2.75 | **11.69** |
| 919 | 18.97 | 25.27 | 25.99 | 20.94 | **91.17** |
| 920 | 160.88 | 158.62 | 144.43 | 150.49 | **614.41** |
| 921 | 186.88 | 101.93 | 116.25 | 174.17 | **579.23** |
| 922 | 97.42 | 101.69 | 95.64 | 102.73 | **397.48** |
| 923 | 137.64 | 137.87 | 147.30 | 152.08 | **574.89** |
| 924 | 16.24 | 12.93 | 13.15 | 18.11 | **60.42** |
| 925 | 169.21 | 123.29 | 126.20 | 133.09 | **551.80** |
| 926 | 156.19 | 151.72 | 149.01 | 147.65 | **604.58** |
| 927 | 27.20 | 25.71 | 31.66 | 27.59 | **112.16** |
| 928 | 126.60 | 122.60 | 129.23 | 136.38 | **514.81** |
| 930 | 60.72 | 60.44 | 53.92 | 56.89 | **231.96** |
| 931 | 14.52 | 14.40 | 11.14 | 12.33 | **52.39** |
| 932 | 66.82 | 62.48 | 65.46 | 68.42 | **263.19** |
| 933 | 75.25 | 76.07 | 77.42 | 69.67 | **298.41** |
| 934 | 71.17 | 59.12 | 58.64 | 61.08 | **250.01** |
| 935 | 45.28 | 47.17 | 42.63 | 49.81 | **184.89** |
| 936 | 31.45 | 31.07 | 37.73 | 38.73 | **138.98** |
| 937 | 64.18 | 66.98 | 64.39 | 63.43 | **258.98** |
| 939 | 14.24 | 13.41 | 14.52 | 14.68 | **56.86** |
| 940 | 74.17 | 59.21 | 62.62 | 63.49 | **259.49** |
| 941 | 69.45 | 69.80 | 69.70 | 67.05 | **276.00** |
| 943 | 23.08 | 23.98 | 22.00 | 28.19 | **97.25** |
| 944 | 9.21 | 11.45 | 8.29 | 11.11 | **40.06** |
| 945 | 226.67 | 222.59 | 214.81 | 218.54 | **882.60** |
| 946 | 28.39 | 29.80 | 29.42 | 27.28 | **114.89** |
| 947 | 11.95 | 11.73 | 10.96 | 10.03 | **44.67** |
| 948 | 5.41 | 4.28 | 4.33 | 3.51 | **17.53** |
| 949 | 27.73 | 29.02 | 30.88 | 31.60 | **119.23** |
| 950 | 54.24 | 49.87 | 51.07 | 51.80 | **206.98** |
| 951 | 44.94 | 44.83 | 40.91 | 37.61 | **168.30** |
| 952 | 53.56 | 48.46 | 60.46 | 53.83 | **216.31** |
| 953 | 126.24 | 114.69 | 121.81 | 116.90 | **479.65** |
| 954 | 57.88 | 46.93 | 46.67 | 47.65 | **199.13** |
| 955 | 26.20 | 22.63 | 22.80 | 21.24 | **92.86** |
| 956 | 160.18 | 146.88 | 140.20 | 140.67 | **587.93** |
| 957 | 19.58 | 18.91 | 18.32 | 17.73 | **74.54** |
| 958 | 84.44 | 80.35 | 80.34 | 88.23 | **333.36** |
| 959 | 61.82 | 54.23 | 51.93 | 48.72 | **216.70** |
| 960 | 39.63 | 36.72 | 36.88 | 36.91 | **150.14** |
| 961 | 7.64 | 6.50 | 5.92 | 5.08 | **25.15** |
| **CA - CALIFORNIA - Total** | **3,674.48** | **3,559.16** | **3,447.63** | **3,602.60** | **14,283.87** |

**STATE: CO - COLORADO**

| REGISTRANT ZIP CODE 3 | 1ST QUARTER | 2ND QUARTER | 3RD QUARTER | 4TH QUARTER | TOTAL GRAMS |
|---|---|---|---|---|---|
| 800 | 120.48 | 124.56 | 113.54 | 125.14 | **483.71** |
| 801 | 101.18 | 108.78 | 98.17 | 101.08 | **409.21** |
| 802 | 103.02 | 108.05 | 89.91 | 93.53 | **394.52** |
| 803 | 12.82 | 11.61 | 13.14 | 13.97 | **51.55** |
| 804 | 22.63 | 24.93 | 23.39 | 19.49 | **90.43** |
| 805 | 70.49 | 69.64 | 68.36 | 71.52 | **280.01** |
| 806 | 24.20 | 22.17 | 23.29 | 26.04 | **95.70** |
| 807 | 10.75 | 11.16 | 11.46 | 12.36 | **45.73** |

| | | | | | |
|---|---|---|---|---|---|
| 808 | 16.36 | 15.70 | 15.79 | 14.27 | **62.12** |
| 809 | 68.31 | 72.40 | 66.63 | 71.12 | **278.46** |
| 810 | 39.58 | 37.08 | 37.06 | 41.06 | **154.78** |
| 811 | 9.16 | 9.79 | 8.63 | 8.99 | **36.57** |
| 812 | 12.32 | 12.51 | 14.07 | 13.09 | **51.99** |
| 813 | 7.97 | 7.13 | 7.00 | 6.05 | **28.15** |
| 814 | 7.69 | 7.20 | 8.21 | 7.58 | **30.68** |
| 815 | 24.16 | 21.59 | 29.27 | 21.41 | **96.43** |
| 816 | 13.14 | 12.94 | 11.64 | 11.68 | **49.40** |
| **CO - COLORADO - Total** | **664.26** | **677.24** | **639.56** | **658.40** | **2,639.45** |

**STATE: CT - CONNECTICUT**

| REGISTRANT ZIP CODE 3 | 1ST QUARTER | 2ND QUARTER | 3RD QUARTER | 4TH QUARTER | TOTAL GRAMS |
|---|---|---|---|---|---|
| 060 | 81.45 | 90.23 | 86.89 | 86.25 | **344.82** |
| 061 | 27.60 | 30.73 | 26.10 | 22.29 | **106.72** |
| 062 | 24.94 | 23.52 | 20.06 | 23.72 | **92.23** |
| 063 | 38.94 | 39.51 | 37.30 | 39.40 | **155.14** |
| 064 | 134.72 | 125.91 | 126.45 | 147.84 | **534.93** |
| 065 | 60.08 | 50.90 | 57.50 | 64.67 | **233.16** |
| 066 | 22.09 | 26.61 | 22.06 | 30.30 | **101.07** |
| 067 | 55.24 | 56.22 | 50.72 | 50.38 | **212.55** |
| 068 | 54.66 | 53.85 | 46.21 | 47.94 | **202.66** |
| 069 | 7.59 | 7.29 | 5.92 | 7.51 | **28.32** |
| **CT - CONNECTICUT - Total** | **507.31** | **504.77** | **479.22** | **520.31** | **2,011.60** |

**STATE: DC - DISTRICT OF COLUMBIA**

| REGISTRANT ZIP CODE 3 | 1ST QUARTER | 2ND QUARTER | 3RD QUARTER | 4TH QUARTER | TOTAL GRAMS |
|---|---|---|---|---|---|
| 200 | 51.51 | 63.51 | 45.66 | 40.01 | **200.70** |
| 203 | 0.02 | 0 | 0 | 0 | **0.02** |
| 204 | 1.30 | 3.69 | 3.13 | 1.27 | **9.39** |
| 205 | 0 | 0.01 | 0 | 0 | **0.01** |
| **DC - DISTRICT OF COLUMBIA - Total** | **52.84** | **67.20** | **48.79** | **41.28** | **210.11** |

**STATE: DE - DELAWARE**

| REGISTRANT ZIP CODE 3 | 1ST QUARTER | 2ND QUARTER | 3RD QUARTER | 4TH QUARTER | TOTAL GRAMS |
|---|---|---|---|---|---|
| 197 | 66.69 | 62.15 | 64.13 | 52.98 | **245.95** |
| 198 | 30.43 | 23.93 | 22.15 | 23.32 | **99.84** |
| 199 | 51.86 | 50.58 | 45.61 | 47.27 | **195.32** |
| **DE - DELAWARE - Total** | **148.98** | **136.66** | **131.90** | **123.57** | **541.11** |

**STATE: FL - FLORIDA**

| REGISTRANT ZIP CODE 3 | 1ST QUARTER | 2ND QUARTER | 3RD QUARTER | 4TH QUARTER | TOTAL GRAMS |
|---|---|---|---|---|---|
| 320 | 104.04 | 95.69 | 100.00 | 94.45 | **394.18** |
| 321 | 124.32 | 114.78 | 114.63 | 107.14 | **460.87** |
| 322 | 182.12 | 156.07 | 173.10 | 158.12 | **669.40** |
| 323 | 64.22 | 54.88 | 56.13 | 53.24 | **228.46** |
| 324 | 73.01 | 65.56 | 64.79 | 63.82 | **267.19** |
| 325 | 79.39 | 72.22 | 78.93 | 74.33 | **304.87** |
| 326 | 80.23 | 68.56 | 76.65 | 63.57 | **289.01** |
| 327 | 178.69 | 161.46 | 175.41 | 159.97 | **675.53** |
| 328 | 109.01 | 100.53 | 122.85 | 101.83 | **434.21** |
| 329 | 177.27 | 178.04 | 171.80 | 172.98 | **700.09** |
| 330 | 151.83 | 144.48 | 169.00 | 146.51 | **611.81** |
| 331 | 128.01 | 157.83 | 167.67 | 136.98 | **590.50** |
| 333 | 143.65 | 136.21 | 140.25 | 129.98 | **550.09** |
| 334 | 313.98 | 306.59 | 296.71 | 276.93 | **1,194.20** |
| 335 | 107.90 | 99.43 | 104.63 | 99.89 | **411.85** |
| 336 | 413.95 | 652.22 | 142.55 | 129.56 | **1,338.28** |
| 337 | 150.78 | 135.74 | 133.66 | 134.89 | **555.07** |
| 338 | 94.55 | 87.11 | 81.84 | 86.21 | **349.70** |
| 339 | 165.27 | 130.67 | 131.41 | 129.04 | **556.40** |
| 341 | 64.49 | 56.81 | 52.30 | 56.22 | **229.82** |
| 342 | 148.42 | 144.96 | 141.04 | 129.20 | **563.61** |
| 344 | 73.15 | 65.48 | 67.00 | 62.93 | **268.55** |

| | | | | | |
|---|---|---|---|---|---|
| 346 | 154.73 | 147.74 | 147.67 | 145.39 | **595.52** |
| 347 | 78.19 | 75.97 | 82.02 | 70.40 | **306.58** |
| 349 | 112.60 | 104.52 | 105.51 | 107.05 | **429.68** |
| **FL - FLORIDA - Total** | **3,473.79** | **3,513.55** | **3,097.53** | **2,890.62** | **12,975.48** |

**STATE: GA - GEORGIA**

| REGISTRANT ZIP CODE 3 | 1ST QUARTER | 2ND QUARTER | 3RD QUARTER | 4TH QUARTER | TOTAL GRAMS |
|---|---|---|---|---|---|
| 300 | 295.42 | 283.54 | 290.78 | 258.23 | **1,127.97** |
| 301 | 191.39 | 245.97 | 190.26 | 182.93 | **810.56** |
| 302 | 135.62 | 126.68 | 137.68 | 131.73 | **531.70** |
| 303 | 131.05 | 188.23 | 183.56 | 140.33 | **643.18** |
| 304 | 34.02 | 35.07 | 32.77 | 34.26 | **136.12** |
| 305 | 132.12 | 129.55 | 127.78 | 127.06 | **516.51** |
| 306 | 64.64 | 59.30 | 61.40 | 61.37 | **246.71** |
| 307 | 31.80 | 34.29 | 38.80 | 36.40 | **141.29** |
| 308 | 35.75 | 33.09 | 29.91 | 28.99 | **127.74** |
| 309 | 61.50 | 58.67 | 60.73 | 53.99 | **234.89** |
| 310 | 99.65 | 100.17 | 101.13 | 95.69 | **396.64** |
| 312 | 38.97 | 37.90 | 42.08 | 37.55 | **156.50** |
| 313 | 24.45 | 20.12 | 20.55 | 18.40 | **83.52** |
| 314 | 42.60 | 52.77 | 36.90 | 47.77 | **180.03** |
| 315 | 69.25 | 64.62 | 72.71 | 65.46 | **272.04** |
| 316 | 42.11 | 36.09 | 38.68 | 40.97 | **157.85** |
| 317 | 92.86 | 92.47 | 81.44 | 76.40 | **343.16** |
| 318 | 3.18 | 2.04 | 1.92 | 2.47 | **9.61** |
| 319 | 26.54 | 32.57 | 27.64 | 27.81 | **114.56** |
| 398 | 24.77 | 23.14 | 20.56 | 21.80 | **90.28** |
| **GA - GEORGIA - Total** | **1,577.69** | **1,656.28** | **1,597.29** | **1,489.63** | **6,320.88** |

**STATE: GU - GUAM**

| REGISTRANT ZIP CODE 3 | 1ST QUARTER | 2ND QUARTER | 3RD QUARTER | 4TH QUARTER | TOTAL GRAMS |
|---|---|---|---|---|---|
| 969 | 9.05 | 8.65 | 8.15 | 9.80 | **35.65** |
| **GU - GUAM - Total** | **9.05** | **8.65** | **8.15** | **9.80** | **35.65** |

**STATE: HI - HAWAII**

| REGISTRANT ZIP CODE 3 | 1ST QUARTER | 2ND QUARTER | 3RD QUARTER | 4TH QUARTER | TOTAL GRAMS |
|---|---|---|---|---|---|
| 967 | 64.81 | 59.39 | 63.06 | 60.03 | **247.29** |
| 968 | 49.10 | 42.03 | 44.52 | 41.04 | **176.70** |
| **HI - HAWAII - Total** | **113.91** | **101.42** | **107.59** | **101.07** | **423.99** |

**STATE: IA - IOWA**

| REGISTRANT ZIP CODE 3 | 1ST QUARTER | 2ND QUARTER | 3RD QUARTER | 4TH QUARTER | TOTAL GRAMS |
|---|---|---|---|---|---|
| 500 | 48.15 | 43.18 | 45.29 | 43.37 | **179.99** |
| 501 | 43.25 | 40.74 | 41.72 | 37.28 | **162.99** |
| 502 | 45.09 | 39.31 | 37.79 | 37.94 | **160.14** |
| 503 | 58.54 | 60.17 | 56.52 | 56.35 | **231.58** |
| 504 | 24.36 | 25.36 | 22.69 | 29.54 | **101.95** |
| 505 | 29.68 | 29.58 | 27.36 | 27.86 | **114.47** |
| 506 | 30.61 | 25.02 | 26.95 | 23.03 | **105.61** |
| 507 | 13.69 | 12.93 | 10.64 | 12.55 | **49.81** |
| 508 | 5.93 | 5.34 | 6.64 | 5.70 | **23.61** |
| 510 | 7.49 | 5.68 | 5.80 | 6.18 | **25.15** |
| 511 | 16.24 | 16.79 | 11.78 | 11.60 | **56.41** |
| 512 | 6.29 | 5.68 | 5.81 | 5.14 | **22.91** |
| 513 | 8.24 | 8.38 | 7.56 | 7.09 | **31.27** |
| 514 | 18.38 | 17.70 | 16.05 | 15.21 | **67.33** |
| 515 | 16.27 | 14.20 | 14.26 | 14.41 | **59.14** |
| 516 | 3.89 | 4.30 | 3.90 | 3.67 | **15.76** |
| 520 | 22.66 | 22.27 | 19.16 | 21.31 | **85.40** |
| 521 | 10.50 | 8.81 | 9.87 | 8.21 | **37.39** |
| 522 | 26.65 | 22.51 | 24.90 | 27.07 | **101.13** |
| 523 | 12.38 | 9.88 | 9.92 | 10.75 | **42.93** |
| 524 | 22.50 | 18.74 | 15.47 | 18.68 | **75.40** |
| 525 | 26.06 | 24.34 | 23.79 | 22.68 | **96.87** |

| | | | | | |
|---|---|---|---|---|---|
| 526 | 17.49 | 14.15 | 16.98 | 13.92 | **62.53** |
| 527 | 13.47 | 12.55 | 12.35 | 11.79 | **50.15** |
| 528 | 16.82 | 17.69 | 17.12 | 16.95 | **68.57** |
| **IA - IOWA - Total** | **544.61** | **505.31** | **490.30** | **488.28** | **2,028.51** |

**STATE: ID - IDAHO**

| REGISTRANT ZIP CODE 3 | 1ST QUARTER | 2ND QUARTER | 3RD QUARTER | 4TH QUARTER | TOTAL GRAMS |
|---|---|---|---|---|---|
| 832 | 42.63 | 35.66 | 41.30 | 39.55 | **159.15** |
| 833 | 33.15 | 29.72 | 26.21 | 27.97 | **117.05** |
| 834 | 45.30 | 43.57 | 45.65 | 42.02 | **176.55** |
| 835 | 24.72 | 21.68 | 22.86 | 22.98 | **92.24** |
| 836 | 85.78 | 86.09 | 84.30 | 84.73 | **340.89** |
| 837 | 71.64 | 67.92 | 66.32 | 69.34 | **275.22** |
| 838 | 48.37 | 40.51 | 44.97 | 42.73 | **176.58** |
| **ID - IDAHO - Total** | **351.60** | **325.15** | **331.60** | **329.33** | **1,337.68** |

**STATE: IL - ILLINOIS**

| REGISTRANT ZIP CODE 3 | 1ST QUARTER | 2ND QUARTER | 3RD QUARTER | 4TH QUARTER | TOTAL GRAMS |
|---|---|---|---|---|---|
| 600 | 268.60 | 279.19 | 245.30 | 244.03 | **1,037.11** |
| 601 | 151.92 | 167.58 | 141.86 | 144.67 | **606.03** |
| 602 | 8.85 | 9.81 | 7.30 | 7.68 | **33.63** |
| 603 | 7.48 | 9.15 | 6.54 | 11.06 | **34.23** |
| 604 | 191.11 | 194.35 | 172.78 | 181.13 | **739.38** |
| 605 | 125.64 | 118.16 | 115.48 | 119.60 | **478.88** |
| 606 | 175.90 | 234.03 | 144.65 | 167.11 | **721.69** |
| 607 | 17.13 | 14.44 | 14.61 | 13.73 | **59.91** |
| 608 | 7.25 | 10.52 | 5.48 | 8.31 | **31.57** |
| 609 | 24.82 | 19.64 | 21.12 | 21.17 | **86.75** |
| 610 | 64.01 | 62.82 | 61.77 | 62.52 | **251.12** |
| 611 | 67.26 | 64.44 | 67.99 | 66.56 | **266.25** |
| 612 | 16.70 | 15.40 | 18.50 | 17.62 | **68.23** |
| 613 | 16.36 | 16.09 | 19.35 | 16.51 | **68.32** |
| 614 | 22.53 | 21.88 | 21.56 | 23.66 | **89.63** |
| 615 | 30.75 | 30.45 | 30.42 | 27.00 | **118.63** |
| 616 | 81.35 | 74.17 | 74.67 | 70.36 | **300.55** |
| 617 | 47.05 | 44.23 | 46.51 | 42.89 | **180.67** |
| 618 | 45.21 | 39.49 | 42.37 | 43.16 | **170.23** |
| 619 | 17.97 | 17.69 | 15.72 | 14.82 | **66.20** |
| 620 | 49.75 | 49.26 | 47.42 | 47.84 | **194.27** |
| 622 | 49.63 | 49.94 | 44.86 | 44.77 | **189.21** |
| 623 | 30.06 | 26.00 | 26.42 | 31.16 | **113.64** |
| 624 | 23.45 | 22.64 | 21.06 | 18.41 | **85.56** |
| 625 | 34.75 | 31.60 | 31.29 | 31.40 | **129.05** |
| 626 | 27.81 | 27.69 | 26.30 | 24.27 | **106.07** |
| 627 | 41.57 | 37.43 | 36.00 | 37.60 | **152.60** |
| 628 | 28.30 | 28.86 | 31.60 | 27.75 | **116.51** |
| 629 | 46.91 | 42.44 | 42.76 | 43.17 | **175.29** |
| **IL - ILLINOIS - Total** | **1,720.12** | **1,759.41** | **1,581.70** | **1,609.96** | **6,671.19** |

**STATE: IN - INDIANA**

| REGISTRANT ZIP CODE 3 | 1ST QUARTER | 2ND QUARTER | 3RD QUARTER | 4TH QUARTER | TOTAL GRAMS |
|---|---|---|---|---|---|
| 460 | 82.18 | 76.99 | 72.69 | 72.95 | **304.81** |
| 461 | 84.97 | 79.67 | 75.43 | 81.01 | **321.08** |
| 462 | 191.61 | 166.49 | 172.02 | 174.68 | **704.80** |
| 463 | 109.97 | 102.85 | 104.39 | 94.89 | **412.10** |
| 464 | 9.28 | 7.14 | 8.11 | 8.74 | **33.27** |
| 465 | 74.61 | 72.98 | 70.57 | 72.03 | **290.19** |
| 466 | 17.89 | 17.40 | 17.55 | 17.30 | **70.13** |
| 467 | 41.82 | 39.58 | 42.77 | 42.48 | **166.65** |
| 468 | 59.79 | 65.91 | 56.24 | 65.14 | **247.07** |
| 469 | 47.62 | 47.29 | 44.56 | 47.26 | **186.72** |
| 470 | 23.58 | 27.13 | 22.25 | 22.15 | **95.12** |
| 471 | 51.88 | 46.36 | 42.88 | 45.62 | **186.75** |

| | | | | | |
|---|---|---|---|---|---|
| 472 | 28.47 | 26.47 | 27.19 | 27.23 | **109.37** |
| 473 | 58.66 | 57.62 | 60.27 | 56.60 | **233.15** |
| 474 | 41.18 | 37.18 | 36.48 | 37.60 | **152.45** |
| 475 | 27.99 | 26.82 | 26.30 | 24.03 | **105.14** |
| 476 | 12.56 | 9.68 | 11.68 | 11.74 | **45.66** |
| 477 | 51.04 | 30.48 | 65.23 | 33.81 | **180.56** |
| 478 | 19.17 | 17.62 | 18.79 | 17.87 | **73.44** |
| 479 | 25.35 | 27.13 | 23.68 | 26.22 | **102.38** |
| **IN - INDIANA - Total** | **1,059.64** | **982.77** | **999.07** | **979.33** | **4,020.81** |

**STATE: KS - KANSAS**

| REGISTRANT ZIP CODE 3 | 1ST QUARTER | 2ND QUARTER | 3RD QUARTER | 4TH QUARTER | TOTAL GRAMS |
|---|---|---|---|---|---|
| 660 | 87.27 | 83.13 | 81.92 | 82.31 | **334.63** |
| 661 | 28.00 | 22.83 | 22.30 | 24.20 | **97.33** |
| 662 | 89.04 | 84.88 | 84.61 | 80.77 | **339.30** |
| 664 | 16.60 | 15.93 | 15.63 | 13.45 | **61.61** |
| 665 | 27.41 | 24.23 | 25.02 | 26.67 | **103.33** |
| 666 | 41.54 | 39.36 | 38.70 | 40.64 | **160.25** |
| 667 | 36.36 | 37.12 | 35.35 | 37.08 | **145.90** |
| 668 | 4.61 | 4.39 | 4.15 | 4.30 | **17.45** |
| 669 | 11.66 | 11.27 | 10.91 | 9.90 | **43.74** |
| 670 | 31.52 | 31.96 | 28.75 | 30.91 | **123.14** |
| 671 | 17.30 | 16.77 | 14.90 | 15.54 | **64.51** |
| 672 | 67.57 | 58.76 | 64.70 | 72.87 | **263.90** |
| 673 | 10.96 | 11.14 | 10.91 | 10.28 | **43.30** |
| 674 | 45.31 | 43.91 | 45.20 | 44.10 | **178.52** |
| 675 | 24.31 | 26.23 | 22.30 | 24.67 | **97.50** |
| 676 | 21.57 | 18.49 | 18.12 | 15.25 | **73.43** |
| 677 | 10.80 | 11.23 | 11.88 | 10.47 | **44.37** |
| 678 | 31.45 | 28.57 | 27.98 | 28.88 | **116.88** |
| 679 | 10.09 | 10.30 | 10.60 | 9.46 | **40.45** |
| **KS - KANSAS - Total** | **613.33** | **580.52** | **573.94** | **581.74** | **2,349.54** |

**STATE: KY - KENTUCKY**

| REGISTRANT ZIP CODE 3 | 1ST QUARTER | 2ND QUARTER | 3RD QUARTER | 4TH QUARTER | TOTAL GRAMS |
|---|---|---|---|---|---|
| 400 | 25.47 | 25.03 | 25.34 | 23.18 | **99.03** |
| 401 | 15.54 | 15.82 | 16.02 | 13.33 | **60.71** |
| 402 | 128.55 | 110.69 | 102.47 | 115.94 | **457.64** |
| 403 | 40.91 | 36.50 | 35.79 | 35.68 | **148.88** |
| 404 | 21.03 | 16.52 | 21.51 | 20.12 | **79.19** |
| 405 | 52.67 | 44.55 | 44.72 | 46.69 | **188.63** |
| 406 | 4.72 | 4.29 | 4.59 | 4.85 | **18.45** |
| 407 | 23.57 | 20.97 | 17.33 | 18.54 | **80.41** |
| 408 | 3.24 | 2.97 | 2.88 | 2.79 | **11.88** |
| 409 | 10.29 | 8.13 | 8.48 | 9.07 | **35.97** |
| 410 | 71.66 | 83.85 | 62.01 | 65.52 | **283.05** |
| 411 | 13.74 | 13.49 | 11.38 | 17.02 | **55.62** |
| 412 | 7.50 | 6.98 | 8.08 | 7.19 | **29.75** |
| 413 | 6.29 | 6.00 | 4.87 | 4.85 | **22.00** |
| 414 | 1.52 | 1.79 | 2.47 | 2.61 | **8.39** |
| 415 | 10.26 | 8.53 | 12.66 | 12.91 | **44.36** |
| 416 | 7.19 | 6.83 | 7.42 | 5.92 | **27.35** |
| 417 | 26.36 | 23.68 | 24.52 | 26.11 | **100.67** |
| 418 | 2.19 | 2.14 | 1.72 | 2.05 | **8.11** |
| 420 | 36.75 | 61.66 | 32.55 | 32.80 | **163.77** |
| 421 | 36.48 | 34.36 | 35.79 | 34.72 | **141.34** |
| 422 | 12.63 | 12.30 | 12.48 | 11.04 | **48.45** |
| 423 | 16.44 | 18.43 | 19.60 | 17.58 | **72.05** |
| 424 | 19.38 | 15.59 | 16.71 | 15.30 | **66.98** |
| 425 | 12.81 | 11.27 | 15.31 | 14.62 | **54.01** |
| 426 | 9.30 | 8.35 | 8.00 | 7.18 | **32.84** |
| 427 | 30.88 | 27.61 | 25.79 | 27.56 | **111.83** |

| KY - KENTUCKY - Total | 647.36 | 628.32 | 580.50 | 595.18 | 2,451.37 |
|---|---|---|---|---|---|

**STATE: LA - LOUISIANA**

| REGISTRANT ZIP CODE 3 | 1ST QUARTER | 2ND QUARTER | 3RD QUARTER | 4TH QUARTER | TOTAL GRAMS |
|---|---|---|---|---|---|
| 700 | 64.21 | 55.71 | 58.88 | 56.54 | **235.33** |
| 701 | 50.23 | 38.00 | 43.66 | 45.14 | **177.04** |
| 703 | 34.23 | 31.99 | 31.94 | 30.06 | **128.21** |
| 704 | 72.38 | 67.03 | 62.53 | 69.18 | **271.12** |
| 705 | 97.24 | 90.74 | 93.57 | 100.82 | **382.37** |
| 706 | 26.46 | 26.81 | 22.52 | 26.94 | **102.74** |
| 707 | 55.11 | 58.39 | 52.36 | 46.70 | **212.56** |
| 708 | 71.54 | 55.88 | 55.24 | 55.72 | **238.38** |
| 710 | 17.18 | 17.29 | 15.42 | 16.15 | **66.04** |
| 711 | 49.33 | 44.99 | 44.41 | 45.45 | **184.18** |
| 712 | 74.60 | 73.96 | 68.77 | 68.18 | **285.51** |
| 713 | 39.88 | 37.43 | 37.76 | 34.53 | **149.59** |
| 714 | 21.36 | 19.74 | 22.24 | 19.10 | **82.44** |
| **LA - LOUISIANA - Total** | **673.75** | **617.95** | **609.29** | **614.54** | **2,515.52** |

**STATE: MA - MASSACHUSETTS**

| REGISTRANT ZIP CODE 3 | 1ST QUARTER | 2ND QUARTER | 3RD QUARTER | 4TH QUARTER | TOTAL GRAMS |
|---|---|---|---|---|---|
| 010 | 81.76 | 74.80 | 74.38 | 70.72 | **301.66** |
| 011 | 26.91 | 27.02 | 28.53 | 30.07 | **112.53** |
| 012 | 18.83 | 21.19 | 19.44 | 18.12 | **77.59** |
| 013 | 11.33 | 9.51 | 9.82 | 10.33 | **40.99** |
| 014 | 21.89 | 23.35 | 21.87 | 21.55 | **88.66** |
| 015 | 37.71 | 33.34 | 33.05 | 33.40 | **137.51** |
| 016 | 31.20 | 39.31 | 23.33 | 27.90 | **121.74** |
| 017 | 48.94 | 50.67 | 47.05 | 45.78 | **192.45** |
| 018 | 113.94 | 110.33 | 87.69 | 97.37 | **409.33** |
| 019 | 46.48 | 41.97 | 38.04 | 40.45 | **166.94** |
| 020 | 33.04 | 33.51 | 28.79 | 27.97 | **123.30** |
| 021 | 110.58 | 158.72 | 91.70 | 109.61 | **470.61** |
| 022 | 16.45 | 32.91 | 13.29 | 13.91 | **76.56** |
| 023 | 49.66 | 57.26 | 45.97 | 44.07 | **196.96** |
| 024 | 22.28 | 23.50 | 23.66 | 24.14 | **93.58** |
| 025 | 14.54 | 15.37 | 14.25 | 12.51 | **56.68** |
| 026 | 16.61 | 18.25 | 16.74 | 16.48 | **68.08** |
| 027 | 112.03 | 111.23 | 103.75 | 96.37 | **423.38** |
| **MA - MASSACHUSETTS - Total** | **814.19** | **882.23** | **721.36** | **740.77** | **3,158.55** |

**STATE: MD - MARYLAND**

| REGISTRANT ZIP CODE 3 | 1ST QUARTER | 2ND QUARTER | 3RD QUARTER | 4TH QUARTER | TOTAL GRAMS |
|---|---|---|---|---|---|
| 206 | 36.26 | 35.01 | 33.46 | 31.62 | **136.35** |
| 207 | 57.73 | 62.72 | 53.50 | 51.15 | **225.09** |
| 208 | 82.25 | 81.91 | 76.81 | 77.98 | **318.95** |
| 209 | 22.34 | 36.38 | 16.08 | 20.44 | **95.24** |
| 210 | 107.25 | 110.00 | 100.43 | 115.54 | **433.23** |
| 211 | 48.45 | 48.53 | 45.52 | 46.23 | **188.73** |
| 212 | 165.35 | 186.09 | 133.08 | 148.14 | **632.65** |
| 214 | 15.88 | 13.69 | 10.37 | 10.11 | **50.05** |
| 215 | 20.32 | 20.31 | 18.33 | 18.70 | **77.66** |
| 216 | 22.02 | 20.96 | 18.87 | 19.41 | **81.24** |
| 217 | 89.10 | 84.30 | 85.99 | 78.10 | **337.48** |
| 218 | 25.86 | 24.06 | 25.94 | 23.88 | **99.74** |
| 219 | 10.69 | 11.63 | 8.21 | 9.28 | **39.81** |
| **MD - MARYLAND - Total** | **703.50** | **735.58** | **626.58** | **650.57** | **2,716.22** |

**STATE: ME - MAINE**

| REGISTRANT ZIP CODE 3 | 1ST QUARTER | 2ND QUARTER | 3RD QUARTER | 4TH QUARTER | TOTAL GRAMS |
|---|---|---|---|---|---|
| 039 | 2.59 | 2.50 | 3.22 | 3.77 | **12.08** |
| 040 | 59.94 | 54.29 | 53.42 | 51.79 | **219.45** |
| 041 | 24.18 | 23.80 | 24.58 | 22.08 | **94.64** |
| 042 | 28.46 | 24.35 | 22.06 | 28.07 | **102.94** |

| | | | | | |
|---|---|---|---|---|---|
| 043 | 26.29 | 25.99 | 24.53 | 23.53 | **100.34** |
| 044 | 31.06 | 31.36 | 24.24 | 23.39 | **110.05** |
| 045 | 5.22 | 4.68 | 5.67 | 3.88 | **19.44** |
| 046 | 9.98 | 10.54 | 8.71 | 8.17 | **37.41** |
| 047 | 12.91 | 11.69 | 10.63 | 9.59 | **44.82** |
| 048 | 4.85 | 3.52 | 4.81 | 5.02 | **18.20** |
| 049 | 24.71 | 26.05 | 24.75 | 24.01 | **99.51** |
| **ME - MAINE - Total** | **230.19** | **218.78** | **206.62** | **203.29** | **858.88** |

**STATE: MI - MICHIGAN**

| REGISTRANT ZIP CODE 3 | 1ST QUARTER | 2ND QUARTER | 3RD QUARTER | 4TH QUARTER | TOTAL GRAMS |
|---|---|---|---|---|---|
| 480 | 184.26 | 200.37 | 169.61 | 174.28 | **728.51** |
| 481 | 217.58 | 235.95 | 199.78 | 206.52 | **859.83** |
| 482 | 67.26 | 93.57 | 60.30 | 74.25 | **295.38** |
| 483 | 151.14 | 160.85 | 138.91 | 152.28 | **603.18** |
| 484 | 67.21 | 61.45 | 61.47 | 67.84 | **257.96** |
| 485 | 27.73 | 27.35 | 26.60 | 33.42 | **115.10** |
| 486 | 103.73 | 94.25 | 96.46 | 98.90 | **393.34** |
| 487 | 49.25 | 44.91 | 44.37 | 45.04 | **183.57** |
| 488 | 68.63 | 70.22 | 66.95 | 69.18 | **274.99** |
| 489 | 32.67 | 28.57 | 31.24 | 36.47 | **128.96** |
| 490 | 180.16 | 163.29 | 164.46 | 163.94 | **671.85** |
| 491 | 18.92 | 19.08 | 19.24 | 18.21 | **75.45** |
| 492 | 46.16 | 46.37 | 42.55 | 43.73 | **178.80** |
| 493 | 22.73 | 23.03 | 24.13 | 21.72 | **91.61** |
| 494 | 87.97 | 87.37 | 87.19 | 82.93 | **345.47** |
| 495 | 90.78 | 106.90 | 80.04 | 101.70 | **379.41** |
| 496 | 67.46 | 79.57 | 68.97 | 67.92 | **283.92** |
| 497 | 54.55 | 51.15 | 49.52 | 50.64 | **205.86** |
| 498 | 41.38 | 38.79 | 40.88 | 38.61 | **159.66** |
| 499 | 13.30 | 14.47 | 13.27 | 13.58 | **54.62** |
| **MI - MICHIGAN - Total** | **1,592.86** | **1,647.51** | **1,485.95** | **1,561.17** | **6,287.48** |

**STATE: MN - MINNESOTA**

| REGISTRANT ZIP CODE 3 | 1ST QUARTER | 2ND QUARTER | 3RD QUARTER | 4TH QUARTER | TOTAL GRAMS |
|---|---|---|---|---|---|
| 550 | 55.78 | 52.77 | 52.37 | 51.27 | **212.19** |
| 551 | 78.16 | 76.72 | 70.40 | 79.27 | **304.55** |
| 553 | 62.63 | 55.41 | 59.22 | 55.15 | **232.41** |
| 554 | 197.21 | 150.40 | 155.91 | 143.62 | **647.13** |
| 556 | 1.22 | 1.34 | 1.35 | 1.24 | **5.15** |
| 557 | 24.05 | 22.20 | 23.63 | 22.12 | **91.99** |
| 558 | 15.41 | 13.36 | 18.02 | 16.49 | **63.29** |
| 559 | 38.14 | 31.81 | 31.90 | 36.36 | **138.21** |
| 560 | 24.25 | 23.29 | 22.47 | 23.55 | **93.56** |
| 561 | 9.76 | 7.59 | 8.12 | 7.75 | **33.22** |
| 562 | 20.21 | 17.33 | 16.48 | 19.38 | **73.39** |
| 563 | 41.39 | 39.17 | 34.73 | 39.81 | **155.09** |
| 564 | 19.11 | 14.91 | 16.23 | 18.09 | **68.35** |
| 565 | 14.35 | 13.96 | 14.08 | 13.19 | **55.58** |
| 566 | 9.55 | 9.71 | 8.40 | 8.93 | **36.59** |
| 567 | 19.91 | 18.53 | 17.37 | 17.24 | **73.04** |
| **MN - MINNESOTA - Total** | **631.13** | **548.47** | **550.69** | **553.46** | **2,283.74** |

**STATE: MO - MISSOURI**

| REGISTRANT ZIP CODE 3 | 1ST QUARTER | 2ND QUARTER | 3RD QUARTER | 4TH QUARTER | TOTAL GRAMS |
|---|---|---|---|---|---|
| 630 | 99.88 | 93.70 | 89.12 | 97.94 | **380.64** |
| 631 | 200.46 | 248.86 | 205.18 | 219.72 | **874.23** |
| 633 | 56.77 | 53.88 | 52.38 | 50.51 | **213.55** |
| 634 | 17.78 | 16.08 | 15.53 | 15.76 | **65.15** |
| 635 | 18.12 | 17.54 | 18.55 | 18.33 | **72.54** |
| 636 | 26.44 | 25.03 | 24.39 | 22.60 | **98.46** |
| 637 | 16.37 | 13.25 | 15.22 | 19.68 | **64.52** |
| 638 | 21.44 | 22.60 | 19.68 | 19.96 | **83.68** |

| | | | | | |
|---|---|---|---|---|---|
| 639 | 8.85 | 8.77 | 8.55 | 11.42 | **37.59** |
| 640 | 116.76 | 113.45 | 111.82 | 116.88 | **458.92** |
| 641 | 85.76 | 73.55 | 80.55 | 87.85 | **327.71** |
| 644 | 15.67 | 15.30 | 15.79 | 15.83 | **62.59** |
| 645 | 21.04 | 19.23 | 19.18 | 17.17 | **76.62** |
| 646 | 11.73 | 12.47 | 11.47 | 11.61 | **47.27** |
| 647 | 28.36 | 25.33 | 25.75 | 25.62 | **105.07** |
| 648 | 42.46 | 37.74 | 39.58 | 40.94 | **160.73** |
| 650 | 26.71 | 27.26 | 25.84 | 26.13 | **105.94** |
| 651 | 17.54 | 15.70 | 19.50 | 16.84 | **69.58** |
| 652 | 55.85 | 50.26 | 96.32 | 229.19 | **431.62** |
| 653 | 19.63 | 17.25 | 16.71 | 18.56 | **72.14** |
| 654 | 20.21 | 19.21 | 18.67 | 19.82 | **77.92** |
| 655 | 17.49 | 17.04 | 17.23 | 18.69 | **70.46** |
| 656 | 48.96 | 46.62 | 47.60 | 44.62 | **187.81** |
| 657 | 74.40 | 69.64 | 68.58 | 69.25 | **281.86** |
| 658 | 70.86 | 69.23 | 67.89 | 63.84 | **271.82** |
| **MO - MISSOURI - Total** | **1,139.55** | **1,129.02** | **1,131.08** | **1,298.75** | **4,698.40** |

**STATE: MS - MISSISSIPPI**

| REGISTRANT ZIP CODE 3 | 1ST QUARTER | 2ND QUARTER | 3RD QUARTER | 4TH QUARTER | TOTAL GRAMS |
|---|---|---|---|---|---|
| 386 | 55.99 | 54.36 | 59.17 | 59.00 | **228.53** |
| 387 | 14.23 | 13.82 | 15.52 | 16.38 | **59.94** |
| 388 | 61.56 | 53.20 | 52.07 | 57.69 | **224.51** |
| 389 | 15.67 | 14.60 | 12.55 | 12.44 | **55.27** |
| 390 | 36.77 | 35.80 | 35.19 | 36.39 | **144.14** |
| 391 | 296.71 | 464.21 | 317.43 | 301.10 | **1,379.44** |
| 392 | 65.62 | 62.13 | 74.71 | 74.81 | **277.27** |
| 393 | 38.31 | 35.39 | 35.58 | 38.01 | **147.30** |
| 394 | 79.89 | 75.35 | 75.16 | 71.25 | **301.64** |
| 395 | 52.98 | 50.07 | 55.04 | 51.88 | **209.97** |
| 396 | 27.19 | 25.91 | 26.36 | 24.69 | **104.15** |
| 397 | 24.37 | 21.16 | 14.41 | 13.31 | **73.26** |
| **MS - MISSISSIPPI - Total** | **769.30** | **905.99** | **773.19** | **756.94** | **3,205.42** |

**STATE: MT - MONTANA**

| REGISTRANT ZIP CODE 3 | 1ST QUARTER | 2ND QUARTER | 3RD QUARTER | 4TH QUARTER | TOTAL GRAMS |
|---|---|---|---|---|---|
| 590 | 11.08 | 8.48 | 9.28 | 10.02 | **38.86** |
| 591 | 39.56 | 36.20 | 39.31 | 48.50 | **163.57** |
| 592 | 8.61 | 8.42 | 8.23 | 7.80 | **33.06** |
| 593 | 9.59 | 7.79 | 8.32 | 8.01 | **33.72** |
| 594 | 28.41 | 28.11 | 28.15 | 28.72 | **113.38** |
| 595 | 7.42 | 6.76 | 7.04 | 5.70 | **26.93** |
| 596 | 18.77 | 18.47 | 20.32 | 21.65 | **79.22** |
| 597 | 21.70 | 21.08 | 17.80 | 19.13 | **79.72** |
| 598 | 30.11 | 30.35 | 31.54 | 29.26 | **121.27** |
| 599 | 22.99 | 22.31 | 20.96 | 20.85 | **87.09** |
| **MT - MONTANA - Total** | **198.24** | **187.98** | **190.95** | **199.65** | **776.81** |

**STATE: NC - NORTH CAROLINA**

| REGISTRANT ZIP CODE 3 | 1ST QUARTER | 2ND QUARTER | 3RD QUARTER | 4TH QUARTER | TOTAL GRAMS |
|---|---|---|---|---|---|
| 270 | 36.62 | 35.30 | 35.64 | 33.34 | **140.89** |
| 271 | 58.72 | 49.89 | 55.19 | 54.79 | **218.59** |
| 272 | 86.72 | 82.50 | 77.82 | 73.64 | **320.68** |
| 273 | 52.53 | 53.92 | 53.25 | 51.75 | **211.44** |
| 274 | 35.51 | 36.40 | 42.39 | 41.38 | **155.68** |
| 275 | 160.36 | 145.39 | 148.71 | 143.43 | **597.89** |
| 276 | 78.30 | 73.42 | 68.00 | 81.88 | **301.59** |
| 277 | 57.77 | 52.63 | 42.91 | 40.05 | **193.36** |
| 278 | 105.91 | 100.86 | 99.71 | 100.72 | **407.20** |
| 279 | 37.42 | 34.95 | 34.51 | 33.37 | **140.25** |
| 280 | 122.10 | 117.51 | 113.68 | 115.30 | **468.59** |
| 281 | 94.26 | 89.72 | 89.79 | 93.29 | **367.06** |

| | | | | | |
|---|---|---|---|---|---|
| 282 | 92.03 | 92.93 | 90.36 | 86.95 | **362.27** |
| 283 | 103.57 | 110.87 | 100.23 | 99.87 | **414.54** |
| 284 | 93.26 | 98.69 | 89.10 | 91.58 | **372.64** |
| 285 | 109.96 | 107.73 | 108.54 | 107.49 | **433.73** |
| 286 | 140.59 | 130.16 | 131.33 | 133.03 | **535.11** |
| 287 | 76.59 | 72.65 | 73.30 | 71.46 | **294.00** |
| 288 | 32.73 | 32.12 | 31.83 | 35.28 | **131.96** |
| 289 | 11.02 | 9.61 | 9.15 | 9.31 | **39.08** |
| **NC - NORTH CAROLINA - Total** | **1,585.99** | **1,527.25** | **1,495.42** | **1,497.90** | **6,106.55** |

**STATE: ND - NORTH DAKOTA**

| REGISTRANT ZIP CODE 3 | 1ST QUARTER | 2ND QUARTER | 3RD QUARTER | 4TH QUARTER | TOTAL GRAMS |
|---|---|---|---|---|---|
| 580 | 5.15 | 4.64 | 4.52 | 5.04 | **19.35** |
| 581 | 29.27 | 32.43 | 19.61 | 34.79 | **116.10** |
| 582 | 26.92 | 21.73 | 19.42 | 24.41 | **92.48** |
| 583 | 10.02 | 10.11 | 9.57 | 9.02 | **38.71** |
| 584 | 7.22 | 7.44 | 6.94 | 6.90 | **28.50** |
| 585 | 35.34 | 27.55 | 25.81 | 28.50 | **117.20** |
| 586 | 9.27 | 8.98 | 8.15 | 8.45 | **34.84** |
| 587 | 12.17 | 13.50 | 11.35 | 12.19 | **49.21** |
| 588 | 8.10 | 7.52 | 6.71 | 7.62 | **29.95** |
| **ND - NORTH DAKOTA - Total** | **143.44** | **133.90** | **112.08** | **136.92** | **526.34** |

**STATE: NE - NEBRASKA**

| REGISTRANT ZIP CODE 3 | 1ST QUARTER | 2ND QUARTER | 3RD QUARTER | 4TH QUARTER | TOTAL GRAMS |
|---|---|---|---|---|---|
| 680 | 41.69 | 40.42 | 38.02 | 34.89 | **155.02** |
| 681 | 81.07 | 70.99 | 66.62 | 74.55 | **293.23** |
| 683 | 16.95 | 15.97 | 15.32 | 15.75 | **63.98** |
| 684 | 8.92 | 9.00 | 8.08 | 8.22 | **34.22** |
| 685 | 36.43 | 32.48 | 33.72 | 33.80 | **136.43** |
| 686 | 8.35 | 8.03 | 8.94 | 8.01 | **33.33** |
| 687 | 24.39 | 21.97 | 21.80 | 20.52 | **88.68** |
| 688 | 41.33 | 40.61 | 38.83 | 38.02 | **158.78** |
| 689 | 20.52 | 19.77 | 18.69 | 22.00 | **80.98** |
| 690 | 12.14 | 12.49 | 12.01 | 11.23 | **47.88** |
| 691 | 21.28 | 17.87 | 16.98 | 15.86 | **72.00** |
| 692 | 1.08 | 0.53 | 1.05 | 0.98 | **3.65** |
| 693 | 23.28 | 22.64 | 21.36 | 19.71 | **86.99** |
| **NE - NEBRASKA - Total** | **337.44** | **312.77** | **301.44** | **303.53** | **1,255.18** |

**STATE: NH - NEW HAMPSHIRE**

| REGISTRANT ZIP CODE 3 | 1ST QUARTER | 2ND QUARTER | 3RD QUARTER | 4TH QUARTER | TOTAL GRAMS |
|---|---|---|---|---|---|
| 030 | 59.67 | 54.15 | 48.53 | 47.08 | **209.44** |
| 031 | 25.36 | 25.66 | 24.74 | 24.62 | **100.37** |
| 032 | 18.18 | 16.77 | 15.67 | 17.65 | **68.27** |
| 033 | 25.45 | 27.45 | 28.10 | 32.38 | **113.39** |
| 034 | 8.25 | 8.24 | 6.71 | 7.45 | **30.65** |
| 035 | 6.78 | 6.77 | 7.34 | 7.84 | **28.73** |
| 036 | 1.25 | 1.21 | 1.24 | 0.81 | **4.51** |
| 037 | 13.89 | 12.15 | 13.68 | 15.16 | **54.89** |
| 038 | 41.26 | 41.11 | 40.63 | 44.77 | **167.78** |
| **NH - NEW HAMPSHIRE - Total** | **200.09** | **193.52** | **186.64** | **197.77** | **778.02** |

**STATE: NJ - NEW JERSEY**

| REGISTRANT ZIP CODE 3 | 1ST QUARTER | 2ND QUARTER | 3RD QUARTER | 4TH QUARTER | TOTAL GRAMS |
|---|---|---|---|---|---|
| 070 | 190.75 | 193.08 | 167.78 | 168.00 | **719.61** |
| 071 | 24.58 | 44.12 | 23.18 | 30.33 | **122.22** |
| 072 | 9.50 | 13.33 | 9.49 | 7.57 | **39.89** |
| 073 | 8.42 | 15.14 | 5.89 | 9.84 | **39.29** |
| 074 | 111.01 | 90.03 | 76.28 | 84.16 | **361.48** |
| 075 | 17.57 | 22.30 | 11.22 | 12.32 | **63.41** |
| 076 | 148.48 | 143.91 | 113.76 | 138.68 | **544.83** |
| 077 | 99.48 | 116.87 | 82.10 | 86.35 | **384.79** |
| 078 | 45.92 | 47.41 | 41.96 | 40.94 | **176.24** |

| | | | | | |
|---|---|---|---|---|---|
| 079 | 37.85 | 50.20 | 36.94 | 50.63 | **175.61** |
| 080 | 191.52 | 206.53 | 174.48 | 180.17 | **752.70** |
| 081 | 17.57 | 19.48 | 18.38 | 22.93 | **78.36** |
| 082 | 29.16 | 31.01 | 31.11 | 26.52 | **117.79** |
| 083 | 15.81 | 18.05 | 13.31 | 15.47 | **62.65** |
| 084 | 5.83 | 8.49 | 6.13 | 5.59 | **26.03** |
| 085 | 33.39 | 33.45 | 27.59 | 31.48 | **125.91** |
| 086 | 31.57 | 36.13 | 26.98 | 28.67 | **123.35** |
| 087 | 71.72 | 77.18 | 67.02 | 63.70 | **279.62** |
| 088 | 148.95 | 142.22 | 132.02 | 122.77 | **545.96** |
| 089 | 29.23 | 28.29 | 20.68 | 29.38 | **107.59** |
| **NJ - NEW JERSEY - Total** | **1,268.33** | **1,337.20** | **1,086.31** | **1,155.50** | **4,847.34** |

**STATE: NM - NEW MEXICO**

| REGISTRANT ZIP CODE 3 | 1ST QUARTER | 2ND QUARTER | 3RD QUARTER | 4TH QUARTER | TOTAL GRAMS |
|---|---|---|---|---|---|
| 870 | 12.16 | 12.27 | 12.12 | 10.85 | **47.39** |
| 871 | 108.42 | 95.71 | 96.59 | 104.02 | **404.75** |
| 873 | 5.92 | 6.93 | 5.52 | 6.14 | **24.51** |
| 874 | 15.64 | 15.58 | 12.96 | 15.42 | **59.59** |
| 875 | 31.01 | 30.32 | 28.65 | 29.63 | **119.61** |
| 877 | 4.55 | 3.67 | 3.13 | 3.10 | **14.46** |
| 878 | 0.43 | 0.45 | 0.52 | 0.41 | **1.80** |
| 879 | 1.12 | 1.37 | 1.85 | 1.11 | **5.45** |
| 880 | 23.01 | 20.79 | 20.41 | 25.22 | **89.43** |
| 881 | 6.56 | 7.74 | 7.39 | 7.17 | **28.85** |
| 882 | 28.89 | 25.92 | 24.19 | 26.49 | **105.49** |
| 883 | 7.49 | 6.86 | 6.71 | 7.03 | **28.09** |
| 884 | 2.81 | 3.02 | 3.12 | 3.01 | **11.96** |
| **NM - NEW MEXICO - Total** | **248.00** | **230.64** | **223.15** | **239.60** | **941.39** |

**STATE: NV - NEVADA**

| REGISTRANT ZIP CODE 3 | 1ST QUARTER | 2ND QUARTER | 3RD QUARTER | 4TH QUARTER | TOTAL GRAMS |
|---|---|---|---|---|---|
| 890 | 69.37 | 182.97 | 131.01 | 103.99 | **487.34** |
| 891 | 159.62 | 164.04 | 169.01 | 165.84 | **658.51** |
| 893 | 0.87 | 0.75 | 0.76 | 0.99 | **3.37** |
| 894 | 18.54 | 19.83 | 20.55 | 20.66 | **79.58** |
| 895 | 32.78 | 29.54 | 33.19 | 37.46 | **132.97** |
| 897 | 7.38 | 7.31 | 7.59 | 7.33 | **29.61** |
| 898 | 8.98 | 8.29 | 5.37 | 6.04 | **28.68** |
| **NV - NEVADA - Total** | **297.55** | **412.72** | **367.47** | **342.32** | **1,420.06** |

**STATE: NY - NEW YORK**

| REGISTRANT ZIP CODE 3 | 1ST QUARTER | 2ND QUARTER | 3RD QUARTER | 4TH QUARTER | TOTAL GRAMS |
|---|---|---|---|---|---|
| 100 | 316.05 | 392.36 | 225.32 | 222.58 | **1,156.31** |
| 101 | 3.52 | 2.94 | 3.65 | 3.55 | **13.65** |
| 102 | 0.01 | 0.03 | 0.09 | 0.37 | **0.51** |
| 103 | 80.03 | 109.83 | 74.96 | 82.44 | **347.26** |
| 104 | 102.42 | 171.82 | 67.47 | 80.49 | **422.19** |
| 105 | 76.49 | 86.39 | 60.73 | 63.49 | **287.10** |
| 106 | 11.69 | 17.38 | 10.22 | 8.29 | **47.58** |
| 107 | 24.98 | 27.93 | 21.51 | 20.37 | **94.78** |
| 108 | 4.03 | 6.89 | 3.17 | 3.23 | **17.31** |
| 109 | 68.22 | 70.72 | 65.28 | 63.64 | **267.86** |
| 110 | 66.24 | 95.10 | 60.91 | 59.42 | **281.67** |
| 111 | 12.37 | 11.76 | 8.79 | 7.83 | **40.75** |
| 112 | 131.86 | 217.65 | 128.53 | 129.57 | **607.62** |
| 113 | 69.53 | 88.44 | 57.53 | 56.95 | **272.44** |
| 114 | 33.63 | 39.79 | 30.45 | 28.79 | **132.66** |
| 115 | 88.26 | 140.71 | 94.96 | 67.48 | **391.41** |
| 116 | 5.01 | 5.98 | 3.85 | 4.13 | **18.97** |
| 117 | 300.74 | 314.87 | 254.10 | 248.37 | **1,118.08** |
| 118 | 11.53 | 12.64 | 9.20 | 11.68 | **45.04** |
| 119 | 34.98 | 36.00 | 31.60 | 30.05 | **132.63** |

| | | | | | |
|---|---|---|---|---|---|
| 120 | 63.85 | 57.87 | 62.30 | 54.56 | **238.59** |
| 121 | 28.73 | 30.11 | 26.43 | 28.49 | **113.77** |
| 122 | 47.53 | 45.05 | 33.33 | 37.07 | **162.99** |
| 123 | 24.37 | 25.02 | 28.49 | 27.25 | **105.13** |
| 124 | 32.15 | 32.59 | 31.06 | 28.34 | **124.14** |
| 125 | 56.51 | 62.72 | 55.51 | 59.81 | **234.54** |
| 126 | 24.51 | 26.84 | 19.38 | 21.79 | **92.53** |
| 127 | 11.44 | 12.98 | 11.00 | 9.28 | **44.70** |
| 128 | 52.19 | 54.08 | 48.99 | 44.22 | **199.49** |
| 129 | 24.05 | 23.24 | 25.28 | 23.09 | **95.66** |
| 130 | 59.88 | 50.77 | 53.52 | 51.69 | **215.86** |
| 131 | 16.37 | 13.06 | 14.44 | 15.70 | **59.57** |
| 132 | 52.11 | 46.64 | 49.99 | 53.54 | **202.28** |
| 133 | 19.54 | 20.59 | 21.93 | 21.63 | **83.68** |
| 134 | 31.54 | 26.29 | 25.59 | 24.50 | **107.93** |
| 135 | 10.67 | 9.85 | 8.41 | 9.68 | **38.61** |
| 136 | 36.53 | 27.82 | 34.19 | 29.86 | **128.40** |
| 137 | 18.04 | 18.37 | 17.21 | 14.60 | **68.22** |
| 138 | 38.65 | 38.98 | 32.53 | 33.90 | **144.06** |
| 139 | 13.50 | 10.66 | 11.61 | 12.20 | **47.97** |
| 140 | 57.24 | 48.72 | 45.00 | 44.22 | **195.18** |
| 141 | 30.03 | 26.40 | 25.34 | 26.47 | **108.24** |
| 142 | 94.35 | 97.03 | 84.93 | 88.08 | **364.39** |
| 143 | 8.50 | 7.22 | 7.27 | 8.75 | **31.75** |
| 144 | 43.28 | 44.31 | 44.33 | 40.95 | **172.86** |
| 145 | 38.62 | 35.74 | 33.90 | 33.43 | **141.70** |
| 146 | 100.83 | 106.84 | 85.12 | 103.35 | **396.15** |
| 147 | 27.90 | 28.21 | 26.67 | 26.03 | **108.81** |
| 148 | 49.69 | 45.68 | 44.60 | 41.73 | **181.71** |
| 149 | 14.41 | 12.81 | 12.10 | 13.01 | **52.32** |
| **NY - NEW YORK - Total** | **2,568.61** | **2,935.72** | **2,232.76** | **2,219.96** | **9,957.05** |

**STATE: OH - OHIO**

| REGISTRANT ZIP CODE 3 | 1ST QUARTER | 2ND QUARTER | 3RD QUARTER | 4TH QUARTER | TOTAL GRAMS |
|---|---|---|---|---|---|
| 430 | 79.58 | 78.21 | 72.54 | 76.38 | **306.71** |
| 431 | 40.52 | 39.11 | 40.32 | 41.97 | **161.92** |
| 432 | 96.99 | 92.29 | 82.97 | 95.45 | **367.69** |
| 433 | 19.43 | 17.50 | 18.18 | 18.83 | **73.94** |
| 434 | 12.22 | 12.38 | 11.56 | 12.59 | **48.75** |
| 435 | 38.78 | 39.72 | 36.70 | 39.45 | **154.64** |
| 436 | 72.68 | 75.76 | 67.41 | 72.45 | **288.30** |
| 437 | 19.11 | 21.46 | 17.84 | 20.97 | **79.38** |
| 438 | 1.88 | 2.23 | 2.57 | 2.68 | **9.36** |
| 439 | 14.59 | 16.72 | 13.20 | 13.28 | **57.79** |
| 440 | 56.31 | 58.63 | 55.03 | 57.26 | **227.23** |
| 441 | 194.35 | 117.86 | 98.85 | 123.78 | **534.84** |
| 442 | 34.16 | 37.58 | 33.72 | 37.56 | **143.02** |
| 443 | 22.99 | 28.57 | 19.58 | 22.29 | **93.43** |
| 444 | 27.04 | 28.92 | 27.65 | 28.46 | **112.07** |
| 445 | 36.08 | 56.24 | 26.69 | 31.40 | **150.41** |
| 446 | 35.23 | 34.51 | 29.55 | 32.73 | **132.02** |
| 447 | 29.43 | 32.31 | 27.89 | 32.40 | **122.04** |
| 448 | 27.81 | 28.11 | 30.06 | 26.69 | **112.67** |
| 449 | 10.78 | 10.21 | 10.65 | 11.98 | **43.63** |
| 450 | 86.07 | 84.97 | 78.61 | 79.89 | **329.53** |
| 451 | 18.37 | 21.38 | 19.74 | 20.88 | **80.37** |
| 452 | 91.02 | 114.35 | 84.12 | 96.57 | **386.06** |
| 453 | 68.62 | 63.18 | 58.25 | 59.74 | **249.80** |
| 454 | 53.24 | 50.80 | 51.77 | 58.36 | **214.16** |
| 455 | 10.62 | 17.24 | 10.78 | 11.38 | **50.02** |
| 456 | 56.26 | 51.95 | 54.24 | 53.44 | **215.90** |

| | | | | | |
|---|---|---|---|---|---|
| 457 | 14.80 | 14.47 | 13.03 | 13.92 | **56.22** |
| 458 | 28.89 | 30.29 | 28.40 | 28.62 | **116.21** |
| **OH - OHIO - Total** | **1,297.88** | **1,276.94** | **1,121.88** | **1,221.41** | **4,918.11** |

**STATE: OK - OKLAHOMA**

| REGISTRANT ZIP CODE 3 | 1ST QUARTER | 2ND QUARTER | 3RD QUARTER | 4TH QUARTER | TOTAL GRAMS |
|---|---|---|---|---|---|
| 730 | 106.79 | 92.34 | 92.13 | 82.95 | **374.21** |
| 731 | 127.45 | 109.19 | 110.14 | 120.48 | **467.26** |
| 734 | 16.94 | 17.19 | 14.38 | 14.47 | **62.98** |
| 735 | 56.49 | 53.96 | 59.71 | 55.00 | **225.15** |
| 736 | 17.84 | 14.94 | 16.41 | 15.86 | **65.05** |
| 737 | 20.40 | 18.09 | 17.62 | 16.72 | **72.83** |
| 738 | 5.19 | 5.69 | 4.86 | 4.79 | **20.53** |
| 739 | 2.43 | 2.41 | 2.12 | 2.04 | **9.00** |
| 740 | 82.08 | 79.50 | 76.56 | 75.91 | **314.05** |
| 741 | 80.78 | 71.80 | 75.93 | 76.89 | **305.40** |
| 743 | 14.85 | 14.60 | 12.92 | 12.47 | **54.84** |
| 744 | 36.49 | 33.82 | 28.82 | 27.66 | **126.80** |
| 745 | 10.38 | 11.54 | 10.15 | 10.56 | **42.62** |
| 746 | 10.58 | 10.40 | 13.48 | 14.06 | **48.52** |
| 747 | 12.39 | 12.95 | 11.04 | 11.90 | **48.28** |
| 748 | 29.17 | 26.41 | 24.60 | 23.91 | **104.09** |
| 749 | 8.37 | 8.03 | 8.37 | 8.68 | **33.45** |
| **OK - OKLAHOMA - Total** | **638.63** | **582.87** | **579.21** | **574.35** | **2,375.05** |

**STATE: OR - OREGON**

| REGISTRANT ZIP CODE 3 | 1ST QUARTER | 2ND QUARTER | 3RD QUARTER | 4TH QUARTER | TOTAL GRAMS |
|---|---|---|---|---|---|
| 970 | 78.55 | 77.36 | 77.36 | 75.00 | **308.26** |
| 971 | 39.49 | 38.15 | 37.87 | 39.12 | **154.63** |
| 972 | 114.58 | 115.04 | 112.61 | 106.75 | **448.98** |
| 973 | 70.62 | 59.14 | 65.16 | 65.27 | **260.18** |
| 974 | 75.83 | 69.59 | 71.19 | 72.09 | **288.69** |
| 975 | 55.12 | 56.14 | 51.59 | 49.21 | **212.06** |
| 976 | 7.04 | 6.10 | 7.60 | 7.53 | **28.27** |
| 977 | 27.51 | 25.23 | 29.10 | 33.69 | **115.53** |
| 978 | 24.39 | 24.17 | 22.26 | 23.39 | **94.20** |
| 979 | 7.20 | 6.34 | 6.49 | 7.40 | **27.44** |
| **OR - OREGON - Total** | **500.33** | **477.26** | **481.23** | **479.43** | **1,938.25** |

**STATE: PA - PENNSYLVANIA**

| REGISTRANT ZIP CODE 3 | 1ST QUARTER | 2ND QUARTER | 3RD QUARTER | 4TH QUARTER | TOTAL GRAMS |
|---|---|---|---|---|---|
| 150 | 106.88 | 93.45 | 91.14 | 86.57 | **378.04** |
| 151 | 57.94 | 54.70 | 51.41 | 52.80 | **216.86** |
| 152 | 153.05 | 129.85 | 127.71 | 136.99 | **547.60** |
| 153 | 25.54 | 23.60 | 22.49 | 24.02 | **95.64** |
| 154 | 19.46 | 20.50 | 18.10 | 17.09 | **75.16** |
| 155 | 18.84 | 16.17 | 15.68 | 11.62 | **62.31** |
| 156 | 57.19 | 48.82 | 47.95 | 47.22 | **201.19** |
| 157 | 38.72 | 32.49 | 35.18 | 33.06 | **139.46** |
| 158 | 34.59 | 31.38 | 24.65 | 25.93 | **116.56** |
| 159 | 35.06 | 32.68 | 31.97 | 32.40 | **132.10** |
| 160 | 51.88 | 45.09 | 48.53 | 45.61 | **191.11** |
| 161 | 40.42 | 36.81 | 34.41 | 37.14 | **148.78** |
| 162 | 15.77 | 13.80 | 13.43 | 13.61 | **56.61** |
| 163 | 48.66 | 47.94 | 46.50 | 42.88 | **185.97** |
| 164 | 14.02 | 14.18 | 15.04 | 14.83 | **58.07** |
| 165 | 43.98 | 37.94 | 37.19 | 41.06 | **160.16** |
| 166 | 57.54 | 50.44 | 51.15 | 57.79 | **216.92** |
| 167 | 7.95 | 6.86 | 6.93 | 7.41 | **29.15** |
| 168 | 19.01 | 19.09 | 20.59 | 18.78 | **77.47** |
| 169 | 10.22 | 8.49 | 9.31 | 8.64 | **36.66** |
| 170 | 108.13 | 106.51 | 91.46 | 101.82 | **407.92** |
| 171 | 20.72 | 28.46 | 17.55 | 14.25 | **80.98** |

| | | | | | |
|---|---|---|---|---|---|
| 172 | 31.63 | 29.67 | 27.43 | 28.62 | **117.35** |
| 173 | 50.34 | 48.54 | 46.88 | 51.44 | **197.19** |
| 174 | 30.36 | 30.58 | 27.06 | 30.19 | **118.20** |
| 175 | 33.58 | 32.71 | 31.60 | 33.07 | **130.96** |
| 176 | 34.06 | 30.82 | 29.88 | 30.76 | **125.52** |
| 177 | 36.40 | 37.87 | 35.43 | 40.43 | **150.14** |
| 178 | 56.30 | 48.44 | 51.72 | 53.52 | **209.98** |
| 179 | 22.61 | 21.31 | 20.25 | 21.25 | **85.42** |
| 180 | 74.12 | 78.20 | 77.46 | 73.79 | **303.56** |
| 181 | 49.17 | 61.53 | 43.64 | 49.54 | **203.87** |
| 182 | 24.64 | 26.38 | 21.76 | 23.80 | **96.58** |
| 183 | 23.84 | 23.32 | 22.47 | 23.92 | **93.54** |
| 184 | 38.12 | 36.78 | 35.35 | 33.86 | **144.11** |
| 185 | 35.65 | 33.95 | 29.85 | 34.74 | **134.19** |
| 186 | 35.34 | 35.89 | 35.72 | 34.92 | **141.87** |
| 187 | 129.84 | 149.48 | 143.95 | 144.88 | **568.14** |
| 188 | 17.92 | 16.49 | 17.76 | 17.42 | **69.58** |
| 189 | 57.13 | 57.84 | 52.84 | 58.50 | **226.31** |
| 190 | 236.38 | 420.34 | 442.32 | 373.46 | **1,472.50** |
| 191 | 169.58 | 185.86 | 153.12 | 177.43 | **685.98** |
| 193 | 53.76 | 51.17 | 48.67 | 53.80 | **207.40** |
| 194 | 90.16 | 87.67 | 75.10 | 74.99 | **327.92** |
| 195 | 25.65 | 26.76 | 25.45 | 25.61 | **103.47** |
| 196 | 25.21 | 25.06 | 22.03 | 26.82 | **99.12** |
| **PA - PENNSYLVANIA - Total** | **2,367.35** | **2,495.90** | **2,376.08** | **2,388.29** | **9,627.61** |

**STATE: PR - PUERTO RICO**

| REGISTRANT ZIP CODE 3 | 1ST QUARTER | 2ND QUARTER | 3RD QUARTER | 4TH QUARTER | TOTAL GRAMS |
|---|---|---|---|---|---|
| 006 | 52.54 | 35.06 | 50.13 | 45.71 | **183.44** |
| 007 | 54.26 | 40.87 | 53.56 | 55.96 | **204.65** |
| 009 | 64.87 | 50.82 | 70.71 | 61.81 | **248.22** |
| **PR - PUERTO RICO - Total** | **171.68** | **126.74** | **174.40** | **163.48** | **636.31** |

**STATE: RI - RHODE ISLAND**

| REGISTRANT ZIP CODE 3 | 1ST QUARTER | 2ND QUARTER | 3RD QUARTER | 4TH QUARTER | TOTAL GRAMS |
|---|---|---|---|---|---|
| 028 | 63.65 | 61.51 | 63.49 | 58.13 | **246.77** |
| 029 | 41.29 | 48.11 | 43.57 | 41.45 | **174.43** |
| **RI - RHODE ISLAND - Total** | **104.94** | **109.62** | **107.06** | **99.58** | **421.20** |

**STATE: SC - SOUTH CAROLINA**

| REGISTRANT ZIP CODE 3 | 1ST QUARTER | 2ND QUARTER | 3RD QUARTER | 4TH QUARTER | TOTAL GRAMS |
|---|---|---|---|---|---|
| 290 | 49.79 | 51.84 | 50.15 | 47.46 | **199.24** |
| 291 | 60.46 | 56.54 | 57.32 | 55.78 | **230.10** |
| 292 | 58.90 | 63.80 | 45.51 | 49.09 | **217.31** |
| 293 | 46.48 | 38.00 | 41.61 | 41.85 | **167.95** |
| 294 | 123.64 | 115.15 | 111.96 | 104.16 | **454.90** |
| 295 | 135.57 | 128.29 | 132.15 | 118.85 | **514.87** |
| 296 | 131.98 | 132.05 | 136.39 | 136.18 | **536.59** |
| 297 | 30.58 | 30.63 | 29.07 | 30.72 | **120.99** |
| 298 | 49.05 | 48.30 | 46.58 | 48.00 | **191.93** |
| 299 | 47.98 | 42.31 | 41.27 | 45.01 | **176.57** |
| **SC - SOUTH CAROLINA - Total** | **734.42** | **706.91** | **692.01** | **677.12** | **2,810.45** |

**STATE: SD - SOUTH DAKOTA**

| REGISTRANT ZIP CODE 3 | 1ST QUARTER | 2ND QUARTER | 3RD QUARTER | 4TH QUARTER | TOTAL GRAMS |
|---|---|---|---|---|---|
| 570 | 31.43 | 29.98 | 27.92 | 29.94 | **119.27** |
| 571 | 57.83 | 59.17 | 54.87 | 56.45 | **228.31** |
| 572 | 10.26 | 9.99 | 10.35 | 9.80 | **40.40** |
| 573 | 13.86 | 12.50 | 14.70 | 12.28 | **53.34** |
| 574 | 10.03 | 10.04 | 10.16 | 10.29 | **40.52** |
| 575 | 12.19 | 10.89 | 11.09 | 11.24 | **45.41** |
| 576 | 2.21 | 1.76 | 2.17 | 2.37 | **8.51** |
| 577 | 17.65 | 19.02 | 13.40 | 14.15 | **64.22** |
| **SD - SOUTH DAKOTA - Total** | **155.44** | **153.34** | **144.67** | **146.52** | **599.98** |

**STATE: TN - TENNESSEE**

| REGISTRANT ZIP CODE 3 | 1ST QUARTER | 2ND QUARTER | 3RD QUARTER | 4TH QUARTER | TOTAL GRAMS |
|---|---|---|---|---|---|
| 370 | 88.28 | 77.12 | 76.82 | 75.62 | **317.84** |
| 371 | 81.88 | 68.79 | 67.65 | 72.61 | **290.93** |
| 372 | 93.54 | 98.49 | 91.85 | 104.02 | **387.89** |
| 373 | 74.28 | 69.00 | 71.24 | 65.59 | **280.11** |
| 374 | 33.60 | 31.75 | 33.15 | 36.03 | **134.52** |
| 376 | 69.64 | 64.84 | 63.51 | 68.15 | **266.14** |
| 377 | 37.93 | 34.97 | 34.41 | 35.28 | **142.59** |
| 378 | 74.97 | 69.06 | 71.86 | 68.83 | **284.72** |
| 379 | 63.78 | 58.35 | 58.68 | 59.87 | **240.69** |
| 380 | 48.63 | 52.65 | 51.97 | 46.27 | **199.52** |
| 381 | 77.84 | 285.48 | 352.00 | 260.91 | **976.23** |
| 382 | 14.77 | 15.93 | 13.12 | 13.41 | **57.23** |
| 383 | 42.30 | 42.92 | 46.72 | 45.39 | **177.33** |
| 384 | 19.70 | 18.93 | 16.85 | 23.11 | **78.59** |
| 385 | 39.67 | 36.88 | 40.06 | 39.66 | **156.27** |
| **TN - TENNESSEE - Total** | **860.83** | **1,025.15** | **1,089.89** | **1,014.74** | **3,990.60** |

**STATE: TX - TEXAS**

| REGISTRANT ZIP CODE 3 | 1ST QUARTER | 2ND QUARTER | 3RD QUARTER | 4TH QUARTER | TOTAL GRAMS |
|---|---|---|---|---|---|
| 750 | 275.98 | 258.64 | 259.32 | 255.68 | **1,049.61** |
| 751 | 78.14 | 74.86 | 75.75 | 80.80 | **309.55** |
| 752 | 235.81 | 184.91 | 211.87 | 208.61 | **841.19** |
| 753 | 10.45 | 6.91 | 9.72 | 9.94 | **37.01** |
| 754 | 71.01 | 68.72 | 68.01 | 69.98 | **277.72** |
| 755 | 23.10 | 22.26 | 21.98 | 22.83 | **90.16** |
| 756 | 85.19 | 82.69 | 83.26 | 83.69 | **334.83** |
| 757 | 85.71 | 77.94 | 88.05 | 80.66 | **332.36** |
| 758 | 9.51 | 11.05 | 10.10 | 10.10 | **40.75** |
| 759 | 29.81 | 27.01 | 27.43 | 26.95 | **111.20** |
| 760 | 159.91 | 154.78 | 154.71 | 168.97 | **638.37** |
| 761 | 145.98 | 140.57 | 143.05 | 152.41 | **582.00** |
| 762 | 66.81 | 62.62 | 60.02 | 67.47 | **256.91** |
| 763 | 40.71 | 35.58 | 35.37 | 38.65 | **150.31** |
| 764 | 26.03 | 24.72 | 26.98 | 23.78 | **101.51** |
| 765 | 45.72 | 40.99 | 43.71 | 38.74 | **169.16** |
| 766 | 10.24 | 9.39 | 8.69 | 8.94 | **37.26** |
| 767 | 19.76 | 20.93 | 19.33 | 16.83 | **76.85** |
| 768 | 24.30 | 21.29 | 21.88 | 19.87 | **87.35** |
| 769 | 14.00 | 13.17 | 14.16 | 14.24 | **55.57** |
| 770 | 225.48 | 218.08 | 234.71 | 200.21 | **878.48** |
| 772 | 0 | 0 | 0.01 | 0 | **0.01** |
| 773 | 128.36 | 124.92 | 133.73 | 128.61 | **515.62** |
| 774 | 95.28 | 91.05 | 92.75 | 87.96 | **367.04** |
| 775 | 132.74 | 132.03 | 139.92 | 131.21 | **535.90** |
| 776 | 36.92 | 29.02 | 32.29 | 28.11 | **126.34** |
| 777 | 22.06 | 23.29 | 21.38 | 22.43 | **89.15** |
| 778 | 41.43 | 36.38 | 35.44 | 36.09 | **149.34** |
| 779 | 27.42 | 26.85 | 29.65 | 29.23 | **113.14** |
| 780 | 47.56 | 47.34 | 58.37 | 56.44 | **209.70** |
| 781 | 46.46 | 40.56 | 41.79 | 41.46 | **170.28** |
| 782 | 231.56 | 219.96 | 266.39 | 235.02 | **952.93** |
| 783 | 21.27 | 22.58 | 22.70 | 20.61 | **87.17** |
| 784 | 51.11 | 49.56 | 70.31 | 43.87 | **214.86** |
| 785 | 87.49 | 83.78 | 144.60 | 89.92 | **405.79** |
| 786 | 89.02 | 96.76 | 100.51 | 101.29 | **387.58** |
| 787 | 95.19 | 88.73 | 97.98 | 91.58 | **373.47** |
| 788 | 7.57 | 9.08 | 10.00 | 7.77 | **34.42** |
| 789 | 5.85 | 6.33 | 6.51 | 5.61 | **24.29** |
| 790 | 30.97 | 27.40 | 28.47 | 29.87 | **116.71** |

| | | | | | |
|---|---|---|---|---|---|
| 791 | 55.99 | 48.05 | 53.77 | 61.02 | **218.82** |
| 792 | 7.53 | 6.21 | 6.21 | 6.10 | **26.05** |
| 793 | 13.74 | 13.68 | 14.30 | 14.55 | **56.28** |
| 794 | 71.98 | 64.02 | 65.89 | 77.76 | **279.65** |
| 795 | 20.11 | 15.93 | 16.63 | 15.96 | **68.64** |
| 796 | 42.67 | 38.38 | 40.30 | 39.31 | **160.66** |
| 797 | 23.51 | 23.69 | 24.66 | 33.23 | **105.10** |
| 798 | 3.36 | 2.47 | 2.87 | 3.32 | **12.02** |
| 799 | 69.82 | 62.71 | 72.43 | 96.09 | **301.04** |
| **TX - TEXAS - Total** | **3,190.63** | **2,987.86** | **3,247.94** | **3,133.76** | **12,560.18** |

**STATE: UT - UTAH**

| REGISTRANT ZIP CODE 3 | 1ST QUARTER | 2ND QUARTER | 3RD QUARTER | 4TH QUARTER | **TOTAL GRAMS** |
|---|---|---|---|---|---|
| 840 | 186.36 | 172.31 | 170.88 | 168.99 | **698.54** |
| 841 | 126.58 | 132.60 | 111.65 | 105.67 | **476.50** |
| 843 | 29.12 | 26.77 | 32.15 | 27.96 | **116.00** |
| 844 | 36.62 | 38.87 | 39.19 | 43.67 | **158.35** |
| 845 | 16.75 | 16.82 | 16.12 | 16.69 | **66.38** |
| 846 | 47.78 | 51.14 | 45.52 | 47.19 | **191.64** |
| 847 | 85.84 | 78.13 | 77.25 | 78.69 | **319.91** |
| **UT - UTAH - Total** | **529.05** | **516.65** | **492.78** | **488.86** | **2,027.33** |

**STATE: VA - VIRGINIA**

| REGISTRANT ZIP CODE 3 | 1ST QUARTER | 2ND QUARTER | 3RD QUARTER | 4TH QUARTER | **TOTAL GRAMS** |
|---|---|---|---|---|---|
| 201 | 72.52 | 71.35 | 65.20 | 68.90 | **277.97** |
| 220 | 41.09 | 44.94 | 33.40 | 28.34 | **147.77** |
| 221 | 22.59 | 23.51 | 21.21 | 26.03 | **93.34** |
| 222 | 8.00 | 11.12 | 4.64 | 7.71 | **31.46** |
| 223 | 19.56 | 23.21 | 18.77 | 16.70 | **78.24** |
| 224 | 26.78 | 26.02 | 22.50 | 20.79 | **96.09** |
| 225 | 8.34 | 10.48 | 7.68 | 7.75 | **34.25** |
| 226 | 29.57 | 29.10 | 25.39 | 29.33 | **113.38** |
| 227 | 7.93 | 7.74 | 6.25 | 6.40 | **28.32** |
| 228 | 27.62 | 29.11 | 24.84 | 26.41 | **107.98** |
| 229 | 50.13 | 47.09 | 46.99 | 48.01 | **192.22** |
| 230 | 34.04 | 29.13 | 27.23 | 24.03 | **114.43** |
| 231 | 53.49 | 64.23 | 49.00 | 48.11 | **214.82** |
| 232 | 98.28 | 90.94 | 78.78 | 84.40 | **352.40** |
| 233 | 43.85 | 40.37 | 42.35 | 41.14 | **167.72** |
| 234 | 81.01 | 85.25 | 78.27 | 77.52 | **322.04** |
| 235 | 34.49 | 32.50 | 33.25 | 27.46 | **127.70** |
| 236 | 40.83 | 47.70 | 41.60 | 43.89 | **174.03** |
| 237 | 17.73 | 25.15 | 21.74 | 16.08 | **80.71** |
| 238 | 28.51 | 28.32 | 28.21 | 28.65 | **113.68** |
| 239 | 8.41 | 10.15 | 10.45 | 10.54 | **39.54** |
| 240 | 62.34 | 59.30 | 57.30 | 58.65 | **237.60** |
| 241 | 40.21 | 39.27 | 38.05 | 40.55 | **158.09** |
| 242 | 45.10 | 38.84 | 34.24 | 36.85 | **155.03** |
| 243 | 24.30 | 20.41 | 19.98 | 18.66 | **83.35** |
| 244 | 26.40 | 25.74 | 27.37 | 25.02 | **104.53** |
| 245 | 54.24 | 51.93 | 50.63 | 46.19 | **202.99** |
| 246 | 11.78 | 11.36 | 11.86 | 11.70 | **46.70** |
| **VA - VIRGINIA - Total** | **1,019.15** | **1,024.25** | **927.17** | **925.79** | **3,896.36** |

**STATE: VI - VIRGIN ISLANDS**

| REGISTRANT ZIP CODE 3 | 1ST QUARTER | 2ND QUARTER | 3RD QUARTER | 4TH QUARTER | **TOTAL GRAMS** |
|---|---|---|---|---|---|
| 008 | 3.86 | 5.22 | 4.05 | 4.76 | **17.90** |
| **VI - VIRGIN ISLANDS - Total** | **3.86** | **5.22** | **4.05** | **4.76** | **17.90** |

**STATE: VT - VERMONT**

| REGISTRANT ZIP CODE 3 | 1ST QUARTER | 2ND QUARTER | 3RD QUARTER | 4TH QUARTER | **TOTAL GRAMS** |
|---|---|---|---|---|---|
| 050 | 4.25 | 4.22 | 5.03 | 5.58 | **19.09** |
| 051 | 3.05 | 2.76 | 3.67 | 2.81 | **12.30** |
| 052 | 8.39 | 7.57 | 7.58 | 8.49 | **32.03** |

| | | | | | |
|---|---|---|---|---|---|
| 053 | 2.92 | 1.81 | 2.07 | 2.09 | **8.88** |
| 054 | 39.90 | 38.61 | 37.82 | 37.03 | **153.36** |
| 056 | 14.45 | 12.21 | 12.05 | 12.85 | **51.56** |
| 057 | 17.69 | 15.76 | 18.11 | 18.46 | **70.02** |
| 058 | 9.96 | 9.95 | 11.68 | 9.60 | **41.19** |
| **VT - VERMONT - Total** | **100.62** | **92.90** | **98.00** | **96.91** | **388.43** |

**STATE: WA - WASHINGTON**

| REGISTRANT ZIP CODE 3 | 1ST QUARTER | 2ND QUARTER | 3RD QUARTER | 4TH QUARTER | TOTAL GRAMS |
|---|---|---|---|---|---|
| 980 | 120.72 | 119.77 | 114.30 | 125.41 | **480.20** |
| 981 | 103.98 | 101.92 | 102.67 | 109.43 | **418.01** |
| 982 | 83.93 | 75.02 | 81.83 | 91.28 | **332.07** |
| 983 | 87.96 | 80.29 | 86.79 | 89.15 | **344.19** |
| 984 | 47.77 | 41.62 | 48.99 | 47.08 | **185.45** |
| 985 | 47.93 | 43.66 | 41.05 | 43.46 | **176.11** |
| 986 | 101.27 | 93.16 | 91.37 | 94.07 | **379.87** |
| 988 | 22.33 | 21.35 | 24.75 | 21.35 | **89.78** |
| 989 | 34.13 | 32.49 | 29.30 | 32.71 | **128.63** |
| 990 | 13.10 | 14.82 | 14.59 | 12.06 | **54.57** |
| 991 | 19.57 | 19.20 | 18.41 | 19.40 | **76.58** |
| 992 | 96.63 | 79.57 | 82.19 | 81.46 | **339.85** |
| 993 | 68.60 | 64.29 | 56.09 | 57.24 | **246.23** |
| 994 | 16.79 | 14.29 | 14.13 | 12.18 | **57.39** |
| **WA - WASHINGTON - Total** | **864.73** | **801.47** | **806.47** | **836.28** | **3,308.95** |

**STATE: WI - WISCONSIN**

| REGISTRANT ZIP CODE 3 | 1ST QUARTER | 2ND QUARTER | 3RD QUARTER | 4TH QUARTER | TOTAL GRAMS |
|---|---|---|---|---|---|
| 530 | 73.94 | 74.53 | 69.04 | 68.01 | **285.52** |
| 531 | 80.01 | 79.79 | 89.29 | 75.05 | **324.14** |
| 532 | 134.99 | 109.15 | 88.40 | 97.84 | **430.38** |
| 534 | 12.88 | 11.35 | 11.12 | 11.26 | **46.61** |
| 535 | 68.26 | 61.20 | 62.98 | 64.44 | **256.88** |
| 537 | 59.23 | 45.92 | 49.32 | 51.41 | **205.88** |
| 538 | 13.14 | 10.81 | 12.20 | 10.59 | **46.74** |
| 539 | 36.43 | 36.58 | 35.64 | 36.83 | **145.49** |
| 540 | 11.31 | 10.35 | 9.42 | 10.47 | **41.56** |
| 541 | 25.72 | 23.60 | 23.48 | 23.13 | **95.94** |
| 542 | 13.98 | 13.72 | 13.64 | 14.19 | **55.54** |
| 543 | 28.61 | 29.83 | 23.75 | 25.50 | **107.69** |
| 544 | 43.64 | 38.92 | 36.15 | 40.03 | **158.74** |
| 545 | 6.23 | 7.71 | 7.24 | 6.46 | **27.66** |
| 546 | 44.58 | 37.44 | 38.31 | 35.71 | **156.04** |
| 547 | 46.63 | 37.33 | 40.90 | 42.90 | **167.76** |
| 548 | 24.08 | 21.98 | 23.37 | 23.33 | **92.76** |
| 549 | 101.50 | 93.55 | 87.50 | 84.74 | **367.29** |
| **WI - WISCONSIN - Total** | **825.16** | **743.78** | **721.77** | **721.90** | **3,012.62** |

**STATE: WV - WEST VIRGINIA**

| REGISTRANT ZIP CODE 3 | 1ST QUARTER | 2ND QUARTER | 3RD QUARTER | 4TH QUARTER | TOTAL GRAMS |
|---|---|---|---|---|---|
| 247 | 9.03 | 9.94 | 9.86 | 11.50 | **40.33** |
| 248 | 3.40 | 3.07 | 3.07 | 2.89 | **12.43** |
| 249 | 2.83 | 2.74 | 3.03 | 3.33 | **11.92** |
| 250 | 9.33 | 8.53 | 9.56 | 9.29 | **36.70** |
| 251 | 6.60 | 6.27 | 6.51 | 5.80 | **25.19** |
| 252 | 4.84 | 4.40 | 4.14 | 4.17 | **17.54** |
| 253 | 43.59 | 37.44 | 33.63 | 34.18 | **148.83** |
| 254 | 36.09 | 32.20 | 30.78 | 31.78 | **130.86** |
| 255 | 16.92 | 14.72 | 13.87 | 13.75 | **59.25** |
| 256 | 5.70 | 6.57 | 6.65 | 4.42 | **23.34** |
| 257 | 29.49 | 30.47 | 26.19 | 35.22 | **121.38** |
| 258 | 14.10 | 15.38 | 13.92 | 14.81 | **58.22** |
| 259 | 3.70 | 3.56 | 4.04 | 4.04 | **15.35** |
| 260 | 17.46 | 20.19 | 14.83 | 17.24 | **69.72** |

| | | | | | |
|---|---|---|---|---|---|
| 261 | 10.48 | 9.77 | 10.33 | 11.94 | **42.52** |
| 262 | 9.70 | 10.31 | 8.77 | 9.03 | **37.82** |
| 263 | 16.87 | 14.81 | 13.93 | 12.19 | **57.80** |
| 264 | 6.19 | 5.15 | 5.38 | 6.05 | **22.77** |
| 265 | 30.77 | 28.07 | 31.09 | 36.92 | **126.85** |
| 266 | 4.23 | 3.95 | 4.53 | 5.19 | **17.90** |
| 267 | 11.27 | 12.64 | 13.21 | 12.22 | **49.34** |
| 268 | 2.53 | 3.07 | 2.45 | 3.03 | **11.08** |
| **WV - WEST VIRGINIA - Total** | **295.09** | **283.27** | **269.78** | **288.97** | **1,137.12** |

**STATE: WY - WYOMING**

| REGISTRANT ZIP CODE 3 | 1ST QUARTER | 2ND QUARTER | 3RD QUARTER | 4TH QUARTER | TOTAL GRAMS |
|---|---|---|---|---|---|
| 820 | 13.92 | 13.58 | 13.66 | 14.58 | **55.73** |
| 822 | 3.53 | 3.80 | 2.72 | 2.10 | **12.14** |
| 823 | 0.53 | 0.57 | 0.66 | 0.48 | **2.23** |
| 824 | 14.83 | 13.61 | 13.66 | 13.32 | **55.41** |
| 825 | 4.46 | 4.78 | 3.66 | 5.01 | **17.91** |
| 826 | 13.68 | 15.38 | 16.06 | 16.58 | **61.70** |
| 827 | 3.62 | 3.25 | 3.87 | 3.31 | **14.05** |
| 828 | 4.70 | 5.61 | 7.02 | 8.04 | **25.36** |
| 829 | 21.85 | 20.06 | 19.52 | 20.20 | **81.63** |
| 830 | 2.71 | 3.21 | 2.86 | 2.63 | **11.41** |
| 831 | 3.58 | 4.09 | 3.23 | 3.47 | **14.37** |
| **WY - WYOMING - Total** | **87.40** | **87.92** | **86.91** | **89.71** | **351.95** |

**DRUG: 1100 - AMPHETAMINE**

| STATE | 1ST QUARTER | 2ND QUARTER | 3RD QUARTER | 4TH QUARTER | TOTAL GRAMS |
|---|---|---|---|---|---|
| ALABAMA | 138,913.28 | 134,435.90 | 139,860.25 | 144,394.96 | 557,604.40 |
| ALASKA | 8,776.37 | 8,523.53 | 9,218.04 | 9,033.97 | 35,551.91 |
| AMERICAN SAMOA | 1.25 | 0 | 1.25 | 1.25 | 3.75 |
| ARIZONA | 107,068.17 | 96,999.79 | 104,146.34 | 110,051.71 | 418,266.00 |
| ARKANSAS | 57,040.92 | 56,368.21 | 57,988.40 | 61,321.73 | 232,719.27 |
| CALIFORNIA | 330,919.44 | 307,724.58 | 318,691.84 | 331,841.77 | 1,289,177.62 |
| COLORADO | 103,383.61 | 95,069.58 | 104,350.53 | 108,690.40 | 411,494.11 |
| CONNECTICUT | 62,817.84 | 62,771.85 | 64,278.31 | 66,877.82 | 256,745.82 |
| DELAWARE | 21,658.81 | 21,771.61 | 21,811.24 | 22,857.81 | 88,099.48 |
| DISTRICT OF COLUMBIA | 13,391.96 | 11,172.94 | 12,188.38 | 12,028.15 | 48,781.43 |
| FLORIDA | 378,588.81 | 334,488.72 | 371,613.02 | 383,560.88 | 1,468,251.43 |
| GEORGIA | 216,879.29 | 202,161.25 | 214,834.94 | 221,179.33 | 855,054.81 |
| GUAM | 458.80 | 400.33 | 605.21 | 409.16 | 1,873.50 |
| HAWAII | 11,482.97 | 10,094.69 | 11,097.70 | 11,559.24 | 44,234.60 |
| IDAHO | 32,977.75 | 32,530.29 | 34,212.68 | 36,213.55 | 135,934.28 |
| ILLINOIS | 217,089.27 | 209,128.24 | 213,963.66 | 223,632.15 | 863,813.33 |
| INDIANA | 150,900.67 | 145,023.48 | 149,895.04 | 155,391.39 | 601,210.59 |
| IOWA | 56,595.88 | 54,938.91 | 57,441.57 | 59,309.02 | 228,285.38 |
| KANSAS | 69,780.28 | 68,449.76 | 71,268.15 | 74,176.17 | 283,674.35 |
| KENTUCKY | 80,599.95 | 75,952.84 | 78,486.75 | 81,752.42 | 316,791.97 |
| LOUISIANA | 181,712.87 | 173,101.92 | 179,219.74 | 189,521.44 | 723,555.98 |
| MAINE | 27,918.87 | 25,756.25 | 28,221.38 | 28,692.72 | 110,589.22 |
| MARYLAND | 106,957.83 | 101,211.66 | 103,696.43 | 106,855.15 | 418,721.06 |
| MASSACHUSETTS | 194,068.35 | 186,353.92 | 191,984.25 | 198,160.44 | 770,566.96 |
| MICHIGAN | 257,881.27 | 248,610.24 | 257,471.78 | 265,417.53 | 1,029,380.82 |
| MINNESOTA | 119,220.03 | 118,362.87 | 120,122.93 | 126,221.49 | 483,927.32 |
| MISSISSIPPI | 65,427.47 | 62,708.08 | 65,773.81 | 68,867.07 | 262,776.43 |
| MISSOURI | 151,785.72 | 152,917.79 | 167,555.88 | 164,391.48 | 636,650.88 |
| MONTANA | 16,968.49 | 16,780.24 | 17,442.63 | 18,034.25 | 69,225.61 |
| NEBRASKA | 31,275.37 | 30,487.00 | 31,315.27 | 33,509.16 | 126,586.80 |
| NEVADA | 31,317.49 | 28,676.38 | 30,890.05 | 31,933.33 | 122,817.25 |
| NEW HAMPSHIRE | 44,015.90 | 44,543.88 | 45,457.51 | 47,134.87 | 181,152.17 |
| NEW JERSEY | 123,810.17 | 118,799.48 | 122,372.09 | 129,502.66 | 494,484.40 |
| NEW MEXICO | 18,589.03 | 14,388.35 | 17,170.33 | 17,403.44 | 67,551.14 |
| NEW YORK | 279,794.59 | 249,730.54 | 262,787.64 | 276,988.28 | 1,069,301.05 |
| NORTH CAROLINA | 237,825.31 | 228,009.09 | 238,184.17 | 247,329.39 | 951,347.95 |
| NORTH DAKOTA | 10,561.24 | 9,985.92 | 9,957.21 | 10,966.29 | 41,470.66 |
| OHIO | 222,338.66 | 216,275.59 | 221,886.36 | 232,997.60 | 893,498.21 |
| OKLAHOMA | 58,786.14 | 57,232.59 | 59,364.66 | 61,534.64 | 236,918.03 |
| OREGON | 70,862.47 | 71,640.65 | 73,693.95 | 77,788.88 | 293,985.95 |
| PENNSYLVANIA | 229,008.50 | 227,828.06 | 229,107.57 | 234,241.73 | 920,185.86 |
| PUERTO RICO | 8,330.60 | 6,823.61 | 8,508.26 | 8,330.77 | 31,993.24 |
| RHODE ISLAND | 40,274.83 | 39,390.00 | 40,615.32 | 40,987.09 | 161,267.23 |
| SOUTH CAROLINA | 151,958.92 | 146,004.30 | 151,913.29 | 157,981.93 | 607,858.44 |
| SOUTH DAKOTA | 14,906.94 | 14,151.81 | 14,296.48 | 14,198.33 | 57,553.56 |
| TENNESSEE | 142,208.40 | 139,292.03 | 144,326.77 | 150,684.68 | 576,511.88 |
| TEXAS | 502,601.16 | 492,077.50 | 508,174.54 | 527,569.66 | 2,030,422.85 |
| UTAH | 101,386.17 | 102,304.35 | 106,077.15 | 110,331.08 | 420,098.76 |
| VERMONT | 11,501.54 | 10,447.96 | 10,990.64 | 11,600.36 | 44,540.51 |
| VIRGIN ISLANDS | 447.40 | 337.41 | 356.19 | 345.12 | 1,486.11 |
| VIRGINIA | 195,083.74 | 188,121.88 | 193,279.10 | 197,838.79 | 774,323.52 |
| WASHINGTON | 124,971.25 | 123,231.04 | 126,896.00 | 132,004.39 | 507,102.69 |
| WEST VIRGINIA | 24,879.94 | 22,416.16 | 23,426.49 | 24,266.27 | 94,988.86 |
| WISCONSIN | 129,724.86 | 126,919.43 | 129,032.35 | 133,571.04 | 519,247.69 |
| WYOMING | 7,937.45 | 7,303.55 | 7,855.10 | 8,187.23 | 31,283.34 |
| **1100 - AMPHETAMINE - Total** | **5,995,664.29** | **5,730,228.04** | **5,975,376.63** | **6,199,671.49** | **23,900,940.45** |

**DRUG: 1105 - METHAMPHETAMINE**

| STATE | 1ST QUARTER | 2ND QUARTER | 3RD QUARTER | 4TH QUARTER | TOTAL GRAMS |
|---|---|---|---|---|---|

| | | | | | |
|---|---|---|---|---|---|
| ALABAMA | 30.53 | 34.95 | 34.55 | 24.91 | 124.94 |
| ALASKA | 2.01 | 3.21 | 1.21 | 0 | 6.43 |
| ARIZONA | 27.32 | 32.55 | 27.72 | 30.94 | 118.52 |
| ARKANSAS | 1.61 | 3.21 | 2.81 | 3.21 | 10.85 |
| CALIFORNIA | 230.21 | 229.00 | 224.18 | 218.16 | 901.54 |
| COLORADO | 37.76 | 41.38 | 40.58 | 44.99 | 164.72 |
| CONNECTICUT | 15.67 | 22.10 | 22.10 | 22.90 | 82.76 |
| DELAWARE | 5.22 | 5.62 | 6.83 | 3.62 | 21.29 |
| DISTRICT OF COLUMBIA | 4.82 | 6.43 | 5.22 | 10.45 | 26.92 |
| FLORIDA | 51.43 | 51.03 | 52.23 | 52.23 | 206.91 |
| GEORGIA | 28.52 | 24.51 | 25.71 | 24.51 | 103.25 |
| HAWAII | 4.02 | 4.02 | 2.41 | 3.21 | 13.66 |
| IDAHO | 11.25 | 12.46 | 13.26 | 14.06 | 51.02 |
| ILLINOIS | 44.59 | 30.53 | 40.98 | 31.74 | 147.83 |
| INDIANA | 27.72 | 36.16 | 24.11 | 17.68 | 105.66 |
| IOWA | 10.44 | 16.87 | 16.47 | 16.47 | 60.26 |
| KANSAS | 13.26 | 9.64 | 10.44 | 8.84 | 42.18 |
| KENTUCKY | 10.45 | 9.64 | 9.24 | 10.04 | 39.37 |
| LOUISIANA | 11.25 | 12.05 | 16.47 | 14.46 | 54.24 |
| MAINE | 8.44 | 9.64 | 10.45 | 7.63 | 36.16 |
| MARYLAND | 31.74 | 34.15 | 34.55 | 35.36 | 135.80 |
| MASSACHUSETTS | 26.12 | 18.48 | 28.12 | 18.08 | 90.80 |
| MICHIGAN | 18.08 | 13.66 | 13.66 | 14.46 | 59.86 |
| MINNESOTA | 47.81 | 47.01 | 47.81 | 53.43 | 196.06 |
| MISSISSIPPI | 5.22 | 9.24 | 7.23 | 7.23 | 28.92 |
| MISSOURI | 39.37 | 43.79 | 50.22 | 57.04 | 190.42 |
| MONTANA | 0 | 0 | 2.01 | 0 | 2.01 |
| NEBRASKA | 6.83 | 8.84 | 5.62 | 1.21 | 22.50 |
| NEVADA | 6.43 | 10.04 | 8.84 | 8.44 | 33.75 |
| NEW HAMPSHIRE | 7.63 | 5.22 | 6.43 | 5.22 | 24.51 |
| NEW JERSEY | 36.16 | 28.12 | 26.11 | 27.72 | 118.12 |
| NEW MEXICO | 6.03 | 9.24 | 10.85 | 10.45 | 36.56 |
| NEW YORK | 81.15 | 87.18 | 85.98 | 78.74 | 333.06 |
| NORTH CAROLINA | 48.61 | 34.55 | 29.33 | 30.53 | 143.02 |
| NORTH DAKOTA | 3.21 | 6.03 | 4.42 | 2.81 | 16.47 |
| OHIO | 28.52 | 31.34 | 32.54 | 29.73 | 122.13 |
| OKLAHOMA | 6.83 | 12.05 | 12.05 | 12.45 | 43.39 |
| OREGON | 47.41 | 42.99 | 34.55 | 38.57 | 163.51 |
| PENNSYLVANIA | 88.39 | 89.60 | 96.03 | 93.22 | 367.23 |
| RHODE ISLAND | 3.62 | 8.04 | 3.62 | 2.41 | 17.68 |
| SOUTH CAROLINA | 6.03 | 8.84 | 9.24 | 6.03 | 30.13 |
| SOUTH DAKOTA | 5.62 | 4.02 | 8.44 | 8.84 | 26.91 |
| TENNESSEE | 12.86 | 15.27 | 17.28 | 16.07 | 61.47 |
| TEXAS | 92.40 | 77.94 | 94.81 | 91.59 | 356.75 |
| UTAH | 24.10 | 23.30 | 30.13 | 29.33 | 106.85 |
| VERMONT | 3.21 | 1.21 | 1.61 | 1.61 | 7.63 |
| VIRGINIA | 21.29 | 28.12 | 25.71 | 23.70 | 98.84 |
| WASHINGTON | 92.39 | 75.92 | 66.28 | 68.29 | 302.89 |
| WEST VIRGINIA | 3.21 | 1.61 | 3.62 | 3.21 | 11.65 |
| WISCONSIN | 20.49 | 22.10 | 20.89 | 19.29 | 82.77 |
| WYOMING | 5.62 | 3.21 | 5.22 | 5.62 | 19.68 |
| **1105 - METHAMPHETAMINE - Total** | **1,402.92** | **1,396.10** | **1,410.16** | **1,360.72** | **5,569.91** |

**DRUG: 1205 - LISDEXAMFETAMINE**

| STATE | 1ST QUARTER | 2ND QUARTER | 3RD QUARTER | 4TH QUARTER | TOTAL GRAMS |
|---|---|---|---|---|---|
| ALABAMA | 73,984.08 | 64,814.84 | 68,506.58 | 70,751.35 | 278,056.85 |
| ALASKA | 3,107.87 | 2,969.70 | 3,205.56 | 3,298.64 | 12,581.77 |
| ARIZONA | 42,559.14 | 40,061.17 | 41,158.99 | 43,821.71 | 167,601.02 |
| ARKANSAS | 29,358.23 | 24,666.37 | 26,438.83 | 27,251.06 | 107,714.48 |
| CALIFORNIA | 117,544.49 | 107,981.56 | 110,591.11 | 114,695.62 | 450,812.79 |

| | | | | | |
|---|---|---|---|---|---|
| COLORADO | 31,520.32 | 28,866.85 | 31,279.26 | 32,859.78 | 124,526.21 |
| CONNECTICUT | 36,771.78 | 34,838.62 | 35,923.71 | 37,376.48 | 144,910.59 |
| DELAWARE | 13,624.66 | 12,665.59 | 12,825.15 | 13,151.20 | 52,266.60 |
| DISTRICT OF COLUMBIA | 6,209.95 | 5,086.70 | 5,720.30 | 5,683.88 | 22,700.83 |
| FLORIDA | 141,459.91 | 126,692.35 | 127,764.15 | 135,310.66 | 531,227.07 |
| GEORGIA | 97,524.31 | 84,296.23 | 89,714.18 | 92,804.71 | 364,339.43 |
| GUAM | 21.97 | 27.17 | 13.87 | 12.72 | 75.73 |
| HAWAII | 1,961.49 | 1,742.97 | 1,860.90 | 1,945.88 | 7,511.25 |
| IDAHO | 10,849.78 | 10,317.35 | 11,006.45 | 11,656.23 | 43,829.81 |
| ILLINOIS | 76,589.00 | 70,322.40 | 74,431.53 | 77,841.17 | 299,184.09 |
| INDIANA | 83,362.02 | 74,074.27 | 78,761.50 | 81,445.62 | 317,643.40 |
| IOWA | 34,307.92 | 30,930.66 | 32,323.88 | 33,455.23 | 131,017.69 |
| KANSAS | 32,976.56 | 30,412.11 | 32,030.79 | 32,778.27 | 128,197.72 |
| KENTUCKY | 39,812.56 | 35,326.53 | 36,522.05 | 37,544.13 | 149,205.27 |
| LOUISIANA | 91,533.46 | 77,470.60 | 83,760.33 | 87,380.39 | 340,144.79 |
| MAINE | 17,625.11 | 16,772.42 | 18,106.09 | 18,596.90 | 71,100.52 |
| MARYLAND | 60,073.84 | 55,029.92 | 55,407.99 | 58,579.45 | 229,091.20 |
| MASSACHUSETTS | 42,654.53 | 39,140.84 | 40,430.58 | 41,931.29 | 164,157.24 |
| MICHIGAN | 65,758.88 | 57,630.79 | 59,902.72 | 63,817.04 | 247,109.42 |
| MINNESOTA | 34,488.87 | 32,886.95 | 34,237.97 | 36,722.65 | 138,336.44 |
| MISSISSIPPI | 37,753.40 | 29,898.75 | 32,980.03 | 34,431.64 | 135,063.82 |
| MISSOURI | 67,722.10 | 63,536.08 | 65,663.49 | 67,916.92 | 264,838.59 |
| MONTANA | 7,176.53 | 7,089.24 | 7,098.49 | 7,439.57 | 28,803.83 |
| NEBRASKA | 18,573.20 | 16,875.90 | 18,135.00 | 18,913.70 | 72,497.79 |
| NEVADA | 8,358.75 | 7,399.10 | 7,100.80 | 7,195.61 | 30,054.26 |
| NEW HAMPSHIRE | 14,643.27 | 14,210.28 | 14,713.22 | 15,104.60 | 58,671.37 |
| NEW JERSEY | 49,684.80 | 45,871.08 | 45,696.49 | 48,392.17 | 189,644.55 |
| NEW MEXICO | 3,671.51 | 3,674.98 | 3,782.51 | 3,688.86 | 14,817.86 |
| NEW YORK | 93,379.91 | 79,861.61 | 83,374.74 | 89,589.30 | 346,205.56 |
| NORTH CAROLINA | 112,073.36 | 100,768.03 | 106,352.48 | 109,711.24 | 428,905.11 |
| NORTH DAKOTA | 6,570.11 | 6,234.23 | 5,981.02 | 6,778.22 | 25,563.58 |
| OHIO | 96,140.34 | 85,775.59 | 90,231.00 | 93,369.51 | 365,516.44 |
| OKLAHOMA | 39,256.46 | 33,504.36 | 35,820.81 | 37,404.23 | 145,985.86 |
| OREGON | 16,809.99 | 15,264.73 | 16,390.87 | 17,001.34 | 65,466.93 |
| PENNSYLVANIA | 94,326.26 | 87,264.77 | 88,587.47 | 91,717.88 | 361,896.38 |
| PUERTO RICO | 2,196.20 | 1,711.18 | 1,851.08 | 2,010.63 | 7,769.09 |
| RHODE ISLAND | 7,935.00 | 7,395.06 | 7,834.99 | 7,352.85 | 30,517.90 |
| SOUTH CAROLINA | 63,760.73 | 55,343.94 | 57,605.90 | 60,162.71 | 236,873.27 |
| SOUTH DAKOTA | 9,709.77 | 8,534.49 | 8,637.97 | 8,773.25 | 35,655.47 |
| TENNESSEE | 67,393.74 | 58,278.84 | 63,188.64 | 66,197.65 | 255,058.88 |
| TEXAS | 262,676.50 | 224,767.01 | 239,329.35 | 248,455.82 | 975,228.69 |
| UTAH | 23,265.63 | 22,762.11 | 23,347.15 | 24,529.94 | 93,904.83 |
| VERMONT | 7,855.80 | 7,477.15 | 7,628.61 | 7,995.12 | 30,956.68 |
| VIRGIN ISLANDS | 64.75 | 98.28 | 75.15 | 68.22 | 306.39 |
| VIRGINIA | 72,687.98 | 62,749.29 | 66,174.53 | 67,200.66 | 268,812.45 |
| WASHINGTON | 38,889.94 | 35,652.01 | 37,845.32 | 38,744.84 | 151,132.10 |
| WEST VIRGINIA | 19,181.94 | 16,716.92 | 17,095.00 | 17,813.00 | 70,806.84 |
| WISCONSIN | 80,013.92 | 74,290.79 | 77,293.70 | 82,157.14 | 313,755.57 |
| WYOMING | 2,829.80 | 2,563.87 | 2,921.14 | 2,942.53 | 11,257.34 |
| **1205 - LISDEXAMFETAMINE - Total** | **2,510,282.47** | **2,240,594.61** | **2,344,661.42** | **2,443,771.17** | **9,539,309.67** |

**DRUG: 1724 - METHYLPHENIDATE (DL;D;L;ISOMERS)**

| STATE | 1ST QUARTER | 2ND QUARTER | 3RD QUARTER | 4TH QUARTER | TOTAL GRAMS |
|---|---|---|---|---|---|
| ALABAMA | 77,712.65 | 60,307.15 | 65,336.42 | 71,485.05 | 274,841.27 |
| ALASKA | 8,213.75 | 7,093.54 | 7,493.00 | 7,719.89 | 30,520.19 |
| AMERICAN SAMOA | 1.73 | 4.32 | 6.92 | 2.59 | 15.57 |
| ARIZONA | 80,465.43 | 68,109.67 | 70,987.30 | 76,102.07 | 295,664.47 |
| ARKANSAS | 39,371.34 | 31,250.31 | 33,976.92 | 37,063.06 | 141,661.62 |
| CALIFORNIA | 269,470.13 | 230,838.34 | 233,837.75 | 247,632.87 | 981,779.10 |
| COLORADO | 66,720.41 | 57,448.30 | 61,586.42 | 65,301.70 | 251,056.84 |

| | | | | | |
|---|---|---|---|---|---|
| CONNECTICUT | 56,488.00 | 48,867.42 | 49,098.39 | 52,920.75 | 207,374.56 |
| DELAWARE | 18,737.51 | 15,181.09 | 14,854.22 | 16,847.98 | 65,620.81 |
| DISTRICT OF COLUMBIA | 9,178.64 | 6,764.92 | 6,600.07 | 7,193.09 | 29,736.72 |
| FLORIDA | 213,392.92 | 173,960.75 | 178,240.06 | 197,384.04 | 762,977.76 |
| GEORGIA | 169,690.52 | 125,196.48 | 138,014.94 | 150,136.86 | 583,038.80 |
| GUAM | 417.60 | 412.90 | 346.18 | 482.07 | 1,658.75 |
| HAWAII | 10,867.20 | 8,565.04 | 9,187.59 | 9,543.33 | 38,163.16 |
| IDAHO | 30,026.38 | 26,447.61 | 27,085.32 | 29,641.44 | 113,200.75 |
| ILLINOIS | 170,295.93 | 138,818.33 | 145,907.92 | 159,256.21 | 614,278.40 |
| INDIANA | 124,582.71 | 97,982.47 | 107,469.02 | 116,842.93 | 446,877.14 |
| IOWA | 81,698.92 | 69,059.22 | 72,999.72 | 78,659.37 | 302,417.24 |
| KANSAS | 51,811.08 | 44,359.97 | 45,376.48 | 50,721.27 | 192,268.80 |
| KENTUCKY | 79,040.91 | 63,143.59 | 65,633.32 | 69,955.57 | 277,773.39 |
| LOUISIANA | 83,144.88 | 58,945.61 | 65,310.52 | 73,747.11 | 281,148.12 |
| MAINE | 36,600.41 | 33,436.15 | 34,600.14 | 35,533.15 | 140,169.85 |
| MARYLAND | 106,029.40 | 87,883.90 | 86,779.24 | 95,100.16 | 375,792.69 |
| MASSACHUSETTS | 147,341.82 | 123,353.48 | 123,834.08 | 134,895.12 | 529,424.49 |
| MICHIGAN | 176,927.74 | 146,667.10 | 150,172.68 | 165,708.03 | 639,475.55 |
| MINNESOTA | 104,799.72 | 90,880.06 | 91,036.15 | 100,378.88 | 387,094.81 |
| MISSISSIPPI | 45,365.63 | 32,311.22 | 36,818.24 | 41,307.41 | 155,802.50 |
| MISSOURI | 137,242.64 | 120,891.16 | 124,932.22 | 141,138.00 | 524,204.02 |
| MONTANA | 17,581.15 | 15,846.31 | 16,379.06 | 17,458.03 | 67,264.56 |
| NEBRASKA | 32,603.57 | 27,138.59 | 29,614.75 | 32,416.61 | 121,773.51 |
| NEVADA | 15,848.91 | 13,744.76 | 13,750.92 | 14,908.92 | 58,253.51 |
| NEW HAMPSHIRE | 35,038.47 | 32,076.53 | 32,587.55 | 34,649.60 | 134,352.15 |
| NEW JERSEY | 94,877.24 | 81,066.91 | 77,975.47 | 89,646.71 | 343,566.33 |
| NEW MEXICO | 18,385.37 | 15,732.77 | 16,409.74 | 16,421.96 | 66,949.84 |
| NEW YORK | 234,468.84 | 194,162.93 | 191,325.12 | 215,414.57 | 835,371.46 |
| NORTH CAROLINA | 194,761.90 | 156,941.27 | 163,365.00 | 178,000.33 | 693,068.50 |
| NORTH DAKOTA | 16,134.84 | 13,579.49 | 12,958.86 | 15,354.71 | 58,027.91 |
| OHIO | 205,917.54 | 170,381.63 | 177,572.28 | 195,467.45 | 749,338.89 |
| OKLAHOMA | 49,291.23 | 39,438.16 | 43,469.57 | 45,211.90 | 177,410.86 |
| OREGON | 66,697.81 | 59,143.12 | 59,811.22 | 64,237.88 | 249,890.03 |
| PENNSYLVANIA | 228,356.43 | 197,855.63 | 203,184.30 | 217,129.45 | 846,525.81 |
| PUERTO RICO | 13,110.85 | 8,416.74 | 9,813.70 | 9,872.29 | 41,213.58 |
| RHODE ISLAND | 19,331.98 | 16,355.84 | 16,477.89 | 16,419.10 | 68,584.81 |
| SOUTH CAROLINA | 93,802.03 | 73,368.68 | 75,037.95 | 84,563.83 | 326,772.50 |
| SOUTH DAKOTA | 19,387.42 | 16,444.78 | 16,589.38 | 17,580.97 | 70,002.56 |
| TENNESSEE | 86,314.12 | 69,602.62 | 75,589.93 | 82,473.43 | 313,980.10 |
| TEXAS | 350,523.40 | 258,295.68 | 281,179.60 | 321,940.89 | 1,211,939.57 |
| UTAH | 50,605.97 | 46,079.68 | 47,206.44 | 51,748.95 | 195,641.04 |
| VERMONT | 19,444.07 | 17,656.08 | 17,942.62 | 19,308.38 | 74,351.15 |
| VIRGIN ISLANDS | 126.17 | 78.47 | 100.47 | 85.25 | 390.37 |
| VIRGINIA | 138,454.85 | 110,650.38 | 116,130.16 | 122,049.17 | 487,284.55 |
| WASHINGTON | 118,559.42 | 104,058.93 | 104,390.27 | 112,286.02 | 439,294.64 |
| WEST VIRGINIA | 30,481.01 | 25,547.18 | 25,141.93 | 26,957.44 | 108,127.56 |
| WISCONSIN | 108,413.02 | 93,430.83 | 96,049.66 | 103,887.51 | 401,781.02 |
| WYOMING | 7,834.74 | 6,990.44 | 7,100.03 | 7,510.26 | 29,435.46 |
| **1724 - METHYLPHENIDATE (DL;D;L;IS** | **4,661,988.37** | **3,832,264.52** | **3,974,675.12** | **4,345,701.64** | **16,814,629.65** |

**DRUG: 9050 - CODEINE**

| STATE | 1ST QUARTER | 2ND QUARTER | 3RD QUARTER | 4TH QUARTER | TOTAL GRAMS |
|---|---|---|---|---|---|
| ALABAMA | 43,089.65 | 41,641.00 | 40,137.68 | 43,313.83 | 168,182.16 |
| ALASKA | 4,808.13 | 4,179.20 | 4,875.41 | 4,397.22 | 18,259.96 |
| AMERICAN SAMOA | 360.25 | 320.46 | 395.61 | 278.47 | 1,354.79 |
| ARIZONA | 47,215.13 | 42,905.06 | 44,614.94 | 43,669.04 | 178,404.18 |
| ARKANSAS | 48,108.33 | 46,202.27 | 46,521.82 | 45,509.30 | 186,341.73 |
| CALIFORNIA | 305,815.29 | 296,733.11 | 299,620.29 | 297,198.18 | 1,199,366.88 |
| COLORADO | 15,569.21 | 14,747.60 | 15,319.22 | 15,083.39 | 60,719.42 |
| CONNECTICUT | 15,182.75 | 14,548.25 | 14,498.38 | 14,590.89 | 58,820.26 |

| | | | | | |
|---|---|---|---|---|---|
| DELAWARE | 6,179.10 | 5,821.66 | 6,316.42 | 6,308.66 | 24,625.84 |
| DISTRICT OF COLUMBIA | 7,227.78 | 6,370.34 | 6,268.65 | 6,765.93 | 26,632.70 |
| FLORIDA | 129,699.36 | 116,584.06 | 121,816.09 | 127,856.02 | 495,955.54 |
| GEORGIA | 76,776.06 | 73,033.50 | 74,599.49 | 77,944.71 | 302,353.76 |
| GUAM | 919.77 | 876.63 | 897.74 | 769.11 | 3,463.26 |
| HAWAII | 9,016.67 | 8,506.29 | 8,690.72 | 8,691.81 | 34,905.49 |
| IDAHO | 10,431.47 | 10,201.69 | 10,629.50 | 10,450.46 | 41,713.13 |
| ILLINOIS | 114,355.60 | 112,206.31 | 113,221.47 | 115,063.71 | 454,847.09 |
| INDIANA | 31,158.90 | 30,961.66 | 31,939.43 | 32,949.77 | 127,009.76 |
| IOWA | 12,909.09 | 11,818.69 | 12,270.50 | 11,768.46 | 48,766.74 |
| KANSAS | 21,006.12 | 21,544.59 | 21,430.00 | 21,654.90 | 85,635.60 |
| KENTUCKY | 34,609.66 | 33,504.47 | 33,644.74 | 33,665.74 | 135,424.61 |
| LOUISIANA | 30,344.13 | 28,992.56 | 30,255.42 | 30,845.49 | 120,437.60 |
| MAINE | 6,882.49 | 6,599.70 | 6,679.49 | 6,735.26 | 26,896.94 |
| MARYLAND | 42,461.58 | 38,077.93 | 41,148.10 | 41,626.31 | 163,313.92 |
| MASSACHUSETTS | 32,932.79 | 29,589.55 | 29,857.88 | 31,476.80 | 123,857.02 |
| MICHIGAN | 162,137.09 | 150,358.86 | 159,156.71 | 158,970.75 | 630,623.41 |
| MINNESOTA | 23,494.05 | 22,196.88 | 22,695.25 | 23,298.46 | 91,684.63 |
| MISSISSIPPI | 17,721.51 | 17,156.32 | 16,740.17 | 18,791.74 | 70,409.74 |
| MISSOURI | 56,735.16 | 54,210.35 | 53,887.13 | 56,205.27 | 221,037.90 |
| MONTANA | 7,141.64 | 6,696.87 | 6,701.08 | 6,818.72 | 27,358.30 |
| NEBRASKA | 7,709.07 | 7,891.47 | 7,626.35 | 7,778.81 | 31,005.70 |
| NEVADA | 16,057.17 | 15,703.30 | 15,591.27 | 15,345.22 | 62,696.95 |
| NEW HAMPSHIRE | 5,720.71 | 5,875.14 | 5,256.63 | 5,737.10 | 22,589.58 |
| NEW JERSEY | 56,234.18 | 48,967.33 | 55,877.69 | 56,692.47 | 217,771.67 |
| NEW MEXICO | 13,652.71 | 13,232.55 | 13,006.74 | 13,391.78 | 53,283.78 |
| NEW YORK | 104,114.82 | 90,090.35 | 94,223.53 | 96,406.11 | 384,834.80 |
| NORTH CAROLINA | 32,991.28 | 32,280.93 | 30,891.15 | 33,012.42 | 129,175.78 |
| NORTH DAKOTA | 3,780.02 | 3,263.00 | 3,330.78 | 3,664.10 | 14,037.91 |
| OHIO | 56,974.04 | 52,956.64 | 54,770.58 | 54,797.81 | 219,499.06 |
| OKLAHOMA | 68,371.42 | 65,707.25 | 73,756.91 | 72,013.18 | 279,848.77 |
| OREGON | 24,276.75 | 23,801.62 | 24,529.32 | 23,773.89 | 96,381.58 |
| PENNSYLVANIA | 82,477.79 | 78,034.72 | 80,144.62 | 81,978.34 | 322,635.48 |
| PUERTO RICO | 9,299.00 | 6,274.47 | 9,198.44 | 9,003.95 | 33,775.85 |
| RHODE ISLAND | 4,169.89 | 4,066.84 | 3,698.75 | 3,830.91 | 15,766.40 |
| SOUTH CAROLINA | 54,303.90 | 54,605.99 | 50,799.37 | 58,814.65 | 218,523.90 |
| SOUTH DAKOTA | 4,261.24 | 4,091.11 | 4,057.14 | 4,073.04 | 16,482.53 |
| TENNESSEE | 51,150.27 | 53,284.21 | 49,322.72 | 47,041.03 | 200,798.23 |
| TEXAS | 769,470.88 | 707,778.85 | 740,263.11 | 732,952.20 | 2,950,465.05 |
| UTAH | 12,039.91 | 11,749.93 | 12,232.60 | 11,975.70 | 47,998.14 |
| VERMONT | 2,318.06 | 2,055.04 | 2,501.75 | 2,082.42 | 8,957.27 |
| VIRGIN ISLANDS | 644.24 | 537.05 | 690.66 | 539.26 | 2,411.22 |
| VIRGINIA | 37,773.56 | 37,390.60 | 34,809.14 | 37,080.24 | 147,053.55 |
| WASHINGTON | 42,311.00 | 41,100.27 | 41,750.34 | 40,165.95 | 165,327.56 |
| WEST VIRGINIA | 34,328.68 | 33,055.88 | 34,464.29 | 34,083.16 | 135,932.01 |
| WISCONSIN | 34,235.11 | 32,681.92 | 33,679.83 | 32,337.66 | 132,934.53 |
| WYOMING | 2,837.77 | 2,762.98 | 2,868.34 | 2,904.58 | 11,373.68 |
| **9050 - CODEINE - Total** | **2,845,792.25** | **2,675,825.30** | **2,754,171.42** | **2,774,174.38** | **11,049,963.34** |

**DRUG: 9064 - BUPRENORPHINE**

| STATE | 1ST QUARTER | 2ND QUARTER | 3RD QUARTER | 4TH QUARTER | TOTAL GRAMS |
|---|---|---|---|---|---|
| ALABAMA | 33,975.14 | 35,737.30 | 36,715.00 | 37,255.07 | 143,682.51 |
| ALASKA | 2,687.85 | 2,663.53 | 2,813.89 | 2,958.94 | 11,124.21 |
| ARIZONA | 15,893.16 | 15,407.16 | 15,971.14 | 16,505.78 | 63,777.24 |
| ARKANSAS | 9,909.01 | 10,645.60 | 11,262.67 | 11,513.63 | 43,330.91 |
| CALIFORNIA | 59,322.45 | 61,422.71 | 65,665.16 | 71,469.89 | 257,880.21 |
| COLORADO | 12,305.85 | 11,787.94 | 12,191.77 | 12,666.06 | 48,951.62 |
| CONNECTICUT | 19,160.79 | 19,303.56 | 18,923.07 | 19,453.06 | 76,840.48 |
| DELAWARE | 4,639.19 | 4,716.66 | 4,799.58 | 4,771.24 | 18,926.66 |
| DISTRICT OF COLUMBIA | 3,637.37 | 3,607.89 | 3,929.87 | 4,000.11 | 15,175.24 |

| | | | | | |
|---|---|---|---|---|---|
| FLORIDA | 63,260.47 | 63,770.88 | 65,083.61 | 67,463.68 | 259,578.64 |
| GEORGIA | 20,797.85 | 21,786.05 | 21,985.93 | 22,354.48 | 86,924.31 |
| GUAM | 0.01 | 0 | 2.99 | 0.48 | 3.47 |
| HAWAII | 2,347.18 | 2,306.74 | 2,520.22 | 2,503.06 | 9,677.20 |
| IDAHO | 5,466.73 | 5,851.54 | 5,925.07 | 6,039.82 | 23,283.15 |
| ILLINOIS | 21,382.14 | 21,676.64 | 22,287.89 | 23,133.16 | 88,479.83 |
| INDIANA | 34,268.41 | 35,946.33 | 37,398.72 | 38,632.83 | 146,246.28 |
| IOWA | 3,027.86 | 2,944.40 | 3,068.91 | 3,161.09 | 12,202.26 |
| KANSAS | 3,439.72 | 3,502.72 | 3,748.25 | 3,907.19 | 14,597.88 |
| KENTUCKY | 48,527.73 | 49,703.12 | 53,370.40 | 54,353.78 | 205,955.04 |
| LOUISIANA | 25,882.27 | 27,697.63 | 28,697.76 | 28,907.54 | 111,185.20 |
| MAINE | 15,207.53 | 15,201.26 | 15,742.69 | 16,069.80 | 62,221.29 |
| MARYLAND | 36,141.08 | 35,290.28 | 36,650.75 | 38,137.87 | 146,219.99 |
| MASSACHUSETTS | 51,123.74 | 50,943.53 | 50,331.42 | 51,598.85 | 203,997.54 |
| MICHIGAN | 36,721.68 | 37,596.92 | 38,972.98 | 40,399.20 | 153,690.78 |
| MINNESOTA | 12,360.78 | 12,273.37 | 12,706.26 | 13,278.01 | 50,618.41 |
| MISSISSIPPI | 12,327.92 | 12,813.16 | 13,032.02 | 13,287.94 | 51,461.04 |
| MISSOURI | 19,340.84 | 19,802.37 | 20,369.60 | 21,335.10 | 80,847.92 |
| MONTANA | 4,342.76 | 4,484.77 | 4,650.04 | 4,858.79 | 18,336.37 |
| NEBRASKA | 2,121.64 | 2,149.20 | 2,234.06 | 2,397.42 | 8,902.33 |
| NEVADA | 4,905.44 | 4,713.37 | 5,063.22 | 5,293.08 | 19,975.11 |
| NEW HAMPSHIRE | 11,131.21 | 10,981.54 | 11,280.80 | 11,711.83 | 45,105.37 |
| NEW JERSEY | 35,642.68 | 34,906.13 | 34,727.87 | 36,131.25 | 141,407.92 |
| NEW MEXICO | 10,108.79 | 10,256.74 | 10,273.13 | 10,166.87 | 40,805.54 |
| NEW YORK | 77,598.23 | 78,159.57 | 79,161.05 | 80,566.31 | 315,485.16 |
| NORTH CAROLINA | 55,357.18 | 56,210.40 | 58,510.26 | 59,785.61 | 229,863.45 |
| NORTH DAKOTA | 1,826.67 | 1,725.49 | 1,815.68 | 1,821.29 | 7,189.14 |
| OHIO | 65,113.70 | 65,560.84 | 67,213.60 | 68,197.61 | 266,085.76 |
| OKLAHOMA | 18,963.98 | 19,354.23 | 20,416.80 | 20,718.99 | 79,453.99 |
| OREGON | 17,702.87 | 17,499.17 | 18,162.29 | 17,844.17 | 71,208.49 |
| PENNSYLVANIA | 89,328.27 | 91,319.68 | 92,451.36 | 95,901.23 | 369,000.54 |
| PUERTO RICO | 2,332.59 | 1,949.95 | 2,245.15 | 2,308.75 | 8,836.44 |
| RHODE ISLAND | 9,249.89 | 9,395.60 | 8,727.50 | 9,530.18 | 36,903.18 |
| SOUTH CAROLINA | 16,990.43 | 18,076.80 | 18,120.02 | 19,287.30 | 72,474.56 |
| SOUTH DAKOTA | 1,211.52 | 1,263.40 | 1,357.62 | 1,401.52 | 5,234.06 |
| TENNESSEE | 45,775.51 | 48,977.21 | 50,425.13 | 50,573.48 | 195,751.34 |
| TEXAS | 33,316.90 | 33,269.17 | 34,408.50 | 35,693.23 | 136,687.79 |
| UTAH | 12,225.11 | 12,036.96 | 12,219.63 | 12,507.31 | 48,989.01 |
| VERMONT | 8,306.21 | 8,068.36 | 8,912.38 | 9,155.98 | 34,442.93 |
| VIRGIN ISLANDS | 28.43 | 13.14 | 29.81 | 31.94 | 103.33 |
| VIRGINIA | 28,554.98 | 29,415.62 | 29,951.80 | 30,812.56 | 118,734.95 |
| WASHINGTON | 35,311.51 | 35,234.59 | 35,646.34 | 36,124.56 | 142,317.00 |
| WEST VIRGINIA | 21,133.96 | 22,119.25 | 23,430.45 | 23,865.89 | 90,549.56 |
| WISCONSIN | 15,673.63 | 15,849.75 | 16,680.25 | 17,255.36 | 65,458.99 |
| WYOMING | 1,583.97 | 1,552.11 | 1,644.04 | 1,602.28 | 6,382.40 |
| **9064 - BUPRENORPHINE - Total** | **1,198,992.85** | **1,218,942.36** | **1,253,926.36** | **1,290,702.45** | **4,962,564.02** |

**DRUG: 9143 - OXYCODONE**

| STATE | 1ST QUARTER | 2ND QUARTER | 3RD QUARTER | 4TH QUARTER | TOTAL GRAMS |
|---|---|---|---|---|---|
| ALABAMA | 163,861.22 | 156,016.14 | 162,188.95 | 160,307.32 | 642,373.62 |
| ALASKA | 19,695.59 | 18,871.76 | 19,611.63 | 19,420.23 | 77,599.20 |
| AMERICAN SAMOA | 20.62 | 14.34 | 7.17 | 8.97 | 51.10 |
| ARIZONA | 268,760.73 | 260,309.26 | 261,017.39 | 262,759.27 | 1,052,846.65 |
| ARKANSAS | 79,654.28 | 76,033.80 | 76,415.74 | 75,853.81 | 307,957.64 |
| CALIFORNIA | 551,181.82 | 523,429.08 | 534,853.07 | 526,715.01 | 2,136,178.98 |
| COLORADO | 155,811.48 | 149,547.46 | 153,796.25 | 152,668.87 | 611,824.06 |
| CONNECTICUT | 116,788.24 | 111,826.14 | 115,451.16 | 114,609.27 | 458,674.80 |
| DELAWARE | 45,112.05 | 43,964.30 | 44,870.62 | 44,927.53 | 178,874.50 |
| DISTRICT OF COLUMBIA | 10,797.62 | 9,268.95 | 9,892.37 | 9,887.24 | 39,846.18 |
| FLORIDA | 849,652.64 | 821,613.32 | 852,896.15 | 856,584.90 | 3,380,747.01 |

| | | | | | |
|---|---|---|---|---|---|
| GEORGIA | 344,510.47 | 328,950.02 | 340,665.07 | 338,666.54 | 1,352,792.10 |
| GUAM | 672.29 | 609.08 | 751.00 | 708.50 | 2,740.87 |
| HAWAII | 28,239.19 | 26,370.16 | 26,261.46 | 24,814.97 | 105,685.78 |
| IDAHO | 45,660.61 | 43,697.23 | 45,399.03 | 44,971.95 | 179,728.81 |
| ILLINOIS | 104,985.04 | 99,250.52 | 102,715.68 | 103,212.35 | 410,163.59 |
| INDIANA | 169,062.67 | 163,284.66 | 168,448.77 | 169,705.60 | 670,501.70 |
| IOWA | 35,115.06 | 33,185.56 | 34,990.03 | 35,037.12 | 138,327.77 |
| KANSAS | 86,671.05 | 83,129.73 | 86,368.85 | 85,721.08 | 341,890.72 |
| KENTUCKY | 162,135.14 | 152,088.24 | 158,679.16 | 155,894.94 | 628,797.49 |
| LOUISIANA | 138,928.22 | 131,875.41 | 136,665.68 | 137,011.46 | 544,480.77 |
| MAINE | 36,604.45 | 35,536.35 | 37,186.81 | 36,745.60 | 146,073.21 |
| MARYLAND | 231,606.11 | 221,758.90 | 225,809.20 | 223,732.67 | 902,906.87 |
| MASSACHUSETTS | 172,445.93 | 162,080.81 | 170,758.69 | 168,381.18 | 673,666.61 |
| MICHIGAN | 188,576.62 | 177,185.58 | 182,311.12 | 183,464.92 | 731,538.25 |
| MINNESOTA | 91,908.18 | 86,743.51 | 91,540.20 | 90,740.23 | 360,932.12 |
| MISSISSIPPI | 77,069.73 | 73,632.22 | 77,007.47 | 77,758.70 | 305,468.13 |
| MISSOURI | 177,464.07 | 170,166.97 | 175,697.99 | 172,941.10 | 696,270.13 |
| MONTANA | 26,627.13 | 25,674.61 | 26,911.93 | 26,748.88 | 105,962.55 |
| NEBRASKA | 28,231.39 | 26,291.98 | 27,404.00 | 27,299.99 | 109,227.36 |
| NEVADA | 130,880.68 | 128,034.45 | 129,731.04 | 128,821.07 | 517,467.24 |
| NEW HAMPSHIRE | 42,026.61 | 39,848.19 | 41,896.56 | 41,943.63 | 165,714.99 |
| NEW JERSEY | 336,690.17 | 319,951.95 | 331,233.89 | 329,668.00 | 1,317,544.01 |
| NEW MEXICO | 63,901.26 | 60,551.02 | 61,524.64 | 60,355.14 | 246,332.07 |
| NEW YORK | 488,159.02 | 457,731.11 | 475,872.75 | 473,052.67 | 1,894,815.55 |
| NORTH CAROLINA | 424,533.59 | 409,100.93 | 423,184.77 | 421,045.68 | 1,677,864.97 |
| NORTH DAKOTA | 10,058.75 | 9,706.62 | 9,830.06 | 10,315.60 | 39,911.02 |
| OHIO | 306,745.45 | 291,654.33 | 306,798.41 | 306,360.25 | 1,211,558.44 |
| OKLAHOMA | 135,180.60 | 131,573.42 | 132,854.17 | 131,885.58 | 531,493.77 |
| OREGON | 119,099.66 | 115,073.98 | 120,896.89 | 119,931.17 | 475,001.70 |
| PENNSYLVANIA | 461,027.84 | 449,381.25 | 463,456.14 | 447,686.20 | 1,821,551.43 |
| PUERTO RICO | 11,447.09 | 9,127.49 | 11,313.96 | 10,742.04 | 42,630.59 |
| RHODE ISLAND | 28,398.35 | 26,550.77 | 28,312.24 | 25,968.96 | 109,230.32 |
| SOUTH CAROLINA | 159,510.27 | 152,105.35 | 157,296.80 | 158,615.35 | 627,527.78 |
| SOUTH DAKOTA | 10,453.79 | 9,493.98 | 10,194.15 | 10,023.99 | 40,165.91 |
| TENNESSEE | 318,051.32 | 313,938.44 | 319,165.89 | 310,194.28 | 1,261,349.93 |
| TEXAS | 238,776.06 | 237,195.76 | 242,515.64 | 228,447.39 | 946,934.86 |
| UTAH | 127,790.26 | 126,196.61 | 127,036.89 | 124,862.87 | 505,886.63 |
| VERMONT | 15,780.56 | 15,173.37 | 15,621.33 | 15,436.11 | 62,011.36 |
| VIRGIN ISLANDS | 431.22 | 409.25 | 374.06 | 340.00 | 1,554.53 |
| VIRGINIA | 216,639.31 | 206,765.97 | 210,805.73 | 206,147.72 | 840,358.73 |
| WASHINGTON | 202,369.80 | 195,171.01 | 202,273.24 | 199,567.03 | 799,381.08 |
| WEST VIRGINIA | 49,576.70 | 47,101.48 | 48,585.18 | 47,182.39 | 192,445.76 |
| WISCONSIN | 118,490.82 | 111,776.90 | 116,385.34 | 115,267.31 | 461,920.37 |
| WYOMING | 19,050.56 | 18,122.38 | 18,561.10 | 18,489.01 | 74,223.05 |
| **9143 - OXYCODONE - Total** | **8,442,919.41** | **8,094,142.10** | **8,352,293.54** | **8,269,649.65** | **33,159,004.69** |

**DRUG: 9150 - HYDROMORPHONE**

| STATE | 1ST QUARTER | 2ND QUARTER | 3RD QUARTER | 4TH QUARTER | TOTAL GRAMS |
|---|---|---|---|---|---|
| ALABAMA | 2,246.08 | 2,178.47 | 2,336.67 | 2,334.94 | 9,096.16 |
| ALASKA | 526.30 | 517.99 | 562.47 | 502.73 | 2,109.49 |
| AMERICAN SAMOA | 0.53 | 0.53 | 0.18 | 0.44 | 1.68 |
| ARIZONA | 5,386.03 | 5,357.41 | 5,475.13 | 5,338.77 | 21,557.34 |
| ARKANSAS | 1,884.37 | 1,820.07 | 1,761.02 | 1,797.10 | 7,262.57 |
| CALIFORNIA | 27,563.99 | 26,536.57 | 26,325.41 | 25,381.01 | 105,806.98 |
| COLORADO | 5,905.07 | 5,835.41 | 5,798.01 | 5,227.46 | 22,765.94 |
| CONNECTICUT | 4,990.33 | 4,552.84 | 4,306.73 | 4,266.20 | 18,116.11 |
| DELAWARE | 1,265.00 | 1,214.55 | 1,178.90 | 1,144.26 | 4,802.71 |
| DISTRICT OF COLUMBIA | 446.31 | 363.96 | 379.32 | 381.28 | 1,570.87 |
| FLORIDA | 34,298.37 | 40,697.97 | 28,787.52 | 27,989.64 | 131,773.51 |
| GEORGIA | 6,267.70 | 6,227.08 | 5,551.83 | 5,467.90 | 23,514.51 |

| | | | | | |
|---|---|---|---|---|---|
| GUAM | 28.81 | 31.78 | 28.73 | 17.12 | 106.44 |
| HAWAII | 559.66 | 526.62 | 483.98 | 459.36 | 2,029.61 |
| IDAHO | 1,467.43 | 1,437.15 | 1,460.46 | 1,386.51 | 5,751.55 |
| ILLINOIS | 7,851.79 | 7,637.95 | 7,092.41 | 6,897.18 | 29,479.33 |
| INDIANA | 5,236.95 | 5,566.22 | 5,084.40 | 4,323.29 | 20,210.86 |
| IOWA | 2,045.41 | 1,908.33 | 1,889.26 | 1,912.81 | 7,755.82 |
| KANSAS | 2,681.87 | 2,497.59 | 2,478.02 | 2,450.06 | 10,107.53 |
| KENTUCKY | 2,480.24 | 2,241.61 | 2,192.71 | 2,238.84 | 9,153.39 |
| LOUISIANA | 2,392.79 | 1,927.24 | 2,219.28 | 2,174.57 | 8,713.88 |
| MAINE | 1,603.34 | 1,646.13 | 1,582.01 | 1,691.59 | 6,523.07 |
| MARYLAND | 6,075.78 | 5,805.93 | 5,493.39 | 5,331.35 | 22,706.44 |
| MASSACHUSETTS | 6,500.63 | 7,372.03 | 6,003.67 | 6,564.19 | 26,440.52 |
| MICHIGAN | 6,546.42 | 6,208.78 | 5,944.06 | 5,432.92 | 24,132.18 |
| MINNESOTA | 4,809.99 | 4,839.14 | 4,746.60 | 4,670.59 | 19,066.30 |
| MISSISSIPPI | 4,011.13 | 5,783.13 | 5,772.14 | 5,753.14 | 21,319.53 |
| MISSOURI | 3,900.69 | 3,859.12 | 3,504.50 | 3,740.98 | 15,005.29 |
| MONTANA | 1,436.22 | 1,419.21 | 1,449.80 | 1,345.51 | 5,650.73 |
| NEBRASKA | 914.10 | 976.79 | 934.11 | 1,019.78 | 3,844.78 |
| NEVADA | 2,314.26 | 2,413.68 | 2,356.63 | 2,293.76 | 9,378.32 |
| NEW HAMPSHIRE | 1,610.20 | 1,336.32 | 1,439.63 | 1,415.28 | 5,801.43 |
| NEW JERSEY | 7,138.29 | 7,568.71 | 6,488.22 | 6,560.59 | 27,755.80 |
| NEW MEXICO | 1,284.76 | 990.26 | 980.30 | 1,055.65 | 4,310.97 |
| NEW YORK | 16,419.63 | 20,152.93 | 13,490.84 | 13,900.32 | 63,963.71 |
| NORTH CAROLINA | 9,453.32 | 8,795.99 | 8,892.66 | 8,335.58 | 35,477.56 |
| NORTH DAKOTA | 587.99 | 460.48 | 431.37 | 416.61 | 1,896.45 |
| OHIO | 7,356.09 | 6,664.11 | 6,865.84 | 6,774.50 | 27,660.54 |
| OKLAHOMA | 2,924.73 | 2,677.19 | 2,706.47 | 2,729.58 | 11,037.97 |
| OREGON | 3,966.07 | 3,866.17 | 3,974.15 | 3,815.28 | 15,621.67 |
| PENNSYLVANIA | 10,670.90 | 12,531.85 | 10,536.30 | 10,156.82 | 43,895.86 |
| PUERTO RICO | 47.30 | 41.49 | 25.62 | 32.98 | 147.40 |
| RHODE ISLAND | 988.61 | 888.28 | 876.71 | 807.39 | 3,560.99 |
| SOUTH CAROLINA | 5,529.80 | 5,861.92 | 6,796.37 | 5,233.61 | 23,421.69 |
| SOUTH DAKOTA | 405.75 | 483.80 | 444.99 | 394.52 | 1,729.06 |
| TENNESSEE | 5,083.91 | 6,265.23 | 3,788.12 | 4,207.15 | 19,344.41 |
| TEXAS | 19,132.03 | 17,878.33 | 19,071.14 | 17,843.57 | 73,925.07 |
| UTAH | 2,737.14 | 2,419.82 | 2,360.69 | 2,330.18 | 9,847.83 |
| VERMONT | 1,046.68 | 927.99 | 930.24 | 871.77 | 3,776.68 |
| VIRGIN ISLANDS | 17.42 | 10.52 | 18.08 | 16.98 | 62.99 |
| VIRGINIA | 8,360.44 | 7,679.64 | 7,820.22 | 7,782.38 | 31,642.67 |
| WASHINGTON | 7,989.69 | 7,888.71 | 8,668.94 | 8,052.48 | 32,599.82 |
| WEST VIRGINIA | 1,022.95 | 947.63 | 992.25 | 932.82 | 3,895.65 |
| WISCONSIN | 3,909.17 | 3,344.37 | 3,298.57 | 2,922.07 | 13,474.18 |
| WYOMING | 498.71 | 589.42 | 546.58 | 515.70 | 2,150.41 |
| **9150 - HYDROMORPHONE - Total** | **271,819.15** | **279,672.41** | **254,623.64** | **246,639.09** | **1,052,754.29** |

**DRUG: 9193 - HYDROCODONE**

| STATE | 1ST QUARTER | 2ND QUARTER | 3RD QUARTER | 4TH QUARTER | TOTAL GRAMS |
|---|---|---|---|---|---|
| ALABAMA | 181,957.55 | 171,303.17 | 176,392.46 | 174,810.69 | 704,463.87 |
| ALASKA | 7,431.60 | 7,010.30 | 7,634.84 | 7,403.95 | 29,480.70 |
| AMERICAN SAMOA | 1.82 | 3.63 | 4.84 | 0 | 10.29 |
| ARIZONA | 72,844.89 | 69,935.74 | 70,789.24 | 71,395.16 | 284,965.03 |
| ARKANSAS | 98,666.90 | 94,820.61 | 97,774.11 | 98,275.73 | 389,537.35 |
| CALIFORNIA | 626,760.44 | 598,544.57 | 611,397.50 | 606,040.19 | 2,442,742.71 |
| COLORADO | 44,730.22 | 42,401.37 | 43,785.14 | 43,723.39 | 174,640.12 |
| CONNECTICUT | 13,843.06 | 12,719.15 | 13,378.14 | 12,956.23 | 52,896.58 |
| DELAWARE | 4,059.16 | 3,831.49 | 4,092.90 | 4,172.84 | 16,156.38 |
| DISTRICT OF COLUMBIA | 891.40 | 682.21 | 780.13 | 699.79 | 3,053.53 |
| FLORIDA | 261,087.28 | 243,358.40 | 248,899.54 | 247,805.11 | 1,001,150.34 |
| GEORGIA | 181,577.60 | 169,005.49 | 176,352.55 | 174,164.86 | 701,100.50 |
| GUAM | 103.05 | 88.09 | 108.52 | 111.39 | 411.05 |

| | | | | | |
|---|---|---|---|---|---|
| HAWAII | 12,203.65 | 11,758.26 | 11,289.60 | 11,175.07 | 46,426.58 |
| IDAHO | 43,083.20 | 42,310.34 | 43,137.64 | 43,400.14 | 171,931.31 |
| ILLINOIS | 211,106.51 | 196,960.59 | 208,028.68 | 205,845.72 | 821,941.50 |
| INDIANA | 167,572.24 | 159,250.97 | 165,497.80 | 164,941.79 | 657,262.81 |
| IOWA | 37,799.33 | 35,585.55 | 37,218.48 | 37,265.94 | 147,869.30 |
| KANSAS | 58,294.09 | 55,819.69 | 57,683.70 | 56,787.97 | 228,585.45 |
| KENTUCKY | 162,433.38 | 155,360.44 | 160,965.97 | 159,422.02 | 638,181.81 |
| LOUISIANA | 116,527.59 | 108,708.91 | 113,224.68 | 111,913.78 | 450,374.96 |
| MAINE | 12,609.69 | 12,131.00 | 12,525.92 | 12,435.28 | 49,701.88 |
| MARYLAND | 24,372.82 | 22,012.34 | 23,551.05 | 22,903.33 | 92,839.54 |
| MASSACHUSETTS | 21,592.15 | 19,548.95 | 20,724.45 | 20,000.43 | 81,865.98 |
| MICHIGAN | 241,957.37 | 229,852.72 | 238,444.37 | 237,836.43 | 948,090.89 |
| MINNESOTA | 28,756.30 | 27,171.98 | 28,694.76 | 28,445.78 | 113,068.82 |
| MISSISSIPPI | 74,545.51 | 70,357.22 | 73,233.65 | 73,286.45 | 291,422.83 |
| MISSOURI | 123,188.40 | 114,768.01 | 120,116.06 | 118,828.24 | 476,900.71 |
| MONTANA | 19,213.00 | 18,624.27 | 18,993.96 | 18,896.59 | 75,727.82 |
| NEBRASKA | 21,895.07 | 20,410.41 | 21,322.35 | 20,991.36 | 84,619.19 |
| NEVADA | 64,899.14 | 63,708.72 | 65,177.46 | 63,081.32 | 256,866.63 |
| NEW HAMPSHIRE | 6,792.94 | 6,193.85 | 6,515.19 | 6,556.37 | 26,058.35 |
| NEW JERSEY | 24,673.01 | 21,951.28 | 23,245.44 | 23,029.50 | 92,899.23 |
| NEW MEXICO | 25,653.08 | 24,375.13 | 25,144.51 | 24,926.49 | 100,099.20 |
| NEW YORK | 119,954.27 | 112,286.20 | 115,458.76 | 113,809.81 | 461,509.04 |
| NORTH CAROLINA | 139,424.82 | 127,986.31 | 131,646.05 | 132,031.14 | 531,088.32 |
| NORTH DAKOTA | 7,140.67 | 6,558.66 | 6,591.69 | 6,887.15 | 27,178.17 |
| OHIO | 120,297.26 | 114,619.85 | 120,321.86 | 119,591.30 | 474,830.27 |
| OKLAHOMA | 124,976.79 | 120,301.25 | 123,912.60 | 122,369.92 | 491,560.56 |
| OREGON | 68,270.10 | 66,138.20 | 67,988.50 | 67,368.96 | 269,765.75 |
| PENNSYLVANIA | 104,844.13 | 99,169.77 | 102,327.61 | 101,032.83 | 407,374.34 |
| PUERTO RICO | 384.66 | 233.06 | 277.33 | 263.84 | 1,158.90 |
| RHODE ISLAND | 7,117.89 | 6,785.16 | 7,127.67 | 6,749.46 | 27,780.18 |
| SOUTH CAROLINA | 94,967.78 | 87,574.46 | 90,809.17 | 91,293.44 | 364,644.86 |
| SOUTH DAKOTA | 9,550.42 | 8,491.70 | 9,000.24 | 8,862.03 | 35,904.38 |
| TENNESSEE | 172,647.48 | 163,521.98 | 167,247.19 | 167,602.22 | 671,018.86 |
| TEXAS | 481,739.26 | 459,172.40 | 472,361.04 | 465,558.29 | 1,878,831.00 |
| UTAH | 43,950.07 | 42,531.06 | 42,689.07 | 42,319.09 | 171,489.29 |
| VERMONT | 3,559.23 | 3,397.60 | 3,441.64 | 3,416.70 | 13,815.17 |
| VIRGIN ISLANDS | 250.73 | 138.69 | 161.69 | 151.27 | 702.39 |
| VIRGINIA | 73,643.77 | 67,202.64 | 69,564.74 | 68,409.58 | 278,820.72 |
| WASHINGTON | 92,786.62 | 88,423.17 | 91,593.97 | 90,422.34 | 363,226.10 |
| WEST VIRGINIA | 35,328.32 | 33,770.38 | 34,239.73 | 33,271.98 | 136,610.40 |
| WISCONSIN | 55,829.13 | 52,515.61 | 55,579.75 | 54,607.87 | 218,532.35 |
| WYOMING | 8,070.20 | 7,920.53 | 8,026.31 | 7,970.49 | 31,987.54 |
| **9193 - HYDROCODONE - Total** | **4,757,857.03** | **4,499,303.55** | **4,646,492.22** | **4,607,519.05** | **18,511,171.84** |

**DRUG: 9230 - MEPERIDINE (PETHIDINE)**

| STATE | 1ST QUARTER | 2ND QUARTER | 3RD QUARTER | 4TH QUARTER | TOTAL GRAMS |
|---|---|---|---|---|---|
| ALABAMA | 1,426.10 | 1,539.61 | 2,012.62 | 1,421.74 | 6,400.08 |
| ALASKA | 105.89 | 107.74 | 118.42 | 155.24 | 487.28 |
| ARIZONA | 1,391.65 | 1,280.34 | 1,721.74 | 1,719.16 | 6,112.90 |
| ARKANSAS | 1,253.43 | 1,138.83 | 1,415.21 | 1,266.29 | 5,073.76 |
| CALIFORNIA | 5,463.00 | 4,353.25 | 8,133.82 | 6,313.91 | 24,263.98 |
| COLORADO | 166.02 | 198.48 | 344.42 | 269.40 | 978.32 |
| CONNECTICUT | 29.63 | 25.16 | 63.73 | 60.24 | 178.77 |
| DELAWARE | 46.19 | 19.39 | 86.61 | 15.69 | 167.87 |
| DISTRICT OF COLUMBIA | 72.99 | 31.05 | 68.63 | 91.51 | 264.17 |
| FLORIDA | 1,837.78 | 1,904.66 | 2,358.50 | 1,941.38 | 8,042.31 |
| GEORGIA | 1,272.61 | 1,269.56 | 1,836.58 | 1,521.09 | 5,899.84 |
| GUAM | 11.44 | 67.54 | 39.22 | 26.15 | 144.34 |
| HAWAII | 51.96 | 51.31 | 122.66 | 70.05 | 295.98 |
| IDAHO | 188.14 | 212.86 | 154.91 | 148.81 | 704.72 |

| | | | | | |
|---|---|---|---|---|---|
| ILLINOIS | 410.26 | 343.92 | 617.78 | 544.69 | 1,916.65 |
| INDIANA | 688.81 | 466.14 | 937.52 | 1,219.77 | 3,312.24 |
| IOWA | 592.18 | 320.71 | 758.53 | 667.46 | 2,338.89 |
| KANSAS | 295.55 | 262.32 | 445.66 | 324.52 | 1,328.06 |
| KENTUCKY | 449.26 | 544.58 | 671.49 | 501.22 | 2,166.55 |
| LOUISIANA | 1,125.22 | 1,315.42 | 1,507.59 | 1,273.15 | 5,221.37 |
| MAINE | 85.30 | 60.46 | 25.16 | 69.50 | 240.43 |
| MARYLAND | 442.61 | 351.87 | 467.45 | 436.08 | 1,698.01 |
| MASSACHUSETTS | 317.88 | 212.10 | 394.46 | 280.41 | 1,204.85 |
| MICHIGAN | 353.07 | 395.77 | 677.05 | 472.14 | 1,898.02 |
| MINNESOTA | 138.68 | 117.22 | 146.41 | 109.59 | 511.90 |
| MISSISSIPPI | 1,055.71 | 1,055.28 | 1,292.00 | 1,124.89 | 4,527.88 |
| MISSOURI | 541.86 | 555.36 | 717.14 | 543.92 | 2,358.28 |
| MONTANA | 107.09 | 61.99 | 116.02 | 88.57 | 373.66 |
| NEBRASKA | 105.78 | 100.55 | 186.94 | 140.09 | 533.36 |
| NEVADA | 412.87 | 336.51 | 579.22 | 514.95 | 1,843.55 |
| NEW HAMPSHIRE | 41.29 | 47.39 | 28.98 | 26.58 | 144.23 |
| NEW JERSEY | 258.94 | 185.85 | 310.04 | 344.24 | 1,099.07 |
| NEW MEXICO | 162.10 | 186.07 | 208.29 | 190.97 | 747.42 |
| NEW YORK | 610.05 | 541.96 | 909.30 | 832.17 | 2,893.49 |
| NORTH CAROLINA | 558.96 | 564.84 | 787.62 | 773.57 | 2,684.98 |
| NORTH DAKOTA | 42.27 | 24.84 | 37.80 | 36.82 | 141.73 |
| OHIO | 969.65 | 788.38 | 1,742.13 | 1,296.68 | 4,796.84 |
| OKLAHOMA | 639.46 | 520.29 | 942.85 | 830.76 | 2,933.36 |
| OREGON | 248.05 | 252.41 | 388.69 | 385.53 | 1,274.68 |
| PENNSYLVANIA | 507.54 | 568.98 | 746.98 | 631.95 | 2,455.45 |
| PUERTO RICO | 1,612.28 | 1,337.97 | 3,744.94 | 2,153.59 | 8,848.78 |
| RHODE ISLAND | 50.11 | 22.99 | 41.83 | 63.40 | 178.33 |
| SOUTH CAROLINA | 485.32 | 431.50 | 567.78 | 503.84 | 1,988.44 |
| SOUTH DAKOTA | 88.68 | 70.05 | 98.04 | 75.39 | 332.16 |
| TENNESSEE | 1,041.77 | 943.83 | 1,327.40 | 1,283.17 | 4,596.18 |
| TEXAS | 3,488.61 | 3,118.12 | 4,926.48 | 3,613.02 | 15,146.23 |
| UTAH | 523.66 | 710.82 | 695.89 | 1,012.79 | 2,943.16 |
| VERMONT | 135.63 | 114.17 | 223.65 | 201.97 | 675.41 |
| VIRGIN ISLANDS | 2.18 | 6.54 | 16.89 | 22.88 | 48.48 |
| VIRGINIA | 747.75 | 561.79 | 852.11 | 741.32 | 2,902.97 |
| WASHINGTON | 566.48 | 477.91 | 662.45 | 603.30 | 2,310.13 |
| WEST VIRGINIA | 118.52 | 90.42 | 114.71 | 91.83 | 415.49 |
| WISCONSIN | 315.16 | 283.67 | 336.29 | 298.92 | 1,234.04 |
| WYOMING | 11.55 | 27.56 | 24.18 | 34.97 | 98.26 |
| **9230 - MEPERIDINE (PETHIDINE) - Tot** | **33,664.93** | **30,578.32** | **47,752.80** | **39,381.26** | **151,377.31** |

**DRUG: 9250B - METHADONE**

| STATE | 1ST QUARTER | 2ND QUARTER | 3RD QUARTER | 4TH QUARTER | TOTAL GRAMS |
|---|---|---|---|---|---|
| ALABAMA | 82,657.50 | 70,790.51 | 49,908.68 | 50,618.66 | 253,975.34 |
| ALASKA | 11,828.98 | 8,905.18 | 8,561.38 | 9,140.32 | 38,435.86 |
| ARIZONA | 129,480.94 | 103,990.64 | 117,891.39 | 122,725.63 | 474,088.60 |
| ARKANSAS | 14,422.65 | 10,104.57 | 11,765.03 | 11,543.48 | 47,835.73 |
| CALIFORNIA | 331,256.05 | 288,493.06 | 299,033.04 | 291,435.70 | 1,210,217.84 |
| COLORADO | 66,987.51 | 48,490.95 | 50,774.18 | 54,127.11 | 220,379.75 |
| CONNECTICUT | 156,122.17 | 108,171.51 | 106,875.34 | 125,421.44 | 496,590.47 |
| DELAWARE | 44,686.73 | 31,622.05 | 42,476.49 | 37,734.47 | 156,519.73 |
| DISTRICT OF COLUMBIA | 10,471.76 | 9,706.11 | 10,195.11 | 9,740.83 | 40,113.80 |
| FLORIDA | 230,703.17 | 197,396.24 | 106,431.84 | 109,094.06 | 643,625.30 |
| GEORGIA | 128,318.23 | 101,293.13 | 96,792.15 | 96,564.07 | 422,967.57 |
| GUAM | 10.73 | 10.73 | 20.12 | 10.73 | 52.32 |
| HAWAII | 8,164.98 | 6,540.03 | 6,847.53 | 7,298.57 | 28,851.11 |
| IDAHO | 12,080.57 | 10,003.86 | 10,788.61 | 11,772.81 | 44,645.85 |
| ILLINOIS | 142,505.17 | 114,466.97 | 117,615.62 | 124,312.20 | 498,899.97 |
| INDIANA | 94,905.92 | 79,080.61 | 78,642.09 | 78,840.40 | 331,469.02 |

| | | | | | |
|---|---|---|---|---|---|
| IOWA | 15,642.61 | 11,292.60 | 13,225.14 | 13,730.11 | 53,890.46 |
| KANSAS | 20,185.62 | 19,339.61 | 19,774.70 | 19,259.09 | 78,559.02 |
| KENTUCKY | 73,722.48 | 48,380.90 | 56,517.01 | 60,062.81 | 238,683.19 |
| LOUISIANA | 45,536.37 | 29,958.52 | 33,453.31 | 37,562.83 | 146,511.04 |
| MAINE | 38,262.52 | 25,596.03 | 28,447.55 | 27,247.63 | 119,553.73 |
| MARYLAND | 241,391.02 | 178,564.06 | 171,951.91 | 182,451.72 | 774,358.72 |
| MASSACHUSETTS | 180,017.37 | 142,957.70 | 149,885.53 | 160,783.38 | 633,643.98 |
| MICHIGAN | 137,279.49 | 107,225.01 | 117,058.44 | 119,386.32 | 480,949.26 |
| MINNESOTA | 63,410.55 | 49,499.58 | 45,459.96 | 48,257.03 | 206,627.12 |
| MISSISSIPPI | 9,380.29 | 7,550.88 | 5,028.81 | 5,086.36 | 27,046.34 |
| MISSOURI | 38,016.74 | 30,876.31 | 23,426.80 | 25,832.87 | 118,152.73 |
| MONTANA | 9,690.11 | 7,952.47 | 9,699.05 | 8,243.86 | 35,585.49 |
| NEBRASKA | 8,531.27 | 5,942.70 | 5,280.65 | 4,943.73 | 24,698.36 |
| NEVADA | 39,278.38 | 32,500.98 | 32,876.36 | 33,971.55 | 138,627.26 |
| NEW HAMPSHIRE | 39,078.10 | 30,496.30 | 17,138.60 | 16,672.14 | 103,385.14 |
| NEW JERSEY | 154,752.93 | 126,536.58 | 133,650.82 | 128,121.55 | 543,061.88 |
| NEW MEXICO | 61,071.60 | 44,322.26 | 43,995.71 | 42,126.24 | 191,515.81 |
| NEW YORK | 377,797.40 | 311,832.33 | 320,877.68 | 335,133.28 | 1,345,640.69 |
| NORTH CAROLINA | 154,995.37 | 131,549.22 | 107,489.54 | 98,108.99 | 492,143.12 |
| NORTH DAKOTA | 5,835.69 | 5,611.64 | 6,087.47 | 6,027.63 | 23,562.43 |
| OHIO | 94,116.99 | 85,928.89 | 95,034.31 | 99,937.70 | 375,017.89 |
| OKLAHOMA | 43,133.16 | 31,024.23 | 32,938.16 | 36,746.51 | 143,842.06 |
| OREGON | 70,632.99 | 59,704.95 | 60,821.87 | 57,755.87 | 248,915.66 |
| PENNSYLVANIA | 234,418.73 | 179,603.62 | 190,266.82 | 208,576.28 | 812,865.45 |
| PUERTO RICO | 27,210.33 | 22,686.46 | 15,860.40 | 23,434.62 | 89,191.80 |
| RHODE ISLAND | 61,694.46 | 33,221.50 | 43,751.81 | 40,049.71 | 178,717.49 |
| SOUTH CAROLINA | 62,234.77 | 54,970.50 | 56,765.26 | 49,663.88 | 223,634.41 |
| SOUTH DAKOTA | 1,935.75 | 1,489.80 | 2,018.39 | 1,588.45 | 7,032.40 |
| TENNESSEE | 53,817.27 | 41,439.94 | 47,649.49 | 50,157.86 | 193,064.56 |
| TEXAS | 155,069.64 | 120,520.59 | 111,693.69 | 114,650.29 | 501,934.21 |
| UTAH | 34,340.40 | 28,471.17 | 25,790.92 | 28,945.29 | 117,547.77 |
| VERMONT | 22,440.85 | 18,828.37 | 20,654.29 | 21,634.28 | 83,557.80 |
| VIRGIN ISLANDS | 9.84 | 13.42 | 8.94 | 17.89 | 50.09 |
| VIRGINIA | 110,567.80 | 92,407.46 | 86,062.93 | 88,738.87 | 377,777.06 |
| WASHINGTON | 126,420.04 | 106,868.46 | 89,643.30 | 94,329.42 | 417,261.21 |
| WEST VIRGINIA | 28,525.28 | 23,037.60 | 24,231.17 | 24,305.50 | 100,099.55 |
| WISCONSIN | 73,082.22 | 56,496.18 | 63,397.75 | 65,586.85 | 258,563.01 |
| WYOMING | 698.88 | 648.44 | 641.73 | 584.40 | 2,573.46 |
| **9250B - METHADONE - Total** | **4,378,828.35** | **3,494,413.41** | **3,423,174.87** | **3,520,093.30** | **14,816,509.92** |

**DRUG: 9300 - MORPHINE**

| STATE | 1ST QUARTER | 2ND QUARTER | 3RD QUARTER | 4TH QUARTER | TOTAL GRAMS |
|---|---|---|---|---|---|
| ALABAMA | 49,356.36 | 46,181.20 | 46,797.49 | 45,716.05 | 188,051.10 |
| ALASKA | 6,962.18 | 6,740.22 | 6,752.82 | 6,347.33 | 26,802.55 |
| AMERICAN SAMOA | 6.77 | 9.85 | 0.30 | 5.79 | 22.71 |
| ARIZONA | 85,334.77 | 80,954.71 | 80,253.35 | 78,165.53 | 324,708.36 |
| ARKANSAS | 25,234.47 | 24,062.10 | 24,176.56 | 23,701.08 | 97,174.21 |
| CALIFORNIA | 282,483.68 | 264,423.71 | 262,665.18 | 258,527.06 | 1,068,099.63 |
| COLORADO | 50,615.85 | 48,433.15 | 48,536.27 | 48,364.01 | 195,949.27 |
| CONNECTICUT | 25,332.00 | 25,753.67 | 23,731.07 | 24,104.86 | 98,921.60 |
| DELAWARE | 14,365.39 | 13,446.55 | 13,892.38 | 13,974.27 | 55,678.60 |
| DISTRICT OF COLUMBIA | 2,709.70 | 2,867.17 | 2,431.15 | 2,317.96 | 10,325.98 |
| FLORIDA | 277,443.67 | 268,768.21 | 266,778.24 | 262,027.20 | 1,075,017.32 |
| GEORGIA | 79,418.53 | 77,118.09 | 77,936.13 | 75,543.45 | 310,016.19 |
| GUAM | 673.81 | 663.97 | 682.85 | 670.71 | 2,691.33 |
| HAWAII | 13,125.34 | 11,938.42 | 11,278.33 | 11,421.26 | 47,763.35 |
| IDAHO | 19,085.79 | 18,778.67 | 18,754.20 | 18,731.66 | 75,350.32 |
| ILLINOIS | 78,578.71 | 74,852.06 | 73,522.26 | 75,670.94 | 302,623.97 |
| INDIANA | 59,087.56 | 56,593.54 | 56,094.89 | 56,153.17 | 227,929.16 |
| IOWA | 23,473.75 | 22,464.85 | 22,798.79 | 23,456.61 | 92,194.00 |

| | | | | | |
|---|---|---|---|---|---|
| KANSAS | 30,800.45 | 29,515.34 | 29,462.90 | 30,685.81 | 120,464.50 |
| KENTUCKY | 31,773.11 | 29,813.49 | 29,920.36 | 29,565.68 | 121,072.64 |
| LOUISIANA | 38,542.70 | 35,614.13 | 36,950.95 | 34,617.14 | 145,724.93 |
| MAINE | 11,661.50 | 11,881.31 | 11,925.17 | 12,142.30 | 47,610.28 |
| MARYLAND | 48,033.49 | 46,985.23 | 45,807.40 | 44,639.08 | 185,465.20 |
| MASSACHUSETTS | 48,177.56 | 47,944.37 | 46,860.05 | 49,953.59 | 192,935.58 |
| MICHIGAN | 117,590.60 | 112,899.18 | 112,901.55 | 112,962.60 | 456,353.92 |
| MINNESOTA | 29,638.80 | 30,236.52 | 27,897.28 | 29,991.80 | 117,764.39 |
| MISSISSIPPI | 24,758.71 | 25,253.74 | 24,047.66 | 26,069.39 | 100,129.51 |
| MISSOURI | 61,900.12 | 61,330.21 | 60,441.64 | 60,984.73 | 244,656.70 |
| MONTANA | 11,805.37 | 11,483.89 | 11,679.15 | 11,335.15 | 46,303.56 |
| NEBRASKA | 14,005.46 | 13,564.99 | 13,690.96 | 14,160.51 | 55,421.92 |
| NEVADA | 36,374.78 | 34,229.12 | 34,736.43 | 33,185.23 | 138,525.56 |
| NEW HAMPSHIRE | 13,425.45 | 13,251.87 | 12,994.49 | 13,446.52 | 53,118.33 |
| NEW JERSEY | 59,269.30 | 56,335.36 | 56,136.15 | 55,589.96 | 227,330.77 |
| NEW MEXICO | 17,818.87 | 16,748.92 | 17,199.16 | 17,356.64 | 69,123.58 |
| NEW YORK | 108,842.08 | 106,951.65 | 103,169.69 | 103,055.13 | 422,018.55 |
| NORTH CAROLINA | 92,590.15 | 88,652.46 | 88,873.16 | 87,825.97 | 357,941.74 |
| NORTH DAKOTA | 4,505.77 | 4,227.34 | 3,908.38 | 4,572.03 | 17,213.51 |
| OHIO | 79,034.32 | 76,713.28 | 77,315.92 | 79,497.42 | 312,560.94 |
| OKLAHOMA | 46,525.69 | 44,572.73 | 44,428.37 | 44,039.43 | 179,566.23 |
| OREGON | 48,698.05 | 46,975.27 | 47,764.00 | 46,614.59 | 190,051.91 |
| PENNSYLVANIA | 143,553.21 | 150,848.67 | 141,779.20 | 140,518.92 | 576,699.99 |
| PUERTO RICO | 1,551.93 | 1,194.30 | 1,485.29 | 1,411.28 | 5,642.81 |
| RHODE ISLAND | 8,098.86 | 7,906.86 | 8,237.58 | 7,546.43 | 31,789.73 |
| SOUTH CAROLINA | 50,200.75 | 47,366.00 | 47,514.72 | 47,957.14 | 193,038.60 |
| SOUTH DAKOTA | 5,394.99 | 5,040.47 | 5,025.65 | 5,196.25 | 20,657.36 |
| TENNESSEE | 95,370.06 | 98,174.50 | 99,452.35 | 96,667.75 | 389,664.66 |
| TEXAS | 146,208.62 | 135,344.55 | 139,874.73 | 138,735.18 | 560,163.09 |
| UTAH | 35,550.93 | 35,072.34 | 35,070.91 | 33,928.43 | 139,622.61 |
| VERMONT | 7,616.38 | 7,305.95 | 6,902.82 | 7,286.03 | 29,111.18 |
| VIRGIN ISLANDS | 71.55 | 64.07 | 69.14 | 68.21 | 272.97 |
| VIRGINIA | 58,499.56 | 56,490.56 | 57,287.93 | 56,208.62 | 228,486.67 |
| WASHINGTON | 73,491.80 | 70,587.23 | 72,484.04 | 69,730.59 | 286,293.65 |
| WEST VIRGINIA | 13,921.15 | 13,995.43 | 13,990.38 | 13,983.88 | 55,890.85 |
| WISCONSIN | 50,768.96 | 48,754.92 | 49,296.13 | 49,364.10 | 198,184.11 |
| WYOMING | 6,254.88 | 5,824.53 | 6,027.63 | 6,150.94 | 24,257.98 |
| **9300 - MORPHINE - Total** | **2,765,614.29** | **2,673,200.93** | **2,659,691.59** | **2,641,943.36** | **10,740,450.16** |

**DRUG: 9652 - OXYMORPHONE**

| STATE | 1ST QUARTER | 2ND QUARTER | 3RD QUARTER | 4TH QUARTER | TOTAL GRAMS |
|---|---|---|---|---|---|
| ALABAMA | 1,038.67 | 938.49 | 918.06 | 799.28 | 3,694.50 |
| ALASKA | 37.74 | 29.71 | 26.23 | 21.95 | 115.62 |
| ARIZONA | 1,181.01 | 1,054.24 | 1,064.68 | 1,002.63 | 4,302.55 |
| ARKANSAS | 127.44 | 115.66 | 114.41 | 84.97 | 442.48 |
| CALIFORNIA | 3,095.59 | 2,834.87 | 2,711.85 | 2,693.87 | 11,336.18 |
| COLORADO | 847.72 | 748.52 | 786.65 | 736.07 | 3,118.96 |
| CONNECTICUT | 846.87 | 824.21 | 713.64 | 721.08 | 3,105.80 |
| DELAWARE | 850.53 | 732.55 | 677.95 | 665.11 | 2,926.13 |
| DISTRICT OF COLUMBIA | 52.28 | 34.08 | 37.65 | 30.96 | 154.96 |
| FLORIDA | 5,976.13 | 5,325.75 | 5,080.20 | 4,883.85 | 21,265.92 |
| GEORGIA | 1,799.14 | 1,663.72 | 1,532.40 | 1,343.86 | 6,339.13 |
| HAWAII | 135.87 | 159.51 | 116.15 | 131.14 | 542.66 |
| IDAHO | 495.03 | 437.58 | 435.48 | 408.49 | 1,776.57 |
| ILLINOIS | 1,311.61 | 1,019.27 | 1,034.66 | 1,046.61 | 4,412.15 |
| INDIANA | 1,177.84 | 968.78 | 896.61 | 903.79 | 3,947.01 |
| IOWA | 199.07 | 201.70 | 207.23 | 182.12 | 790.13 |
| KANSAS | 502.16 | 453.77 | 473.66 | 401.36 | 1,830.95 |
| KENTUCKY | 1,355.46 | 1,239.08 | 969.94 | 951.83 | 4,516.30 |
| LOUISIANA | 2,154.64 | 1,840.09 | 1,784.07 | 1,586.33 | 7,365.13 |

| | | | | | |
|---|---|---|---|---|---|
| MAINE | 247.33 | 242.25 | 227.40 | 218.74 | 935.72 |
| MARYLAND | 2,954.06 | 2,373.70 | 2,209.78 | 2,138.50 | 9,676.03 |
| MASSACHUSETTS | 680.76 | 641.60 | 640.30 | 647.31 | 2,609.97 |
| MICHIGAN | 5,351.75 | 4,915.92 | 4,027.65 | 3,654.44 | 17,949.77 |
| MINNESOTA | 164.95 | 155.05 | 163.34 | 134.89 | 618.23 |
| MISSISSIPPI | 604.44 | 550.11 | 591.37 | 567.91 | 2,313.84 |
| MISSOURI | 1,184.49 | 1,155.63 | 1,102.46 | 1,039.65 | 4,482.22 |
| MONTANA | 83.32 | 78.77 | 76.01 | 58.61 | 296.71 |
| NEBRASKA | 303.94 | 264.73 | 258.89 | 242.07 | 1,069.63 |
| NEVADA | 876.93 | 696.95 | 680.05 | 697.85 | 2,951.78 |
| NEW HAMPSHIRE | 176.64 | 157.81 | 171.82 | 149.16 | 655.43 |
| NEW JERSEY | 2,043.53 | 1,799.14 | 1,790.18 | 1,698.34 | 7,331.19 |
| NEW MEXICO | 227.66 | 227.49 | 211.16 | 216.65 | 882.96 |
| NEW YORK | 5,428.87 | 4,619.03 | 4,611.22 | 4,485.48 | 19,144.60 |
| NORTH CAROLINA | 5,674.83 | 5,065.83 | 4,735.36 | 4,557.99 | 20,034.01 |
| NORTH DAKOTA | 44.25 | 34.88 | 50.23 | 69.49 | 198.85 |
| OHIO | 2,161.15 | 1,937.73 | 1,865.25 | 1,741.07 | 7,705.20 |
| OKLAHOMA | 717.11 | 658.37 | 483.25 | 460.90 | 2,319.64 |
| OREGON | 524.42 | 471.25 | 455.73 | 427.09 | 1,878.49 |
| PENNSYLVANIA | 5,931.35 | 5,116.64 | 5,017.57 | 4,773.45 | 20,839.01 |
| RHODE ISLAND | 155.85 | 81.18 | 116.60 | 124.54 | 478.17 |
| SOUTH CAROLINA | 1,991.21 | 1,790.94 | 1,878.05 | 1,735.22 | 7,395.42 |
| SOUTH DAKOTA | 167.22 | 127.04 | 139.70 | 137.74 | 571.70 |
| TENNESSEE | 5,970.60 | 4,812.83 | 4,499.66 | 4,312.50 | 19,595.60 |
| TEXAS | 2,305.77 | 2,122.31 | 2,114.86 | 1,901.47 | 8,444.40 |
| UTAH | 1,918.77 | 1,690.35 | 1,705.43 | 1,512.96 | 6,827.51 |
| VERMONT | 83.50 | 75.47 | 88.94 | 76.81 | 324.72 |
| VIRGINIA | 3,201.52 | 2,608.28 | 2,650.16 | 2,350.42 | 10,810.38 |
| WASHINGTON | 871.22 | 843.93 | 884.87 | 863.20 | 3,463.22 |
| WEST VIRGINIA | 499.93 | 453.23 | 393.77 | 367.14 | 1,714.08 |
| WISCONSIN | 467.95 | 404.30 | 393.33 | 380.84 | 1,646.42 |
| WYOMING | 64.77 | 67.93 | 60.35 | 53.84 | 246.89 |
| **9652 - OXYMORPHONE - Total** | **76,264.91** | **66,862.23** | **63,876.23** | **60,391.54** | **267,394.91** |

**DRUG: 9780 - TAPENTADOL**

| STATE | 1ST QUARTER | 2ND QUARTER | 3RD QUARTER | 4TH QUARTER | TOTAL GRAMS |
|---|---|---|---|---|---|
| ALABAMA | 25,830.00 | 24,507.00 | 24,404.50 | 22,882.50 | 97,624.00 |
| ALASKA | 1,332.50 | 1,426.50 | 1,608.00 | 1,577.50 | 5,944.50 |
| ARIZONA | 21,389.00 | 19,660.50 | 19,631.00 | 20,787.00 | 81,467.50 |
| ARKANSAS | 8,475.50 | 8,078.50 | 8,345.00 | 8,915.00 | 33,814.00 |
| CALIFORNIA | 59,263.00 | 57,086.00 | 56,932.40 | 56,028.50 | 229,309.90 |
| COLORADO | 20,763.50 | 19,612.00 | 20,544.00 | 21,375.50 | 82,295.00 |
| CONNECTICUT | 6,378.50 | 5,954.50 | 5,951.50 | 6,203.00 | 24,487.50 |
| DELAWARE | 4,430.00 | 3,718.50 | 3,508.50 | 3,876.50 | 15,533.50 |
| DISTRICT OF COLUMBIA | 536.50 | 576.00 | 441.00 | 492.00 | 2,045.50 |
| FLORIDA | 54,780.50 | 52,046.50 | 51,461.50 | 52,213.00 | 210,501.50 |
| GEORGIA | 40,705.50 | 37,870.50 | 38,737.00 | 37,724.00 | 155,037.00 |
| HAWAII | 1,316.50 | 1,250.00 | 1,191.00 | 1,228.00 | 4,985.50 |
| IDAHO | 3,986.00 | 3,801.50 | 4,094.00 | 4,284.00 | 16,165.50 |
| ILLINOIS | 21,297.50 | 18,843.00 | 19,174.50 | 20,460.00 | 79,775.00 |
| INDIANA | 23,495.50 | 22,216.50 | 22,454.00 | 22,572.50 | 90,738.50 |
| IOWA | 6,840.50 | 6,134.50 | 6,289.50 | 5,923.00 | 25,187.50 |
| KANSAS | 4,400.00 | 4,350.00 | 4,577.00 | 4,618.00 | 17,945.00 |
| KENTUCKY | 7,802.50 | 7,561.00 | 8,134.00 | 7,647.50 | 31,145.00 |
| LOUISIANA | 10,803.00 | 10,254.00 | 10,391.50 | 10,166.00 | 41,614.50 |
| MAINE | 1,014.50 | 1,017.50 | 1,108.00 | 909.50 | 4,049.50 |
| MARYLAND | 22,117.00 | 20,992.50 | 21,731.00 | 20,973.50 | 85,814.00 |
| MASSACHUSETTS | 15,089.50 | 14,767.50 | 14,222.00 | 13,562.50 | 57,641.50 |
| MICHIGAN | 12,375.50 | 11,731.50 | 11,737.00 | 11,515.00 | 47,359.00 |
| MINNESOTA | 2,833.50 | 2,829.50 | 2,854.00 | 2,901.00 | 11,418.00 |

| | | | | | |
|---|---|---|---|---|---|
| MISSISSIPPI | 7,122.50 | 6,714.50 | 6,942.00 | 6,799.50 | 27,578.50 |
| MISSOURI | 15,041.00 | 14,618.50 | 15,014.00 | 14,572.50 | 59,246.00 |
| MONTANA | 1,205.00 | 1,105.50 | 1,097.00 | 1,030.50 | 4,438.00 |
| NEBRASKA | 7,191.50 | 7,088.00 | 7,598.50 | 8,057.00 | 29,935.00 |
| NEVADA | 5,617.50 | 5,732.00 | 5,472.50 | 5,724.00 | 22,546.00 |
| NEW HAMPSHIRE | 5,177.00 | 5,833.50 | 5,299.00 | 5,680.50 | 21,990.00 |
| NEW JERSEY | 27,616.00 | 26,435.50 | 26,864.50 | 26,694.00 | 107,610.00 |
| NEW MEXICO | 1,840.00 | 1,783.00 | 1,810.00 | 1,740.50 | 7,173.50 |
| NEW YORK | 58,017.50 | 55,997.50 | 56,354.50 | 57,194.50 | 227,564.00 |
| NORTH CAROLINA | 69,647.00 | 67,498.40 | 69,179.85 | 67,811.50 | 274,136.75 |
| NORTH DAKOTA | 1,060.00 | 1,007.00 | 846.50 | 1,082.00 | 3,995.50 |
| OHIO | 32,327.00 | 31,186.00 | 31,199.50 | 30,845.50 | 125,558.00 |
| OKLAHOMA | 7,285.50 | 7,406.50 | 6,854.00 | 6,660.00 | 28,206.00 |
| OREGON | 4,288.00 | 3,918.50 | 3,915.50 | 3,960.50 | 16,082.50 |
| PENNSYLVANIA | 40,346.00 | 38,315.50 | 39,363.50 | 37,830.00 | 155,855.00 |
| PUERTO RICO | 290.50 | 254.50 | 328.50 | 369.50 | 1,243.00 |
| RHODE ISLAND | 1,549.50 | 1,611.50 | 1,558.00 | 1,439.50 | 6,158.50 |
| SOUTH CAROLINA | 25,088.50 | 23,781.00 | 24,377.00 | 25,098.50 | 98,345.00 |
| SOUTH DAKOTA | 2,593.50 | 2,482.00 | 2,462.00 | 2,227.00 | 9,764.50 |
| TENNESSEE | 21,363.50 | 20,935.00 | 20,229.00 | 20,882.00 | 83,409.50 |
| TEXAS | 52,950.00 | 50,467.50 | 52,333.00 | 51,606.00 | 207,356.50 |
| UTAH | 8,073.50 | 8,178.50 | 7,931.00 | 7,730.50 | 31,913.50 |
| VERMONT | 759.00 | 675.00 | 663.50 | 855.00 | 2,952.50 |
| VIRGIN ISLANDS | 7.50 | 15.00 | 15.00 | 7.50 | 45.00 |
| VIRGINIA | 24,331.00 | 24,180.50 | 24,232.00 | 23,262.50 | 96,006.00 |
| WASHINGTON | 11,010.50 | 10,528.00 | 10,948.00 | 10,655.50 | 43,142.00 |
| WEST VIRGINIA | 3,476.50 | 3,348.00 | 3,352.50 | 3,205.00 | 13,382.00 |
| WISCONSIN | 7,449.25 | 7,303.50 | 7,775.50 | 7,657.25 | 30,185.50 |
| WYOMING | 1,020.00 | 940.00 | 980.00 | 906.50 | 3,846.50 |
| **9780 - TAPENTADOL - Total** | **821,000.25** | **785,625.40** | **794,517.75** | **790,419.75** | **3,191,563.15** |

**DRUG: 9801 - FENTANYL BASE**

| STATE | 1ST QUARTER | 2ND QUARTER | 3RD QUARTER | 4TH QUARTER | TOTAL GRAMS |
|---|---|---|---|---|---|
| ALABAMA | 790.96 | 776.82 | 753.16 | 773.68 | 3,094.63 |
| ALASKA | 98.72 | 84.63 | 81.84 | 79.76 | 344.96 |
| AMERICAN SAMOA | 0.11 | 0.08 | 0.03 | 0.03 | 0.24 |
| ARIZONA | 824.45 | 800.72 | 711.74 | 769.25 | 3,106.16 |
| ARKANSAS | 436.55 | 410.38 | 423.02 | 418.90 | 1,688.86 |
| CALIFORNIA | 3,674.48 | 3,559.16 | 3,447.63 | 3,602.60 | 14,283.87 |
| COLORADO | 664.26 | 677.24 | 639.56 | 658.40 | 2,639.45 |
| CONNECTICUT | 507.31 | 504.77 | 479.22 | 520.31 | 2,011.60 |
| DELAWARE | 148.98 | 136.66 | 131.90 | 123.57 | 541.11 |
| DISTRICT OF COLUMBIA | 52.84 | 67.20 | 48.79 | 41.28 | 210.11 |
| FLORIDA | 3,473.79 | 3,513.55 | 3,097.53 | 2,890.62 | 12,975.48 |
| GEORGIA | 1,577.69 | 1,656.28 | 1,597.29 | 1,489.63 | 6,320.88 |
| GUAM | 9.05 | 8.65 | 8.15 | 9.80 | 35.65 |
| HAWAII | 113.91 | 101.42 | 107.59 | 101.07 | 423.99 |
| IDAHO | 351.60 | 325.15 | 331.60 | 329.33 | 1,337.68 |
| ILLINOIS | 1,720.12 | 1,759.41 | 1,581.70 | 1,609.96 | 6,671.19 |
| INDIANA | 1,059.64 | 982.77 | 999.07 | 979.33 | 4,020.81 |
| IOWA | 544.61 | 505.31 | 490.30 | 488.28 | 2,028.51 |
| KANSAS | 613.33 | 580.52 | 573.94 | 581.74 | 2,349.54 |
| KENTUCKY | 647.36 | 628.32 | 580.50 | 595.18 | 2,451.37 |
| LOUISIANA | 673.75 | 617.95 | 609.29 | 614.54 | 2,515.52 |
| MAINE | 230.19 | 218.78 | 206.62 | 203.29 | 858.88 |
| MARYLAND | 703.50 | 735.58 | 626.58 | 650.57 | 2,716.22 |
| MASSACHUSETTS | 814.19 | 882.23 | 721.36 | 740.77 | 3,158.55 |
| MICHIGAN | 1,592.86 | 1,647.51 | 1,485.95 | 1,561.17 | 6,287.48 |
| MINNESOTA | 631.13 | 548.47 | 550.69 | 553.46 | 2,283.74 |
| MISSISSIPPI | 769.30 | 905.99 | 773.19 | 756.94 | 3,205.42 |

| | | | | | |
|---|---|---|---|---|---|
| MISSOURI | 1,139.55 | 1,129.02 | 1,131.08 | 1,298.75 | 4,698.40 |
| MONTANA | 198.24 | 187.98 | 190.95 | 199.65 | 776.81 |
| NEBRASKA | 337.44 | 312.77 | 301.44 | 303.53 | 1,255.18 |
| NEVADA | 297.55 | 412.72 | 367.47 | 342.32 | 1,420.06 |
| NEW HAMPSHIRE | 200.09 | 193.52 | 186.64 | 197.77 | 778.02 |
| NEW JERSEY | 1,268.33 | 1,337.20 | 1,086.31 | 1,155.50 | 4,847.34 |
| NEW MEXICO | 248.00 | 230.64 | 223.15 | 239.60 | 941.39 |
| NEW YORK | 2,568.61 | 2,935.72 | 2,232.76 | 2,219.96 | 9,957.05 |
| NORTH CAROLINA | 1,585.99 | 1,527.25 | 1,495.42 | 1,497.90 | 6,106.55 |
| NORTH DAKOTA | 143.44 | 133.90 | 112.08 | 136.92 | 526.34 |
| OHIO | 1,297.88 | 1,276.94 | 1,121.88 | 1,221.41 | 4,918.11 |
| OKLAHOMA | 638.63 | 582.87 | 579.21 | 574.35 | 2,375.05 |
| OREGON | 500.33 | 477.26 | 481.23 | 479.43 | 1,938.25 |
| PENNSYLVANIA | 2,367.35 | 2,495.90 | 2,376.08 | 2,388.29 | 9,627.61 |
| PUERTO RICO | 171.68 | 126.74 | 174.40 | 163.48 | 636.31 |
| RHODE ISLAND | 104.94 | 109.62 | 107.06 | 99.58 | 421.20 |
| SOUTH CAROLINA | 734.42 | 706.91 | 692.01 | 677.12 | 2,810.45 |
| SOUTH DAKOTA | 155.44 | 153.34 | 144.67 | 146.52 | 599.98 |
| TENNESSEE | 860.83 | 1,025.15 | 1,089.89 | 1,014.74 | 3,990.60 |
| TEXAS | 3,190.63 | 2,987.86 | 3,247.94 | 3,133.76 | 12,560.18 |
| UTAH | 529.05 | 516.65 | 492.78 | 488.86 | 2,027.33 |
| VERMONT | 100.62 | 92.90 | 98.00 | 96.91 | 388.43 |
| VIRGIN ISLANDS | 3.86 | 5.22 | 4.05 | 4.76 | 17.90 |
| VIRGINIA | 1,019.15 | 1,024.25 | 927.17 | 925.79 | 3,896.36 |
| WASHINGTON | 864.73 | 801.47 | 806.47 | 836.28 | 3,308.95 |
| WEST VIRGINIA | 295.09 | 283.27 | 269.78 | 288.97 | 1,137.12 |
| WISCONSIN | 825.16 | 743.78 | 721.77 | 721.90 | 3,012.62 |
| WYOMING | 87.40 | 87.92 | 86.91 | 89.71 | 351.95 |
| **9801 - FENTANYL BASE - Total** | **44,459.12** | **44,534.31** | **41,806.82** | **42,087.22** | **172,887.46** |

**DRUG: 1100 - AMPHETAMINE**

| STATE NAME | 1ST QUARTER | 2ND QUARTER | 3RD QUARTER | 4TH QUARTER | TOTAL GRAMS |
|---|---|---|---|---|---|
| ALABAMA | 2,906.30 | 2,812.62 | 2,926.11 | 3,020.98 | 11,666.01 |
| ALASKA | 1,235.71 | 1,200.11 | 1,297.89 | 1,271.98 | 5,005.68 |
| AMERICAN SAMOA | 1.91 | 0.00 | 1.91 | 1.91 | 5.72 |
| ARIZONA | 1,675.03 | 1,517.51 | 1,629.32 | 1,721.71 | 6,543.57 |
| ARKANSAS | 1,956.19 | 1,933.12 | 1,988.68 | 2,103.00 | 7,980.99 |
| CALIFORNIA | 888.28 | 826.02 | 855.46 | 890.76 | 3,460.51 |
| COLORADO | 2,055.67 | 1,890.35 | 2,074.89 | 2,161.19 | 8,182.11 |
| CONNECTICUT | 1,757.59 | 1,756.30 | 1,798.45 | 1,871.18 | 7,183.52 |
| DELAWARE | 2,412.07 | 2,424.63 | 2,429.05 | 2,545.60 | 9,811.35 |
| DISTRICT OF COLUMBIA | 2,225.60 | 1,856.82 | 2,025.58 | 1,998.95 | 8,106.96 |
| FLORIDA | 2,013.63 | 1,779.07 | 1,976.53 | 2,040.08 | 7,809.30 |
| GEORGIA | 2,238.72 | 2,086.79 | 2,217.62 | 2,283.11 | 8,826.23 |
| GUAM | 257.13 | 224.36 | 339.19 | 229.31 | 1,049.99 |
| HAWAII | 844.15 | 742.09 | 815.83 | 849.76 | 3,251.82 |
| IDAHO | 2,103.73 | 2,075.19 | 2,182.51 | 2,310.15 | 8,671.59 |
| ILLINOIS | 1,691.96 | 1,629.91 | 1,667.60 | 1,742.96 | 6,732.43 |
| INDIANA | 2,327.35 | 2,236.70 | 2,311.84 | 2,396.61 | 9,272.50 |
| IOWA | 1,857.82 | 1,803.43 | 1,885.58 | 1,946.88 | 7,493.72 |
| KANSAS | 2,445.75 | 2,399.12 | 2,497.90 | 2,599.83 | 9,942.61 |
| KENTUCKY | 1,857.41 | 1,750.32 | 1,808.71 | 1,883.97 | 7,300.42 |
| LOUISIANA | 4,008.34 | 3,818.39 | 3,953.34 | 4,180.58 | 15,960.66 |
| MAINE | 2,101.75 | 1,938.95 | 2,124.53 | 2,160.01 | 8,325.24 |
| MARYLAND | 1,852.55 | 1,753.02 | 1,796.06 | 1,850.77 | 7,252.40 |
| MASSACHUSETTS | 2,963.95 | 2,846.13 | 2,932.12 | 3,026.45 | 11,768.64 |
| MICHIGAN | 2,609.17 | 2,515.37 | 2,605.03 | 2,685.42 | 10,415.00 |
| MINNESOTA | 2,247.77 | 2,231.61 | 2,264.79 | 2,379.78 | 9,123.95 |
| MISSISSIPPI | 2,204.95 | 2,113.31 | 2,216.62 | 2,320.87 | 8,855.75 |
| MISSOURI | 2,534.44 | 2,553.34 | 2,797.76 | 2,744.92 | 10,630.47 |
| MONTANA | 1,715.00 | 1,695.98 | 1,762.92 | 1,822.72 | 6,996.62 |
| NEBRASKA | 1,712.46 | 1,669.29 | 1,714.65 | 1,834.77 | 6,931.17 |
| NEVADA | 1,159.67 | 1,061.87 | 1,143.84 | 1,182.47 | 4,547.86 |
| NEW HAMPSHIRE | 3,343.48 | 3,383.58 | 3,452.98 | 3,580.40 | 13,760.45 |
| NEW JERSEY | 1,408.23 | 1,351.24 | 1,391.87 | 1,472.98 | 5,624.32 |
| NEW MEXICO | 902.74 | 698.74 | 833.84 | 845.16 | 3,280.49 |
| NEW YORK | 1,443.87 | 1,288.73 | 1,356.11 | 1,429.39 | 5,518.09 |
| NORTH CAROLINA | 2,494.11 | 2,391.16 | 2,497.87 | 2,593.78 | 9,976.92 |
| NORTH DAKOTA | 1,570.23 | 1,484.69 | 1,480.43 | 1,630.45 | 6,165.81 |
| OHIO | 1,927.26 | 1,874.71 | 1,923.34 | 2,019.66 | 7,744.97 |
| OKLAHOMA | 1,567.07 | 1,525.65 | 1,582.49 | 1,640.33 | 6,315.54 |
| OREGON | 1,849.68 | 1,869.99 | 1,923.58 | 2,030.47 | 7,673.72 |
| PENNSYLVANIA | 1,802.88 | 1,793.59 | 1,803.66 | 1,844.08 | 7,244.20 |
| PUERTO RICO | 223.59 | 183.15 | 228.36 | 223.60 | 858.70 |
| RHODE ISLAND | 3,826.34 | 3,742.28 | 3,858.69 | 3,894.01 | 15,321.33 |
| SOUTH CAROLINA | 3,285.34 | 3,156.60 | 3,284.35 | 3,415.56 | 13,141.85 |
| SOUTH DAKOTA | 1,830.91 | 1,738.17 | 1,755.94 | 1,743.88 | 7,068.90 |
| TENNESSEE | 2,240.88 | 2,194.92 | 2,274.26 | 2,374.44 | 9,084.50 |
| TEXAS | 1,998.77 | 1,956.92 | 2,020.93 | 2,098.06 | 8,074.68 |
| UTAH | 3,668.25 | 3,701.47 | 3,837.97 | 3,991.88 | 15,199.57 |
| VERMONT | 1,838.07 | 1,669.69 | 1,756.42 | 1,853.86 | 7,118.04 |
| VIRGIN ISLANDS | 407.37 | 307.22 | 324.32 | 314.24 | 1,353.16 |
| VIRGINIA | 2,438.23 | 2,351.22 | 2,415.68 | 2,472.67 | 9,677.81 |
| WASHINGTON | 1,858.44 | 1,832.56 | 1,887.06 | 1,963.02 | 7,541.08 |
| WEST VIRGINIA | 1,342.69 | 1,209.73 | 1,264.25 | 1,309.57 | 5,126.24 |
| WISCONSIN | 2,281.08 | 2,231.75 | 2,268.91 | 2,348.71 | 9,130.45 |
| WYOMING | 1,408.28 | 1,295.82 | 1,393.67 | 1,452.60 | 5,550.37 |
| **US TOTAL** | **1,916.62** | **1,831.77** | **1,910.13** | **1,981.83** | **7,640.35** |

**DRUG: 1105 - METHAMPHETAMINE**

| STATE NAME | 1ST QUARTER | 2ND QUARTER | 3RD QUARTER | 4TH QUARTER | TOTAL GRAMS |
|---|---|---|---|---|---|
| ALABAMA | 0.64 | 0.73 | 0.72 | 0.52 | 2.61 |

| | | | | | |
|---|---|---|---|---|---|
| ALASKA | 0.28 | 0.45 | 0.17 | 0.00 | 0.90 |
| ARIZONA | 0.43 | 0.51 | 0.43 | 0.48 | 1.85 |
| ARKANSAS | 0.06 | 0.11 | 0.10 | 0.11 | 0.37 |
| CALIFORNIA | 0.62 | 0.61 | 0.60 | 0.59 | 2.42 |
| COLORADO | 0.75 | 0.82 | 0.81 | 0.89 | 3.28 |
| CONNECTICUT | 0.44 | 0.62 | 0.62 | 0.64 | 2.32 |
| DELAWARE | 0.58 | 0.63 | 0.76 | 0.40 | 2.37 |
| DISTRICT OF COLUMBIA | 0.80 | 1.07 | 0.87 | 1.74 | 4.47 |
| FLORIDA | 0.27 | 0.27 | 0.28 | 0.28 | 1.10 |
| GEORGIA | 0.29 | 0.25 | 0.27 | 0.25 | 1.07 |
| HAWAII | 0.30 | 0.30 | 0.18 | 0.24 | 1.00 |
| IDAHO | 0.72 | 0.79 | 0.85 | 0.90 | 3.25 |
| ILLINOIS | 0.35 | 0.24 | 0.32 | 0.25 | 1.15 |
| INDIANA | 0.43 | 0.56 | 0.37 | 0.27 | 1.63 |
| IOWA | 0.34 | 0.55 | 0.54 | 0.54 | 1.98 |
| KANSAS | 0.46 | 0.34 | 0.37 | 0.31 | 1.48 |
| KENTUCKY | 0.24 | 0.22 | 0.21 | 0.23 | 0.91 |
| LOUISIANA | 0.25 | 0.27 | 0.36 | 0.32 | 1.20 |
| MAINE | 0.64 | 0.73 | 0.79 | 0.57 | 2.72 |
| MARYLAND | 0.55 | 0.59 | 0.60 | 0.61 | 2.35 |
| MASSACHUSETTS | 0.40 | 0.28 | 0.43 | 0.28 | 1.39 |
| MICHIGAN | 0.18 | 0.14 | 0.14 | 0.15 | 0.61 |
| MINNESOTA | 0.90 | 0.89 | 0.90 | 1.01 | 3.70 |
| MISSISSIPPI | 0.18 | 0.31 | 0.24 | 0.24 | 0.97 |
| MISSOURI | 0.66 | 0.73 | 0.84 | 0.95 | 3.18 |
| MONTANA | 0.00 | 0.00 | 0.20 | 0.00 | 0.20 |
| NEBRASKA | 0.37 | 0.48 | 0.31 | 0.07 | 1.23 |
| NEVADA | 0.24 | 0.37 | 0.33 | 0.31 | 1.25 |
| NEW HAMPSHIRE | 0.58 | 0.40 | 0.49 | 0.40 | 1.86 |
| NEW JERSEY | 0.41 | 0.32 | 0.30 | 0.32 | 1.34 |
| NEW MEXICO | 0.29 | 0.45 | 0.53 | 0.51 | 1.78 |
| NEW YORK | 0.42 | 0.45 | 0.44 | 0.41 | 1.72 |
| NORTH CAROLINA | 0.51 | 0.36 | 0.31 | 0.32 | 1.50 |
| NORTH DAKOTA | 0.48 | 0.90 | 0.66 | 0.42 | 2.45 |
| OHIO | 0.25 | 0.27 | 0.28 | 0.26 | 1.06 |
| OKLAHOMA | 0.18 | 0.32 | 0.32 | 0.33 | 1.16 |
| OREGON | 1.24 | 1.12 | 0.90 | 1.01 | 4.27 |
| PENNSYLVANIA | 0.70 | 0.71 | 0.76 | 0.73 | 2.89 |
| RHODE ISLAND | 0.34 | 0.76 | 0.34 | 0.23 | 1.68 |
| SOUTH CAROLINA | 0.13 | 0.19 | 0.20 | 0.13 | 0.65 |
| SOUTH DAKOTA | 0.69 | 0.49 | 1.04 | 1.09 | 3.31 |
| TENNESSEE | 0.20 | 0.24 | 0.27 | 0.25 | 0.97 |
| TEXAS | 0.37 | 0.31 | 0.38 | 0.36 | 1.42 |
| UTAH | 0.87 | 0.84 | 1.09 | 1.06 | 3.87 |
| VERMONT | 0.51 | 0.19 | 0.26 | 0.26 | 1.22 |
| VIRGINIA | 0.27 | 0.35 | 0.32 | 0.30 | 1.24 |
| WASHINGTON | 1.37 | 1.13 | 0.99 | 1.02 | 4.50 |
| WEST VIRGINIA | 0.17 | 0.09 | 0.20 | 0.17 | 0.63 |
| WISCONSIN | 0.36 | 0.39 | 0.37 | 0.34 | 1.46 |
| WYOMING | 1.00 | 0.57 | 0.93 | 1.00 | 3.49 |
| **US TOTAL** | **0.45** | **0.45** | **0.45** | **0.43** | **1.78** |

**DRUG: 1205 - LISDEXAMFETAMINE**

| STATE NAME | 1ST QUARTER | 2ND QUARTER | 3RD QUARTER | 4TH QUARTER | TOTAL GRAMS |
|---|---|---|---|---|---|
| ALABAMA | 1,547.87 | 1,356.03 | 1,433.27 | 1,480.24 | 5,817.41 |
| ALASKA | 437.59 | 418.13 | 451.34 | 464.45 | 1,771.50 |
| ARIZONA | 665.82 | 626.74 | 643.91 | 685.57 | 2,622.04 |
| ARKANSAS | 1,006.83 | 845.92 | 906.71 | 934.56 | 3,694.02 |
| CALIFORNIA | 315.52 | 289.85 | 296.86 | 307.88 | 1,210.11 |
| COLORADO | 626.75 | 573.99 | 621.95 | 653.38 | 2,476.07 |
| CONNECTICUT | 1,028.84 | 974.75 | 1,005.11 | 1,045.76 | 4,054.47 |
| DELAWARE | 1,517.33 | 1,410.53 | 1,428.30 | 1,464.61 | 5,820.76 |

| | | | | | |
|---|---|---|---|---|---|
| DISTRICT OF COLUMBIA | 1,032.03 | 845.36 | 950.65 | 944.60 | 3,772.64 |
| FLORIDA | 752.39 | 673.85 | 679.55 | 719.69 | 2,825.48 |
| GEORGIA | 1,006.69 | 870.14 | 926.07 | 957.97 | 3,760.86 |
| GUAM | 12.31 | 15.23 | 7.78 | 7.13 | 42.44 |
| HAWAII | 144.20 | 128.13 | 136.80 | 143.05 | 552.18 |
| IDAHO | 692.13 | 658.17 | 702.13 | 743.58 | 2,796.01 |
| ILLINOIS | 596.92 | 548.08 | 580.11 | 606.68 | 2,331.80 |
| INDIANA | 1,285.70 | 1,142.45 | 1,214.74 | 1,256.14 | 4,899.03 |
| IOWA | 1,126.20 | 1,015.33 | 1,061.07 | 1,098.21 | 4,300.80 |
| KANSAS | 1,155.81 | 1,065.93 | 1,122.66 | 1,148.86 | 4,493.25 |
| KENTUCKY | 917.47 | 814.09 | 841.64 | 865.20 | 3,438.41 |
| LOUISIANA | 2,019.10 | 1,708.90 | 1,847.64 | 1,927.49 | 7,503.13 |
| MAINE | 1,326.83 | 1,262.64 | 1,363.04 | 1,399.99 | 5,352.50 |
| MARYLAND | 1,040.50 | 953.14 | 959.69 | 1,014.62 | 3,967.94 |
| MASSACHUSETTS | 651.45 | 597.79 | 617.48 | 640.40 | 2,507.12 |
| MICHIGAN | 665.33 | 583.09 | 606.08 | 645.68 | 2,500.19 |
| MINNESOTA | 650.25 | 620.05 | 645.52 | 692.37 | 2,608.19 |
| MISSISSIPPI | 1,272.32 | 1,007.61 | 1,111.45 | 1,160.37 | 4,551.75 |
| MISSOURI | 1,130.79 | 1,060.89 | 1,096.41 | 1,134.04 | 4,422.14 |
| MONTANA | 725.33 | 716.51 | 717.44 | 751.92 | 2,911.20 |
| NEBRASKA | 1,016.96 | 924.03 | 992.97 | 1,035.61 | 3,969.56 |
| NEVADA | 309.52 | 273.98 | 262.94 | 266.45 | 1,112.89 |
| NEW HAMPSHIRE | 1,112.31 | 1,079.42 | 1,117.63 | 1,147.36 | 4,456.72 |
| NEW JERSEY | 565.12 | 521.74 | 519.76 | 550.42 | 2,157.04 |
| NEW MEXICO | 178.30 | 178.47 | 183.69 | 179.14 | 719.60 |
| NEW YORK | 481.88 | 412.12 | 430.25 | 462.32 | 1,786.58 |
| NORTH CAROLINA | 1,175.33 | 1,056.77 | 1,115.33 | 1,150.56 | 4,497.99 |
| NORTH DAKOTA | 976.84 | 926.90 | 889.25 | 1,007.78 | 3,800.76 |
| OHIO | 833.36 | 743.51 | 782.13 | 809.34 | 3,168.35 |
| OKLAHOMA | 1,046.46 | 893.13 | 954.88 | 997.09 | 3,891.55 |
| OREGON | 438.78 | 398.45 | 427.84 | 443.77 | 1,708.84 |
| PENNSYLVANIA | 742.59 | 687.00 | 697.41 | 722.05 | 2,849.04 |
| PUERTO RICO | 58.95 | 45.93 | 49.68 | 53.97 | 208.52 |
| RHODE ISLAND | 753.87 | 702.57 | 744.37 | 698.56 | 2,899.38 |
| SOUTH CAROLINA | 1,378.50 | 1,196.53 | 1,245.43 | 1,300.71 | 5,121.18 |
| SOUTH DAKOTA | 1,192.58 | 1,048.23 | 1,060.94 | 1,077.56 | 4,379.31 |
| TENNESSEE | 1,061.97 | 918.34 | 995.71 | 1,043.12 | 4,019.14 |
| TEXAS | 1,044.62 | 893.86 | 951.78 | 988.07 | 3,878.33 |
| UTAH | 841.77 | 823.55 | 844.72 | 887.52 | 3,397.57 |
| VERMONT | 1,255.44 | 1,194.93 | 1,219.13 | 1,277.70 | 4,947.20 |
| VIRGIN ISLANDS | 58.95 | 89.49 | 68.43 | 62.11 | 278.98 |
| VIRGINIA | 908.48 | 784.27 | 827.08 | 839.90 | 3,359.73 |
| WASHINGTON | 578.33 | 530.18 | 562.79 | 576.17 | 2,247.47 |
| WEST VIRGINIA | 1,035.19 | 902.16 | 922.56 | 961.31 | 3,821.21 |
| WISCONSIN | 1,406.97 | 1,306.33 | 1,359.13 | 1,444.65 | 5,517.08 |
| WYOMING | 502.07 | 454.89 | 518.28 | 522.07 | 1,997.31 |
| **US TOTAL** | **802.46** | **716.24** | **749.51** | **781.19** | **3,049.41** |

**DRUG: 1724 - METHYLPHENIDATE (DL;D;L;ISOMERS)**

| STATE NAME | 1ST QUARTER | 2ND QUARTER | 3RD QUARTER | 4TH QUARTER | TOTAL GRAMS |
|---|---|---|---|---|---|
| ALABAMA | 1,625.88 | 1,261.73 | 1,366.95 | 1,495.59 | 5,750.14 |
| ALASKA | 1,156.49 | 998.77 | 1,055.01 | 1,086.95 | 4,297.22 |
| AMERICAN SAMOA | 2.64 | 6.59 | 10.54 | 3.95 | 23.72 |
| ARIZONA | 1,258.84 | 1,065.54 | 1,110.56 | 1,190.58 | 4,625.53 |
| ARKANSAS | 1,350.22 | 1,071.71 | 1,165.22 | 1,271.06 | 4,858.22 |
| CALIFORNIA | 723.33 | 619.63 | 627.69 | 664.72 | 2,635.37 |
| COLORADO | 1,326.66 | 1,142.30 | 1,224.58 | 1,298.45 | 4,991.99 |
| CONNECTICUT | 1,580.48 | 1,367.27 | 1,373.73 | 1,480.67 | 5,802.15 |
| DELAWARE | 2,086.74 | 1,690.67 | 1,654.27 | 1,876.31 | 7,307.98 |
| DISTRICT OF COLUMBIA | 1,525.39 | 1,124.26 | 1,096.86 | 1,195.42 | 4,941.93 |
| FLORIDA | 1,134.99 | 925.26 | 948.02 | 1,049.84 | 4,058.11 |
| GEORGIA | 1,751.62 | 1,292.33 | 1,424.65 | 1,549.78 | 6,018.37 |

| | | | | | |
|---|---|---|---|---|---|
| GUAM | 234.04 | 231.41 | 194.01 | 270.17 | 929.64 |
| HAWAII | 798.88 | 629.64 | 675.41 | 701.56 | 2,805.49 |
| IDAHO | 1,915.46 | 1,687.16 | 1,727.84 | 1,890.90 | 7,221.36 |
| ILLINOIS | 1,327.26 | 1,081.93 | 1,137.18 | 1,241.22 | 4,787.59 |
| INDIANA | 1,921.45 | 1,511.19 | 1,657.50 | 1,802.07 | 6,892.21 |
| IOWA | 2,681.86 | 2,266.95 | 2,396.30 | 2,582.08 | 9,927.18 |
| KANSAS | 1,815.95 | 1,554.79 | 1,590.42 | 1,777.75 | 6,738.90 |
| KENTUCKY | 1,821.48 | 1,455.13 | 1,512.51 | 1,612.11 | 6,401.24 |
| LOUISIANA | 1,834.06 | 1,300.26 | 1,440.66 | 1,626.76 | 6,201.74 |
| MAINE | 2,755.31 | 2,517.10 | 2,604.72 | 2,674.96 | 10,552.09 |
| MARYLAND | 1,836.47 | 1,522.18 | 1,503.05 | 1,647.17 | 6,508.86 |
| MASSACHUSETTS | 2,250.31 | 1,883.94 | 1,891.28 | 2,060.21 | 8,085.74 |
| MICHIGAN | 1,790.11 | 1,483.94 | 1,519.41 | 1,676.59 | 6,470.04 |
| MINNESOTA | 1,975.89 | 1,713.45 | 1,716.39 | 1,892.54 | 7,298.27 |
| MISSISSIPPI | 1,528.85 | 1,088.91 | 1,240.80 | 1,392.09 | 5,250.65 |
| MISSOURI | 2,291.61 | 2,018.58 | 2,086.05 | 2,356.65 | 8,752.89 |
| MONTANA | 1,776.92 | 1,601.58 | 1,655.43 | 1,764.48 | 6,798.42 |
| NEBRASKA | 1,785.19 | 1,485.95 | 1,621.53 | 1,774.95 | 6,667.62 |
| NEVADA | 586.88 | 508.96 | 509.19 | 552.07 | 2,157.10 |
| NEW HAMPSHIRE | 2,661.55 | 2,436.56 | 2,475.37 | 2,632.01 | 10,205.49 |
| NEW JERSEY | 1,079.14 | 922.06 | 886.90 | 1,019.65 | 3,907.76 |
| NEW MEXICO | 892.85 | 764.03 | 796.91 | 797.50 | 3,251.29 |
| NEW YORK | 1,209.97 | 1,001.97 | 987.33 | 1,111.64 | 4,310.90 |
| NORTH CAROLINA | 2,042.50 | 1,645.87 | 1,713.23 | 1,866.72 | 7,268.31 |
| NORTH DAKOTA | 2,398.91 | 2,018.98 | 1,926.71 | 2,282.92 | 8,627.52 |
| OHIO | 1,784.92 | 1,476.89 | 1,539.22 | 1,694.34 | 6,495.37 |
| OKLAHOMA | 1,313.96 | 1,051.31 | 1,158.77 | 1,205.22 | 4,729.25 |
| OREGON | 1,740.97 | 1,543.77 | 1,561.21 | 1,676.76 | 6,522.71 |
| PENNSYLVANIA | 1,797.75 | 1,557.63 | 1,599.58 | 1,709.36 | 6,664.31 |
| PUERTO RICO | 351.89 | 225.90 | 263.40 | 264.97 | 1,106.17 |
| RHODE ISLAND | 1,836.65 | 1,553.90 | 1,565.50 | 1,559.91 | 6,515.96 |
| SOUTH CAROLINA | 2,027.99 | 1,586.23 | 1,622.31 | 1,828.26 | 7,064.80 |
| SOUTH DAKOTA | 2,381.22 | 2,019.80 | 2,037.56 | 2,159.35 | 8,597.92 |
| TENNESSEE | 1,360.11 | 1,096.78 | 1,191.12 | 1,299.59 | 4,947.60 |
| TEXAS | 1,393.98 | 1,027.20 | 1,118.21 | 1,280.31 | 4,819.70 |
| UTAH | 1,830.97 | 1,667.21 | 1,707.97 | 1,872.33 | 7,078.48 |
| VERMONT | 3,107.37 | 2,821.63 | 2,867.42 | 3,085.68 | 11,882.10 |
| VIRGIN ISLANDS | 114.89 | 71.45 | 91.48 | 77.63 | 355.45 |
| VIRGINIA | 1,730.46 | 1,382.95 | 1,451.44 | 1,525.42 | 6,090.28 |
| WASHINGTON | 1,763.09 | 1,547.45 | 1,552.38 | 1,669.79 | 6,532.71 |
| WEST VIRGINIA | 1,644.96 | 1,378.70 | 1,356.83 | 1,454.80 | 5,835.29 |
| WISCONSIN | 1,906.34 | 1,642.89 | 1,688.94 | 1,826.76 | 7,064.92 |
| WYOMING | 1,390.06 | 1,240.26 | 1,259.71 | 1,332.49 | 5,222.52 |
| **US TOTAL** | **1,490.29** | **1,225.05** | **1,270.57** | **1,389.18** | **5,375.09** |

**DRUG: 9050 - CODEINE**

| STATE NAME | 1ST QUARTER | 2ND QUARTER | 3RD QUARTER | 4TH QUARTER | TOTAL GRAMS |
|---|---|---|---|---|---|
| ALABAMA | 901.51 | 871.20 | 839.75 | 906.20 | 3,518.65 |
| ALASKA | 676.98 | 588.43 | 686.45 | 619.13 | 2,570.99 |
| AMERICAN SAMOA | 548.92 | 488.30 | 602.80 | 424.32 | 2,064.35 |
| ARIZONA | 738.66 | 671.23 | 697.98 | 683.18 | 2,791.05 |
| ARKANSAS | 1,649.85 | 1,584.48 | 1,595.44 | 1,560.72 | 6,390.50 |
| CALIFORNIA | 820.89 | 796.51 | 804.26 | 797.76 | 3,219.43 |
| COLORADO | 309.58 | 293.24 | 304.61 | 299.92 | 1,207.34 |
| CONNECTICUT | 424.80 | 407.05 | 405.65 | 408.24 | 1,645.74 |
| DELAWARE | 688.15 | 648.34 | 703.44 | 702.57 | 2,742.50 |
| DISTRICT OF COLUMBIA | 1,201.18 | 1,058.68 | 1,041.78 | 1,124.43 | 4,426.07 |
| FLORIDA | 689.84 | 620.08 | 647.91 | 680.04 | 2,637.88 |
| GEORGIA | 792.51 | 753.88 | 770.05 | 804.58 | 3,121.02 |
| GUAM | 515.48 | 491.30 | 503.13 | 431.05 | 1,940.96 |
| HAWAII | 662.84 | 625.32 | 638.88 | 638.96 | 2,566.01 |
| IDAHO | 665.45 | 650.79 | 678.08 | 666.66 | 2,660.99 |

| | | | | | |
|---|---|---|---|---|---|
| ILLINOIS | 891.27 | 874.52 | 882.43 | 896.79 | 3,545.01 |
| INDIANA | 480.57 | 477.52 | 492.60 | 508.19 | 1,958.88 |
| IOWA | 423.76 | 387.96 | 402.79 | 386.31 | 1,600.82 |
| KANSAS | 736.25 | 755.12 | 751.11 | 758.99 | 3,001.47 |
| KENTUCKY | 797.57 | 772.10 | 775.34 | 775.82 | 3,120.84 |
| LOUISIANA | 669.35 | 639.54 | 667.39 | 680.41 | 2,656.69 |
| MAINE | 518.12 | 496.83 | 502.84 | 507.04 | 2,024.82 |
| MARYLAND | 735.45 | 659.52 | 712.70 | 720.98 | 2,828.66 |
| MASSACHUSETTS | 502.97 | 451.91 | 456.01 | 480.74 | 1,891.63 |
| MICHIGAN | 1,640.46 | 1,521.29 | 1,610.30 | 1,608.42 | 6,380.48 |
| MINNESOTA | 442.96 | 418.50 | 427.90 | 439.27 | 1,728.62 |
| MISSISSIPPI | 597.23 | 578.18 | 564.16 | 633.29 | 2,372.86 |
| MISSOURI | 947.33 | 905.18 | 899.78 | 938.49 | 3,690.78 |
| MONTANA | 721.80 | 676.85 | 677.28 | 689.17 | 2,765.10 |
| NEBRASKA | 422.10 | 432.09 | 417.58 | 425.92 | 1,697.69 |
| NEVADA | 594.59 | 581.49 | 577.34 | 568.23 | 2,321.64 |
| NEW HAMPSHIRE | 434.55 | 446.28 | 399.30 | 435.79 | 1,715.92 |
| NEW JERSEY | 639.61 | 556.96 | 635.56 | 644.83 | 2,476.96 |
| NEW MEXICO | 663.02 | 642.61 | 631.65 | 650.35 | 2,587.62 |
| NEW YORK | 537.28 | 464.91 | 486.24 | 497.50 | 1,985.93 |
| NORTH CAROLINA | 345.98 | 338.53 | 323.96 | 346.21 | 1,354.69 |
| NORTH DAKOTA | 562.01 | 485.14 | 495.22 | 544.77 | 2,087.14 |
| OHIO | 493.86 | 459.04 | 474.76 | 474.99 | 1,902.65 |
| OKLAHOMA | 1,822.58 | 1,751.56 | 1,966.14 | 1,919.66 | 7,459.95 |
| OREGON | 633.68 | 621.28 | 640.27 | 620.55 | 2,515.78 |
| PENNSYLVANIA | 649.31 | 614.33 | 630.94 | 645.38 | 2,539.96 |
| PUERTO RICO | 249.58 | 168.41 | 246.89 | 241.67 | 906.54 |
| RHODE ISLAND | 396.16 | 386.37 | 351.40 | 363.96 | 1,497.90 |
| SOUTH CAROLINA | 1,174.05 | 1,180.58 | 1,098.28 | 1,271.57 | 4,724.47 |
| SOUTH DAKOTA | 523.38 | 502.48 | 498.31 | 500.26 | 2,024.43 |
| TENNESSEE | 806.01 | 839.64 | 777.21 | 741.26 | 3,164.12 |
| TEXAS | 3,060.07 | 2,814.73 | 2,943.91 | 2,914.84 | 11,733.54 |
| UTAH | 435.62 | 425.12 | 442.59 | 433.29 | 1,736.62 |
| VERMONT | 370.45 | 328.42 | 399.81 | 332.79 | 1,431.47 |
| VIRGIN ISLANDS | 586.61 | 489.01 | 628.87 | 491.02 | 2,195.51 |
| VIRGINIA | 472.11 | 467.32 | 435.06 | 463.44 | 1,837.93 |
| WASHINGTON | 629.20 | 611.20 | 620.87 | 597.30 | 2,458.57 |
| WEST VIRGINIA | 1,852.61 | 1,783.92 | 1,859.92 | 1,839.36 | 7,335.80 |
| WISCONSIN | 601.99 | 574.68 | 592.23 | 568.63 | 2,337.52 |
| WYOMING | 503.48 | 490.22 | 508.91 | 515.34 | 2,017.95 |
| **US TOTAL** | **909.71** | **855.37** | **880.42** | **886.81** | **3,532.31** |

**DRUG: 9064 - BUPRENORPHINE**

| STATE NAME | 1ST QUARTER | 2ND QUARTER | 3RD QUARTER | 4TH QUARTER | TOTAL GRAMS |
|---|---|---|---|---|---|
| ALABAMA | 710.82 | 747.68 | 768.14 | 779.44 | 3,006.08 |
| ALASKA | 378.45 | 375.02 | 396.19 | 416.62 | 1,566.28 |
| ARIZONA | 248.64 | 241.04 | 249.86 | 258.22 | 997.76 |
| ARKANSAS | 339.82 | 365.09 | 386.25 | 394.85 | 1,486.01 |
| CALIFORNIA | 159.24 | 164.88 | 176.26 | 191.85 | 692.22 |
| COLORADO | 244.69 | 234.39 | 242.42 | 251.85 | 973.35 |
| CONNECTICUT | 536.10 | 540.10 | 529.45 | 544.28 | 2,149.93 |
| DELAWARE | 516.65 | 525.28 | 534.51 | 531.36 | 2,107.80 |
| DISTRICT OF COLUMBIA | 604.49 | 599.59 | 653.10 | 664.78 | 2,521.96 |
| FLORIDA | 336.47 | 339.18 | 346.17 | 358.82 | 1,380.64 |
| GEORGIA | 214.68 | 224.88 | 226.95 | 230.75 | 897.27 |
| GUAM | 0.00 | 0.00 | 1.67 | 0.27 | 1.95 |
| HAWAII | 172.55 | 169.58 | 185.27 | 184.01 | 711.40 |
| IDAHO | 348.74 | 373.28 | 377.97 | 385.30 | 1,485.29 |
| ILLINOIS | 166.65 | 168.94 | 173.71 | 180.30 | 689.60 |
| INDIANA | 528.52 | 554.40 | 576.80 | 595.84 | 2,255.56 |
| IOWA | 99.39 | 96.65 | 100.74 | 103.77 | 400.55 |
| KANSAS | 120.56 | 122.77 | 131.37 | 136.94 | 511.65 |

| | | | | | |
|---|---|---|---|---|---|
| KENTUCKY | 1,118.31 | 1,145.40 | 1,229.91 | 1,252.57 | 4,746.20 |
| LOUISIANA | 570.93 | 610.97 | 633.03 | 637.66 | 2,452.59 |
| MAINE | 1,144.83 | 1,144.36 | 1,185.12 | 1,209.75 | 4,684.06 |
| MARYLAND | 625.98 | 611.24 | 634.80 | 660.56 | 2,532.58 |
| MASSACHUSETTS | 780.80 | 778.05 | 768.70 | 788.05 | 3,115.59 |
| MICHIGAN | 371.54 | 380.40 | 394.32 | 408.75 | 1,555.00 |
| MINNESOTA | 233.05 | 231.40 | 239.56 | 250.34 | 954.36 |
| MISSISSIPPI | 415.46 | 431.81 | 439.19 | 447.81 | 1,734.27 |
| MISSOURI | 322.94 | 330.65 | 340.12 | 356.24 | 1,349.96 |
| MONTANA | 438.92 | 453.28 | 469.98 | 491.08 | 1,853.25 |
| NEBRASKA | 116.17 | 117.68 | 122.32 | 131.27 | 487.44 |
| NEVADA | 181.65 | 174.53 | 187.49 | 196.00 | 739.67 |
| NEW HAMPSHIRE | 845.53 | 834.17 | 856.90 | 889.64 | 3,426.24 |
| NEW JERSEY | 405.40 | 397.03 | 395.00 | 410.96 | 1,608.39 |
| NEW MEXICO | 490.91 | 498.10 | 498.89 | 493.73 | 1,981.64 |
| NEW YORK | 400.44 | 403.34 | 408.51 | 415.76 | 1,628.05 |
| NORTH CAROLINA | 580.54 | 589.49 | 613.61 | 626.98 | 2,410.61 |
| NORTH DAKOTA | 271.59 | 256.54 | 269.95 | 270.79 | 1,068.87 |
| OHIO | 564.41 | 568.29 | 582.62 | 591.15 | 2,306.47 |
| OKLAHOMA | 505.52 | 515.93 | 544.25 | 552.31 | 2,118.01 |
| OREGON | 462.09 | 456.77 | 474.08 | 465.77 | 1,858.71 |
| PENNSYLVANIA | 703.24 | 718.92 | 727.83 | 754.99 | 2,904.97 |
| PUERTO RICO | 62.61 | 52.34 | 60.26 | 61.97 | 237.17 |
| RHODE ISLAND | 878.79 | 892.64 | 829.16 | 905.42 | 3,506.02 |
| SOUTH CAROLINA | 367.33 | 390.82 | 391.75 | 416.99 | 1,566.89 |
| SOUTH DAKOTA | 148.80 | 155.17 | 166.75 | 172.14 | 642.86 |
| TENNESSEE | 721.32 | 771.77 | 794.58 | 796.92 | 3,084.59 |
| TEXAS | 132.50 | 132.31 | 136.84 | 141.95 | 543.59 |
| UTAH | 442.32 | 435.51 | 442.12 | 452.53 | 1,772.47 |
| VERMONT | 1,327.42 | 1,289.41 | 1,424.29 | 1,463.22 | 5,504.34 |
| VIRGIN ISLANDS | 25.89 | 11.97 | 27.14 | 29.08 | 94.08 |
| VIRGINIA | 356.89 | 367.65 | 374.35 | 385.11 | 1,484.00 |
| WASHINGTON | 525.11 | 523.97 | 530.09 | 537.20 | 2,116.38 |
| WEST VIRGINIA | 1,140.53 | 1,193.70 | 1,264.46 | 1,287.96 | 4,886.66 |
| WISCONSIN | 275.61 | 278.70 | 293.31 | 303.42 | 1,151.03 |
| WYOMING | 281.03 | 275.38 | 291.69 | 284.28 | 1,132.38 |
| **US TOTAL** | **383.28** | **389.66** | **400.84** | **412.60** | **1,586.37** |

**DRUG: 9143 - OXYCODONE**

| STATE NAME | 1ST QUARTER | 2ND QUARTER | 3RD QUARTER | 4TH QUARTER | TOTAL GRAMS |
|---|---|---|---|---|---|
| ALABAMA | 3,428.25 | 3,264.12 | 3,393.26 | 3,353.89 | 13,439.52 |
| ALASKA | 2,773.12 | 2,657.13 | 2,761.30 | 2,734.35 | 10,925.91 |
| AMERICAN SAMOA | 31.42 | 21.86 | 10.93 | 13.66 | 77.86 |
| ARIZONA | 4,204.63 | 4,072.41 | 4,083.49 | 4,110.74 | 16,471.27 |
| ARKANSAS | 2,731.71 | 2,607.54 | 2,620.64 | 2,601.37 | 10,561.26 |
| CALIFORNIA | 1,479.53 | 1,405.03 | 1,435.69 | 1,413.85 | 5,734.10 |
| COLORADO | 3,098.14 | 2,973.59 | 3,058.07 | 3,035.65 | 12,165.44 |
| CONNECTICUT | 3,267.63 | 3,128.79 | 3,230.22 | 3,206.66 | 12,833.31 |
| DELAWARE | 5,023.98 | 4,896.16 | 4,997.10 | 5,003.43 | 19,920.67 |
| DISTRICT OF COLUMBIA | 1,794.45 | 1,540.40 | 1,644.01 | 1,643.15 | 6,622.01 |
| FLORIDA | 4,519.11 | 4,369.98 | 4,536.37 | 4,555.99 | 17,981.44 |
| GEORGIA | 3,556.18 | 3,395.56 | 3,516.49 | 3,495.86 | 13,964.09 |
| GUAM | 376.78 | 341.36 | 420.89 | 397.08 | 1,536.10 |
| HAWAII | 2,075.95 | 1,938.55 | 1,930.56 | 1,824.23 | 7,769.29 |
| IDAHO | 2,912.80 | 2,787.56 | 2,896.12 | 2,868.87 | 11,465.35 |
| ILLINOIS | 818.24 | 773.54 | 800.55 | 804.42 | 3,196.75 |
| INDIANA | 2,607.46 | 2,518.35 | 2,597.99 | 2,617.38 | 10,341.18 |
| IOWA | 1,152.69 | 1,089.35 | 1,148.59 | 1,150.13 | 4,540.76 |
| KANSAS | 3,037.77 | 2,913.65 | 3,027.17 | 3,004.47 | 11,983.06 |
| KENTUCKY | 3,736.38 | 3,504.85 | 3,656.74 | 3,592.57 | 14,490.53 |
| LOUISIANA | 3,064.57 | 2,908.99 | 3,014.66 | 3,022.29 | 12,010.50 |
| MAINE | 2,755.61 | 2,675.20 | 2,799.45 | 2,766.24 | 10,996.50 |

| | | | | | |
|---|---|---|---|---|---|
| MARYLAND | 4,011.50 | 3,840.94 | 3,911.10 | 3,875.13 | 15,638.67 |
| MASSACHUSETTS | 2,633.72 | 2,475.41 | 2,607.95 | 2,571.64 | 10,288.71 |
| MICHIGAN | 1,907.97 | 1,792.72 | 1,844.57 | 1,856.25 | 7,401.51 |
| MINNESOTA | 1,732.83 | 1,635.46 | 1,725.90 | 1,710.81 | 6,805.00 |
| MISSISSIPPI | 2,597.30 | 2,481.46 | 2,595.21 | 2,620.52 | 10,294.49 |
| MISSOURI | 2,963.20 | 2,841.36 | 2,933.71 | 2,887.68 | 11,625.96 |
| MONTANA | 2,691.20 | 2,594.93 | 2,719.98 | 2,703.50 | 10,709.62 |
| NEBRASKA | 1,545.79 | 1,439.60 | 1,500.49 | 1,494.79 | 5,980.67 |
| NEVADA | 4,846.44 | 4,741.05 | 4,803.87 | 4,770.18 | 19,161.54 |
| NEW HAMPSHIRE | 3,192.37 | 3,026.90 | 3,182.49 | 3,186.07 | 12,587.83 |
| NEW JERSEY | 3,829.55 | 3,639.17 | 3,767.49 | 3,749.68 | 14,985.90 |
| NEW MEXICO | 3,103.24 | 2,940.54 | 2,987.82 | 2,931.03 | 11,962.64 |
| NEW YORK | 2,519.13 | 2,362.10 | 2,455.72 | 2,441.17 | 9,778.13 |
| NORTH CAROLINA | 4,452.15 | 4,290.30 | 4,438.00 | 4,415.57 | 17,596.01 |
| NORTH DAKOTA | 1,495.52 | 1,443.17 | 1,461.52 | 1,533.71 | 5,933.92 |
| OHIO | 2,658.91 | 2,528.10 | 2,659.37 | 2,655.57 | 10,501.95 |
| OKLAHOMA | 3,603.52 | 3,507.36 | 3,541.50 | 3,515.68 | 14,168.06 |
| OREGON | 3,108.78 | 3,003.70 | 3,155.69 | 3,130.48 | 12,398.66 |
| PENNSYLVANIA | 3,629.46 | 3,537.77 | 3,648.58 | 3,524.43 | 14,340.24 |
| PUERTO RICO | 307.24 | 244.98 | 303.67 | 288.32 | 1,144.20 |
| RHODE ISLAND | 2,698.01 | 2,522.48 | 2,689.83 | 2,467.20 | 10,377.52 |
| SOUTH CAROLINA | 3,448.60 | 3,288.51 | 3,400.74 | 3,429.25 | 13,567.10 |
| SOUTH DAKOTA | 1,283.97 | 1,166.08 | 1,252.08 | 1,231.18 | 4,933.30 |
| TENNESSEE | 5,011.76 | 4,946.95 | 5,029.32 | 4,887.95 | 19,875.97 |
| TEXAS | 949.58 | 943.29 | 964.45 | 908.50 | 3,765.81 |
| UTAH | 4,623.57 | 4,565.91 | 4,596.32 | 4,517.66 | 18,303.46 |
| VERMONT | 2,521.90 | 2,424.86 | 2,496.45 | 2,466.85 | 9,910.07 |
| VIRGIN ISLANDS | 392.64 | 372.64 | 340.60 | 309.58 | 1,415.46 |
| VIRGINIA | 2,707.64 | 2,584.24 | 2,634.73 | 2,576.52 | 10,503.14 |
| WASHINGTON | 3,009.42 | 2,902.37 | 3,007.99 | 2,967.74 | 11,887.52 |
| WEST VIRGINIA | 2,675.49 | 2,541.91 | 2,621.98 | 2,546.28 | 10,385.67 |
| WISCONSIN | 2,083.54 | 1,965.49 | 2,046.52 | 2,026.86 | 8,122.41 |
| WYOMING | 3,380.00 | 3,215.32 | 3,293.16 | 3,280.37 | 13,168.85 |
| **US TOTAL** | **2,698.93** | **2,587.43** | **2,669.96** | **2,643.54** | **10,599.85** |

**DRUG: 9150 - HYDROMORPHONE**

| STATE NAME | 1ST QUARTER | 2ND QUARTER | 3RD QUARTER | 4TH QUARTER | TOTAL GRAMS |
|---|---|---|---|---|---|
| ALABAMA | 46.99 | 45.58 | 48.89 | 48.85 | 190.31 |
| ALASKA | 74.10 | 72.93 | 79.20 | 70.78 | 297.01 |
| AMERICAN SAMOA | 0.81 | 0.81 | 0.27 | 0.68 | 2.57 |
| ARIZONA | 84.26 | 83.81 | 85.66 | 83.52 | 337.25 |
| ARKANSAS | 64.62 | 62.42 | 60.39 | 61.63 | 249.07 |
| CALIFORNIA | 73.99 | 71.23 | 70.66 | 68.13 | 284.02 |
| COLORADO | 117.42 | 116.03 | 115.29 | 103.94 | 452.68 |
| CONNECTICUT | 139.62 | 127.38 | 120.50 | 119.36 | 506.87 |
| DELAWARE | 140.88 | 135.26 | 131.29 | 127.43 | 534.86 |
| DISTRICT OF COLUMBIA | 74.17 | 60.49 | 63.04 | 63.36 | 261.06 |
| FLORIDA | 182.43 | 216.46 | 153.11 | 148.87 | 700.87 |
| GEORGIA | 64.70 | 64.28 | 57.31 | 56.44 | 242.73 |
| GUAM | 16.15 | 17.81 | 16.10 | 9.59 | 59.65 |
| HAWAII | 41.14 | 38.71 | 35.58 | 33.77 | 149.20 |
| IDAHO | 93.61 | 91.68 | 93.17 | 88.45 | 366.91 |
| ILLINOIS | 61.20 | 59.53 | 55.28 | 53.76 | 229.76 |
| INDIANA | 80.77 | 85.85 | 78.42 | 66.68 | 311.71 |
| IOWA | 67.14 | 62.64 | 62.02 | 62.79 | 254.59 |
| KANSAS | 94.00 | 87.54 | 86.85 | 85.87 | 354.26 |
| KENTUCKY | 57.16 | 51.66 | 50.53 | 51.59 | 210.94 |
| LOUISIANA | 52.78 | 42.51 | 48.95 | 47.97 | 192.22 |
| MAINE | 120.70 | 123.92 | 119.10 | 127.34 | 491.06 |
| MARYLAND | 105.23 | 100.56 | 95.15 | 92.34 | 393.28 |
| MASSACHUSETTS | 99.28 | 112.59 | 91.69 | 100.25 | 403.82 |
| MICHIGAN | 66.23 | 62.82 | 60.14 | 54.97 | 244.16 |

| | | | | | |
|---|---|---|---|---|---|
| MINNESOTA | 90.69 | 91.24 | 89.49 | 88.06 | 359.48 |
| MISSISSIPPI | 135.18 | 194.90 | 194.53 | 193.88 | 718.48 |
| MISSOURI | 65.13 | 64.44 | 58.52 | 62.46 | 250.55 |
| MONTANA | 145.16 | 143.44 | 146.53 | 135.99 | 571.12 |
| NEBRASKA | 50.05 | 53.48 | 51.15 | 55.84 | 210.52 |
| NEVADA | 85.70 | 89.38 | 87.26 | 84.94 | 347.27 |
| NEW HAMPSHIRE | 122.31 | 101.51 | 109.36 | 107.51 | 440.68 |
| NEW JERSEY | 81.19 | 86.09 | 73.80 | 74.62 | 315.70 |
| NEW MEXICO | 62.39 | 48.09 | 47.61 | 51.27 | 209.35 |
| NEW YORK | 84.73 | 104.00 | 69.62 | 71.73 | 330.08 |
| NORTH CAROLINA | 99.14 | 92.24 | 93.26 | 87.42 | 372.06 |
| NORTH DAKOTA | 87.42 | 68.46 | 64.14 | 61.94 | 281.96 |
| OHIO | 63.76 | 57.77 | 59.51 | 58.72 | 239.77 |
| OKLAHOMA | 77.96 | 71.37 | 72.15 | 72.76 | 294.24 |
| OREGON | 103.52 | 100.92 | 103.73 | 99.59 | 407.76 |
| PENNSYLVANIA | 84.01 | 98.66 | 82.95 | 79.96 | 345.57 |
| PUERTO RICO | 1.27 | 1.11 | 0.69 | 0.89 | 3.96 |
| RHODE ISLAND | 93.92 | 84.39 | 83.29 | 76.71 | 338.31 |
| SOUTH CAROLINA | 119.55 | 126.73 | 146.94 | 113.15 | 506.38 |
| SOUTH DAKOTA | 49.84 | 59.42 | 54.65 | 48.46 | 212.37 |
| TENNESSEE | 80.11 | 98.73 | 59.69 | 66.30 | 304.82 |
| TEXAS | 76.09 | 71.10 | 75.84 | 70.96 | 293.99 |
| UTAH | 99.03 | 87.55 | 85.41 | 84.31 | 356.30 |
| VERMONT | 167.27 | 148.30 | 148.66 | 139.32 | 603.55 |
| VIRGIN ISLANDS | 15.86 | 9.57 | 16.46 | 15.46 | 57.36 |
| VIRGINIA | 104.49 | 95.98 | 97.74 | 97.27 | 395.48 |
| WASHINGTON | 118.81 | 117.31 | 128.91 | 119.75 | 484.79 |
| WEST VIRGINIA | 55.21 | 51.14 | 53.55 | 50.34 | 210.24 |
| WISCONSIN | 68.74 | 58.81 | 58.00 | 51.38 | 236.93 |
| WYOMING | 88.48 | 104.58 | 96.98 | 91.50 | 381.53 |
| US TOTAL | 86.89 | 89.40 | 81.39 | 78.84 | 336.53 |

**DRUG: 9193 - HYDROCODONE**

| STATE NAME | 1ST QUARTER | 2ND QUARTER | 3RD QUARTER | 4TH QUARTER | TOTAL GRAMS |
|---|---|---|---|---|---|
| ALABAMA | 3,806.85 | 3,583.95 | 3,690.42 | 3,657.33 | 14,738.55 |
| ALASKA | 1,046.36 | 987.05 | 1,074.98 | 1,042.47 | 4,150.86 |
| AMERICAN SAMOA | 2.77 | 5.53 | 7.38 | 0.00 | 15.68 |
| ARIZONA | 1,139.62 | 1,094.11 | 1,107.46 | 1,116.94 | 4,458.14 |
| ARKANSAS | 3,383.73 | 3,251.83 | 3,353.12 | 3,370.32 | 13,359.00 |
| CALIFORNIA | 1,682.40 | 1,606.66 | 1,641.16 | 1,626.78 | 6,557.00 |
| COLORADO | 889.41 | 843.10 | 870.62 | 869.39 | 3,472.53 |
| CONNECTICUT | 387.32 | 355.87 | 374.31 | 362.50 | 1,480.00 |
| DELAWARE | 452.05 | 426.70 | 455.81 | 464.72 | 1,799.28 |
| DISTRICT OF COLUMBIA | 148.14 | 113.38 | 129.65 | 116.30 | 507.46 |
| FLORIDA | 1,388.67 | 1,294.37 | 1,323.84 | 1,318.02 | 5,324.90 |
| GEORGIA | 1,874.32 | 1,744.55 | 1,820.38 | 1,797.80 | 7,237.05 |
| GUAM | 57.76 | 49.37 | 60.82 | 62.43 | 230.37 |
| HAWAII | 897.13 | 864.39 | 829.93 | 821.51 | 3,412.96 |
| IDAHO | 2,748.39 | 2,699.08 | 2,751.86 | 2,768.60 | 10,967.93 |
| ILLINOIS | 1,645.33 | 1,535.08 | 1,621.34 | 1,604.33 | 6,406.09 |
| INDIANA | 2,584.48 | 2,456.14 | 2,552.48 | 2,543.91 | 10,137.00 |
| IOWA | 1,240.81 | 1,168.14 | 1,221.74 | 1,223.30 | 4,853.97 |
| KANSAS | 2,043.17 | 1,956.45 | 2,021.78 | 1,990.38 | 8,011.78 |
| KENTUCKY | 3,743.25 | 3,580.26 | 3,709.43 | 3,673.85 | 14,706.79 |
| LOUISIANA | 2,570.44 | 2,397.97 | 2,497.58 | 2,468.67 | 9,934.66 |
| MAINE | 949.27 | 913.23 | 942.96 | 936.14 | 3,741.59 |
| MARYLAND | 422.15 | 381.26 | 407.91 | 396.69 | 1,608.01 |
| MASSACHUSETTS | 329.77 | 298.57 | 316.52 | 305.46 | 1,250.31 |
| MICHIGAN | 2,448.06 | 2,325.59 | 2,412.52 | 2,406.36 | 9,592.53 |
| MINNESOTA | 542.17 | 512.30 | 541.01 | 536.32 | 2,131.80 |
| MISSISSIPPI | 2,512.24 | 2,371.09 | 2,468.03 | 2,469.80 | 9,821.15 |
| MISSOURI | 2,056.94 | 1,916.34 | 2,005.64 | 1,984.13 | 7,963.04 |

| | | | | | |
|---|---|---|---|---|---|
| MONTANA | 1,941.85 | 1,882.35 | 1,919.72 | 1,909.88 | 7,653.80 |
| NEBRASKA | 1,198.85 | 1,117.56 | 1,167.49 | 1,149.37 | 4,633.26 |
| NEVADA | 2,403.18 | 2,359.10 | 2,413.49 | 2,335.87 | 9,511.64 |
| NEW HAMPSHIRE | 516.00 | 470.49 | 494.90 | 498.03 | 1,979.41 |
| NEW JERSEY | 280.63 | 249.68 | 264.40 | 261.94 | 1,056.65 |
| NEW MEXICO | 1,245.79 | 1,183.73 | 1,221.09 | 1,210.51 | 4,861.12 |
| NEW YORK | 619.02 | 579.45 | 595.82 | 587.31 | 2,381.60 |
| NORTH CAROLINA | 1,462.17 | 1,342.21 | 1,380.59 | 1,384.63 | 5,569.60 |
| NORTH DAKOTA | 1,061.67 | 975.13 | 980.04 | 1,023.97 | 4,040.82 |
| OHIO | 1,042.75 | 993.54 | 1,042.97 | 1,036.63 | 4,115.89 |
| OKLAHOMA | 3,331.51 | 3,206.88 | 3,303.15 | 3,262.02 | 13,103.56 |
| OREGON | 1,782.01 | 1,726.36 | 1,774.66 | 1,758.49 | 7,041.52 |
| PENNSYLVANIA | 825.39 | 780.72 | 805.58 | 795.39 | 3,207.07 |
| PUERTO RICO | 10.32 | 6.26 | 7.44 | 7.08 | 31.10 |
| RHODE ISLAND | 676.24 | 644.63 | 677.17 | 641.24 | 2,639.28 |
| SOUTH CAROLINA | 2,053.20 | 1,893.35 | 1,963.29 | 1,973.76 | 7,883.59 |
| SOUTH DAKOTA | 1,173.01 | 1,042.98 | 1,105.44 | 1,088.46 | 4,409.88 |
| TENNESSEE | 2,720.53 | 2,576.73 | 2,635.43 | 2,641.02 | 10,573.71 |
| TEXAS | 1,915.80 | 1,826.06 | 1,878.51 | 1,851.45 | 7,471.82 |
| UTAH | 1,590.16 | 1,538.81 | 1,544.53 | 1,531.14 | 6,204.65 |
| VERMONT | 568.80 | 542.97 | 550.01 | 546.02 | 2,207.81 |
| VIRGIN ISLANDS | 228.30 | 126.28 | 147.23 | 137.74 | 639.55 |
| VIRGINIA | 920.43 | 839.93 | 869.45 | 855.01 | 3,484.81 |
| WASHINGTON | 1,379.82 | 1,314.93 | 1,362.09 | 1,344.66 | 5,401.50 |
| WEST VIRGINIA | 1,906.55 | 1,822.48 | 1,847.81 | 1,795.58 | 7,372.41 |
| WISCONSIN | 981.70 | 923.43 | 977.31 | 960.23 | 3,842.67 |
| WYOMING | 1,431.84 | 1,405.28 | 1,424.05 | 1,414.15 | 5,675.31 |
| US TOTAL | 1,520.93 | 1,438.28 | 1,485.33 | 1,472.87 | 5,917.42 |

**DRUG: 9230 - MEPERIDINE (PETHIDINE)**

| STATE NAME | 1ST QUARTER | 2ND QUARTER | 3RD QUARTER | 4TH QUARTER | TOTAL GRAMS |
|---|---|---|---|---|---|
| ALABAMA | 29.84 | 32.21 | 42.11 | 29.75 | 133.90 |
| ALASKA | 14.91 | 15.17 | 16.67 | 21.86 | 68.61 |
| ARIZONA | 21.77 | 20.03 | 26.94 | 26.90 | 95.63 |
| ARKANSAS | 42.99 | 39.06 | 48.53 | 43.43 | 174.00 |
| CALIFORNIA | 14.66 | 11.69 | 21.83 | 16.95 | 65.13 |
| COLORADO | 3.30 | 3.95 | 6.85 | 5.36 | 19.45 |
| CONNECTICUT | 0.83 | 0.70 | 1.78 | 1.69 | 5.00 |
| DELAWARE | 5.14 | 2.16 | 9.64 | 1.75 | 18.70 |
| DISTRICT OF COLUMBIA | 12.13 | 5.16 | 11.41 | 15.21 | 43.90 |
| FLORIDA | 9.77 | 10.13 | 12.54 | 10.33 | 42.78 |
| GEORGIA | 13.14 | 13.10 | 18.96 | 15.70 | 60.90 |
| GUAM | 6.41 | 37.85 | 21.98 | 14.65 | 80.90 |
| HAWAII | 3.82 | 3.77 | 9.02 | 5.15 | 21.76 |
| IDAHO | 12.00 | 13.58 | 9.88 | 9.49 | 44.96 |
| ILLINOIS | 3.20 | 2.68 | 4.81 | 4.25 | 14.94 |
| INDIANA | 10.62 | 7.19 | 14.46 | 18.81 | 51.08 |
| IOWA | 19.44 | 10.53 | 24.90 | 21.91 | 76.78 |
| KANSAS | 10.36 | 9.19 | 15.62 | 11.37 | 46.55 |
| KENTUCKY | 10.35 | 12.55 | 15.47 | 11.55 | 49.93 |
| LOUISIANA | 24.82 | 29.02 | 33.26 | 28.08 | 115.18 |
| MAINE | 6.42 | 4.55 | 1.89 | 5.23 | 18.10 |
| MARYLAND | 7.67 | 6.09 | 8.10 | 7.55 | 29.41 |
| MASSACHUSETTS | 4.85 | 3.24 | 6.02 | 4.28 | 18.40 |
| MICHIGAN | 3.57 | 4.00 | 6.85 | 4.78 | 19.20 |
| MINNESOTA | 2.61 | 2.21 | 2.76 | 2.07 | 9.65 |
| MISSISSIPPI | 35.58 | 35.56 | 43.54 | 37.91 | 152.59 |
| MISSOURI | 9.05 | 9.27 | 11.97 | 9.08 | 39.38 |
| MONTANA | 10.82 | 6.26 | 11.73 | 8.95 | 37.77 |
| NEBRASKA | 5.79 | 5.51 | 10.24 | 7.67 | 29.20 |
| NEVADA | 15.29 | 12.46 | 21.45 | 19.07 | 68.27 |
| NEW HAMPSHIRE | 3.14 | 3.60 | 2.20 | 2.02 | 10.96 |

| | | | | | |
|---|---|---|---|---|---|
| NEW JERSEY | 2.95 | 2.11 | 3.53 | 3.92 | 12.50 |
| NEW MEXICO | 7.87 | 9.04 | 10.12 | 9.27 | 36.30 |
| NEW YORK | 3.15 | 2.80 | 4.69 | 4.29 | 14.93 |
| NORTH CAROLINA | 5.86 | 5.92 | 8.26 | 8.11 | 28.16 |
| NORTH DAKOTA | 6.28 | 3.69 | 5.62 | 5.47 | 21.07 |
| OHIO | 8.41 | 6.83 | 15.10 | 11.24 | 41.58 |
| OKLAHOMA | 17.05 | 13.87 | 25.13 | 22.15 | 78.19 |
| OREGON | 6.47 | 6.59 | 10.15 | 10.06 | 33.27 |
| PENNSYLVANIA | 4.00 | 4.48 | 5.88 | 4.98 | 19.33 |
| PUERTO RICO | 43.27 | 35.91 | 100.51 | 57.80 | 237.50 |
| RHODE ISLAND | 4.76 | 2.18 | 3.97 | 6.02 | 16.94 |
| SOUTH CAROLINA | 10.49 | 9.33 | 12.28 | 10.89 | 42.99 |
| SOUTH DAKOTA | 10.89 | 8.60 | 12.04 | 9.26 | 40.80 |
| TENNESSEE | 16.42 | 14.87 | 20.92 | 20.22 | 72.43 |
| TEXAS | 13.87 | 12.40 | 19.59 | 14.37 | 60.23 |
| UTAH | 18.95 | 25.72 | 25.18 | 36.64 | 106.49 |
| VERMONT | 21.67 | 18.25 | 35.74 | 32.28 | 107.94 |
| VIRGIN ISLANDS | 1.98 | 5.95 | 15.37 | 20.83 | 44.14 |
| VIRGINIA | 9.35 | 7.02 | 10.65 | 9.27 | 36.28 |
| WASHINGTON | 8.42 | 7.11 | 9.85 | 8.97 | 34.35 |
| WEST VIRGINIA | 6.40 | 4.88 | 6.19 | 4.96 | 22.42 |
| WISCONSIN | 5.54 | 4.99 | 5.91 | 5.26 | 21.70 |
| WYOMING | 2.05 | 4.89 | 4.29 | 6.20 | 17.43 |
| **US TOTAL** | **10.76** | **9.77** | **15.27** | **12.59** | **48.39** |

**DRUG: 9250B - METHADONE**

| STATE NAME | 1ST QUARTER | 2ND QUARTER | 3RD QUARTER | 4TH QUARTER | TOTAL GRAMS |
|---|---|---|---|---|---|
| ALABAMA | 1,729.33 | 1,481.05 | 1,044.17 | 1,059.03 | 5,313.59 |
| ALASKA | 1,665.51 | 1,253.84 | 1,205.44 | 1,286.95 | 5,411.74 |
| ARIZONA | 2,025.67 | 1,626.88 | 1,844.35 | 1,919.98 | 7,416.89 |
| ARKANSAS | 494.62 | 346.53 | 403.48 | 395.88 | 1,640.50 |
| CALIFORNIA | 889.18 | 774.40 | 802.69 | 782.29 | 3,248.56 |
| COLORADO | 1,331.97 | 964.19 | 1,009.59 | 1,076.26 | 4,382.01 |
| CONNECTICUT | 4,368.16 | 3,026.54 | 2,990.28 | 3,509.18 | 13,894.15 |
| DELAWARE | 4,976.62 | 3,521.65 | 4,730.47 | 4,202.37 | 17,431.10 |
| DISTRICT OF COLUMBIA | 1,740.30 | 1,613.05 | 1,694.32 | 1,618.82 | 6,666.49 |
| FLORIDA | 1,227.06 | 1,049.91 | 566.09 | 580.25 | 3,423.30 |
| GEORGIA | 1,324.55 | 1,045.59 | 999.13 | 996.77 | 4,366.05 |
| GUAM | 6.02 | 6.02 | 11.28 | 6.02 | 29.32 |
| HAWAII | 600.23 | 480.78 | 503.38 | 536.54 | 2,120.94 |
| IDAHO | 770.65 | 638.17 | 688.23 | 751.02 | 2,848.07 |
| ILLINOIS | 1,110.66 | 892.14 | 916.68 | 968.87 | 3,888.35 |
| INDIANA | 1,463.74 | 1,219.66 | 1,212.90 | 1,215.96 | 5,112.26 |
| IOWA | 513.49 | 370.69 | 434.13 | 450.71 | 1,769.01 |
| KANSAS | 707.49 | 677.84 | 693.09 | 675.02 | 2,753.44 |
| KENTUCKY | 1,698.92 | 1,114.93 | 1,302.43 | 1,384.14 | 5,500.41 |
| LOUISIANA | 1,004.47 | 660.84 | 737.93 | 828.58 | 3,231.83 |
| MAINE | 2,880.43 | 1,926.89 | 2,141.55 | 2,051.22 | 9,000.09 |
| MARYLAND | 4,180.98 | 3,092.79 | 2,978.27 | 3,160.13 | 13,412.17 |
| MASSACHUSETTS | 2,749.35 | 2,183.35 | 2,289.16 | 2,455.60 | 9,677.46 |
| MICHIGAN | 1,388.96 | 1,084.87 | 1,184.37 | 1,207.92 | 4,866.11 |
| MINNESOTA | 1,195.54 | 933.26 | 857.10 | 909.84 | 3,895.74 |
| MISSISSIPPI | 316.12 | 254.47 | 169.47 | 171.41 | 911.48 |
| MISSOURI | 634.78 | 515.56 | 391.17 | 431.34 | 1,972.85 |
| MONTANA | 979.38 | 803.75 | 980.28 | 833.21 | 3,596.62 |
| NEBRASKA | 467.12 | 325.39 | 289.14 | 270.69 | 1,352.34 |
| NEVADA | 1,454.46 | 1,203.49 | 1,217.39 | 1,257.95 | 5,133.30 |
| NEW HAMPSHIRE | 2,968.40 | 2,316.52 | 1,301.86 | 1,266.43 | 7,853.21 |
| NEW JERSEY | 1,760.18 | 1,439.24 | 1,520.16 | 1,457.27 | 6,176.85 |
| NEW MEXICO | 2,965.82 | 2,152.42 | 2,136.57 | 2,045.78 | 9,300.59 |
| NEW YORK | 1,949.61 | 1,609.20 | 1,655.88 | 1,729.44 | 6,944.13 |
| NORTH CAROLINA | 1,625.46 | 1,379.58 | 1,127.26 | 1,028.88 | 5,161.18 |

| | | | | | |
|---|---|---|---|---|---|
| NORTH DAKOTA | 867.64 | 834.33 | 905.08 | 896.18 | 3,503.23 |
| OHIO | 815.82 | 744.84 | 823.77 | 866.27 | 3,250.71 |
| OKLAHOMA | 1,149.80 | 827.01 | 878.03 | 979.55 | 3,834.41 |
| OREGON | 1,843.69 | 1,558.44 | 1,587.59 | 1,507.56 | 6,497.28 |
| PENNSYLVANIA | 1,845.47 | 1,413.94 | 1,497.88 | 1,642.03 | 6,399.32 |
| PUERTO RICO | 730.32 | 608.90 | 425.69 | 628.98 | 2,393.90 |
| RHODE ISLAND | 5,861.33 | 3,156.24 | 4,156.68 | 3,804.96 | 16,979.20 |
| SOUTH CAROLINA | 1,345.51 | 1,188.46 | 1,227.26 | 1,073.73 | 4,834.96 |
| SOUTH DAKOTA | 237.75 | 182.98 | 247.90 | 195.10 | 863.74 |
| TENNESSEE | 848.04 | 653.00 | 750.85 | 790.37 | 3,042.25 |
| TEXAS | 616.69 | 479.29 | 444.19 | 455.95 | 1,996.11 |
| UTAH | 1,242.47 | 1,030.11 | 933.14 | 1,047.27 | 4,252.99 |
| VERMONT | 3,586.28 | 3,008.97 | 3,300.77 | 3,457.39 | 13,353.42 |
| VIRGIN ISLANDS | 8.96 | 12.22 | 8.14 | 16.29 | 45.61 |
| VIRGINIA | 1,381.92 | 1,154.95 | 1,075.65 | 1,109.09 | 4,721.61 |
| WASHINGTON | 1,879.98 | 1,589.23 | 1,333.08 | 1,402.76 | 6,205.05 |
| WEST VIRGINIA | 1,539.42 | 1,243.26 | 1,307.68 | 1,311.69 | 5,402.04 |
| WISCONSIN | 1,285.08 | 993.43 | 1,114.79 | 1,153.28 | 4,546.57 |
| WYOMING | 124.00 | 115.05 | 113.86 | 103.69 | 456.59 |
| **US TOTAL** | **1,399.77** | **1,117.05** | **1,094.28** | **1,125.26** | **4,736.35** |

**DRUG: 9300 - MORPHINE**

| STATE NAME | 1ST QUARTER | 2ND QUARTER | 3RD QUARTER | 4TH QUARTER | TOTAL GRAMS |
|---|---|---|---|---|---|
| ALABAMA | 1,032.62 | 966.19 | 979.08 | 956.46 | 3,934.34 |
| ALASKA | 980.27 | 949.02 | 950.79 | 893.70 | 3,773.78 |
| AMERICAN SAMOA | 10.31 | 15.01 | 0.46 | 8.82 | 34.60 |
| ARIZONA | 1,335.02 | 1,266.50 | 1,255.52 | 1,222.86 | 5,079.90 |
| ARKANSAS | 865.40 | 825.20 | 829.12 | 812.82 | 3,332.54 |
| CALIFORNIA | 758.26 | 709.79 | 705.07 | 693.96 | 2,867.08 |
| COLORADO | 1,006.44 | 963.04 | 965.09 | 961.66 | 3,896.23 |
| CONNECTICUT | 708.77 | 720.56 | 663.97 | 674.43 | 2,767.74 |
| DELAWARE | 1,599.83 | 1,497.50 | 1,547.15 | 1,556.27 | 6,200.74 |
| DISTRICT OF COLUMBIA | 450.32 | 476.49 | 404.03 | 385.22 | 1,716.07 |
| FLORIDA | 1,475.66 | 1,429.52 | 1,418.93 | 1,393.66 | 5,717.78 |
| GEORGIA | 819.79 | 796.05 | 804.49 | 779.79 | 3,200.12 |
| GUAM | 377.63 | 372.12 | 382.70 | 375.89 | 1,508.34 |
| HAWAII | 964.89 | 877.63 | 829.11 | 839.61 | 3,511.23 |
| IDAHO | 1,217.53 | 1,197.94 | 1,196.38 | 1,194.94 | 4,806.79 |
| ILLINOIS | 612.43 | 583.39 | 573.02 | 589.77 | 2,358.61 |
| INDIANA | 911.31 | 872.84 | 865.15 | 866.05 | 3,515.36 |
| IOWA | 770.55 | 737.43 | 748.40 | 769.99 | 3,026.37 |
| KANSAS | 1,079.54 | 1,034.49 | 1,032.66 | 1,075.52 | 4,222.21 |
| KENTUCKY | 732.21 | 687.05 | 689.51 | 681.34 | 2,790.10 |
| LOUISIANA | 850.20 | 785.60 | 815.09 | 763.61 | 3,214.49 |
| MAINE | 877.89 | 894.43 | 897.74 | 914.08 | 3,584.14 |
| MARYLAND | 831.96 | 813.80 | 793.40 | 773.16 | 3,212.32 |
| MASSACHUSETTS | 735.80 | 732.24 | 715.68 | 762.93 | 2,946.65 |
| MICHIGAN | 1,189.75 | 1,142.28 | 1,142.31 | 1,142.93 | 4,617.27 |
| MINNESOTA | 558.81 | 570.08 | 525.97 | 565.46 | 2,220.33 |
| MISSISSIPPI | 834.39 | 851.07 | 810.42 | 878.56 | 3,374.44 |
| MISSOURI | 1,033.58 | 1,024.06 | 1,009.22 | 1,018.29 | 4,085.15 |
| MONTANA | 1,193.17 | 1,160.67 | 1,180.41 | 1,145.64 | 4,679.89 |
| NEBRASKA | 766.86 | 742.74 | 749.64 | 775.35 | 3,034.59 |
| NEVADA | 1,346.94 | 1,267.49 | 1,286.27 | 1,228.83 | 5,129.53 |
| NEW HAMPSHIRE | 1,019.81 | 1,006.62 | 987.07 | 1,021.41 | 4,034.91 |
| NEW JERSEY | 674.14 | 640.76 | 638.50 | 632.29 | 2,585.69 |
| NEW MEXICO | 865.34 | 813.38 | 835.24 | 842.89 | 3,356.85 |
| NEW YORK | 561.68 | 551.92 | 532.40 | 531.81 | 2,177.81 |
| NORTH CAROLINA | 971.01 | 929.71 | 932.03 | 921.04 | 3,753.79 |
| NORTH DAKOTA | 669.91 | 628.52 | 581.09 | 679.76 | 2,559.28 |
| OHIO | 685.08 | 664.96 | 670.18 | 689.09 | 2,709.32 |
| OKLAHOMA | 1,240.24 | 1,188.18 | 1,184.33 | 1,173.96 | 4,786.71 |

| | | | | | |
|---|---|---|---|---|---|
| OREGON | 1,271.13 | 1,226.16 | 1,246.75 | 1,216.75 | 4,960.80 |
| PENNSYLVANIA | 1,130.13 | 1,187.56 | 1,116.16 | 1,106.24 | 4,540.09 |
| PUERTO RICO | 41.65 | 32.05 | 39.87 | 37.88 | 151.45 |
| RHODE ISLAND | 769.44 | 751.20 | 782.62 | 716.95 | 3,020.21 |
| SOUTH CAROLINA | 1,085.34 | 1,024.05 | 1,027.26 | 1,036.83 | 4,173.48 |
| SOUTH DAKOTA | 662.63 | 619.09 | 617.27 | 638.22 | 2,537.20 |
| TENNESSEE | 1,502.81 | 1,547.00 | 1,567.14 | 1,523.26 | 6,140.22 |
| TEXAS | 581.45 | 538.24 | 556.26 | 551.73 | 2,227.68 |
| UTAH | 1,286.27 | 1,268.95 | 1,268.90 | 1,227.56 | 5,051.68 |
| VERMONT | 1,217.18 | 1,167.57 | 1,103.14 | 1,164.38 | 4,652.27 |
| VIRGIN ISLANDS | 65.15 | 58.34 | 62.96 | 62.10 | 248.55 |
| VIRGINIA | 731.15 | 706.04 | 716.01 | 702.52 | 2,855.72 |
| WASHINGTON | 1,092.89 | 1,049.70 | 1,077.90 | 1,036.96 | 4,257.45 |
| WEST VIRGINIA | 751.28 | 755.29 | 755.02 | 754.66 | 3,016.25 |
| WISCONSIN | 892.72 | 857.31 | 866.82 | 868.02 | 3,484.87 |
| WYOMING | 1,109.76 | 1,033.40 | 1,069.44 | 1,091.32 | 4,303.91 |
| **US TOTAL** | **884.08** | **854.54** | **850.22** | **844.54** | **3,433.37** |
| **DRUG: 9652 - OXYMORPHONE** | | | | | |
| STATE NAME | 1ST QUARTER | 2ND QUARTER | 3RD QUARTER | 4TH QUARTER | TOTAL GRAMS |
| ALABAMA | 21.73 | 19.63 | 19.21 | 16.72 | 77.30 |
| ALASKA | 5.31 | 4.18 | 3.69 | 3.09 | 16.28 |
| ARIZONA | 18.48 | 16.49 | 16.66 | 15.69 | 67.31 |
| ARKANSAS | 4.37 | 3.97 | 3.92 | 2.91 | 15.17 |
| CALIFORNIA | 8.31 | 7.61 | 7.28 | 7.23 | 30.43 |
| COLORADO | 16.86 | 14.88 | 15.64 | 14.64 | 62.02 |
| CONNECTICUT | 23.69 | 23.06 | 19.97 | 20.18 | 86.90 |
| DELAWARE | 94.72 | 81.58 | 75.50 | 74.07 | 325.87 |
| DISTRICT OF COLUMBIA | 8.69 | 5.66 | 6.26 | 5.14 | 25.75 |
| FLORIDA | 31.79 | 28.33 | 27.02 | 25.98 | 113.11 |
| GEORGIA | 18.57 | 17.17 | 15.82 | 13.87 | 65.44 |
| HAWAII | 9.99 | 11.73 | 8.54 | 9.64 | 39.89 |
| IDAHO | 31.58 | 27.91 | 27.78 | 26.06 | 113.33 |
| ILLINOIS | 10.22 | 7.94 | 8.06 | 8.16 | 34.39 |
| INDIANA | 18.17 | 14.94 | 13.83 | 13.94 | 60.87 |
| IOWA | 6.53 | 6.62 | 6.80 | 5.98 | 25.94 |
| KANSAS | 17.60 | 15.90 | 16.60 | 14.07 | 64.17 |
| KENTUCKY | 31.24 | 28.55 | 22.35 | 21.93 | 104.08 |
| LOUISIANA | 47.53 | 40.59 | 39.35 | 34.99 | 162.46 |
| MAINE | 18.62 | 18.24 | 17.12 | 16.47 | 70.44 |
| MARYLAND | 51.17 | 41.11 | 38.27 | 37.04 | 167.59 |
| MASSACHUSETTS | 10.40 | 9.80 | 9.78 | 9.89 | 39.86 |
| MICHIGAN | 54.15 | 49.74 | 40.75 | 36.97 | 181.61 |
| MINNESOTA | 3.11 | 2.92 | 3.08 | 2.54 | 11.66 |
| MISSISSIPPI | 20.37 | 18.54 | 19.93 | 19.14 | 77.98 |
| MISSOURI | 19.78 | 19.30 | 18.41 | 17.36 | 74.84 |
| MONTANA | 8.42 | 7.96 | 7.68 | 5.92 | 29.99 |
| NEBRASKA | 16.64 | 14.50 | 14.18 | 13.25 | 58.57 |
| NEVADA | 32.47 | 25.81 | 25.18 | 25.84 | 109.30 |
| NEW HAMPSHIRE | 13.42 | 11.99 | 13.05 | 11.33 | 49.79 |
| NEW JERSEY | 23.24 | 20.46 | 20.36 | 19.32 | 83.39 |
| NEW MEXICO | 11.06 | 11.05 | 10.25 | 10.52 | 42.88 |
| NEW YORK | 28.02 | 23.84 | 23.80 | 23.15 | 98.80 |
| NORTH CAROLINA | 59.51 | 53.13 | 49.66 | 47.80 | 210.10 |
| NORTH DAKOTA | 6.58 | 5.19 | 7.47 | 10.33 | 29.56 |
| OHIO | 18.73 | 16.80 | 16.17 | 15.09 | 66.79 |
| OKLAHOMA | 19.12 | 17.55 | 12.88 | 12.29 | 61.83 |
| OREGON | 13.69 | 12.30 | 11.90 | 11.15 | 49.03 |
| PENNSYLVANIA | 46.69 | 40.28 | 39.50 | 37.58 | 164.06 |
| RHODE ISLAND | 14.81 | 7.71 | 11.08 | 11.83 | 45.43 |
| SOUTH CAROLINA | 43.05 | 38.72 | 40.60 | 37.52 | 159.89 |
| SOUTH DAKOTA | 20.54 | 15.60 | 17.16 | 16.92 | 70.22 |

| | | | | | |
|---|---|---|---|---|---|
| TENNESSEE | 94.08 | 75.84 | 70.90 | 67.96 | 308.78 |
| TEXAS | 9.17 | 8.44 | 8.41 | 7.56 | 33.58 |
| UTAH | 69.42 | 61.16 | 61.70 | 54.74 | 247.03 |
| VERMONT | 13.34 | 12.06 | 14.21 | 12.28 | 51.89 |
| VIRGINIA | 40.01 | 32.60 | 33.12 | 29.38 | 135.11 |
| WASHINGTON | 12.96 | 12.55 | 13.16 | 12.84 | 51.50 |
| WEST VIRGINIA | 26.98 | 24.46 | 21.25 | 19.81 | 92.50 |
| WISCONSIN | 8.23 | 7.11 | 6.92 | 6.70 | 28.95 |
| WYOMING | 11.49 | 12.05 | 10.71 | 9.55 | 43.80 |
| **US TOTAL** | **24.38** | **21.37** | **20.42** | **19.31** | **85.48** |

**DRUG: 9780 - TAPENTADOL**

| STATE NAME | 1ST QUARTER | 2ND QUARTER | 3RD QUARTER | 4TH QUARTER | TOTAL GRAMS |
|---|---|---|---|---|---|
| ALABAMA | 540.41 | 512.73 | 510.58 | 478.74 | 2,042.46 |
| ALASKA | 187.62 | 200.85 | 226.41 | 222.11 | 836.98 |
| ARIZONA | 334.62 | 307.58 | 307.12 | 325.20 | 1,274.52 |
| ARKANSAS | 290.66 | 277.05 | 286.19 | 305.74 | 1,159.63 |
| CALIFORNIA | 159.08 | 153.23 | 152.82 | 150.40 | 615.53 |
| COLORADO | 412.86 | 389.96 | 408.49 | 425.03 | 1,636.35 |
| CONNECTICUT | 178.46 | 166.60 | 166.52 | 173.55 | 685.14 |
| DELAWARE | 493.35 | 414.12 | 390.73 | 431.71 | 1,729.92 |
| DISTRICT OF COLUMBIA | 89.16 | 95.73 | 73.29 | 81.77 | 339.94 |
| FLORIDA | 291.37 | 276.82 | 273.71 | 277.71 | 1,119.61 |
| GEORGIA | 420.18 | 390.92 | 399.86 | 389.40 | 1,600.36 |
| HAWAII | 96.78 | 91.89 | 87.55 | 90.27 | 366.50 |
| IDAHO | 254.28 | 242.51 | 261.17 | 273.29 | 1,031.24 |
| ILLINOIS | 165.99 | 146.86 | 149.44 | 159.46 | 621.75 |
| INDIANA | 362.37 | 342.65 | 346.31 | 348.14 | 1,399.46 |
| IOWA | 224.55 | 201.37 | 206.46 | 194.43 | 826.81 |
| KANSAS | 154.22 | 152.46 | 160.42 | 161.86 | 628.96 |
| KENTUCKY | 179.81 | 174.24 | 187.45 | 176.24 | 717.73 |
| LOUISIANA | 238.30 | 226.19 | 229.22 | 224.25 | 917.96 |
| MAINE | 76.37 | 76.60 | 83.41 | 68.47 | 304.85 |
| MARYLAND | 383.07 | 363.60 | 376.39 | 363.27 | 1,486.33 |
| MASSACHUSETTS | 230.46 | 225.54 | 217.21 | 207.14 | 880.34 |
| MICHIGAN | 125.21 | 118.70 | 118.75 | 116.51 | 479.17 |
| MINNESOTA | 53.42 | 53.35 | 53.81 | 54.70 | 215.27 |
| MISSISSIPPI | 240.03 | 226.28 | 233.95 | 229.15 | 929.41 |
| MISSOURI | 251.15 | 244.09 | 250.70 | 243.32 | 989.26 |
| MONTANA | 121.79 | 111.73 | 110.87 | 104.15 | 448.55 |
| NEBRASKA | 393.77 | 388.10 | 416.05 | 441.16 | 1,639.07 |
| NEVADA | 208.01 | 212.25 | 202.64 | 211.96 | 834.87 |
| NEW HAMPSHIRE | 393.25 | 443.12 | 402.52 | 431.49 | 1,670.38 |
| NEW JERSEY | 314.11 | 300.68 | 305.56 | 303.62 | 1,223.97 |
| NEW MEXICO | 89.36 | 86.59 | 87.90 | 84.52 | 348.37 |
| NEW YORK | 299.40 | 288.97 | 290.82 | 295.15 | 1,174.34 |
| NORTH CAROLINA | 730.40 | 707.87 | 725.50 | 711.15 | 2,874.91 |
| NORTH DAKOTA | 157.60 | 149.72 | 125.86 | 160.87 | 594.05 |
| OHIO | 280.21 | 270.32 | 270.44 | 267.37 | 1,088.35 |
| OKLAHOMA | 194.21 | 197.44 | 182.71 | 177.54 | 751.89 |
| OREGON | 111.93 | 102.28 | 102.20 | 103.38 | 419.79 |
| PENNSYLVANIA | 317.63 | 301.64 | 309.89 | 297.82 | 1,226.97 |
| PUERTO RICO | 7.80 | 6.83 | 8.82 | 9.92 | 33.36 |
| RHODE ISLAND | 147.21 | 153.10 | 148.02 | 136.76 | 585.09 |
| SOUTH CAROLINA | 542.41 | 514.14 | 527.03 | 542.63 | 2,126.21 |
| SOUTH DAKOTA | 318.54 | 304.85 | 302.39 | 273.53 | 1,199.30 |
| TENNESSEE | 336.64 | 329.89 | 318.76 | 329.05 | 1,314.34 |
| TEXAS | 210.57 | 200.70 | 208.12 | 205.23 | 824.62 |
| UTAH | 292.11 | 295.91 | 286.95 | 279.70 | 1,154.66 |
| VERMONT | 121.30 | 107.87 | 106.03 | 136.64 | 471.84 |
| VIRGIN ISLANDS | 6.83 | 13.66 | 13.66 | 6.83 | 40.97 |
| VIRGINIA | 304.10 | 302.22 | 302.86 | 290.74 | 1,199.92 |

| | | | | | |
|---|---|---|---|---|---|
| WASHINGTON | 163.74 | 156.56 | 162.81 | 158.46 | 641.56 |
| WEST VIRGINIA | 187.62 | 180.68 | 180.92 | 172.96 | 722.18 |
| WISCONSIN | 130.99 | 128.42 | 136.72 | 134.65 | 530.78 |
| WYOMING | 180.97 | 166.78 | 173.87 | 160.83 | 682.46 |
| US TOTAL | 262.45 | 251.14 | 253.98 | 252.67 | 1,020.24 |

**DRUG: 9801 - FENTANYL BASE**

| STATE NAME | 1ST QUARTER | 2ND QUARTER | 3RD QUARTER | 4TH QUARTER | TOTAL GRAMS |
|---|---|---|---|---|---|
| ALABAMA | 16.55 | 16.25 | 15.76 | 16.19 | 64.74 |
| ALASKA | 13.90 | 11.92 | 11.52 | 11.23 | 48.57 |
| AMERICAN SAMOA | 0.16 | 0.12 | 0.04 | 0.04 | 0.36 |
| ARIZONA | 12.90 | 12.53 | 11.13 | 12.03 | 48.59 |
| ARKANSAS | 14.97 | 14.07 | 14.51 | 14.37 | 57.92 |
| CALIFORNIA | 9.86 | 9.55 | 9.25 | 9.67 | 38.34 |
| COLORADO | 13.21 | 13.47 | 12.72 | 13.09 | 52.48 |
| CONNECTICUT | 14.19 | 14.12 | 13.41 | 14.56 | 56.28 |
| DELAWARE | 16.59 | 15.22 | 14.69 | 13.76 | 60.26 |
| DISTRICT OF COLUMBIA | 8.78 | 11.17 | 8.11 | 6.86 | 34.92 |
| FLORIDA | 18.48 | 18.69 | 16.48 | 15.37 | 69.01 |
| GEORGIA | 16.29 | 17.10 | 16.49 | 15.38 | 65.25 |
| GUAM | 5.07 | 4.85 | 4.57 | 5.49 | 19.98 |
| HAWAII | 8.37 | 7.46 | 7.91 | 7.43 | 31.17 |
| IDAHO | 22.43 | 20.74 | 21.15 | 21.01 | 85.33 |
| ILLINOIS | 13.41 | 13.71 | 12.33 | 12.55 | 51.99 |
| INDIANA | 16.34 | 15.16 | 15.41 | 15.10 | 62.01 |
| IOWA | 17.88 | 16.59 | 16.09 | 16.03 | 66.59 |
| KANSAS | 21.50 | 20.35 | 20.12 | 20.39 | 82.35 |
| KENTUCKY | 14.92 | 14.48 | 13.38 | 13.72 | 56.49 |
| LOUISIANA | 14.86 | 13.63 | 13.44 | 13.56 | 55.49 |
| MAINE | 17.33 | 16.47 | 15.55 | 15.30 | 64.66 |
| MARYLAND | 12.18 | 12.74 | 10.85 | 11.27 | 47.05 |
| MASSACHUSETTS | 12.43 | 13.47 | 11.02 | 11.31 | 48.24 |
| MICHIGAN | 16.12 | 16.67 | 15.03 | 15.80 | 63.62 |
| MINNESOTA | 11.90 | 10.34 | 10.38 | 10.43 | 43.06 |
| MISSISSIPPI | 25.93 | 30.53 | 26.06 | 25.51 | 108.03 |
| MISSOURI | 19.03 | 18.85 | 18.89 | 21.69 | 78.45 |
| MONTANA | 20.04 | 19.00 | 19.30 | 20.18 | 78.51 |
| NEBRASKA | 18.48 | 17.13 | 16.50 | 16.62 | 68.73 |
| NEVADA | 11.02 | 15.28 | 13.61 | 12.68 | 52.58 |
| NEW HAMPSHIRE | 15.20 | 14.70 | 14.18 | 15.02 | 59.10 |
| NEW JERSEY | 14.43 | 15.21 | 12.36 | 13.14 | 55.13 |
| NEW MEXICO | 12.04 | 11.20 | 10.84 | 11.64 | 45.72 |
| NEW YORK | 13.26 | 15.15 | 11.52 | 11.46 | 51.38 |
| NORTH CAROLINA | 16.63 | 16.02 | 15.68 | 15.71 | 64.04 |
| NORTH DAKOTA | 21.33 | 19.91 | 16.66 | 20.36 | 78.26 |
| OHIO | 11.25 | 11.07 | 9.72 | 10.59 | 42.63 |
| OKLAHOMA | 17.02 | 15.54 | 15.44 | 15.31 | 63.31 |
| OREGON | 13.06 | 12.46 | 12.56 | 12.51 | 50.59 |
| PENNSYLVANIA | 18.64 | 19.65 | 18.71 | 18.80 | 75.79 |
| PUERTO RICO | 4.61 | 3.40 | 4.68 | 4.39 | 17.08 |
| RHODE ISLAND | 9.97 | 10.41 | 10.17 | 9.46 | 40.02 |
| SOUTH CAROLINA | 15.88 | 15.28 | 14.96 | 14.64 | 60.76 |
| SOUTH DAKOTA | 19.09 | 18.83 | 17.77 | 18.00 | 73.69 |
| TENNESSEE | 13.56 | 16.15 | 17.17 | 15.99 | 62.88 |
| TEXAS | 12.69 | 11.88 | 12.92 | 12.46 | 49.95 |
| UTAH | 19.14 | 18.69 | 17.83 | 17.69 | 73.35 |
| VERMONT | 16.08 | 14.85 | 15.66 | 15.49 | 62.07 |
| VIRGIN ISLANDS | 3.51 | 4.76 | 3.69 | 4.34 | 16.30 |
| VIRGINIA | 12.74 | 12.80 | 11.59 | 11.57 | 48.70 |
| WASHINGTON | 12.86 | 11.92 | 11.99 | 12.44 | 49.21 |
| WEST VIRGINIA | 15.93 | 15.29 | 14.56 | 15.60 | 61.37 |
| WISCONSIN | 14.51 | 13.08 | 12.69 | 12.69 | 52.97 |

| | | | | | |
|---|---|---|---|---|---|
| WYOMING | 15.51 | 15.60 | 15.42 | 15.92 | 62.44 |
| **US TOTAL** | **14.21** | **14.24** | **13.36** | **13.45** | **55.27** |

**DRUG: 1100 - AMPHETAMINE**

| RANK | STATE NAME | STATE POPULATION | TOTAL GRAMS | TOTAL GRAMS/100K POPULATION |
|---|---|---|---|---|
| 1 | LOUISIANA | 4,533,372 | 723,555.98 | 15,960.66 |
| 2 | RHODE ISLAND | 1,052,567 | 161,267.23 | 15,321.33 |
| 3 | UTAH | 2,763,885 | 420,098.76 | 15,199.57 |
| 4 | NEW HAMPSHIRE | 1,316,470 | 181,152.17 | 13,760.45 |
| 5 | SOUTH CAROLINA | 4,625,364 | 607,858.44 | 13,141.85 |
| 6 | MASSACHUSETTS | 6,547,629 | 770,566.96 | 11,768.64 |
| 7 | ALABAMA | 4,779,736 | 557,604.40 | 11,666.01 |
| 8 | MISSOURI | 5,988,927 | 636,650.88 | 10,630.47 |
| 9 | MICHIGAN | 9,883,640 | 1,029,380.82 | 10,415.00 |
| 10 | NORTH CAROLINA | 9,535,483 | 951,347.95 | 9,976.92 |
| 11 | KANSAS | 2,853,118 | 283,674.35 | 9,942.61 |
| 12 | DELAWARE | 897,934 | 88,099.48 | 9,811.35 |
| 13 | VIRGINIA | 8,001,024 | 774,323.52 | 9,677.81 |
| 14 | INDIANA | 6,483,802 | 601,210.59 | 9,272.50 |
| 15 | WISCONSIN | 5,686,986 | 519,247.69 | 9,130.45 |
| 16 | MINNESOTA | 5,303,925 | 483,927.32 | 9,123.95 |
| 17 | TENNESSEE | 6,346,105 | 576,511.88 | 9,084.50 |
| 18 | MISSISSIPPI | 2,967,297 | 262,776.43 | 8,855.75 |
| 19 | GEORGIA | 9,687,653 | 855,054.81 | 8,826.23 |
| 20 | IDAHO | 1,567,582 | 135,934.28 | 8,671.59 |
| 21 | MAINE | 1,328,361 | 110,589.22 | 8,325.24 |
| 22 | COLORADO | 5,029,196 | 411,494.11 | 8,182.11 |
| 23 | DISTRICT OF COLUMBIA | 601,723 | 48,781.43 | 8,106.96 |
| 24 | TEXAS | 25,145,561 | 2,030,422.85 | 8,074.68 |
| 25 | ARKANSAS | 2,915,918 | 232,719.27 | 7,980.99 |
| 26 | FLORIDA | 18,801,310 | 1,468,251.43 | 7,809.30 |
| 27 | OHIO | 11,536,504 | 893,498.21 | 7,744.97 |
| 28 | OREGON | 3,831,074 | 293,985.95 | 7,673.72 |
| 29 | WASHINGTON | 6,724,540 | 507,102.69 | 7,541.08 |
| 30 | IOWA | 3,046,355 | 228,285.38 | 7,493.72 |
| 31 | KENTUCKY | 4,339,367 | 316,791.97 | 7,300.42 |
| 32 | MARYLAND | 5,773,552 | 418,721.06 | 7,252.40 |
| 33 | PENNSYLVANIA | 12,702,379 | 920,185.86 | 7,244.20 |
| 34 | CONNECTICUT | 3,574,097 | 256,745.82 | 7,183.52 |
| 35 | VERMONT | 625,741 | 44,540.51 | 7,118.04 |
| 36 | SOUTH DAKOTA | 814,180 | 57,553.56 | 7,068.90 |
| 37 | MONTANA | 989,415 | 69,225.61 | 6,996.62 |
| 38 | NEBRASKA | 1,826,341 | 126,586.80 | 6,931.17 |
| 39 | ILLINOIS | 12,830,632 | 863,813.33 | 6,732.43 |
| 40 | ARIZONA | 6,392,017 | 418,266.00 | 6,543.57 |
| 41 | OKLAHOMA | 3,751,351 | 236,918.03 | 6,315.54 |
| 42 | NORTH DAKOTA | 672,591 | 41,470.66 | 6,165.81 |
| 43 | NEW JERSEY | 8,791,894 | 494,484.40 | 5,624.32 |
| 44 | WYOMING | 563,626 | 31,283.34 | 5,550.37 |
| 45 | NEW YORK | 19,378,102 | 1,069,301.05 | 5,518.09 |
| 46 | WEST VIRGINIA | 1,852,994 | 94,988.86 | 5,126.24 |
| 47 | ALASKA | 710,231 | 35,551.91 | 5,005.68 |
| 48 | NEVADA | 2,700,551 | 122,817.25 | 4,547.86 |
| 49 | CALIFORNIA | 37,253,956 | 1,289,177.62 | 3,460.51 |
| 50 | NEW MEXICO | 2,059,179 | 67,551.14 | 3,280.49 |
| 51 | HAWAII | 1,360,301 | 44,234.60 | 3,251.82 |
| 52 | VIRGIN ISLANDS | 109,825 | 1,486.11 | 1,353.16 |
| 53 | GUAM | 178,430 | 1,873.50 | 1,049.99 |

| | | | | |
|---|---|---|---|---|
| 54 | PUERTO RICO | 3,725,789 | 31,993.24 | 858.70 |
| 55 | AMERICAN SAMOA | 65,628 | 3.75 | 5.72 |
| **US TOTAL** | | **312,825,210** | 23,900,940.45 | **7,640.35** |

**DRUG: 1105 - METHAMPHETAMINE**

| RANK | STATE NAME | STATE POPULATION | TOTAL GRAMS | TOTAL GRAMS/100K POPULATION |
|---|---|---|---|---|
| 1 | WASHINGTON | 6,724,540 | 302.89 | 4.50 |
| 2 | DISTRICT OF COLUMBIA | 601,723 | 26.92 | 4.47 |
| 3 | OREGON | 3,831,074 | 163.51 | 4.27 |
| 4 | UTAH | 2,763,885 | 106.85 | 3.87 |
| 5 | MINNESOTA | 5,303,925 | 196.06 | 3.70 |
| 6 | WYOMING | 563,626 | 19.68 | 3.49 |
| 7 | SOUTH DAKOTA | 814,180 | 26.91 | 3.31 |
| 8 | COLORADO | 5,029,196 | 164.72 | 3.28 |
| 9 | IDAHO | 1,567,582 | 51.02 | 3.25 |
| 10 | MISSOURI | 5,988,927 | 190.42 | 3.18 |
| 11 | PENNSYLVANIA | 12,702,379 | 367.23 | 2.89 |
| 12 | MAINE | 1,328,361 | 36.16 | 2.72 |
| 13 | ALABAMA | 4,779,736 | 124.94 | 2.61 |
| 14 | NORTH DAKOTA | 672,591 | 16.47 | 2.45 |
| 15 | CALIFORNIA | 37,253,956 | 901.54 | 2.42 |
| 16 | DELAWARE | 897,934 | 21.29 | 2.37 |
| 17 | MARYLAND | 5,773,552 | 135.80 | 2.35 |
| 18 | CONNECTICUT | 3,574,097 | 82.76 | 2.32 |
| 19 | IOWA | 3,046,355 | 60.26 | 1.98 |
| 20 | NEW HAMPSHIRE | 1,316,470 | 24.51 | 1.86 |
| 21 | ARIZONA | 6,392,017 | 118.52 | 1.85 |
| 22 | NEW MEXICO | 2,059,179 | 36.56 | 1.78 |
| 23 | NEW YORK | 19,378,102 | 333.06 | 1.72 |
| 24 | RHODE ISLAND | 1,052,567 | 17.68 | 1.68 |
| 25 | INDIANA | 6,483,802 | 105.66 | 1.63 |
| 26 | NORTH CAROLINA | 9,535,483 | 143.02 | 1.50 |
| 27 | KANSAS | 2,853,118 | 42.18 | 1.48 |
| 28 | WISCONSIN | 5,686,986 | 82.77 | 1.46 |
| 29 | TEXAS | 25,145,561 | 356.75 | 1.42 |
| 30 | MASSACHUSETTS | 6,547,629 | 90.80 | 1.39 |
| 31 | NEW JERSEY | 8,791,894 | 118.12 | 1.34 |
| 32 | NEVADA | 2,700,551 | 33.75 | 1.25 |
| 33 | VIRGINIA | 8,001,024 | 98.84 | 1.24 |
| 34 | NEBRASKA | 1,826,341 | 22.50 | 1.23 |
| 35 | VERMONT | 625,741 | 7.63 | 1.22 |
| 36 | LOUISIANA | 4,533,372 | 54.24 | 1.20 |
| 37 | OKLAHOMA | 3,751,351 | 43.39 | 1.16 |
| 38 | ILLINOIS | 12,830,632 | 147.83 | 1.15 |
| 39 | FLORIDA | 18,801,310 | 206.91 | 1.10 |
| 40 | GEORGIA | 9,687,653 | 103.25 | 1.07 |
| 41 | OHIO | 11,536,504 | 122.13 | 1.06 |
| 42 | HAWAII | 1,360,301 | 13.66 | 1.00 |
| 43 | MISSISSIPPI | 2,967,297 | 28.92 | 0.97 |
| 44 | TENNESSEE | 6,346,105 | 61.47 | 0.97 |
| 45 | KENTUCKY | 4,339,367 | 39.37 | 0.91 |
| 46 | ALASKA | 710,231 | 6.43 | 0.90 |
| 47 | SOUTH CAROLINA | 4,625,364 | 30.13 | 0.65 |
| 48 | WEST VIRGINIA | 1,852,994 | 11.65 | 0.63 |

| | | | | |
|---|---|---|---|---|
| 49 | MICHIGAN | 9,883,640 | 59.86 | 0.61 |
| 50 | ARKANSAS | 2,915,918 | 10.85 | 0.37 |
| 51 | MONTANA | 989,415 | 2.01 | 0.20 |
| **US TOTAL** | | **312,825,210** | **5,569.91** | **1.78** |

**DRUG: 1205 - LISDEXAMFETAMINE**

| RANK | STATE NAME | STATE POPULATION | TOTAL GRAMS | TOTAL GRAMS/100K POPULATION |
|---|---|---|---|---|
| 1 | LOUISIANA | 4,533,372 | 340,144.79 | 7,503.13 |
| 2 | DELAWARE | 897,934 | 52,266.60 | 5,820.76 |
| 3 | ALABAMA | 4,779,736 | 278,056.85 | 5,817.41 |
| 4 | WISCONSIN | 5,686,986 | 313,755.57 | 5,517.08 |
| 5 | MAINE | 1,328,361 | 71,100.52 | 5,352.50 |
| 6 | SOUTH CAROLINA | 4,625,364 | 236,873.27 | 5,121.18 |
| 7 | VERMONT | 625,741 | 30,956.68 | 4,947.20 |
| 8 | INDIANA | 6,483,802 | 317,643.40 | 4,899.03 |
| 9 | MISSISSIPPI | 2,967,297 | 135,063.82 | 4,551.75 |
| 10 | NORTH CAROLINA | 9,535,483 | 428,905.11 | 4,497.99 |
| 11 | KANSAS | 2,853,118 | 128,197.72 | 4,493.25 |
| 12 | NEW HAMPSHIRE | 1,316,470 | 58,671.37 | 4,456.72 |
| 13 | MISSOURI | 5,988,927 | 264,838.59 | 4,422.14 |
| 14 | SOUTH DAKOTA | 814,180 | 35,655.47 | 4,379.31 |
| 15 | IOWA | 3,046,355 | 131,017.69 | 4,300.80 |
| 16 | CONNECTICUT | 3,574,097 | 144,910.59 | 4,054.47 |
| 17 | TENNESSEE | 6,346,105 | 255,058.88 | 4,019.14 |
| 18 | NEBRASKA | 1,826,341 | 72,497.79 | 3,969.56 |
| 19 | MARYLAND | 5,773,552 | 229,091.20 | 3,967.94 |
| 20 | OKLAHOMA | 3,751,351 | 145,985.86 | 3,891.55 |
| 21 | TEXAS | 25,145,561 | 975,228.69 | 3,878.33 |
| 22 | WEST VIRGINIA | 1,852,994 | 70,806.84 | 3,821.21 |
| 23 | NORTH DAKOTA | 672,591 | 25,563.58 | 3,800.76 |
| 24 | DISTRICT OF COLUMBIA | 601,723 | 22,700.83 | 3,772.64 |
| 25 | GEORGIA | 9,687,653 | 364,339.43 | 3,760.86 |
| 26 | ARKANSAS | 2,915,918 | 107,714.48 | 3,694.02 |
| 27 | KENTUCKY | 4,339,367 | 149,205.27 | 3,438.41 |
| 28 | UTAH | 2,763,885 | 93,904.83 | 3,397.57 |
| 29 | VIRGINIA | 8,001,024 | 268,812.45 | 3,359.73 |
| 30 | OHIO | 11,536,504 | 365,516.44 | 3,168.35 |
| 31 | MONTANA | 989,415 | 28,803.83 | 2,911.20 |
| 32 | RHODE ISLAND | 1,052,567 | 30,517.90 | 2,899.38 |
| 33 | PENNSYLVANIA | 12,702,379 | 361,896.38 | 2,849.04 |
| 34 | FLORIDA | 18,801,310 | 531,227.07 | 2,825.48 |
| 35 | IDAHO | 1,567,582 | 43,829.81 | 2,796.01 |
| 36 | ARIZONA | 6,392,017 | 167,601.02 | 2,622.04 |
| 37 | MINNESOTA | 5,303,925 | 138,336.44 | 2,608.19 |
| 38 | MASSACHUSETTS | 6,547,629 | 164,157.24 | 2,507.12 |
| 39 | MICHIGAN | 9,883,640 | 247,109.42 | 2,500.19 |
| 40 | COLORADO | 5,029,196 | 124,526.21 | 2,476.07 |
| 41 | ILLINOIS | 12,830,632 | 299,184.09 | 2,331.80 |
| 42 | WASHINGTON | 6,724,540 | 151,132.10 | 2,247.47 |
| 43 | NEW JERSEY | 8,791,894 | 189,644.55 | 2,157.04 |
| 44 | WYOMING | 563,626 | 11,257.34 | 1,997.31 |
| 45 | NEW YORK | 19,378,102 | 346,205.56 | 1,786.58 |
| 46 | ALASKA | 710,231 | 12,581.77 | 1,771.50 |
| 47 | OREGON | 3,831,074 | 65,466.93 | 1,708.84 |

| | | | | |
|---|---|---|---|---|
| 48 | CALIFORNIA | 37,253,956 | 450,812.79 | 1,210.11 |
| 49 | NEVADA | 2,700,551 | 30,054.26 | 1,112.89 |
| 50 | NEW MEXICO | 2,059,179 | 14,817.86 | 719.60 |
| 51 | HAWAII | 1,360,301 | 7,511.25 | 552.18 |
| 52 | VIRGIN ISLANDS | 109,825 | 306.39 | 278.98 |
| 53 | PUERTO RICO | 3,725,789 | 7,769.09 | 208.52 |
| 54 | GUAM | 178,430 | 75.73 | 42.44 |
| **US TOTAL** | | **312,825,210** | 9,539,309.67 | **3,049.41** |

**DRUG: 1724 - METHYLPHENIDATE (DL;D;L;ISOMERS)**

| RANK | STATE NAME | STATE POPULATION | TOTAL GRAMS | TOTAL GRAMS/100K POPULATION |
|---|---|---|---|---|
| 1 | VERMONT | 625,741 | 74,351.15 | 11,882.10 |
| 2 | MAINE | 1,328,361 | 140,169.85 | 10,552.09 |
| 3 | NEW HAMPSHIRE | 1,316,470 | 134,352.15 | 10,205.49 |
| 4 | IOWA | 3,046,355 | 302,417.24 | 9,927.18 |
| 5 | MISSOURI | 5,988,927 | 524,204.02 | 8,752.89 |
| 6 | NORTH DAKOTA | 672,591 | 58,027.91 | 8,627.52 |
| 7 | SOUTH DAKOTA | 814,180 | 70,002.56 | 8,597.92 |
| 8 | MASSACHUSETTS | 6,547,629 | 529,424.49 | 8,085.74 |
| 9 | DELAWARE | 897,934 | 65,620.81 | 7,307.98 |
| 10 | MINNESOTA | 5,303,925 | 387,094.81 | 7,298.27 |
| 11 | NORTH CAROLINA | 9,535,483 | 693,068.50 | 7,268.31 |
| 12 | IDAHO | 1,567,582 | 113,200.75 | 7,221.36 |
| 13 | UTAH | 2,763,885 | 195,641.04 | 7,078.48 |
| 14 | WISCONSIN | 5,686,986 | 401,781.02 | 7,064.92 |
| 15 | SOUTH CAROLINA | 4,625,364 | 326,772.50 | 7,064.80 |
| 16 | INDIANA | 6,483,802 | 446,877.14 | 6,892.21 |
| 17 | MONTANA | 989,415 | 67,264.56 | 6,798.42 |
| 18 | KANSAS | 2,853,118 | 192,268.80 | 6,738.90 |
| 19 | NEBRASKA | 1,826,341 | 121,773.51 | 6,667.62 |
| 20 | PENNSYLVANIA | 12,702,379 | 846,525.81 | 6,664.31 |
| 21 | WASHINGTON | 6,724,540 | 439,294.64 | 6,532.71 |
| 22 | OREGON | 3,831,074 | 249,890.03 | 6,522.71 |
| 23 | RHODE ISLAND | 1,052,567 | 68,584.81 | 6,515.96 |
| 24 | MARYLAND | 5,773,552 | 375,792.69 | 6,508.86 |
| 25 | OHIO | 11,536,504 | 749,338.89 | 6,495.37 |
| 26 | MICHIGAN | 9,883,640 | 639,475.55 | 6,470.04 |
| 27 | KENTUCKY | 4,339,367 | 277,773.39 | 6,401.24 |
| 28 | LOUISIANA | 4,533,372 | 281,148.12 | 6,201.74 |
| 29 | VIRGINIA | 8,001,024 | 487,284.55 | 6,090.28 |
| 30 | GEORGIA | 9,687,653 | 583,038.80 | 6,018.37 |
| 31 | WEST VIRGINIA | 1,852,994 | 108,127.56 | 5,835.29 |
| 32 | CONNECTICUT | 3,574,097 | 207,374.56 | 5,802.15 |
| 33 | ALABAMA | 4,779,736 | 274,841.27 | 5,750.14 |
| 34 | MISSISSIPPI | 2,967,297 | 155,802.50 | 5,250.65 |
| 35 | WYOMING | 563,626 | 29,435.46 | 5,222.52 |
| 36 | COLORADO | 5,029,196 | 251,056.84 | 4,991.99 |
| 37 | TENNESSEE | 6,346,105 | 313,980.10 | 4,947.60 |
| 38 | DISTRICT OF COLUMBIA | 601,723 | 29,736.72 | 4,941.93 |
| 39 | ARKANSAS | 2,915,918 | 141,661.62 | 4,858.22 |
| 40 | TEXAS | 25,145,561 | 1,211,939.57 | 4,819.70 |
| 41 | ILLINOIS | 12,830,632 | 614,278.40 | 4,787.59 |
| 42 | OKLAHOMA | 3,751,351 | 177,410.86 | 4,729.25 |
| 43 | ARIZONA | 6,392,017 | 295,664.47 | 4,625.53 |

| | | | | |
|---|---|---|---|---|
| 44 | NEW YORK | 19,378,102 | 835,371.46 | 4,310.90 |
| 45 | ALASKA | 710,231 | 30,520.19 | 4,297.22 |
| 46 | FLORIDA | 18,801,310 | 762,977.76 | 4,058.11 |
| 47 | NEW JERSEY | 8,791,894 | 343,566.33 | 3,907.76 |
| 48 | NEW MEXICO | 2,059,179 | 66,949.84 | 3,251.29 |
| 49 | HAWAII | 1,360,301 | 38,163.16 | 2,805.49 |
| 50 | CALIFORNIA | 37,253,956 | 981,779.10 | 2,635.37 |
| 51 | NEVADA | 2,700,551 | 58,253.51 | 2,157.10 |
| 52 | PUERTO RICO | 3,725,789 | 41,213.58 | 1,106.17 |
| 53 | GUAM | 178,430 | 1,658.75 | 929.64 |
| 54 | VIRGIN ISLANDS | 109,825 | 390.37 | 355.45 |
| 55 | AMERICAN SAMOA | 65,628 | 15.57 | 23.72 |
| **US TOTAL** | | **312,825,210** | 16,814,629.65 | **5,375.09** |

**DRUG: 9050 - CODEINE**

| RANK | STATE NAME | STATE POPULATION | TOTAL GRAMS | TOTAL GRAMS/100K POPULATION |
|---|---|---|---|---|
| 1 | TEXAS | 25,145,561 | 2,950,465.05 | 11,733.54 |
| 2 | OKLAHOMA | 3,751,351 | 279,848.77 | 7,459.95 |
| 3 | WEST VIRGINIA | 1,852,994 | 135,932.01 | 7,335.80 |
| 4 | ARKANSAS | 2,915,918 | 186,341.73 | 6,390.50 |
| 5 | MICHIGAN | 9,883,640 | 630,623.41 | 6,380.48 |
| 6 | SOUTH CAROLINA | 4,625,364 | 218,523.90 | 4,724.47 |
| 7 | DISTRICT OF COLUMBIA | 601,723 | 26,632.70 | 4,426.07 |
| 8 | MISSOURI | 5,988,927 | 221,037.90 | 3,690.78 |
| 9 | ILLINOIS | 12,830,632 | 454,847.09 | 3,545.01 |
| 10 | ALABAMA | 4,779,736 | 168,182.16 | 3,518.65 |
| 11 | CALIFORNIA | 37,253,956 | 1,199,366.88 | 3,219.43 |
| 12 | TENNESSEE | 6,346,105 | 200,798.23 | 3,164.12 |
| 13 | GEORGIA | 9,687,653 | 302,353.76 | 3,121.02 |
| 14 | KENTUCKY | 4,339,367 | 135,424.61 | 3,120.84 |
| 15 | KANSAS | 2,853,118 | 85,635.60 | 3,001.47 |
| 16 | MARYLAND | 5,773,552 | 163,313.92 | 2,828.66 |
| 17 | ARIZONA | 6,392,017 | 178,404.18 | 2,791.05 |
| 18 | MONTANA | 989,415 | 27,358.30 | 2,765.10 |
| 19 | DELAWARE | 897,934 | 24,625.84 | 2,742.50 |
| 20 | IDAHO | 1,567,582 | 41,713.13 | 2,660.99 |
| 21 | LOUISIANA | 4,533,372 | 120,437.60 | 2,656.69 |
| 22 | FLORIDA | 18,801,310 | 495,955.54 | 2,637.88 |
| 23 | NEW MEXICO | 2,059,179 | 53,283.78 | 2,587.62 |
| 24 | ALASKA | 710,231 | 18,259.96 | 2,570.99 |
| 25 | HAWAII | 1,360,301 | 34,905.49 | 2,566.01 |
| 26 | PENNSYLVANIA | 12,702,379 | 322,635.48 | 2,539.96 |
| 27 | OREGON | 3,831,074 | 96,381.58 | 2,515.78 |
| 28 | NEW JERSEY | 8,791,894 | 217,771.67 | 2,476.96 |
| 29 | WASHINGTON | 6,724,540 | 165,327.56 | 2,458.57 |
| 30 | MISSISSIPPI | 2,967,297 | 70,409.74 | 2,372.86 |
| 31 | WISCONSIN | 5,686,986 | 132,934.53 | 2,337.52 |
| 32 | NEVADA | 2,700,551 | 62,696.95 | 2,321.64 |
| 33 | VIRGIN ISLANDS | 109,825 | 2,411.22 | 2,195.51 |
| 34 | NORTH DAKOTA | 672,591 | 14,037.91 | 2,087.14 |
| 35 | AMERICAN SAMOA | 65,628 | 1,354.79 | 2,064.35 |
| 36 | MAINE | 1,328,361 | 26,896.94 | 2,024.82 |
| 37 | SOUTH DAKOTA | 814,180 | 16,482.53 | 2,024.43 |
| 38 | WYOMING | 563,626 | 11,373.68 | 2,017.95 |

| | | | | |
|---|---|---|---|---|
| 39 | NEW YORK | 19,378,102 | 384,834.80 | 1,985.93 |
| 40 | INDIANA | 6,483,802 | 127,009.76 | 1,958.88 |
| 41 | GUAM | 178,430 | 3,463.26 | 1,940.96 |
| 42 | OHIO | 11,536,504 | 219,499.06 | 1,902.65 |
| 43 | MASSACHUSETTS | 6,547,629 | 123,857.02 | 1,891.63 |
| 44 | VIRGINIA | 8,001,024 | 147,053.55 | 1,837.93 |
| 45 | UTAH | 2,763,885 | 47,998.14 | 1,736.62 |
| 46 | MINNESOTA | 5,303,925 | 91,684.63 | 1,728.62 |
| 47 | NEW HAMPSHIRE | 1,316,470 | 22,589.58 | 1,715.92 |
| 48 | NEBRASKA | 1,826,341 | 31,005.70 | 1,697.69 |
| 49 | CONNECTICUT | 3,574,097 | 58,820.26 | 1,645.74 |
| 50 | IOWA | 3,046,355 | 48,766.74 | 1,600.82 |
| 51 | RHODE ISLAND | 1,052,567 | 15,766.40 | 1,497.90 |
| 52 | VERMONT | 625,741 | 8,957.27 | 1,431.47 |
| 53 | NORTH CAROLINA | 9,535,483 | 129,175.78 | 1,354.69 |
| 54 | COLORADO | 5,029,196 | 60,719.42 | 1,207.34 |
| 55 | PUERTO RICO | 3,725,789 | 33,775.85 | 906.54 |
| US TOTAL | | 312,825,210 | 11,049,963.34 | 3,532.31 |

**DRUG: 9064 - BUPRENORPHINE**

| RANK | STATE NAME | STATE POPULATION | TOTAL GRAMS | TOTAL GRAMS/100K POPULATION |
|---|---|---|---|---|
| 1 | VERMONT | 625,741 | 34,442.93 | 5,504.34 |
| 2 | WEST VIRGINIA | 1,852,994 | 90,549.56 | 4,886.66 |
| 3 | KENTUCKY | 4,339,367 | 205,955.04 | 4,746.20 |
| 4 | MAINE | 1,328,361 | 62,221.29 | 4,684.06 |
| 5 | RHODE ISLAND | 1,052,567 | 36,903.18 | 3,506.02 |
| 6 | NEW HAMPSHIRE | 1,316,470 | 45,105.37 | 3,426.24 |
| 7 | MASSACHUSETTS | 6,547,629 | 203,997.54 | 3,115.59 |
| 8 | TENNESSEE | 6,346,105 | 195,751.34 | 3,084.59 |
| 9 | ALABAMA | 4,779,736 | 143,682.51 | 3,006.08 |
| 10 | PENNSYLVANIA | 12,702,379 | 369,000.54 | 2,904.97 |
| 11 | MARYLAND | 5,773,552 | 146,219.99 | 2,532.58 |
| 12 | DISTRICT OF COLUMBIA | 601,723 | 15,175.24 | 2,521.96 |
| 13 | LOUISIANA | 4,533,372 | 111,185.20 | 2,452.59 |
| 14 | NORTH CAROLINA | 9,535,483 | 229,863.45 | 2,410.61 |
| 15 | OHIO | 11,536,504 | 266,085.76 | 2,306.47 |
| 16 | INDIANA | 6,483,802 | 146,246.28 | 2,255.56 |
| 17 | CONNECTICUT | 3,574,097 | 76,840.48 | 2,149.93 |
| 18 | OKLAHOMA | 3,751,351 | 79,453.99 | 2,118.01 |
| 19 | WASHINGTON | 6,724,540 | 142,317.00 | 2,116.38 |
| 20 | DELAWARE | 897,934 | 18,926.66 | 2,107.80 |
| 21 | NEW MEXICO | 2,059,179 | 40,805.54 | 1,981.64 |
| 22 | OREGON | 3,831,074 | 71,208.49 | 1,858.71 |
| 23 | MONTANA | 989,415 | 18,336.37 | 1,853.25 |
| 24 | UTAH | 2,763,885 | 48,989.01 | 1,772.47 |
| 25 | MISSISSIPPI | 2,967,297 | 51,461.04 | 1,734.27 |
| 26 | NEW YORK | 19,378,102 | 315,485.16 | 1,628.05 |
| 27 | NEW JERSEY | 8,791,894 | 141,407.92 | 1,608.39 |
| 28 | SOUTH CAROLINA | 4,625,364 | 72,474.56 | 1,566.89 |
| 29 | ALASKA | 710,231 | 11,124.21 | 1,566.28 |
| 30 | MICHIGAN | 9,883,640 | 153,690.78 | 1,555.00 |
| 31 | ARKANSAS | 2,915,918 | 43,330.91 | 1,486.01 |
| 32 | IDAHO | 1,567,582 | 23,283.15 | 1,485.29 |
| 33 | VIRGINIA | 8,001,024 | 118,734.95 | 1,484.00 |

| | | | | |
|---|---|---|---|---|
| 34 | FLORIDA | 18,801,310 | 259,578.64 | 1,380.64 |
| 35 | MISSOURI | 5,988,927 | 80,847.92 | 1,349.96 |
| 36 | WISCONSIN | 5,686,986 | 65,458.99 | 1,151.03 |
| 37 | WYOMING | 563,626 | 6,382.40 | 1,132.38 |
| 38 | NORTH DAKOTA | 672,591 | 7,189.14 | 1,068.87 |
| 39 | ARIZONA | 6,392,017 | 63,777.24 | 997.76 |
| 40 | COLORADO | 5,029,196 | 48,951.62 | 973.35 |
| 41 | MINNESOTA | 5,303,925 | 50,618.41 | 954.36 |
| 42 | GEORGIA | 9,687,653 | 86,924.31 | 897.27 |
| 43 | NEVADA | 2,700,551 | 19,975.11 | 739.67 |
| 44 | HAWAII | 1,360,301 | 9,677.20 | 711.40 |
| 45 | CALIFORNIA | 37,253,956 | 257,880.21 | 692.22 |
| 46 | ILLINOIS | 12,830,632 | 88,479.83 | 689.60 |
| 47 | SOUTH DAKOTA | 814,180 | 5,234.06 | 642.86 |
| 48 | TEXAS | 25,145,561 | 136,687.79 | 543.59 |
| 49 | KANSAS | 2,853,118 | 14,597.88 | 511.65 |
| 50 | NEBRASKA | 1,826,341 | 8,902.33 | 487.44 |
| 51 | IOWA | 3,046,355 | 12,202.26 | 400.55 |
| 52 | PUERTO RICO | 3,725,789 | 8,836.44 | 237.17 |
| 53 | VIRGIN ISLANDS | 109,825 | 103.33 | 94.08 |
| 54 | GUAM | 178,430 | 3.47 | 1.95 |
| **US TOTAL** | | **312,825,210** | 4,962,564.02 | **1,586.37** |

**DRUG: 9143 - OXYCODONE**

| RANK | STATE NAME | STATE POPULATION | TOTAL GRAMS | TOTAL GRAMS/100K POPULATION |
|---|---|---|---|---|
| 1 | DELAWARE | 897,934 | 178,874.50 | 19,920.67 |
| 2 | TENNESSEE | 6,346,105 | 1,261,349.93 | 19,875.97 |
| 3 | NEVADA | 2,700,551 | 517,467.24 | 19,161.54 |
| 4 | UTAH | 2,763,885 | 505,886.63 | 18,303.46 |
| 5 | FLORIDA | 18,801,310 | 3,380,747.01 | 17,981.44 |
| 6 | NORTH CAROLINA | 9,535,483 | 1,677,864.97 | 17,596.01 |
| 7 | ARIZONA | 6,392,017 | 1,052,846.65 | 16,471.27 |
| 8 | MARYLAND | 5,773,552 | 902,906.87 | 15,638.67 |
| 9 | NEW JERSEY | 8,791,894 | 1,317,544.01 | 14,985.90 |
| 10 | KENTUCKY | 4,339,367 | 628,797.49 | 14,490.53 |
| 11 | PENNSYLVANIA | 12,702,379 | 1,821,551.43 | 14,340.24 |
| 12 | OKLAHOMA | 3,751,351 | 531,493.77 | 14,168.06 |
| 13 | GEORGIA | 9,687,653 | 1,352,792.10 | 13,964.09 |
| 14 | SOUTH CAROLINA | 4,625,364 | 627,527.78 | 13,567.10 |
| 15 | ALABAMA | 4,779,736 | 642,373.62 | 13,439.52 |
| 16 | WYOMING | 563,626 | 74,223.05 | 13,168.85 |
| 17 | CONNECTICUT | 3,574,097 | 458,674.80 | 12,833.31 |
| 18 | NEW HAMPSHIRE | 1,316,470 | 165,714.99 | 12,587.83 |
| 19 | OREGON | 3,831,074 | 475,001.70 | 12,398.66 |
| 20 | COLORADO | 5,029,196 | 611,824.06 | 12,165.44 |
| 21 | LOUISIANA | 4,533,372 | 544,480.77 | 12,010.50 |
| 22 | KANSAS | 2,853,118 | 341,890.72 | 11,983.06 |
| 23 | NEW MEXICO | 2,059,179 | 246,332.07 | 11,962.64 |
| 24 | WASHINGTON | 6,724,540 | 799,381.08 | 11,887.52 |
| 25 | MISSOURI | 5,988,927 | 696,270.13 | 11,625.96 |
| 26 | IDAHO | 1,567,582 | 179,728.81 | 11,465.35 |
| 27 | MAINE | 1,328,361 | 146,073.21 | 10,996.50 |
| 28 | ALASKA | 710,231 | 77,599.20 | 10,925.91 |
| 29 | MONTANA | 989,415 | 105,962.55 | 10,709.62 |

| | | | | |
|---|---|---|---|---|
| 30 | ARKANSAS | 2,915,918 | 307,957.64 | 10,561.26 |
| 31 | VIRGINIA | 8,001,024 | 840,358.73 | 10,503.14 |
| 32 | OHIO | 11,536,504 | 1,211,558.44 | 10,501.95 |
| 33 | WEST VIRGINIA | 1,852,994 | 192,445.76 | 10,385.67 |
| 34 | RHODE ISLAND | 1,052,567 | 109,230.32 | 10,377.52 |
| 35 | INDIANA | 6,483,802 | 670,501.70 | 10,341.18 |
| 36 | MISSISSIPPI | 2,967,297 | 305,468.13 | 10,294.49 |
| 37 | MASSACHUSETTS | 6,547,629 | 673,666.61 | 10,288.71 |
| 38 | VERMONT | 625,741 | 62,011.36 | 9,910.07 |
| 39 | NEW YORK | 19,378,102 | 1,894,815.55 | 9,778.13 |
| 40 | WISCONSIN | 5,686,986 | 461,920.37 | 8,122.41 |
| 41 | HAWAII | 1,360,301 | 105,685.78 | 7,769.29 |
| 42 | MICHIGAN | 9,883,640 | 731,538.25 | 7,401.51 |
| 43 | MINNESOTA | 5,303,925 | 360,932.12 | 6,805.00 |
| 44 | DISTRICT OF COLUMBIA | 601,723 | 39,846.18 | 6,622.01 |
| 45 | NEBRASKA | 1,826,341 | 109,227.36 | 5,980.67 |
| 46 | NORTH DAKOTA | 672,591 | 39,911.02 | 5,933.92 |
| 47 | CALIFORNIA | 37,253,956 | 2,136,178.98 | 5,734.10 |
| 48 | SOUTH DAKOTA | 814,180 | 40,165.91 | 4,933.30 |
| 49 | IOWA | 3,046,355 | 138,327.77 | 4,540.76 |
| 50 | TEXAS | 25,145,561 | 946,934.86 | 3,765.81 |
| 51 | ILLINOIS | 12,830,632 | 410,163.59 | 3,196.75 |
| 52 | GUAM | 178,430 | 2,740.87 | 1,536.10 |
| 53 | VIRGIN ISLANDS | 109,825 | 1,554.53 | 1,415.46 |
| 54 | PUERTO RICO | 3,725,789 | 42,630.59 | 1,144.20 |
| 55 | AMERICAN SAMOA | 65,628 | 51.10 | 77.86 |
| **US TOTAL** | | **312,825,210** | 33,159,004.69 | **10,599.85** |

**DRUG: 9150 - HYDROMORPHONE**

| RANK | STATE NAME | STATE POPULATION | TOTAL GRAMS | TOTAL GRAMS/100K POPULATION |
|---|---|---|---|---|
| 1 | MISSISSIPPI | 2,967,297 | 21,319.53 | 718.48 |
| 2 | FLORIDA | 18,801,310 | 131,773.51 | 700.87 |
| 3 | VERMONT | 625,741 | 3,776.68 | 603.55 |
| 4 | MONTANA | 989,415 | 5,650.73 | 571.12 |
| 5 | DELAWARE | 897,934 | 4,802.71 | 534.86 |
| 6 | CONNECTICUT | 3,574,097 | 18,116.11 | 506.87 |
| 7 | SOUTH CAROLINA | 4,625,364 | 23,421.69 | 506.38 |
| 8 | MAINE | 1,328,361 | 6,523.07 | 491.06 |
| 9 | WASHINGTON | 6,724,540 | 32,599.82 | 484.79 |
| 10 | COLORADO | 5,029,196 | 22,765.94 | 452.68 |
| 11 | NEW HAMPSHIRE | 1,316,470 | 5,801.43 | 440.68 |
| 12 | OREGON | 3,831,074 | 15,621.67 | 407.76 |
| 13 | MASSACHUSETTS | 6,547,629 | 26,440.52 | 403.82 |
| 14 | VIRGINIA | 8,001,024 | 31,642.67 | 395.48 |
| 15 | MARYLAND | 5,773,552 | 22,706.44 | 393.28 |
| 16 | WYOMING | 563,626 | 2,150.41 | 381.53 |
| 17 | NORTH CAROLINA | 9,535,483 | 35,477.56 | 372.06 |
| 18 | IDAHO | 1,567,582 | 5,751.55 | 366.91 |
| 19 | MINNESOTA | 5,303,925 | 19,066.30 | 359.48 |
| 20 | UTAH | 2,763,885 | 9,847.83 | 356.30 |
| 21 | KANSAS | 2,853,118 | 10,107.53 | 354.26 |
| 22 | NEVADA | 2,700,551 | 9,378.32 | 347.27 |
| 23 | PENNSYLVANIA | 12,702,379 | 43,895.86 | 345.57 |
| 24 | RHODE ISLAND | 1,052,567 | 3,560.99 | 338.31 |

| | | | | |
|---|---|---|---|---|
| 25 | ARIZONA | 6,392,017 | 21,557.34 | 337.25 |
| 26 | NEW YORK | 19,378,102 | 63,963.71 | 330.08 |
| 27 | NEW JERSEY | 8,791,894 | 27,755.80 | 315.70 |
| 28 | INDIANA | 6,483,802 | 20,210.86 | 311.71 |
| 29 | TENNESSEE | 6,346,105 | 19,344.41 | 304.82 |
| 30 | ALASKA | 710,231 | 2,109.49 | 297.01 |
| 31 | OKLAHOMA | 3,751,351 | 11,037.97 | 294.24 |
| 32 | TEXAS | 25,145,561 | 73,925.07 | 293.99 |
| 33 | CALIFORNIA | 37,253,956 | 105,806.98 | 284.02 |
| 34 | NORTH DAKOTA | 672,591 | 1,896.45 | 281.96 |
| 35 | DISTRICT OF COLUMBIA | 601,723 | 1,570.87 | 261.06 |
| 36 | IOWA | 3,046,355 | 7,755.82 | 254.59 |
| 37 | MISSOURI | 5,988,927 | 15,005.29 | 250.55 |
| 38 | ARKANSAS | 2,915,918 | 7,262.57 | 249.07 |
| 39 | MICHIGAN | 9,883,640 | 24,132.18 | 244.16 |
| 40 | GEORGIA | 9,687,653 | 23,514.51 | 242.73 |
| 41 | OHIO | 11,536,504 | 27,660.54 | 239.77 |
| 42 | WISCONSIN | 5,686,986 | 13,474.18 | 236.93 |
| 43 | ILLINOIS | 12,830,632 | 29,479.33 | 229.76 |
| 44 | SOUTH DAKOTA | 814,180 | 1,729.06 | 212.37 |
| 45 | KENTUCKY | 4,339,367 | 9,153.39 | 210.94 |
| 46 | NEBRASKA | 1,826,341 | 3,844.78 | 210.52 |
| 47 | WEST VIRGINIA | 1,852,994 | 3,895.65 | 210.24 |
| 48 | NEW MEXICO | 2,059,179 | 4,310.97 | 209.35 |
| 49 | LOUISIANA | 4,533,372 | 8,713.88 | 192.22 |
| 50 | ALABAMA | 4,779,736 | 9,096.16 | 190.31 |
| 51 | HAWAII | 1,360,301 | 2,029.61 | 149.20 |
| 52 | GUAM | 178,430 | 106.44 | 59.65 |
| 53 | VIRGIN ISLANDS | 109,825 | 62.99 | 57.36 |
| 54 | PUERTO RICO | 3,725,789 | 147.40 | 3.96 |
| 55 | AMERICAN SAMOA | 65,628 | 1.68 | 2.57 |
| **US TOTAL** | | **312,825,210** | 1,052,754.29 | **336.53** |

**DRUG: 9193 - HYDROCODONE**

| RANK | STATE NAME | STATE POPULATION | TOTAL GRAMS | TOTAL GRAMS/100K POPULATION |
|---|---|---|---|---|
| 1 | ALABAMA | 4,779,736 | 704,463.87 | 14,738.55 |
| 2 | KENTUCKY | 4,339,367 | 638,181.81 | 14,706.79 |
| 3 | ARKANSAS | 2,915,918 | 389,537.35 | 13,359.00 |
| 4 | OKLAHOMA | 3,751,351 | 491,560.56 | 13,103.56 |
| 5 | IDAHO | 1,567,582 | 171,931.31 | 10,967.93 |
| 6 | TENNESSEE | 6,346,105 | 671,018.86 | 10,573.71 |
| 7 | INDIANA | 6,483,802 | 657,262.81 | 10,137.00 |
| 8 | LOUISIANA | 4,533,372 | 450,374.96 | 9,934.66 |
| 9 | MISSISSIPPI | 2,967,297 | 291,422.83 | 9,821.15 |
| 10 | MICHIGAN | 9,883,640 | 948,090.89 | 9,592.53 |
| 11 | NEVADA | 2,700,551 | 256,866.63 | 9,511.64 |
| 12 | KANSAS | 2,853,118 | 228,585.45 | 8,011.78 |
| 13 | MISSOURI | 5,988,927 | 476,900.71 | 7,963.04 |
| 14 | SOUTH CAROLINA | 4,625,364 | 364,644.86 | 7,883.59 |
| 15 | MONTANA | 989,415 | 75,727.82 | 7,653.80 |
| 16 | TEXAS | 25,145,561 | 1,878,831.00 | 7,471.82 |
| 17 | WEST VIRGINIA | 1,852,994 | 136,610.40 | 7,372.41 |
| 18 | GEORGIA | 9,687,653 | 701,100.50 | 7,237.05 |
| 19 | OREGON | 3,831,074 | 269,765.75 | 7,041.52 |

| | | | | |
|---|---|---|---|---|
| 20 | CALIFORNIA | 37,253,956 | 2,442,742.71 | 6,557.00 |
| 21 | ILLINOIS | 12,830,632 | 821,941.50 | 6,406.09 |
| 22 | UTAH | 2,763,885 | 171,489.29 | 6,204.65 |
| 23 | WYOMING | 563,626 | 31,987.54 | 5,675.31 |
| 24 | NORTH CAROLINA | 9,535,483 | 531,088.32 | 5,569.60 |
| 25 | WASHINGTON | 6,724,540 | 363,226.10 | 5,401.50 |
| 26 | FLORIDA | 18,801,310 | 1,001,150.34 | 5,324.90 |
| 27 | NEW MEXICO | 2,059,179 | 100,099.20 | 4,861.12 |
| 28 | IOWA | 3,046,355 | 147,869.30 | 4,853.97 |
| 29 | NEBRASKA | 1,826,341 | 84,619.19 | 4,633.26 |
| 30 | ARIZONA | 6,392,017 | 284,965.03 | 4,458.14 |
| 31 | SOUTH DAKOTA | 814,180 | 35,904.38 | 4,409.88 |
| 32 | ALASKA | 710,231 | 29,480.70 | 4,150.86 |
| 33 | OHIO | 11,536,504 | 474,830.27 | 4,115.89 |
| 34 | NORTH DAKOTA | 672,591 | 27,178.17 | 4,040.82 |
| 35 | WISCONSIN | 5,686,986 | 218,532.35 | 3,842.67 |
| 36 | MAINE | 1,328,361 | 49,701.88 | 3,741.59 |
| 37 | VIRGINIA | 8,001,024 | 278,820.72 | 3,484.81 |
| 38 | COLORADO | 5,029,196 | 174,640.12 | 3,472.53 |
| 39 | HAWAII | 1,360,301 | 46,426.58 | 3,412.96 |
| 40 | PENNSYLVANIA | 12,702,379 | 407,374.34 | 3,207.07 |
| 41 | RHODE ISLAND | 1,052,567 | 27,780.18 | 2,639.28 |
| 42 | NEW YORK | 19,378,102 | 461,509.04 | 2,381.60 |
| 43 | VERMONT | 625,741 | 13,815.17 | 2,207.81 |
| 44 | MINNESOTA | 5,303,925 | 113,068.82 | 2,131.80 |
| 45 | NEW HAMPSHIRE | 1,316,470 | 26,058.35 | 1,979.41 |
| 46 | DELAWARE | 897,934 | 16,156.38 | 1,799.28 |
| 47 | MARYLAND | 5,773,552 | 92,839.54 | 1,608.01 |
| 48 | CONNECTICUT | 3,574,097 | 52,896.58 | 1,480.00 |
| 49 | MASSACHUSETTS | 6,547,629 | 81,865.98 | 1,250.31 |
| 50 | NEW JERSEY | 8,791,894 | 92,899.23 | 1,056.65 |
| 51 | VIRGIN ISLANDS | 109,825 | 702.39 | 639.55 |
| 52 | DISTRICT OF COLUMBIA | 601,723 | 3,053.53 | 507.46 |
| 53 | GUAM | 178,430 | 411.05 | 230.37 |
| 54 | PUERTO RICO | 3,725,789 | 1,158.90 | 31.10 |
| 55 | AMERICAN SAMOA | 65,628 | 10.29 | 15.68 |
| **US TOTAL** | | **312,825,210** | 18,511,171.84 | **5,917.42** |

**DRUG: 9230 - MEPERIDINE (PETHIDINE)**

| RANK | STATE NAME | STATE POPULATION | TOTAL GRAMS | TOTAL GRAMS/100K POPULATION |
|---|---|---|---|---|
| 1 | PUERTO RICO | 3,725,789 | 8,848.78 | 237.50 |
| 2 | ARKANSAS | 2,915,918 | 5,073.76 | 174.00 |
| 3 | MISSISSIPPI | 2,967,297 | 4,527.88 | 152.59 |
| 4 | ALABAMA | 4,779,736 | 6,400.08 | 133.90 |
| 5 | LOUISIANA | 4,533,372 | 5,221.37 | 115.18 |
| 6 | VERMONT | 625,741 | 675.41 | 107.94 |
| 7 | UTAH | 2,763,885 | 2,943.16 | 106.49 |
| 8 | ARIZONA | 6,392,017 | 6,112.90 | 95.63 |
| 9 | GUAM | 178,430 | 144.34 | 80.90 |
| 10 | OKLAHOMA | 3,751,351 | 2,933.36 | 78.19 |
| 11 | IOWA | 3,046,355 | 2,338.89 | 76.78 |
| 12 | TENNESSEE | 6,346,105 | 4,596.18 | 72.43 |
| 13 | ALASKA | 710,231 | 487.28 | 68.61 |
| 14 | NEVADA | 2,700,551 | 1,843.55 | 68.27 |

| | | | | |
|---|---|---|---|---|
| 15 | CALIFORNIA | 37,253,956 | 24,263.98 | 65.13 |
| 16 | GEORGIA | 9,687,653 | 5,899.84 | 60.90 |
| 17 | TEXAS | 25,145,561 | 15,146.23 | 60.23 |
| 18 | INDIANA | 6,483,802 | 3,312.24 | 51.08 |
| 19 | KENTUCKY | 4,339,367 | 2,166.55 | 49.93 |
| 20 | KANSAS | 2,853,118 | 1,328.06 | 46.55 |
| 21 | IDAHO | 1,567,582 | 704.72 | 44.96 |
| 22 | VIRGIN ISLANDS | 109,825 | 48.48 | 44.14 |
| 23 | DISTRICT OF COLUMBIA | 601,723 | 264.17 | 43.90 |
| 24 | SOUTH CAROLINA | 4,625,364 | 1,988.44 | 42.99 |
| 25 | FLORIDA | 18,801,310 | 8,042.31 | 42.78 |
| 26 | OHIO | 11,536,504 | 4,796.84 | 41.58 |
| 27 | SOUTH DAKOTA | 814,180 | 332.16 | 40.80 |
| 28 | MISSOURI | 5,988,927 | 2,358.28 | 39.38 |
| 29 | MONTANA | 989,415 | 373.66 | 37.77 |
| 30 | NEW MEXICO | 2,059,179 | 747.42 | 36.30 |
| 31 | VIRGINIA | 8,001,024 | 2,902.97 | 36.28 |
| 32 | WASHINGTON | 6,724,540 | 2,310.13 | 34.35 |
| 33 | OREGON | 3,831,074 | 1,274.68 | 33.27 |
| 34 | MARYLAND | 5,773,552 | 1,698.01 | 29.41 |
| 35 | NEBRASKA | 1,826,341 | 533.36 | 29.20 |
| 36 | NORTH CAROLINA | 9,535,483 | 2,684.98 | 28.16 |
| 37 | WEST VIRGINIA | 1,852,994 | 415.49 | 22.42 |
| 38 | HAWAII | 1,360,301 | 295.98 | 21.76 |
| 39 | WISCONSIN | 5,686,986 | 1,234.04 | 21.70 |
| 40 | NORTH DAKOTA | 672,591 | 141.73 | 21.07 |
| 41 | COLORADO | 5,029,196 | 978.32 | 19.45 |
| 42 | PENNSYLVANIA | 12,702,379 | 2,455.45 | 19.33 |
| 43 | MICHIGAN | 9,883,640 | 1,898.02 | 19.20 |
| 44 | DELAWARE | 897,934 | 167.87 | 18.70 |
| 45 | MASSACHUSETTS | 6,547,629 | 1,204.85 | 18.40 |
| 46 | MAINE | 1,328,361 | 240.43 | 18.10 |
| 47 | WYOMING | 563,626 | 98.26 | 17.43 |
| 48 | RHODE ISLAND | 1,052,567 | 178.33 | 16.94 |
| 49 | ILLINOIS | 12,830,632 | 1,916.65 | 14.94 |
| 50 | NEW YORK | 19,378,102 | 2,893.49 | 14.93 |
| 51 | NEW JERSEY | 8,791,894 | 1,099.07 | 12.50 |
| 52 | NEW HAMPSHIRE | 1,316,470 | 144.23 | 10.96 |
| 53 | MINNESOTA | 5,303,925 | 511.90 | 9.65 |
| 54 | CONNECTICUT | 3,574,097 | 178.77 | 5.00 |
| US TOTAL | | 312,825,210 | 151,377.31 | 48.39 |

**DRUG: 9250B - METHADONE**

| RANK | STATE NAME | STATE POPULATION | TOTAL GRAMS | TOTAL GRAMS/100K POPULATION |
|---|---|---|---|---|
| 1 | DELAWARE | 897,934 | 156,519.73 | 17,431.10 |
| 2 | RHODE ISLAND | 1,052,567 | 178,717.49 | 16,979.20 |
| 3 | CONNECTICUT | 3,574,097 | 496,590.47 | 13,894.15 |
| 4 | MARYLAND | 5,773,552 | 774,358.72 | 13,412.17 |
| 5 | VERMONT | 625,741 | 83,557.80 | 13,353.42 |
| 6 | MASSACHUSETTS | 6,547,629 | 633,643.98 | 9,677.46 |
| 7 | NEW MEXICO | 2,059,179 | 191,515.81 | 9,300.59 |
| 8 | MAINE | 1,328,361 | 119,553.73 | 9,000.09 |
| 9 | NEW HAMPSHIRE | 1,316,470 | 103,385.14 | 7,853.21 |
| 10 | ARIZONA | 6,392,017 | 474,088.60 | 7,416.89 |

| | | | | |
|---|---|---|---|---|
| 11 | NEW YORK | 19,378,102 | 1,345,640.69 | 6,944.13 |
| 12 | DISTRICT OF COLUMBIA | 601,723 | 40,113.80 | 6,666.49 |
| 13 | OREGON | 3,831,074 | 248,915.66 | 6,497.28 |
| 14 | PENNSYLVANIA | 12,702,379 | 812,865.45 | 6,399.32 |
| 15 | WASHINGTON | 6,724,540 | 417,261.21 | 6,205.05 |
| 16 | NEW JERSEY | 8,791,894 | 543,061.88 | 6,176.85 |
| 17 | KENTUCKY | 4,339,367 | 238,683.19 | 5,500.41 |
| 18 | ALASKA | 710,231 | 38,435.86 | 5,411.74 |
| 19 | WEST VIRGINIA | 1,852,994 | 100,099.55 | 5,402.04 |
| 20 | ALABAMA | 4,779,736 | 253,975.34 | 5,313.59 |
| 21 | NORTH CAROLINA | 9,535,483 | 492,143.12 | 5,161.18 |
| 22 | NEVADA | 2,700,551 | 138,627.26 | 5,133.30 |
| 23 | INDIANA | 6,483,802 | 331,469.02 | 5,112.26 |
| 24 | MICHIGAN | 9,883,640 | 480,949.26 | 4,866.11 |
| 25 | SOUTH CAROLINA | 4,625,364 | 223,634.41 | 4,834.96 |
| 26 | VIRGINIA | 8,001,024 | 377,777.06 | 4,721.61 |
| 27 | WISCONSIN | 5,686,986 | 258,563.01 | 4,546.57 |
| 28 | COLORADO | 5,029,196 | 220,379.75 | 4,382.01 |
| 29 | GEORGIA | 9,687,653 | 422,967.57 | 4,366.05 |
| 30 | UTAH | 2,763,885 | 117,547.77 | 4,252.99 |
| 31 | MINNESOTA | 5,303,925 | 206,627.12 | 3,895.74 |
| 32 | ILLINOIS | 12,830,632 | 498,899.97 | 3,888.35 |
| 33 | OKLAHOMA | 3,751,351 | 143,842.06 | 3,834.41 |
| 34 | MONTANA | 989,415 | 35,585.49 | 3,596.62 |
| 35 | NORTH DAKOTA | 672,591 | 23,562.43 | 3,503.23 |
| 36 | FLORIDA | 18,801,310 | 643,625.30 | 3,423.30 |
| 37 | OHIO | 11,536,504 | 375,017.89 | 3,250.71 |
| 38 | CALIFORNIA | 37,253,956 | 1,210,217.84 | 3,248.56 |
| 39 | LOUISIANA | 4,533,372 | 146,511.04 | 3,231.83 |
| 40 | TENNESSEE | 6,346,105 | 193,064.56 | 3,042.25 |
| 41 | IDAHO | 1,567,582 | 44,645.85 | 2,848.07 |
| 42 | KANSAS | 2,853,118 | 78,559.02 | 2,753.44 |
| 43 | PUERTO RICO | 3,725,789 | 89,191.80 | 2,393.90 |
| 44 | HAWAII | 1,360,301 | 28,851.11 | 2,120.94 |
| 45 | TEXAS | 25,145,561 | 501,934.21 | 1,996.11 |
| 46 | MISSOURI | 5,988,927 | 118,152.73 | 1,972.85 |
| 47 | IOWA | 3,046,355 | 53,890.46 | 1,769.01 |
| 48 | ARKANSAS | 2,915,918 | 47,835.73 | 1,640.50 |
| 49 | NEBRASKA | 1,826,341 | 24,698.36 | 1,352.34 |
| 50 | MISSISSIPPI | 2,967,297 | 27,046.34 | 911.48 |
| 51 | SOUTH DAKOTA | 814,180 | 7,032.40 | 863.74 |
| 52 | WYOMING | 563,626 | 2,573.46 | 456.59 |
| 53 | VIRGIN ISLANDS | 109,825 | 50.09 | 45.61 |
| 54 | GUAM | 178,430 | 52.32 | 29.32 |
| **US TOTAL** | | **312,825,210** | 14,816,509.92 | **4,736.35** |

**DRUG: 9300 - MORPHINE**

| RANK | STATE NAME | STATE POPULATION | TOTAL GRAMS | TOTAL GRAMS/100K POPULATION |
|---|---|---|---|---|
| 1 | DELAWARE | 897,934 | 55,678.60 | 6,200.74 |
| 2 | TENNESSEE | 6,346,105 | 389,664.66 | 6,140.22 |
| 3 | FLORIDA | 18,801,310 | 1,075,017.32 | 5,717.78 |
| 4 | NEVADA | 2,700,551 | 138,525.56 | 5,129.53 |
| 5 | ARIZONA | 6,392,017 | 324,708.36 | 5,079.90 |
| 6 | UTAH | 2,763,885 | 139,622.61 | 5,051.68 |

| | | | | |
|---|---|---|---|---|
| 7 | OREGON | 3,831,074 | 190,051.91 | 4,960.80 |
| 8 | IDAHO | 1,567,582 | 75,350.32 | 4,806.79 |
| 9 | OKLAHOMA | 3,751,351 | 179,566.23 | 4,786.71 |
| 10 | MONTANA | 989,415 | 46,303.56 | 4,679.89 |
| 11 | VERMONT | 625,741 | 29,111.18 | 4,652.27 |
| 12 | MICHIGAN | 9,883,640 | 456,353.92 | 4,617.27 |
| 13 | PENNSYLVANIA | 12,702,379 | 576,699.99 | 4,540.09 |
| 14 | WYOMING | 563,626 | 24,257.98 | 4,303.91 |
| 15 | WASHINGTON | 6,724,540 | 286,293.65 | 4,257.45 |
| 16 | KANSAS | 2,853,118 | 120,464.50 | 4,222.21 |
| 17 | SOUTH CAROLINA | 4,625,364 | 193,038.60 | 4,173.48 |
| 18 | MISSOURI | 5,988,927 | 244,656.70 | 4,085.15 |
| 19 | NEW HAMPSHIRE | 1,316,470 | 53,118.33 | 4,034.91 |
| 20 | ALABAMA | 4,779,736 | 188,051.10 | 3,934.34 |
| 21 | COLORADO | 5,029,196 | 195,949.27 | 3,896.23 |
| 22 | ALASKA | 710,231 | 26,802.55 | 3,773.78 |
| 23 | NORTH CAROLINA | 9,535,483 | 357,941.74 | 3,753.79 |
| 24 | MAINE | 1,328,361 | 47,610.28 | 3,584.14 |
| 25 | INDIANA | 6,483,802 | 227,929.16 | 3,515.36 |
| 26 | HAWAII | 1,360,301 | 47,763.35 | 3,511.23 |
| 27 | WISCONSIN | 5,686,986 | 198,184.11 | 3,484.87 |
| 28 | MISSISSIPPI | 2,967,297 | 100,129.51 | 3,374.44 |
| 29 | NEW MEXICO | 2,059,179 | 69,123.58 | 3,356.85 |
| 30 | ARKANSAS | 2,915,918 | 97,174.21 | 3,332.54 |
| 31 | LOUISIANA | 4,533,372 | 145,724.93 | 3,214.49 |
| 32 | MARYLAND | 5,773,552 | 185,465.20 | 3,212.32 |
| 33 | GEORGIA | 9,687,653 | 310,016.19 | 3,200.12 |
| 34 | NEBRASKA | 1,826,341 | 55,421.92 | 3,034.59 |
| 35 | IOWA | 3,046,355 | 92,194.00 | 3,026.37 |
| 36 | RHODE ISLAND | 1,052,567 | 31,789.73 | 3,020.21 |
| 37 | WEST VIRGINIA | 1,852,994 | 55,890.85 | 3,016.25 |
| 38 | MASSACHUSETTS | 6,547,629 | 192,935.58 | 2,946.65 |
| 39 | CALIFORNIA | 37,253,956 | 1,068,099.63 | 2,867.08 |
| 40 | VIRGINIA | 8,001,024 | 228,486.67 | 2,855.72 |
| 41 | KENTUCKY | 4,339,367 | 121,072.64 | 2,790.10 |
| 42 | CONNECTICUT | 3,574,097 | 98,921.60 | 2,767.74 |
| 43 | OHIO | 11,536,504 | 312,560.94 | 2,709.32 |
| 44 | NEW JERSEY | 8,791,894 | 227,330.77 | 2,585.69 |
| 45 | NORTH DAKOTA | 672,591 | 17,213.51 | 2,559.28 |
| 46 | SOUTH DAKOTA | 814,180 | 20,657.36 | 2,537.20 |
| 47 | ILLINOIS | 12,830,632 | 302,623.97 | 2,358.61 |
| 48 | TEXAS | 25,145,561 | 560,163.09 | 2,227.68 |
| 49 | MINNESOTA | 5,303,925 | 117,764.39 | 2,220.33 |
| 50 | NEW YORK | 19,378,102 | 422,018.55 | 2,177.81 |
| 51 | DISTRICT OF COLUMBIA | 601,723 | 10,325.98 | 1,716.07 |
| 52 | GUAM | 178,430 | 2,691.33 | 1,508.34 |
| 53 | VIRGIN ISLANDS | 109,825 | 272.97 | 248.55 |
| 54 | PUERTO RICO | 3,725,789 | 5,642.81 | 151.45 |
| 55 | AMERICAN SAMOA | 65,628 | 22.71 | 34.60 |
| **US TOTAL** | | **312,825,210** | 10,740,450.16 | **3,433.37** |

**DRUG: 9652 - OXYMORPHONE**

| RANK | STATE NAME | STATE POPULATION | TOTAL GRAMS | TOTAL GRAMS/100K POPULATION |
|---|---|---|---|---|
| 1 | DELAWARE | 897,934 | 2,926.13 | 325.87 |

| | | | | |
|---|---|---|---|---|
| 2 | TENNESSEE | 6,346,105 | 19,595.60 | 308.78 |
| 3 | UTAH | 2,763,885 | 6,827.51 | 247.03 |
| 4 | NORTH CAROLINA | 9,535,483 | 20,034.01 | 210.10 |
| 5 | MICHIGAN | 9,883,640 | 17,949.77 | 181.61 |
| 6 | MARYLAND | 5,773,552 | 9,676.03 | 167.59 |
| 7 | PENNSYLVANIA | 12,702,379 | 20,839.01 | 164.06 |
| 8 | LOUISIANA | 4,533,372 | 7,365.13 | 162.46 |
| 9 | SOUTH CAROLINA | 4,625,364 | 7,395.42 | 159.89 |
| 10 | VIRGINIA | 8,001,024 | 10,810.38 | 135.11 |
| 11 | IDAHO | 1,567,582 | 1,776.57 | 113.33 |
| 12 | FLORIDA | 18,801,310 | 21,265.92 | 113.11 |
| 13 | NEVADA | 2,700,551 | 2,951.78 | 109.30 |
| 14 | KENTUCKY | 4,339,367 | 4,516.30 | 104.08 |
| 15 | NEW YORK | 19,378,102 | 19,144.60 | 98.80 |
| 16 | WEST VIRGINIA | 1,852,994 | 1,714.08 | 92.50 |
| 17 | CONNECTICUT | 3,574,097 | 3,105.80 | 86.90 |
| 18 | NEW JERSEY | 8,791,894 | 7,331.19 | 83.39 |
| 19 | MISSISSIPPI | 2,967,297 | 2,313.84 | 77.98 |
| 20 | ALABAMA | 4,779,736 | 3,694.50 | 77.30 |
| 21 | MISSOURI | 5,988,927 | 4,482.22 | 74.84 |
| 22 | MAINE | 1,328,361 | 935.72 | 70.44 |
| 23 | SOUTH DAKOTA | 814,180 | 571.70 | 70.22 |
| 24 | ARIZONA | 6,392,017 | 4,302.55 | 67.31 |
| 25 | OHIO | 11,536,504 | 7,705.20 | 66.79 |
| 26 | GEORGIA | 9,687,653 | 6,339.13 | 65.44 |
| 27 | KANSAS | 2,853,118 | 1,830.95 | 64.17 |
| 28 | COLORADO | 5,029,196 | 3,118.96 | 62.02 |
| 29 | OKLAHOMA | 3,751,351 | 2,319.64 | 61.83 |
| 30 | INDIANA | 6,483,802 | 3,947.01 | 60.87 |
| 31 | NEBRASKA | 1,826,341 | 1,069.63 | 58.57 |
| 32 | VERMONT | 625,741 | 324.72 | 51.89 |
| 33 | WASHINGTON | 6,724,540 | 3,463.22 | 51.50 |
| 34 | NEW HAMPSHIRE | 1,316,470 | 655.43 | 49.79 |
| 35 | OREGON | 3,831,074 | 1,878.49 | 49.03 |
| 36 | RHODE ISLAND | 1,052,567 | 478.17 | 45.43 |
| 37 | WYOMING | 563,626 | 246.89 | 43.80 |
| 38 | NEW MEXICO | 2,059,179 | 882.96 | 42.88 |
| 39 | HAWAII | 1,360,301 | 542.66 | 39.89 |
| 40 | MASSACHUSETTS | 6,547,629 | 2,609.97 | 39.86 |
| 41 | ILLINOIS | 12,830,632 | 4,412.15 | 34.39 |
| 42 | TEXAS | 25,145,561 | 8,444.40 | 33.58 |
| 43 | CALIFORNIA | 37,253,956 | 11,336.18 | 30.43 |
| 44 | MONTANA | 989,415 | 296.71 | 29.99 |
| 45 | NORTH DAKOTA | 672,591 | 198.85 | 29.56 |
| 46 | WISCONSIN | 5,686,986 | 1,646.42 | 28.95 |
| 47 | IOWA | 3,046,355 | 790.13 | 25.94 |
| 48 | DISTRICT OF COLUMBIA | 601,723 | 154.96 | 25.75 |
| 49 | ALASKA | 710,231 | 115.62 | 16.28 |
| 50 | ARKANSAS | 2,915,918 | 442.48 | 15.17 |
| 51 | MINNESOTA | 5,303,925 | 618.23 | 11.66 |
| **US TOTAL** | | **312,825,210** | 267,394.91 | **85.48** |

**DRUG: 9780 - TAPENTADOL**

| RANK | STATE NAME | STATE POPULATION | TOTAL GRAMS | TOTAL GRAMS/100K POPULATION |
|---|---|---|---|---|

| | | | | |
|---|---|---|---|---|
| 1 | NORTH CAROLINA | 9,535,483 | 274,136.75 | 2,874.91 |
| 2 | SOUTH CAROLINA | 4,625,364 | 98,345.00 | 2,126.21 |
| 3 | ALABAMA | 4,779,736 | 97,624.00 | 2,042.46 |
| 4 | DELAWARE | 897,934 | 15,533.50 | 1,729.92 |
| 5 | NEW HAMPSHIRE | 1,316,470 | 21,990.00 | 1,670.38 |
| 6 | NEBRASKA | 1,826,341 | 29,935.00 | 1,639.07 |
| 7 | COLORADO | 5,029,196 | 82,295.00 | 1,636.35 |
| 8 | GEORGIA | 9,687,653 | 155,037.00 | 1,600.36 |
| 9 | MARYLAND | 5,773,552 | 85,814.00 | 1,486.33 |
| 10 | INDIANA | 6,483,802 | 90,738.50 | 1,399.46 |
| 11 | TENNESSEE | 6,346,105 | 83,409.50 | 1,314.34 |
| 12 | ARIZONA | 6,392,017 | 81,467.50 | 1,274.52 |
| 13 | PENNSYLVANIA | 12,702,379 | 155,855.00 | 1,226.97 |
| 14 | NEW JERSEY | 8,791,894 | 107,610.00 | 1,223.97 |
| 15 | VIRGINIA | 8,001,024 | 96,006.00 | 1,199.92 |
| 16 | SOUTH DAKOTA | 814,180 | 9,764.50 | 1,199.30 |
| 17 | NEW YORK | 19,378,102 | 227,564.00 | 1,174.34 |
| 18 | ARKANSAS | 2,915,918 | 33,814.00 | 1,159.63 |
| 19 | UTAH | 2,763,885 | 31,913.50 | 1,154.66 |
| 20 | FLORIDA | 18,801,310 | 210,501.50 | 1,119.61 |
| 21 | OHIO | 11,536,504 | 125,558.00 | 1,088.35 |
| 22 | IDAHO | 1,567,582 | 16,165.50 | 1,031.24 |
| 23 | MISSOURI | 5,988,927 | 59,246.00 | 989.26 |
| 24 | MISSISSIPPI | 2,967,297 | 27,578.50 | 929.41 |
| 25 | LOUISIANA | 4,533,372 | 41,614.50 | 917.96 |
| 26 | MASSACHUSETTS | 6,547,629 | 57,641.50 | 880.34 |
| 27 | ALASKA | 710,231 | 5,944.50 | 836.98 |
| 28 | NEVADA | 2,700,551 | 22,546.00 | 834.87 |
| 29 | IOWA | 3,046,355 | 25,187.50 | 826.81 |
| 30 | TEXAS | 25,145,561 | 207,356.50 | 824.62 |
| 31 | OKLAHOMA | 3,751,351 | 28,206.00 | 751.89 |
| 32 | WEST VIRGINIA | 1,852,994 | 13,382.00 | 722.18 |
| 33 | KENTUCKY | 4,339,367 | 31,145.00 | 717.73 |
| 34 | CONNECTICUT | 3,574,097 | 24,487.50 | 685.14 |
| 35 | WYOMING | 563,626 | 3,846.50 | 682.46 |
| 36 | WASHINGTON | 6,724,540 | 43,142.00 | 641.56 |
| 37 | KANSAS | 2,853,118 | 17,945.00 | 628.96 |
| 38 | ILLINOIS | 12,830,632 | 79,775.00 | 621.75 |
| 39 | CALIFORNIA | 37,253,956 | 229,309.90 | 615.53 |
| 40 | NORTH DAKOTA | 672,591 | 3,995.50 | 594.05 |
| 41 | RHODE ISLAND | 1,052,567 | 6,158.50 | 585.09 |
| 42 | WISCONSIN | 5,686,986 | 30,185.50 | 530.78 |
| 43 | MICHIGAN | 9,883,640 | 47,359.00 | 479.17 |
| 44 | VERMONT | 625,741 | 2,952.50 | 471.84 |
| 45 | MONTANA | 989,415 | 4,438.00 | 448.55 |
| 46 | OREGON | 3,831,074 | 16,082.50 | 419.79 |
| 47 | HAWAII | 1,360,301 | 4,985.50 | 366.50 |
| 48 | NEW MEXICO | 2,059,179 | 7,173.50 | 348.37 |
| 49 | DISTRICT OF COLUMBIA | 601,723 | 2,045.50 | 339.94 |
| 50 | MAINE | 1,328,361 | 4,049.50 | 304.85 |
| 51 | MINNESOTA | 5,303,925 | 11,418.00 | 215.27 |
| 52 | VIRGIN ISLANDS | 109,825 | 45.00 | 40.97 |
| 53 | PUERTO RICO | 3,725,789 | 1,243.00 | 33.36 |

| US TOTAL | | 312,825,210 | 3,191,563.15 | 1,020.24 |
|---|---|---|---|---|
| **DRUG: 9801 - FENTANYL BASE** | | | | |
| RANK | STATE NAME | STATE POPULATION | TOTAL GRAMS | TOTAL GRAMS/100K POPULATION |
| 1 | MISSISSIPPI | 2,967,297 | 3,205.42 | 108.03 |
| 2 | IDAHO | 1,567,582 | 1,337.68 | 85.33 |
| 3 | KANSAS | 2,853,118 | 2,349.54 | 82.35 |
| 4 | MONTANA | 989,415 | 776.81 | 78.51 |
| 5 | MISSOURI | 5,988,927 | 4,698.40 | 78.45 |
| 6 | NORTH DAKOTA | 672,591 | 526.34 | 78.26 |
| 7 | PENNSYLVANIA | 12,702,379 | 9,627.61 | 75.79 |
| 8 | SOUTH DAKOTA | 814,180 | 599.98 | 73.69 |
| 9 | UTAH | 2,763,885 | 2,027.33 | 73.35 |
| 10 | FLORIDA | 18,801,310 | 12,975.48 | 69.01 |
| 11 | NEBRASKA | 1,826,341 | 1,255.18 | 68.73 |
| 12 | IOWA | 3,046,355 | 2,028.51 | 66.59 |
| 13 | GEORGIA | 9,687,653 | 6,320.88 | 65.25 |
| 14 | ALABAMA | 4,779,736 | 3,094.63 | 64.74 |
| 15 | MAINE | 1,328,361 | 858.88 | 64.66 |
| 16 | NORTH CAROLINA | 9,535,483 | 6,106.55 | 64.04 |
| 17 | MICHIGAN | 9,883,640 | 6,287.48 | 63.62 |
| 18 | OKLAHOMA | 3,751,351 | 2,375.05 | 63.31 |
| 19 | TENNESSEE | 6,346,105 | 3,990.60 | 62.88 |
| 20 | WYOMING | 563,626 | 351.95 | 62.44 |
| 21 | VERMONT | 625,741 | 388.43 | 62.07 |
| 22 | INDIANA | 6,483,802 | 4,020.81 | 62.01 |
| 23 | WEST VIRGINIA | 1,852,994 | 1,137.12 | 61.37 |
| 24 | SOUTH CAROLINA | 4,625,364 | 2,810.45 | 60.76 |
| 25 | DELAWARE | 897,934 | 541.11 | 60.26 |
| 26 | NEW HAMPSHIRE | 1,316,470 | 778.02 | 59.10 |
| 27 | ARKANSAS | 2,915,918 | 1,688.86 | 57.92 |
| 28 | KENTUCKY | 4,339,367 | 2,451.37 | 56.49 |
| 29 | CONNECTICUT | 3,574,097 | 2,011.60 | 56.28 |
| 30 | LOUISIANA | 4,533,372 | 2,515.52 | 55.49 |
| 31 | NEW JERSEY | 8,791,894 | 4,847.34 | 55.13 |
| 32 | WISCONSIN | 5,686,986 | 3,012.62 | 52.97 |
| 33 | NEVADA | 2,700,551 | 1,420.06 | 52.58 |
| 34 | COLORADO | 5,029,196 | 2,639.45 | 52.48 |
| 35 | ILLINOIS | 12,830,632 | 6,671.19 | 51.99 |
| 36 | NEW YORK | 19,378,102 | 9,957.05 | 51.38 |
| 37 | OREGON | 3,831,074 | 1,938.25 | 50.59 |
| 38 | TEXAS | 25,145,561 | 12,560.18 | 49.95 |
| 39 | WASHINGTON | 6,724,540 | 3,308.95 | 49.21 |
| 40 | VIRGINIA | 8,001,024 | 3,896.36 | 48.70 |
| 41 | ARIZONA | 6,392,017 | 3,106.16 | 48.59 |
| 42 | ALASKA | 710,231 | 344.96 | 48.57 |
| 43 | MASSACHUSETTS | 6,547,629 | 3,158.55 | 48.24 |
| 44 | MARYLAND | 5,773,552 | 2,716.22 | 47.05 |
| 45 | NEW MEXICO | 2,059,179 | 941.39 | 45.72 |
| 46 | MINNESOTA | 5,303,925 | 2,283.74 | 43.06 |
| 47 | OHIO | 11,536,504 | 4,918.11 | 42.63 |
| 48 | RHODE ISLAND | 1,052,567 | 421.20 | 40.02 |
| 49 | CALIFORNIA | 37,253,956 | 14,283.87 | 38.34 |
| 50 | DISTRICT OF COLUMBIA | 601,723 | 210.11 | 34.92 |

| | | | | |
|---|---|---|---|---|
| 51 | HAWAII | 1,360,301 | 423.99 | 31.17 |
| 52 | GUAM | 178,430 | 35.65 | 19.98 |
| 53 | PUERTO RICO | 3,725,789 | 636.31 | 17.08 |
| 54 | VIRGIN ISLANDS | 109,825 | 17.90 | 16.30 |
| 55 | AMERICAN SAMOA | 65,628 | 0.24 | 0.36 |
| **US TOTAL** | | **312,825,210** | 172,887.46 | **55.27** |

**STATE: AK - ALASKA**

**BUSINESS ACTIVITY: A - PHARMACIES**

| DRUG NAME | DRUG CODE | NUMBER OF REGISTRANT SOLD TO | TOTAL GRAMS | AVG GRAMS |
|---|---|---|---|---|
| AMPHETAMINE | 1100 | 106 | **30,081.00** | 283.78 |
| METHAMPHETAMINE | 1105 | 2 | **6.43** | 3.21 |
| LISDEXAMFETAMINE | 1205 | 94 | **11,160.80** | 118.73 |
| METHYLPHENIDATE (DL;D;L;ISOMERS) | 1724 | 107 | **23,958.73** | 223.91 |
| CODEINE | 9050 | 105 | **13,004.15** | 123.85 |
| BUPRENORPHINE | 9064 | 102 | **9,277.52** | 90.96 |
| OXYCODONE | 9143 | 108 | **67,739.09** | 627.21 |
| HYDROMORPHONE | 9150 | 90 | **1,232.51** | 13.69 |
| HYDROCODONE | 9193 | 108 | **26,235.55** | 242.92 |
| MEPERIDINE (PETHIDINE) | 9230 | 10 | **394.57** | 39.46 |
| METHADONE | 9250B | 83 | **5,606.55** | 67.55 |
| MORPHINE | 9300 | 107 | **22,068.62** | 206.25 |
| OXYMORPHONE | 9652 | 12 | **115.62** | 9.63 |
| TAPENTADOL | 9780 | 48 | **5,536.50** | 115.34 |
| FENTANYL BASE | 9801 | 95 | **249.85** | 2.63 |

**BUSINESS ACTIVITY: B - HOSPITALS**

| DRUG NAME | DRUG CODE | NUMBER OF REGISTRANT SOLD TO | TOTAL GRAMS | AVG GRAMS |
|---|---|---|---|---|
| AMPHETAMINE | 1100 | 28 | **5,470.91** | 195.39 |
| LISDEXAMFETAMINE | 1205 | 17 | **1,420.97** | 83.59 |
| METHYLPHENIDATE (DL;D;L;ISOMERS) | 1724 | 32 | **6,561.46** | 205.05 |
| CODEINE | 9050 | 37 | **5,035.03** | 136.08 |
| BUPRENORPHINE | 9064 | 34 | **1,730.66** | 50.90 |
| OXYCODONE | 9143 | 47 | **9,855.67** | 209.70 |
| HYDROMORPHONE | 9150 | 42 | **386.25** | 9.20 |
| HYDROCODONE | 9193 | 47 | **3,189.08** | 67.85 |
| MEPERIDINE (PETHIDINE) | 9230 | 18 | **57.41** | 3.19 |
| METHADONE | 9250B | 26 | **843.78** | 32.45 |
| MORPHINE | 9300 | 46 | **4,694.99** | 102.06 |
| TAPENTADOL | 9780 | 7 | **408.00** | 58.29 |
| FENTANYL BASE | 9801 | 51 | **88.00** | 1.73 |

**BUSINESS ACTIVITY: C - PRACTITIONERS**

| DRUG NAME | DRUG CODE | NUMBER OF REGISTRANT SOLD TO | TOTAL GRAMS | AVG GRAMS |
|---|---|---|---|---|
| CODEINE | 9050 | 6 | **218.57** | 36.43 |
| BUPRENORPHINE | 9064 | 60 | **11.82** | 0.20 |
| OXYCODONE | 9143 | 3 | **4.44** | 1.48 |
| HYDROMORPHONE | 9150 | 46 | **490.73** | 10.67 |
| HYDROCODONE | 9193 | 21 | **51.43** | 2.45 |
| MEPERIDINE (PETHIDINE) | 9230 | 7 | **35.30** | 5.04 |
| METHADONE | 9250B | 3 | **1.79** | 0.60 |
| MORPHINE | 9300 | 30 | **38.57** | 1.29 |
| FENTANYL BASE | 9801 | 89 | **7.11** | 0.08 |

**BUSINESS ACTIVITY: M - MID-LEVEL PRACTITIONERS**

| DRUG NAME | DRUG CODE | NUMBER OF REGISTRANT SOLD TO | TOTAL GRAMS | AVG GRAMS |
|---|---|---|---|---|
| CODEINE | 9050 | 1 | **2.21** | 2.21 |
| BUPRENORPHINE | 9064 | 3 | **50.39** | 16.80 |
| HYDROCODONE | 9193 | 4 | **4.63** | 1.16 |
| MORPHINE | 9300 | 1 | **0.38** | 0.38 |
| FENTANYL BASE | 9801 | 2 | **0.01** | 0.00 |

**BUSINESS ACTIVITY: NTP - NARCOTIC TREATMENT PROGRAMS**

| DRUG NAME | DRUG CODE | NUMBER OF REGISTRANT SOLD TO | TOTAL GRAMS | AVG GRAMS |
|---|---|---|---|---|
| BUPRENORPHINE | 9064 | 2 | **53.82** | 26.91 |
| METHADONE | 9250B | 4 | **31,983.74** | 7,995.94 |

**STATE: AL - ALABAMA**

**BUSINESS ACTIVITY: A - PHARMACIES**

| DRUG NAME | DRUG CODE | NUMBER OF REGISTRANT SOLD TO | TOTAL GRAMS | AVG GRAMS |
|---|---|---|---|---|
| AMPHETAMINE | 1100 | 1,263 | **549,651.35** | 435.20 |
| METHAMPHETAMINE | 1105 | 35 | **124.94** | 3.57 |
| LISDEXAMFETAMINE | 1205 | 1,235 | **275,459.45** | 223.04 |

| | | | | |
|---|---|---|---|---|
| METHYLPHENIDATE (DL;D;L;ISOMERS) | 1724 | 1,256 | **269,127.34** | 214.27 |
| CODEINE | 9050 | 1,234 | **163,693.69** | 132.65 |
| BUPRENORPHINE | 9064 | 1,219 | **141,200.52** | 115.83 |
| OXYCODONE | 9143 | 1,278 | **616,478.97** | 482.38 |
| HYDROMORPHONE | 9150 | 972 | **5,745.81** | 5.91 |
| HYDROCODONE | 9193 | 1,288 | **678,464.54** | 526.76 |
| MEPERIDINE (PETHIDINE) | 9230 | 143 | **1,570.77** | 10.98 |
| METHADONE | 9250B | 1,018 | **38,335.15** | 37.66 |
| MORPHINE | 9300 | 1,235 | **174,447.22** | 141.25 |
| OXYMORPHONE | 9652 | 333 | **3,674.25** | 11.03 |
| TAPENTADOL | 9780 | 802 | **96,830.50** | 120.74 |
| FENTANYL BASE | 9801 | 1,163 | **2,539.70** | 2.18 |

**BUSINESS ACTIVITY: B - HOSPITALS**

| DRUG NAME | DRUG CODE | NUMBER OF REGISTRANT SOLD TO | TOTAL GRAMS | AVG GRAMS |
|---|---|---|---|---|
| AMPHETAMINE | 1100 | 68 | **7,953.05** | 116.96 |
| LISDEXAMFETAMINE | 1205 | 29 | **2,597.40** | 89.57 |
| METHYLPHENIDATE (DL;D;L;ISOMERS) | 1724 | 82 | **5,713.93** | 69.68 |
| CODEINE | 9050 | 108 | **2,477.93** | 22.94 |
| BUPRENORPHINE | 9064 | 102 | **1,791.72** | 17.57 |
| OXYCODONE | 9143 | 144 | **25,652.10** | 178.14 |
| HYDROMORPHONE | 9150 | 131 | **2,784.04** | 21.25 |
| HYDROCODONE | 9193 | 148 | **25,381.47** | 171.50 |
| MEPERIDINE (PETHIDINE) | 9230 | 100 | **3,964.34** | 39.64 |
| METHADONE | 9250B | 104 | **1,841.93** | 17.71 |
| MORPHINE | 9300 | 146 | **13,281.92** | 90.97 |
| OXYMORPHONE | 9652 | 6 | **20.25** | 3.38 |
| TAPENTADOL | 9780 | 20 | **793.50** | 39.68 |
| FENTANYL BASE | 9801 | 130 | **524.38** | 4.03 |

**BUSINESS ACTIVITY: C - PRACTITIONERS**

| DRUG NAME | DRUG CODE | NUMBER OF REGISTRANT SOLD TO | TOTAL GRAMS | AVG GRAMS |
|---|---|---|---|---|
| CODEINE | 9050 | 47 | **2,007.74** | 42.72 |
| BUPRENORPHINE | 9064 | 326 | **38.78** | 0.12 |
| OXYCODONE | 9143 | 28 | **242.55** | 8.66 |
| HYDROMORPHONE | 9150 | 101 | **552.13** | 5.47 |
| HYDROCODONE | 9193 | 120 | **617.86** | 5.15 |
| MEPERIDINE (PETHIDINE) | 9230 | 83 | **864.96** | 10.42 |
| METHADONE | 9250B | 3 | **4.47** | 1.49 |
| MORPHINE | 9300 | 107 | **321.96** | 3.01 |
| FENTANYL BASE | 9801 | 253 | **30.24** | 0.12 |

**BUSINESS ACTIVITY: D - TEACHING INSTITUTIONS**

| DRUG NAME | DRUG CODE | NUMBER OF REGISTRANT SOLD TO | TOTAL GRAMS | AVG GRAMS |
|---|---|---|---|---|
| BUPRENORPHINE | 9064 | 1 | **0.01** | 0.01 |
| HYDROMORPHONE | 9150 | 1 | **14.19** | 14.19 |
| FENTANYL BASE | 9801 | 1 | **0.30** | 0.30 |

**BUSINESS ACTIVITY: M - MID-LEVEL PRACTITIONERS**

| DRUG NAME | DRUG CODE | NUMBER OF REGISTRANT SOLD TO | TOTAL GRAMS | AVG GRAMS |
|---|---|---|---|---|
| CODEINE | 9050 | 4 | **2.80** | 0.70 |
| BUPRENORPHINE | 9064 | 1 | **13.86** | 13.86 |

**BUSINESS ACTIVITY: NTP - NARCOTIC TREATMENT PROGRAMS**

| DRUG NAME | DRUG CODE | NUMBER OF REGISTRANT SOLD TO | TOTAL GRAMS | AVG GRAMS |
|---|---|---|---|---|
| BUPRENORPHINE | 9064 | 15 | **637.63** | 42.51 |
| METHADONE | 9250B | 23 | **213,793.80** | 9,295.38 |

**STATE: AR - ARKANSAS**

**BUSINESS ACTIVITY: A - PHARMACIES**

| DRUG NAME | DRUG CODE | NUMBER OF REGISTRANT SOLD TO | TOTAL GRAMS | AVG GRAMS |
|---|---|---|---|---|
| AMPHETAMINE | 1100 | 725 | **227,813.84** | 314.23 |
| METHAMPHETAMINE | 1105 | 7 | **10.85** | 1.55 |
| LISDEXAMFETAMINE | 1205 | 720 | **106,581.41** | 148.03 |
| METHYLPHENIDATE (DL;D;L;ISOMERS) | 1724 | 730 | **137,795.83** | 188.76 |
| CODEINE | 9050 | 725 | **183,230.70** | 252.73 |
| BUPRENORPHINE | 9064 | 700 | **42,166.82** | 60.24 |

| | | | | |
|---|---|---|---|---|
| OXYCODONE | 9143 | 731 | **296,106.67** | 405.07 |
| HYDROMORPHONE | 9150 | 668 | **5,561.76** | 8.33 |
| HYDROCODONE | 9193 | 736 | **377,489.91** | 512.89 |
| MEPERIDINE (PETHIDINE) | 9230 | 238 | **2,687.60** | 11.29 |
| METHADONE | 9250B | 597 | **13,002.16** | 21.78 |
| MORPHINE | 9300 | 715 | **89,565.06** | 125.27 |
| OXYMORPHONE | 9652 | 81 | **436.59** | 5.39 |
| TAPENTADOL | 9780 | 398 | **33,380.50** | 83.87 |
| FENTANYL BASE | 9801 | 676 | **1,352.20** | 2.00 |

**BUSINESS ACTIVITY: B - HOSPITALS**

| DRUG NAME | DRUG CODE | NUMBER OF REGISTRANT SOLD TO | TOTAL GRAMS | AVG GRAMS |
|---|---|---|---|---|
| AMPHETAMINE | 1100 | 50 | **4,905.42** | 98.11 |
| LISDEXAMFETAMINE | 1205 | 21 | **1,133.08** | 53.96 |
| METHYLPHENIDATE (DL;D;L;ISOMERS) | 1724 | 67 | **3,865.80** | 57.70 |
| CODEINE | 9050 | 86 | **2,853.66** | 33.18 |
| BUPRENORPHINE | 9064 | 55 | **1,141.28** | 20.75 |
| OXYCODONE | 9143 | 117 | **11,831.52** | 101.12 |
| HYDROMORPHONE | 9150 | 107 | **1,371.52** | 12.82 |
| HYDROCODONE | 9193 | 122 | **11,884.92** | 97.42 |
| MEPERIDINE (PETHIDINE) | 9230 | 82 | **2,066.33** | 25.20 |
| METHADONE | 9250B | 83 | **1,382.12** | 16.65 |
| MORPHINE | 9300 | 118 | **7,535.30** | 63.86 |
| OXYMORPHONE | 9652 | 2 | **5.89** | 2.94 |
| TAPENTADOL | 9780 | 17 | **433.50** | 25.50 |
| FENTANYL BASE | 9801 | 121 | **325.02** | 2.69 |

**BUSINESS ACTIVITY: C - PRACTITIONERS**

| DRUG NAME | DRUG CODE | NUMBER OF REGISTRANT SOLD TO | TOTAL GRAMS | AVG GRAMS |
|---|---|---|---|---|
| CODEINE | 9050 | 38 | **250.31** | 6.59 |
| BUPRENORPHINE | 9064 | 180 | **18.26** | 0.10 |
| OXYCODONE | 9143 | 7 | **19.45** | 2.78 |
| HYDROMORPHONE | 9150 | 77 | **329.29** | 4.28 |
| HYDROCODONE | 9193 | 74 | **162.51** | 2.20 |
| MEPERIDINE (PETHIDINE) | 9230 | 36 | **319.84** | 8.88 |
| METHADONE | 9250B | 1 | **0.89** | 0.89 |
| MORPHINE | 9300 | 58 | **73.85** | 1.27 |
| FENTANYL BASE | 9801 | 147 | **11.64** | 0.08 |

**BUSINESS ACTIVITY: M - MID-LEVEL PRACTITIONERS**

| DRUG NAME | DRUG CODE | NUMBER OF REGISTRANT SOLD TO | TOTAL GRAMS | AVG GRAMS |
|---|---|---|---|---|
| CODEINE | 9050 | 11 | **7.06** | 0.64 |

**BUSINESS ACTIVITY: NTP - NARCOTIC TREATMENT PROGRAMS**

| DRUG NAME | DRUG CODE | NUMBER OF REGISTRANT SOLD TO | TOTAL GRAMS | AVG GRAMS |
|---|---|---|---|---|
| BUPRENORPHINE | 9064 | 1 | **4.56** | 4.56 |
| METHADONE | 9250B | 6 | **33,450.56** | 5,575.09 |

**STATE: AS - AMERICAN SAMOA**

**BUSINESS ACTIVITY: B - HOSPITALS**

| DRUG NAME | DRUG CODE | NUMBER OF REGISTRANT SOLD TO | TOTAL GRAMS | AVG GRAMS |
|---|---|---|---|---|
| AMPHETAMINE | 1100 | 1 | **3.75** | 3.75 |
| METHYLPHENIDATE (DL;D;L;ISOMERS) | 1724 | 1 | **15.57** | 15.57 |
| CODEINE | 9050 | 1 | **1,354.79** | 1,354.79 |
| OXYCODONE | 9143 | 1 | **51.10** | 51.10 |
| HYDROMORPHONE | 9150 | 1 | **1.68** | 1.68 |
| HYDROCODONE | 9193 | 1 | **10.29** | 10.29 |
| MORPHINE | 9300 | 1 | **22.71** | 22.71 |
| FENTANYL BASE | 9801 | 1 | **0.24** | 0.24 |

**STATE: AZ - ARIZONA**

**BUSINESS ACTIVITY: A - PHARMACIES**

| DRUG NAME | DRUG CODE | NUMBER OF REGISTRANT SOLD TO | TOTAL GRAMS | AVG GRAMS |
|---|---|---|---|---|
| AMPHETAMINE | 1100 | 1,100 | **409,095.40** | 371.90 |
| METHAMPHETAMINE | 1105 | 29 | **118.52** | 4.09 |
| LISDEXAMFETAMINE | 1205 | 1,046 | **165,952.85** | 158.65 |
| METHYLPHENIDATE (DL;D;L;ISOMERS) | 1724 | 1,091 | **284,332.98** | 260.62 |

| | | | | |
|---|---|---|---|---|
| CODEINE | 9050 | 1,088 | **167,362.57** | 153.83 |
| BUPRENORPHINE | 9064 | 1,096 | **58,144.11** | 53.05 |
| OXYCODONE | 9143 | 1,116 | **1,011,728.56** | 906.57 |
| HYDROMORPHONE | 9150 | 984 | **15,806.27** | 16.06 |
| HYDROCODONE | 9193 | 1,125 | **272,872.55** | 242.55 |
| MEPERIDINE (PETHIDINE) | 9230 | 110 | **1,762.06** | 16.02 |
| METHADONE | 9250B | 805 | **22,444.52** | 27.88 |
| MORPHINE | 9300 | 1,095 | **300,556.09** | 274.48 |
| OXYMORPHONE | 9652 | 342 | **4,293.72** | 12.55 |
| TAPENTADOL | 9780 | 704 | **80,804.00** | 114.78 |
| FENTANYL BASE | 9801 | 980 | **2,450.95** | 2.50 |

**BUSINESS ACTIVITY: B - HOSPITALS**

| DRUG NAME | DRUG CODE | NUMBER OF REGISTRANT SOLD TO | TOTAL GRAMS | AVG GRAMS |
|---|---|---|---|---|
| AMPHETAMINE | 1100 | 97 | **9,169.24** | 94.53 |
| LISDEXAMFETAMINE | 1205 | 38 | **1,648.16** | 43.37 |
| METHYLPHENIDATE (DL;D;L;ISOMERS) | 1724 | 107 | **11,331.49** | 105.90 |
| CODEINE | 9050 | 122 | **8,648.44** | 70.89 |
| BUPRENORPHINE | 9064 | 101 | **3,760.84** | 37.24 |
| OXYCODONE | 9143 | 168 | **40,783.17** | 242.76 |
| HYDROMORPHONE | 9150 | 124 | **2,806.43** | 22.63 |
| HYDROCODONE | 9193 | 157 | **11,233.59** | 71.55 |
| MEPERIDINE (PETHIDINE) | 9230 | 77 | **2,306.64** | 29.96 |
| METHADONE | 9250B | 114 | **3,678.83** | 32.27 |
| MORPHINE | 9300 | 159 | **23,954.39** | 150.66 |
| OXYMORPHONE | 9652 | 4 | **8.83** | 2.21 |
| TAPENTADOL | 9780 | 11 | **663.50** | 60.32 |
| FENTANYL BASE | 9801 | 135 | **599.04** | 4.44 |

**BUSINESS ACTIVITY: C - PRACTITIONERS**

| DRUG NAME | DRUG CODE | NUMBER OF REGISTRANT SOLD TO | TOTAL GRAMS | AVG GRAMS |
|---|---|---|---|---|
| AMPHETAMINE | 1100 | 1 | **1.35** | 1.35 |
| CODEINE | 9050 | 50 | **2,365.98** | 47.32 |
| BUPRENORPHINE | 9064 | 698 | **124.43** | 0.18 |
| OXYCODONE | 9143 | 111 | **334.03** | 3.01 |
| HYDROMORPHONE | 9150 | 439 | **2,944.64** | 6.71 |
| HYDROCODONE | 9193 | 265 | **858.89** | 3.24 |
| MEPERIDINE (PETHIDINE) | 9230 | 234 | **2,044.19** | 8.74 |
| METHADONE | 9250B | 17 | **123.52** | 7.27 |
| MORPHINE | 9300 | 267 | **197.69** | 0.74 |
| FENTANYL BASE | 9801 | 792 | **56.10** | 0.07 |

**BUSINESS ACTIVITY: M - MID-LEVEL PRACTITIONERS**

| DRUG NAME | DRUG CODE | NUMBER OF REGISTRANT SOLD TO | TOTAL GRAMS | AVG GRAMS |
|---|---|---|---|---|
| CODEINE | 9050 | 4 | **27.18** | 6.80 |
| BUPRENORPHINE | 9064 | 3 | **6.72** | 2.24 |
| OXYCODONE | 9143 | 2 | **0.90** | 0.45 |
| MORPHINE | 9300 | 1 | **0.19** | 0.19 |
| FENTANYL BASE | 9801 | 2 | **0.08** | 0.04 |

**BUSINESS ACTIVITY: NTP - NARCOTIC TREATMENT PROGRAMS**

| DRUG NAME | DRUG CODE | NUMBER OF REGISTRANT SOLD TO | TOTAL GRAMS | AVG GRAMS |
|---|---|---|---|---|
| BUPRENORPHINE | 9064 | 32 | **1,741.15** | 54.41 |
| METHADONE | 9250B | 66 | **447,841.73** | 6,785.48 |

**STATE: CA - CALIFORNIA**

**BUSINESS ACTIVITY: A - PHARMACIES**

| DRUG NAME | DRUG CODE | NUMBER OF REGISTRANT SOLD TO | TOTAL GRAMS | AVG GRAMS |
|---|---|---|---|---|
| AMPHETAMINE | 1100 | 5,288 | **1,261,411.13** | 238.54 |
| METHAMPHETAMINE | 1105 | 221 | **901.14** | 4.08 |
| LISDEXAMFETAMINE | 1205 | 4,602 | **447,492.76** | 97.24 |
| METHYLPHENIDATE (DL;D;L;ISOMERS) | 1724 | 5,144 | **957,893.00** | 186.22 |
| CODEINE | 9050 | 5,906 | **1,154,270.75** | 195.44 |
| BUPRENORPHINE | 9064 | 5,130 | **223,766.43** | 43.62 |
| OXYCODONE | 9143 | 5,592 | **2,038,222.84** | 364.49 |
| HYDROMORPHONE | 9150 | 4,726 | **78,160.45** | 16.54 |

| | | | | |
|---|---|---|---|---|
| HYDROCODONE | 9193 | 5,887 | **2,352,250.19** | 399.57 |
| MEPERIDINE (PETHIDINE) | 9230 | 252 | **4,851.20** | 19.25 |
| METHADONE | 9250B | 4,155 | **173,680.88** | 41.80 |
| MORPHINE | 9300 | 5,156 | **985,364.95** | 191.11 |
| OXYMORPHONE | 9652 | 706 | **11,299.16** | 16.00 |
| TAPENTADOL | 9780 | 2,261 | **227,192.00** | 100.48 |
| FENTANYL BASE | 9801 | 4,194 | **10,985.08** | 2.62 |

**BUSINESS ACTIVITY: B - HOSPITALS**

| DRUG NAME | DRUG CODE | NUMBER OF REGISTRANT SOLD TO | TOTAL GRAMS | AVG GRAMS |
|---|---|---|---|---|
| AMPHETAMINE | 1100 | 253 | **27,399.30** | 108.30 |
| METHAMPHETAMINE | 1105 | 1 | **0.40** | 0.40 |
| LISDEXAMFETAMINE | 1205 | 92 | **3,320.03** | 36.09 |
| METHYLPHENIDATE (DL;D;L;ISOMERS) | 1724 | 362 | **23,765.72** | 65.65 |
| CODEINE | 9050 | 509 | **25,891.58** | 50.87 |
| BUPRENORPHINE | 9064 | 487 | **26,430.56** | 54.27 |
| OXYCODONE | 9143 | 644 | **97,429.38** | 151.29 |
| HYDROMORPHONE | 9150 | 663 | **18,100.30** | 27.30 |
| HYDROCODONE | 9193 | 694 | **81,686.68** | 117.70 |
| MEPERIDINE (PETHIDINE) | 9230 | 530 | **13,962.52** | 26.34 |
| METHADONE | 9250B | 511 | **22,815.33** | 44.65 |
| MORPHINE | 9300 | 738 | **82,078.93** | 111.22 |
| OXYMORPHONE | 9652 | 11 | **37.02** | 3.37 |
| TAPENTADOL | 9780 | 54 | **2,117.90** | 39.22 |
| FENTANYL BASE | 9801 | 824 | **2,881.26** | 3.50 |

**BUSINESS ACTIVITY: C - PRACTITIONERS**

| DRUG NAME | DRUG CODE | NUMBER OF REGISTRANT SOLD TO | TOTAL GRAMS | AVG GRAMS |
|---|---|---|---|---|
| AMPHETAMINE | 1100 | 6 | **367.19** | 61.20 |
| METHYLPHENIDATE (DL;D;L;ISOMERS) | 1724 | 4 | **120.38** | 30.10 |
| CODEINE | 9050 | 282 | **19,089.84** | 67.69 |
| BUPRENORPHINE | 9064 | 3,097 | **953.21** | 0.31 |
| OXYCODONE | 9143 | 191 | **522.28** | 2.73 |
| HYDROMORPHONE | 9150 | 1,633 | **9,534.17** | 5.84 |
| HYDROCODONE | 9193 | 1,768 | **8,805.84** | 4.98 |
| MEPERIDINE (PETHIDINE) | 9230 | 558 | **5,450.25** | 9.77 |
| METHADONE | 9250B | 164 | **329.77** | 2.01 |
| MORPHINE | 9300 | 747 | **652.74** | 0.87 |
| FENTANYL BASE | 9801 | 2,313 | **417.53** | 0.18 |

**BUSINESS ACTIVITY: D - TEACHING INSTITUTIONS**

| DRUG NAME | DRUG CODE | NUMBER OF REGISTRANT SOLD TO | TOTAL GRAMS | AVG GRAMS |
|---|---|---|---|---|
| BUPRENORPHINE | 9064 | 7 | **0.05** | 0.01 |
| OXYCODONE | 9143 | 1 | **4.48** | 4.48 |
| HYDROMORPHONE | 9150 | 3 | **12.06** | 4.02 |
| MORPHINE | 9300 | 1 | **3.01** | 3.01 |

**BUSINESS ACTIVITY: M - MID-LEVEL PRACTITIONERS**

| DRUG NAME | DRUG CODE | NUMBER OF REGISTRANT SOLD TO | TOTAL GRAMS | AVG GRAMS |
|---|---|---|---|---|
| CODEINE | 9050 | 4 | **114.70** | 28.68 |
| BUPRENORPHINE | 9064 | 3 | **8.94** | 2.98 |

**BUSINESS ACTIVITY: NTP - NARCOTIC TREATMENT PROGRAMS**

| DRUG NAME | DRUG CODE | NUMBER OF REGISTRANT SOLD TO | TOTAL GRAMS | AVG GRAMS |
|---|---|---|---|---|
| BUPRENORPHINE | 9064 | 117 | **6,721.02** | 57.44 |
| METHADONE | 9250B | 157 | **1,013,391.87** | 6,454.73 |

**STATE: CO - COLORADO**

**BUSINESS ACTIVITY: A - PHARMACIES**

| DRUG NAME | DRUG CODE | NUMBER OF REGISTRANT SOLD TO | TOTAL GRAMS | AVG GRAMS |
|---|---|---|---|---|
| AMPHETAMINE | 1100 | 790 | **360,729.70** | 456.62 |
| METHAMPHETAMINE | 1105 | 63 | **150.66** | 2.39 |
| LISDEXAMFETAMINE | 1205 | 750 | **117,479.17** | 156.64 |
| METHYLPHENIDATE (DL;D;L;ISOMERS) | 1724 | 785 | **209,104.58** | 266.38 |
| CODEINE | 9050 | 757 | **51,253.15** | 67.71 |
| BUPRENORPHINE | 9064 | 784 | **41,309.03** | 52.69 |
| OXYCODONE | 9143 | 797 | **523,753.98** | 657.16 |

| | | | | |
|---|---|---|---|---|
| HYDROMORPHONE | 9150 | 768 | **14,760.53** | 19.22 |
| HYDROCODONE | 9193 | 801 | **150,763.22** | 188.22 |
| MEPERIDINE (PETHIDINE) | 9230 | 53 | **392.68** | 7.41 |
| METHADONE | 9250B | 629 | **16,498.91** | 26.23 |
| MORPHINE | 9300 | 780 | **159,960.79** | 205.08 |
| OXYMORPHONE | 9652 | 251 | **2,938.53** | 11.71 |
| TAPENTADOL | 9780 | 576 | **78,230.50** | 135.82 |
| FENTANYL BASE | 9801 | 734 | **1,885.41** | 2.57 |

**BUSINESS ACTIVITY: B - HOSPITALS**

| DRUG NAME | DRUG CODE | NUMBER OF REGISTRANT SOLD TO | TOTAL GRAMS | AVG GRAMS |
|---|---|---|---|---|
| AMPHETAMINE | 1100 | 123 | **50,763.79** | 412.71 |
| METHAMPHETAMINE | 1105 | 1 | **14.06** | 14.06 |
| LISDEXAMFETAMINE | 1205 | 75 | **7,047.04** | 93.96 |
| METHYLPHENIDATE (DL;D;L;ISOMERS) | 1724 | 133 | **41,952.26** | 315.43 |
| CODEINE | 9050 | 133 | **8,297.89** | 62.39 |
| BUPRENORPHINE | 9064 | 145 | **5,856.56** | 40.39 |
| OXYCODONE | 9143 | 204 | **87,762.12** | 430.21 |
| HYDROMORPHONE | 9150 | 193 | **4,281.20** | 22.18 |
| HYDROCODONE | 9193 | 211 | **23,253.20** | 110.20 |
| MEPERIDINE (PETHIDINE) | 9230 | 80 | **382.26** | 4.78 |
| METHADONE | 9250B | 134 | **4,693.23** | 35.02 |
| MORPHINE | 9300 | 199 | **35,700.41** | 179.40 |
| OXYMORPHONE | 9652 | 29 | **180.43** | 6.22 |
| TAPENTADOL | 9780 | 56 | **4,064.50** | 72.58 |
| FENTANYL BASE | 9801 | 197 | **697.18** | 3.54 |

**BUSINESS ACTIVITY: C - PRACTITIONERS**

| DRUG NAME | DRUG CODE | NUMBER OF REGISTRANT SOLD TO | TOTAL GRAMS | AVG GRAMS |
|---|---|---|---|---|
| AMPHETAMINE | 1100 | 1 | **0.63** | 0.63 |
| CODEINE | 9050 | 43 | **1,165.07** | 27.09 |
| BUPRENORPHINE | 9064 | 799 | **721.59** | 0.90 |
| OXYCODONE | 9143 | 87 | **307.96** | 3.54 |
| HYDROMORPHONE | 9150 | 547 | **3,723.57** | 6.81 |
| HYDROCODONE | 9193 | 278 | **623.70** | 2.24 |
| MEPERIDINE (PETHIDINE) | 9230 | 56 | **200.12** | 3.57 |
| METHADONE | 9250B | 30 | **58.40** | 1.95 |
| MORPHINE | 9300 | 218 | **278.82** | 1.28 |
| FENTANYL BASE | 9801 | 462 | **55.25** | 0.12 |

**BUSINESS ACTIVITY: M - MID-LEVEL PRACTITIONERS**

| DRUG NAME | DRUG CODE | NUMBER OF REGISTRANT SOLD TO | TOTAL GRAMS | AVG GRAMS |
|---|---|---|---|---|
| CODEINE | 9050 | 1 | **3.32** | 3.32 |
| BUPRENORPHINE | 9064 | 5 | **30.74** | 6.15 |
| HYDROMORPHONE | 9150 | 11 | **0.65** | 0.06 |
| MEPERIDINE (PETHIDINE) | 9230 | 1 | **3.27** | 3.27 |
| MORPHINE | 9300 | 28 | **9.24** | 0.33 |
| FENTANYL BASE | 9801 | 56 | **1.61** | 0.03 |

**BUSINESS ACTIVITY: NTP - NARCOTIC TREATMENT PROGRAMS**

| DRUG NAME | DRUG CODE | NUMBER OF REGISTRANT SOLD TO | TOTAL GRAMS | AVG GRAMS |
|---|---|---|---|---|
| BUPRENORPHINE | 9064 | 17 | **1,033.70** | 60.81 |
| METHADONE | 9250B | 28 | **199,129.22** | 7,111.76 |

**STATE: CT - CONNECTICUT**

**BUSINESS ACTIVITY: A - PHARMACIES**

| DRUG NAME | DRUG CODE | NUMBER OF REGISTRANT SOLD TO | TOTAL GRAMS | AVG GRAMS |
|---|---|---|---|---|
| AMPHETAMINE | 1100 | 676 | **252,020.81** | 372.81 |
| METHAMPHETAMINE | 1105 | 23 | **73.92** | 3.21 |
| LISDEXAMFETAMINE | 1205 | 660 | **143,390.77** | 217.26 |
| METHYLPHENIDATE (DL;D;L;ISOMERS) | 1724 | 682 | **203,144.04** | 297.87 |
| CODEINE | 9050 | 660 | **57,139.67** | 86.58 |
| BUPRENORPHINE | 9064 | 669 | **73,673.65** | 110.13 |
| OXYCODONE | 9143 | 682 | **444,690.66** | 652.04 |
| HYDROMORPHONE | 9150 | 651 | **13,775.74** | 21.16 |
| HYDROCODONE | 9193 | 673 | **51,587.31** | 76.65 |

| | | | | |
|---|---|---|---|---|
| MEPERIDINE (PETHIDINE) | 9230 | 5 | **13.94** | 2.79 |
| METHADONE | 9250B | 485 | **14,266.57** | 29.42 |
| MORPHINE | 9300 | 646 | **90,814.00** | 140.58 |
| OXYMORPHONE | 9652 | 175 | **3,104.91** | 17.74 |
| TAPENTADOL | 9780 | 318 | **24,280.50** | 76.35 |
| FENTANYL BASE | 9801 | 582 | **1,692.22** | 2.91 |

**BUSINESS ACTIVITY: B - HOSPITALS**

| DRUG NAME | DRUG CODE | NUMBER OF REGISTRANT SOLD TO | TOTAL GRAMS | AVG GRAMS |
|---|---|---|---|---|
| AMPHETAMINE | 1100 | 41 | **4,725.01** | 115.24 |
| METHAMPHETAMINE | 1105 | 1 | **8.84** | 8.84 |
| LISDEXAMFETAMINE | 1205 | 23 | **1,519.82** | 66.08 |
| METHYLPHENIDATE (DL;D;L;ISOMERS) | 1724 | 51 | **4,229.65** | 82.93 |
| CODEINE | 9050 | 59 | **1,555.92** | 26.37 |
| BUPRENORPHINE | 9064 | 62 | **2,864.72** | 46.21 |
| OXYCODONE | 9143 | 78 | **13,918.16** | 178.44 |
| HYDROMORPHONE | 9150 | 58 | **3,578.71** | 61.70 |
| HYDROCODONE | 9193 | 56 | **1,186.74** | 21.19 |
| MEPERIDINE (PETHIDINE) | 9230 | 27 | **132.58** | 4.91 |
| METHADONE | 9250B | 46 | **3,934.82** | 85.54 |
| MORPHINE | 9300 | 59 | **8,013.75** | 135.83 |
| OXYMORPHONE | 9652 | 1 | **0.89** | 0.89 |
| TAPENTADOL | 9780 | 7 | **207.00** | 29.57 |
| FENTANYL BASE | 9801 | 63 | **286.99** | 4.56 |

**BUSINESS ACTIVITY: C - PRACTITIONERS**

| DRUG NAME | DRUG CODE | NUMBER OF REGISTRANT SOLD TO | TOTAL GRAMS | AVG GRAMS |
|---|---|---|---|---|
| METHYLPHENIDATE (DL;D;L;ISOMERS) | 1724 | 1 | **0.86** | 0.86 |
| CODEINE | 9050 | 7 | **124.68** | 17.81 |
| BUPRENORPHINE | 9064 | 327 | **49.26** | 0.15 |
| OXYCODONE | 9143 | 26 | **65.98** | 2.54 |
| HYDROMORPHONE | 9150 | 154 | **761.66** | 4.95 |
| HYDROCODONE | 9193 | 67 | **122.53** | 1.83 |
| MEPERIDINE (PETHIDINE) | 9230 | 12 | **32.25** | 2.69 |
| METHADONE | 9250B | 14 | **54.56** | 3.90 |
| MORPHINE | 9300 | 89 | **93.86** | 1.05 |
| FENTANYL BASE | 9801 | 215 | **32.40** | 0.15 |

**BUSINESS ACTIVITY: NTP - NARCOTIC TREATMENT PROGRAMS**

| DRUG NAME | DRUG CODE | NUMBER OF REGISTRANT SOLD TO | TOTAL GRAMS | AVG GRAMS |
|---|---|---|---|---|
| BUPRENORPHINE | 9064 | 8 | **252.86** | 31.61 |
| METHADONE | 9250B | 43 | **478,334.51** | 11,124.06 |

**STATE: DC - DISTRICT OF COLUMBIA**

**BUSINESS ACTIVITY: A - PHARMACIES**

| DRUG NAME | DRUG CODE | NUMBER OF REGISTRANT SOLD TO | TOTAL GRAMS | AVG GRAMS |
|---|---|---|---|---|
| AMPHETAMINE | 1100 | 143 | **45,849.12** | 320.62 |
| METHAMPHETAMINE | 1105 | 7 | **26.92** | 3.85 |
| LISDEXAMFETAMINE | 1205 | 130 | **22,359.17** | 171.99 |
| METHYLPHENIDATE (DL;D;L;ISOMERS) | 1724 | 136 | **27,134.08** | 199.52 |
| CODEINE | 9050 | 151 | **25,904.81** | 171.56 |
| BUPRENORPHINE | 9064 | 137 | **14,533.87** | 106.09 |
| OXYCODONE | 9143 | 145 | **33,487.31** | 230.95 |
| HYDROMORPHONE | 9150 | 98 | **624.34** | 6.37 |
| HYDROCODONE | 9193 | 139 | **2,688.12** | 19.34 |
| MEPERIDINE (PETHIDINE) | 9230 | 6 | **33.77** | 5.63 |
| METHADONE | 9250B | 47 | **717.93** | 15.28 |
| MORPHINE | 9300 | 110 | **6,271.64** | 57.01 |
| OXYMORPHONE | 9652 | 12 | **151.84** | 12.65 |
| TAPENTADOL | 9780 | 32 | **1,997.00** | 62.41 |
| FENTANYL BASE | 9801 | 76 | **91.93** | 1.21 |

**BUSINESS ACTIVITY: B - HOSPITALS**

| DRUG NAME | DRUG CODE | NUMBER OF REGISTRANT SOLD TO | TOTAL GRAMS | AVG GRAMS |
|---|---|---|---|---|
| AMPHETAMINE | 1100 | 10 | **2,932.31** | 293.23 |
| LISDEXAMFETAMINE | 1205 | 6 | **341.66** | 56.94 |

| | | | | |
|---|---|---|---|---|
| METHYLPHENIDATE (DL;D;L;ISOMERS) | 1724 | 13 | **2,602.64** | 200.20 |
| CODEINE | 9050 | 15 | **727.89** | 48.53 |
| BUPRENORPHINE | 9064 | 25 | **638.27** | 25.53 |
| OXYCODONE | 9143 | 20 | **6,356.63** | 317.83 |
| HYDROMORPHONE | 9150 | 22 | **927.55** | 42.16 |
| HYDROCODONE | 9193 | 16 | **363.29** | 22.71 |
| MEPERIDINE (PETHIDINE) | 9230 | 6 | **217.88** | 36.31 |
| METHADONE | 9250B | 15 | **739.90** | 49.33 |
| MORPHINE | 9300 | 23 | **4,051.77** | 176.16 |
| OXYMORPHONE | 9652 | 2 | **3.12** | 1.56 |
| TAPENTADOL | 9780 | 2 | **48.50** | 24.25 |
| FENTANYL BASE | 9801 | 19 | **117.67** | 6.19 |

**BUSINESS ACTIVITY: C - PRACTITIONERS**

| DRUG NAME | DRUG CODE | NUMBER OF REGISTRANT SOLD TO | TOTAL GRAMS | AVG GRAMS |
|---|---|---|---|---|
| BUPRENORPHINE | 9064 | 20 | **1.65** | 0.08 |
| OXYCODONE | 9143 | 4 | **2.24** | 0.56 |
| HYDROMORPHONE | 9150 | 6 | **18.98** | 3.16 |
| HYDROCODONE | 9193 | 3 | **2.12** | 0.71 |
| MEPERIDINE (PETHIDINE) | 9230 | 3 | **12.53** | 4.18 |
| MORPHINE | 9300 | 6 | **2.56** | 0.43 |
| FENTANYL BASE | 9801 | 18 | **0.44** | 0.02 |

**BUSINESS ACTIVITY: D - TEACHING INSTITUTIONS**

| DRUG NAME | DRUG CODE | NUMBER OF REGISTRANT SOLD TO | TOTAL GRAMS | AVG GRAMS |
|---|---|---|---|---|
| BUPRENORPHINE | 9064 | 1 | **0.00** | 0.00 |

**BUSINESS ACTIVITY: M - MID-LEVEL PRACTITIONERS**

| DRUG NAME | DRUG CODE | NUMBER OF REGISTRANT SOLD TO | TOTAL GRAMS | AVG GRAMS |
|---|---|---|---|---|
| FENTANYL BASE | 9801 | 1 | **0.08** | 0.08 |

**BUSINESS ACTIVITY: NTP - NARCOTIC TREATMENT PROGRAMS**

| DRUG NAME | DRUG CODE | NUMBER OF REGISTRANT SOLD TO | TOTAL GRAMS | AVG GRAMS |
|---|---|---|---|---|
| BUPRENORPHINE | 9064 | 1 | **1.44** | 1.44 |
| METHADONE | 9250B | 5 | **38,655.97** | 7,731.19 |

**STATE: DE - DELAWARE**

**BUSINESS ACTIVITY: A - PHARMACIES**

| DRUG NAME | DRUG CODE | NUMBER OF REGISTRANT SOLD TO | TOTAL GRAMS | AVG GRAMS |
|---|---|---|---|---|
| AMPHETAMINE | 1100 | 200 | **85,737.55** | 428.69 |
| METHAMPHETAMINE | 1105 | 4 | **21.29** | 5.32 |
| LISDEXAMFETAMINE | 1205 | 193 | **51,659.02** | 267.66 |
| METHYLPHENIDATE (DL;D;L;ISOMERS) | 1724 | 198 | **64,062.03** | 323.55 |
| CODEINE | 9050 | 196 | **23,506.61** | 119.93 |
| BUPRENORPHINE | 9064 | 196 | **16,181.43** | 82.56 |
| OXYCODONE | 9143 | 201 | **171,448.69** | 852.98 |
| HYDROMORPHONE | 9150 | 196 | **3,973.50** | 20.27 |
| HYDROCODONE | 9193 | 200 | **15,481.51** | 77.41 |
| MEPERIDINE (PETHIDINE) | 9230 | 10 | **24.18** | 2.42 |
| METHADONE | 9250B | 153 | **5,167.04** | 33.77 |
| MORPHINE | 9300 | 196 | **51,964.55** | 265.13 |
| OXYMORPHONE | 9652 | 89 | **2,918.91** | 32.80 |
| TAPENTADOL | 9780 | 138 | **15,378.00** | 111.43 |
| FENTANYL BASE | 9801 | 173 | **452.13** | 2.61 |

**BUSINESS ACTIVITY: B - HOSPITALS**

| DRUG NAME | DRUG CODE | NUMBER OF REGISTRANT SOLD TO | TOTAL GRAMS | AVG GRAMS |
|---|---|---|---|---|
| AMPHETAMINE | 1100 | 15 | **2,361.92** | 157.46 |
| LISDEXAMFETAMINE | 1205 | 8 | **607.58** | 75.95 |
| METHYLPHENIDATE (DL;D;L;ISOMERS) | 1724 | 17 | **1,558.78** | 91.69 |
| CODEINE | 9050 | 16 | **617.83** | 38.61 |
| BUPRENORPHINE | 9064 | 18 | **1,045.59** | 58.09 |
| OXYCODONE | 9143 | 27 | **7,425.81** | 275.03 |
| HYDROMORPHONE | 9150 | 27 | **601.50** | 22.28 |
| HYDROCODONE | 9193 | 24 | **560.63** | 23.36 |
| MEPERIDINE (PETHIDINE) | 9230 | 10 | **135.08** | 13.51 |
| METHADONE | 9250B | 15 | **1,380.42** | 92.03 |

| | | | | |
|---|---|---|---|---|
| MORPHINE | 9300 | 26 | **3,711.14** | 142.74 |
| OXYMORPHONE | 9652 | 3 | **7.23** | 2.41 |
| TAPENTADOL | 9780 | 4 | **155.50** | 38.88 |
| FENTANYL BASE | 9801 | 32 | **85.49** | 2.67 |

**BUSINESS ACTIVITY: C - PRACTITIONERS**

| DRUG NAME | DRUG CODE | NUMBER OF REGISTRANT SOLD TO | TOTAL GRAMS | AVG GRAMS |
|---|---|---|---|---|
| CODEINE | 9050 | 12 | **496.98** | 41.41 |
| BUPRENORPHINE | 9064 | 68 | **7.13** | 0.10 |
| HYDROMORPHONE | 9150 | 41 | **227.71** | 5.55 |
| HYDROCODONE | 9193 | 30 | **114.24** | 3.81 |
| MEPERIDINE (PETHIDINE) | 9230 | 3 | **8.61** | 2.87 |
| METHADONE | 9250B | 6 | **12.70** | 2.12 |
| MORPHINE | 9300 | 10 | **2.71** | 0.27 |
| FENTANYL BASE | 9801 | 56 | **3.50** | 0.06 |

**BUSINESS ACTIVITY: M - MID-LEVEL PRACTITIONERS**

| DRUG NAME | DRUG CODE | NUMBER OF REGISTRANT SOLD TO | TOTAL GRAMS | AVG GRAMS |
|---|---|---|---|---|
| CODEINE | 9050 | 1 | **4.42** | 4.42 |
| MORPHINE | 9300 | 1 | **0.19** | 0.19 |

**BUSINESS ACTIVITY: NTP - NARCOTIC TREATMENT PROGRAMS**

| DRUG NAME | DRUG CODE | NUMBER OF REGISTRANT SOLD TO | TOTAL GRAMS | AVG GRAMS |
|---|---|---|---|---|
| BUPRENORPHINE | 9064 | 15 | **1,692.51** | 112.83 |
| METHADONE | 9250B | 14 | **149,959.58** | 10,711.40 |

**STATE: FL - FLORIDA**

**BUSINESS ACTIVITY: A - PHARMACIES**

| DRUG NAME | DRUG CODE | NUMBER OF REGISTRANT SOLD TO | TOTAL GRAMS | AVG GRAMS |
|---|---|---|---|---|
| AMPHETAMINE | 1100 | 4,318 | **1,435,144.72** | 332.36 |
| METHAMPHETAMINE | 1105 | 91 | **206.91** | 2.27 |
| LISDEXAMFETAMINE | 1205 | 3,915 | **523,937.81** | 133.83 |
| METHYLPHENIDATE (DL;D;L;ISOMERS) | 1724 | 4,136 | **737,941.88** | 178.42 |
| CODEINE | 9050 | 4,295 | **485,385.84** | 113.01 |
| BUPRENORPHINE | 9064 | 4,008 | **248,284.53** | 61.95 |
| OXYCODONE | 9143 | 4,477 | **3,285,213.15** | 733.80 |
| HYDROMORPHONE | 9150 | 3,889 | **116,488.82** | 29.95 |
| HYDROCODONE | 9193 | 4,395 | **965,362.54** | 219.65 |
| MEPERIDINE (PETHIDINE) | 9230 | 196 | **2,341.61** | 11.95 |
| METHADONE | 9250B | 3,204 | **133,165.07** | 41.56 |
| MORPHINE | 9300 | 4,119 | **1,019,026.89** | 247.40 |
| OXYMORPHONE | 9652 | 1,104 | **21,238.58** | 19.24 |
| TAPENTADOL | 9780 | 2,178 | **207,788.50** | 95.40 |
| FENTANYL BASE | 9801 | 3,671 | **10,893.01** | 2.97 |

**BUSINESS ACTIVITY: B - HOSPITALS**

| DRUG NAME | DRUG CODE | NUMBER OF REGISTRANT SOLD TO | TOTAL GRAMS | AVG GRAMS |
|---|---|---|---|---|
| AMPHETAMINE | 1100 | 202 | **33,106.21** | 163.89 |
| LISDEXAMFETAMINE | 1205 | 89 | **7,289.26** | 81.90 |
| METHYLPHENIDATE (DL;D;L;ISOMERS) | 1724 | 263 | **25,035.88** | 95.19 |
| CODEINE | 9050 | 358 | **9,283.91** | 25.93 |
| BUPRENORPHINE | 9064 | 300 | **8,729.94** | 29.10 |
| OXYCODONE | 9143 | 617 | **95,389.34** | 154.60 |
| HYDROMORPHONE | 9150 | 623 | **10,526.37** | 16.90 |
| HYDROCODONE | 9193 | 539 | **32,618.60** | 60.52 |
| MEPERIDINE (PETHIDINE) | 9230 | 374 | **3,651.48** | 9.76 |
| METHADONE | 9250B | 312 | **7,156.85** | 22.94 |
| MORPHINE | 9300 | 640 | **55,322.61** | 86.44 |
| OXYMORPHONE | 9652 | 11 | **27.34** | 2.49 |
| TAPENTADOL | 9780 | 61 | **2,713.00** | 44.48 |
| FENTANYL BASE | 9801 | 815 | **1,999.99** | 2.45 |

**BUSINESS ACTIVITY: C - PRACTITIONERS**

| DRUG NAME | DRUG CODE | NUMBER OF REGISTRANT SOLD TO | TOTAL GRAMS | AVG GRAMS |
|---|---|---|---|---|
| AMPHETAMINE | 1100 | 1 | **0.51** | 0.51 |
| CODEINE | 9050 | 56 | **1,283.57** | 22.92 |
| BUPRENORPHINE | 9064 | 2,265 | **686.86** | 0.30 |

| | | | | |
|---|---|---|---|---|
| OXYCODONE | 9143 | 62 | **144.52** | 2.33 |
| HYDROMORPHONE | 9150 | 744 | **4,757.61** | 6.39 |
| HYDROCODONE | 9193 | 763 | **3,169.20** | 4.15 |
| MEPERIDINE (PETHIDINE) | 9230 | 195 | **2,049.22** | 10.51 |
| METHADONE | 9250B | 33 | **72.36** | 2.19 |
| MORPHINE | 9300 | 325 | **667.83** | 2.05 |
| FENTANYL BASE | 9801 | 1,235 | **82.48** | 0.07 |

**BUSINESS ACTIVITY: D - TEACHING INSTITUTIONS**

| DRUG NAME | DRUG CODE | NUMBER OF REGISTRANT SOLD TO | TOTAL GRAMS | AVG GRAMS |
|---|---|---|---|---|
| BUPRENORPHINE | 9064 | 1 | **0.02** | 0.02 |
| HYDROMORPHONE | 9150 | 1 | **0.71** | 0.71 |

**BUSINESS ACTIVITY: M - MID-LEVEL PRACTITIONERS**

| DRUG NAME | DRUG CODE | NUMBER OF REGISTRANT SOLD TO | TOTAL GRAMS | AVG GRAMS |
|---|---|---|---|---|
| CODEINE | 9050 | 1 | **2.21** | 2.21 |
| BUPRENORPHINE | 9064 | 8 | **24.50** | 3.06 |
| FENTANYL BASE | 9801 | 1 | **0.01** | 0.01 |

**BUSINESS ACTIVITY: NTP - NARCOTIC TREATMENT PROGRAMS**

| DRUG NAME | DRUG CODE | NUMBER OF REGISTRANT SOLD TO | TOTAL GRAMS | AVG GRAMS |
|---|---|---|---|---|
| BUPRENORPHINE | 9064 | 58 | **1,852.79** | 31.94 |
| METHADONE | 9250B | 63 | **503,231.03** | 7,987.79 |

**STATE: GA - GEORGIA**

**BUSINESS ACTIVITY: A - PHARMACIES**

| DRUG NAME | DRUG CODE | NUMBER OF REGISTRANT SOLD TO | TOTAL GRAMS | AVG GRAMS |
|---|---|---|---|---|
| AMPHETAMINE | 1100 | 2,108 | **840,531.57** | 398.73 |
| METHAMPHETAMINE | 1105 | 30 | **98.03** | 3.27 |
| LISDEXAMFETAMINE | 1205 | 2,005 | **360,161.50** | 179.63 |
| METHYLPHENIDATE (DL;D;L;ISOMERS) | 1724 | 2,081 | **568,988.59** | 273.42 |
| CODEINE | 9050 | 2,086 | **294,804.50** | 141.33 |
| BUPRENORPHINE | 9064 | 2,021 | **81,972.36** | 40.56 |
| OXYCODONE | 9143 | 2,134 | **1,306,434.72** | 612.20 |
| HYDROMORPHONE | 9150 | 1,839 | **17,073.92** | 9.28 |
| HYDROCODONE | 9193 | 2,141 | **676,036.74** | 315.76 |
| MEPERIDINE (PETHIDINE) | 9230 | 288 | **1,991.81** | 6.92 |
| METHADONE | 9250B | 1,622 | **53,913.90** | 33.24 |
| MORPHINE | 9300 | 2,029 | **285,799.45** | 140.86 |
| OXYMORPHONE | 9652 | 465 | **6,330.83** | 13.61 |
| TAPENTADOL | 9780 | 1,311 | **151,749.00** | 115.75 |
| FENTANYL BASE | 9801 | 1,908 | **4,875.11** | 2.56 |

**BUSINESS ACTIVITY: B - HOSPITALS**

| DRUG NAME | DRUG CODE | NUMBER OF REGISTRANT SOLD TO | TOTAL GRAMS | AVG GRAMS |
|---|---|---|---|---|
| AMPHETAMINE | 1100 | 93 | **14,506.35** | 155.98 |
| METHAMPHETAMINE | 1105 | 2 | **5.22** | 2.61 |
| LISDEXAMFETAMINE | 1205 | 52 | **4,177.93** | 80.34 |
| METHYLPHENIDATE (DL;D;L;ISOMERS) | 1724 | 137 | **14,049.78** | 102.55 |
| CODEINE | 9050 | 179 | **6,097.11** | 34.06 |
| BUPRENORPHINE | 9064 | 120 | **2,110.61** | 17.59 |
| OXYCODONE | 9143 | 224 | **45,622.93** | 203.67 |
| HYDROMORPHONE | 9150 | 199 | **4,561.85** | 22.92 |
| HYDROCODONE | 9193 | 219 | **23,616.57** | 107.84 |
| MEPERIDINE (PETHIDINE) | 9230 | 136 | **2,765.92** | 20.34 |
| METHADONE | 9250B | 157 | **2,577.03** | 16.41 |
| MORPHINE | 9300 | 219 | **23,841.25** | 108.86 |
| OXYMORPHONE | 9652 | 3 | **8.30** | 2.77 |
| TAPENTADOL | 9780 | 52 | **3,288.00** | 63.23 |
| FENTANYL BASE | 9801 | 203 | **1,390.02** | 6.85 |

**BUSINESS ACTIVITY: C - PRACTITIONERS**

| DRUG NAME | DRUG CODE | NUMBER OF REGISTRANT SOLD TO | TOTAL GRAMS | AVG GRAMS |
|---|---|---|---|---|
| AMPHETAMINE | 1100 | 2 | **16.89** | 8.45 |
| METHYLPHENIDATE (DL;D;L;ISOMERS) | 1724 | 1 | **0.43** | 0.43 |
| CODEINE | 9050 | 73 | **1,450.91** | 19.88 |
| BUPRENORPHINE | 9064 | 858 | **679.44** | 0.79 |

| | | | | |
|---|---|---|---|---|
| OXYCODONE | 9143 | 102 | **734.46** | 7.20 |
| HYDROMORPHONE | 9150 | 412 | **1,878.75** | 4.56 |
| HYDROCODONE | 9193 | 348 | **1,447.19** | 4.16 |
| MEPERIDINE (PETHIDINE) | 9230 | 138 | **1,142.10** | 8.28 |
| METHADONE | 9250B | 11 | **53.93** | 4.90 |
| MORPHINE | 9300 | 255 | **375.49** | 1.47 |
| FENTANYL BASE | 9801 | 874 | **55.75** | 0.06 |

**BUSINESS ACTIVITY: M - MID-LEVEL PRACTITIONERS**

| DRUG NAME | DRUG CODE | NUMBER OF REGISTRANT SOLD TO | TOTAL GRAMS | AVG GRAMS |
|---|---|---|---|---|
| CODEINE | 9050 | 2 | **1.25** | 0.63 |
| BUPRENORPHINE | 9064 | 3 | **304.15** | 101.38 |

**BUSINESS ACTIVITY: NTP - NARCOTIC TREATMENT PROGRAMS**

| DRUG NAME | DRUG CODE | NUMBER OF REGISTRANT SOLD TO | TOTAL GRAMS | AVG GRAMS |
|---|---|---|---|---|
| BUPRENORPHINE | 9064 | 37 | **1,857.75** | 50.21 |
| METHADONE | 9250B | 75 | **366,422.71** | 4,885.64 |

**STATE: GU - GUAM**

**BUSINESS ACTIVITY: A - PHARMACIES**

| DRUG NAME | DRUG CODE | NUMBER OF REGISTRANT SOLD TO | TOTAL GRAMS | AVG GRAMS |
|---|---|---|---|---|
| AMPHETAMINE | 1100 | 13 | **994.33** | 76.49 |
| LISDEXAMFETAMINE | 1205 | 3 | **18.50** | 6.17 |
| METHYLPHENIDATE (DL;D;L;ISOMERS) | 1724 | 11 | **611.40** | 55.58 |
| CODEINE | 9050 | 21 | **3,047.73** | 145.13 |
| OXYCODONE | 9143 | 14 | **2,092.88** | 149.49 |
| HYDROMORPHONE | 9150 | 7 | **45.39** | 6.48 |
| HYDROCODONE | 9193 | 13 | **311.18** | 23.94 |
| METHADONE | 9250B | 2 | **34.88** | 17.44 |
| MORPHINE | 9300 | 11 | **2,256.90** | 205.17 |
| FENTANYL BASE | 9801 | 7 | **14.71** | 2.10 |

**BUSINESS ACTIVITY: B - HOSPITALS**

| DRUG NAME | DRUG CODE | NUMBER OF REGISTRANT SOLD TO | TOTAL GRAMS | AVG GRAMS |
|---|---|---|---|---|
| AMPHETAMINE | 1100 | 3 | **879.17** | 293.06 |
| LISDEXAMFETAMINE | 1205 | 2 | **57.23** | 28.62 |
| METHYLPHENIDATE (DL;D;L;ISOMERS) | 1724 | 3 | **1,047.35** | 349.12 |
| CODEINE | 9050 | 5 | **406.69** | 81.34 |
| BUPRENORPHINE | 9064 | 2 | **3.37** | 1.68 |
| OXYCODONE | 9143 | 6 | **647.99** | 108.00 |
| HYDROMORPHONE | 9150 | 7 | **25.54** | 3.65 |
| HYDROCODONE | 9193 | 6 | **99.88** | 16.65 |
| MEPERIDINE (PETHIDINE) | 9230 | 3 | **5.99** | 2.00 |
| METHADONE | 9250B | 3 | **17.44** | 5.81 |
| MORPHINE | 9300 | 6 | **432.29** | 72.05 |
| FENTANYL BASE | 9801 | 5 | **20.48** | 4.10 |

**BUSINESS ACTIVITY: C - PRACTITIONERS**

| DRUG NAME | DRUG CODE | NUMBER OF REGISTRANT SOLD TO | TOTAL GRAMS | AVG GRAMS |
|---|---|---|---|---|
| CODEINE | 9050 | 1 | **8.84** | 8.84 |
| BUPRENORPHINE | 9064 | 1 | **0.11** | 0.11 |
| HYDROMORPHONE | 9150 | 2 | **35.51** | 17.75 |
| MEPERIDINE (PETHIDINE) | 9230 | 2 | **138.35** | 69.18 |
| MORPHINE | 9300 | 3 | **2.14** | 0.71 |
| FENTANYL BASE | 9801 | 5 | **0.46** | 0.09 |

**STATE: HI - HAWAII**

**BUSINESS ACTIVITY: A - PHARMACIES**

| DRUG NAME | DRUG CODE | NUMBER OF REGISTRANT SOLD TO | TOTAL GRAMS | AVG GRAMS |
|---|---|---|---|---|
| AMPHETAMINE | 1100 | 210 | **35,782.39** | 170.39 |
| METHAMPHETAMINE | 1105 | 6 | **13.66** | 2.28 |
| LISDEXAMFETAMINE | 1205 | 163 | **6,075.25** | 37.27 |
| METHYLPHENIDATE (DL;D;L;ISOMERS) | 1724 | 206 | **28,748.54** | 139.56 |
| CODEINE | 9050 | 220 | **32,749.98** | 148.86 |
| BUPRENORPHINE | 9064 | 195 | **9,390.21** | 48.15 |
| OXYCODONE | 9143 | 217 | **92,781.16** | 427.56 |
| HYDROMORPHONE | 9150 | 134 | **1,417.22** | 10.58 |

| | | | | |
|---|---|---|---|---|
| HYDROCODONE | 9193 | 219 | **41,903.92** | 191.34 |
| MEPERIDINE (PETHIDINE) | 9230 | 3 | **9.15** | 3.05 |
| METHADONE | 9250B | 143 | **6,671.06** | 46.65 |
| MORPHINE | 9300 | 195 | **42,893.46** | 219.97 |
| OXYMORPHONE | 9652 | 22 | **342.16** | 15.55 |
| TAPENTADOL | 9780 | 49 | **4,527.50** | 92.40 |
| FENTANYL BASE | 9801 | 157 | **320.32** | 2.04 |

**BUSINESS ACTIVITY: B - HOSPITALS**

| DRUG NAME | DRUG CODE | NUMBER OF REGISTRANT SOLD TO | TOTAL GRAMS | AVG GRAMS |
|---|---|---|---|---|
| AMPHETAMINE | 1100 | 16 | **8,427.81** | 526.74 |
| LISDEXAMFETAMINE | 1205 | 12 | **1,436.00** | 119.67 |
| METHYLPHENIDATE (DL;D;L;ISOMERS) | 1724 | 22 | **9,414.62** | 427.94 |
| CODEINE | 9050 | 28 | **1,904.12** | 68.00 |
| BUPRENORPHINE | 9064 | 20 | **279.73** | 13.99 |
| OXYCODONE | 9143 | 37 | **11,401.33** | 308.14 |
| HYDROMORPHONE | 9150 | 34 | **341.77** | 10.05 |
| HYDROCODONE | 9193 | 31 | **3,306.29** | 106.65 |
| MEPERIDINE (PETHIDINE) | 9230 | 17 | **185.74** | 10.93 |
| METHADONE | 9250B | 23 | **621.97** | 27.04 |
| MORPHINE | 9300 | 36 | **4,729.74** | 131.38 |
| OXYMORPHONE | 9652 | 3 | **200.50** | 66.83 |
| TAPENTADOL | 9780 | 7 | **458.00** | 65.43 |
| FENTANYL BASE | 9801 | 42 | **100.49** | 2.39 |

**BUSINESS ACTIVITY: C - PRACTITIONERS**

| DRUG NAME | DRUG CODE | NUMBER OF REGISTRANT SOLD TO | TOTAL GRAMS | AVG GRAMS |
|---|---|---|---|---|
| AMPHETAMINE | 1100 | 1 | **24.40** | 24.40 |
| CODEINE | 9050 | 13 | **251.38** | 19.34 |
| BUPRENORPHINE | 9064 | 104 | **7.26** | 0.07 |
| OXYCODONE | 9143 | 18 | **1,503.30** | 83.52 |
| HYDROMORPHONE | 9150 | 46 | **270.63** | 5.88 |
| HYDROCODONE | 9193 | 38 | **1,216.37** | 32.01 |
| MEPERIDINE (PETHIDINE) | 9230 | 18 | **101.09** | 5.62 |
| METHADONE | 9250B | 3 | **6.62** | 2.21 |
| MORPHINE | 9300 | 21 | **139.40** | 6.64 |
| FENTANYL BASE | 9801 | 55 | **3.16** | 0.06 |

**BUSINESS ACTIVITY: M - MID-LEVEL PRACTITIONERS**

| DRUG NAME | DRUG CODE | NUMBER OF REGISTRANT SOLD TO | TOTAL GRAMS | AVG GRAMS |
|---|---|---|---|---|
| MORPHINE | 9300 | 1 | **0.75** | 0.75 |
| FENTANYL BASE | 9801 | 1 | **0.02** | 0.02 |

**BUSINESS ACTIVITY: NTP - NARCOTIC TREATMENT PROGRAMS**

| DRUG NAME | DRUG CODE | NUMBER OF REGISTRANT SOLD TO | TOTAL GRAMS | AVG GRAMS |
|---|---|---|---|---|
| METHADONE | 9250B | 4 | **21,551.46** | 5,387.87 |

**STATE: IA - IOWA**

**BUSINESS ACTIVITY: A - PHARMACIES**

| DRUG NAME | DRUG CODE | NUMBER OF REGISTRANT SOLD TO | TOTAL GRAMS | AVG GRAMS |
|---|---|---|---|---|
| AMPHETAMINE | 1100 | 666 | **222,714.33** | 334.41 |
| METHAMPHETAMINE | 1105 | 13 | **60.26** | 4.64 |
| LISDEXAMFETAMINE | 1205 | 646 | **129,198.99** | 200.00 |
| METHYLPHENIDATE (DL;D;L;ISOMERS) | 1724 | 667 | **295,944.62** | 443.70 |
| CODEINE | 9050 | 639 | **47,204.62** | 73.87 |
| BUPRENORPHINE | 9064 | 576 | **10,321.63** | 17.92 |
| OXYCODONE | 9143 | 671 | **128,028.02** | 190.80 |
| HYDROMORPHONE | 9150 | 613 | **5,695.95** | 9.29 |
| HYDROCODONE | 9193 | 679 | **139,205.28** | 205.02 |
| MEPERIDINE (PETHIDINE) | 9230 | 22 | **225.28** | 10.24 |
| METHADONE | 9250B | 466 | **10,179.52** | 21.84 |
| MORPHINE | 9300 | 657 | **83,424.99** | 126.98 |
| OXYMORPHONE | 9652 | 75 | **790.13** | 10.54 |
| TAPENTADOL | 9780 | 312 | **24,900.00** | 79.81 |
| FENTANYL BASE | 9801 | 635 | **1,752.92** | 2.76 |

**BUSINESS ACTIVITY: B - HOSPITALS**

| DRUG NAME | DRUG CODE | NUMBER OF REGISTRANT SOLD TO | TOTAL GRAMS | AVG GRAMS |
|---|---|---|---|---|
| AMPHETAMINE | 1100 | 61 | **5,568.24** | 91.28 |
| LISDEXAMFETAMINE | 1205 | 34 | **1,818.70** | 53.49 |
| METHYLPHENIDATE (DL;D;L;ISOMERS) | 1724 | 84 | **6,472.62** | 77.06 |
| CODEINE | 9050 | 97 | **1,342.50** | 13.84 |
| BUPRENORPHINE | 9064 | 75 | **789.46** | 10.53 |
| OXYCODONE | 9143 | 145 | **10,169.40** | 70.13 |
| HYDROMORPHONE | 9150 | 135 | **1,606.66** | 11.90 |
| HYDROCODONE | 9193 | 148 | **8,362.41** | 56.50 |
| MEPERIDINE (PETHIDINE) | 9230 | 77 | **1,225.44** | 15.91 |
| METHADONE | 9250B | 86 | **752.91** | 8.75 |
| MORPHINE | 9300 | 144 | **8,740.11** | 60.70 |
| TAPENTADOL | 9780 | 25 | **287.50** | 11.50 |
| FENTANYL BASE | 9801 | 152 | **259.07** | 1.70 |

**BUSINESS ACTIVITY: C - PRACTITIONERS**

| DRUG NAME | DRUG CODE | NUMBER OF REGISTRANT SOLD TO | TOTAL GRAMS | AVG GRAMS |
|---|---|---|---|---|
| AMPHETAMINE | 1100 | 1 | **2.82** | 2.82 |
| CODEINE | 9050 | 12 | **219.62** | 18.30 |
| BUPRENORPHINE | 9064 | 303 | **30.97** | 0.10 |
| OXYCODONE | 9143 | 8 | **130.35** | 16.29 |
| HYDROMORPHONE | 9150 | 88 | **451.80** | 5.13 |
| HYDROCODONE | 9193 | 109 | **301.61** | 2.77 |
| MEPERIDINE (PETHIDINE) | 9230 | 19 | **888.17** | 46.75 |
| MORPHINE | 9300 | 55 | **28.90** | 0.53 |
| FENTANYL BASE | 9801 | 108 | **16.51** | 0.15 |

**BUSINESS ACTIVITY: D - TEACHING INSTITUTIONS**

| DRUG NAME | DRUG CODE | NUMBER OF REGISTRANT SOLD TO | TOTAL GRAMS | AVG GRAMS |
|---|---|---|---|---|
| HYDROMORPHONE | 9150 | 1 | **1.42** | 1.42 |

**BUSINESS ACTIVITY: M - MID-LEVEL PRACTITIONERS**

| DRUG NAME | DRUG CODE | NUMBER OF REGISTRANT SOLD TO | TOTAL GRAMS | AVG GRAMS |
|---|---|---|---|---|
| BUPRENORPHINE | 9064 | 1 | **0.00** | 0.00 |
| FENTANYL BASE | 9801 | 1 | **0.00** | 0.00 |

**BUSINESS ACTIVITY: NTP - NARCOTIC TREATMENT PROGRAMS**

| DRUG NAME | DRUG CODE | NUMBER OF REGISTRANT SOLD TO | TOTAL GRAMS | AVG GRAMS |
|---|---|---|---|---|
| BUPRENORPHINE | 9064 | 19 | **1,060.20** | 55.80 |
| METHADONE | 9250B | 18 | **42,958.03** | 2,386.56 |

**STATE: ID - IDAHO**

**BUSINESS ACTIVITY: A - PHARMACIES**

| DRUG NAME | DRUG CODE | NUMBER OF REGISTRANT SOLD TO | TOTAL GRAMS | AVG GRAMS |
|---|---|---|---|---|
| AMPHETAMINE | 1100 | 305 | **134,243.49** | 440.14 |
| METHAMPHETAMINE | 1105 | 14 | **51.02** | 3.64 |
| LISDEXAMFETAMINE | 1205 | 291 | **43,575.44** | 149.74 |
| METHYLPHENIDATE (DL;D;L;ISOMERS) | 1724 | 306 | **111,376.74** | 363.98 |
| CODEINE | 9050 | 303 | **37,284.49** | 123.05 |
| BUPRENORPHINE | 9064 | 304 | **22,331.05** | 73.46 |
| OXYCODONE | 9143 | 308 | **171,335.32** | 556.28 |
| HYDROMORPHONE | 9150 | 285 | **4,567.07** | 16.02 |
| HYDROCODONE | 9193 | 313 | **164,838.91** | 526.64 |
| MEPERIDINE (PETHIDINE) | 9230 | 30 | **275.83** | 9.19 |
| METHADONE | 9250B | 272 | **13,677.97** | 50.29 |
| MORPHINE | 9300 | 304 | **69,513.59** | 228.66 |
| OXYMORPHONE | 9652 | 113 | **1,756.01** | 15.54 |
| TAPENTADOL | 9780 | 168 | **16,003.50** | 95.26 |
| FENTANYL BASE | 9801 | 292 | **1,163.35** | 3.98 |

**BUSINESS ACTIVITY: B - HOSPITALS**

| DRUG NAME | DRUG CODE | NUMBER OF REGISTRANT SOLD TO | TOTAL GRAMS | AVG GRAMS |
|---|---|---|---|---|
| AMPHETAMINE | 1100 | 28 | **1,690.79** | 60.39 |
| LISDEXAMFETAMINE | 1205 | 11 | **254.36** | 23.12 |
| METHYLPHENIDATE (DL;D;L;ISOMERS) | 1724 | 37 | **1,824.01** | 49.30 |
| CODEINE | 9050 | 37 | **1,111.85** | 30.05 |
| BUPRENORPHINE | 9064 | 42 | **688.02** | 16.38 |

| | | | | |
|---|---|---|---|---|
| OXYCODONE | 9143 | 62 | **8,356.63** | 134.78 |
| HYDROMORPHONE | 9150 | 53 | **566.09** | 10.68 |
| HYDROCODONE | 9193 | 64 | **6,877.28** | 107.46 |
| MEPERIDINE (PETHIDINE) | 9230 | 35 | **386.62** | 11.05 |
| METHADONE | 9250B | 41 | **789.22** | 19.25 |
| MORPHINE | 9300 | 60 | **5,760.89** | 96.01 |
| OXYMORPHONE | 9652 | 4 | **20.56** | 5.14 |
| TAPENTADOL | 9780 | 7 | **162.00** | 23.14 |
| FENTANYL BASE | 9801 | 58 | **150.33** | 2.59 |

**BUSINESS ACTIVITY: C - PRACTITIONERS**

| DRUG NAME | DRUG CODE | NUMBER OF REGISTRANT SOLD TO | TOTAL GRAMS | AVG GRAMS |
|---|---|---|---|---|
| CODEINE | 9050 | 19 | **2,092.17** | 110.11 |
| BUPRENORPHINE | 9064 | 155 | **32.23** | 0.21 |
| OXYCODONE | 9143 | 17 | **36.87** | 2.17 |
| HYDROMORPHONE | 9150 | 94 | **617.65** | 6.57 |
| HYDROCODONE | 9193 | 77 | **215.12** | 2.79 |
| MEPERIDINE (PETHIDINE) | 9230 | 17 | **41.72** | 2.45 |
| METHADONE | 9250B | 8 | **19.50** | 2.44 |
| MORPHINE | 9300 | 66 | **75.39** | 1.14 |
| FENTANYL BASE | 9801 | 130 | **23.84** | 0.18 |

**BUSINESS ACTIVITY: D - TEACHING INSTITUTIONS**

| DRUG NAME | DRUG CODE | NUMBER OF REGISTRANT SOLD TO | TOTAL GRAMS | AVG GRAMS |
|---|---|---|---|---|
| BUPRENORPHINE | 9064 | 1 | **0.00** | 0.00 |

**BUSINESS ACTIVITY: M - MID-LEVEL PRACTITIONERS**

| DRUG NAME | DRUG CODE | NUMBER OF REGISTRANT SOLD TO | TOTAL GRAMS | AVG GRAMS |
|---|---|---|---|---|
| CODEINE | 9050 | 4 | **1,224.62** | 306.15 |
| BUPRENORPHINE | 9064 | 4 | **4.25** | 1.06 |
| HYDROMORPHONE | 9150 | 1 | **0.74** | 0.74 |
| MEPERIDINE (PETHIDINE) | 9230 | 1 | **0.54** | 0.54 |
| MORPHINE | 9300 | 2 | **0.45** | 0.23 |
| FENTANYL BASE | 9801 | 4 | **0.17** | 0.04 |

**BUSINESS ACTIVITY: NTP - NARCOTIC TREATMENT PROGRAMS**

| DRUG NAME | DRUG CODE | NUMBER OF REGISTRANT SOLD TO | TOTAL GRAMS | AVG GRAMS |
|---|---|---|---|---|
| BUPRENORPHINE | 9064 | 4 | **227.61** | 56.90 |
| METHADONE | 9250B | 4 | **30,159.17** | 7,539.79 |

**STATE: IL - ILLINOIS**

**BUSINESS ACTIVITY: A - PHARMACIES**

| DRUG NAME | DRUG CODE | NUMBER OF REGISTRANT SOLD TO | TOTAL GRAMS | AVG GRAMS |
|---|---|---|---|---|
| AMPHETAMINE | 1100 | 2,046 | **853,569.06** | 417.19 |
| METHAMPHETAMINE | 1105 | 49 | **147.03** | 3.00 |
| LISDEXAMFETAMINE | 1205 | 1,895 | **296,556.05** | 156.49 |
| METHYLPHENIDATE (DL;D;L;ISOMERS) | 1724 | 2,024 | **604,697.93** | 298.76 |
| CODEINE | 9050 | 2,109 | **445,479.29** | 211.23 |
| BUPRENORPHINE | 9064 | 1,938 | **83,434.25** | 43.05 |
| OXYCODONE | 9143 | 2,006 | **391,149.35** | 194.99 |
| HYDROMORPHONE | 9150 | 1,700 | **21,526.18** | 12.66 |
| HYDROCODONE | 9193 | 2,119 | **782,606.35** | 369.33 |
| MEPERIDINE (PETHIDINE) | 9230 | 42 | **192.93** | 4.59 |
| METHADONE | 9250B | 1,318 | **29,163.79** | 22.13 |
| MORPHINE | 9300 | 1,901 | **277,610.91** | 146.03 |
| OXYMORPHONE | 9652 | 408 | **4,260.13** | 10.44 |
| TAPENTADOL | 9780 | 923 | **78,401.00** | 84.94 |
| FENTANYL BASE | 9801 | 1,801 | **5,448.33** | 3.03 |

**BUSINESS ACTIVITY: B - HOSPITALS**

| DRUG NAME | DRUG CODE | NUMBER OF REGISTRANT SOLD TO | TOTAL GRAMS | AVG GRAMS |
|---|---|---|---|---|
| AMPHETAMINE | 1100 | 131 | **10,244.27** | 78.20 |
| METHAMPHETAMINE | 1105 | 1 | **0.80** | 0.80 |
| LISDEXAMFETAMINE | 1205 | 58 | **2,628.04** | 45.31 |
| METHYLPHENIDATE (DL;D;L;ISOMERS) | 1724 | 187 | **9,580.47** | 51.23 |
| CODEINE | 9050 | 226 | **7,108.24** | 31.45 |
| BUPRENORPHINE | 9064 | 177 | **2,900.29** | 16.39 |

| | | | | |
|---|---|---|---|---|
| OXYCODONE | 9143 | 223 | **18,913.65** | 84.81 |
| HYDROMORPHONE | 9150 | 231 | **5,632.70** | 24.38 |
| HYDROCODONE | 9193 | 247 | **37,157.32** | 150.43 |
| MEPERIDINE (PETHIDINE) | 9230 | 128 | **1,309.21** | 10.23 |
| METHADONE | 9250B | 188 | **4,795.13** | 25.51 |
| MORPHINE | 9300 | 244 | **24,835.84** | 101.79 |
| OXYMORPHONE | 9652 | 12 | **152.01** | 12.67 |
| TAPENTADOL | 9780 | 40 | **1,374.00** | 34.35 |
| FENTANYL BASE | 9801 | 236 | **1,174.14** | 4.98 |

**BUSINESS ACTIVITY: C - PRACTITIONERS**

| DRUG NAME | DRUG CODE | NUMBER OF REGISTRANT SOLD TO | TOTAL GRAMS | AVG GRAMS |
|---|---|---|---|---|
| CODEINE | 9050 | 102 | **2,242.82** | 21.99 |
| BUPRENORPHINE | 9064 | 954 | **137.93** | 0.14 |
| OXYCODONE | 9143 | 38 | **100.58** | 2.65 |
| HYDROMORPHONE | 9150 | 427 | **2,309.37** | 5.41 |
| HYDROCODONE | 9193 | 455 | **2,174.19** | 4.78 |
| MEPERIDINE (PETHIDINE) | 9230 | 85 | **414.51** | 4.88 |
| METHADONE | 9250B | 36 | **72.09** | 2.00 |
| MORPHINE | 9300 | 178 | **177.15** | 1.00 |
| FENTANYL BASE | 9801 | 603 | **48.41** | 0.08 |

**BUSINESS ACTIVITY: D - TEACHING INSTITUTIONS**

| DRUG NAME | DRUG CODE | NUMBER OF REGISTRANT SOLD TO | TOTAL GRAMS | AVG GRAMS |
|---|---|---|---|---|
| CODEINE | 9050 | 1 | **16.75** | 16.75 |
| BUPRENORPHINE | 9064 | 2 | **0.11** | 0.06 |
| HYDROMORPHONE | 9150 | 1 | **11.08** | 11.08 |
| HYDROCODONE | 9193 | 1 | **3.63** | 3.63 |
| METHADONE | 9250B | 1 | **2.33** | 2.33 |
| MORPHINE | 9300 | 1 | **0.08** | 0.08 |
| FENTANYL BASE | 9801 | 2 | **0.31** | 0.15 |

**BUSINESS ACTIVITY: NTP - NARCOTIC TREATMENT PROGRAMS**

| DRUG NAME | DRUG CODE | NUMBER OF REGISTRANT SOLD TO | TOTAL GRAMS | AVG GRAMS |
|---|---|---|---|---|
| BUPRENORPHINE | 9064 | 15 | **2,007.25** | 133.82 |
| METHADONE | 9250B | 92 | **464,866.64** | 5,052.90 |

**STATE: IN - INDIANA**

**BUSINESS ACTIVITY: A - PHARMACIES**

| DRUG NAME | DRUG CODE | NUMBER OF REGISTRANT SOLD TO | TOTAL GRAMS | AVG GRAMS |
|---|---|---|---|---|
| AMPHETAMINE | 1100 | 1,120 | **591,979.26** | 528.55 |
| METHAMPHETAMINE | 1105 | 31 | **105.66** | 3.41 |
| LISDEXAMFETAMINE | 1205 | 1,111 | **312,853.85** | 281.60 |
| METHYLPHENIDATE (DL;D;L;ISOMERS) | 1724 | 1,118 | **439,467.21** | 393.08 |
| CODEINE | 9050 | 1,090 | **123,997.94** | 113.76 |
| BUPRENORPHINE | 9064 | 1,106 | **142,563.32** | 128.90 |
| OXYCODONE | 9143 | 1,131 | **643,222.56** | 568.72 |
| HYDROMORPHONE | 9150 | 982 | **13,463.61** | 13.71 |
| HYDROCODONE | 9193 | 1,149 | **628,244.64** | 546.78 |
| MEPERIDINE (PETHIDINE) | 9230 | 47 | **200.66** | 4.27 |
| METHADONE | 9250B | 905 | **33,601.80** | 37.13 |
| MORPHINE | 9300 | 1,098 | **210,369.95** | 191.59 |
| OXYMORPHONE | 9652 | 297 | **3,896.16** | 13.12 |
| TAPENTADOL | 9780 | 665 | **89,589.00** | 134.72 |
| FENTANYL BASE | 9801 | 1,050 | **3,403.15** | 3.24 |

**BUSINESS ACTIVITY: B - HOSPITALS**

| DRUG NAME | DRUG CODE | NUMBER OF REGISTRANT SOLD TO | TOTAL GRAMS | AVG GRAMS |
|---|---|---|---|---|
| AMPHETAMINE | 1100 | 128 | **9,208.18** | 71.94 |
| LISDEXAMFETAMINE | 1205 | 83 | **4,789.56** | 57.71 |
| METHYLPHENIDATE (DL;D;L;ISOMERS) | 1724 | 140 | **7,409.92** | 52.93 |
| CODEINE | 9050 | 141 | **2,687.40** | 19.06 |
| BUPRENORPHINE | 9064 | 161 | **3,058.74** | 19.00 |
| OXYCODONE | 9143 | 248 | **27,258.75** | 109.91 |
| HYDROMORPHONE | 9150 | 241 | **4,919.22** | 20.41 |
| HYDROCODONE | 9193 | 262 | **28,506.44** | 108.80 |

| | | | | |
|---|---|---|---|---|
| MEPERIDINE (PETHIDINE) | 9230 | 159 | **3,042.95** | 19.14 |
| METHADONE | 9250B | 148 | **5,473.01** | 36.98 |
| MORPHINE | 9300 | 258 | **17,477.68** | 67.74 |
| OXYMORPHONE | 9652 | 10 | **50.85** | 5.08 |
| TAPENTADOL | 9780 | 24 | **1,149.50** | 47.90 |
| FENTANYL BASE | 9801 | 293 | **602.33** | 2.06 |

**BUSINESS ACTIVITY: C - PRACTITIONERS**

| DRUG NAME | DRUG CODE | NUMBER OF REGISTRANT SOLD TO | TOTAL GRAMS | AVG GRAMS |
|---|---|---|---|---|
| AMPHETAMINE | 1100 | 1 | **23.15** | 23.15 |
| CODEINE | 9050 | 25 | **320.78** | 12.83 |
| BUPRENORPHINE | 9064 | 511 | **98.95** | 0.19 |
| OXYCODONE | 9143 | 14 | **20.40** | 1.46 |
| HYDROMORPHONE | 9150 | 200 | **1,828.04** | 9.14 |
| HYDROCODONE | 9193 | 200 | **511.73** | 2.56 |
| MEPERIDINE (PETHIDINE) | 9230 | 33 | **68.63** | 2.08 |
| METHADONE | 9250B | 4 | **5.90** | 1.48 |
| MORPHINE | 9300 | 95 | **81.54** | 0.86 |
| FENTANYL BASE | 9801 | 229 | **15.33** | 0.07 |

**BUSINESS ACTIVITY: M - MID-LEVEL PRACTITIONERS**

| DRUG NAME | DRUG CODE | NUMBER OF REGISTRANT SOLD TO | TOTAL GRAMS | AVG GRAMS |
|---|---|---|---|---|
| CODEINE | 9050 | 1 | **3.65** | 3.65 |

**BUSINESS ACTIVITY: NTP - NARCOTIC TREATMENT PROGRAMS**

| DRUG NAME | DRUG CODE | NUMBER OF REGISTRANT SOLD TO | TOTAL GRAMS | AVG GRAMS |
|---|---|---|---|---|
| BUPRENORPHINE | 9064 | 9 | **525.27** | 58.36 |
| METHADONE | 9250B | 23 | **292,388.30** | 12,712.53 |

**STATE: KS - KANSAS**

**BUSINESS ACTIVITY: A - PHARMACIES**

| DRUG NAME | DRUG CODE | NUMBER OF REGISTRANT SOLD TO | TOTAL GRAMS | AVG GRAMS |
|---|---|---|---|---|
| AMPHETAMINE | 1100 | 606 | **275,081.40** | 453.93 |
| METHAMPHETAMINE | 1105 | 10 | **42.18** | 4.22 |
| LISDEXAMFETAMINE | 1205 | 595 | **126,680.79** | 212.91 |
| METHYLPHENIDATE (DL;D;L;ISOMERS) | 1724 | 601 | **185,070.81** | 307.94 |
| CODEINE | 9050 | 596 | **82,323.77** | 138.13 |
| BUPRENORPHINE | 9064 | 542 | **13,510.23** | 24.93 |
| OXYCODONE | 9143 | 608 | **320,986.34** | 527.94 |
| HYDROMORPHONE | 9150 | 544 | **7,702.68** | 14.16 |
| HYDROCODONE | 9193 | 612 | **217,558.26** | 355.49 |
| MEPERIDINE (PETHIDINE) | 9230 | 68 | **619.53** | 9.11 |
| METHADONE | 9250B | 473 | **16,260.73** | 34.38 |
| MORPHINE | 9300 | 591 | **111,239.08** | 188.22 |
| OXYMORPHONE | 9652 | 141 | **1,739.06** | 12.33 |
| TAPENTADOL | 9780 | 236 | **17,372.50** | 73.61 |
| FENTANYL BASE | 9801 | 578 | **2,030.87** | 3.51 |

**BUSINESS ACTIVITY: B - HOSPITALS**

| DRUG NAME | DRUG CODE | NUMBER OF REGISTRANT SOLD TO | TOTAL GRAMS | AVG GRAMS |
|---|---|---|---|---|
| AMPHETAMINE | 1100 | 55 | **8,592.96** | 156.24 |
| LISDEXAMFETAMINE | 1205 | 24 | **1,516.93** | 63.21 |
| METHYLPHENIDATE (DL;D;L;ISOMERS) | 1724 | 86 | **7,197.99** | 83.70 |
| CODEINE | 9050 | 136 | **2,723.58** | 20.03 |
| BUPRENORPHINE | 9064 | 46 | **674.42** | 14.66 |
| OXYCODONE | 9143 | 194 | **20,890.44** | 107.68 |
| HYDROMORPHONE | 9150 | 174 | **1,655.65** | 9.52 |
| HYDROCODONE | 9193 | 203 | **10,838.02** | 53.39 |
| MEPERIDINE (PETHIDINE) | 9230 | 103 | **645.78** | 6.27 |
| METHADONE | 9250B | 82 | **924.57** | 11.28 |
| MORPHINE | 9300 | 204 | **9,170.67** | 44.95 |
| OXYMORPHONE | 9652 | 10 | **91.89** | 9.19 |
| TAPENTADOL | 9780 | 20 | **572.50** | 28.63 |
| FENTANYL BASE | 9801 | 208 | **307.83** | 1.48 |

**BUSINESS ACTIVITY: C - PRACTITIONERS**

| DRUG NAME | DRUG CODE | NUMBER OF REGISTRANT SOLD TO | TOTAL GRAMS | AVG GRAMS |
|---|---|---|---|---|

| | | | | |
|---|---|---|---|---|
| CODEINE | 9050 | 28 | **588.25** | 21.01 |
| BUPRENORPHINE | 9064 | 290 | **34.75** | 0.12 |
| OXYCODONE | 9143 | 6 | **13.94** | 2.32 |
| HYDROMORPHONE | 9150 | 104 | **748.41** | 7.20 |
| HYDROCODONE | 9193 | 101 | **189.17** | 1.87 |
| MEPERIDINE (PETHIDINE) | 9230 | 14 | **62.75** | 4.48 |
| MORPHINE | 9300 | 35 | **47.59** | 1.36 |
| FENTANYL BASE | 9801 | 111 | **10.34** | 0.09 |

**BUSINESS ACTIVITY: M - MID-LEVEL PRACTITIONERS**

| DRUG NAME | DRUG CODE | NUMBER OF REGISTRANT SOLD TO | TOTAL GRAMS | AVG GRAMS |
|---|---|---|---|---|
| BUPRENORPHINE | 9064 | 1 | **5.94** | 5.94 |
| HYDROMORPHONE | 9150 | 15 | **0.78** | 0.05 |
| MORPHINE | 9300 | 29 | **7.16** | 0.25 |
| FENTANYL BASE | 9801 | 35 | **0.50** | 0.01 |

**BUSINESS ACTIVITY: NTP - NARCOTIC TREATMENT PROGRAMS**

| DRUG NAME | DRUG CODE | NUMBER OF REGISTRANT SOLD TO | TOTAL GRAMS | AVG GRAMS |
|---|---|---|---|---|
| BUPRENORPHINE | 9064 | 6 | **372.54** | 62.09 |
| METHADONE | 9250B | 9 | **61,373.73** | 6,819.30 |

**STATE: KY - KENTUCKY**

**BUSINESS ACTIVITY: A - PHARMACIES**

| DRUG NAME | DRUG CODE | NUMBER OF REGISTRANT SOLD TO | TOTAL GRAMS | AVG GRAMS |
|---|---|---|---|---|
| AMPHETAMINE | 1100 | 1,076 | **309,756.85** | 287.88 |
| METHAMPHETAMINE | 1105 | 17 | **39.37** | 2.32 |
| LISDEXAMFETAMINE | 1205 | 1,019 | **147,776.23** | 145.02 |
| METHYLPHENIDATE (DL;D;L;ISOMERS) | 1724 | 1,075 | **271,409.80** | 252.47 |
| CODEINE | 9050 | 1,062 | **132,625.03** | 124.88 |
| BUPRENORPHINE | 9064 | 1,035 | **202,857.18** | 196.00 |
| OXYCODONE | 9143 | 1,085 | **604,951.83** | 557.56 |
| HYDROMORPHONE | 9150 | 753 | **6,167.95** | 8.19 |
| HYDROCODONE | 9193 | 1,094 | **618,006.01** | 564.90 |
| MEPERIDINE (PETHIDINE) | 9230 | 64 | **521.16** | 8.14 |
| METHADONE | 9250B | 845 | **27,916.37** | 33.04 |
| MORPHINE | 9300 | 1,028 | **108,473.31** | 105.52 |
| OXYMORPHONE | 9652 | 307 | **4,503.54** | 14.67 |
| TAPENTADOL | 9780 | 378 | **30,272.50** | 80.09 |
| FENTANYL BASE | 9801 | 954 | **1,944.79** | 2.04 |

**BUSINESS ACTIVITY: B - HOSPITALS**

| DRUG NAME | DRUG CODE | NUMBER OF REGISTRANT SOLD TO | TOTAL GRAMS | AVG GRAMS |
|---|---|---|---|---|
| AMPHETAMINE | 1100 | 54 | **7,035.11** | 130.28 |
| LISDEXAMFETAMINE | 1205 | 27 | **1,429.03** | 52.93 |
| METHYLPHENIDATE (DL;D;L;ISOMERS) | 1724 | 83 | **6,363.59** | 76.67 |
| CODEINE | 9050 | 118 | **2,366.85** | 20.06 |
| BUPRENORPHINE | 9064 | 105 | **2,561.14** | 24.39 |
| OXYCODONE | 9143 | 144 | **23,821.27** | 165.43 |
| HYDROMORPHONE | 9150 | 132 | **2,396.92** | 18.16 |
| HYDROCODONE | 9193 | 147 | **19,617.86** | 133.45 |
| MEPERIDINE (PETHIDINE) | 9230 | 90 | **1,423.92** | 15.82 |
| METHADONE | 9250B | 101 | **2,334.68** | 23.12 |
| MORPHINE | 9300 | 141 | **12,233.35** | 86.76 |
| OXYMORPHONE | 9652 | 9 | **12.76** | 1.42 |
| TAPENTADOL | 9780 | 15 | **872.50** | 58.17 |
| FENTANYL BASE | 9801 | 142 | **483.90** | 3.41 |

**BUSINESS ACTIVITY: C - PRACTITIONERS**

| DRUG NAME | DRUG CODE | NUMBER OF REGISTRANT SOLD TO | TOTAL GRAMS | AVG GRAMS |
|---|---|---|---|---|
| CODEINE | 9050 | 12 | **432.15** | 36.01 |
| BUPRENORPHINE | 9064 | 304 | **154.82** | 0.51 |
| OXYCODONE | 9143 | 16 | **24.38** | 1.52 |
| HYDROMORPHONE | 9150 | 101 | **586.40** | 5.81 |
| HYDROCODONE | 9193 | 134 | **557.94** | 4.16 |
| MEPERIDINE (PETHIDINE) | 9230 | 25 | **221.47** | 8.86 |
| METHADONE | 9250B | 3 | **3.40** | 1.13 |

| | | | | |
|---|---|---|---|---|
| MORPHINE | 9300 | 107 | **365.99** | 3.42 |
| FENTANYL BASE | 9801 | 274 | **22.67** | 0.08 |

**BUSINESS ACTIVITY: D - TEACHING INSTITUTIONS**

| DRUG NAME | DRUG CODE | NUMBER OF REGISTRANT SOLD TO | TOTAL GRAMS | AVG GRAMS |
|---|---|---|---|---|
| HYDROMORPHONE | 9150 | 1 | **2.13** | 2.13 |

**BUSINESS ACTIVITY: M - MID-LEVEL PRACTITIONERS**

| DRUG NAME | DRUG CODE | NUMBER OF REGISTRANT SOLD TO | TOTAL GRAMS | AVG GRAMS |
|---|---|---|---|---|
| CODEINE | 9050 | 1 | **0.59** | 0.59 |

**BUSINESS ACTIVITY: NTP - NARCOTIC TREATMENT PROGRAMS**

| DRUG NAME | DRUG CODE | NUMBER OF REGISTRANT SOLD TO | TOTAL GRAMS | AVG GRAMS |
|---|---|---|---|---|
| BUPRENORPHINE | 9064 | 19 | **381.90** | 20.10 |
| METHADONE | 9250B | 28 | **208,428.74** | 7,443.88 |

**STATE: LA - LOUISIANA**

**BUSINESS ACTIVITY: A - PHARMACIES**

| DRUG NAME | DRUG CODE | NUMBER OF REGISTRANT SOLD TO | TOTAL GRAMS | AVG GRAMS |
|---|---|---|---|---|
| AMPHETAMINE | 1100 | 1,127 | **715,278.92** | 634.68 |
| METHAMPHETAMINE | 1105 | 24 | **54.24** | 2.26 |
| LISDEXAMFETAMINE | 1205 | 1,093 | **338,024.90** | 309.26 |
| METHYLPHENIDATE (DL;D;L;ISOMERS) | 1724 | 1,110 | **277,806.73** | 250.28 |
| CODEINE | 9050 | 1,101 | **118,121.14** | 107.29 |
| BUPRENORPHINE | 9064 | 991 | **108,878.02** | 109.87 |
| OXYCODONE | 9143 | 1,120 | **522,983.71** | 466.95 |
| HYDROMORPHONE | 9150 | 833 | **6,158.26** | 7.39 |
| HYDROCODONE | 9193 | 1,134 | **426,803.69** | 376.37 |
| MEPERIDINE (PETHIDINE) | 9230 | 204 | **2,073.73** | 10.17 |
| METHADONE | 9250B | 687 | **19,252.23** | 28.02 |
| MORPHINE | 9300 | 1,024 | **135,436.76** | 132.26 |
| OXYMORPHONE | 9652 | 270 | **7,356.75** | 27.25 |
| TAPENTADOL | 9780 | 496 | **40,967.50** | 82.60 |
| FENTANYL BASE | 9801 | 930 | **2,082.79** | 2.24 |

**BUSINESS ACTIVITY: B - HOSPITALS**

| DRUG NAME | DRUG CODE | NUMBER OF REGISTRANT SOLD TO | TOTAL GRAMS | AVG GRAMS |
|---|---|---|---|---|
| AMPHETAMINE | 1100 | 57 | **8,277.05** | 145.21 |
| LISDEXAMFETAMINE | 1205 | 31 | **2,119.89** | 68.38 |
| METHYLPHENIDATE (DL;D;L;ISOMERS) | 1724 | 103 | **3,341.40** | 32.44 |
| CODEINE | 9050 | 123 | **1,859.12** | 15.11 |
| BUPRENORPHINE | 9064 | 105 | **1,425.94** | 13.58 |
| OXYCODONE | 9143 | 200 | **21,472.05** | 107.36 |
| HYDROMORPHONE | 9150 | 187 | **2,062.05** | 11.03 |
| HYDROCODONE | 9193 | 209 | **23,087.27** | 110.47 |
| MEPERIDINE (PETHIDINE) | 9230 | 136 | **2,604.59** | 19.15 |
| METHADONE | 9250B | 124 | **1,577.27** | 12.72 |
| MORPHINE | 9300 | 200 | **10,233.81** | 51.17 |
| OXYMORPHONE | 9652 | 6 | **8.39** | 1.40 |
| TAPENTADOL | 9780 | 13 | **647.00** | 49.77 |
| FENTANYL BASE | 9801 | 194 | **411.23** | 2.12 |

**BUSINESS ACTIVITY: C - PRACTITIONERS**

| DRUG NAME | DRUG CODE | NUMBER OF REGISTRANT SOLD TO | TOTAL GRAMS | AVG GRAMS |
|---|---|---|---|---|
| CODEINE | 9050 | 28 | **452.93** | 16.18 |
| BUPRENORPHINE | 9064 | 349 | **18.45** | 0.05 |
| OXYCODONE | 9143 | 19 | **25.01** | 1.32 |
| HYDROMORPHONE | 9150 | 122 | **493.57** | 4.05 |
| HYDROCODONE | 9193 | 181 | **484.00** | 2.67 |
| MEPERIDINE (PETHIDINE) | 9230 | 85 | **543.05** | 6.39 |
| METHADONE | 9250B | 4 | **6.26** | 1.57 |
| MORPHINE | 9300 | 105 | **54.35** | 0.52 |
| FENTANYL BASE | 9801 | 253 | **21.49** | 0.08 |

**BUSINESS ACTIVITY: D - TEACHING INSTITUTIONS**

| DRUG NAME | DRUG CODE | NUMBER OF REGISTRANT SOLD TO | TOTAL GRAMS | AVG GRAMS |
|---|---|---|---|---|
| FENTANYL BASE | 9801 | 1 | **0.02** | 0.02 |

**BUSINESS ACTIVITY: M - MID-LEVEL PRACTITIONERS**

| DRUG NAME | DRUG CODE | NUMBER OF REGISTRANT SOLD TO | TOTAL GRAMS | AVG GRAMS |
|---|---|---|---|---|
| CODEINE | 9050 | 7 | **4.41** | 0.63 |

**BUSINESS ACTIVITY: NTP - NARCOTIC TREATMENT PROGRAMS**

| DRUG NAME | DRUG CODE | NUMBER OF REGISTRANT SOLD TO | TOTAL GRAMS | AVG GRAMS |
|---|---|---|---|---|
| BUPRENORPHINE | 9064 | 9 | **862.79** | 95.87 |
| METHADONE | 9250B | 10 | **125,675.27** | 12,567.53 |

**STATE: MA - MASSACHUSETTS**

**BUSINESS ACTIVITY: A - PHARMACIES**

| DRUG NAME | DRUG CODE | NUMBER OF REGISTRANT SOLD TO | TOTAL GRAMS | AVG GRAMS |
|---|---|---|---|---|
| AMPHETAMINE | 1100 | 1,090 | **748,022.25** | 686.26 |
| METHAMPHETAMINE | 1105 | 31 | **90.80** | 2.93 |
| LISDEXAMFETAMINE | 1205 | 1,059 | **160,793.31** | 151.84 |
| METHYLPHENIDATE (DL;D;L;ISOMERS) | 1724 | 1,087 | **511,486.16** | 470.55 |
| CODEINE | 9050 | 1,060 | **117,911.32** | 111.24 |
| BUPRENORPHINE | 9064 | 1,094 | **190,466.49** | 174.10 |
| OXYCODONE | 9143 | 1,098 | **627,152.90** | 571.18 |
| HYDROMORPHONE | 9150 | 1,030 | **15,737.74** | 15.28 |
| HYDROCODONE | 9193 | 1,080 | **79,080.04** | 73.22 |
| MEPERIDINE (PETHIDINE) | 9230 | 38 | **725.20** | 19.08 |
| METHADONE | 9250B | 913 | **45,685.50** | 50.04 |
| MORPHINE | 9300 | 1,022 | **173,760.02** | 170.02 |
| OXYMORPHONE | 9652 | 114 | **2,319.10** | 20.34 |
| TAPENTADOL | 9780 | 370 | **55,769.00** | 150.73 |
| FENTANYL BASE | 9801 | 924 | **2,503.52** | 2.71 |

**BUSINESS ACTIVITY: B - HOSPITALS**

| DRUG NAME | DRUG CODE | NUMBER OF REGISTRANT SOLD TO | TOTAL GRAMS | AVG GRAMS |
|---|---|---|---|---|
| AMPHETAMINE | 1100 | 153 | **22,544.71** | 147.35 |
| LISDEXAMFETAMINE | 1205 | 80 | **3,363.93** | 42.05 |
| METHYLPHENIDATE (DL;D;L;ISOMERS) | 1724 | 161 | **17,938.34** | 111.42 |
| CODEINE | 9050 | 138 | **5,497.47** | 39.84 |
| BUPRENORPHINE | 9064 | 163 | **11,542.46** | 70.81 |
| OXYCODONE | 9143 | 195 | **46,483.18** | 238.38 |
| HYDROMORPHONE | 9150 | 172 | **9,355.66** | 54.39 |
| HYDROCODONE | 9193 | 142 | **2,398.75** | 16.89 |
| MEPERIDINE (PETHIDINE) | 9230 | 60 | **442.18** | 7.37 |
| METHADONE | 9250B | 158 | **15,159.49** | 95.95 |
| MORPHINE | 9300 | 190 | **19,130.08** | 100.68 |
| OXYMORPHONE | 9652 | 15 | **290.87** | 19.39 |
| TAPENTADOL | 9780 | 16 | **1,872.50** | 117.03 |
| FENTANYL BASE | 9801 | 182 | **634.32** | 3.49 |

**BUSINESS ACTIVITY: C - PRACTITIONERS**

| DRUG NAME | DRUG CODE | NUMBER OF REGISTRANT SOLD TO | TOTAL GRAMS | AVG GRAMS |
|---|---|---|---|---|
| CODEINE | 9050 | 12 | **448.22** | 37.35 |
| BUPRENORPHINE | 9064 | 599 | **94.62** | 0.16 |
| OXYCODONE | 9143 | 22 | **30.53** | 1.39 |
| HYDROMORPHONE | 9150 | 236 | **1,347.13** | 5.71 |
| HYDROCODONE | 9193 | 132 | **387.20** | 2.93 |
| MEPERIDINE (PETHIDINE) | 9230 | 11 | **37.47** | 3.41 |
| METHADONE | 9250B | 32 | **104.82** | 3.28 |
| MORPHINE | 9300 | 81 | **45.47** | 0.56 |
| FENTANYL BASE | 9801 | 270 | **20.71** | 0.08 |

**BUSINESS ACTIVITY: M - MID-LEVEL PRACTITIONERS**

| DRUG NAME | DRUG CODE | NUMBER OF REGISTRANT SOLD TO | TOTAL GRAMS | AVG GRAMS |
|---|---|---|---|---|
| BUPRENORPHINE | 9064 | 1 | **4.92** | 4.92 |

**BUSINESS ACTIVITY: NTP - NARCOTIC TREATMENT PROGRAMS**

| DRUG NAME | DRUG CODE | NUMBER OF REGISTRANT SOLD TO | TOTAL GRAMS | AVG GRAMS |
|---|---|---|---|---|
| BUPRENORPHINE | 9064 | 44 | **1,889.05** | 42.93 |
| METHADONE | 9250B | 85 | **572,694.16** | 6,737.58 |

**STATE: MD - MARYLAND**

**BUSINESS ACTIVITY: A - PHARMACIES**

| DRUG NAME | DRUG CODE | NUMBER OF REGISTRANT SOLD TO | TOTAL GRAMS | AVG GRAMS |
|---|---|---|---|---|

| | | | | |
|---|---|---|---|---|
| AMPHETAMINE | 1100 | 1,161 | **409,338.25** | 352.57 |
| METHAMPHETAMINE | 1105 | 33 | **135.80** | 4.12 |
| LISDEXAMFETAMINE | 1205 | 1,122 | **226,784.58** | 202.13 |
| METHYLPHENIDATE (DL;D;L;ISOMERS) | 1724 | 1,152 | **361,801.81** | 314.06 |
| CODEINE | 9050 | 1,155 | **153,531.85** | 132.93 |
| BUPRENORPHINE | 9064 | 1,143 | **135,840.93** | 118.85 |
| OXYCODONE | 9143 | 1,184 | **862,552.50** | 728.51 |
| HYDROMORPHONE | 9150 | 1,069 | **16,707.64** | 15.63 |
| HYDROCODONE | 9193 | 1,150 | **89,805.26** | 78.09 |
| MEPERIDINE (PETHIDINE) | 9230 | 76 | **941.22** | 12.38 |
| METHADONE | 9250B | 843 | **26,227.30** | 31.11 |
| MORPHINE | 9300 | 1,109 | **172,480.42** | 155.53 |
| OXYMORPHONE | 9652 | 558 | **9,514.47** | 17.05 |
| TAPENTADOL | 9780 | 712 | **82,562.00** | 115.96 |
| FENTANYL BASE | 9801 | 968 | **2,122.06** | 2.19 |

**BUSINESS ACTIVITY: B - HOSPITALS**

| DRUG NAME | DRUG CODE | NUMBER OF REGISTRANT SOLD TO | TOTAL GRAMS | AVG GRAMS |
|---|---|---|---|---|
| AMPHETAMINE | 1100 | 58 | **9,382.81** | 161.77 |
| LISDEXAMFETAMINE | 1205 | 30 | **2,306.62** | 76.89 |
| METHYLPHENIDATE (DL;D;L;ISOMERS) | 1724 | 67 | **13,990.88** | 208.82 |
| CODEINE | 9050 | 80 | **3,251.79** | 40.65 |
| BUPRENORPHINE | 9064 | 87 | **2,183.41** | 25.10 |
| OXYCODONE | 9143 | 131 | **38,319.18** | 292.51 |
| HYDROMORPHONE | 9150 | 129 | **4,600.43** | 35.66 |
| HYDROCODONE | 9193 | 100 | **1,984.83** | 19.85 |
| MEPERIDINE (PETHIDINE) | 9230 | 64 | **507.87** | 7.94 |
| METHADONE | 9250B | 70 | **6,950.28** | 99.29 |
| MORPHINE | 9300 | 125 | **12,459.87** | 99.68 |
| OXYMORPHONE | 9652 | 6 | **121.01** | 20.17 |
| TAPENTADOL | 9780 | 17 | **3,150.00** | 185.29 |
| FENTANYL BASE | 9801 | 214 | **556.36** | 2.60 |

**BUSINESS ACTIVITY: C - PRACTITIONERS**

| DRUG NAME | DRUG CODE | NUMBER OF REGISTRANT SOLD TO | TOTAL GRAMS | AVG GRAMS |
|---|---|---|---|---|
| CODEINE | 9050 | 84 | **6,530.28** | 77.74 |
| BUPRENORPHINE | 9064 | 568 | **132.48** | 0.23 |
| OXYCODONE | 9143 | 46 | **2,035.19** | 44.24 |
| HYDROMORPHONE | 9150 | 260 | **1,398.38** | 5.38 |
| HYDROCODONE | 9193 | 213 | **1,049.45** | 4.93 |
| MEPERIDINE (PETHIDINE) | 9230 | 46 | **244.56** | 5.32 |
| METHADONE | 9250B | 24 | **75.93** | 3.16 |
| MORPHINE | 9300 | 103 | **524.88** | 5.10 |
| OXYMORPHONE | 9652 | 4 | **40.55** | 10.14 |
| TAPENTADOL | 9780 | 5 | **102.00** | 20.40 |
| FENTANYL BASE | 9801 | 335 | **37.79** | 0.11 |

**BUSINESS ACTIVITY: D - TEACHING INSTITUTIONS**

| DRUG NAME | DRUG CODE | NUMBER OF REGISTRANT SOLD TO | TOTAL GRAMS | AVG GRAMS |
|---|---|---|---|---|
| MEPERIDINE (PETHIDINE) | 9230 | 1 | **4.36** | 4.36 |
| MORPHINE | 9300 | 1 | **0.03** | 0.03 |
| FENTANYL BASE | 9801 | 1 | **0.01** | 0.01 |

**BUSINESS ACTIVITY: M - MID-LEVEL PRACTITIONERS**

| DRUG NAME | DRUG CODE | NUMBER OF REGISTRANT SOLD TO | TOTAL GRAMS | AVG GRAMS |
|---|---|---|---|---|
| BUPRENORPHINE | 9064 | 2 | **1.92** | 0.96 |

**BUSINESS ACTIVITY: NTP - NARCOTIC TREATMENT PROGRAMS**

| DRUG NAME | DRUG CODE | NUMBER OF REGISTRANT SOLD TO | TOTAL GRAMS | AVG GRAMS |
|---|---|---|---|---|
| BUPRENORPHINE | 9064 | 67 | **8,061.24** | 120.32 |
| METHADONE | 9250B | 89 | **741,105.21** | 8,327.02 |

**STATE: ME - MAINE**

**BUSINESS ACTIVITY: A - PHARMACIES**

| DRUG NAME | DRUG CODE | NUMBER OF REGISTRANT SOLD TO | TOTAL GRAMS | AVG GRAMS |
|---|---|---|---|---|
| AMPHETAMINE | 1100 | 284 | **108,672.43** | 382.65 |
| METHAMPHETAMINE | 1105 | 7 | **36.16** | 5.17 |

| | | | | |
|---|---|---|---|---|
| LISDEXAMFETAMINE | 1205 | 285 | **70,636.88** | 247.85 |
| METHYLPHENIDATE (DL;D;L;ISOMERS) | 1724 | 289 | **137,732.14** | 476.58 |
| CODEINE | 9050 | 286 | **26,222.62** | 91.69 |
| BUPRENORPHINE | 9064 | 288 | **59,620.66** | 207.02 |
| OXYCODONE | 9143 | 288 | **138,713.69** | 481.64 |
| HYDROMORPHONE | 9150 | 281 | **5,131.40** | 18.26 |
| HYDROCODONE | 9193 | 291 | **47,138.42** | 161.99 |
| MEPERIDINE (PETHIDINE) | 9230 | 17 | **156.98** | 9.23 |
| METHADONE | 9250B | 265 | **11,634.62** | 43.90 |
| MORPHINE | 9300 | 290 | **43,976.72** | 151.64 |
| OXYMORPHONE | 9652 | 71 | **934.12** | 13.16 |
| TAPENTADOL | 9780 | 78 | **4,028.50** | 51.65 |
| FENTANYL BASE | 9801 | 282 | **734.01** | 2.60 |

**BUSINESS ACTIVITY: B - HOSPITALS**

| DRUG NAME | DRUG CODE | NUMBER OF REGISTRANT SOLD TO | TOTAL GRAMS | AVG GRAMS |
|---|---|---|---|---|
| AMPHETAMINE | 1100 | 26 | **1,916.79** | 73.72 |
| LISDEXAMFETAMINE | 1205 | 14 | **463.64** | 33.12 |
| METHYLPHENIDATE (DL;D;L;ISOMERS) | 1724 | 40 | **2,437.72** | 60.94 |
| CODEINE | 9050 | 34 | **431.34** | 12.69 |
| BUPRENORPHINE | 9064 | 45 | **2,111.01** | 46.91 |
| OXYCODONE | 9143 | 48 | **7,342.26** | 152.96 |
| HYDROMORPHONE | 9150 | 44 | **993.90** | 22.59 |
| HYDROCODONE | 9193 | 46 | **2,485.89** | 54.04 |
| MEPERIDINE (PETHIDINE) | 9230 | 23 | **50.76** | 2.21 |
| METHADONE | 9250B | 38 | **1,223.99** | 32.21 |
| MORPHINE | 9300 | 45 | **3,610.28** | 80.23 |
| OXYMORPHONE | 9652 | 1 | **1.61** | 1.61 |
| TAPENTADOL | 9780 | 1 | **21.00** | 21.00 |
| FENTANYL BASE | 9801 | 46 | **121.75** | 2.65 |

**BUSINESS ACTIVITY: C - PRACTITIONERS**

| DRUG NAME | DRUG CODE | NUMBER OF REGISTRANT SOLD TO | TOTAL GRAMS | AVG GRAMS |
|---|---|---|---|---|
| CODEINE | 9050 | 8 | **111.92** | 13.99 |
| BUPRENORPHINE | 9064 | 170 | **127.00** | 0.75 |
| OXYCODONE | 9143 | 5 | **17.26** | 3.45 |
| HYDROMORPHONE | 9150 | 76 | **397.77** | 5.23 |
| HYDROCODONE | 9193 | 44 | **77.57** | 1.76 |
| MEPERIDINE (PETHIDINE) | 9230 | 4 | **32.68** | 8.17 |
| METHADONE | 9250B | 2 | **8.41** | 4.20 |
| MORPHINE | 9300 | 27 | **23.28** | 0.86 |
| FENTANYL BASE | 9801 | 39 | **3.12** | 0.08 |

**BUSINESS ACTIVITY: D - TEACHING INSTITUTIONS**

| DRUG NAME | DRUG CODE | NUMBER OF REGISTRANT SOLD TO | TOTAL GRAMS | AVG GRAMS |
|---|---|---|---|---|
| BUPRENORPHINE | 9064 | 1 | **0.01** | 0.01 |

**BUSINESS ACTIVITY: M - MID-LEVEL PRACTITIONERS**

| DRUG NAME | DRUG CODE | NUMBER OF REGISTRANT SOLD TO | TOTAL GRAMS | AVG GRAMS |
|---|---|---|---|---|
| CODEINE | 9050 | 5 | **131.06** | 26.21 |
| BUPRENORPHINE | 9064 | 6 | **148.05** | 24.67 |

**BUSINESS ACTIVITY: NTP - NARCOTIC TREATMENT PROGRAMS**

| DRUG NAME | DRUG CODE | NUMBER OF REGISTRANT SOLD TO | TOTAL GRAMS | AVG GRAMS |
|---|---|---|---|---|
| BUPRENORPHINE | 9064 | 2 | **214.56** | 107.28 |
| METHADONE | 9250B | 10 | **106,686.72** | 10,668.67 |

**STATE: MI - MICHIGAN**

**BUSINESS ACTIVITY: A - PHARMACIES**

| DRUG NAME | DRUG CODE | NUMBER OF REGISTRANT SOLD TO | TOTAL GRAMS | AVG GRAMS |
|---|---|---|---|---|
| AMPHETAMINE | 1100 | 2,286 | **1,011,136.31** | 442.32 |
| METHAMPHETAMINE | 1105 | 24 | **59.86** | 2.49 |
| LISDEXAMFETAMINE | 1205 | 2,049 | **243,909.06** | 119.04 |
| METHYLPHENIDATE (DL;D;L;ISOMERS) | 1724 | 2,181 | **625,804.03** | 286.93 |
| CODEINE | 9050 | 2,320 | **618,004.82** | 266.38 |
| BUPRENORPHINE | 9064 | 2,166 | **149,451.46** | 69.00 |
| OXYCODONE | 9143 | 2,315 | **692,254.39** | 299.03 |

| | | | | |
|---|---|---|---|---|
| HYDROMORPHONE | 9150 | 1,691 | **16,619.61** | 9.83 |
| HYDROCODONE | 9193 | 2,356 | **911,733.04** | 386.98 |
| MEPERIDINE (PETHIDINE) | 9230 | 43 | **661.03** | 15.37 |
| METHADONE | 9250B | 1,679 | **69,104.47** | 41.16 |
| MORPHINE | 9300 | 2,136 | **424,986.62** | 198.96 |
| OXYMORPHONE | 9652 | 513 | **17,763.23** | 34.63 |
| TAPENTADOL | 9780 | 597 | **46,523.50** | 77.93 |
| FENTANYL BASE | 9801 | 1,882 | **5,145.36** | 2.73 |

**BUSINESS ACTIVITY: B - HOSPITALS**

| DRUG NAME | DRUG CODE | NUMBER OF REGISTRANT SOLD TO | TOTAL GRAMS | AVG GRAMS |
|---|---|---|---|---|
| AMPHETAMINE | 1100 | 114 | **18,242.63** | 160.02 |
| LISDEXAMFETAMINE | 1205 | 58 | **3,200.36** | 55.18 |
| METHYLPHENIDATE (DL;D;L;ISOMERS) | 1724 | 149 | **13,668.06** | 91.73 |
| CODEINE | 9050 | 177 | **10,930.81** | 61.76 |
| BUPRENORPHINE | 9064 | 142 | **3,570.52** | 25.14 |
| OXYCODONE | 9143 | 191 | **38,798.24** | 203.13 |
| HYDROMORPHONE | 9150 | 187 | **4,844.08** | 25.90 |
| HYDROCODONE | 9193 | 200 | **35,649.40** | 178.25 |
| MEPERIDINE (PETHIDINE) | 9230 | 92 | **927.93** | 10.09 |
| METHADONE | 9250B | 157 | **6,324.72** | 40.28 |
| MORPHINE | 9300 | 193 | **31,081.05** | 161.04 |
| OXYMORPHONE | 9652 | 13 | **186.54** | 14.35 |
| TAPENTADOL | 9780 | 15 | **835.50** | 55.70 |
| FENTANYL BASE | 9801 | 187 | **1,076.75** | 5.76 |

**BUSINESS ACTIVITY: C - PRACTITIONERS**

| DRUG NAME | DRUG CODE | NUMBER OF REGISTRANT SOLD TO | TOTAL GRAMS | AVG GRAMS |
|---|---|---|---|---|
| AMPHETAMINE | 1100 | 1 | **1.88** | 1.88 |
| METHYLPHENIDATE (DL;D;L;ISOMERS) | 1724 | 1 | **3.46** | 3.46 |
| CODEINE | 9050 | 87 | **1,470.52** | 16.90 |
| BUPRENORPHINE | 9064 | 800 | **449.99** | 0.56 |
| OXYCODONE | 9143 | 40 | **485.63** | 12.14 |
| HYDROMORPHONE | 9150 | 402 | **2,662.29** | 6.62 |
| HYDROCODONE | 9193 | 229 | **681.01** | 2.97 |
| MEPERIDINE (PETHIDINE) | 9230 | 71 | **309.06** | 4.35 |
| METHADONE | 9250B | 12 | **61.71** | 5.14 |
| MORPHINE | 9300 | 175 | **280.54** | 1.60 |
| FENTANYL BASE | 9801 | 498 | **62.50** | 0.13 |

**BUSINESS ACTIVITY: D - TEACHING INSTITUTIONS**

| DRUG NAME | DRUG CODE | NUMBER OF REGISTRANT SOLD TO | TOTAL GRAMS | AVG GRAMS |
|---|---|---|---|---|
| CODEINE | 9050 | 1 | **197.36** | 197.36 |
| BUPRENORPHINE | 9064 | 1 | **1.24** | 1.24 |
| HYDROMORPHONE | 9150 | 1 | **6.21** | 6.21 |
| HYDROCODONE | 9193 | 1 | **27.44** | 27.44 |
| METHADONE | 9250B | 1 | **47.40** | 47.40 |
| MORPHINE | 9300 | 1 | **5.72** | 5.72 |
| FENTANYL BASE | 9801 | 2 | **2.87** | 1.44 |

**BUSINESS ACTIVITY: M - MID-LEVEL PRACTITIONERS**

| DRUG NAME | DRUG CODE | NUMBER OF REGISTRANT SOLD TO | TOTAL GRAMS | AVG GRAMS |
|---|---|---|---|---|
| CODEINE | 9050 | 1 | **19.89** | 19.89 |
| BUPRENORPHINE | 9064 | 4 | **4.99** | 1.25 |

**BUSINESS ACTIVITY: NTP - NARCOTIC TREATMENT PROGRAMS**

| DRUG NAME | DRUG CODE | NUMBER OF REGISTRANT SOLD TO | TOTAL GRAMS | AVG GRAMS |
|---|---|---|---|---|
| BUPRENORPHINE | 9064 | 11 | **212.59** | 19.33 |
| METHADONE | 9250B | 45 | **405,410.95** | 9,009.13 |

**STATE: MN - MINNESOTA**

**BUSINESS ACTIVITY: A - PHARMACIES**

| DRUG NAME | DRUG CODE | NUMBER OF REGISTRANT SOLD TO | TOTAL GRAMS | AVG GRAMS |
|---|---|---|---|---|
| AMPHETAMINE | 1100 | 952 | **475,920.60** | 499.92 |
| METHAMPHETAMINE | 1105 | 39 | **173.55** | 4.45 |
| LISDEXAMFETAMINE | 1205 | 934 | **136,844.94** | 146.51 |
| METHYLPHENIDATE (DL;D;L;ISOMERS) | 1724 | 954 | **379,518.40** | 397.82 |

| | | | | |
|---|---|---|---|---|
| CODEINE | 9050 | 934 | **87,661.52** | 93.86 |
| BUPRENORPHINE | 9064 | 907 | **45,337.00** | 49.99 |
| OXYCODONE | 9143 | 960 | **335,871.05** | 349.87 |
| HYDROMORPHONE | 9150 | 895 | **12,872.64** | 14.38 |
| HYDROCODONE | 9193 | 960 | **105,608.19** | 110.01 |
| MEPERIDINE (PETHIDINE) | 9230 | 26 | **140.97** | 5.42 |
| METHADONE | 9250B | 773 | **18,365.88** | 23.76 |
| MORPHINE | 9300 | 934 | **106,506.88** | 114.03 |
| OXYMORPHONE | 9652 | 79 | **608.77** | 7.71 |
| TAPENTADOL | 9780 | 216 | **11,349.50** | 52.54 |
| FENTANYL BASE | 9801 | 833 | **1,857.88** | 2.23 |

**BUSINESS ACTIVITY: B - HOSPITALS**

| DRUG NAME | DRUG CODE | NUMBER OF REGISTRANT SOLD TO | TOTAL GRAMS | AVG GRAMS |
|---|---|---|---|---|
| AMPHETAMINE | 1100 | 78 | **8,004.34** | 102.62 |
| METHAMPHETAMINE | 1105 | 3 | **22.50** | 7.50 |
| LISDEXAMFETAMINE | 1205 | 47 | **1,491.50** | 31.73 |
| METHYLPHENIDATE (DL;D;L;ISOMERS) | 1724 | 97 | **7,576.41** | 78.11 |
| CODEINE | 9050 | 111 | **3,082.45** | 27.77 |
| BUPRENORPHINE | 9064 | 92 | **2,583.60** | 28.08 |
| OXYCODONE | 9143 | 148 | **24,862.56** | 167.99 |
| HYDROMORPHONE | 9150 | 152 | **4,129.76** | 27.17 |
| HYDROCODONE | 9193 | 147 | **7,013.60** | 47.71 |
| MEPERIDINE (PETHIDINE) | 9230 | 60 | **265.48** | 4.42 |
| METHADONE | 9250B | 102 | **2,712.68** | 26.59 |
| MORPHINE | 9300 | 152 | **11,164.21** | 73.45 |
| OXYMORPHONE | 9652 | 2 | **9.46** | 4.73 |
| TAPENTADOL | 9780 | 8 | **68.50** | 8.56 |
| FENTANYL BASE | 9801 | 149 | **393.12** | 2.64 |

**BUSINESS ACTIVITY: C - PRACTITIONERS**

| DRUG NAME | DRUG CODE | NUMBER OF REGISTRANT SOLD TO | TOTAL GRAMS | AVG GRAMS |
|---|---|---|---|---|
| AMPHETAMINE | 1100 | 1 | **2.38** | 2.38 |
| CODEINE | 9050 | 36 | **938.67** | 26.07 |
| BUPRENORPHINE | 9064 | 603 | **783.70** | 1.30 |
| OXYCODONE | 9143 | 62 | **198.51** | 3.20 |
| HYDROMORPHONE | 9150 | 353 | **2,063.02** | 5.84 |
| HYDROCODONE | 9193 | 234 | **447.04** | 1.91 |
| MEPERIDINE (PETHIDINE) | 9230 | 31 | **105.45** | 3.40 |
| METHADONE | 9250B | 3 | **2.15** | 0.72 |
| MORPHINE | 9300 | 133 | **93.30** | 0.70 |
| FENTANYL BASE | 9801 | 348 | **32.73** | 0.09 |

**BUSINESS ACTIVITY: D - TEACHING INSTITUTIONS**

| DRUG NAME | DRUG CODE | NUMBER OF REGISTRANT SOLD TO | TOTAL GRAMS | AVG GRAMS |
|---|---|---|---|---|
| BUPRENORPHINE | 9064 | 1 | **0.01** | 0.01 |
| HYDROMORPHONE | 9150 | 1 | **0.89** | 0.89 |

**BUSINESS ACTIVITY: M - MID-LEVEL PRACTITIONERS**

| DRUG NAME | DRUG CODE | NUMBER OF REGISTRANT SOLD TO | TOTAL GRAMS | AVG GRAMS |
|---|---|---|---|---|
| CODEINE | 9050 | 1 | **1.99** | 1.99 |
| BUPRENORPHINE | 9064 | 4 | **12.06** | 3.02 |
| FENTANYL BASE | 9801 | 2 | **0.01** | 0.01 |

**BUSINESS ACTIVITY: NTP - NARCOTIC TREATMENT PROGRAMS**

| DRUG NAME | DRUG CODE | NUMBER OF REGISTRANT SOLD TO | TOTAL GRAMS | AVG GRAMS |
|---|---|---|---|---|
| BUPRENORPHINE | 9064 | 16 | **1,902.03** | 118.88 |
| METHADONE | 9250B | 16 | **185,546.41** | 11,596.65 |

**STATE: MO - MISSOURI**

**BUSINESS ACTIVITY: A - PHARMACIES**

| DRUG NAME | DRUG CODE | NUMBER OF REGISTRANT SOLD TO | TOTAL GRAMS | AVG GRAMS |
|---|---|---|---|---|
| AMPHETAMINE | 1100 | 1,312 | **627,175.68** | 478.03 |
| METHAMPHETAMINE | 1105 | 14 | **183.59** | 13.11 |
| LISDEXAMFETAMINE | 1205 | 1,276 | **261,815.71** | 205.18 |
| METHYLPHENIDATE (DL;D;L;ISOMERS) | 1724 | 1,313 | **516,227.65** | 393.17 |
| CODEINE | 9050 | 1,295 | **213,293.95** | 164.71 |

| | | | | |
|---|---|---|---|---|
| BUPRENORPHINE | 9064 | 1,271 | **77,600.67** | 61.05 |
| OXYCODONE | 9143 | 1,322 | **661,279.50** | 500.21 |
| HYDROMORPHONE | 9150 | 1,068 | **10,722.84** | 10.04 |
| HYDROCODONE | 9193 | 1,322 | **458,877.39** | 347.11 |
| MEPERIDINE (PETHIDINE) | 9230 | 66 | **875.31** | 13.26 |
| METHADONE | 9250B | 1,005 | **25,794.59** | 25.67 |
| MORPHINE | 9300 | 1,268 | **225,366.05** | 177.73 |
| OXYMORPHONE | 9652 | 254 | **4,468.84** | 17.59 |
| TAPENTADOL | 9780 | 646 | **58,386.00** | 90.38 |
| FENTANYL BASE | 9801 | 1,178 | **3,765.98** | 3.20 |

**BUSINESS ACTIVITY: B - HOSPITALS**

| DRUG NAME | DRUG CODE | NUMBER OF REGISTRANT SOLD TO | TOTAL GRAMS | AVG GRAMS |
|---|---|---|---|---|
| AMPHETAMINE | 1100 | 105 | **9,475.20** | 90.24 |
| METHAMPHETAMINE | 1105 | 1 | **6.83** | 6.83 |
| LISDEXAMFETAMINE | 1205 | 50 | **3,022.88** | 60.46 |
| METHYLPHENIDATE (DL;D;L;ISOMERS) | 1724 | 124 | **7,976.37** | 64.33 |
| CODEINE | 9050 | 154 | **3,503.66** | 22.75 |
| BUPRENORPHINE | 9064 | 111 | **2,235.94** | 20.14 |
| OXYCODONE | 9143 | 220 | **34,910.08** | 158.68 |
| HYDROMORPHONE | 9150 | 179 | **3,158.21** | 17.64 |
| HYDROCODONE | 9193 | 225 | **16,553.13** | 73.57 |
| MEPERIDINE (PETHIDINE) | 9230 | 131 | **1,464.66** | 11.18 |
| METHADONE | 9250B | 114 | **2,373.28** | 20.82 |
| MORPHINE | 9300 | 225 | **19,170.58** | 85.20 |
| OXYMORPHONE | 9652 | 9 | **13.38** | 1.49 |
| TAPENTADOL | 9780 | 25 | **860.00** | 34.40 |
| FENTANYL BASE | 9801 | 257 | **916.71** | 3.57 |

**BUSINESS ACTIVITY: C - PRACTITIONERS**

| DRUG NAME | DRUG CODE | NUMBER OF REGISTRANT SOLD TO | TOTAL GRAMS | AVG GRAMS |
|---|---|---|---|---|
| CODEINE | 9050 | 56 | **4,039.17** | 72.13 |
| BUPRENORPHINE | 9064 | 490 | **228.88** | 0.47 |
| OXYCODONE | 9143 | 15 | **80.55** | 5.37 |
| HYDROMORPHONE | 9150 | 167 | **1,114.09** | 6.67 |
| HYDROCODONE | 9193 | 219 | **1,452.39** | 6.63 |
| MEPERIDINE (PETHIDINE) | 9230 | 14 | **18.30** | 1.31 |
| METHADONE | 9250B | 5 | **26.12** | 5.22 |
| MORPHINE | 9300 | 86 | **89.69** | 1.04 |
| FENTANYL BASE | 9801 | 151 | **12.99** | 0.09 |

**BUSINESS ACTIVITY: D - TEACHING INSTITUTIONS**

| DRUG NAME | DRUG CODE | NUMBER OF REGISTRANT SOLD TO | TOTAL GRAMS | AVG GRAMS |
|---|---|---|---|---|
| CODEINE | 9050 | 1 | **201.12** | 201.12 |
| BUPRENORPHINE | 9064 | 1 | **0.79** | 0.79 |
| HYDROMORPHONE | 9150 | 1 | **9.97** | 9.97 |
| HYDROCODONE | 9193 | 1 | **17.80** | 17.80 |
| MORPHINE | 9300 | 1 | **14.21** | 14.21 |
| FENTANYL BASE | 9801 | 3 | **1.86** | 0.62 |

**BUSINESS ACTIVITY: M - MID-LEVEL PRACTITIONERS**

| DRUG NAME | DRUG CODE | NUMBER OF REGISTRANT SOLD TO | TOTAL GRAMS | AVG GRAMS |
|---|---|---|---|---|
| HYDROMORPHONE | 9150 | 5 | **0.17** | 0.03 |
| MORPHINE | 9300 | 40 | **16.16** | 0.40 |
| FENTANYL BASE | 9801 | 63 | **0.86** | 0.01 |

**BUSINESS ACTIVITY: NTP - NARCOTIC TREATMENT PROGRAMS**

| DRUG NAME | DRUG CODE | NUMBER OF REGISTRANT SOLD TO | TOTAL GRAMS | AVG GRAMS |
|---|---|---|---|---|
| BUPRENORPHINE | 9064 | 16 | **781.64** | 48.85 |
| METHADONE | 9250B | 16 | **89,958.75** | 5,622.42 |

**STATE: MS - MISSISSIPPI**

**BUSINESS ACTIVITY: A - PHARMACIES**

| DRUG NAME | DRUG CODE | NUMBER OF REGISTRANT SOLD TO | TOTAL GRAMS | AVG GRAMS |
|---|---|---|---|---|
| AMPHETAMINE | 1100 | 665 | **255,014.24** | 383.48 |
| METHAMPHETAMINE | 1105 | 4 | **28.92** | 7.23 |
| LISDEXAMFETAMINE | 1205 | 636 | **130,969.13** | 205.93 |

| | | | | |
|---|---|---|---|---|
| METHYLPHENIDATE (DL;D;L;ISOMERS) | 1724 | 659 | **151,152.37** | 229.37 |
| CODEINE | 9050 | 664 | **66,278.16** | 99.82 |
| BUPRENORPHINE | 9064 | 610 | **49,713.47** | 81.50 |
| OXYCODONE | 9143 | 675 | **293,788.48** | 435.24 |
| HYDROMORPHONE | 9150 | 518 | **19,823.53** | 38.27 |
| HYDROCODONE | 9193 | 684 | **279,196.62** | 408.18 |
| MEPERIDINE (PETHIDINE) | 9230 | 85 | **1,549.74** | 18.23 |
| METHADONE | 9250B | 347 | **5,853.94** | 16.87 |
| MORPHINE | 9300 | 631 | **93,417.85** | 148.05 |
| OXYMORPHONE | 9652 | 121 | **2,296.71** | 18.98 |
| TAPENTADOL | 9780 | 316 | **26,616.50** | 84.23 |
| FENTANYL BASE | 9801 | 618 | **2,847.49** | 4.61 |

**BUSINESS ACTIVITY: B - HOSPITALS**

| DRUG NAME | DRUG CODE | NUMBER OF REGISTRANT SOLD TO | TOTAL GRAMS | AVG GRAMS |
|---|---|---|---|---|
| AMPHETAMINE | 1100 | 48 | **7,762.18** | 161.71 |
| LISDEXAMFETAMINE | 1205 | 35 | **4,094.68** | 116.99 |
| METHYLPHENIDATE (DL;D;L;ISOMERS) | 1724 | 63 | **4,650.14** | 73.81 |
| CODEINE | 9050 | 96 | **3,145.88** | 32.77 |
| BUPRENORPHINE | 9064 | 61 | **1,544.34** | 25.32 |
| OXYCODONE | 9143 | 132 | **11,645.27** | 88.22 |
| HYDROMORPHONE | 9150 | 123 | **1,367.41** | 11.12 |
| HYDROCODONE | 9193 | 141 | **12,141.68** | 86.11 |
| MEPERIDINE (PETHIDINE) | 9230 | 89 | **2,677.68** | 30.09 |
| METHADONE | 9250B | 54 | **259.51** | 4.81 |
| MORPHINE | 9300 | 137 | **6,656.39** | 48.59 |
| OXYMORPHONE | 9652 | 5 | **17.13** | 3.43 |
| TAPENTADOL | 9780 | 9 | **962.00** | 106.89 |
| FENTANYL BASE | 9801 | 133 | **350.36** | 2.63 |

**BUSINESS ACTIVITY: C - PRACTITIONERS**

| DRUG NAME | DRUG CODE | NUMBER OF REGISTRANT SOLD TO | TOTAL GRAMS | AVG GRAMS |
|---|---|---|---|---|
| CODEINE | 9050 | 15 | **985.70** | 65.71 |
| BUPRENORPHINE | 9064 | 122 | **4.37** | 0.04 |
| OXYCODONE | 9143 | 14 | **34.38** | 2.46 |
| HYDROMORPHONE | 9150 | 42 | **127.17** | 3.03 |
| HYDROCODONE | 9193 | 36 | **84.52** | 2.35 |
| MEPERIDINE (PETHIDINE) | 9230 | 35 | **300.45** | 8.58 |
| METHADONE | 9250B | 1 | **0.36** | 0.36 |
| MORPHINE | 9300 | 58 | **55.26** | 0.95 |
| FENTANYL BASE | 9801 | 111 | **7.57** | 0.07 |

**BUSINESS ACTIVITY: D - TEACHING INSTITUTIONS**

| DRUG NAME | DRUG CODE | NUMBER OF REGISTRANT SOLD TO | TOTAL GRAMS | AVG GRAMS |
|---|---|---|---|---|
| BUPRENORPHINE | 9064 | 1 | **0.00** | 0.00 |
| HYDROMORPHONE | 9150 | 1 | **1.42** | 1.42 |

**BUSINESS ACTIVITY: NTP - NARCOTIC TREATMENT PROGRAMS**

| DRUG NAME | DRUG CODE | NUMBER OF REGISTRANT SOLD TO | TOTAL GRAMS | AVG GRAMS |
|---|---|---|---|---|
| BUPRENORPHINE | 9064 | 4 | **198.86** | 49.71 |
| METHADONE | 9250B | 5 | **20,932.54** | 4,186.51 |

**STATE: MT - MONTANA**

**BUSINESS ACTIVITY: A - PHARMACIES**

| DRUG NAME | DRUG CODE | NUMBER OF REGISTRANT SOLD TO | TOTAL GRAMS | AVG GRAMS |
|---|---|---|---|---|
| AMPHETAMINE | 1100 | 228 | **65,290.03** | 286.36 |
| METHAMPHETAMINE | 1105 | 2 | **2.01** | 1.00 |
| LISDEXAMFETAMINE | 1205 | 220 | **27,057.39** | 122.99 |
| METHYLPHENIDATE (DL;D;L;ISOMERS) | 1724 | 225 | **62,113.48** | 276.06 |
| CODEINE | 9050 | 223 | **24,568.52** | 110.17 |
| BUPRENORPHINE | 9064 | 220 | **16,205.52** | 73.66 |
| OXYCODONE | 9143 | 230 | **96,288.69** | 418.65 |
| HYDROMORPHONE | 9150 | 215 | **4,368.35** | 20.32 |
| HYDROCODONE | 9193 | 232 | **67,179.00** | 289.56 |
| MEPERIDINE (PETHIDINE) | 9230 | 13 | **61.66** | 4.74 |
| METHADONE | 9250B | 172 | **4,282.21** | 24.90 |

| | | | | |
|---|---|---|---|---|
| MORPHINE | 9300 | 229 | **39,927.83** | 174.36 |
| OXYMORPHONE | 9652 | 36 | **295.02** | 8.19 |
| TAPENTADOL | 9780 | 68 | **4,354.00** | 64.03 |
| FENTANYL BASE | 9801 | 220 | **611.00** | 2.78 |

**BUSINESS ACTIVITY: B - HOSPITALS**

| DRUG NAME | DRUG CODE | NUMBER OF REGISTRANT SOLD TO | TOTAL GRAMS | AVG GRAMS |
|---|---|---|---|---|
| AMPHETAMINE | 1100 | 33 | **3,935.59** | 119.26 |
| LISDEXAMFETAMINE | 1205 | 26 | **1,746.44** | 67.17 |
| METHYLPHENIDATE (DL;D;L;ISOMERS) | 1724 | 42 | **5,151.08** | 122.64 |
| CODEINE | 9050 | 48 | **2,321.29** | 48.36 |
| BUPRENORPHINE | 9064 | 36 | **1,276.18** | 35.45 |
| OXYCODONE | 9143 | 84 | **9,671.17** | 115.13 |
| HYDROMORPHONE | 9150 | 70 | **736.70** | 10.52 |
| HYDROCODONE | 9193 | 87 | **8,412.94** | 96.70 |
| MEPERIDINE (PETHIDINE) | 9230 | 36 | **292.39** | 8.12 |
| METHADONE | 9250B | 27 | **457.22** | 16.93 |
| MORPHINE | 9300 | 87 | **6,299.48** | 72.41 |
| OXYMORPHONE | 9652 | 2 | **1.69** | 0.85 |
| TAPENTADOL | 9780 | 5 | **84.00** | 16.80 |
| FENTANYL BASE | 9801 | 87 | **158.19** | 1.82 |

**BUSINESS ACTIVITY: C - PRACTITIONERS**

| DRUG NAME | DRUG CODE | NUMBER OF REGISTRANT SOLD TO | TOTAL GRAMS | AVG GRAMS |
|---|---|---|---|---|
| CODEINE | 9050 | 19 | **468.50** | 24.66 |
| BUPRENORPHINE | 9064 | 141 | **8.90** | 0.06 |
| OXYCODONE | 9143 | 2 | **2.69** | 1.34 |
| HYDROMORPHONE | 9150 | 84 | **545.68** | 6.50 |
| HYDROCODONE | 9193 | 61 | **135.88** | 2.23 |
| MEPERIDINE (PETHIDINE) | 9230 | 3 | **19.61** | 6.54 |
| METHADONE | 9250B | 1 | **7.16** | 7.16 |
| MORPHINE | 9300 | 62 | **76.17** | 1.23 |
| FENTANYL BASE | 9801 | 87 | **7.62** | 0.09 |

**BUSINESS ACTIVITY: M - MID-LEVEL PRACTITIONERS**

| DRUG NAME | DRUG CODE | NUMBER OF REGISTRANT SOLD TO | TOTAL GRAMS | AVG GRAMS |
|---|---|---|---|---|
| BUPRENORPHINE | 9064 | 2 | **6.05** | 3.02 |
| MORPHINE | 9300 | 1 | **0.08** | 0.08 |
| FENTANYL BASE | 9801 | 3 | **0.01** | 0.00 |

**BUSINESS ACTIVITY: NTP - NARCOTIC TREATMENT PROGRAMS**

| DRUG NAME | DRUG CODE | NUMBER OF REGISTRANT SOLD TO | TOTAL GRAMS | AVG GRAMS |
|---|---|---|---|---|
| BUPRENORPHINE | 9064 | 4 | **839.72** | 209.93 |
| METHADONE | 9250B | 4 | **30,838.91** | 7,709.73 |

**STATE: NC - NORTH CAROLINA**

**BUSINESS ACTIVITY: A - PHARMACIES**

| DRUG NAME | DRUG CODE | NUMBER OF REGISTRANT SOLD TO | TOTAL GRAMS | AVG GRAMS |
|---|---|---|---|---|
| AMPHETAMINE | 1100 | 1,985 | **921,638.21** | 464.30 |
| METHAMPHETAMINE | 1105 | 44 | **137.80** | 3.13 |
| LISDEXAMFETAMINE | 1205 | 1,933 | **420,603.59** | 217.59 |
| METHYLPHENIDATE (DL;D;L;ISOMERS) | 1724 | 1,975 | **666,182.10** | 337.31 |
| CODEINE | 9050 | 1,860 | **125,238.75** | 67.33 |
| BUPRENORPHINE | 9064 | 1,966 | **208,615.27** | 106.11 |
| OXYCODONE | 9143 | 1,999 | **1,600,192.81** | 800.50 |
| HYDROMORPHONE | 9150 | 1,841 | **25,157.38** | 13.67 |
| HYDROCODONE | 9193 | 2,009 | **503,107.78** | 250.43 |
| MEPERIDINE (PETHIDINE) | 9230 | 135 | **1,654.32** | 12.25 |
| METHADONE | 9250B | 1,680 | **62,569.36** | 37.24 |
| MORPHINE | 9300 | 1,945 | **327,246.98** | 168.25 |
| OXYMORPHONE | 9652 | 896 | **19,742.13** | 22.03 |
| TAPENTADOL | 9780 | 1,536 | **264,910.00** | 172.47 |
| FENTANYL BASE | 9801 | 1,826 | **5,098.21** | 2.79 |

**BUSINESS ACTIVITY: B - HOSPITALS**

| DRUG NAME | DRUG CODE | NUMBER OF REGISTRANT SOLD TO | TOTAL GRAMS | AVG GRAMS |
|---|---|---|---|---|
| AMPHETAMINE | 1100 | 154 | **29,709.75** | 192.92 |

| | | | | |
|---|---|---|---|---|
| METHAMPHETAMINE | 1105 | 1 | **5.22** | 5.22 |
| LISDEXAMFETAMINE | 1205 | 87 | **8,301.52** | 95.42 |
| METHYLPHENIDATE (DL;D;L;ISOMERS) | 1724 | 176 | **26,886.40** | 152.76 |
| CODEINE | 9050 | 135 | **3,730.66** | 27.63 |
| BUPRENORPHINE | 9064 | 305 | **6,451.68** | 21.15 |
| OXYCODONE | 9143 | 265 | **77,655.58** | 293.04 |
| HYDROMORPHONE | 9150 | 338 | **8,015.19** | 23.71 |
| HYDROCODONE | 9193 | 282 | **27,634.27** | 97.99 |
| MEPERIDINE (PETHIDINE) | 9230 | 127 | **832.17** | 6.55 |
| METHADONE | 9250B | 184 | **6,423.91** | 34.91 |
| MORPHINE | 9300 | 311 | **30,511.70** | 98.11 |
| OXYMORPHONE | 9652 | 48 | **291.88** | 6.08 |
| TAPENTADOL | 9780 | 76 | **9,226.75** | 121.40 |
| FENTANYL BASE | 9801 | 337 | **973.41** | 2.89 |

**BUSINESS ACTIVITY: C - PRACTITIONERS**

| DRUG NAME | DRUG CODE | NUMBER OF REGISTRANT SOLD TO | TOTAL GRAMS | AVG GRAMS |
|---|---|---|---|---|
| CODEINE | 9050 | 12 | **206.38** | 17.20 |
| BUPRENORPHINE | 9064 | 1,008 | **192.37** | 0.19 |
| OXYCODONE | 9143 | 12 | **16.59** | 1.38 |
| HYDROMORPHONE | 9150 | 346 | **2,284.05** | 6.60 |
| HYDROCODONE | 9193 | 151 | **346.27** | 2.29 |
| MEPERIDINE (PETHIDINE) | 9230 | 53 | **198.48** | 3.74 |
| METHADONE | 9250B | 5 | **14.58** | 2.92 |
| MORPHINE | 9300 | 126 | **130.18** | 1.03 |
| FENTANYL BASE | 9801 | 421 | **32.76** | 0.08 |

**BUSINESS ACTIVITY: D - TEACHING INSTITUTIONS**

| DRUG NAME | DRUG CODE | NUMBER OF REGISTRANT SOLD TO | TOTAL GRAMS | AVG GRAMS |
|---|---|---|---|---|
| BUPRENORPHINE | 9064 | 5 | **0.02** | 0.00 |
| HYDROMORPHONE | 9150 | 3 | **4.70** | 1.57 |

**BUSINESS ACTIVITY: M - MID-LEVEL PRACTITIONERS**

| DRUG NAME | DRUG CODE | NUMBER OF REGISTRANT SOLD TO | TOTAL GRAMS | AVG GRAMS |
|---|---|---|---|---|
| BUPRENORPHINE | 9064 | 5 | **262.93** | 52.59 |
| HYDROMORPHONE | 9150 | 7 | **16.24** | 2.32 |
| MORPHINE | 9300 | 71 | **52.89** | 0.74 |
| FENTANYL BASE | 9801 | 77 | **2.16** | 0.03 |

**BUSINESS ACTIVITY: NTP - NARCOTIC TREATMENT PROGRAMS**

| DRUG NAME | DRUG CODE | NUMBER OF REGISTRANT SOLD TO | TOTAL GRAMS | AVG GRAMS |
|---|---|---|---|---|
| BUPRENORPHINE | 9064 | 68 | **14,341.18** | 210.90 |
| METHADONE | 9250B | 82 | **423,135.27** | 5,160.19 |

**STATE: ND - NORTH DAKOTA**

**BUSINESS ACTIVITY: A - PHARMACIES**

| DRUG NAME | DRUG CODE | NUMBER OF REGISTRANT SOLD TO | TOTAL GRAMS | AVG GRAMS |
|---|---|---|---|---|
| AMPHETAMINE | 1100 | 177 | **38,558.51** | 217.84 |
| METHAMPHETAMINE | 1105 | 4 | **15.67** | 3.92 |
| LISDEXAMFETAMINE | 1205 | 175 | **24,086.54** | 137.64 |
| METHYLPHENIDATE (DL;D;L;ISOMERS) | 1724 | 177 | **52,948.14** | 299.14 |
| CODEINE | 9050 | 166 | **12,147.48** | 73.18 |
| BUPRENORPHINE | 9064 | 150 | **6,241.58** | 41.61 |
| OXYCODONE | 9143 | 175 | **33,923.05** | 193.85 |
| HYDROMORPHONE | 9150 | 145 | **1,217.25** | 8.39 |
| HYDROCODONE | 9193 | 180 | **22,435.50** | 124.64 |
| MEPERIDINE (PETHIDINE) | 9230 | 9 | **33.44** | 3.72 |
| METHADONE | 9250B | 115 | **2,062.04** | 17.93 |
| MORPHINE | 9300 | 171 | **13,383.26** | 78.26 |
| OXYMORPHONE | 9652 | 17 | **194.30** | 11.43 |
| TAPENTADOL | 9780 | 57 | **3,474.00** | 60.95 |
| FENTANYL BASE | 9801 | 161 | **381.53** | 2.37 |

**BUSINESS ACTIVITY: B - HOSPITALS**

| DRUG NAME | DRUG CODE | NUMBER OF REGISTRANT SOLD TO | TOTAL GRAMS | AVG GRAMS |
|---|---|---|---|---|
| AMPHETAMINE | 1100 | 17 | **2,909.65** | 171.16 |
| METHAMPHETAMINE | 1105 | 1 | **0.80** | 0.80 |

| | | | | |
|---|---|---|---|---|
| LISDEXAMFETAMINE | 1205 | 15 | **1,468.95** | 97.93 |
| METHYLPHENIDATE (DL;D;L;ISOMERS) | 1724 | 29 | **5,079.77** | 175.16 |
| CODEINE | 9050 | 48 | **1,803.40** | 37.57 |
| BUPRENORPHINE | 9064 | 26 | **800.94** | 30.81 |
| OXYCODONE | 9143 | 54 | **5,979.05** | 110.72 |
| HYDROMORPHONE | 9150 | 54 | **572.85** | 10.61 |
| HYDROCODONE | 9193 | 57 | **4,688.77** | 82.26 |
| MEPERIDINE (PETHIDINE) | 9230 | 16 | **85.84** | 5.37 |
| METHADONE | 9250B | 24 | **310.45** | 12.94 |
| MORPHINE | 9300 | 59 | **3,809.44** | 64.57 |
| OXYMORPHONE | 9652 | 1 | **4.55** | 4.55 |
| TAPENTADOL | 9780 | 5 | **521.50** | 104.30 |
| FENTANYL BASE | 9801 | 53 | **139.88** | 2.64 |

**BUSINESS ACTIVITY: C - PRACTITIONERS**

| DRUG NAME | DRUG CODE | NUMBER OF REGISTRANT SOLD TO | TOTAL GRAMS | AVG GRAMS |
|---|---|---|---|---|
| AMPHETAMINE | 1100 | 1 | **2.50** | 2.50 |
| LISDEXAMFETAMINE | 1205 | 1 | **6.36** | 6.36 |
| CODEINE | 9050 | 6 | **87.03** | 14.50 |
| BUPRENORPHINE | 9064 | 62 | **23.50** | 0.38 |
| OXYCODONE | 9143 | 4 | **7.80** | 1.95 |
| HYDROMORPHONE | 9150 | 26 | **106.26** | 4.09 |
| HYDROCODONE | 9193 | 22 | **53.44** | 2.43 |
| MEPERIDINE (PETHIDINE) | 9230 | 5 | **22.44** | 4.49 |
| METHADONE | 9250B | 2 | **10.55** | 5.28 |
| MORPHINE | 9300 | 18 | **19.94** | 1.11 |
| FENTANYL BASE | 9801 | 42 | **4.81** | 0.11 |

**BUSINESS ACTIVITY: M - MID-LEVEL PRACTITIONERS**

| DRUG NAME | DRUG CODE | NUMBER OF REGISTRANT SOLD TO | TOTAL GRAMS | AVG GRAMS |
|---|---|---|---|---|
| LISDEXAMFETAMINE | 1205 | 1 | **1.73** | 1.73 |
| BUPRENORPHINE | 9064 | 3 | **123.12** | 41.04 |
| OXYCODONE | 9143 | 1 | **1.12** | 1.12 |
| HYDROMORPHONE | 9150 | 1 | **0.09** | 0.09 |
| HYDROCODONE | 9193 | 1 | **0.45** | 0.45 |
| MORPHINE | 9300 | 1 | **0.86** | 0.86 |
| FENTANYL BASE | 9801 | 1 | **0.12** | 0.12 |

**BUSINESS ACTIVITY: NTP - NARCOTIC TREATMENT PROGRAMS**

| DRUG NAME | DRUG CODE | NUMBER OF REGISTRANT SOLD TO | TOTAL GRAMS | AVG GRAMS |
|---|---|---|---|---|
| METHADONE | 9250B | 3 | **21,179.39** | 7,059.80 |

**STATE: NE - NEBRASKA**

**BUSINESS ACTIVITY: A - PHARMACIES**

| DRUG NAME | DRUG CODE | NUMBER OF REGISTRANT SOLD TO | TOTAL GRAMS | AVG GRAMS |
|---|---|---|---|---|
| AMPHETAMINE | 1100 | 420 | **122,868.94** | 292.55 |
| METHAMPHETAMINE | 1105 | 12 | **22.50** | 1.87 |
| LISDEXAMFETAMINE | 1205 | 408 | **71,474.55** | 175.18 |
| METHYLPHENIDATE (DL;D;L;ISOMERS) | 1724 | 419 | **118,102.05** | 281.87 |
| CODEINE | 9050 | 394 | **28,346.80** | 71.95 |
| BUPRENORPHINE | 9064 | 384 | **8,460.68** | 22.03 |
| OXYCODONE | 9143 | 425 | **101,954.50** | 239.89 |
| HYDROMORPHONE | 9150 | 365 | **2,659.55** | 7.29 |
| HYDROCODONE | 9193 | 428 | **80,343.59** | 187.72 |
| MEPERIDINE (PETHIDINE) | 9230 | 21 | **73.10** | 3.48 |
| METHADONE | 9250B | 302 | **6,083.80** | 20.15 |
| MORPHINE | 9300 | 421 | **49,041.84** | 116.49 |
| OXYMORPHONE | 9652 | 97 | **1,046.79** | 10.79 |
| TAPENTADOL | 9780 | 273 | **29,042.00** | 106.38 |
| FENTANYL BASE | 9801 | 382 | **1,051.27** | 2.75 |

**BUSINESS ACTIVITY: B - HOSPITALS**

| DRUG NAME | DRUG CODE | NUMBER OF REGISTRANT SOLD TO | TOTAL GRAMS | AVG GRAMS |
|---|---|---|---|---|
| AMPHETAMINE | 1100 | 41 | **3,717.86** | 90.68 |
| LISDEXAMFETAMINE | 1205 | 31 | **1,023.24** | 33.01 |
| METHYLPHENIDATE (DL;D;L;ISOMERS) | 1724 | 57 | **3,671.20** | 64.41 |

| | | | | |
|---|---|---|---|---|
| CODEINE | 9050 | 63 | **1,410.11** | 22.38 |
| BUPRENORPHINE | 9064 | 53 | **382.15** | 7.21 |
| OXYCODONE | 9143 | 125 | **7,152.27** | 57.22 |
| HYDROMORPHONE | 9150 | 110 | **612.22** | 5.57 |
| HYDROCODONE | 9193 | 126 | **4,173.22** | 33.12 |
| MEPERIDINE (PETHIDINE) | 9230 | 75 | **422.57** | 5.63 |
| METHADONE | 9250B | 58 | **600.00** | 10.34 |
| MORPHINE | 9300 | 126 | **6,297.52** | 49.98 |
| OXYMORPHONE | 9652 | 4 | **22.84** | 5.71 |
| TAPENTADOL | 9780 | 33 | **833.00** | 25.24 |
| FENTANYL BASE | 9801 | 132 | **193.45** | 1.47 |

**BUSINESS ACTIVITY: C - PRACTITIONERS**

| DRUG NAME | DRUG CODE | NUMBER OF REGISTRANT SOLD TO | TOTAL GRAMS | AVG GRAMS |
|---|---|---|---|---|
| METHYLPHENIDATE (DL;D;L;ISOMERS) | 1724 | 1 | **0.26** | 0.26 |
| CODEINE | 9050 | 19 | **1,248.79** | 65.73 |
| BUPRENORPHINE | 9064 | 181 | **15.25** | 0.08 |
| OXYCODONE | 9143 | 11 | **120.58** | 10.96 |
| HYDROMORPHONE | 9150 | 82 | **572.88** | 6.99 |
| HYDROCODONE | 9193 | 53 | **102.38** | 1.93 |
| MEPERIDINE (PETHIDINE) | 9230 | 12 | **37.69** | 3.14 |
| METHADONE | 9250B | 3 | **19.23** | 6.41 |
| MORPHINE | 9300 | 42 | **77.07** | 1.84 |
| TAPENTADOL | 9780 | 1 | **60.00** | 60.00 |
| FENTANYL BASE | 9801 | 95 | **9.91** | 0.10 |

**BUSINESS ACTIVITY: D - TEACHING INSTITUTIONS**

| DRUG NAME | DRUG CODE | NUMBER OF REGISTRANT SOLD TO | TOTAL GRAMS | AVG GRAMS |
|---|---|---|---|---|
| BUPRENORPHINE | 9064 | 1 | **0.00** | 0.00 |

**BUSINESS ACTIVITY: M - MID-LEVEL PRACTITIONERS**

| DRUG NAME | DRUG CODE | NUMBER OF REGISTRANT SOLD TO | TOTAL GRAMS | AVG GRAMS |
|---|---|---|---|---|
| HYDROMORPHONE | 9150 | 5 | **0.12** | 0.02 |
| MORPHINE | 9300 | 25 | **5.49** | 0.22 |
| FENTANYL BASE | 9801 | 39 | **0.55** | 0.01 |

**BUSINESS ACTIVITY: NTP - NARCOTIC TREATMENT PROGRAMS**

| DRUG NAME | DRUG CODE | NUMBER OF REGISTRANT SOLD TO | TOTAL GRAMS | AVG GRAMS |
|---|---|---|---|---|
| BUPRENORPHINE | 9064 | 2 | **44.25** | 22.13 |
| METHADONE | 9250B | 3 | **17,995.33** | 5,998.44 |

**STATE: NH - NEW HAMPSHIRE**

**BUSINESS ACTIVITY: A - PHARMACIES**

| DRUG NAME | DRUG CODE | NUMBER OF REGISTRANT SOLD TO | TOTAL GRAMS | AVG GRAMS |
|---|---|---|---|---|
| AMPHETAMINE | 1100 | 262 | **178,197.19** | 680.14 |
| METHAMPHETAMINE | 1105 | 5 | **24.51** | 4.90 |
| LISDEXAMFETAMINE | 1205 | 261 | **58,319.31** | 223.45 |
| METHYLPHENIDATE (DL;D;L;ISOMERS) | 1724 | 262 | **132,510.71** | 505.77 |
| CODEINE | 9050 | 255 | **22,172.54** | 86.95 |
| BUPRENORPHINE | 9064 | 263 | **43,599.59** | 165.78 |
| OXYCODONE | 9143 | 261 | **158,502.57** | 607.29 |
| HYDROMORPHONE | 9150 | 253 | **4,472.68** | 17.68 |
| HYDROCODONE | 9193 | 262 | **25,258.47** | 96.41 |
| MEPERIDINE (PETHIDINE) | 9230 | 9 | **24.29** | 2.70 |
| METHADONE | 9250B | 236 | **12,746.27** | 54.01 |
| MORPHINE | 9300 | 258 | **49,811.65** | 193.07 |
| OXYMORPHONE | 9652 | 63 | **655.43** | 10.40 |
| TAPENTADOL | 9780 | 169 | **21,458.00** | 126.97 |
| FENTANYL BASE | 9801 | 245 | **669.08** | 2.73 |

**BUSINESS ACTIVITY: B - HOSPITALS**

| DRUG NAME | DRUG CODE | NUMBER OF REGISTRANT SOLD TO | TOTAL GRAMS | AVG GRAMS |
|---|---|---|---|---|
| AMPHETAMINE | 1100 | 25 | **2,954.98** | 118.20 |
| LISDEXAMFETAMINE | 1205 | 13 | **352.06** | 27.08 |
| METHYLPHENIDATE (DL;D;L;ISOMERS) | 1724 | 34 | **1,841.44** | 54.16 |
| CODEINE | 9050 | 26 | **399.46** | 15.36 |
| BUPRENORPHINE | 9064 | 36 | **1,167.72** | 32.44 |

| | | | | |
|---|---|---|---|---|
| OXYCODONE | 9143 | 45 | **7,211.49** | 160.26 |
| HYDROMORPHONE | 9150 | 44 | **907.09** | 20.62 |
| HYDROCODONE | 9193 | 45 | **795.67** | 17.68 |
| MEPERIDINE (PETHIDINE) | 9230 | 17 | **119.94** | 7.06 |
| METHADONE | 9250B | 35 | **1,137.51** | 32.50 |
| MORPHINE | 9300 | 42 | **3,289.22** | 78.31 |
| TAPENTADOL | 9780 | 4 | **532.00** | 133.00 |
| FENTANYL BASE | 9801 | 50 | **100.68** | 2.01 |

**BUSINESS ACTIVITY: C - PRACTITIONERS**

| DRUG NAME | DRUG CODE | NUMBER OF REGISTRANT SOLD TO | TOTAL GRAMS | AVG GRAMS |
|---|---|---|---|---|
| CODEINE | 9050 | 1 | **17.58** | 17.58 |
| BUPRENORPHINE | 9064 | 201 | **15.68** | 0.08 |
| OXYCODONE | 9143 | 3 | **0.93** | 0.31 |
| HYDROMORPHONE | 9150 | 88 | **421.66** | 4.79 |
| HYDROCODONE | 9193 | 8 | **4.21** | 0.53 |
| METHADONE | 9250B | 9 | **7.69** | 0.85 |
| MORPHINE | 9300 | 33 | **17.45** | 0.53 |
| FENTANYL BASE | 9801 | 81 | **8.26** | 0.10 |

**BUSINESS ACTIVITY: M - MID-LEVEL PRACTITIONERS**

| DRUG NAME | DRUG CODE | NUMBER OF REGISTRANT SOLD TO | TOTAL GRAMS | AVG GRAMS |
|---|---|---|---|---|
| BUPRENORPHINE | 9064 | 3 | **5.35** | 1.78 |
| MORPHINE | 9300 | 1 | **0.02** | 0.02 |
| FENTANYL BASE | 9801 | 1 | **0.00** | 0.00 |

**BUSINESS ACTIVITY: NTP - NARCOTIC TREATMENT PROGRAMS**

| DRUG NAME | DRUG CODE | NUMBER OF REGISTRANT SOLD TO | TOTAL GRAMS | AVG GRAMS |
|---|---|---|---|---|
| BUPRENORPHINE | 9064 | 6 | **317.04** | 52.84 |
| METHADONE | 9250B | 10 | **89,493.66** | 8,949.37 |

**STATE: NJ - NEW JERSEY**

**BUSINESS ACTIVITY: A - PHARMACIES**

| DRUG NAME | DRUG CODE | NUMBER OF REGISTRANT SOLD TO | TOTAL GRAMS | AVG GRAMS |
|---|---|---|---|---|
| AMPHETAMINE | 1100 | 1,954 | **490,816.22** | 251.19 |
| METHAMPHETAMINE | 1105 | 46 | **118.12** | 2.57 |
| LISDEXAMFETAMINE | 1205 | 1,701 | **188,460.02** | 110.79 |
| METHYLPHENIDATE (DL;D;L;ISOMERS) | 1724 | 1,892 | **340,139.89** | 179.78 |
| CODEINE | 9050 | 1,965 | **213,314.35** | 108.56 |
| BUPRENORPHINE | 9064 | 1,848 | **131,533.03** | 71.18 |
| OXYCODONE | 9143 | 2,020 | **1,291,395.02** | 639.30 |
| HYDROMORPHONE | 9150 | 1,636 | **20,730.69** | 12.67 |
| HYDROCODONE | 9193 | 1,899 | **91,026.97** | 47.93 |
| MEPERIDINE (PETHIDINE) | 9230 | 41 | **526.82** | 12.85 |
| METHADONE | 9250B | 1,089 | **28,281.11** | 25.97 |
| MORPHINE | 9300 | 1,735 | **214,704.90** | 123.75 |
| OXYMORPHONE | 9652 | 467 | **7,320.48** | 15.68 |
| TAPENTADOL | 9780 | 1,042 | **106,727.00** | 102.43 |
| FENTANYL BASE | 9801 | 1,586 | **3,939.93** | 2.48 |

**BUSINESS ACTIVITY: B - HOSPITALS**

| DRUG NAME | DRUG CODE | NUMBER OF REGISTRANT SOLD TO | TOTAL GRAMS | AVG GRAMS |
|---|---|---|---|---|
| AMPHETAMINE | 1100 | 59 | **3,664.42** | 62.11 |
| LISDEXAMFETAMINE | 1205 | 23 | **1,184.53** | 51.50 |
| METHYLPHENIDATE (DL;D;L;ISOMERS) | 1724 | 98 | **3,426.44** | 34.96 |
| CODEINE | 9050 | 96 | **2,485.62** | 25.89 |
| BUPRENORPHINE | 9064 | 102 | **1,681.72** | 16.49 |
| OXYCODONE | 9143 | 120 | **26,043.16** | 217.03 |
| HYDROMORPHONE | 9150 | 109 | **5,435.35** | 49.87 |
| HYDROCODONE | 9193 | 83 | **1,009.51** | 12.16 |
| MEPERIDINE (PETHIDINE) | 9230 | 58 | **428.89** | 7.39 |
| METHADONE | 9250B | 100 | **4,292.49** | 42.92 |
| MORPHINE | 9300 | 117 | **12,323.42** | 105.33 |
| OXYMORPHONE | 9652 | 2 | **10.71** | 5.35 |
| TAPENTADOL | 9780 | 26 | **883.00** | 33.96 |
| FENTANYL BASE | 9801 | 108 | **863.79** | 8.00 |

**BUSINESS ACTIVITY: C - PRACTITIONERS**

| DRUG NAME | DRUG CODE | NUMBER OF REGISTRANT SOLD TO | TOTAL GRAMS | AVG GRAMS |
|---|---|---|---|---|
| AMPHETAMINE | 1100 | 1 | **3.75** | 3.75 |
| CODEINE | 9050 | 81 | **1,971.70** | 24.34 |
| BUPRENORPHINE | 9064 | 634 | **5,861.25** | 9.24 |
| OXYCODONE | 9143 | 77 | **105.83** | 1.37 |
| HYDROMORPHONE | 9150 | 269 | **1,589.76** | 5.91 |
| HYDROCODONE | 9193 | 225 | **862.76** | 3.83 |
| MEPERIDINE (PETHIDINE) | 9230 | 49 | **143.36** | 2.93 |
| METHADONE | 9250B | 36 | **141.85** | 3.94 |
| MORPHINE | 9300 | 132 | **302.44** | 2.29 |
| FENTANYL BASE | 9801 | 464 | **43.61** | 0.09 |

**BUSINESS ACTIVITY: D - TEACHING INSTITUTIONS**

| DRUG NAME | DRUG CODE | NUMBER OF REGISTRANT SOLD TO | TOTAL GRAMS | AVG GRAMS |
|---|---|---|---|---|
| BUPRENORPHINE | 9064 | 2 | **0.01** | 0.00 |

**BUSINESS ACTIVITY: M - MID-LEVEL PRACTITIONERS**

| DRUG NAME | DRUG CODE | NUMBER OF REGISTRANT SOLD TO | TOTAL GRAMS | AVG GRAMS |
|---|---|---|---|---|
| BUPRENORPHINE | 9064 | 2 | **3.30** | 1.65 |

**BUSINESS ACTIVITY: NTP - NARCOTIC TREATMENT PROGRAMS**

| DRUG NAME | DRUG CODE | NUMBER OF REGISTRANT SOLD TO | TOTAL GRAMS | AVG GRAMS |
|---|---|---|---|---|
| BUPRENORPHINE | 9064 | 33 | **2,328.62** | 70.56 |
| METHADONE | 9250B | 44 | **510,346.43** | 11,598.78 |

**STATE: NM - NEW MEXICO**

**BUSINESS ACTIVITY: A - PHARMACIES**

| DRUG NAME | DRUG CODE | NUMBER OF REGISTRANT SOLD TO | TOTAL GRAMS | AVG GRAMS |
|---|---|---|---|---|
| AMPHETAMINE | 1100 | 299 | **65,146.14** | 217.88 |
| METHAMPHETAMINE | 1105 | 16 | **36.56** | 2.29 |
| LISDEXAMFETAMINE | 1205 | 270 | **14,455.39** | 53.54 |
| METHYLPHENIDATE (DL;D;L;ISOMERS) | 1724 | 300 | **61,852.19** | 206.17 |
| CODEINE | 9050 | 300 | **47,688.57** | 158.96 |
| BUPRENORPHINE | 9064 | 293 | **38,320.92** | 130.79 |
| OXYCODONE | 9143 | 307 | **230,095.75** | 749.50 |
| HYDROMORPHONE | 9150 | 272 | **3,027.86** | 11.13 |
| HYDROCODONE | 9193 | 304 | **92,684.70** | 304.88 |
| MEPERIDINE (PETHIDINE) | 9230 | 41 | **539.24** | 13.15 |
| METHADONE | 9250B | 223 | **5,563.26** | 24.95 |
| MORPHINE | 9300 | 302 | **59,966.74** | 198.57 |
| OXYMORPHONE | 9652 | 81 | **870.29** | 10.74 |
| TAPENTADOL | 9780 | 104 | **6,929.00** | 66.63 |
| FENTANYL BASE | 9801 | 272 | **715.85** | 2.63 |

**BUSINESS ACTIVITY: B - HOSPITALS**

| DRUG NAME | DRUG CODE | NUMBER OF REGISTRANT SOLD TO | TOTAL GRAMS | AVG GRAMS |
|---|---|---|---|---|
| AMPHETAMINE | 1100 | 39 | **2,397.50** | 61.47 |
| LISDEXAMFETAMINE | 1205 | 12 | **362.47** | 30.21 |
| METHYLPHENIDATE (DL;D;L;ISOMERS) | 1724 | 58 | **5,097.65** | 87.89 |
| CODEINE | 9050 | 57 | **5,572.78** | 97.77 |
| BUPRENORPHINE | 9064 | 66 | **2,000.49** | 30.31 |
| OXYCODONE | 9143 | 89 | **16,229.59** | 182.35 |
| HYDROMORPHONE | 9150 | 66 | **682.94** | 10.35 |
| HYDROCODONE | 9193 | 91 | **7,349.83** | 80.77 |
| MEPERIDINE (PETHIDINE) | 9230 | 42 | **201.10** | 4.79 |
| METHADONE | 9250B | 50 | **1,488.10** | 29.76 |
| MORPHINE | 9300 | 91 | **9,107.47** | 100.08 |
| OXYMORPHONE | 9652 | 3 | **12.67** | 4.22 |
| TAPENTADOL | 9780 | 3 | **244.50** | 81.50 |
| FENTANYL BASE | 9801 | 97 | **208.88** | 2.15 |

**BUSINESS ACTIVITY: C - PRACTITIONERS**

| DRUG NAME | DRUG CODE | NUMBER OF REGISTRANT SOLD TO | TOTAL GRAMS | AVG GRAMS |
|---|---|---|---|---|
| CODEINE | 9050 | 4 | **19.11** | 4.78 |
| BUPRENORPHINE | 9064 | 142 | **10.97** | 0.08 |
| OXYCODONE | 9143 | 1 | **4.03** | 4.03 |

| | | | | |
|---|---|---|---|---|
| HYDROMORPHONE | 9150 | 71 | **565.85** | 7.97 |
| HYDROCODONE | 9193 | 37 | **59.81** | 1.62 |
| MEPERIDINE (PETHIDINE) | 9230 | 2 | **7.08** | 3.54 |
| METHADONE | 9250B | 3 | **3.40** | 1.13 |
| MORPHINE | 9300 | 27 | **36.48** | 1.35 |
| FENTANYL BASE | 9801 | 93 | **14.40** | 0.15 |

**BUSINESS ACTIVITY: D - TEACHING INSTITUTIONS**

| DRUG NAME | DRUG CODE | NUMBER OF REGISTRANT SOLD TO | TOTAL GRAMS | AVG GRAMS |
|---|---|---|---|---|
| BUPRENORPHINE | 9064 | 1 | **0.01** | 0.01 |
| OXYCODONE | 9143 | 1 | **2.69** | 2.69 |

**BUSINESS ACTIVITY: M - MID-LEVEL PRACTITIONERS**

| DRUG NAME | DRUG CODE | NUMBER OF REGISTRANT SOLD TO | TOTAL GRAMS | AVG GRAMS |
|---|---|---|---|---|
| AMPHETAMINE | 1100 | 1 | **7.51** | 7.51 |
| CODEINE | 9050 | 1 | **3.32** | 3.32 |
| BUPRENORPHINE | 9064 | 4 | **0.87** | 0.22 |
| HYDROMORPHONE | 9150 | 7 | **34.31** | 4.90 |
| HYDROCODONE | 9193 | 3 | **4.86** | 1.62 |
| MORPHINE | 9300 | 20 | **12.89** | 0.64 |
| FENTANYL BASE | 9801 | 47 | **2.26** | 0.05 |

**BUSINESS ACTIVITY: NTP - NARCOTIC TREATMENT PROGRAMS**

| DRUG NAME | DRUG CODE | NUMBER OF REGISTRANT SOLD TO | TOTAL GRAMS | AVG GRAMS |
|---|---|---|---|---|
| BUPRENORPHINE | 9064 | 6 | **472.28** | 78.71 |
| METHADONE | 9250B | 24 | **184,461.06** | 7,685.88 |

**STATE: NV - NEVADA**

**BUSINESS ACTIVITY: A - PHARMACIES**

| DRUG NAME | DRUG CODE | NUMBER OF REGISTRANT SOLD TO | TOTAL GRAMS | AVG GRAMS |
|---|---|---|---|---|
| AMPHETAMINE | 1100 | 440 | **119,142.17** | 270.78 |
| METHAMPHETAMINE | 1105 | 10 | **33.75** | 3.37 |
| LISDEXAMFETAMINE | 1205 | 394 | **29,648.44** | 75.25 |
| METHYLPHENIDATE (DL;D;L;ISOMERS) | 1724 | 418 | **54,244.50** | 129.77 |
| CODEINE | 9050 | 433 | **60,365.50** | 139.41 |
| BUPRENORPHINE | 9064 | 427 | **19,016.14** | 44.53 |
| OXYCODONE | 9143 | 446 | **492,723.04** | 1,104.76 |
| HYDROMORPHONE | 9150 | 397 | **5,615.08** | 14.14 |
| HYDROCODONE | 9193 | 447 | **240,981.85** | 539.11 |
| MEPERIDINE (PETHIDINE) | 9230 | 21 | **316.35** | 15.06 |
| METHADONE | 9250B | 406 | **23,037.69** | 56.74 |
| MORPHINE | 9300 | 441 | **125,578.71** | 284.76 |
| OXYMORPHONE | 9652 | 165 | **2,948.75** | 17.87 |
| TAPENTADOL | 9780 | 248 | **22,317.50** | 89.99 |
| FENTANYL BASE | 9801 | 378 | **1,025.94** | 2.71 |

**BUSINESS ACTIVITY: B - HOSPITALS**

| DRUG NAME | DRUG CODE | NUMBER OF REGISTRANT SOLD TO | TOTAL GRAMS | AVG GRAMS |
|---|---|---|---|---|
| AMPHETAMINE | 1100 | 38 | **3,675.08** | 96.71 |
| LISDEXAMFETAMINE | 1205 | 11 | **405.83** | 36.89 |
| METHYLPHENIDATE (DL;D;L;ISOMERS) | 1724 | 48 | **4,009.01** | 83.52 |
| CODEINE | 9050 | 43 | **2,232.23** | 51.91 |
| BUPRENORPHINE | 9064 | 38 | **691.36** | 18.19 |
| OXYCODONE | 9143 | 93 | **24,562.79** | 264.12 |
| HYDROMORPHONE | 9150 | 84 | **1,964.40** | 23.39 |
| HYDROCODONE | 9193 | 97 | **15,537.86** | 160.18 |
| MEPERIDINE (PETHIDINE) | 9230 | 64 | **946.99** | 14.80 |
| METHADONE | 9250B | 48 | **1,255.83** | 26.16 |
| MORPHINE | 9300 | 104 | **12,879.87** | 123.84 |
| OXYMORPHONE | 9652 | 3 | **3.03** | 1.01 |
| TAPENTADOL | 9780 | 8 | **228.50** | 28.56 |
| FENTANYL BASE | 9801 | 124 | **368.32** | 2.97 |

**BUSINESS ACTIVITY: C - PRACTITIONERS**

| DRUG NAME | DRUG CODE | NUMBER OF REGISTRANT SOLD TO | TOTAL GRAMS | AVG GRAMS |
|---|---|---|---|---|
| CODEINE | 9050 | 5 | **79.34** | 15.87 |
| BUPRENORPHINE | 9064 | 252 | **62.63** | 0.25 |

| | | | | |
|---|---|---|---|---|
| OXYCODONE | 9143 | 16 | **181.41** | 11.34 |
| HYDROMORPHONE | 9150 | 133 | **1,798.84** | 13.53 |
| HYDROCODONE | 9193 | 90 | **346.92** | 3.85 |
| MEPERIDINE (PETHIDINE) | 9230 | 46 | **580.20** | 12.61 |
| METHADONE | 9250B | 12 | **13.77** | 1.15 |
| MORPHINE | 9300 | 69 | **66.60** | 0.97 |
| FENTANYL BASE | 9801 | 167 | **25.79** | 0.15 |

**BUSINESS ACTIVITY: M - MID-LEVEL PRACTITIONERS**

| DRUG NAME | DRUG CODE | NUMBER OF REGISTRANT SOLD TO | TOTAL GRAMS | AVG GRAMS |
|---|---|---|---|---|
| CODEINE | 9050 | 1 | **19.89** | 19.89 |
| BUPRENORPHINE | 9064 | 5 | **32.47** | 6.49 |
| MORPHINE | 9300 | 1 | **0.38** | 0.38 |

**BUSINESS ACTIVITY: NTP - NARCOTIC TREATMENT PROGRAMS**

| DRUG NAME | DRUG CODE | NUMBER OF REGISTRANT SOLD TO | TOTAL GRAMS | AVG GRAMS |
|---|---|---|---|---|
| BUPRENORPHINE | 9064 | 8 | **172.51** | 21.56 |
| METHADONE | 9250B | 16 | **114,319.97** | 7,145.00 |

**STATE: NY - NEW YORK**

**BUSINESS ACTIVITY: A - PHARMACIES**

| DRUG NAME | DRUG CODE | NUMBER OF REGISTRANT SOLD TO | TOTAL GRAMS | AVG GRAMS |
|---|---|---|---|---|
| AMPHETAMINE | 1100 | 4,290 | **1,058,792.42** | 246.80 |
| METHAMPHETAMINE | 1105 | 92 | **333.06** | 3.62 |
| LISDEXAMFETAMINE | 1205 | 3,393 | **344,404.81** | 101.50 |
| METHYLPHENIDATE (DL;D;L;ISOMERS) | 1724 | 4,118 | **825,841.53** | 200.54 |
| CODEINE | 9050 | 4,454 | **374,926.42** | 84.18 |
| BUPRENORPHINE | 9064 | 3,842 | **305,777.13** | 79.59 |
| OXYCODONE | 9143 | 4,682 | **1,837,158.34** | 392.39 |
| HYDROMORPHONE | 9150 | 3,430 | **42,124.89** | 12.28 |
| HYDROCODONE | 9193 | 4,186 | **444,642.74** | 106.22 |
| MEPERIDINE (PETHIDINE) | 9230 | 48 | **734.67** | 15.31 |
| METHADONE | 9250B | 2,741 | **90,070.02** | 32.86 |
| MORPHINE | 9300 | 3,573 | **380,573.62** | 106.51 |
| OXYMORPHONE | 9652 | 983 | **18,973.23** | 19.30 |
| TAPENTADOL | 9780 | 1,931 | **223,441.00** | 115.71 |
| FENTANYL BASE | 9801 | 3,324 | **7,353.72** | 2.21 |

**BUSINESS ACTIVITY: B - HOSPITALS**

| DRUG NAME | DRUG CODE | NUMBER OF REGISTRANT SOLD TO | TOTAL GRAMS | AVG GRAMS |
|---|---|---|---|---|
| AMPHETAMINE | 1100 | 183 | **10,504.97** | 57.40 |
| LISDEXAMFETAMINE | 1205 | 87 | **1,800.75** | 20.70 |
| METHYLPHENIDATE (DL;D;L;ISOMERS) | 1724 | 238 | **9,528.20** | 40.03 |
| CODEINE | 9050 | 237 | **6,960.28** | 29.37 |
| BUPRENORPHINE | 9064 | 254 | **6,060.56** | 23.86 |
| OXYCODONE | 9143 | 317 | **57,297.49** | 180.75 |
| HYDROMORPHONE | 9150 | 281 | **18,549.70** | 66.01 |
| HYDROCODONE | 9193 | 237 | **15,476.73** | 65.30 |
| MEPERIDINE (PETHIDINE) | 9230 | 142 | **1,561.07** | 10.99 |
| METHADONE | 9250B | 252 | **20,712.57** | 82.19 |
| MORPHINE | 9300 | 312 | **40,910.86** | 131.12 |
| OXYMORPHONE | 9652 | 22 | **171.37** | 7.79 |
| TAPENTADOL | 9780 | 73 | **4,123.00** | 56.48 |
| FENTANYL BASE | 9801 | 280 | **2,486.12** | 8.88 |

**BUSINESS ACTIVITY: C - PRACTITIONERS**

| DRUG NAME | DRUG CODE | NUMBER OF REGISTRANT SOLD TO | TOTAL GRAMS | AVG GRAMS |
|---|---|---|---|---|
| METHYLPHENIDATE (DL;D;L;ISOMERS) | 1724 | 1 | **1.73** | 1.73 |
| CODEINE | 9050 | 91 | **2,942.79** | 32.34 |
| BUPRENORPHINE | 9064 | 1,149 | **353.51** | 0.31 |
| OXYCODONE | 9143 | 101 | **359.72** | 3.56 |
| HYDROMORPHONE | 9150 | 605 | **3,289.13** | 5.44 |
| HYDROCODONE | 9193 | 335 | **1,389.57** | 4.15 |
| MEPERIDINE (PETHIDINE) | 9230 | 93 | **595.56** | 6.40 |
| METHADONE | 9250B | 54 | **116.81** | 2.16 |
| MORPHINE | 9300 | 303 | **534.06** | 1.76 |

| | | | | |
|---|---|---|---|---|
| FENTANYL BASE | 9801 | 1,017 | **116.78** | 0.11 |

**BUSINESS ACTIVITY: D - TEACHING INSTITUTIONS**

| DRUG NAME | DRUG CODE | NUMBER OF REGISTRANT SOLD TO | TOTAL GRAMS | AVG GRAMS |
|---|---|---|---|---|
| AMPHETAMINE | 1100 | 1 | **3.67** | 3.67 |
| BUPRENORPHINE | 9064 | 7 | **0.07** | 0.01 |
| FENTANYL BASE | 9801 | 1 | **0.00** | 0.00 |

**BUSINESS ACTIVITY: M - MID-LEVEL PRACTITIONERS**

| DRUG NAME | DRUG CODE | NUMBER OF REGISTRANT SOLD TO | TOTAL GRAMS | AVG GRAMS |
|---|---|---|---|---|
| CODEINE | 9050 | 2 | **5.30** | 2.65 |
| BUPRENORPHINE | 9064 | 13 | **509.07** | 39.16 |
| MEPERIDINE (PETHIDINE) | 9230 | 1 | **2.18** | 2.18 |
| FENTANYL BASE | 9801 | 2 | **0.44** | 0.22 |

**BUSINESS ACTIVITY: NTP - NARCOTIC TREATMENT PROGRAMS**

| DRUG NAME | DRUG CODE | NUMBER OF REGISTRANT SOLD TO | TOTAL GRAMS | AVG GRAMS |
|---|---|---|---|---|
| BUPRENORPHINE | 9064 | 53 | **2,784.82** | 52.54 |
| METHADONE | 9250B | 122 | **1,234,741.29** | 10,120.83 |

**STATE: OH - OHIO**

**BUSINESS ACTIVITY: A - PHARMACIES**

| DRUG NAME | DRUG CODE | NUMBER OF REGISTRANT SOLD TO | TOTAL GRAMS | AVG GRAMS |
|---|---|---|---|---|
| AMPHETAMINE | 1100 | 2,177 | **876,793.54** | 402.75 |
| METHAMPHETAMINE | 1105 | 41 | **119.32** | 2.91 |
| LISDEXAMFETAMINE | 1205 | 2,118 | **361,502.12** | 170.68 |
| METHYLPHENIDATE (DL;D;L;ISOMERS) | 1724 | 2,167 | **735,151.78** | 339.25 |
| CODEINE | 9050 | 2,099 | **214,058.31** | 101.98 |
| BUPRENORPHINE | 9064 | 2,103 | **225,605.50** | 107.28 |
| OXYCODONE | 9143 | 2,170 | **1,145,214.54** | 527.75 |
| HYDROMORPHONE | 9150 | 1,789 | **17,155.72** | 9.59 |
| HYDROCODONE | 9193 | 2,180 | **458,335.46** | 210.25 |
| MEPERIDINE (PETHIDINE) | 9230 | 87 | **761.91** | 8.76 |
| METHADONE | 9250B | 1,756 | **47,761.41** | 27.20 |
| MORPHINE | 9300 | 2,064 | **286,111.61** | 138.62 |
| OXYMORPHONE | 9652 | 574 | **7,614.65** | 13.27 |
| TAPENTADOL | 9780 | 1,274 | **123,328.00** | 96.80 |
| FENTANYL BASE | 9801 | 1,877 | **3,735.99** | 1.99 |

**BUSINESS ACTIVITY: B - HOSPITALS**

| DRUG NAME | DRUG CODE | NUMBER OF REGISTRANT SOLD TO | TOTAL GRAMS | AVG GRAMS |
|---|---|---|---|---|
| AMPHETAMINE | 1100 | 168 | **16,704.67** | 99.43 |
| METHAMPHETAMINE | 1105 | 1 | **2.81** | 2.81 |
| LISDEXAMFETAMINE | 1205 | 71 | **4,014.33** | 56.54 |
| METHYLPHENIDATE (DL;D;L;ISOMERS) | 1724 | 220 | **14,187.12** | 64.49 |
| CODEINE | 9050 | 226 | **4,360.75** | 19.30 |
| BUPRENORPHINE | 9064 | 282 | **11,372.90** | 40.33 |
| OXYCODONE | 9143 | 383 | **66,277.83** | 173.05 |
| HYDROMORPHONE | 9150 | 362 | **8,247.10** | 22.78 |
| HYDROCODONE | 9193 | 374 | **15,811.55** | 42.28 |
| MEPERIDINE (PETHIDINE) | 9230 | 222 | **3,823.49** | 17.22 |
| METHADONE | 9250B | 258 | **10,793.34** | 41.83 |
| MORPHINE | 9300 | 391 | **26,263.71** | 67.17 |
| OXYMORPHONE | 9652 | 30 | **90.54** | 3.02 |
| TAPENTADOL | 9780 | 62 | **2,230.00** | 35.97 |
| FENTANYL BASE | 9801 | 454 | **1,156.91** | 2.55 |

**BUSINESS ACTIVITY: C - PRACTITIONERS**

| DRUG NAME | DRUG CODE | NUMBER OF REGISTRANT SOLD TO | TOTAL GRAMS | AVG GRAMS |
|---|---|---|---|---|
| CODEINE | 9050 | 17 | **1,080.00** | 63.53 |
| BUPRENORPHINE | 9064 | 965 | **307.41** | 0.32 |
| OXYCODONE | 9143 | 15 | **66.07** | 4.40 |
| HYDROMORPHONE | 9150 | 337 | **2,255.16** | 6.69 |
| HYDROCODONE | 9193 | 258 | **683.26** | 2.65 |
| MEPERIDINE (PETHIDINE) | 9230 | 33 | **211.45** | 6.41 |
| METHADONE | 9250B | 12 | **21.73** | 1.81 |
| MORPHINE | 9300 | 134 | **176.17** | 1.31 |

| | | | | |
|---|---|---|---|---|
| FENTANYL BASE | 9801 | 363 | **24.19** | 0.07 |

**BUSINESS ACTIVITY: D - TEACHING INSTITUTIONS**

| DRUG NAME | DRUG CODE | NUMBER OF REGISTRANT SOLD TO | TOTAL GRAMS | AVG GRAMS |
|---|---|---|---|---|
| BUPRENORPHINE | 9064 | 2 | **0.04** | 0.02 |
| HYDROMORPHONE | 9150 | 2 | **2.13** | 1.06 |
| MORPHINE | 9300 | 1 | **0.11** | 0.11 |
| FENTANYL BASE | 9801 | 1 | **0.14** | 0.14 |

**BUSINESS ACTIVITY: M - MID-LEVEL PRACTITIONERS**

| DRUG NAME | DRUG CODE | NUMBER OF REGISTRANT SOLD TO | TOTAL GRAMS | AVG GRAMS |
|---|---|---|---|---|
| BUPRENORPHINE | 9064 | 10 | **35.31** | 3.53 |
| HYDROMORPHONE | 9150 | 4 | **0.43** | 0.11 |
| MORPHINE | 9300 | 48 | **9.33** | 0.19 |
| FENTANYL BASE | 9801 | 101 | **0.87** | 0.01 |

**BUSINESS ACTIVITY: NTP - NARCOTIC TREATMENT PROGRAMS**

| DRUG NAME | DRUG CODE | NUMBER OF REGISTRANT SOLD TO | TOTAL GRAMS | AVG GRAMS |
|---|---|---|---|---|
| BUPRENORPHINE | 9064 | 68 | **28,764.60** | 423.01 |
| METHADONE | 9250B | 56 | **316,441.40** | 5,650.74 |

**STATE: OK - OKLAHOMA**

**BUSINESS ACTIVITY: A - PHARMACIES**

| DRUG NAME | DRUG CODE | NUMBER OF REGISTRANT SOLD TO | TOTAL GRAMS | AVG GRAMS |
|---|---|---|---|---|
| AMPHETAMINE | 1100 | 855 | **229,699.45** | 268.65 |
| METHAMPHETAMINE | 1105 | 13 | **42.98** | 3.31 |
| LISDEXAMFETAMINE | 1205 | 828 | **142,329.95** | 171.90 |
| METHYLPHENIDATE (DL;D;L;ISOMERS) | 1724 | 847 | **169,593.47** | 200.23 |
| CODEINE | 9050 | 870 | **267,819.30** | 307.84 |
| BUPRENORPHINE | 9064 | 852 | **74,540.84** | 87.49 |
| OXYCODONE | 9143 | 865 | **510,804.75** | 590.53 |
| HYDROMORPHONE | 9150 | 728 | **8,547.84** | 11.74 |
| HYDROCODONE | 9193 | 873 | **466,113.20** | 533.92 |
| MEPERIDINE (PETHIDINE) | 9230 | 99 | **1,190.25** | 12.02 |
| METHADONE | 9250B | 608 | **12,963.79** | 21.32 |
| MORPHINE | 9300 | 839 | **164,298.65** | 195.83 |
| OXYMORPHONE | 9652 | 219 | **2,277.89** | 10.40 |
| TAPENTADOL | 9780 | 391 | **27,918.00** | 71.40 |
| FENTANYL BASE | 9801 | 785 | **1,984.05** | 2.53 |

**BUSINESS ACTIVITY: B - HOSPITALS**

| DRUG NAME | DRUG CODE | NUMBER OF REGISTRANT SOLD TO | TOTAL GRAMS | AVG GRAMS |
|---|---|---|---|---|
| AMPHETAMINE | 1100 | 70 | **7,218.58** | 103.12 |
| METHAMPHETAMINE | 1105 | 1 | **0.40** | 0.40 |
| LISDEXAMFETAMINE | 1205 | 40 | **3,655.90** | 91.40 |
| METHYLPHENIDATE (DL;D;L;ISOMERS) | 1724 | 87 | **7,817.40** | 89.86 |
| CODEINE | 9050 | 142 | **11,542.58** | 81.29 |
| BUPRENORPHINE | 9064 | 94 | **2,967.75** | 31.57 |
| OXYCODONE | 9143 | 193 | **20,569.30** | 106.58 |
| HYDROMORPHONE | 9150 | 177 | **1,789.92** | 10.11 |
| HYDROCODONE | 9193 | 205 | **25,044.10** | 122.17 |
| MEPERIDINE (PETHIDINE) | 9230 | 119 | **1,569.24** | 13.19 |
| METHADONE | 9250B | 75 | **1,037.68** | 13.84 |
| MORPHINE | 9300 | 196 | **14,795.44** | 75.49 |
| OXYMORPHONE | 9652 | 10 | **41.75** | 4.18 |
| TAPENTADOL | 9780 | 22 | **288.00** | 13.09 |
| FENTANYL BASE | 9801 | 198 | **373.68** | 1.89 |

**BUSINESS ACTIVITY: C - PRACTITIONERS**

| DRUG NAME | DRUG CODE | NUMBER OF REGISTRANT SOLD TO | TOTAL GRAMS | AVG GRAMS |
|---|---|---|---|---|
| CODEINE | 9050 | 22 | **486.89** | 22.13 |
| BUPRENORPHINE | 9064 | 269 | **210.43** | 0.78 |
| OXYCODONE | 9143 | 8 | **119.73** | 14.97 |
| HYDROMORPHONE | 9150 | 80 | **635.16** | 7.94 |
| HYDROCODONE | 9193 | 50 | **399.66** | 7.99 |
| MEPERIDINE (PETHIDINE) | 9230 | 27 | **173.86** | 6.44 |
| METHADONE | 9250B | 2 | **4.83** | 2.41 |

| | | | | |
|---|---|---|---|---|
| MORPHINE | 9300 | 41 | **373.64** | 9.11 |
| FENTANYL BASE | 9801 | 158 | **14.56** | 0.09 |

**BUSINESS ACTIVITY: D - TEACHING INSTITUTIONS**

| DRUG NAME | DRUG CODE | NUMBER OF REGISTRANT SOLD TO | TOTAL GRAMS | AVG GRAMS |
|---|---|---|---|---|
| BUPRENORPHINE | 9064 | 1 | **0.22** | 0.22 |
| HYDROMORPHONE | 9150 | 1 | **64.54** | 64.54 |
| HYDROCODONE | 9193 | 1 | **3.60** | 3.60 |
| METHADONE | 9250B | 1 | **10.02** | 10.02 |
| MORPHINE | 9300 | 1 | **84.34** | 84.34 |
| FENTANYL BASE | 9801 | 1 | **1.32** | 1.32 |

**BUSINESS ACTIVITY: M - MID-LEVEL PRACTITIONERS**

| DRUG NAME | DRUG CODE | NUMBER OF REGISTRANT SOLD TO | TOTAL GRAMS | AVG GRAMS |
|---|---|---|---|---|
| HYDROMORPHONE | 9150 | 7 | **0.51** | 0.07 |
| MORPHINE | 9300 | 39 | **14.16** | 0.36 |
| FENTANYL BASE | 9801 | 52 | **1.44** | 0.03 |

**BUSINESS ACTIVITY: NTP - NARCOTIC TREATMENT PROGRAMS**

| DRUG NAME | DRUG CODE | NUMBER OF REGISTRANT SOLD TO | TOTAL GRAMS | AVG GRAMS |
|---|---|---|---|---|
| BUPRENORPHINE | 9064 | 14 | **1,734.75** | 123.91 |
| METHADONE | 9250B | 21 | **129,825.74** | 6,182.18 |

**STATE: OR - OREGON**

**BUSINESS ACTIVITY: A - PHARMACIES**

| DRUG NAME | DRUG CODE | NUMBER OF REGISTRANT SOLD TO | TOTAL GRAMS | AVG GRAMS |
|---|---|---|---|---|
| AMPHETAMINE | 1100 | 644 | **289,656.23** | 449.78 |
| METHAMPHETAMINE | 1105 | 32 | **163.51** | 5.11 |
| LISDEXAMFETAMINE | 1205 | 604 | **64,895.19** | 107.44 |
| METHYLPHENIDATE (DL;D;L;ISOMERS) | 1724 | 644 | **245,371.73** | 381.01 |
| CODEINE | 9050 | 631 | **89,223.80** | 141.40 |
| BUPRENORPHINE | 9064 | 648 | **64,309.09** | 99.24 |
| OXYCODONE | 9143 | 652 | **449,655.62** | 689.66 |
| HYDROMORPHONE | 9150 | 612 | **11,486.05** | 18.77 |
| HYDROCODONE | 9193 | 654 | **256,162.93** | 391.69 |
| MEPERIDINE (PETHIDINE) | 9230 | 60 | **690.12** | 11.50 |
| METHADONE | 9250B | 591 | **32,532.28** | 55.05 |
| MORPHINE | 9300 | 639 | **177,566.21** | 277.88 |
| OXYMORPHONE | 9652 | 183 | **1,878.05** | 10.26 |
| TAPENTADOL | 9780 | 223 | **16,062.50** | 72.03 |
| FENTANYL BASE | 9801 | 590 | **1,660.20** | 2.81 |

**BUSINESS ACTIVITY: B - HOSPITALS**

| DRUG NAME | DRUG CODE | NUMBER OF REGISTRANT SOLD TO | TOTAL GRAMS | AVG GRAMS |
|---|---|---|---|---|
| AMPHETAMINE | 1100 | 52 | **4,327.85** | 83.23 |
| LISDEXAMFETAMINE | 1205 | 20 | **571.74** | 28.59 |
| METHYLPHENIDATE (DL;D;L;ISOMERS) | 1724 | 61 | **4,518.30** | 74.07 |
| CODEINE | 9050 | 81 | **2,279.14** | 28.14 |
| BUPRENORPHINE | 9064 | 93 | **2,038.96** | 21.92 |
| OXYCODONE | 9143 | 92 | **25,165.66** | 273.54 |
| HYDROMORPHONE | 9150 | 80 | **2,427.18** | 30.34 |
| HYDROCODONE | 9193 | 98 | **12,954.97** | 132.19 |
| MEPERIDINE (PETHIDINE) | 9230 | 51 | **357.32** | 7.01 |
| METHADONE | 9250B | 69 | **2,514.01** | 36.43 |
| MORPHINE | 9300 | 93 | **12,347.32** | 132.77 |
| OXYMORPHONE | 9652 | 1 | **0.45** | 0.45 |
| TAPENTADOL | 9780 | 3 | **20.00** | 6.67 |
| FENTANYL BASE | 9801 | 79 | **248.85** | 3.15 |

**BUSINESS ACTIVITY: C - PRACTITIONERS**

| DRUG NAME | DRUG CODE | NUMBER OF REGISTRANT SOLD TO | TOTAL GRAMS | AVG GRAMS |
|---|---|---|---|---|
| CODEINE | 9050 | 75 | **4,832.22** | 64.43 |
| BUPRENORPHINE | 9064 | 614 | **314.18** | 0.51 |
| OXYCODONE | 9143 | 54 | **180.42** | 3.34 |
| HYDROMORPHONE | 9150 | 357 | **1,708.36** | 4.79 |
| HYDROCODONE | 9193 | 264 | **643.30** | 2.44 |
| MEPERIDINE (PETHIDINE) | 9230 | 55 | **227.24** | 4.13 |

| | | | | |
|---|---|---|---|---|
| METHADONE | 9250B | 13 | **28.62** | 2.20 |
| MORPHINE | 9300 | 152 | **138.38** | 0.91 |
| FENTANYL BASE | 9801 | 336 | **29.19** | 0.09 |

**BUSINESS ACTIVITY: D - TEACHING INSTITUTIONS**

| DRUG NAME | DRUG CODE | NUMBER OF REGISTRANT SOLD TO | TOTAL GRAMS | AVG GRAMS |
|---|---|---|---|---|
| BUPRENORPHINE | 9064 | 1 | **0.02** | 0.02 |
| FENTANYL BASE | 9801 | 1 | **0.01** | 0.01 |

**BUSINESS ACTIVITY: M - MID-LEVEL PRACTITIONERS**

| DRUG NAME | DRUG CODE | NUMBER OF REGISTRANT SOLD TO | TOTAL GRAMS | AVG GRAMS |
|---|---|---|---|---|
| AMPHETAMINE | 1100 | 1 | **1.88** | 1.88 |
| CODEINE | 9050 | 3 | **46.41** | 15.47 |
| BUPRENORPHINE | 9064 | 8 | **283.71** | 35.46 |
| HYDROMORPHONE | 9150 | 2 | **0.09** | 0.04 |
| HYDROCODONE | 9193 | 4 | **4.54** | 1.14 |
| FENTANYL BASE | 9801 | 1 | **0.00** | 0.00 |

**BUSINESS ACTIVITY: NTP - NARCOTIC TREATMENT PROGRAMS**

| DRUG NAME | DRUG CODE | NUMBER OF REGISTRANT SOLD TO | TOTAL GRAMS | AVG GRAMS |
|---|---|---|---|---|
| BUPRENORPHINE | 9064 | 19 | **4,262.52** | 224.34 |
| METHADONE | 9250B | 22 | **213,840.75** | 9,720.03 |

**STATE: PA - PENNSYLVANIA**

**BUSINESS ACTIVITY: A - PHARMACIES**

| DRUG NAME | DRUG CODE | NUMBER OF REGISTRANT SOLD TO | TOTAL GRAMS | AVG GRAMS |
|---|---|---|---|---|
| AMPHETAMINE | 1100 | 2,828 | **906,978.59** | 320.71 |
| METHAMPHETAMINE | 1105 | 65 | **367.23** | 5.65 |
| LISDEXAMFETAMINE | 1205 | 2,578 | **357,909.23** | 138.83 |
| METHYLPHENIDATE (DL;D;L;ISOMERS) | 1724 | 2,794 | **834,801.28** | 298.78 |
| CODEINE | 9050 | 2,830 | **312,391.47** | 110.39 |
| BUPRENORPHINE | 9064 | 2,755 | **355,244.50** | 128.95 |
| OXYCODONE | 9143 | 2,892 | **1,736,692.57** | 600.52 |
| HYDROMORPHONE | 9150 | 2,356 | **31,050.25** | 13.18 |
| HYDROCODONE | 9193 | 2,844 | **388,784.31** | 136.70 |
| MEPERIDINE (PETHIDINE) | 9230 | 83 | **1,282.52** | 15.45 |
| METHADONE | 9250B | 1,972 | **62,729.10** | 31.81 |
| MORPHINE | 9300 | 2,742 | **536,211.67** | 195.55 |
| OXYMORPHONE | 9652 | 952 | **20,570.09** | 21.61 |
| TAPENTADOL | 9780 | 1,226 | **154,944.50** | 126.38 |
| FENTANYL BASE | 9801 | 2,515 | **8,247.58** | 3.28 |

**BUSINESS ACTIVITY: B - HOSPITALS**

| DRUG NAME | DRUG CODE | NUMBER OF REGISTRANT SOLD TO | TOTAL GRAMS | AVG GRAMS |
|---|---|---|---|---|
| AMPHETAMINE | 1100 | 177 | **13,207.28** | 74.62 |
| LISDEXAMFETAMINE | 1205 | 64 | **3,987.16** | 62.30 |
| METHYLPHENIDATE (DL;D;L;ISOMERS) | 1724 | 230 | **11,724.53** | 50.98 |
| CODEINE | 9050 | 220 | **7,408.84** | 33.68 |
| BUPRENORPHINE | 9064 | 245 | **10,991.12** | 44.86 |
| OXYCODONE | 9143 | 313 | **84,761.71** | 270.80 |
| HYDROMORPHONE | 9150 | 289 | **9,793.50** | 33.89 |
| HYDROCODONE | 9193 | 273 | **17,452.33** | 63.93 |
| MEPERIDINE (PETHIDINE) | 9230 | 139 | **984.90** | 7.09 |
| METHADONE | 9250B | 235 | **12,885.20** | 54.83 |
| MORPHINE | 9300 | 307 | **40,335.11** | 131.38 |
| OXYMORPHONE | 9652 | 45 | **268.92** | 5.98 |
| TAPENTADOL | 9780 | 47 | **910.50** | 19.37 |
| FENTANYL BASE | 9801 | 343 | **1,333.74** | 3.89 |

**BUSINESS ACTIVITY: C - PRACTITIONERS**

| DRUG NAME | DRUG CODE | NUMBER OF REGISTRANT SOLD TO | TOTAL GRAMS | AVG GRAMS |
|---|---|---|---|---|
| CODEINE | 9050 | 95 | **2,492.08** | 26.23 |
| BUPRENORPHINE | 9064 | 1,088 | **1,355.87** | 1.25 |
| OXYCODONE | 9143 | 45 | **97.15** | 2.16 |
| HYDROMORPHONE | 9150 | 428 | **3,039.17** | 7.10 |
| HYDROCODONE | 9193 | 354 | **1,054.62** | 2.98 |
| MEPERIDINE (PETHIDINE) | 9230 | 49 | **188.03** | 3.84 |

| | | | | |
|---|---|---|---|---|
| METHADONE | 9250B | 48 | **147.75** | 3.08 |
| MORPHINE | 9300 | 152 | **91.28** | 0.60 |
| FENTANYL BASE | 9801 | 508 | **45.09** | 0.09 |

**BUSINESS ACTIVITY: D - TEACHING INSTITUTIONS**

| DRUG NAME | DRUG CODE | NUMBER OF REGISTRANT SOLD TO | TOTAL GRAMS | AVG GRAMS |
|---|---|---|---|---|
| CODEINE | 9050 | 1 | **340.89** | 340.89 |
| BUPRENORPHINE | 9064 | 3 | **0.92** | 0.31 |
| HYDROMORPHONE | 9150 | 1 | **12.94** | 12.94 |
| HYDROCODONE | 9193 | 2 | **83.08** | 41.54 |
| METHADONE | 9250B | 1 | **29.69** | 29.69 |
| MORPHINE | 9300 | 2 | **61.93** | 30.96 |
| FENTANYL BASE | 9801 | 2 | **1.19** | 0.60 |

**BUSINESS ACTIVITY: M - MID-LEVEL PRACTITIONERS**

| DRUG NAME | DRUG CODE | NUMBER OF REGISTRANT SOLD TO | TOTAL GRAMS | AVG GRAMS |
|---|---|---|---|---|
| CODEINE | 9050 | 1 | **2.21** | 2.21 |
| BUPRENORPHINE | 9064 | 1 | **6.00** | 6.00 |

**BUSINESS ACTIVITY: NTP - NARCOTIC TREATMENT PROGRAMS**

| DRUG NAME | DRUG CODE | NUMBER OF REGISTRANT SOLD TO | TOTAL GRAMS | AVG GRAMS |
|---|---|---|---|---|
| BUPRENORPHINE | 9064 | 25 | **1,402.13** | 56.09 |
| METHADONE | 9250B | 90 | **737,073.70** | 8,189.71 |

**STATE: PR - PUERTO RICO**

**BUSINESS ACTIVITY: A - PHARMACIES**

| DRUG NAME | DRUG CODE | NUMBER OF REGISTRANT SOLD TO | TOTAL GRAMS | AVG GRAMS |
|---|---|---|---|---|
| AMPHETAMINE | 1100 | 608 | **31,810.85** | 52.32 |
| LISDEXAMFETAMINE | 1205 | 239 | **7,763.88** | 32.48 |
| METHYLPHENIDATE (DL;D;L;ISOMERS) | 1724 | 631 | **40,992.86** | 64.96 |
| CODEINE | 9050 | 947 | **32,102.81** | 33.90 |
| BUPRENORPHINE | 9064 | 241 | **7,946.45** | 32.97 |
| OXYCODONE | 9143 | 760 | **38,747.67** | 50.98 |
| HYDROMORPHONE | 9150 | 13 | **75.89** | 5.84 |
| HYDROCODONE | 9193 | 335 | **1,062.62** | 3.17 |
| MEPERIDINE (PETHIDINE) | 9230 | 531 | **2,707.64** | 5.10 |
| METHADONE | 9250B | 10 | **113.59** | 11.36 |
| MORPHINE | 9300 | 122 | **3,303.33** | 27.08 |
| TAPENTADOL | 9780 | 14 | **595.00** | 42.50 |
| FENTANYL BASE | 9801 | 707 | **513.09** | 0.73 |

**BUSINESS ACTIVITY: B - HOSPITALS**

| DRUG NAME | DRUG CODE | NUMBER OF REGISTRANT SOLD TO | TOTAL GRAMS | AVG GRAMS |
|---|---|---|---|---|
| AMPHETAMINE | 1100 | 9 | **182.39** | 20.27 |
| LISDEXAMFETAMINE | 1205 | 2 | **5.20** | 2.60 |
| METHYLPHENIDATE (DL;D;L;ISOMERS) | 1724 | 14 | **220.71** | 15.77 |
| CODEINE | 9050 | 63 | **1,673.05** | 26.56 |
| BUPRENORPHINE | 9064 | 20 | **887.96** | 44.40 |
| OXYCODONE | 9143 | 68 | **3,882.92** | 57.10 |
| HYDROMORPHONE | 9150 | 5 | **71.50** | 14.30 |
| HYDROCODONE | 9193 | 25 | **96.27** | 3.85 |
| MEPERIDINE (PETHIDINE) | 9230 | 89 | **6,105.73** | 68.60 |
| METHADONE | 9250B | 5 | **67.53** | 13.51 |
| MORPHINE | 9300 | 90 | **2,337.14** | 25.97 |
| TAPENTADOL | 9780 | 4 | **648.00** | 162.00 |
| FENTANYL BASE | 9801 | 80 | **121.48** | 1.52 |

**BUSINESS ACTIVITY: C - PRACTITIONERS**

| DRUG NAME | DRUG CODE | NUMBER OF REGISTRANT SOLD TO | TOTAL GRAMS | AVG GRAMS |
|---|---|---|---|---|
| BUPRENORPHINE | 9064 | 19 | **2.03** | 0.11 |
| MEPERIDINE (PETHIDINE) | 9230 | 4 | **35.40** | 8.85 |
| MORPHINE | 9300 | 2 | **2.33** | 1.17 |
| FENTANYL BASE | 9801 | 7 | **1.73** | 0.25 |

**BUSINESS ACTIVITY: NTP - NARCOTIC TREATMENT PROGRAMS**

| DRUG NAME | DRUG CODE | NUMBER OF REGISTRANT SOLD TO | TOTAL GRAMS | AVG GRAMS |
|---|---|---|---|---|
| METHADONE | 9250B | 5 | **89,010.69** | 17,802.14 |

**STATE: RI - RHODE ISLAND**

**BUSINESS ACTIVITY: A - PHARMACIES**

| DRUG NAME | DRUG CODE | NUMBER OF REGISTRANT SOLD TO | TOTAL GRAMS | AVG GRAMS |
|---|---|---|---|---|
| AMPHETAMINE | 1100 | 171 | **158,464.11** | 926.69 |
| METHAMPHETAMINE | 1105 | 7 | **17.68** | 2.53 |
| LISDEXAMFETAMINE | 1205 | 158 | **30,116.70** | 190.61 |
| METHYLPHENIDATE (DL;D;L;ISOMERS) | 1724 | 165 | **66,349.44** | 402.12 |
| CODEINE | 9050 | 160 | **15,382.37** | 96.14 |
| BUPRENORPHINE | 9064 | 172 | **35,335.75** | 205.44 |
| OXYCODONE | 9143 | 170 | **102,220.09** | 601.29 |
| HYDROMORPHONE | 9150 | 146 | **2,544.20** | 17.43 |
| HYDROCODONE | 9193 | 169 | **26,290.20** | 155.56 |
| MEPERIDINE (PETHIDINE) | 9230 | 4 | **46.84** | 11.71 |
| METHADONE | 9250B | 141 | **4,652.22** | 32.99 |
| MORPHINE | 9300 | 161 | **27,815.24** | 172.77 |
| OXYMORPHONE | 9652 | 39 | **477.10** | 12.23 |
| TAPENTADOL | 9780 | 70 | **6,148.50** | 87.84 |
| FENTANYL BASE | 9801 | 142 | **324.81** | 2.29 |

**BUSINESS ACTIVITY: B - HOSPITALS**

| DRUG NAME | DRUG CODE | NUMBER OF REGISTRANT SOLD TO | TOTAL GRAMS | AVG GRAMS |
|---|---|---|---|---|
| AMPHETAMINE | 1100 | 18 | **2,803.13** | 155.73 |
| LISDEXAMFETAMINE | 1205 | 8 | **401.20** | 50.15 |
| METHYLPHENIDATE (DL;D;L;ISOMERS) | 1724 | 18 | **2,235.37** | 124.19 |
| CODEINE | 9050 | 17 | **362.86** | 21.34 |
| BUPRENORPHINE | 9064 | 16 | **953.57** | 59.60 |
| OXYCODONE | 9143 | 22 | **7,001.08** | 318.23 |
| HYDROMORPHONE | 9150 | 17 | **658.73** | 38.75 |
| HYDROCODONE | 9193 | 19 | **1,410.65** | 74.24 |
| MEPERIDINE (PETHIDINE) | 9230 | 10 | **107.30** | 10.73 |
| METHADONE | 9250B | 15 | **1,644.00** | 109.60 |
| MORPHINE | 9300 | 19 | **3,969.17** | 208.90 |
| OXYMORPHONE | 9652 | 1 | **1.07** | 1.07 |
| TAPENTADOL | 9780 | 1 | **10.00** | 10.00 |
| FENTANYL BASE | 9801 | 15 | **89.58** | 5.97 |

**BUSINESS ACTIVITY: C - PRACTITIONERS**

| DRUG NAME | DRUG CODE | NUMBER OF REGISTRANT SOLD TO | TOTAL GRAMS | AVG GRAMS |
|---|---|---|---|---|
| CODEINE | 9050 | 1 | **21.17** | 21.17 |
| BUPRENORPHINE | 9064 | 90 | **8.20** | 0.09 |
| OXYCODONE | 9143 | 4 | **9.14** | 2.29 |
| HYDROMORPHONE | 9150 | 38 | **358.06** | 9.42 |
| HYDROCODONE | 9193 | 20 | **79.34** | 3.97 |
| MEPERIDINE (PETHIDINE) | 9230 | 3 | **24.18** | 8.06 |
| METHADONE | 9250B | 7 | **26.12** | 3.73 |
| MORPHINE | 9300 | 10 | **5.32** | 0.53 |
| FENTANYL BASE | 9801 | 35 | **6.81** | 0.19 |

**BUSINESS ACTIVITY: D - TEACHING INSTITUTIONS**

| DRUG NAME | DRUG CODE | NUMBER OF REGISTRANT SOLD TO | TOTAL GRAMS | AVG GRAMS |
|---|---|---|---|---|
| BUPRENORPHINE | 9064 | 1 | **0.00** | 0.00 |

**BUSINESS ACTIVITY: NTP - NARCOTIC TREATMENT PROGRAMS**

| DRUG NAME | DRUG CODE | NUMBER OF REGISTRANT SOLD TO | TOTAL GRAMS | AVG GRAMS |
|---|---|---|---|---|
| BUPRENORPHINE | 9064 | 16 | **605.66** | 37.85 |
| METHADONE | 9250B | 23 | **172,395.15** | 7,495.44 |

**STATE: SC - SOUTH CAROLINA**

**BUSINESS ACTIVITY: A - PHARMACIES**

| DRUG NAME | DRUG CODE | NUMBER OF REGISTRANT SOLD TO | TOTAL GRAMS | AVG GRAMS |
|---|---|---|---|---|
| AMPHETAMINE | 1100 | 1,054 | **598,263.89** | 567.61 |
| METHAMPHETAMINE | 1105 | 12 | **24.11** | 2.01 |
| LISDEXAMFETAMINE | 1205 | 1,035 | **235,450.88** | 227.49 |
| METHYLPHENIDATE (DL;D;L;ISOMERS) | 1724 | 1,047 | **318,640.18** | 304.34 |
| CODEINE | 9050 | 1,021 | **215,890.76** | 211.45 |
| BUPRENORPHINE | 9064 | 1,017 | **68,546.39** | 67.40 |
| OXYCODONE | 9143 | 1,051 | **606,174.13** | 576.76 |

| | | | | |
|---|---|---|---|---|
| HYDROMORPHONE | 9150 | 949 | **12,898.13** | 13.59 |
| HYDROCODONE | 9193 | 1,065 | **355,301.63** | 333.62 |
| MEPERIDINE (PETHIDINE) | 9230 | 92 | **974.45** | 10.59 |
| METHADONE | 9250B | 793 | **22,074.33** | 27.84 |
| MORPHINE | 9300 | 1,030 | **178,782.34** | 173.58 |
| OXYMORPHONE | 9652 | 398 | **7,389.80** | 18.57 |
| TAPENTADOL | 9780 | 724 | **97,591.00** | 134.79 |
| FENTANYL BASE | 9801 | 961 | **2,362.31** | 2.46 |

**BUSINESS ACTIVITY: B - HOSPITALS**

| DRUG NAME | DRUG CODE | NUMBER OF REGISTRANT SOLD TO | TOTAL GRAMS | AVG GRAMS |
|---|---|---|---|---|
| AMPHETAMINE | 1100 | 62 | **9,594.54** | 154.75 |
| METHAMPHETAMINE | 1105 | 1 | **6.03** | 6.03 |
| LISDEXAMFETAMINE | 1205 | 25 | **1,422.39** | 56.90 |
| METHYLPHENIDATE (DL;D;L;ISOMERS) | 1724 | 83 | **8,132.32** | 97.98 |
| CODEINE | 9050 | 69 | **1,996.19** | 28.93 |
| BUPRENORPHINE | 9064 | 86 | **1,822.40** | 21.19 |
| OXYCODONE | 9143 | 146 | **21,342.44** | 146.18 |
| HYDROMORPHONE | 9150 | 151 | **9,088.16** | 60.19 |
| HYDROCODONE | 9193 | 144 | **9,243.43** | 64.19 |
| MEPERIDINE (PETHIDINE) | 9230 | 78 | **749.82** | 9.61 |
| METHADONE | 9250B | 90 | **1,392.55** | 15.47 |
| MORPHINE | 9300 | 151 | **14,212.92** | 94.13 |
| OXYMORPHONE | 9652 | 2 | **5.62** | 2.81 |
| TAPENTADOL | 9780 | 25 | **754.00** | 30.16 |
| FENTANYL BASE | 9801 | 178 | **436.40** | 2.45 |

**BUSINESS ACTIVITY: C - PRACTITIONERS**

| DRUG NAME | DRUG CODE | NUMBER OF REGISTRANT SOLD TO | TOTAL GRAMS | AVG GRAMS |
|---|---|---|---|---|
| CODEINE | 9050 | 10 | **636.95** | 63.70 |
| BUPRENORPHINE | 9064 | 445 | **90.82** | 0.20 |
| OXYCODONE | 9143 | 5 | **11.21** | 2.24 |
| HYDROMORPHONE | 9150 | 154 | **1,435.41** | 9.32 |
| HYDROCODONE | 9193 | 44 | **99.80** | 2.27 |
| MEPERIDINE (PETHIDINE) | 9230 | 37 | **264.17** | 7.14 |
| METHADONE | 9250B | 2 | **4.83** | 2.41 |
| MORPHINE | 9300 | 50 | **28.83** | 0.58 |
| FENTANYL BASE | 9801 | 245 | **10.76** | 0.04 |

**BUSINESS ACTIVITY: D - TEACHING INSTITUTIONS**

| DRUG NAME | DRUG CODE | NUMBER OF REGISTRANT SOLD TO | TOTAL GRAMS | AVG GRAMS |
|---|---|---|---|---|
| BUPRENORPHINE | 9064 | 1 | **0.01** | 0.01 |

**BUSINESS ACTIVITY: M - MID-LEVEL PRACTITIONERS**

| DRUG NAME | DRUG CODE | NUMBER OF REGISTRANT SOLD TO | TOTAL GRAMS | AVG GRAMS |
|---|---|---|---|---|
| MORPHINE | 9300 | 47 | **14.52** | 0.31 |
| FENTANYL BASE | 9801 | 65 | **0.98** | 0.02 |

**BUSINESS ACTIVITY: NTP - NARCOTIC TREATMENT PROGRAMS**

| DRUG NAME | DRUG CODE | NUMBER OF REGISTRANT SOLD TO | TOTAL GRAMS | AVG GRAMS |
|---|---|---|---|---|
| BUPRENORPHINE | 9064 | 17 | **2,014.95** | 118.53 |
| METHADONE | 9250B | 24 | **200,162.70** | 8,340.11 |

**STATE: SD - SOUTH DAKOTA**

**BUSINESS ACTIVITY: A - PHARMACIES**

| DRUG NAME | DRUG CODE | NUMBER OF REGISTRANT SOLD TO | TOTAL GRAMS | AVG GRAMS |
|---|---|---|---|---|
| AMPHETAMINE | 1100 | 181 | **54,979.16** | 303.75 |
| METHAMPHETAMINE | 1105 | 3 | **26.51** | 8.84 |
| LISDEXAMFETAMINE | 1205 | 179 | **34,889.49** | 194.91 |
| METHYLPHENIDATE (DL;D;L;ISOMERS) | 1724 | 181 | **66,642.67** | 368.19 |
| CODEINE | 9050 | 175 | **15,106.94** | 86.33 |
| BUPRENORPHINE | 9064 | 148 | **4,427.75** | 29.92 |
| OXYCODONE | 9143 | 182 | **35,461.20** | 194.84 |
| HYDROMORPHONE | 9150 | 163 | **1,103.68** | 6.77 |
| HYDROCODONE | 9193 | 185 | **29,721.41** | 160.66 |
| MEPERIDINE (PETHIDINE) | 9230 | 8 | **39.76** | 4.97 |
| METHADONE | 9250B | 133 | **2,777.11** | 20.88 |

| | | | | |
|---|---|---|---|---|
| MORPHINE | 9300 | 174 | **16,629.19** | 95.57 |
| OXYMORPHONE | 9652 | 37 | **567.15** | 15.33 |
| TAPENTADOL | 9780 | 111 | **9,207.50** | 82.95 |
| FENTANYL BASE | 9801 | 173 | **470.05** | 2.72 |

**BUSINESS ACTIVITY: B - HOSPITALS**

| DRUG NAME | DRUG CODE | NUMBER OF REGISTRANT SOLD TO | TOTAL GRAMS | AVG GRAMS |
|---|---|---|---|---|
| AMPHETAMINE | 1100 | 25 | **2,574.40** | 102.98 |
| METHAMPHETAMINE | 1105 | 1 | **0.40** | 0.40 |
| LISDEXAMFETAMINE | 1205 | 18 | **765.98** | 42.55 |
| METHYLPHENIDATE (DL;D;L;ISOMERS) | 1724 | 42 | **3,359.89** | 80.00 |
| CODEINE | 9050 | 44 | **1,314.15** | 29.87 |
| BUPRENORPHINE | 9064 | 26 | **610.69** | 23.49 |
| OXYCODONE | 9143 | 71 | **4,698.89** | 66.18 |
| HYDROMORPHONE | 9150 | 62 | **434.83** | 7.01 |
| HYDROCODONE | 9193 | 73 | **6,123.82** | 83.89 |
| MEPERIDINE (PETHIDINE) | 9230 | 25 | **240.76** | 9.63 |
| METHADONE | 9250B | 30 | **1,371.38** | 45.71 |
| MORPHINE | 9300 | 78 | **3,972.60** | 50.93 |
| OXYMORPHONE | 9652 | 2 | **4.55** | 2.27 |
| TAPENTADOL | 9780 | 16 | **557.00** | 34.81 |
| FENTANYL BASE | 9801 | 75 | **124.91** | 1.67 |

**BUSINESS ACTIVITY: C - PRACTITIONERS**

| DRUG NAME | DRUG CODE | NUMBER OF REGISTRANT SOLD TO | TOTAL GRAMS | AVG GRAMS |
|---|---|---|---|---|
| CODEINE | 9050 | 3 | **57.46** | 19.15 |
| BUPRENORPHINE | 9064 | 77 | **194.90** | 2.53 |
| OXYCODONE | 9143 | 2 | **5.83** | 2.91 |
| HYDROMORPHONE | 9150 | 34 | **190.49** | 5.60 |
| HYDROCODONE | 9193 | 23 | **59.15** | 2.57 |
| MEPERIDINE (PETHIDINE) | 9230 | 8 | **51.64** | 6.45 |
| METHADONE | 9250B | 1 | **0.36** | 0.36 |
| MORPHINE | 9300 | 22 | **55.01** | 2.50 |
| FENTANYL BASE | 9801 | 47 | **4.91** | 0.10 |

**BUSINESS ACTIVITY: D - TEACHING INSTITUTIONS**

| DRUG NAME | DRUG CODE | NUMBER OF REGISTRANT SOLD TO | TOTAL GRAMS | AVG GRAMS |
|---|---|---|---|---|
| BUPRENORPHINE | 9064 | 1 | **0.00** | 0.00 |

**BUSINESS ACTIVITY: M - MID-LEVEL PRACTITIONERS**

| DRUG NAME | DRUG CODE | NUMBER OF REGISTRANT SOLD TO | TOTAL GRAMS | AVG GRAMS |
|---|---|---|---|---|
| CODEINE | 9050 | 1 | **3.98** | 3.98 |
| BUPRENORPHINE | 9064 | 1 | **0.72** | 0.72 |
| HYDROMORPHONE | 9150 | 2 | **0.06** | 0.03 |
| MORPHINE | 9300 | 2 | **0.56** | 0.28 |
| FENTANYL BASE | 9801 | 4 | **0.11** | 0.03 |

**BUSINESS ACTIVITY: NTP - NARCOTIC TREATMENT PROGRAMS**

| DRUG NAME | DRUG CODE | NUMBER OF REGISTRANT SOLD TO | TOTAL GRAMS | AVG GRAMS |
|---|---|---|---|---|
| METHADONE | 9250B | 1 | **2,883.55** | 2,883.55 |

**STATE: TN - TENNESSEE**

**BUSINESS ACTIVITY: A - PHARMACIES**

| DRUG NAME | DRUG CODE | NUMBER OF REGISTRANT SOLD TO | TOTAL GRAMS | AVG GRAMS |
|---|---|---|---|---|
| AMPHETAMINE | 1100 | 1,418 | **569,140.79** | 401.37 |
| METHAMPHETAMINE | 1105 | 18 | **61.47** | 3.41 |
| LISDEXAMFETAMINE | 1205 | 1,355 | **251,867.76** | 185.88 |
| METHYLPHENIDATE (DL;D;L;ISOMERS) | 1724 | 1,402 | **308,995.41** | 220.40 |
| CODEINE | 9050 | 1,355 | **195,951.96** | 144.61 |
| BUPRENORPHINE | 9064 | 1,329 | **189,788.26** | 142.81 |
| OXYCODONE | 9143 | 1,428 | **1,208,764.28** | 846.47 |
| HYDROMORPHONE | 9150 | 1,146 | **14,404.23** | 12.57 |
| HYDROCODONE | 9193 | 1,435 | **639,893.32** | 445.92 |
| MEPERIDINE (PETHIDINE) | 9230 | 260 | **2,946.00** | 11.33 |
| METHADONE | 9250B | 861 | **21,850.10** | 25.38 |
| MORPHINE | 9300 | 1,400 | **369,982.79** | 264.27 |
| OXYMORPHONE | 9652 | 589 | **19,536.72** | 33.17 |

| | | | | |
|---|---|---|---|---|
| TAPENTADOL | 9780 | 923 | **80,874.50** | 87.62 |
| FENTANYL BASE | 9801 | 1,251 | **3,316.52** | 2.65 |

**BUSINESS ACTIVITY: B - HOSPITALS**

| DRUG NAME | DRUG CODE | NUMBER OF REGISTRANT SOLD TO | TOTAL GRAMS | AVG GRAMS |
|---|---|---|---|---|
| AMPHETAMINE | 1100 | 90 | **6,555.84** | 72.84 |
| LISDEXAMFETAMINE | 1205 | 41 | **2,767.94** | 67.51 |
| METHYLPHENIDATE (DL;D;L;ISOMERS) | 1724 | 110 | **4,451.84** | 40.47 |
| CODEINE | 9050 | 119 | **2,264.45** | 19.03 |
| BUPRENORPHINE | 9064 | 145 | **4,743.19** | 32.71 |
| OXYCODONE | 9143 | 185 | **52,020.44** | 281.19 |
| HYDROMORPHONE | 9150 | 170 | **3,275.63** | 19.27 |
| HYDROCODONE | 9193 | 190 | **30,255.73** | 159.24 |
| MEPERIDINE (PETHIDINE) | 9230 | 107 | **1,135.67** | 10.61 |
| METHADONE | 9250B | 129 | **1,725.56** | 13.38 |
| MORPHINE | 9300 | 189 | **19,520.91** | 103.29 |
| OXYMORPHONE | 9652 | 14 | **58.88** | 4.21 |
| TAPENTADOL | 9780 | 26 | **2,535.00** | 97.50 |
| FENTANYL BASE | 9801 | 171 | **634.01** | 3.71 |

**BUSINESS ACTIVITY: C - PRACTITIONERS**

| DRUG NAME | DRUG CODE | NUMBER OF REGISTRANT SOLD TO | TOTAL GRAMS | AVG GRAMS |
|---|---|---|---|---|
| AMPHETAMINE | 1100 | 7 | **815.25** | 116.46 |
| LISDEXAMFETAMINE | 1205 | 3 | **423.17** | 141.06 |
| METHYLPHENIDATE (DL;D;L;ISOMERS) | 1724 | 1 | **532.85** | 532.85 |
| CODEINE | 9050 | 63 | **2,534.63** | 40.23 |
| BUPRENORPHINE | 9064 | 608 | **108.24** | 0.18 |
| OXYCODONE | 9143 | 69 | **565.20** | 8.19 |
| HYDROMORPHONE | 9150 | 260 | **1,664.24** | 6.40 |
| HYDROCODONE | 9193 | 235 | **869.82** | 3.70 |
| MEPERIDINE (PETHIDINE) | 9230 | 97 | **508.52** | 5.24 |
| METHADONE | 9250B | 10 | **17.08** | 1.71 |
| MORPHINE | 9300 | 138 | **134.07** | 0.97 |
| FENTANYL BASE | 9801 | 429 | **38.25** | 0.09 |

**BUSINESS ACTIVITY: D - TEACHING INSTITUTIONS**

| DRUG NAME | DRUG CODE | NUMBER OF REGISTRANT SOLD TO | TOTAL GRAMS | AVG GRAMS |
|---|---|---|---|---|
| FENTANYL BASE | 9801 | 1 | **0.02** | 0.02 |

**BUSINESS ACTIVITY: M - MID-LEVEL PRACTITIONERS**

| DRUG NAME | DRUG CODE | NUMBER OF REGISTRANT SOLD TO | TOTAL GRAMS | AVG GRAMS |
|---|---|---|---|---|
| CODEINE | 9050 | 16 | **47.20** | 2.95 |
| HYDROMORPHONE | 9150 | 2 | **0.31** | 0.16 |
| MEPERIDINE (PETHIDINE) | 9230 | 3 | **5.99** | 2.00 |
| MORPHINE | 9300 | 75 | **26.89** | 0.36 |
| FENTANYL BASE | 9801 | 90 | **1.80** | 0.02 |

**BUSINESS ACTIVITY: NTP - NARCOTIC TREATMENT PROGRAMS**

| DRUG NAME | DRUG CODE | NUMBER OF REGISTRANT SOLD TO | TOTAL GRAMS | AVG GRAMS |
|---|---|---|---|---|
| BUPRENORPHINE | 9064 | 12 | **1,111.65** | 92.64 |
| METHADONE | 9250B | 16 | **169,471.81** | 10,591.99 |

**STATE: TX - TEXAS**

**BUSINESS ACTIVITY: A - PHARMACIES**

| DRUG NAME | DRUG CODE | NUMBER OF REGISTRANT SOLD TO | TOTAL GRAMS | AVG GRAMS |
|---|---|---|---|---|
| AMPHETAMINE | 1100 | 4,560 | **1,993,123.43** | 437.09 |
| METHAMPHETAMINE | 1105 | 136 | **356.75** | 2.62 |
| LISDEXAMFETAMINE | 1205 | 4,344 | **966,676.27** | 222.53 |
| METHYLPHENIDATE (DL;D;L;ISOMERS) | 1724 | 4,504 | **1,173,573.72** | 260.56 |
| CODEINE | 9050 | 4,916 | **2,877,692.50** | 585.37 |
| BUPRENORPHINE | 9064 | 4,161 | **128,821.23** | 30.96 |
| OXYCODONE | 9143 | 4,524 | **905,390.85** | 200.13 |
| HYDROMORPHONE | 9150 | 3,772 | **57,563.96** | 15.26 |
| HYDROCODONE | 9193 | 4,889 | **1,789,747.72** | 366.08 |
| MEPERIDINE (PETHIDINE) | 9230 | 285 | **4,767.65** | 16.73 |
| METHADONE | 9250B | 3,212 | **73,687.74** | 22.94 |
| MORPHINE | 9300 | 4,258 | **502,164.99** | 117.93 |

| | | | | |
|---|---|---|---|---|
| OXYMORPHONE | 9652 | 716 | **8,170.70** | 11.41 |
| TAPENTADOL | 9780 | 2,143 | **204,700.00** | 95.52 |
| FENTANYL BASE | 9801 | 3,921 | **9,725.34** | 2.48 |

**BUSINESS ACTIVITY: B - HOSPITALS**

| DRUG NAME | DRUG CODE | NUMBER OF REGISTRANT SOLD TO | TOTAL GRAMS | AVG GRAMS |
|---|---|---|---|---|
| AMPHETAMINE | 1100 | 287 | **37,299.42** | 129.96 |
| LISDEXAMFETAMINE | 1205 | 122 | **8,552.41** | 70.10 |
| METHYLPHENIDATE (DL;D;L;ISOMERS) | 1724 | 402 | **38,365.85** | 95.44 |
| CODEINE | 9050 | 871 | **70,884.12** | 81.38 |
| BUPRENORPHINE | 9064 | 337 | **4,990.85** | 14.81 |
| OXYCODONE | 9143 | 748 | **41,488.02** | 55.47 |
| HYDROMORPHONE | 9150 | 1,080 | **11,772.90** | 10.90 |
| HYDROCODONE | 9193 | 1,165 | **87,066.31** | 74.74 |
| MEPERIDINE (PETHIDINE) | 9230 | 633 | **8,657.59** | 13.68 |
| METHADONE | 9250B | 493 | **7,467.76** | 15.15 |
| MORPHINE | 9300 | 1,225 | **57,592.43** | 47.01 |
| OXYMORPHONE | 9652 | 17 | **273.70** | 16.10 |
| TAPENTADOL | 9780 | 70 | **2,646.50** | 37.81 |
| FENTANYL BASE | 9801 | 1,283 | **2,739.65** | 2.14 |

**BUSINESS ACTIVITY: C - PRACTITIONERS**

| DRUG NAME | DRUG CODE | NUMBER OF REGISTRANT SOLD TO | TOTAL GRAMS | AVG GRAMS |
|---|---|---|---|---|
| CODEINE | 9050 | 107 | **1,812.63** | 16.94 |
| BUPRENORPHINE | 9064 | 2,182 | **487.54** | 0.22 |
| OXYCODONE | 9143 | 23 | **55.99** | 2.43 |
| HYDROMORPHONE | 9150 | 710 | **4,502.62** | 6.34 |
| HYDROCODONE | 9193 | 788 | **2,015.18** | 2.56 |
| MEPERIDINE (PETHIDINE) | 9230 | 249 | **1,720.12** | 6.91 |
| METHADONE | 9250B | 41 | **62.61** | 1.53 |
| MORPHINE | 9300 | 382 | **267.55** | 0.70 |
| TAPENTADOL | 9780 | 1 | **10.00** | 10.00 |
| FENTANYL BASE | 9801 | 1,366 | **77.33** | 0.06 |

**BUSINESS ACTIVITY: D - TEACHING INSTITUTIONS**

| DRUG NAME | DRUG CODE | NUMBER OF REGISTRANT SOLD TO | TOTAL GRAMS | AVG GRAMS |
|---|---|---|---|---|
| CODEINE | 9050 | 1 | **39.78** | 39.78 |
| BUPRENORPHINE | 9064 | 1 | **0.70** | 0.70 |
| HYDROMORPHONE | 9150 | 1 | **84.40** | 84.40 |
| HYDROCODONE | 9193 | 1 | **1.78** | 1.78 |
| METHADONE | 9250B | 1 | **18.07** | 18.07 |
| MORPHINE | 9300 | 2 | **7.03** | 3.52 |
| FENTANYL BASE | 9801 | 3 | **4.29** | 1.43 |

**BUSINESS ACTIVITY: M - MID-LEVEL PRACTITIONERS**

| DRUG NAME | DRUG CODE | NUMBER OF REGISTRANT SOLD TO | TOTAL GRAMS | AVG GRAMS |
|---|---|---|---|---|
| CODEINE | 9050 | 32 | **36.02** | 1.13 |
| HYDROMORPHONE | 9150 | 12 | **1.19** | 0.10 |
| MEPERIDINE (PETHIDINE) | 9230 | 1 | **0.87** | 0.87 |
| MORPHINE | 9300 | 194 | **131.08** | 0.68 |
| FENTANYL BASE | 9801 | 359 | **13.57** | 0.04 |

**BUSINESS ACTIVITY: NTP - NARCOTIC TREATMENT PROGRAMS**

| DRUG NAME | DRUG CODE | NUMBER OF REGISTRANT SOLD TO | TOTAL GRAMS | AVG GRAMS |
|---|---|---|---|---|
| BUPRENORPHINE | 9064 | 45 | **2,387.47** | 53.05 |
| METHADONE | 9250B | 96 | **420,698.03** | 4,382.27 |

**STATE: UT - UTAH**

**BUSINESS ACTIVITY: A - PHARMACIES**

| DRUG NAME | DRUG CODE | NUMBER OF REGISTRANT SOLD TO | TOTAL GRAMS | AVG GRAMS |
|---|---|---|---|---|
| AMPHETAMINE | 1100 | 511 | **415,933.41** | 813.96 |
| METHAMPHETAMINE | 1105 | 23 | **106.85** | 4.65 |
| LISDEXAMFETAMINE | 1205 | 501 | **93,123.24** | 185.87 |
| METHYLPHENIDATE (DL;D;L;ISOMERS) | 1724 | 513 | **191,748.00** | 373.78 |
| CODEINE | 9050 | 506 | **46,845.20** | 92.58 |
| BUPRENORPHINE | 9064 | 507 | **46,959.61** | 92.62 |
| OXYCODONE | 9143 | 516 | **487,329.94** | 944.44 |

| | | | | |
|---|---|---|---|---|
| HYDROMORPHONE | 9150 | 480 | **7,605.57** | 15.84 |
| HYDROCODONE | 9193 | 514 | **163,810.40** | 318.70 |
| MEPERIDINE (PETHIDINE) | 9230 | 89 | **1,956.74** | 21.99 |
| METHADONE | 9250B | 451 | **22,190.51** | 49.20 |
| MORPHINE | 9300 | 509 | **130,832.92** | 257.04 |
| OXYMORPHONE | 9652 | 273 | **6,794.14** | 24.89 |
| TAPENTADOL | 9780 | 288 | **29,705.00** | 103.14 |
| FENTANYL BASE | 9801 | 478 | **1,807.89** | 3.78 |

**BUSINESS ACTIVITY: B - HOSPITALS**

| DRUG NAME | DRUG CODE | NUMBER OF REGISTRANT SOLD TO | TOTAL GRAMS | AVG GRAMS |
|---|---|---|---|---|
| AMPHETAMINE | 1100 | 33 | **4,165.35** | 126.22 |
| LISDEXAMFETAMINE | 1205 | 14 | **781.59** | 55.83 |
| METHYLPHENIDATE (DL;D;L;ISOMERS) | 1724 | 40 | **3,893.05** | 97.33 |
| CODEINE | 9050 | 29 | **887.72** | 30.61 |
| BUPRENORPHINE | 9064 | 50 | **1,601.78** | 32.04 |
| OXYCODONE | 9143 | 87 | **18,538.09** | 213.08 |
| HYDROMORPHONE | 9150 | 83 | **1,458.68** | 17.57 |
| HYDROCODONE | 9193 | 110 | **7,379.75** | 67.09 |
| MEPERIDINE (PETHIDINE) | 9230 | 68 | **838.38** | 12.33 |
| METHADONE | 9250B | 49 | **987.69** | 20.16 |
| MORPHINE | 9300 | 99 | **8,737.06** | 88.25 |
| OXYMORPHONE | 9652 | 11 | **33.36** | 3.03 |
| TAPENTADOL | 9780 | 14 | **2,208.50** | 157.75 |
| FENTANYL BASE | 9801 | 101 | **203.70** | 2.02 |

**BUSINESS ACTIVITY: C - PRACTITIONERS**

| DRUG NAME | DRUG CODE | NUMBER OF REGISTRANT SOLD TO | TOTAL GRAMS | AVG GRAMS |
|---|---|---|---|---|
| CODEINE | 9050 | 6 | **265.21** | 44.20 |
| BUPRENORPHINE | 9064 | 224 | **56.60** | 0.25 |
| OXYCODONE | 9143 | 8 | **18.60** | 2.33 |
| HYDROMORPHONE | 9150 | 111 | **783.56** | 7.06 |
| HYDROCODONE | 9193 | 82 | **299.13** | 3.65 |
| MEPERIDINE (PETHIDINE) | 9230 | 22 | **118.63** | 5.39 |
| METHADONE | 9250B | 10 | **21.11** | 2.11 |
| MORPHINE | 9300 | 84 | **52.48** | 0.62 |
| FENTANYL BASE | 9801 | 210 | **15.22** | 0.07 |

**BUSINESS ACTIVITY: M - MID-LEVEL PRACTITIONERS**

| DRUG NAME | DRUG CODE | NUMBER OF REGISTRANT SOLD TO | TOTAL GRAMS | AVG GRAMS |
|---|---|---|---|---|
| BUPRENORPHINE | 9064 | 1 | **52.77** | 52.77 |
| HYDROMORPHONE | 9150 | 1 | **0.02** | 0.02 |
| MEPERIDINE (PETHIDINE) | 9230 | 3 | **29.41** | 9.80 |
| MORPHINE | 9300 | 1 | **0.15** | 0.15 |
| FENTANYL BASE | 9801 | 9 | **0.52** | 0.06 |

**BUSINESS ACTIVITY: NTP - NARCOTIC TREATMENT PROGRAMS**

| DRUG NAME | DRUG CODE | NUMBER OF REGISTRANT SOLD TO | TOTAL GRAMS | AVG GRAMS |
|---|---|---|---|---|
| BUPRENORPHINE | 9064 | 8 | **318.26** | 39.78 |
| METHADONE | 9250B | 18 | **94,348.47** | 5,241.58 |

**STATE: VA - VIRGINIA**

**BUSINESS ACTIVITY: A - PHARMACIES**

| DRUG NAME | DRUG CODE | NUMBER OF REGISTRANT SOLD TO | TOTAL GRAMS | AVG GRAMS |
|---|---|---|---|---|
| AMPHETAMINE | 1100 | 1,489 | **747,375.24** | 501.93 |
| METHAMPHETAMINE | 1105 | 38 | **98.84** | 2.60 |
| LISDEXAMFETAMINE | 1205 | 1,440 | **263,421.09** | 182.93 |
| METHYLPHENIDATE (DL;D;L;ISOMERS) | 1724 | 1,477 | **461,019.90** | 312.13 |
| CODEINE | 9050 | 1,431 | **140,168.91** | 97.95 |
| BUPRENORPHINE | 9064 | 1,433 | **111,507.79** | 77.81 |
| OXYCODONE | 9143 | 1,494 | **792,193.92** | 530.25 |
| HYDROMORPHONE | 9150 | 1,392 | **23,875.19** | 17.15 |
| HYDROCODONE | 9193 | 1,498 | **267,026.87** | 178.26 |
| MEPERIDINE (PETHIDINE) | 9230 | 97 | **1,025.21** | 10.57 |
| METHADONE | 9250B | 1,163 | **38,025.06** | 32.70 |
| MORPHINE | 9300 | 1,435 | **210,339.08** | 146.58 |

| | | | | |
|---|---|---|---|---|
| OXYMORPHONE | 9652 | 604 | **10,762.12** | 17.82 |
| TAPENTADOL | 9780 | 900 | **94,498.00** | 105.00 |
| FENTANYL BASE | 9801 | 1,327 | **3,151.49** | 2.37 |

**BUSINESS ACTIVITY: B - HOSPITALS**

| DRUG NAME | DRUG CODE | NUMBER OF REGISTRANT SOLD TO | TOTAL GRAMS | AVG GRAMS |
|---|---|---|---|---|
| AMPHETAMINE | 1100 | 95 | **26,948.27** | 283.67 |
| LISDEXAMFETAMINE | 1205 | 52 | **5,391.36** | 103.68 |
| METHYLPHENIDATE (DL;D;L;ISOMERS) | 1724 | 124 | **26,264.65** | 211.81 |
| CODEINE | 9050 | 116 | **3,688.68** | 31.80 |
| BUPRENORPHINE | 9064 | 125 | **2,499.33** | 19.99 |
| OXYCODONE | 9143 | 189 | **48,033.83** | 254.15 |
| HYDROMORPHONE | 9150 | 180 | **5,587.78** | 31.04 |
| HYDROCODONE | 9193 | 177 | **11,239.39** | 63.50 |
| MEPERIDINE (PETHIDINE) | 9230 | 90 | **1,349.19** | 14.99 |
| METHADONE | 9250B | 124 | **3,182.93** | 25.67 |
| MORPHINE | 9300 | 190 | **18,021.14** | 94.85 |
| OXYMORPHONE | 9652 | 9 | **38.90** | 4.32 |
| TAPENTADOL | 9780 | 29 | **1,472.00** | 50.76 |
| FENTANYL BASE | 9801 | 197 | **711.20** | 3.61 |

**BUSINESS ACTIVITY: C - PRACTITIONERS**

| DRUG NAME | DRUG CODE | NUMBER OF REGISTRANT SOLD TO | TOTAL GRAMS | AVG GRAMS |
|---|---|---|---|---|
| CODEINE | 9050 | 63 | **2,975.72** | 47.23 |
| BUPRENORPHINE | 9064 | 904 | **101.80** | 0.11 |
| OXYCODONE | 9143 | 21 | **130.98** | 6.24 |
| HYDROMORPHONE | 9150 | 357 | **2,162.32** | 6.06 |
| HYDROCODONE | 9193 | 170 | **553.57** | 3.26 |
| MEPERIDINE (PETHIDINE) | 9230 | 75 | **528.56** | 7.05 |
| METHADONE | 9250B | 24 | **45.35** | 1.89 |
| MORPHINE | 9300 | 158 | **110.60** | 0.70 |
| OXYMORPHONE | 9652 | 1 | **9.37** | 9.37 |
| TAPENTADOL | 9780 | 1 | **36.00** | 36.00 |
| FENTANYL BASE | 9801 | 423 | **32.59** | 0.08 |

**BUSINESS ACTIVITY: D - TEACHING INSTITUTIONS**

| DRUG NAME | DRUG CODE | NUMBER OF REGISTRANT SOLD TO | TOTAL GRAMS | AVG GRAMS |
|---|---|---|---|---|
| CODEINE | 9050 | 1 | **173.17** | 173.17 |
| BUPRENORPHINE | 9064 | 1 | **0.50** | 0.50 |
| HYDROMORPHONE | 9150 | 1 | **17.38** | 17.38 |
| HYDROCODONE | 9193 | 1 | **0.89** | 0.89 |
| METHADONE | 9250B | 1 | **3.04** | 3.04 |
| MORPHINE | 9300 | 1 | **13.91** | 13.91 |
| FENTANYL BASE | 9801 | 1 | **0.87** | 0.87 |

**BUSINESS ACTIVITY: M - MID-LEVEL PRACTITIONERS**

| DRUG NAME | DRUG CODE | NUMBER OF REGISTRANT SOLD TO | TOTAL GRAMS | AVG GRAMS |
|---|---|---|---|---|
| CODEINE | 9050 | 5 | **47.08** | 9.42 |
| BUPRENORPHINE | 9064 | 5 | **4.40** | 0.88 |
| MORPHINE | 9300 | 9 | **1.94** | 0.22 |
| FENTANYL BASE | 9801 | 16 | **0.21** | 0.01 |

**BUSINESS ACTIVITY: NTP - NARCOTIC TREATMENT PROGRAMS**

| DRUG NAME | DRUG CODE | NUMBER OF REGISTRANT SOLD TO | TOTAL GRAMS | AVG GRAMS |
|---|---|---|---|---|
| BUPRENORPHINE | 9064 | 31 | **4,621.12** | 149.07 |
| METHADONE | 9250B | 39 | **336,520.68** | 8,628.74 |

**STATE: VI - VIRGIN ISLANDS**

**BUSINESS ACTIVITY: A - PHARMACIES**

| DRUG NAME | DRUG CODE | NUMBER OF REGISTRANT SOLD TO | TOTAL GRAMS | AVG GRAMS |
|---|---|---|---|---|
| AMPHETAMINE | 1100 | 19 | **1,486.11** | 78.22 |
| LISDEXAMFETAMINE | 1205 | 11 | **306.39** | 27.85 |
| METHYLPHENIDATE (DL;D;L;ISOMERS) | 1724 | 16 | **390.37** | 24.40 |
| CODEINE | 9050 | 21 | **2,360.39** | 112.40 |
| BUPRENORPHINE | 9064 | 12 | **103.12** | 8.59 |
| OXYCODONE | 9143 | 21 | **1,534.81** | 73.09 |
| HYDROMORPHONE | 9150 | 11 | **50.36** | 4.58 |

| | | | | |
|---|---|---|---|---|
| HYDROCODONE | 9193 | 21 | **678.29** | 32.30 |
| MEPERIDINE (PETHIDINE) | 9230 | 2 | **5.45** | 2.72 |
| METHADONE | 9250B | 4 | **50.09** | 12.52 |
| MORPHINE | 9300 | 13 | **240.11** | 18.47 |
| TAPENTADOL | 9780 | 1 | **45.00** | 45.00 |
| FENTANYL BASE | 9801 | 12 | **13.38** | 1.12 |

**BUSINESS ACTIVITY: B - HOSPITALS**

| DRUG NAME | DRUG CODE | NUMBER OF REGISTRANT SOLD TO | TOTAL GRAMS | AVG GRAMS |
|---|---|---|---|---|
| CODEINE | 9050 | 2 | **50.83** | 25.42 |
| OXYCODONE | 9143 | 3 | **19.72** | 6.57 |
| HYDROMORPHONE | 9150 | 4 | **9.71** | 2.43 |
| HYDROCODONE | 9193 | 3 | **24.09** | 8.03 |
| MEPERIDINE (PETHIDINE) | 9230 | 4 | **41.94** | 10.49 |
| MORPHINE | 9300 | 4 | **32.07** | 8.02 |
| FENTANYL BASE | 9801 | 5 | **4.48** | 0.90 |

**BUSINESS ACTIVITY: C - PRACTITIONERS**

| DRUG NAME | DRUG CODE | NUMBER OF REGISTRANT SOLD TO | TOTAL GRAMS | AVG GRAMS |
|---|---|---|---|---|
| BUPRENORPHINE | 9064 | 4 | **0.21** | 0.05 |
| HYDROMORPHONE | 9150 | 2 | **2.93** | 1.46 |
| MEPERIDINE (PETHIDINE) | 9230 | 1 | **1.09** | 1.09 |
| MORPHINE | 9300 | 1 | **0.79** | 0.79 |
| FENTANYL BASE | 9801 | 1 | **0.03** | 0.03 |

**STATE: VT - VERMONT**

**BUSINESS ACTIVITY: A - PHARMACIES**

| DRUG NAME | DRUG CODE | NUMBER OF REGISTRANT SOLD TO | TOTAL GRAMS | AVG GRAMS |
|---|---|---|---|---|
| AMPHETAMINE | 1100 | 141 | **43,313.57** | 307.19 |
| METHAMPHETAMINE | 1105 | 4 | **7.63** | 1.91 |
| LISDEXAMFETAMINE | 1205 | 140 | **30,511.54** | 217.94 |
| METHYLPHENIDATE (DL;D;L;ISOMERS) | 1724 | 141 | **72,012.35** | 510.73 |
| CODEINE | 9050 | 134 | **8,693.76** | 64.88 |
| BUPRENORPHINE | 9064 | 138 | **28,989.52** | 210.07 |
| OXYCODONE | 9143 | 139 | **57,660.42** | 414.82 |
| HYDROMORPHONE | 9150 | 136 | **2,878.62** | 21.17 |
| HYDROCODONE | 9193 | 140 | **13,060.65** | 93.29 |
| MEPERIDINE (PETHIDINE) | 9230 | 6 | **120.05** | 20.01 |
| METHADONE | 9250B | 129 | **8,615.13** | 66.78 |
| MORPHINE | 9300 | 137 | **26,199.24** | 191.24 |
| OXYMORPHONE | 9652 | 18 | **324.72** | 18.04 |
| TAPENTADOL | 9780 | 35 | **2,887.50** | 82.50 |
| FENTANYL BASE | 9801 | 132 | **352.73** | 2.67 |

**BUSINESS ACTIVITY: B - HOSPITALS**

| DRUG NAME | DRUG CODE | NUMBER OF REGISTRANT SOLD TO | TOTAL GRAMS | AVG GRAMS |
|---|---|---|---|---|
| AMPHETAMINE | 1100 | 13 | **1,226.94** | 94.38 |
| LISDEXAMFETAMINE | 1205 | 6 | **445.14** | 74.19 |
| METHYLPHENIDATE (DL;D;L;ISOMERS) | 1724 | 13 | **2,338.80** | 179.91 |
| CODEINE | 9050 | 18 | **243.73** | 13.54 |
| BUPRENORPHINE | 9064 | 24 | **2,597.95** | 108.25 |
| OXYCODONE | 9143 | 17 | **4,349.68** | 255.86 |
| HYDROMORPHONE | 9150 | 16 | **599.04** | 37.44 |
| HYDROCODONE | 9193 | 15 | **726.73** | 48.45 |
| MEPERIDINE (PETHIDINE) | 9230 | 8 | **554.27** | 69.28 |
| METHADONE | 9250B | 16 | **561.24** | 35.08 |
| MORPHINE | 9300 | 17 | **2,898.45** | 170.50 |
| TAPENTADOL | 9780 | 2 | **65.00** | 32.50 |
| FENTANYL BASE | 9801 | 17 | **34.41** | 2.02 |

**BUSINESS ACTIVITY: C - PRACTITIONERS**

| DRUG NAME | DRUG CODE | NUMBER OF REGISTRANT SOLD TO | TOTAL GRAMS | AVG GRAMS |
|---|---|---|---|---|
| CODEINE | 9050 | 2 | **13.16** | 6.58 |
| BUPRENORPHINE | 9064 | 96 | **8.51** | 0.09 |
| OXYCODONE | 9143 | 2 | **1.26** | 0.63 |
| HYDROMORPHONE | 9150 | 52 | **299.02** | 5.75 |

| | | | | |
|---|---|---|---|---|
| HYDROCODONE | 9193 | 23 | **27.79** | 1.21 |
| MEPERIDINE (PETHIDINE) | 9230 | 1 | **1.09** | 1.09 |
| METHADONE | 9250B | 2 | **6.26** | 3.13 |
| MORPHINE | 9300 | 14 | **13.50** | 0.96 |
| FENTANYL BASE | 9801 | 31 | **1.29** | 0.04 |

**BUSINESS ACTIVITY: M - MID-LEVEL PRACTITIONERS**

| DRUG NAME | DRUG CODE | NUMBER OF REGISTRANT SOLD TO | TOTAL GRAMS | AVG GRAMS |
|---|---|---|---|---|
| CODEINE | 9050 | 2 | **6.63** | 3.32 |

**BUSINESS ACTIVITY: NTP - NARCOTIC TREATMENT PROGRAMS**

| DRUG NAME | DRUG CODE | NUMBER OF REGISTRANT SOLD TO | TOTAL GRAMS | AVG GRAMS |
|---|---|---|---|---|
| BUPRENORPHINE | 9064 | 8 | **2,846.95** | 355.87 |
| METHADONE | 9250B | 8 | **74,375.17** | 9,296.90 |

**STATE: WA - WASHINGTON**

**BUSINESS ACTIVITY: A - PHARMACIES**

| DRUG NAME | DRUG CODE | NUMBER OF REGISTRANT SOLD TO | TOTAL GRAMS | AVG GRAMS |
|---|---|---|---|---|
| AMPHETAMINE | 1100 | 1,182 | **491,852.33** | 416.12 |
| METHAMPHETAMINE | 1105 | 88 | **302.89** | 3.44 |
| LISDEXAMFETAMINE | 1205 | 1,115 | **147,821.90** | 132.58 |
| METHYLPHENIDATE (DL;D;L;ISOMERS) | 1724 | 1,184 | **419,688.73** | 354.47 |
| CODEINE | 9050 | 1,173 | **154,281.00** | 131.53 |
| BUPRENORPHINE | 9064 | 1,198 | **133,918.00** | 111.78 |
| OXYCODONE | 9143 | 1,196 | **749,096.59** | 626.33 |
| HYDROMORPHONE | 9150 | 1,136 | **23,947.36** | 21.08 |
| HYDROCODONE | 9193 | 1,205 | **342,830.84** | 284.51 |
| MEPERIDINE (PETHIDINE) | 9230 | 103 | **1,143.52** | 11.10 |
| METHADONE | 9250B | 1,072 | **57,001.27** | 53.17 |
| MORPHINE | 9300 | 1,179 | **263,637.35** | 223.61 |
| OXYMORPHONE | 9652 | 357 | **3,407.20** | 9.54 |
| TAPENTADOL | 9780 | 519 | **41,092.00** | 79.18 |
| FENTANYL BASE | 9801 | 1,043 | **2,736.46** | 2.62 |

**BUSINESS ACTIVITY: B - HOSPITALS**

| DRUG NAME | DRUG CODE | NUMBER OF REGISTRANT SOLD TO | TOTAL GRAMS | AVG GRAMS |
|---|---|---|---|---|
| AMPHETAMINE | 1100 | 98 | **15,250.36** | 155.62 |
| LISDEXAMFETAMINE | 1205 | 42 | **3,310.20** | 78.81 |
| METHYLPHENIDATE (DL;D;L;ISOMERS) | 1724 | 110 | **19,605.90** | 178.24 |
| CODEINE | 9050 | 130 | **4,363.46** | 33.57 |
| BUPRENORPHINE | 9064 | 154 | **5,262.31** | 34.17 |
| OXYCODONE | 9143 | 231 | **50,223.82** | 217.42 |
| HYDROMORPHONE | 9150 | 206 | **5,240.60** | 25.44 |
| HYDROCODONE | 9193 | 215 | **19,391.95** | 90.20 |
| MEPERIDINE (PETHIDINE) | 9230 | 112 | **784.46** | 7.00 |
| METHADONE | 9250B | 127 | **6,566.06** | 51.70 |
| MORPHINE | 9300 | 238 | **22,295.39** | 93.68 |
| OXYMORPHONE | 9652 | 7 | **56.02** | 8.00 |
| TAPENTADOL | 9780 | 18 | **2,050.00** | 113.89 |
| FENTANYL BASE | 9801 | 229 | **537.69** | 2.35 |

**BUSINESS ACTIVITY: C - PRACTITIONERS**

| DRUG NAME | DRUG CODE | NUMBER OF REGISTRANT SOLD TO | TOTAL GRAMS | AVG GRAMS |
|---|---|---|---|---|
| CODEINE | 9050 | 119 | **6,682.44** | 56.15 |
| BUPRENORPHINE | 9064 | 889 | **752.39** | 0.85 |
| OXYCODONE | 9143 | 33 | **58.42** | 1.77 |
| HYDROMORPHONE | 9150 | 481 | **3,411.60** | 7.09 |
| HYDROCODONE | 9193 | 337 | **1,003.13** | 2.98 |
| MEPERIDINE (PETHIDINE) | 9230 | 75 | **372.68** | 4.97 |
| METHADONE | 9250B | 46 | **102.68** | 2.23 |
| MORPHINE | 9300 | 217 | **358.47** | 1.65 |
| FENTANYL BASE | 9801 | 612 | **34.67** | 0.06 |

**BUSINESS ACTIVITY: D - TEACHING INSTITUTIONS**

| DRUG NAME | DRUG CODE | NUMBER OF REGISTRANT SOLD TO | TOTAL GRAMS | AVG GRAMS |
|---|---|---|---|---|
| BUPRENORPHINE | 9064 | 1 | **0.00** | 0.00 |

**BUSINESS ACTIVITY: M - MID-LEVEL PRACTITIONERS**

| DRUG NAME | DRUG CODE | NUMBER OF REGISTRANT SOLD TO | TOTAL GRAMS | AVG GRAMS |
|---|---|---|---|---|
| CODEINE | 9050 | 1 | **0.66** | 0.66 |
| BUPRENORPHINE | 9064 | 17 | **421.26** | 24.78 |
| OXYCODONE | 9143 | 2 | **2.24** | 1.12 |
| HYDROMORPHONE | 9150 | 4 | **0.25** | 0.06 |
| HYDROCODONE | 9193 | 1 | **0.18** | 0.18 |
| MEPERIDINE (PETHIDINE) | 9230 | 2 | **9.48** | 4.74 |
| METHADONE | 9250B | 1 | **0.89** | 0.89 |
| MORPHINE | 9300 | 2 | **2.44** | 1.22 |
| FENTANYL BASE | 9801 | 8 | **0.13** | 0.02 |

**BUSINESS ACTIVITY: NTP - NARCOTIC TREATMENT PROGRAMS**

| DRUG NAME | DRUG CODE | NUMBER OF REGISTRANT SOLD TO | TOTAL GRAMS | AVG GRAMS |
|---|---|---|---|---|
| BUPRENORPHINE | 9064 | 21 | **1,963.03** | 93.48 |
| METHADONE | 9250B | 30 | **353,590.31** | 11,786.34 |

**STATE: WI - WISCONSIN**

**BUSINESS ACTIVITY: A - PHARMACIES**

| DRUG NAME | DRUG CODE | NUMBER OF REGISTRANT SOLD TO | TOTAL GRAMS | AVG GRAMS |
|---|---|---|---|---|
| AMPHETAMINE | 1100 | 1,048 | **509,682.87** | 486.34 |
| METHAMPHETAMINE | 1105 | 21 | **82.77** | 3.94 |
| LISDEXAMFETAMINE | 1205 | 1,034 | **311,254.82** | 301.02 |
| METHYLPHENIDATE (DL;D;L;ISOMERS) | 1724 | 1,042 | **395,314.85** | 379.38 |
| CODEINE | 9050 | 1,026 | **130,194.85** | 126.90 |
| BUPRENORPHINE | 9064 | 1,006 | **60,236.15** | 59.88 |
| OXYCODONE | 9143 | 1,051 | **438,406.34** | 417.13 |
| HYDROMORPHONE | 9150 | 836 | **7,588.61** | 9.08 |
| HYDROCODONE | 9193 | 1,054 | **208,456.12** | 197.78 |
| MEPERIDINE (PETHIDINE) | 9230 | 23 | **133.99** | 5.83 |
| METHADONE | 9250B | 747 | **16,348.92** | 21.89 |
| MORPHINE | 9300 | 1,034 | **181,921.53** | 175.94 |
| OXYMORPHONE | 9652 | 187 | **1,644.99** | 8.80 |
| TAPENTADOL | 9780 | 475 | **29,997.00** | 63.15 |
| FENTANYL BASE | 9801 | 932 | **2,497.69** | 2.68 |

**BUSINESS ACTIVITY: B - HOSPITALS**

| DRUG NAME | DRUG CODE | NUMBER OF REGISTRANT SOLD TO | TOTAL GRAMS | AVG GRAMS |
|---|---|---|---|---|
| AMPHETAMINE | 1100 | 97 | **9,564.81** | 98.61 |
| LISDEXAMFETAMINE | 1205 | 71 | **2,500.74** | 35.22 |
| METHYLPHENIDATE (DL;D;L;ISOMERS) | 1724 | 118 | **6,466.17** | 54.80 |
| CODEINE | 9050 | 164 | **2,127.33** | 12.97 |
| BUPRENORPHINE | 9064 | 115 | **2,854.70** | 24.82 |
| OXYCODONE | 9143 | 169 | **23,195.53** | 137.25 |
| HYDROMORPHONE | 9150 | 153 | **3,314.04** | 21.66 |
| HYDROCODONE | 9193 | 168 | **9,566.59** | 56.94 |
| MEPERIDINE (PETHIDINE) | 9230 | 84 | **937.19** | 11.16 |
| METHADONE | 9250B | 127 | **1,943.19** | 15.30 |
| MORPHINE | 9300 | 164 | **16,129.76** | 98.35 |
| OXYMORPHONE | 9652 | 2 | **1.43** | 0.71 |
| TAPENTADOL | 9780 | 16 | **188.50** | 11.78 |
| FENTANYL BASE | 9801 | 155 | **484.39** | 3.13 |

**BUSINESS ACTIVITY: C - PRACTITIONERS**

| DRUG NAME | DRUG CODE | NUMBER OF REGISTRANT SOLD TO | TOTAL GRAMS | AVG GRAMS |
|---|---|---|---|---|
| CODEINE | 9050 | 43 | **485.04** | 11.28 |
| BUPRENORPHINE | 9064 | 572 | **284.52** | 0.50 |
| OXYCODONE | 9143 | 49 | **318.50** | 6.50 |
| HYDROMORPHONE | 9150 | 372 | **2,571.54** | 6.91 |
| HYDROCODONE | 9193 | 222 | **509.64** | 2.30 |
| MEPERIDINE (PETHIDINE) | 9230 | 47 | **162.86** | 3.47 |
| METHADONE | 9250B | 16 | **26.12** | 1.63 |
| MORPHINE | 9300 | 128 | **132.80** | 1.04 |
| FENTANYL BASE | 9801 | 299 | **30.52** | 0.10 |

**BUSINESS ACTIVITY: M - MID-LEVEL PRACTITIONERS**

| DRUG NAME | DRUG CODE | NUMBER OF REGISTRANT SOLD TO | TOTAL GRAMS | AVG GRAMS |
|---|---|---|---|---|

| | | | | |
|---|---|---|---|---|
| CODEINE | 9050 | 3 | **127.30** | 42.43 |
| BUPRENORPHINE | 9064 | 3 | **0.72** | 0.24 |
| MORPHINE | 9300 | 1 | **0.02** | 0.02 |
| FENTANYL BASE | 9801 | 3 | **0.01** | 0.00 |

**BUSINESS ACTIVITY: NTP - NARCOTIC TREATMENT PROGRAMS**

| DRUG NAME | DRUG CODE | NUMBER OF REGISTRANT SOLD TO | TOTAL GRAMS | AVG GRAMS |
|---|---|---|---|---|
| BUPRENORPHINE | 9064 | 19 | **2,082.90** | 109.63 |
| METHADONE | 9250B | 22 | **240,244.78** | 10,920.22 |

**STATE: WV - WEST VIRGINIA**

**BUSINESS ACTIVITY: A - PHARMACIES**

| DRUG NAME | DRUG CODE | NUMBER OF REGISTRANT SOLD TO | TOTAL GRAMS | AVG GRAMS |
|---|---|---|---|---|
| AMPHETAMINE | 1100 | 521 | **92,648.17** | 177.83 |
| METHAMPHETAMINE | 1105 | 4 | **11.65** | 2.91 |
| LISDEXAMFETAMINE | 1205 | 512 | **70,058.78** | 136.83 |
| METHYLPHENIDATE (DL;D;L;ISOMERS) | 1724 | 521 | **105,912.22** | 203.29 |
| CODEINE | 9050 | 512 | **134,365.51** | 262.43 |
| BUPRENORPHINE | 9064 | 477 | **86,643.62** | 181.64 |
| OXYCODONE | 9143 | 523 | **181,134.45** | 346.34 |
| HYDROMORPHONE | 9150 | 335 | **2,734.23** | 8.16 |
| HYDROCODONE | 9193 | 525 | **125,503.10** | 239.05 |
| MEPERIDINE (PETHIDINE) | 9230 | 10 | **61.44** | 6.14 |
| METHADONE | 9250B | 303 | **6,497.46** | 21.44 |
| MORPHINE | 9300 | 494 | **49,236.49** | 99.67 |
| OXYMORPHONE | 9652 | 99 | **1,708.33** | 17.26 |
| TAPENTADOL | 9780 | 152 | **13,324.50** | 87.66 |
| FENTANYL BASE | 9801 | 459 | **928.21** | 2.02 |

**BUSINESS ACTIVITY: B - HOSPITALS**

| DRUG NAME | DRUG CODE | NUMBER OF REGISTRANT SOLD TO | TOTAL GRAMS | AVG GRAMS |
|---|---|---|---|---|
| AMPHETAMINE | 1100 | 30 | **2,340.68** | 78.02 |
| LISDEXAMFETAMINE | 1205 | 17 | **748.06** | 44.00 |
| METHYLPHENIDATE (DL;D;L;ISOMERS) | 1724 | 48 | **2,215.34** | 46.15 |
| CODEINE | 9050 | 62 | **1,519.42** | 24.51 |
| BUPRENORPHINE | 9064 | 65 | **3,221.52** | 49.56 |
| OXYCODONE | 9143 | 78 | **11,310.86** | 145.01 |
| HYDROMORPHONE | 9150 | 66 | **699.84** | 10.60 |
| HYDROCODONE | 9193 | 78 | **10,980.00** | 140.77 |
| MEPERIDINE (PETHIDINE) | 9230 | 37 | **269.62** | 7.29 |
| METHADONE | 9250B | 47 | **691.82** | 14.72 |
| MORPHINE | 9300 | 76 | **6,630.60** | 87.24 |
| OXYMORPHONE | 9652 | 4 | **5.75** | 1.44 |
| TAPENTADOL | 9780 | 6 | **57.50** | 9.58 |
| FENTANYL BASE | 9801 | 77 | **204.30** | 2.65 |

**BUSINESS ACTIVITY: C - PRACTITIONERS**

| DRUG NAME | DRUG CODE | NUMBER OF REGISTRANT SOLD TO | TOTAL GRAMS | AVG GRAMS |
|---|---|---|---|---|
| CODEINE | 9050 | 2 | **47.08** | 23.54 |
| BUPRENORPHINE | 9064 | 106 | **195.24** | 1.84 |
| OXYCODONE | 9143 | 1 | **0.45** | 0.45 |
| HYDROMORPHONE | 9150 | 37 | **461.57** | 12.47 |
| HYDROCODONE | 9193 | 33 | **127.31** | 3.86 |
| MEPERIDINE (PETHIDINE) | 9230 | 4 | **84.43** | 21.11 |
| MORPHINE | 9300 | 23 | **9.62** | 0.42 |
| FENTANYL BASE | 9801 | 49 | **4.04** | 0.08 |

**BUSINESS ACTIVITY: M - MID-LEVEL PRACTITIONERS**

| DRUG NAME | DRUG CODE | NUMBER OF REGISTRANT SOLD TO | TOTAL GRAMS | AVG GRAMS |
|---|---|---|---|---|
| MORPHINE | 9300 | 43 | **14.15** | 0.33 |
| FENTANYL BASE | 9801 | 42 | **0.56** | 0.01 |

**BUSINESS ACTIVITY: NTP - NARCOTIC TREATMENT PROGRAMS**

| DRUG NAME | DRUG CODE | NUMBER OF REGISTRANT SOLD TO | TOTAL GRAMS | AVG GRAMS |
|---|---|---|---|---|
| BUPRENORPHINE | 9064 | 7 | **489.18** | 69.88 |
| METHADONE | 9250B | 9 | **92,910.27** | 10,323.36 |

**STATE: WY - WYOMING**

**BUSINESS ACTIVITY: A - PHARMACIES**

| DRUG NAME | DRUG CODE | NUMBER OF REGISTRANT SOLD TO | TOTAL GRAMS | AVG GRAMS |
|---|---|---|---|---|
| AMPHETAMINE | 1100 | 111 | **29,706.36** | 267.62 |
| METHAMPHETAMINE | 1105 | 7 | **19.68** | 2.81 |
| LISDEXAMFETAMINE | 1205 | 109 | **10,811.63** | 99.19 |
| METHYLPHENIDATE (DL;D;L;ISOMERS) | 1724 | 111 | **27,648.00** | 249.08 |
| CODEINE | 9050 | 111 | **10,662.27** | 96.06 |
| BUPRENORPHINE | 9064 | 112 | **5,936.62** | 53.01 |
| OXYCODONE | 9143 | 113 | **69,264.84** | 612.96 |
| HYDROMORPHONE | 9150 | 103 | **1,702.95** | 16.53 |
| HYDROCODONE | 9193 | 113 | **28,564.00** | 252.78 |
| MEPERIDINE (PETHIDINE) | 9230 | 8 | **15.25** | 1.91 |
| METHADONE | 9250B | 91 | **2,365.69** | 26.00 |
| MORPHINE | 9300 | 112 | **20,029.15** | 178.83 |
| OXYMORPHONE | 9652 | 24 | **246.89** | 10.29 |
| TAPENTADOL | 9780 | 55 | **3,809.00** | 69.25 |
| FENTANYL BASE | 9801 | 106 | **301.61** | 2.85 |

**BUSINESS ACTIVITY: B - HOSPITALS**

| DRUG NAME | DRUG CODE | NUMBER OF REGISTRANT SOLD TO | TOTAL GRAMS | AVG GRAMS |
|---|---|---|---|---|
| AMPHETAMINE | 1100 | 18 | **1,576.98** | 87.61 |
| LISDEXAMFETAMINE | 1205 | 10 | **445.72** | 44.57 |
| METHYLPHENIDATE (DL;D;L;ISOMERS) | 1724 | 22 | **1,787.45** | 81.25 |
| CODEINE | 9050 | 27 | **639.80** | 23.70 |
| BUPRENORPHINE | 9064 | 20 | **439.77** | 21.99 |
| OXYCODONE | 9143 | 44 | **4,940.46** | 112.28 |
| HYDROMORPHONE | 9150 | 39 | **138.93** | 3.56 |
| HYDROCODONE | 9193 | 45 | **3,343.35** | 74.30 |
| MEPERIDINE (PETHIDINE) | 9230 | 20 | **73.42** | 3.67 |
| METHADONE | 9250B | 18 | **207.77** | 11.54 |
| MORPHINE | 9300 | 44 | **4,190.46** | 95.24 |
| TAPENTADOL | 9780 | 4 | **37.50** | 9.38 |
| FENTANYL BASE | 9801 | 44 | **46.62** | 1.06 |

**BUSINESS ACTIVITY: C - PRACTITIONERS**

| DRUG NAME | DRUG CODE | NUMBER OF REGISTRANT SOLD TO | TOTAL GRAMS | AVG GRAMS |
|---|---|---|---|---|
| CODEINE | 9050 | 5 | **70.28** | 14.06 |
| BUPRENORPHINE | 9064 | 65 | **5.65** | 0.09 |
| OXYCODONE | 9143 | 1 | **17.75** | 17.75 |
| HYDROMORPHONE | 9150 | 33 | **308.53** | 9.35 |
| HYDROCODONE | 9193 | 27 | **80.19** | 2.97 |
| MEPERIDINE (PETHIDINE) | 9230 | 1 | **9.59** | 9.59 |
| MORPHINE | 9300 | 32 | **38.37** | 1.20 |
| FENTANYL BASE | 9801 | 41 | **3.72** | 0.09 |

**BUSINESS ACTIVITY: M - MID-LEVEL PRACTITIONERS**

| DRUG NAME | DRUG CODE | NUMBER OF REGISTRANT SOLD TO | TOTAL GRAMS | AVG GRAMS |
|---|---|---|---|---|
| CODEINE | 9050 | 2 | **1.33** | 0.66 |
| BUPRENORPHINE | 9064 | 1 | **0.36** | 0.36 |

ENTIRE UNITED STATES — BUSINESS ACTIVITY: A - PHARMACIES

| DRUG NAME | DRUG CODE | NUMBER OF REGISTRANT SOLD TO | TOTAL GRAMS | AVG GRAMS |
|---|---|---|---|---|
| AMPHETAMINE | 1100 | 61,209 | **23,374,103.91** | 381.87 |
| METHAMPHETAMINE | 1105 | 1,571 | **5,495.58** | 3.50 |
| LISDEXAMFETAMINE | 1205 | 56,582 | **9,417,369.24** | 166.44 |
| METHYLPHENIDATE (DL;D;L;ISOMERS) | 1724 | 60,272 | **16,330,120.95** | 270.94 |
| CODEINE | 9050 | 62,026 | **10,697,255.74** | 172.46 |
| BUPRENORPHINE | 9064 | 57,930 | **4,658,456.87** | 80.42 |
| OXYCODONE | 9143 | 62,555 | **31,642,429.10** | 505.83 |
| HYDROMORPHONE | 9150 | 51,941 | **774,149.89** | 14.90 |
| HYDROCODONE | 9193 | 62,314 | **17,689,153.07** | 283.87 |
| MEPERIDINE (PETHIDINE) | 9230 | 4,327 | **49,065.62** | 11.34 |
| METHADONE | 9250B | 44,735 | **1,503,154.87** | 33.60 |
| MORPHINE | 9300 | 57,804 | **9,873,090.20** | 170.80 |
| OXYMORPHONE | 9652 | 14,987 | **264,469.07** | 17.65 |
| TAPENTADOL | 9780 | 29,880 | **3,129,815.50** | 104.75 |
| FENTANYL BASE | 9801 | 53,146 | **139,581.06** | 2.63 |

ENTIRE UNITED STATES — BUSINESS ACTIVITY: B - HOSPITALS

| DRUG NAME | DRUG CODE | NUMBER OF REGISTRANT SOLD TO | TOTAL GRAMS | AVG GRAMS |
|---|---|---|---|---|
| AMPHETAMINE | 1100 | 4,027 | **525,560.79** | 130.51 |
| METHAMPHETAMINE | 1105 | 16 | **74.33** | 4.65 |
| LISDEXAMFETAMINE | 1205 | 1,974 | **121,509.16** | 61.55 |
| METHYLPHENIDATE (DL;D;L;ISOMERS) | 1724 | 5,233 | **483,848.73** | 92.46 |
| CODEINE | 9050 | 6,477 | **268,718.63** | 41.49 |
| BUPRENORPHINE | 9064 | 5,696 | **174,622.69** | 30.66 |
| OXYCODONE | 9143 | 8,846 | **1,506,993.09** | 170.36 |
| HYDROMORPHONE | 9150 | 8,666 | **199,667.96** | 23.04 |
| HYDROCODONE | 9193 | 9,034 | **784,254.92** | 86.81 |
| MEPERIDINE (PETHIDINE) | 9230 | 5,180 | **80,280.52** | 15.50 |
| METHADONE | 9250B | 5,691 | **195,052.16** | 34.27 |
| MORPHINE | 9300 | 9,554 | **858,806.70** | 89.89 |
| OXYMORPHONE | 9652 | 417 | **2,875.93** | 6.90 |
| TAPENTADOL | 9780 | 1,131 | **61,539.65** | 54.41 |
| FENTANYL BASE | 9801 | 10,058 | **31,533.19** | 3.14 |

ENTIRE UNITED STATES — BUSINESS ACTIVITY: C - PRACTITIONERS

| DRUG NAME | DRUG CODE | NUMBER OF REGISTRANT SOLD TO | TOTAL GRAMS | AVG GRAMS |
|---|---|---|---|---|
| AMPHETAMINE | 1100 | 25 | **1,262.69** | 50.51 |
| LISDEXAMFETAMINE | 1205 | 4 | **429.53** | 107.38 |
| METHYLPHENIDATE (DL;D;L;ISOMERS) | 1724 | 10 | **659.97** | 66.00 |
| CODEINE | 9050 | 2,021 | **81,121.24** | 40.14 |
| BUPRENORPHINE | 9064 | 27,925 | **16,657.50** | 0.60 |
| OXYCODONE | 9143 | 1,528 | **9,571.07** | 6.26 |
| HYDROMORPHONE | 9150 | 12,460 | **78,634.31** | 6.31 |
| HYDROCODONE | 9193 | 10,047 | **37,610.96** | 3.74 |
| MEPERIDINE (PETHIDINE) | 9230 | 2,885 | **21,975.07** | 7.62 |
| METHADONE | 9250B | 776 | **1,956.14** | 2.52 |
| MORPHINE | 9300 | 5,991 | **8,040.54** | 1.34 |
| OXYMORPHONE | 9652 | 5 | **49.91** | 9.98 |
| TAPENTADOL | 9780 | 8 | **208.00** | 26.00 |
| FENTANYL BASE | 9801 | 17,593 | **1,730.93** | 0.10 |

ENTIRE UNITED STATES — BUSINESS ACTIVITY: D - TEACHING INSTITUTIONS

| DRUG NAME | DRUG CODE | NUMBER OF REGISTRANT SOLD TO | TOTAL GRAMS | AVG GRAMS |
|---|---|---|---|---|
| AMPHETAMINE | 1100 | 1 | **3.67** | 3.67 |
| CODEINE | 9050 | 6 | **969.07** | 161.51 |
| BUPRENORPHINE | 9064 | 47 | **4.78** | 0.10 |
| OXYCODONE | 9143 | 2 | **7.17** | 3.59 |
| HYDROMORPHONE | 9150 | 21 | **246.16** | 11.72 |
| HYDROCODONE | 9193 | 8 | **138.23** | 17.28 |
| MEPERIDINE (PETHIDINE) | 9230 | 1 | **4.36** | 4.36 |
| METHADONE | 9250B | 6 | **110.55** | 18.42 |
| MORPHINE | 9300 | 12 | **190.36** | 15.86 |
| FENTANYL BASE | 9801 | 21 | **13.21** | 0.63 |

ENTIRE UNITED STATES — BUSINESS ACTIVITY: M - MID-LEVEL PRACTITIONERS

| DRUG NAME | DRUG CODE | NUMBER OF REGISTRANT SOLD TO | TOTAL GRAMS | AVG GRAMS |
|---|---|---|---|---|
| AMPHETAMINE | 1100 | 2 | **9.38** | 4.69 |

| | | | | |
|---|---|---|---|---|
| LISDEXAMFETAMINE | 1205 | 1 | **1.73** | 1.73 |
| CODEINE | 9050 | 119 | **1,898.67** | 15.96 |
| BUPRENORPHINE | 9064 | 131 | **2,369.84** | 18.09 |
| OXYCODONE | 9143 | 5 | **4.26** | 0.85 |
| HYDROMORPHONE | 9150 | 86 | **55.96** | 0.65 |
| HYDROCODONE | 9193 | 13 | **14.67** | 1.13 |
| MEPERIDINE (PETHIDINE) | 9230 | 12 | **51.75** | 4.31 |
| METHADONE | 9250B | 1 | **0.89** | 0.89 |
| MORPHINE | 9300 | 684 | **322.37** | 0.47 |
| FENTANYL BASE | 9801 | 1,088 | **29.08** | 0.03 |

ENTIRE UNITED STATES

BUSINESS ACTIVITY: NTP - NARCOTIC TREATMENT PROGRAMS

| DRUG NAME | DRUG CODE | NUMBER OF REGISTRANT SOLD TO | **TOTAL GRAMS** | AVG GRAMS |
|---|---|---|---|---|
| BUPRENORPHINE | 9064 | 1,034 | **110,452.34** | 106.82 |
| METHADONE | 9250B | 1,726 | **13,116,235.31** | 7,599.21 |
